# Mapping pQTLs of circulating inflammatory proteins identifies drivers of immune-related disease risk and novel therapeutic targets

**DOI:** 10.1101/2023.03.24.23287680

**Authors:** The SCALLOP consortium, Jing Hua Zhao, David Stacey, Niclas Eriksson, Erin Macdonald-Dunlop, Åsa K Hedman, Anette Kalnapenkis, Stefan Enroth, Domenico Cozzetto, Jonathan Digby-Bell, Jonathan Marten, Lasse Folkersen, Christian Herder, Lina Jonsson, Sarah E Bergen, Christian Geiger, Elise J Needham, Praveen Surendran, Estonian Biobank Research Team, Dirk S Paul, Ozren Polasek, Barbara Thorand, Harald Grallert, Michael Roden, Urmo Võsa, Tonu Esko, Caroline Hayward, Åsa Johansson, Ulf Gyllensten, Nicholas Powell, Oskar Hansson, Niklas Mattsson-Carlgren, Peter K Joshi, John Danesh, Leonid Padyukov, Lars Klareskog, Mikael Landén, James F Wilson, Agneta Siegbahn, Lars Wallentin, Anders Mälarstig, Adam S Butterworth, James E Peters

## Abstract

Circulating proteins play key roles in inflammation and a broad range of diseases. To identify genetic influences on inflammation-related proteins, we conducted a genome-wide protein quantitative trait locus (pQTL) study of 91 plasma proteins measured using the Olink Target platform in 15,150 participants. We identified 180 pQTLs, of which 50 were novel. Integration of pQTL data with eQTL and disease GWAS provided insights into pathogenesis, implicating lymphotoxin-alpha (LTA) in multiple sclerosis. Using Mendelian randomisation (MR), we identified both shared and distinct effects of specific proteins across immune-mediated diseases, including directionally discordant causal roles for CD40 in rheumatoid arthritis, multiple sclerosis and inflammatory bowel disease. Our results highlight novel potential therapeutic avenues, including CXCL5 in ulcerative colitis (UC), a finding supported by elevated gut *CXCL5* expression in UC patients. Our data provide a powerful resource to facilitate future drug target prioritization.

## INTRODUCTION

Inflammation is a key physiological host response to infection or injury. However, aberrant inflammatory responses result in tissue damage and are central to the pathophysiology of multiple diseases including sepsis, autoimmunity and atherothrombosis. Inflammatory responses are orchestrated by a complex network of cells and mediators, including circulating proteins such as cytokines and soluble receptors. Therefore, discovery of the genetic influences on abundance of inflammation-related circulating proteins should yield valuable insights into both physiology and the aetiology of a broad range of diseases.

Proteomic studies are particularly informative as proteins are the effector molecules of most biological processes, and from a translational biomedical perspective, proteins are the targets of most drugs. The development of high-throughput proteomic technologies now allows for profiling of the plasma proteome at epidemiological scale. Coupling genomic and proteomic data enables identification of genetic variants associated with protein abundance, protein quantitative trait loci (pQTLs). pQTLs provide valuable insights into the molecular basis of complex traits and diseases, by identifying proteins that lie between genotype and phenotype. Recent years have seen a rapid increase in both the number and size of pQTL studies, transforming our understanding of the genetic architecture of the circulating proteome. Most studies have used either the antibody-based Olink platform or the aptamer-based SomaScan platform^1–6^.

Here we extend this body of work by performing pQTL mapping for 91 inflammation-related proteins in 15,150 participants. We use these data to identify proteins that are the molecular intermediaries that link the genome and disease risk. Using Mendelian randomization and colocalisation analyses, we identify proteins that play a causal role in disease etiology. Our results reveal both pathways that are known to be therapeutically important, and new putative drug targets, including lymphotoxin-alpha (LTA) in multiple sclerosis and the chemokine CXCL5 in ulcerative colitis.

## RESULTS

### Genetic architecture of circulating inflammatory proteins

We performed pQTL mapping for 91 proteins measured using the Olink Target Inflammation panel in 11 cohorts totaling 15,150 European-ancestry participants (**Supplementary Table 1**), and meta-analysed the results (**Supplementary Figure 1**). In order to provide a succinct and standardised nomenclature, we report proteins by the non-italicised symbols of the genes encoding them (see **Supplementary Table 2** for a mapping of symbols to full protein names and UniProt identifiers). We identified a total of 180 significant (*P*<5×10^−10^) associations between 108 genomic regions and 70 proteins (**Figure 1, Supplementary Table 3, Supplementary Item**). To date, 50 of these associations have not previously been reported in peer-reviewed articles (r^2^≥0.8)^1,5–12^ (**Table 1**). Of the 180 significant associations, 59 (33%) were local-acting (‘*cis*’ pQTLs; defined here as a genetic variant lying within +/- 1 megabase of the gene encoding the associated protein) and 121 (67%) were distant-acting (‘*trans*’). We found evidence of *trans*-pQTL hotspots associated with multiple proteins (e.g. rs3184504 at the *SH2B3* locus was associated with six proteins: CXCL9, CXCL10, CXCL11, CD5, CD244, and IL12B) (**Figure 2a**).

**Figure 1.**
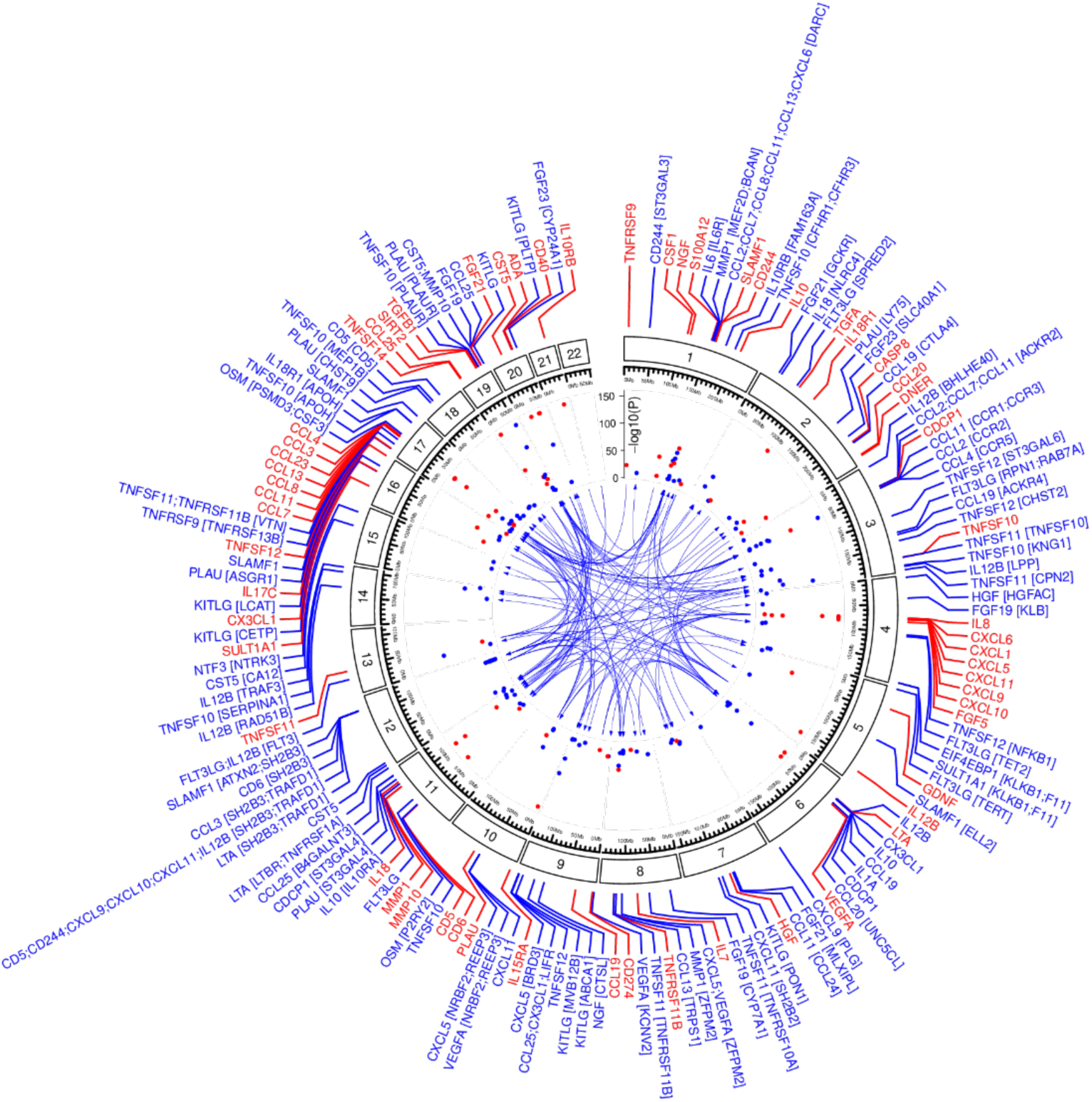
Circos plot showing the location of *cis*- (red) and *trans*- (blue) pQTLs and their associated proteins. Labels for the *cis*-pQTLs (red) indicate the gene encoding the target protein. For the *trans*-pQTLs (blue), the gene symbols of the target proteins are indicated along with the likely mediating gene(s) at the *trans*-pQTLs in square brackets. -log_10_(P-values) are capped at 150 for readability.

**Table 1.**
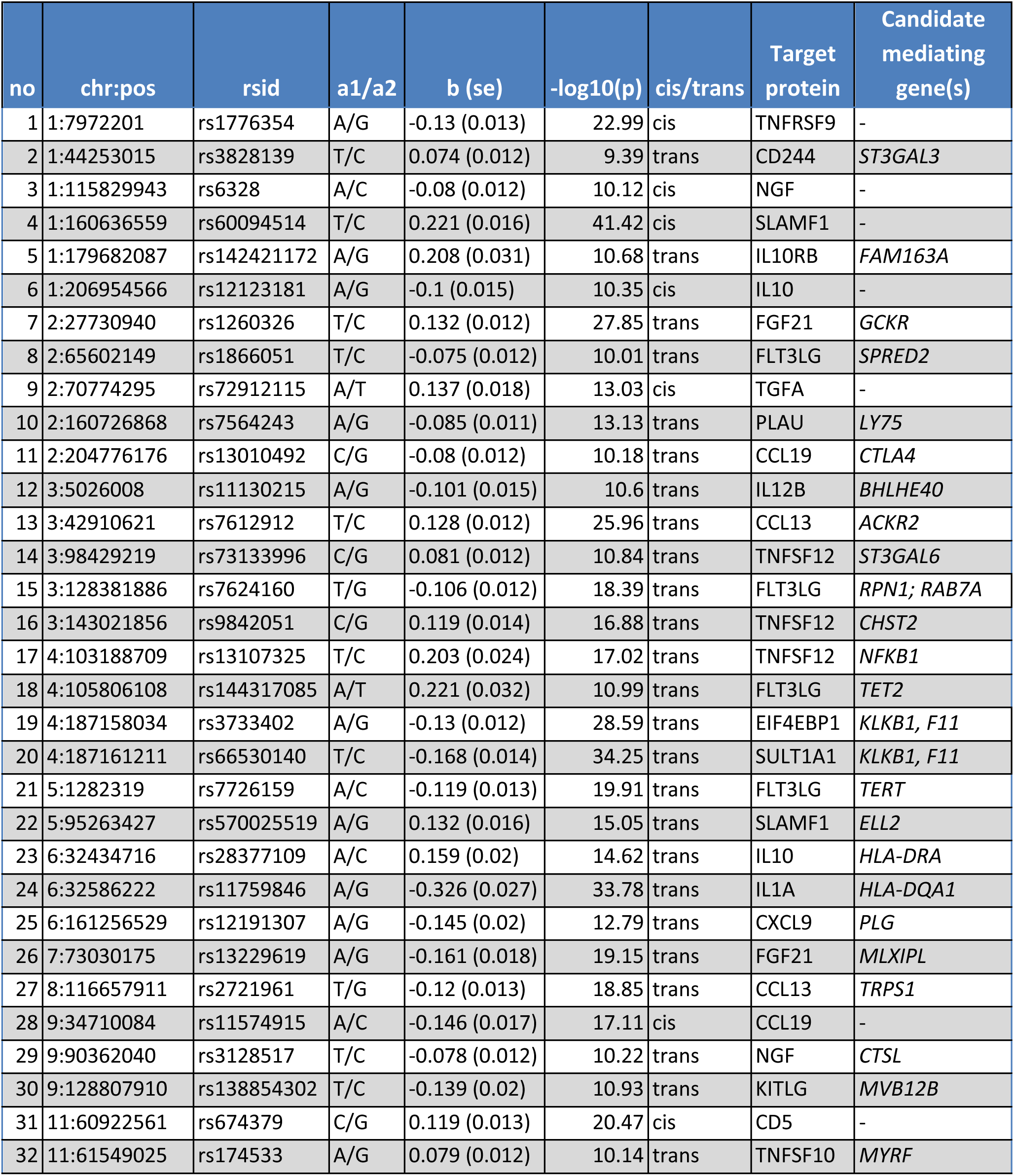
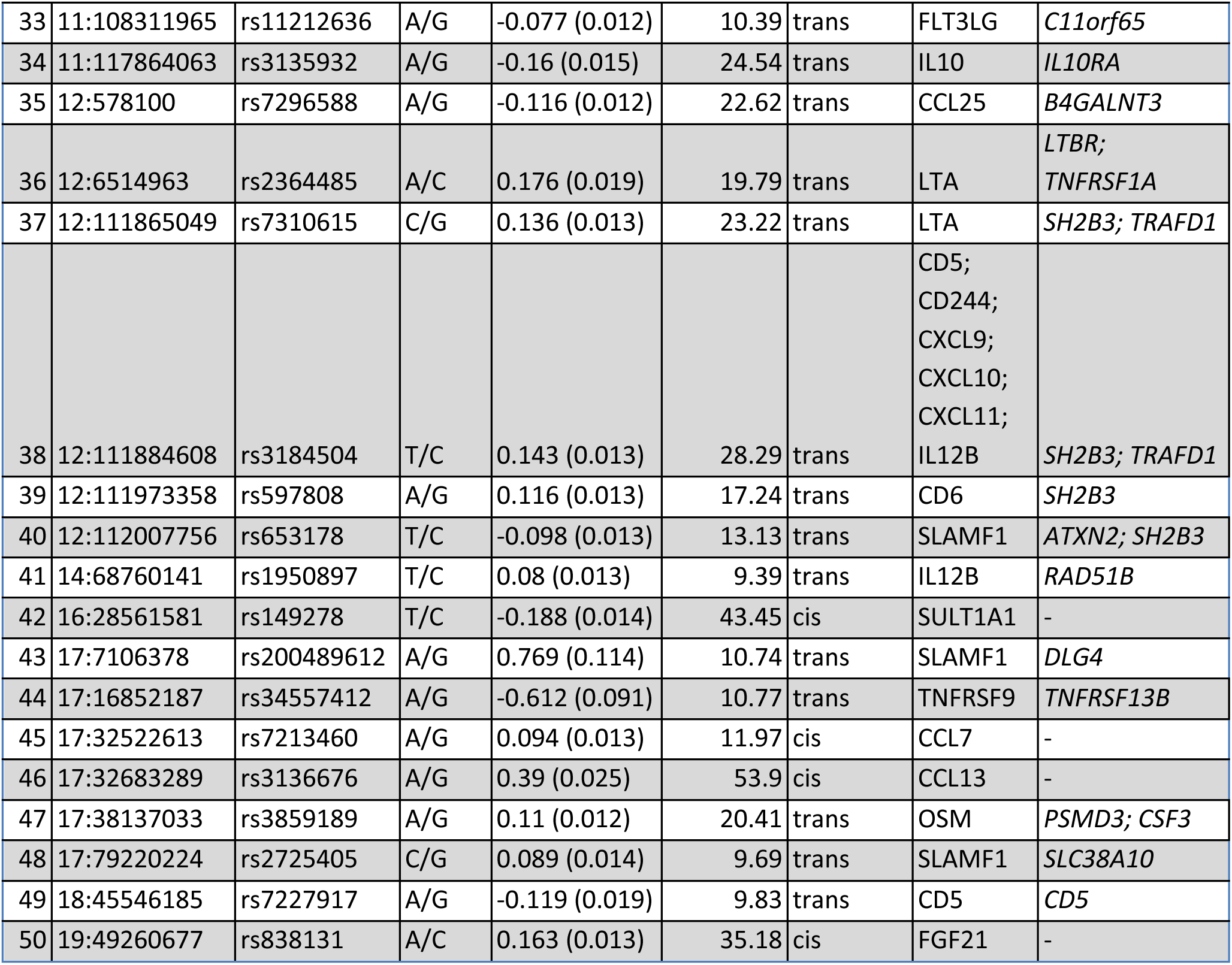
Unreported pQTLs. pQTLs not previously reported to our knowledge. pQTLs are ordered by chromosome and position. Target protein = protein whose abundance is associated with the genetic variant. Target proteins are annotated using the gene symbol of their encoding gene in order to provide a standardised nomenclature. Candidate mediating gene = the leading candidate mediating gene(s) according to ProGeM^23^ for the trans-pQTLs. A1/A2=effect allele/reference allele, b(se)=regression coefficient (standard error).

**Figure 2.**
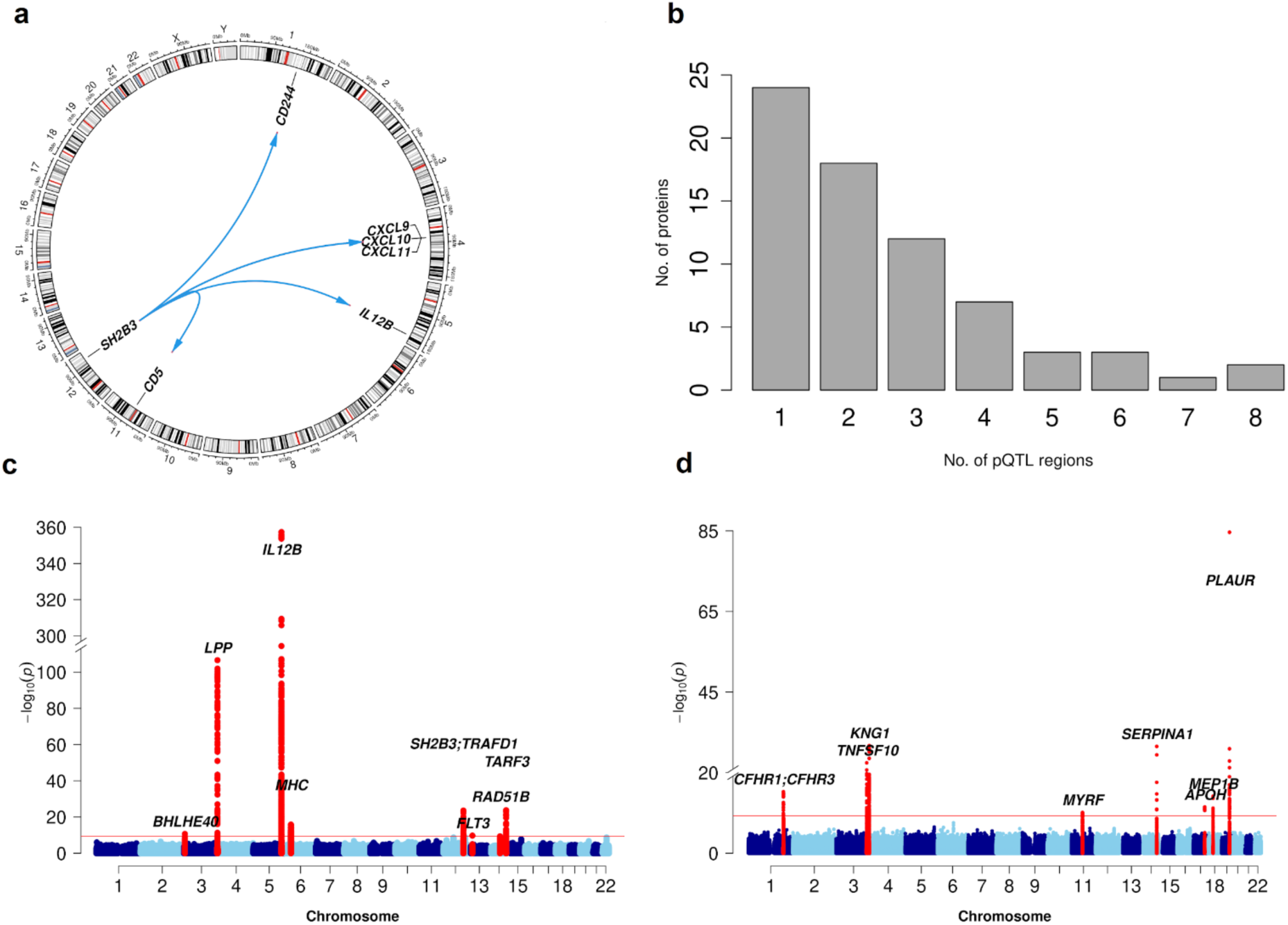
Genetic architecture of 91 inflammation-related proteins. (**a**) Distribution of the number of identified pQTLs per protein. The HLA region was treated as a single region. (**b**) Circos plot showing the six *trans*-pQTLs for the *SH2B3* ‘hotspot’ locus on chromosome 12. (**c**) Manhattan plots showing genetic associations with plasma abundance of IL12B and (**d**) TNFSF10 (TRAIL). The horizontal red line indicates statistical significance (P=5×10^-10^). Nearest genes in the region of pQTL signals are annotated.

For 70 (77%) of the 91 proteins studied, we identified at least 1 significant pQTL, including 59 (65%) proteins that had a *cis*-pQTL. Of these 70 proteins, 19 had only *cis-*pQTL(s), 11 had only *trans*-pQTL(s), and 40 had both *cis*- and *trans*-pQTLs. For 18 of the 21 proteins for which no pQTL was detected, >50% of samples had levels below the lower limit of detection (LLOD) in the INTERVAL study (where we had access to individual-level data), suggesting that the lack of genetic signal is due to low protein abundance in plasma (**Supplementary Figure 2a**). Nevertheless, through inclusion of protein values (‘NPX’) below the LLOD, we were able to identify pQTLs for some low abundance proteins that otherwise would have been missed (e.g., a *cis*-pQTL at IL17C, for which 83% of samples had levels below the LLOD) (**Supplementary Figure 2b**). The number of genomic loci associated with each protein ranged between 1 and 8 (**Figure 2b**), but was less than 4 for the majority of proteins. Examples of multi-locus-regulated proteins include TNFSF10 (TNF-related apoptosis-inducing ligand, TRAIL) and IL12B (interleukin-12B), both of which had 1 *cis*- and 7 *trans*-pQTLs (**Figure 2c,d**). Conditional analyses using GCTA-COJO (**Methods**) revealed the presence of an additional 47 independent signals, which were mostly *cis*. This raised the total number of pQTL signals from 180 (59 *cis*, 108 *trans*) to 227 (99 *cis*, 128 *trans*) (**Supplementary Table 4**).

To validate our genome-wide association study (GWAS) findings, we tested our significant pQTLs for replication in an independent cohort of 1,585 participants recruited as a part of the ARISTOTLE trial^13–15^. Of the 180 pQTL signals, we were able to test 174 in the ARISTOTLE data, of which 168 had a directionally consistent effect estimate. There was a strong correlation (Pearson *r*=0.97) between the pQTL effect estimates in ARISTOTLE and in the discovery meta-analysis; this correlation was consistent for both *cis-* and *trans*-pQTL effect sizes (*r*=0.99 and *r*=0.94) (**Supplementary Figure 3**). Out of the 174 pQTL signals, 32 were replicated at p≤5×10^-10^ and 72 at *p*≤2.9×10^-4^ (a Bonferroni-corrected threshold), respectively (**Supplementary Table 5**).

In line with other GWAS, we observed an inverse relationship between effect size and minor allele frequency (MAF), with pQTLs driven by rarer variants generally showing larger effect sizes (**Supplementary Figure 4**). The proportion of variance explained (PVE) by the significant sentinel variants from our discovery meta-analysis varied from 0.003 for NTF3 (Neurotrophin-3, NT-3) to 0.285 for CCL8 (also known as Monocyte Chemotactic Protein 2, MCP2) (**Supplementary Figure 5**).

### Annotation and characterisation of *cis*-pQTLs

Of the 59 *cis*-pQTLs identified, 11 sentinel variants were protein-altering variants (PAVs) (10 missense and 1 splice acceptor), while a further 10 sentinel variants were in high linkage disequilibrium (r^2^>0.8) with a protein-altering variant (all missense). PAVs can result in false positive *cis*-pQTL signals by altering protein epitopes which affects binding of antibodies used in proteomic assays^16^. However, they can also impact the abundance of plasma proteins through several mechanisms, including protein translation, secretion into the circulation, enzymatic cleavage of pre-proteins, as well as protein clearance and degradation. Alternatively, plasma protein abundance can also be affected by altered transcriptional regulation in blood cells or other tissues.

We next examined the degree to which our 59 *cis*-pQTLs were explained by corresponding *cis*-eQTLs, by comparing our findings with publicly available *cis*-eQTL data. In a meta-analysis of whole blood eQTL data from the eQTLGen Consortium^17^, we found a genome-wide significant (*p*<5×10^-8^) *cis*-eQTL for 32 of the 59 *cis*-pQTLs, where the *cis*-eQTL target gene encodes the *cis-*pQTL target protein. However, systematic statistical colocalization analyses using COLOC showed that only six (rs34790908-*TNFSF12*, rs72912115-*TGFA*, rs471994-*MMP1*, rs674379-*CD5*, rs450373-*CXCL5*, rs5744249-*IL18*) of these *cis*-eQTLs colocalised with their cognate *cis*-pQTLs (**Supplementary Table 6**), indicating that the remaining 26 eQTL-pQTL pairs may not share the same underlying causal genetic variant.

Of the 6 colocalising eQTL-pQTL pairs, 5 were directionally consistent. However, the eQTL and pQTL for IL18 at rs5744249 were oppositely associated with the mRNA and protein levels. rs5744249 resides in intron 2 of *IL18* and is in high LD (r^2^>0.8) with a 3’ UTR variant (rs5744292, r^2^=0.98 | 1000G EUR), but no PAVs. Therefore, the directional discordance is not easily explained either by an artefactual pQTL signal due to altered antibody binding or by a difference in the release of IL18 into the circulation due to differences in protein structure.

To extend our search to tissue- and cell-type-specific *cis*-eQTLs, we explored data from the Genotype-Tissue Expression (GTEx) (v8) project^18^ and the eQTL Catalogue^19^. Systematic COLOC analyses revealed colocalising (PP>0.8) *cis*-eQTLs in at least one tissue or cell type for 30 of the 59 *cis*-pQTLs (**Supplementary Tables 7-8**); 15 were highlighted by both eQTL resources, 10 by GTEx only, and the remaining 5 by the eQTL Catalogue. This included all 6 colocalising *cis*-eQTLs from eQTLGen, as well as several *cis*-eQTLs that did not reach the genome-wide significance threshold, or the significance threshold set by GTEx. Taken together, these findings suggest that at least 50% of our *cis*-pQTLs may be driven by underlying cognate *cis*-eQTLs.

In most cases, colocalization (PP>0.8) between *cis*-eQTL-pQTL pairs was observed across two or more distinct tissues or cell types, up to a maximum of 41 (for rs1883832-*CD40*). In other cases, colocalization was observed in just a single tissue or cell type. For example, the sole colocalising *cis*-eQTL signal (rs62360376) for *GDNF* was in skeletal muscle from GTEx, where, according to the Human Protein Atlas^20^, *GDNF* mRNA is enriched (i.e., tissue-enhanced) relative to other human tissues. GDNF production by skeletal muscle is responsive to physical activity, and has been shown to be a survival factor for peripheral motor neurons^21,22^. Similarly, there were several colocalising *cis*-eQTLs from the eQTL Catalogue highlighting specific cell types, including one for *TNFRSF9* (rs1776354) in Natural Killer Cells, and another for *IL18R1* (rs2270297) in Th17 memory cells.

### Identifying the mediators of *trans*-pQTLs

We sought to identify the most likely gene mediators of the *trans*-pQTLs using the ProGeM bioinformatics tool^23^, which utilises genomic (e.g., *cis*-eQTL) and biological (e.g., gene ontology and pathways) annotation data from multiple sources. For some *trans*-pQTLs, we identified strong evidence to implicate a gene encoded near the pQTL as mediating the distant association with the target protein. Examples included receptor-ligand pairs such as IL6-IL6R, IL10-IL10RA, CCL2-CCR2, CCL4-CCR5, and CCL11-CCR3. We also identified genes mediating pQTLs through intracellular signaling pathways rather than direct ligand-receptor interactions. An example is rs385076, an intronic variant in *NLRC4*, which is a *trans*-pQTL for IL18. IL18 is synthesized as an inactive precursor (pro-IL18), which is cleaved by caspase-1 in the NLRC4 inflammasome to produce the active form of IL18 (**Figure 3a**). Since rs385076 is also a *cis*-eQTL for the inflammasome gene *NLRC4* (**Figure 3b**), together, these QTL data indicate that genetic variation in *NLRC4* alters its expression and likely results in altered inflammasome activity, with consequent effects on circulating IL18 levels.

**Figure 3.**
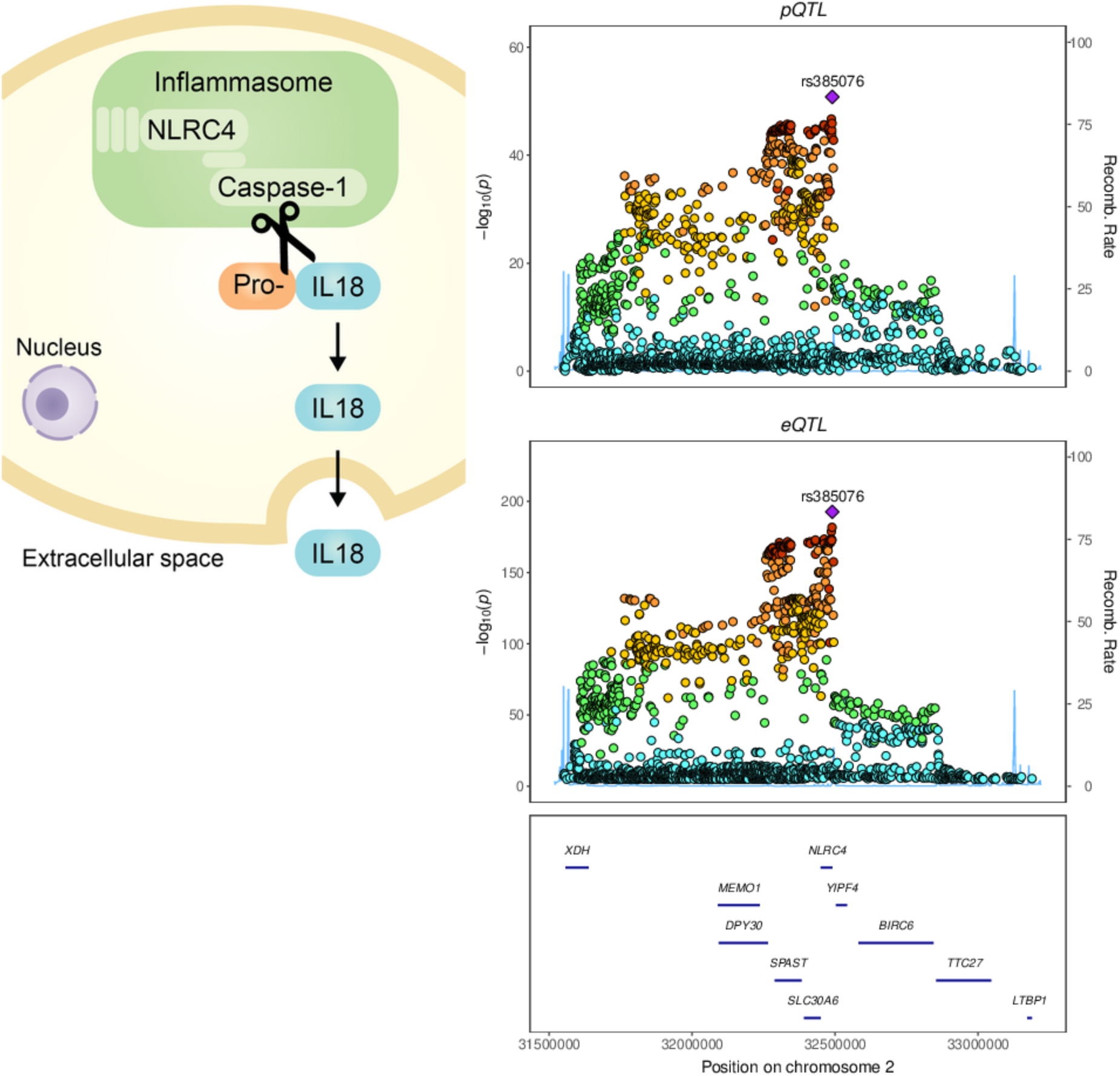
Genetic regulation of the inflammasome impacts plasma IL18 levels. **a)** Schematic illustrating the cleavage of pro-IL18 by caspase 1 and subsequent secretion of mature IL18 from the cell into the extra-cellular space. **b)** Regional association plots around *NLRC4* showing: the *trans*-pQTL signal for plasma IL18 protein (top) from this study (n=15,150), and the *cis*-eQTL signal for *NLRC4* (bottom) in whole blood from the eQTLGen study (n=31,684)^44^.

Following a manual literature review to refine the ProGeM output, we were able to narrow down the most likely mediating gene(s) to either one or two candidates for 100 of the 121 *trans*-pQTLs (**Supplementary Table 9**). For 94 of them, manual review highlighted one of the three nearest genes to the sentinel variant as the primary candidate, many of which are supported by previous biochemical and cellular studies. For example, for the pQTL at rs13010492 associated with CCL19 (C-C Motif Chemokine Ligand 19) levels, we prioritized *CTLA4* as the most likely mediator, which has been shown to regulate the mRNA expression of *CCL19*^24^. Similarly, the prioritized gene at the rs13103023 locus, *KLB*, encodes a necessary cofactor that enables signaling of the *trans*-affected protein, FGF19 (Fibroblast Growth Factor 19)^25^.

For 23 of these 84 loci where either one or two candidate genes were prioritised, ProGeM revealed functional links between both: (i) the sentinel variant and the nearby candidate mediating gene (e.g., *cis*-eQTL), and (ii) the same candidate mediating gene and the *trans*-affected protein(s) (e.g., through protein-protein interaction). We have previously shown that this type of convergence on the same candidate mediator is indicative of a strong candidate^23^. An example of this is the *trans*-pQTL at rs12075, which is associated with multiple chemokines (CCL2, CCL7, CCL8, CCL11, CCL13, CXCL6) that attract and activate leucocytes. rs12075 is a missense variant and a *cis*-eQTL (whole-blood, eQTLGen) for the *DARC* gene, which encodes the atypical chemokine receptor 1 (ACKR1) protein. A STRINGdb analysis revealed experimental evidence to show that ACKR1 is an interacting partner for 3 (CCL2, CCL7, CCL8) of the 6 *trans*-affected chemokines^26–28^. Previous studies have shown that ACKR1 acts as a negative regulator of inflammation by non-specifically binding both the CCL and CXCL family of chemokines^30^, thus explaining the multiple chemokine associations observed at this pQTL. In keeping with the effects on chemokines, rs12075 is also associated with white blood cell count, as well as monocyte and basophil count^31^(**Supplementary Figures 6-7**).

We found that plasma levels of some proteins were associated with numerous genetic loci, with IL12B, KITLG (KIT ligand, also known as stem cell factor, SCF), and TNFSF10 (TRAIL) regulated by seven genetic loci each. We hypothesized that the mediating genes at each of the regulatory loci for a given protein might be functionally related, enabling identification of shared pathways which in turn might facilitate the identification of the most likely mediating gene(s). We therefore generated protein-protein interaction networks for each of these multi-locus-regulated proteins and their respective candidate mediating genes from ProGeM (see Methods) (**Supplementary Table 9**), and visualized the interactions using String-db (**Supplementary Figure 8**). All three networks are significantly enriched with protein-protein interactions relative to the protein-coding genome (TNFSF10: *p*=1.44×10^-9^, KITLG: *p*<1×10^-16^, IL12B: *p*=8.22×10^-9^), and the clusters of interconnected proteins help pinpoint the mediating genes.

For the multi-locus-regulated protein TNFSF10 (TRAIL), we observed a cluster of eight interacting proteins including PLAUR, KNG1, and SERPINA1 (**Supplementary Figure 8a**). This cluster contains proteins encoded by five of the seven loci regulating TNFSF10, each of which converge on the plasminogen-activating system: (i) rs4760 is a missense variant in *PLAUR*, which encodes the plasminogen-activator urokinase receptor; (ii) rs5030044 is intronic in *KNG1*, which encodes the kininogen 1 protein, involved in bradykinin formation, which is regulated by plasmin; (iii) rs28929474 is a missense variant in *SERPINA1*, which cleaves many targets including plasmin; (iv) rs8178824 is intronic to *APOH*, which encodes a cofactor for plasminogen activation^33,34^; and (v) rs654488 is upstream of *MEP1B*, which has been shown to be triggered by the plasminogen-activating system^35^. Together, these findings highlight the most likely mediating genes at 5 of the 7 regulatory loci and indicate that TRAIL is regulated by the plasminogen-activating system.

As a further example, for the multi-locus-regulated protein KITLG (SCF), a key driver of hematopoiesis^36^ encoded by *KITLG*, we found a cluster of interacting proteins (e.g., PON1, ABCA1, PLTP) (**Supplementary Figure 8b**) converging on cholesterol metabolism. To determine whether the 7 *trans*-pQTLs for KITLG were also associated with cholesterol-related traits at the variant level, we performed a phenome-scan of the sentinel variants and proxies in high LD (r^2^>0.8) using Open Targets. We found that 5 of the 7 *trans*-pQTLs were significantly (*p*<5×10^-8^) associated with levels of either HDL or LDL cholesterol, and some with other lipids such as triglycerides (**Supplementary Table 10**). Our findings therefore highlight a potential link between cholesterol metabolism and plasma KITLG levels, which may result in altered hematopoiesis. Indeed, the phenome-scan indicated several genome-wide significant associations between the KITLG *trans*-pQTLs and blood cell traits (**Supplementary Table 10**). Finally, for the interactions in the IL12B network (**Supplementary Figure 8c**) we did not identify a clear biological pattern among the interconnected candidate mediating genes.

### Overlap with GWAS of traits and diseases

Genome-wide association studies (GWAS) have identified thousands of genomic regions associated with common diseases^37^, including immune-mediated diseases (IMDs). Many of these disease-associated loci lie outside protein-coding regions and so the effector molecules and pathways by which these genetic variants confer disease risk are often unclear^38^. Integration of pQTL and GWAS data can help bridge this knowledge gap by linking disease risk loci to specific proteins. To this end, we looked for overlap between pQTLs, or proxy variants in high LD (r^2^≥0.8) with our sentinel variants, in disease GWAS (**Methods**). This revealed overlap between our pQTLs and disease-associated variants for 73 diseases (**Supplementary Figure 9, Supplementary Table 11**). Examples of genetically anchored protein-disease connections included: TNFSF11 (RANKL) with osteoporosis and hypothyroidism; NGF (nerve growth factor) with migraine; TNFSF12 (TWEAK) with hypertension; and FGF5 with hypertension and cardiovascular diseases.

We next focussed on IMDs in more detail, intersecting our pQTLs with IMD GWAS data to identify proteins linking genotype and disease phenotypes. We found that 31 of our pQTLs overlap GWAS hits for at least one common IMD, with 76 unique pQTL-protein-disease associations (**Supplementary Table 12, Supplementary Figure 10**). For example, we observed that a *cis*-pQTL for IL10 was also associated with risk of inflammatory bowel disease (IBD), with the allele associated with higher plasma IL10 also associating with reduced IBD risk, consistent with the anti-inflammatory effects of IL10. Some pQTLs showed diverging directions of effect on different diseases (e.g. the *trans*-pQTL at *IL6R* for plasma IL6 levels described earlier had opposing directions of effect on risk of rheumatoid arthritis and allergic diseases (**Supplementary Figure 10**), as previously described^39–41^). Clustering of these genotype-protein-disease associations recapitulated groupings of diseases with clinical or pathogenic similarity (**Supplementary Figure 10**). For example, ankylosing spondylitis, psoriatic arthritis and psoriasis clustered together. These diseases share IL-17 driven pathology and clinical features (e.g., peripheral joint and spinal inflammation and uveitis). Similarly, we observed clustering of asthma and allergic disease, examples of Th2-mediated pathology. Since the proteomic assay was not proteome-wide and the disease GWAS had a range of sample sizes (and therefore statistical power), we cannot make formal conclusions about IMD similarity from our data. Nevertheless, the clustering provides interesting information about examples of shared etiology related to specific proteins.

### *Trans*-pQTL data implicate the *LTBR*-*LTA* axis in multiple sclerosis pathogenesis

We identified a novel t*rans*-pQTL for lymphotoxin alpha (LTA, also known as TNF-beta) at rs2364485 on chromosome 12 (**Table 1**), an intergenic variant previously found to be associated with multiple sclerosis^42^. We found that the multiple sclerosis risk allele, rs2364485:A, was associated with higher plasma levels of LTA. We next applied the ProGeM algorithm which revealed two candidate genes in the region near the pQTL that might mediate the *trans*-pQTL: *TNFRSF1A* (encoding tumor necrosis factor receptor 1, TNFR1) and *LTBR* (encoding lymphotoxin beta receptor, LTBR). LTA is a ligand for TNFR1, but also can bind the membrane-bound receptor LTBR when bound to LTB as a heterodimer. Functional studies have shown that *TNFRSF1A* is the causal gene underlying a neighboring independent multiple sclerosis association in the region, about 70kb upstream from rs2364485. The sentinel variant at this neighboring signal, rs1800693, results in an alternative *TNFRSF1A* isoform due to skipping of exon 6^43^. We therefore sought to determine if *TNFRSF1A* is also the likely mediating gene for the LTA *trans*-pQTL at rs2364485, or whether *LTBR* is the more likely candidate.

Through mining of eQTL databases, we found that rs2364485 is a *cis*-eQTL for *LTBR* (but not *TNFRSF1A*) in multiple tissues, including in the eQTLGen consortium meta-analysis of whole-blood^44^, with the multiple sclerosis risk allele (rs2364485:A) associated with lower *LTBR* mRNA levels. Pairwise statistical colocalization analyses using conditioned *LTBR* eQTL data (from eQTLGen) and multiple sclerosis GWAS data^42^ (see **Methods**) showed that the rs2364485 *trans*-pQTL signal for LTA colocalises with both *LTBR* mRNA expression in whole blood (PP=0.79) and multiple sclerosis (PP=0.86) (**Figure 4**). Taken together, these data are consistent with a pathogenic model whereby the multiple sclerosis risk allele results in lower abundance of LTBR (the receptor) and consequently higher circulating levels of the ligand LTA.

**Figure 4.**
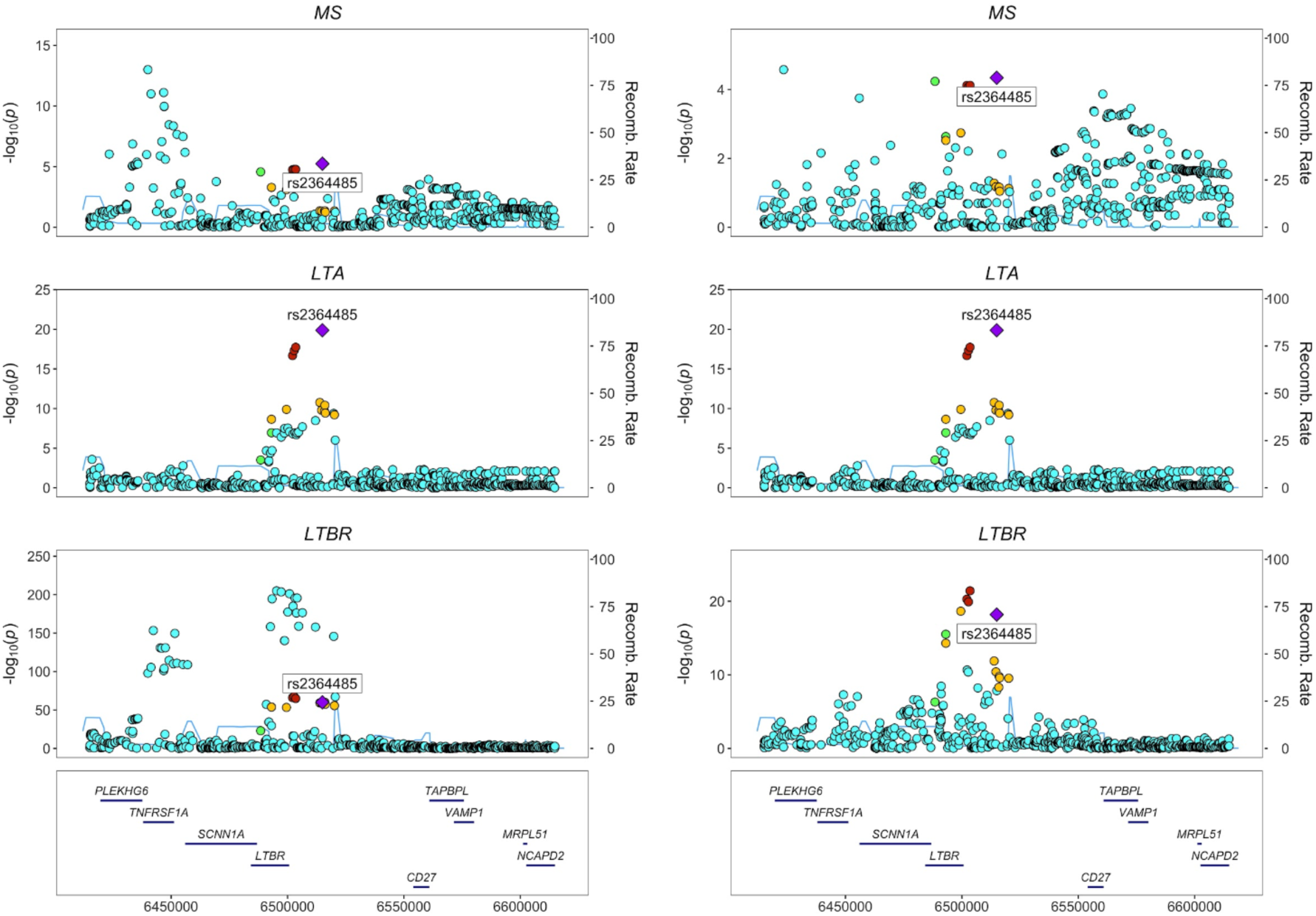
The *LTBR*-*LTA* axis in the etiology of multiple sclerosis (MS). (a) Unconditioned and **(b)** conditioned regional association plots at the *TNFRSF1A*-*LTBR* locus (rs2364485 +/- 100kb) for MS (top), plasma LTA protein levels (middle), and *LTBR* mRNA expression in whole blood from eQTLGen^44^ (bottom). MS associations were conditioned on rs1800693 (the strongest disease signal in the region). *LTBR* mRNA expression levels were conditioned on the following independent eQTLs: rs3759322, rs1800692, rs2228576, rs10849448, rs2364480, and rs12319859.

### Identifying proteins with causal roles in disease using Mendelian randomization (MR)

Observational studies comparing patients with IMDs to healthy controls have identified many proteins that are dysregulated. However, it is often unclear whether such proteins are upstream drivers of the disease process or are merely downstream markers. Distinguishing these possibilities is important therapeutically, as pharmacological targeting of the latter (e.g. C-reactive protein) is unlikely to be beneficial. We therefore applied Mendelian randomisation (MR), an approach that tests the causal role of an exposure in a disease in observational data using genetic variants as instrumental variables^45^. We used the 58 proteins with *cis*-pQTLs outside the HLA region in our study as exposures and 14 IMDs as outcomes (see **Methods)**. By restricting our use of genetic instruments to *cis*-pQTLs, we reduce the likelihood of violating MR assumptions through horizontal pleiotropy. Using Generalised Summary-data-based MR (GSMR)^46^, we found 22 significant (FDR<0.01) putative causal associations (**Figure 5**, **Supplementary Table 13**). To evaluate the robustness of these associations, we first checked the strength of the disease association in the GWAS summary statistics. Of the 22 protein-disease MR associations, we eliminated 5 due to the lack of convincing disease association (smallest p-value at the locus P>1×10^-4^). For the remaining 17 MR associations, we then evaluated whether there might be confounding due to LD by estimating the r^2^ between the sentinel pQTL and the disease-associated variant to determine whether the association signals were likely shared or distinct. For 12 of 17 disease-protein pairs, r^2^ was >0.8 and therefore unlikely to be due to confounding by LD (**Supplementary Table 14**). These 12 were: IL12B with inflammatory bowel disease (IBD) and both its major subtypes, Crohn’s disease (CD) and ulcerative colitis (UC); CD40 with rheumatoid arthritis, multiple sclerosis, IBD and CD; IL18R1 with CD and eczema; CD6 with IBD; CXCL5 with UC; and CD5 with primary sclerosing cholangitis. These results highlighted a number of established links between proteins and inflammatory diseases that are supported by other lines of evidence. For example, we found that genetic predisposition to higher plasma IL12B levels was associated with increased risk of IBD, consistent with the therapeutic benefit of ustekinumab (an anti-IL12/23 monoclonal antibody) in this disease (**Supplementary Table 15)**.

**Figure 5.**
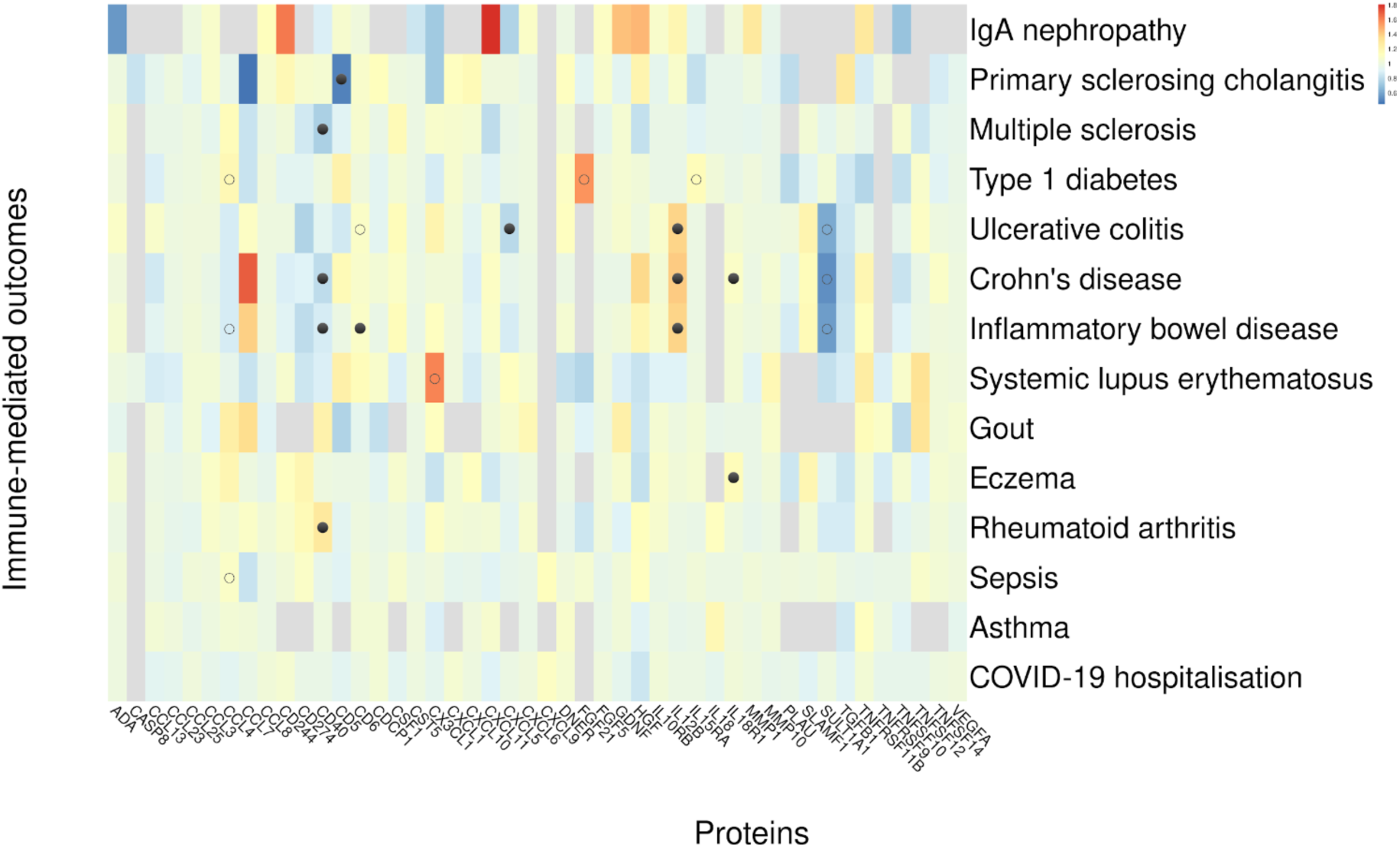
GSMR results for circulating proteins and immune-mediated diseases. Effect estimates are from GSMR analysis^46^ using *cis*-pQTLs which involved at least 10 genome-wide significant and quasi-independent variants required for the GSMR analysis. Cells are coloured according to Z-scores - Red: higher genetically-predicted plasma protein levels are associated with higher risk of disease; blue: higher genetically-predicted plasma protein levels are associated with lower risk of disease; gray: fewer than 10 variants available to run the GSMR analysis. Associations with FDR ≤0.01 are denoted with dots, with filled circles indicating those that are robust to confounding by LD and open circles indicating those that were not.

Another example was our MR finding implicating CXCL5 in the pathogenesis of UC. CXCL5 is a chemokine that acts on neutrophils, and a potential therapeutically tractable target. The plasma *cis*-pQTL for CXCL5, colocalised with *cis-*eQTLs for *CXCL5* in both blood and gut tissue, and with the UC GWAS signal (**Figure 6a**). Furthermore, the importance of CXCL5 in the pathogenesis of UC was supported by analysis of mucosal expression of *CXCL5* transcripts in gut samples from patients with IBD and healthy controls. IBD TAMMA is a publicly available, open access resource for interrogating transcriptomic data across multiple datasets in a meta-analysis framework^47^. Using this platform, we observed that the expression of transcripts encoding *CXCL5* was significantly increased in mucosal biopsies sampled from patients with UC in comparison with biopsies from healthy control participants (log_2_ fold change 7.07, P<1.98×10^-174^) (**Figure 6b**). Indeed, in patients with UC, *CXCL5* was the third most highly upregulated transcript in UC across the entire transcriptome (**Figure 6c**). We replicated these findings in three independent datasets (**Figure 6d**). Of note, our GSMR analysis revealed the association of CXCL5 was restricted to UC (unadjusted P=2.3×10^-6^), with no significant association in CD (unadjusted P 0.4) (**Figures 6a,e**). *CXCL5* gene expression in gut samples from IBD patients was higher in UC than in CD (**Figure 6b**), supporting a pathogenic effect of CXCL5 specific to UC rather than CD. Somewhat counter-intuitively (given the upregulation of *CXCL5* in UC patient tissue samples), evaluation of the direction of MR association effect revealed that genetic susceptibility to higher plasma CXCL5 reduces risk of UC (**Figure 6e**). This effect was consistent across 12 of the 13 the individual genetic variants used in the MR score (**Supplementary Figure 11**). We found consistent direction of effects for the CXCL5 plasma pQTL and the blood and gut eQTLs (**Supplementary Figure 12**), so the lower UC risk with genetic susceptibility to higher plasma CXCL5 levels could not be accounted for by discordant effects between mRNA and protein levels, or by differences between tissue at the site of inflammation and systemically.

**Figure 6.**
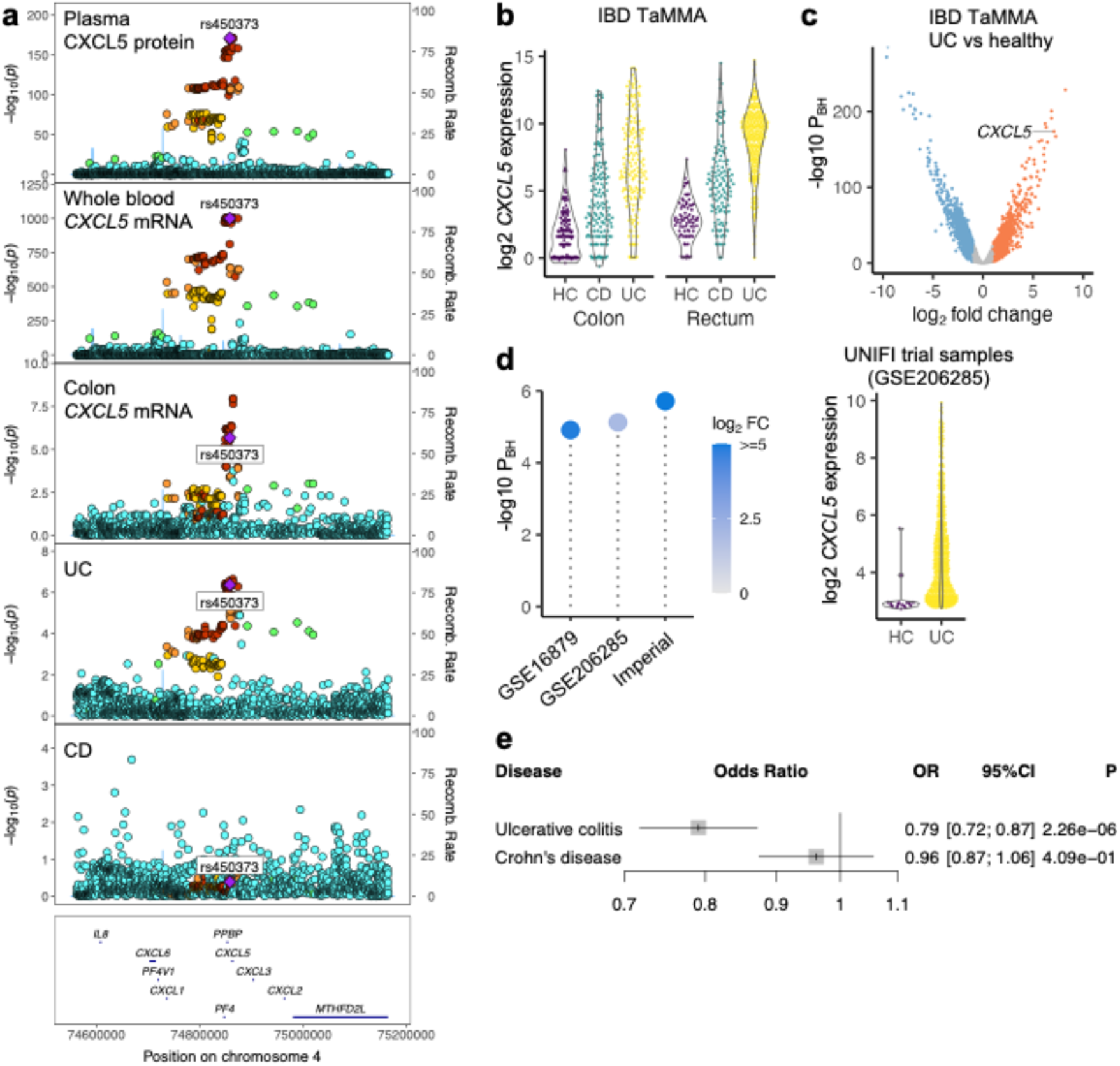
CXCL5 in UC pathogenesis. a) Genetic associations in the *CXCL5* gene region. From top to bottom: plasma CXCL5 pQTL, whole blood eQTL (from eQTLGen data), colon tissue eQTL (from GTEx), ulcerative colitis and Crohn’s disease (from the IBD Genetics Consortium). b) Violin plots showing *CXCL5* expression in gut mucosal samples from patients with UC or CD and from healthy controls (IBD TaMMA). P_BH_ = Benjamini-Hochberg adjusted p-values. c) Volcano plot showing differential expression analysis comparing colonic tissue from UC versus healthy controls (IBD TaMMA). Each point represents a transcript. Red and blue indicate significantly (5% FDR) up- and down-regulated, respectively. d) Replication of CXCL5 differential gene expression patterns in colon biopsies from ulcerative colitis (UC) patients compared to healthy controls (HC). Left hand panel: CXCL5 differential expression results from transcriptome-wide analysis across 3 cohorts. GSE numbers are Gene Expression Omnibus accession numbers. Imperial = Imperial UC cohort. Each lollipop represents a separate cohort. GSE16879 n= 24 UC patients versus n=6 healthy controls. GSE206285 n=550 UC patients versus n=18 healthy control samples. Imperial n=16 UC vs 6 healthy controls. P_BH_ = Benjamini-Hochberg adjusted p-values. Circle colour indicates the log_2_ fold change (FC) in *CXCL5* expression between UC and HC. Right hand panel: CXCL5 expression in colon biopsies sampled at baseline during the UNIFI clinical trial. Each point is associated to one participant in the ulcerative colitis (UC) or healthy control (HC) group, and its y coordinate reflects the normalised expression level of CXCL5. e) Forest plot showing Mendelian randomisation analysis for UC and CD. OR = odds ratio for the risk associated with a 1 standard deviation increase in the level of the protein. Square box indicates point estimate and whiskers indicate 95% confidence intervals (CI).

We observed that genetic predisposition to higher plasma CD40 levels was associated with increased rheumatoid arthritis risk, consistent with evidence from both animal models and humans implicating the CD40 pathway in rheumatoid pathogenesis^48^. In addition, our MR analysis identified a potential causal role for the CD40 pathway in IBD (including both Crohn’s and UC) and multiple sclerosis. Interestingly, however, the MR associations for these diseases had the opposite direction of effect compared to rheumatoid arthritis i.e. genetic predisposition to *lower* plasma CD40 levels was associated with *higher* risk of IBD and multiple sclerosis. These findings highlight how the same pathway can have pleiotropic effects on disease susceptibility, but also point to the complexity of immune-mediated disease pathogenesis, with opposing effects on different diseases.

## DISCUSSION

Here, we performed a large-scale pQTL GWAS of 91 circulating inflammation-related proteins measured using Olink immunoassays in 15,150 participants. We identified 180 significant associations (59 *cis*, 121 *trans*), involving 108 independent loci and 70 proteins. Our results provide validation of previous pQTL studies, with 130 (∼72%) of the 180 significant associations we observed here previously reported in peer-reviewed journals^1, 5–12,49^. We also highlighted 50 pQTLs (11 *cis*, 39 *trans*) that have not yet been reported in peer-reviewed articles, despite recent highly powered pQTL studies including Ferkingstad et al.^10^ (*n*=35,559 participants) and Pietzner et al.^6^ (*n*=10,708 participants), both of which utilized the Somascan platform for discovery. Thus, the novel contributions made by our study and others (e.g., ^5^) underscore the importance of conducting analyses using data not only from large samples but also from complementary proteomic platforms, including immunoassays and mass spectrometry. Each of these technologies have their own relative strengths, with the Olink proximity extension immunoassay system reducing the likelihood of cross-reactivity, while mass spectrometry is robust to epitope effects.

By comparison with eQTL databases, we showed that at least 50% of the *cis*-pQTLs we observed are likely driven by cognate *cis*-eQTLs in a diverse range of tissues and cell-types. eQTL studies in blood have been carried out using large sample sizes of a similar order of magnitude to the sample size in our pQTL study. However, eQTL studies in other tissues tend to be smaller, and so it is likely that some of the plasma cis-pQTLs observed here are underpinned by tissue-specific eQTLs that have not yet been detected due to lack of statistical power. We also observed evidence of directional uncoupling when comparing our observed *cis*-pQTLs with *cis*-eQTLs for cognate genes (e.g., rs5744249 associated with IL18 in plasma and *IL18* in whole blood in opposing directions). Directional uncoupling of eQTL-pQTL pairs has been reported (e.g., ^6,50^). However, it is important to bear in mind that plasma pQTL studies examine genetic effects on extracellular protein levels whereas blood eQTL studies examine the effects on intracellular RNA levels (predominantly in leucocytes). This has several implications. First, a wide range of tissues other than blood cells contribute to the plasma proteome, with the liver the major producer of circulating proteins. eQTL directional discordance has been observed between different tissues^18^ or even within different leucocytes^51^. As it is not always clear which cell types are the principal source of plasma proteins, it is conceivable that tissue-specific eQTLs could underpin discordance between blood eQTLs and plasma pQTLs. Second, abundances of proteins within the plasma are also affected by non-transcriptional mechanisms including cleavage, secretion and clearance.

Our pQTL study identified twice as many *trans* associations compared to *cis* (121 versus 59, respectively), which is typical of well-powered pQTL studies (e.g., ^5,6,10^). The integration of *cis*-pQTLs (and *cis*-eQTLs) with GWAS data provides useful, if sometimes obvious, insights into the upstream mechanisms of disease, since the mediating gene has usually already been suspected by virtue of the location of the GWAS signal. In contrast, *trans*-pQTLs represent a double-edged sword for interpreting genetic associations with disease. On the one hand, they often represent a less direct link from genotype to disease than *cis*-pQTLs, and from the perspective of causal inference analysis, are more vulnerable to violating the assumptions of MR through horizontal pleiotropy. On the other hand, the integration of GWAS data and *trans*-pQTLs can provide unexpected regulatory network-level insights into more distal pathophysiology, thereby highlighting potential therapeutic targets further downstream. For example, our study highlights the multi-locus-regulated protein KIT ligand (stem cell factor), a regulator of hematopoiesis, as a potential nexus point through which perturbed cholesterol metabolism might interface with downstream inflammatory responses. Indeed, deposition of cholesterol in the vascular wall leads to elevated hematopoiesis and a consequent increase in circulating leucocytes, which serves to exacerbate atherosclerosis^52,53^. KIT ligand may therefore be of particular relevance within the context of atherosclerotic cardiovascular disease.

As a further example of the utility of *trans*-pQTLs in understanding disease mechanisms, we identified a novel *trans*-pQTL (rs2364485) for LTA at a multiple sclerosis risk locus on chromosome 12. This multiple sclerosis risk locus contains two plausible causal genes (*TNFRSF1A*, *LTBR*) and two independent signals for multiple sclerosis risk (rs1800693, rs2364485). By integrating whole blood eQTL and multiple sclerosis GWAS data, we showed that *LTBR* is the most likely gene meditating our LTA *trans*-pQTL at rs2364485, and one of the multiple sclerosis signals at the locus. LTA is a member of the TNF superfamily of proteins and is the only member of this superfamily that is generated as a secreted protein rather than through cleavage of a membrane-bound protein. The multiple sclerosis risk allele is associated with lower expression of *LTBR* and higher circulating protein levels of LTA, a component of its ligand. This raises the question as to whether elevated LTA is secondary to lower LTBR, or vice versa (e.g. through compensatory receptor downregulation). The distinction between *cis-* and *trans*-QTLs enables us to address this. Given that the eQTL for *LTBR* is *cis*, and the pQTL for LTA *trans*, it is highly likely that former is the upstream effect, with the higher levels of soluble LTA occurring as a result of reduced binding to its receptor. This demonstrates the value of pairing QTLs for ligands and their receptors for deconvoluting the ordering of biological pathways.

Integration of pQTLs with GWAS disease signals revealed disease-protein connections reflecting both established and plausible putative mechanisms of pathophysiology. For example, a *cis*-pQTL for TNFSF11 (RANKL) overlapped with GWAS signals for osteoporosis and hypothyroidism. The former is consistent with RANKL’s well-established role in bone biology and RANKL is the target of the anti-osteoporosis drug denosumab^54^. However, RANKL also plays a role in the immune system^55^, and these effects may be relevant to risk of autoimmune hypothyroidism. A *cis*-pQTL for TNFSF12 (TWEAK) was associated with risk of hypertension. TWEAK is a cytokine predominantly produced by leucocytes and has pleiotropic actions, including on the endothelium^56,57^, potentially explaining the association with blood pressure. A *cis*-pQTL for NGF (nerve growth factor) was associated with migraine risk, consistent with NGF’s role in the nervous system. A *cis*-pQTL for FGF5 was also associated with susceptibility to hypertension and cardiovascular diseases, with the allele associated with higher plasma FGF5 levels associating with lower risk of cardiovascular diseases. Consistent with this, there are reports that FGF5 has cardioprotective effects in pig models^58^.

31 of our pQTLs overlap GWAS hits for at least one common IMD. Disease-protein links identified from this analysis highlighted commonalities in pathogenesis between specific IMDs (e.g. ankylosing spondylitis and psoriasis), mirroring the overlap in clinical manifestations (e.g. uveitis and spinal inflammation can occur in psoriasis as well as in ankylosing spondylitis). However, the contributions of proteins to IMD risk were sometimes complex, with the same protein conferring risk of one IMD but protecting from another. For example, genetic predisposition to higher levels of soluble IL6 had opposing effects on risk of rheumatoid arthritis and allergic disease. We observed a similar phenomenon for CD40, with genetic predisposition to higher CD40 increasing risk of RA but protecting against IBD and multiple sclerosis.

The development of biologic therapies targeting specific inflammatory proteins has transformed the clinical management of immune-mediated diseases^59^. Understanding which proteins are drivers of disease and distinguishing these from proteins that are simply markers of inflammation is therefore critical for development of new treatments. We used Mendelian randomisation to evaluate the causal contributions of proteins to different IMDs. Our results identify pathways that are already the target of existing drugs (e.g. IL12B in inflammatory bowel disease), providing confirmation of the utility of this approach, and also highlight new potential therapeutic targets.

One such example was CXCL5 in UC. CXCL5 is a chemokine that acts on neutrophils, which are known to play a role in tissue injury in IBD^60^. A previous study reported that serum levels of CXCL5 are higher in IBD patients than controls^61^. Our MR findings extend this by providing evidence to support a causal role for CXCL5 in UC pathogenesis. We provide further support for the importance of CXCL5 in UC pathogenesis by showing that *CXCL5* is one of the most upregulated transcripts in gut tissue samples from UC patients. However, the direction of effect of our MR association presents an apparent paradox, with genetic susceptibility to higher levels of plasma CXCL5 associated with lower UC risk. This could not be explained by differences between plasma protein levels and gene expression, since we observed directionally concordant findings when examining eQTL data from both the blood and gut samples. We hypothesize that the discrepancy between direction of effects from the MR analysis and from analysis of patient samples might reflect differences in the processes of disease initiation and perpetuation. By analogy, a non-coding genetic variant associated with lower gene and protein expression of *TNFSF15* (encoding the inflammatory cytokine TL1A) in monocytes and macrophages increases IBD susceptibility^62^, but in patients with active IBD TL1A is upregulated both systemically and in the gut^63–65^ and anti-TL1A therapies have recently shown efficacy in IBD in phase 2 randomised trials (NCT05013905 and NCT04996797)^66–68^. The MR association at CXCL5 appeared specific to UC, and not CD (the other major subtype of IBD), and in IBD gut samples, *CXCL5* was more upregulated in UC than CD. In UC the extent of neutrophilic inflammation in the gut correlates with disease severity^69,70^. Recent studies have implicated neutrophil recruitment as a key pathogenic event in UC correlating with important histopathological features, such as ulceration, and differentiating patient trajectories, including their responsiveness to different treatments^71,72^. Accordingly, excessive production of CXCL5 in non-immune compartments, including stromal cells and epithelial cells, which is driven by pathogenic cytokines, such as IL1-β^71,72^, represents a conceptually attractive pathogenic axis to target therapeutically, especially in patients refractory to conventional therapies. Consistent with this hypothesis, targeting CXCR2, the receptor for CXCL5, significantly attenuates preclinical models of UC^72^.

Another potential therapeutic avenue highlighted by our data was targeting the CD40 pathway in rheumatoid arthritis. CD40, a member of the TNF receptor superfamily, is a stimulatory receptor constitutively or inducibly expressed on both immune cells (B cells, dendritic cells, macrophages and microglia) and non-immune cells (including endothelial cells, epithelial cells, and keratinocytes)^73^. Its ligand, CD40L, is expressed primarily on activated T cells but also on a range of other immune and non-immune cells. CD40L-CD40 binding triggers immune cell activation and proliferation and inflammatory cytokine production. In B cells, CD40 stimulation leads to differentiation of B cells into IgG-secreting plasma cells, making it central to antibody responses. In a murine model of inflammatory arthritis, knock out or inhibition of the CD40 pathway resulted in reduced inflammation^74^. Observational studies have demonstrated upregulation of CD40L in the blood and tissues of patients with RA and other autoimmune rheumatic diseases^48^, and indicated a role of CD40/CD40L in the longitudinal course of RA from autoimmunity to joint inflammation^75^. These findings previously motivated development of drugs targeting the CD40 pathway in RA and other IMDs, but, unfortunately, anti-CD40L therapy was complicated by thrombosis due to cross-linking CD40L on platelets. Therapeutic targeting of CD40 rather than CD40L may avoid this. Our MR results suggest rheumatoid arthritis as a candidate for this approach. However, the plethora of approved treatments for rheumatoid arthritis make demonstrating value of a new agent is challenging, although the prospect of using such therapy very early in the disease course is an interesting possibility^75^. In addition, the directionally discordant effects we observed of CD40 on RA versus multiple sclerosis and IBD raises the possibility of triggering other forms of immune-mediated diseases as a side-effect of anti-CD40 therapy. This has some parallels with therapies targeting TNF-α (another TNF superfamily member). Anti-TNF-α therapy is effective in rheumatoid arthritis, but not in multiple sclerosis, and indeed can worsen multiple sclerosis or provoke *de novo* central nervous system demyelination^76,77^. Thus, for any anti-CD40 therapy careful monitoring for the development of features of multiple sclerosis or IBD would be required in both clinical trials and post-marketing surveillance.

The observation that genetic predisposition to lower circulating CD40 levels is a risk factor for IBD may at first appear paradoxical given that CD40 is generally considered to be pro-inflammatory. However, accumulating evidence points to the role of defective mucosal homeostasis and host-microbe interactions in the pathogenesis of IBD. Reduced CD40 may reduce the host’s immunological defence against gut microbes and allow them to breach the intestinal wall and thus trigger a chronic inflammatory response. This hypothesis is consistent with the emerging concept that IBD arises from relative immunodeficiency^78^. By analogy, there are coding variants at several other IBD risk loci that result in impaired host antibacterial responses, including *NOD2*^79^ and *ATG16L1*^80–82^, illustrating the importance of the immune system in gut barrier integrity, as well as the example of TL1A discussed earlier^62^.

Our study has several limitations. Our pQTL analysis was restricted to 91 proteins measured on the Olink Inflammation panel. These proteins do not include all the circulating proteins that are important in inflammation, and so this limits the generalisability of our findings, particularly with regards to genetic architecture. Since this was a pQTL meta-analysis, study-level technical variation resulted in heterogeneity, which necessitated filtering out of potentially spurious associations that were inconsistent across cohorts. There is a risk that some true biological signals were also removed in this process. Very large single cohorts with standardized sample processing such as UK Biobank will avoid this issue.

Our meta-analysis consisted predominantly of general population cohorts without inflammatory disease. As a result, some of the proteins measured were below the lower limit of detection in a high proportion of samples, although we were able to partially mitigate this by using untruncated NPX values. In addition, there may be context-specific pQTLs that are only present during infection or inflammation. eQTL studies using human immune cells stimulated *in vitro* (e.g. with lipopolysaccharide or interferon) have already demonstrated eQTLs that are not present in resting cells but become apparent in the context of cellular activation^83,84^. Conducting well-powered pQTL studies in patients with infection or inflammation will be an important future research endeavor. Where proteins exist in both membrane-bound and cleaved states, it is not always clear whether plasma proteomic assays are exclusively capturing the soluble form or also protein from cell membranes (e.g. arising from in vivo sources such as exo-/ectosomes, or *ex vivo* processes such as venepuncture or sample processing). This complicates the interpretation of the direction of effect from MR analysis. Future well-powered studies examining genetic determinants of cell surface proteins measured through flow cytometry would provide valuable complementary information to aid the interpretation of plasma pQTL studies. Finally, as with all epidemiological-scale pQTL studies, proteins were measured in plasma (i.e. the extracellular component of blood), which may not always be the disease-relevant biological matrix. For example, our MR analysis revealed that genetic predisposition to higher plasma IL12B is associated with increased IBD risk and the causal role for IL12B is supported by the therapeutic efficacy of ustekinumab. However, it is more likely that local levels of IL12B in the gut rather than circulating levels are most relevant to IBD pathogenesis. We speculate that in this instance the plasma pQTL mirrors the genetic effect in local tissues (perhaps because it is driven by subsets of leucocytes, which are also a major constituent of blood). However, for other proteins, the genetic determinants of concentration may differ between plasma and tissues, and so future tissue- and cell-specific pQTL studies will be valuable to understand differences in genetic signals across tissues.

In summary, we have used a large international consortium to robustly characterise the genetic determinants of a set of inflammation-related proteins, providing insight into the etiology of immune-mediated diseases. The increasing number of *trans*-pQTLs now being highlighted in proteomic GWAS should facilitate network-level insights into disease pathophysiology. The pQTL summary statistics generated in this study will be a valuable resource for interrogating future disease GWAS and will help guide drug target identification and prioritization.

## METHODS

### Cohorts

We recruited 11 cohorts of participants with genome-wide genetic data and plasma proteomic data measured using the Olink Target Inflammation panel. All participants provided written, informed consent.

### Biomarkers For Identifying Neurodegenerative Disorders Early and Reliably (BioFINDER)

The BioFINDER study^85^ is run by the Clinical Memory Research Unit and The Biomedical center, at Lund University, Sweden. Participants were recruited from the Memory and Neurology clinics at Skåne University Hospital as well as the Memory Clinic at Ängelholm’s Hospital. A total of 1,496 participants with mild cognitive symptoms, dementia or parkinsonian symptoms, as well as cognitively healthy elderly, have so far been enrolled in the study.

### Estonian Biobank (EstBB)

The EstBB cohort is a volunteer-based sample of the Estonian resident adult population (aged ≥ 18 years)^86^. A total of 487 participants contributed to this study. The research conducted in this project was approved by the Estonian Committee on Bioethics and Human Research (1.1-12/624) and by the Research Ethics Committee of the University of Tartu (application number 262/T-3, October 2016), data extraction nr K29.

### INTERVAL

The INTERVAL study^1,87,88^ is a prospective cohort study of blood donors, initially recruited for a randomized trial to determine optimal blood donation intervals. Volunteers were recruited and consented between 2012 and 2014 from 25 NHSBT (National Health Service Blood and Transplant) static donor centers across England. The INTERVAL study was approved by the Cambridge (East) Research Ethics Committee. A subset of 4,994 participants from INTERVAL with Olink inflammation panel data contributed to the present study. These participants had an age range between 49 and 78 years (median 61, IQR=55-66).

### Cooperative Health Research in the Region of Augsburg (KORA)

KORA is a series of independent population-based studies from the general population living in the region of Augsburg, Southern Germany. The KORA F4 study was conducted from 2006-2008 as a follow-up study to KORA S4 (1999-2001)^89^. 1,064 participants from KORA F4 contributed data to this study..

### Orkney Complex Disease Study (ORCADES)

The ORCADES is an ongoing family-based study in the isolated Scottish archipelago of Orkney, part of the Viking Genes studies (ed.ac.uk/viking). A total of 981 individuals with Olink inflammation panel data contributed to this pQTL study. All participants gave written informed consent and the study was approved by Research Ethics Committees in Orkney, Aberdeen (North of Scotland REC), and South East Scotland REC, NHS Lothian (reference: 12/SS/0151).

### Northern Sweden Population Health Study (NSPHS)

The NSPHS is a cross-sectional study conducted in the communities of Karesuando (samples gathered in 2006) and Soppero (2009) in the subarctic region of the County of Norrbotten, Sweden. A total of 866 individuals with Olink inflammation panel data contributed to this pQTL study.

### RECOMBINE/EIRA

The RECOMBINE/EIRA biobank was generated from participants with rheumatoid arthritis according to the American College of Rheumatology (ACR) 1987 or the 2010 ACR/EULAR diagnostic criteria and being part of the Epidemiologic Investigation of Rheumatoid Arthritis (EIRA) study^90^. At the time of recruitment, all participants were undergoing a change to or were at the start of a new treatment regimen between February 2011 and May 2013 at the Rheumatology clinic, Karolinska University Hospital, Stockholm, Sweden. Participants donated blood at baseline and then again at a 3-month follow-up visit. A total of 860 rheumatoid arthritis patients with Olink inflammation panel data contributed to this pQTL study.

### Stabilization of Atherosclerotic Plaque by Initiation of Darapladib Therapy Trial (STABILITY)

STABILITY is a randomized, controlled trial conducted in 15,828 patients with chronic coronary heart disease (CHD) comparing darapladib or placebo, in addition to standard of care (https://clinicaltrials.gov/ct2/show/NCT00799903)^91^. Data from 2,951 individuals with Olink inflammation panel and genotyping data contributed to this pQTL study. All participants gave written informed consent to the biomarker and genetic sub-studies and the study was approved by the National Ethics Committees in the participating countries.

### STANLEY (SWEBIC)

The SWEBIC cohort comprises participants meeting the diagnostic criteria for Bipolar disorder as per the DSM-IV recruited at Swedish sites in affiliation with the Stanley Medical Research Institute. In total, SWEBIC contributed 644 samples (cohort A [swe6]: 300, cohort B [lahl]: 344) with Olink inflammation panel data to this pQTL study. Ethical approval for SWEBIC was granted by the Regional Ethical Review Board in Stockholm, Sweden (2008/2009-31/2).

### VIS

CROATIA-Vis is a family-based, cross-sectional study in the island of Vis, Croatia. 899 participants with Olink Inflammation panel and genotyping data contributed to this pQTL study. All participants gave written informed consent and the study was approved by Research Ethics Committees in Croatia (Institutional Ethics Committee of the University of Split School of Medicine [protocol code 2181-198-03-04/10-11-0008] and Scotland).

### ARISTOTLE

The Apixaban for reduction in stroke and other ThromboemboLic events in atrial fibrillation (ARISTOTLE) trial was designed to investigate the safety and efficacy of the Factor Xa (FXa) inhibitor apixaban for secondary prevention of stroke in atrial fibrillation patients. Full details of the ARISTOTLE cohort are available at^13–15^. In total, ARISTOTLE contributed 1,585 samples with Olink inflammation panel data for comparison with the pQTL discovery meta-analysis findings. All participants gave written informed consent to the biomarker and genetic sub-studies and the study was approved by the National Ethics Committees in the participating countries.

### Protein assays

Plasma proteins were measured using Olink immunoassays. We used the Olink Target-96 Inflammation panel, which measures 92 inflammation-related proteins (https://www.olink.com/resources-support/document-download-center/, accessed: 9^th^ September 2022) from multiplexed antibody-based immunoassays which uses a matched pair of antibodies for a protein linked to paired oligonucleotides^92^. Proteomic data for each cohort were generated at Olink laboratories in Uppsala. Normalized Protein mapping to UniProt (https://www.uniprot.org/) identifiers was provided by Olink with additional information obtained from BioMaRt^93^. Protein eXpression, or (NPX), is Olink’s normalised relative units in log_2_ scale. Olink defines the lower limit of detection (LOD) for quantification of each protein as 3 standard deviations above background (determined using blank control samples). During the course of the project, BDNF was removed from the inflammation panel by Olink due to assay problems therefore 91 proteins were included in our study (**Supplementary Table 2**).

### Genotyping

Each cohort was genotyped on a SNP array and imputed using either a 1000Genomes or Haplotype Reference Consortium (HRC) panel (**Supplementary Table 1**).

### Cohort-level pQTL mapping

A GWAS analysis was run for each protein in each cohort using an additive association model with protein level as the dependent variable. Proteins were inverse-rank normalised as the outcomes in the regression models. Population substructure was adjusted for by including genetic principal components as covariates. We also included age, sex and other study-specific covariates in the model (see **Supplementary Table 1**). To avoid proteins with truncated distributions due to LOD with multiple tied values that would violate linear regression assumptions, pQTL analysis was performed using continuous protein values (including those below the LOD where relevant). We illustrate the value of this approach in recovering biological signals in **Supplementary Figure 2b**.

### pQTL meta-analysis

We meta-analysed pQTL summary statistics from each cohort (**Supplementary Table 1**), representing a total of 15,150 participants. A schematic of our analysis pipeline is shown in **Supplementary Figure 1**. Prior to the meta-analysis, we applied cohort-level filters to pQTL GWAS summary statistics with respect to MAF (≥0.001), HWE (*P*>10^-6^), and imputation score (*r* ≥0.3 or SNPTEST proper_info>=0.4). For each cohort, we generated QQ plots and Manhattan plots for visual examination using the R packages qqman v0.1.4 and QCGWAS v1.0-8. We performed the meta-analysis with the METAL software (version 28.8.2018), using inverse-variance weighted analysis of regression betas and standard errors from the cohort-level summary statistics. We observed no evidence of genomic inflation (mean lambda 0.9945, range 0.9945-1.0485) across the 91 proteins. From the meta-analysis summary statistics, we generated QQ and Manhattan plots for each protein GWAS (**Supplementary Figure 13**). The forest plots were generated using the gap package v1.2.3-6. Regional association plots were generated using LocusZoom 1.4 (**Supplementary Figure 14**). We defined statistical significance as *P*<5×10^−10^ (based on Bonferroni correction of the conventional ‘genome-wide’ significance threshold *P*<5×10^-8^ for approximately 100 proteins).

To remove potentially erroneous meta-analysis signals arising due to an extremely strong association in a single cohort, we examined the meta-analysis results at each sentinel variant by visual inspection of the forest plot and imposed the following criteria: 1) to be included in the meta-analysis, a variant was required to be available in at least three studies and in at least 3,500 participants; 2) in order to be declared significant, we required a meta-analysis P<5×10^−10^, and, if there was evidence of heterogeneity with *I^2^* >30%, then we required the P-value in at least three studies to be <0.05 and the direction of effect in those studies to be consistent with the overall meta-analysis results. These were implemented through modification of METAL source code.

### Replication cohort

We compared the results from our primary meta-analysis to pQTL results generated in an independent set of 1,585 participants from the ARISTOTLE study^13–15^ for validation purposes.

### Definition of pQTL sentinel variants and regions

We defined a pQTL as a genetic locus significantly (*P*<5×10^-10^) associated with protein abundance. We defined the sentinel variant at a locus as the variant with the lowest P-value in the region for a given protein. We used the following approach for each protein to define genomic regions and the sentinel variant in each: 1) we first obtained a list of significant (*P*<5×10^-10^) variants and the flanking region (+/-1Mb) for each variant; 2) overlapping regions were then iteratively merged until no overlapping regions remained; 3) the most significant variant in each resulting region was then defined as the sentinel variant. This approach has the flexibility to cope with long stretches of LD whilst avoiding the drawback of setting a longer than necessary region for all variants. The algorithm was implemented using bedtools v2.27.0. Signals within/beyond 1Mb of the transcription start site (TSS) of the gene encoding the target protein were defined as *cis* and *trans*, respectively.

### Protein variance explained by pQTLs

We used the following equation to estimate the proportion of variance explained (PVE) by (*T*) pQTLs from the meta-analysis summary statistics for each protein:

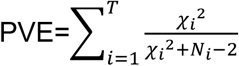

where *χi*^2^ is the chi-squared statistic based on each associated variant’s effect size and its standard error and *N_i_* is the associated sample size. We estimated the variance of the PVE for each variant as 2(*N_i_* -2)/[(*N_i_* -1)^2^(*N_i_* +1)] ^94^. For visualization of effect size in **Supplementary Figure 4**, we used the associated form 2*β*^2^[*f(1-f)*] ^95^, where *β* is the effect size and *f* the effect allele frequency, respectively.

### Conditional analysis

To identify conditionally independent signals within a genomic region, we performed approximate stepwise conditional analyses using GCTA v1.93.0beta with the ‘- -cojo-slct’ option, using effect sizes and standard errors from the meta-analysis. We estimated the correlation between variants using individual-level data from the INTERVAL study as a reference. As GCTA imputes LD from mean genotypes when they are missing, to avoid bias we excluded variants with MAF<1% (unless they were sentinel variants). For stepwise selection, we considered only those variants passing the genome-wide threshold (*P*≤5×10^-10^), rather than all variants in the region^96^. As in certain cases GCTA conditional analysis yielded results involving pairs of variants in relatively high LD (*r^2^*≥0.7), we used independent genetic variants (*r^2^*≤0.1)^97^ in the INTERVAL imputed genotype data whilst forcing the inclusion of the sentinel variants in the pruned set^98^ (**Supplementary Table 4**).

### Identification of known pQTLs

To identify previously reported pQTLs, we manually curated published results from literature obtained from the NCBI web interface (https://pubmed.ncbi.nlm.nih.gov/) through its Entrez programming utility R/rentrez^99^, PhenoScanner v2^39^, and the NHGRI-EBI GWAS Catalog with phenotypes mapped to the Experimental Factor Ontology (EFO)^100^ (EFO_0004747), restricting to associations reported in European-ancestry populations. We selected variants that reached the conventional genome-wide significance threshold P=5×10^-8^ and that were in high LD (*r^2^*=0.8) with the meta-analysis pQTL sentinel variant, paying attention to any possible protein isoforms on the Somascan platform. We also considered significant loci reported from cohorts in this^1,101^ and meta-analyses^5^ as known loci, as were those published at later stage of this analysis^6,10,12,49,102^.

### Variant annotation

We obtained the absolute distance of sentinel variants to the TSS of the gene encoding the target protein using the rGREAT (Genomic Regions Enrichment of Annotations Tool)^103^ R package rGREAT. We annotated sentinel variants and LD proxies (defined as r^2^≥0.8, using the INTERVAL dataset as the LD reference panel) using the Ensembl Variant Effect Predictor (VEP, v98.3)^104^ including the LOFTEE plugin^32^.

### Prioritizing likely mediating genes at *trans*-pQTLs

To prioritize likely mediating genes at *trans*-pQTLs, we used an adapted version of the ProGeM method, which has been described previously^23^. For the *cis*-eQTL lookups of our *trans*-pQTL sentinel variants we utilized eQTLGen^44^, the eQTL Catalogue^19^, and the Genotype-Tissue Expression project (GTEx, v8)^18^ data. To determine whether the *trans*-pQTL sentinels are likely to be causal *cis*-eQTL variants in the eQTL Catalogue and GTEx data, we used the fine-mapped eQTL credible sets available at the eQTL Catalogue (https://www.ebi.ac.uk/eqtl/Data_access/). For the eQTLGen data, where credible sets were not available, we used a manual approach whereby we: (i) first defined a region around each *trans*-pQTL sentinel variant of +/-500kb; (ii) identified the variant with the lowest *cis*-eQTL *p*-value in this region for the *cis*-affected gene(s); and (iii) checked to see whether this sentinel *cis*-eQTL variant is the same sentinel variant for the *trans*-pQTL, or if the two are in high LD (r^2^>0.8).

For the “top-down” component of ProGeM, we first identified locally-encoded genes using a window around each *trans*-pQTL sentinel variant of +/-500kb. We then probed the proteins encoded by these local genes using: (1) protein:protein interaction (PPI) data; and (2) data from functional annotation databases. With the PPI data we aimed to determine whether there is evidence to indicate that genes residing close to each sentinel variant might physically interact with the corresponding *trans*-affected protein. We used the Bioconductor package STRINGdb (v2.8.4) to identify any pairwise interactions. We used data from functional annotation databases to determine whether any local genes encode proteins that might be functionally related to the corresponding *trans*-affected protein(s). For both the *trans*-affected proteins and locally encoded proteins, all assigned Gene Ontology terms, Reactome pathways, and KEGG pathways were extracted using the Bioconductor biomaRt (v2.52) and KEGGREST (v1.36) packages. To assess whether there is significant overlap between the functional annotation terms/pathways assigned to locally encoded proteins and the corresponding *trans*-affected proteins, we determined the number of shared and non-shared terms for each local gene and the corresponding *trans*-affected protein. Fisher’s exact test was then applied for each local gene/*trans*-protein combination, and p-values were Bonferroni-corrected for the number of local genes at each given *trans*-pQTL. The background set of terms for each *trans*-pQTL was composed of all terms assigned to all local genes at the locus (i.e., all protein-coding genes within 500kb from the sentinel variant).

To determine the most likely mediating genes for the multi-locus regulated proteins IL12B, KITLG (SCF), and TNFSF10 (TRAIL), we used the STRINGdb^105^ webtool to identify interactions or functional relationships between genes residing at distinct loci. This is based on the assumption that if the true mediating genes at distinct loci are all associated with plasma levels of the same protein, then they may share some other functional relationship. As input to STRINGdb, we used all proteins encoded by candidate mediating genes identified by ProGeM (**Supplementary Table 9**) at each of the loci for a given protein, as well as the relevant *trans*-affected protein. We deemed clusters of proteins residing at distinct loci with multiple functional interactions to be the most likely mediating genes at their respective loci. We performed a phenome-scan of the *trans*-pQTLs for *KITLG* using the Open Targets Genetics webtool^106^.

### Statistical colocalization analysis

We performed pairwise statistical colocalization analyses of *cis*-pQTLs identified in the meta-analysis with all cognate *cis*-eQTL data derived from eQTLGen^44,107^, the eQTL Catalogue^19^, and GTEx v8^18^. We extracted the meta-analysis summary statistics for each *cis*-pQTL sentinel and their +/-1Mb flanking regions, then extracted the same genomic windows from their cognate *cis*-eQTL data. We performed colocalization analyses using the coloc R package as implemented in v.21.01.1 of the eQTL Catalogue/colocalization workflow^19^ (https://github.com/kauralasoo/eQTL-Catalogue-resources). Briefly, coloc returns posterior probabilities indicating the likelihood that the following scenarios are true: (i) there is no association at the locus with either protein or mRNA (PP0), (ii) there is an association with either the protein (PP1) or the mRNA (PP2), but not both, (iii) there is an association with both the protein and the mRNA but with distinct causal variants (PP3) or with a shared causal variant (PP4). We considered a PP4>=0.8 to be robust evidence of colocalization between a *cis*-pQTL and its cognate *cis*-eQTL. As eQTLGen data only provides allele frequency (*f*) and z-score statistic (z) for a particular variant, we obtained the effect size (*b*) and its standard error (*se*) with *b* = *z*/*d*, *se* = 1/*d*, where 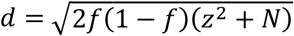, and *N* is the associate sample size^108^.

To investigate potential colocalization between a *trans*-pQTL (rs2364485) for LTA identified in this meta-analysis, a multiple sclerosis GWAS signal^42^, and a *cis*-eQTL for *LTBR* from eQTLGen^44^, we used a tool for multi-trait colocalization called HyPrColoc^109^. We obtained multiple sclerosis summary statistics (MSchip, “discovery_metav3.0.meta.gz”) from Patsopoulos et al. by request to the International Multiple Sclerosis Genetics Consortium (IMSGC). Due to a lack of genotype coverage at the *LTBR*/*TNFRSF1A* locus in the extended and replication samples from Patsopoulos et al., we selected the summary statistics from the “discovery” sample (*n*=41,505) for colocalization analyses, not the full meta-analysis. As a result, the *p*-value for association between the variant of interest (rs2364458) and multiple sclerosis in the discovery subset (*p*=5.78×10^-6^) was higher than reported in Patsopoulos et al. (*p*=2.0×10^-20^)^42^. We then extracted summary statistics for rs2364458 (+/-1Mb) (chr12:5514963-7514963) from each of the 3 datasets, and performed conditional analyses to adjust for any independent signals at the locus using GCTA-COJO. We ran this using a two-step approach: we first used the COJO-slct function to identify independent signals at the locus, and then for datasets with independent signals (i.e., in addition to rs2364485), we used COJO-cond to generate conditioned summary statistics for use in HyPrColoc. HyPrColoc returns the posterior probability that 2 or more traits colocalize, akin to PP4 from coloc. We considered a PP>=0.8 as robust evidence of colocalization between traits.

### Overlap of pQTL and disease traits

We used a **PhenoScanner** v2-based R code to look up associations of our pQTL sentinels and their LD proxies (r^2^≥0.8) in disease GWAS summary statistics.

### Mendelian randomization analyses

We performed MR analyses using the proteins with *cis*-pQTLs identified in this meta-analysis as exposures, and immune-mediated diseases (IMDs) as outcome variables. All MR analyses were run using the Generalized Summary-data-based Mendelian Randomisation (GSMR) method^46^. For each protein analysed, we defined a +/- 1 Mb window around the gene encoding it and extracted pQTL summary statistics for this region. For outcome data, we downloaded GWAS summary statistics for immune-mediated diseases (IMDs) from OpenGWAS (https://gwas.mrcieu.ac.uk/datasets/) and when larger sample and variants are available from the GWAS catalog (https://www.ebi.ac.uk/gwas/downloads). For IMDs with several alternative datasets available, we selected the dataset with the largest case sample size, provided it (i) had genotype data with sufficient coverage at the loci of interest, (ii) it was generated in European-ancestry samples so that it matched the ancestry of the participants in our pQTL meta-analysis, and (ii) betas and standard errors either available or calculable. We excluded UKB GWASs since they generally underestimate effect sizes of causal effects due to relatively small number of cases but large number of controls. We used the GSMR implementation in GCTA with at least 3 (--gsmr-snp-min 3) genome-wide significant (--gwas-thresh 5e-8), quasi-independent variants (--clump-r2 0.1) with specification of difference in allele frequency of each SNP between the GWAS summary datasets and the LD reference sample (--diff-freq 0.4) and p-value threshold for the HEIDI-outlier filtering analysis (--heidi-thresh 0.05) (https://yanglab.westlake.edu.cn/software/gcta/#Mendelianrandomisation), These criteria additionally excluded Celiac disease and primary biliary cholangitis. The final analysis involved 58 proteins and 14 diseases since it is from the *HLA* region whose MR results were unreliable. P-values were corrected for multiple testing using Benjamini-Hochberg correction, applied to the results for 331 models which involved at least 3 variants for both proteins and diseases.

#### *CXCL5* differential expression analysis in UC cohorts

Changes in *CXCL5* gene expression levels were evaluated in four independent cohorts, including the IBD Transcriptome and Metatranscriptome Meta-Analysis (TaMMA) platform^47^, the Gene Expression Omnibus (GEO) series GSE16879, GSE206285, as well as the Imperial UC cohort. IBD TaMMA (https://ibd-meta-analysis.herokuapp.com/) gives access to 3,853 transcriptomic profiles from 26 independent studies including IBD and control samples across different tissues, all processed with the same pipeline and batch-corrected^47^. Pre-computed differential expression results between colon biopsies from UC patients versus healthy donors were downloaded and plotted.

Data from GEO Series GSE16879 used in this study consist of colonic mucosa microarray expression profiles from healthy donors (n=6) and UC patients (n=24) sampled before first infliximab treatment^110^. CEL file import into R, and background correction, normalization and RMA calibration of the raw intensity data were carried out using the oligo package^111^. Only probe sets with median expression greater than 4 and associated to only one ENTREZ gene identifier were kept for analysis. Intensity data for different probe sets mapped to the same ENTREZ gene identifier were combined by taking the geometric mean sample wise. Tests of differential gene expression of ulcerative colitis samples compared to healthy control samples were performed with the limma package. P-values were adjusted for multiple testing with the Benjamini and Hochberg procedure.

GEO Series GSE206285 contains array-based transcriptomic data collected at baseline as part of UNIFI, a randomized placebo-controlled phase 3 clinical trial evaluating the efficacy and safety of ustekinumab^112^. RMA signal intensity profiles and associated donor information were downloaded from NCBI GEO. Only probe sets associated to only one ENTREZ gene identifier were kept for analysis. Intensity data for different probe sets mapped to the same ENTREZ gene identifier were combined by taking the geometric mean sample wise. Genes with median expression greater than 3 across all samples were tested for differential expression between ulcerative colitis samples (n=550) versus healthy control samples (n=18) using the limma package^113^. P-values were adjusted for multiple testing with the Benjamini and Hochberg procedure.

The Imperial UC cohort includes whole tissue biopsies from ulcerative colitis patients (n=16) and healthy volunteers (n=6). RNA was extracted (Qiagen RNeasy mini kit) and sequencing libraries were generated using NEBNext® Ultra™ RNA Library Prep Kit for Illumina® (NEB, USA) following manufacturer’s recommendations. Briefly, mRNA was purified from total RNA using poly-T oligo-attached magnetic beads. Fragmentation was carried out using divalent cations under elevated temperature in NEBNext First Strand Synthesis Reaction Buffer (5X). First strand cDNA was synthesized using random hexamer primer and M-MuLV Reverse Transcriptase (RNase H-). Second strand cDNA synthesis was subsequently performed using DNA Polymerase I and RNase H. Remaining overhangs were converted into blunt ends via exonuclease/polymerase activities. After adenylation of 3’ ends of DNA fragments, NEBNext Adaptor with hairpin loop structure were ligated to prepare for hybridization. Library fragments were purified with AMPure XP system (Beckman Coulter, Beverly, USA) and treated with 3 μl USER Enzyme (NEB, USA) at 37°C for 15 min, followed by 5 min at 95 °C. Then PCR was performed with Phusion High-Fidelity DNA polymerase, Universal PCR primers and Index (X) Primer. Library quality was assessed on Agilent Bioanalyzer 2100 and on Nanodrop ND-1000 Spectrophotometer. The library preparations were sequenced on an Illumina HiSeq platform, generating 150 bp paired end reads.

The resulting fastq files were processed with trimmomatic^1148^ (v. 0.39) to remove adaptor contamination and poor-quality bases. The output read files were mapped to the GRCh38 assembly of the human genome using Hisat2^1159^ (v. 2.2.1) with default parameters. The number of reads mapping to the genomic features annotated in Ensembl^11610^ with a MAPQ score higher than or equal to 10 was calculated for all samples using htseq-count^11711^(v. 0.11.3) with default parameters. Data for Ensembl genes with no associated ENTREZ gene identifier were discarded; the read counts for Ensembl genes mapped to the same ENTREZ gene identifier were summed up sample wise.

Differential expression analysis between ulcerative colitis versus healthy biopsies was performed in R (v. 3.6.1) using the Wald test as implemented in DESeq2^11812^. Only genes with an average read count across samples greater than or equal to 10 were tested for differential expression. P-values were adjusted for multiple testing with the Benjamini and Hochberg procedure.

### Data availability

Full per-protein summary statistics will be made available on publication.

### Code availability

GitHub: https://jinghuazhao.github.io/INF/, cambridge-ceu: +https://cambridge-ceu.github.io/public (modified METAL, pQTLtools).

## Supporting information

Supplementary Item

## Data Availability

All data produced in the present study are available upon reasonable request to the authors. Full per-protein summary statistics will be made available on publication of the peer-reviewed journal article.

## Acknowledgements

We thank:

study participants from the contributing cohorts;
the International Multiple Sclerosis Genetics Consortium, who provided multiple sclerosis GWAS summary statistics used in our analyses;
Aikaterina Siopi for support with SCALLOP Consortium administration;
the authors of the GCTA software for advice;
Bram Prins for help with INTERVAL study genotype data quality control.

## STABILITY and ARISTOTLE

Funding of the GWAS and proteomics studies of STABILITY and ARISTOTLE: These studies were supported by GlaxoSmithKline, BristolMyersSquibb, and the Swedish Foundation for Strategic Research (grant number RB13-0197).

## ORCADES

The Orkney Complex Disease Study (ORCADES) was supported by the Chief Scientist Office of the Scottish Government (CZB/4/276, CZB/4/710), a Royal Society URF to J.F.W., the MRC Human Genetics Unit quinquennial programme “QTL in Health and Disease”, Arthritis Research UK and the European Union framework program 6 EUROSPAN project (contract no. LSHG-CT-2006-018947). DNA extractions were performed at the Edinburgh Clinical Research Facility, University of Edinburgh. We would like to acknowledge the invaluable contributions of the research nurses in Orkney, the administrative team in Edinburgh and the people of Orkney. For the purpose of open access, the author has applied a Creative Commons Attribution (CC BY) licence to any Author Accepted Manuscript version arising from this submission.

## INTERVAL

Participants in the INTERVAL randomised controlled trial were recruited with the active collaboration of NHS Blood and Transplant England (www.nhsbt.nhs.uk), which has supported field work and other elements of the trial. DNA extraction and genotyping were co-funded by the National Institute for Health and Care Research (NIHR), the NIHR BioResource (http://bioresource.nihr.ac.uk) and the NIHR Cambridge Biomedical Research Centre (BRC-1215-20014) [*]. The academic coordinating centre for INTERVAL was supported by core funding from the: NIHR Blood and Transplant Research Unit (BTRU) in Donor Health and Genomics (NIHR BTRU-2014-10024), NIHR BTRU in Donor Health and Behaviour (NIHR203337), UK Medical Research Council (MR/L003120/1), British Heart Foundation (SP/09/002; RG/13/13/30194; RG/18/13/33946) and NIHR Cambridge BRC (BRC-1215-20014; NIHR203312) [*]. A complete list of the investigators and contributors to the INTERVAL trial is provided in reference [**]. The academic coordinating centre would like to thank blood donor centre staff and blood donors for participating in the INTERVAL trial.

This work was supported by Health Data Research UK, which is funded by the UK Medical Research Council, Engineering and Physical Sciences Research Council, Economic and Social Research Council, Department of Health and Social Care (England), Chief Scientist Office of the Scottish Government Health and Social Care Directorates, Health and Social Care Research and Development Division (Welsh Government), Public Health Agency (Northern Ireland), British Heart Foundation and Wellcome.

For the purpose of open access, the author has applied a Creative Commons Attribution (CC BY) licence to any Author Accepted Manuscript version arising from this submission.

*The views expressed are those of the author(s) and not necessarily those of the NHS, the NIHR, NHSBT or the Department of Health and Social Care. **Di Angelantonio E, Thompson SG, Kaptoge SK, Moore C, Walker M, Armitage J, Ouwehand WH, Roberts DJ, Danesh J, INTERVAL Trial Group. Efficiency and safety of varying the frequency of whole blood donation (INTERVAL): a randomised trial of 45,000 donors. Lancet. 2017 Nov 25;390(10110):2360-2371.

## (EstBB)

Estonian Biobank work was supported by the European Regional Development Fund and the programme Mobilitas Pluss (MOBTP108), No. 2014-2020.4.01.15-0012 GENTRANSMED and 2014-2020.4.01.16-0125 This study was also funded by the EU H2020 grant 692145, by the Estonian Research Council Grant PUT1660 and by the Estonian Research Council grant PUT (PRG1291). Data analyzes with Estonian datasets were carried out in part in the High-Performance Computing Center of University of Tartu.

## SWEBIC

The SWEBIC biobank was supported by the Stanley Medical Research Institute. The proteomic analyses in SWEBIC was funded by the Swedish foundation for Strategic Research (KF10-0039). For RECOMBINE and SWEBIC, the data handling and analysis were enabled by resources provided by the Swedish National Infrastructure for Computing (SNIC), partially funded by the Swedish Research Council through grant agreement no. 2018-05973.

## CROATIA-Vis

The CROATIA-Vis study was funded by grants from the Medical Research Council (UK), from the Republic of Croatia Ministry of Science, Education and Sports (108-1080315-0302; 216-1080315-0302) and the Croatian Science Foundation (8875). We thank the staff of several institutions in Croatia that supported the field work, including Zagreb Medical Schools, the Institute for Anthropological Research in Zagreb, the recruitment team from the Croatian Centre for Global Health, University of Split and all the study participants.

## KORA

The KORA study was initiated and financed by the Helmholtz Zentrum München – German Research Center for Environmental Health, which is funded by the German Federal Ministry of Education and Research and by the State of Bavaria. Furthermore, KORA research was supported within the Munich Center of Health Sciences (MC-Health), Ludwig-Maximilians-Universität, as part of LMUinnovativ.

The measurement of inflammatory biomarkers was funded by a grant from the German Center for Diabetes Research (DZD; to C. Herder and B. Thorand). This work was also supported by the Ministry of Culture and Science of the State of North Rhine-Westphalia and the German Federal Ministry of Health. This study was supported in part by a grant from the German Federal Ministry of Education and Research to the German Center for Diabetes Research (DZD).

## Supplementary Information

### Banner authors

Estonian Biobank Research Team

(Estonian Genome Centre, Institute of Genomics, University of Tartu):

Andres Metspalu

Lili Milani

Reedik Mägi

Mari Nelis

Georgi Hudjašov

### Supplementary Table titles

These refer to the accompanying tabs of the Excel file.

**Supplementary Table 1.** Overview of participating cohorts in the SCALLOP INF1 study.

**Supplementary Table 2.** Summary of the 91 proteins measured on the Olink Inflammation panel.

**Supplementary Table 3.** Summary statistics for the sentinel variants at 180 pQTLs.

**Supplementary Table 4.** Summary statistics from conditional analyses of 180 pQTLs using GCTA-COJO.

**Supplementary Table 5.** Summary statistics from the ARISTOTLE study.

**Supplementary Table 6.** Systematic pairwise coloc analyses of 59 cis-pQTLs with whole blood cis-eQTL data from eQTLGen.

**Supplementary Table 7.** Systematic pairwise coloc analyses of 59 cis-pQTLs with multi-tissue cis-eQTL data from GTEx v8.

**Supplementary Table 8.** Systematic pairwise coloc analyses of 59 cis-pQTLs with cell- and tissue-specific cis-eQTL data from the eQTL Catalogue.

**Supplementary Table 9.** Prioritization of candidate mediating genes for trans-pQTLs using ProGeM.

**Supplementary Table 10.** Phenome-wide association scan of 7 trans-pQTLs for stem cell factor (SCF) protein using Open Targets.

**Supplementary Table 11.** Overlap of pQTLs with disease GWAS signals.

**Supplementary Table 12.** Overlap of pQTLs with immune-mediated disease (IMD) GWAS signals.

**Supplementary Table 13.** GSMR analyses.

**Supplementary Table 14.** Prioritisation of signals from GSMR analyses.

**Supplementary Table 15.** Annotations of proteins with pQTLs that are druggable targets.

### Supplementary Figures and Supplementary Item titles

**Supplementary Figure 1.**
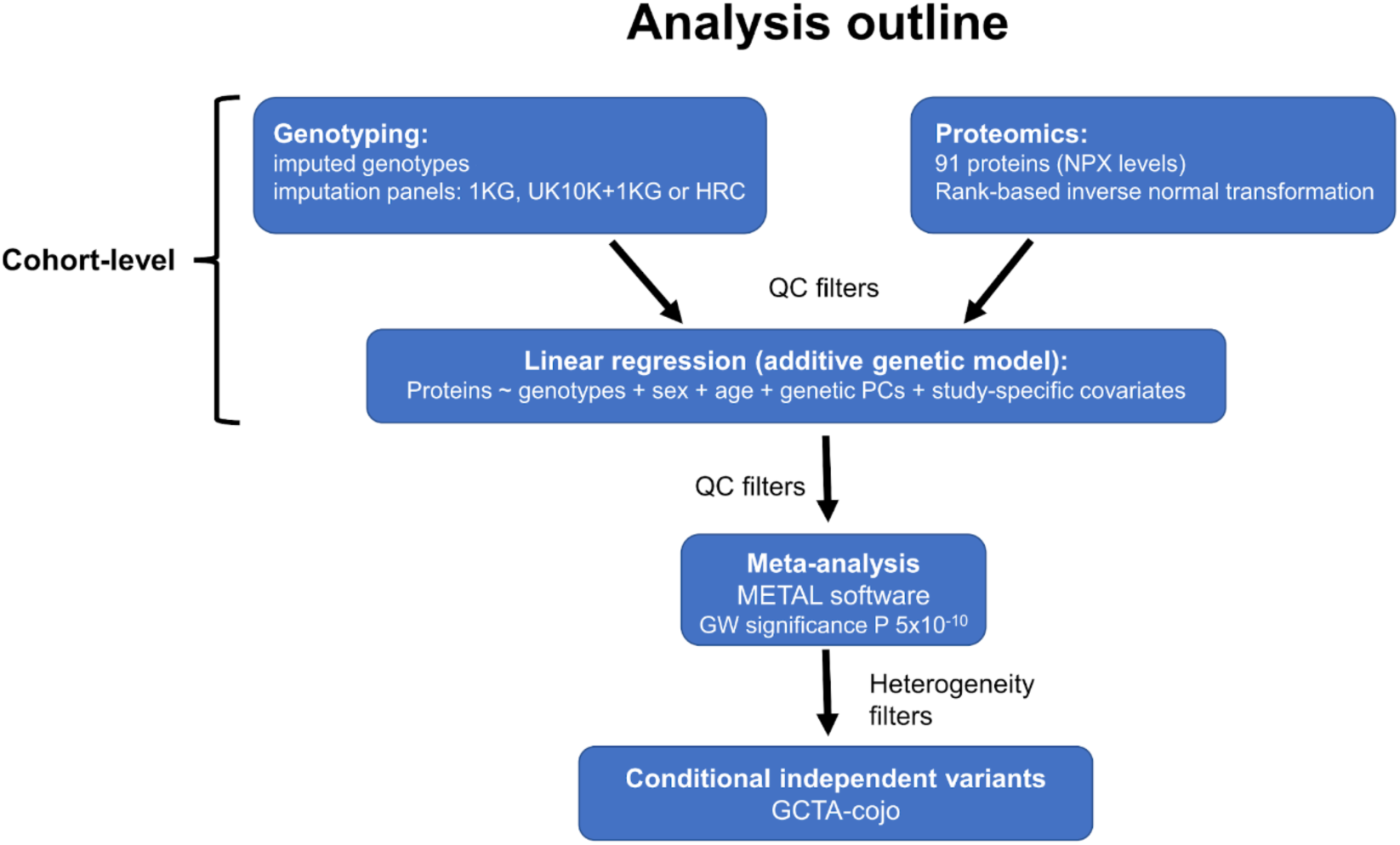
Overview of the pQTL analysis.

**Supplementary Figure 2.**
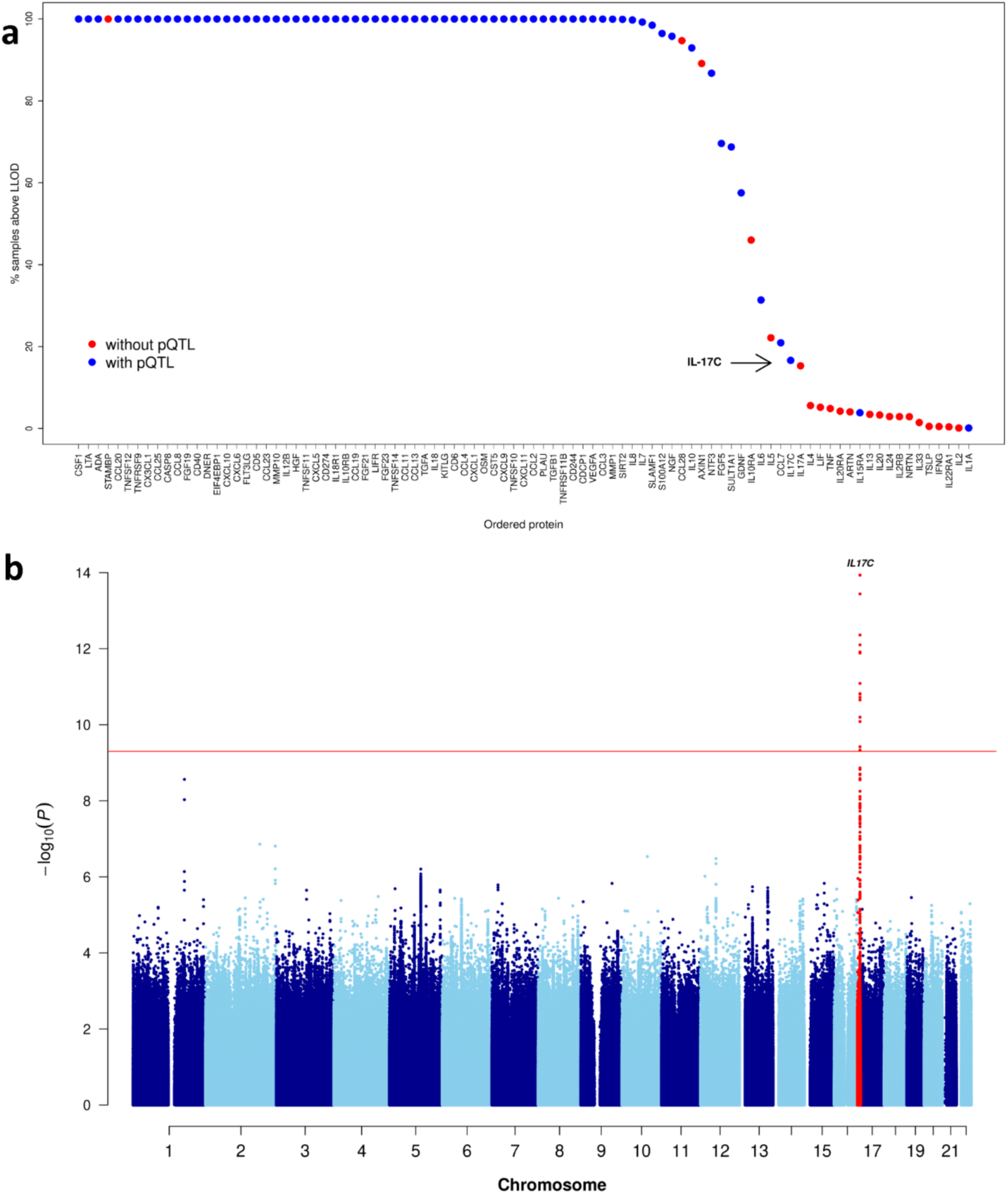
Plasma protein abundance and pQTL detection. **a)** Proteins with low abundance are more likely to have no detectable pQTL. Y-xxis percentage of samples above lower limit of detection for each protein, calculated using the INTERVAL data (n=4994) for which we had individual-level protein data available. Blue and red points indicate presence or absence of at least 1 significant pQTL in the GWAS meta-analysis, respectively. **b)** Manhattan plot for genetic associations with plasma IL17C, where the red horizontal line indicates the statistical significant threshold (5×10^-10^).

**Supplementary Figure 3.**
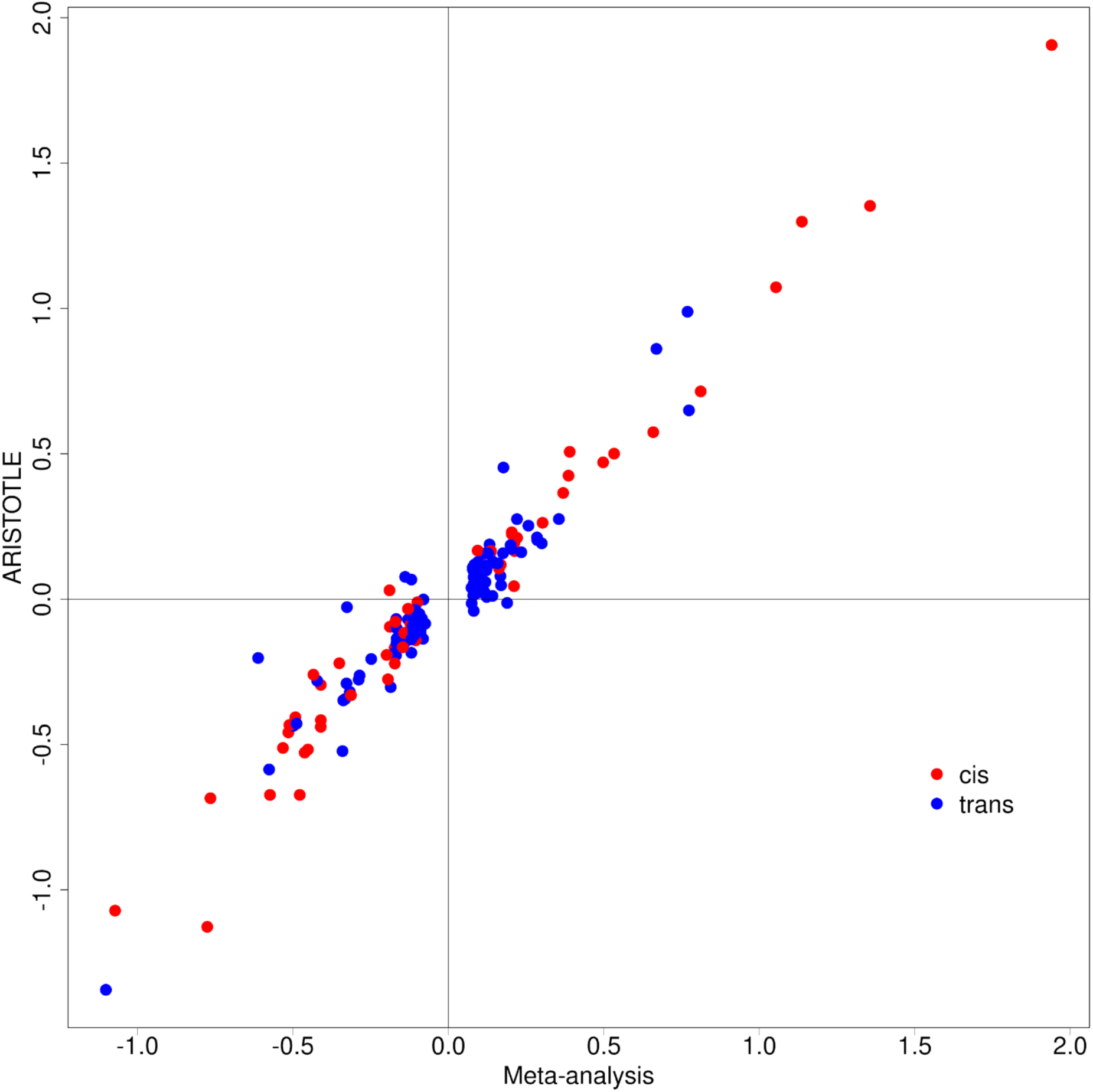
pQTL replication in the ARISTOTLE cohort. Comparison of effect sizes between pQTLs from the discovery pQTL meta-analysis (n=15,150) and the ARISTOTLE cohort (n=1,585). Each point represents a genetic variant that was a significant pQTL in the discovery meta-analysis. Red= cis, Blue= trans.

**Supplementary Figure 4.**
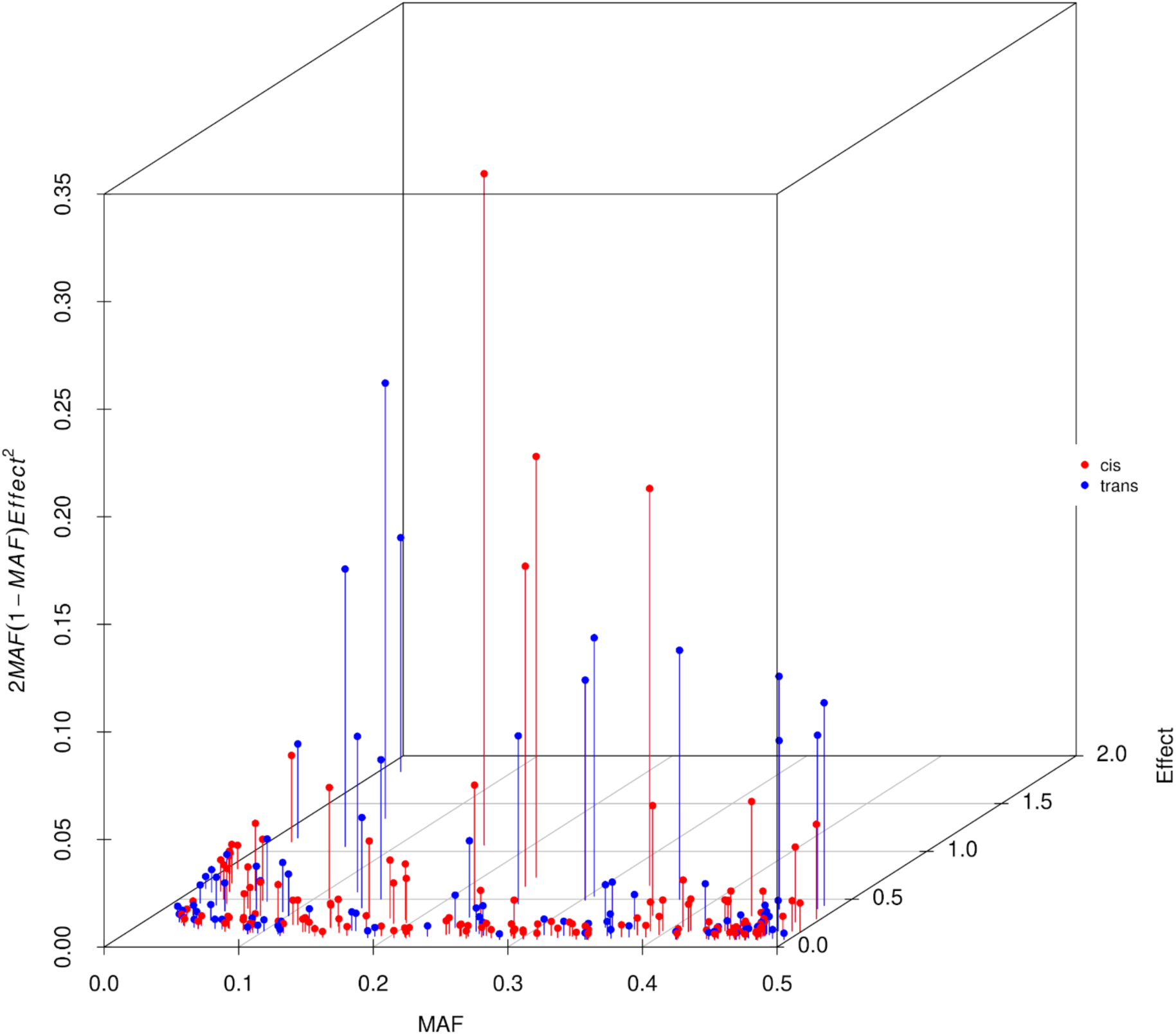
Relationship between allele frequency, pQTL effect size and proportion of variance explained. Effect size versus MAF versus the proportion of variance explained (2MAF(1-MAF)Effect^2^, red=cis, blue=trans) for 227 conditionally independent pQTLs.

**Supplementary Figure 5.**
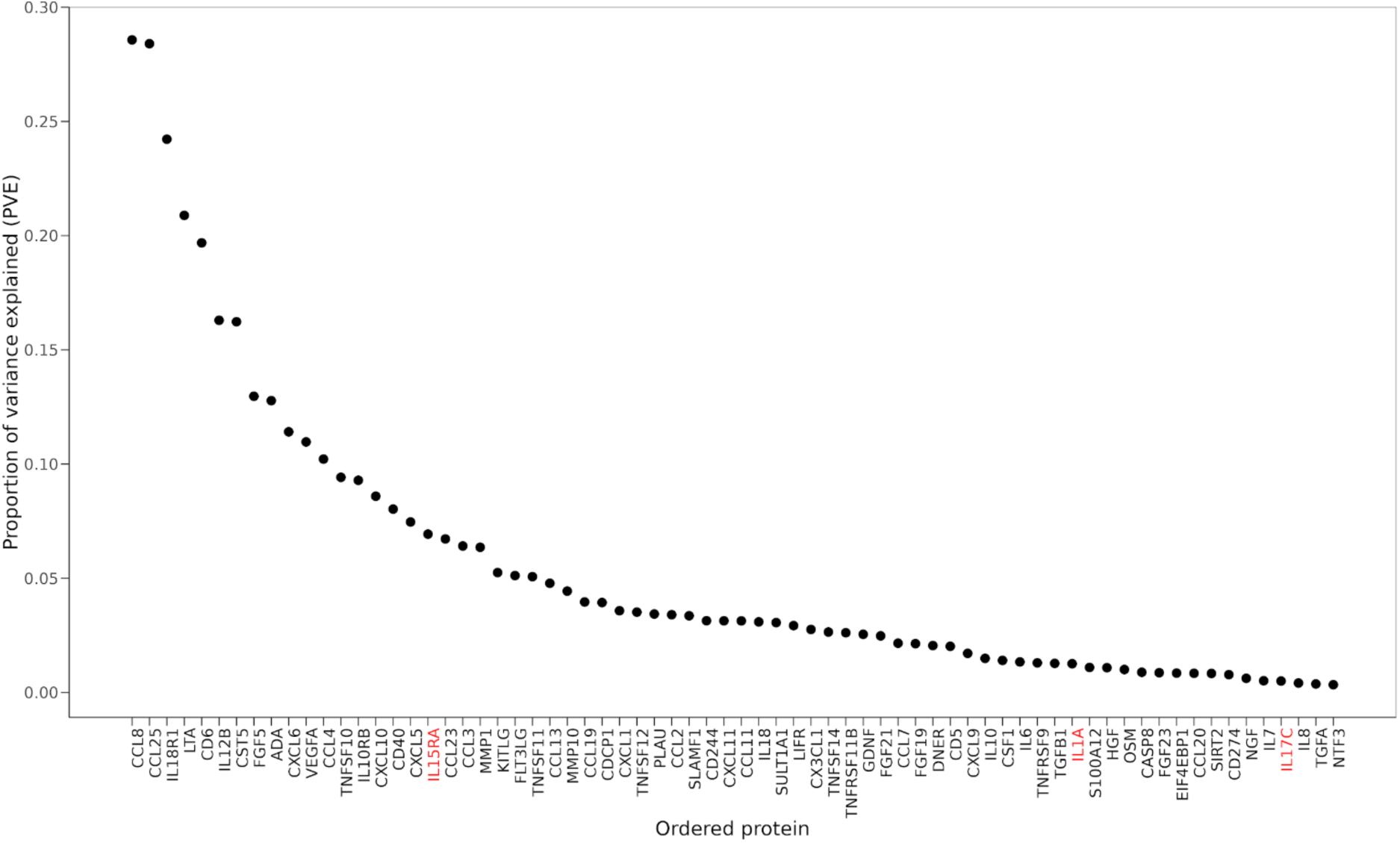
Proportion of variance explained. Proportion of variance explained (PVE) by the conditionally independent variants associated with each protein. Proteins are annotated using the gene symbol of their encoding genes. Protein names are colored in red if over 80% of samples have levels below the lower limit of detection in the INTERVAL dataset.

**Supplementary Figure 6.**
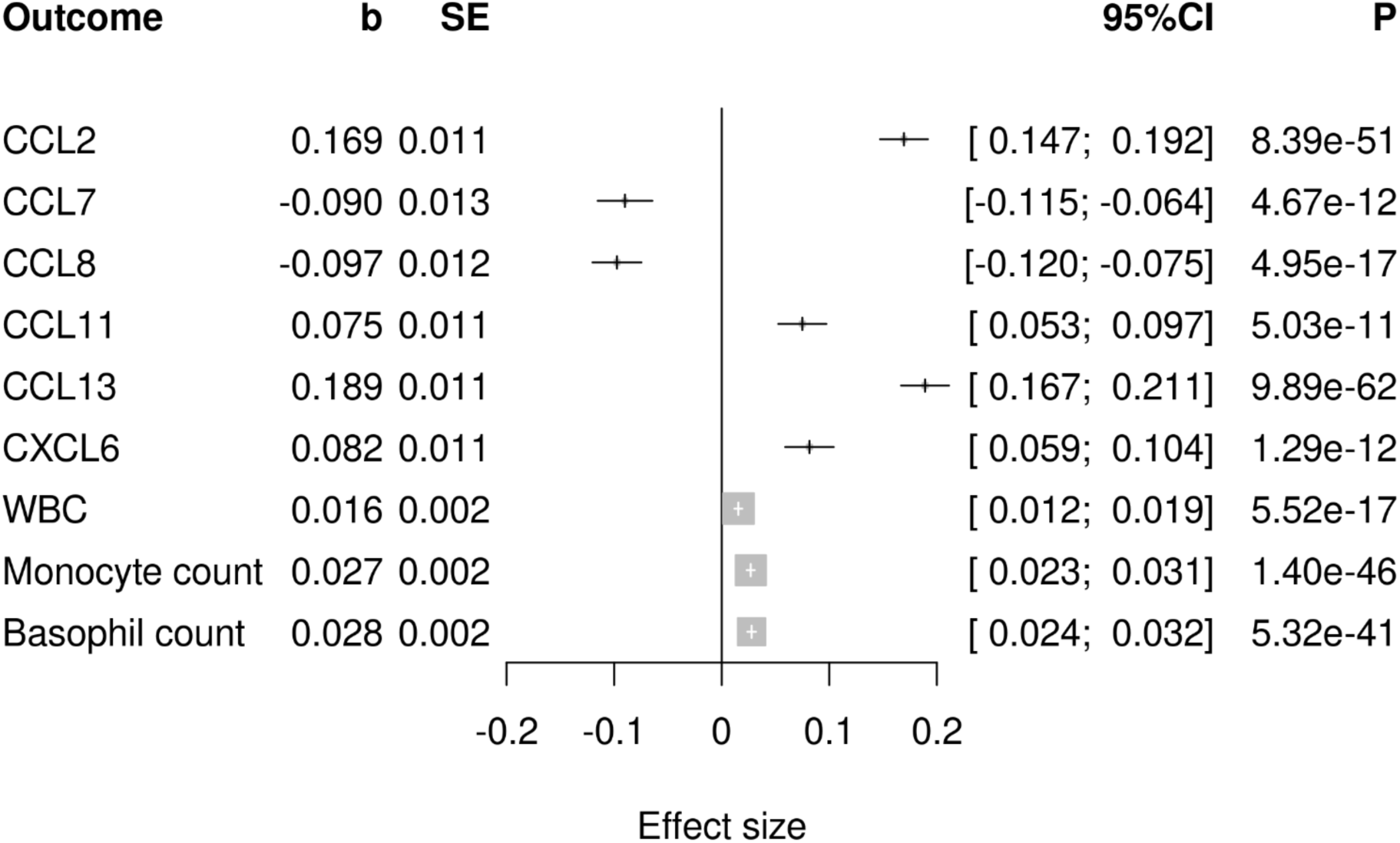
Chemokine *trans*-pQTL hotspot. Forest plot showing the associations for the pleiotropic *trans*-pQTL at rs12075 (GRCh37, 1:158175353-160525679) with plasma levels of chemokines and blood cell counts. WBC = white blood cell count. P = p-value, CI = confidence interval, b= beta (effect size). SE = standard error. Blood cell association data from Chen et al^31^.

**Supplementary Figure 7.**
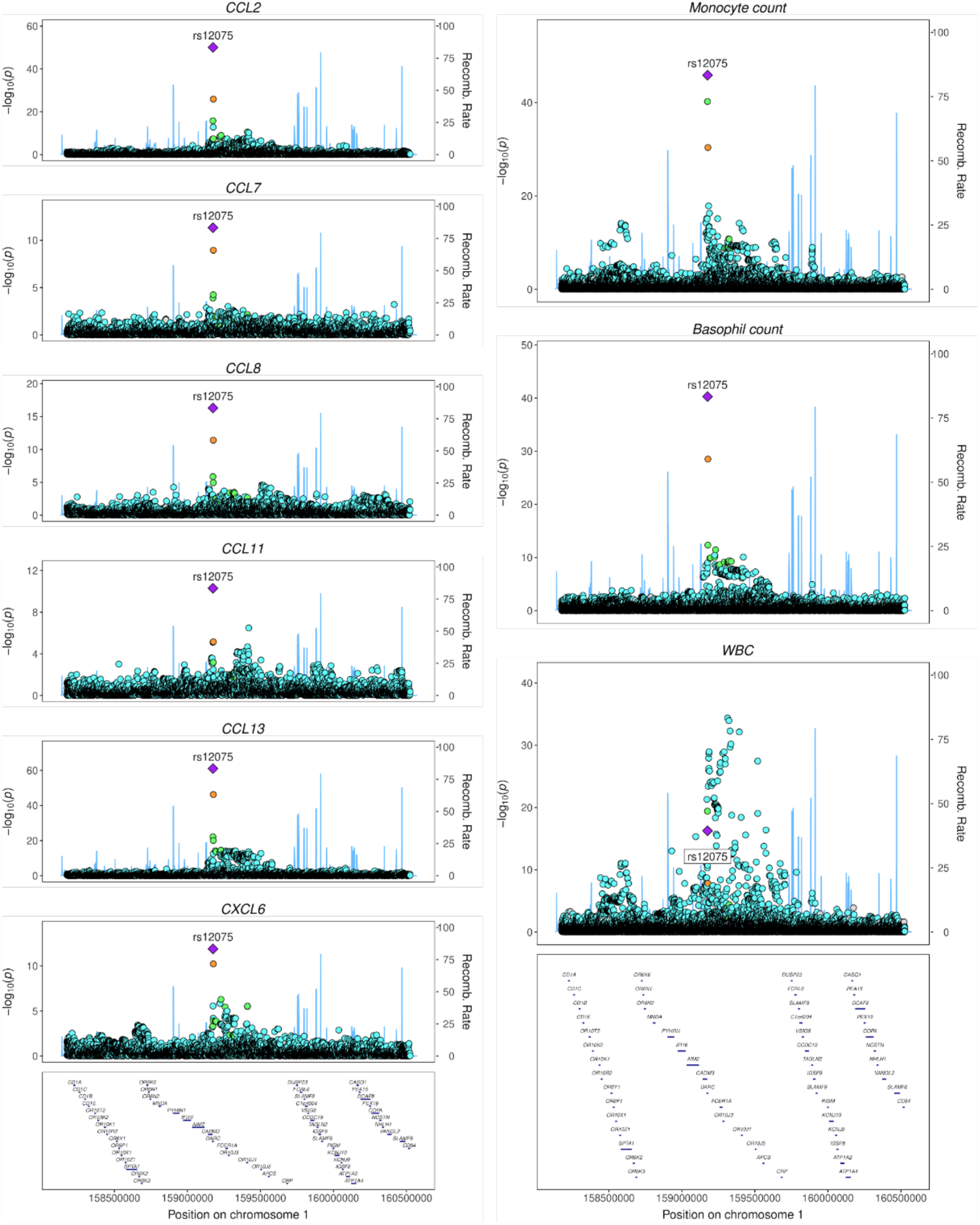
Colocalisation of pleiotropic chemokine trans-pQTL and blood cell count trait signals. Regional association plots in the region around rs12075 (GRCh37, 1:158175353-160525679). Left side: association with plasma chemokine levels. Right: associations with basophil, monocyte and white blood cell (WBC) counts using data from Chen et al^31^.

**Supplementary Figure 8.**
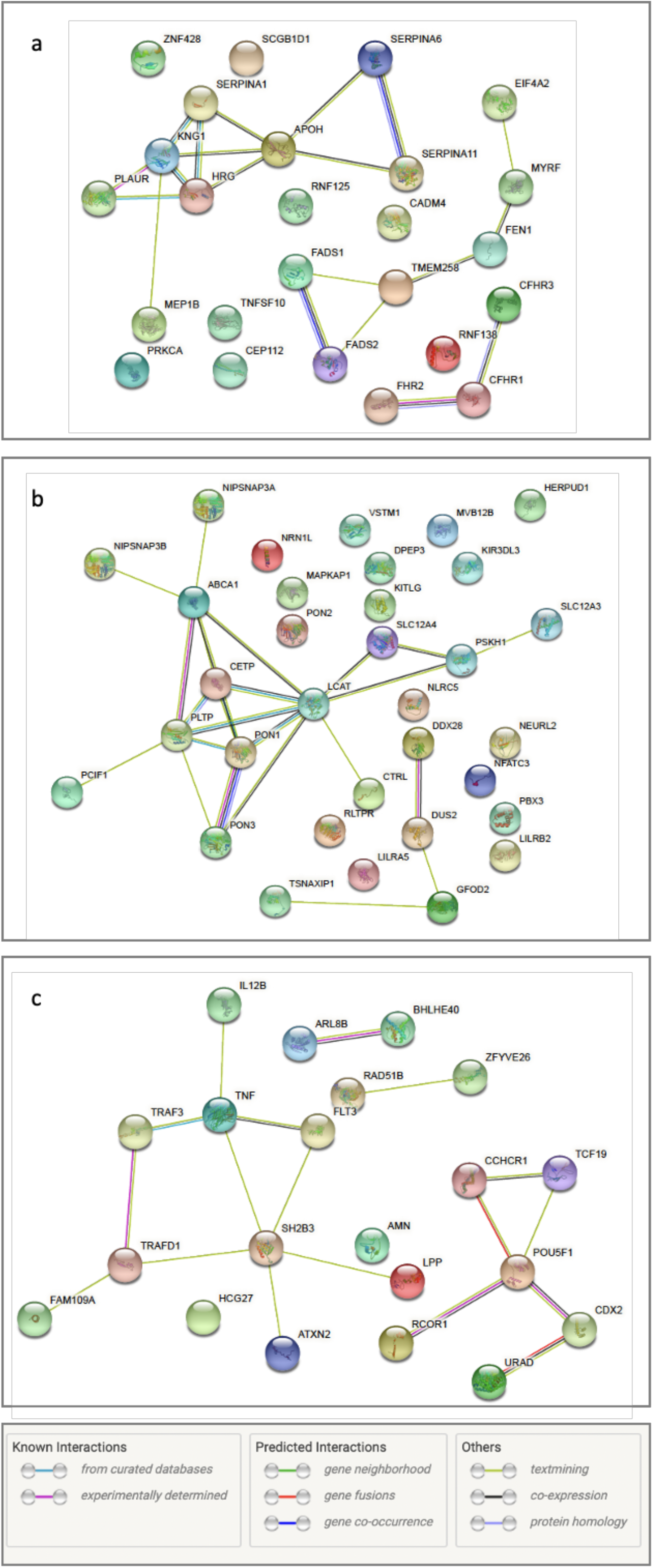
Interactions between the candidate mediators for multi-locus-regulated proteins. (**A**) TNFSF10 (TRAIL), (**B**) KITLG (KIT ligand, also known as stem cell factor), and (**C**) IL-12B. The graphs were generated using the STRINGdb (v11.5) webtool. The coloring of the edges indicates the type of evidence supporting an interaction, as shown in the legend above.

**Supplementary Figure 9.**
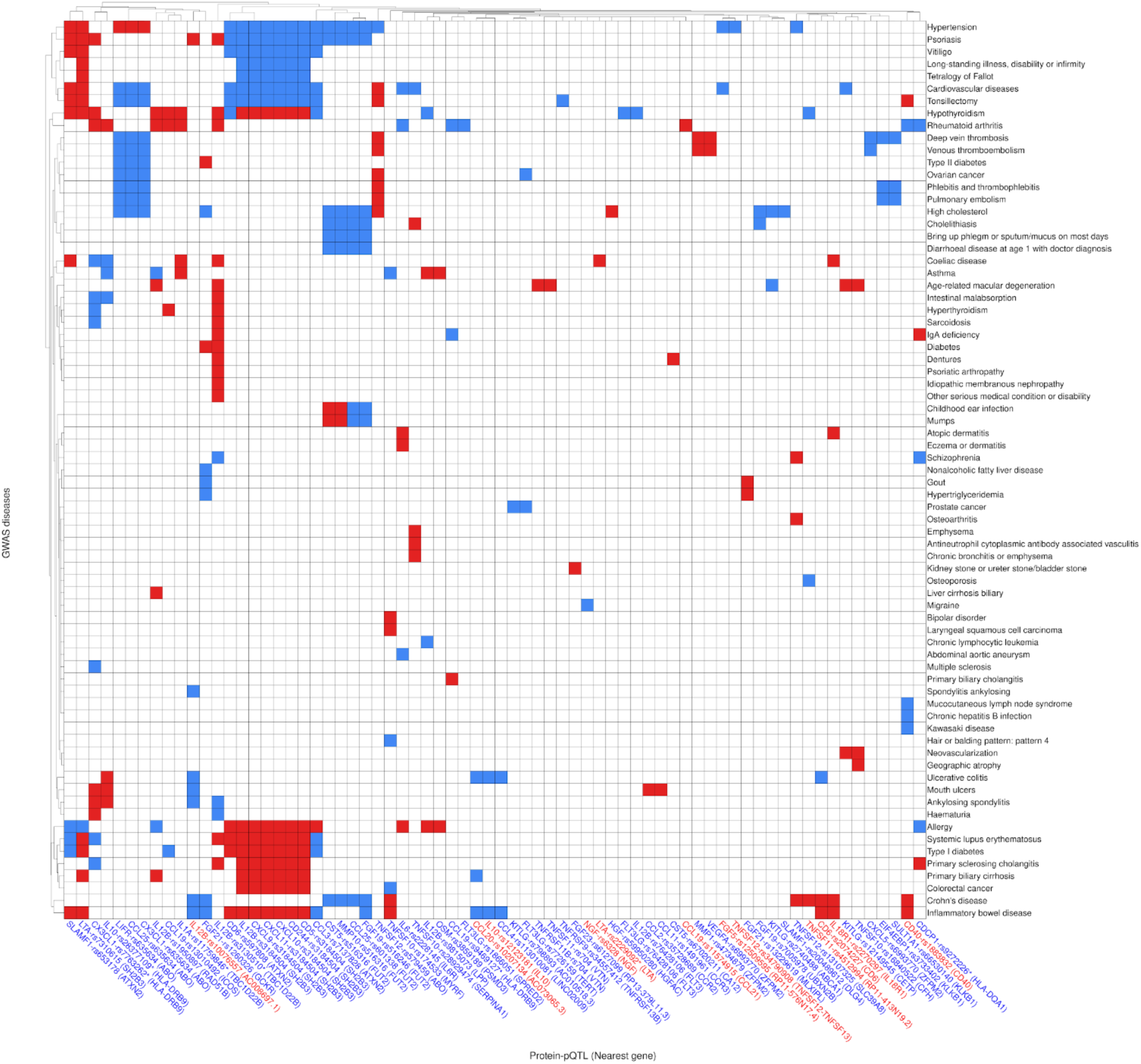
Protein-disease connections from overlap of pQTLs and disease GWAS. The protein and the corresponding pQTL sentinel variant are indicated in the format of protein-rsid. The nearest gene in the genomic region around the pQTL is shown in brackets. Asterix indicates the genetic variant lies in the HLA region. Red squares: genetic susceptibility to increased plasma levels of the protein is associated with increased disease risk. Blue squares: decreased disease risk.

**Supplementary Figure 10.**
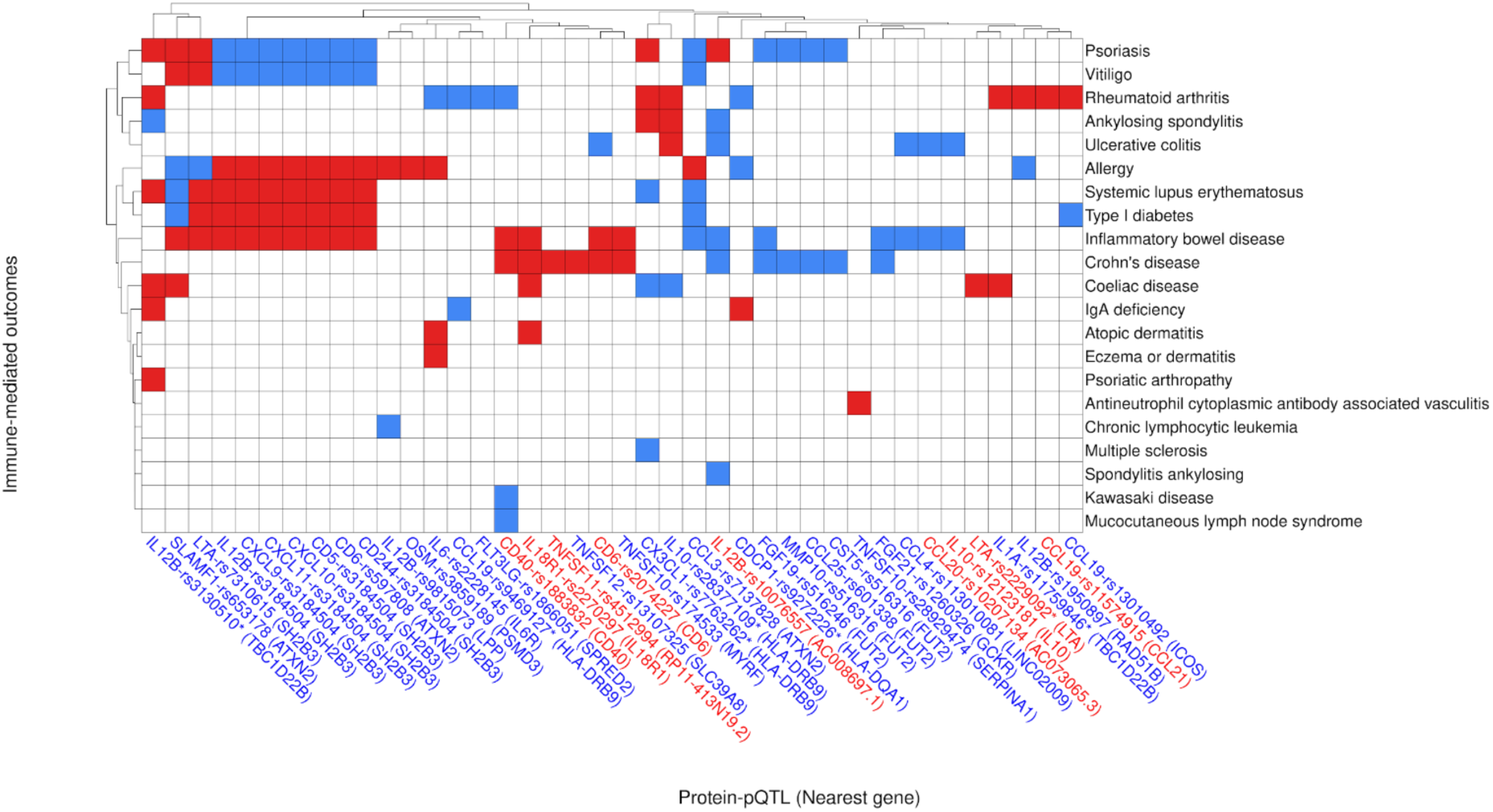
Protein and immune-mediated disease (IMD) connections from overlap of pQTLs and disease GWAS. The protein and the corresponding pQTL sentinel variant are indicated in the format of protein-rsid. The nearest gene in the genomic region around the pQTL is shown in brackets. Asterix indicates the genetic variant lies in the HLA region. Red squares: genetic susceptibility to increased plasma levels of the protein is associated with increased disease risk. Blue squares: decreased disease risk.

**Supplementary Figure 11.**
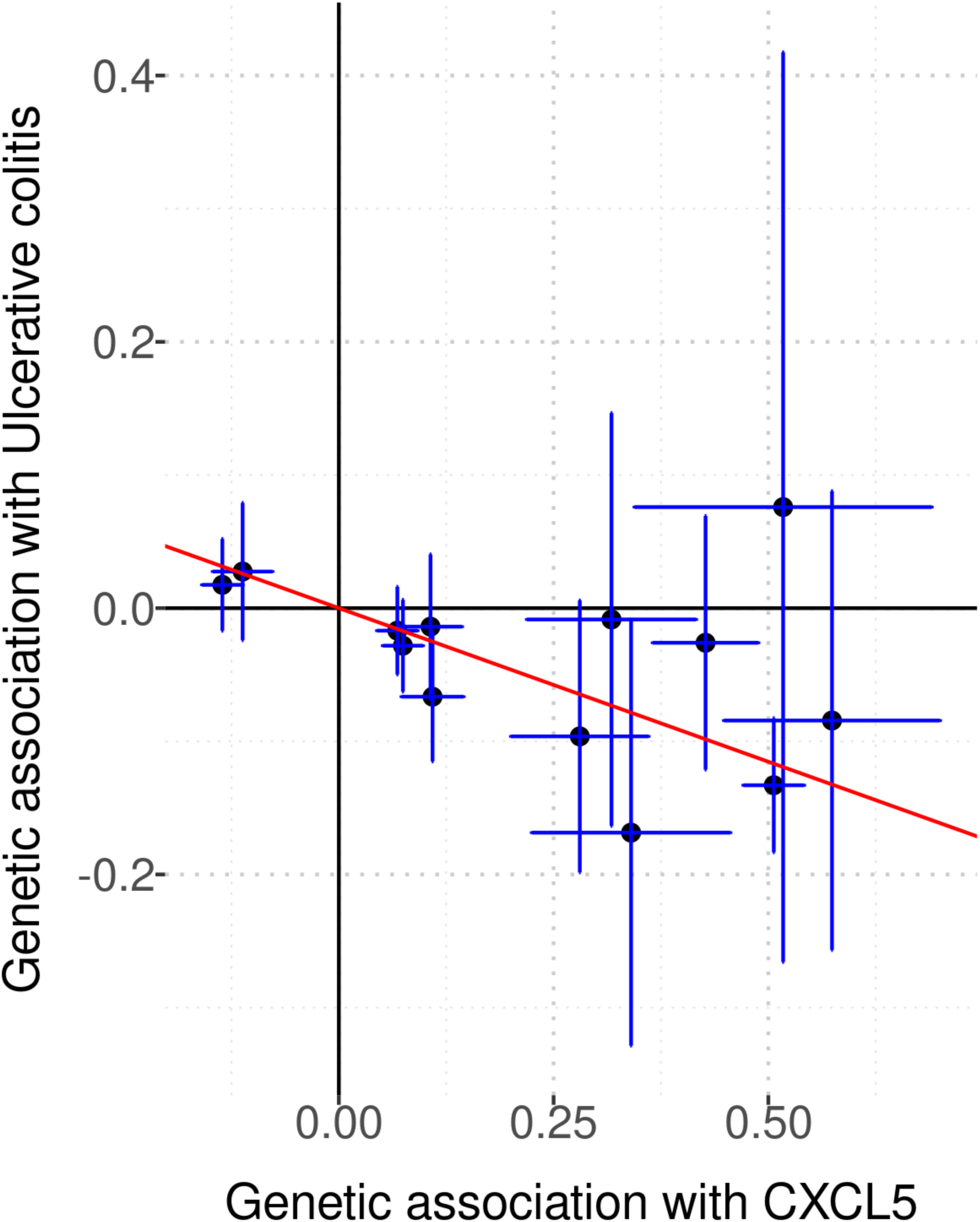
Mendelian randomisation analysis for CXCL5 and ulcerative colitis. Scatterplot showing the 13 variants used in the GSMR analysis assessing the effect of CXCL5 on ulcerative colitis (UC) risk from the GWAS by de Lange et al^119^. Each point represents a genetic variant, and indicates the effect size of the variant on CXCL5 levels versus UC risk (log odds ratio). Vertical and horizontal lines represent 95% confidence intervals.

**Supplementary Figure 12.**
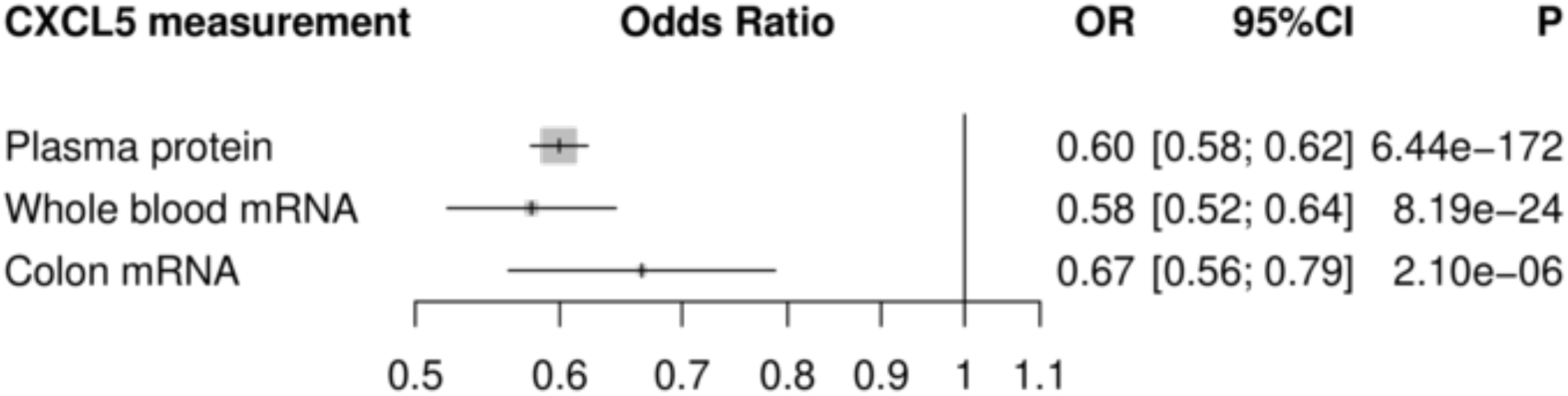
Directional concordance between CXCL5 pQTL and blood and colon tissue eQTLs. Forest plots showing effect size estimates for rs450373 pQTL in plasma (from our discovery meta-analysis) and eQTLs in whole blood and transverse colon tissue (GTEx v8 data). OR= odds ratio, calculated from beta estimate (representing the change in inverse-rank normalised plasma protein level in standard deviations associated with each copy of the effect allele). CI = confidence interval. P = p-value.

**Supplementary Figure 13.** Manhattan and Q-Q plots for the 91 proteins. *λ*_GC_ = lambdaGC, the genomic inflation factor. Proteins are labelled with both the label provided by Olink, and with the encoding gene symbol in parentheses. Pdf file provided separately.

**Supplementary Figure 14.** Forest plots and regional association plots for the 180 pQTLs. Left panels: forest plots show the effect estimate and 95% confidence intervals in each cohort. Metrics of heterogeneity are provided. Right panels: locuszoom plots of the regional associations. Proteins are labelled with both the label provided by Olink, and with the encoding gene symbol in parentheses. Pdf file provided separately.

**Supplementary Item. 3-dimensional interactive genomic map of pQTLs.** The html file shows pQTL sentinel variant position in relation to the gene encoding the target protein and the strength of the statistical association. Hover over a point to see detailed information. The image can be rotated by holding at left clicking the mouse.

