## Supplementary Figure 13 for "Mapping pQTLs of circulating inflammatory proteins identifies drivers of immune-related disease risk and novel therapeutic targets"

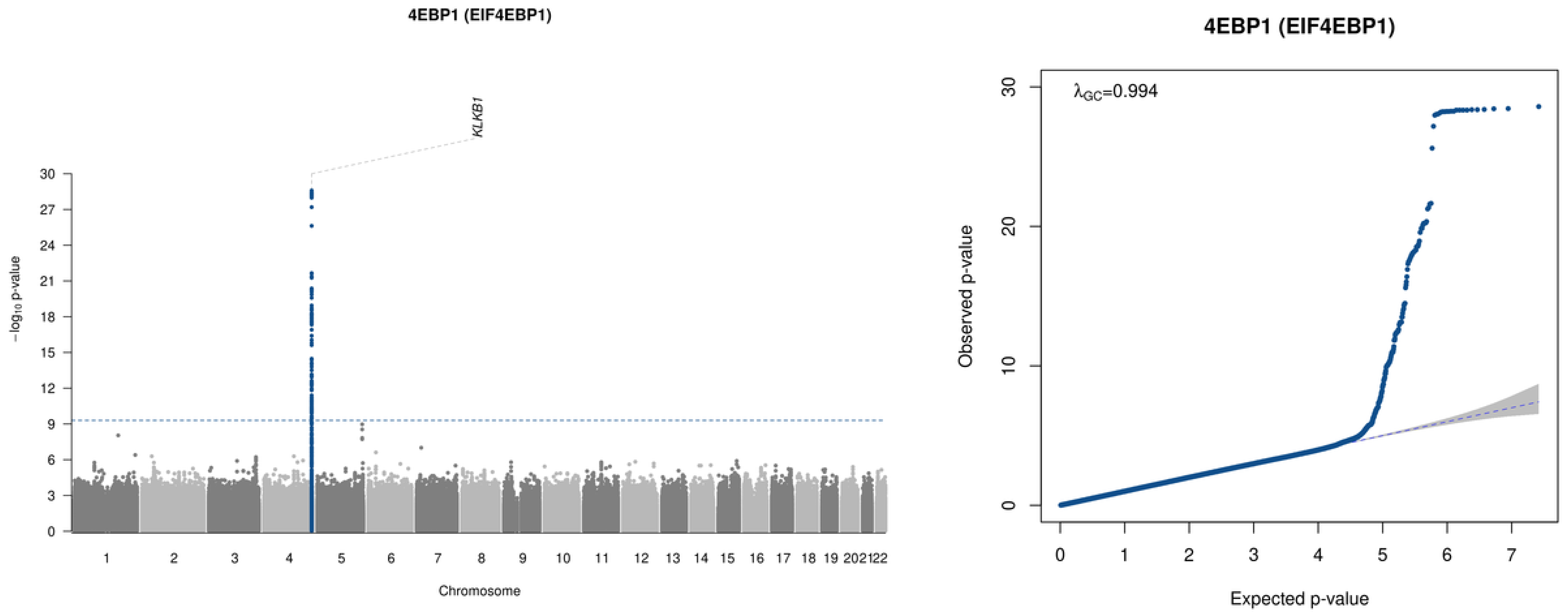
4EBP1 (EIF4EBP1)
KLKB1
−log10 p-value
30
27
24
21
18
15
12
9
6
3
202122
Chromosome
4EBP1 (EIF4EBP1)
λGC=0.994
Observed p-value
Expected p-value
0
10
20
30
0
1
2
3
4
5
6
7

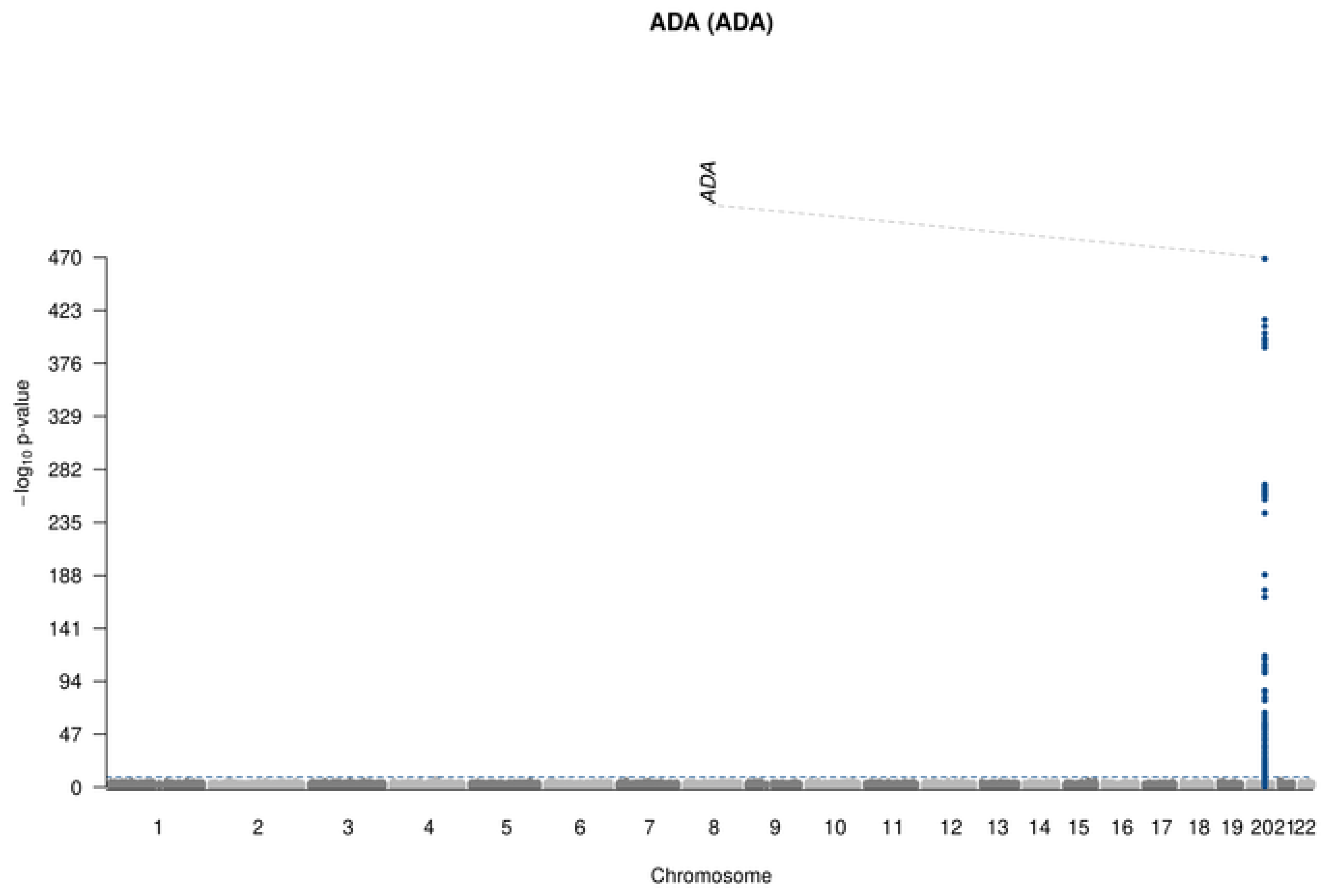

ADA (ADA)
ADA
470
423
376
329
282
235
188
141
94
47
0
−log10 p-value
Chromosome

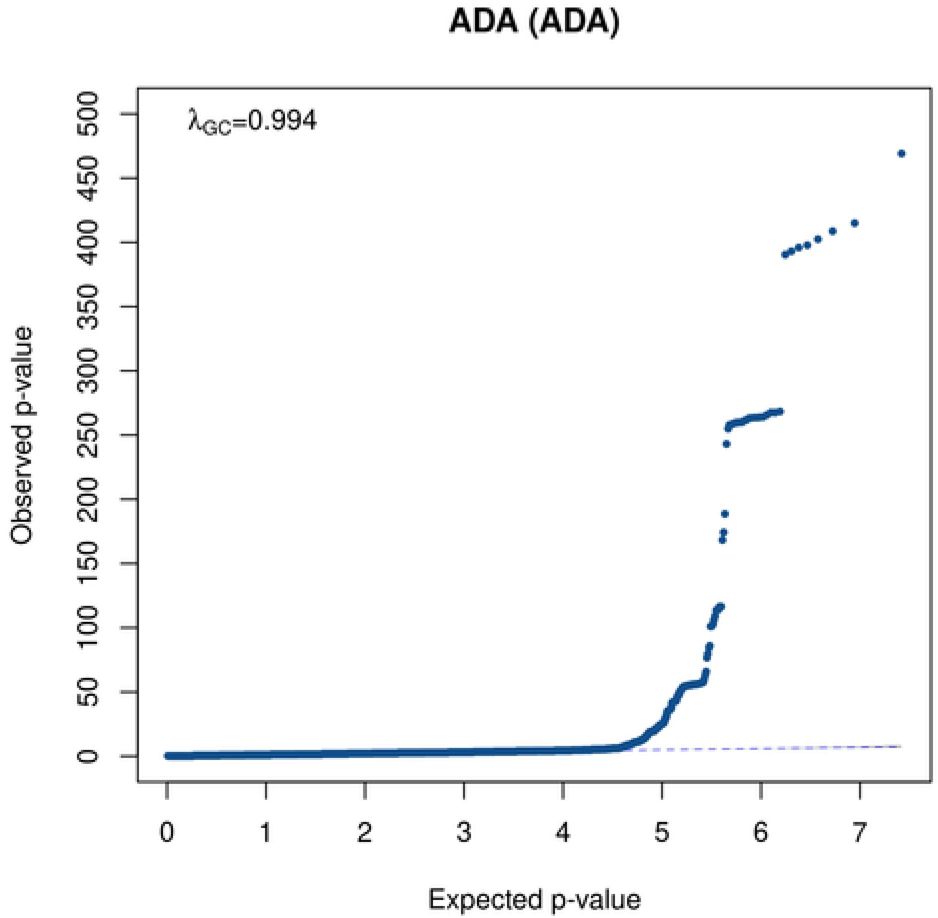

ADA (ADA)
λGC=0.994
500
450
400
350
300
250
200
150
100
50
0
Observed p-value
0
1
2
3
4
5
6
7
Expected p-value

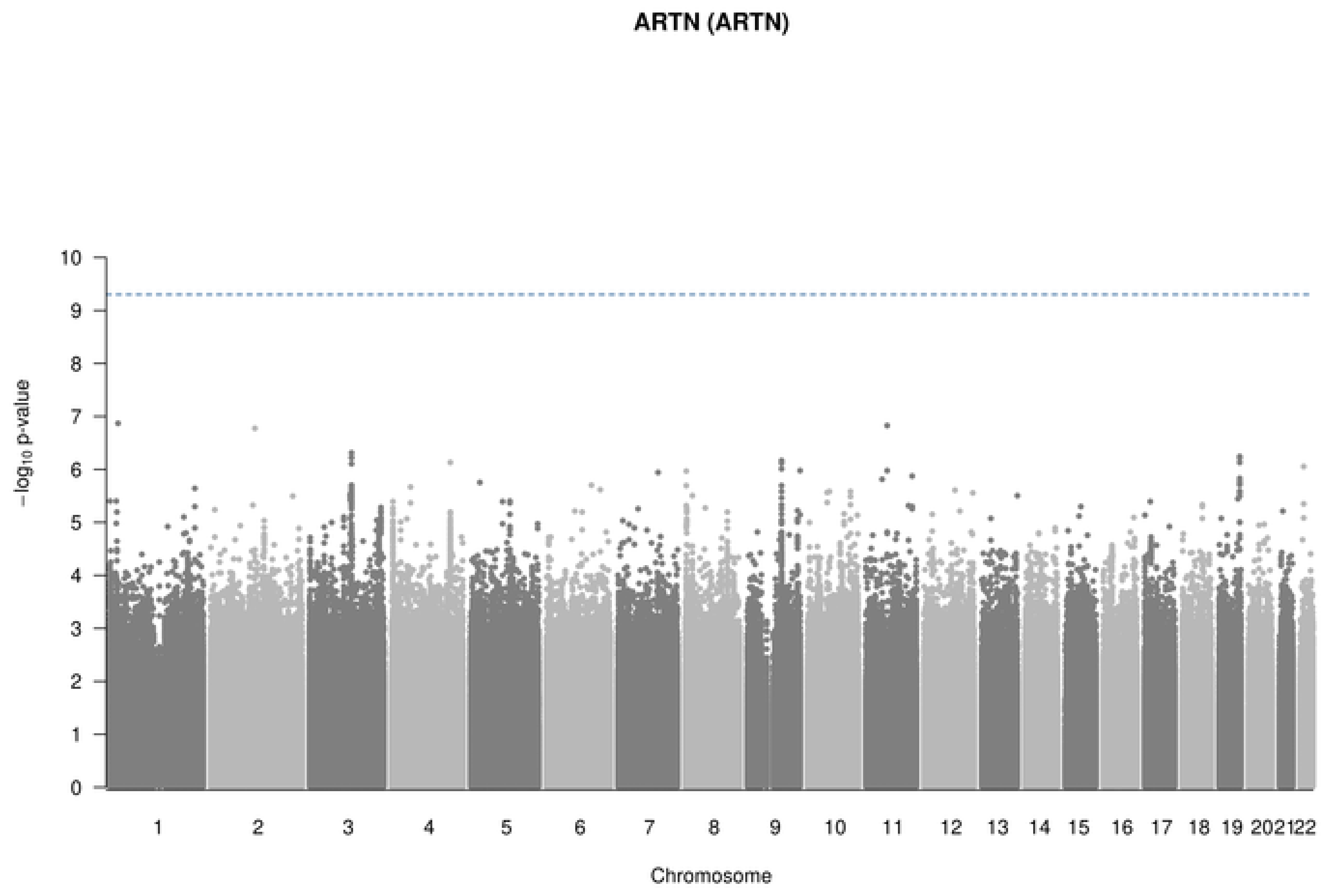
ARTN (ARTN)
−log10 p-value
0
1
2
3
4
5
6
7
8
9
10
202122
Chromosome

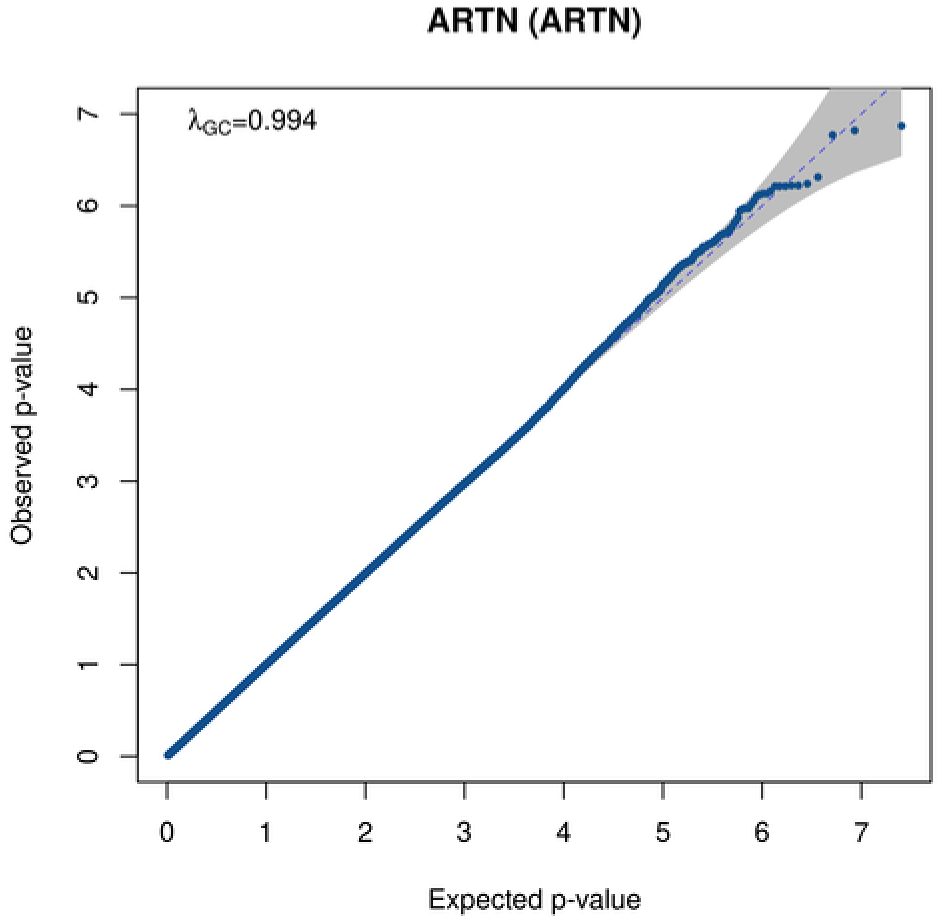
ARTN (ARTN)
λGC=0.994
Observed p-value
Expected p-value
0
1
2
3
4
5
6
7

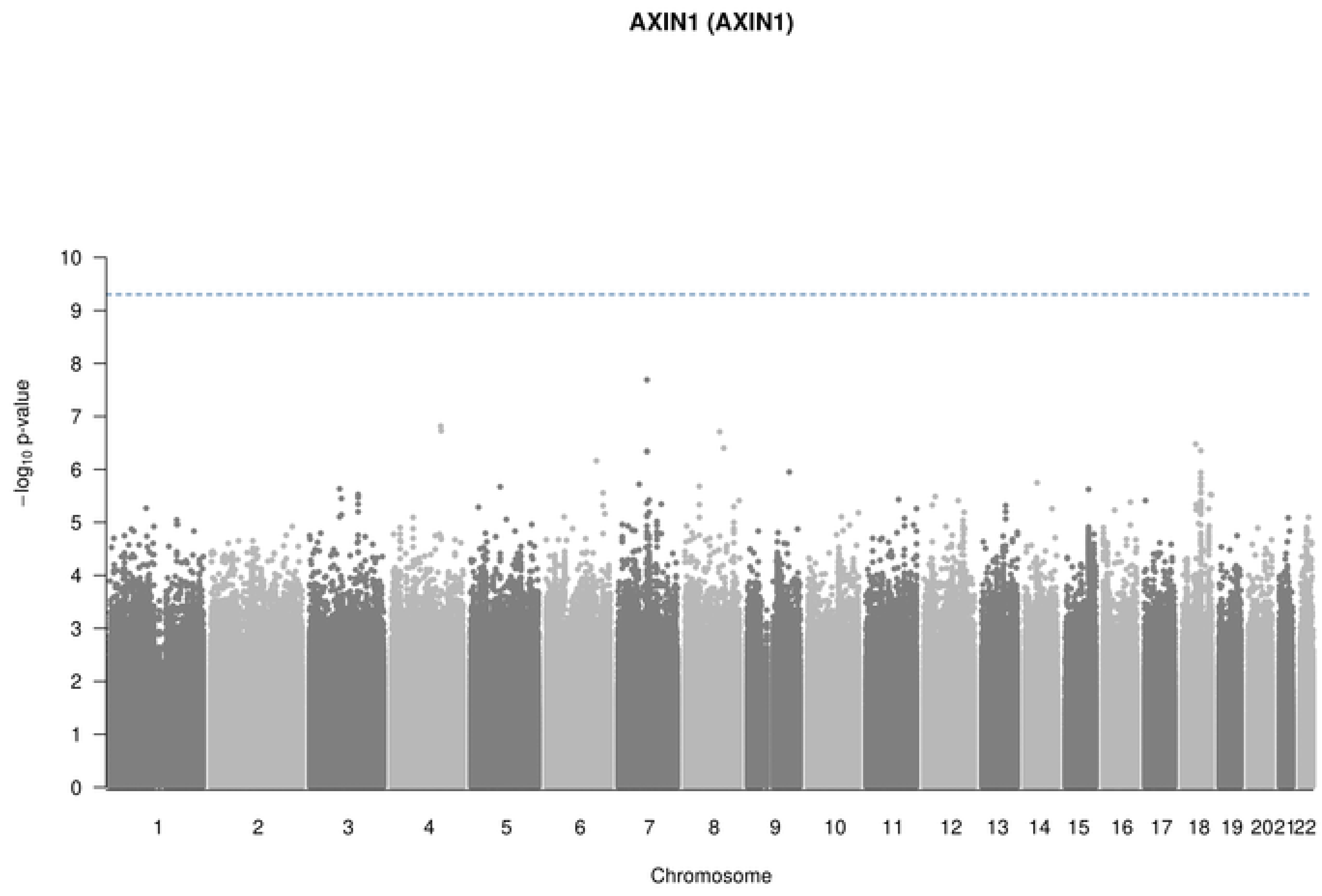

AXIN1 (AXIN1)
−log10 p-value
Chromosome

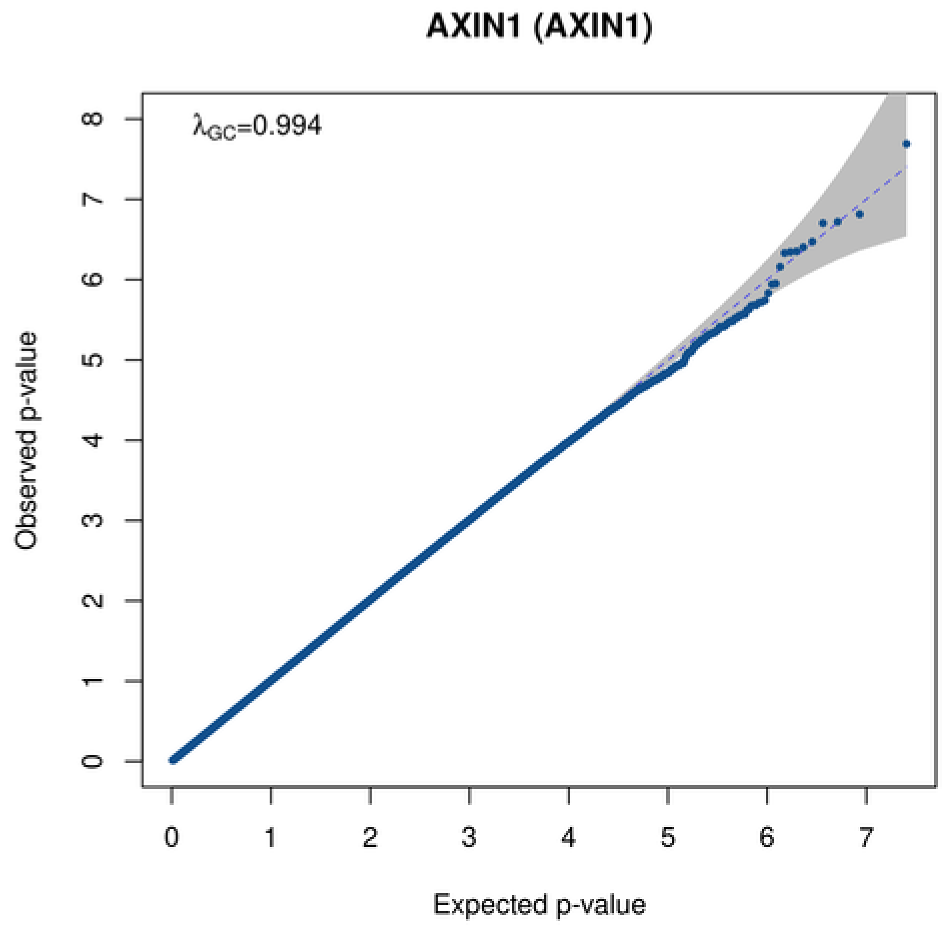

AXIN1 (AXIN1)
λGC=0.994
Observed p-value
Expected p-value

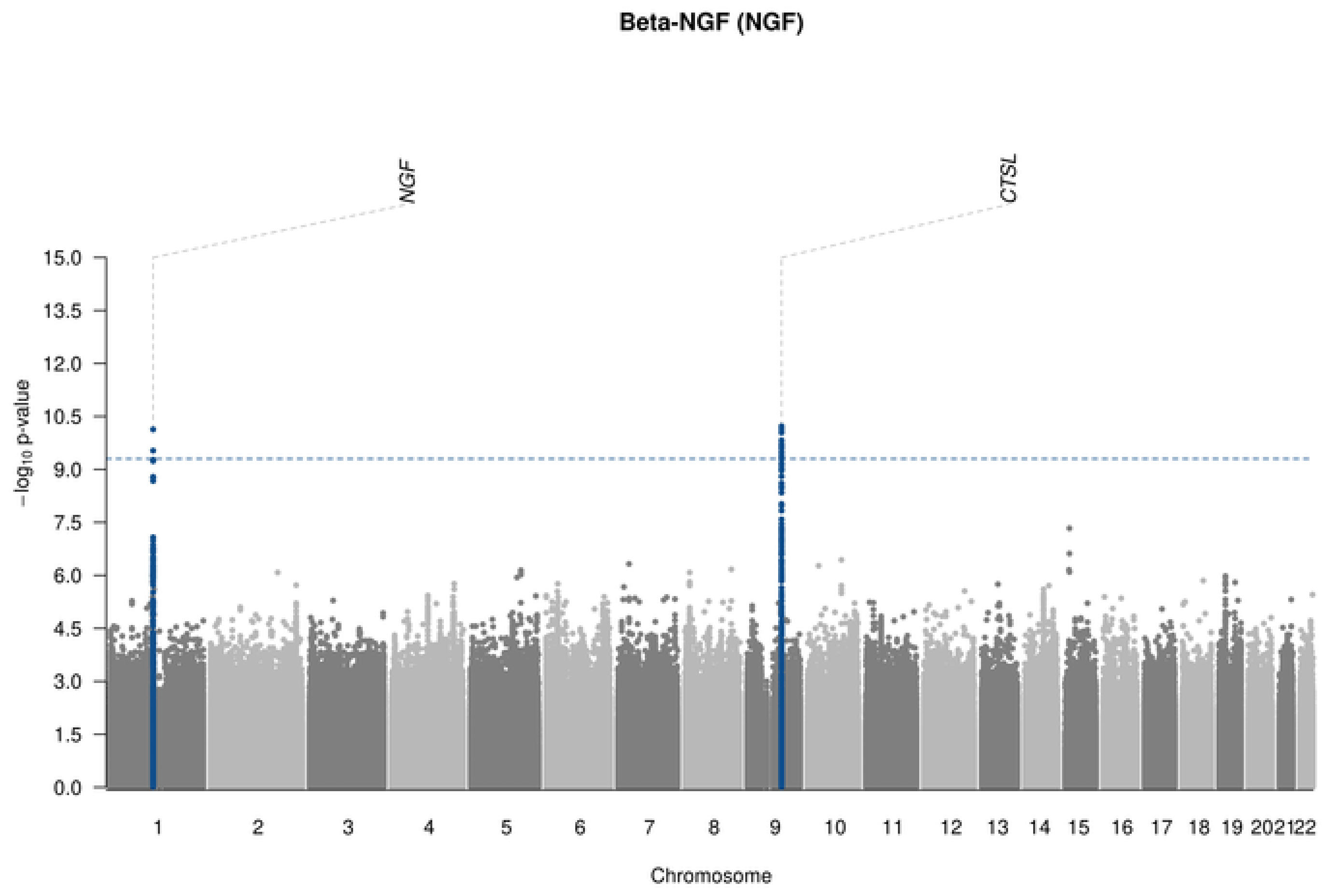

Beta-NGF (NGF)
NGF
CTSL
15.0
13.5
12.0
10.5
9.0
7.5
6.0
4.5
3.0
1.5
0.0
−log10 p-value
1 2 3 4 5 6 7 8 9 10 11 12 13 14 15 16 17 18 19 202122
Chromosome

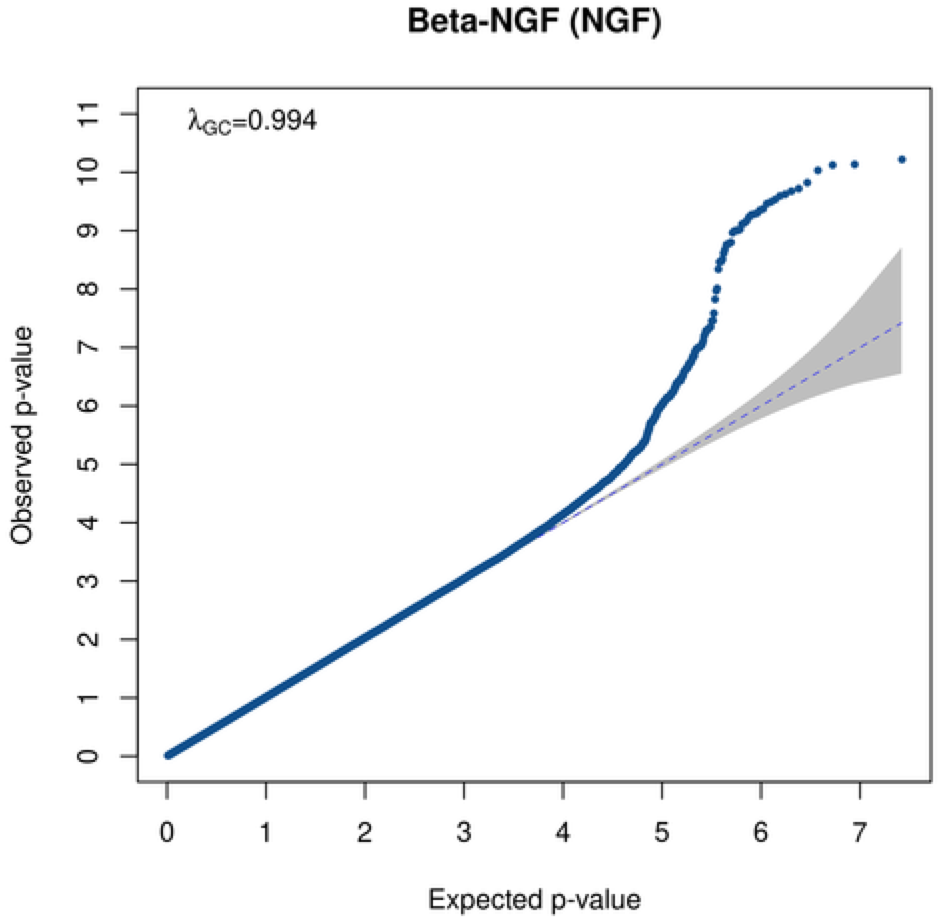

Beta-NGF (NGF)
λGC=0.994
Observed p-value
0 1 2 3 4 5 6 7 8 9 10 11
0 1 2 3 4 5 6 7
Expected p-value

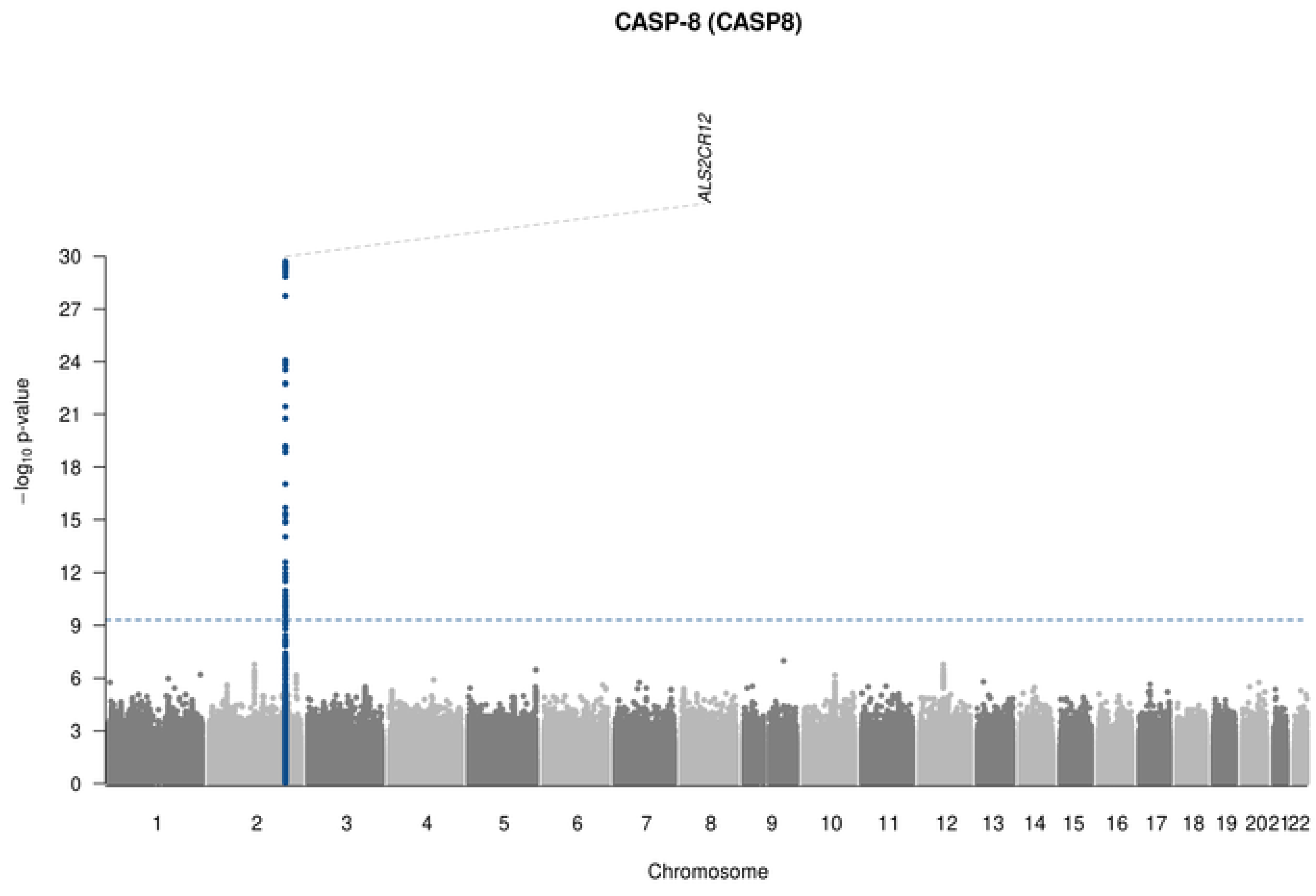

CASP-8 (CASP8)
ALS2CR12
−log10 p-value
Chromosome
202122

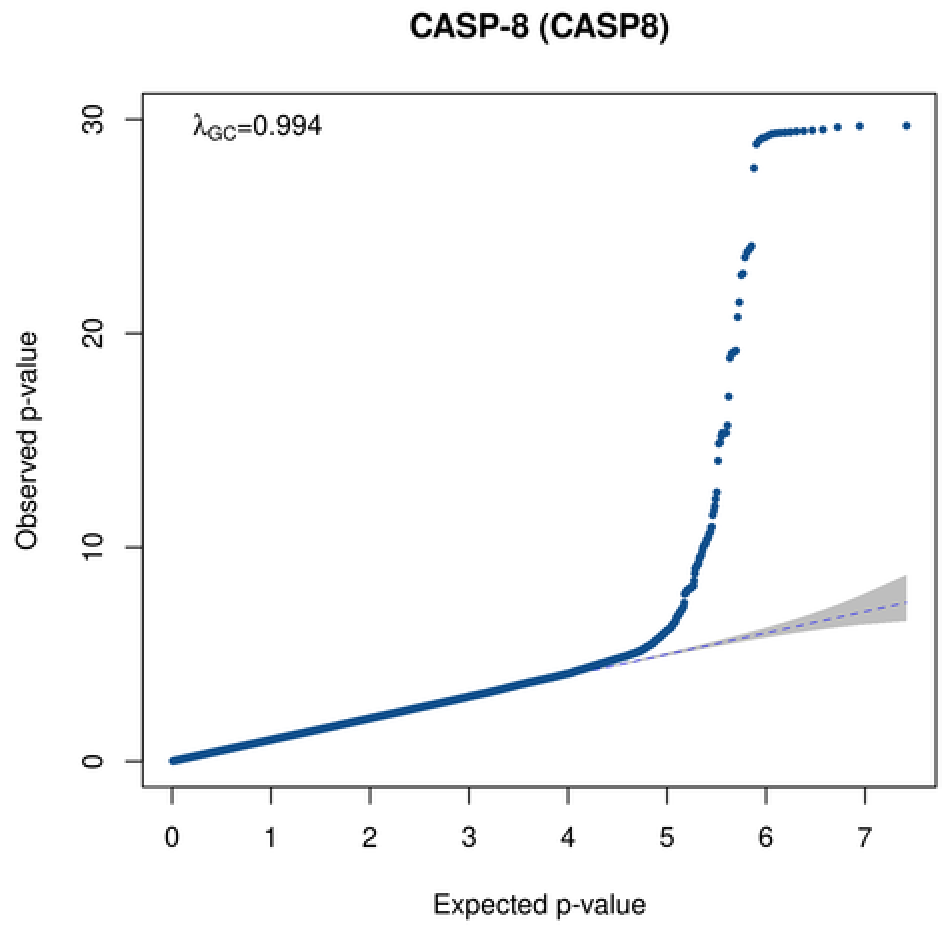

CASP-8 (CASP8)
λGC=0.994
Observed p-value
Expected p-value

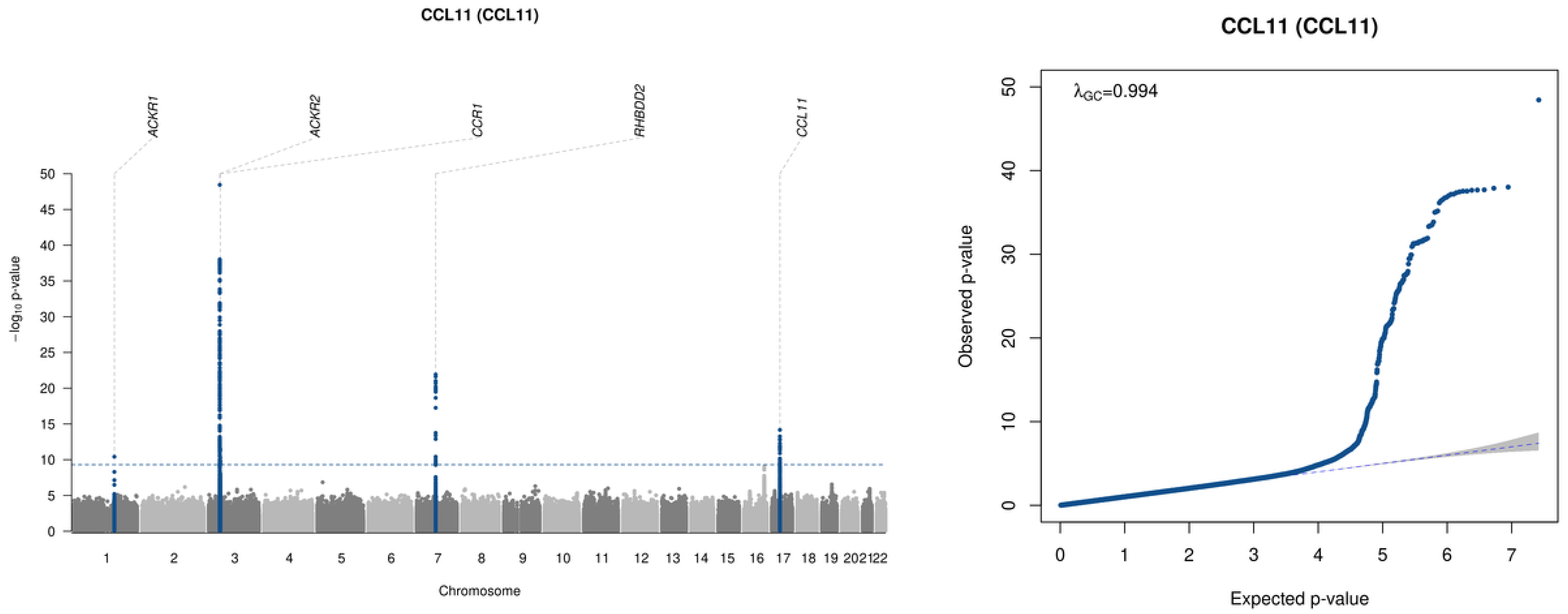
CCL11 (CCL11)
ACKR1
ACKR2
CCR1
RHBDD2
CCL11
−log10 p-value
Chromosome
CCL11 (CCL11)
λGC=0.994
Observed p-value
Expected p-value

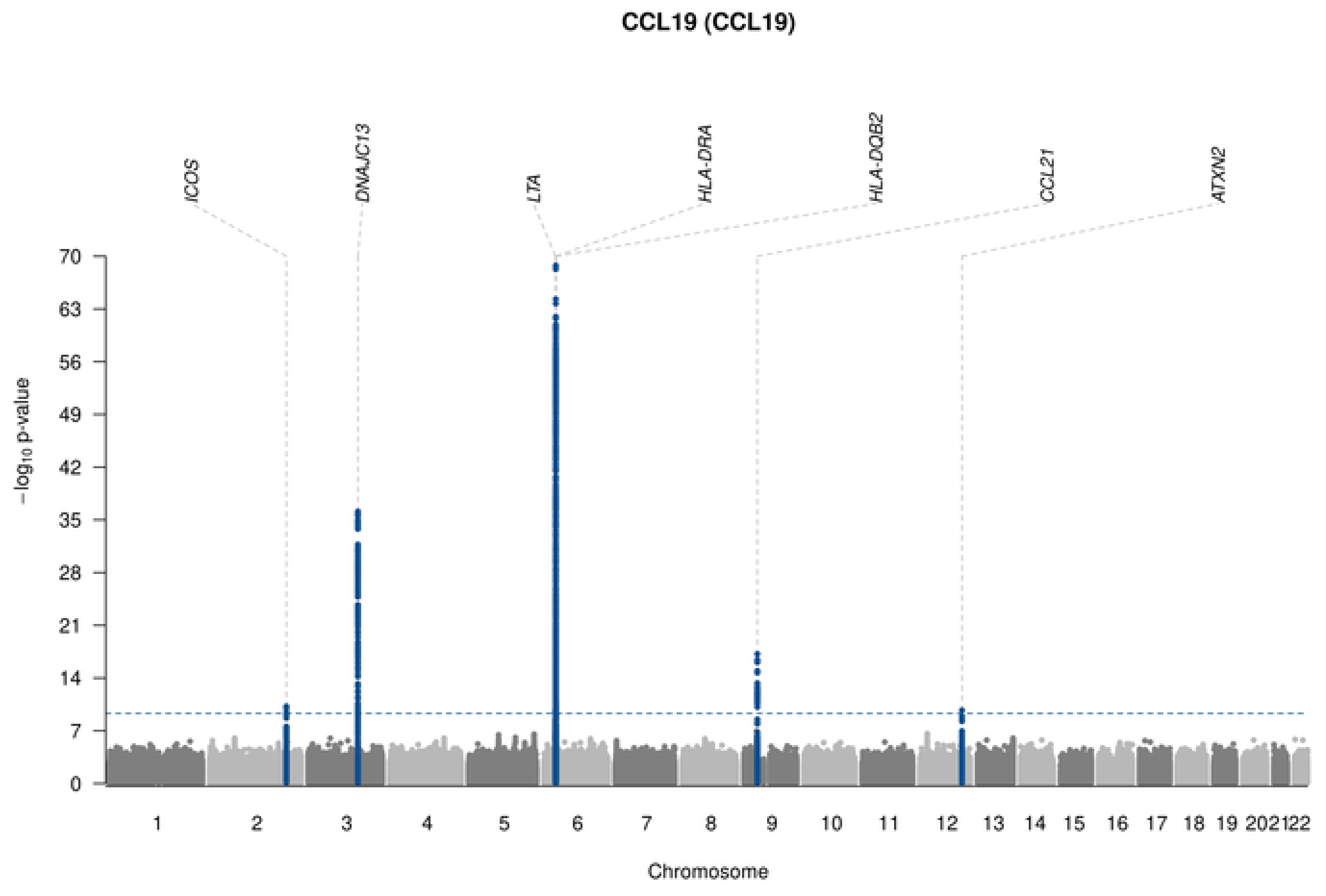
CCL19 (CCL19)
ICOS
DNAJC13
LTA
HLA-DRA
HLA-DQB2
CCL21
ATXN2
70
63
56
49
42
35
28
21
14
7
0
−log10 p-value
Chromosome

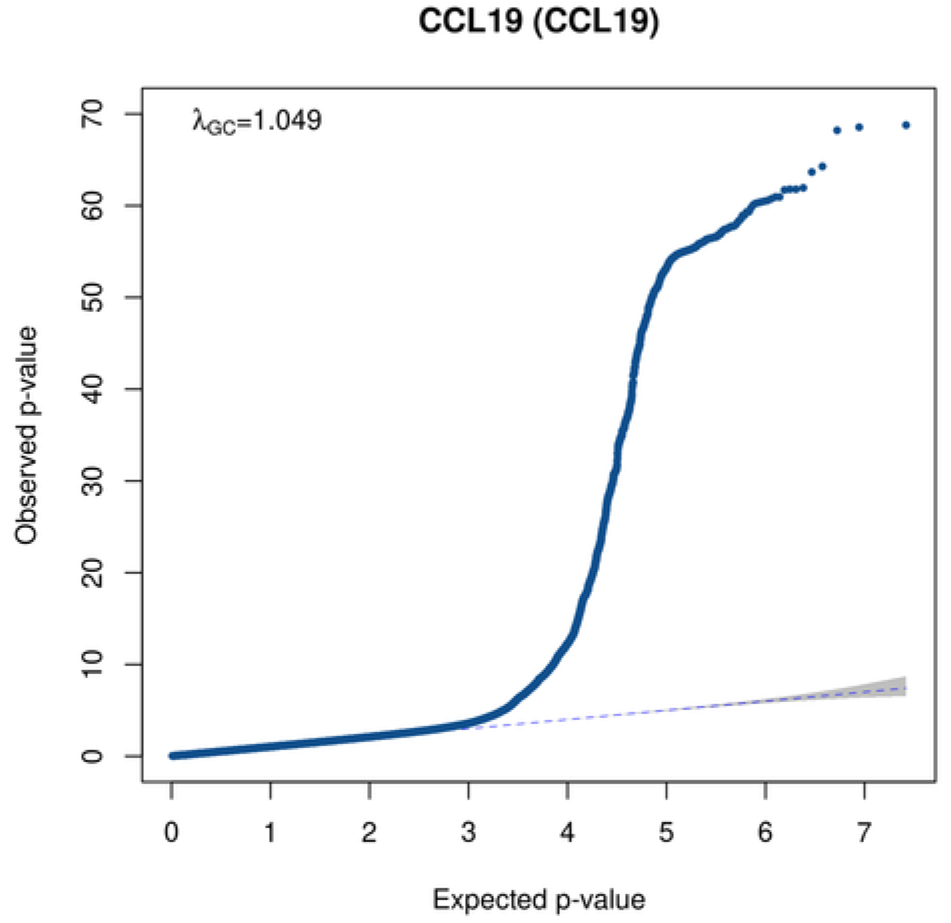
CCL19 (CCL19)
λGC=1.049
70
60
50
40
30
20
10
0
Observed p-value
0
1
2
3
4
5
6
7
Expected p-value

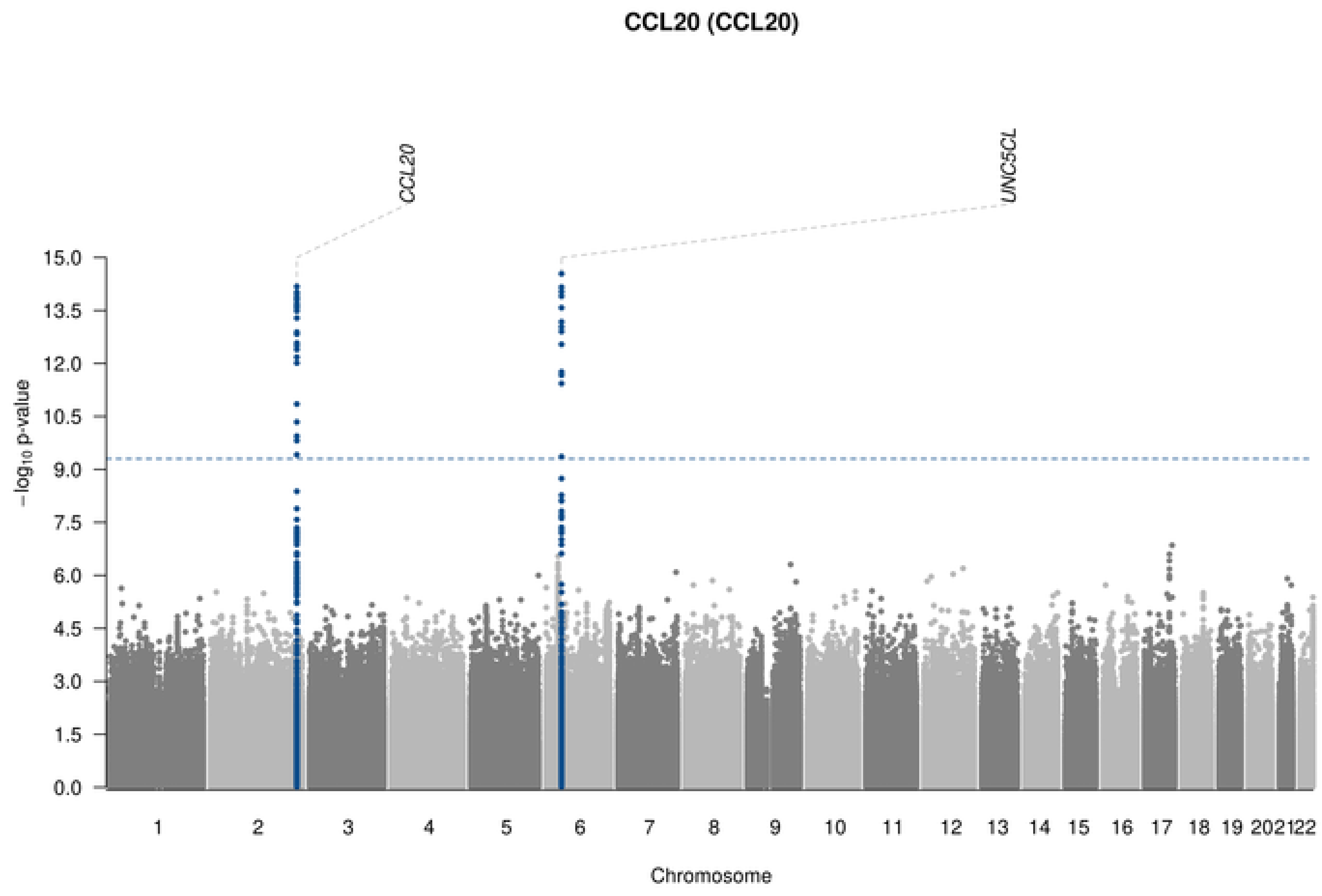

CCL20 (CCL20)
CCL20
UNC5CL
$-\log_{10}$ p-value
15.0
13.5
12.0
10.5
9.0
7.5
6.0
4.5
3.0
1.5
0.0
1 2 3 4 5 6 7 8 9 10 11 12 13 14 15 16 17 18 19 202122
Chromosome

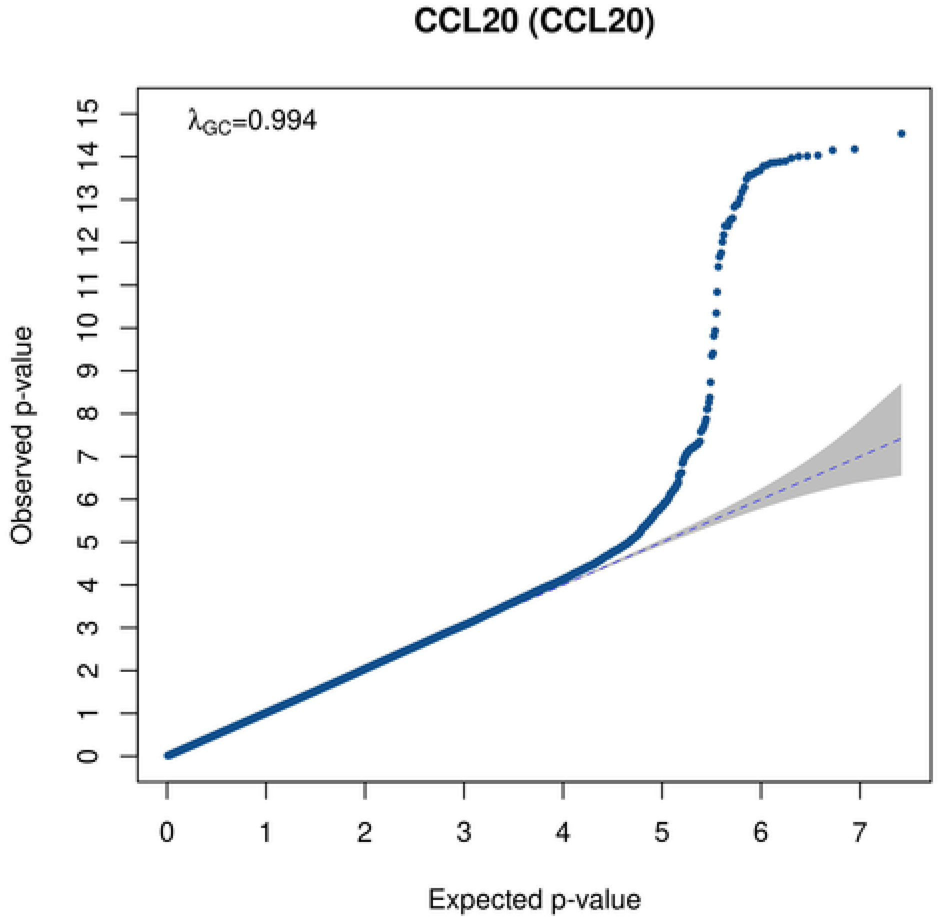

CCL20 (CCL20)
$\lambda_{GC}$=0.994
Observed p-value
0 1 2 3 4 5 6 7 8 9 10 11 12 13 14 15
0 1 2 3 4 5 6 7
Expected p-value

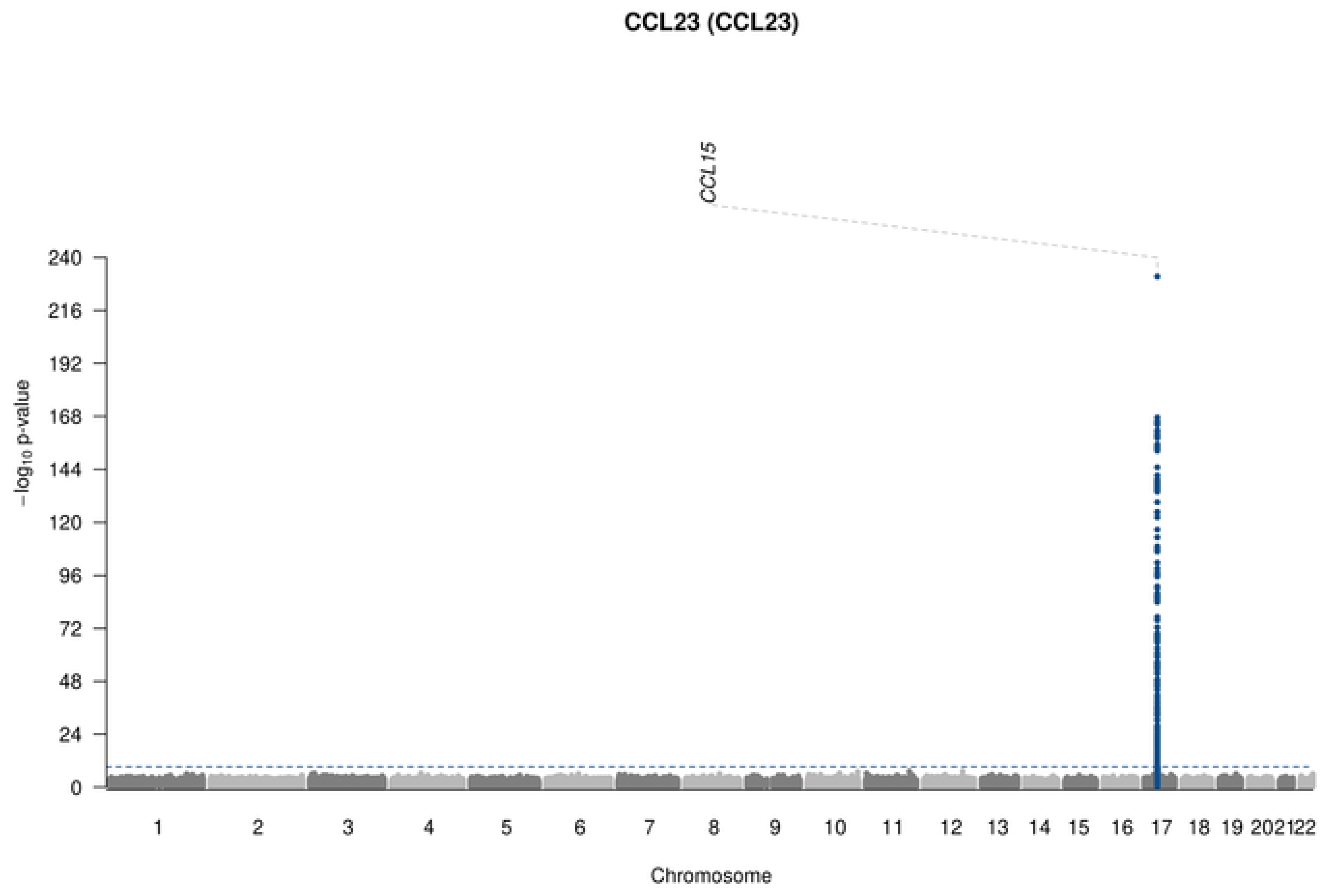
CCL23 (CCL23)
CCL15
−log10 p-value
240
216
192
168
144
120
96
72
48
24
Chromosome

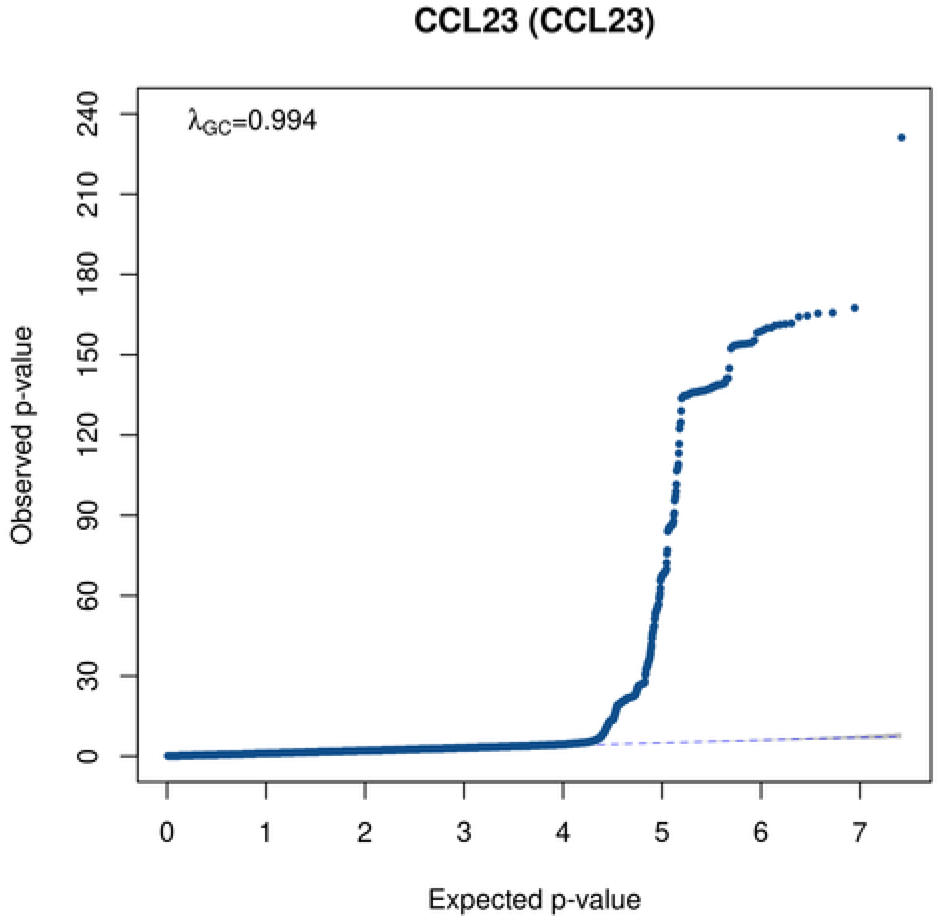
CCL23 (CCL23)
λGC=0.994
Observed p-value
240
210
180
150
120
90
60
30
0
0
1
2
3
4
5
6
7
Expected p-value

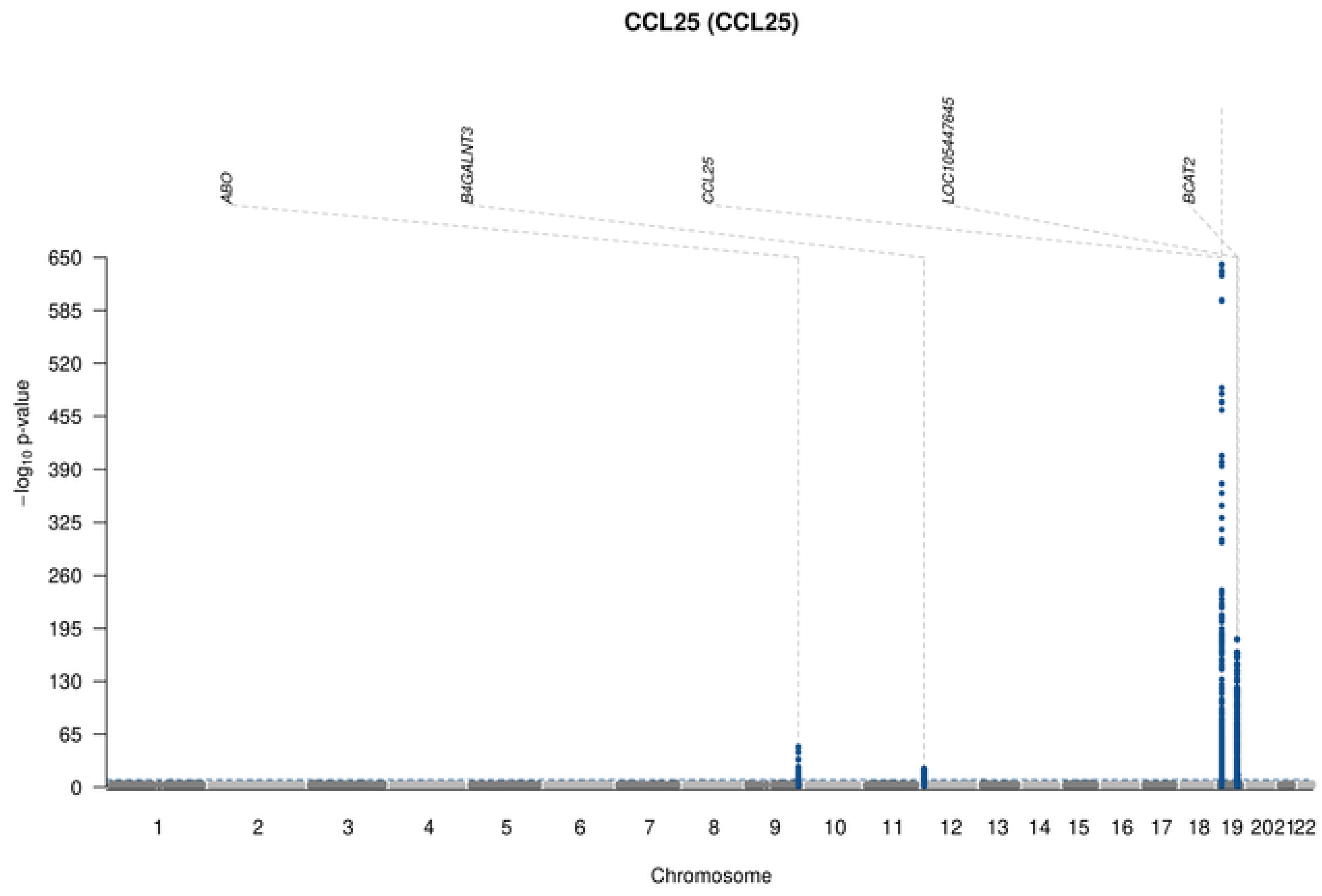

CCL25 (CCL25)
ABO
B4GALNT3
CCL25
LOC105447645
BCAT2
−log10 p-value
650
585
520
455
390
325
260
195
130
65
0
1 2 3 4 5 6 7 8 9 10 11 12 13 14 15 16 17 18 19 202122
Chromosome

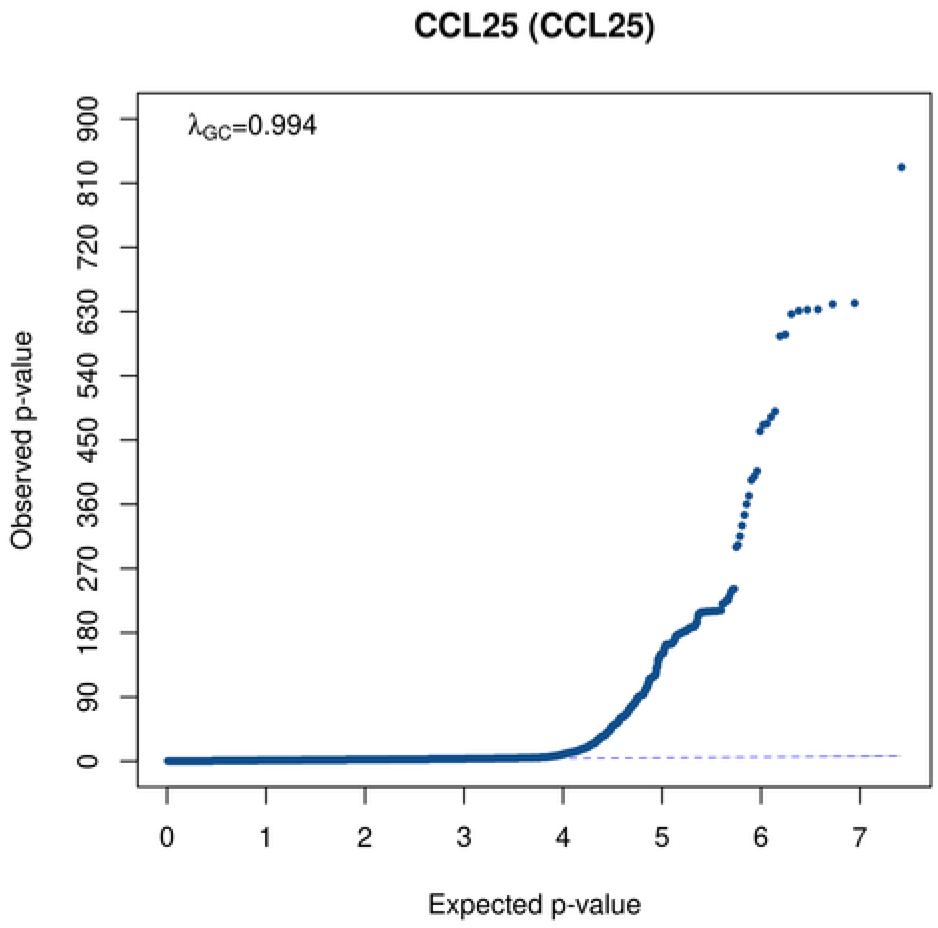

CCL25 (CCL25)
λGC=0.994
Observed p-value
900
810
720
630
540
450
360
270
180
90
0
0 1 2 3 4 5 6 7
Expected p-value

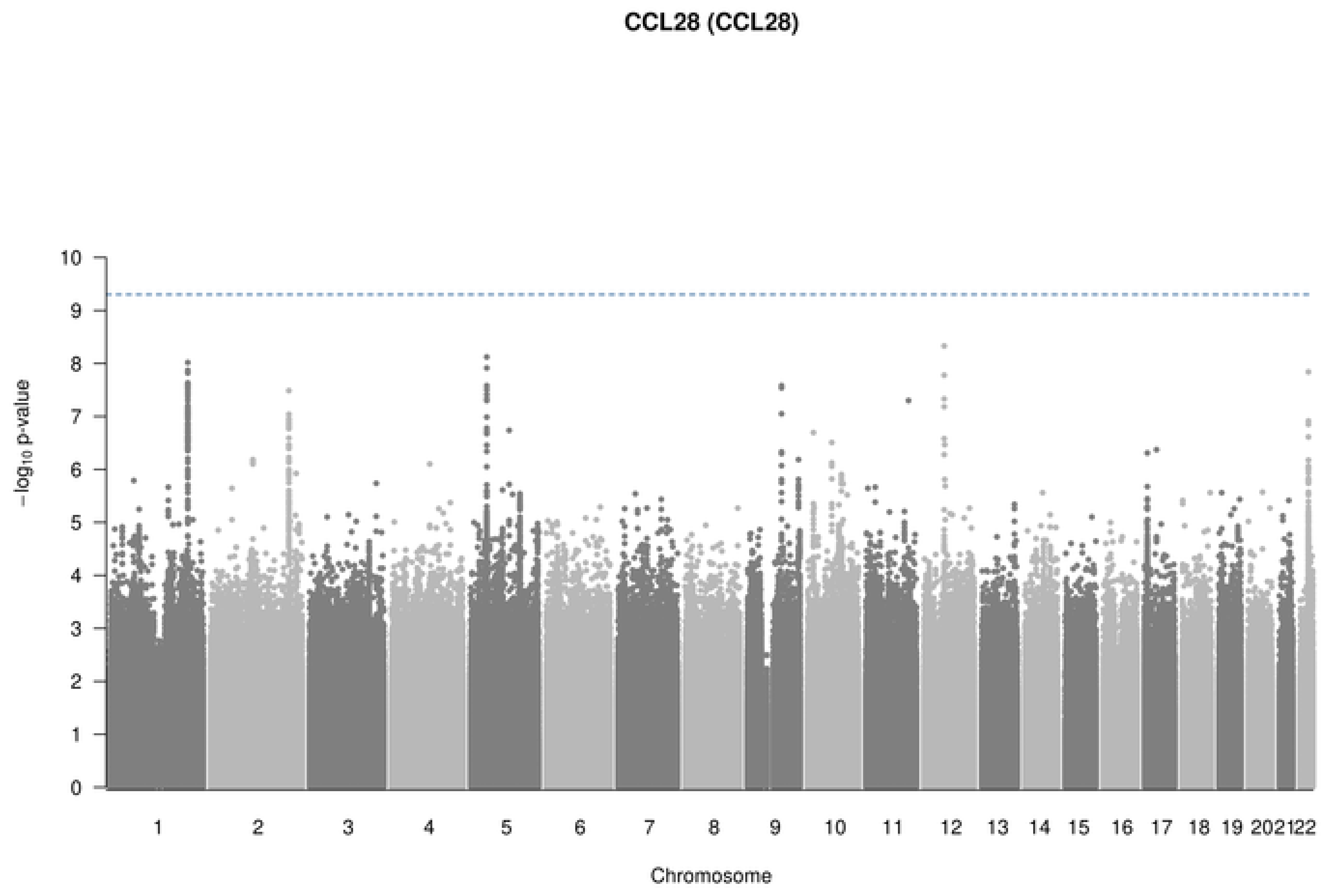
CCL28 (CCL28)
−log10 p-value
Chromosome
10
9
8
7
6
5
4
3
2
1
0
1 2 3 4 5 6 7 8 9 10 11 12 13 14 15 16 17 18 19 202122
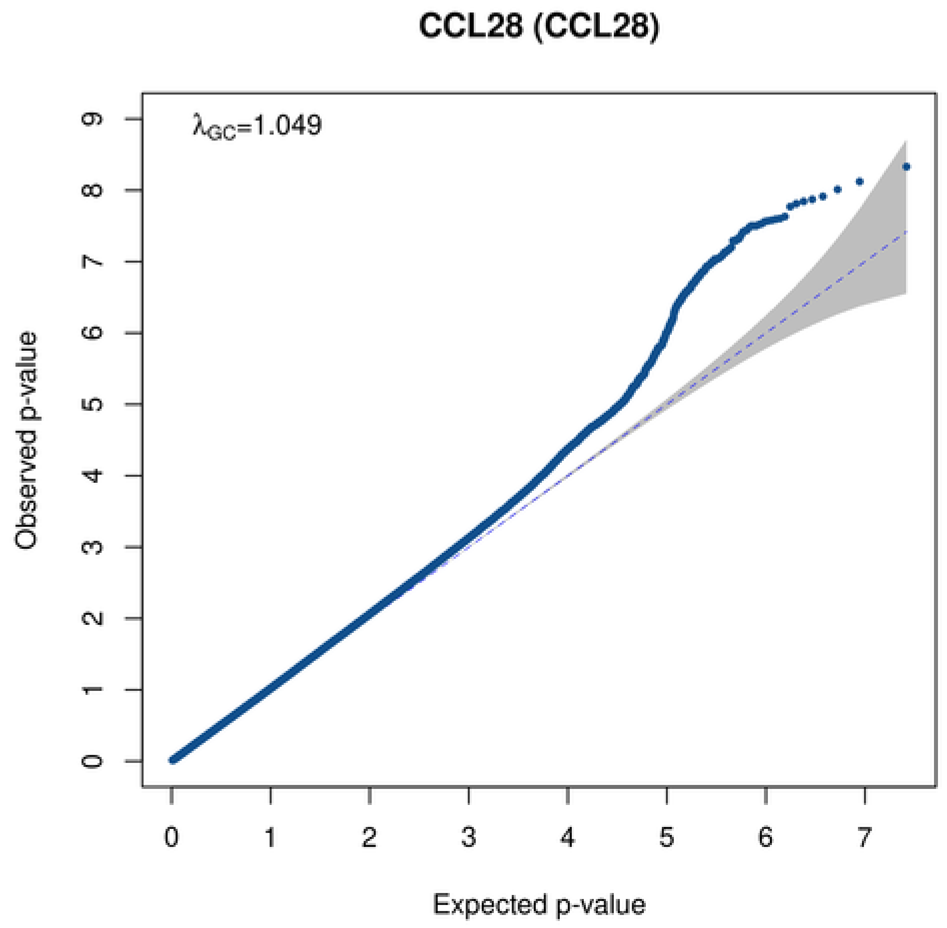
CCL28 (CCL28)
$\lambda_{GC}$=1.049
Observed p-value
Expected p-value
0 1 2 3 4 5 6 7 8 9
0 1 2 3 4 5 6 7

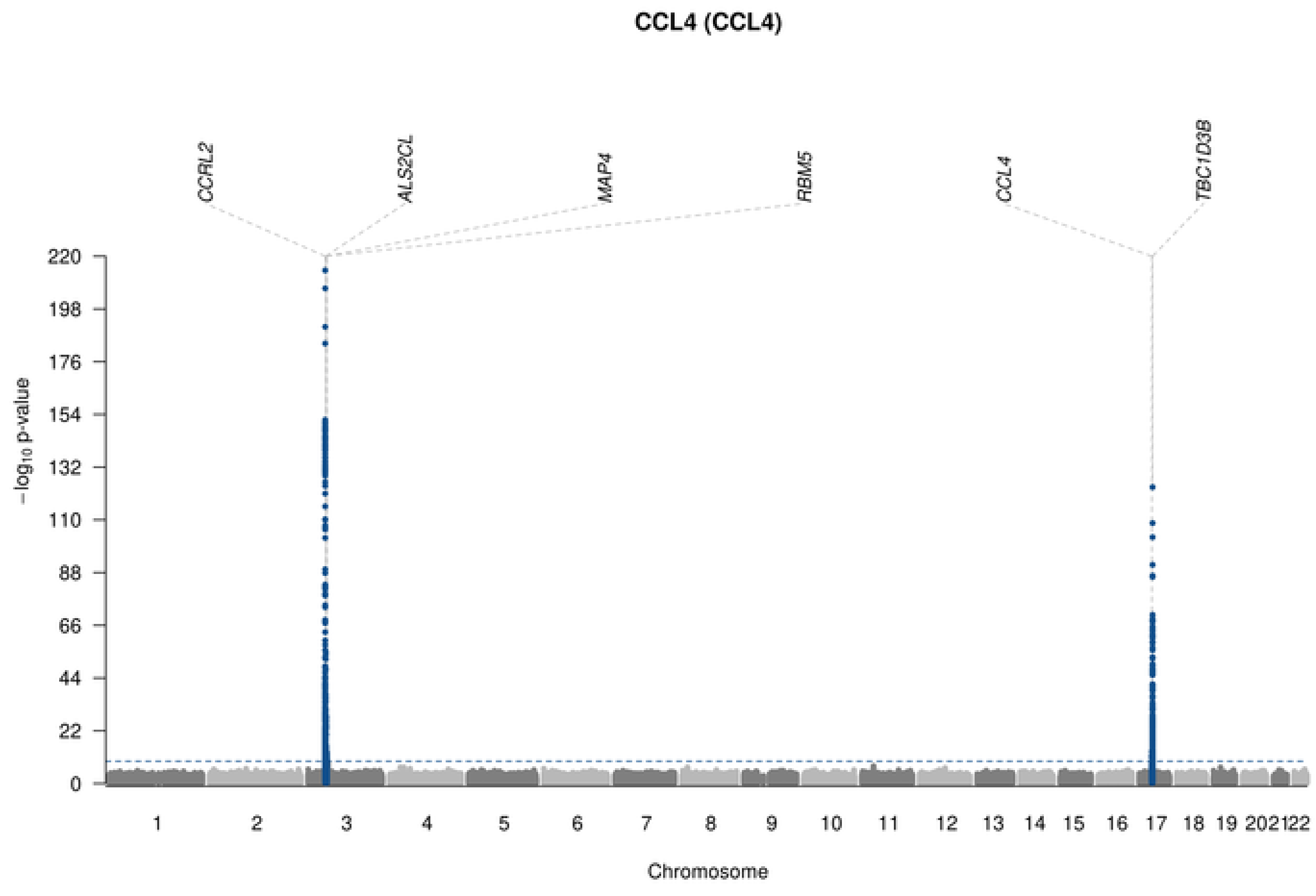
CCL4 (CCL4)
CCRL2
ALS2CL
MAP4
RBM5
CCL4
TBC1D3B
−log10 p-value
220
198
176
154
132
110
88
66
44
22
Chromosome

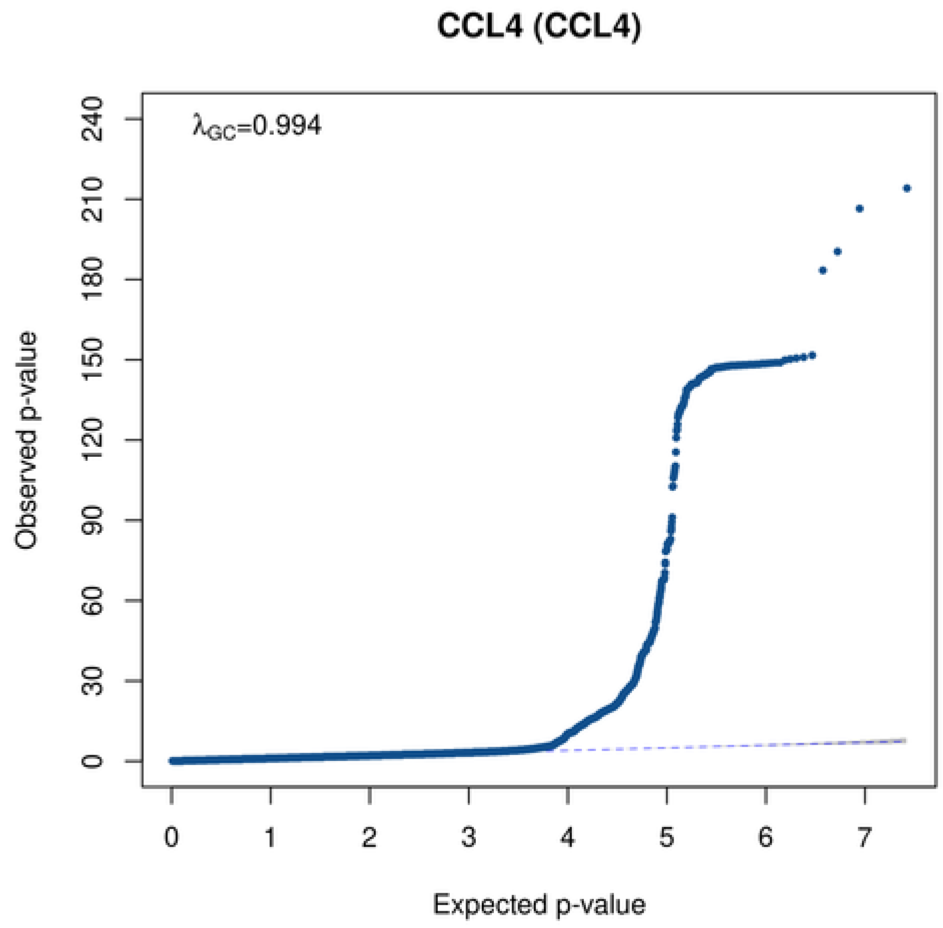
CCL4 (CCL4)
λGC=0.994
Observed p-value
240
210
180
150
120
90
60
30
0
0
1
2
3
4
5
6
7
Expected p-value

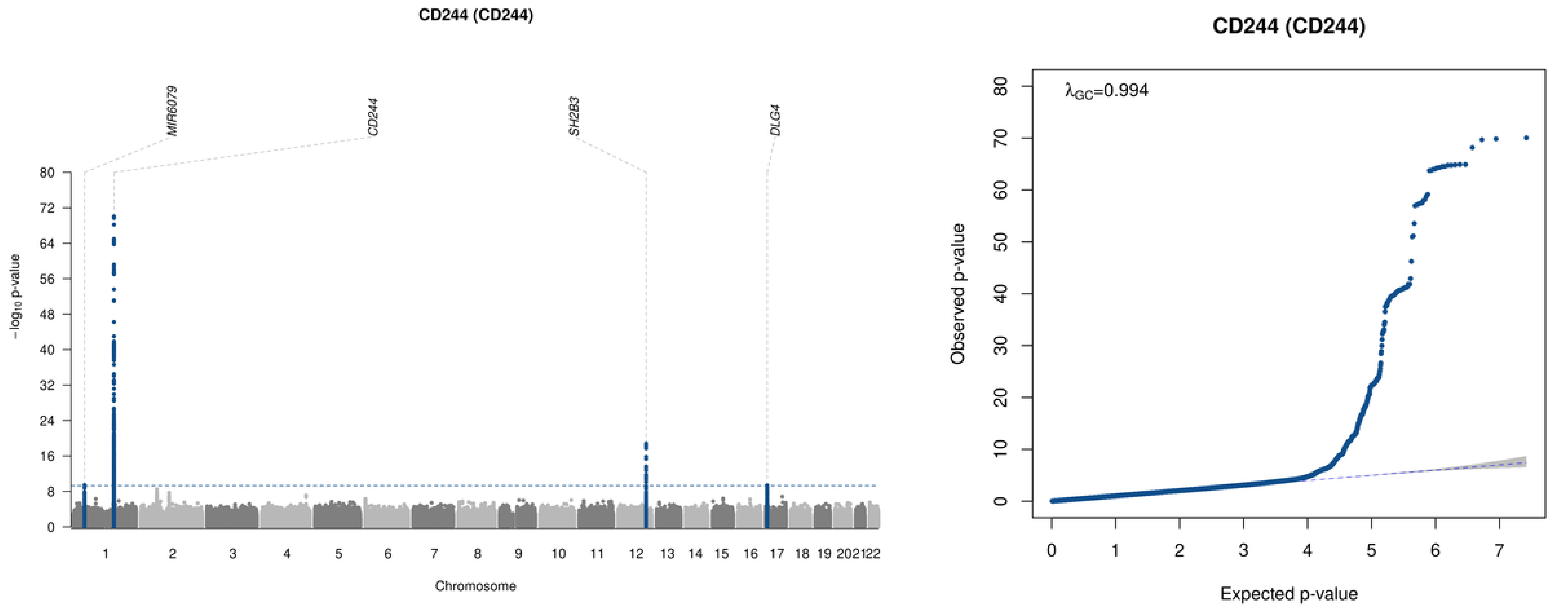

CD244 (CD244)
MIR6079
CD244
SH2B3
DLG4
−log10 p-value
Chromosome
1 2 3 4 5 6 7 8 9 10 11 12 13 14 15 16 17 18 19 20 21 22
0 8 16 24 32 40 48 56 64 72 80
CD244 (CD244)
λGC=0.994
Observed p-value
Expected p-value
0 10 20 30 40 50 60 70 80
0 1 2 3 4 5 6 7

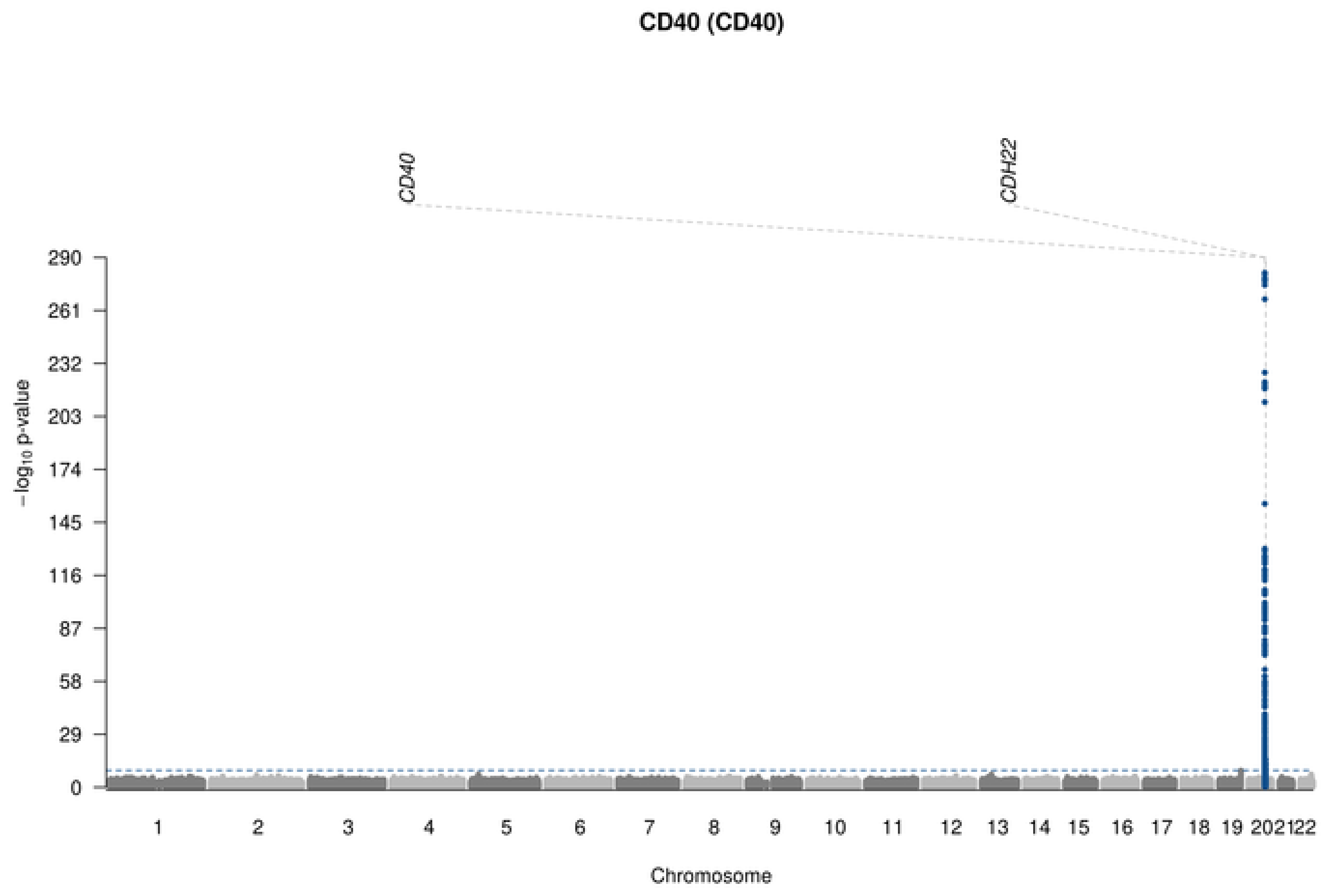
CD40 (CD40)
CD40
CDH22
−log10 p-value
290
261
232
203
174
145
116
87
58
29
Chromosome

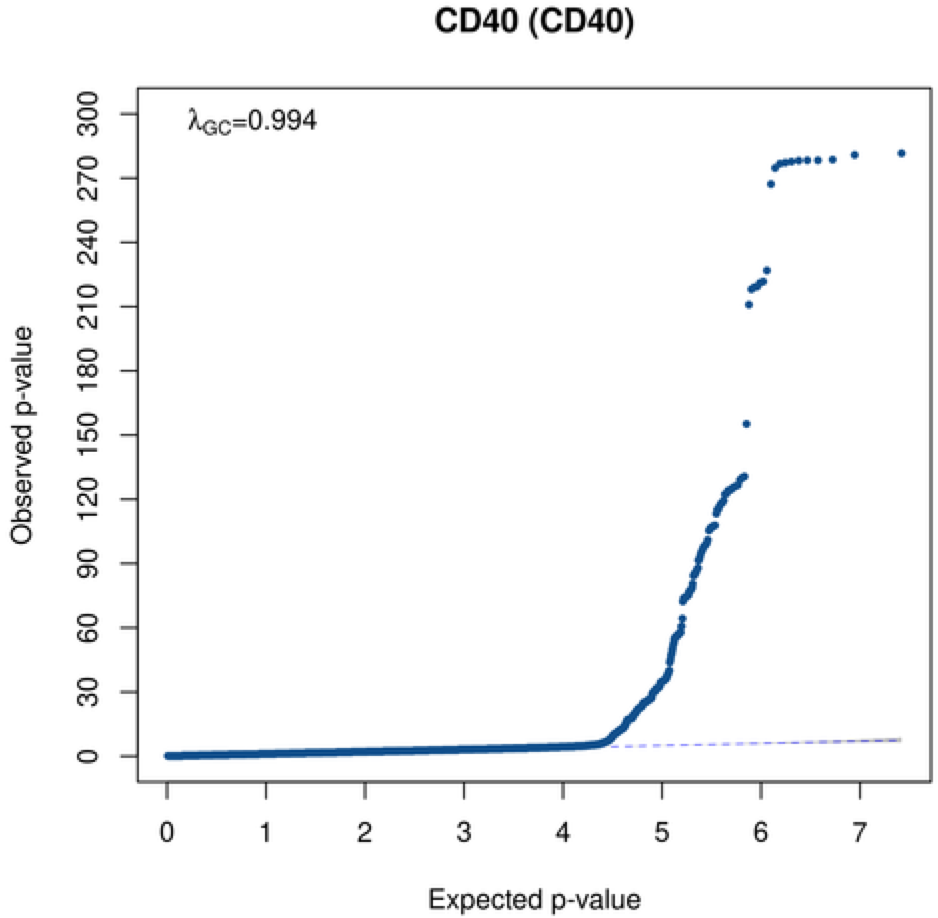
CD40 (CD40)
λGC=0.994
Observed p-value
300
270
240
210
180
150
120
90
60
30
0
Expected p-value
0
1
2
3
4
5
6
7

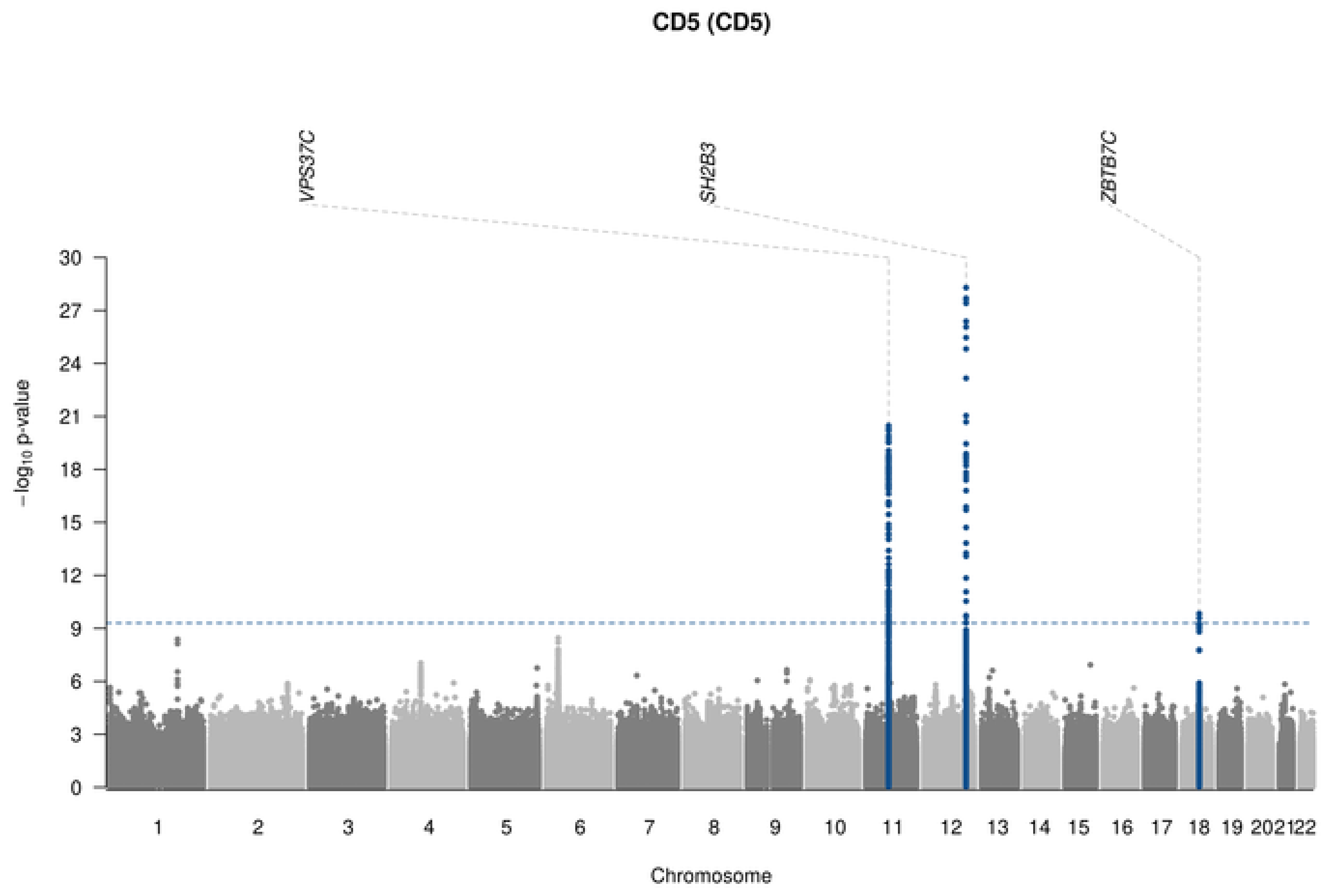

CD5 (CD5)
VPS37C
SH2B3
ZBTB7C
−log10 p-value
Chromosome

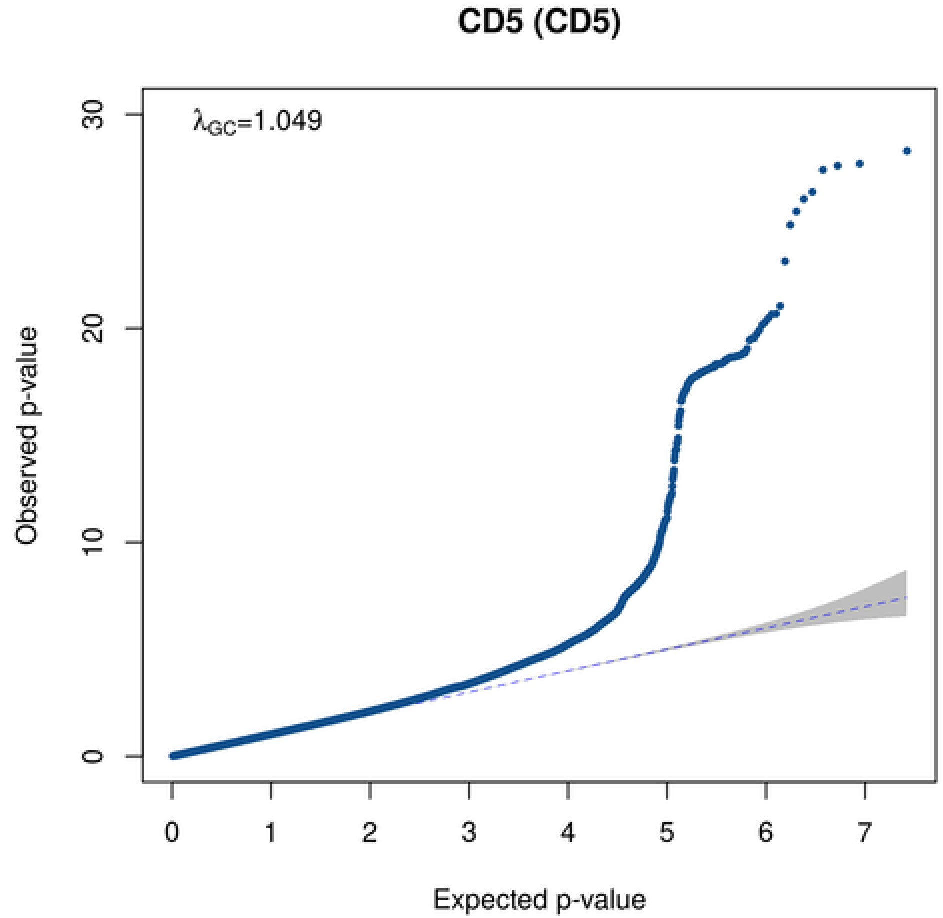

CD5 (CD5)
λGC=1.049
Observed p-value
Expected p-value

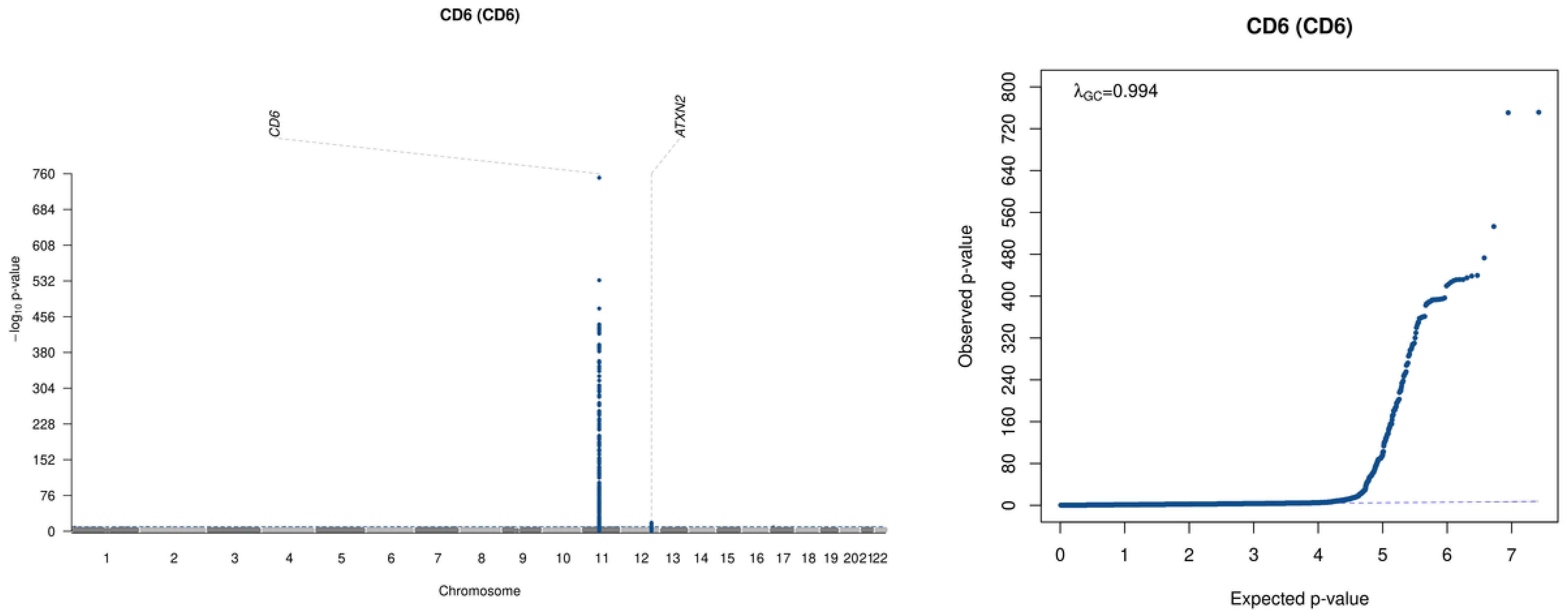

CD6 (CD6)
CD6
ATXN2
760
684
608
532
456
380
304
228
152
76
0
−log10 p-value
202122
Chromosome
CD6 (CD6)
λGC=0.994
800
720
640
560
480
400
320
240
160
80
0
Observed p-value
0
1
2
3
4
5
6
7
Expected p-value

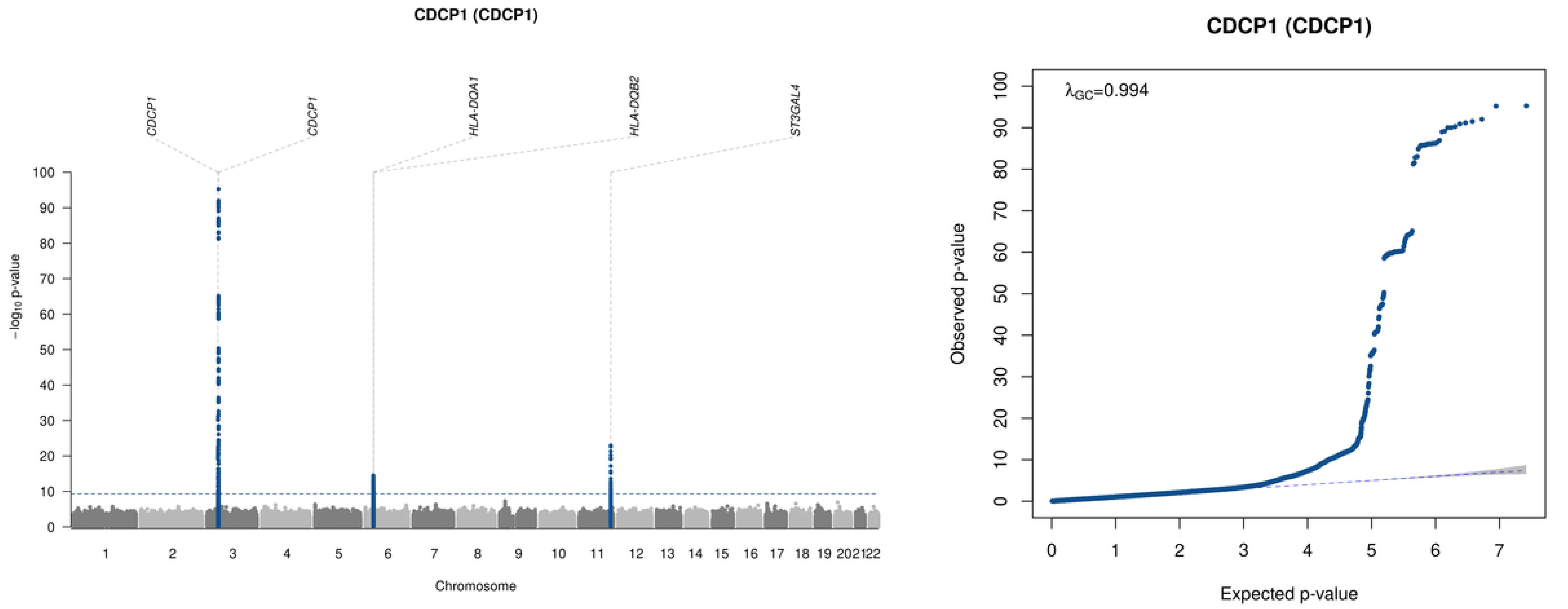

CDCP1 (CDCP1)
CDCP1
CDCP1
HLA-DQA1
HLA-DQB2
ST3GAL4
−log10 p-value
Chromosome
CDCP1 (CDCP1)
λGC=0.994
Observed p-value
Expected p-value

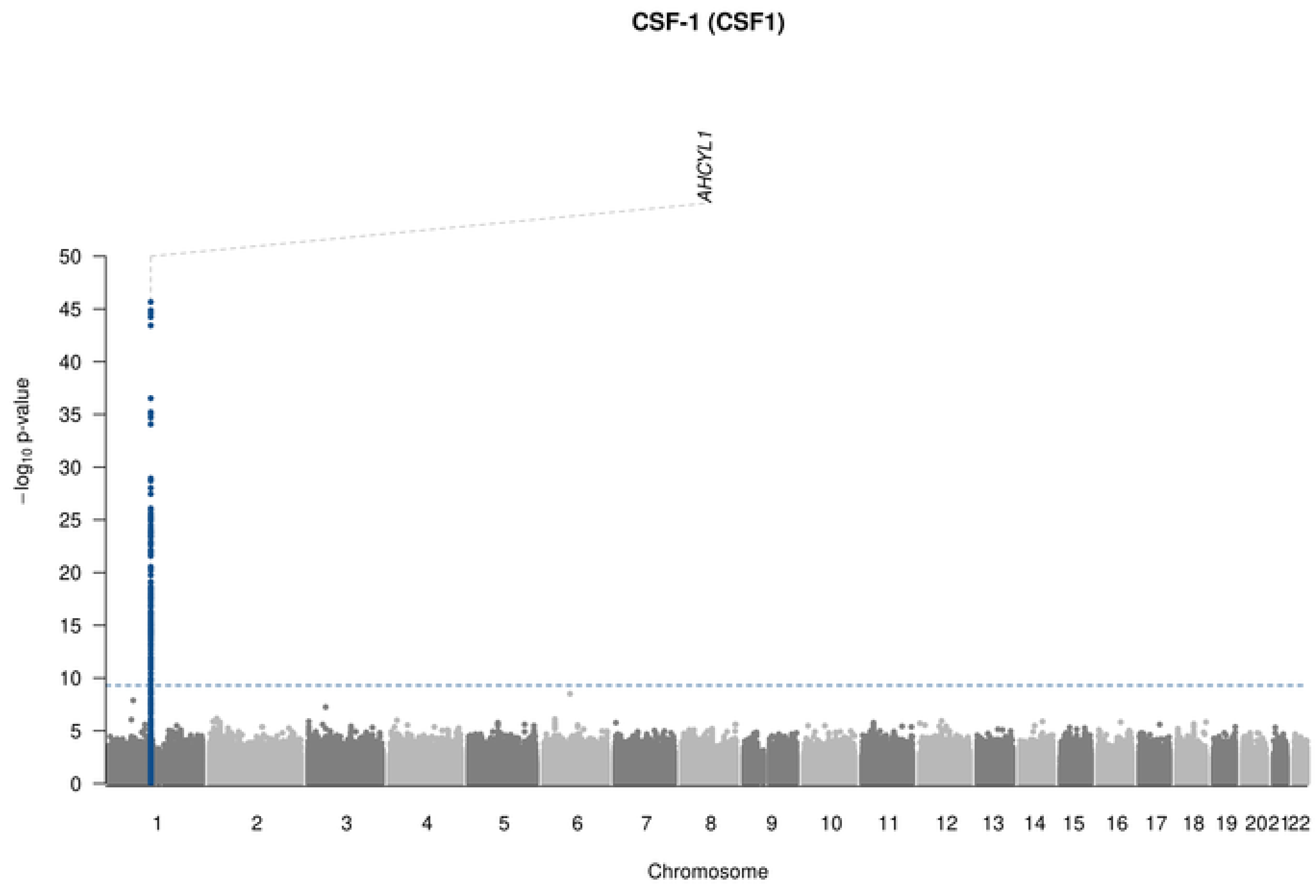

CSF-1 (CSF1)
AHCYL1
−log10 p-value
50
45
40
35
30
25
20
15
10
5
0
1 2 3 4 5 6 7 8 9 10 11 12 13 14 15 16 17 18 19 202122
Chromosome

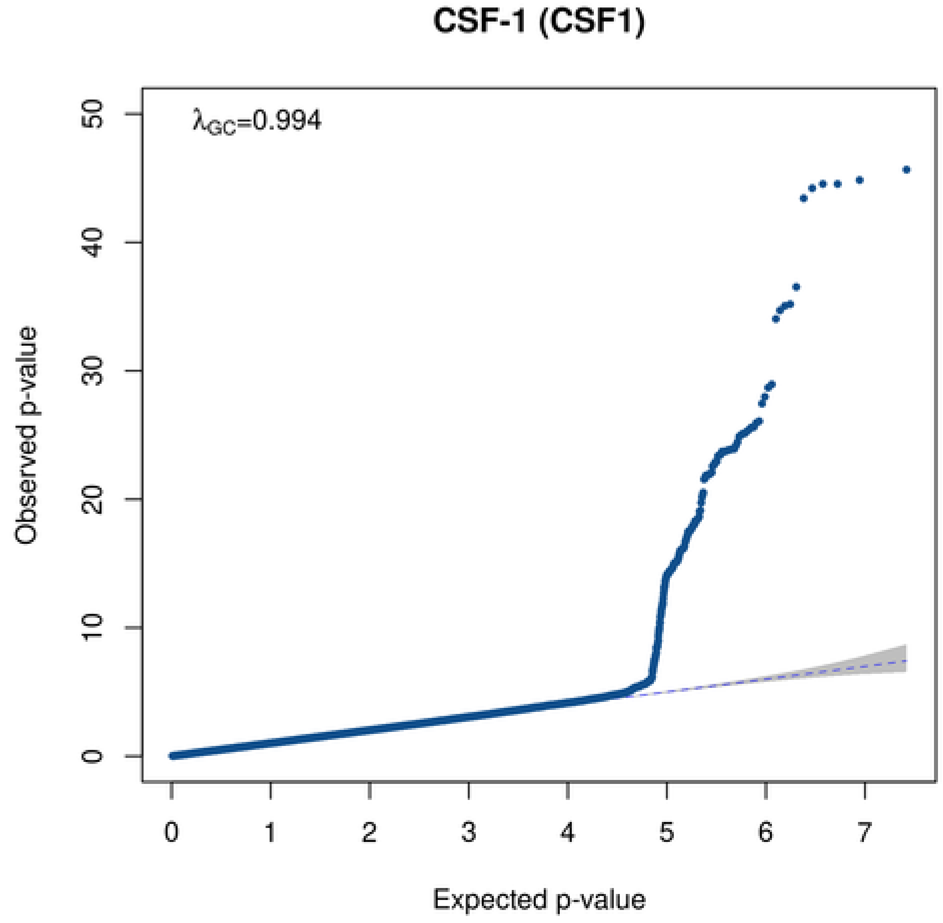

CSF-1 (CSF1)
λGC=0.994
Observed p-value
50
40
30
20
10
0
0 1 2 3 4 5 6 7
Expected p-value

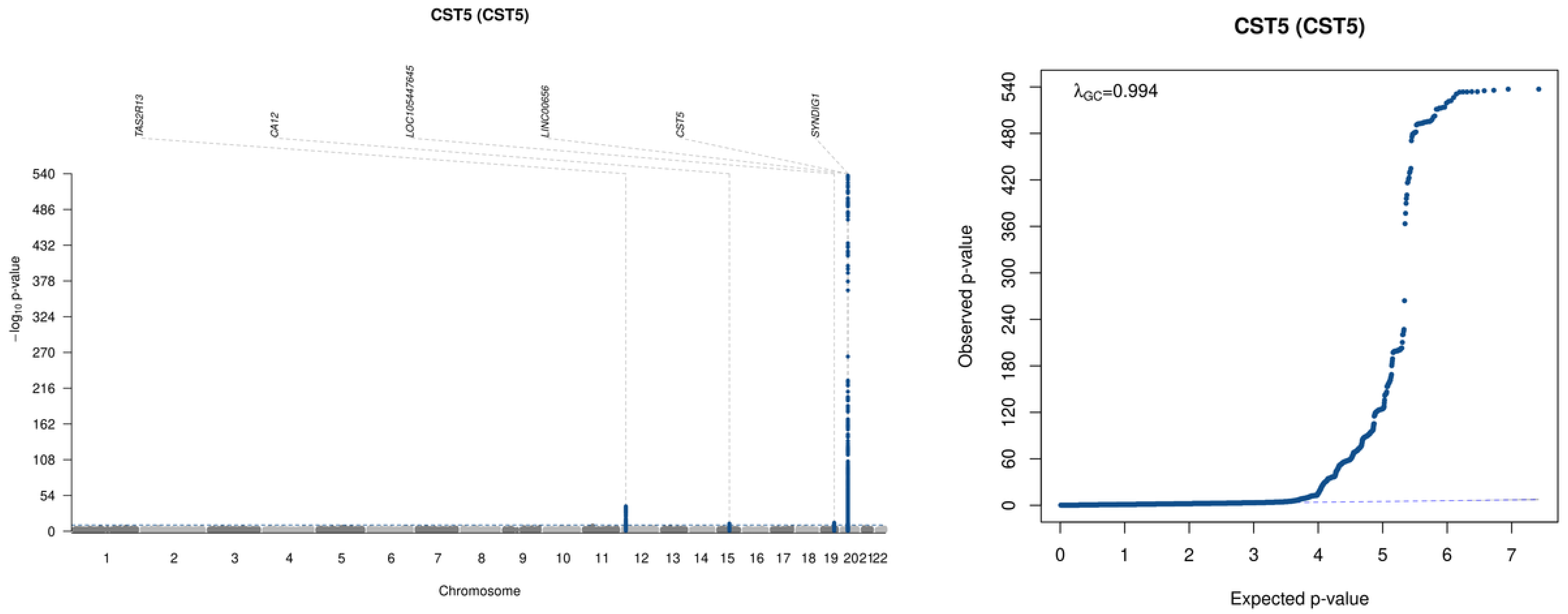

CST5 (CST5)
TAS2R13
CA12
LOC105447645
LINC00656
CST5
SYNDIG1
$-\log_{10}$ p-value
Chromosome
CST5 (CST5)
$\lambda_{GC}$=0.994
Observed p-value
Expected p-value

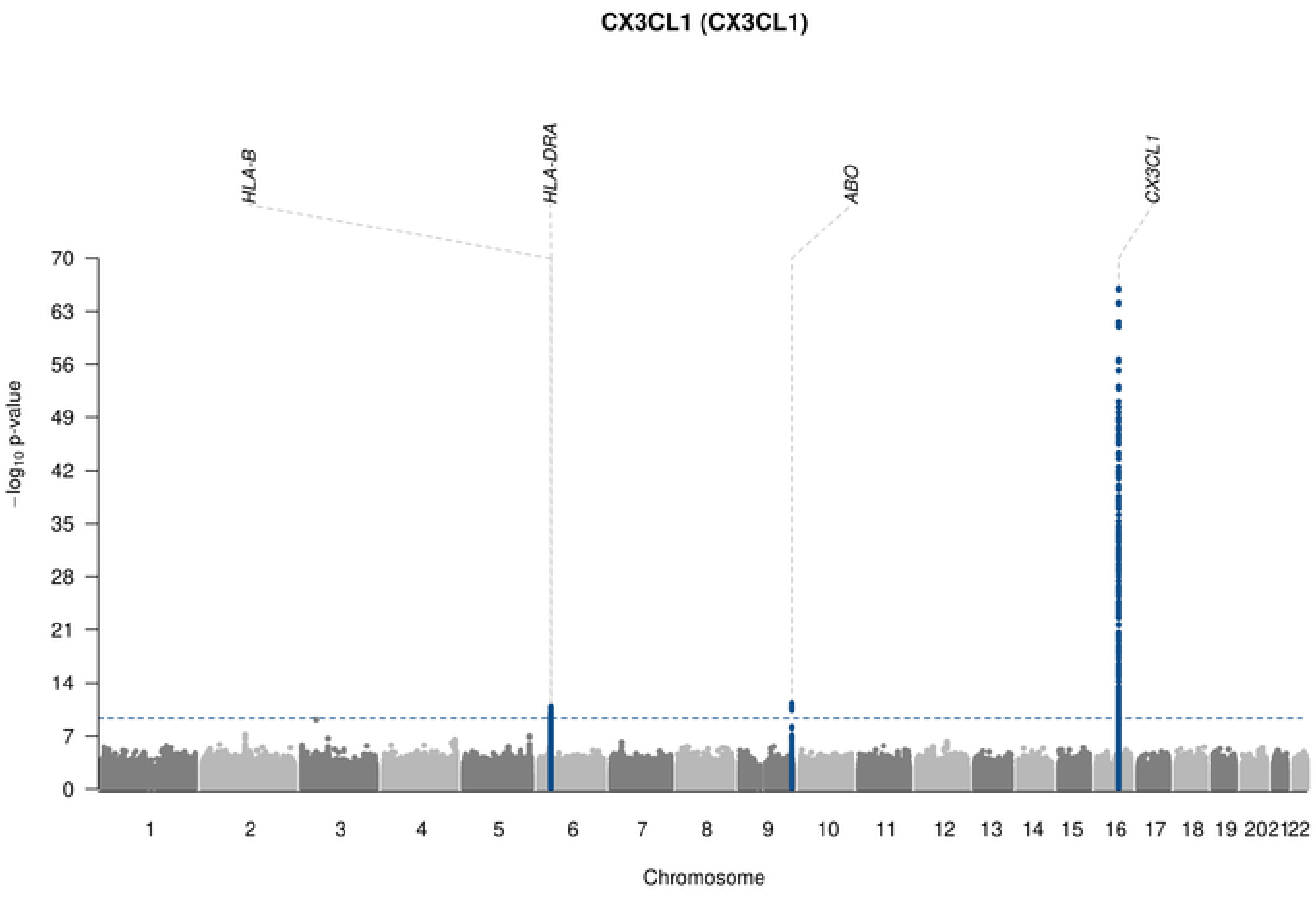

CX3CL1 (CX3CL1)
HLA-B
HLA-DRA
ABO
CX3CL1
70
63
56
49
42
35
28
21
14
7
0
$-\log_{10}$ p-value
1 2 3 4 5 6 7 8 9 10 11 12 13 14 15 16 17 18 19 20 21 22
Chromosome

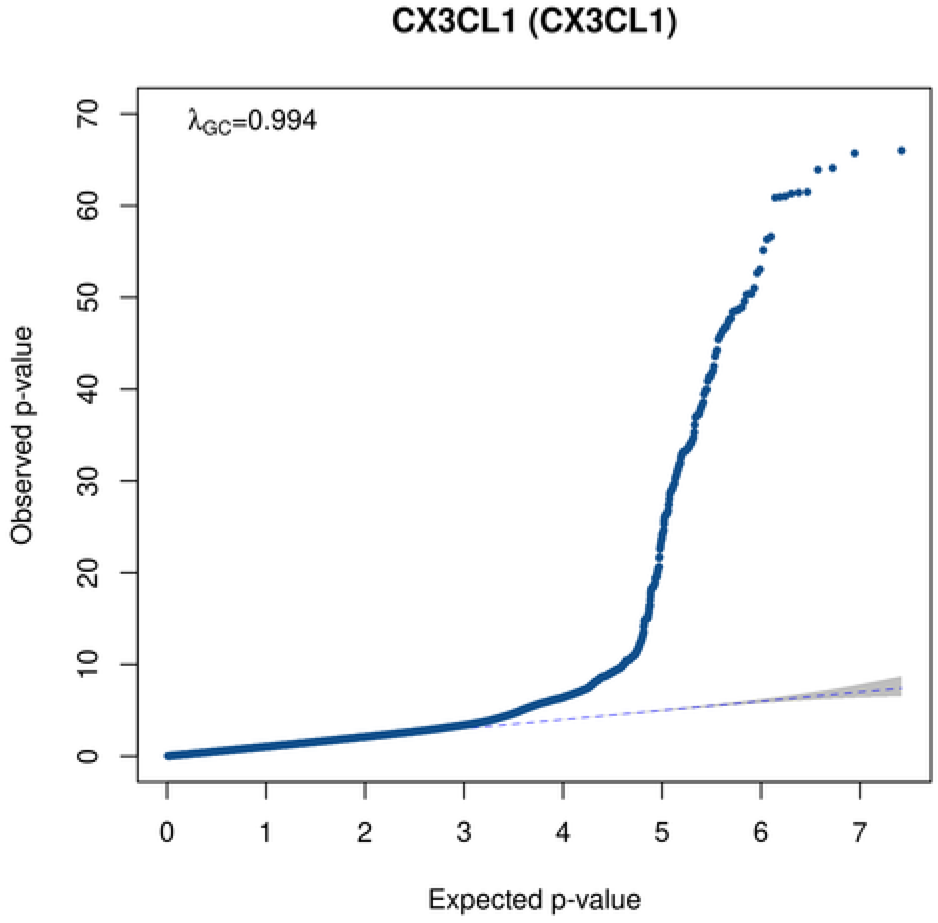

CX3CL1 (CX3CL1)
$\lambda_{GC}$=0.994
0 10 20 30 40 50 60 70
Observed p-value
0 1 2 3 4 5 6 7
Expected p-value

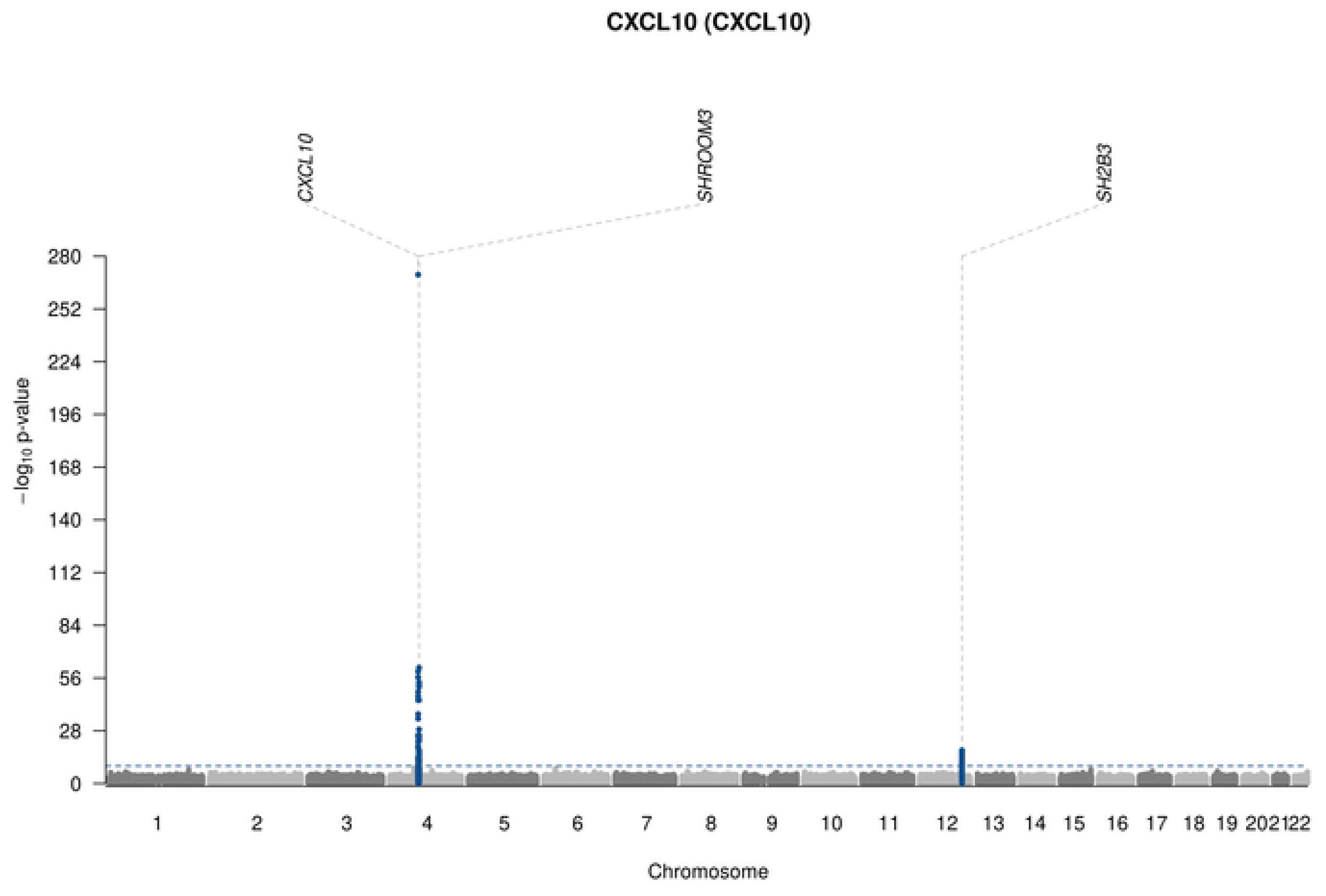
CXCL10 (CXCL10)
CXCL10
SHROOM3
SH2B3
280
252
224
196
168
140
112
84
56
28
0
−log10 p-value
202122
Chromosome

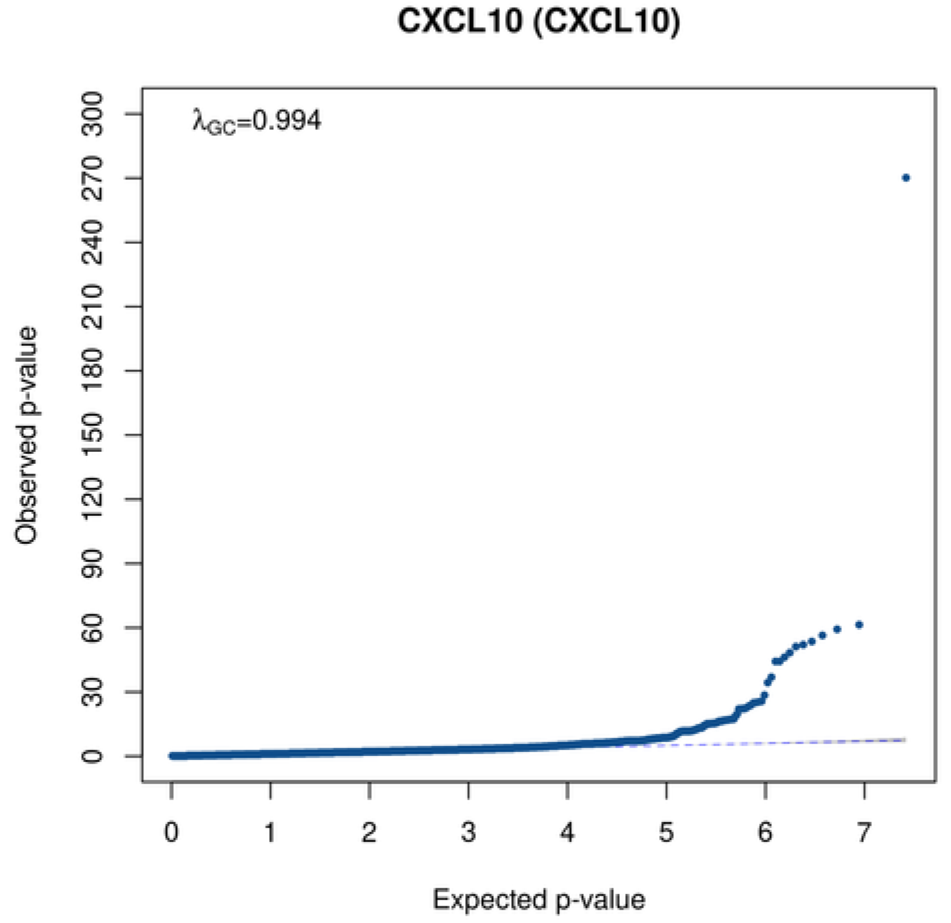
CXCL10 (CXCL10)
λGC=0.994
300
270
240
210
180
150
120
90
60
30
0
Observed p-value
0
1
2
3
4
5
6
7
Expected p-value

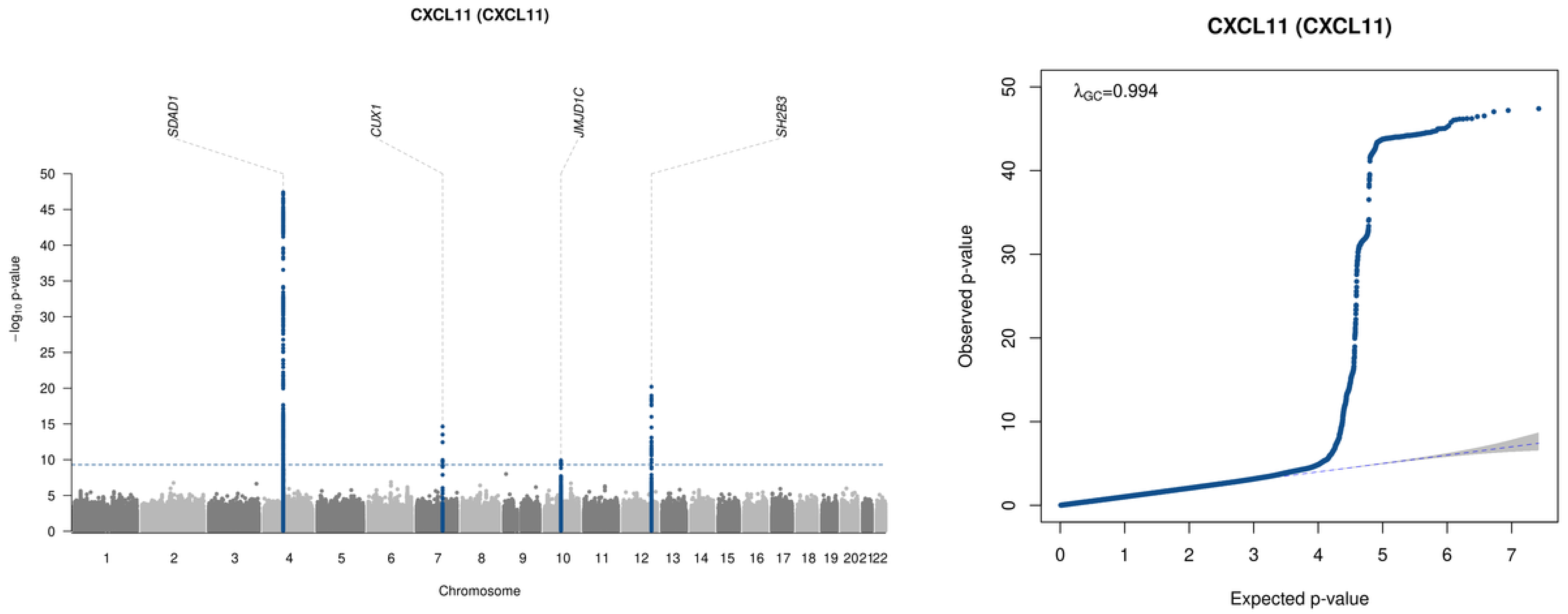

CXCL11 (CXCL11)
SDAD1
CUX1
JMJD1C
SH2B3
−log10 p-value
Chromosome
1 2 3 4 5 6 7 8 9 10 11 12 13 14 15 16 17 18 19 2021 22
CXCL11 (CXCL11)
λGC=0.994
Observed p-value
Expected p-value

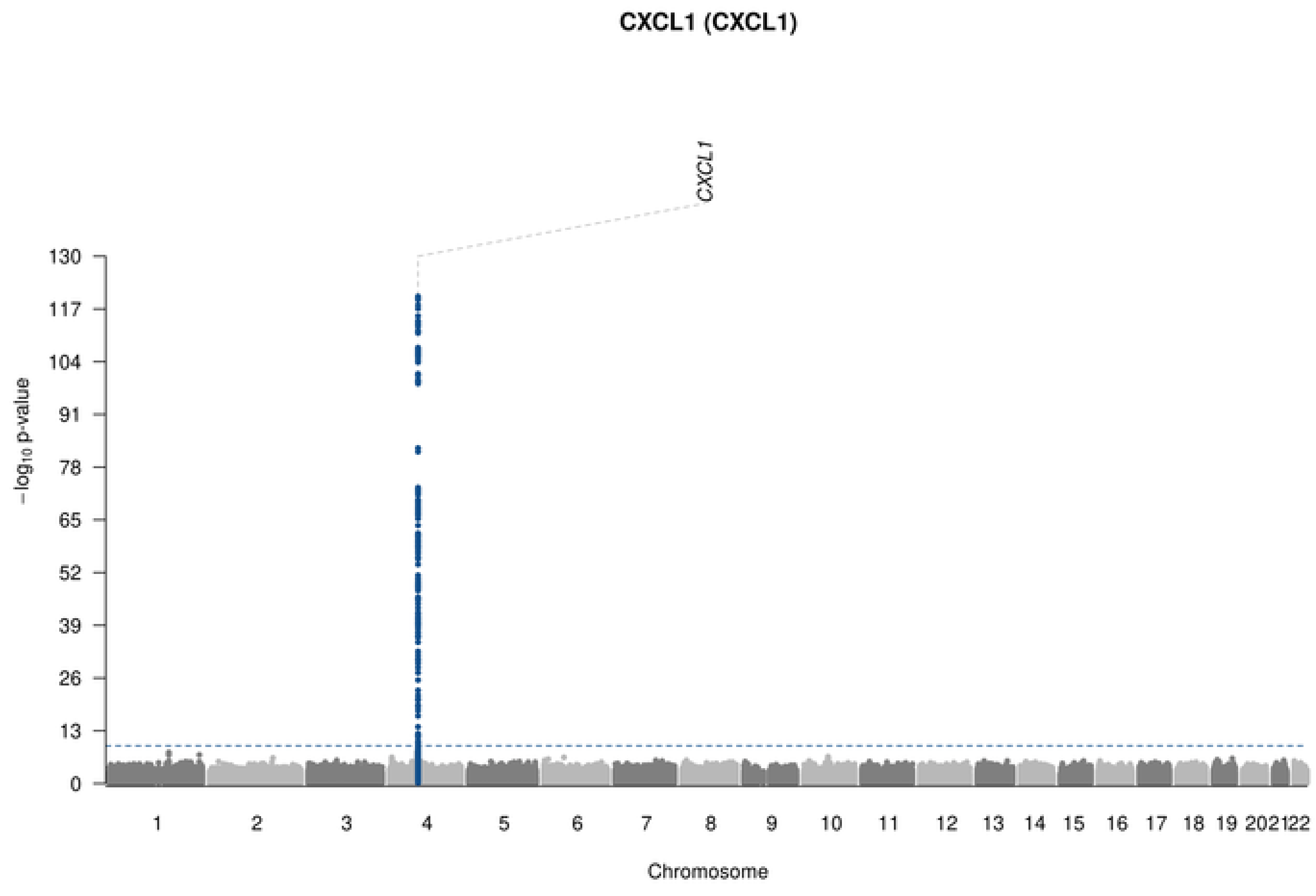
CXCL1 (CXCL1)
CXCL1
−log10 p-value
130
117
104
91
78
65
52
39
26
13
0
1 2 3 4 5 6 7 8 9 10 11 12 13 14 15 16 17 18 19 202122
Chromosome

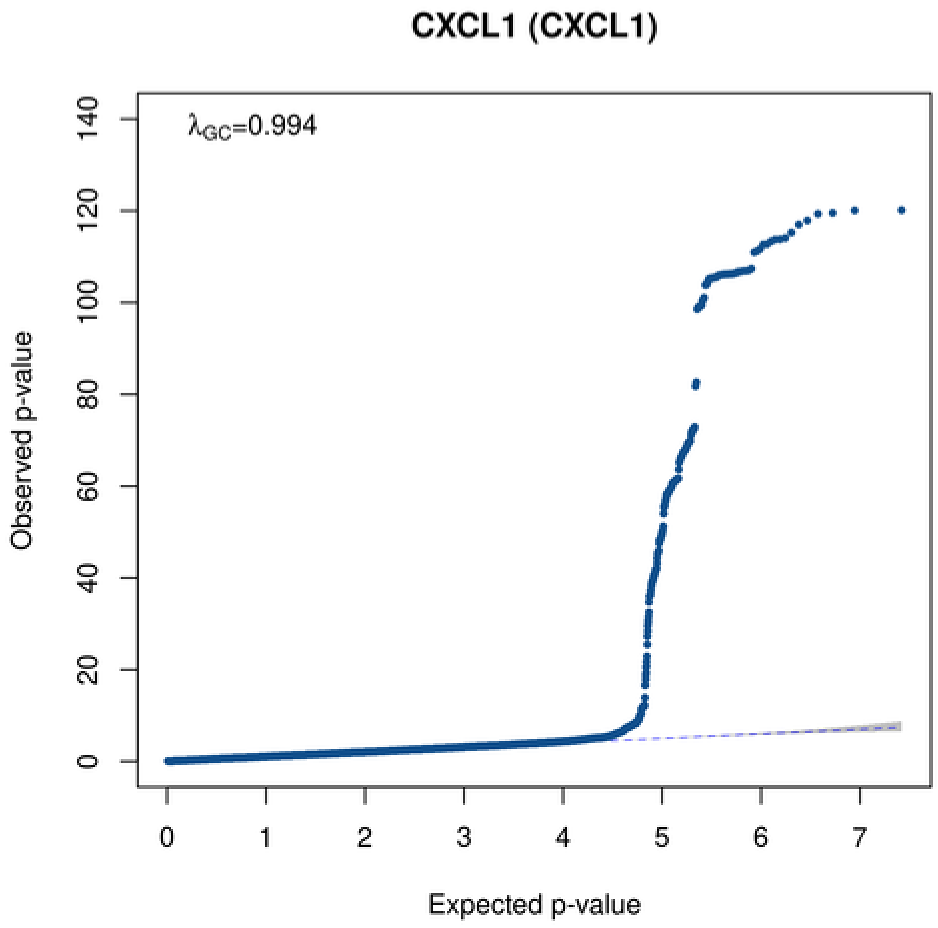
CXCL1 (CXCL1)
λGC=0.994
Observed p-value
140
120
100
80
60
40
20
0
0 1 2 3 4 5 6 7
Expected p-value

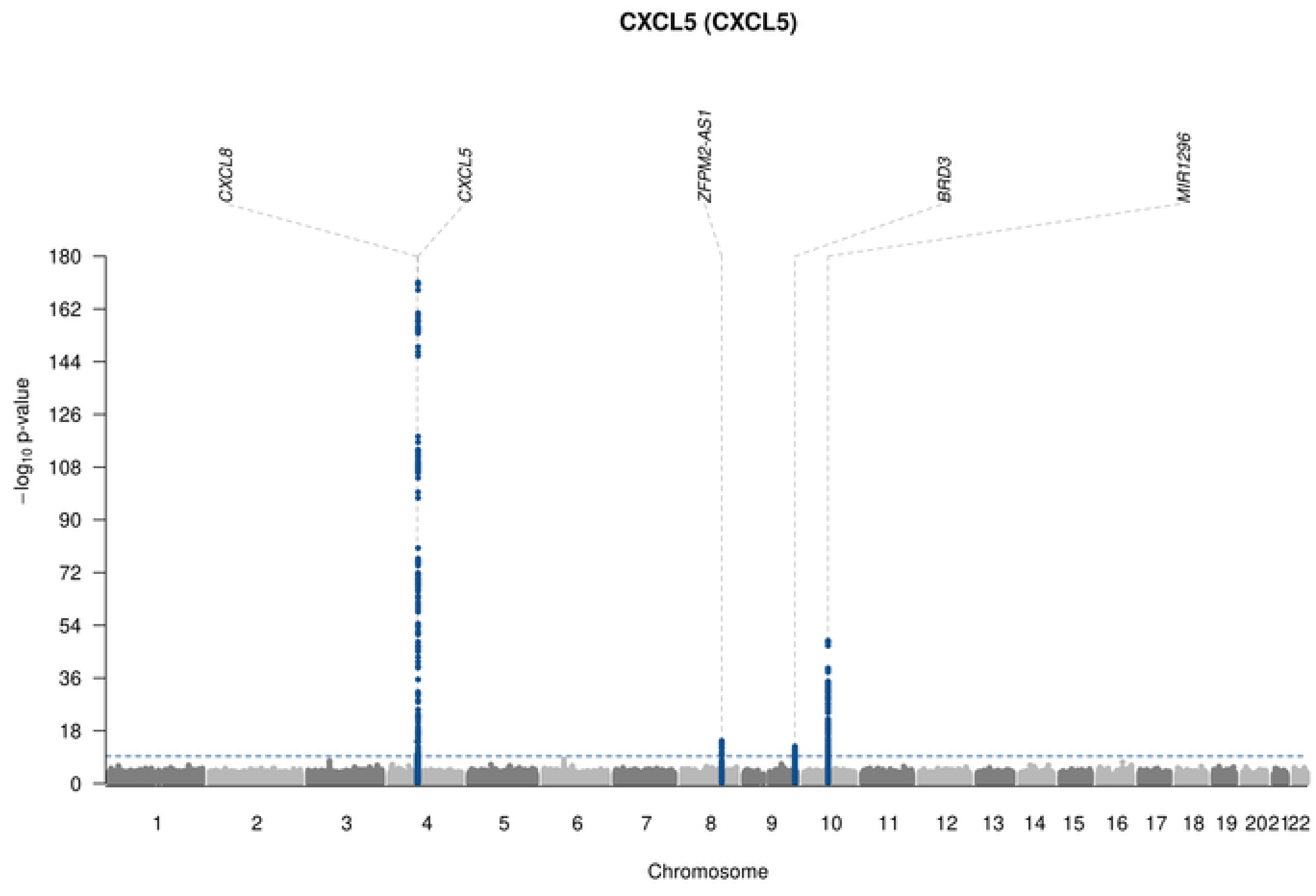
CXCL5 (CXCL5)
CXCL8
CXCL5
ZFPM2-AS1
BRD3
MIR1296
$-\log_{10}$ p-value
Chromosome

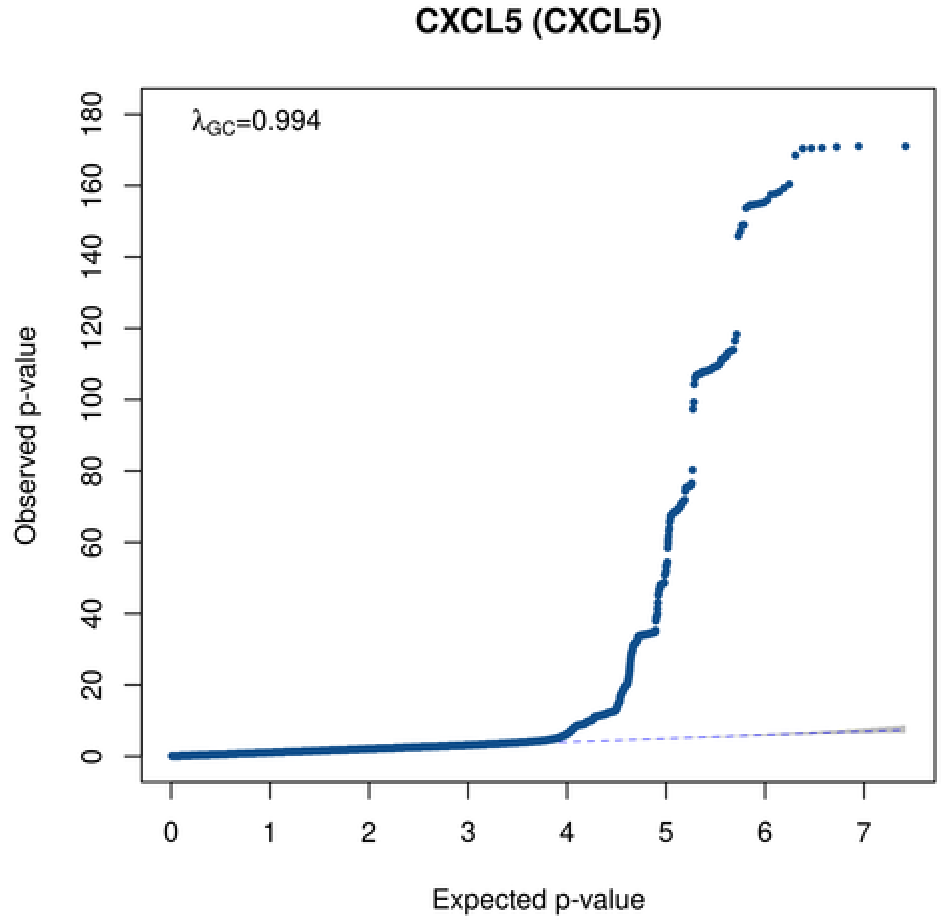
CXCL5 (CXCL5)
$\lambda_{GC}$=0.994
Observed p-value
Expected p-value

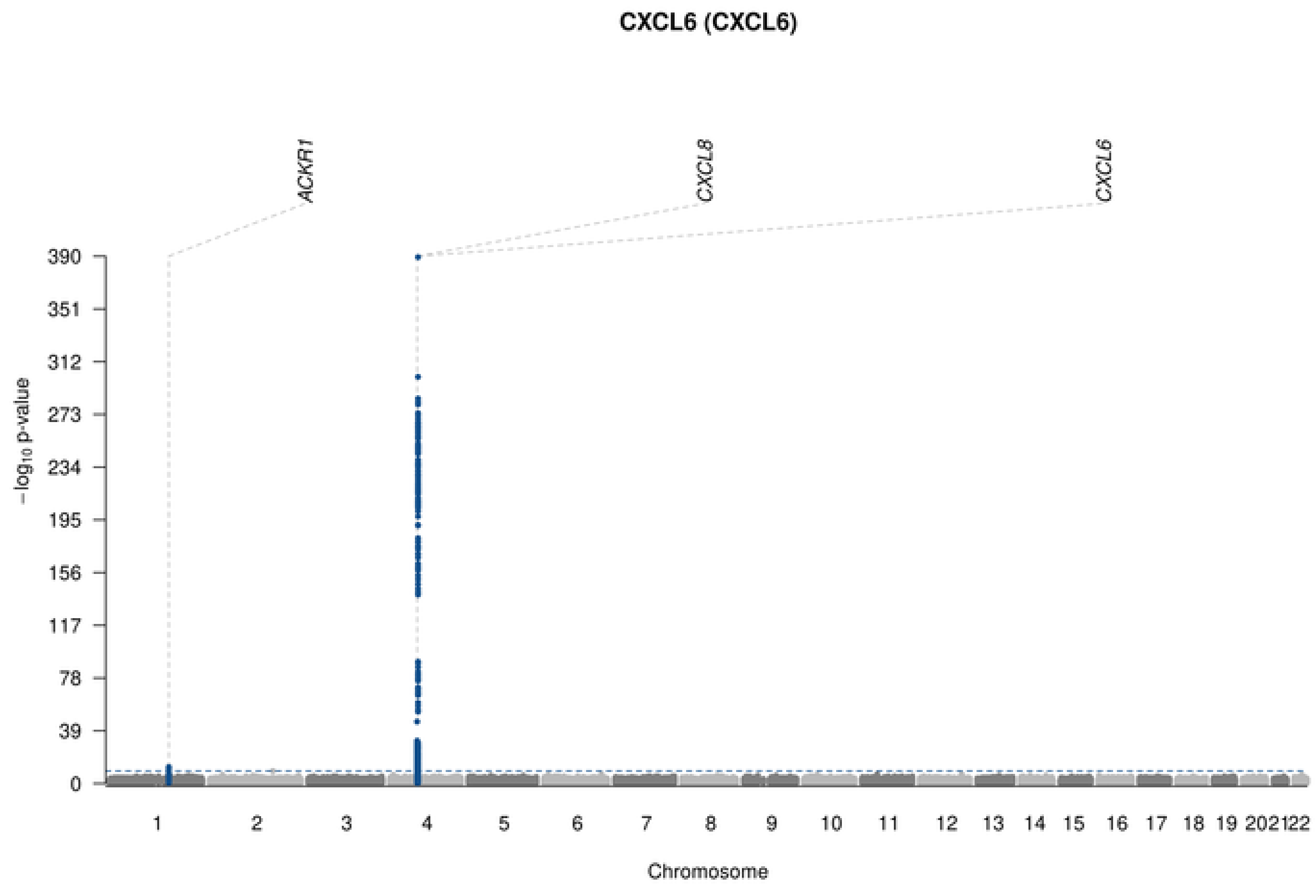

CXCL6 (CXCL6)
ACKR1
CXCL8
CXCL6
-log10 p-value
390
351
312
273
234
195
156
117
78
39
202122
Chromosome

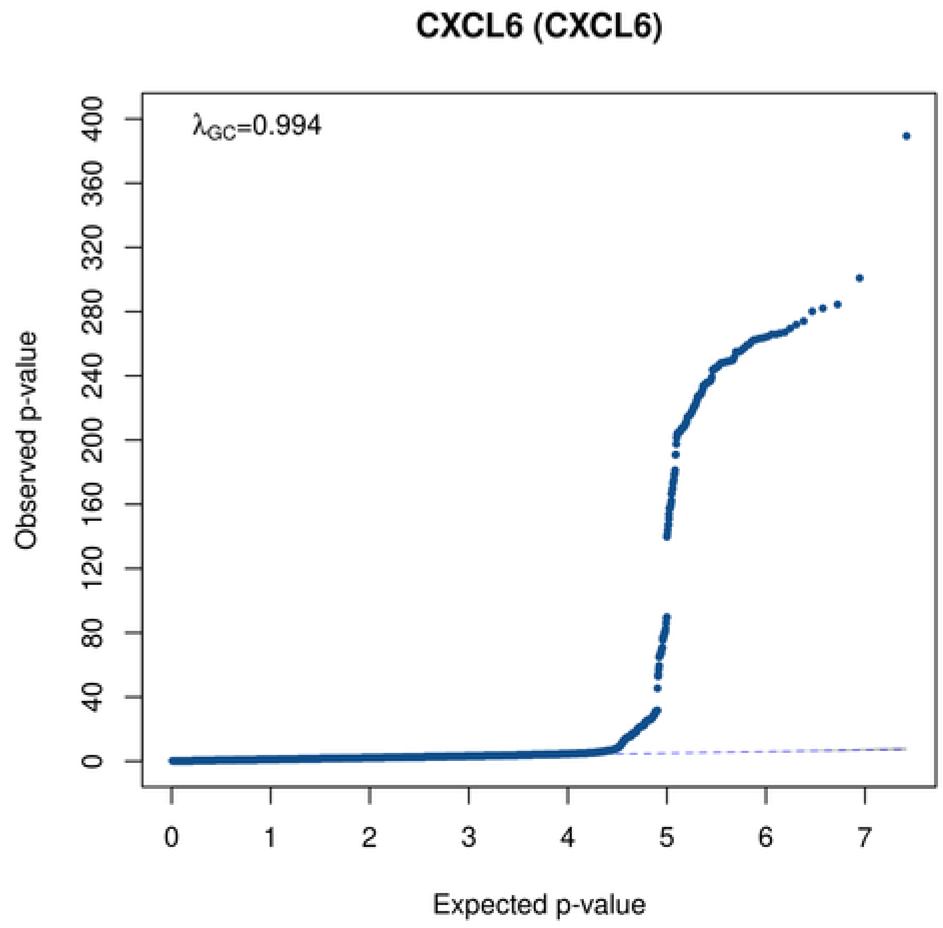

CXCL6 (CXCL6)
λGC=0.994
Observed p-value
400
360
320
280
240
200
160
120
80
40
0
0
1
2
3
4
5
6
7
Expected p-value

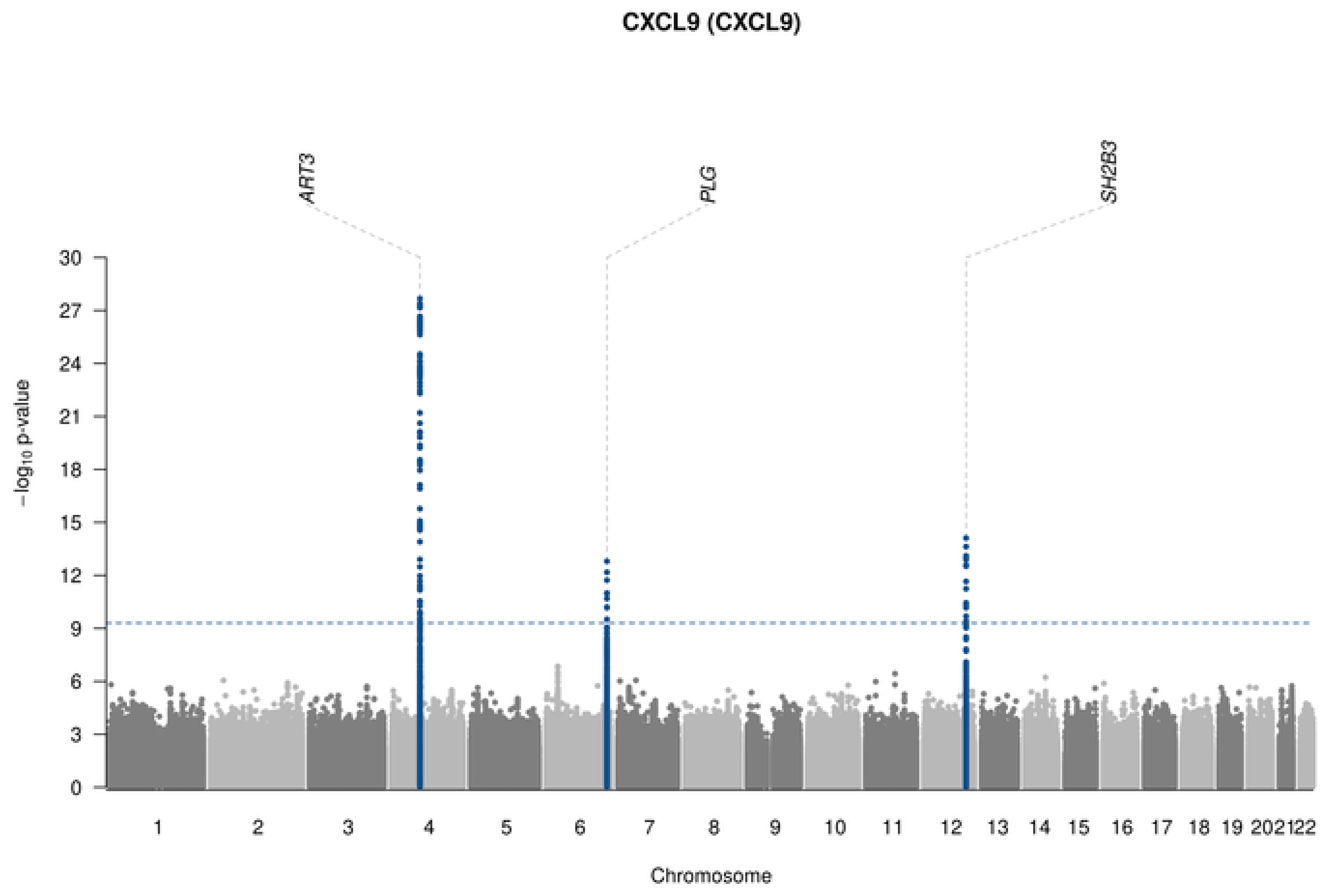
CXCL9 (CXCL9)
ART3
PLG
SH2B3
$-\log_{10}$ p-value
30
27
24
21
18
15
12
9
6
3
0
1 2 3 4 5 6 7 8 9 10 11 12 13 14 15 16 17 18 19 20 21 22
Chromosome

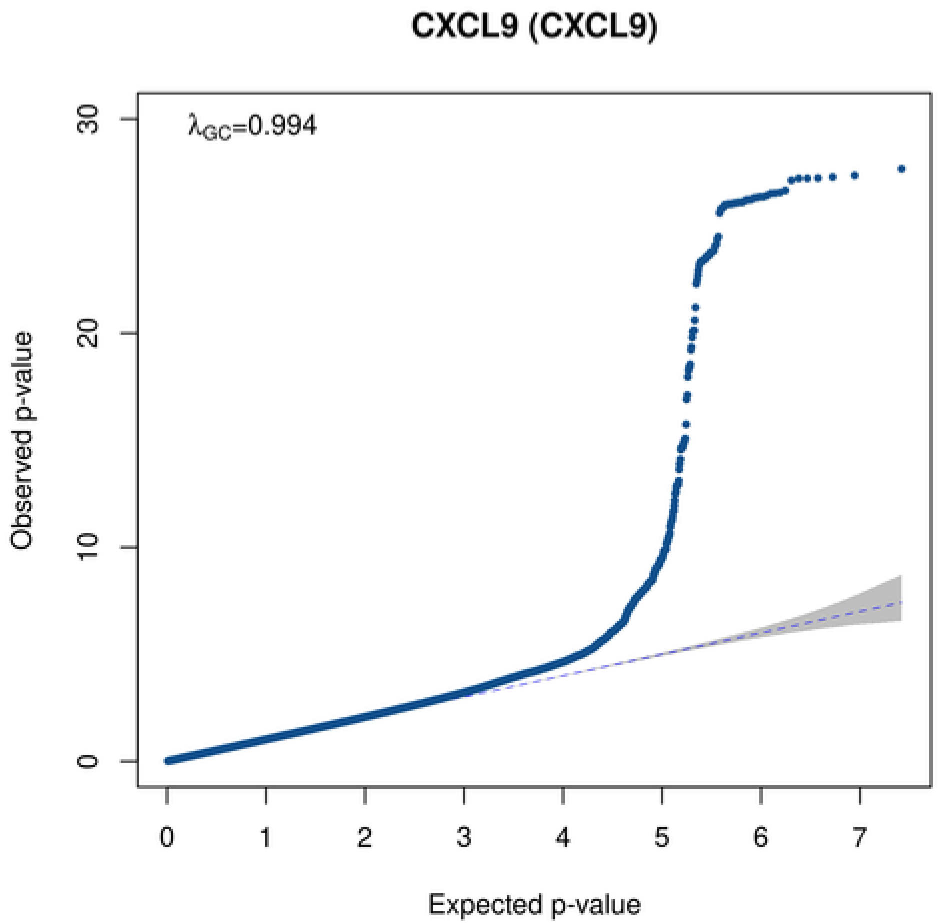
CXCL9 (CXCL9)
$\lambda_{GC}$=0.994
Observed p-value
30
20
10
0
0 1 2 3 4 5 6 7
Expected p-value

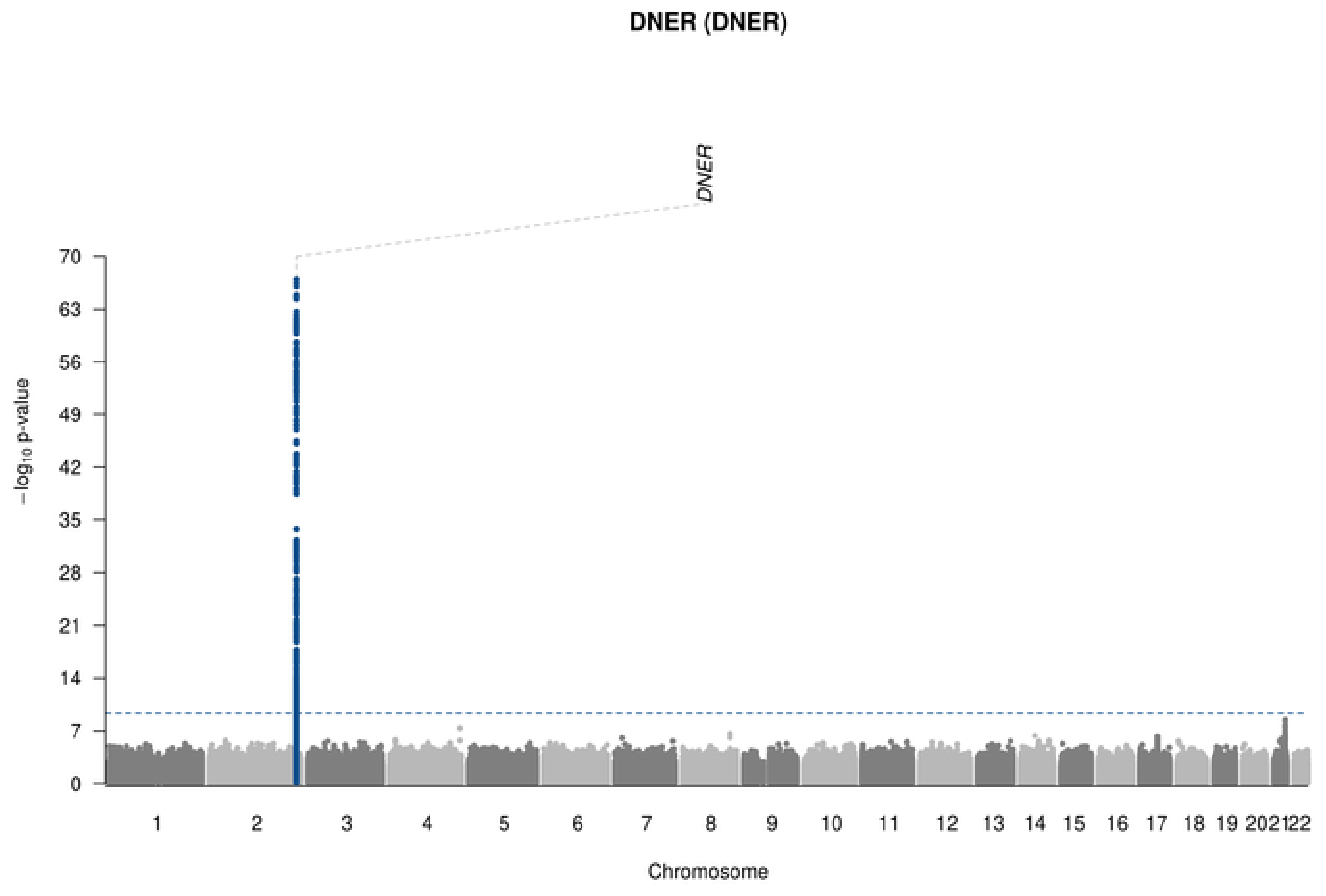
DNER (DNER)
DNER
70
63
56
49
42
35
28
21
14
7
0
−log10 p-value
202122
Chromosome

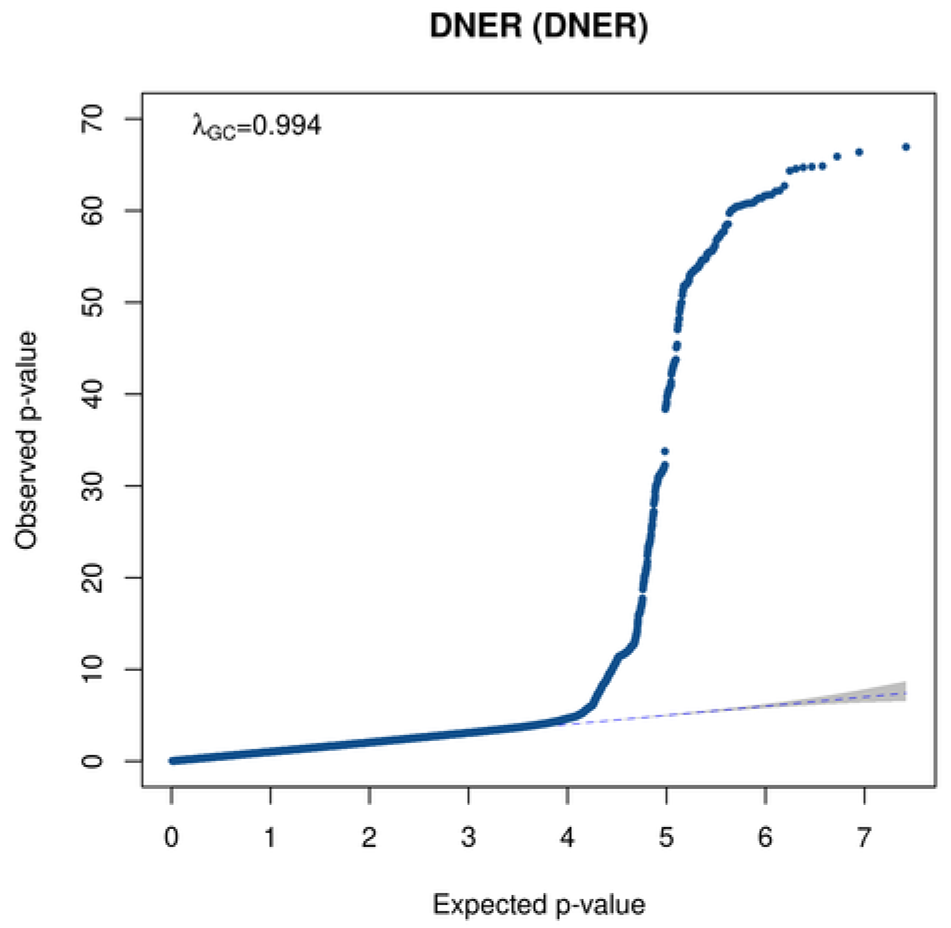
DNER (DNER)
λGC=0.994
70
60
50
40
30
20
10
0
Observed p-value
0
1
2
3
4
5
6
7
Expected p-value

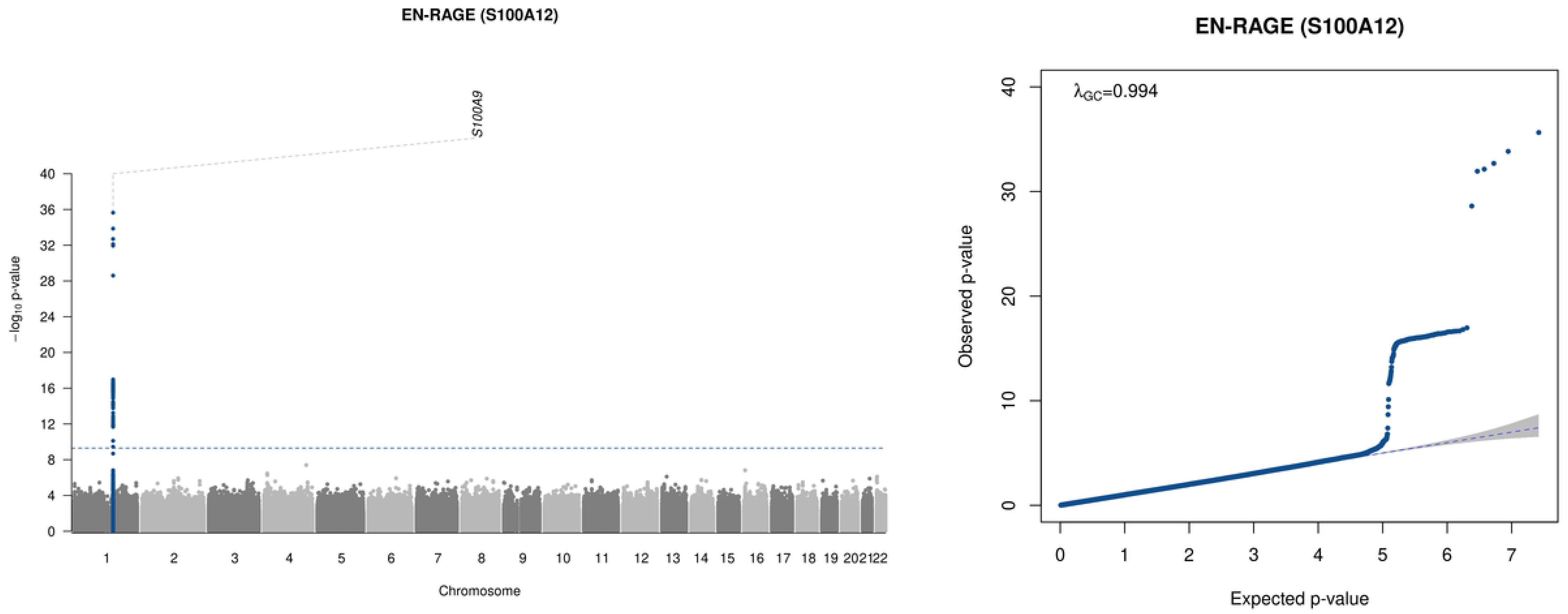

EN-RAGE (S100A12)
S100A9
−log10 p-value
Chromosome
1 2 3 4 5 6 7 8 9 10 11 12 13 14 15 16 17 18 19 20 21 22
0 4 8 12 16 20 24 28 32 36 40
EN-RAGE (S100A12)
λGC=0.994
Observed p-value
Expected p-value
0 1 2 3 4 5 6 7
0 10 20 30 40

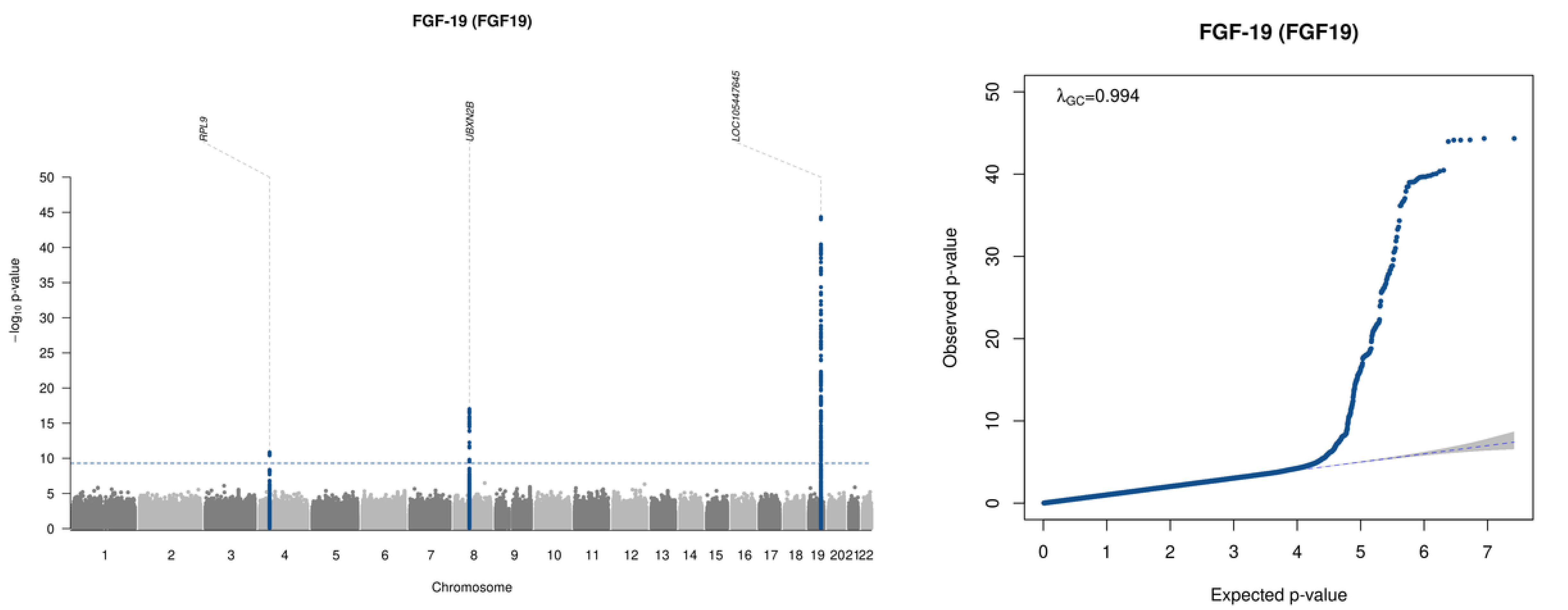

FGF-19 (FGF19)
RPL9
UBXN2B
LOC105447645
−log10 p-value
Chromosome
FGF-19 (FGF19)
λGC=0.994
Observed p-value
Expected p-value

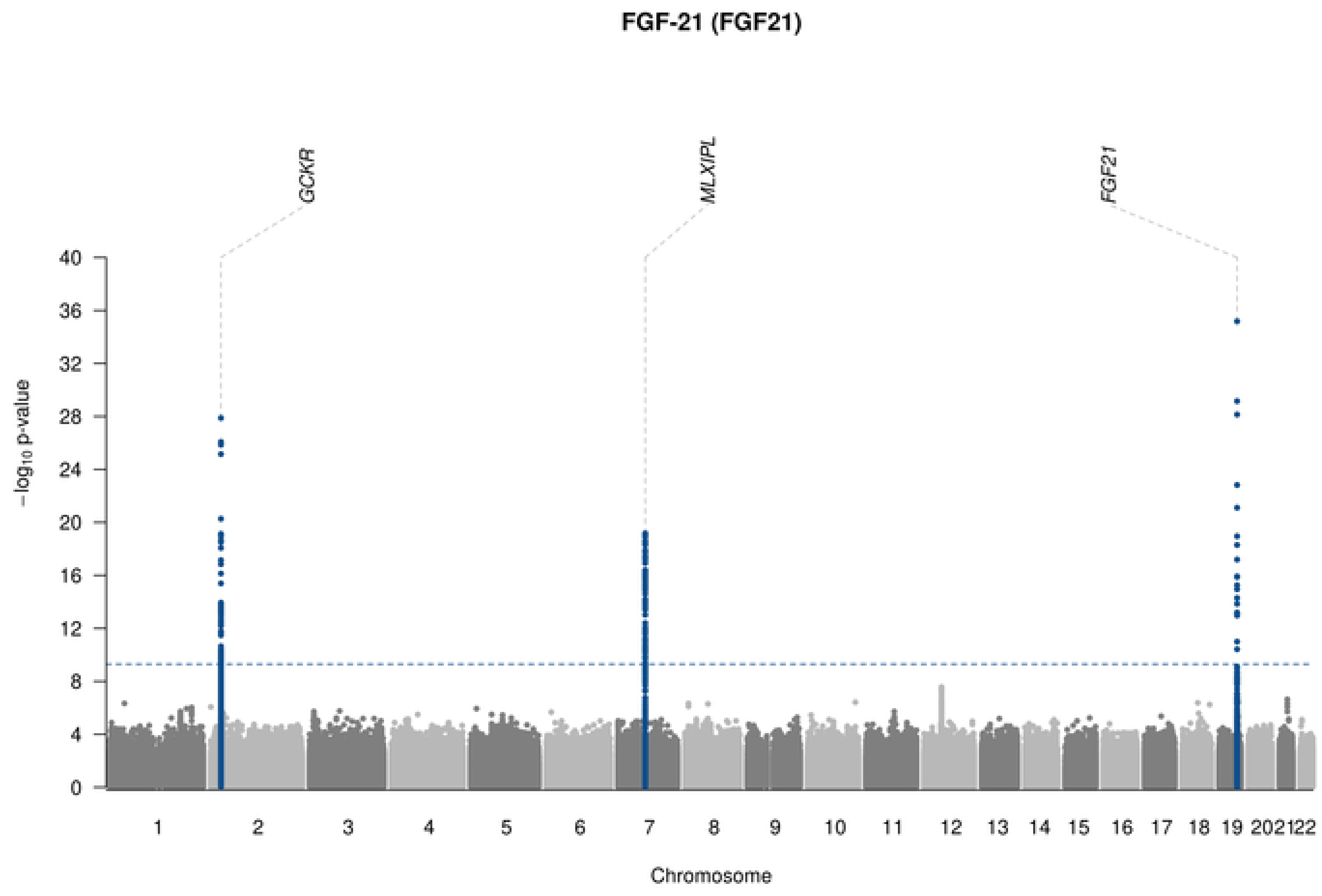

FGF-21 (FGF21)
GCKR
MLXIPL
FGF21
−log10 p-value
Chromosome

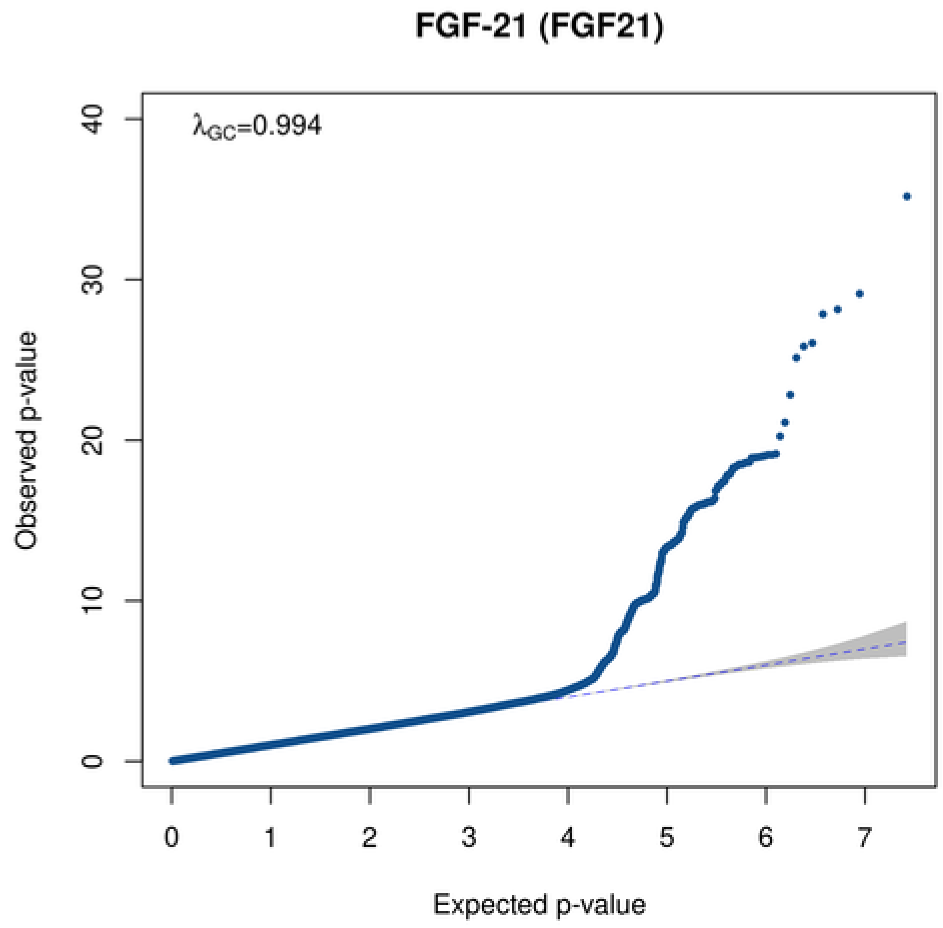

FGF-21 (FGF21)
λGC=0.994
Observed p-value
Expected p-value

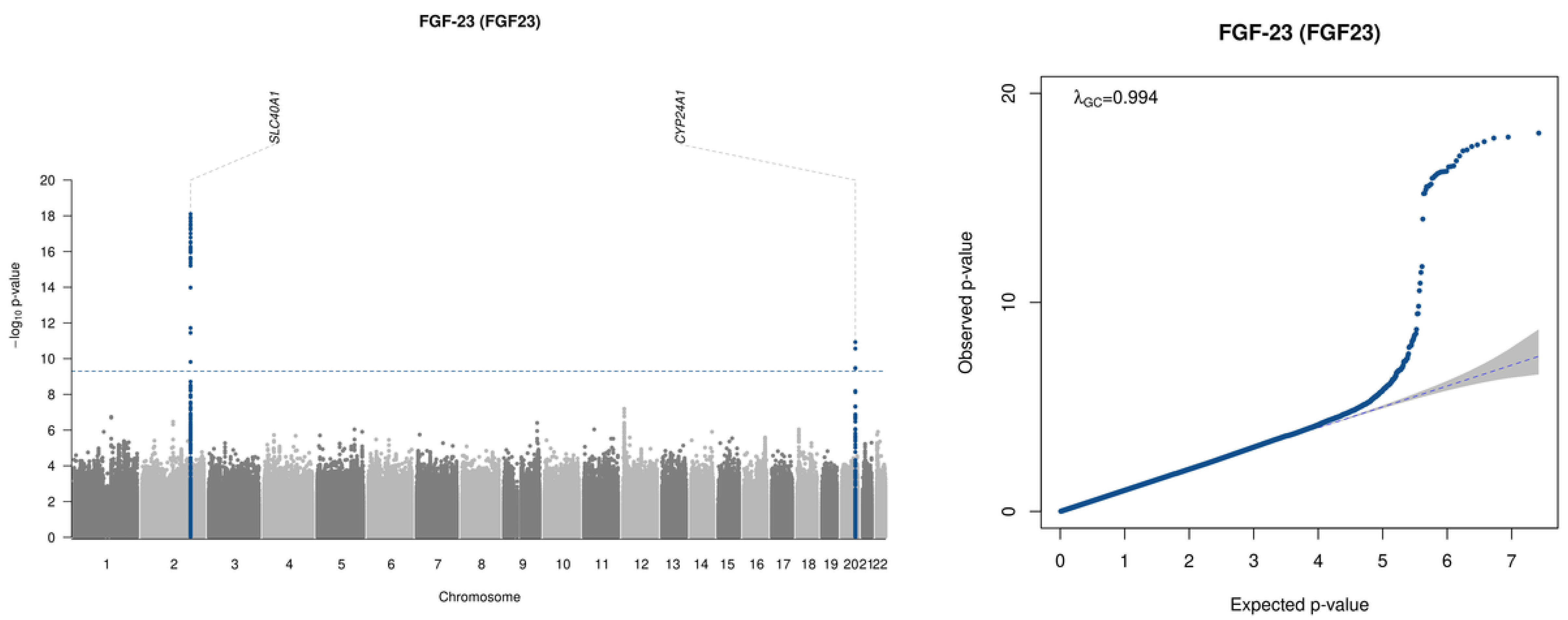

FGF-23 (FGF23)
SLC40A1
CYP24A1
−log10 p-value
Chromosome
FGF-23 (FGF23)
λGC=0.994
Observed p-value
Expected p-value

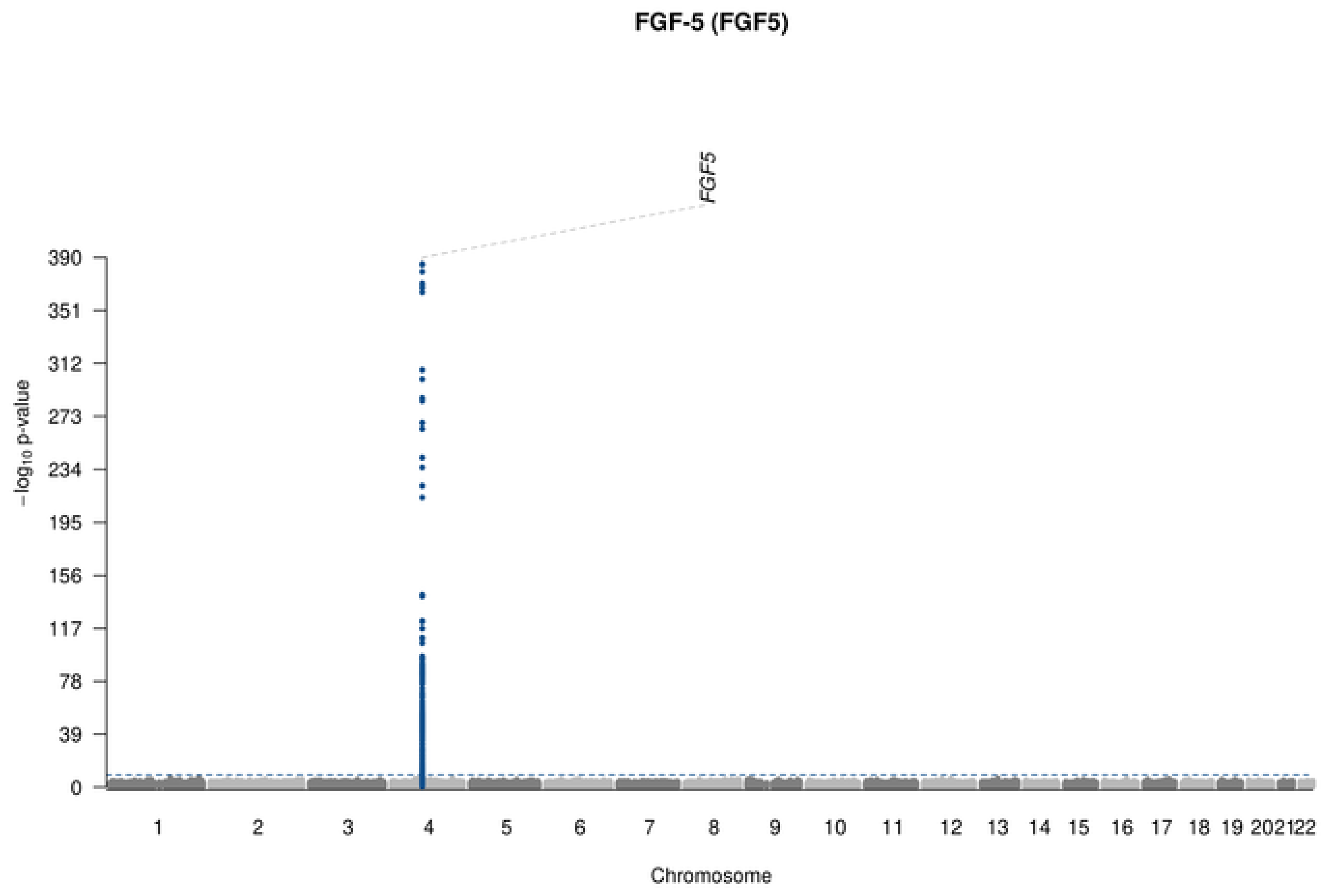

FGF-5 (FGF5)
FGF5
−log10 p-value
390
351
312
273
234
195
156
117
78
39
0
1 2 3 4 5 6 7 8 9 10 11 12 13 14 15 16 17 18 19 202122
Chromosome

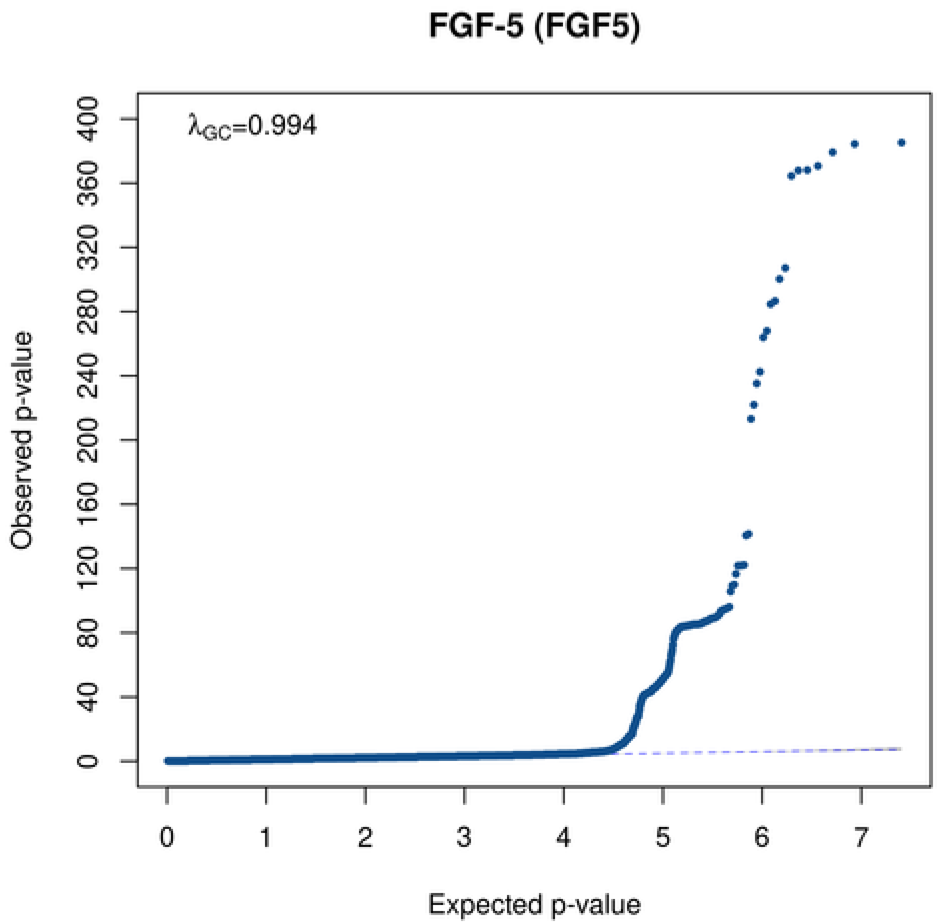

FGF-5 (FGF5)
λGC=0.994
Observed p-value
400
360
320
280
240
200
160
120
80
40
0
0 1 2 3 4 5 6 7
Expected p-value

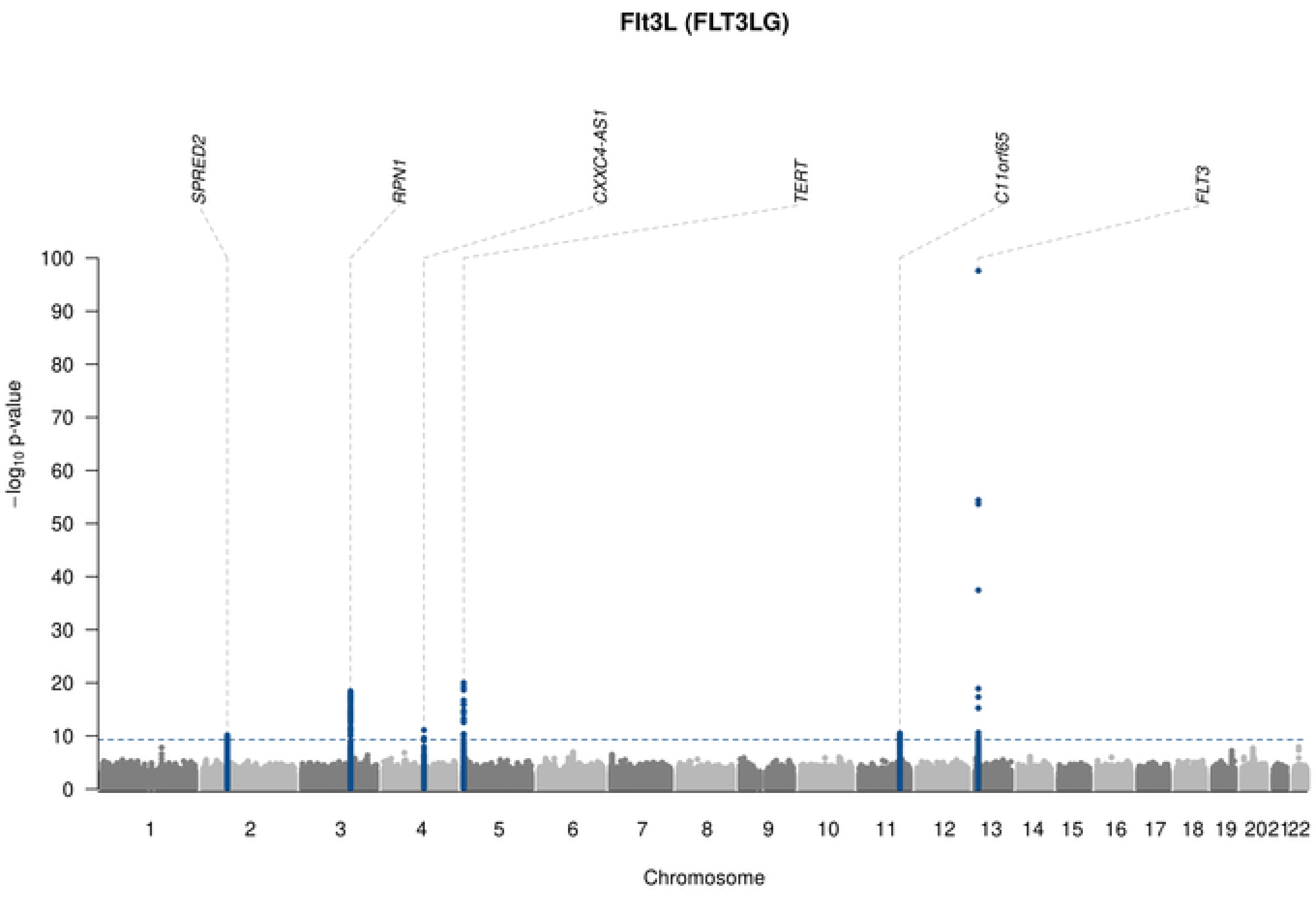
Flt3L (FLT3LG)
SPRED2
RPN1
CXXC4-AS1
TERT
C11orf65
FLT3
−log10 p-value
Chromosome

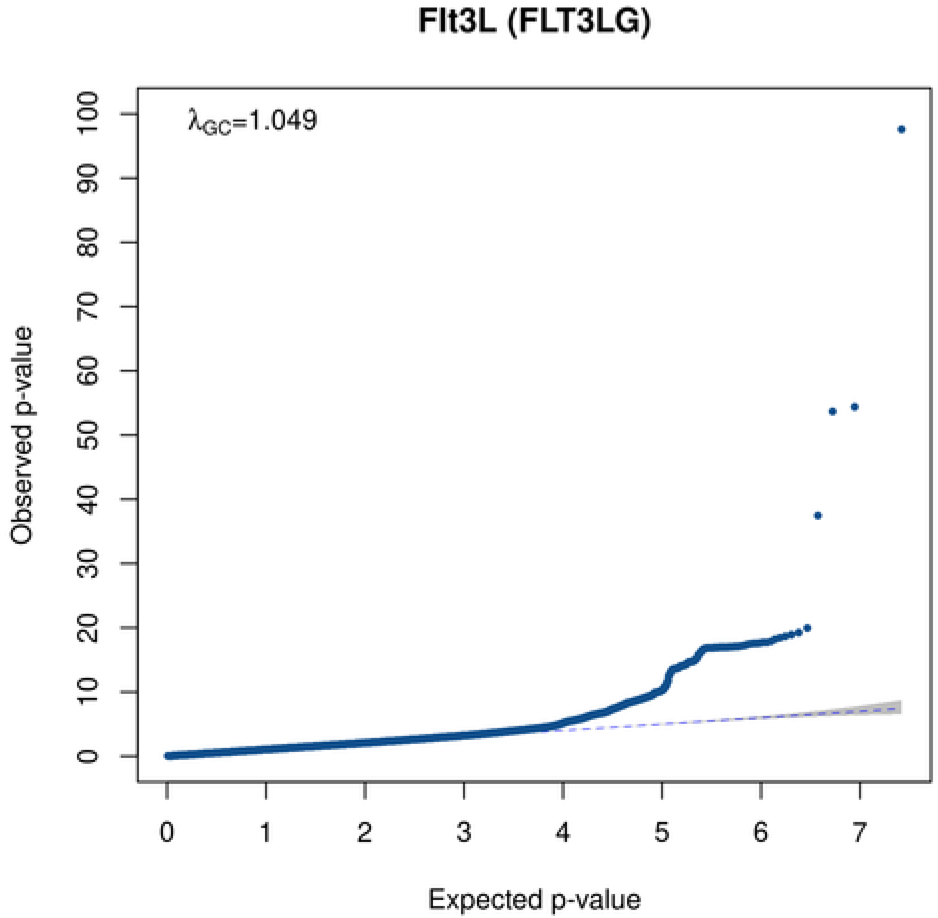
Flt3L (FLT3LG)
λGC=1.049
Observed p-value
Expected p-value

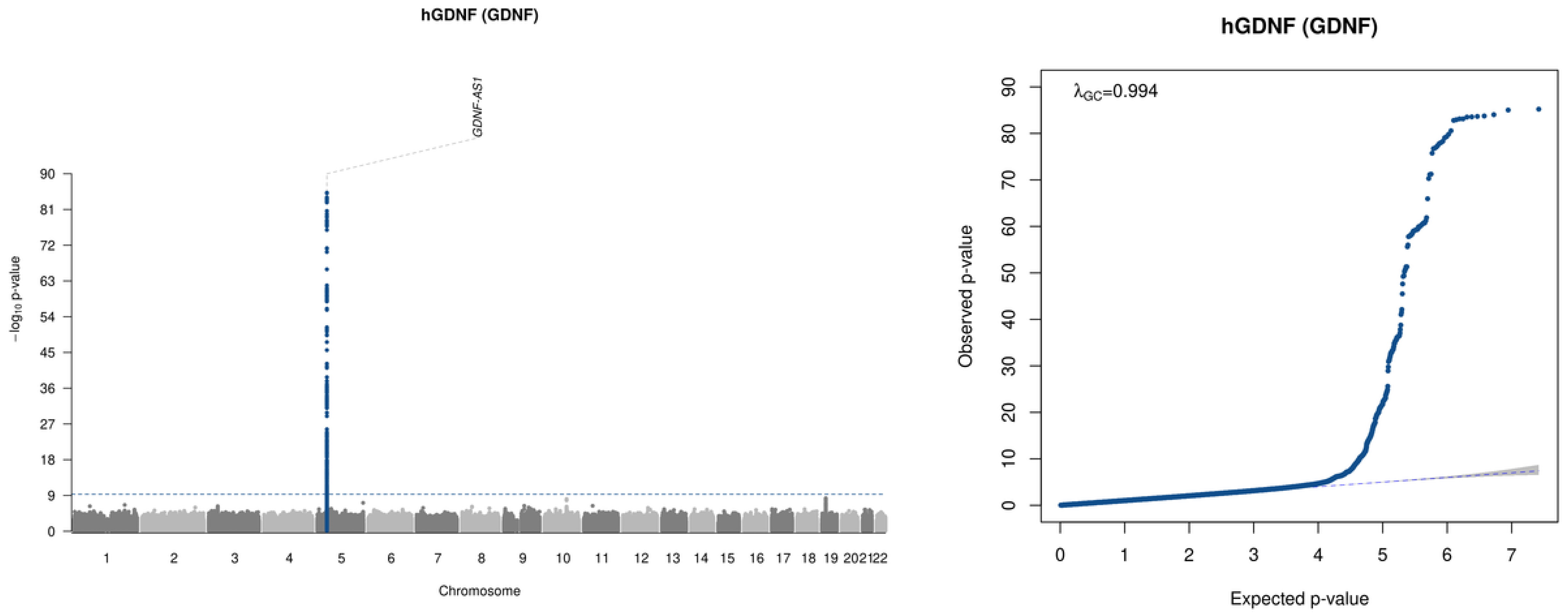
hGDNF (GDNF)
GDNF-AS1
−log10 p-value
Chromosome
hGDNF (GDNF)
λGC=0.994
Observed p-value
Expected p-value

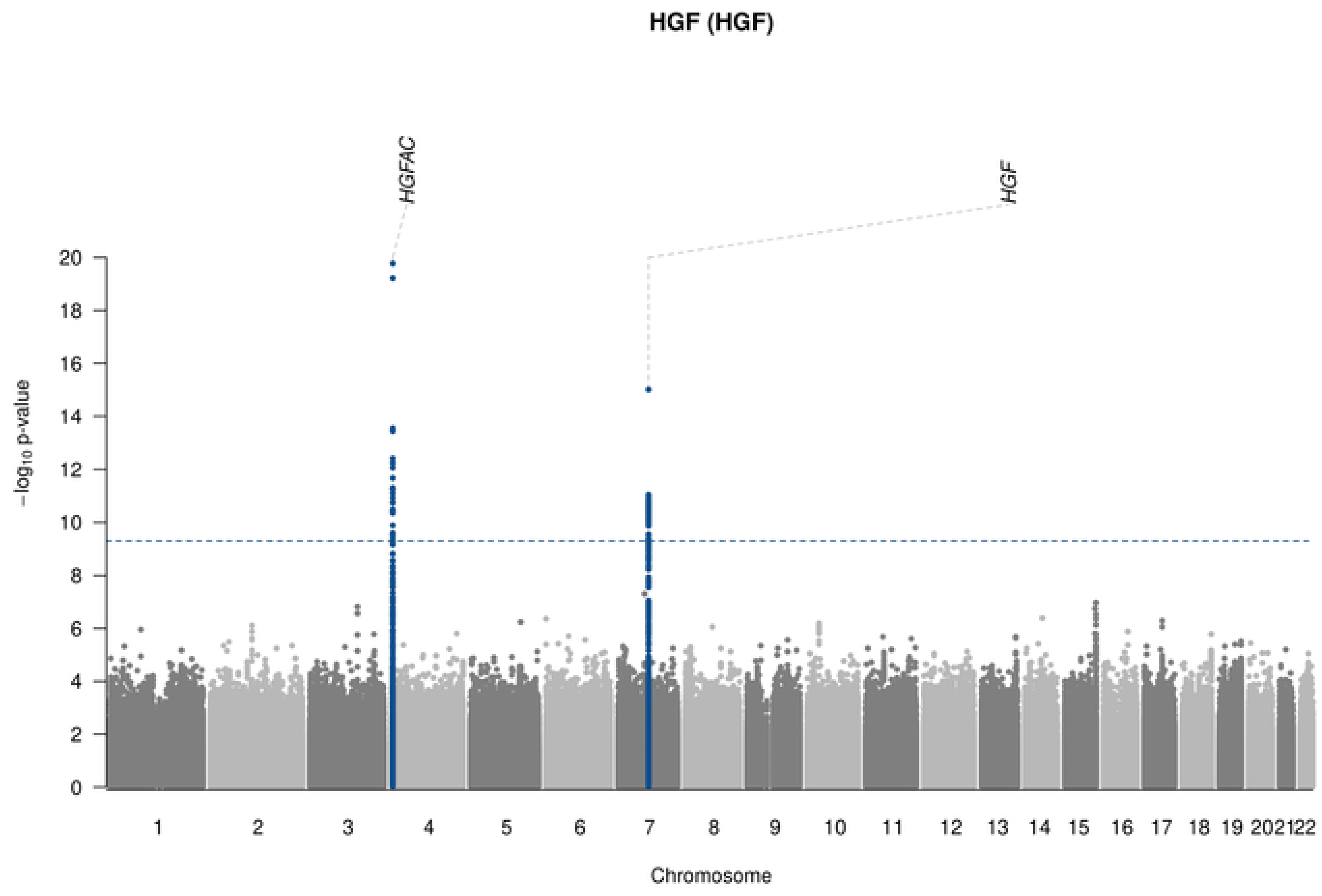
HGF (HGF)
HGFAC
HGF
−log10 p-value
Chromosome

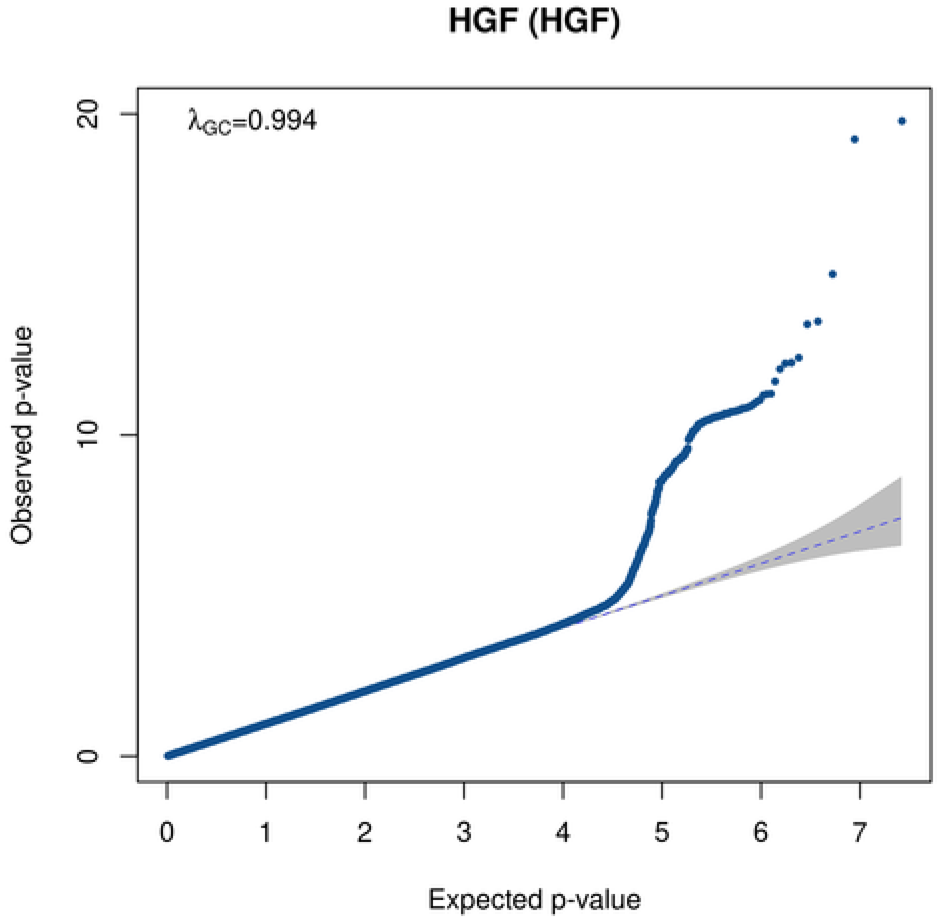
HGF (HGF)
λGC=0.994
Observed p-value
Expected p-value

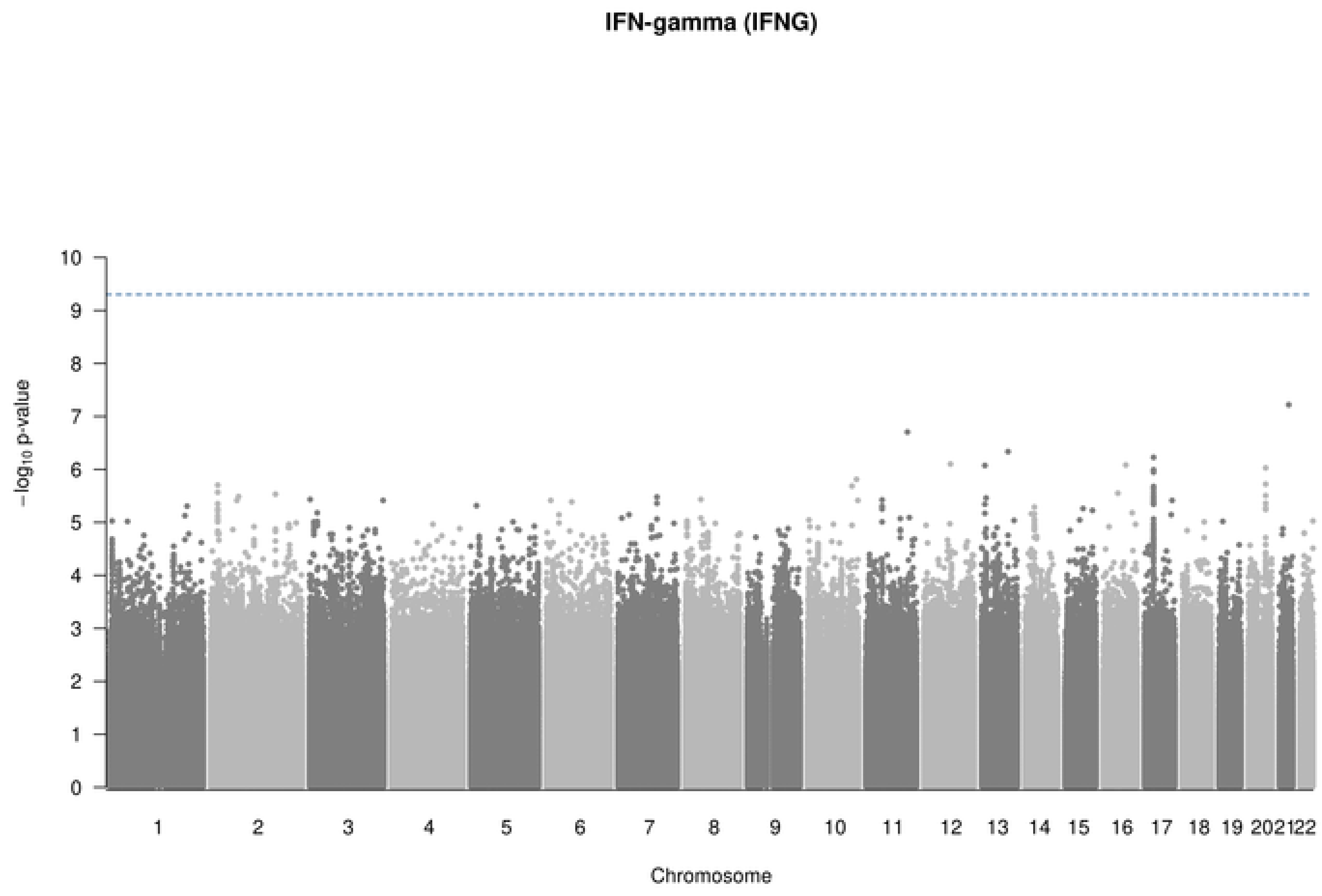
IFN-gamma (IFNG)
$-\log_{10}$ p-value
Chromosome
202122

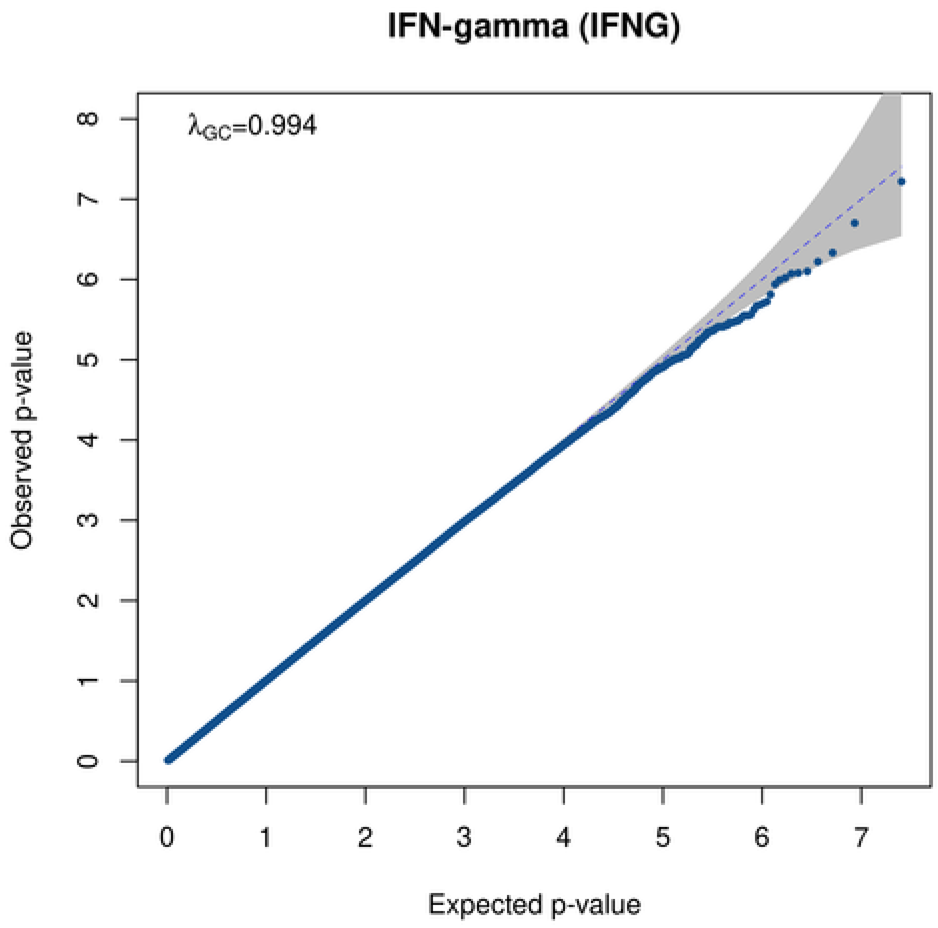
IFN-gamma (IFNG)
$\lambda_{GC}$=0.994
Observed p-value
Expected p-value

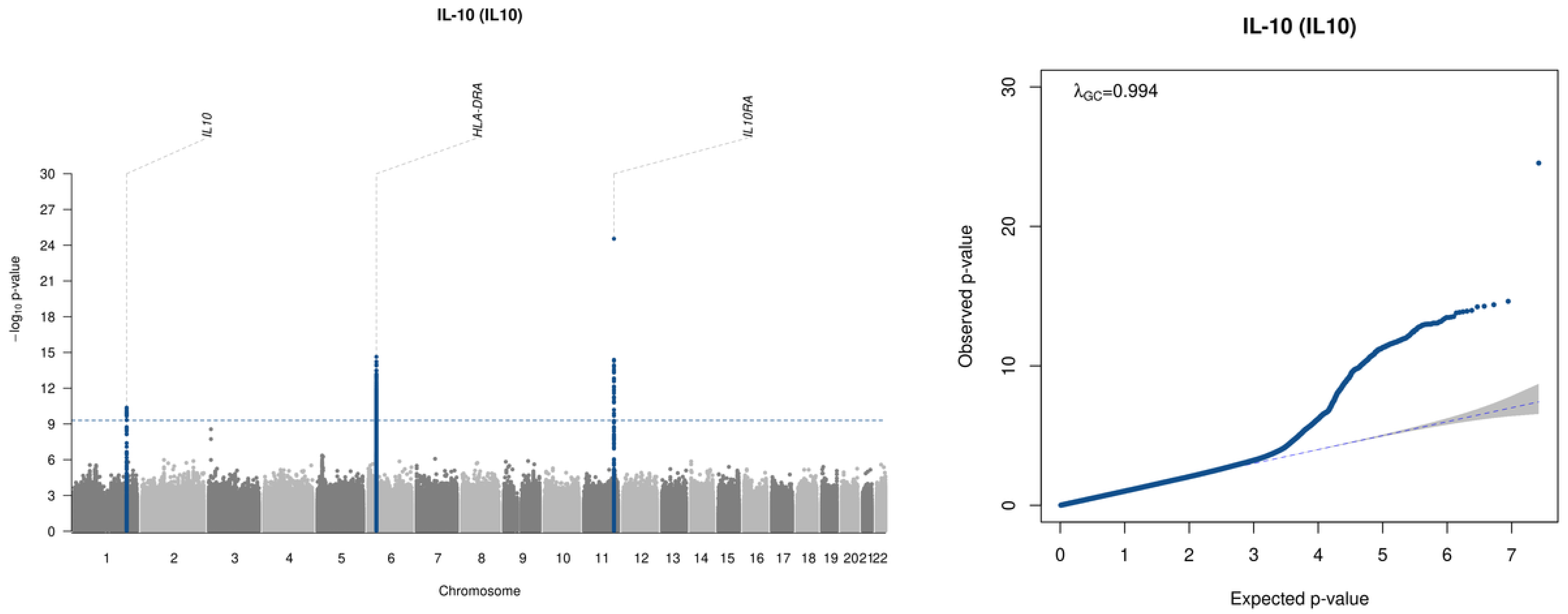
IL-10 (IL10)
IL10
HLA-DRA
IL10RA
−log10 p-value
30
27
24
21
18
15
12
9
6
3
202122
Chromosome
IL-10 (IL10)
λGC=0.994
Observed p-value
Expected p-value
0
10
20
30
0
1
2
3
4
5
6
7

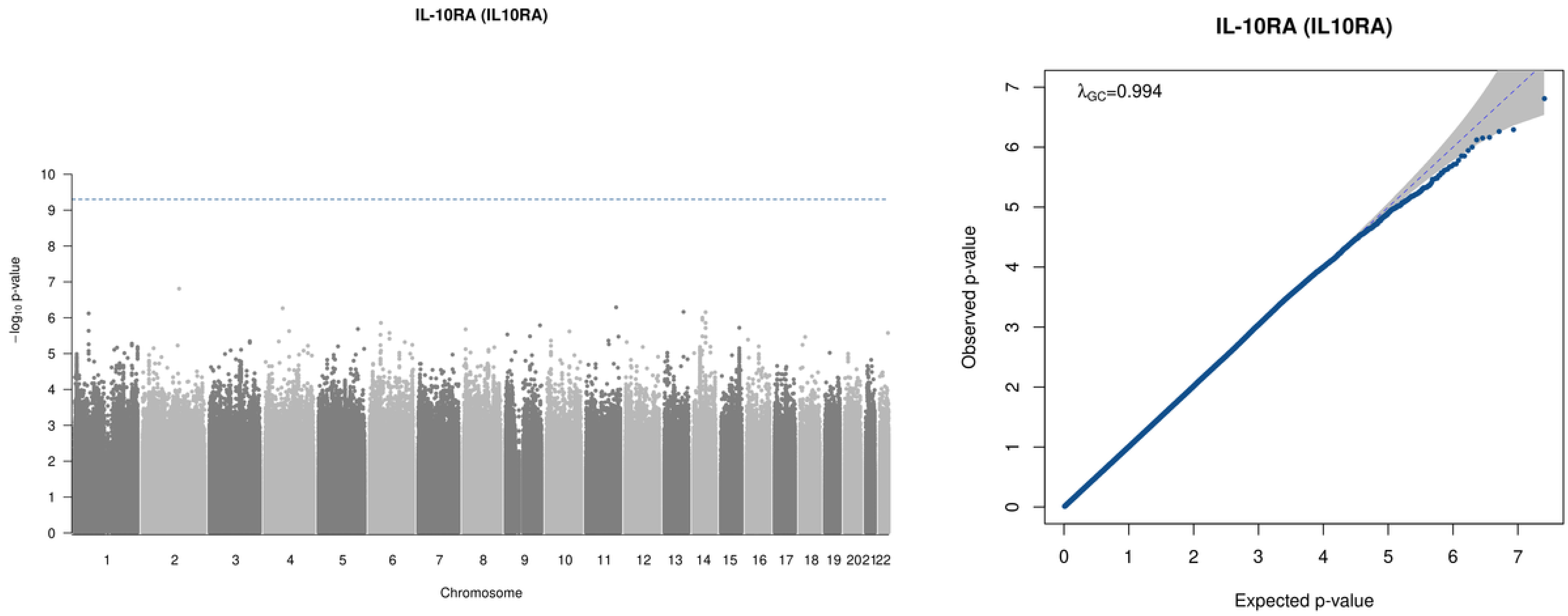

IL-10RA (IL10RA)
−log10 p-value
Chromosome
IL-10RA (IL10RA)
λGC=0.994
Observed p-value
Expected p-value

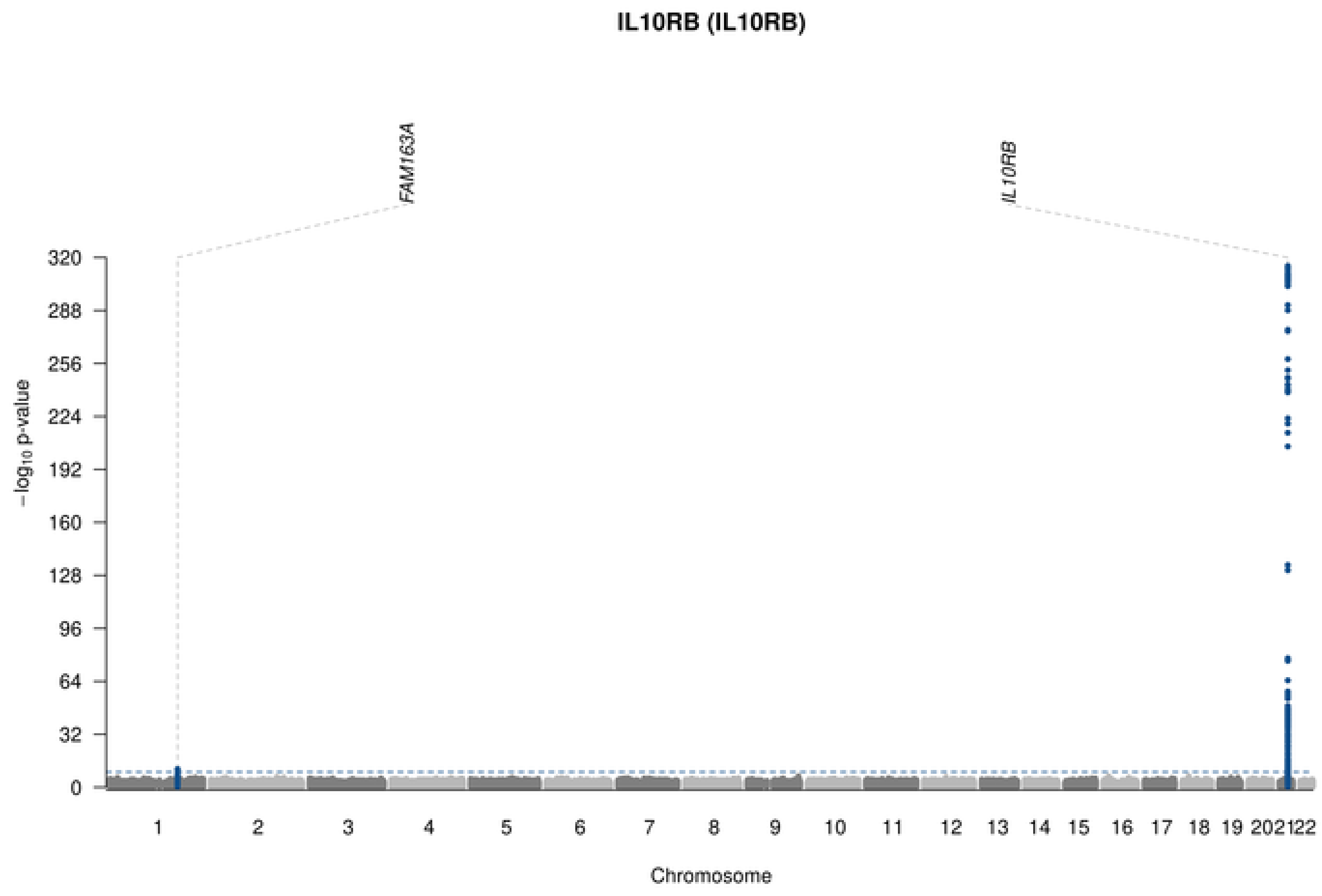
IL10RB (IL10RB)
FAM163A
IL10RB
−log10 p-value
320
288
256
224
192
160
128
96
64
32
202122
Chromosome

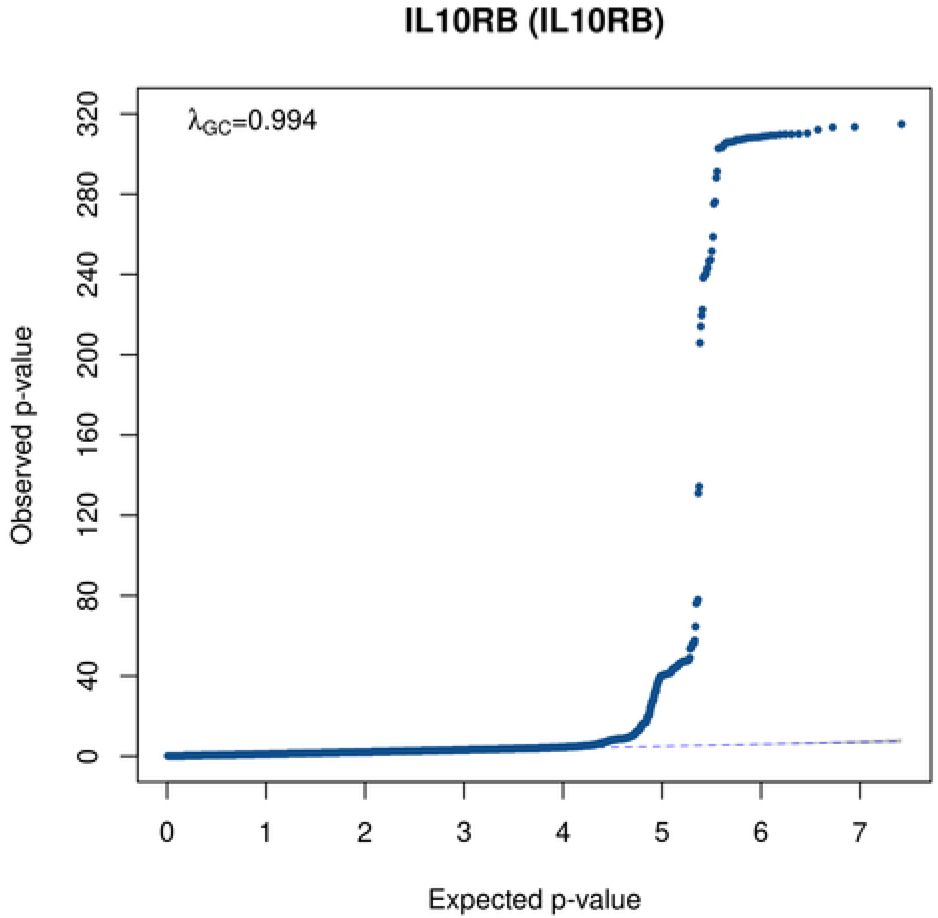
IL10RB (IL10RB)
λGC=0.994
Observed p-value
Expected p-value
0
40
80
120
160
200
240
280
320
0
1
2
3
4
5
6
7

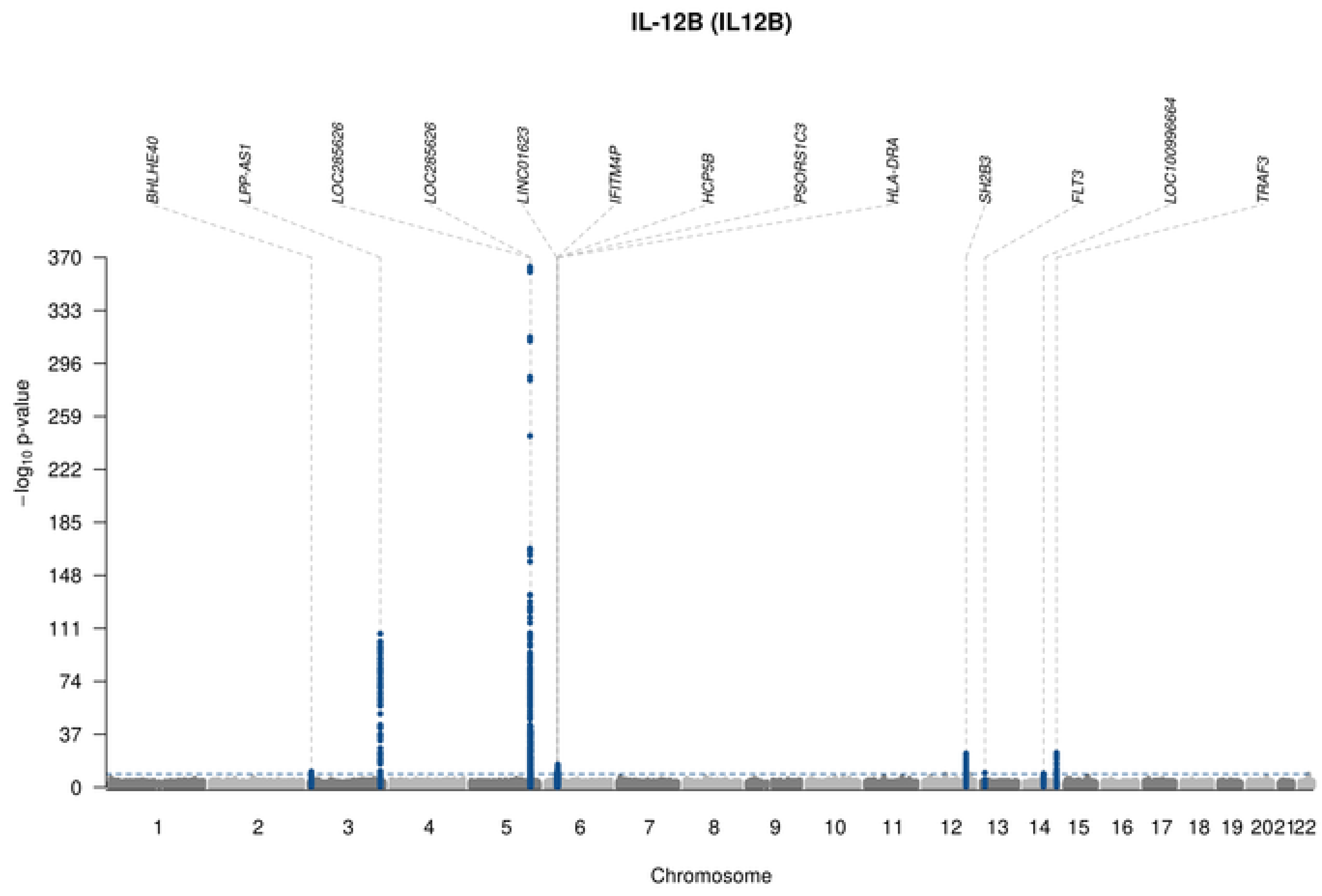

IL-12B (IL12B)
BHLHE40
LPP-AS1
LOC285626
LOC285626
LINC01623
IFITM4P
HCP5B
PSORS1C3
HLA-DRA
SH2B3
FLT3
LOC100996664
TRAF3
370
333
296
259
222
185
148
111
74
37
0
$-\log_{10}$ p-value
1 2 3 4 5 6 7 8 9 10 11 12 13 14 15 16 17 18 19 20 21 22
Chromosome

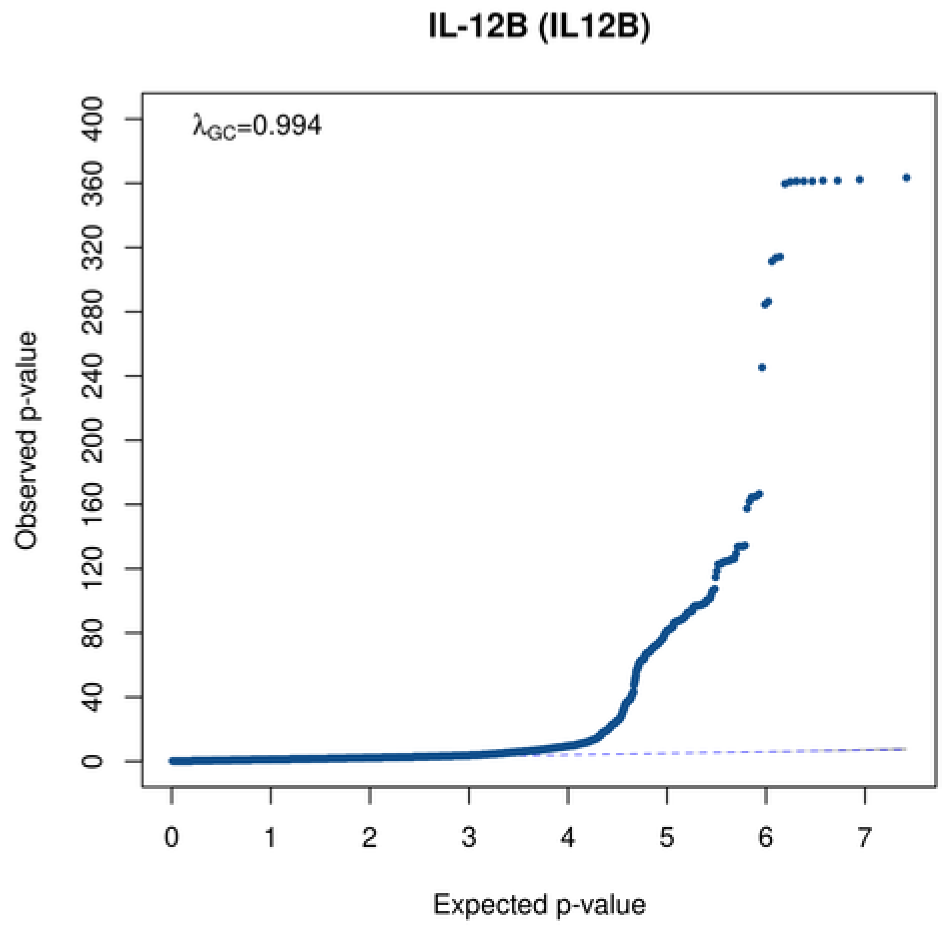

IL-12B (IL12B)
$\lambda_{GC}$=0.994
400
360
320
280
240
200
160
120
80
40
0
Observed p-value
0 1 2 3 4 5 6 7
Expected p-value

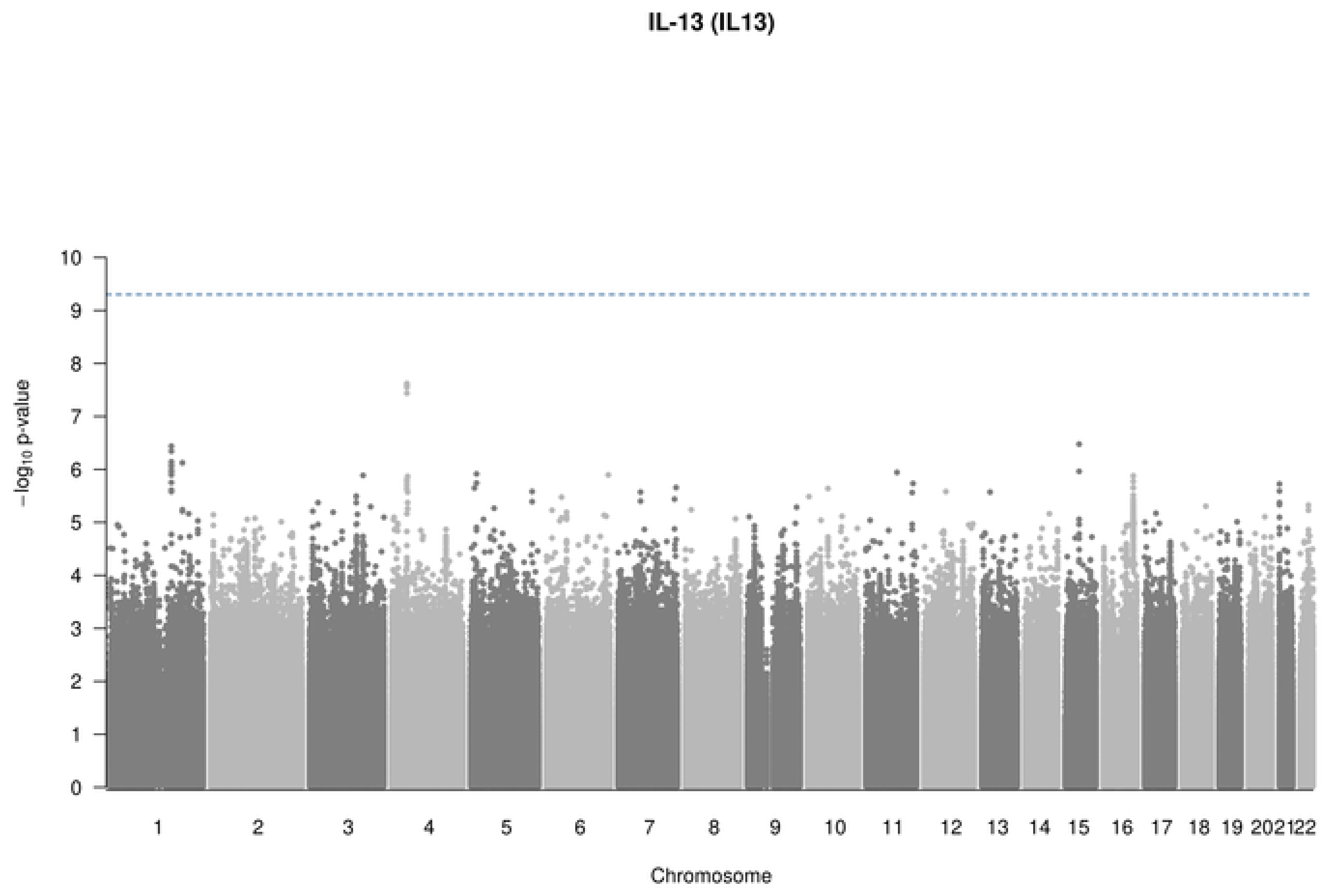
IL-13 (IL13)
−log10 p-value
10
9
8
7
6
5
4
3
2
1
0
1 2 3 4 5 6 7 8 9 10 11 12 13 14 15 16 17 18 19 20 21 22
Chromosome

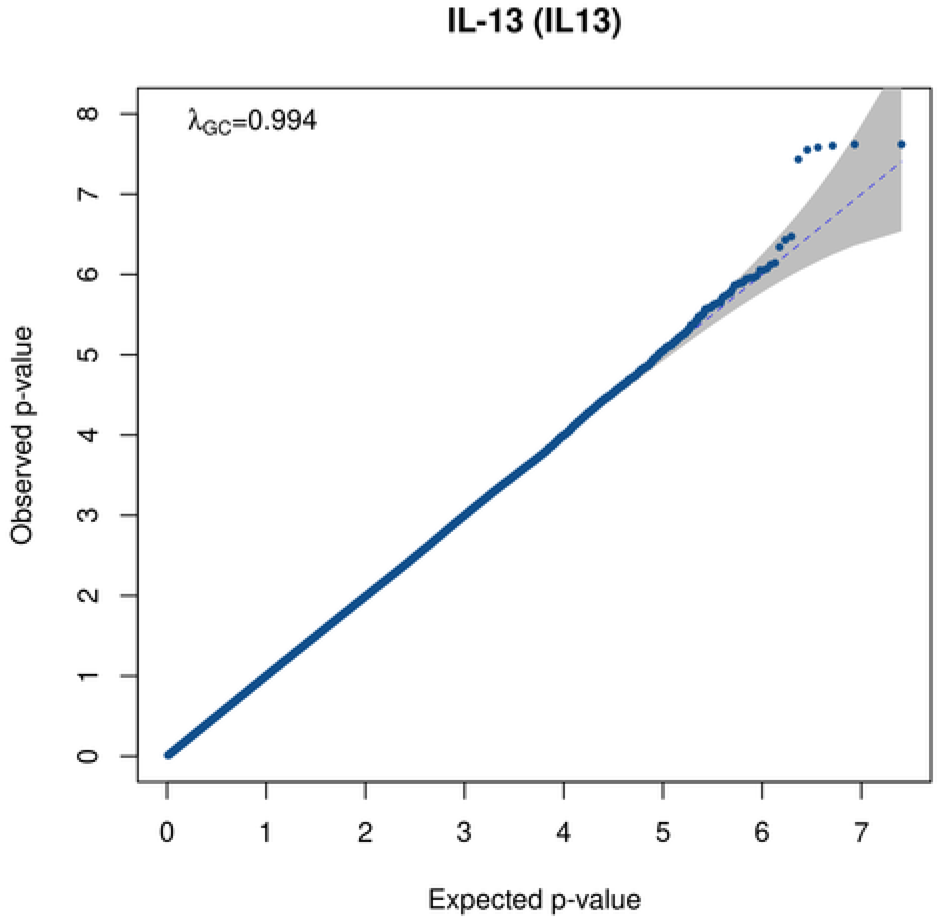
IL-13 (IL13)
λGC=0.994
Observed p-value
Expected p-value
0 1 2 3 4 5 6 7
0 1 2 3 4 5 6 7 8

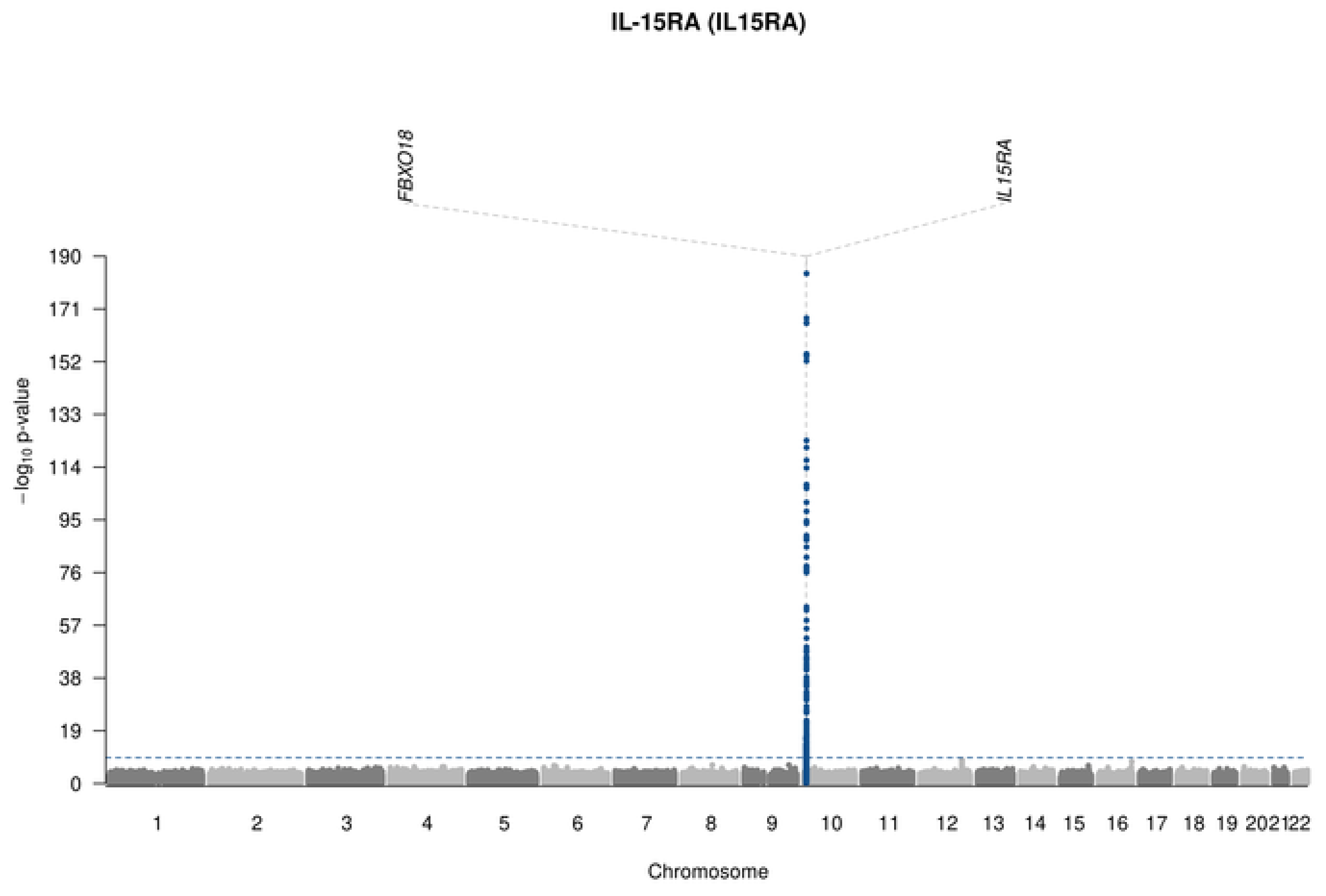
IL-15RA (IL15RA)
FBXO18
IL15RA
−log10 p-value
190
171
152
133
114
95
76
57
38
19
202122
Chromosome
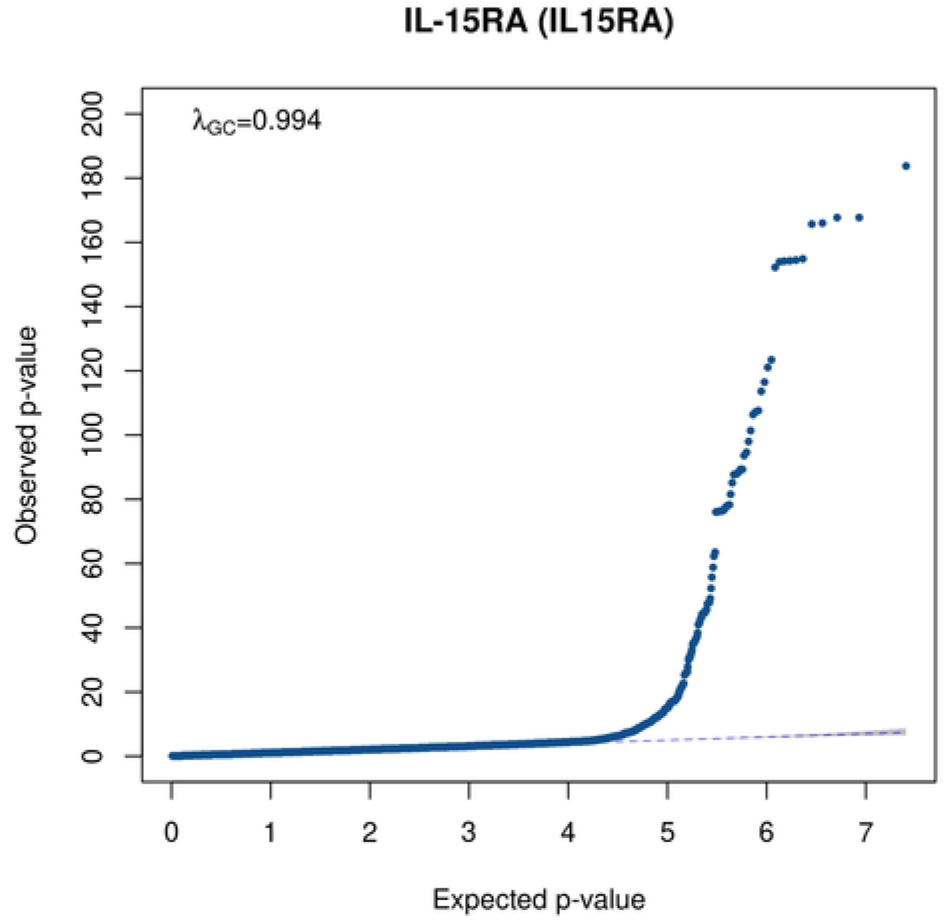
IL-15RA (IL15RA)
λGC=0.994
Observed p-value
Expected p-value
0
20
40
60
80
100
120
140
160
180
200
0
1
2
3
4
5
6
7

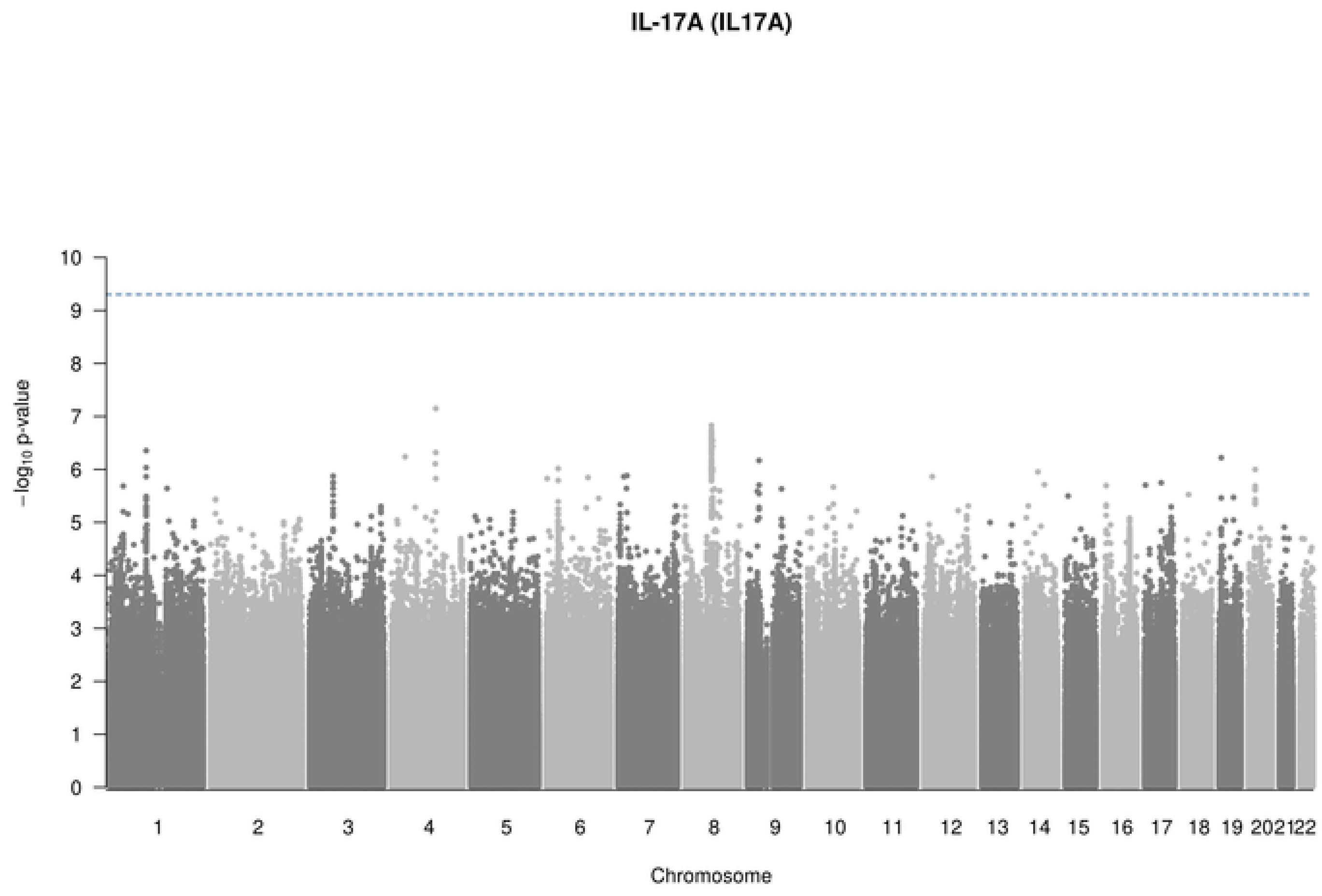

IL-17A (IL17A)
10
9
8
7
6
5
4
3
2
1
0
−log10 p-value
202122
Chromosome

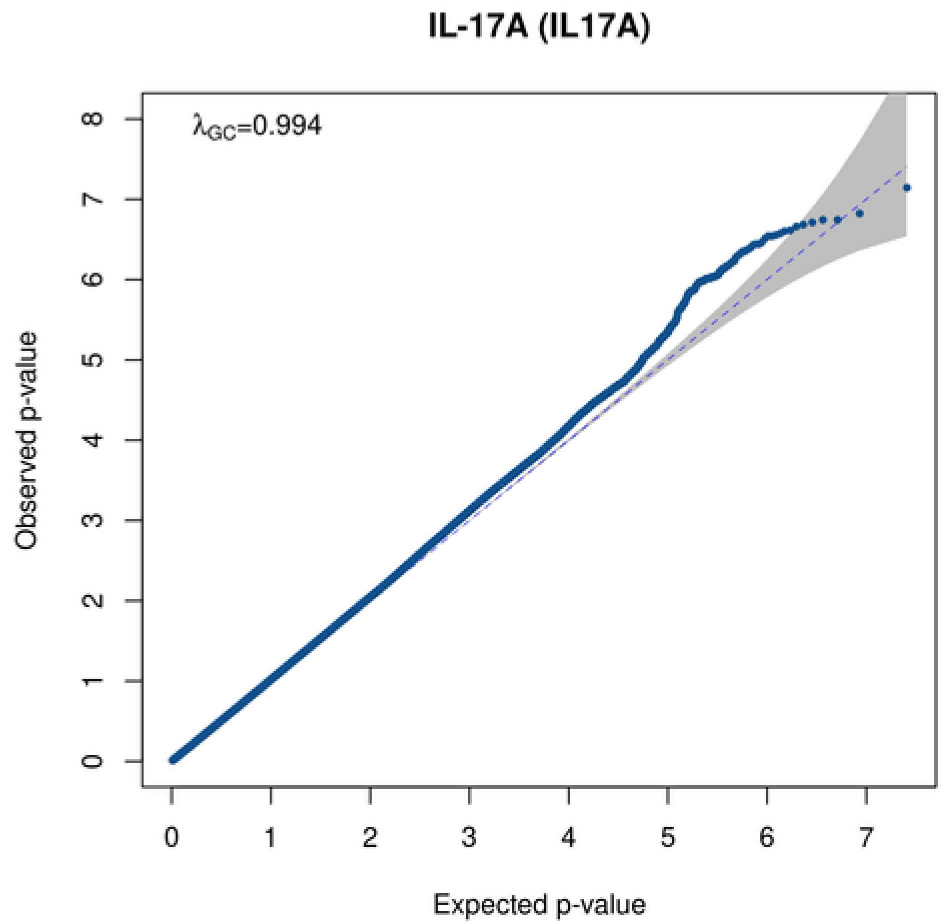

IL-17A (IL17A)
λGC=0.994
0
1
2
3
4
5
6
7
8
Observed p-value
0
1
2
3
4
5
6
7
Expected p-value

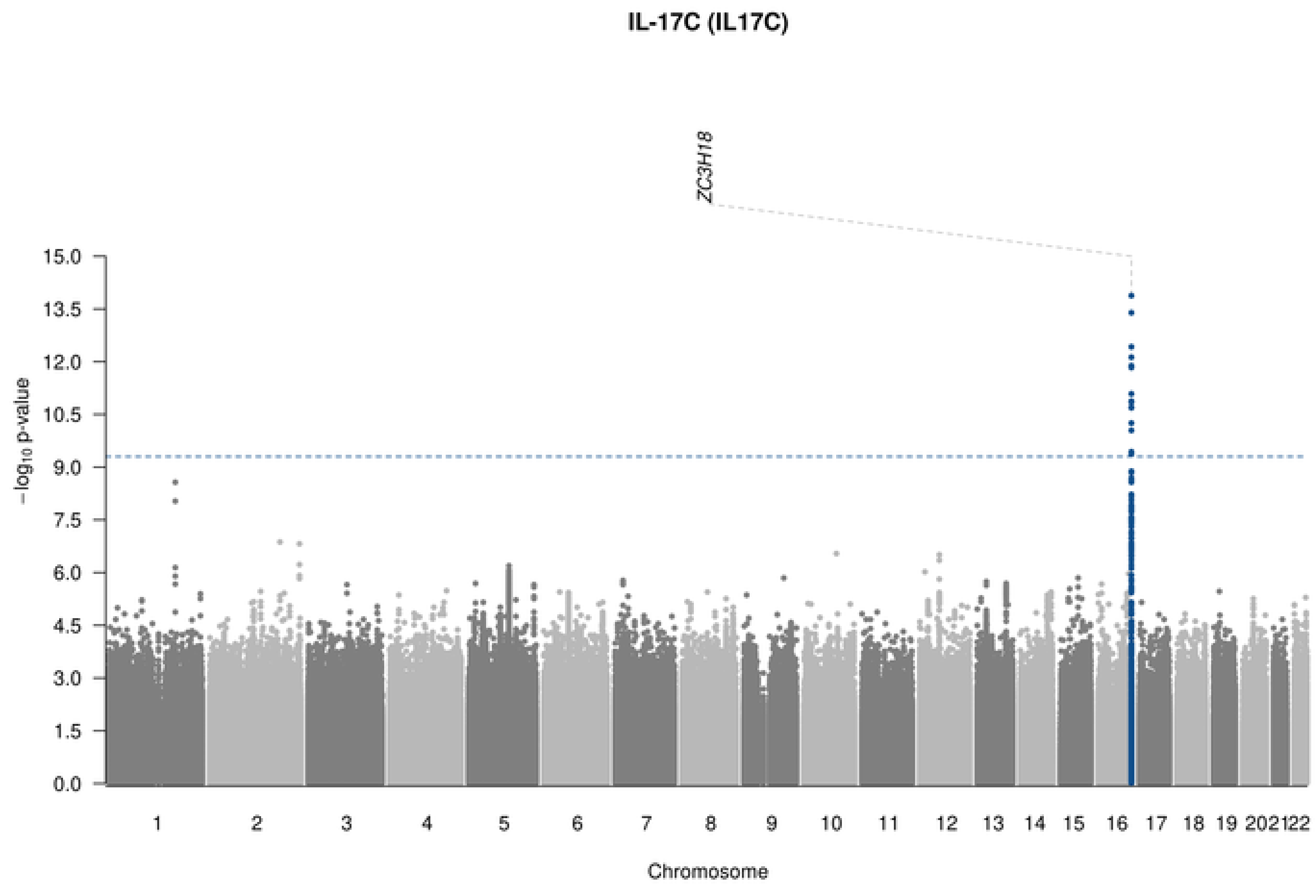

IL-17C (IL17C)
ZC3H18
−log10 p-value
15.0
13.5
12.0
10.5
9.0
7.5
6.0
4.5
3.0
1.5
0.0
202122
Chromosome

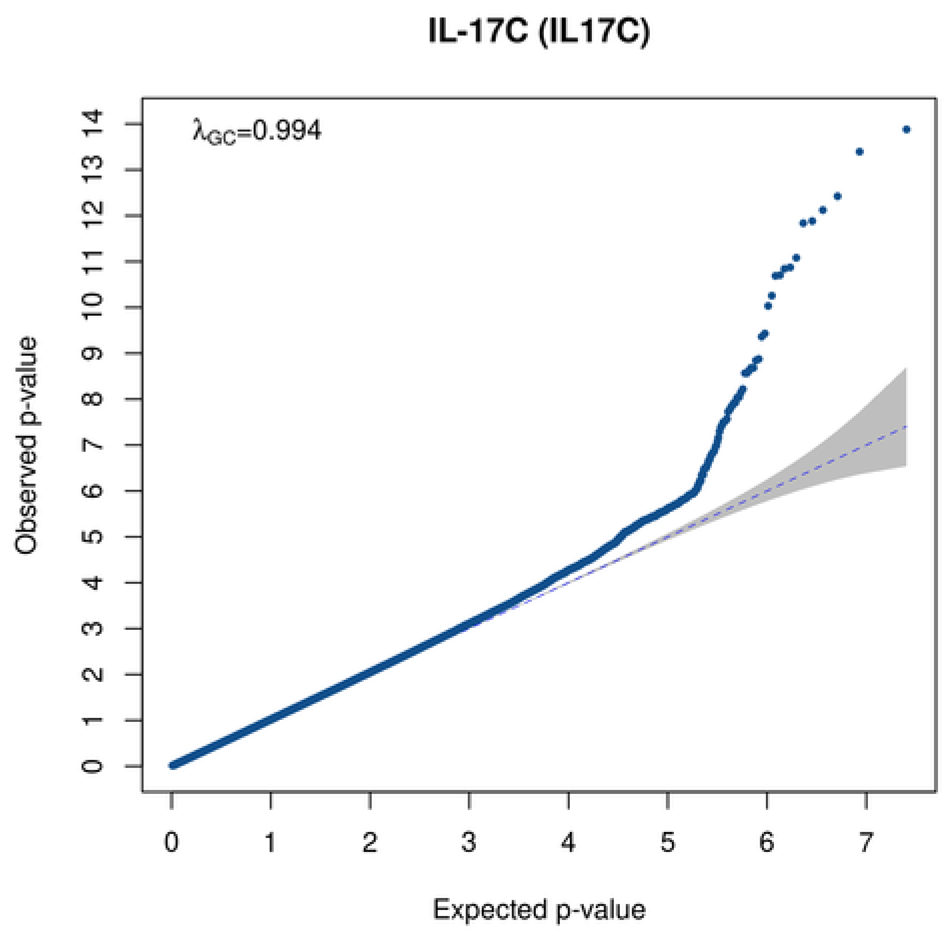

IL-17C (IL17C)
λGC=0.994
Observed p-value
Expected p-value

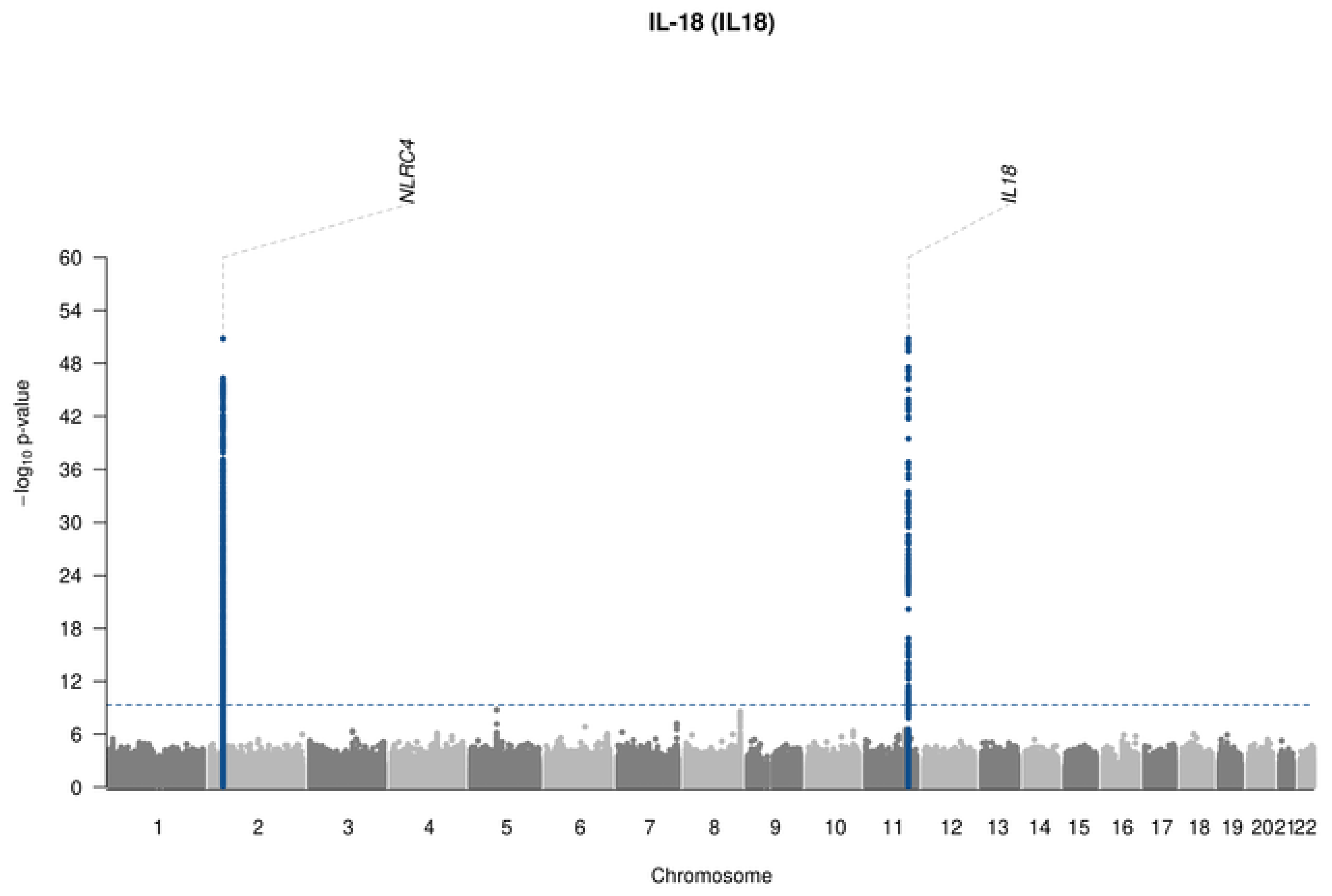
IL-18 (IL18)
NLRC4
IL18
−log10 p-value
60
54
48
42
36
30
24
18
12
6
0
1 2 3 4 5 6 7 8 9 10 11 12 13 14 15 16 17 18 19 202122
Chromosome

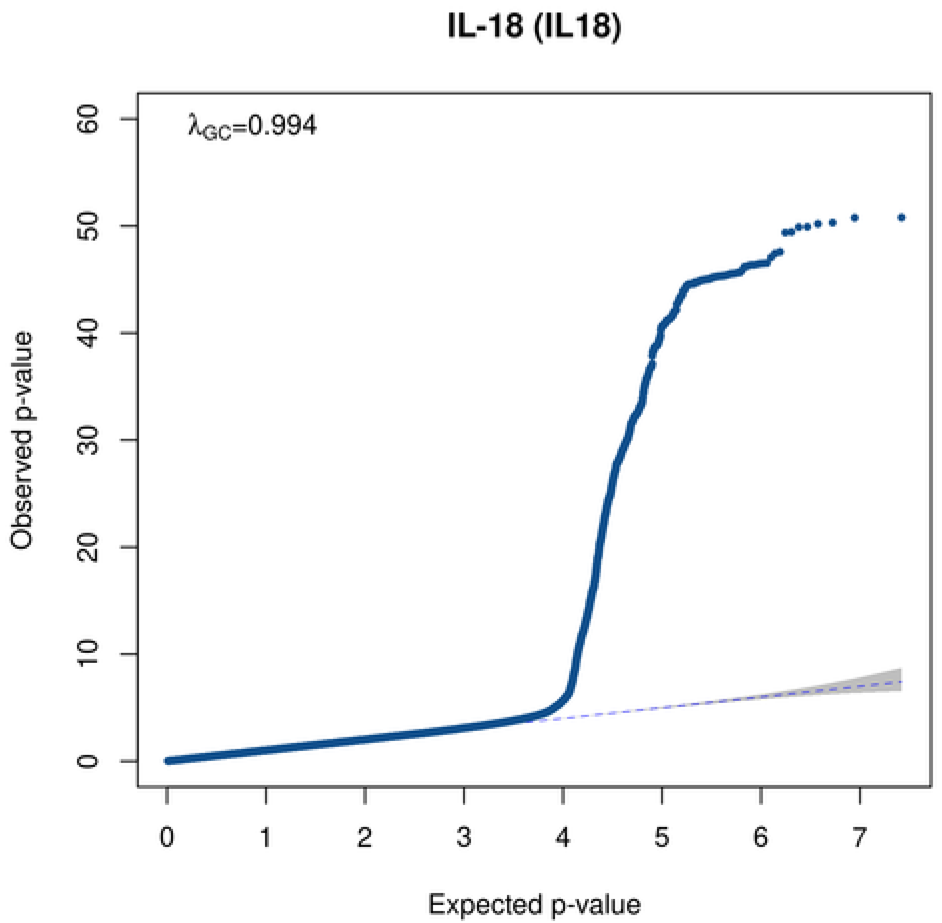
IL-18 (IL18)
λGC=0.994
Observed p-value
60
50
40
30
20
10
0
0 1 2 3 4 5 6 7
Expected p-value

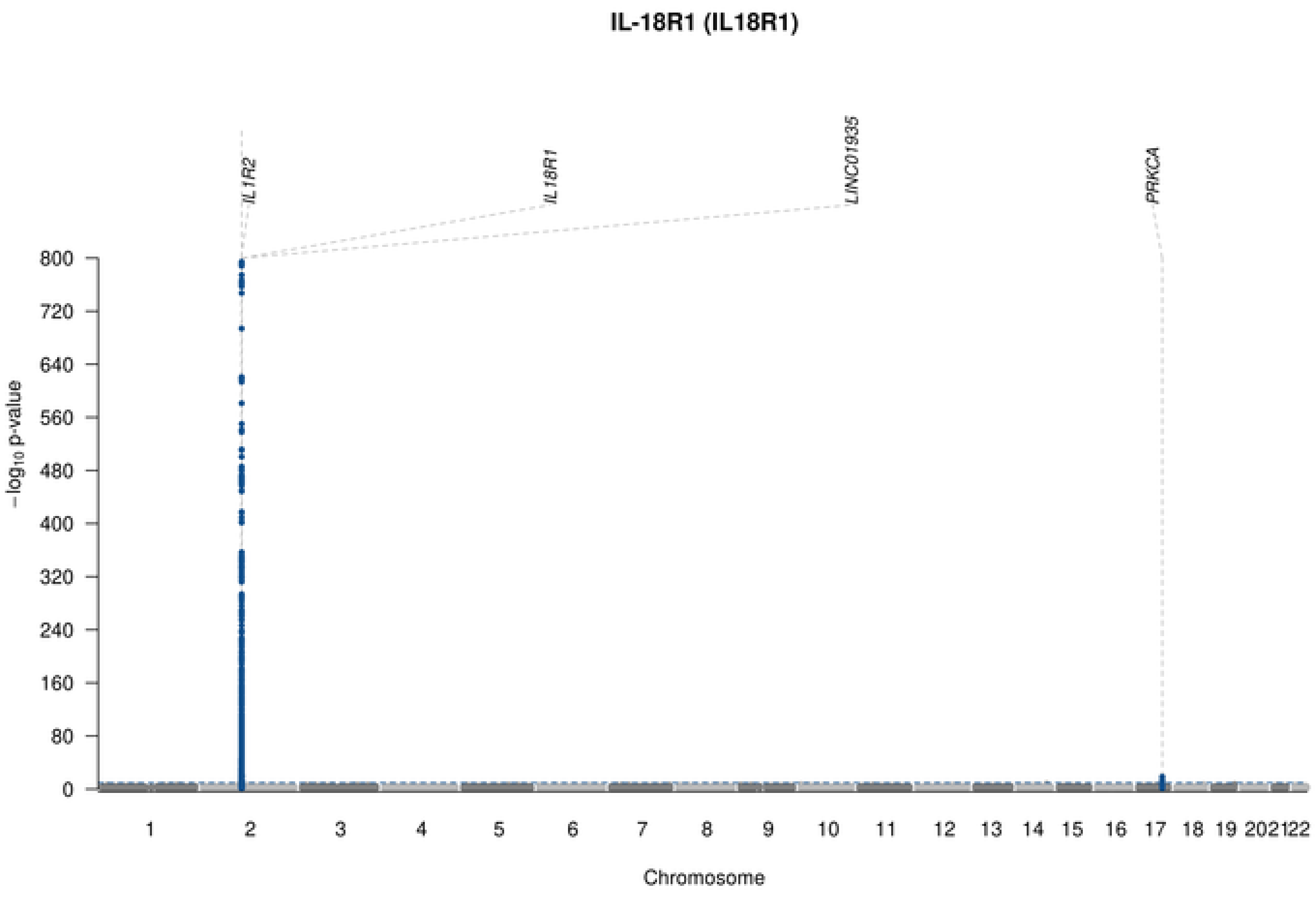

IL-18R1 (IL18R1)
IL1R2
IL18R1
LINC01935
PRKCA
−log10 p-value
Chromosome

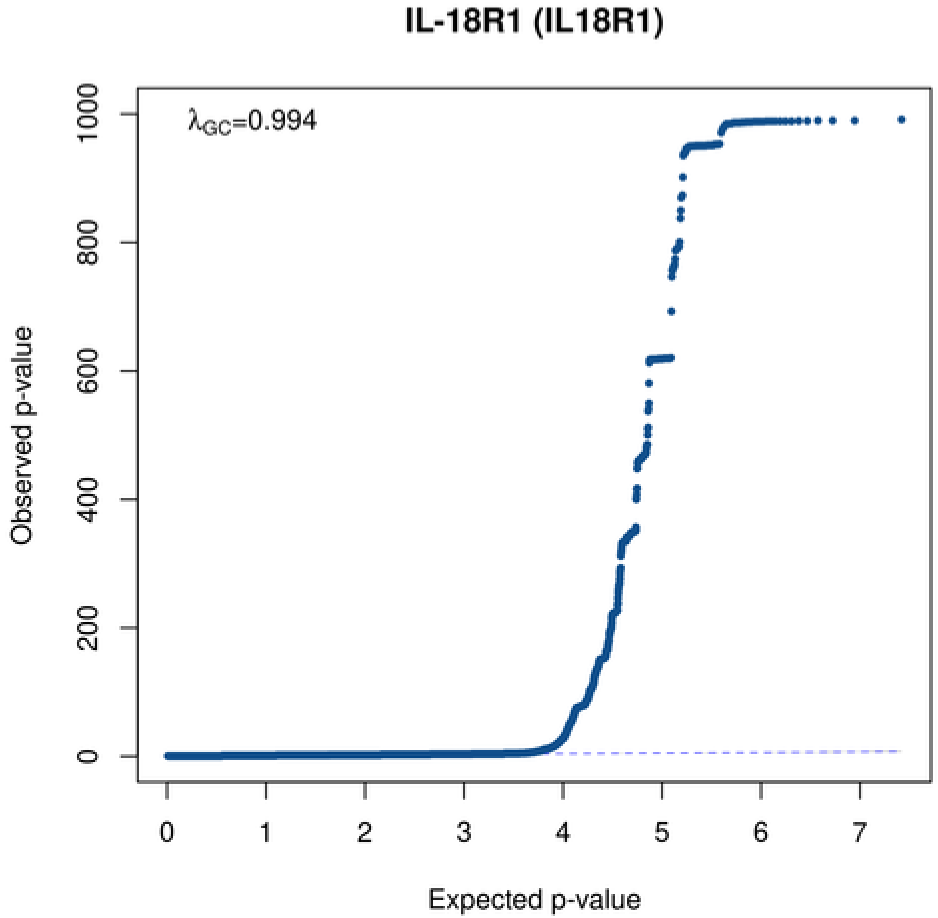

IL-18R1 (IL18R1)
λGC=0.994
Observed p-value
Expected p-value

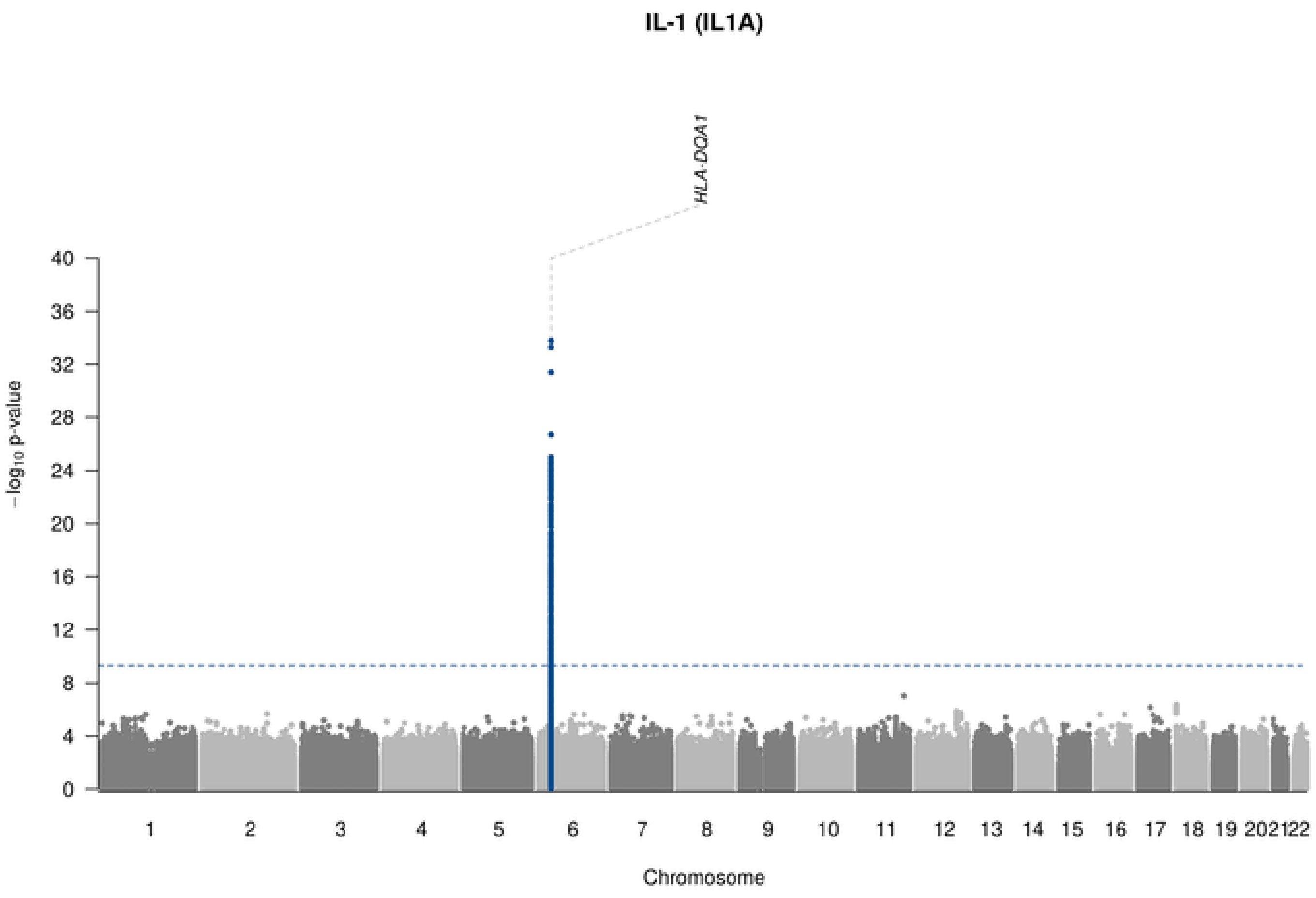
IL-1 (IL1A)
HLA-DQA1
−log10 p-value
0
4
8
12
16
20
24
28
32
36
40
1 2 3 4 5 6 7 8 9 10 11 12 13 14 15 16 17 18 19 2021 22
Chromosome

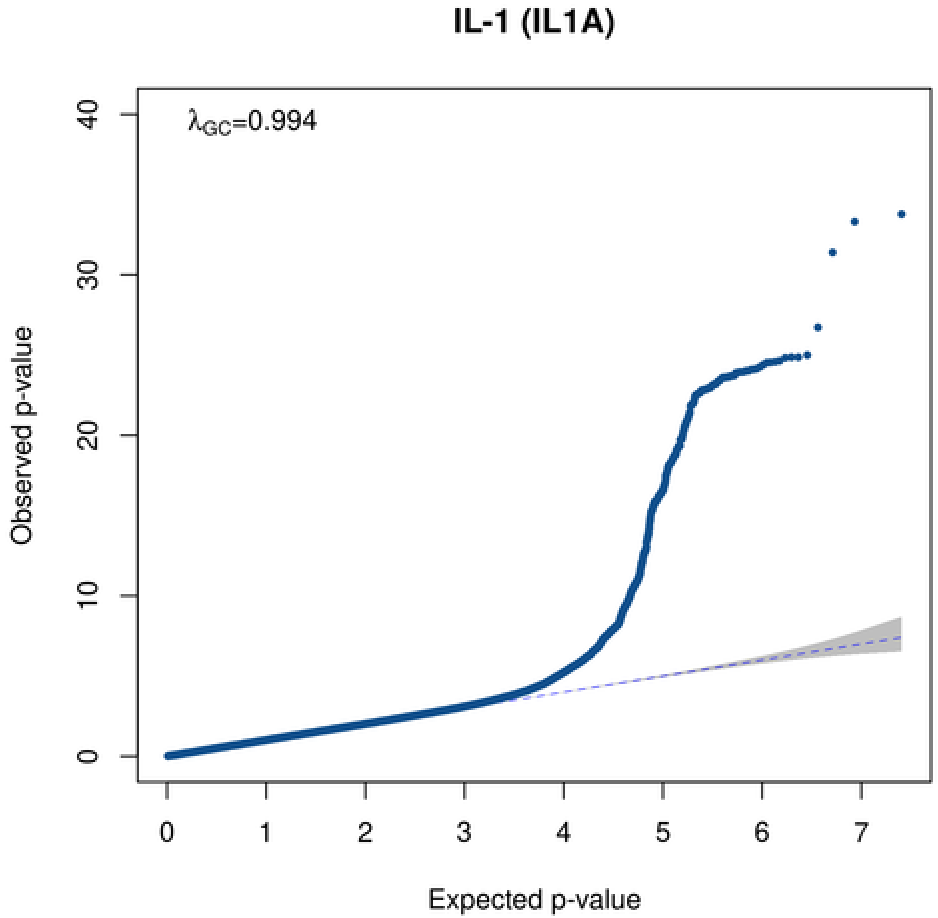
IL-1 (IL1A)
λGC=0.994
Observed p-value
0
10
20
30
40
0 1 2 3 4 5 6 7
Expected p-value

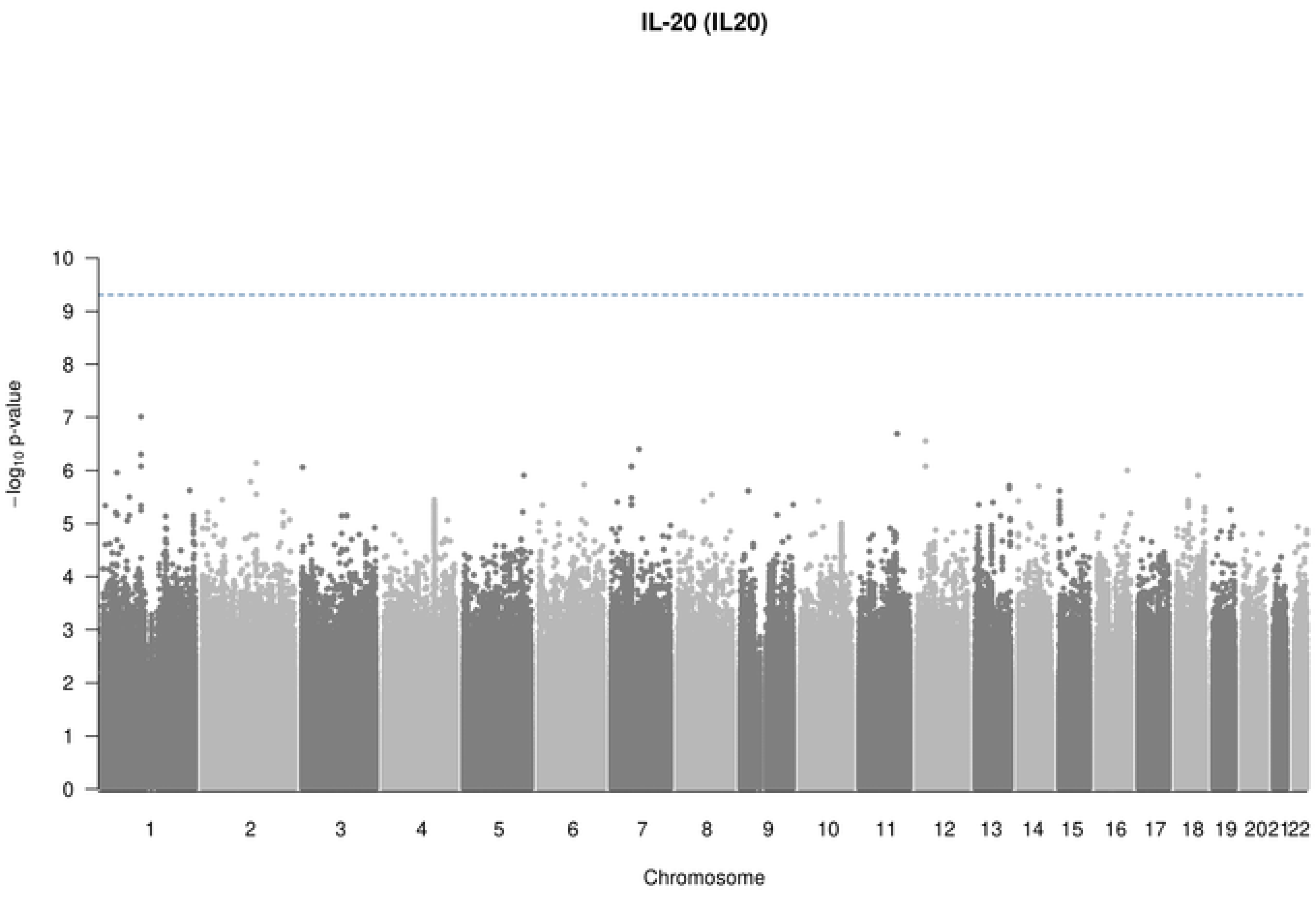

IL-20 (IL20)
−log10 p-value
Chromosome

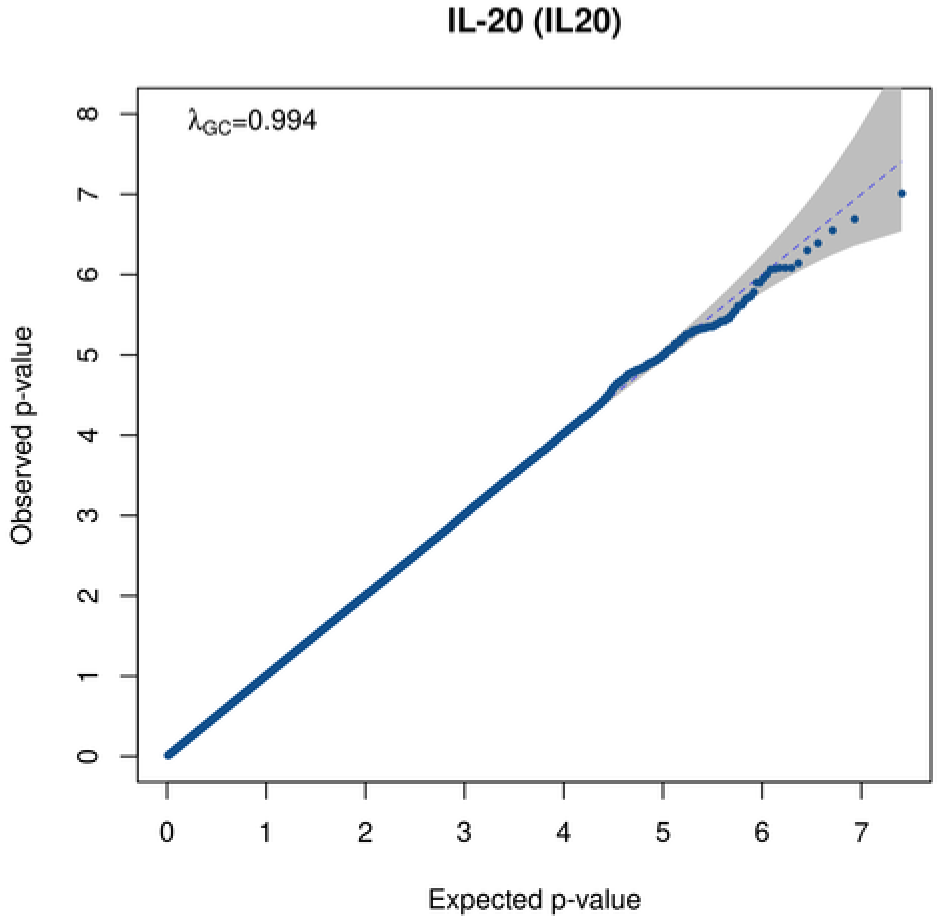

IL-20 (IL20)
$\lambda_{GC}$=0.994
Observed p-value
Expected p-value

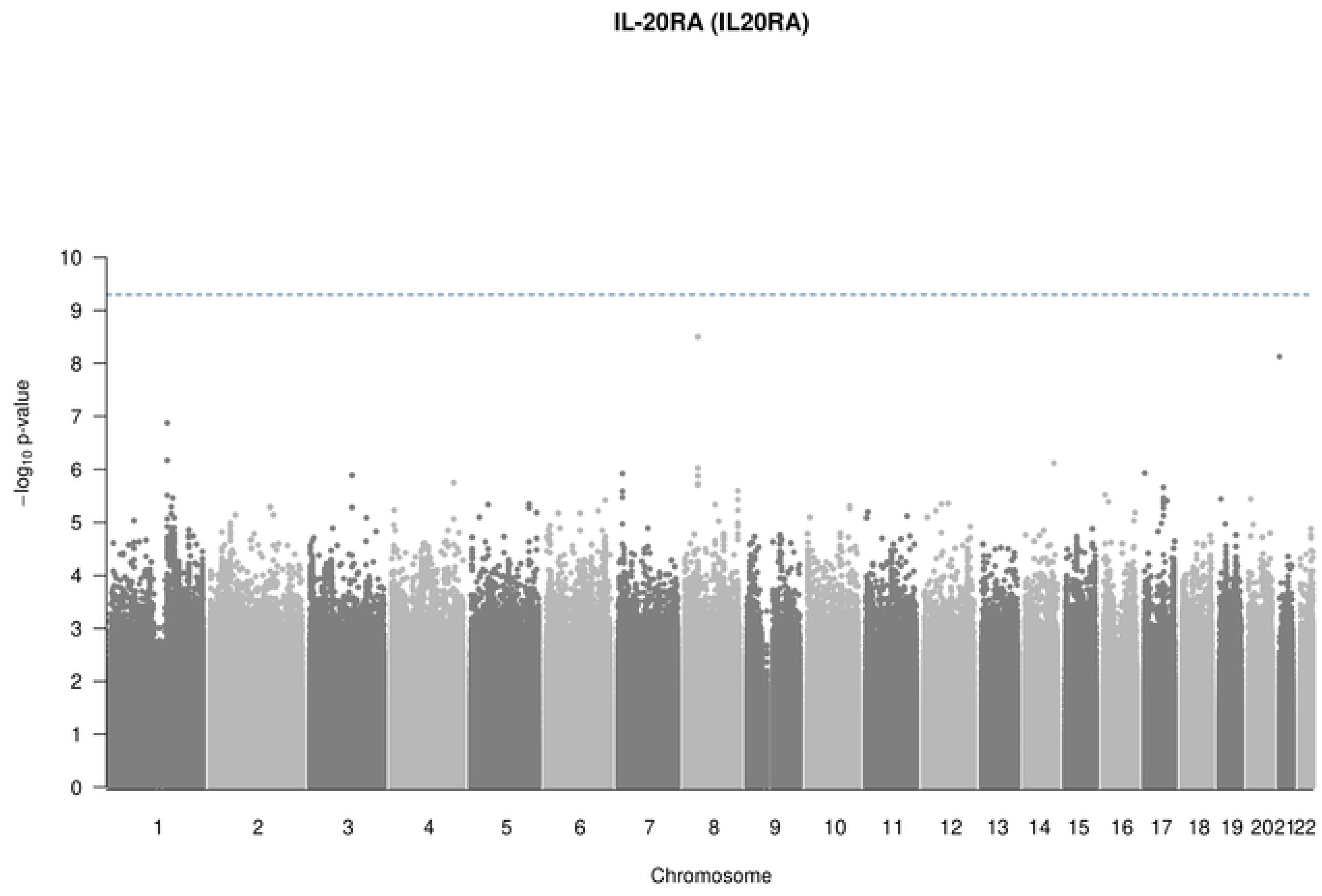

IL-20RA (IL20RA)
$-\log_{10}$ p-value
Chromosome
1 2 3 4 5 6 7 8 9 10 11 12 13 14 15 16 17 18 19 20 21 22

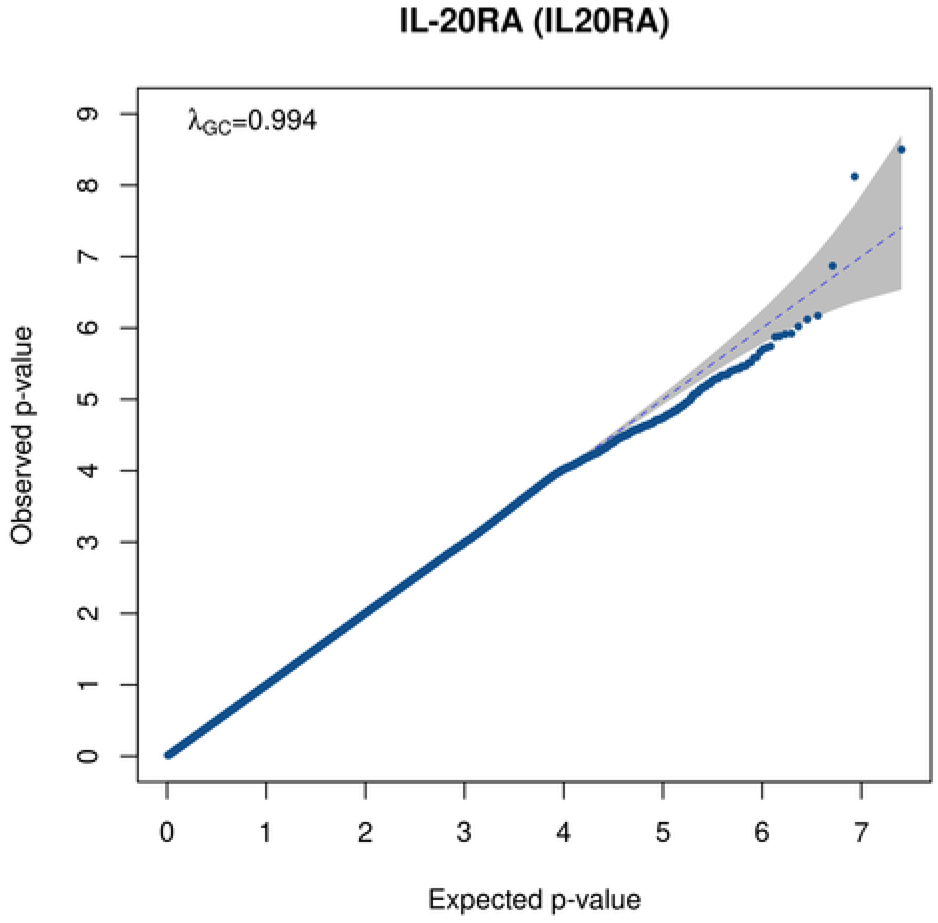

IL-20RA (IL20RA)
$\lambda_{GC}$=0.994
Observed p-value
Expected p-value

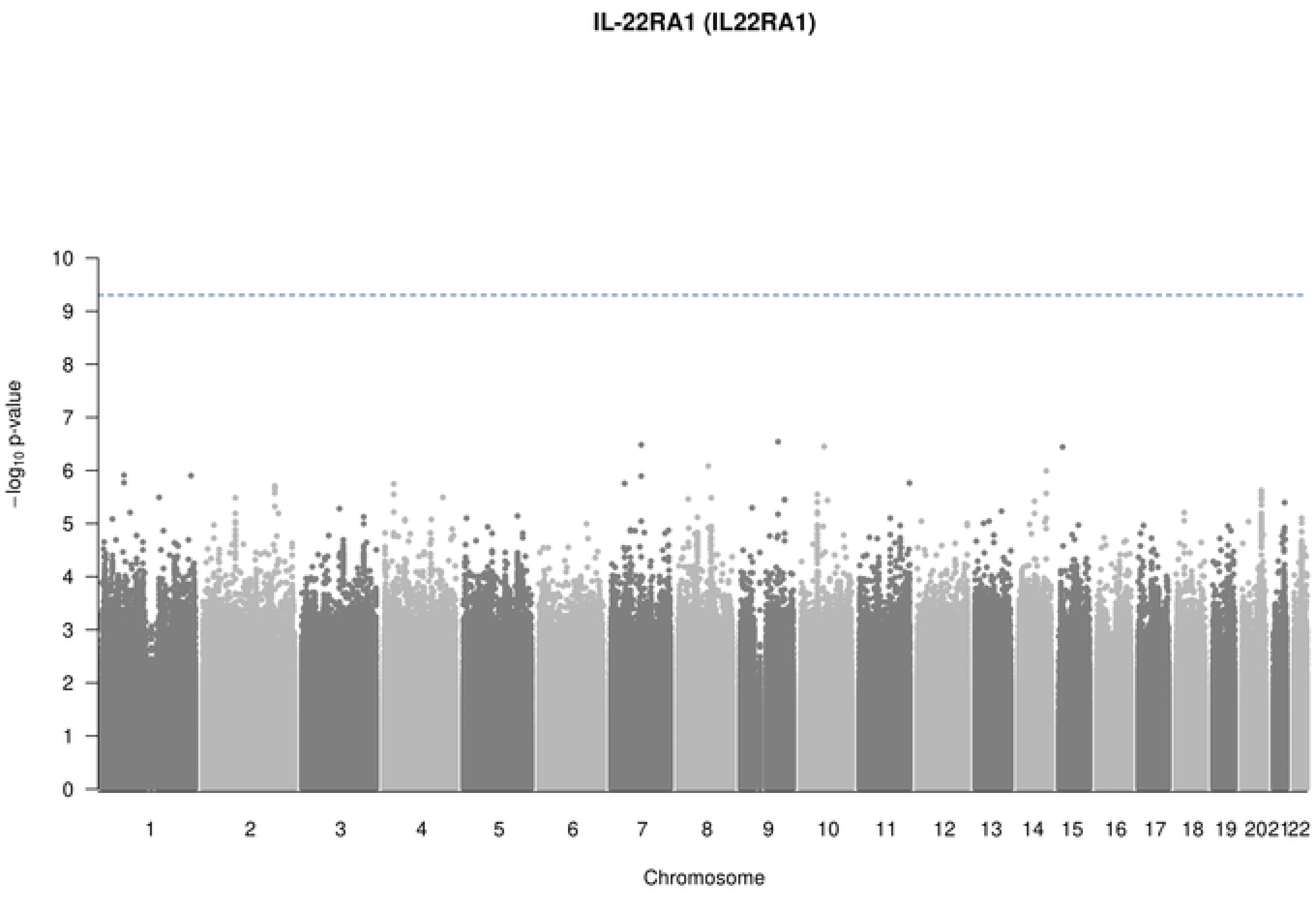
IL-22RA1 (IL22RA1)
-log10 p-value
Chromosome

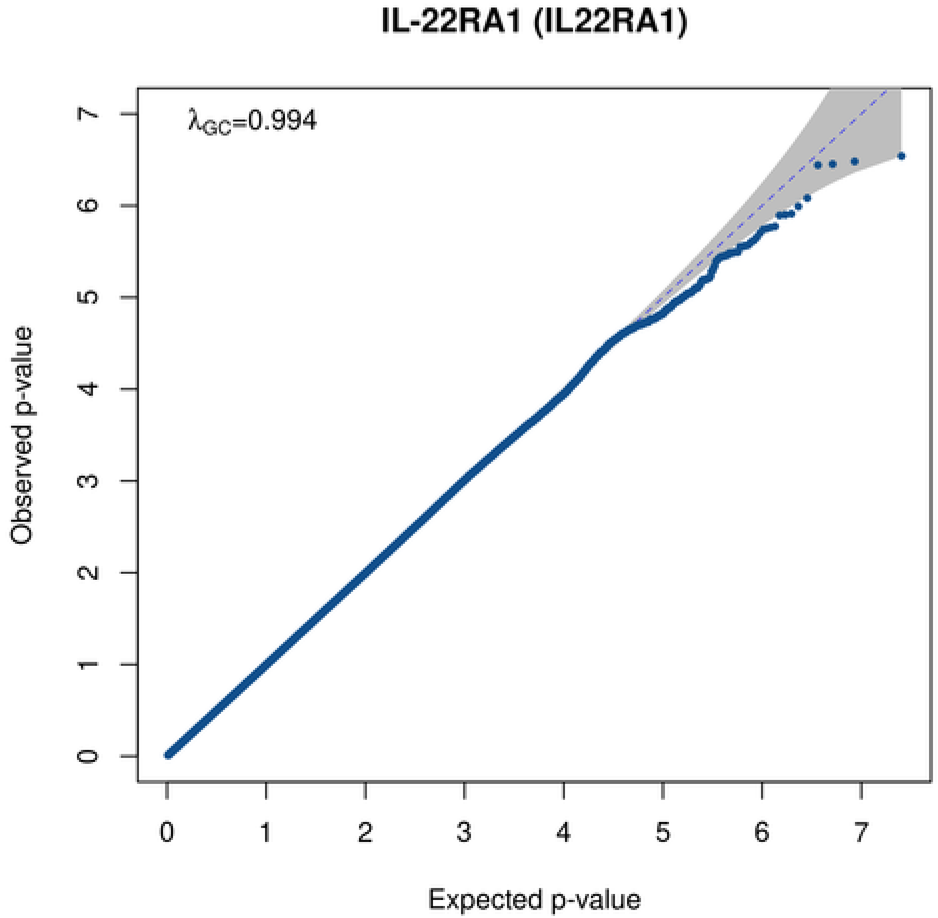
IL-22RA1 (IL22RA1)
$\lambda_{GC}$=0.994
Observed p-value
Expected p-value

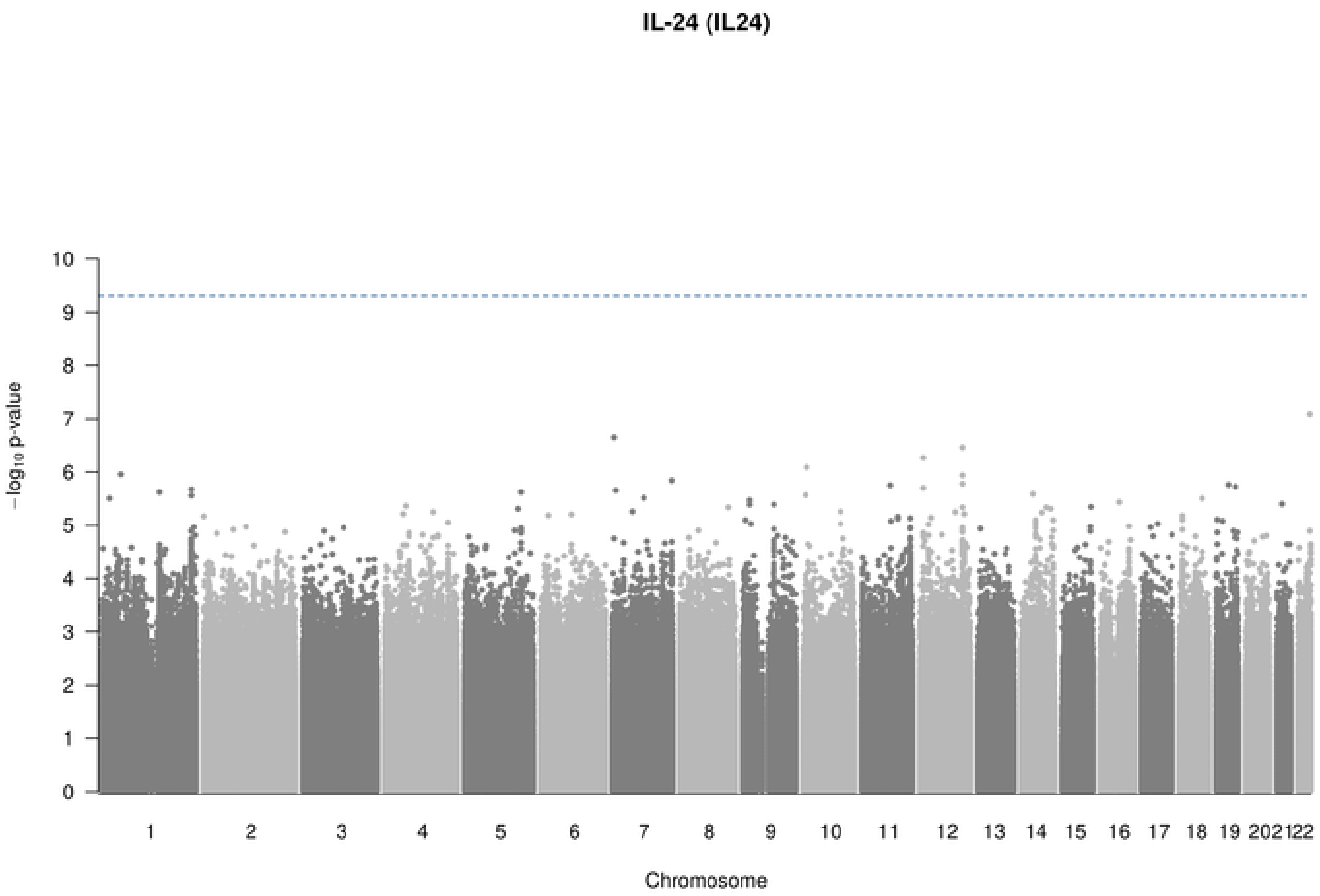
IL-24 (IL24)
10
9
8
7
6
5
4
3
2
1
0
$-\log_{10}$ p-value
202122
Chromosome

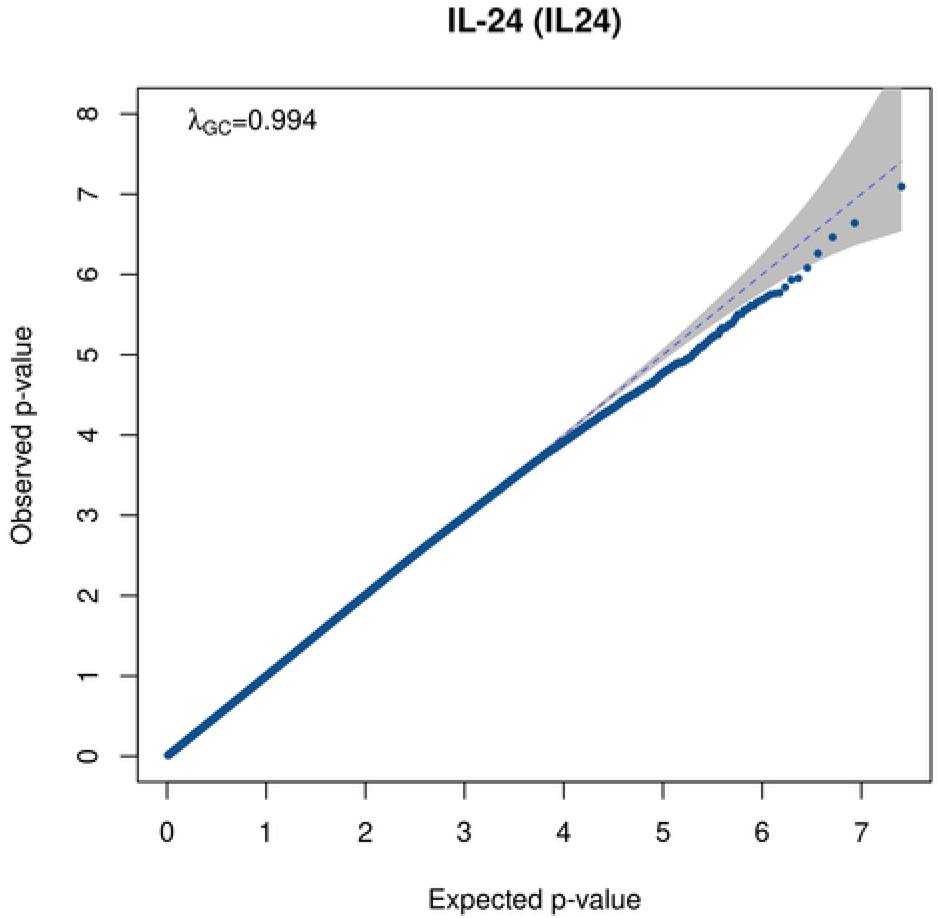
IL-24 (IL24)
$\lambda_{GC}$=0.994
Observed p-value
Expected p-value
0
1
2
3
4
5
6
7
8

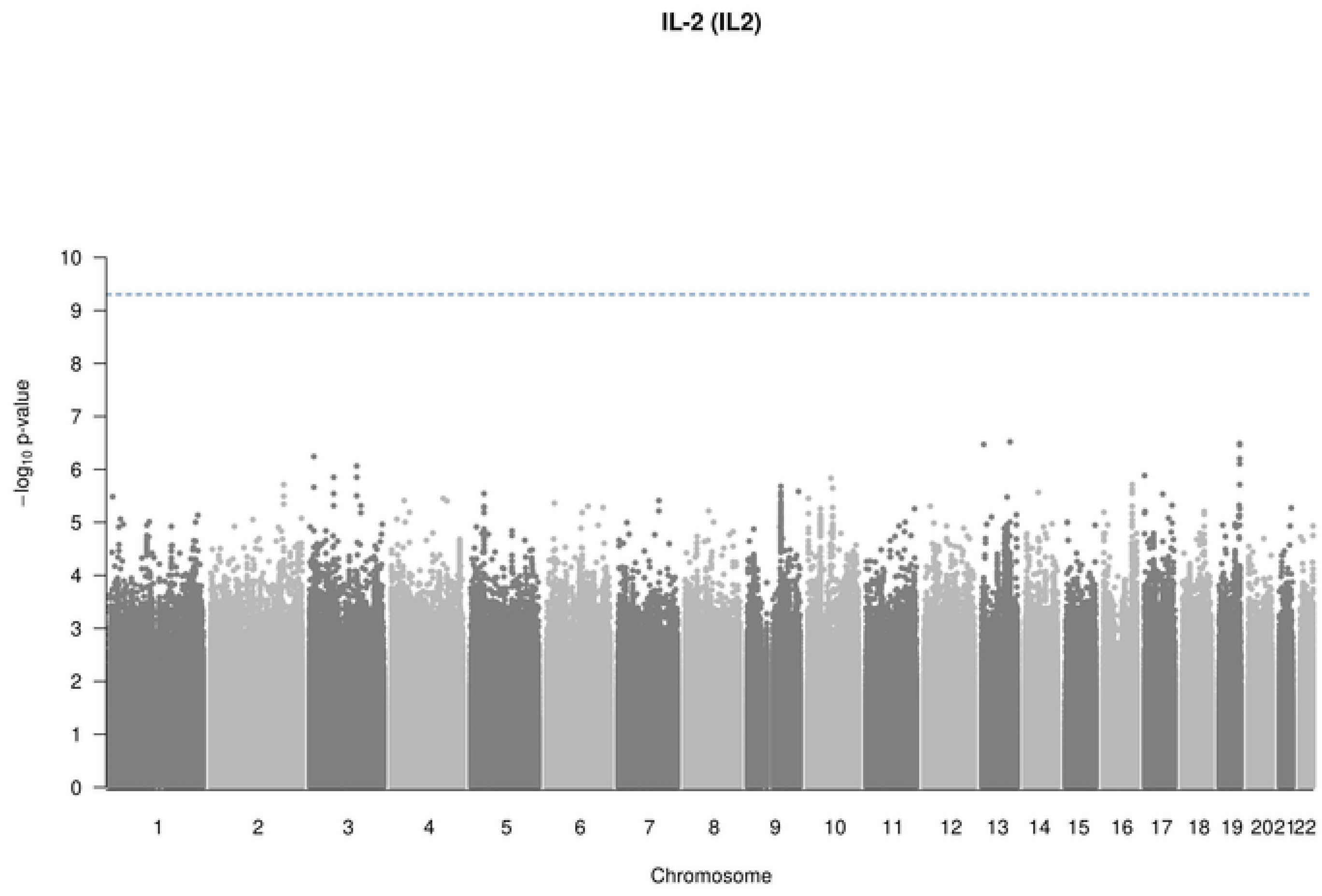
IL-2 (IL2)
−log10 p-value
Chromosome
0 1 2 3 4 5 6 7 8 9 10
1 2 3 4 5 6 7 8 9 10 11 12 13 14 15 16 17 18 19 20 21 22

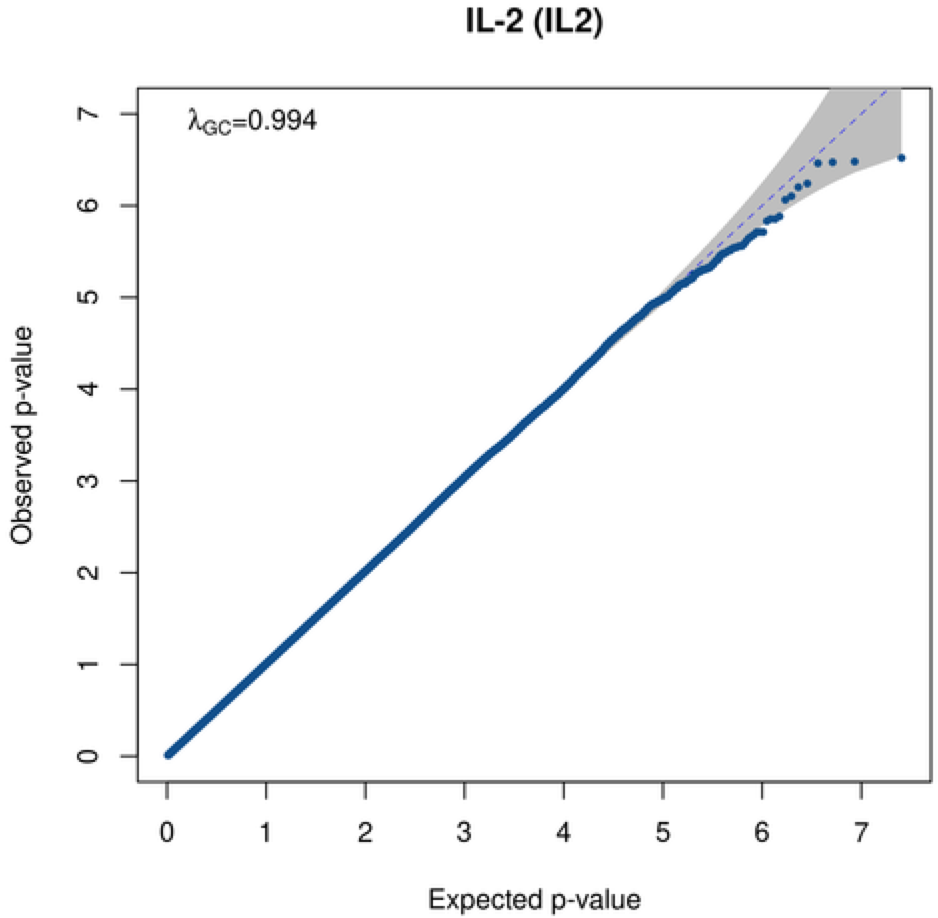
IL-2 (IL2)
λGC=0.994
Observed p-value
Expected p-value
0 1 2 3 4 5 6 7

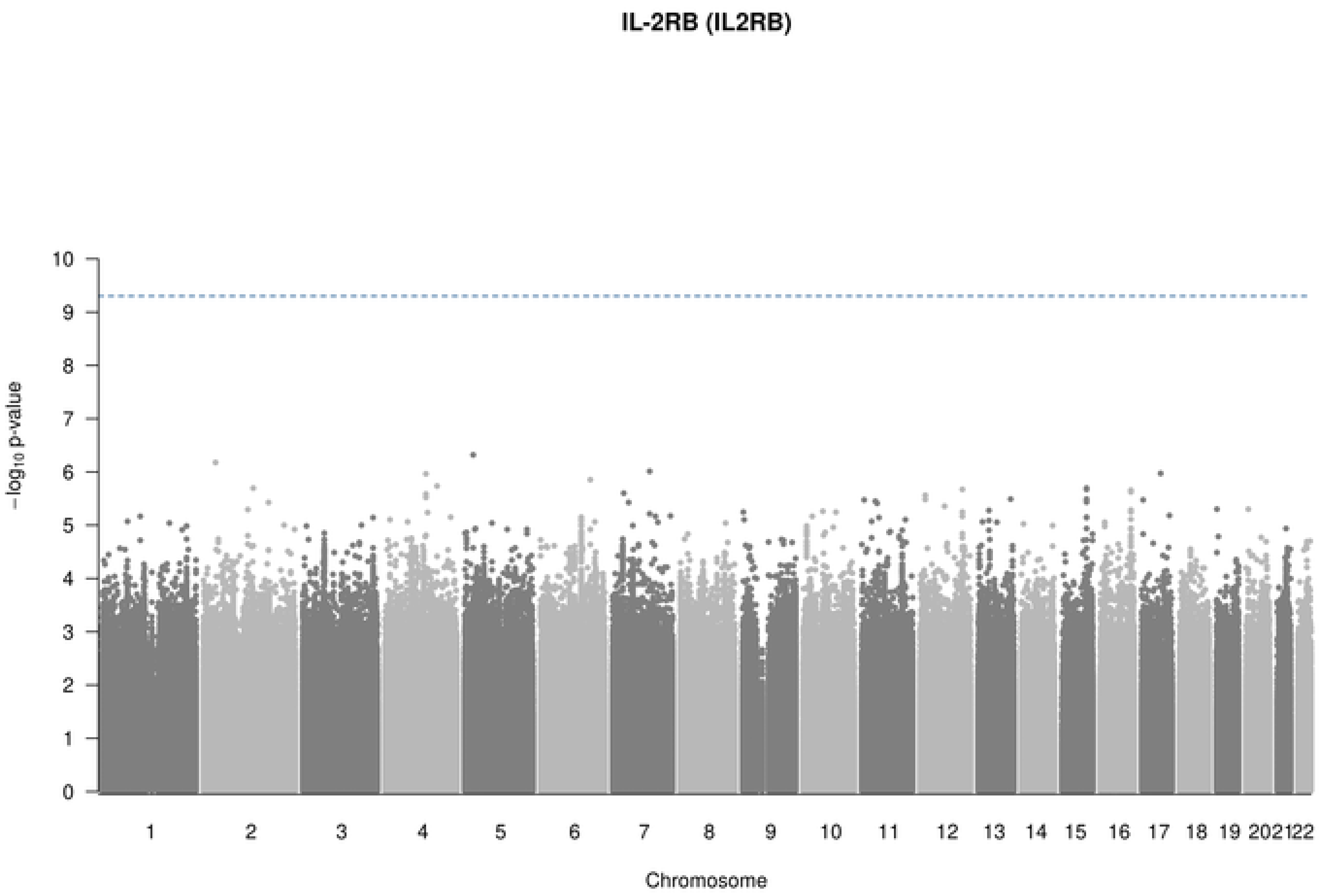
IL-2RB (IL2RB)
$-\log_{10}$ p-value
Chromosome
1 2 3 4 5 6 7 8 9 10 11 12 13 14 15 16 17 18 19 202122

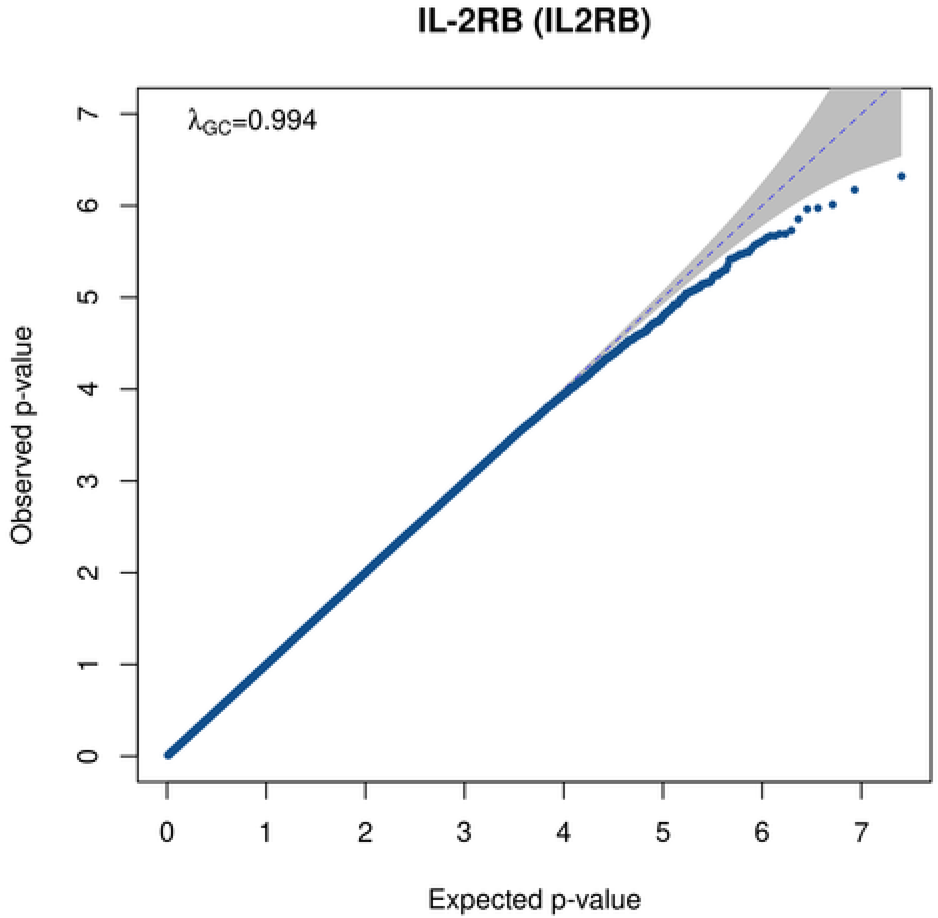
IL-2RB (IL2RB)
$\lambda_{GC}$=0.994
Observed p-value
Expected p-value

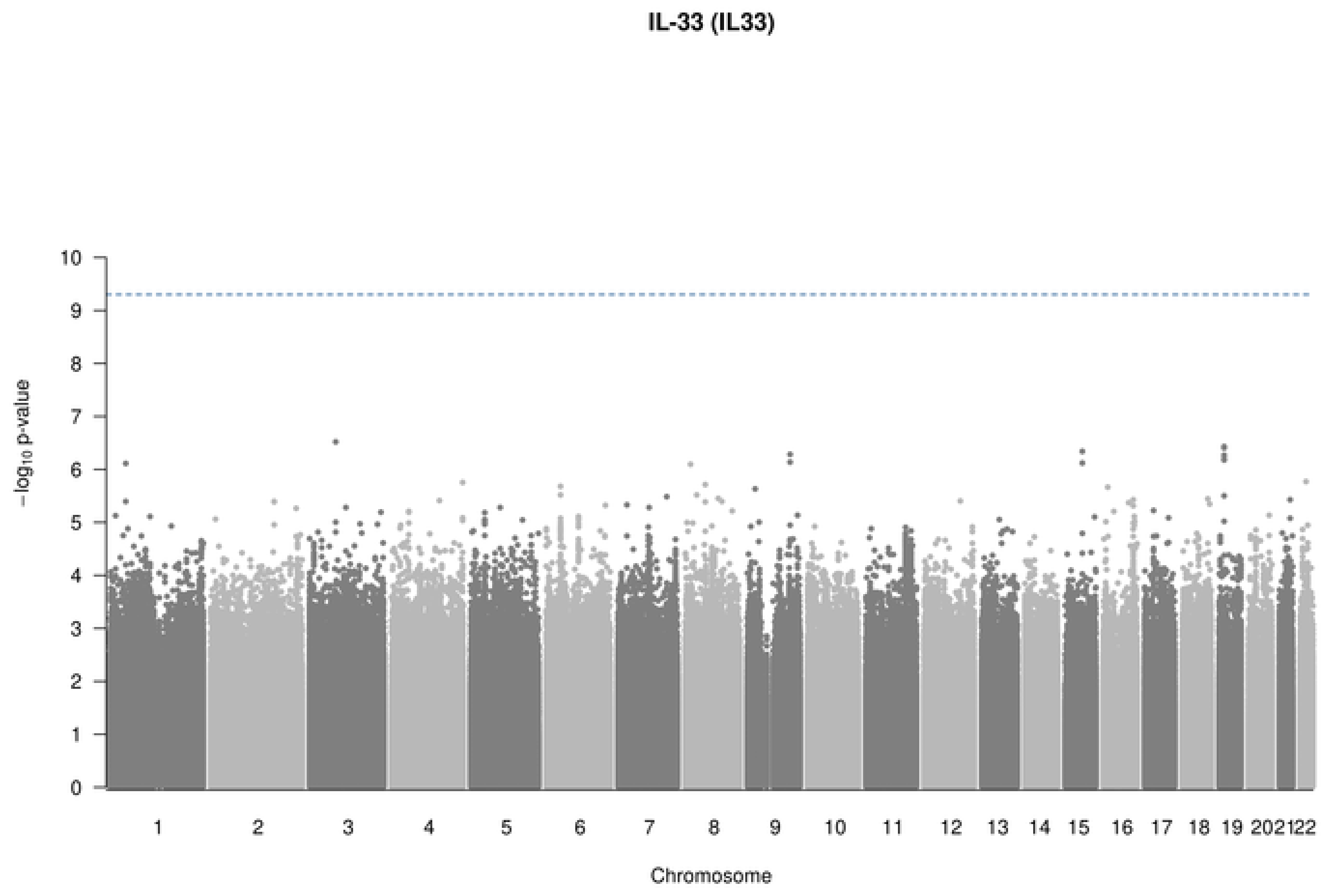

IL-33 (IL33)
−log10 p-value
Chromosome

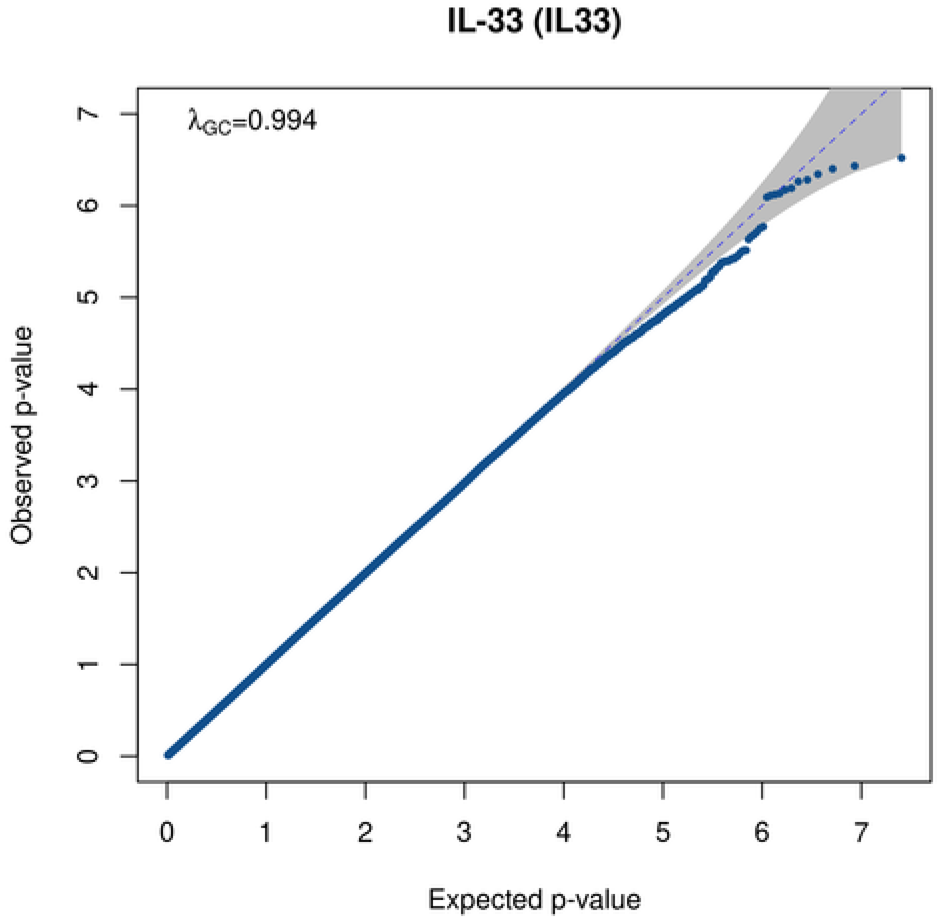

IL-33 (IL33)
λGC=0.994
Observed p-value
Expected p-value

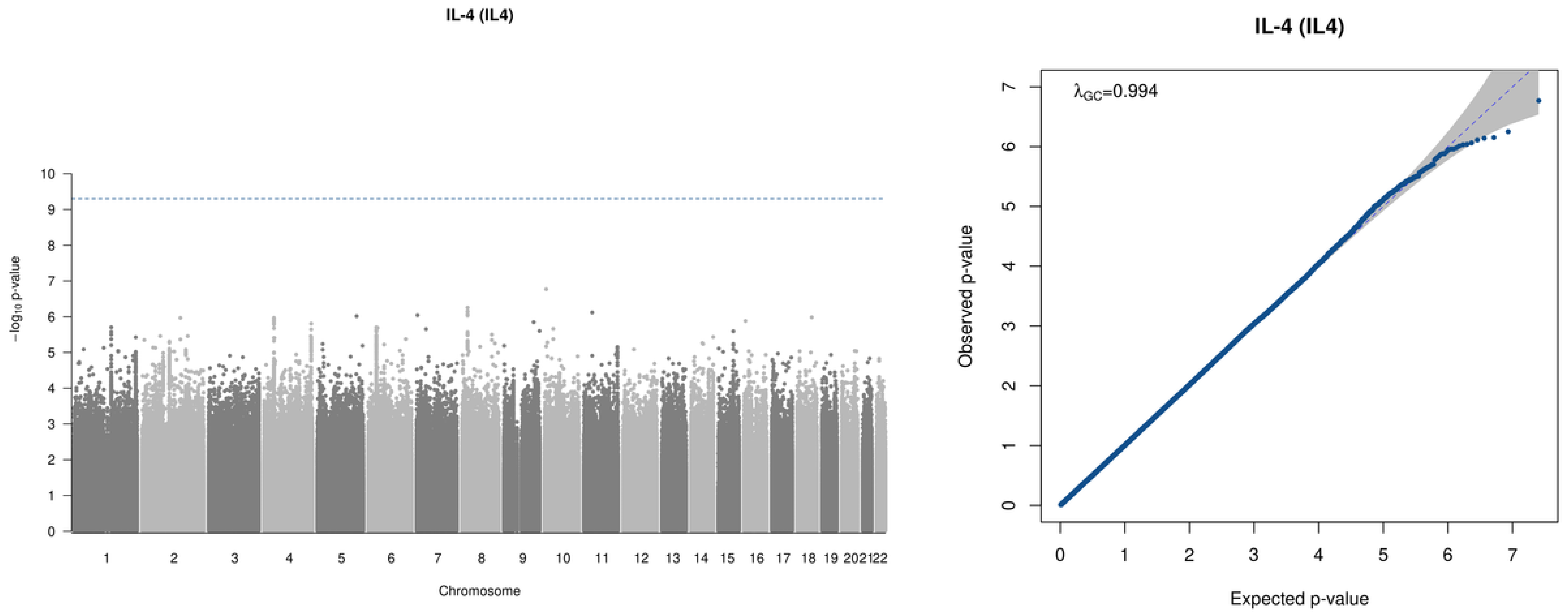
IL-4 (IL4)
−log10 p-value
Chromosome
IL-4 (IL4)
λGC=0.994
Observed p-value
Expected p-value

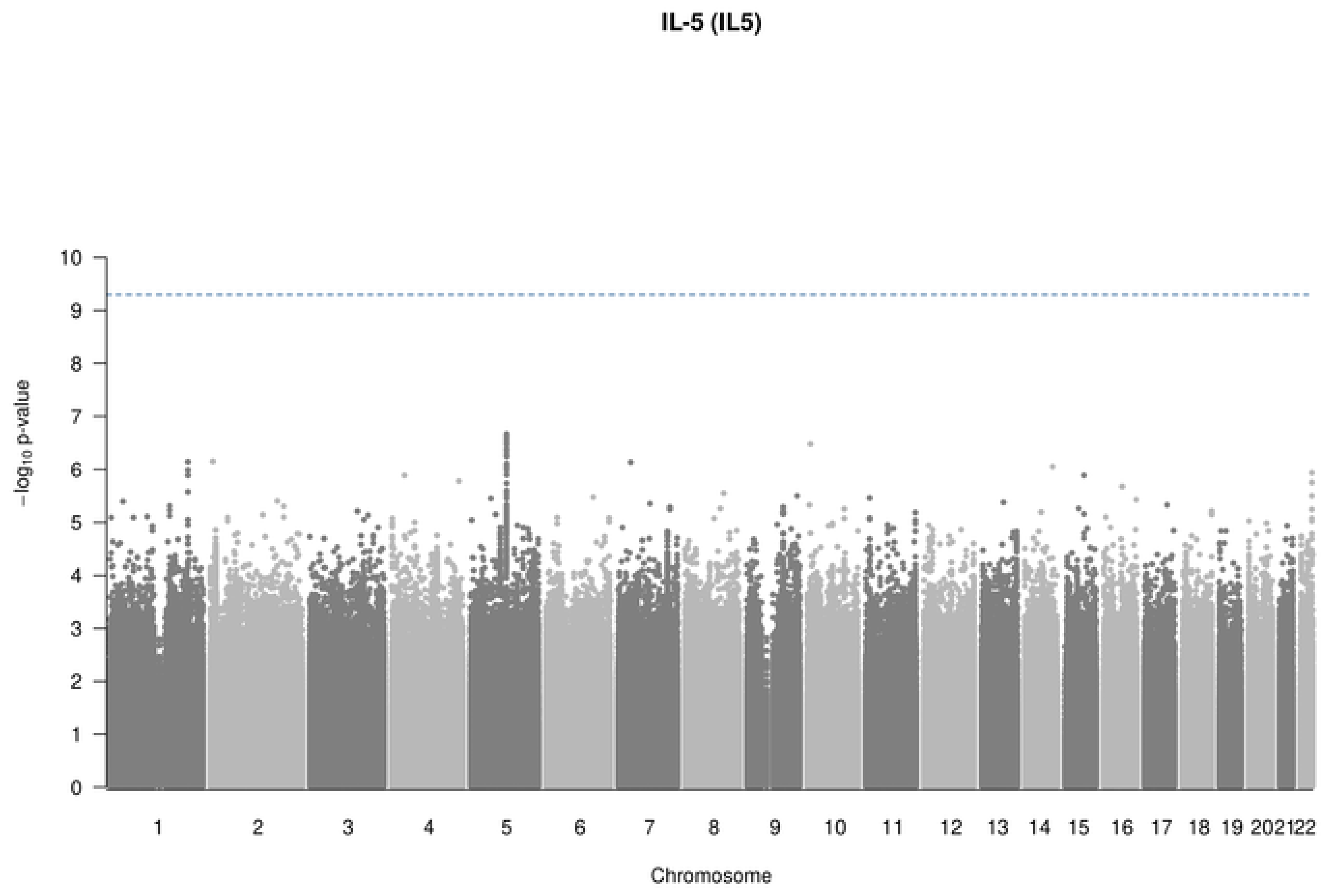

IL-5 (IL5)
10
9
8
7
6
5
4
3
2
1
0
−log10 p-value
202122
Chromosome

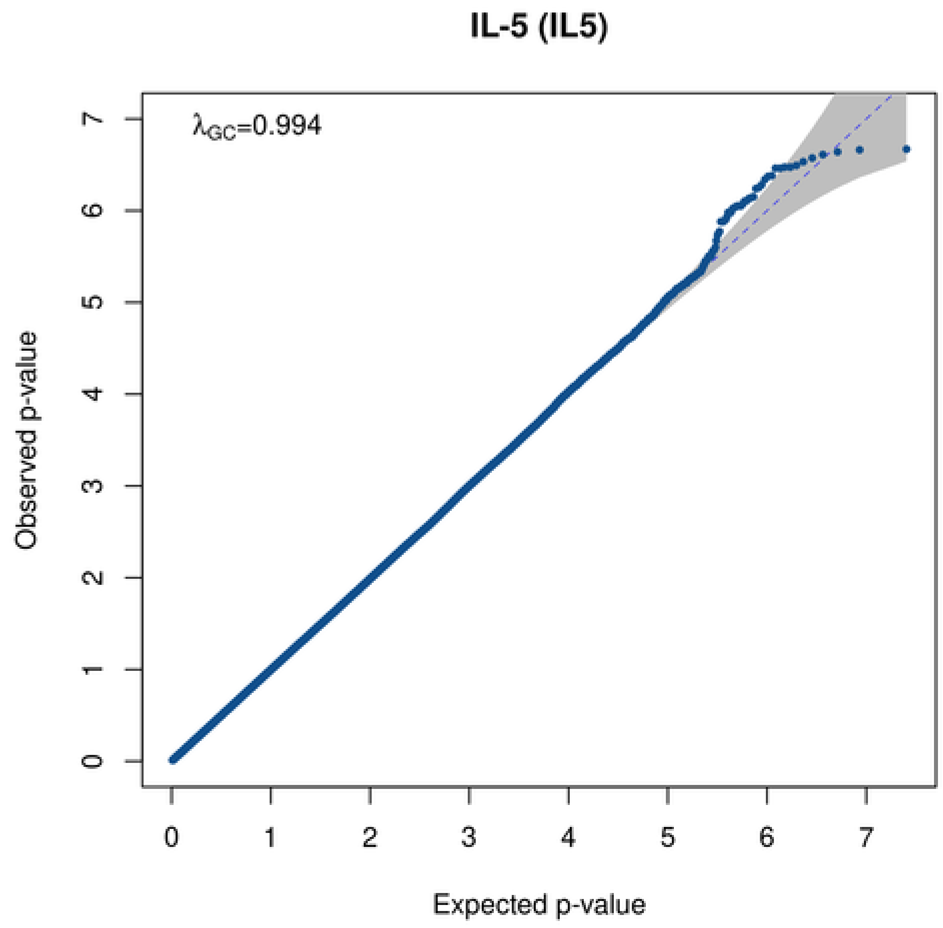

IL-5 (IL5)
λGC=0.994
Observed p-value
0
1
2
3
4
5
6
7
Expected p-value

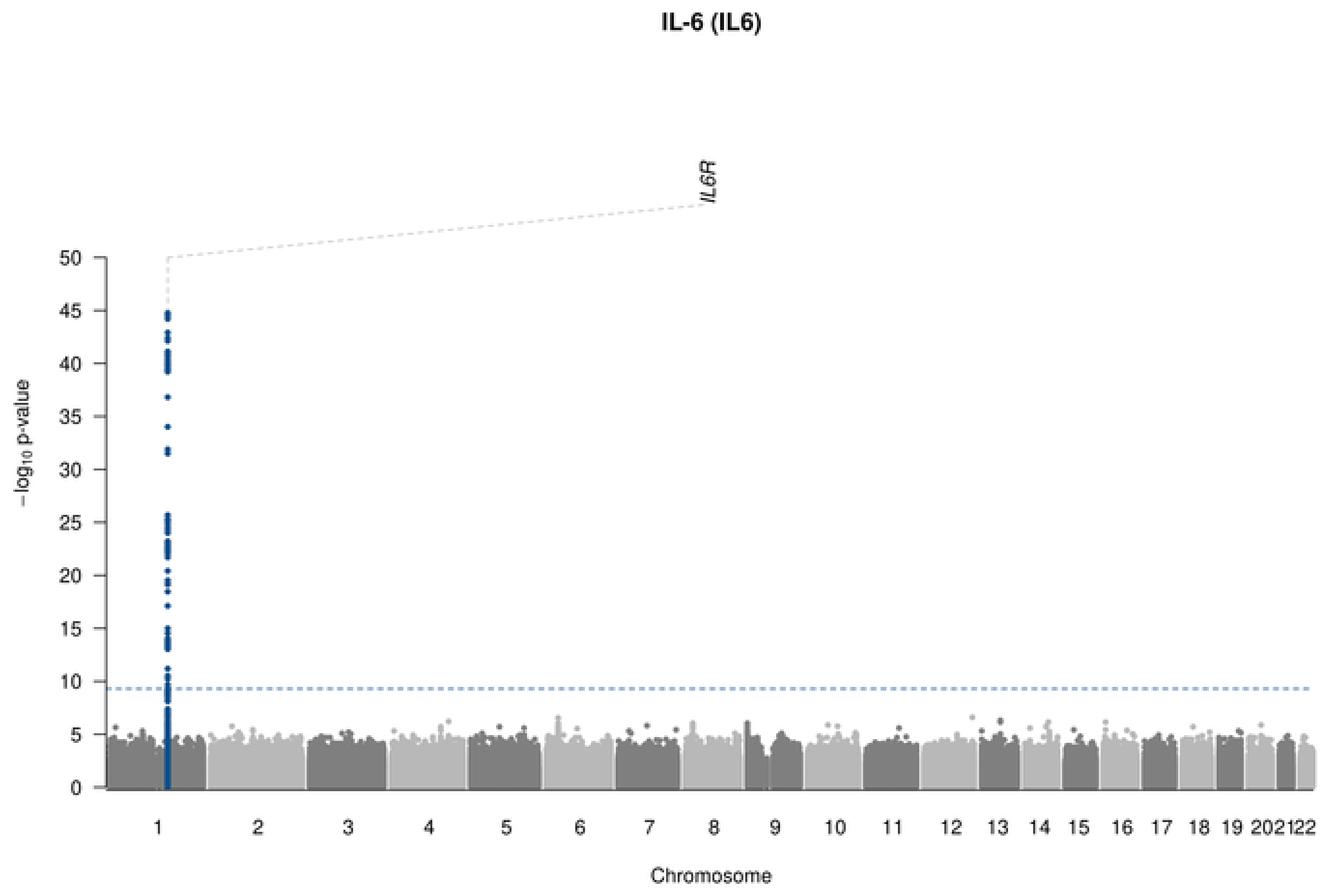
IL-6 (IL6)
IL6R
50
45
40
35
30
25
20
15
10
5
0
−log10 p-value
1 2 3 4 5 6 7 8 9 10 11 12 13 14 15 16 17 18 19 202122
Chromosome

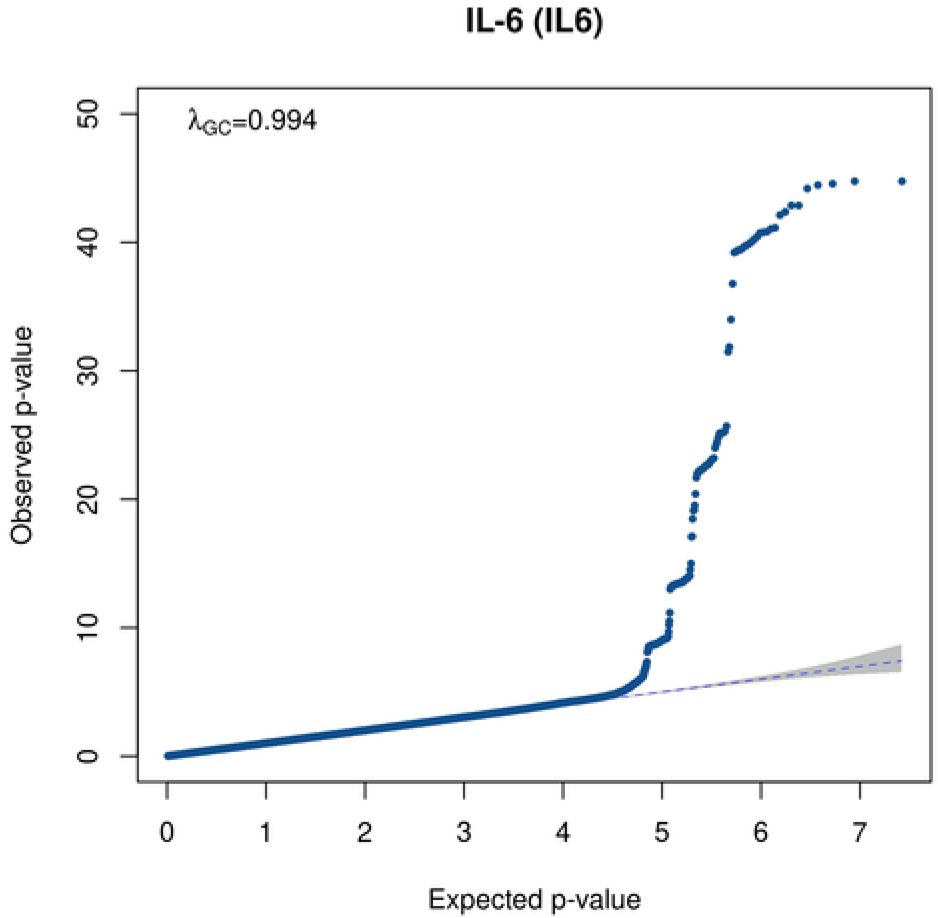
IL-6 (IL6)
λGC=0.994
50
40
30
20
10
0
Observed p-value
0 1 2 3 4 5 6 7
Expected p-value

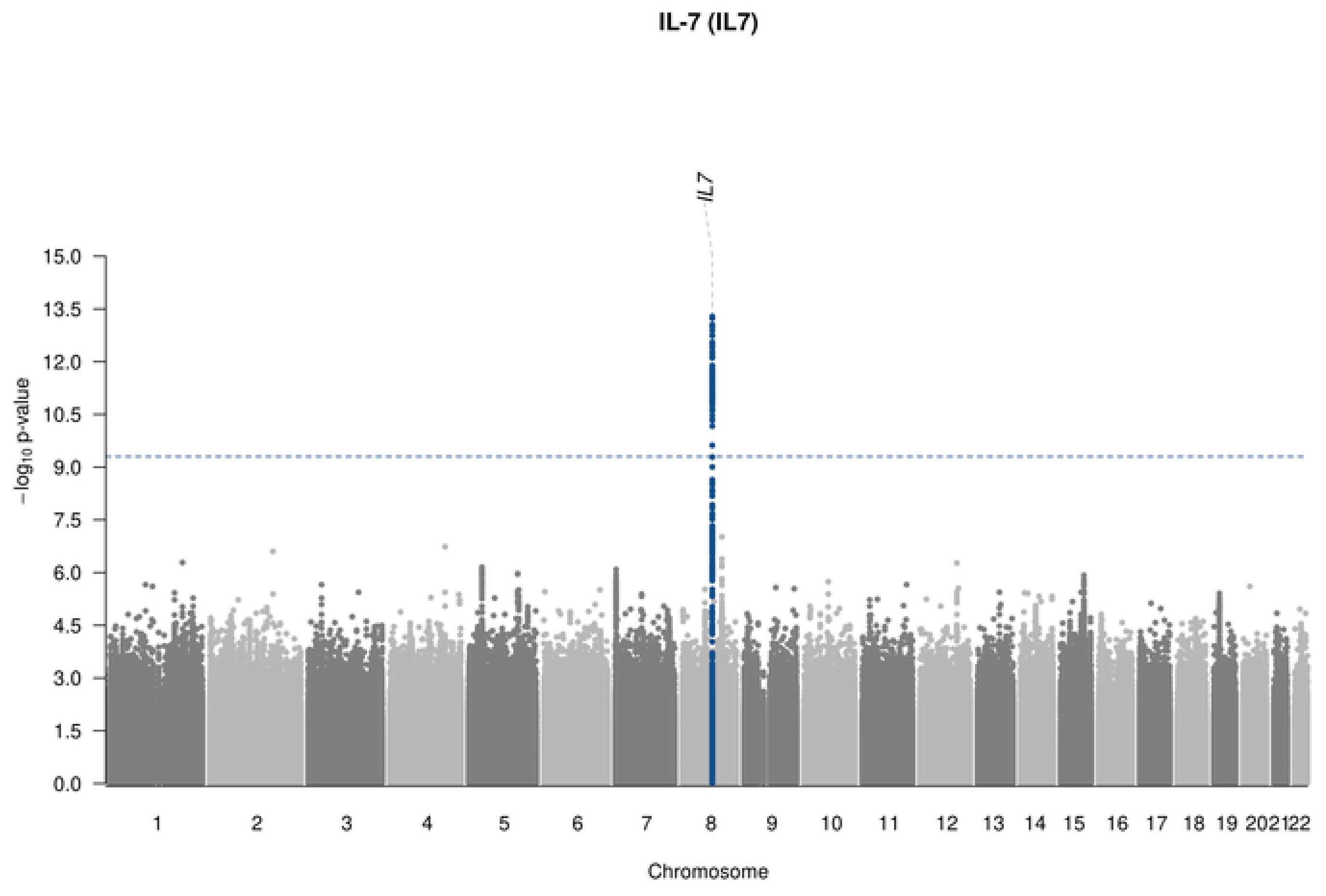
IL-7 (IL7)
IL7
−log10 p-value
15.0
13.5
12.0
10.5
9.0
7.5
6.0
4.5
3.0
1.5
0.0
1 2 3 4 5 6 7 8 9 10 11 12 13 14 15 16 17 18 19 20 21 22
Chromosome

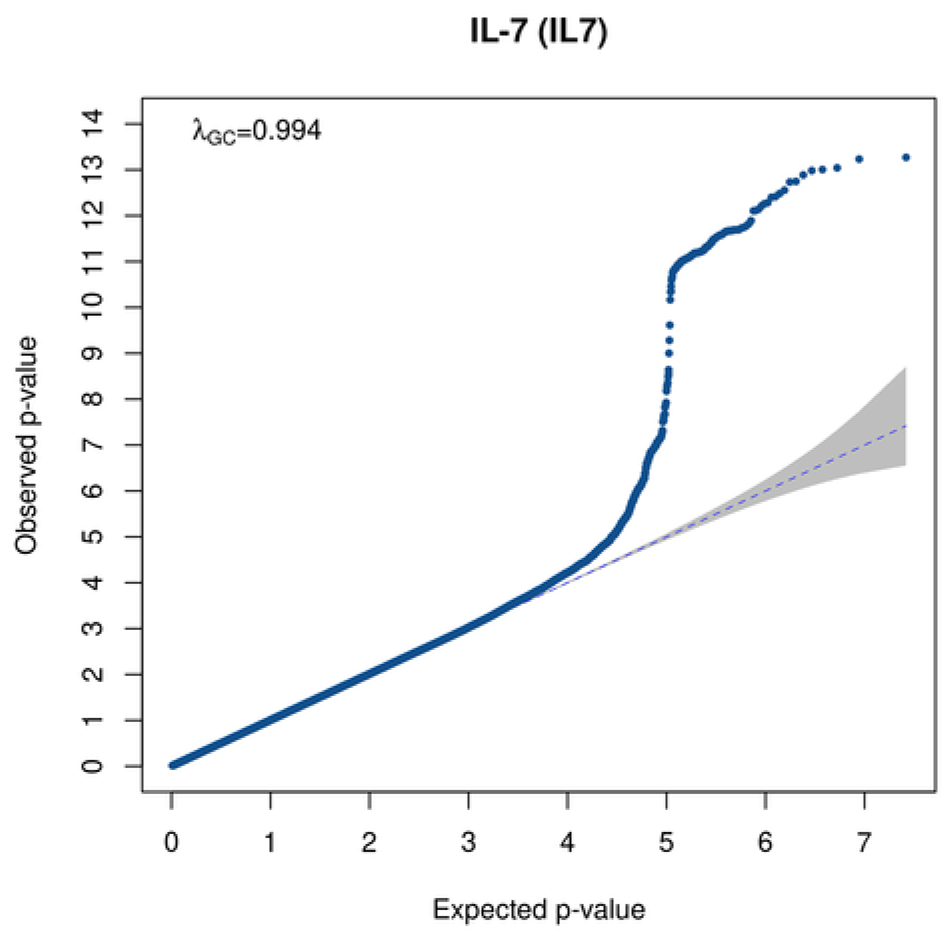
IL-7 (IL7)
λGC=0.994
Observed p-value
Expected p-value
0 1 2 3 4 5 6 7 8 9 10 11 12 13 14
0 1 2 3 4 5 6 7

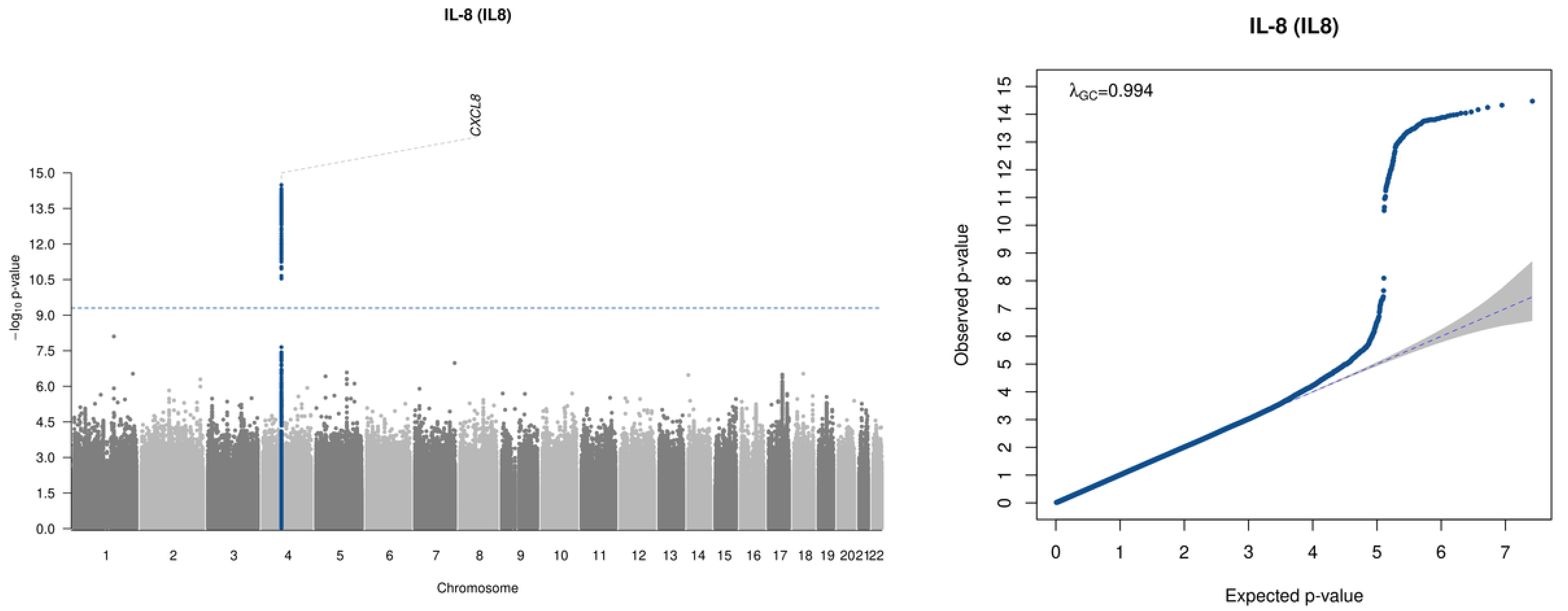

IL-8 (IL8)
CXCL8
15.0
13.5
12.0
10.5
9.0
7.5
6.0
4.5
3.0
1.5
0.0
−log10 p-value
1 2 3 4 5 6 7 8 9 10 11 12 13 14 15 16 17 18 19 2021 22
Chromosome
IL-8 (IL8)
λGC=0.994
Observed p-value
Expected p-value

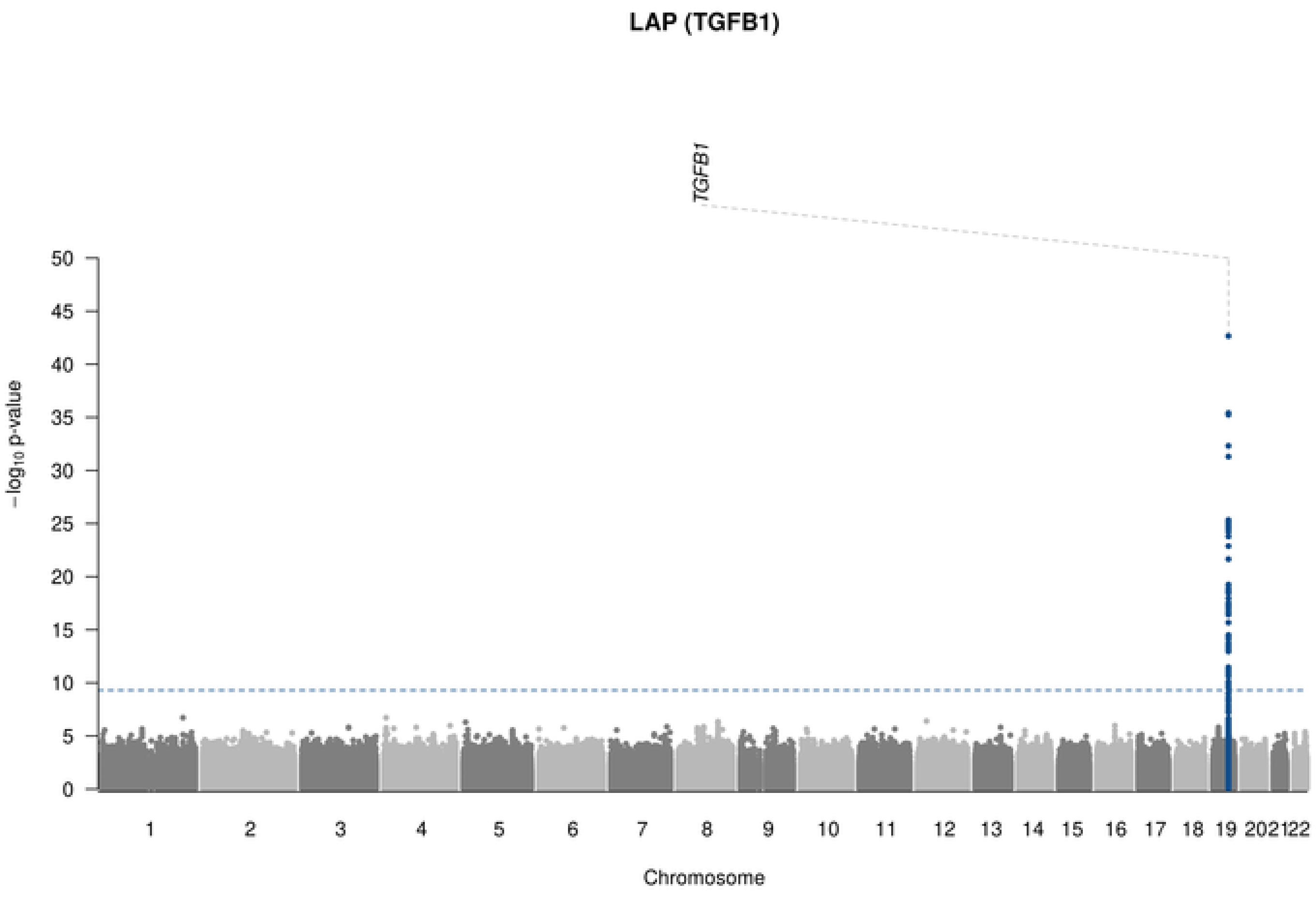

LAP (TGFB1)
TGFB1
$-\log_{10}$ p-value
Chromosome

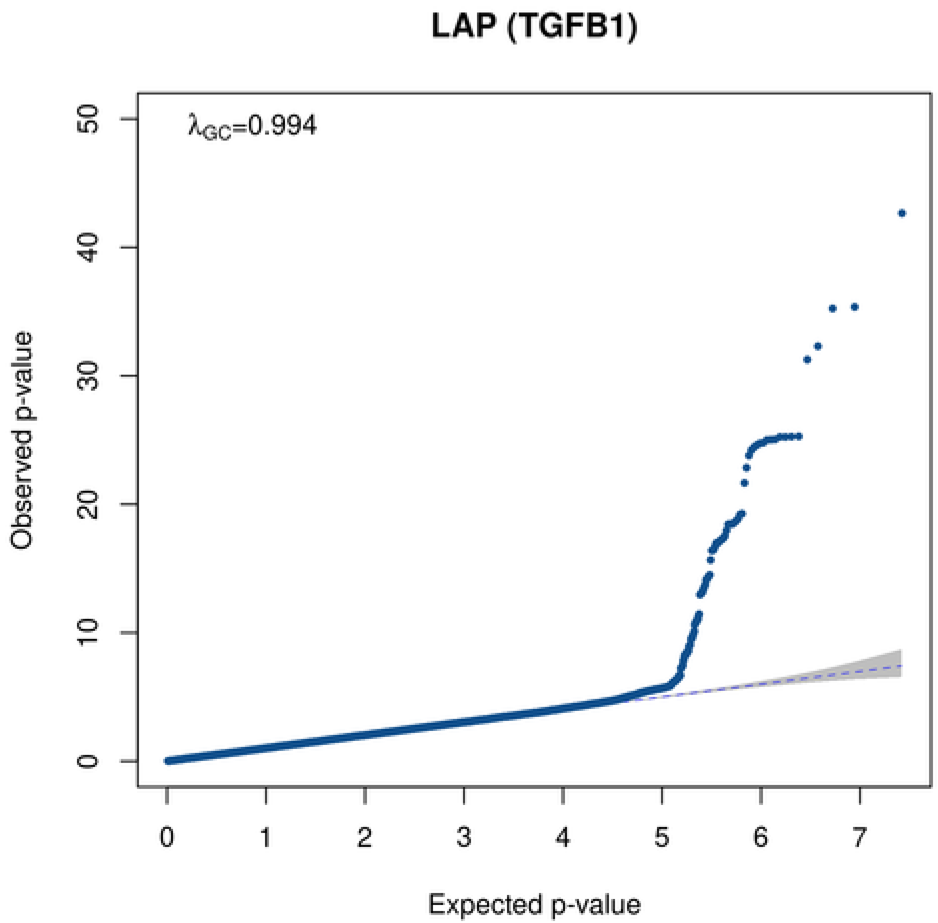

LAP (TGFB1)
$\lambda_{GC}$=0.994
Observed p-value
Expected p-value

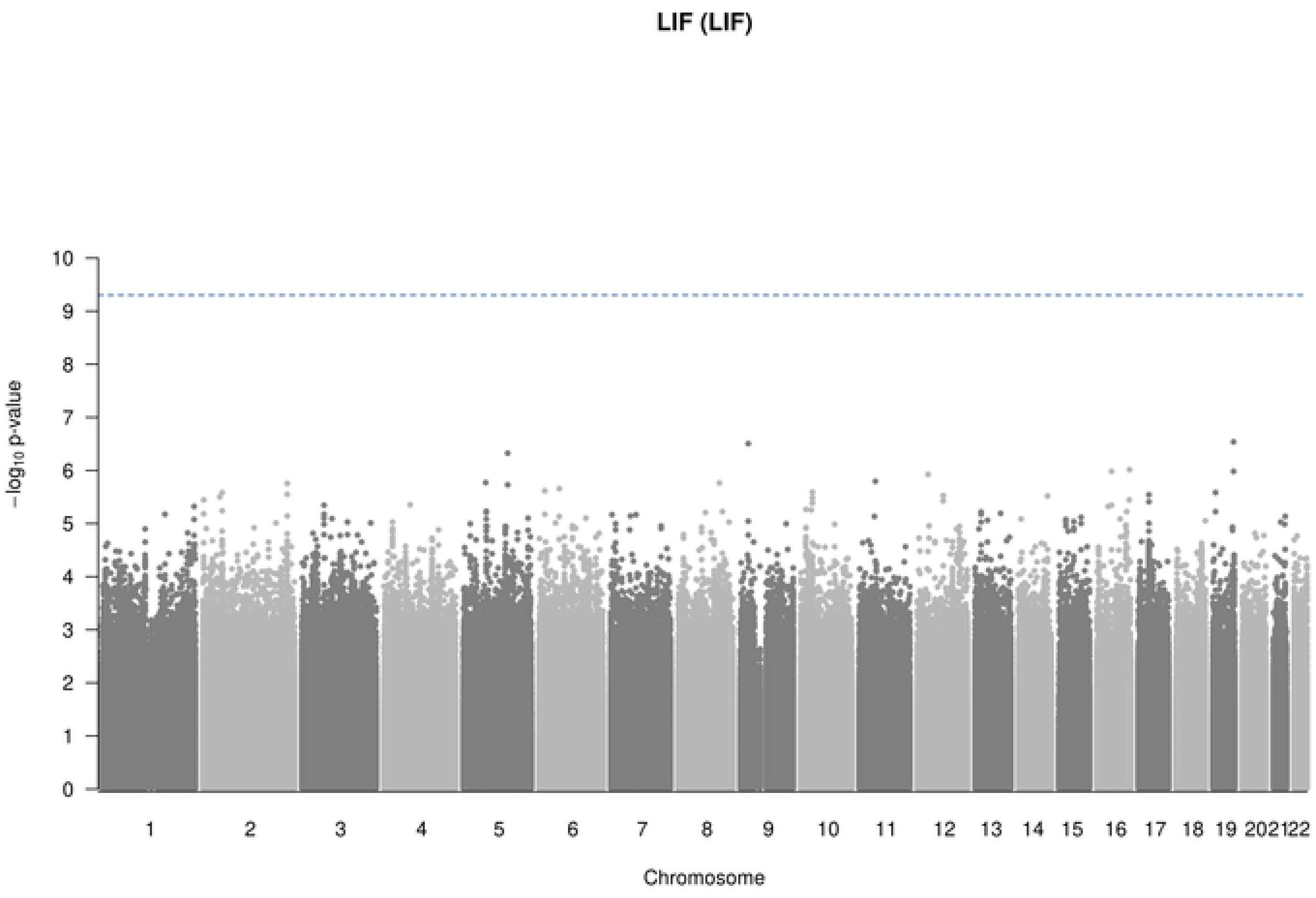

LIF (LIF)
−log10 p-value
Chromosome

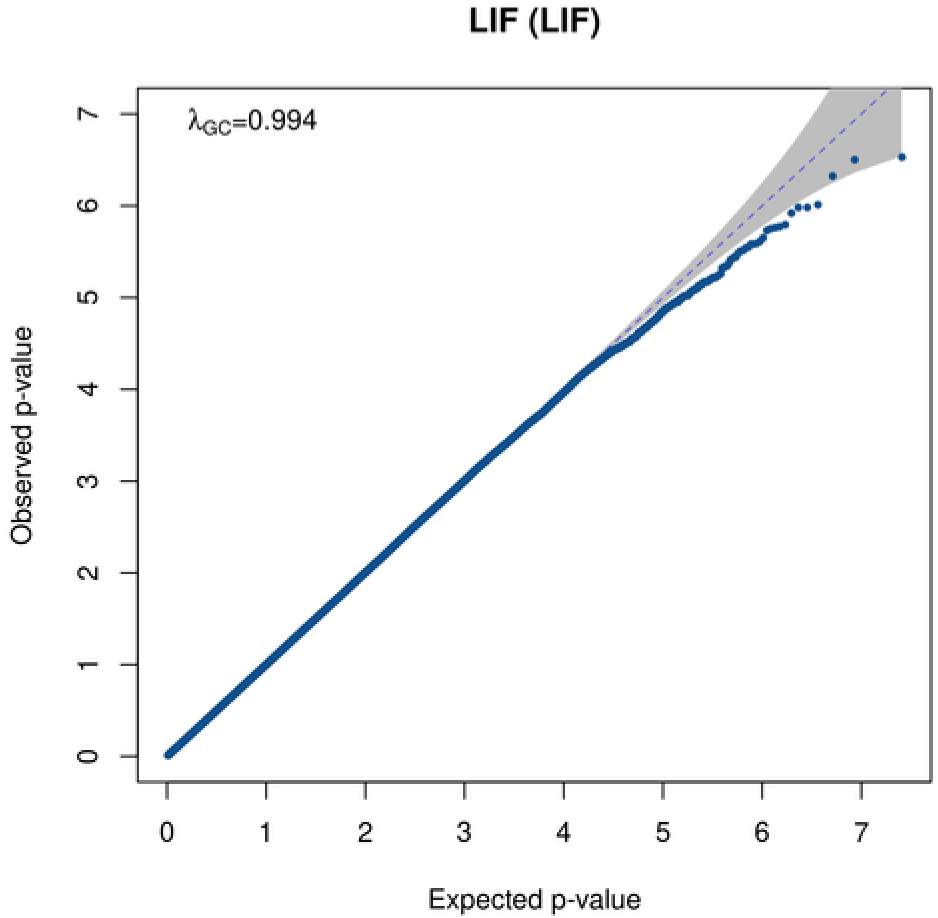

LIF (LIF)
λGC=0.994
Observed p-value
Expected p-value

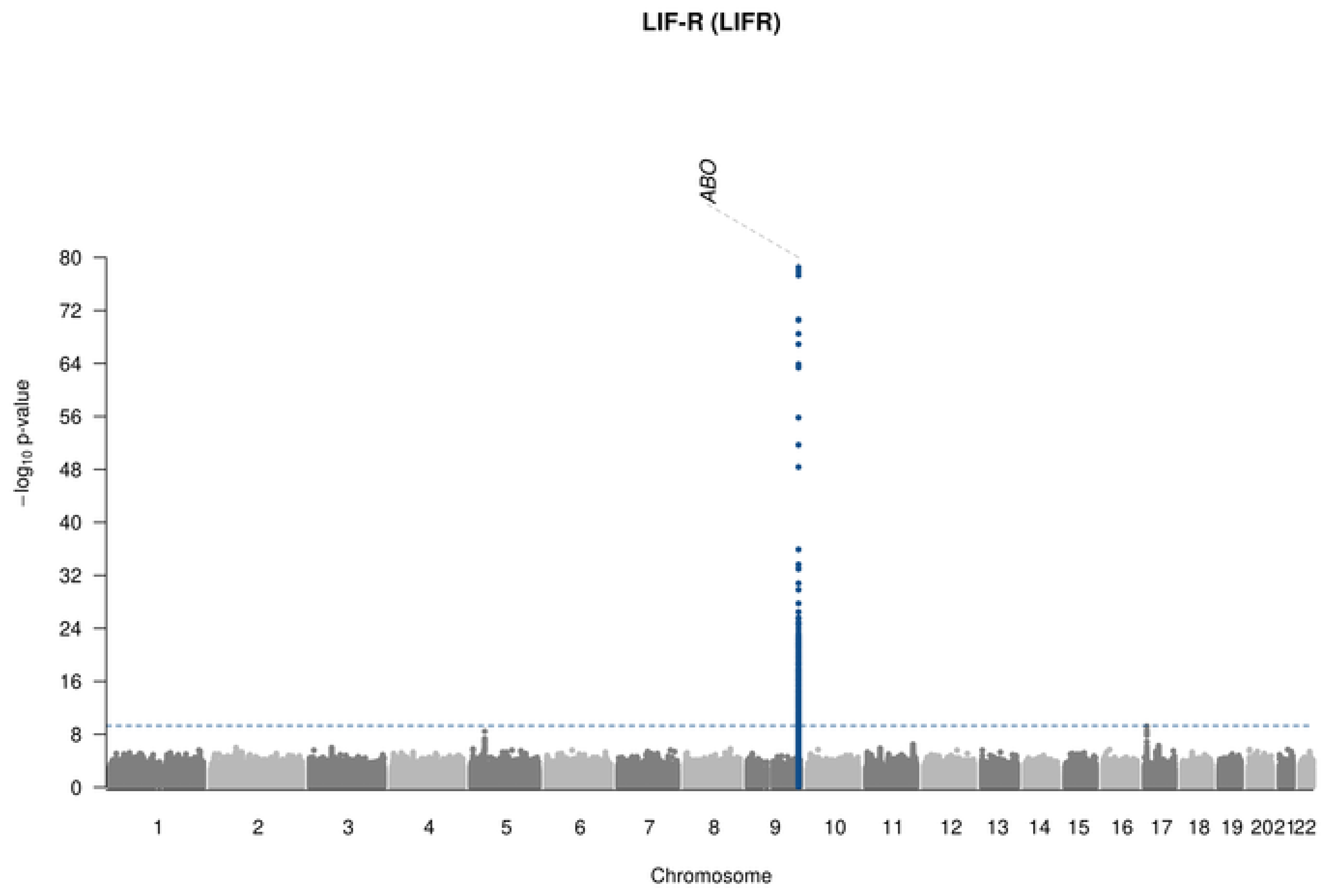

LIF-R (LIFR)
ABO
80
72
64
56
48
40
32
24
16
8
0
−log10 p-value
2021
22
Chromosome

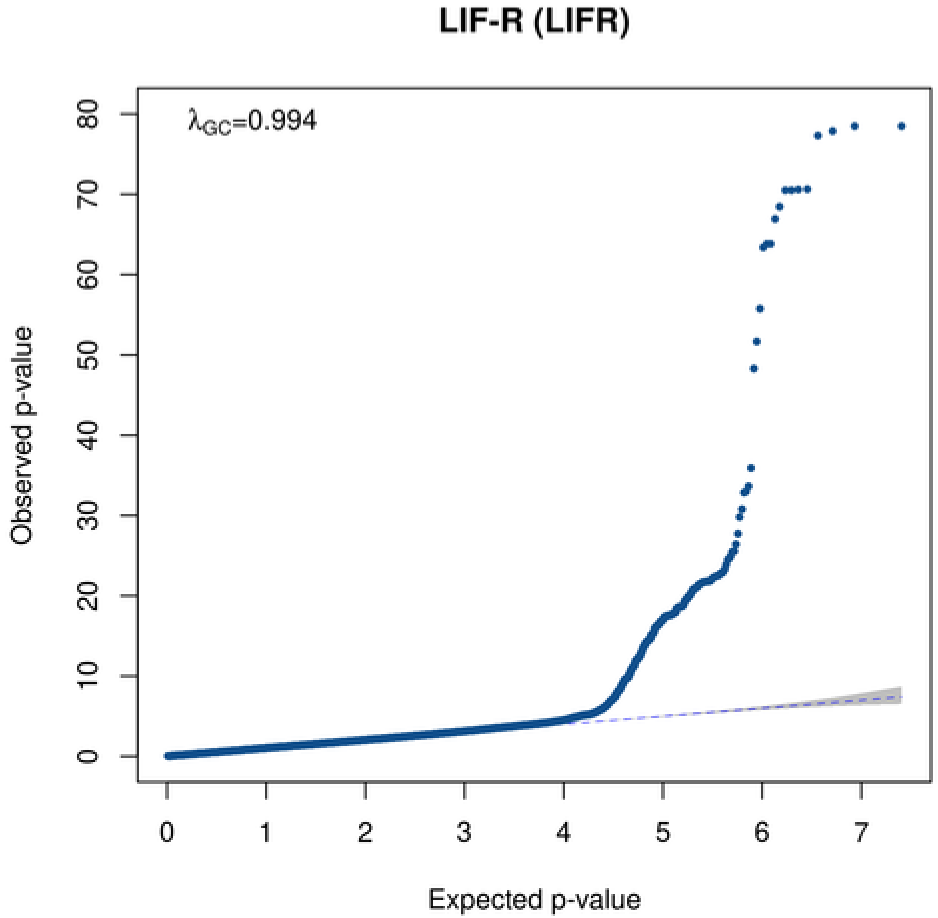

LIF-R (LIFR)
λGC=0.994
80
70
60
50
40
30
20
10
0
Observed p-value
0
1
2
3
4
5
6
7
Expected p-value

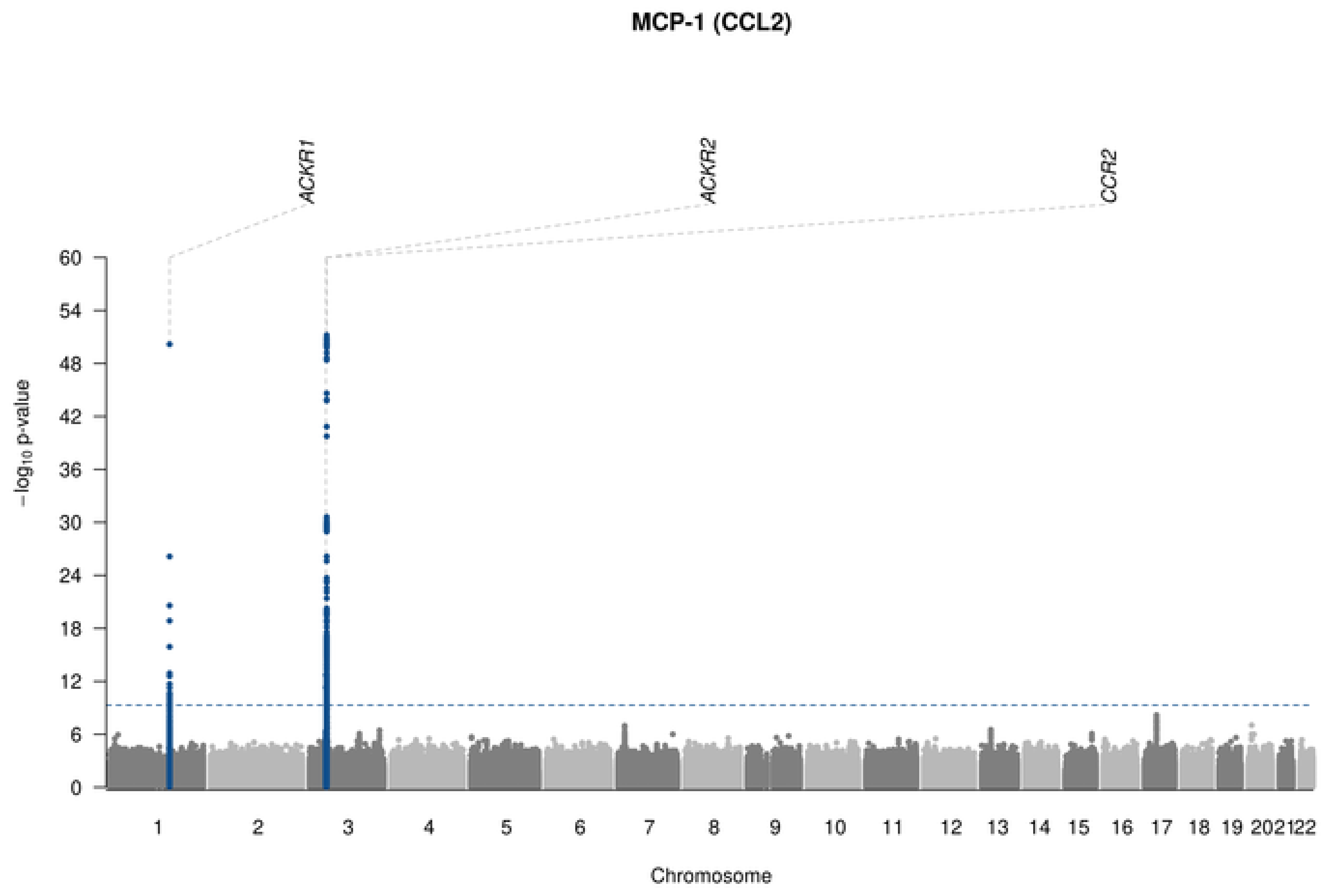

MCP-1 (CCL2)
ACKR1
ACKR2
CCR2
−log10 p-value
Chromosome

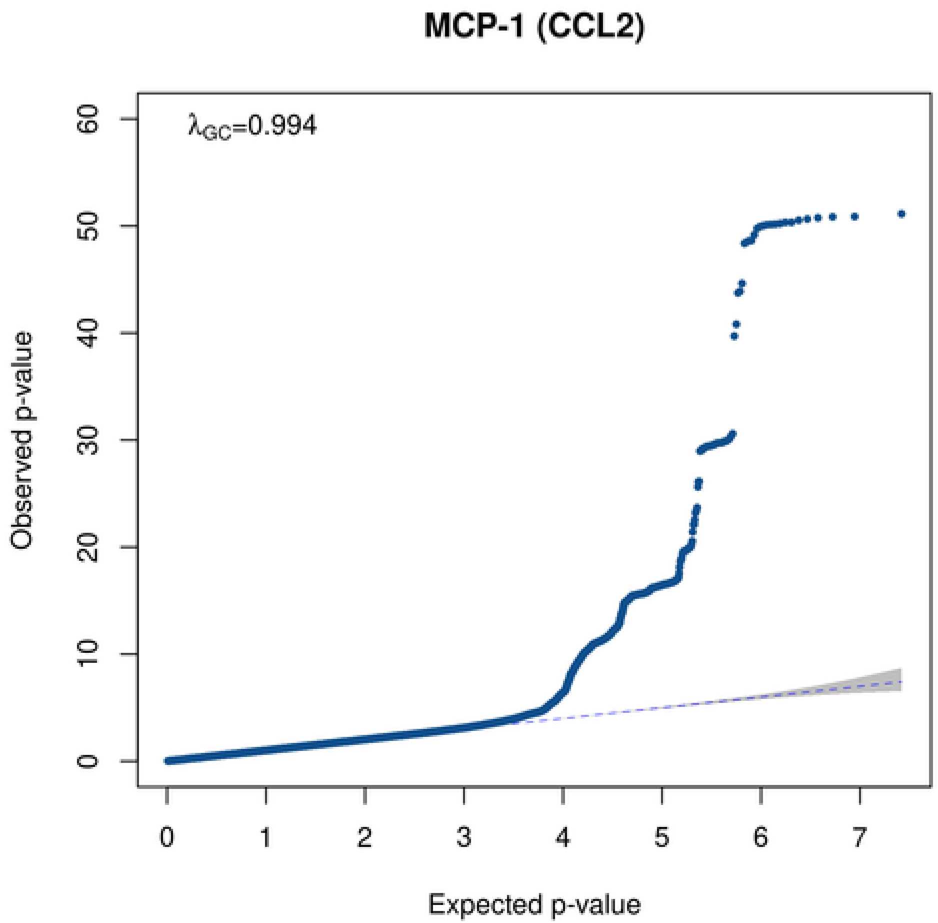

MCP-1 (CCL2)
λGC=0.994
Observed p-value
Expected p-value

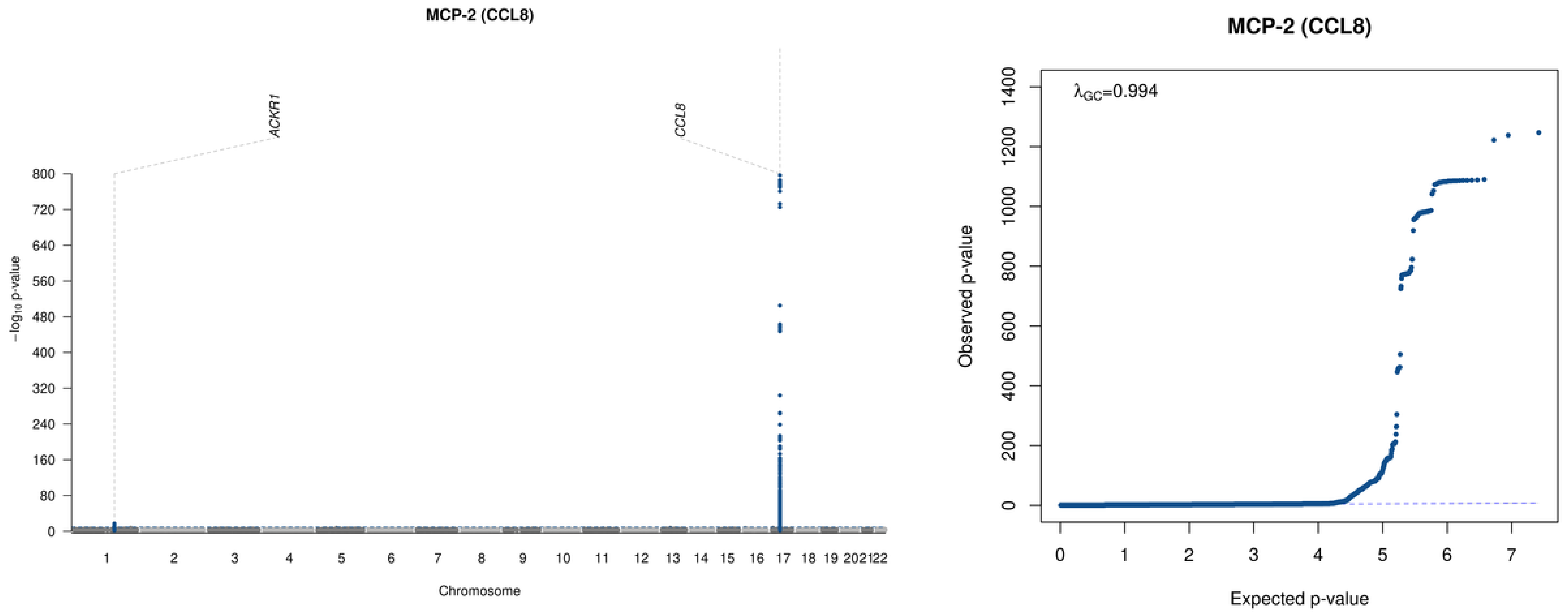

MCP-2 (CCL8)
ACKR1
CCL8
−log10 p-value
800
720
640
560
480
400
320
240
160
80
Chromosome
MCP-2 (CCL8)
λGC=0.994
Observed p-value
Expected p-value
1400
1200
1000
800
600
400
200
0
0
1
2
3
4
5
6
7

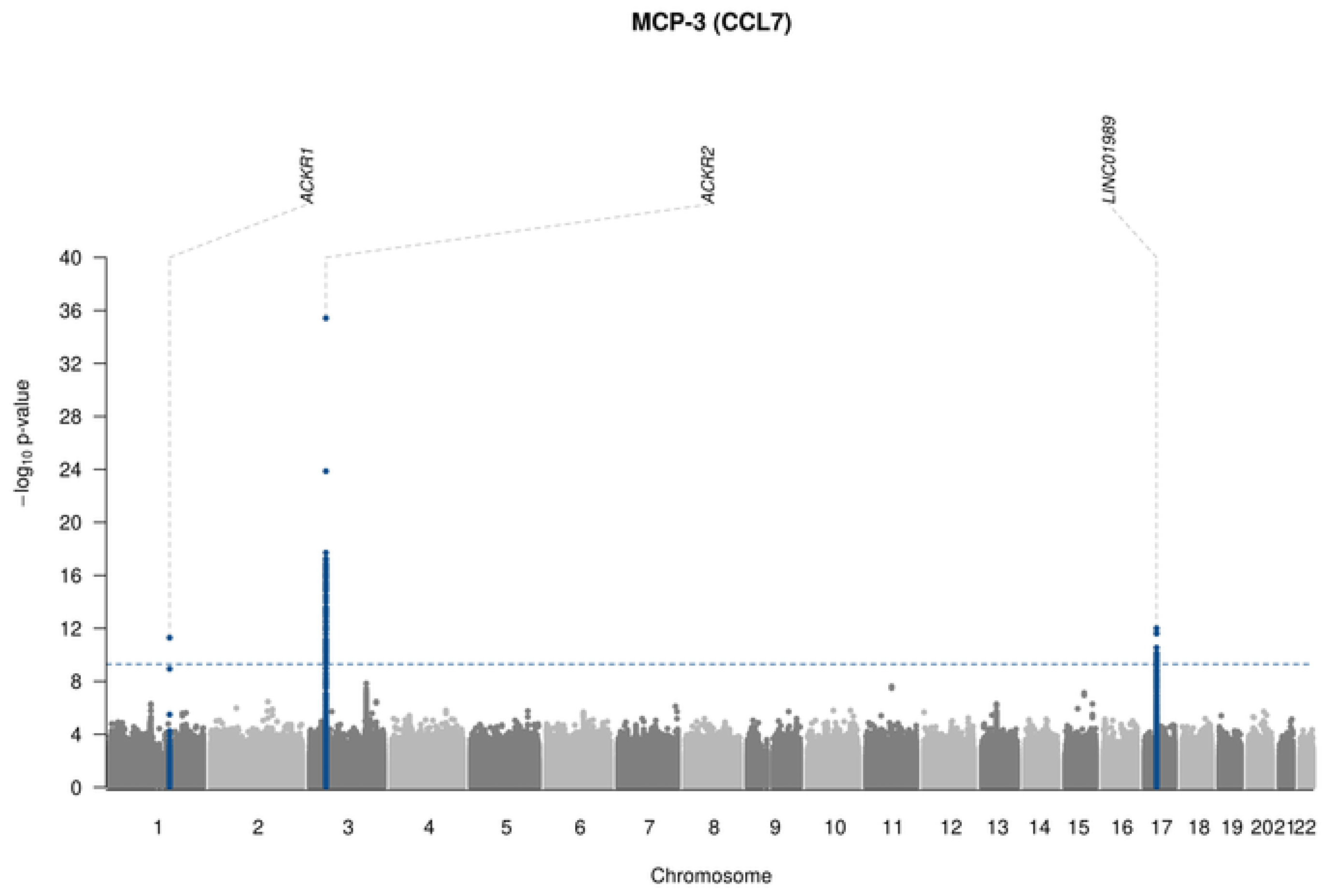
MCP-3 (CCL7)
ACKR1
ACKR2
LINC01989
−log10 p-value
40
36
32
28
24
20
16
12
8
4
0
1 2 3 4 5 6 7 8 9 10 11 12 13 14 15 16 17 18 19 20 21 22
Chromosome

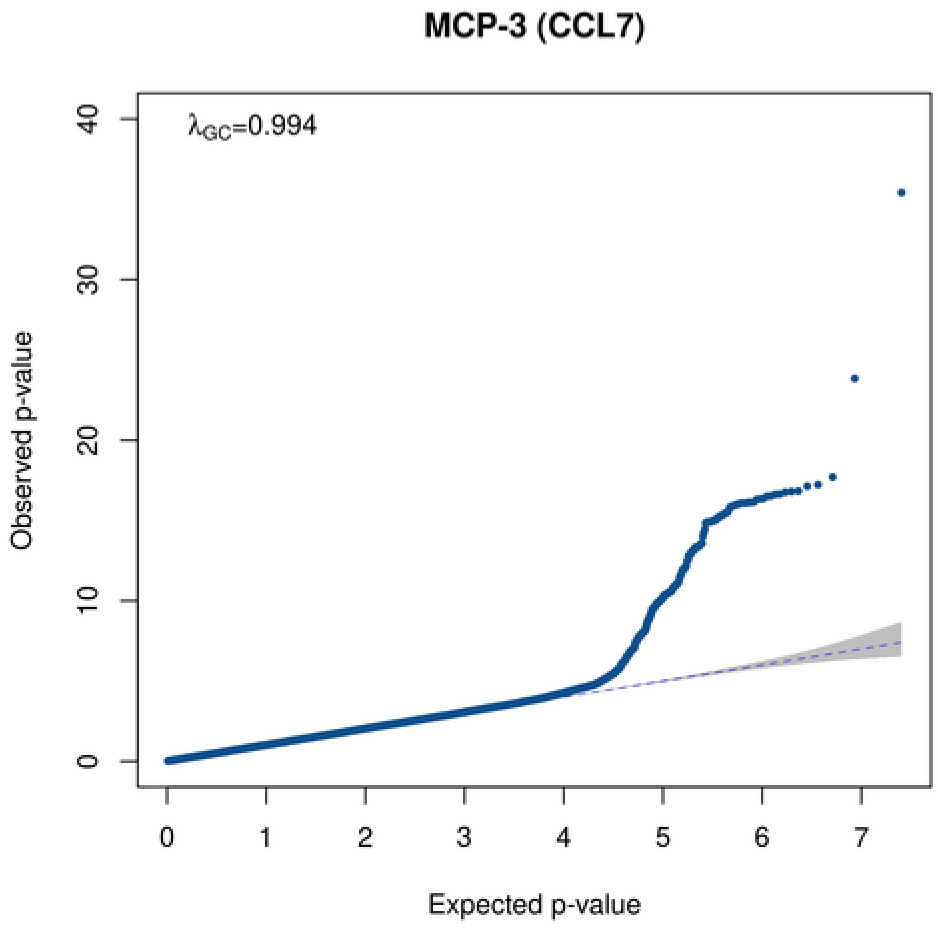
MCP-3 (CCL7)
λGC=0.994
Observed p-value
0 10 20 30 40
0 1 2 3 4 5 6 7
Expected p-value

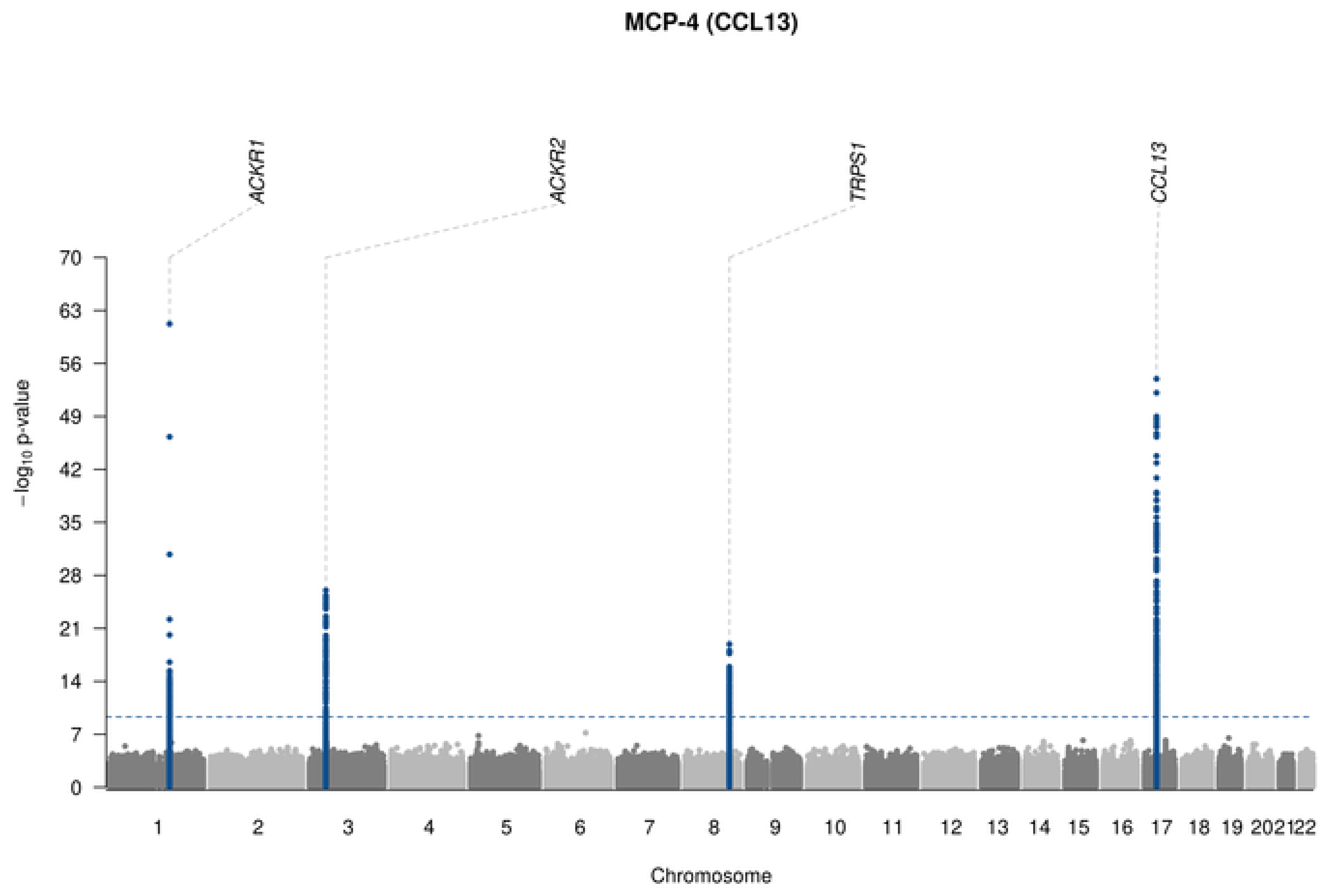

MCP-4 (CCL13)
ACKR1
ACKR2
TRPS1
CCL13
−log10 p-value
70
63
56
49
42
35
28
21
14
7
202122
Chromosome

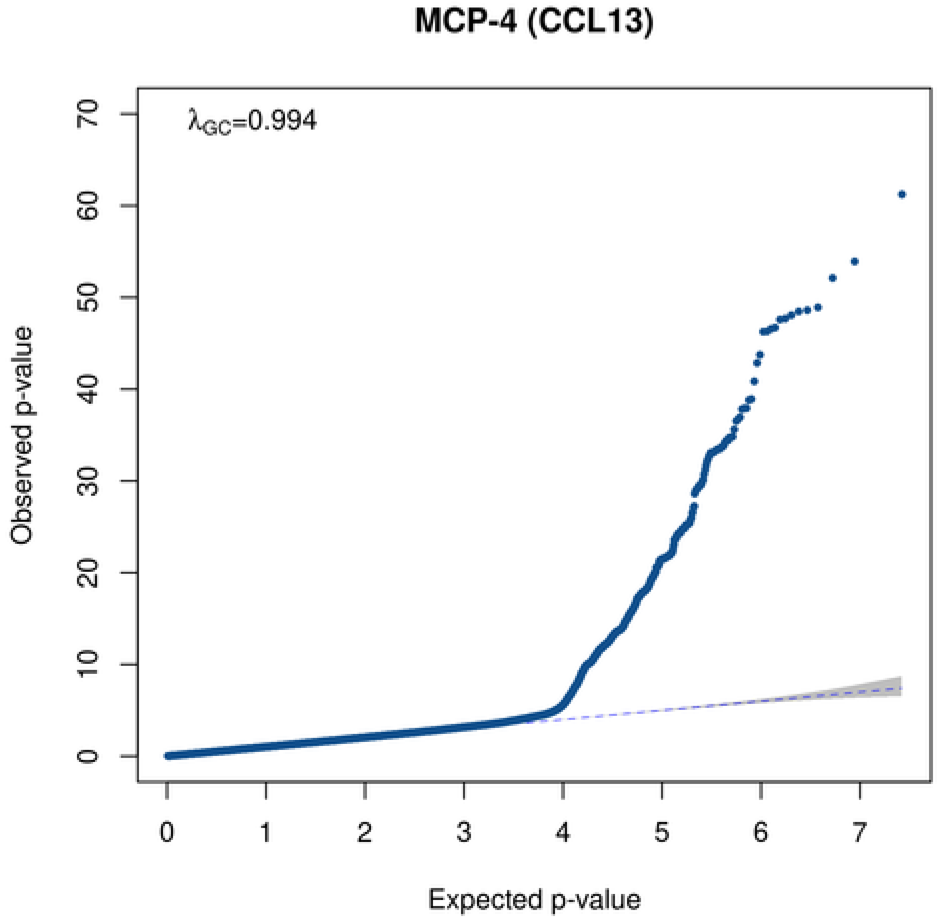

MCP-4 (CCL13)
λGC=0.994
Observed p-value
Expected p-value
70
60
50
40
30
20
10
0
0
1
2
3
4
5
6
7

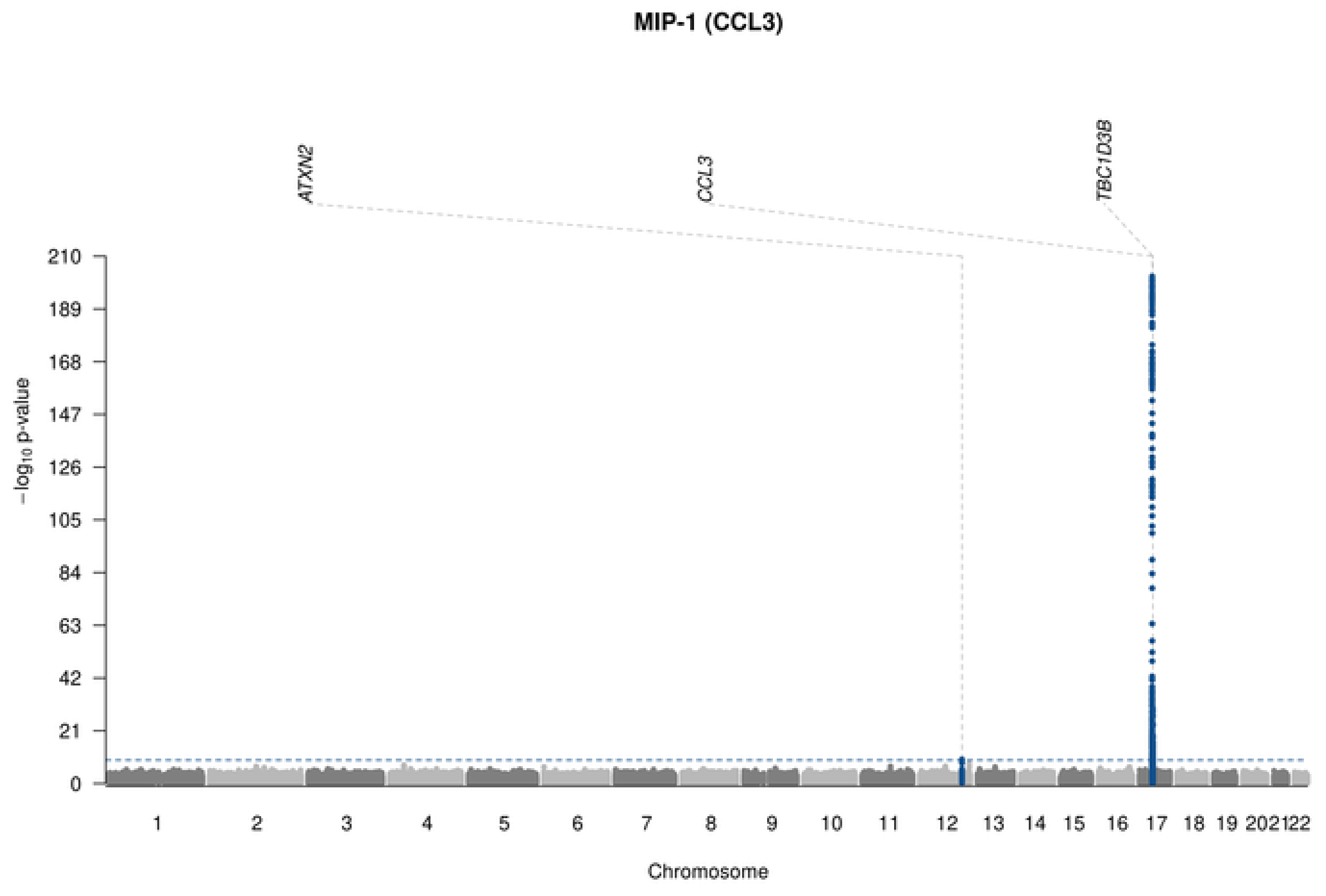

MIP-1 (CCL3)
ATXN2
CCL3
TBC1D3B
210
189
168
147
126
105
84
63
42
21
0
−log10 p-value
2021
22
Chromosome

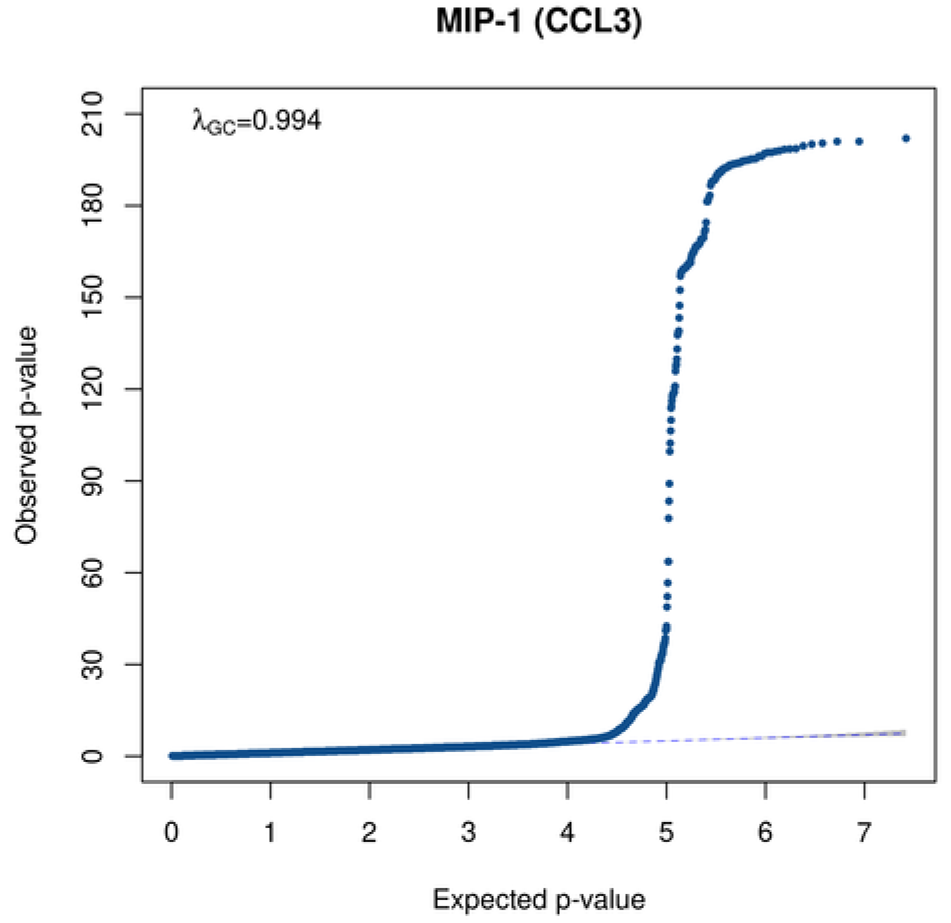

MIP-1 (CCL3)
λGC=0.994
210
180
150
120
90
60
30
0
Observed p-value
0
1
2
3
4
5
6
7
Expected p-value

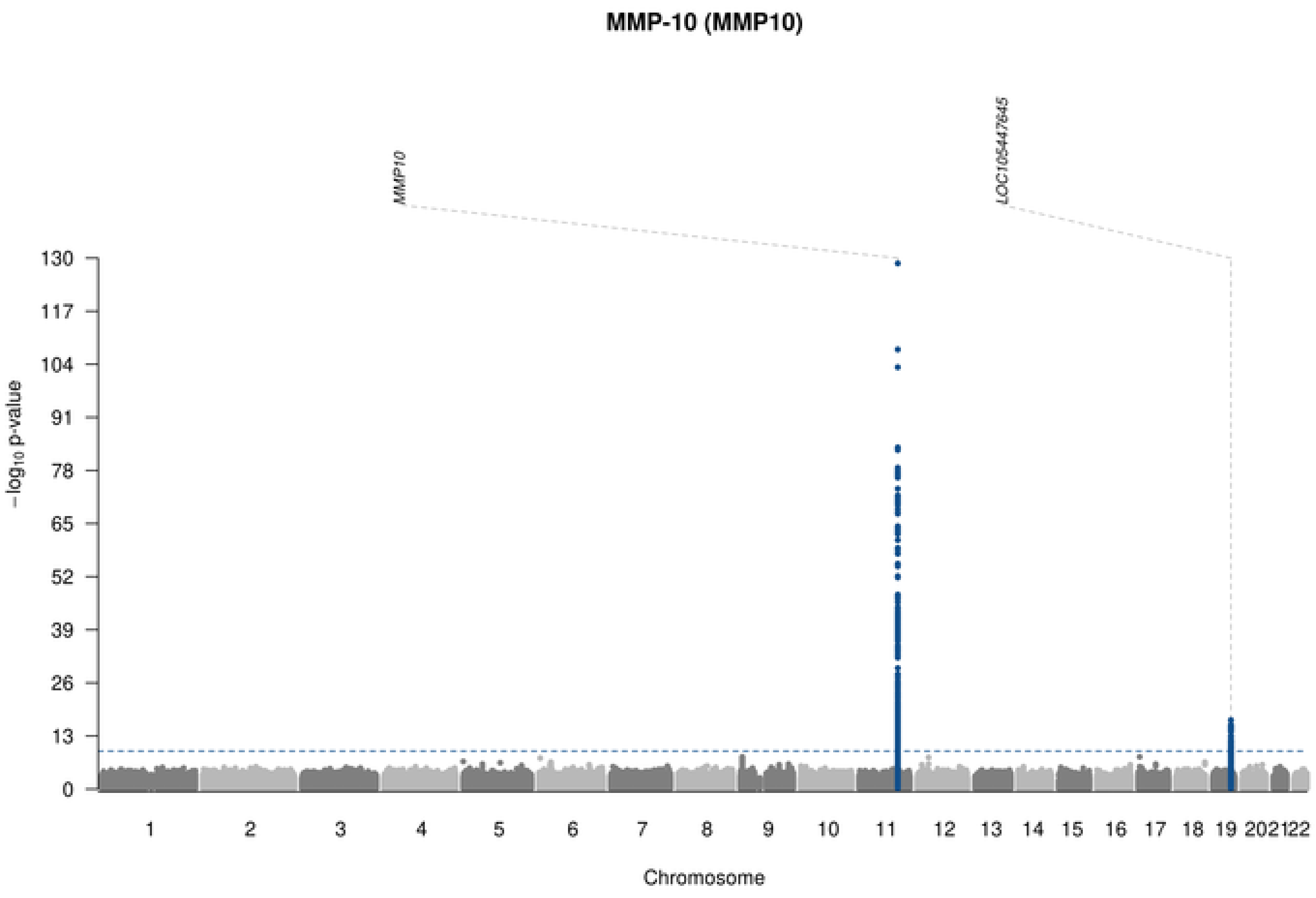
MMP-10 (MMP10)
MMP10
LOC105447645
−log10 p-value
130
117
104
91
78
65
52
39
26
13
Chromosome

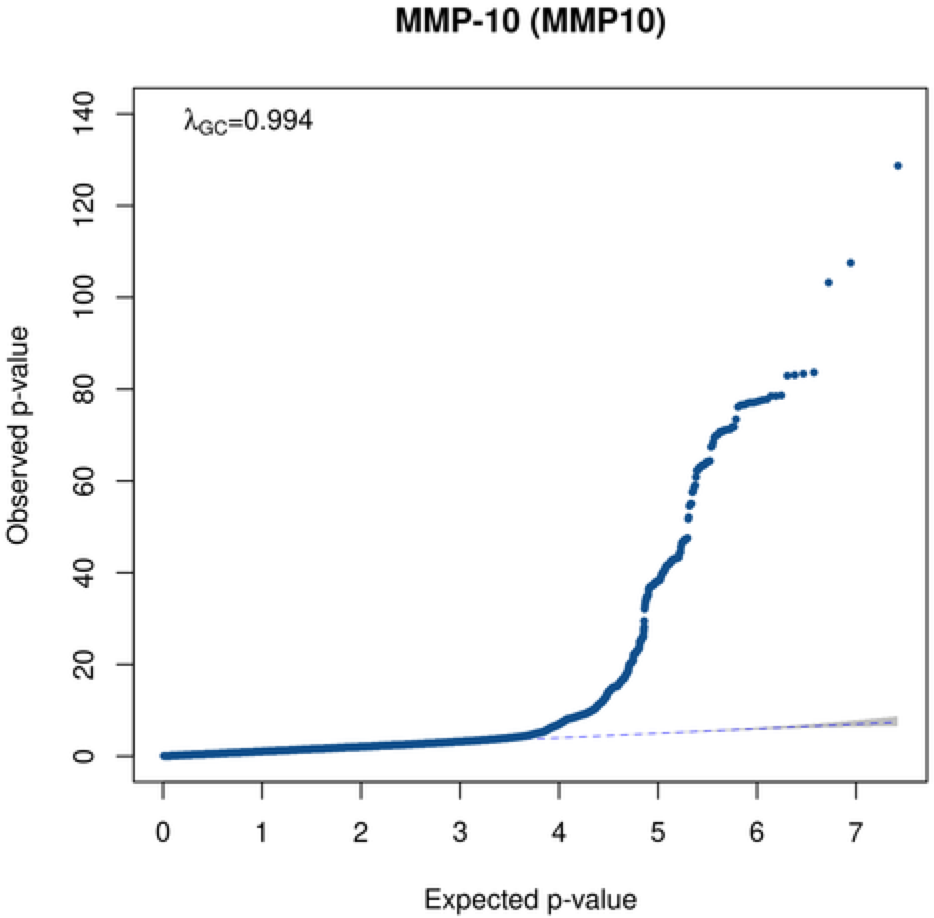
MMP-10 (MMP10)
λGC=0.994
Observed p-value
140
120
100
80
60
40
20
0
1
2
3
4
5
6
7
Expected p-value

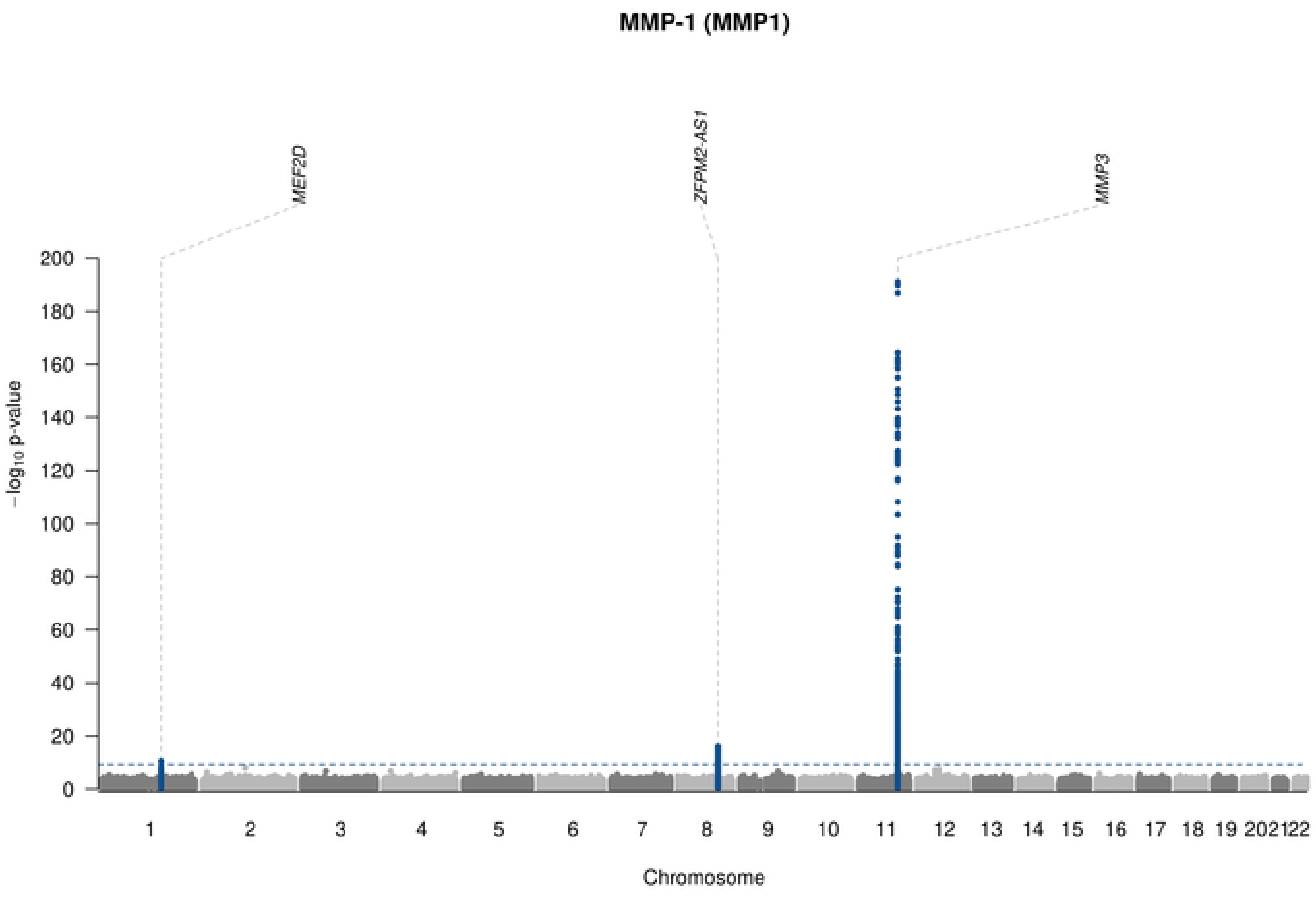

MMP-1 (MMP1)
MEF2D
ZFPM2-AS1
MMP3
−log10 p-value
Chromosome

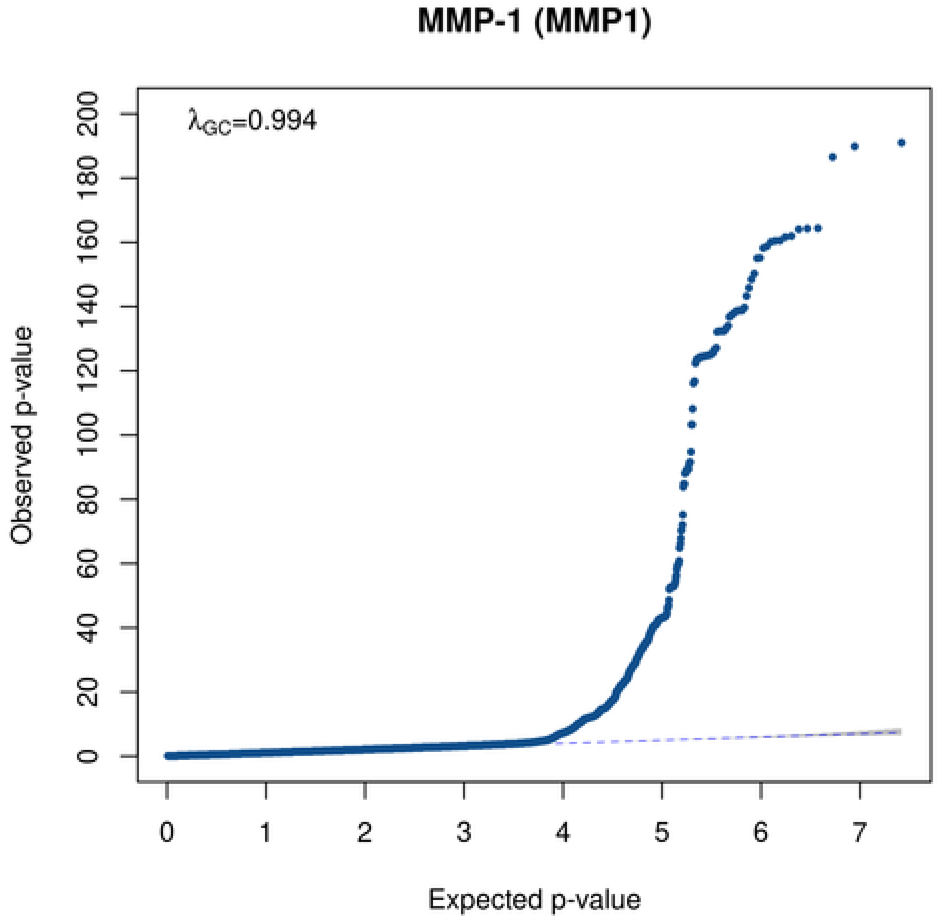

MMP-1 (MMP1)
λGC=0.994
Observed p-value
Expected p-value

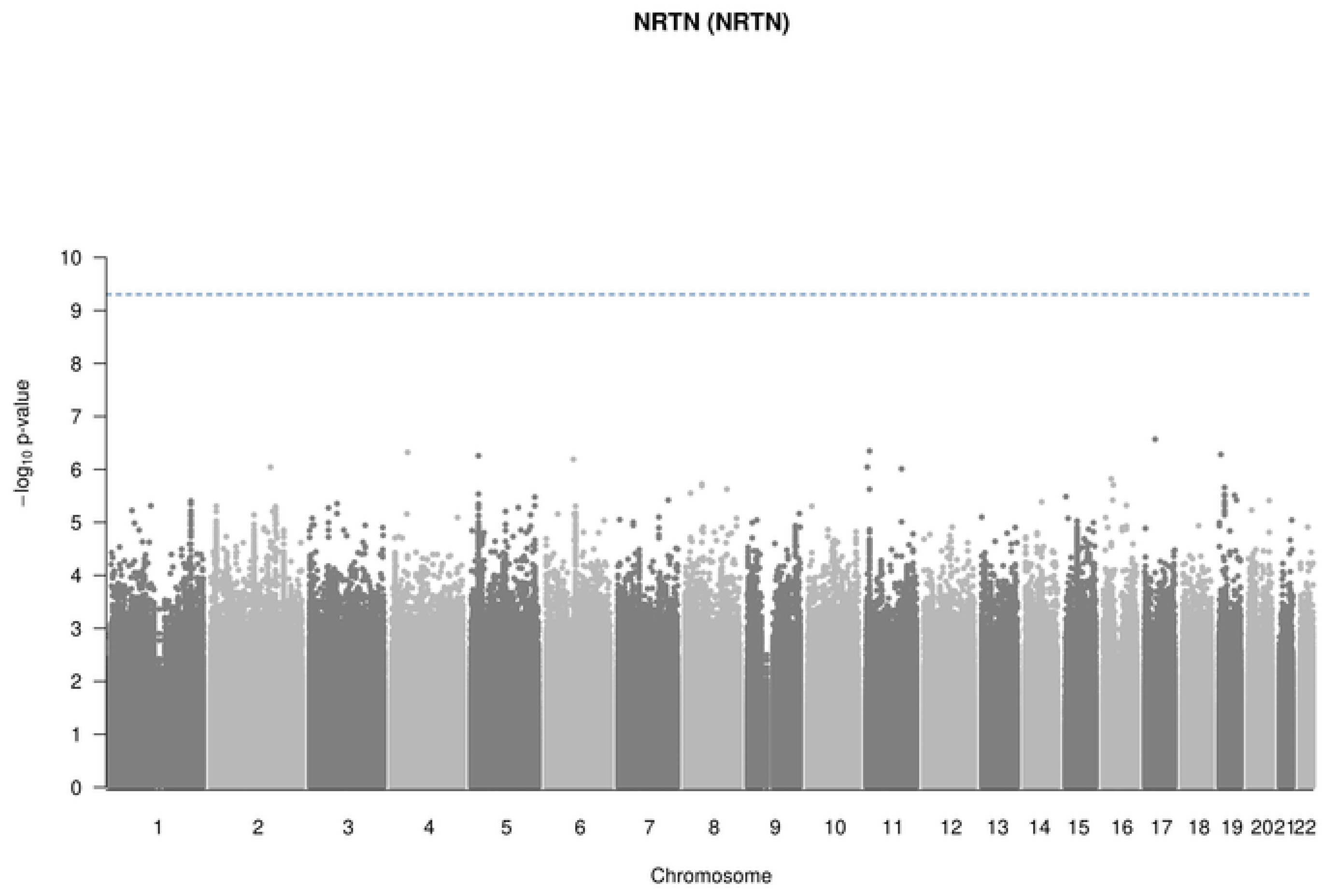
NRTN (NRTN)
-log10 p-value
Chromosome

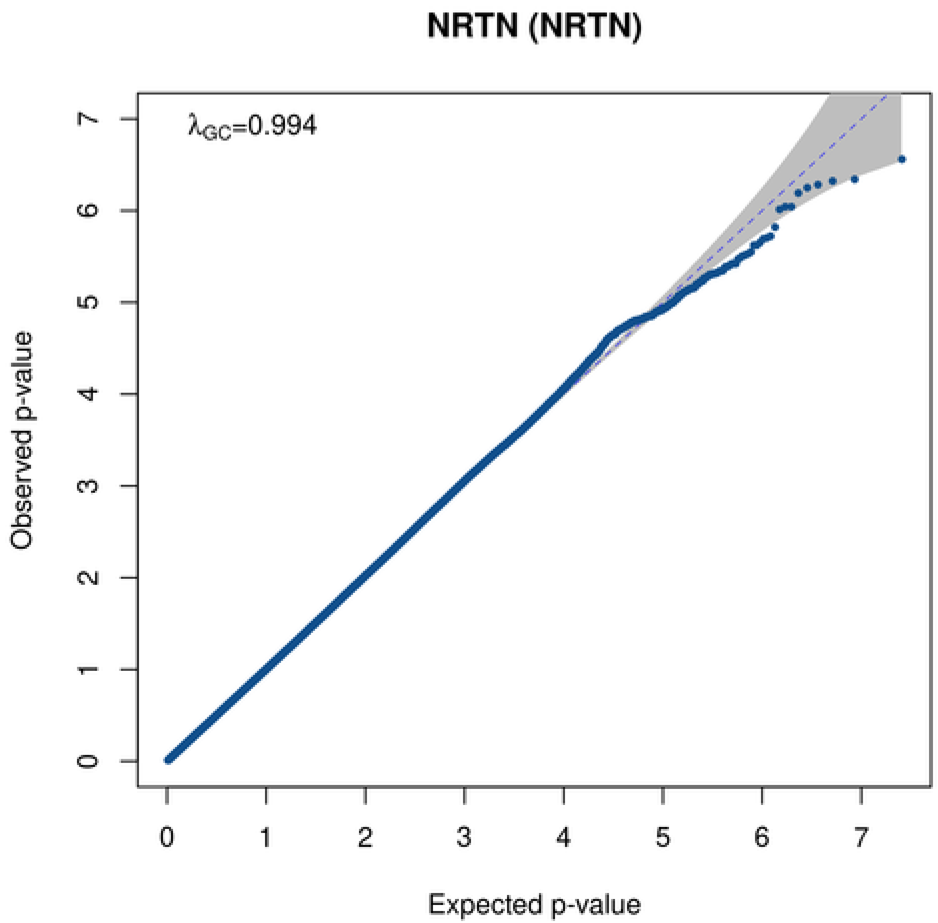
NRTN (NRTN)
λGC=0.994
Observed p-value
Expected p-value

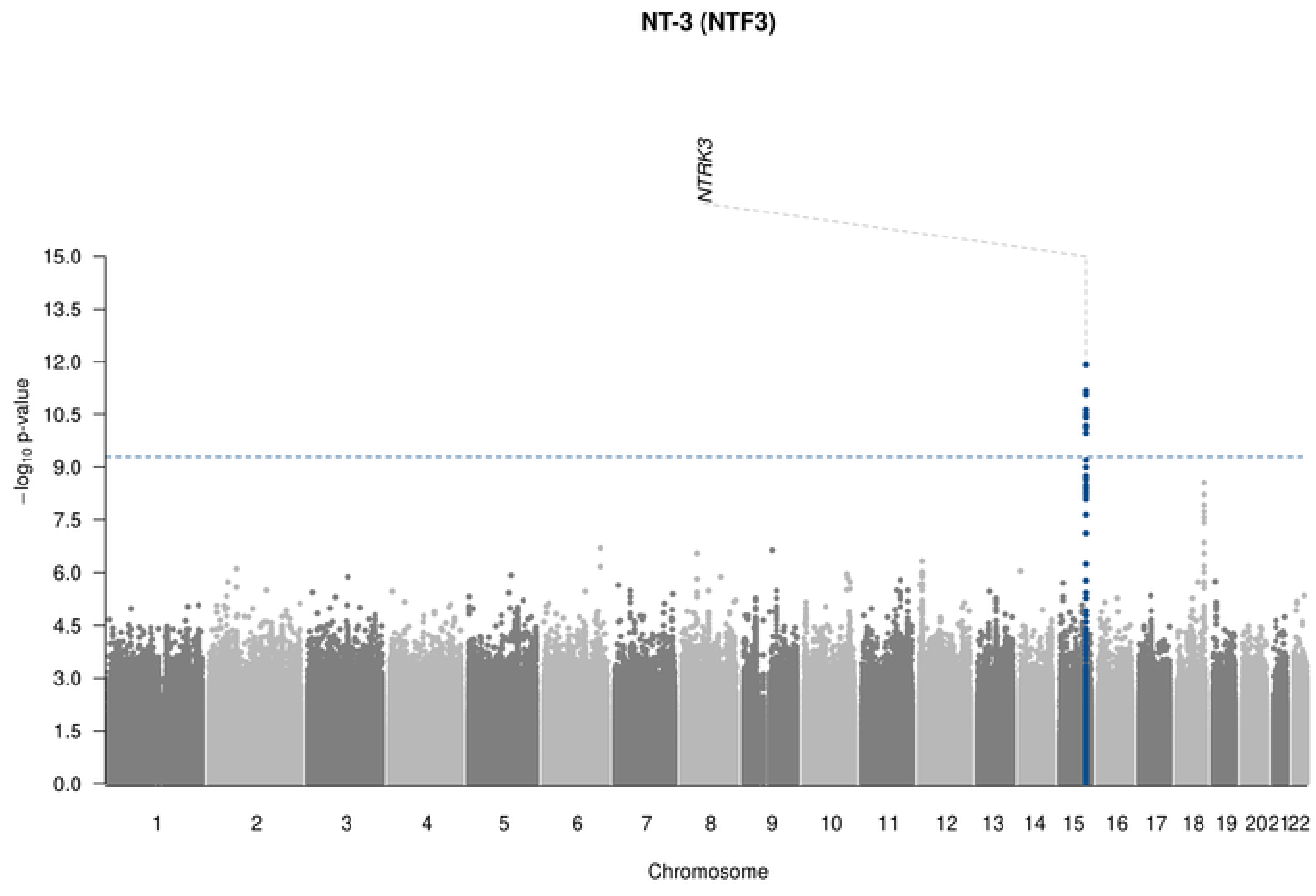

NT-3 (NTF3)
NTRK3
−log10 p-value
15.0
13.5
12.0
10.5
9.0
7.5
6.0
4.5
3.0
1.5
0.0
1 2 3 4 5 6 7 8 9 10 11 12 13 14 15 16 17 18 19 202122
Chromosome

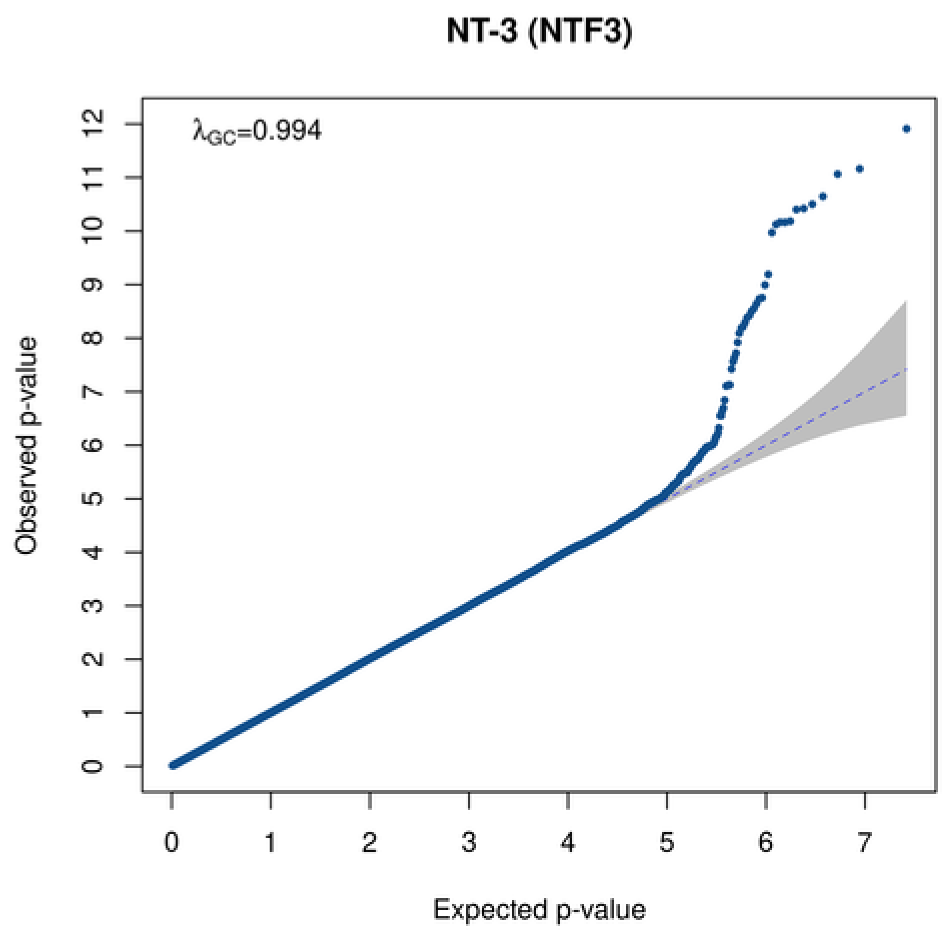

NT-3 (NTF3)
λGC=0.994
Observed p-value
0 1 2 3 4 5 6 7 8 9 10 11 12
0 1 2 3 4 5 6 7
Expected p-value

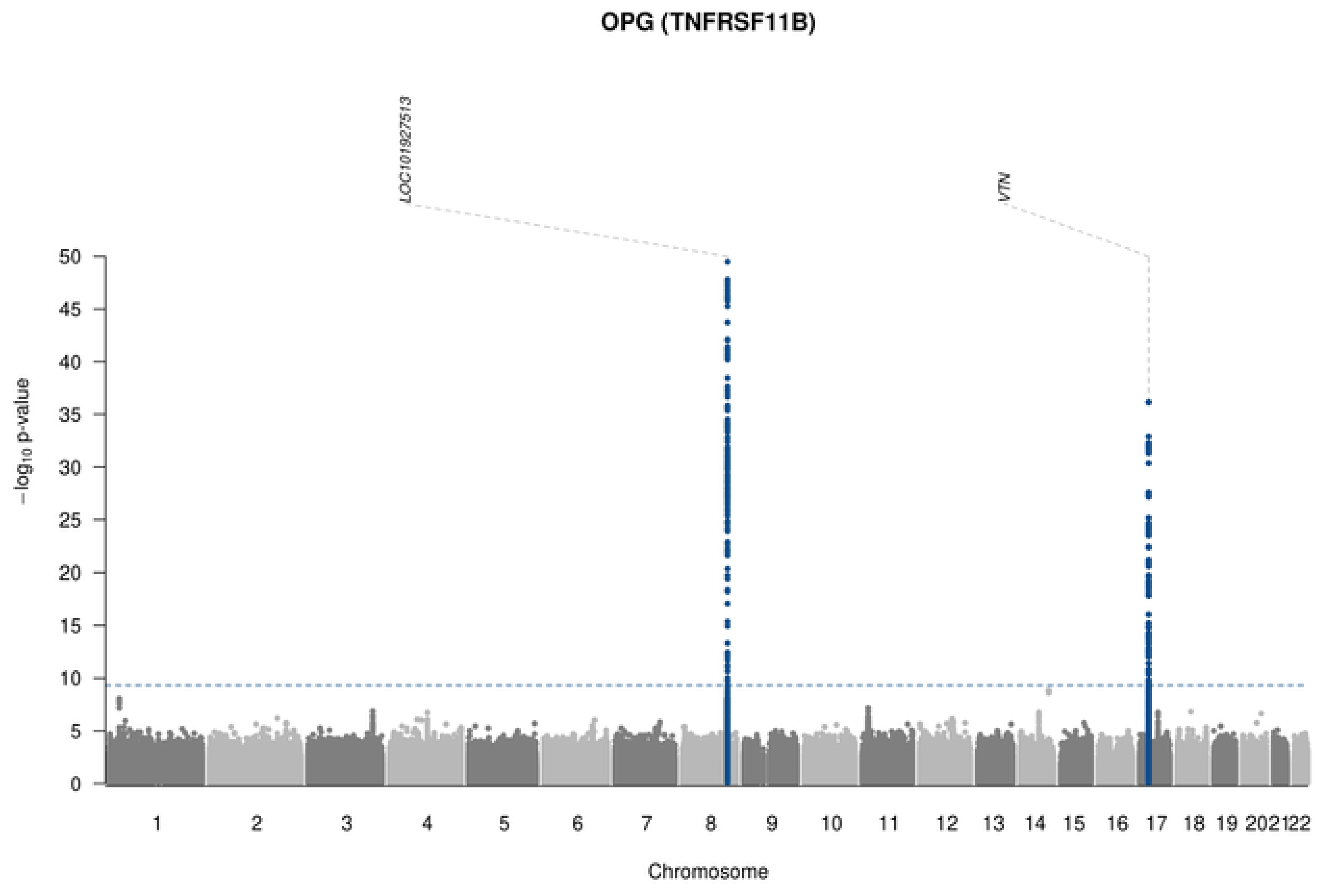
OPG (TNFRSF11B)
LOC101927513
VTN
−log10 p-value
0
5
10
15
20
25
30
35
40
45
50
202122
Chromosome

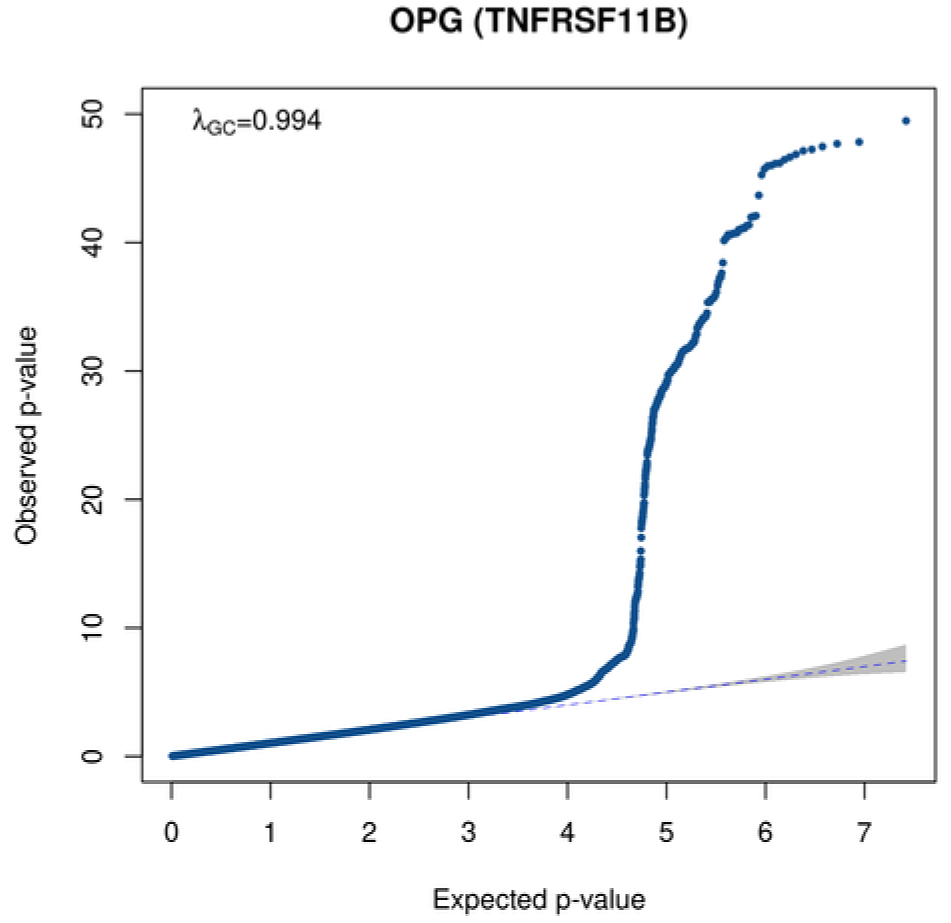
OPG (TNFRSF11B)
λGC=0.994
Observed p-value
0
10
20
30
40
50
0
1
2
3
4
5
6
7
Expected p-value

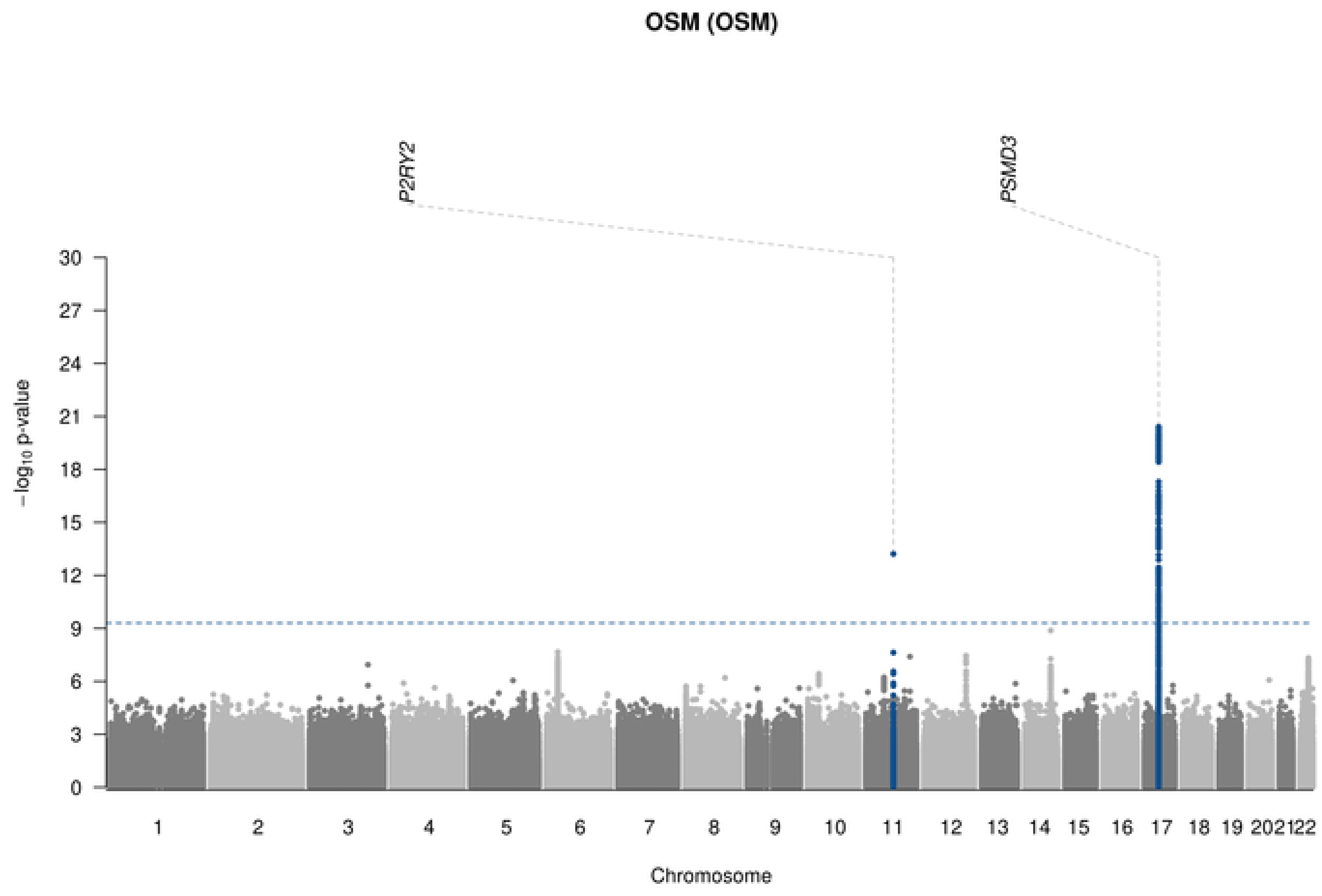

OSM (OSM)
P2RY2
PSMD3
−log10 p-value
Chromosome

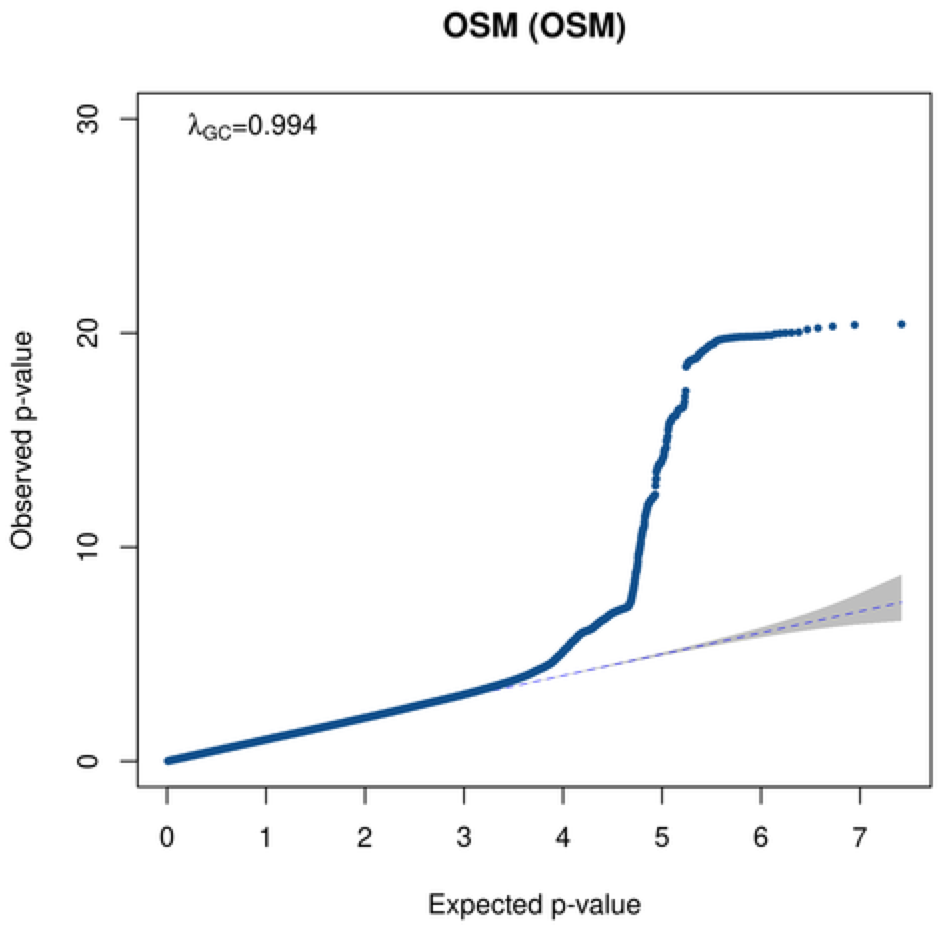

OSM (OSM)
λGC=0.994
Observed p-value
Expected p-value

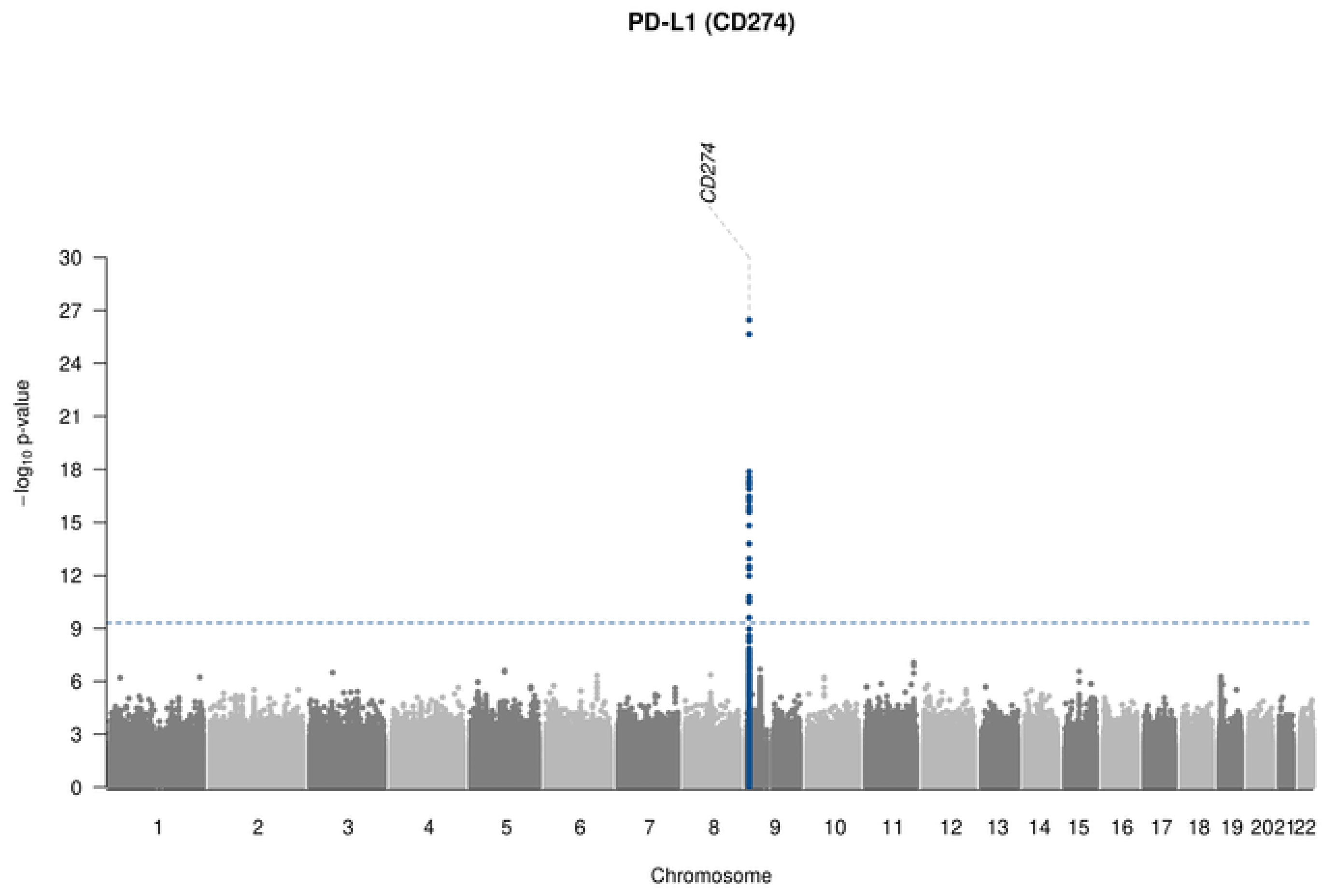

PD-L1 (CD274)
CD274
−log10 p-value
30
27
24
21
18
15
12
9
6
3
Chromosome

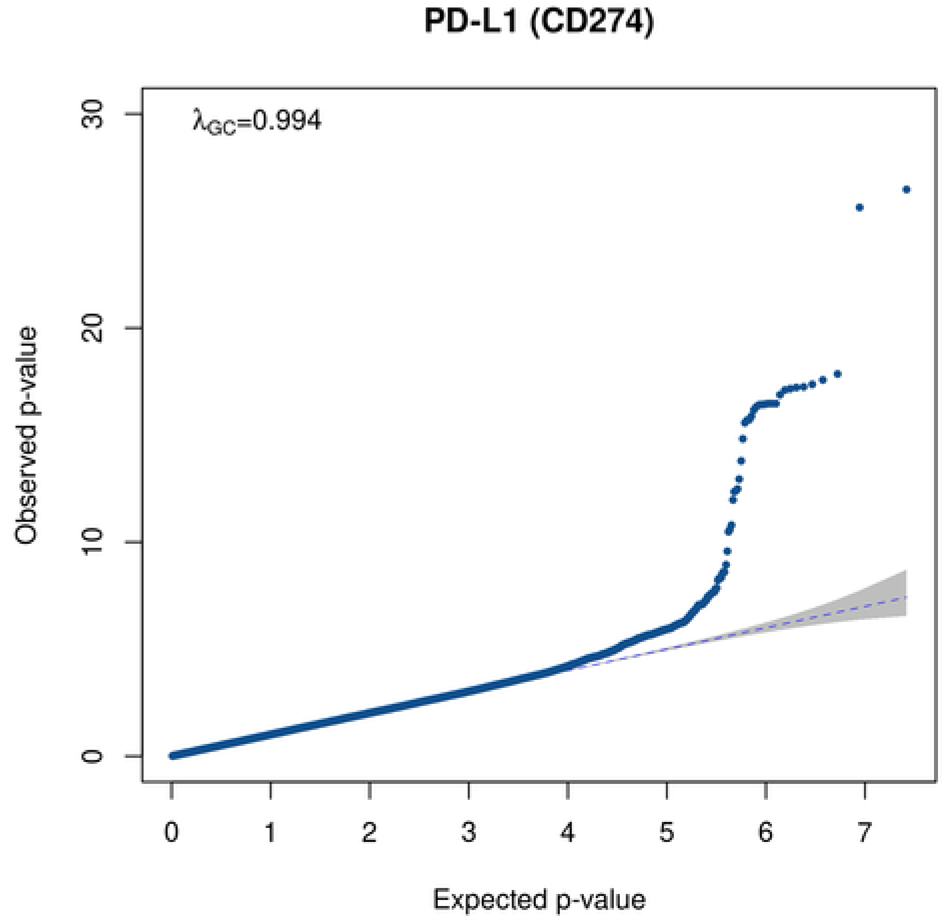

PD-L1 (CD274)
λGC=0.994
Observed p-value
30
20
10
0
0
1
2
3
4
5
6
7
Expected p-value

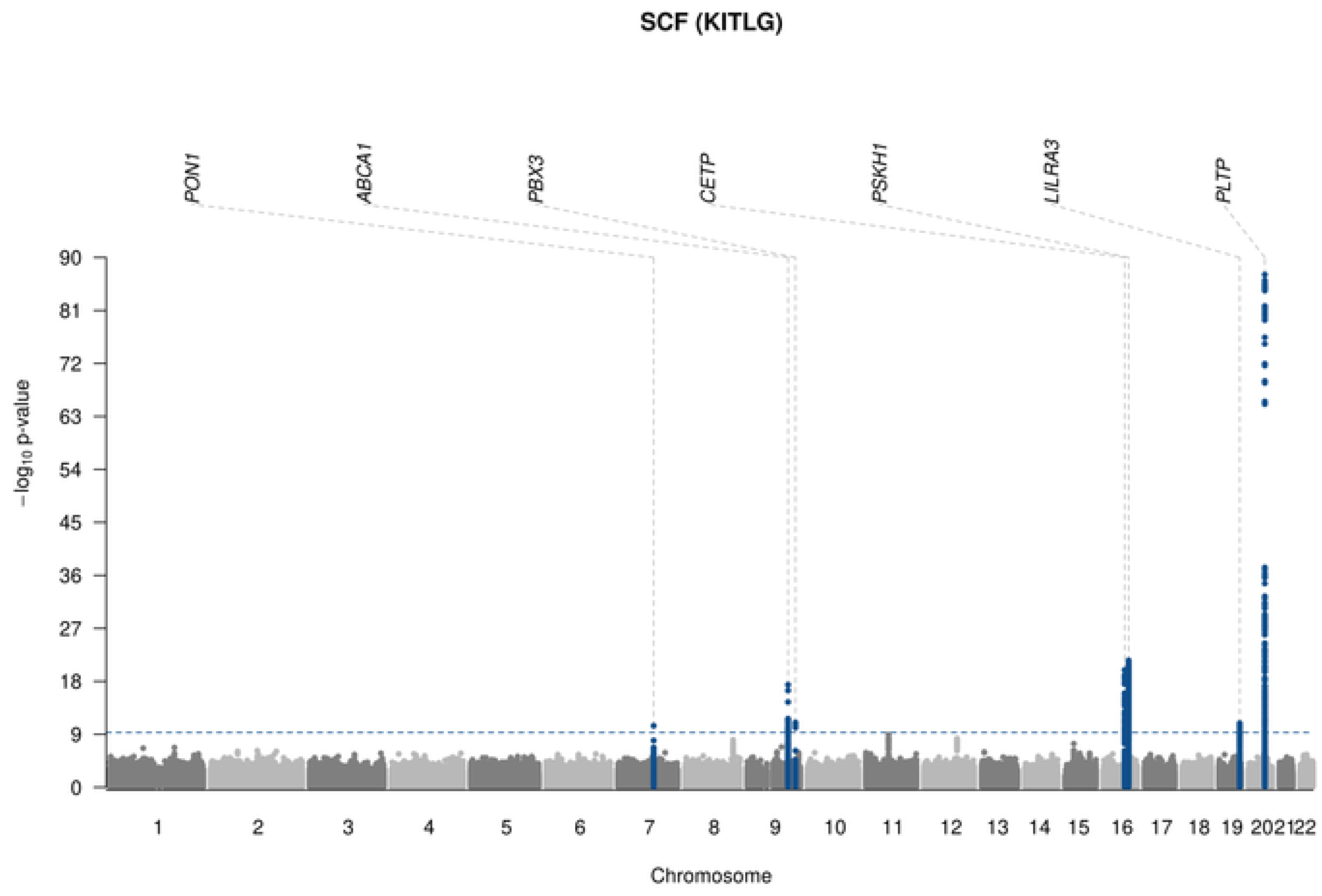
SCF (KITLG)
PON1
ABCA1
PBX3
CETP
PSKH1
LILRA3
PLTP
90
81
72
63
54
45
36
27
18
9
0
−log10 p-value
Chromosome

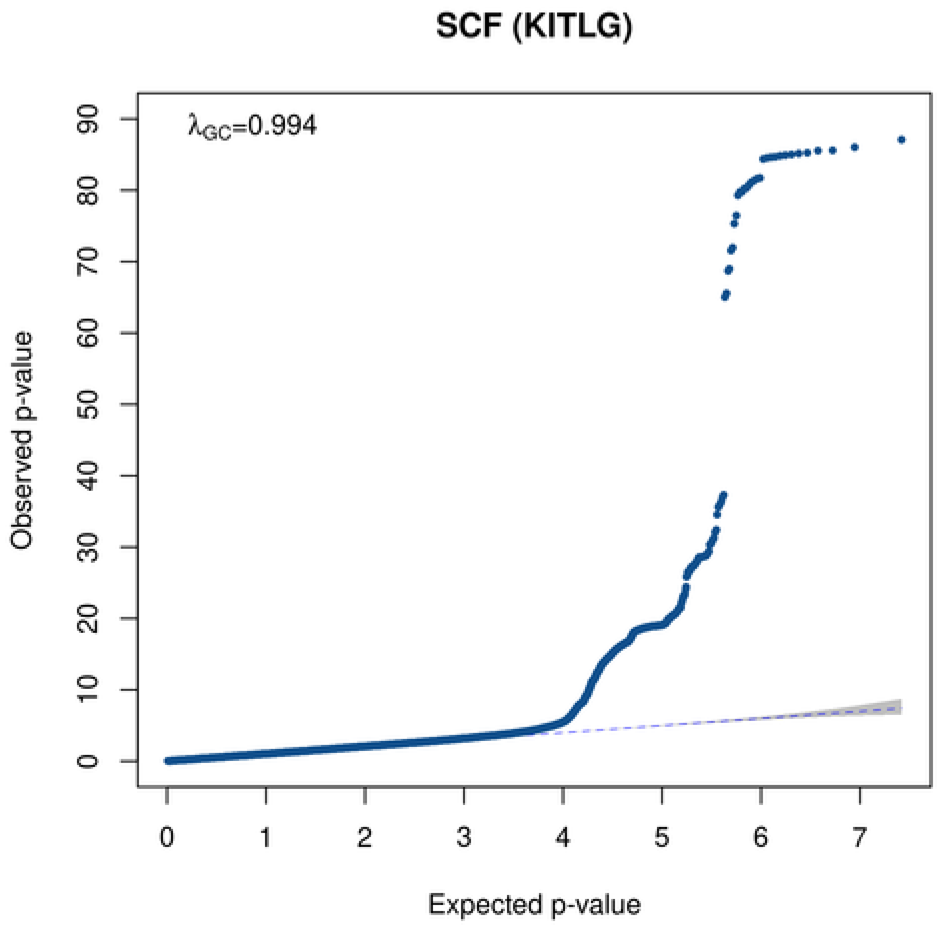
SCF (KITLG)
λGC=0.994
Observed p-value
Expected p-value
0
1
2
3
4
5
6
7
0
10
20
30
40
50
60
70
80
90

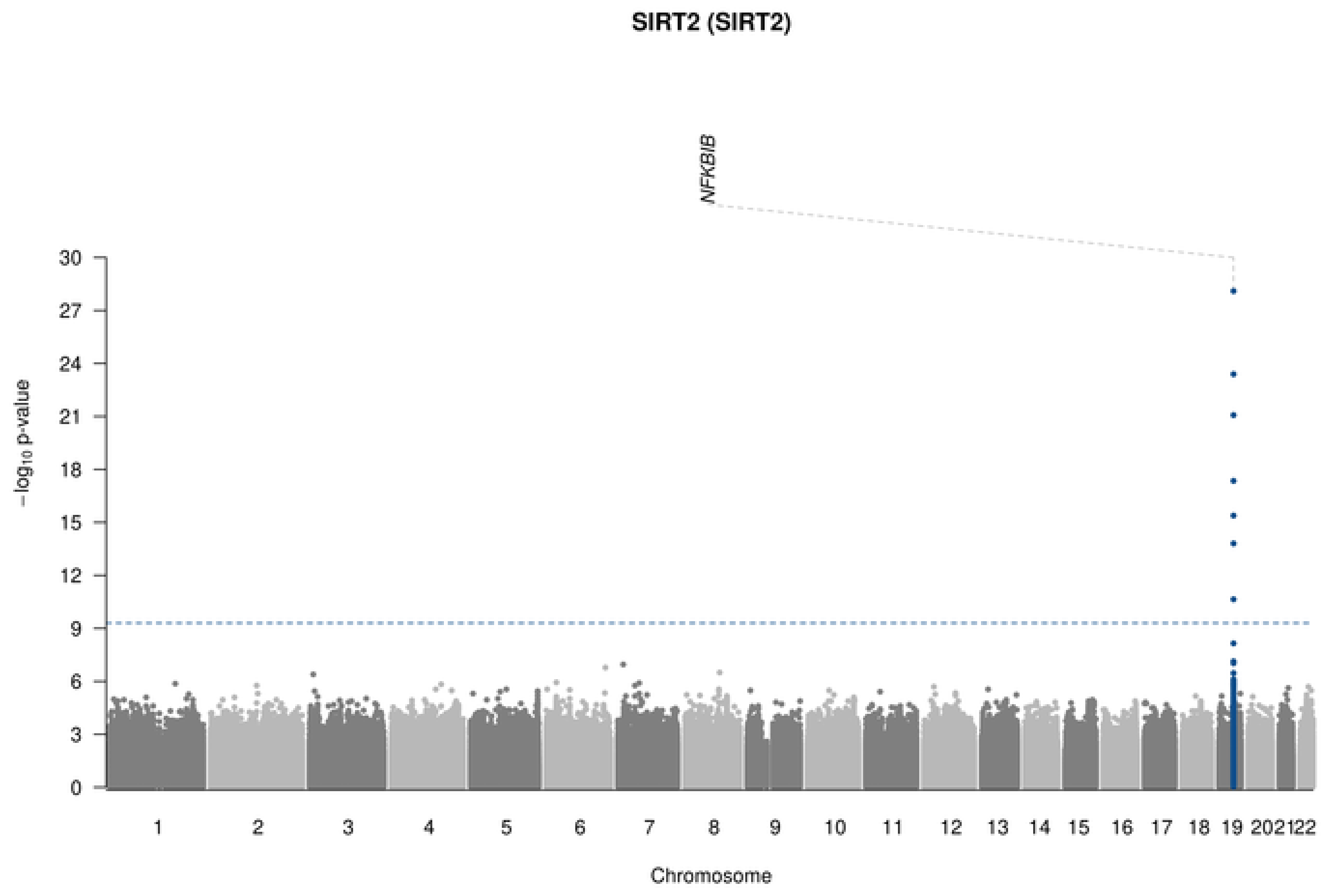

SIRT2 (SIRT2)
NFKBIB
30
27
24
21
18
15
12
9
6
3
0
$-\log_{10}$ p-value
202122
Chromosome

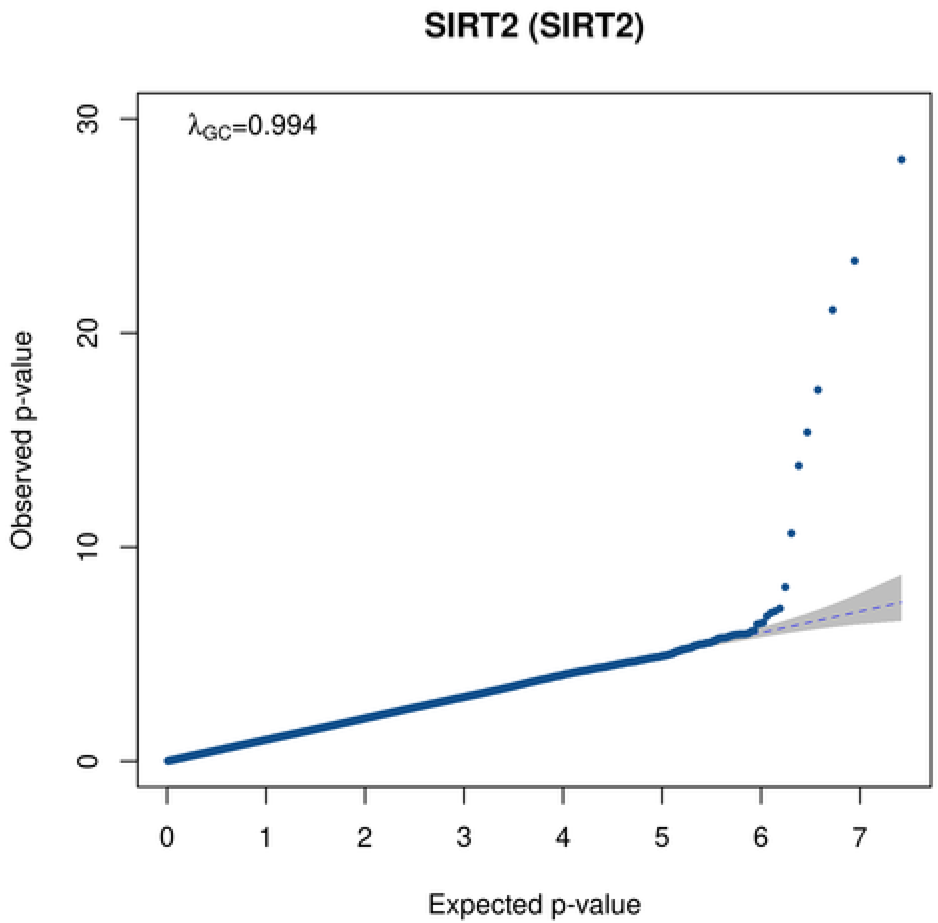

SIRT2 (SIRT2)
$\lambda_{GC}$=0.994
30
20
10
0
Observed p-value
0
1
2
3
4
5
6
7
Expected p-value

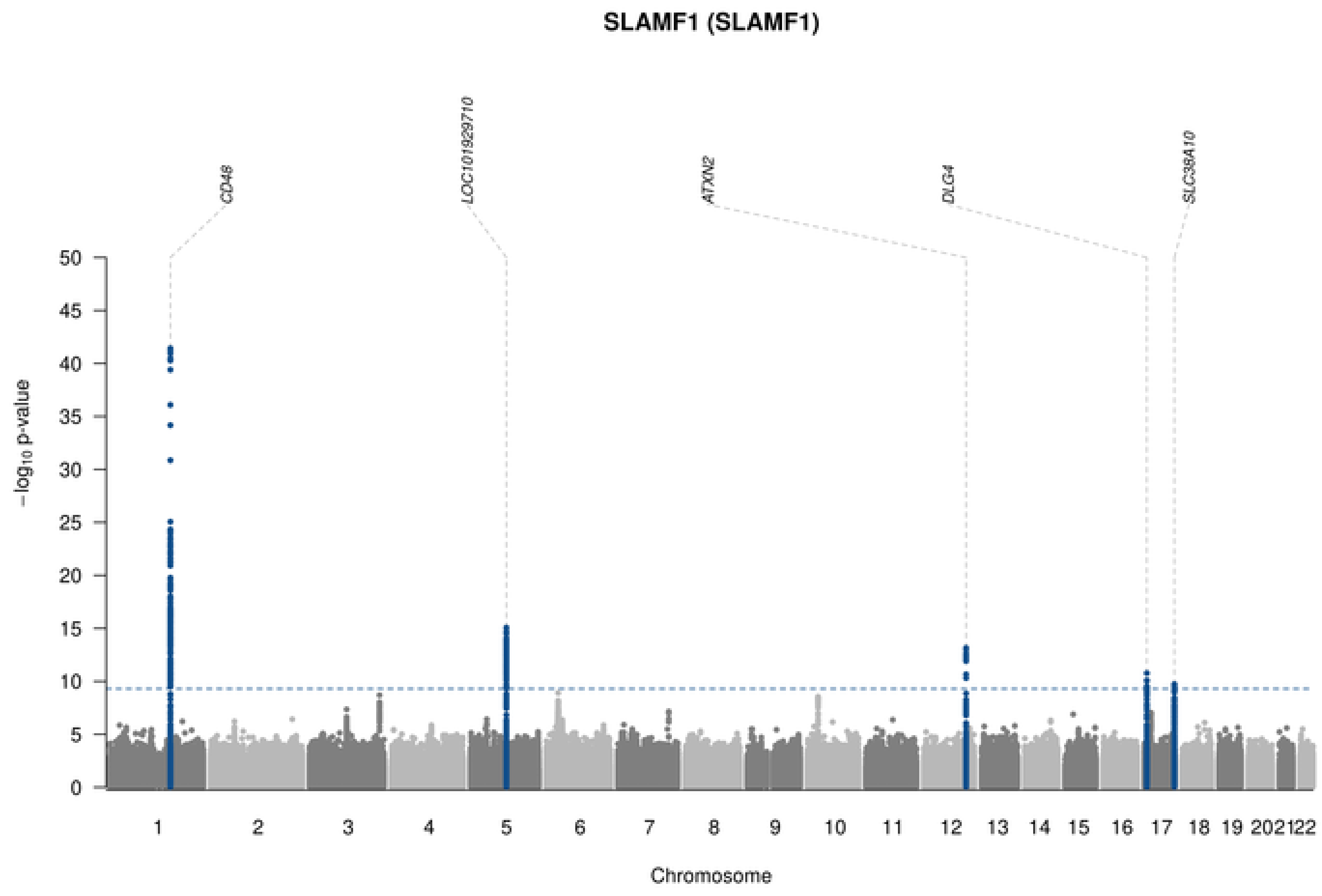
SLAMF1 (SLAMF1)
CD48
LOC101929710
ATXN2
DLG4
SLC38A10
−log10 p-value
Chromosome

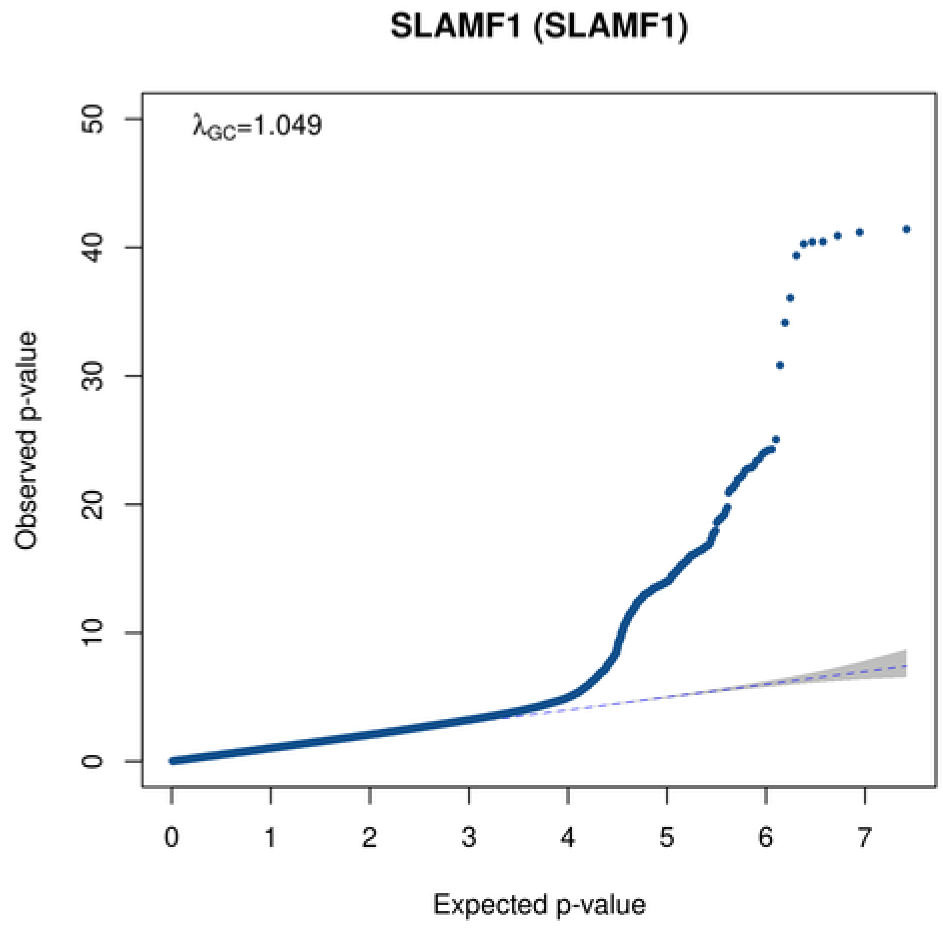
SLAMF1 (SLAMF1)
λGC=1.049
Observed p-value
Expected p-value

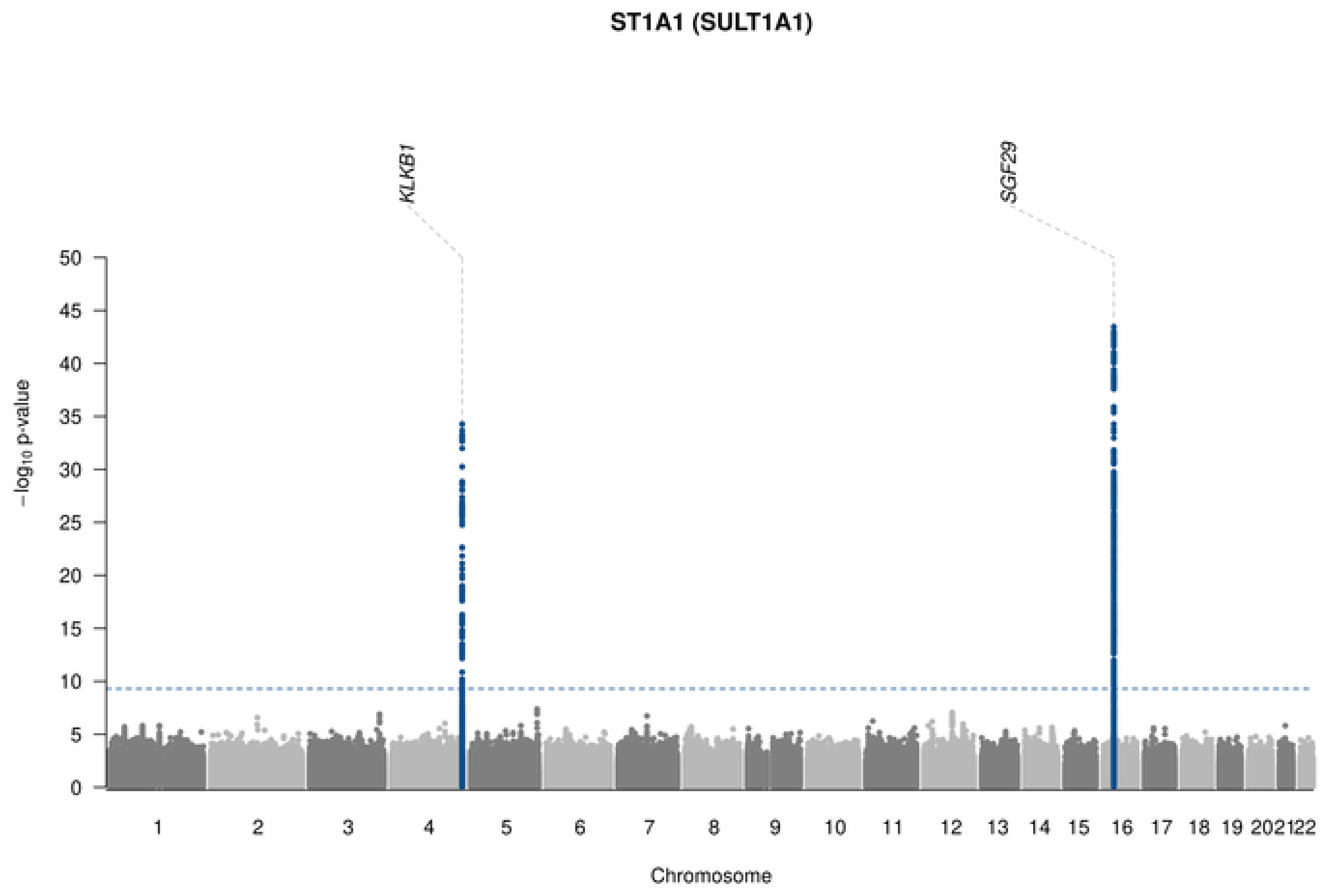
ST1A1 (SULT1A1)
KLKB1
SGF29
−log10 p-value
50
45
40
35
30
25
20
15
10
5
202122
Chromosome

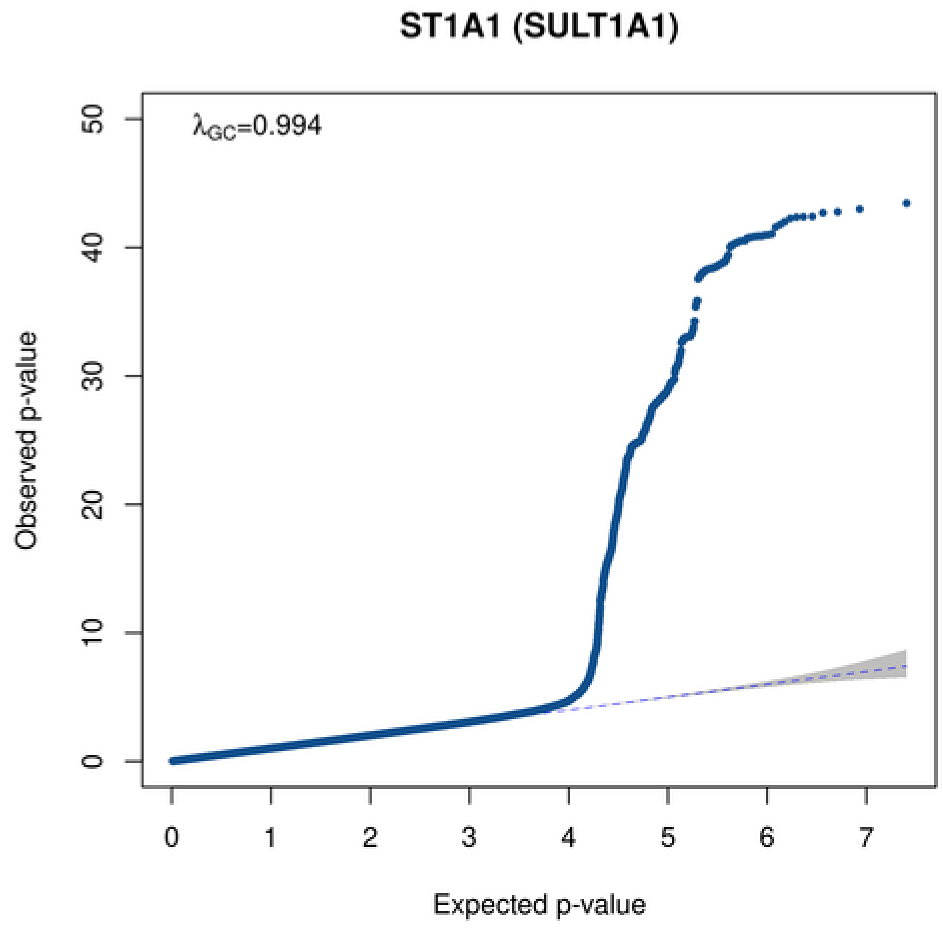
ST1A1 (SULT1A1)
λGC=0.994
Observed p-value
50
40
30
20
10
0
0
1
2
3
4
5
6
7
Expected p-value

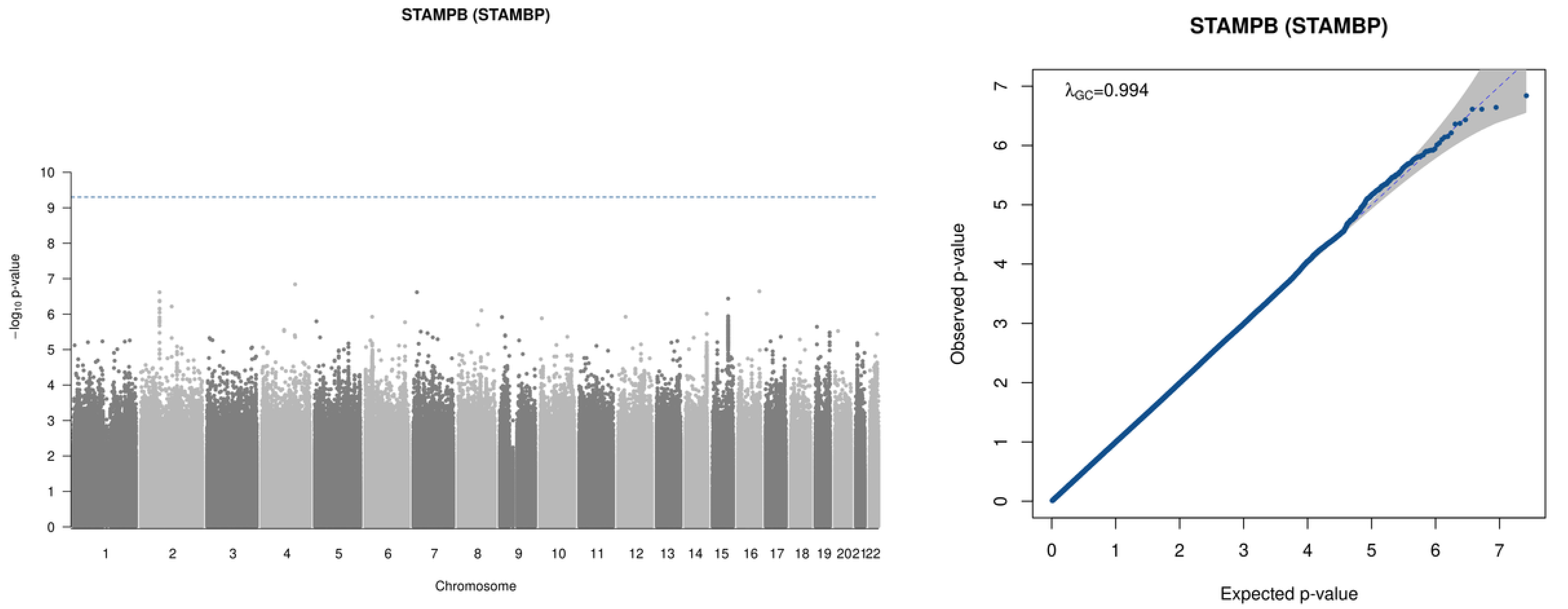

STAMPB (STAMBP)
10
9
8
7
6
5
4
3
2
1
0
−log10 p-value
202122
Chromosome
STAMPB (STAMBP)
λGC=0.994
Observed p-value
Expected p-value
0
1
2
3
4
5
6
7

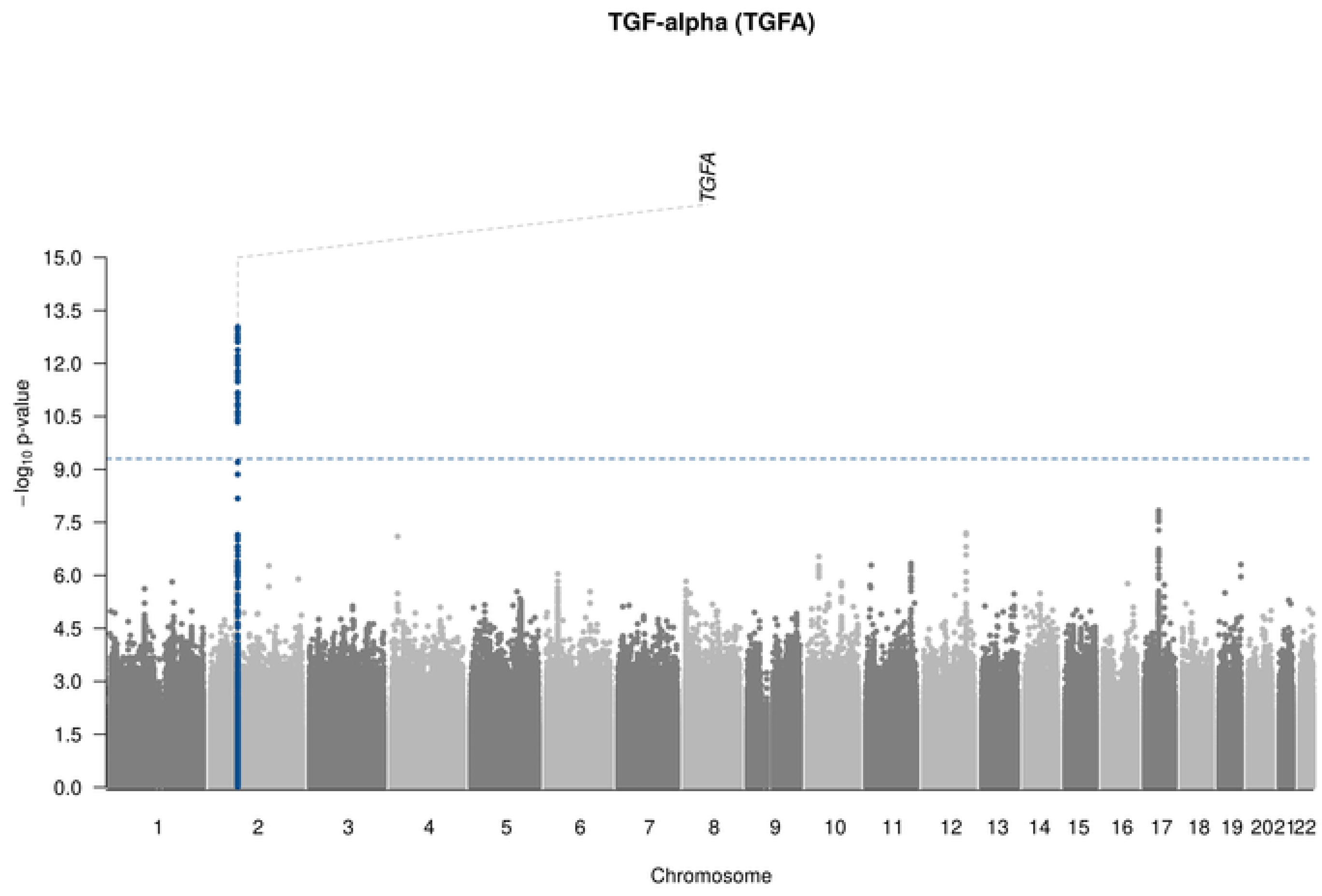
TGF-alpha (TGFA)
TGFA
−log10 p-value
15.0
13.5
12.0
10.5
9.0
7.5
6.0
4.5
3.0
1.5
0.0
1 2 3 4 5 6 7 8 9 10 11 12 13 14 15 16 17 18 19 202122
Chromosome

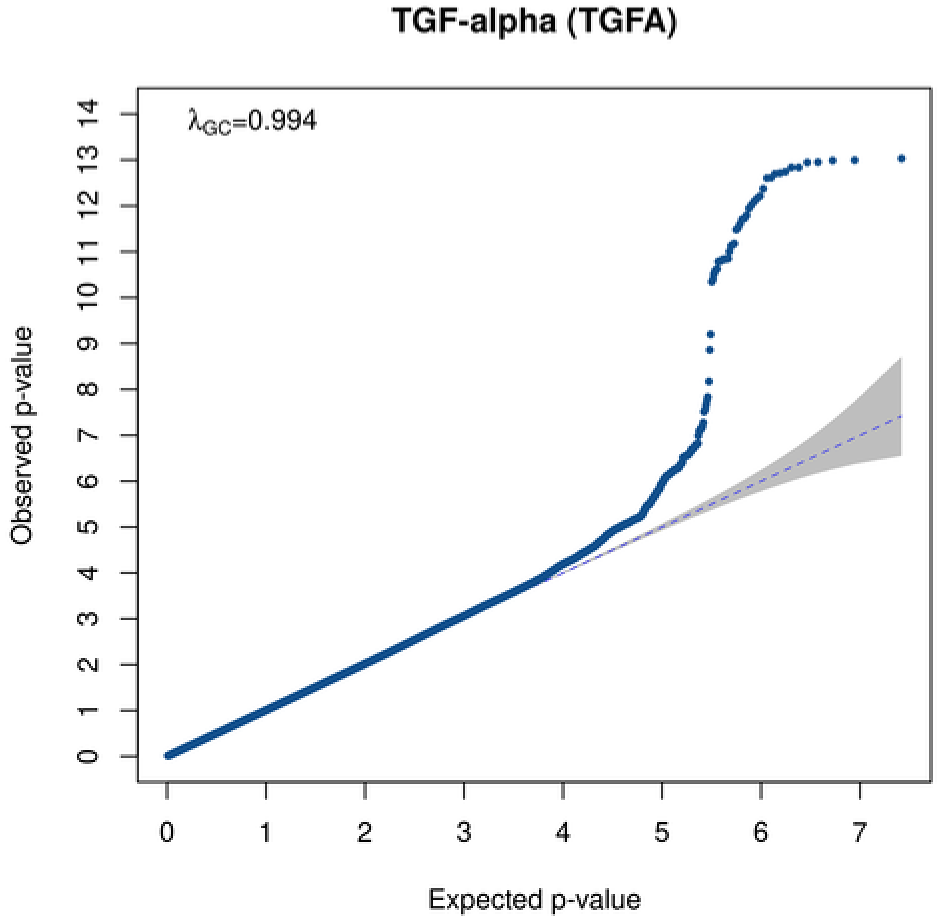
TGF-alpha (TGFA)
λGC=0.994
Observed p-value
0 1 2 3 4 5 6 7 8 9 10 11 12 13 14
0 1 2 3 4 5 6 7
Expected p-value

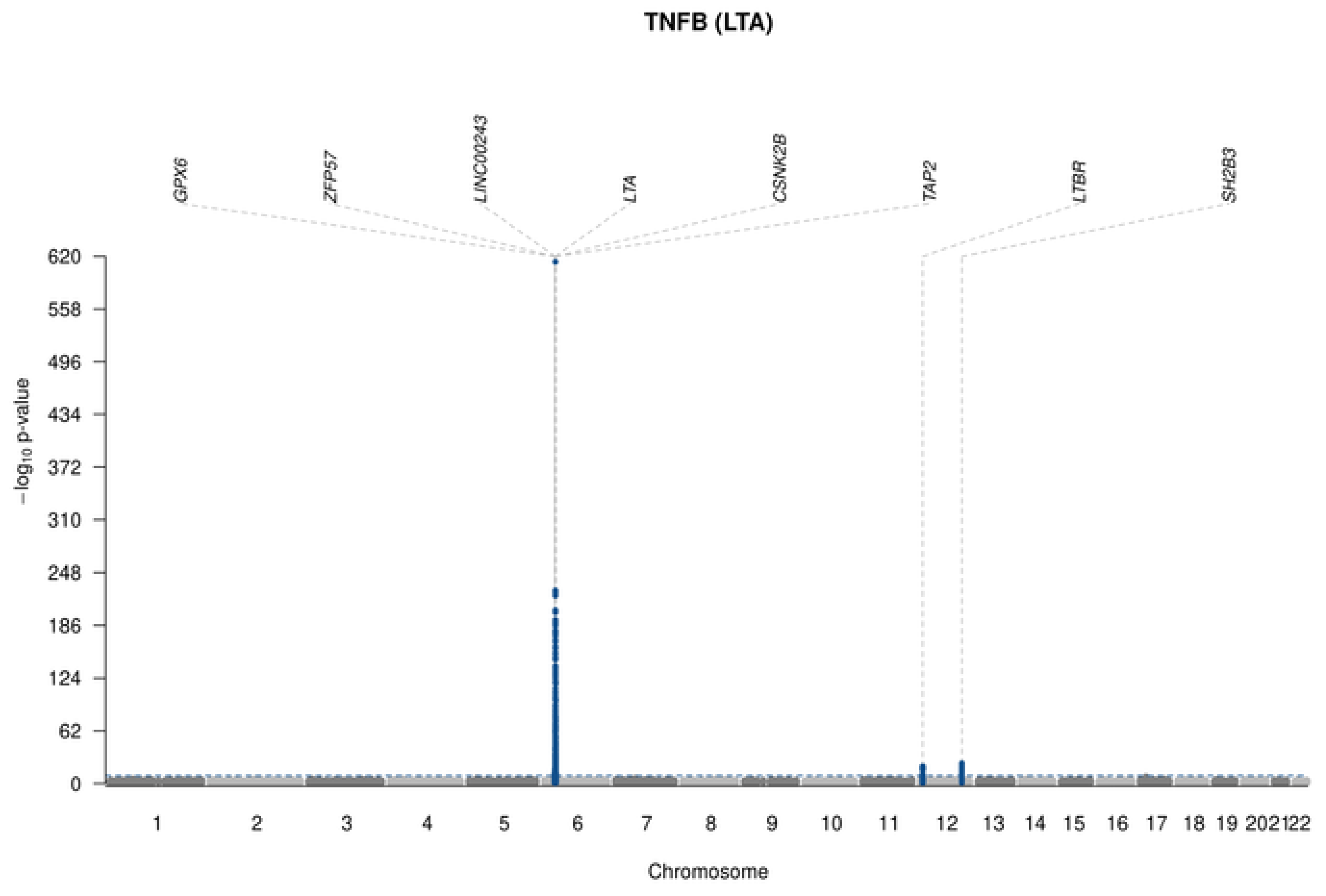

TNFB (LTA)
GPX6
ZFP57
LINC00243
LTA
CSNK2B
TAP2
LTBR
SH2B3
620
558
496
434
372
310
248
186
124
62
0
−log10 p-value
202122
Chromosome

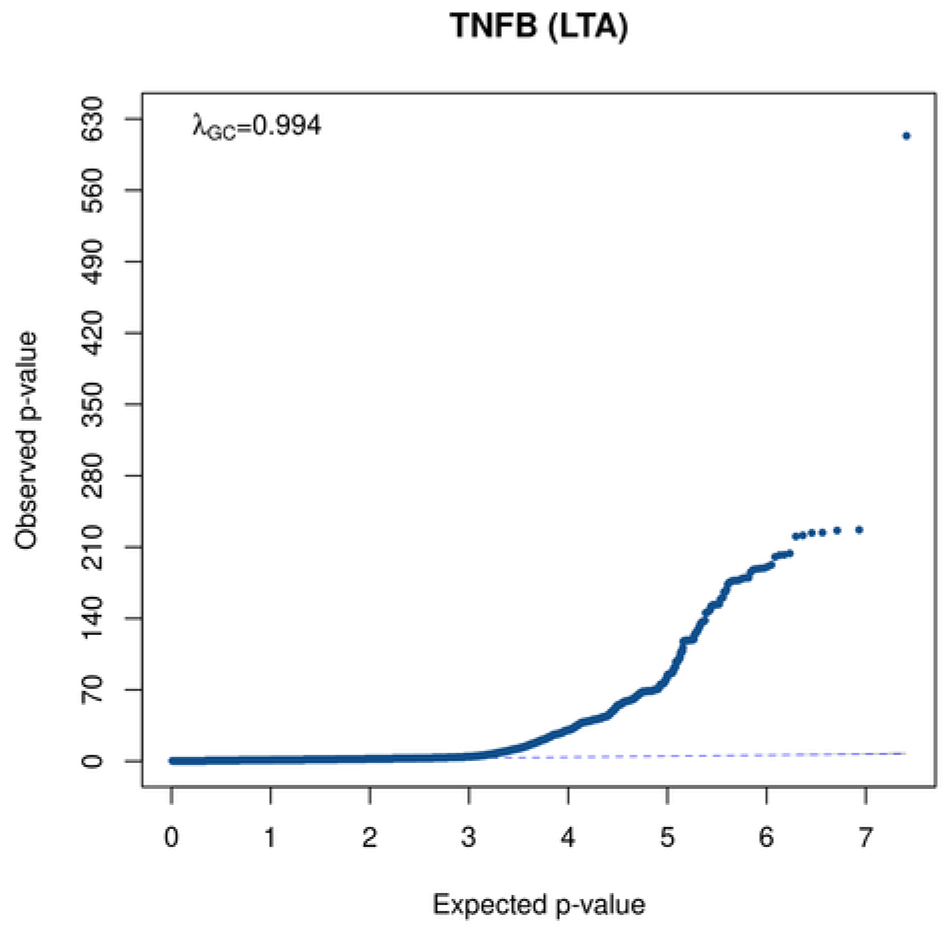

TNFB (LTA)
λGC=0.994
630
560
490
420
350
280
210
140
70
0
Observed p-value
0
1
2
3
4
5
6
7
Expected p-value

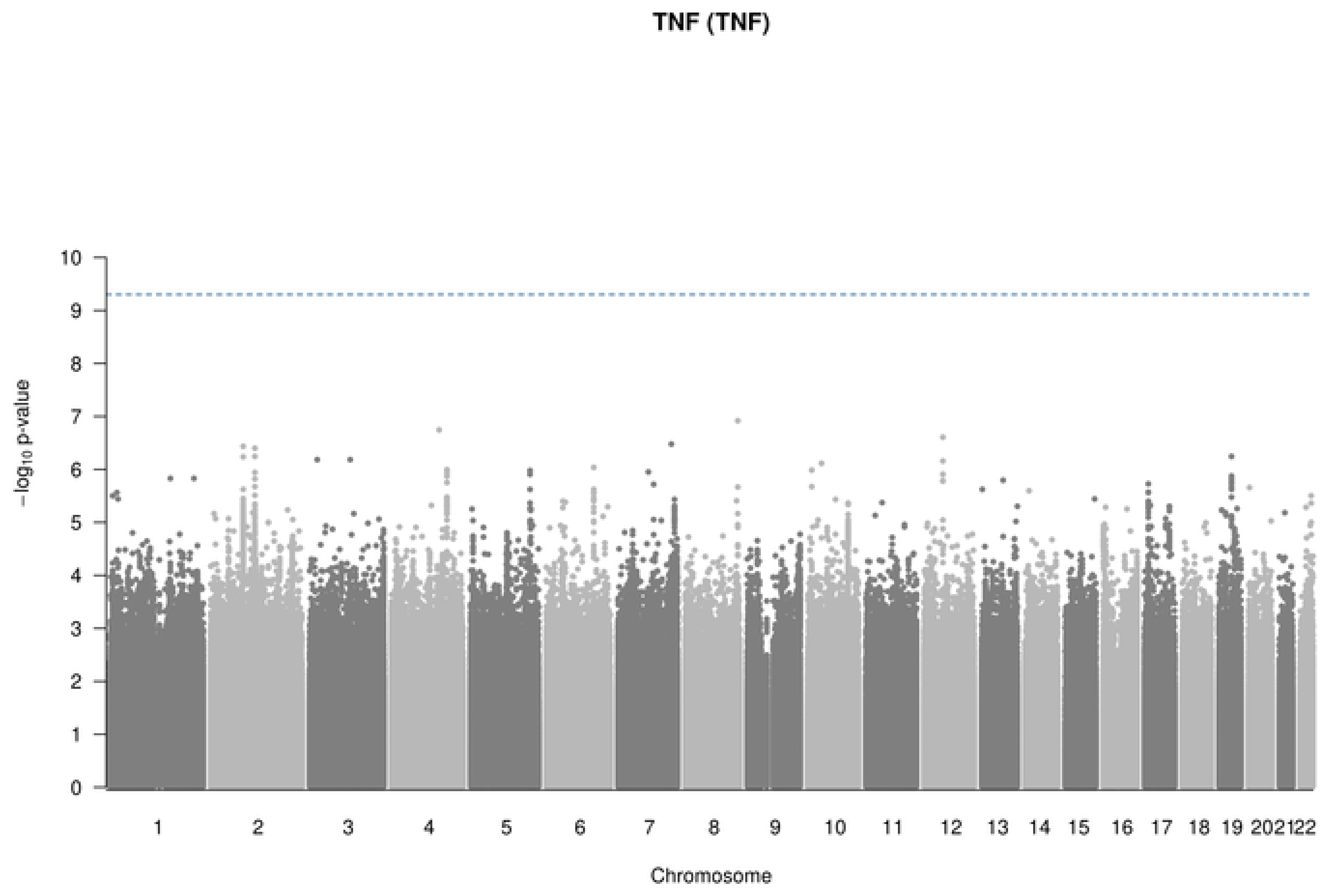

TNF (TNF)
−log10 p-value
10
9
8
7
6
5
4
3
2
1
202122
Chromosome

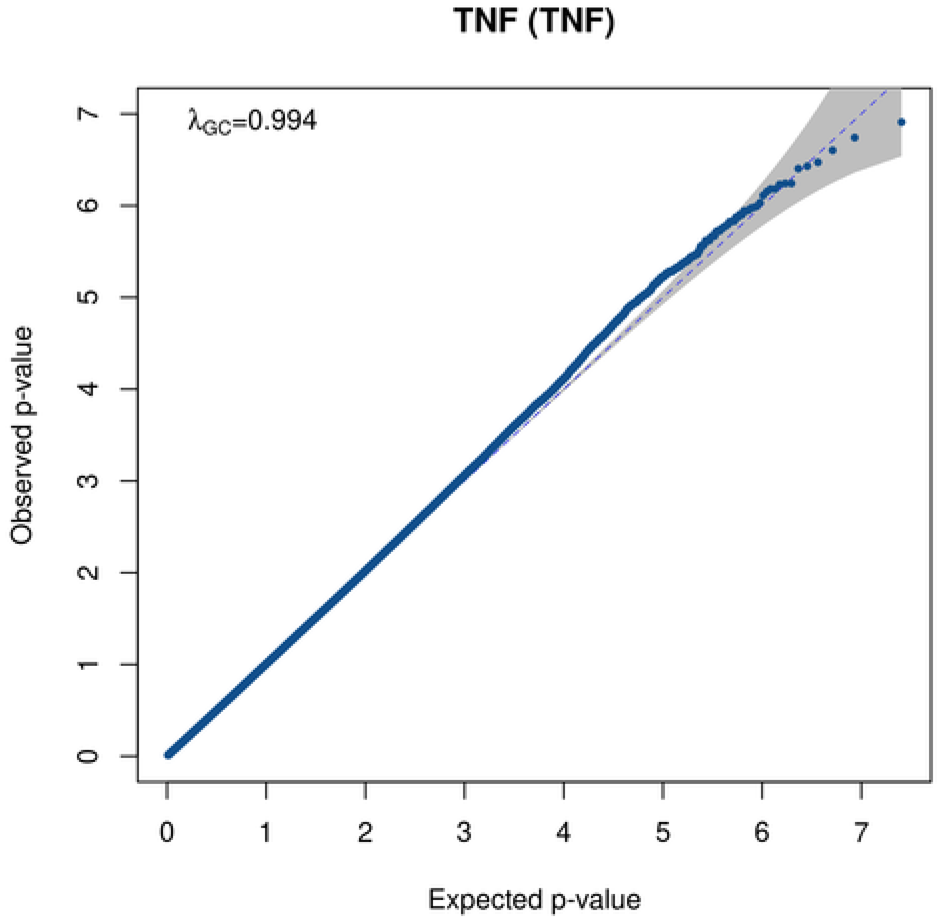

TNF (TNF)
λGC=0.994
Observed p-value
Expected p-value
0
1
2
3
4
5
6
7

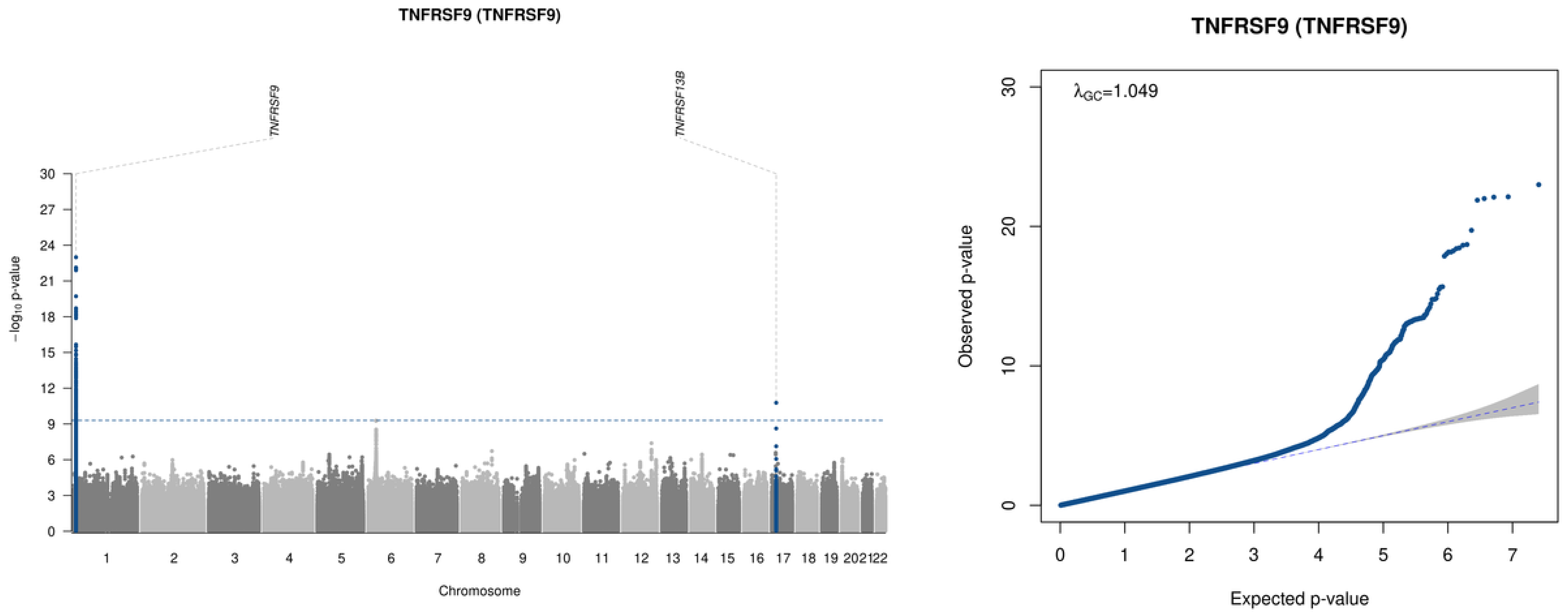

TNFRSF9 (TNFRSF9)
TNFRSF9
TNFRSF13B
−log10 p-value
Chromosome
TNFRSF9 (TNFRSF9)
λGC=1.049
Observed p-value
Expected p-value

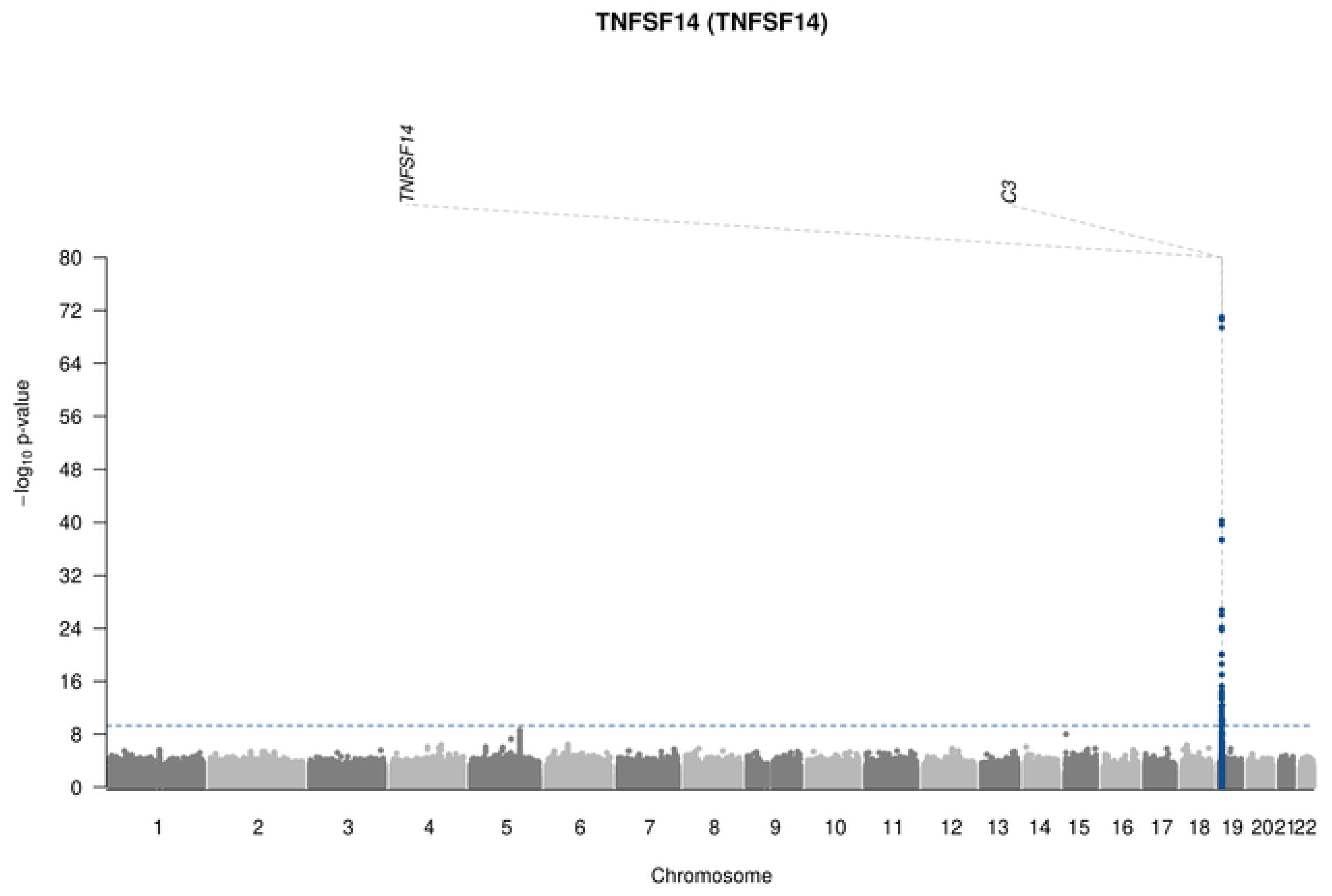

TNFSF14 (TNFSF14)
TNFSF14
C3
80
72
64
56
48
40
32
24
16
8
0
−log10 p-value
Chromosome

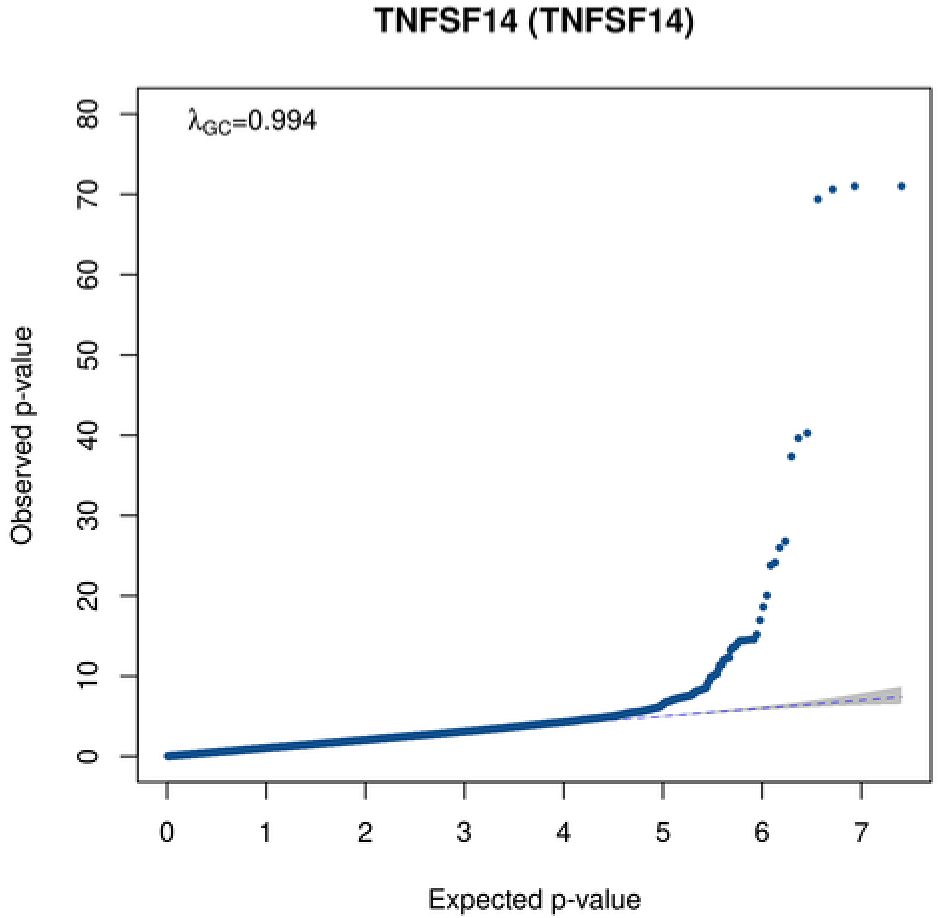

TNFSF14 (TNFSF14)
λGC=0.994
Observed p-value
Expected p-value
0
10
20
30
40
50
60
70
80
0
1
2
3
4
5
6
7

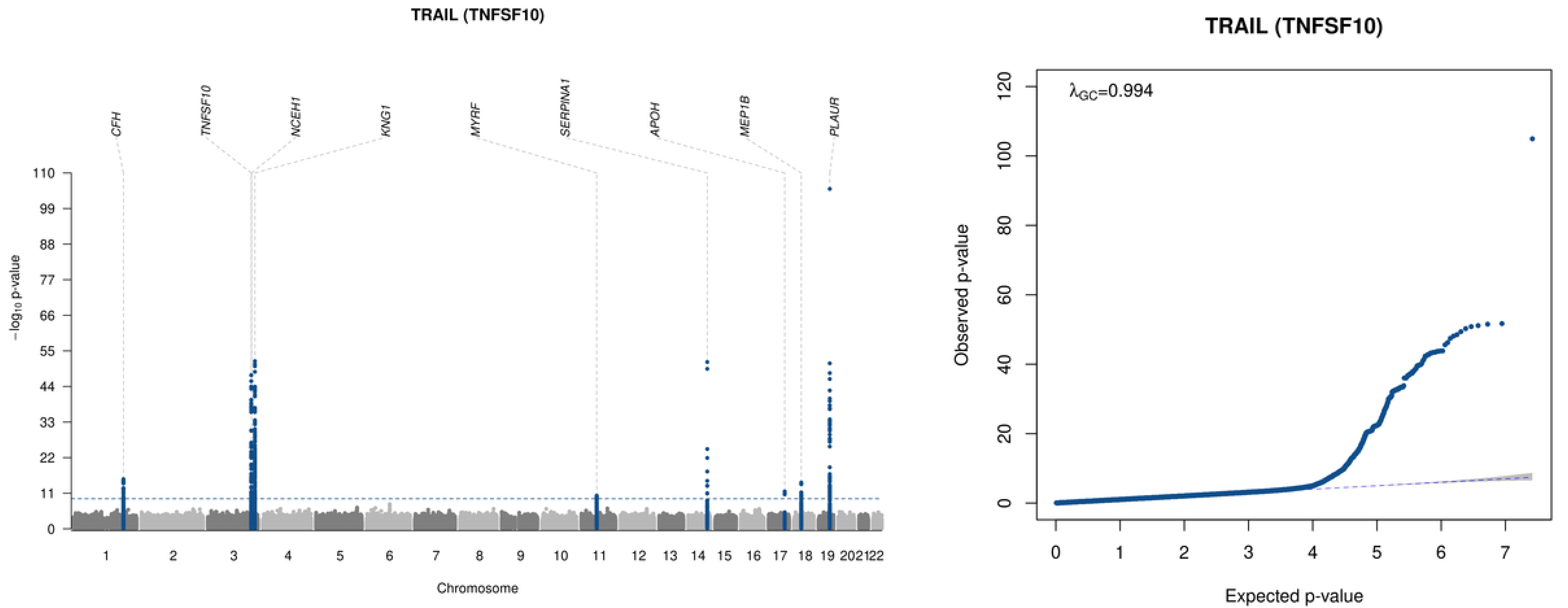
TRAIL (TNFSF10)
CFH
TNFSF10
NCEH1
KNG1
MYRF
SERPINA1
APOH
MEP1B
PLAUR
−log10 p-value
Chromosome
1 2 3 4 5 6 7 8 9 10 11 12 13 14 15 16 17 18 19 20 21 22
0 11 22 33 44 55 66 77 88 99 110
TRAIL (TNFSF10)
λGC=0.994
Observed p-value
Expected p-value
0 20 40 60 80 100 120
0 1 2 3 4 5 6 7

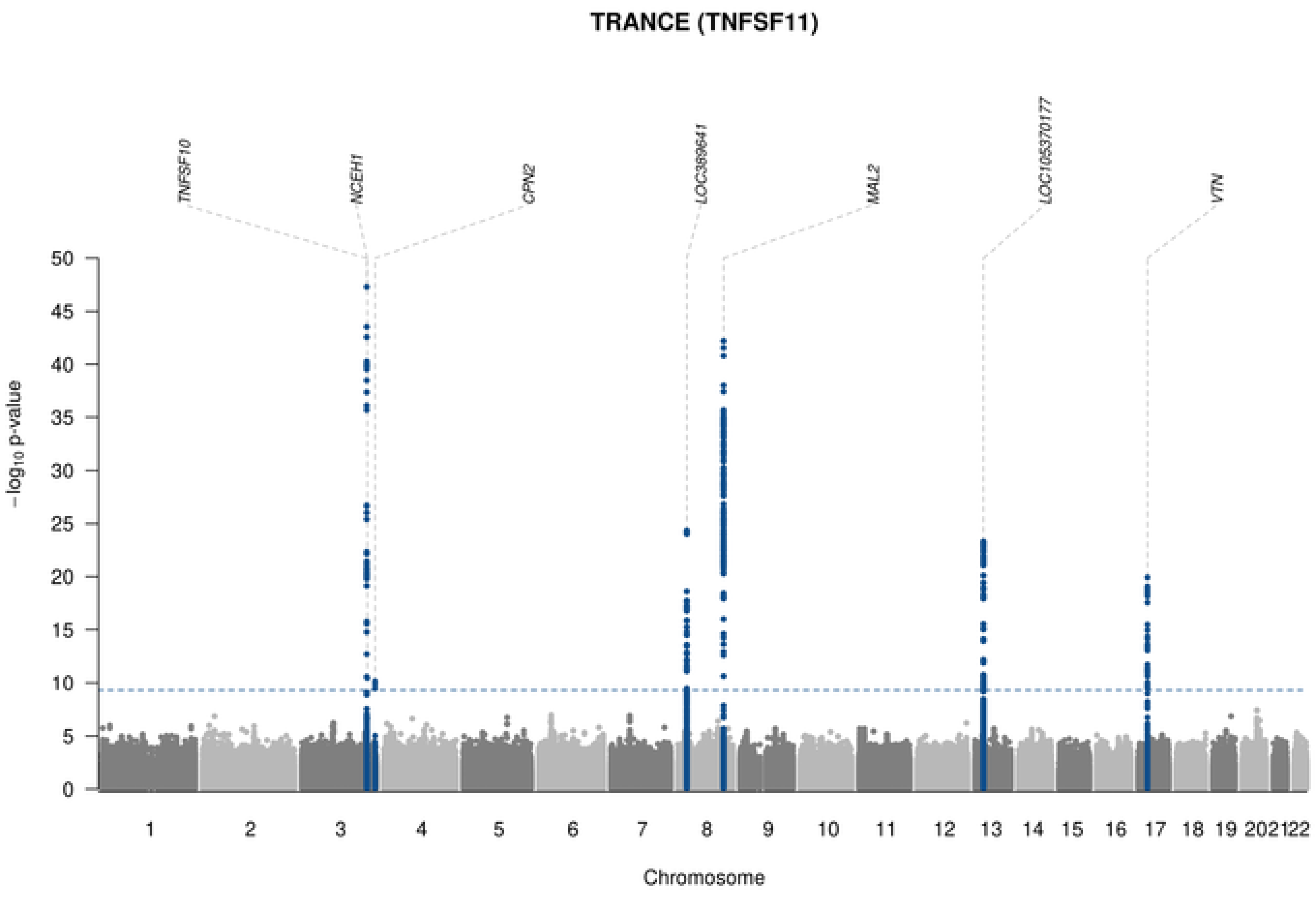

TRANCE (TNFSF11)
TNFSF10
NCEH1
CPN2
LOC389641
MAL2
LOC105370177
VTN
$-\log_{10}$ p-value
Chromosome

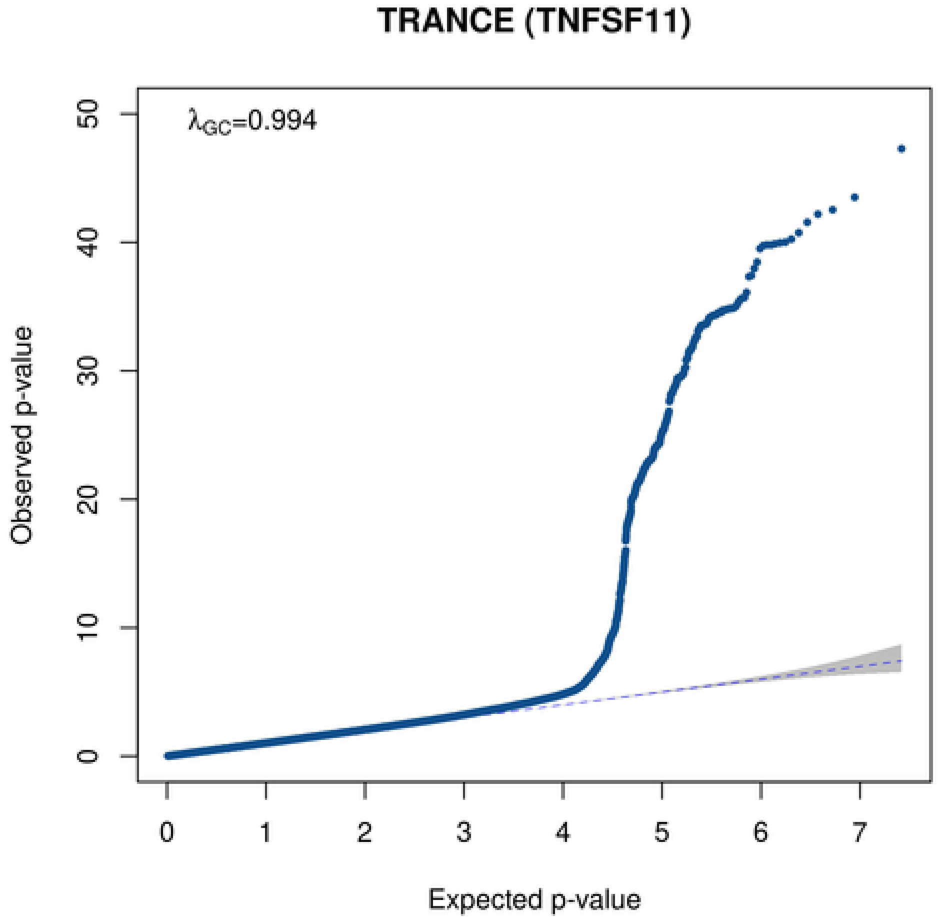

TRANCE (TNFSF11)
$\lambda_{GC}$=0.994
Observed p-value
Expected p-value

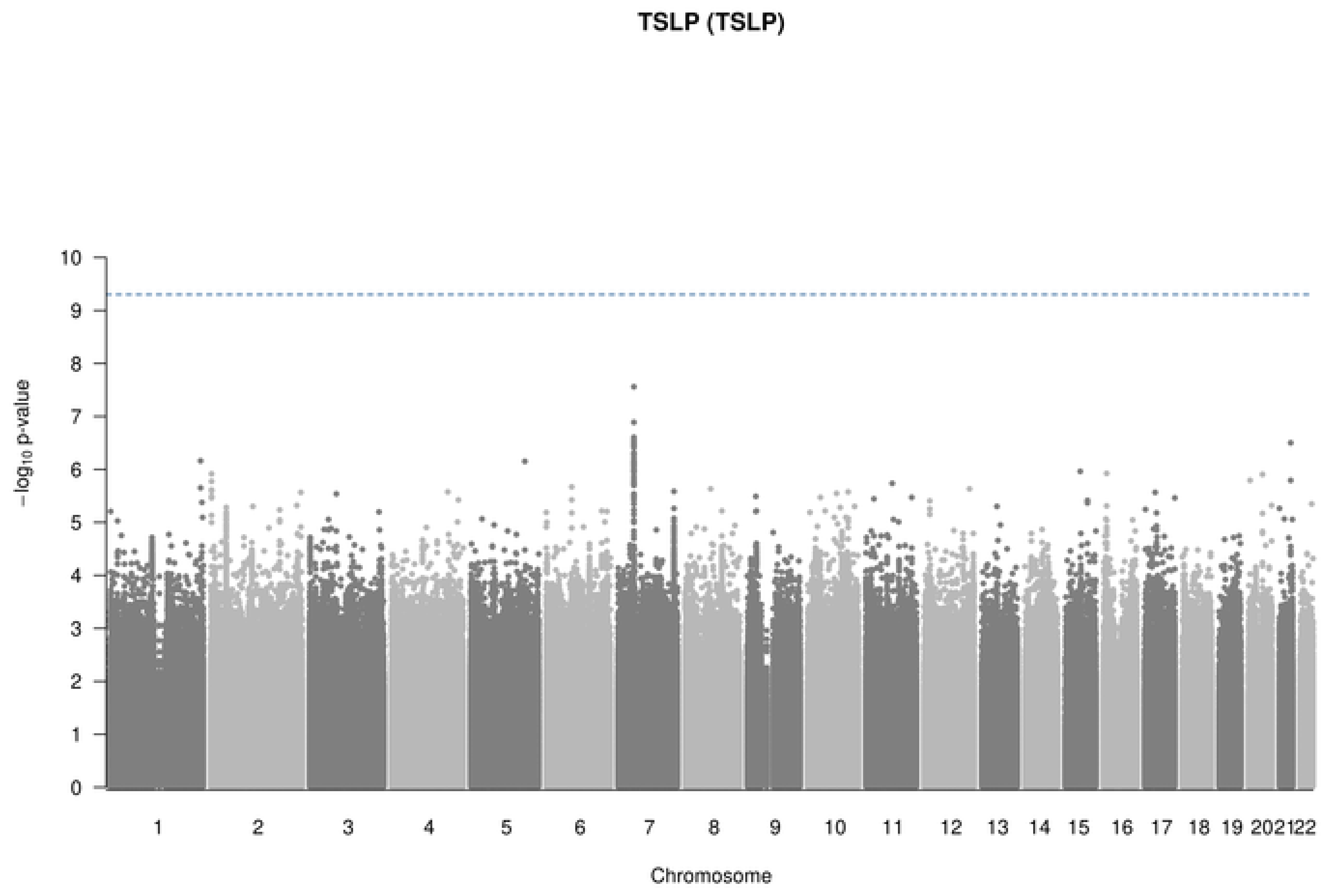
TSLP (TSLP)
−log10 p-value
0
1
2
3
4
5
6
7
8
9
10
202122
Chromosome

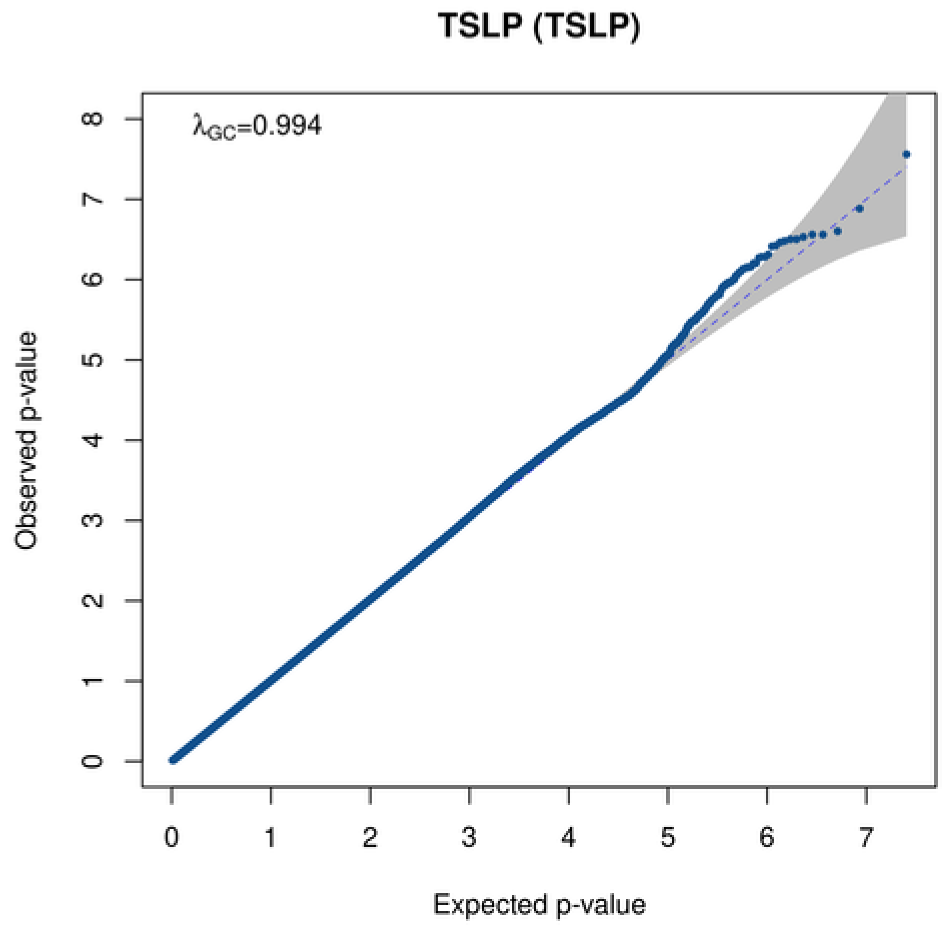
TSLP (TSLP)
λGC=0.994
Observed p-value
0
1
2
3
4
5
6
7
8
Expected p-value

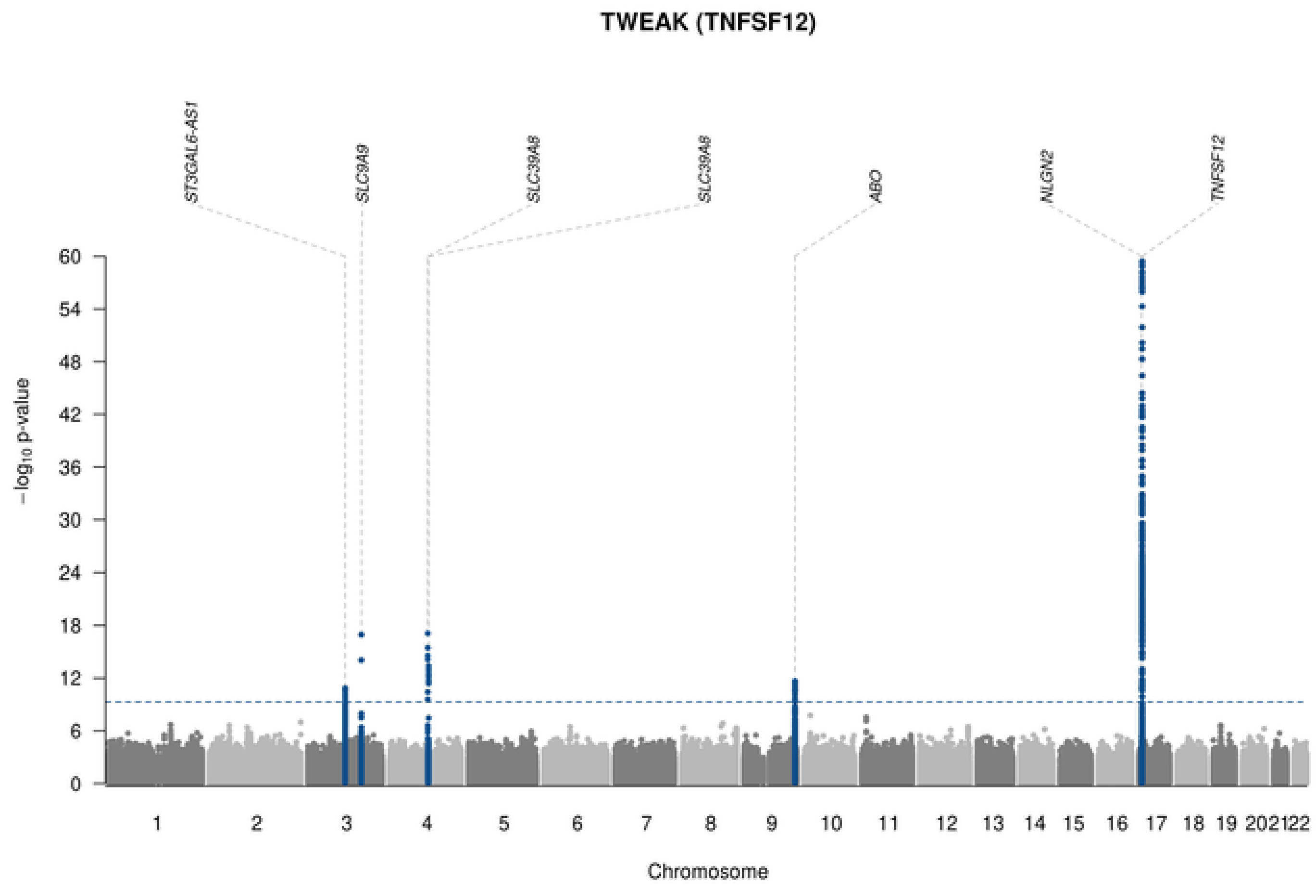
TWEAK (TNFSF12)
ST3GAL6-AS1
SLC9A9
SLC39A8
SLC39A8
ABO
NLGN2
TNFSF12
−log10 p-value
Chromosome

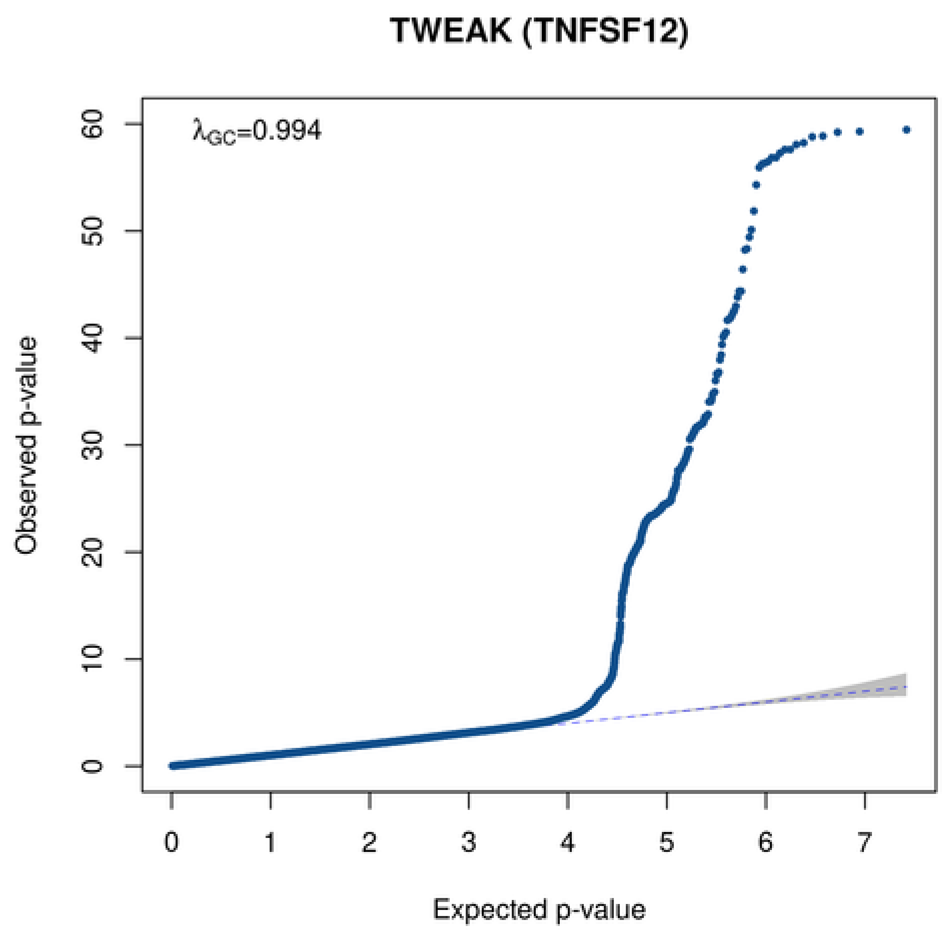
TWEAK (TNFSF12)
λGC=0.994
Observed p-value
Expected p-value

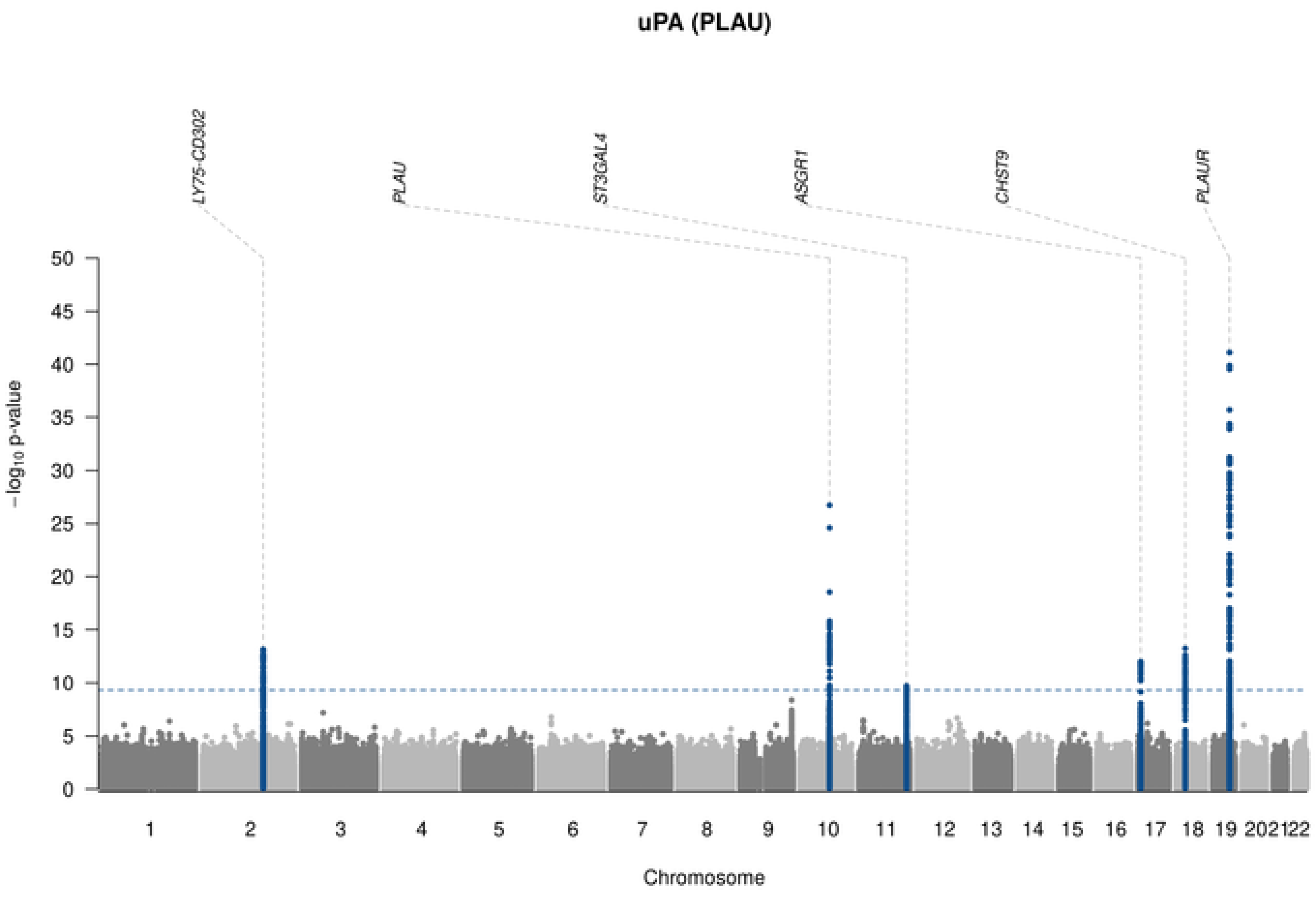

uPA (PLAU)
LY75-CD302
PLAU
ST3GAL4
ASGR1
CHST9
PLAUR
−log10 p-value
Chromosome
1 2 3 4 5 6 7 8 9 10 11 12 13 14 15 16 17 18 19 20 21 22

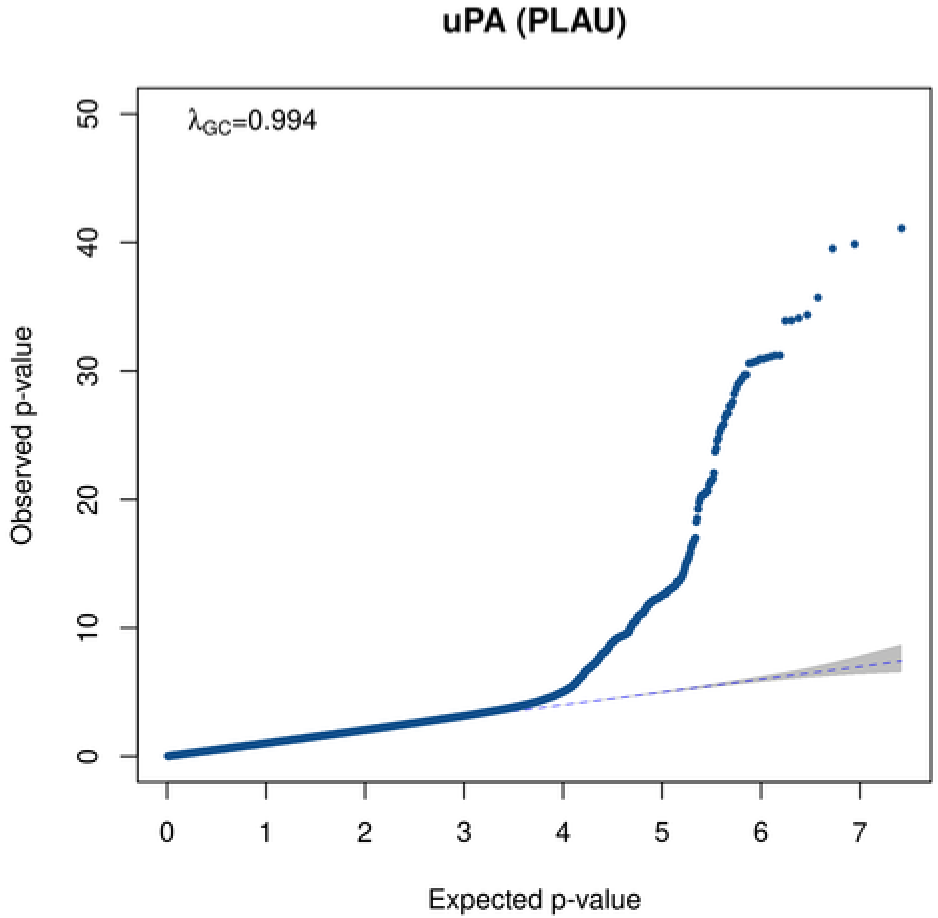

uPA (PLAU)
λGC=0.994
Observed p-value
Expected p-value

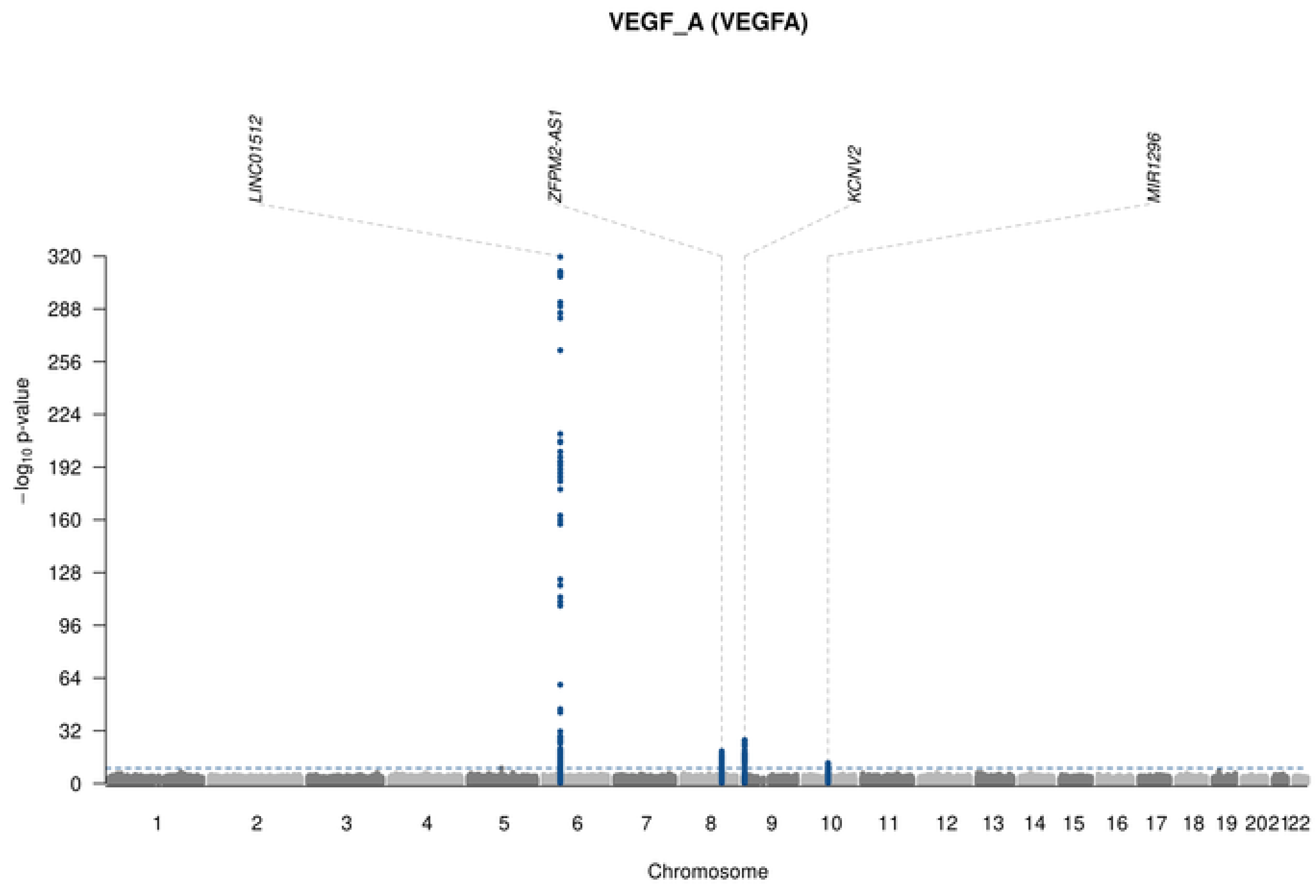
VEGF_A (VEGFA)
LINC01512
ZFPM2-AS1
KCNV2
MIR1296
320
288
256
224
192
160
128
96
64
32
0
$-\log_{10}$ p-value
1 2 3 4 5 6 7 8 9 10 11 12 13 14 15 16 17 18 19 202122
Chromosome

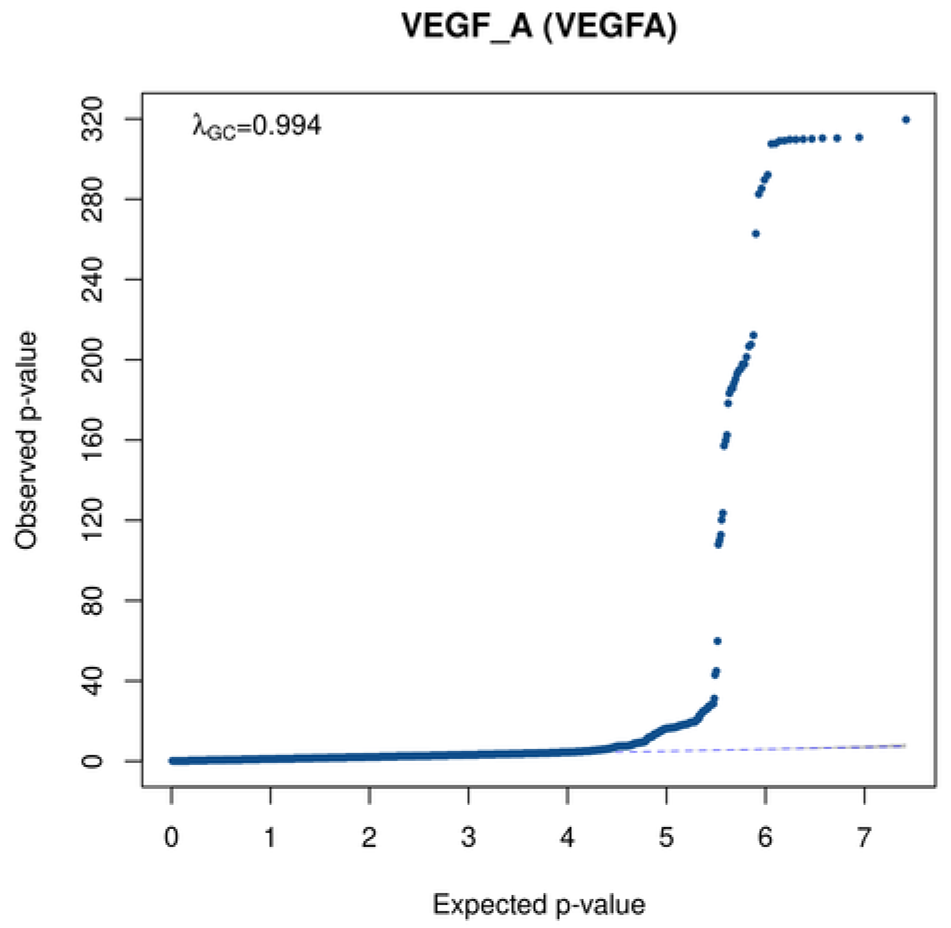
VEGF_A (VEGFA)
$\lambda_{GC}$=0.994
320
280
240
200
160
120
80
40
0
Observed p-value
0 1 2 3 4 5 6 7
Expected p-value
