## Supplementary Figure 14 for "Mapping pQTLs of circulating inflammatory proteins identifies drivers of immune-related disease risk and novel therapeutic targets"

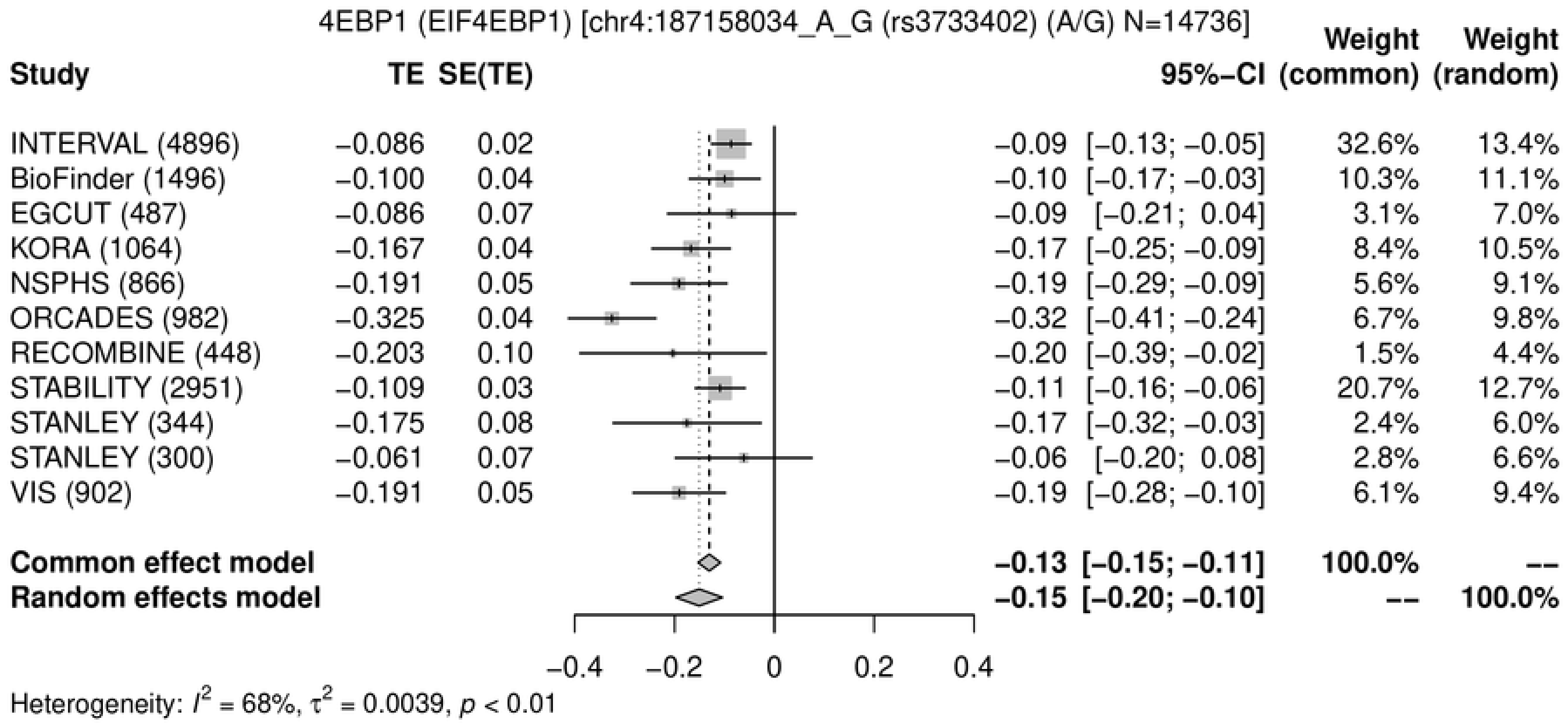
4EBP1 (EIF4EBP1) [chr4:187158034_A_G (rs3733402) (A/G) N=14736]
Study
TE
SE(TE)
95%−CI
Weight (common)
Weight (random)
INTERVAL (4896) −0.086 0.02 −0.09 [−0.13; −0.05] 32.6% 13.4%
BioFinder (1496) −0.100 0.04 −0.10 [−0.17; −0.03] 10.3% 11.1%
EGCUT (487) −0.086 0.07 −0.09 [−0.21; 0.04] 3.1% 7.0%
KORA (1064) −0.167 0.04 −0.17 [−0.25; −0.09] 8.4% 10.5%
NSPHS (866) −0.191 0.05 −0.19 [−0.29; −0.09] 5.6% 9.1%
ORCADES (982) −0.325 0.04 −0.32 [−0.41; −0.24] 6.7% 9.8%
RECOMBINE (448) −0.203 0.10 −0.20 [−0.39; −0.02] 1.5% 4.4%
STABILITY (2951) −0.109 0.03 −0.11 [−0.16; −0.06] 20.7% 12.7%
STANLEY (344) −0.175 0.08 −0.17 [−0.32; −0.03] 2.4% 6.0%
STANLEY (300) −0.061 0.07 −0.06 [−0.20; 0.08] 2.8% 6.6%
VIS (902) −0.191 0.05 −0.19 [−0.28; −0.10] 6.1% 9.4%
Common effect model −0.13 [−0.15; −0.11] 100.0% --
Random effects model −0.15 [−0.20; −0.10] -- 100.0%
−0.4 −0.2 0 0.2 0.4
Heterogeneity: $I^2$ = 68%, $\tau^2$ = 0.0039, $p$ < 0.01

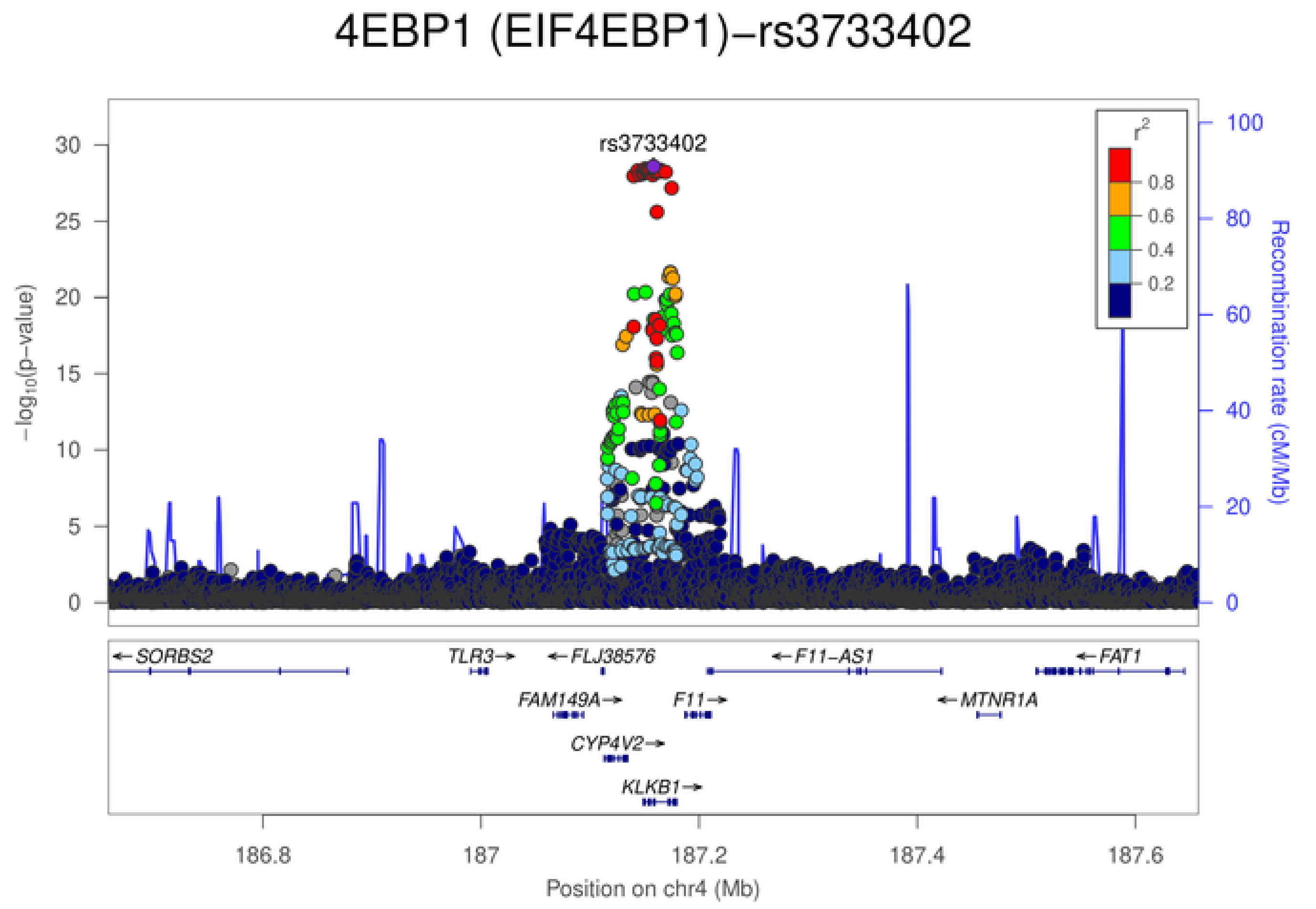
4EBP1 (EIF4EBP1)−rs3733402
rs3733402
$-\log_{10}$(p-value)
Recombination rate (cM/Mb)
$r^2$
0.8
0.6
0.4
0.2
SORBS2
TLR3
FLJ38576
F11-AS1
FAT1
FAM149A
F11
MTNR1A
CYP4V2
KLKB1
186.8 187 187.2 187.4 187.6
Position on chr4 (Mb)

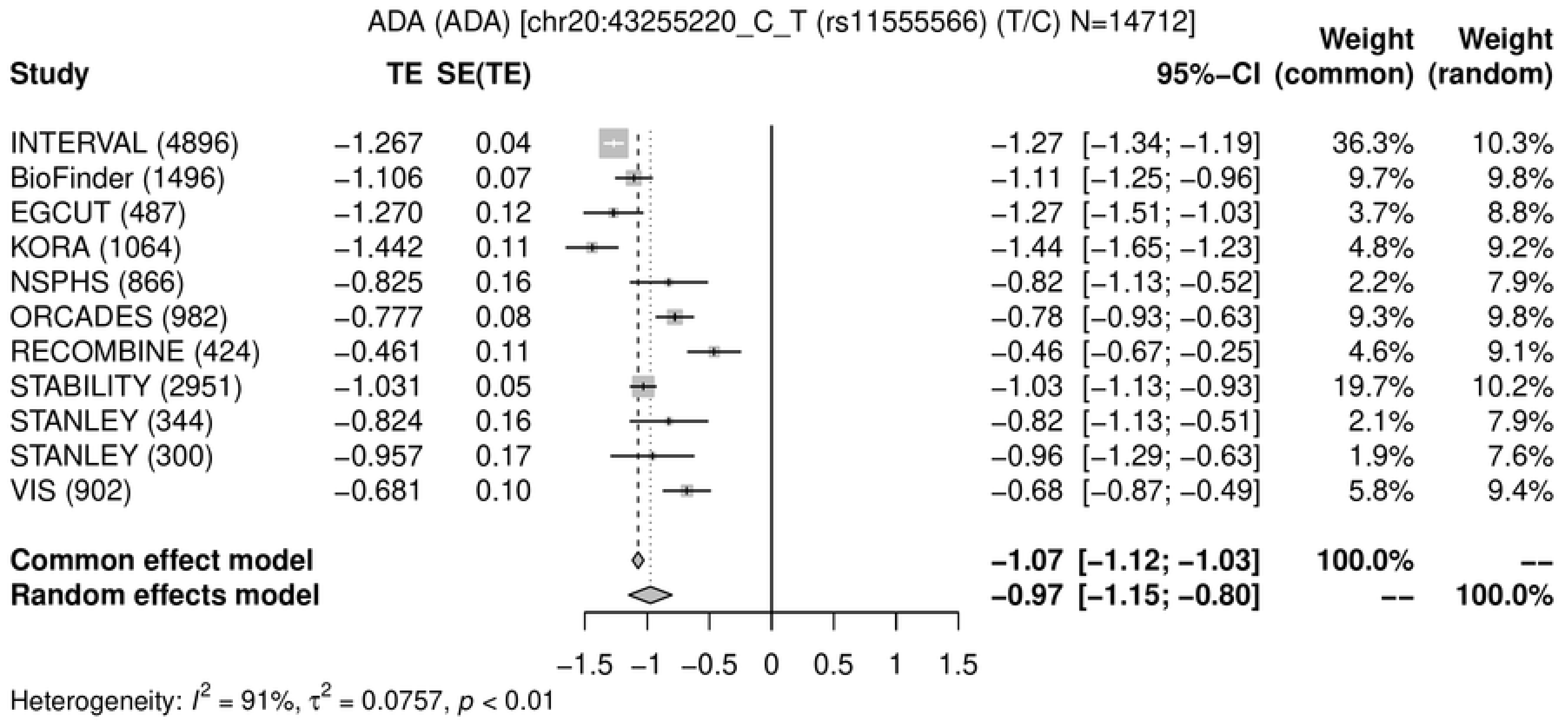
ADA (ADA) [chr20:43255220_C_T (rs11555566) (T/C) N=14712]
Study
TE
SE(TE)
95%−CI
Weight (common)
Weight (random)
INTERVAL (4896) −1.267 0.04 −1.27 [−1.34; −1.19] 36.3% 10.3%
BioFinder (1496) −1.106 0.07 −1.11 [−1.25; −0.96] 9.7% 9.8%
EGCUT (487) −1.270 0.12 −1.27 [−1.51; −1.03] 3.7% 8.8%
KORA (1064) −1.442 0.11 −1.44 [−1.65; −1.23] 4.8% 9.2%
NSPHS (866) −0.825 0.16 −0.82 [−1.13; −0.52] 2.2% 7.9%
ORCADES (982) −0.777 0.08 −0.78 [−0.93; −0.63] 9.3% 9.8%
RECOMBINE (424) −0.461 0.11 −0.46 [−0.67; −0.25] 4.6% 9.1%
STABILITY (2951) −1.031 0.05 −1.03 [−1.13; −0.93] 19.7% 10.2%
STANLEY (344) −0.824 0.16 −0.82 [−1.13; −0.51] 2.1% 7.9%
STANLEY (300) −0.957 0.17 −0.96 [−1.29; −0.63] 1.9% 7.6%
VIS (902) −0.681 0.10 −0.68 [−0.87; −0.49] 5.8% 9.4%
Common effect model −1.07 [−1.12; −1.03] 100.0% --
Random effects model −0.97 [−1.15; −0.80] -- 100.0%
−1.5 −1 −0.5 0 0.5 1 1.5
Heterogeneity: $I^2$ = 91%, $\tau^2$ = 0.0757, $p$ < 0.01

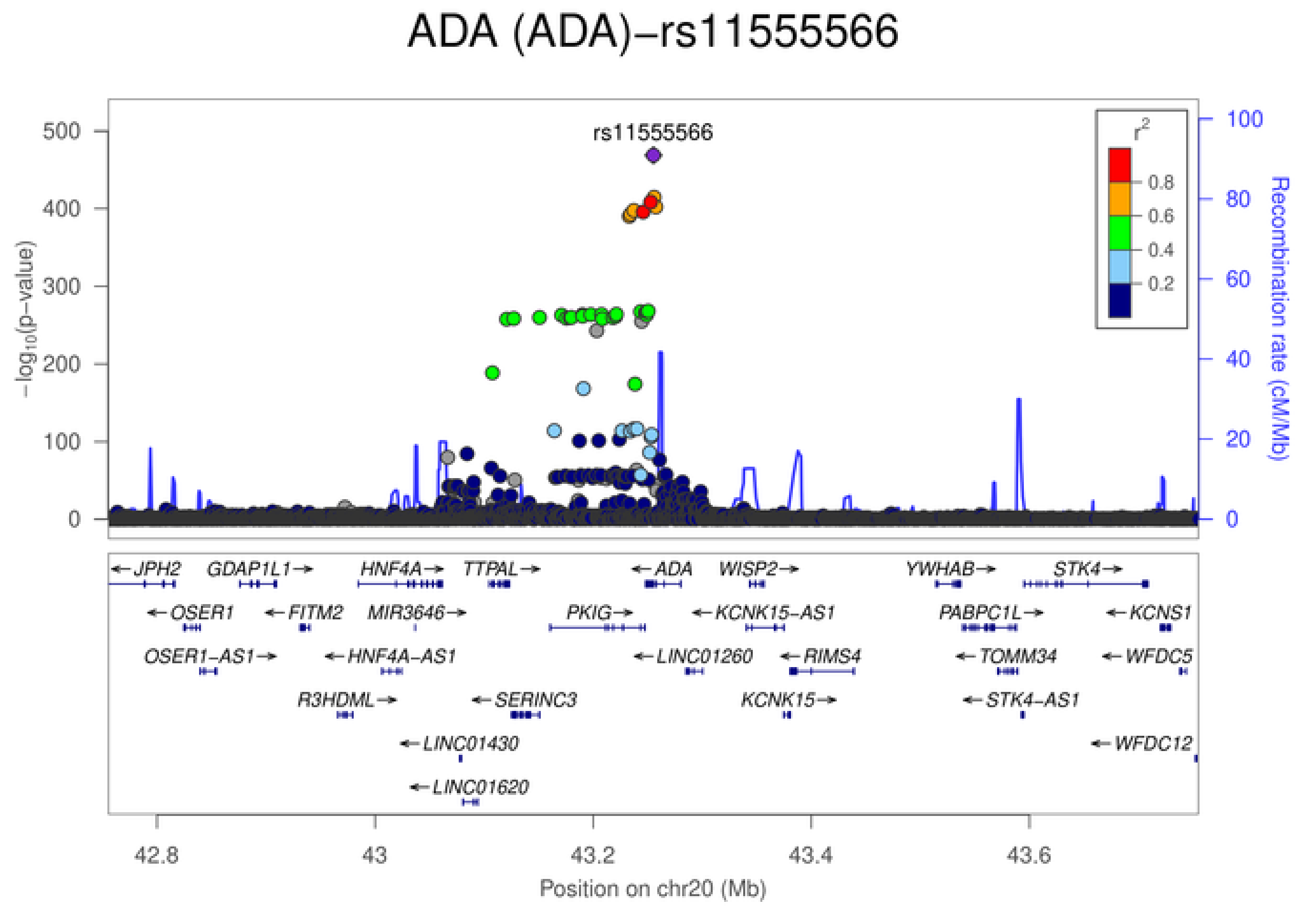
ADA (ADA)−rs11555566
rs11555566
$-\log_{10}$(p-value)
Recombination rate (cM/Mb)
$r^2$
0.8
0.6
0.4
0.2
JPH2
GDAP1L1
HNF4A
TTPAL
ADA
WISP2
YWHAB
STK4
OSER1
FITM2
MIR3646
PKIG
KCNK15−AS1
PABPC1L
KCNS1
OSER1−AS1
HNF4A−AS1
LINC01260
RIMS4
TOMM34
WFDC5
R3HDML
SERINC3
KCNK15
STK4−AS1
LINC01430
WFDC12
LINC01620
42.8
43
43.2
43.4
43.6
Position on chr20 (Mb)

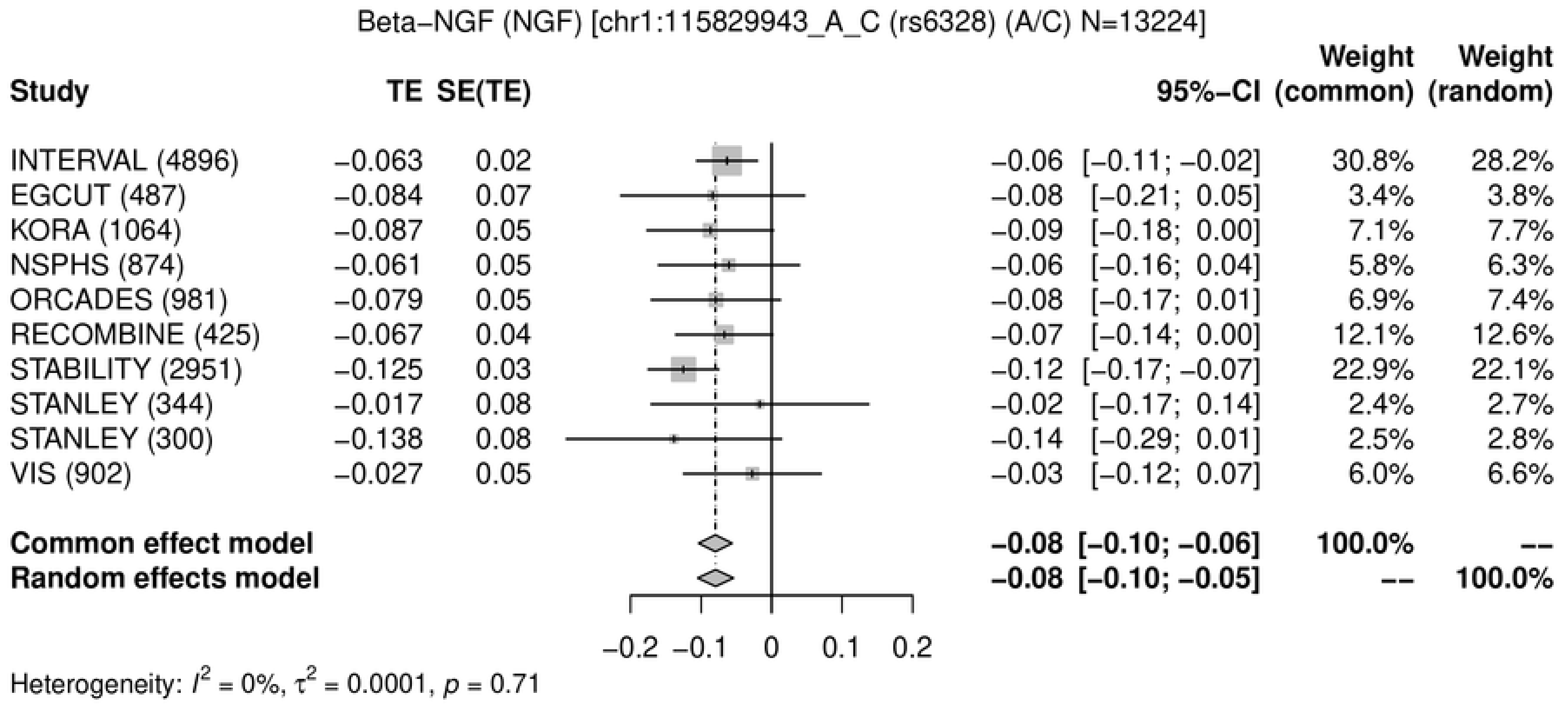
Beta−NGF (NGF) [chr1:115829943_A_C (rs6328) (A/C) N=13224]
Study
TE
SE(TE)
95%−CI
Weight (common)
Weight (random)
INTERVAL (4896) −0.063 0.02 −0.06 [−0.11; −0.02] 30.8% 28.2%
EGCUT (487) −0.084 0.07 −0.08 [−0.21; 0.05] 3.4% 3.8%
KORA (1064) −0.087 0.05 −0.09 [−0.18; 0.00] 7.1% 7.7%
NSPHS (874) −0.061 0.05 −0.06 [−0.16; 0.04] 5.8% 6.3%
ORCADES (981) −0.079 0.05 −0.08 [−0.17; 0.01] 6.9% 7.4%
RECOMBINE (425) −0.067 0.04 −0.07 [−0.14; 0.00] 12.1% 12.6%
STABILITY (2951) −0.125 0.03 −0.12 [−0.17; −0.07] 22.9% 22.1%
STANLEY (344) −0.017 0.08 −0.02 [−0.17; 0.14] 2.4% 2.7%
STANLEY (300) −0.138 0.08 −0.14 [−0.29; 0.01] 2.5% 2.8%
VIS (902) −0.027 0.05 −0.03 [−0.12; 0.07] 6.0% 6.6%
Common effect model −0.08 [−0.10; −0.06] 100.0% --
Random effects model −0.08 [−0.10; −0.05] -- 100.0%
−0.2 −0.1 0 0.1 0.2
Heterogeneity: $I^2$ = 0%, $\tau^2$ = 0.0001, $p$ = 0.71

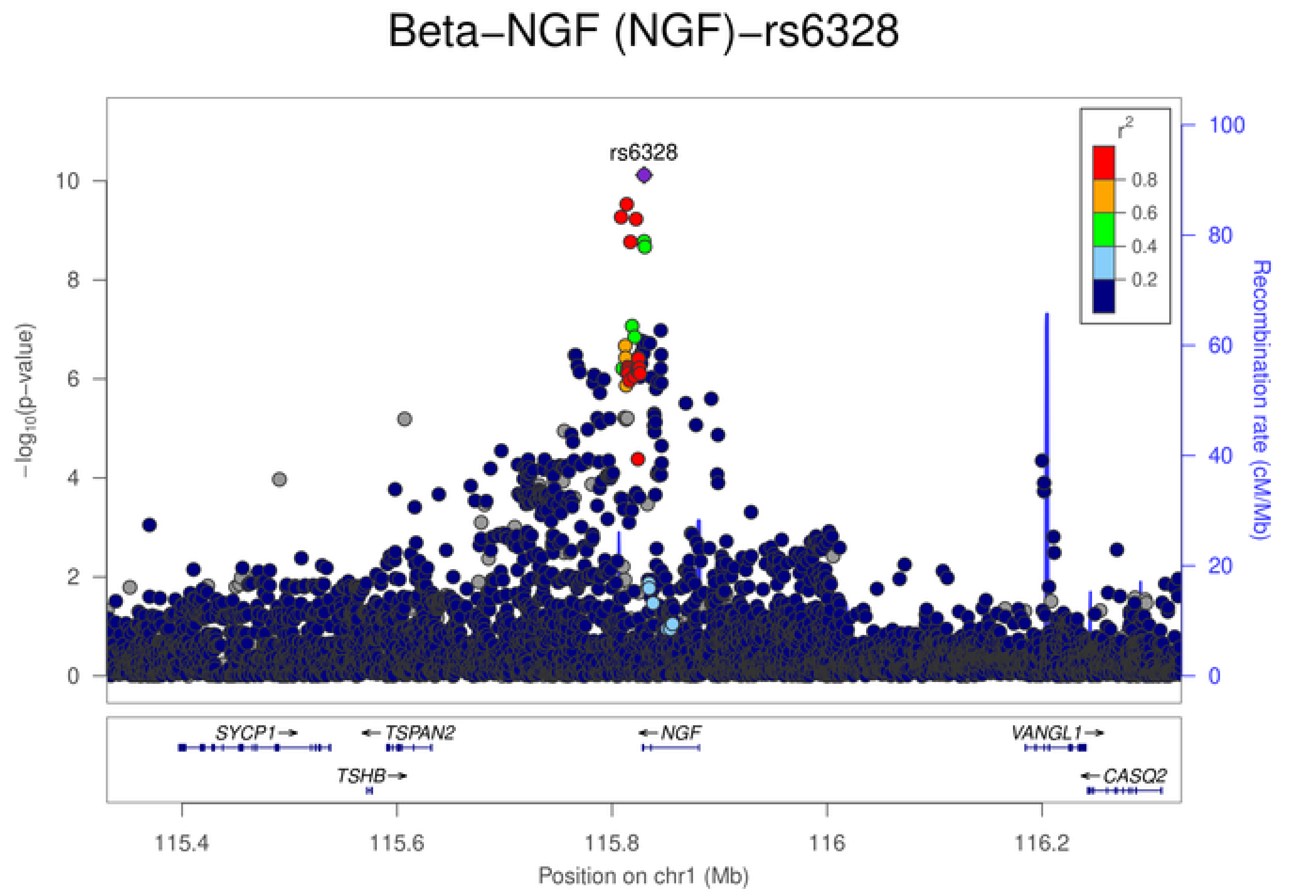
Beta−NGF (NGF)−rs6328
rs6328
$r^2$
0.8
0.6
0.4
0.2
$-\log_{10}$(p−value)
0
2
4
6
8
10
Recombination rate (cM/Mb)
0
20
40
60
80
100
SYCP1
TSPAN2
NGF
VANGL1
TSHB
CASQ2
115.4
115.6
115.8
116
116.2
Position on chr1 (Mb)

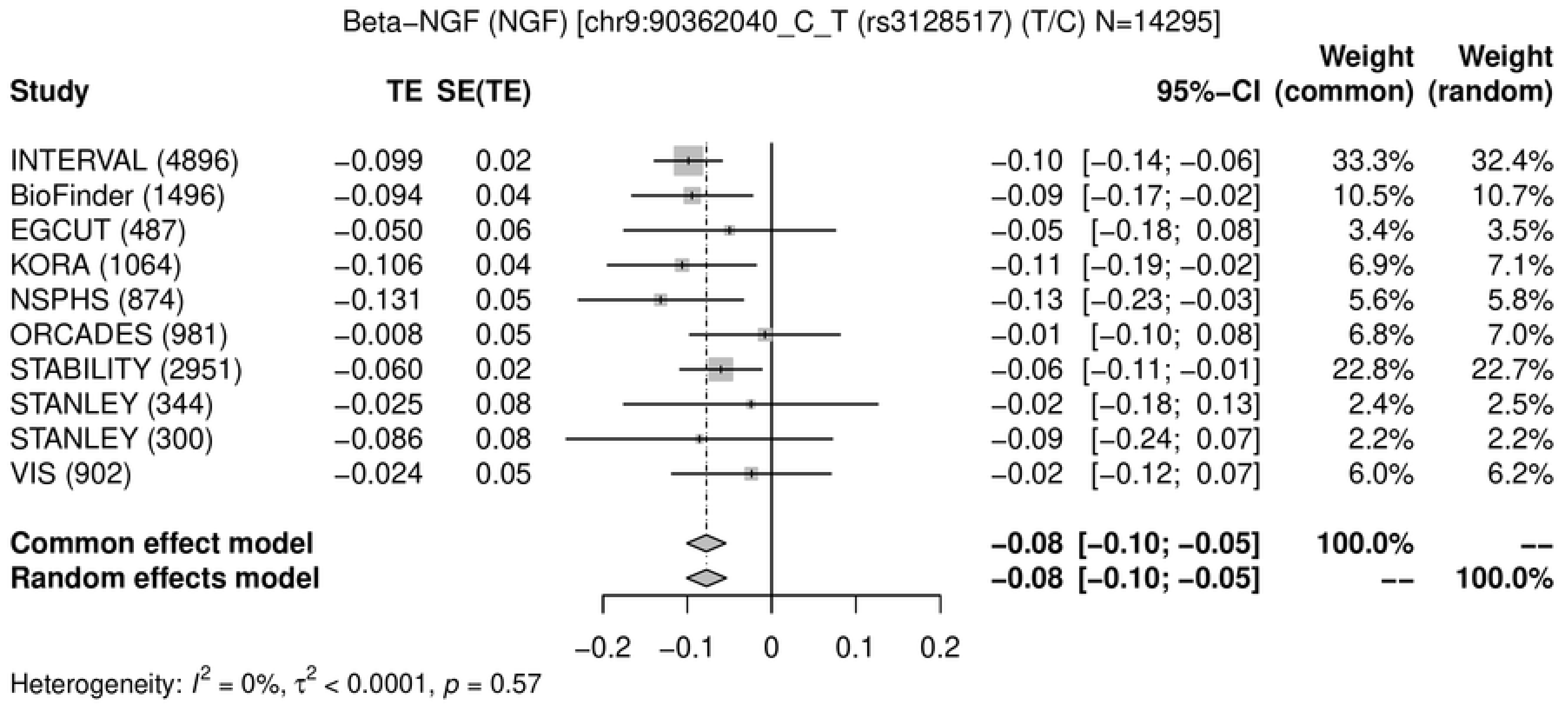
Beta–NGF (NGF) [chr9:90362040_C_T (rs3128517) (T/C) N=14295]
Study
TE
SE(TE)
95%–CI
Weight (common)
Weight (random)
INTERVAL (4896) –0.099 0.02 –0.10 [–0.14; –0.06] 33.3% 32.4%
BioFinder (1496) –0.094 0.04 –0.09 [–0.17; –0.02] 10.5% 10.7%
EGCUT (487) –0.050 0.06 –0.05 [–0.18; 0.08] 3.4% 3.5%
KORA (1064) –0.106 0.04 –0.11 [–0.19; –0.02] 6.9% 7.1%
NSPHS (874) –0.131 0.05 –0.13 [–0.23; –0.03] 5.6% 5.8%
ORCADES (981) –0.008 0.05 –0.01 [–0.10; 0.08] 6.8% 7.0%
STABILITY (2951) –0.060 0.02 –0.06 [–0.11; –0.01] 22.8% 22.7%
STANLEY (344) –0.025 0.08 –0.02 [–0.18; 0.13] 2.4% 2.5%
STANLEY (300) –0.086 0.08 –0.09 [–0.24; 0.07] 2.2% 2.2%
VIS (902) –0.024 0.05 –0.02 [–0.12; 0.07] 6.0% 6.2%
Common effect model –0.08 [–0.10; –0.05] 100.0% --
Random effects model –0.08 [–0.10; –0.05] -- 100.0%
–0.2
–0.1
0
0.1
0.2
Heterogeneity: $I^2$ = 0%, $\tau^2$ < 0.0001, $p$ = 0.57

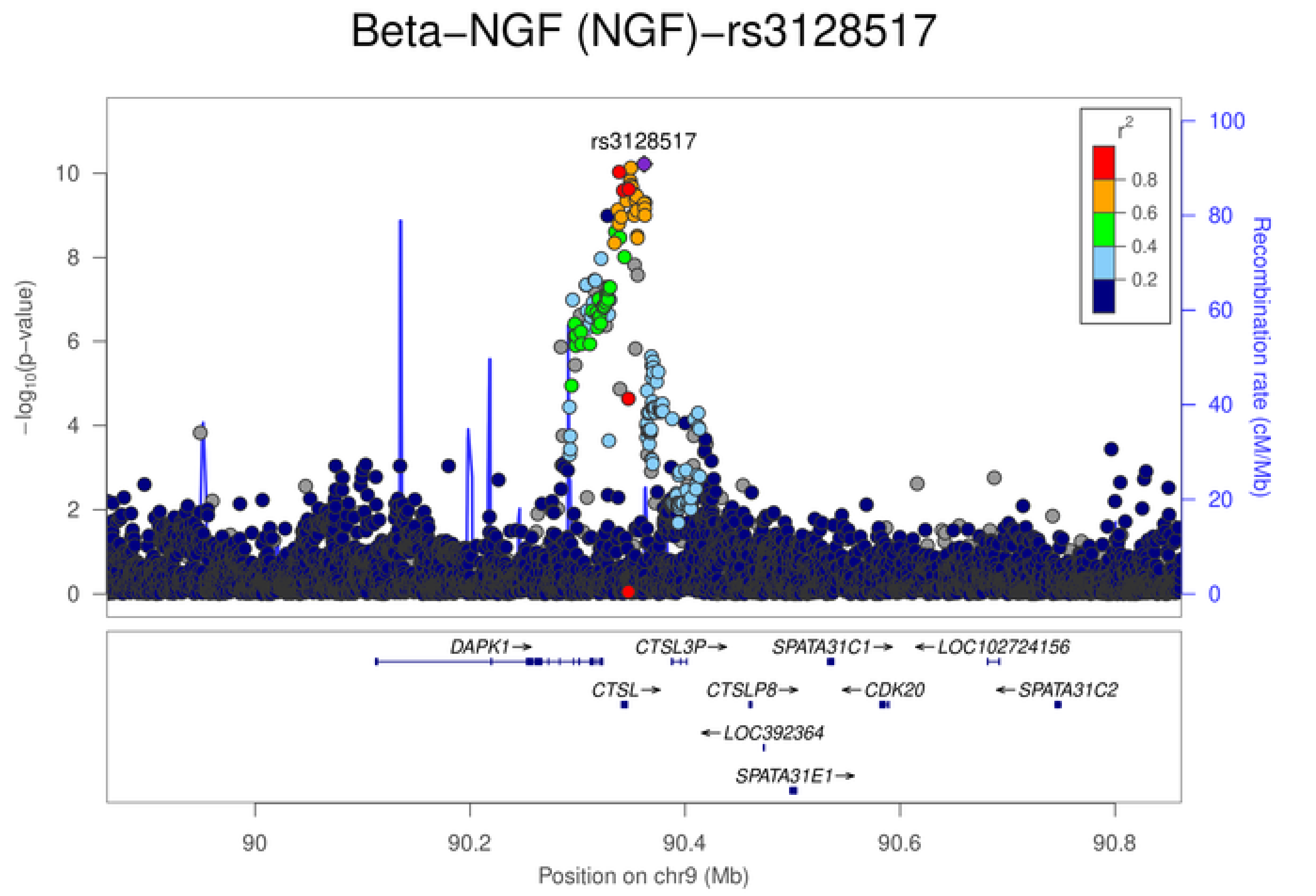
Beta–NGF (NGF)–rs3128517
rs3128517
$-\log_{10}$(p–value)
Recombination rate (cM/Mb)
$r^2$
0.8
0.6
0.4
0.2
DAPK1→
CTSL3P→
SPATA31C1→
←LOC102724156
CTSL→
CTSLP8→
←CDK20
←SPATA31C2
←LOC392364
SPATA31E1→
90
90.2
90.4
90.6
90.8
Position on chr9 (Mb)

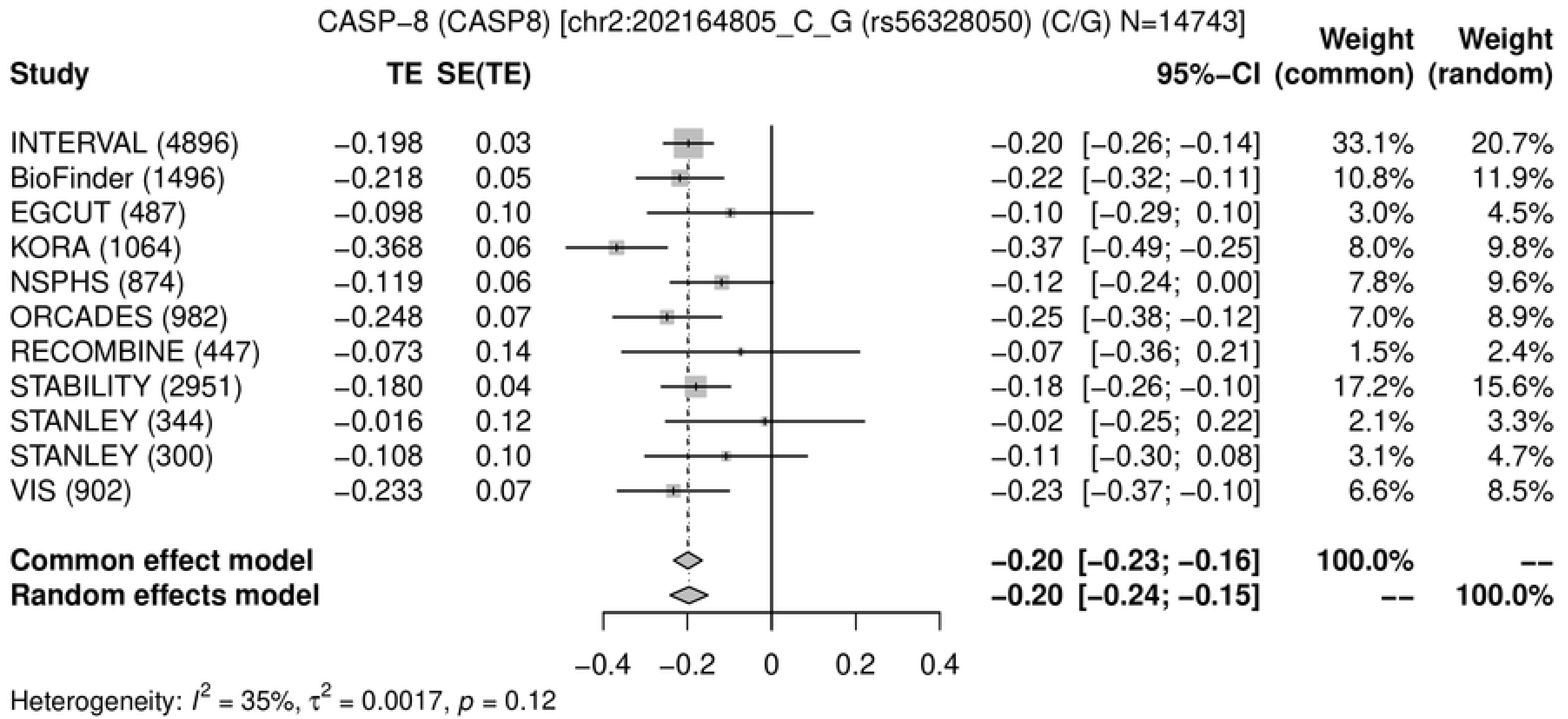
CASP-8 (CASP8) [chr2:202164805_C_G (rs56328050) (C/G) N=14743]
Study
TE
SE(TE)
95%-CI
Weight (common)
Weight (random)
INTERVAL (4896) −0.198 0.03 −0.20 [−0.26; −0.14] 33.1% 20.7%
BioFinder (1496) −0.218 0.05 −0.22 [−0.32; −0.11] 10.8% 11.9%
EGCUT (487) −0.098 0.10 −0.10 [−0.29; 0.10] 3.0% 4.5%
KORA (1064) −0.368 0.06 −0.37 [−0.49; −0.25] 8.0% 9.8%
NSPHS (874) −0.119 0.06 −0.12 [−0.24; 0.00] 7.8% 9.6%
ORCADES (982) −0.248 0.07 −0.25 [−0.38; −0.12] 7.0% 8.9%
RECOMBINE (447) −0.073 0.14 −0.07 [−0.36; 0.21] 1.5% 2.4%
STABILITY (2951) −0.180 0.04 −0.18 [−0.26; −0.10] 17.2% 15.6%
STANLEY (344) −0.016 0.12 −0.02 [−0.25; 0.22] 2.1% 3.3%
STANLEY (300) −0.108 0.10 −0.11 [−0.30; 0.08] 3.1% 4.7%
VIS (902) −0.233 0.07 −0.23 [−0.37; −0.10] 6.6% 8.5%
Common effect model −0.20 [−0.23; −0.16] 100.0% --
Random effects model −0.20 [−0.24; −0.15] -- 100.0%
−0.4 −0.2 0 0.2 0.4
Heterogeneity: $I^2$ = 35%, $\tau^2$ = 0.0017, $p$ = 0.12

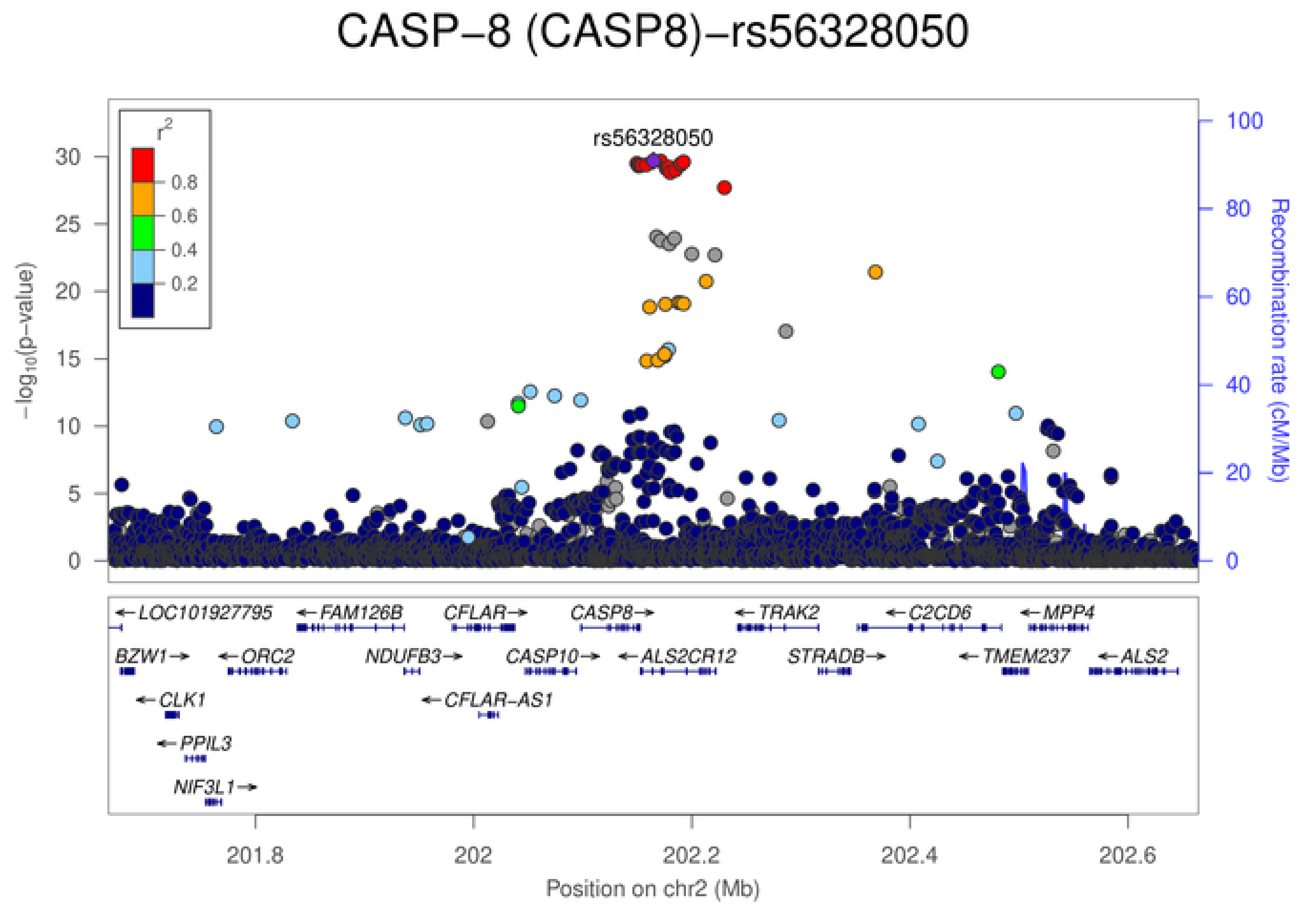
CASP−8 (CASP8)−rs56328050
rs56328050
$r^2$
0.8
0.6
0.4
0.2
$-\log_{10}$(p-value)
Recombination rate (cM/Mb)
LOC101927795
FAM126B
CFLAR
CASP8
TRAK2
C2CD6
MPP4
BZW1
ORC2
NDUFB3
CASP10
ALS2CR12
STRADB
TMEM237
ALS2
CLK1
CFLAR-AS1
PPIL3
NIF3L1
201.8
202
202.2
202.4
202.6
Position on chr2 (Mb)

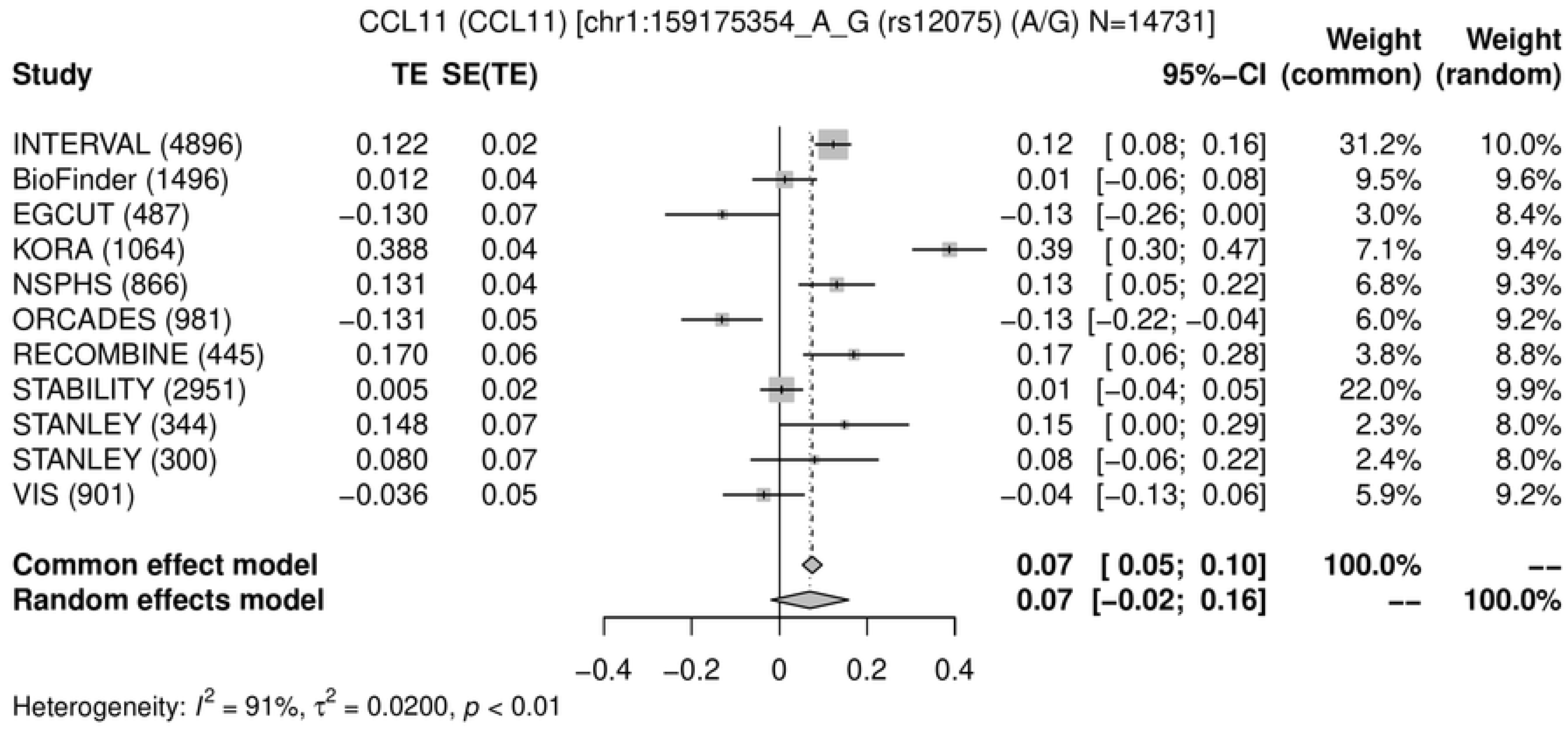
CCL11 (CCL11) [chr1:159175354_A_G (rs12075) (A/G) N=14731]
Study
TE
SE(TE)
95%−CI
Weight (common)
Weight (random)
INTERVAL (4896) 0.122 0.02 0.12 [ 0.08; 0.16] 31.2% 10.0%
BioFinder (1496) 0.012 0.04 0.01 [−0.06; 0.08] 9.5% 9.6%
EGCUT (487) −0.130 0.07 −0.13 [−0.26; 0.00] 3.0% 8.4%
KORA (1064) 0.388 0.04 0.39 [ 0.30; 0.47] 7.1% 9.4%
NSPHS (866) 0.131 0.04 0.13 [ 0.05; 0.22] 6.8% 9.3%
ORCADES (981) −0.131 0.05 −0.13 [−0.22; −0.04] 6.0% 9.2%
RECOMBINE (445) 0.170 0.06 0.17 [ 0.06; 0.28] 3.8% 8.8%
STABILITY (2951) 0.005 0.02 0.01 [−0.04; 0.05] 22.0% 9.9%
STANLEY (344) 0.148 0.07 0.15 [ 0.00; 0.29] 2.3% 8.0%
STANLEY (300) 0.080 0.07 0.08 [−0.06; 0.22] 2.4% 8.0%
VIS (901) −0.036 0.05 −0.04 [−0.13; 0.06] 5.9% 9.2%
Common effect model 0.07 [ 0.05; 0.10] 100.0% --
Random effects model 0.07 [−0.02; 0.16] -- 100.0%
−0.4 −0.2 0 0.2 0.4
Heterogeneity: $I^2$ = 91%, $\tau^2$ = 0.0200, $p$ < 0.01

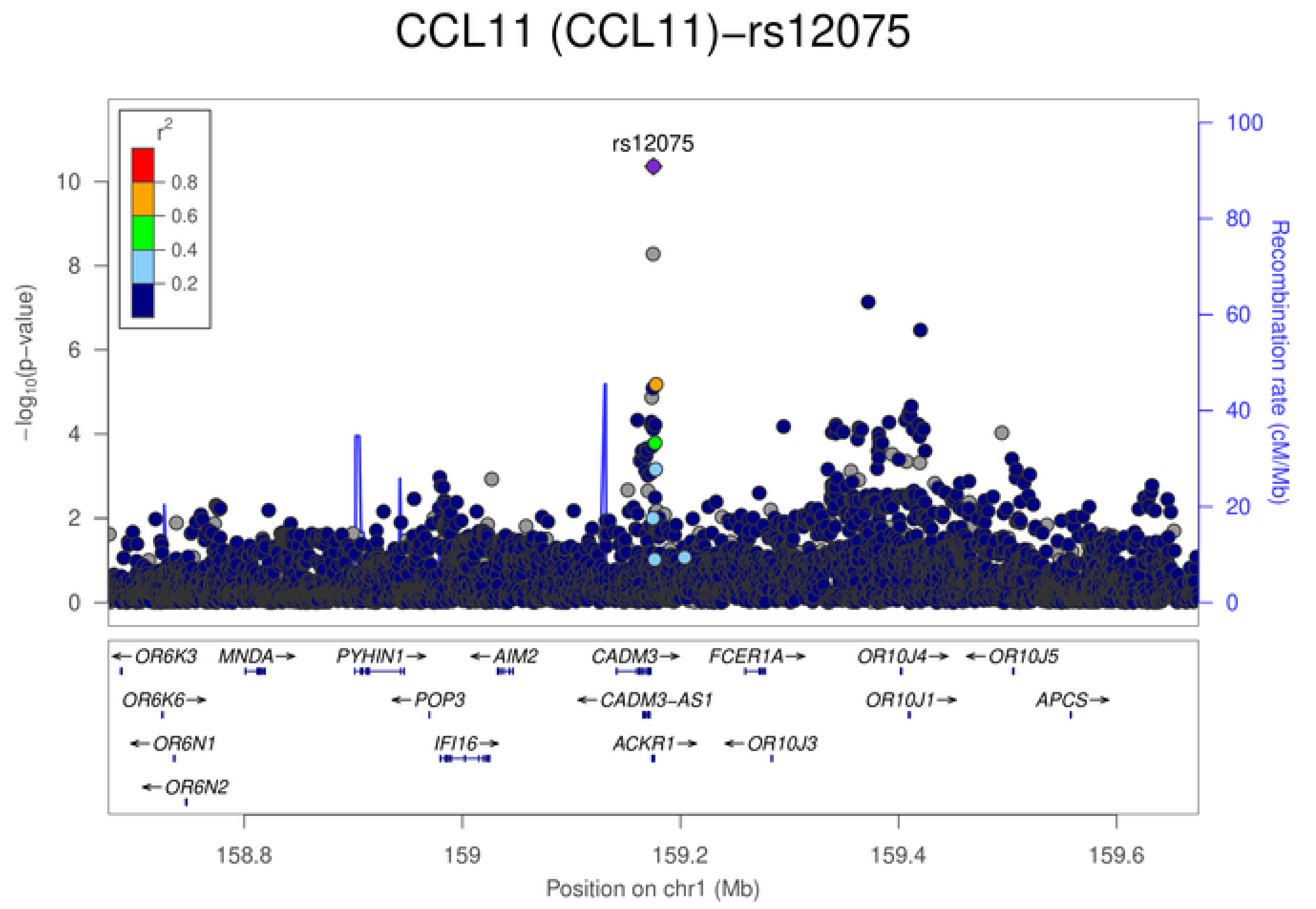
CCL11 (CCL11)−rs12075
rs12075
$r^2$
0.8
0.6
0.4
0.2
$-\log_{10}$(p−value)
Recombination rate (cM/Mb)
OR6K3
MNDA
PYHIN1
AIM2
CADM3
FCER1A
OR10J4
OR10J5
OR6K6
POP3
CADM3-AS1
OR10J1
APCS
OR6N1
IFI16
ACKR1
OR10J3
OR6N2
158.8
159
159.2
159.4
159.6
Position on chr1 (Mb)

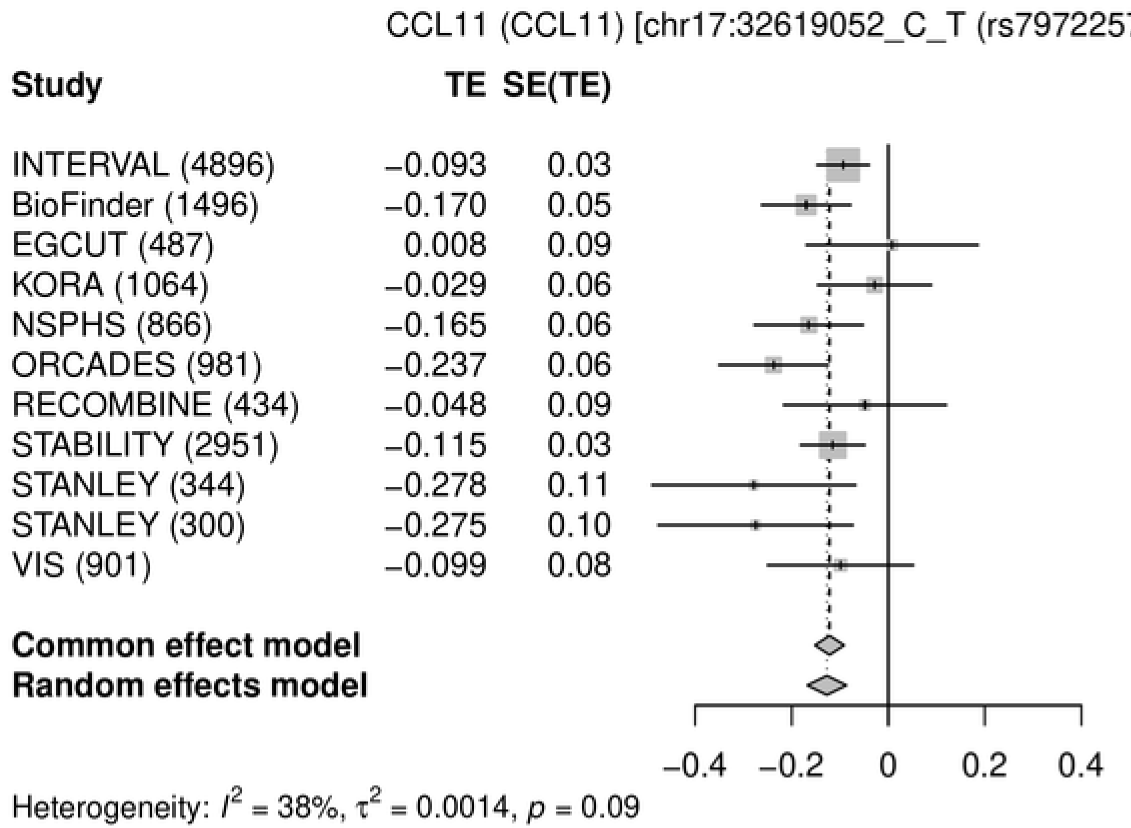
CCL11 (CCL11) [chr17:32619052_C_T (rs79722574) (T/C) N=14720]
Study | TE | SE(TE) | 95%-CI | Weight (common) | Weight (random)
INTERVAL (4896) | −0.093 | 0.03 | −0.09 [−0.15; −0.04] | 31.9% | 20.6%
BioFinder (1496) | −0.170 | 0.05 | −0.17 [−0.26; −0.08] | 10.9% | 12.2%
EGCUT (487) | 0.008 | 0.09 | 0.01 [−0.17; 0.19] | 3.0% | 4.5%
KORA (1064) | −0.029 | 0.06 | −0.03 [−0.15; 0.09] | 6.7% | 8.8%
NSPHS (866) | −0.165 | 0.06 | −0.16 [−0.28; −0.05] | 7.3% | 9.3%
ORCADES (981) | −0.237 | 0.06 | −0.24 [−0.35; −0.12] | 7.3% | 9.3%
RECOMBINE (434) | −0.048 | 0.09 | −0.05 [−0.22; 0.12] | 3.3% | 5.0%
STABILITY (2951) | −0.115 | 0.03 | −0.11 [−0.18; −0.05] | 21.1% | 17.4%
STANLEY (344) | −0.278 | 0.11 | −0.28 [−0.49; −0.07] | 2.1% | 3.4%
STANLEY (300) | −0.275 | 0.10 | −0.27 [−0.48; −0.07] | 2.3% | 3.6%
VIS (901) | −0.099 | 0.08 | −0.10 [−0.25; 0.05] | 4.1% | 6.0%
Common effect model | −0.12 [−0.15; −0.09] | 100.0% | --
Random effects model | −0.13 [−0.17; −0.09] | -- | 100.0%
−0.4 −0.2 0 0.2 0.4
Heterogeneity: $I^2$ = 38%, $\tau^2$ = 0.0014, $p$ = 0.09

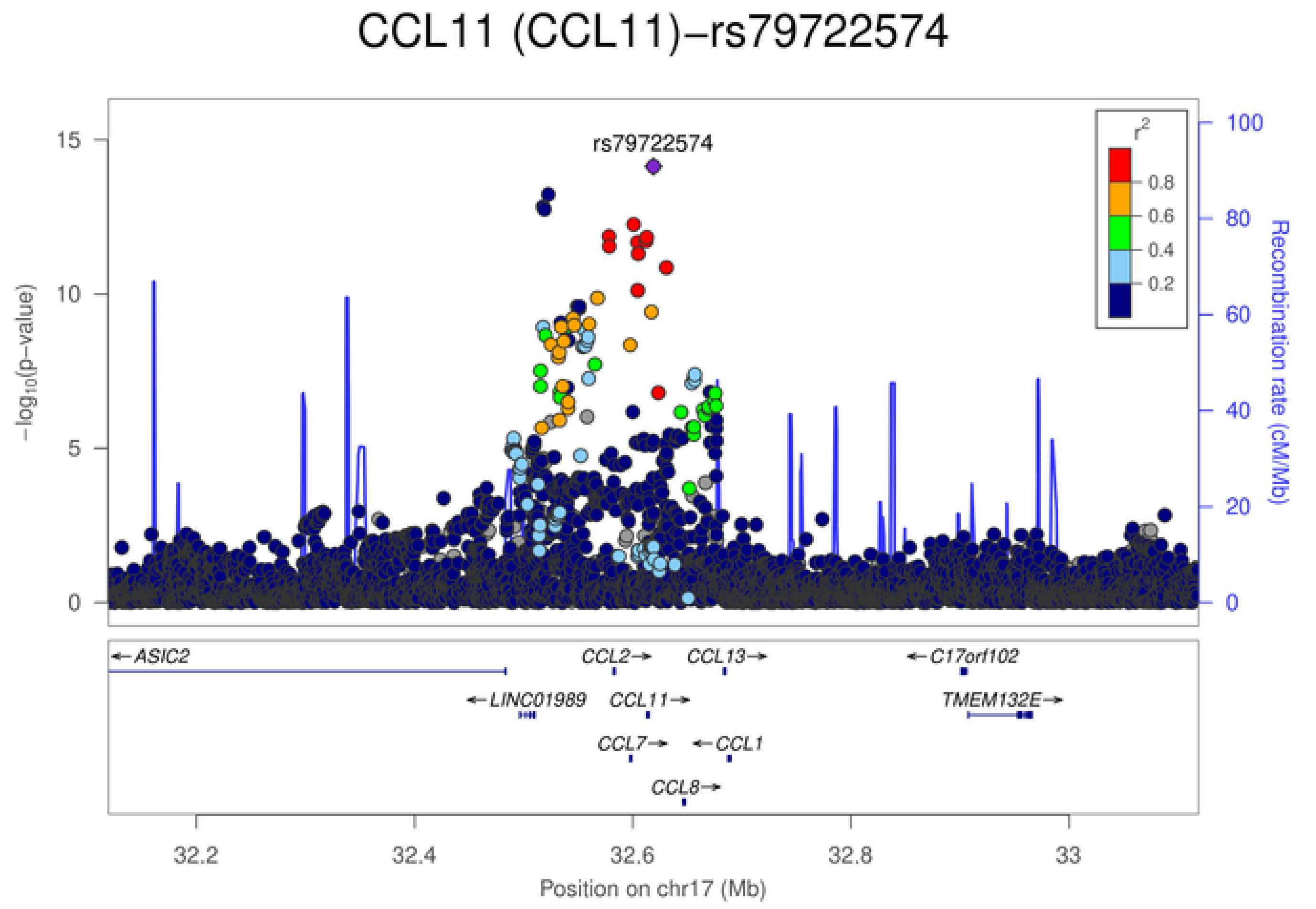
CCL11 (CCL11)−rs79722574
rs79722574
$-\log_{10}$(p-value)
Recombination rate (cM/Mb)
$r^2$
0.8
0.6
0.4
0.2
ASIC2
CCL2
CCL13
C17orf102
LINC01989
CCL11
TMEM132E
CCL7
CCL1
CCL8
32.2 32.4 32.6 32.8 33
Position on chr17 (Mb)

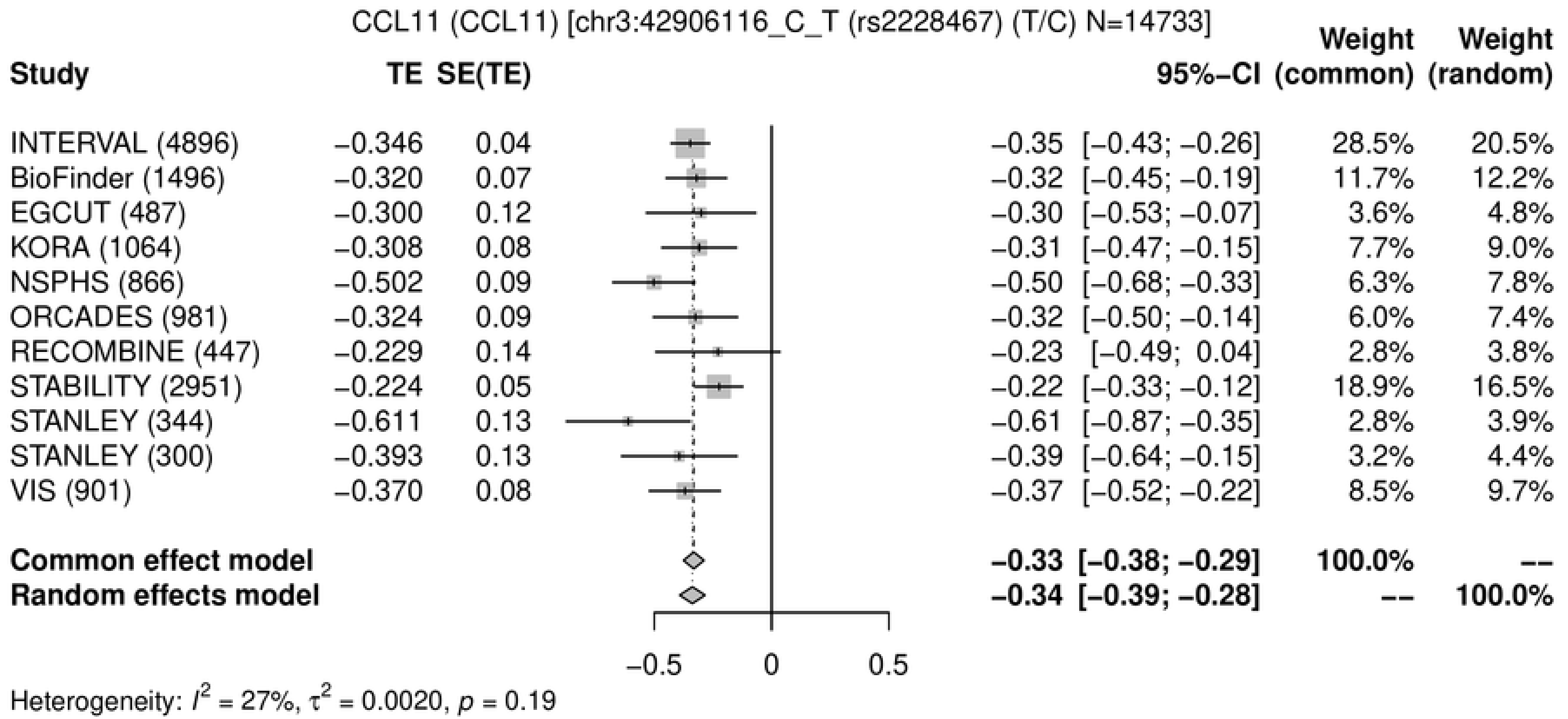
CCL11 (CCL11) [chr3:42906116_C_T (rs2228467) (T/C) N=14733]
Study TE SE(TE) 95%−CI Weight (common) Weight (random)
INTERVAL (4896) −0.346 0.04 −0.35 [−0.43; −0.26] 28.5% 20.5%
BioFinder (1496) −0.320 0.07 −0.32 [−0.45; −0.19] 11.7% 12.2%
EGCUT (487) −0.300 0.12 −0.30 [−0.53; −0.07] 3.6% 4.8%
KORA (1064) −0.308 0.08 −0.31 [−0.47; −0.15] 7.7% 9.0%
NSPHS (866) −0.502 0.09 −0.50 [−0.68; −0.33] 6.3% 7.8%
ORCADES (981) −0.324 0.09 −0.32 [−0.50; −0.14] 6.0% 7.4%
RECOMBINE (447) −0.229 0.14 −0.23 [−0.49; 0.04] 2.8% 3.8%
STABILITY (2951) −0.224 0.05 −0.22 [−0.33; −0.12] 18.9% 16.5%
STANLEY (344) −0.611 0.13 −0.61 [−0.87; −0.35] 2.8% 3.9%
STANLEY (300) −0.393 0.13 −0.39 [−0.64; −0.15] 3.2% 4.4%
VIS (901) −0.370 0.08 −0.37 [−0.52; −0.22] 8.5% 9.7%
Common effect model −0.33 [−0.38; −0.29] 100.0% --
Random effects model −0.34 [−0.39; −0.28] -- 100.0%
−0.5 0 0.5
Heterogeneity: $I^2$ = 27%, $\tau^2$ = 0.0020, $p$ = 0.19

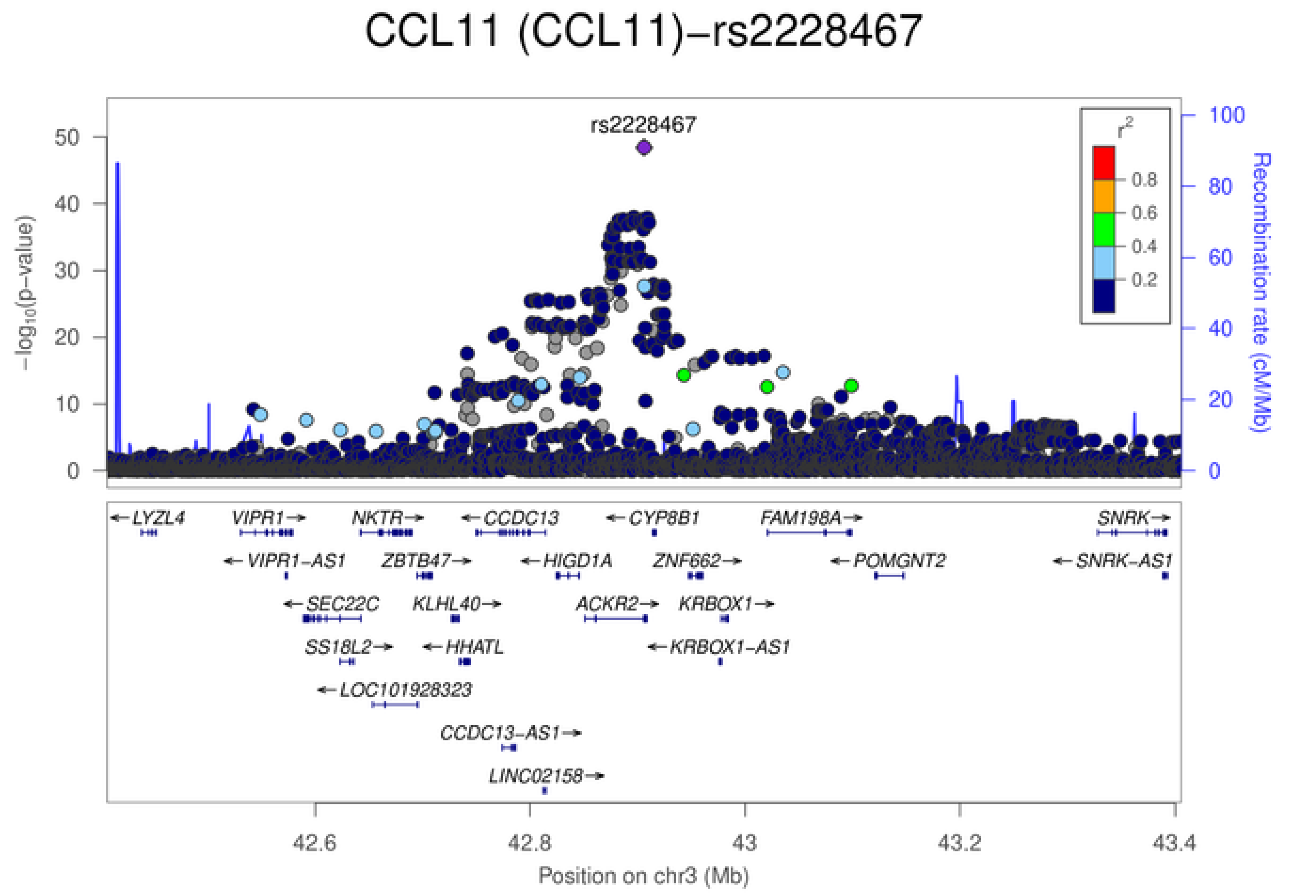
CCL11 (CCL11)−rs2228467
rs2228467
−log10(p−value)
Recombination rate (cM/Mb)
r2
0.8
0.6
0.4
0.2
LYZL4
VIPR1
NKTR
CCDC13
CYP8B1
FAM198A
SNRK
VIPR1−AS1
ZBTB47
HIGD1A
ZNF662
POMGNT2
SNRK−AS1
SEC22C
KLHL40
ACKR2
KRBOX1
SS18L2
HHATL
KRBOX1−AS1
LOC101928323
CCDC13−AS1
LINC02158
42.6
42.8
43
43.2
43.4
Position on chr3 (Mb)

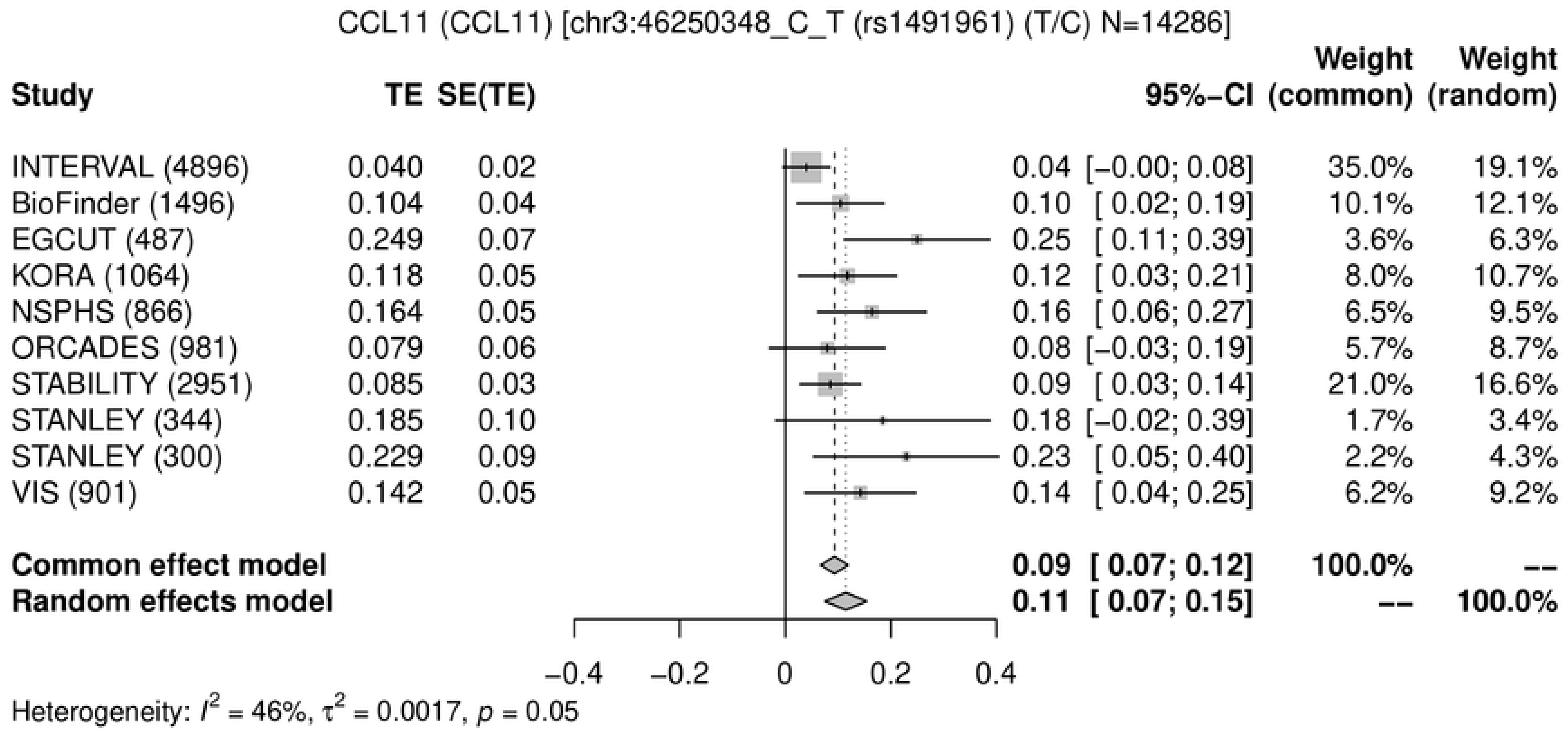
CCL11 (CCL11) [chr3:46250348_C_T (rs1491961) (T/C) N=14286]
Study | TE | SE(TE) | 95%-CI | Weight (common) | Weight (random)
INTERVAL (4896) | 0.040 | 0.02 | 0.04 [−0.00; 0.08] | 35.0% | 19.1%
BioFinder (1496) | 0.104 | 0.04 | 0.10 [ 0.02; 0.19] | 10.1% | 12.1%
EGCUT (487) | 0.249 | 0.07 | 0.25 [ 0.11; 0.39] | 3.6% | 6.3%
KORA (1064) | 0.118 | 0.05 | 0.12 [ 0.03; 0.21] | 8.0% | 10.7%
NSPHS (866) | 0.164 | 0.05 | 0.16 [ 0.06; 0.27] | 6.5% | 9.5%
ORCADES (981) | 0.079 | 0.06 | 0.08 [−0.03; 0.19] | 5.7% | 8.7%
STABILITY (2951) | 0.085 | 0.03 | 0.09 [ 0.03; 0.14] | 21.0% | 16.6%
STANLEY (344) | 0.185 | 0.10 | 0.18 [−0.02; 0.39] | 1.7% | 3.4%
STANLEY (300) | 0.229 | 0.09 | 0.23 [ 0.05; 0.40] | 2.2% | 4.3%
VIS (901) | 0.142 | 0.05 | 0.14 [ 0.04; 0.25] | 6.2% | 9.2%
Common effect model | 0.09 [ 0.07; 0.12] | 100.0% | --
Random effects model | 0.11 [ 0.07; 0.15] | -- | 100.0%
−0.4 −0.2 0 0.2 0.4
Heterogeneity: $I^2$ = 46%, $\tau^2$ = 0.0017, $p$ = 0.05

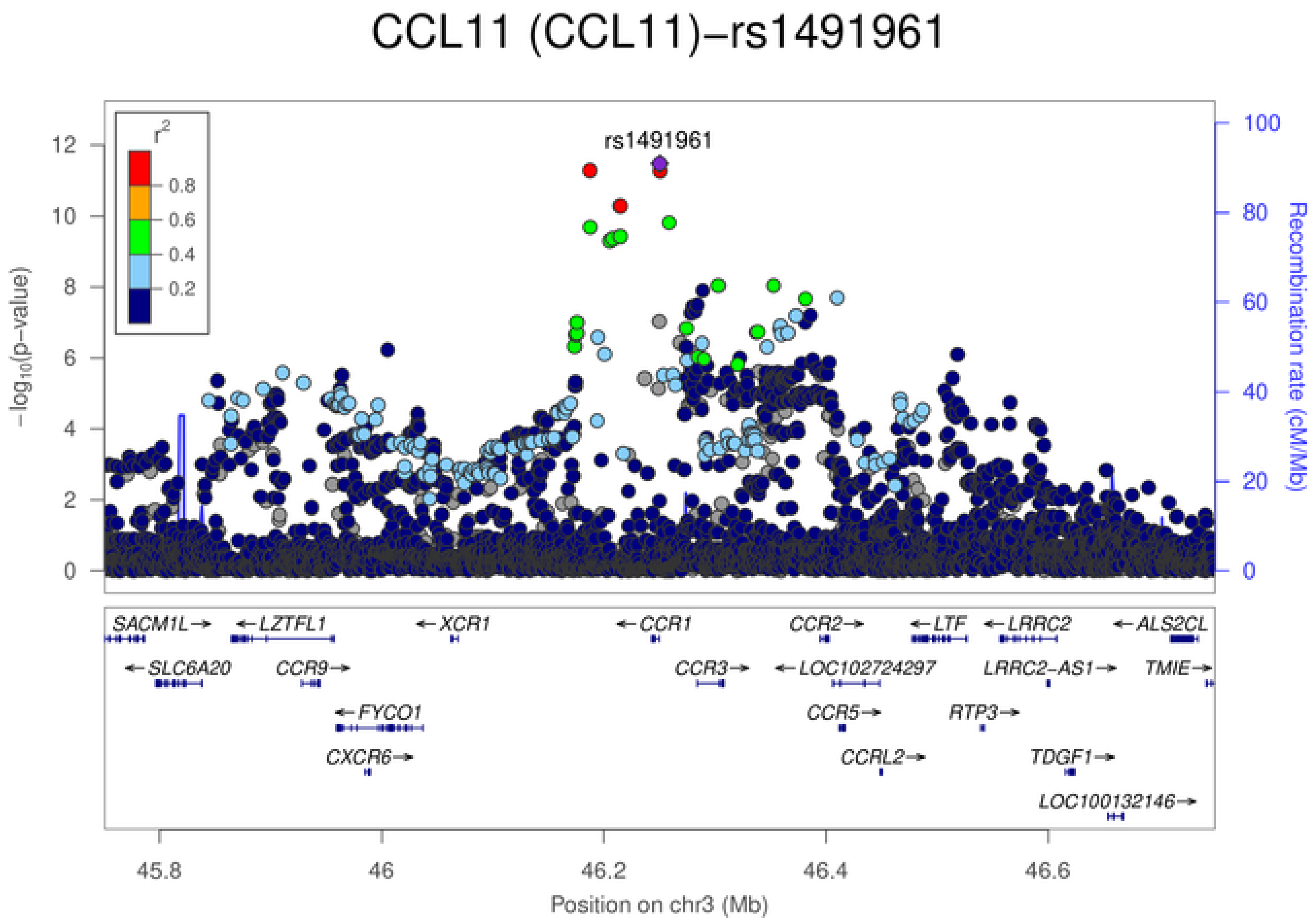
CCL11 (CCL11)−rs1491961
rs1491961
$r^2$
0.8
0.6
0.4
0.2
$-\log_{10}$(p−value)
Recombination rate (cM/Mb)
SACM1L→
←LZTFL1
←XCR1
←CCR1
CCR2→
←LTF
←LRRC2
←ALS2CL
←SLC6A20
CCR9→
CCR3→
←LOC102724297
LRRC2−AS1→
TMIE→
←FYCO1
CCR5→
RTP3→
CXCR6→
CCRL2→
TDGF1→
LOC100132146→
45.8 46 46.2 46.4 46.6
Position on chr3 (Mb)

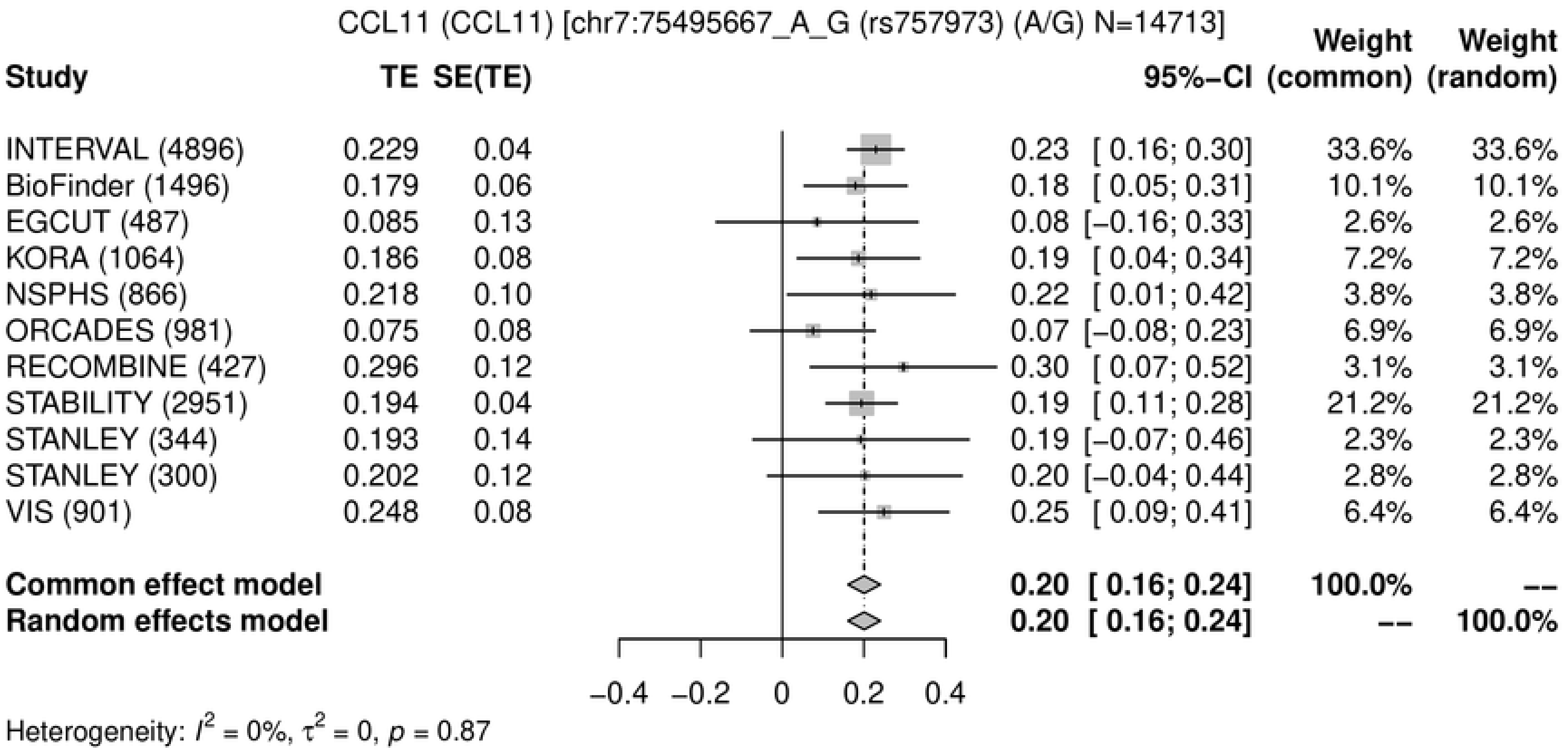
CCL11 (CCL11) [chr7:75495667_A_G (rs757973) (A/G) N=14713]
Study
TE
SE(TE)
95%−CI
Weight (common)
Weight (random)
INTERVAL (4896) 0.229 0.04 0.23 [ 0.16; 0.30] 33.6% 33.6%
BioFinder (1496) 0.179 0.06 0.18 [ 0.05; 0.31] 10.1% 10.1%
EGCUT (487) 0.085 0.13 0.08 [−0.16; 0.33] 2.6% 2.6%
KORA (1064) 0.186 0.08 0.19 [ 0.04; 0.34] 7.2% 7.2%
NSPHS (866) 0.218 0.10 0.22 [ 0.01; 0.42] 3.8% 3.8%
ORCADES (981) 0.075 0.08 0.07 [−0.08; 0.23] 6.9% 6.9%
RECOMBINE (427) 0.296 0.12 0.30 [ 0.07; 0.52] 3.1% 3.1%
STABILITY (2951) 0.194 0.04 0.19 [ 0.11; 0.28] 21.2% 21.2%
STANLEY (344) 0.193 0.14 0.19 [−0.07; 0.46] 2.3% 2.3%
STANLEY (300) 0.202 0.12 0.20 [−0.04; 0.44] 2.8% 2.8%
VIS (901) 0.248 0.08 0.25 [ 0.09; 0.41] 6.4% 6.4%
Common effect model 0.20 [ 0.16; 0.24] 100.0% --
Random effects model 0.20 [ 0.16; 0.24] -- 100.0%
−0.4 −0.2 0 0.2 0.4
Heterogeneity: $I^2$ = 0%, $\tau^2$ = 0, $p$ = 0.87

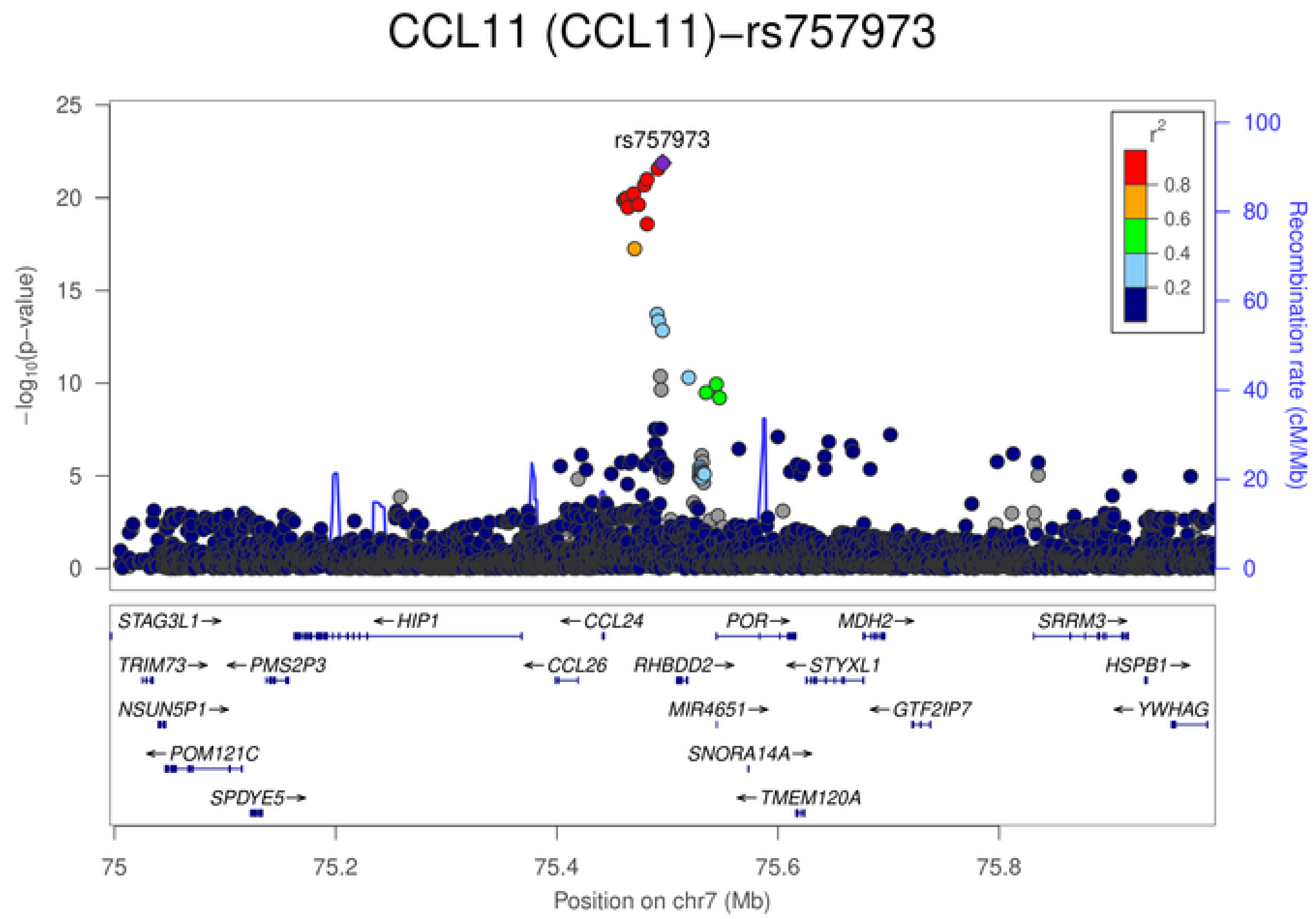
CCL11 (CCL11)−rs757973
rs757973
$-\log_{10}$(p−value)
Recombination rate (cM/Mb)
$r^2$
0.8
0.6
0.4
0.2
STAG3L1→
←HIP1
←CCL24
POR→
MDH2→
SRRM3→
TRIM73→
←PMS2P3
←CCL26
RHBDD2→
←STYXL1
HSPB1→
NSUN5P1→
MIR4651→
←GTF2IP7
←YWHAG
←POM121C
SNORA14A→
SPDYE5→
←TMEM120A
75 75.2 75.4 75.6 75.8
Position on chr7 (Mb)

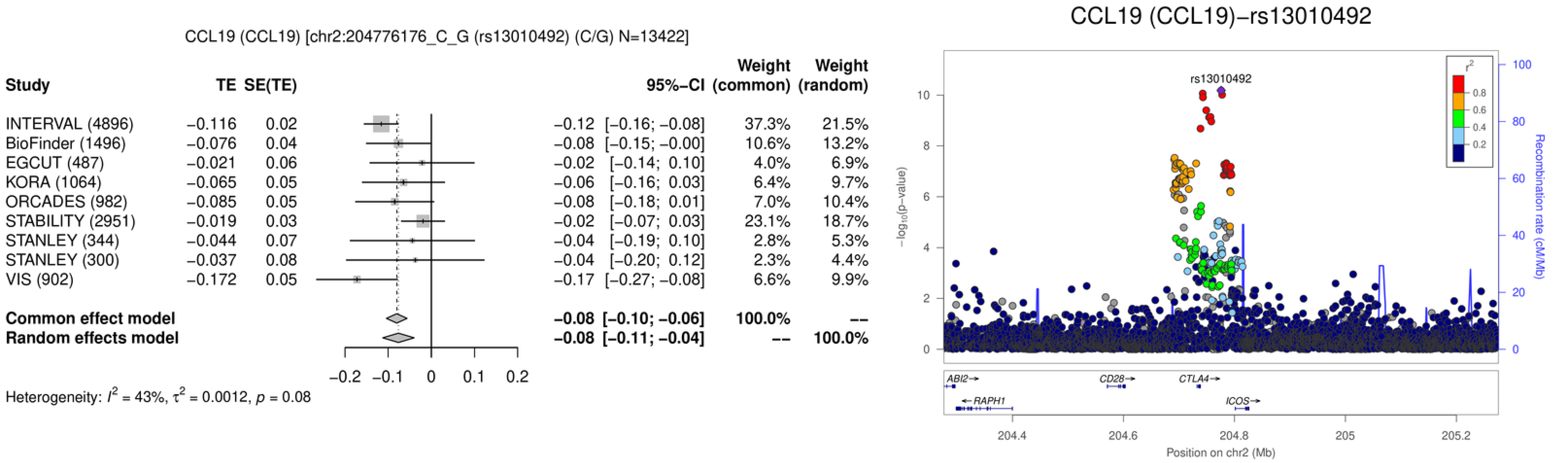
CCL19 (CCL19) [chr2:204776176_C_G (rs13010492) (C/G) N=13422]
Study
TE
SE(TE)
95%−CI
Weight (common)
Weight (random)
INTERVAL (4896) −0.116 0.02 −0.12 [−0.16; −0.08] 37.3% 21.5%
BioFinder (1496) −0.076 0.04 −0.08 [−0.15; −0.00] 10.6% 13.2%
EGCUT (487) −0.021 0.06 −0.02 [−0.14; 0.10] 4.0% 6.9%
KORA (1064) −0.065 0.05 −0.06 [−0.16; 0.03] 6.4% 9.7%
ORCADES (982) −0.085 0.05 −0.08 [−0.18; 0.01] 7.0% 10.4%
STABILITY (2951) −0.019 0.03 −0.02 [−0.07; 0.03] 23.1% 18.7%
STANLEY (344) −0.044 0.07 −0.04 [−0.19; 0.10] 2.8% 5.3%
STANLEY (300) −0.037 0.08 −0.04 [−0.20; 0.12] 2.3% 4.4%
VIS (902) −0.172 0.05 −0.17 [−0.27; −0.08] 6.6% 9.9%
Common effect model −0.08 [−0.10; −0.06] 100.0% --
Random effects model −0.08 [−0.11; −0.04] -- 100.0%
−0.2 −0.1 0 0.1 0.2
Heterogeneity: $I^2$ = 43%, $\tau^2$ = 0.0012, $p$ = 0.08
CCL19 (CCL19)−rs13010492
rs13010492
$-\log_{10}$(p−value)
Recombination rate (cM/Mb)
$r^2$
0.8
0.6
0.4
0.2
0 2 4 6 8 10
0 20 40 60 80 100
ABI2→
CD28→
CTLA4→
←RAPH1
ICOS→
204.4 204.6 204.8 205 205.2
Position on chr2 (Mb)

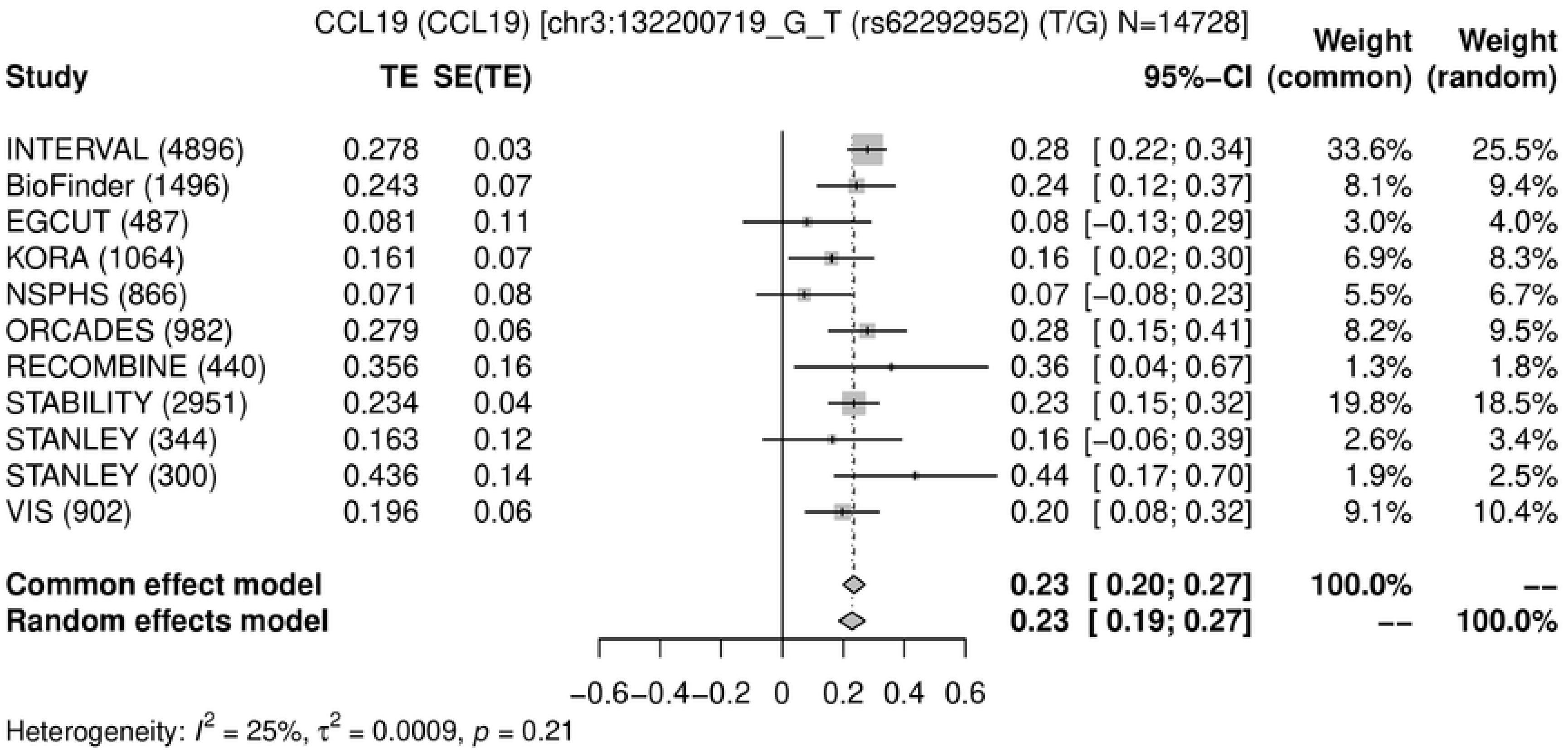
CCL19 (CCL19) [chr3:132200719_G_T (rs62292952) (T/G) N=14728]
Study
TE
SE(TE)
95%-CI
Weight (common)
Weight (random)
INTERVAL (4896) 0.278 0.03 0.28 [ 0.22; 0.34] 33.6% 25.5%
BioFinder (1496) 0.243 0.07 0.24 [ 0.12; 0.37] 8.1% 9.4%
EGCUT (487) 0.081 0.11 0.08 [−0.13; 0.29] 3.0% 4.0%
KORA (1064) 0.161 0.07 0.16 [ 0.02; 0.30] 6.9% 8.3%
NSPHS (866) 0.071 0.08 0.07 [−0.08; 0.23] 5.5% 6.7%
ORCADES (982) 0.279 0.06 0.28 [ 0.15; 0.41] 8.2% 9.5%
RECOMBINE (440) 0.356 0.16 0.36 [ 0.04; 0.67] 1.3% 1.8%
STABILITY (2951) 0.234 0.04 0.23 [ 0.15; 0.32] 19.8% 18.5%
STANLEY (344) 0.163 0.12 0.16 [−0.06; 0.39] 2.6% 3.4%
STANLEY (300) 0.436 0.14 0.44 [ 0.17; 0.70] 1.9% 2.5%
VIS (902) 0.196 0.06 0.20 [ 0.08; 0.32] 9.1% 10.4%
Common effect model 0.23 [ 0.20; 0.27] 100.0% --
Random effects model 0.23 [ 0.19; 0.27] -- 100.0%
−0.6 −0.4 −0.2 0 0.2 0.4 0.6
Heterogeneity: $I^2$ = 25%, $\tau^2$ = 0.0009, $p$ = 0.21

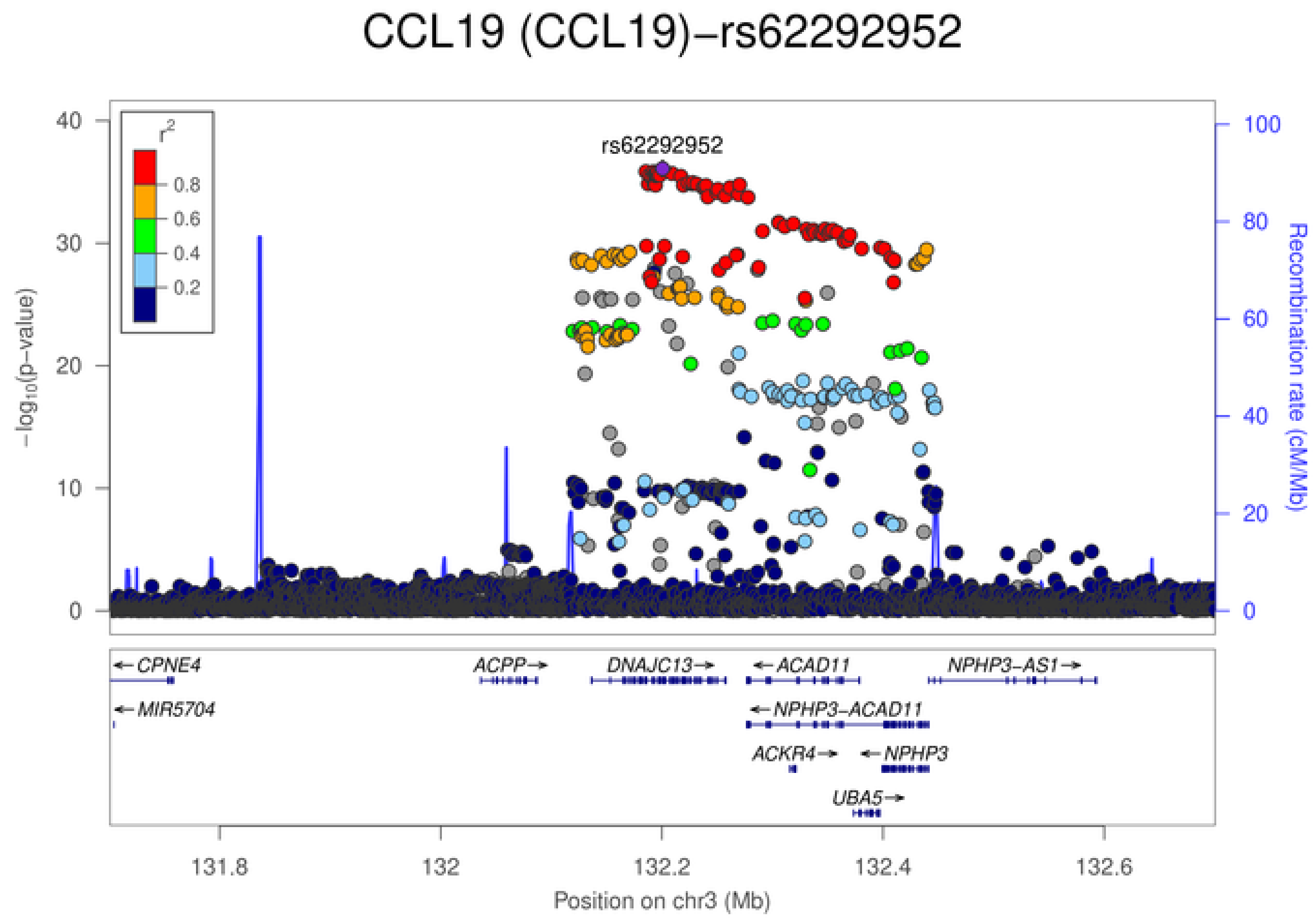
CCL19 (CCL19)−rs62292952
rs62292952
$r^2$
0.8
0.6
0.4
0.2
$-\log_{10}$(p−value)
0
10
20
30
40
Recombination rate (cM/Mb)
0
20
40
60
80
100
←CPNE4
←MIR5704
ACPP→
DNAJC13→
←ACAD11
←NPHP3−ACAD11
ACKR4→
←NPHP3
UBA5→
NPHP3−AS1→
131.8
132
132.2
132.4
132.6
Position on chr3 (Mb)

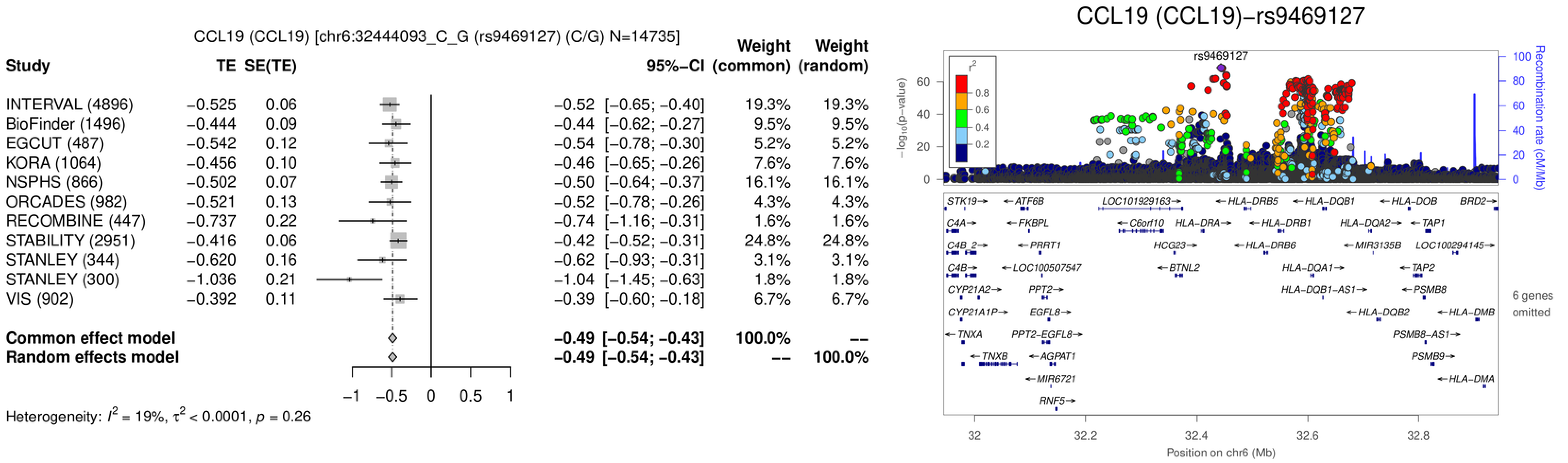
CCL19 (CCL19) [chr6:32444093_C_G (rs9469127) (C/G) N=14735]
Study
TE
SE(TE)
95%−CI
Weight (common)
Weight (random)
INTERVAL (4896) −0.525 0.06 −0.52 [−0.65; −0.40] 19.3% 19.3%
BioFinder (1496) −0.444 0.09 −0.44 [−0.62; −0.27] 9.5% 9.5%
EGCUT (487) −0.542 0.12 −0.54 [−0.78; −0.30] 5.2% 5.2%
KORA (1064) −0.456 0.10 −0.46 [−0.65; −0.26] 7.6% 7.6%
NSPHS (866) −0.502 0.07 −0.50 [−0.64; −0.37] 16.1% 16.1%
ORCADES (982) −0.521 0.13 −0.52 [−0.78; −0.26] 4.3% 4.3%
RECOMBINE (447) −0.737 0.22 −0.74 [−1.16; −0.31] 1.6% 1.6%
STABILITY (2951) −0.416 0.06 −0.42 [−0.52; −0.31] 24.8% 24.8%
STANLEY (344) −0.620 0.16 −0.62 [−0.93; −0.31] 3.1% 3.1%
STANLEY (300) −1.036 0.21 −1.04 [−1.45; −0.63] 1.8% 1.8%
VIS (902) −0.392 0.11 −0.39 [−0.60; −0.18] 6.7% 6.7%
Common effect model −0.49 [−0.54; −0.43] 100.0% --
Random effects model −0.49 [−0.54; −0.43] -- 100.0%
−1 −0.5 0 0.5 1
Heterogeneity: $I^2$ = 19%, $\tau^2$ < 0.0001, $p$ = 0.26
CCL19 (CCL19)−rs9469127
rs9469127
$-\log_{10}$(p−value)
0 20 40 60
$r^2$
0.8 0.6 0.4 0.2
Recombination rate (cM/Mb)
0 20 40 60 80 100
STK19 ATF6B LOC101929163 HLA−DRB5 HLA−DQB1 HLA−DOB BRD2
C4A FKBPL C6orf10 HLA−DRA HLA−DRB1 HLA−DQA2 TAP1
C4B_2 PRRT1 HCG23 HLA−DRB6 MIR3135B LOC100294145
C4B LOC100507547 BTNL2 HLA−DQA1 TAP2
CYP21A2 PPT2 HLA−DQB1−AS1 PSMB8
CYP21A1P EGFL8 HLA−DQB2 HLA−DMB
TNXA PPT2−EGFL8 PSMB8−AS1
TNXB AGPAT1 PSMB9
MIR6721 HLA−DMA
RNF5
6 genes omitted
32 32.2 32.4 32.6 32.8
Position on chr6 (Mb)

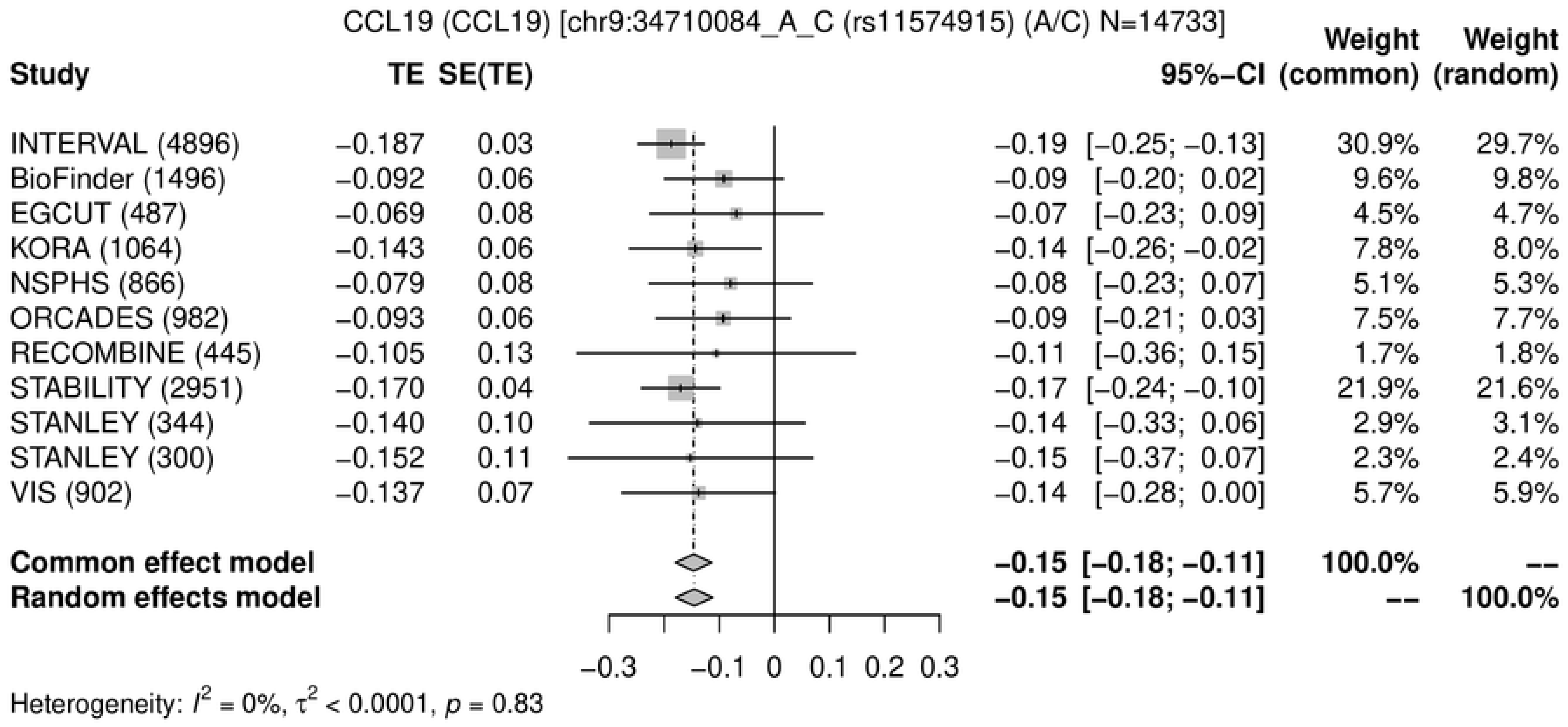
CCL19 (CCL19) [chr9:34710084_A_C (rs11574915) (A/C) N=14733]
Study TE SE(TE) 95%−CI Weight (common) Weight (random)
INTERVAL (4896) −0.187 0.03 −0.19 [−0.25; −0.13] 30.9% 29.7%
BioFinder (1496) −0.092 0.06 −0.09 [−0.20; 0.02] 9.6% 9.8%
EGCUT (487) −0.069 0.08 −0.07 [−0.23; 0.09] 4.5% 4.7%
KORA (1064) −0.143 0.06 −0.14 [−0.26; −0.02] 7.8% 8.0%
NSPHS (866) −0.079 0.08 −0.08 [−0.23; 0.07] 5.1% 5.3%
ORCADES (982) −0.093 0.06 −0.09 [−0.21; 0.03] 7.5% 7.7%
RECOMBINE (445) −0.105 0.13 −0.11 [−0.36; 0.15] 1.7% 1.8%
STABILITY (2951) −0.170 0.04 −0.17 [−0.24; −0.10] 21.9% 21.6%
STANLEY (344) −0.140 0.10 −0.14 [−0.33; 0.06] 2.9% 3.1%
STANLEY (300) −0.152 0.11 −0.15 [−0.37; 0.07] 2.3% 2.4%
VIS (902) −0.137 0.07 −0.14 [−0.28; 0.00] 5.7% 5.9%
Common effect model −0.15 [−0.18; −0.11] 100.0% --
Random effects model −0.15 [−0.18; −0.11] -- 100.0%
−0.3 −0.1 0 0.1 0.2 0.3
Heterogeneity: $I^2 = 0\%$, $\tau^2 < 0.0001$, $p = 0.83$

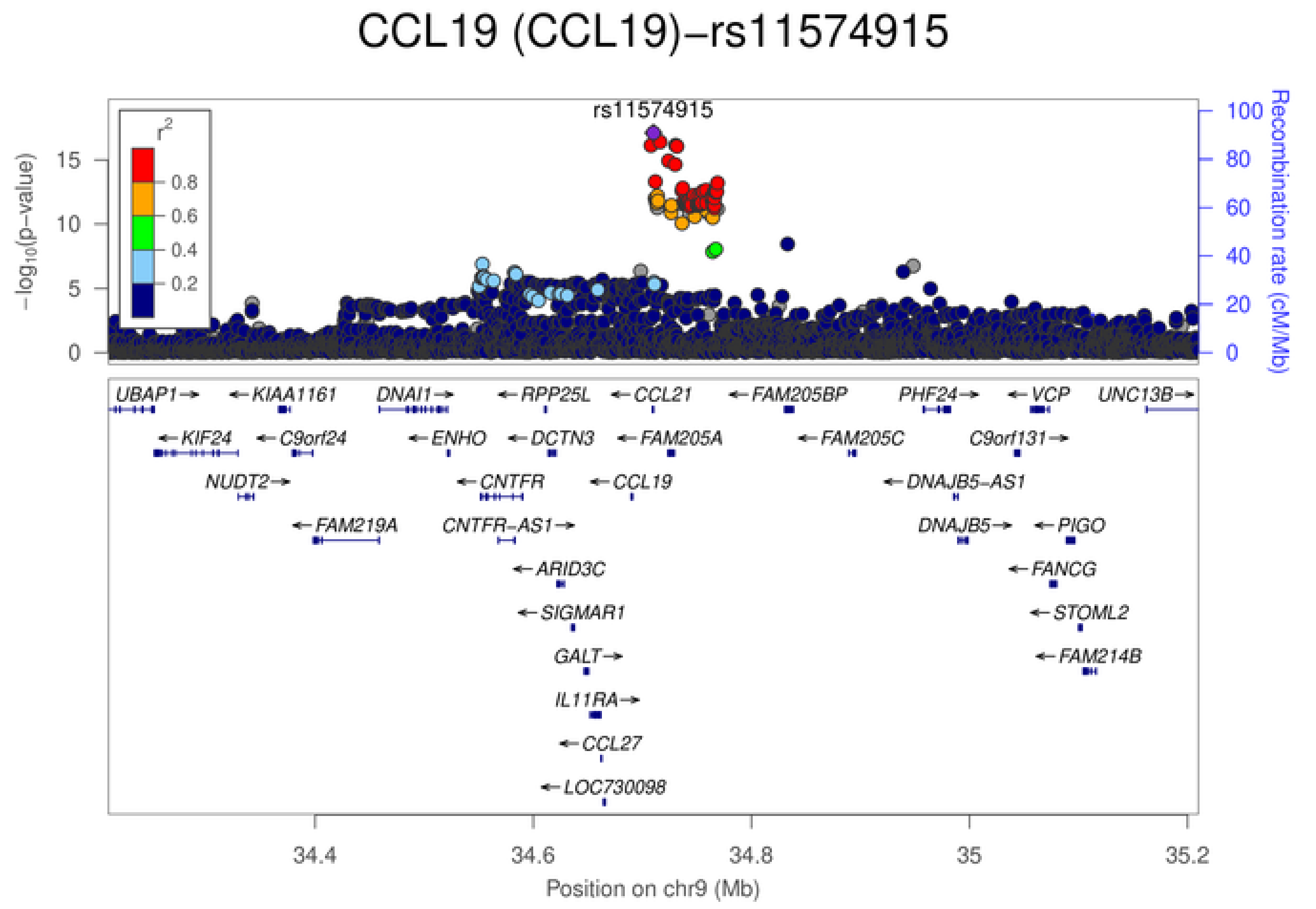
CCL19 (CCL19)−rs11574915
rs11574915
$-\log_{10}$(p−value)
Recombination rate (cM/Mb)
$r^2$
UBAP1 KIAA1161 DNAI1 RPP25L CCL21 FAM205BP PHF24 VCP UNC13B
KIF24 C9orf24 ENHO DCTN3 FAM205A FAM205C C9orf131
NUDT2 CNTFR CCL19 DNAJB5-AS1
FAM219A CNTFR-AS1 DNAJB5 PIGO
ARID3C FANCG
SIGMAR1 STOML2
GALT FAM214B
IL11RA
CCL27
LOC730098
34.4 34.6 34.8 35 35.2
Position on chr9 (Mb)

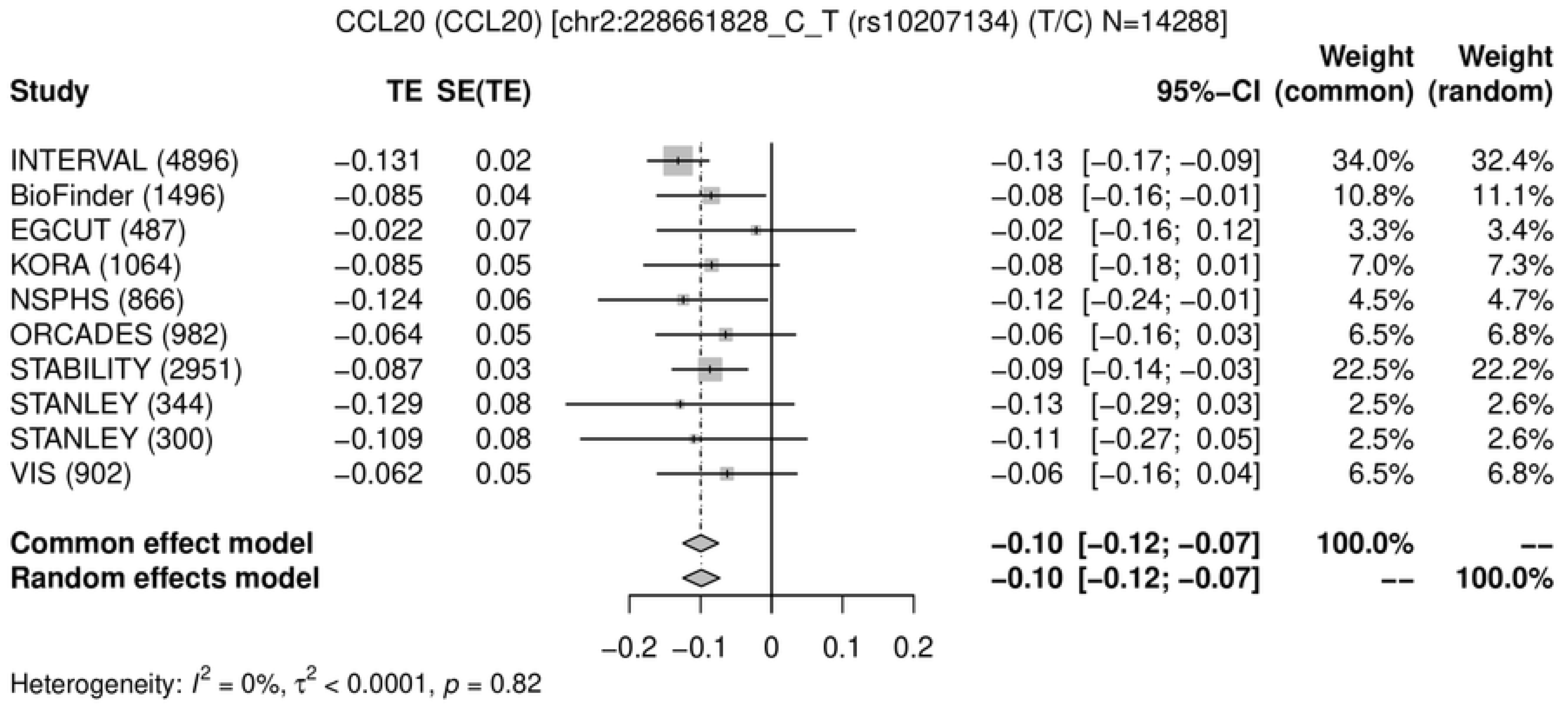
CCL20 (CCL20) [chr2:228661828_C_T (rs10207134) (T/C) N=14288]
Study | TE | SE(TE) | 95%-CI | Weight (common) | Weight (random)
INTERVAL (4896) | −0.131 | 0.02 | −0.13 [−0.17; −0.09] | 34.0% | 32.4%
BioFinder (1496) | −0.085 | 0.04 | −0.08 [−0.16; −0.01] | 10.8% | 11.1%
EGCUT (487) | −0.022 | 0.07 | −0.02 [−0.16; 0.12] | 3.3% | 3.4%
KORA (1064) | −0.085 | 0.05 | −0.08 [−0.18; 0.01] | 7.0% | 7.3%
NSPHS (866) | −0.124 | 0.06 | −0.12 [−0.24; −0.01] | 4.5% | 4.7%
ORCADES (982) | −0.064 | 0.05 | −0.06 [−0.16; 0.03] | 6.5% | 6.8%
STABILITY (2951) | −0.087 | 0.03 | −0.09 [−0.14; −0.03] | 22.5% | 22.2%
STANLEY (344) | −0.129 | 0.08 | −0.13 [−0.29; 0.03] | 2.5% | 2.6%
STANLEY (300) | −0.109 | 0.08 | −0.11 [−0.27; 0.05] | 2.5% | 2.6%
VIS (902) | −0.062 | 0.05 | −0.06 [−0.16; 0.04] | 6.5% | 6.8%
Common effect model | −0.10 [−0.12; −0.07] | 100.0% | --
Random effects model | −0.10 [−0.12; −0.07] | -- | 100.0%
−0.2 −0.1 0 0.1 0.2
Heterogeneity: $I^2 = 0\%$, $\tau^2 < 0.0001$, $p = 0.82$

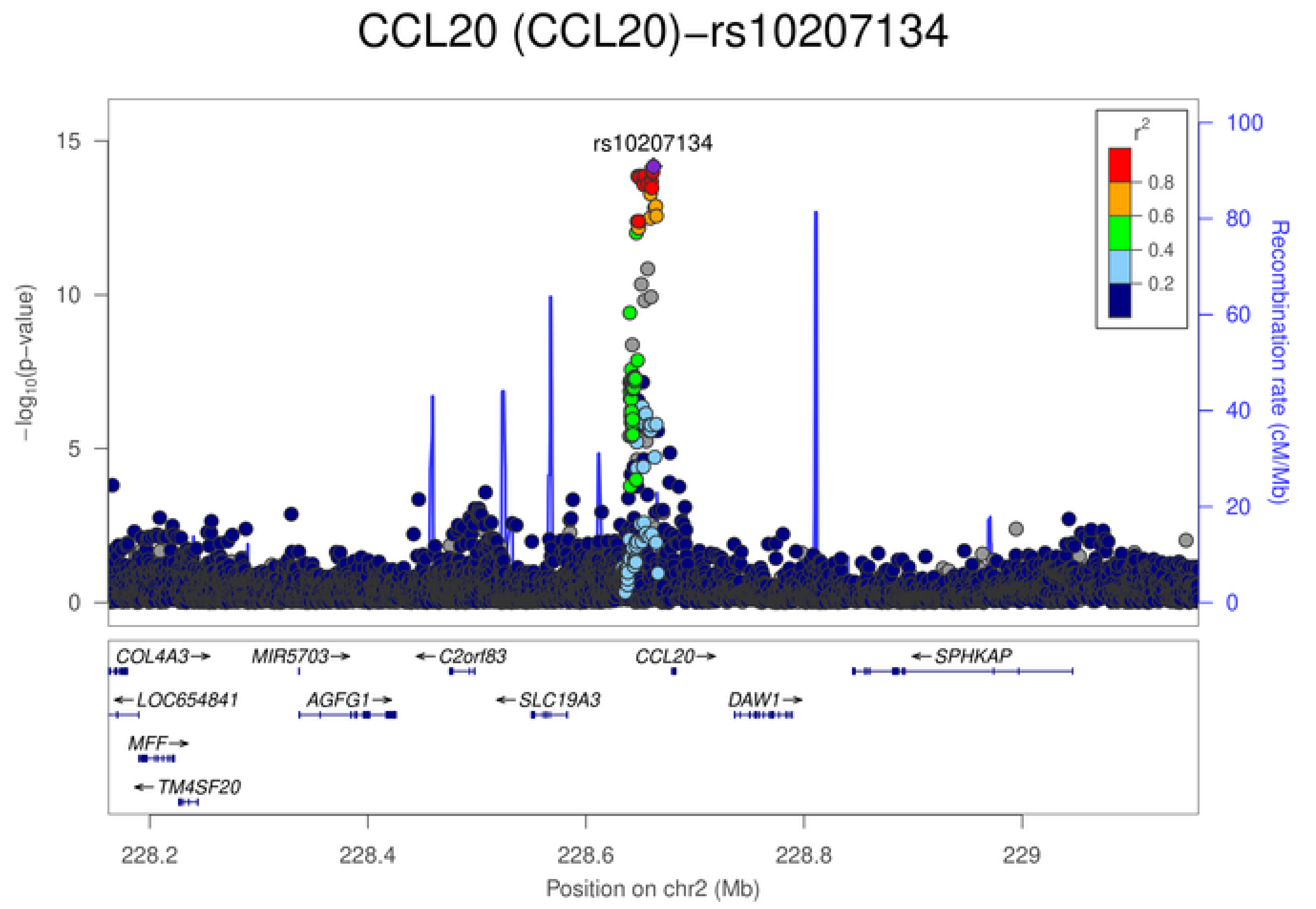
CCL20 (CCL20)−rs10207134
rs10207134
$-\log_{10}$(p−value)
0
5
10
15
Recombination rate (cM/Mb)
0
20
40
60
80
100
$r^2$
0.8
0.6
0.4
0.2
COL4A3→
MIR5703→
←C2orf83
CCL20→
←SPHKAP
←LOC654841
AGFG1→
←SLC19A3
DAW1→
MFF→
←TM4SF20
228.2
228.4
228.6
228.8
229
Position on chr2 (Mb)

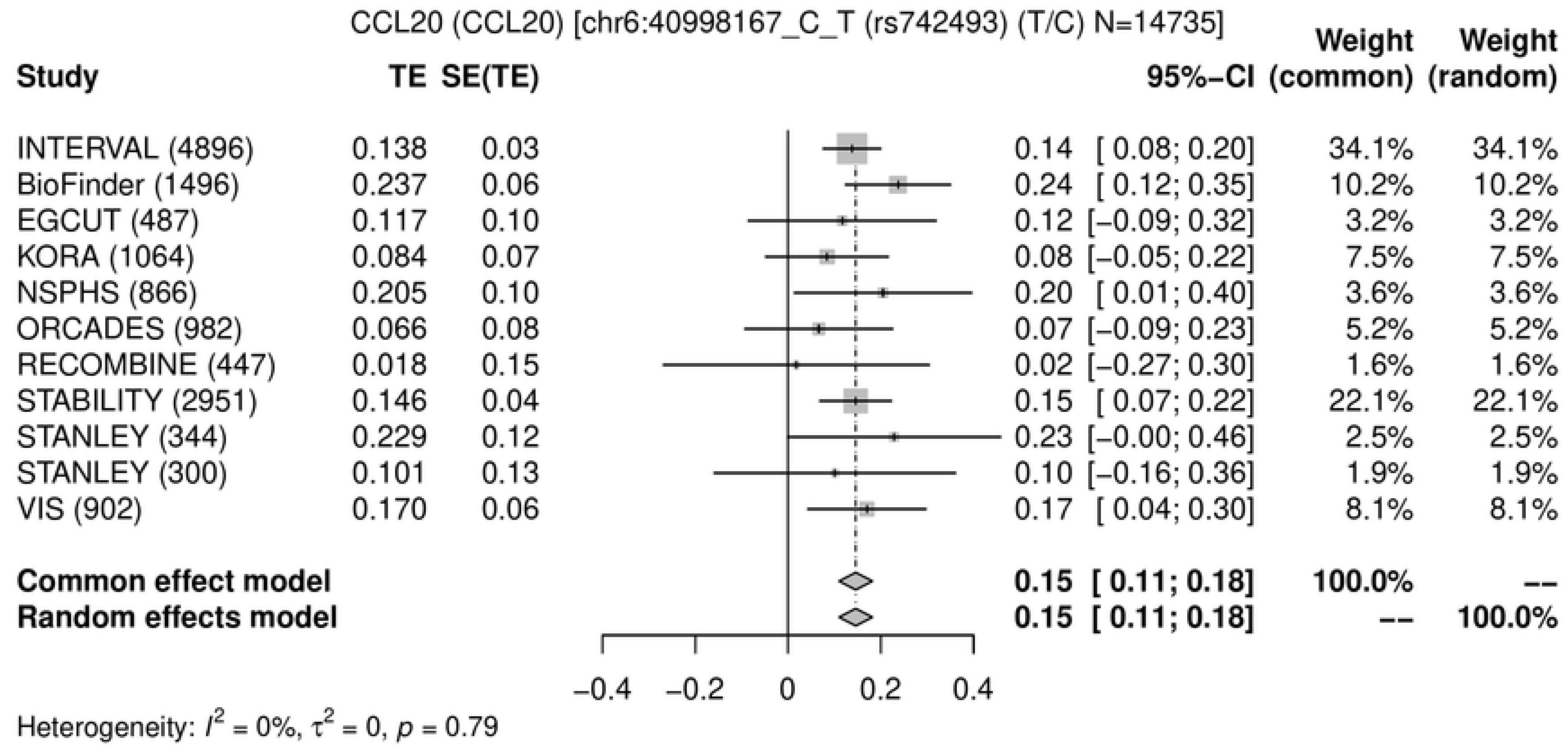
CCL20 (CCL20) [chr6:40998167_C_T (rs742493) (T/C) N=14735]
Study
TE
SE(TE)
95%−CI
Weight (common)
Weight (random)
INTERVAL (4896) 0.138 0.03 0.14 [ 0.08; 0.20] 34.1% 34.1%
BioFinder (1496) 0.237 0.06 0.24 [ 0.12; 0.35] 10.2% 10.2%
EGCUT (487) 0.117 0.10 0.12 [−0.09; 0.32] 3.2% 3.2%
KORA (1064) 0.084 0.07 0.08 [−0.05; 0.22] 7.5% 7.5%
NSPHS (866) 0.205 0.10 0.20 [ 0.01; 0.40] 3.6% 3.6%
ORCADES (982) 0.066 0.08 0.07 [−0.09; 0.23] 5.2% 5.2%
RECOMBINE (447) 0.018 0.15 0.02 [−0.27; 0.30] 1.6% 1.6%
STABILITY (2951) 0.146 0.04 0.15 [ 0.07; 0.22] 22.1% 22.1%
STANLEY (344) 0.229 0.12 0.23 [−0.00; 0.46] 2.5% 2.5%
STANLEY (300) 0.101 0.13 0.10 [−0.16; 0.36] 1.9% 1.9%
VIS (902) 0.170 0.06 0.17 [ 0.04; 0.30] 8.1% 8.1%
Common effect model 0.15 [ 0.11; 0.18] 100.0% --
Random effects model 0.15 [ 0.11; 0.18] -- 100.0%
−0.4 −0.2 0 0.2 0.4
Heterogeneity: $I^2 = 0\%$, $\tau^2 = 0$, $p = 0.79$

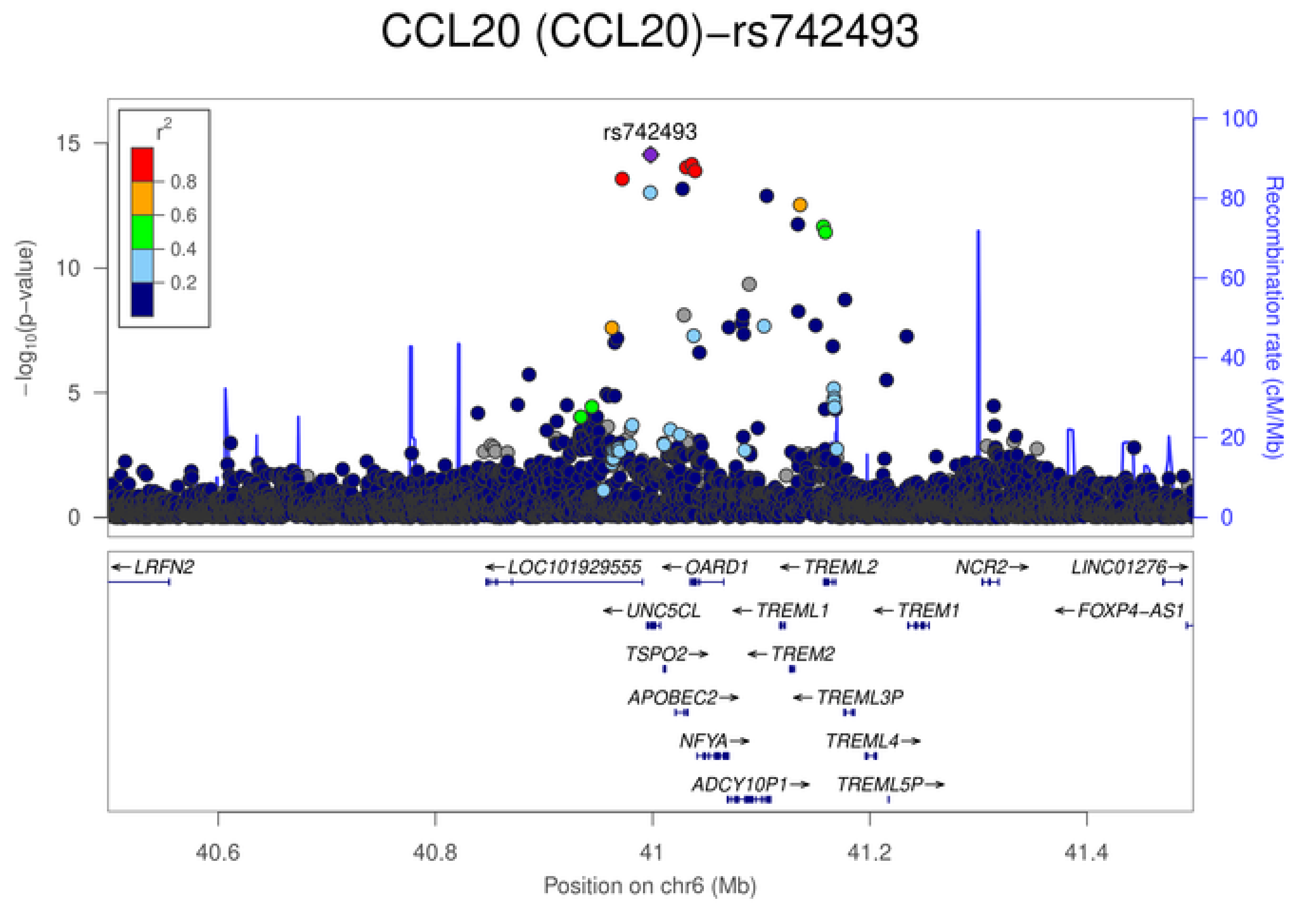
CCL20 (CCL20)−rs742493
rs742493
$r^2$
0.8
0.6
0.4
0.2
$-\log_{10}$(p-value)
Recombination rate (cM/Mb)
LRFN2
LOC101929555
OARD1
TREML2
NCR2
LINC01276
UNC5CL
TREML1
TREM1
FOXP4-AS1
TSPO2
TREM2
APOBEC2
TREML3P
NFYA
TREML4
ADCY10P1
TREML5P
40.6
40.8
41
41.2
41.4
Position on chr6 (Mb)

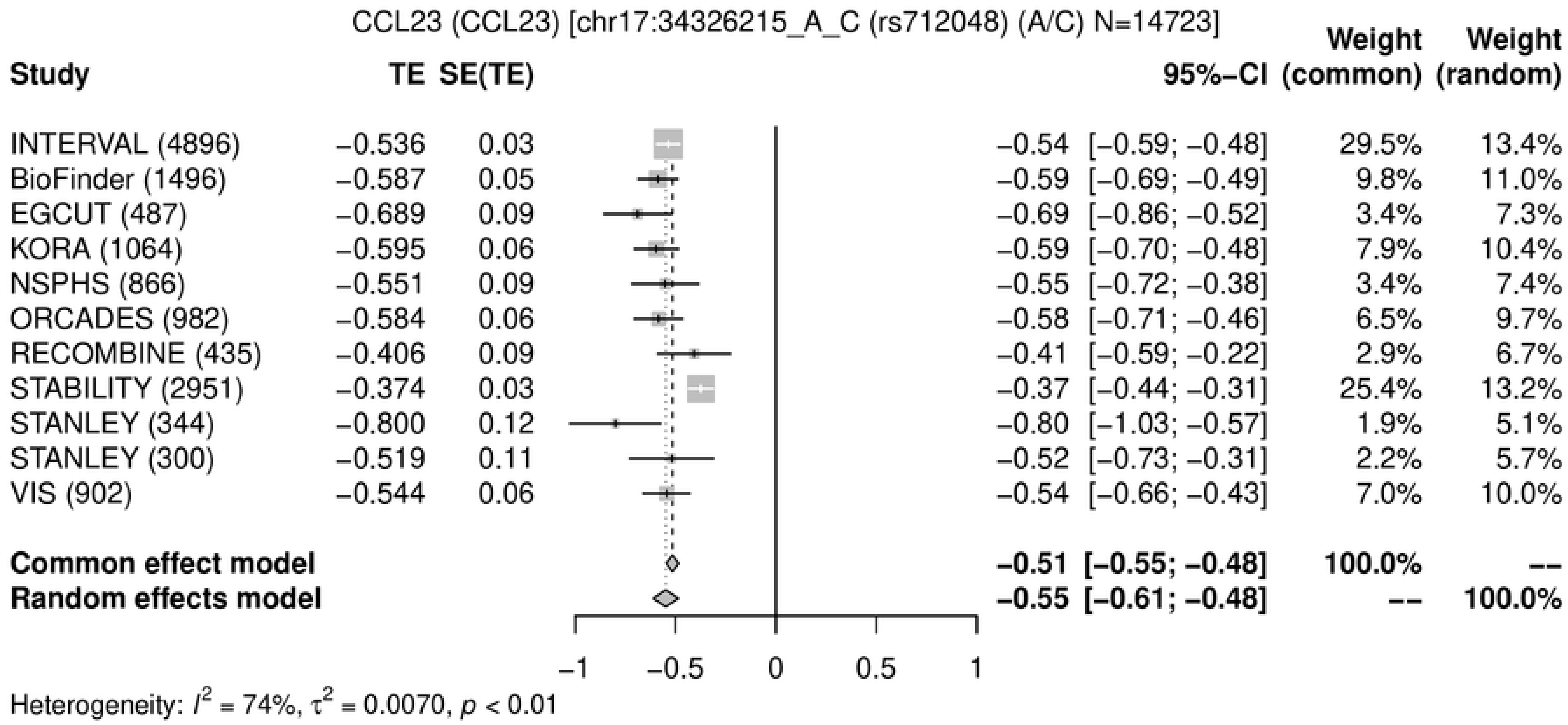
CCL23 (CCL23) [chr17:34326215_A_C (rs712048) (A/C) N=14723]
Study TE SE(TE) 95%−CI Weight (common) Weight (random)
INTERVAL (4896) −0.536 0.03 −0.54 [−0.59; −0.48] 29.5% 13.4%
BioFinder (1496) −0.587 0.05 −0.59 [−0.69; −0.49] 9.8% 11.0%
EGCUT (487) −0.689 0.09 −0.69 [−0.86; −0.52] 3.4% 7.3%
KORA (1064) −0.595 0.06 −0.59 [−0.70; −0.48] 7.9% 10.4%
NSPHS (866) −0.551 0.09 −0.55 [−0.72; −0.38] 3.4% 7.4%
ORCADES (982) −0.584 0.06 −0.58 [−0.71; −0.46] 6.5% 9.7%
RECOMBINE (435) −0.406 0.09 −0.41 [−0.59; −0.22] 2.9% 6.7%
STABILITY (2951) −0.374 0.03 −0.37 [−0.44; −0.31] 25.4% 13.2%
STANLEY (344) −0.800 0.12 −0.80 [−1.03; −0.57] 1.9% 5.1%
STANLEY (300) −0.519 0.11 −0.52 [−0.73; −0.31] 2.2% 5.7%
VIS (902) −0.544 0.06 −0.54 [−0.66; −0.43] 7.0% 10.0%
Common effect model −0.51 [−0.55; −0.48] 100.0% --
Random effects model −0.55 [−0.61; −0.48] -- 100.0%
−1 −0.5 0 0.5 1
Heterogeneity: $I^2$ = 74%, $\tau^2$ = 0.0070, $p$ < 0.01

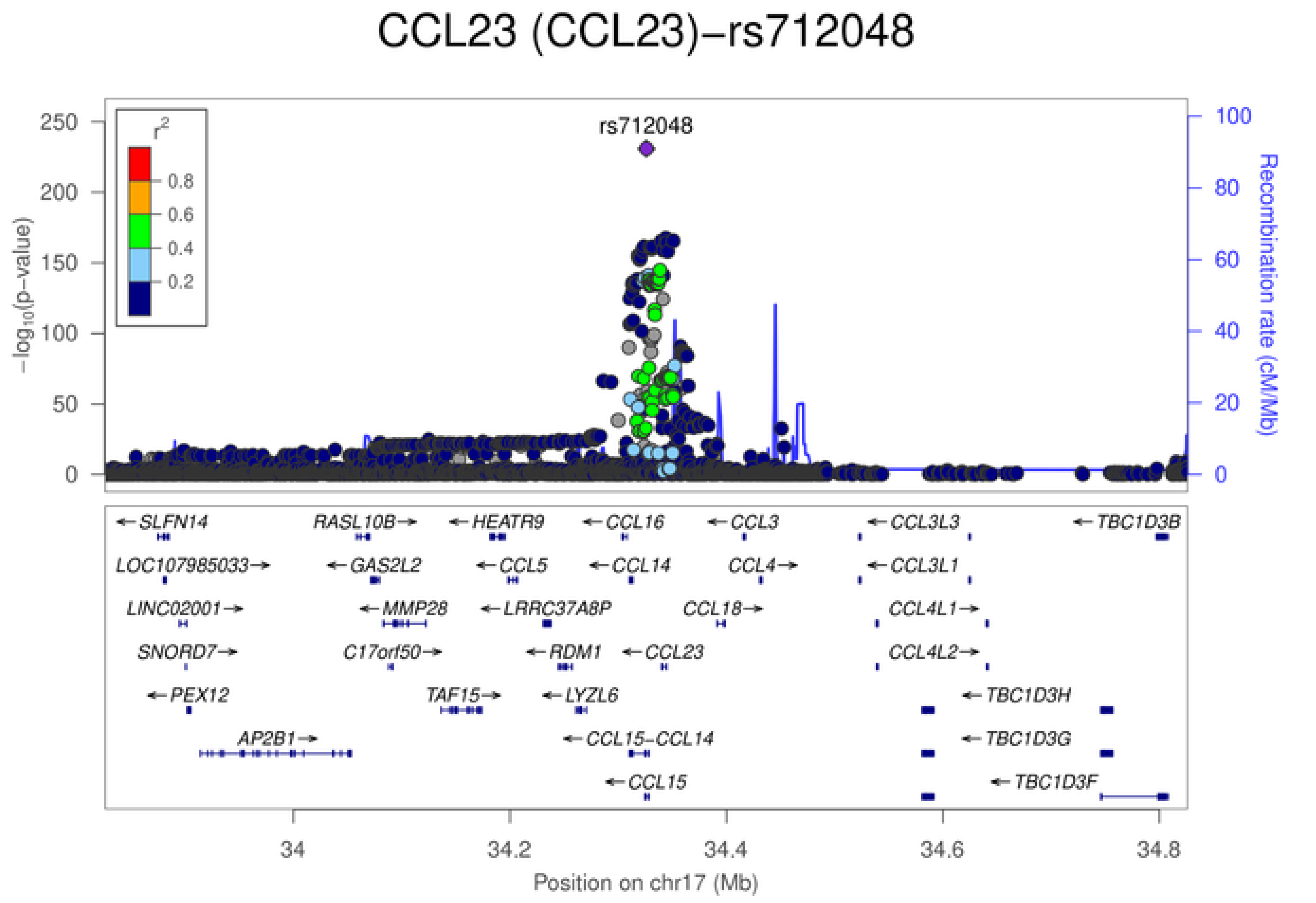
CCL23 (CCL23)−rs712048
rs712048
$r^2$
0.8
0.6
0.4
0.2
$-\log_{10}$(p−value)
Recombination rate (cM/Mb)
SLFN14
RASL10B
HEATR9
CCL16
CCL3
CCL3L3
TBC1D3B
LOC107985033
GAS2L2
CCL5
CCL14
CCL4
CCL3L1
LINC02001
MMP28
LRRC37A8P
CCL18
CCL4L1
SNORD7
C17orf50
RDM1
CCL23
CCL4L2
PEX12
TAF15
LYZL6
TBC1D3H
AP2B1
CCL15−CCL14
TBC1D3G
CCL15
TBC1D3F
34 34.2 34.4 34.6 34.8
Position on chr17 (Mb)

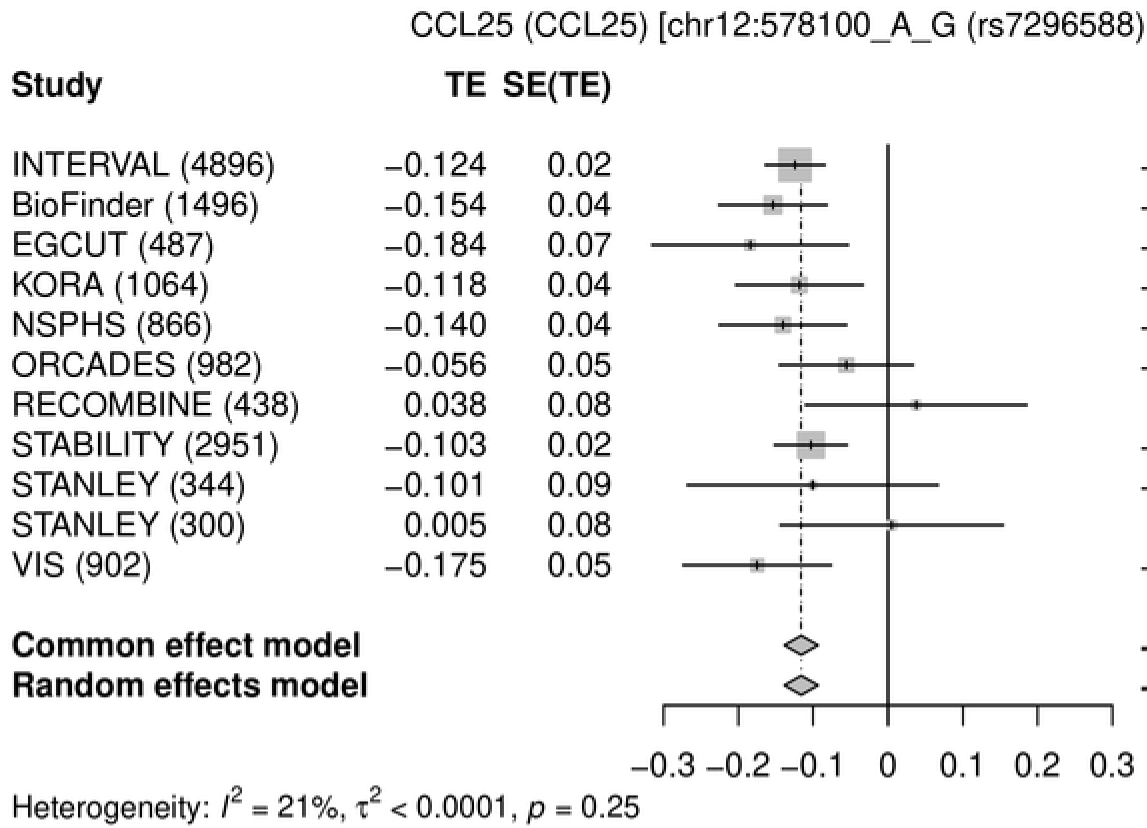
CCL25 (CCL25) [chr12:578100_A_G (rs7296588) (A/G) N=14726]
Study TE SE(TE) 95%−CI Weight (common) Weight (random)
INTERVAL (4896) −0.124 0.02 −0.12 [−0.16; −0.08] 32.5% 32.4%
BioFinder (1496) −0.154 0.04 −0.15 [−0.23; −0.08] 9.9% 9.9%
EGCUT (487) −0.184 0.07 −0.18 [−0.32; −0.05] 3.0% 3.0%
KORA (1064) −0.118 0.04 −0.12 [−0.20; −0.03] 7.2% 7.2%
NSPHS (866) −0.140 0.04 −0.14 [−0.23; −0.06] 7.2% 7.2%
ORCADES (982) −0.056 0.05 −0.06 [−0.15; 0.03] 6.5% 6.5%
RECOMBINE (438) 0.038 0.08 0.04 [−0.11; 0.19] 2.4% 2.4%
STABILITY (2951) −0.103 0.02 −0.10 [−0.15; −0.05] 21.9% 21.9%
STANLEY (344) −0.101 0.09 −0.10 [−0.27; 0.07] 1.8% 1.9%
STANLEY (300) 0.005 0.08 0.01 [−0.14; 0.15] 2.3% 2.4%
VIS (902) −0.175 0.05 −0.17 [−0.27; −0.08] 5.3% 5.3%
Common effect model −0.12 [−0.14; −0.09] 100.0% --
Random effects model −0.12 [−0.14; −0.09] -- 100.0%
−0.3 −0.2 −0.1 0 0.1 0.2 0.3
Heterogeneity: I² = 21%, τ² < 0.0001, p = 0.25
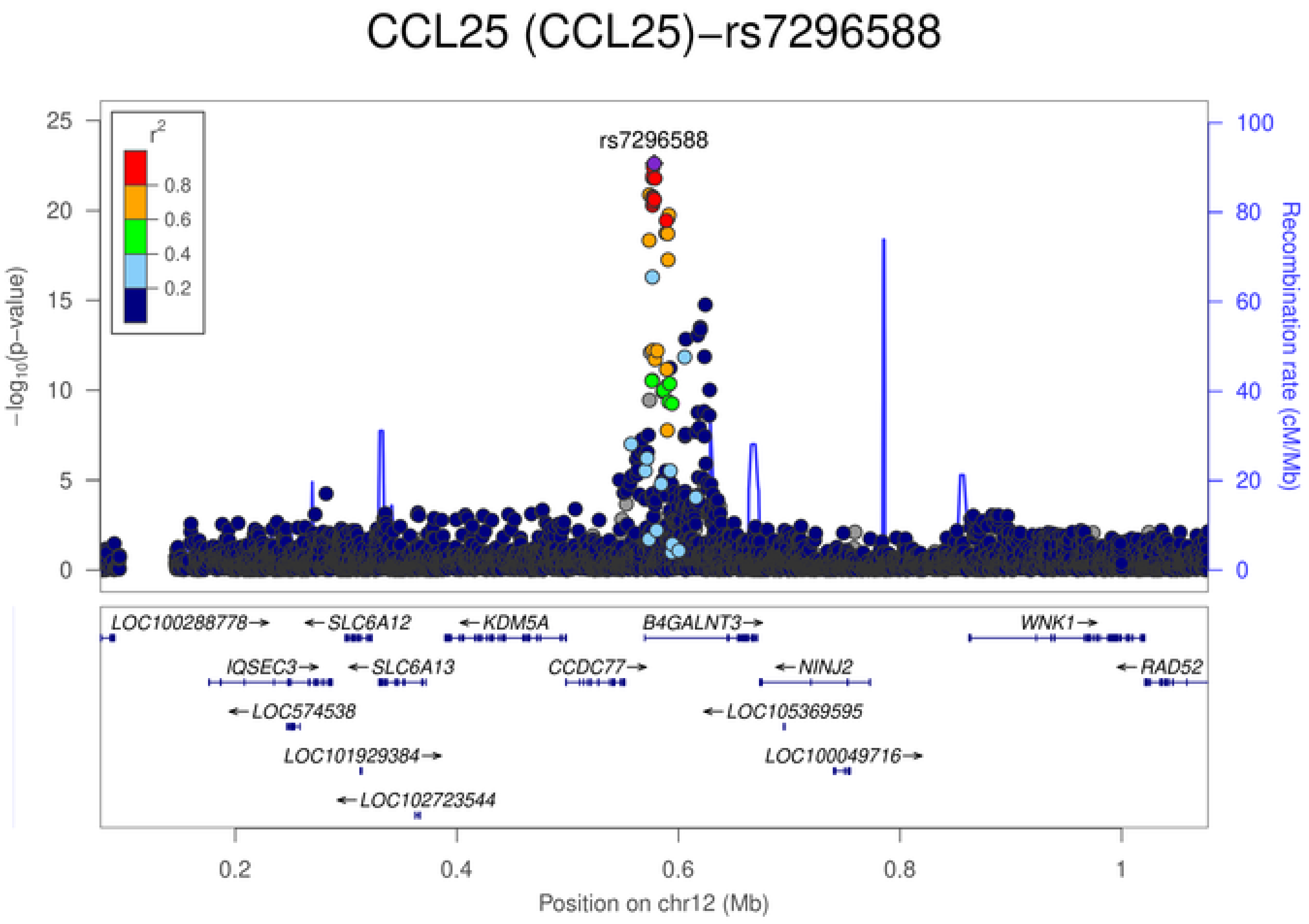
CCL25 (CCL25)−rs7296588
rs7296588
r²
0.8
0.6
0.4
0.2
−log10(p−value)
Recombination rate (cM/Mb)
LOC100288778
SLC6A12
KDM5A
B4GALNT3
WNK1
IQSEC3
SLC6A13
CCDC77
NINJ2
RAD52
LOC574538
LOC105369595
LOC101929384
LOC100049716
LOC102723544
Position on chr12 (Mb)

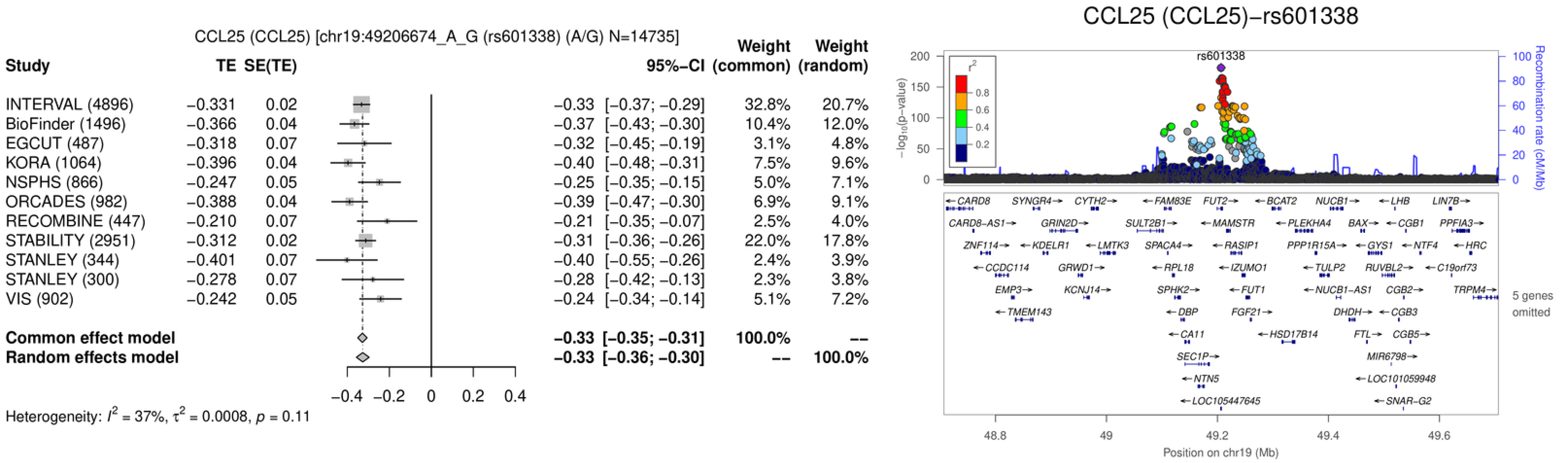

CCL25 (CCL25) [chr19:49206674_A_G (rs601338) (A/G) N=14735]
Study
TE
SE(TE)
95%−CI
Weight (common)
Weight (random)
INTERVAL (4896) −0.331 0.02 −0.33 [−0.37; −0.29] 32.8% 20.7%
BioFinder (1496) −0.366 0.04 −0.37 [−0.43; −0.30] 10.4% 12.0%
EGCUT (487) −0.318 0.07 −0.32 [−0.45; −0.19] 3.1% 4.8%
KORA (1064) −0.396 0.04 −0.40 [−0.48; −0.31] 7.5% 9.6%
NSPHS (866) −0.247 0.05 −0.25 [−0.35; −0.15] 5.0% 7.1%
ORCADES (982) −0.388 0.04 −0.39 [−0.47; −0.30] 6.9% 9.1%
RECOMBINE (447) −0.210 0.07 −0.21 [−0.35; −0.07] 2.5% 4.0%
STABILITY (2951) −0.312 0.02 −0.31 [−0.36; −0.26] 22.0% 17.8%
STANLEY (344) −0.401 0.07 −0.40 [−0.55; −0.26] 2.4% 3.9%
STANLEY (300) −0.278 0.07 −0.28 [−0.42; −0.13] 2.3% 3.8%
VIS (902) −0.242 0.05 −0.24 [−0.34; −0.14] 5.1% 7.2%
Common effect model −0.33 [−0.35; −0.31] 100.0% --
Random effects model −0.33 [−0.36; −0.30] -- 100.0%
−0.4 −0.2 0 0.2 0.4
Heterogeneity: $I^2$ = 37%, $\tau^2$ = 0.0008, $p$ = 0.11
CCL25 (CCL25)−rs601338
rs601338
$-\log_{10}$(p−value)
Recombination rate (cM/Mb)
$r^2$
0.8
0.6
0.4
0.2
CARD8 SYNGR4 CYTH2 FAM83E FUT2 BCAT2 NUCB1 LHB LIN7B
CARD8−AS1 GRIN2D SULT2B1 MAMSTR PLEKHA4 BAX CGB1 PPFIA3
ZNF114 KDELR1 LMTK3 SPACA4 RASIP1 PPP1R15A GYS1 NTF4 HRC
CCDC114 GRWD1 RPL18 IZUMO1 TULP2 RUVBL2 C19orf73
EMP3 KCNJ14 SPHK2 FUT1 NUCB1−AS1 CGB2 TRPM4
TMEM143 DBP FGF21 DHDH CGB3
CA11 HSD17B14 FTL CGB5
SEC1P MIR6798
NTN5 LOC101059948
LOC105447645 SNAR−G2
5 genes omitted
48.8 49 49.2 49.4 49.6
Position on chr19 (Mb)

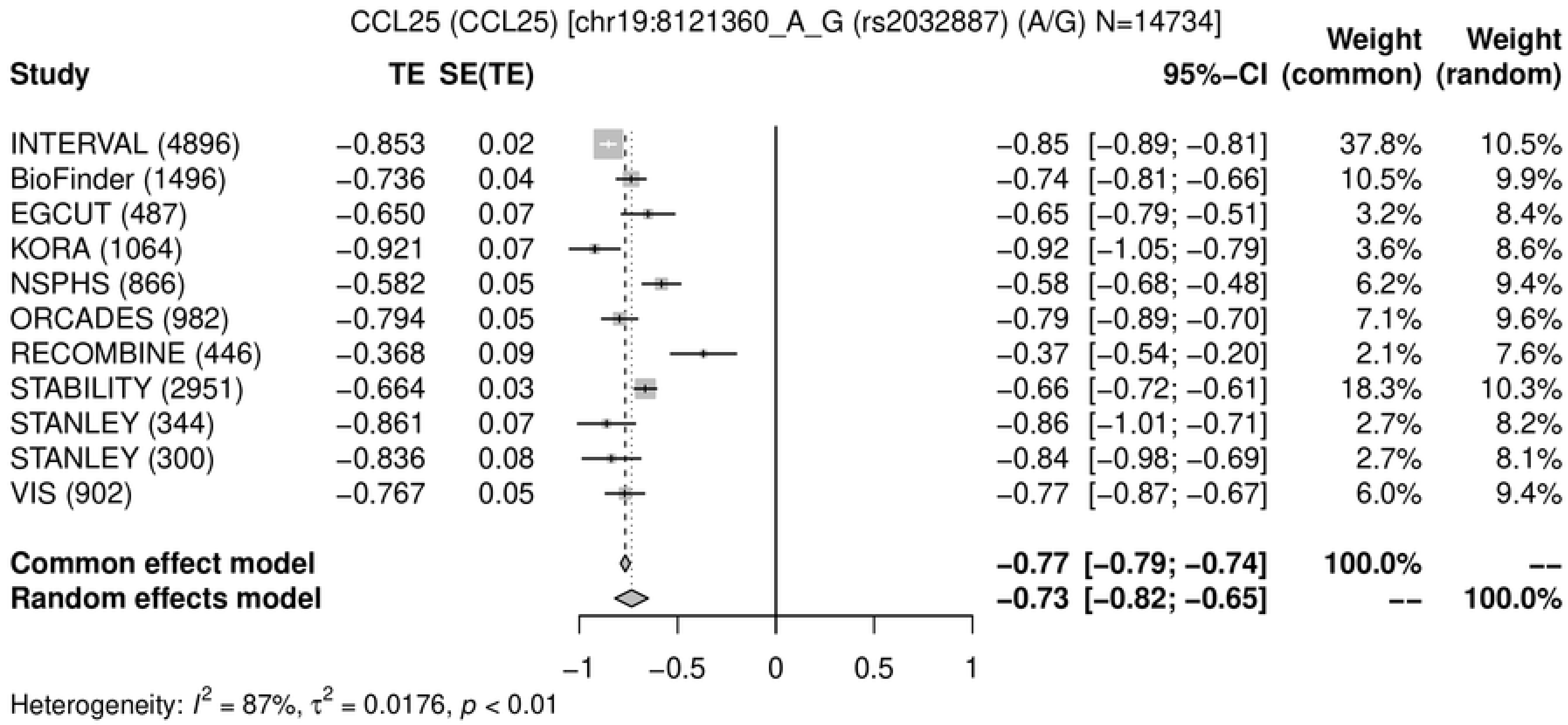
CCL25 (CCL25) [chr19:8121360_A_G (rs2032887) (A/G) N=14734]
Study
TE
SE(TE)
95%−CI
Weight (common)
Weight (random)
INTERVAL (4896) −0.853 0.02 −0.85 [−0.89; −0.81] 37.8% 10.5%
BioFinder (1496) −0.736 0.04 −0.74 [−0.81; −0.66] 10.5% 9.9%
EGCUT (487) −0.650 0.07 −0.65 [−0.79; −0.51] 3.2% 8.4%
KORA (1064) −0.921 0.07 −0.92 [−1.05; −0.79] 3.6% 8.6%
NSPHS (866) −0.582 0.05 −0.58 [−0.68; −0.48] 6.2% 9.4%
ORCADES (982) −0.794 0.05 −0.79 [−0.89; −0.70] 7.1% 9.6%
RECOMBINE (446) −0.368 0.09 −0.37 [−0.54; −0.20] 2.1% 7.6%
STABILITY (2951) −0.664 0.03 −0.66 [−0.72; −0.61] 18.3% 10.3%
STANLEY (344) −0.861 0.07 −0.86 [−1.01; −0.71] 2.7% 8.2%
STANLEY (300) −0.836 0.08 −0.84 [−0.98; −0.69] 2.7% 8.1%
VIS (902) −0.767 0.05 −0.77 [−0.87; −0.67] 6.0% 9.4%
Common effect model −0.77 [−0.79; −0.74] 100.0% --
Random effects model −0.73 [−0.82; −0.65] -- 100.0%
−1 −0.5 0 0.5 1
Heterogeneity: $I^2$ = 87%, $\tau^2$ = 0.0176, $p$ < 0.01

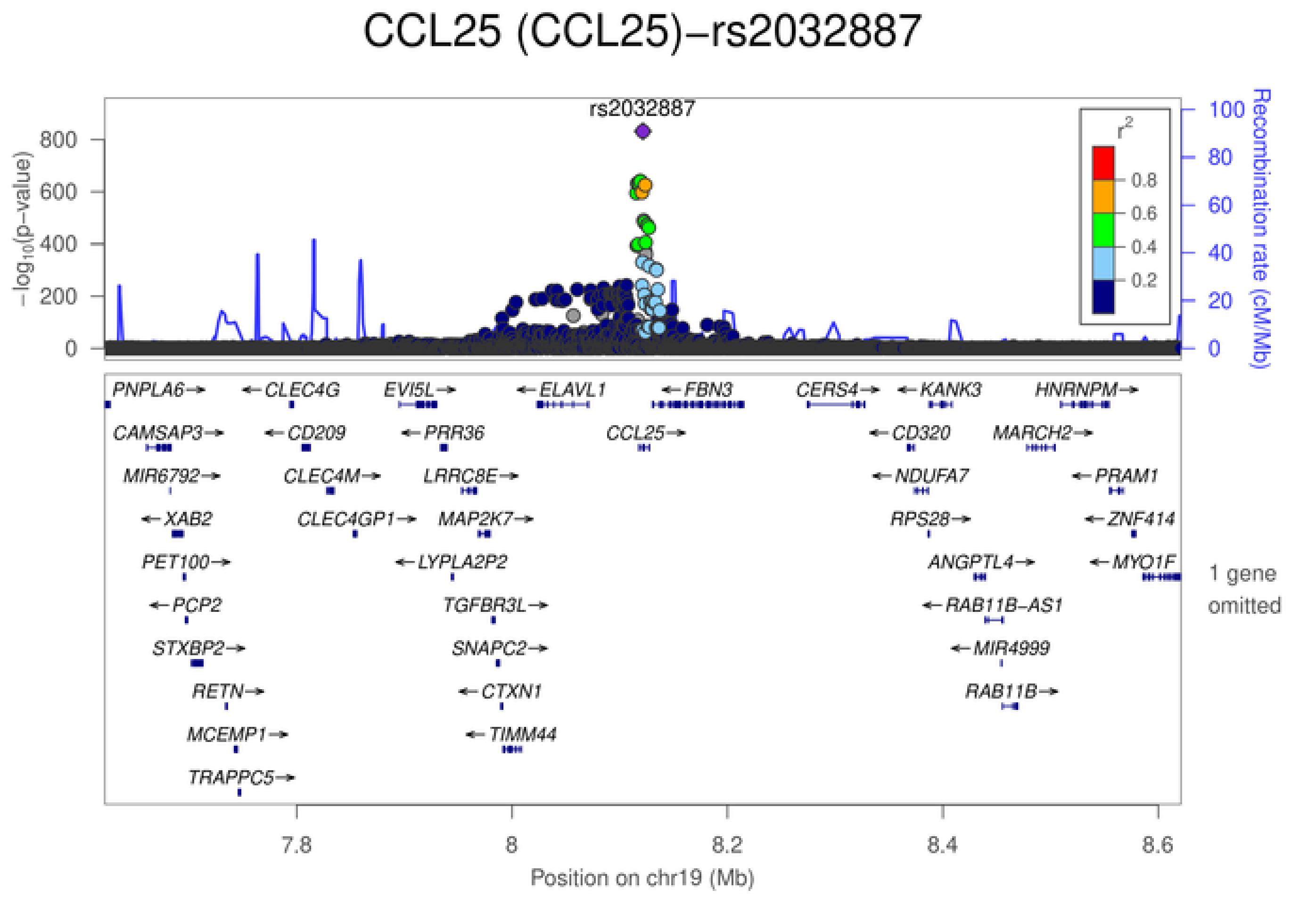
CCL25 (CCL25)−rs2032887
rs2032887
$-\log_{10}$(p−value)
800
600
400
200
0
Recombination rate (cM/Mb)
100
80
60
40
20
0
$r^2$
0.8
0.6
0.4
0.2
PNPLA6→ ←CLEC4G EVI5L→ ←ELAVL1 ←FBN3 CERS4→ ←KANK3 HNRNPM→
CAMSAP3→ ←CD209 ←PRR36 CCL25→ ←CD320 MARCH2→
MIR6792→ CLEC4M→ LRRC8E→ ←NDUFA7 ←PRAM1
←XAB2 CLEC4GP1→ MAP2K7→ RPS28→ ←ZNF414
PET100→ ←LYPLA2P2 ANGPTL4→ ←MYO1F
←PCP2 TGFBR3L→ ←RAB11B−AS1
STXBP2→ SNAPC2→ ←MIR4999
RETN→ ←CTXN1 RAB11B→
MCEMP1→ ←TIMM44
TRAPPC5→
1 gene omitted
7.8
8
8.2
8.4
8.6
Position on chr19 (Mb)

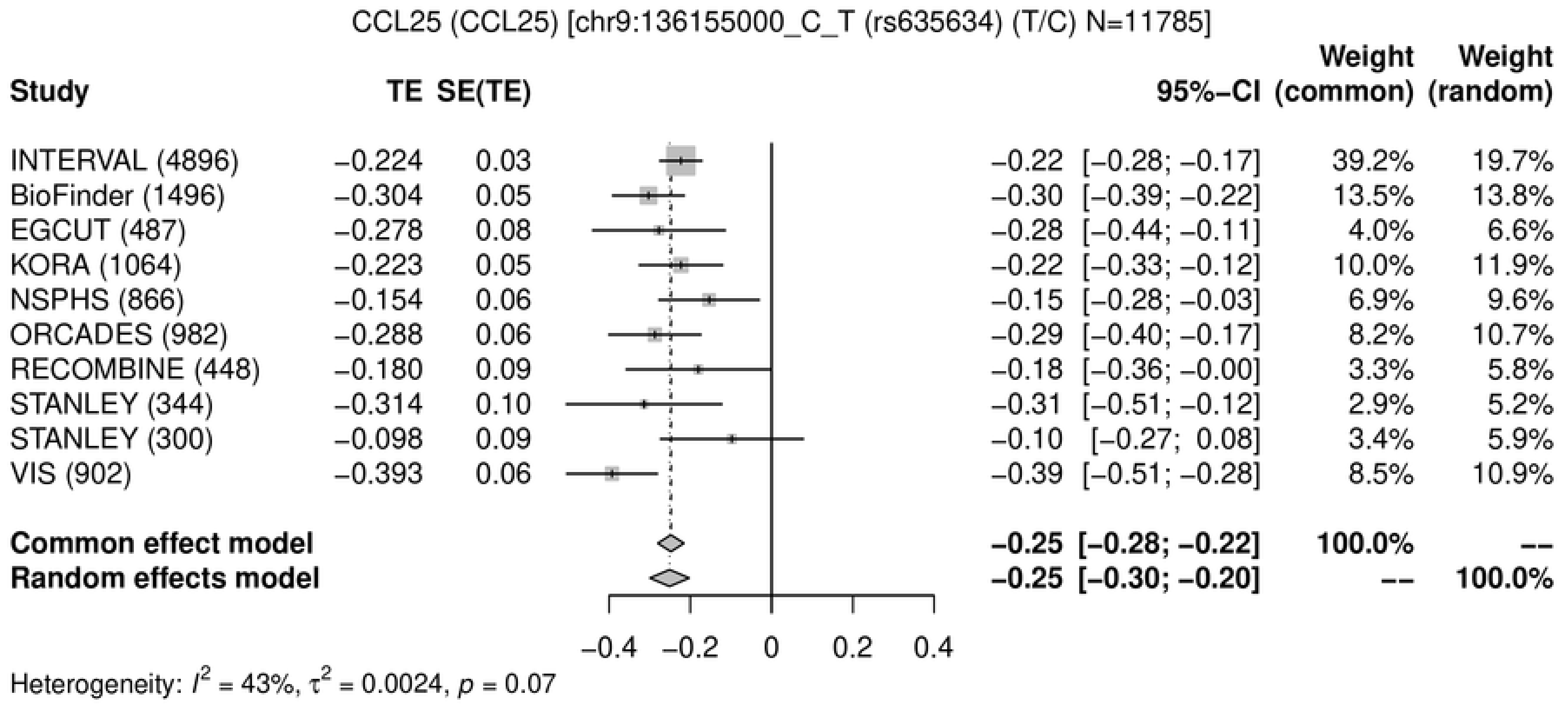

CCL25 (CCL25) [chr9:136155000_C_T (rs635634) (T/C) N=11785]
Study
TE
SE(TE)
95%-CI
Weight (common)
Weight (random)
INTERVAL (4896) −0.224 0.03 −0.22 [−0.28; −0.17] 39.2% 19.7%
BioFinder (1496) −0.304 0.05 −0.30 [−0.39; −0.22] 13.5% 13.8%
EGCUT (487) −0.278 0.08 −0.28 [−0.44; −0.11] 4.0% 6.6%
KORA (1064) −0.223 0.05 −0.22 [−0.33; −0.12] 10.0% 11.9%
NSPHS (866) −0.154 0.06 −0.15 [−0.28; −0.03] 6.9% 9.6%
ORCADES (982) −0.288 0.06 −0.29 [−0.40; −0.17] 8.2% 10.7%
RECOMBINE (448) −0.180 0.09 −0.18 [−0.36; −0.00] 3.3% 5.8%
STANLEY (344) −0.314 0.10 −0.31 [−0.51; −0.12] 2.9% 5.2%
STANLEY (300) −0.098 0.09 −0.10 [−0.27; 0.08] 3.4% 5.9%
VIS (902) −0.393 0.06 −0.39 [−0.51; −0.28] 8.5% 10.9%
Common effect model −0.25 [−0.28; −0.22] 100.0% --
Random effects model −0.25 [−0.30; −0.20] -- 100.0%
−0.4 −0.2 0 0.2 0.4
Heterogeneity: $I^2$ = 43%, $\tau^2$ = 0.0024, $p$ = 0.07


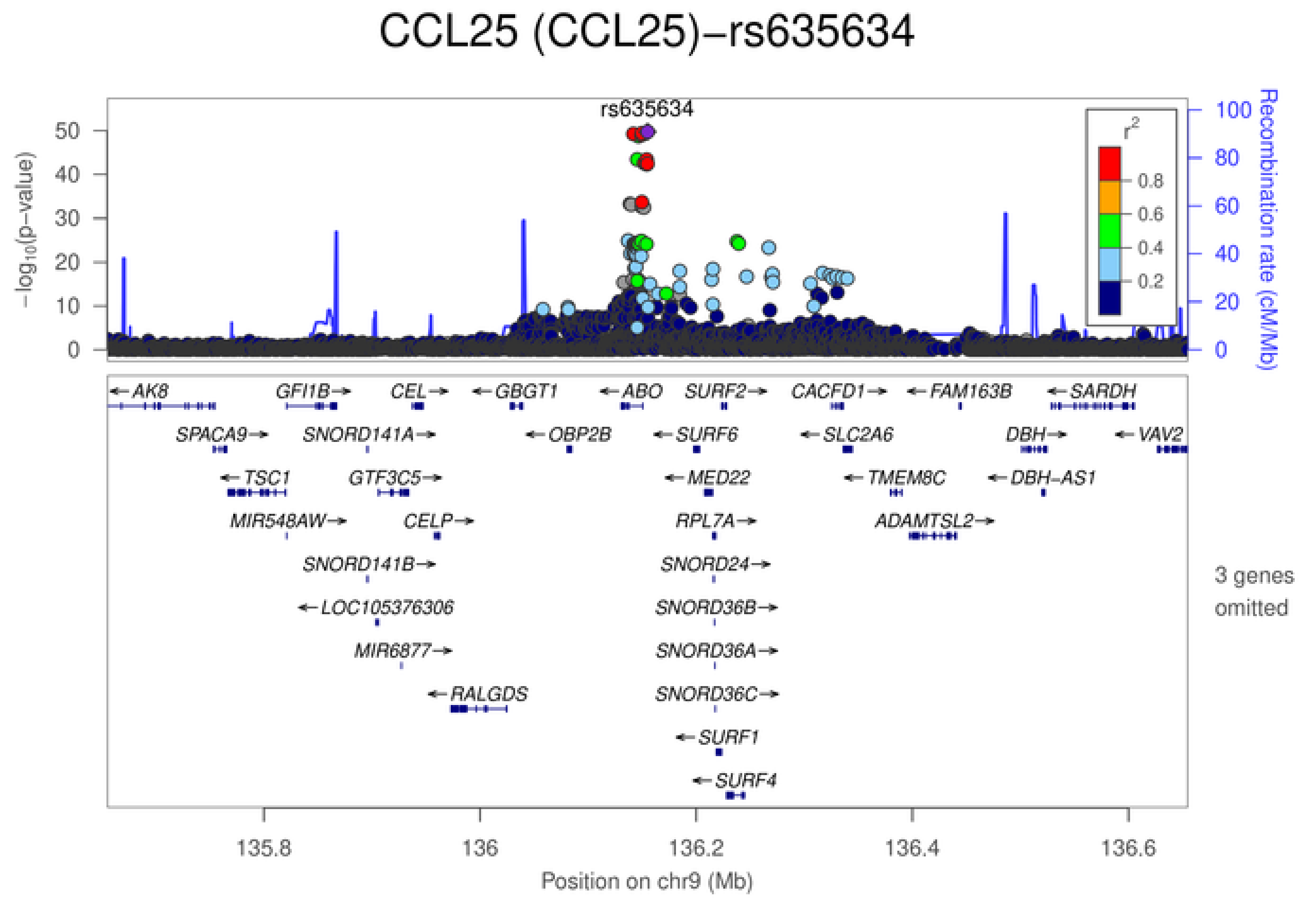

CCL25 (CCL25)−rs635634
rs635634
$-\log_{10}$(p−value)
Recombination rate (cM/Mb)
$r^2$
0.8
0.6
0.4
0.2
AK8
GFI1B
CEL
GBGT1
ABO
SURF2
CACFD1
FAM163B
SARDH
SPACA9
SNORD141A
OBP2B
SURF6
SLC2A6
DBH
VAV2
TSC1
GTF3C5
MED22
TMEM8C
DBH-AS1
MIR548AW
CELP
RPL7A
ADAMTSL2
SNORD141B
SNORD24
LOC105376306
SNORD36B
MIR6877
SNORD36A
RALGDS
SNORD36C
SURF1
SURF4
3 genes omitted
135.8 136 136.2 136.4 136.6
Position on chr9 (Mb)

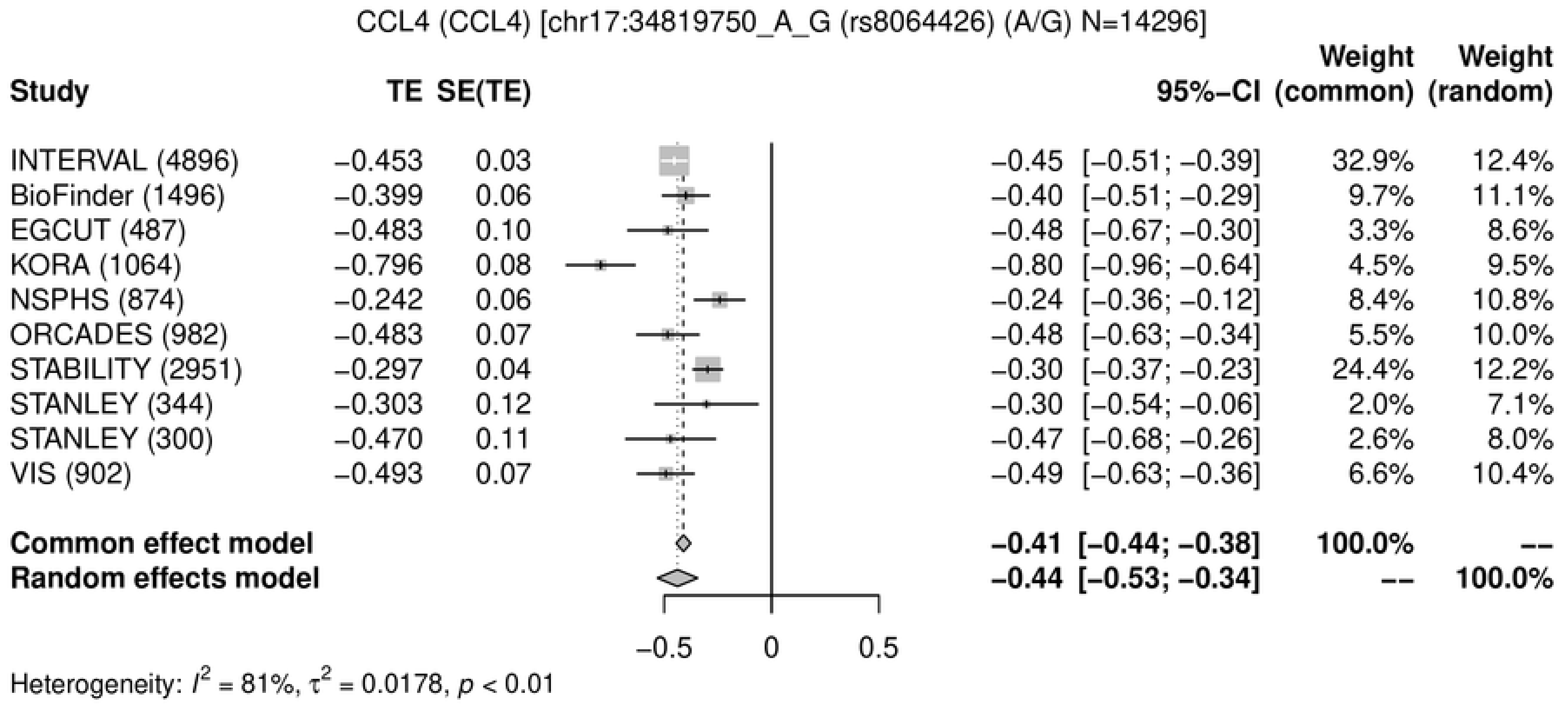
CCL4 (CCL4) [chr17:34819750_A_G (rs8064426) (A/G) N=14296]
Study
TE
SE(TE)
95%−CI
Weight (common)
Weight (random)
INTERVAL (4896) −0.453 0.03 −0.45 [−0.51; −0.39] 32.9% 12.4%
BioFinder (1496) −0.399 0.06 −0.40 [−0.51; −0.29] 9.7% 11.1%
EGCUT (487) −0.483 0.10 −0.48 [−0.67; −0.30] 3.3% 8.6%
KORA (1064) −0.796 0.08 −0.80 [−0.96; −0.64] 4.5% 9.5%
NSPHS (874) −0.242 0.06 −0.24 [−0.36; −0.12] 8.4% 10.8%
ORCADES (982) −0.483 0.07 −0.48 [−0.63; −0.34] 5.5% 10.0%
STABILITY (2951) −0.297 0.04 −0.30 [−0.37; −0.23] 24.4% 12.2%
STANLEY (344) −0.303 0.12 −0.30 [−0.54; −0.06] 2.0% 7.1%
STANLEY (300) −0.470 0.11 −0.47 [−0.68; −0.26] 2.6% 8.0%
VIS (902) −0.493 0.07 −0.49 [−0.63; −0.36] 6.6% 10.4%
Common effect model −0.41 [−0.44; −0.38] 100.0% --
Random effects model −0.44 [−0.53; −0.34] -- 100.0%
−0.5
0
0.5
Heterogeneity: $I^2$ = 81%, $\tau^2$ = 0.0178, $p$ < 0.01

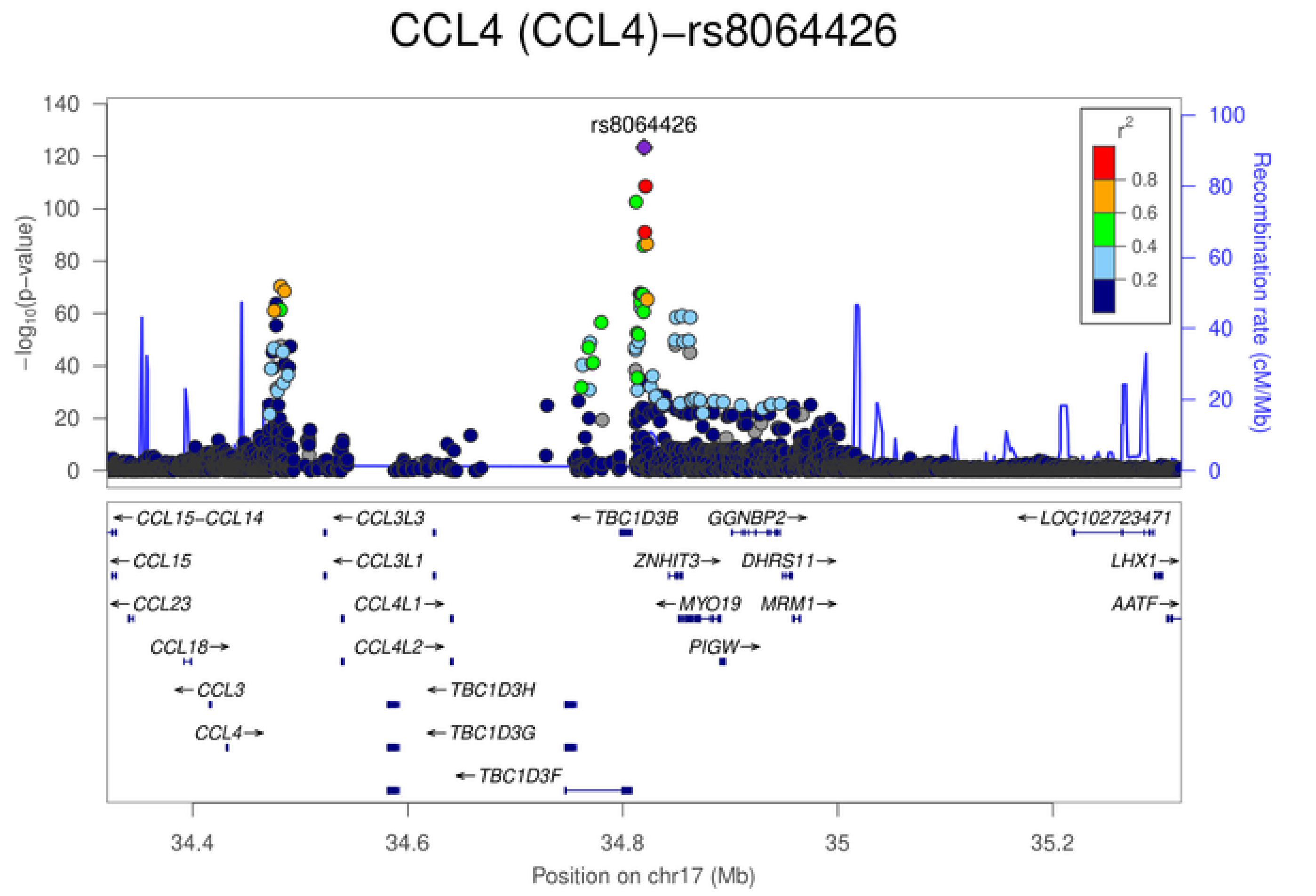
CCL4 (CCL4)−rs8064426
rs8064426
$-\log_{10}$(p−value)
Recombination rate (cM/Mb)
$r^2$
0.8
0.6
0.4
0.2
CCL15−CCL14
CCL15
CCL23
CCL18
CCL3
CCL4
CCL3L3
CCL3L1
CCL4L1
CCL4L2
TBC1D3H
TBC1D3G
TBC1D3F
TBC1D3B
GGNBP2
ZNHIT3
DHRS11
MYO19
MRM1
PIGW
LOC102723471
LHX1
AATF
34.4
34.6
34.8
35
35.2
Position on chr17 (Mb)

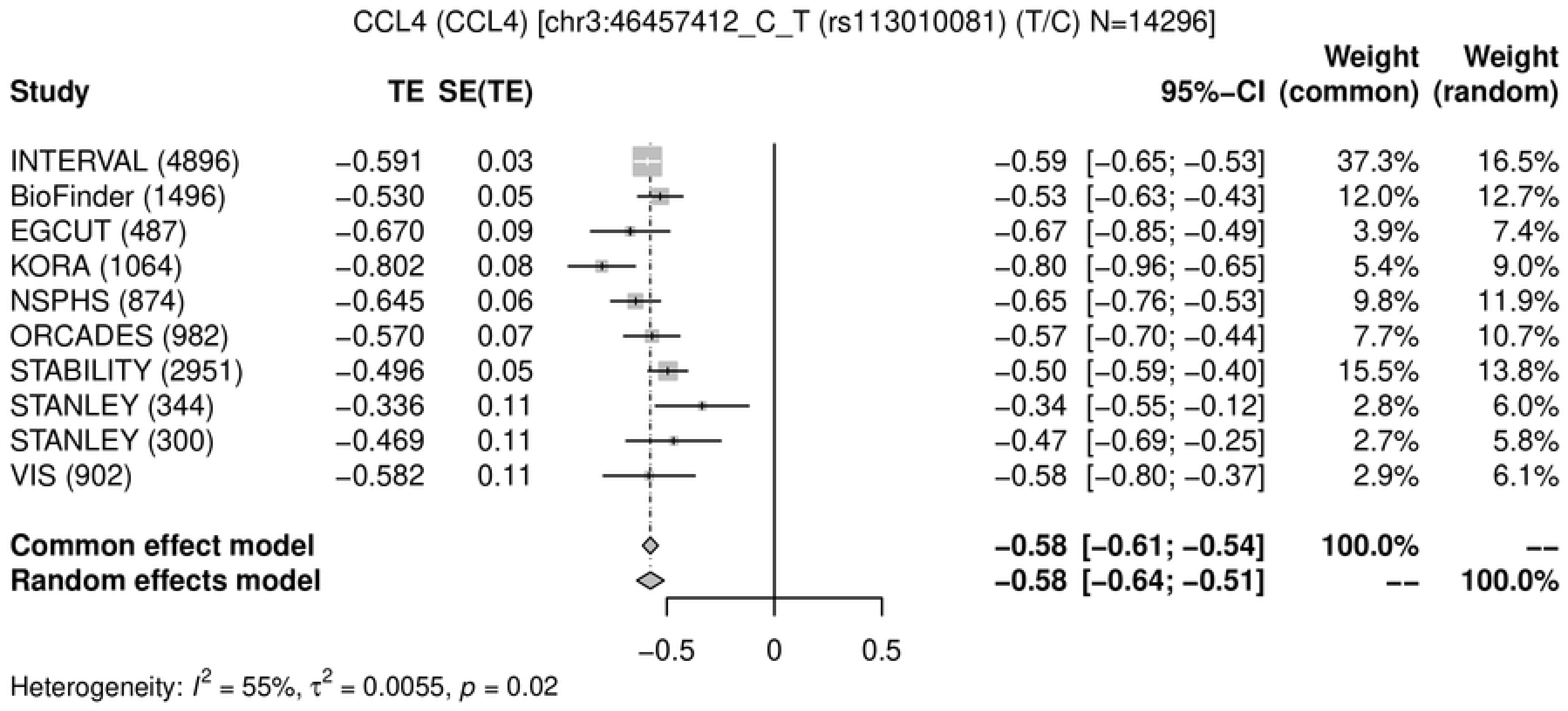
CCL4 (CCL4) [chr3:46457412_C_T (rs113010081) (T/C) N=14296]
Study
TE
SE(TE)
95%−CI
Weight (common)
Weight (random)
INTERVAL (4896) −0.591 0.03 −0.59 [−0.65; −0.53] 37.3% 16.5%
BioFinder (1496) −0.530 0.05 −0.53 [−0.63; −0.43] 12.0% 12.7%
EGCUT (487) −0.670 0.09 −0.67 [−0.85; −0.49] 3.9% 7.4%
KORA (1064) −0.802 0.08 −0.80 [−0.96; −0.65] 5.4% 9.0%
NSPHS (874) −0.645 0.06 −0.65 [−0.76; −0.53] 9.8% 11.9%
ORCADES (982) −0.570 0.07 −0.57 [−0.70; −0.44] 7.7% 10.7%
STABILITY (2951) −0.496 0.05 −0.50 [−0.59; −0.40] 15.5% 13.8%
STANLEY (344) −0.336 0.11 −0.34 [−0.55; −0.12] 2.8% 6.0%
STANLEY (300) −0.469 0.11 −0.47 [−0.69; −0.25] 2.7% 5.8%
VIS (902) −0.582 0.11 −0.58 [−0.80; −0.37] 2.9% 6.1%
Common effect model −0.58 [−0.61; −0.54] 100.0% --
Random effects model −0.58 [−0.64; −0.51] -- 100.0%
−0.5
0
0.5
Heterogeneity: $I^2$ = 55%, $\tau^2$ = 0.0055, $p$ = 0.02

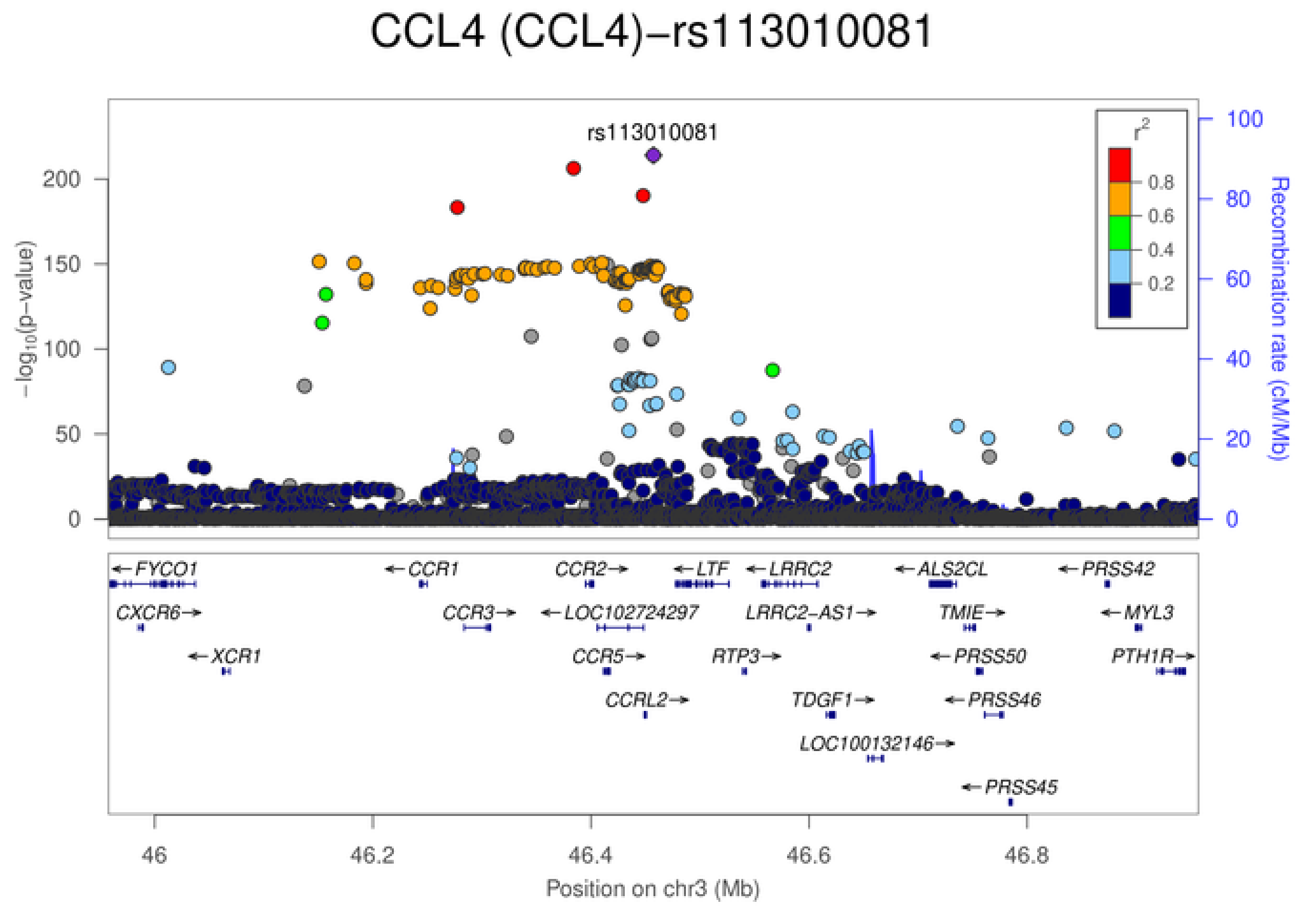
CCL4 (CCL4)−rs113010081
rs113010081
$-\log_{10}$(p-value)
200
150
100
50
0
$r^2$
0.8
0.6
0.4
0.2
100
80
60
40
20
0
Recombination rate (cM/Mb)
FYCO1
CXCR6
XCR1
CCR1
CCR3
CCR2
LOC102724297
CCR5
CCRL2
LTF
LRRC2
LRRC2-AS1
RTP3
TDGF1
LOC100132146
ALS2CL
TMIE
PRSS50
PRSS46
PRSS45
PRSS42
MYL3
PTH1R
46
46.2
46.4
46.6
46.8
Position on chr3 (Mb)

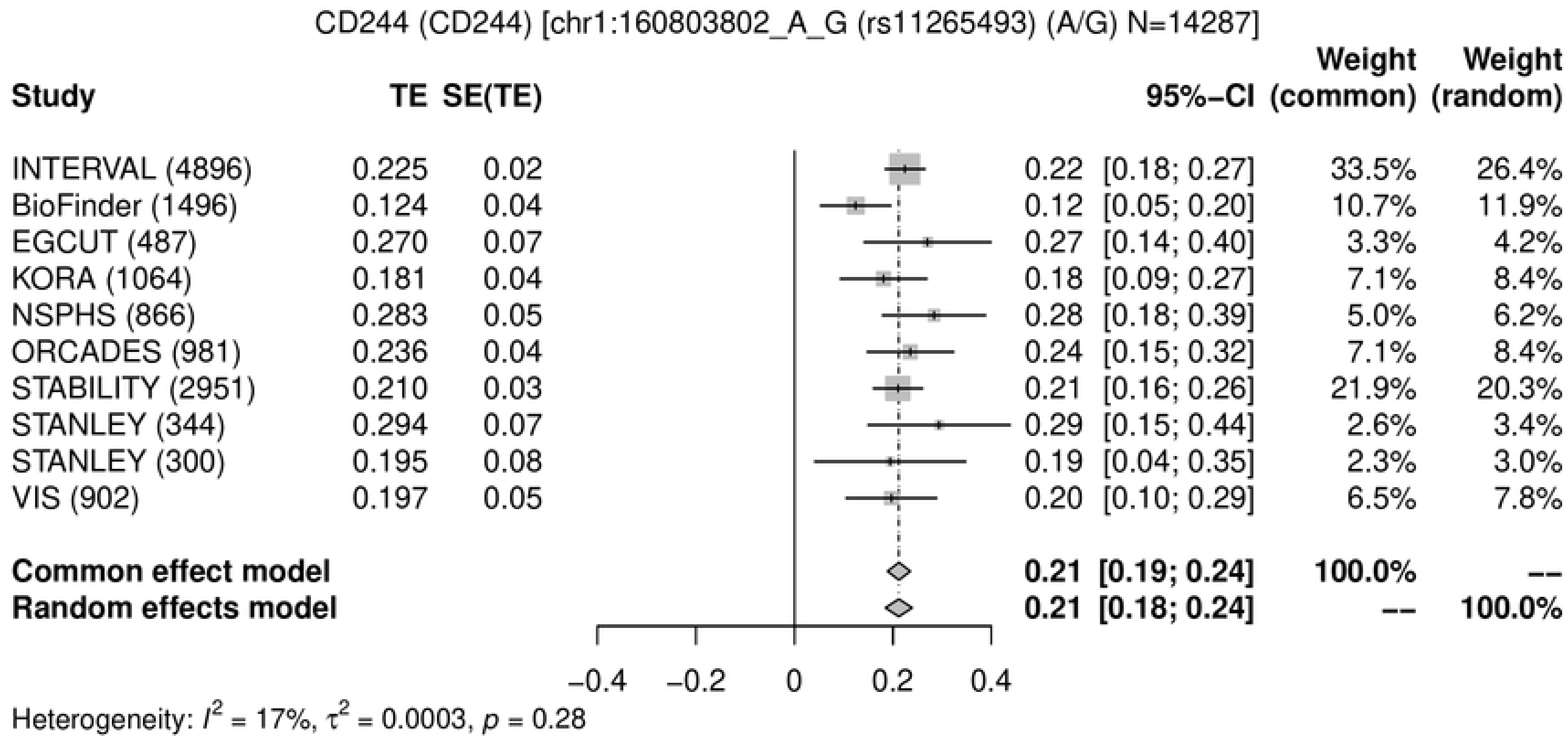
CD244 (CD244) [chr1:160803802_A_G (rs11265493) (A/G) N=14287]
Study
TE
SE(TE)
95%−CI
Weight (common)
Weight (random)
INTERVAL (4896) 0.225 0.02 0.22 [0.18; 0.27] 33.5% 26.4%
BioFinder (1496) 0.124 0.04 0.12 [0.05; 0.20] 10.7% 11.9%
EGCUT (487) 0.270 0.07 0.27 [0.14; 0.40] 3.3% 4.2%
KORA (1064) 0.181 0.04 0.18 [0.09; 0.27] 7.1% 8.4%
NSPHS (866) 0.283 0.05 0.28 [0.18; 0.39] 5.0% 6.2%
ORCADES (981) 0.236 0.04 0.24 [0.15; 0.32] 7.1% 8.4%
STABILITY (2951) 0.210 0.03 0.21 [0.16; 0.26] 21.9% 20.3%
STANLEY (344) 0.294 0.07 0.29 [0.15; 0.44] 2.6% 3.4%
STANLEY (300) 0.195 0.08 0.19 [0.04; 0.35] 2.3% 3.0%
VIS (902) 0.197 0.05 0.20 [0.10; 0.29] 6.5% 7.8%
Common effect model 0.21 [0.19; 0.24] 100.0% --
Random effects model 0.21 [0.18; 0.24] -- 100.0%
−0.4 −0.2 0 0.2 0.4
Heterogeneity: $I^2$ = 17%, $\tau^2$ = 0.0003, $p$ = 0.28

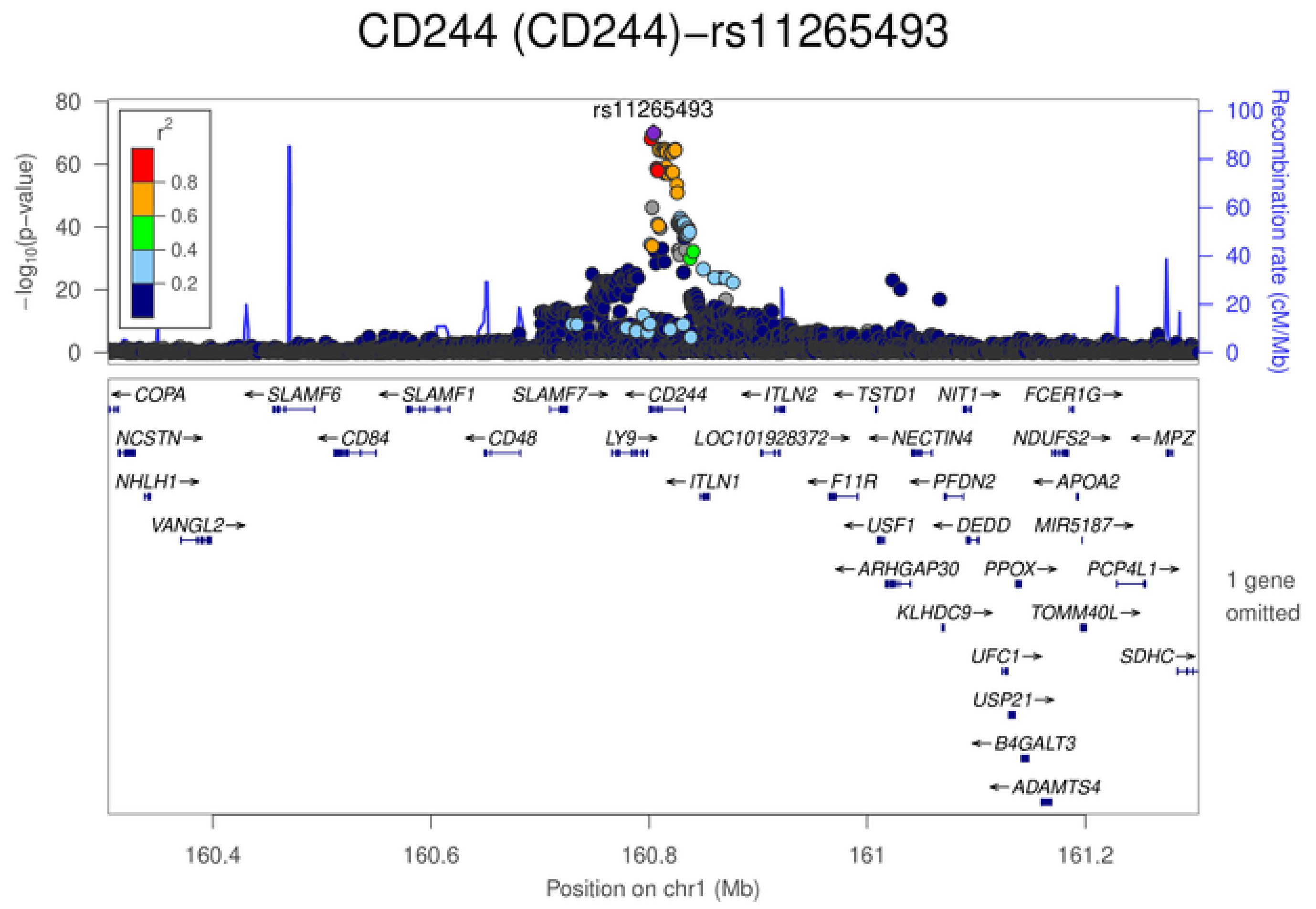
CD244 (CD244)−rs11265493
rs11265493
$-\log_{10}$(p−value)
Recombination rate (cM/Mb)
$r^2$
0.8
0.6
0.4
0.2
0 20 40 60 80
0 20 40 60 80 100
COPA
SLAMF6
SLAMF1
SLAMF7
CD244
ITLN2
TSTD1
NIT1
FCER1G
NCSTN
CD84
CD48
LY9
LOC101928372
NECTIN4
NDUFS2
MPZ
NHLH1
ITLN1
F11R
PFDN2
APOA2
VANGL2
USF1
DEDD
MIR5187
ARHGAP30
PPOX
PCP4L1
KLHDC9
TOMM40L
UFC1
SDHC
USP21
B4GALT3
ADAMTS4
1 gene omitted
160.4 160.6 160.8 161 161.2
Position on chr1 (Mb)

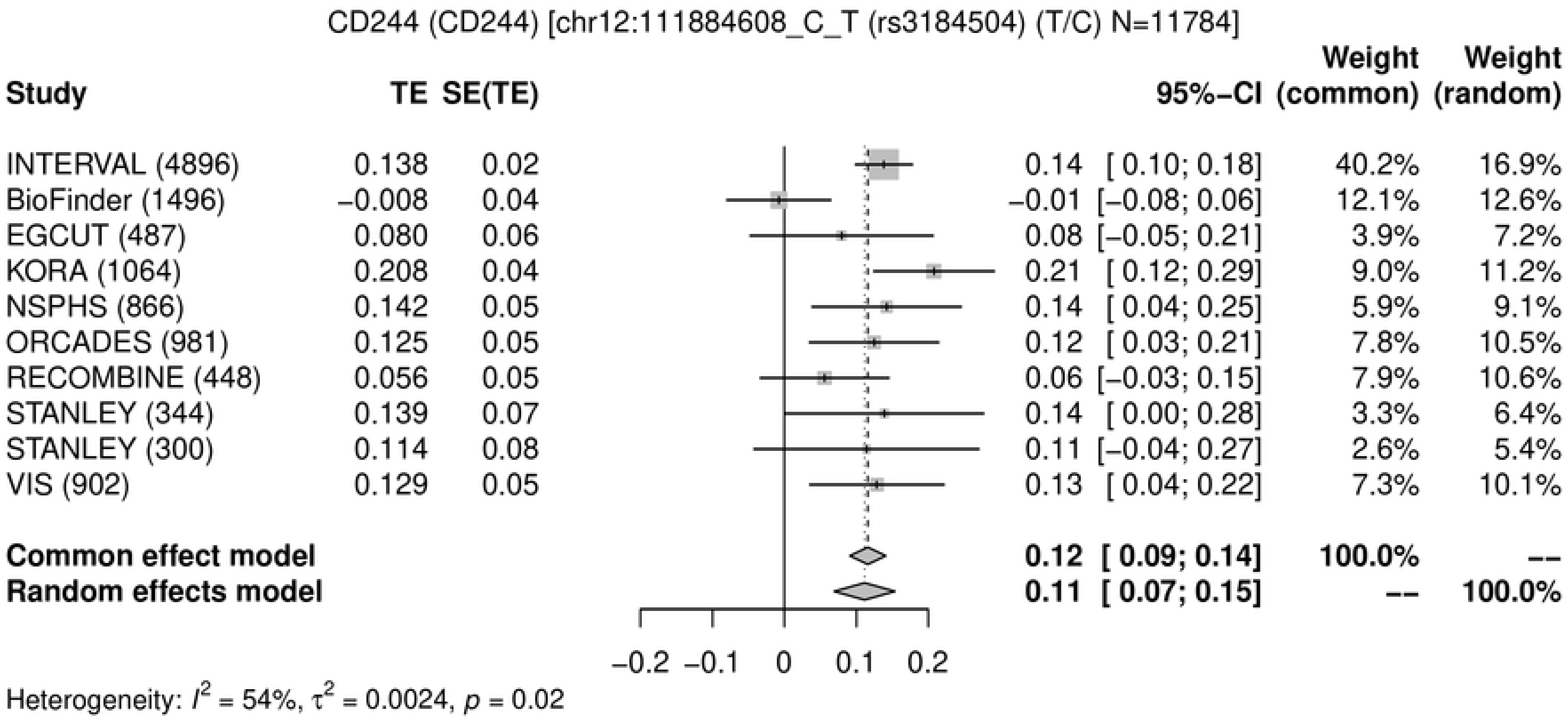
CD244 (CD244) [chr12:111884608_C_T (rs3184504) (T/C) N=11784]
Study
TE
SE(TE)
95%−CI
Weight (common)
Weight (random)
INTERVAL (4896) 0.138 0.02 0.14 [ 0.10; 0.18] 40.2% 16.9%
BioFinder (1496) −0.008 0.04 −0.01 [−0.08; 0.06] 12.1% 12.6%
EGCUT (487) 0.080 0.06 0.08 [−0.05; 0.21] 3.9% 7.2%
KORA (1064) 0.208 0.04 0.21 [ 0.12; 0.29] 9.0% 11.2%
NSPHS (866) 0.142 0.05 0.14 [ 0.04; 0.25] 5.9% 9.1%
ORCADES (981) 0.125 0.05 0.12 [ 0.03; 0.21] 7.8% 10.5%
RECOMBINE (448) 0.056 0.05 0.06 [−0.03; 0.15] 7.9% 10.6%
STANLEY (344) 0.139 0.07 0.14 [ 0.00; 0.28] 3.3% 6.4%
STANLEY (300) 0.114 0.08 0.11 [−0.04; 0.27] 2.6% 5.4%
VIS (902) 0.129 0.05 0.13 [ 0.04; 0.22] 7.3% 10.1%
Common effect model 0.12 [ 0.09; 0.14] 100.0% --
Random effects model 0.11 [ 0.07; 0.15] -- 100.0%
−0.2 −0.1 0 0.1 0.2
Heterogeneity: $I^2$ = 54%, $\tau^2$ = 0.0024, $p$ = 0.02

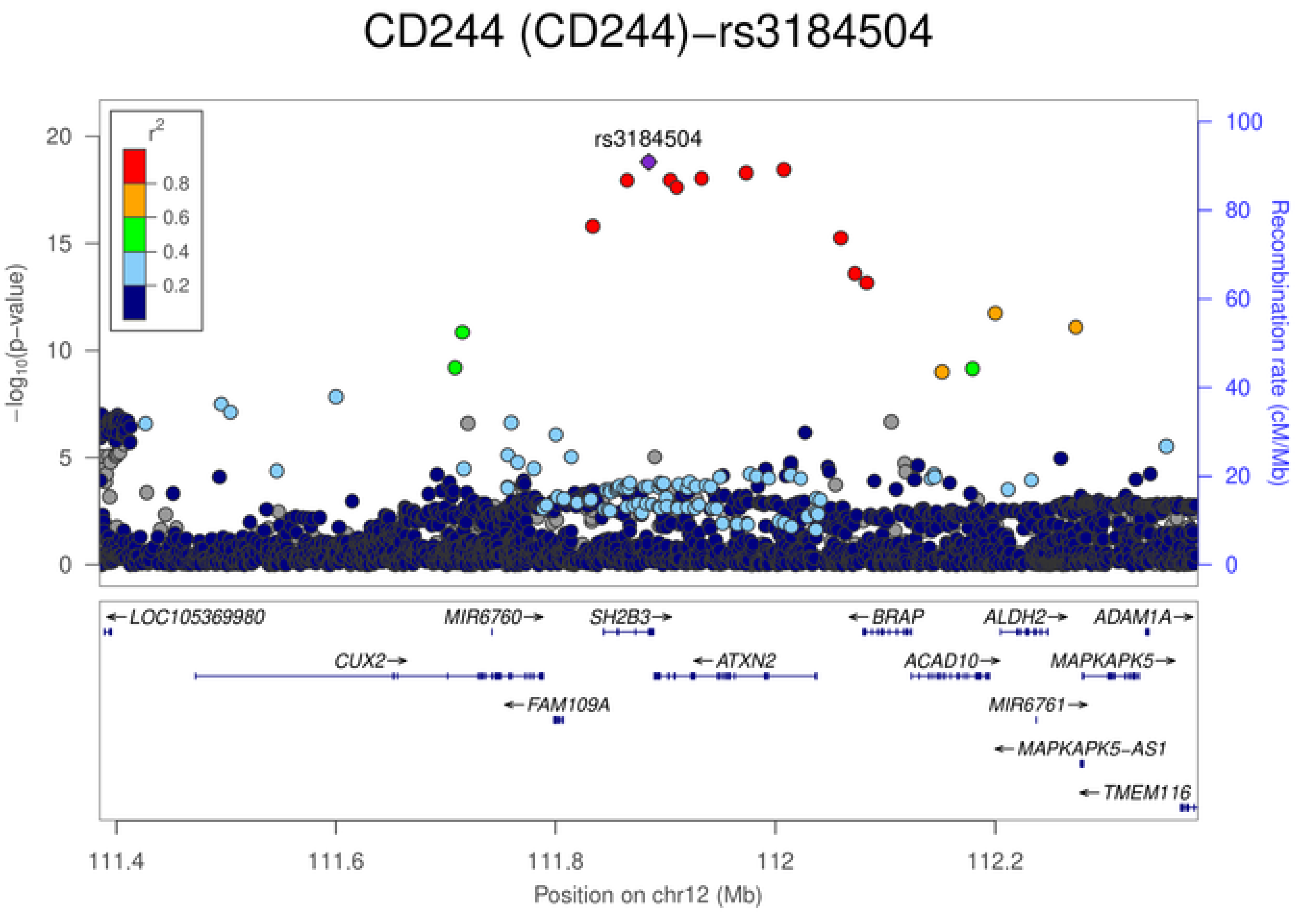
CD244 (CD244)−rs3184504
rs3184504
$r^2$
0.8
0.6
0.4
0.2
$-\log_{10}$(p−value)
Recombination rate (cM/Mb)
LOC105369980
MIR6760
SH2B3
BRAP
ALDH2
ADAM1A
CUX2
ATXN2
ACAD10
MAPKAPK5
FAM109A
MIR6761
MAPKAPK5−AS1
TMEM116
111.4
111.6
111.8
112
112.2
Position on chr12 (Mb)

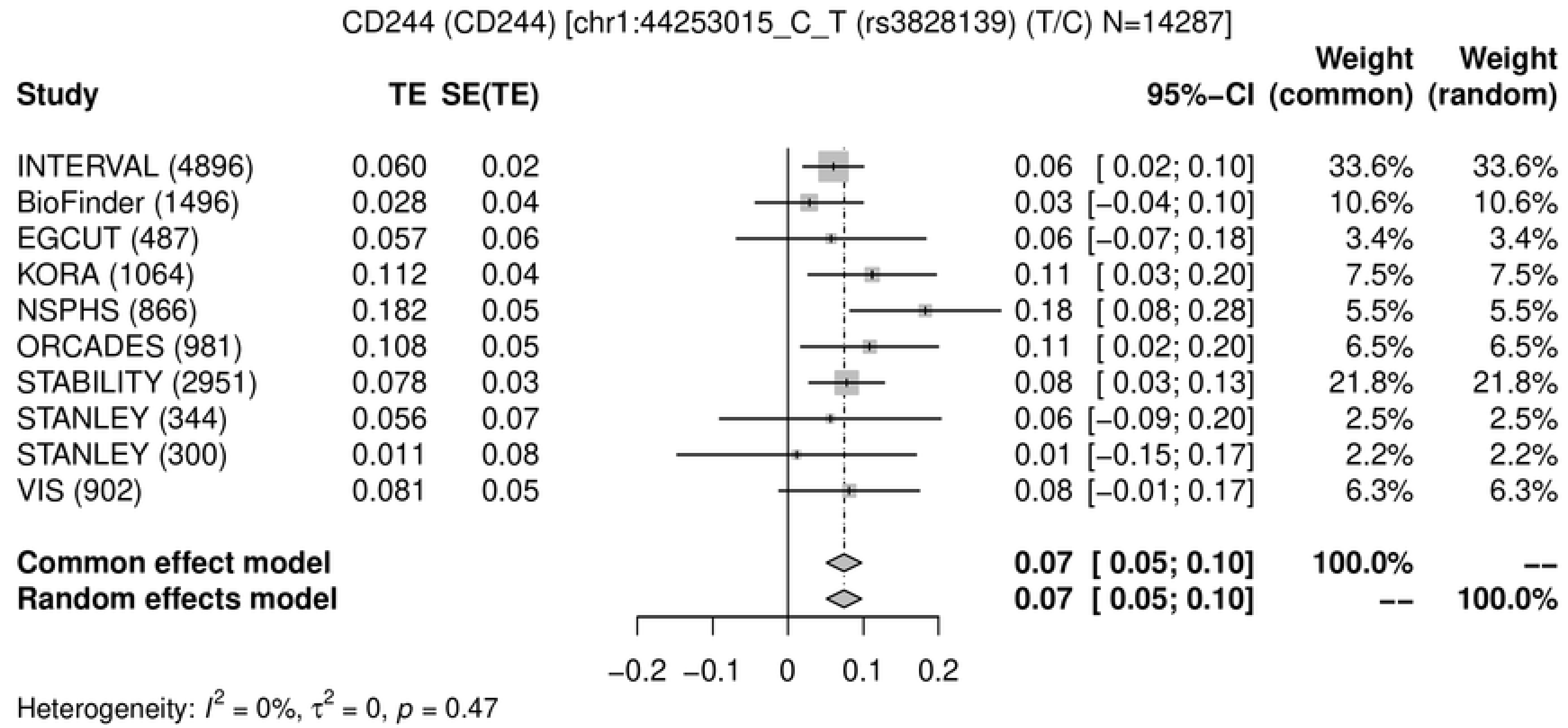
CD244 (CD244) [chr1:44253015_C_T (rs3828139) (T/C) N=14287]
Study
TE
SE(TE)
95%–CI
Weight (common)
Weight (random)
INTERVAL (4896) 0.060 0.02 0.06 [ 0.02; 0.10] 33.6% 33.6%
BioFinder (1496) 0.028 0.04 0.03 [−0.04; 0.10] 10.6% 10.6%
EGCUT (487) 0.057 0.06 0.06 [−0.07; 0.18] 3.4% 3.4%
KORA (1064) 0.112 0.04 0.11 [ 0.03; 0.20] 7.5% 7.5%
NSPHS (866) 0.182 0.05 0.18 [ 0.08; 0.28] 5.5% 5.5%
ORCADES (981) 0.108 0.05 0.11 [ 0.02; 0.20] 6.5% 6.5%
STABILITY (2951) 0.078 0.03 0.08 [ 0.03; 0.13] 21.8% 21.8%
STANLEY (344) 0.056 0.07 0.06 [−0.09; 0.20] 2.5% 2.5%
STANLEY (300) 0.011 0.08 0.01 [−0.15; 0.17] 2.2% 2.2%
VIS (902) 0.081 0.05 0.08 [−0.01; 0.17] 6.3% 6.3%
Common effect model 0.07 [ 0.05; 0.10] 100.0% --
Random effects model 0.07 [ 0.05; 0.10] -- 100.0%
−0.2 −0.1 0 0.1 0.2
Heterogeneity: $I^2$ = 0%, $\tau^2$ = 0, $p$ = 0.47

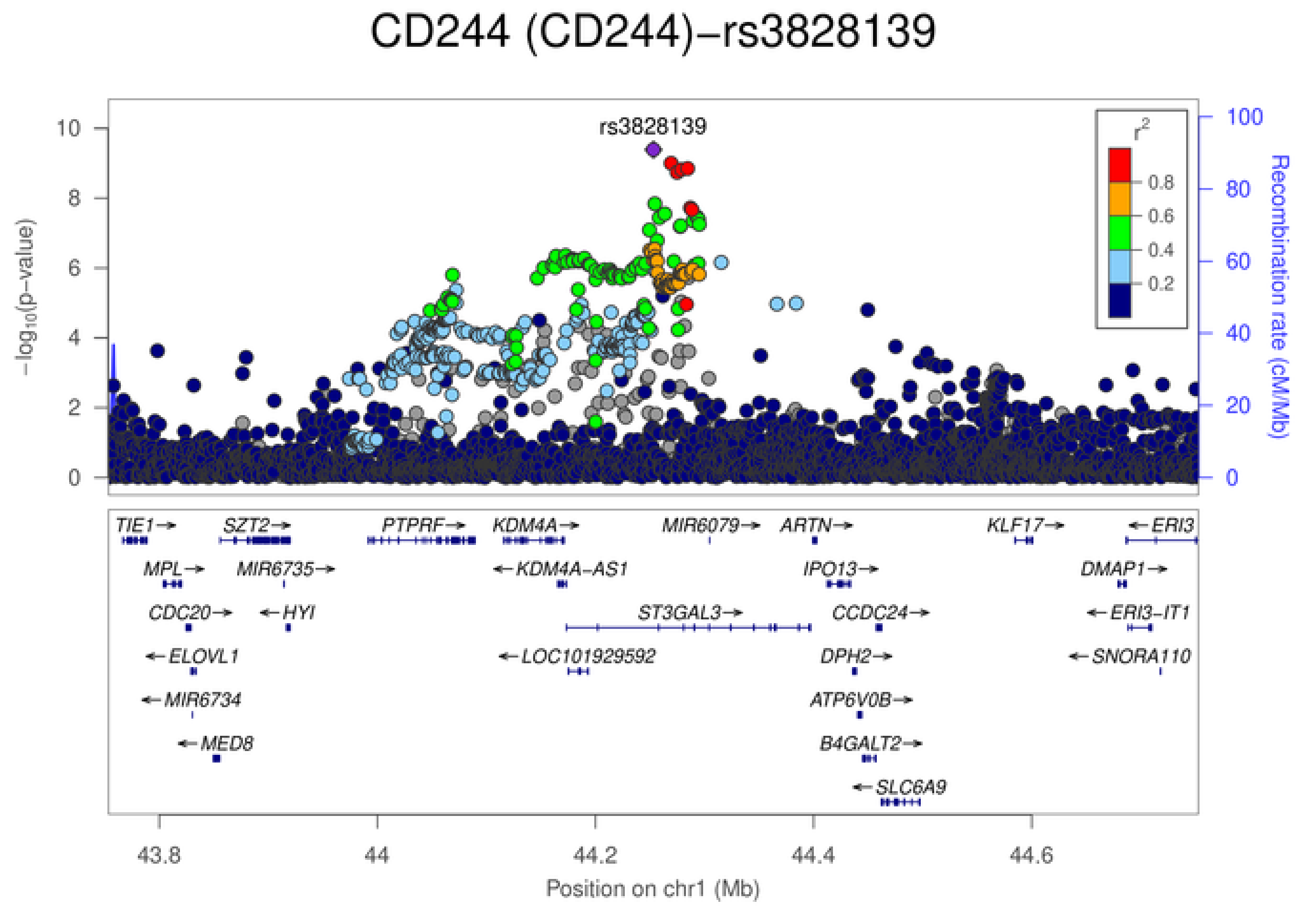
CD244 (CD244)−rs3828139
rs3828139
$-\log_{10}$(p−value)
Recombination rate (cM/Mb)
$r^2$
0.8
0.6
0.4
0.2
TIE1
SZT2
PTPRF
KDM4A
MIR6079
ARTN
KLF17
ERI3
MPL
MIR6735
KDM4A-AS1
IPO13
DMAP1
CDC20
HYI
ST3GAL3
CCDC24
ERI3-IT1
ELOVL1
LOC101929592
DPH2
SNORA110
MIR6734
ATP6V0B
MED8
B4GALT2
SLC6A9
43.8
44
44.2
44.4
44.6
Position on chr1 (Mb)

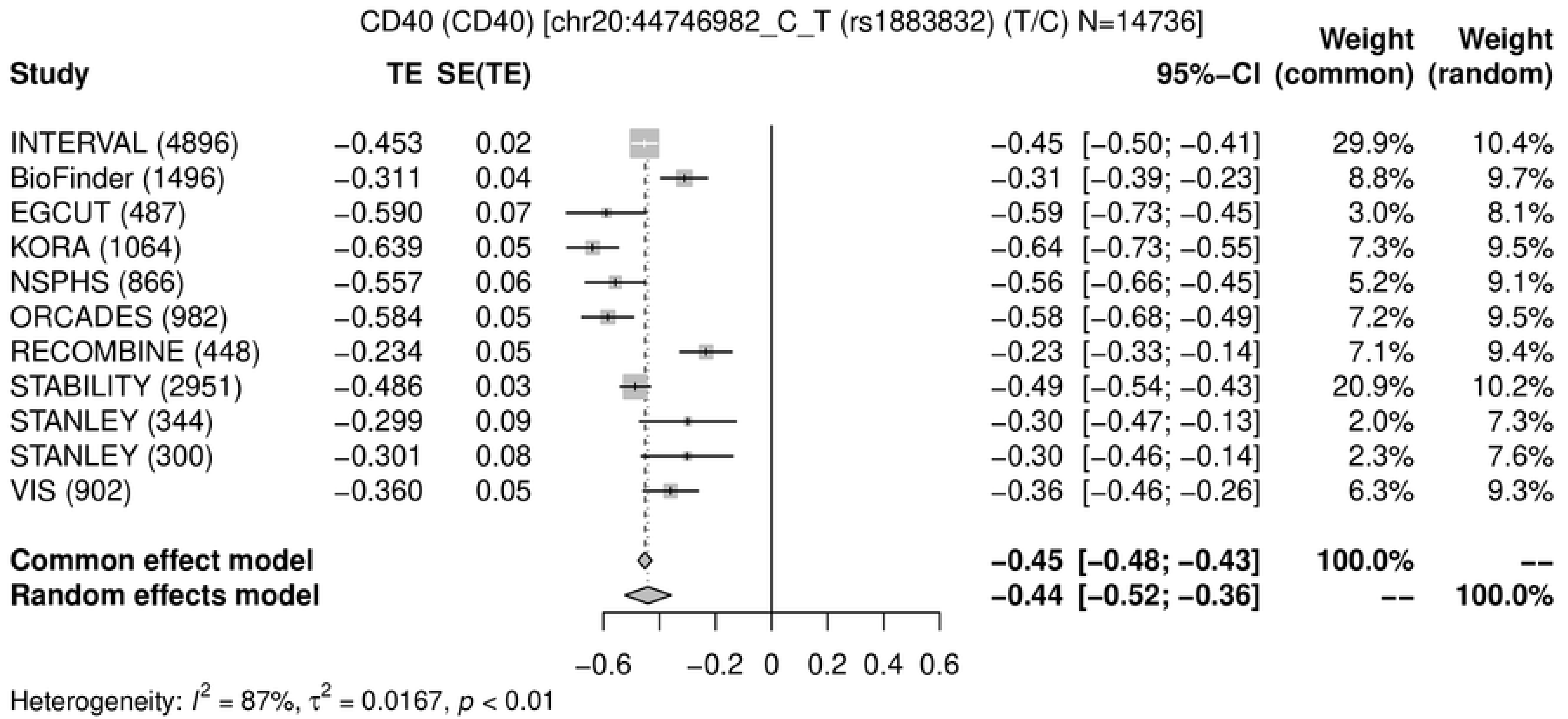
CD40 (CD40) [chr20:44746982_C_T (rs1883832) (T/C) N=14736]
Study
TE
SE(TE)
95%−CI
Weight (common)
Weight (random)
INTERVAL (4896) −0.453 0.02 −0.45 [−0.50; −0.41] 29.9% 10.4%
BioFinder (1496) −0.311 0.04 −0.31 [−0.39; −0.23] 8.8% 9.7%
EGCUT (487) −0.590 0.07 −0.59 [−0.73; −0.45] 3.0% 8.1%
KORA (1064) −0.639 0.05 −0.64 [−0.73; −0.55] 7.3% 9.5%
NSPHS (866) −0.557 0.06 −0.56 [−0.66; −0.45] 5.2% 9.1%
ORCADES (982) −0.584 0.05 −0.58 [−0.68; −0.49] 7.2% 9.5%
RECOMBINE (448) −0.234 0.05 −0.23 [−0.33; −0.14] 7.1% 9.4%
STABILITY (2951) −0.486 0.03 −0.49 [−0.54; −0.43] 20.9% 10.2%
STANLEY (344) −0.299 0.09 −0.30 [−0.47; −0.13] 2.0% 7.3%
STANLEY (300) −0.301 0.08 −0.30 [−0.46; −0.14] 2.3% 7.6%
VIS (902) −0.360 0.05 −0.36 [−0.46; −0.26] 6.3% 9.3%
Common effect model −0.45 [−0.48; −0.43] 100.0% --
Random effects model −0.44 [−0.52; −0.36] -- 100.0%
−0.6 −0.2 0 0.2 0.4 0.6
Heterogeneity: $I^2$ = 87%, $\tau^2$ = 0.0167, $p$ < 0.01

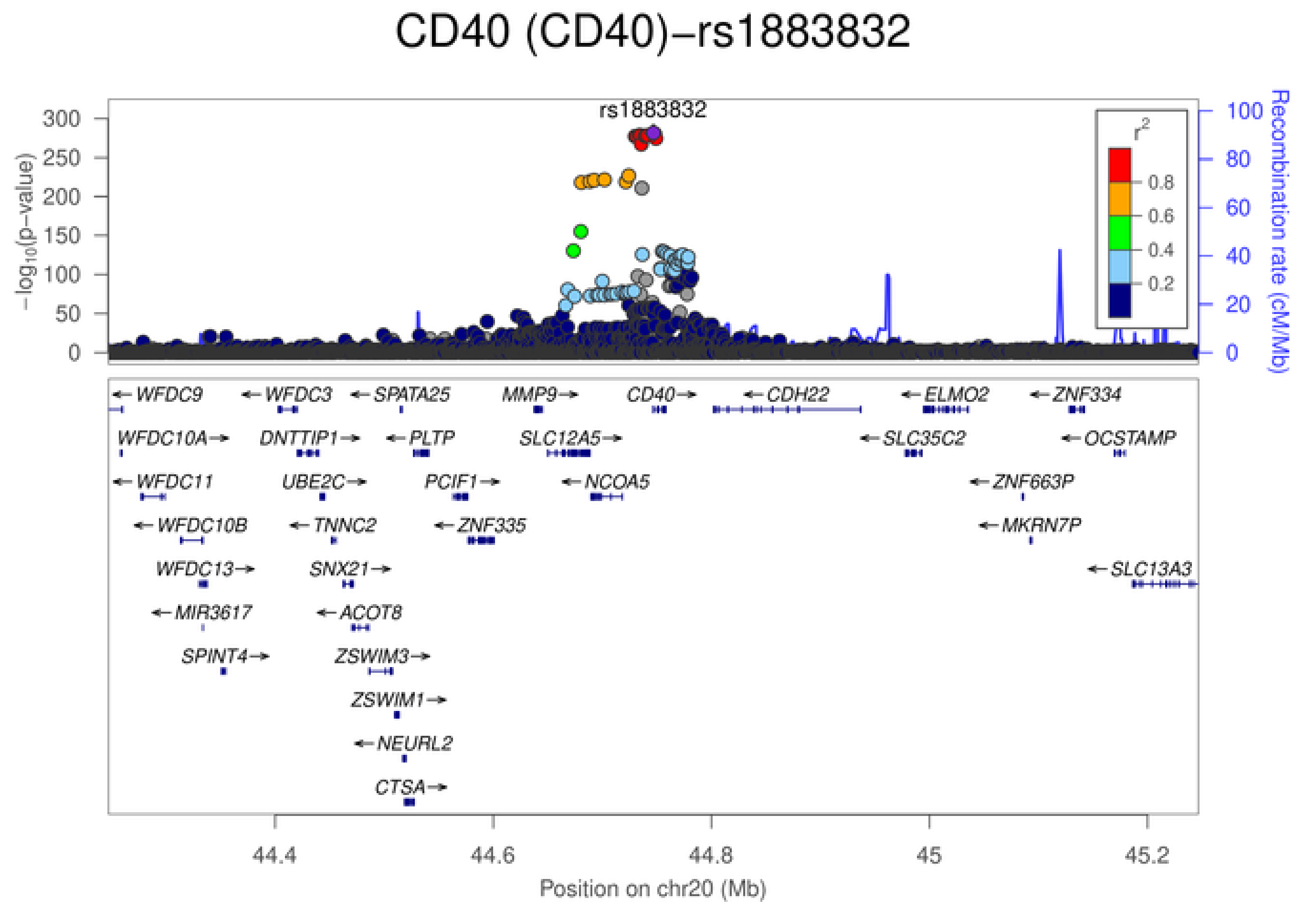
CD40 (CD40)−rs1883832
rs1883832
$-\log_{10}$(p−value)
Recombination rate (cM/Mb)
$r^2$
0.8
0.6
0.4
0.2
WFDC9
WFDC3
SPATA25
MMP9
CD40
CDH22
ELMO2
ZNF334
WFDC10A
DNTTIP1
PLTP
SLC12A5
SLC35C2
OCSTAMP
WFDC11
UBE2C
PCIF1
NCOA5
ZNF663P
WFDC10B
TNNC2
ZNF335
MKRN7P
WFDC13
SNX21
SLC13A3
MIR3617
ACOT8
SPINT4
ZSWIM3
ZSWIM1
NEURL2
CTSA
44.4
44.6
44.8
45
45.2
Position on chr20 (Mb)

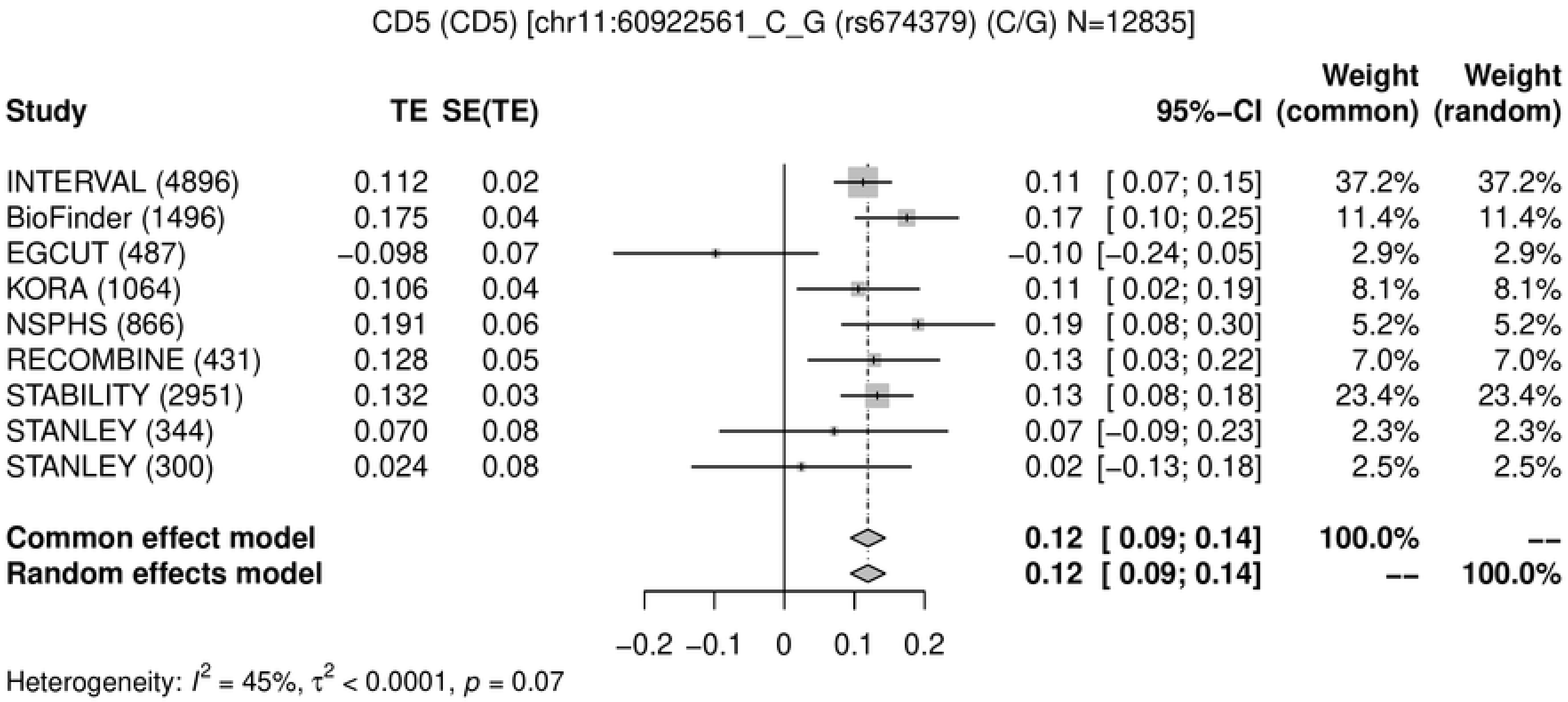
CD5 (CD5) [chr11:60922561_C_G (rs674379) (C/G) N=12835]
Study
TE
SE(TE)
95%-CI
Weight (common)
Weight (random)
INTERVAL (4896) 0.112 0.02 0.11 [ 0.07; 0.15] 37.2% 37.2%
BioFinder (1496) 0.175 0.04 0.17 [ 0.10; 0.25] 11.4% 11.4%
EGCUT (487) −0.098 0.07 −0.10 [−0.24; 0.05] 2.9% 2.9%
KORA (1064) 0.106 0.04 0.11 [ 0.02; 0.19] 8.1% 8.1%
NSPHS (866) 0.191 0.06 0.19 [ 0.08; 0.30] 5.2% 5.2%
RECOMBINE (431) 0.128 0.05 0.13 [ 0.03; 0.22] 7.0% 7.0%
STABILITY (2951) 0.132 0.03 0.13 [ 0.08; 0.18] 23.4% 23.4%
STANLEY (344) 0.070 0.08 0.07 [−0.09; 0.23] 2.3% 2.3%
STANLEY (300) 0.024 0.08 0.02 [−0.13; 0.18] 2.5% 2.5%
Common effect model 0.12 [ 0.09; 0.14] 100.0% --
Random effects model 0.12 [ 0.09; 0.14] -- 100.0%
−0.2 −0.1 0 0.1 0.2
Heterogeneity: $I^2$ = 45%, $\tau^2$ < 0.0001, $p$ = 0.07

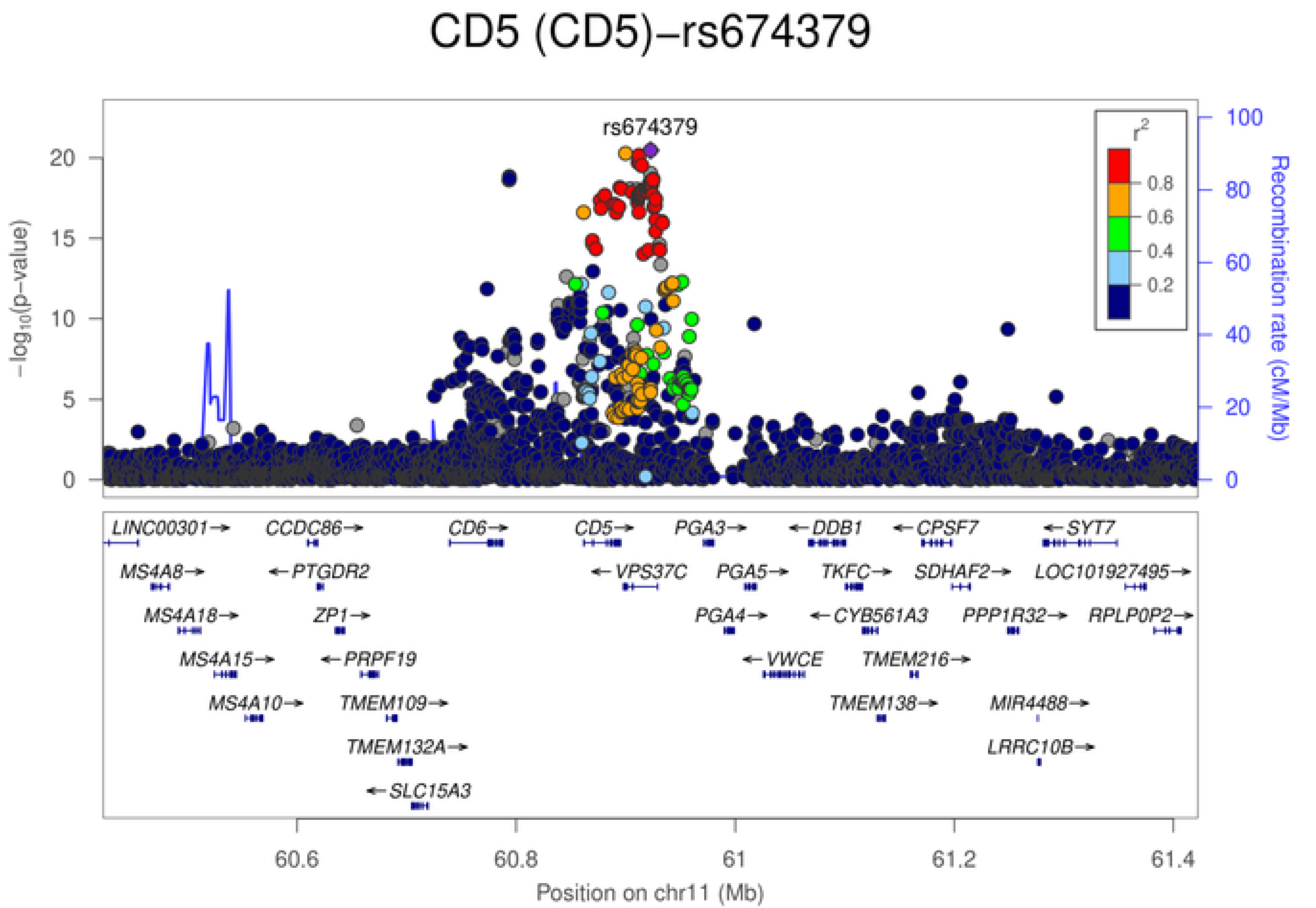
CD5 (CD5)−rs674379
rs674379
$-\log_{10}$(p-value)
Recombination rate (cM/Mb)
$r^2$
0.8
0.6
0.4
0.2
0
5
10
15
20
0
20
40
60
80
100
LINC00301→
CCDC86→
CD6→
CD5→
PGA3→
←DDB1
←CPSF7
←SYT7
MS4A8→
←PTGDR2
←VPS37C
PGA5→
TKFC→
SDHAF2→
LOC101927495→
MS4A18→
ZP1→
PGA4→
←CYB561A3
PPP1R32→
RPLP0P2→
MS4A15→
←PRPF19
←VWCE
TMEM216→
MS4A10→
TMEM109→
TMEM138→
MIR4488→
TMEM132A→
LRRC10B→
←SLC15A3
60.6
60.8
61
61.2
61.4
Position on chr11 (Mb)

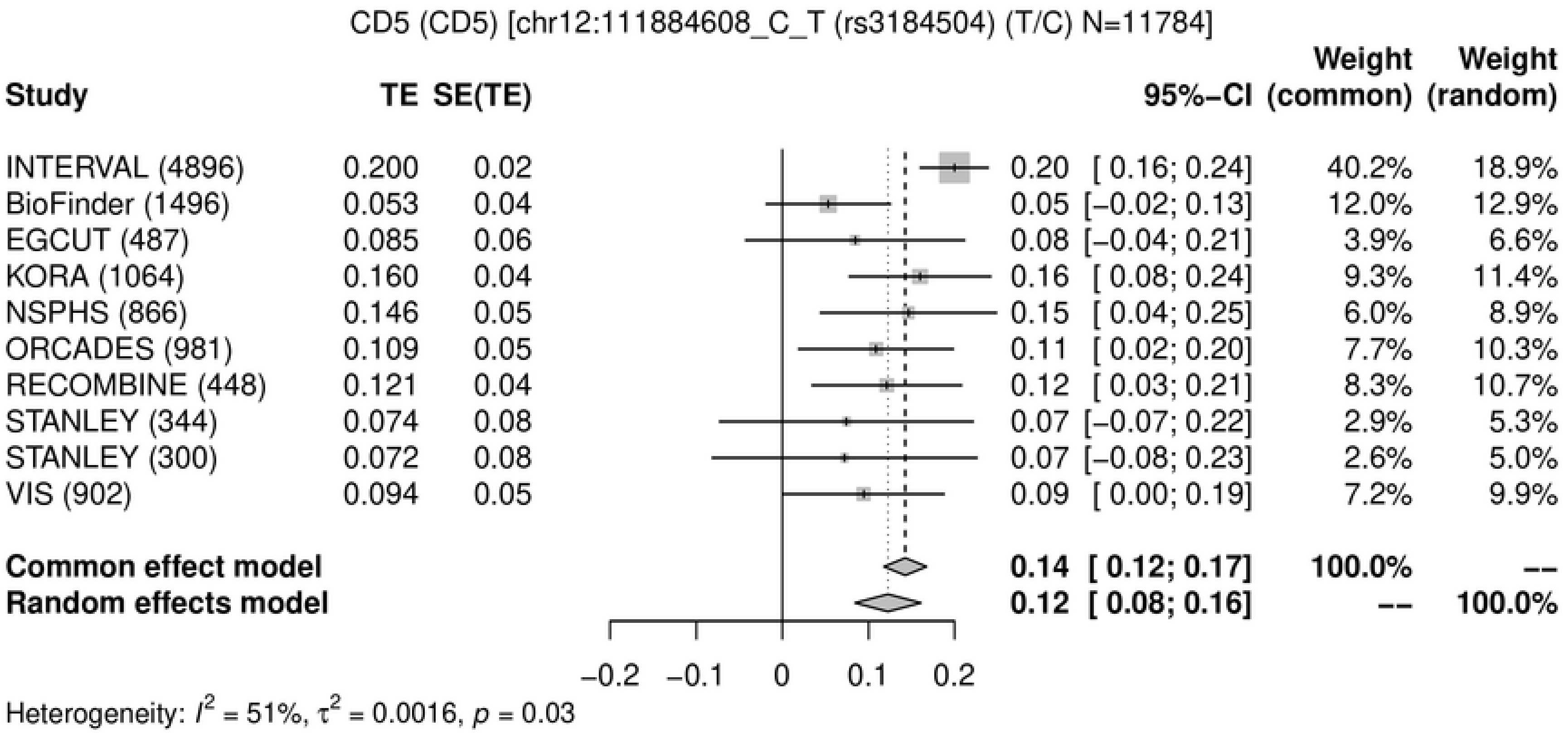
CD5 (CD5) [chr12:111884608_C_T (rs3184504) (T/C) N=11784]
Study
TE
SE(TE)
95%-CI
Weight (common)
Weight (random)
INTERVAL (4896) 0.200 0.02 0.20 [ 0.16; 0.24] 40.2% 18.9%
BioFinder (1496) 0.053 0.04 0.05 [−0.02; 0.13] 12.0% 12.9%
EGCUT (487) 0.085 0.06 0.08 [−0.04; 0.21] 3.9% 6.6%
KORA (1064) 0.160 0.04 0.16 [ 0.08; 0.24] 9.3% 11.4%
NSPHS (866) 0.146 0.05 0.15 [ 0.04; 0.25] 6.0% 8.9%
ORCADES (981) 0.109 0.05 0.11 [ 0.02; 0.20] 7.7% 10.3%
RECOMBINE (448) 0.121 0.04 0.12 [ 0.03; 0.21] 8.3% 10.7%
STANLEY (344) 0.074 0.08 0.07 [−0.07; 0.22] 2.9% 5.3%
STANLEY (300) 0.072 0.08 0.07 [−0.08; 0.23] 2.6% 5.0%
VIS (902) 0.094 0.05 0.09 [ 0.00; 0.19] 7.2% 9.9%
Common effect model 0.14 [ 0.12; 0.17] 100.0% --
Random effects model 0.12 [ 0.08; 0.16] -- 100.0%
−0.2 −0.1 0 0.1 0.2
Heterogeneity: $I^2$ = 51%, $\tau^2$ = 0.0016, $p$ = 0.03

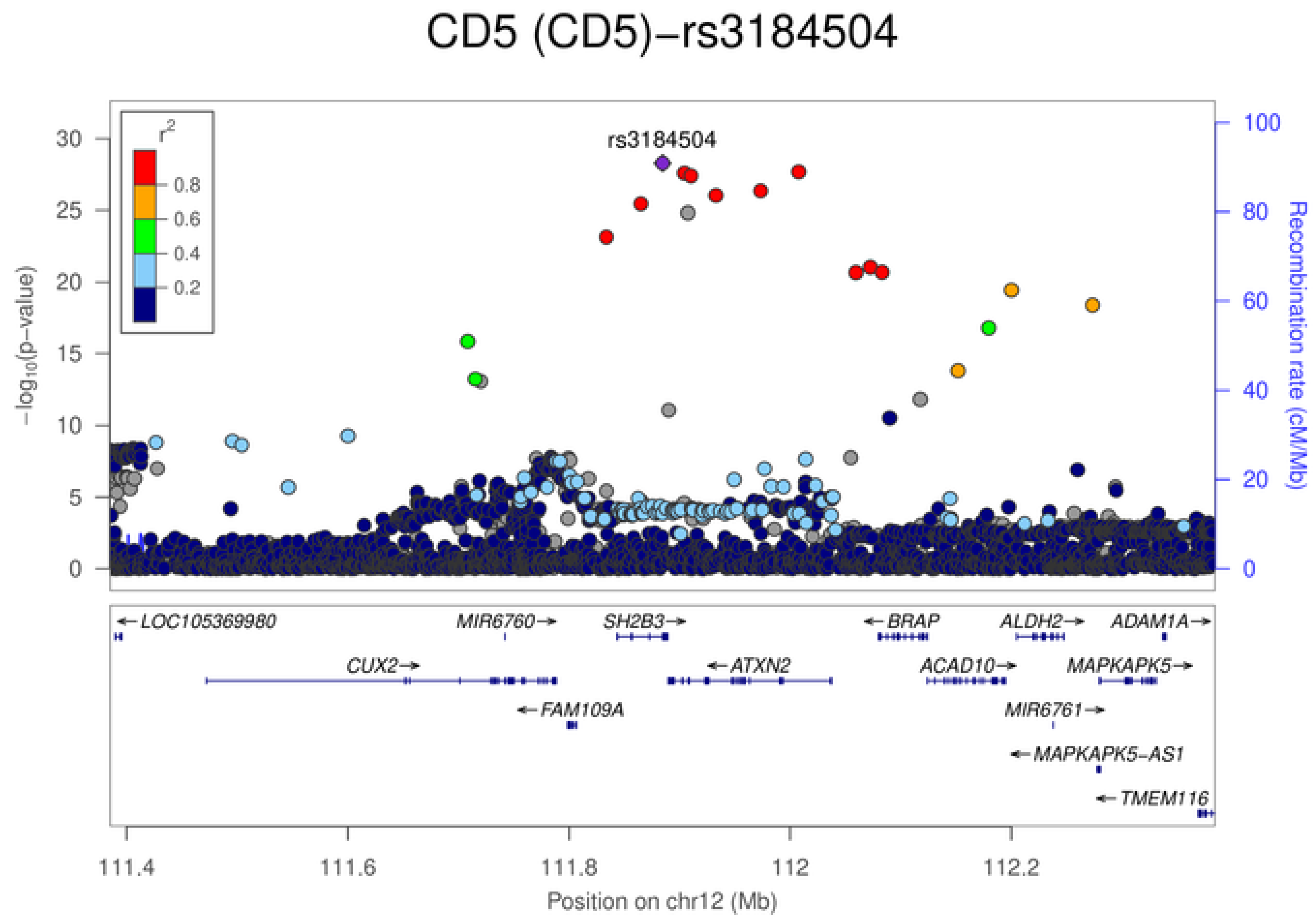
CD5 (CD5)−rs3184504
rs3184504
$r^2$
0.8
0.6
0.4
0.2
$-\log_{10}$(p-value)
Recombination rate (cM/Mb)
0 5 10 15 20 25 30
0 20 40 60 80 100
LOC105369980
MIR6760
SH2B3
BRAP
ALDH2
ADAM1A
CUX2
ATXN2
ACAD10
MAPKAPK5
FAM109A
MIR6761
MAPKAPK5-AS1
TMEM116
111.4 111.6 111.8 112 112.2
Position on chr12 (Mb)

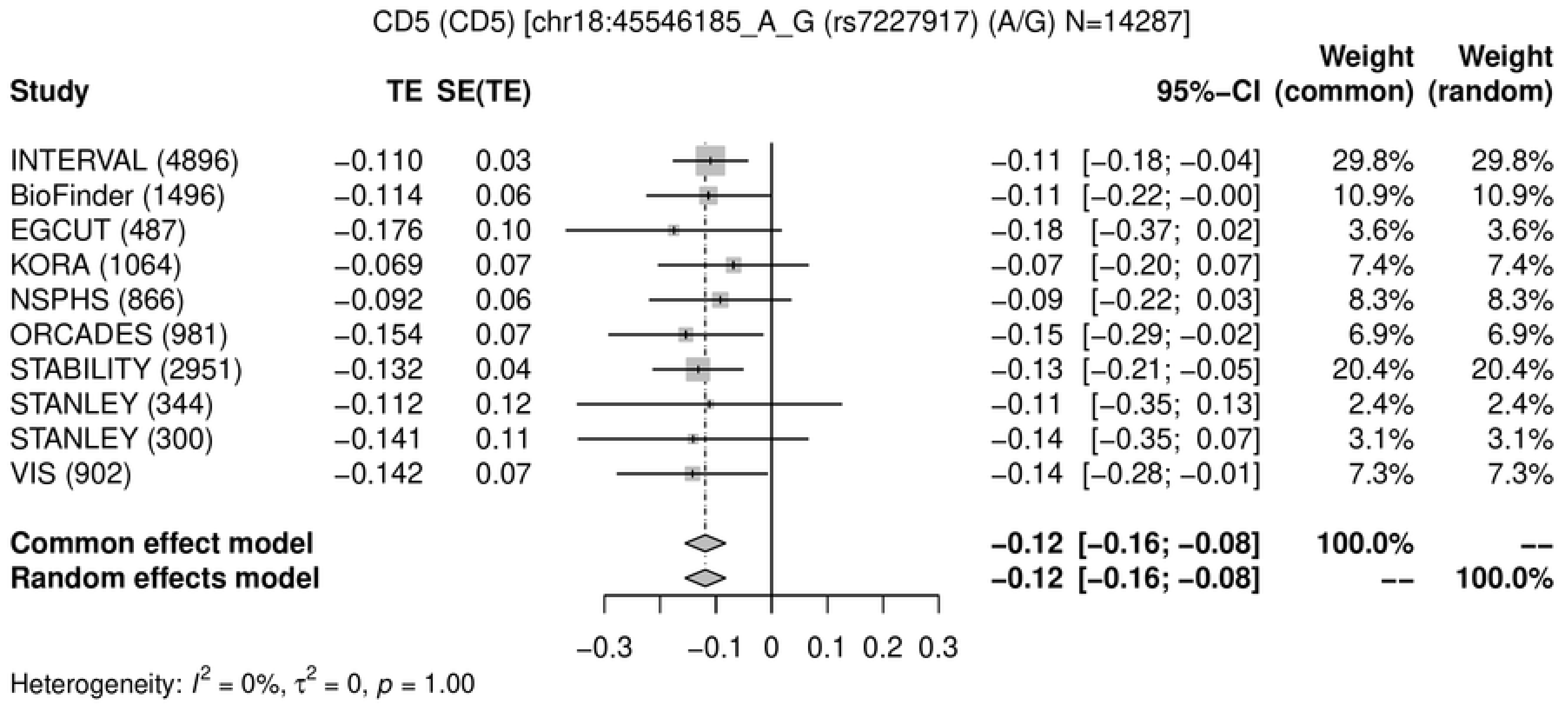
CD5 (CD5) [chr18:45546185_A_G (rs7227917) (A/G) N=14287]
Study
TE
SE(TE)
95%–CI
Weight (common)
Weight (random)
INTERVAL (4896) −0.110 0.03 −0.11 [−0.18; −0.04] 29.8% 29.8%
BioFinder (1496) −0.114 0.06 −0.11 [−0.22; −0.00] 10.9% 10.9%
EGCUT (487) −0.176 0.10 −0.18 [−0.37; 0.02] 3.6% 3.6%
KORA (1064) −0.069 0.07 −0.07 [−0.20; 0.07] 7.4% 7.4%
NSPHS (866) −0.092 0.06 −0.09 [−0.22; 0.03] 8.3% 8.3%
ORCADES (981) −0.154 0.07 −0.15 [−0.29; −0.02] 6.9% 6.9%
STABILITY (2951) −0.132 0.04 −0.13 [−0.21; −0.05] 20.4% 20.4%
STANLEY (344) −0.112 0.12 −0.11 [−0.35; 0.13] 2.4% 2.4%
STANLEY (300) −0.141 0.11 −0.14 [−0.35; 0.07] 3.1% 3.1%
VIS (902) −0.142 0.07 −0.14 [−0.28; −0.01] 7.3% 7.3%
Common effect model −0.12 [−0.16; −0.08] 100.0% --
Random effects model −0.12 [−0.16; −0.08] -- 100.0%
−0.3 −0.1 0 0.1 0.2 0.3
Heterogeneity: $I^2$ = 0%, $\tau^2$ = 0, $p$ = 1.00

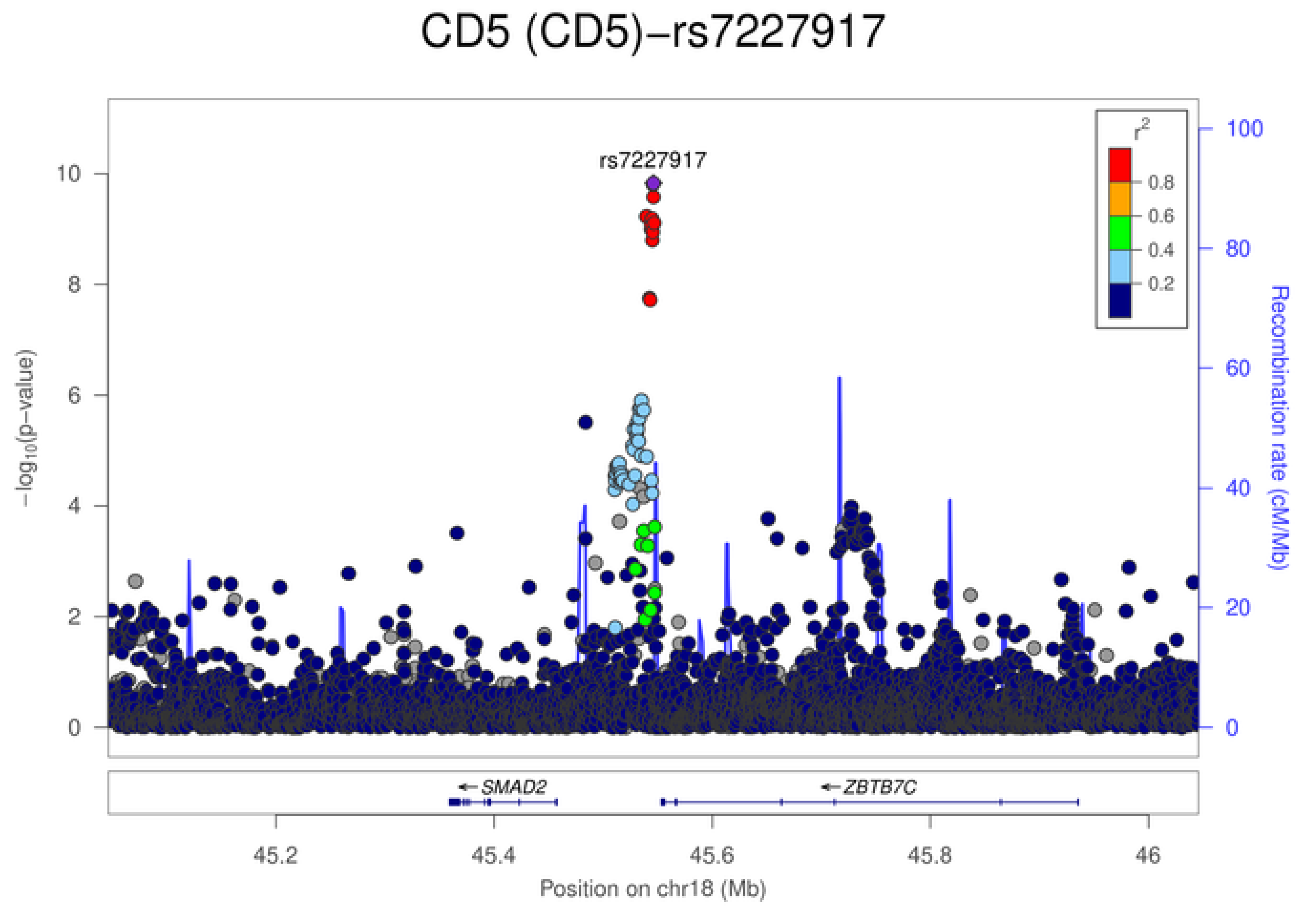
CD5 (CD5)−rs7227917
rs7227917
$-\log_{10}$(p−value)
Recombination rate (cM/Mb)
$r^2$
0.8
0.6
0.4
0.2
SMAD2
ZBTB7C
45.2 45.4 45.6 45.8 46
Position on chr18 (Mb)

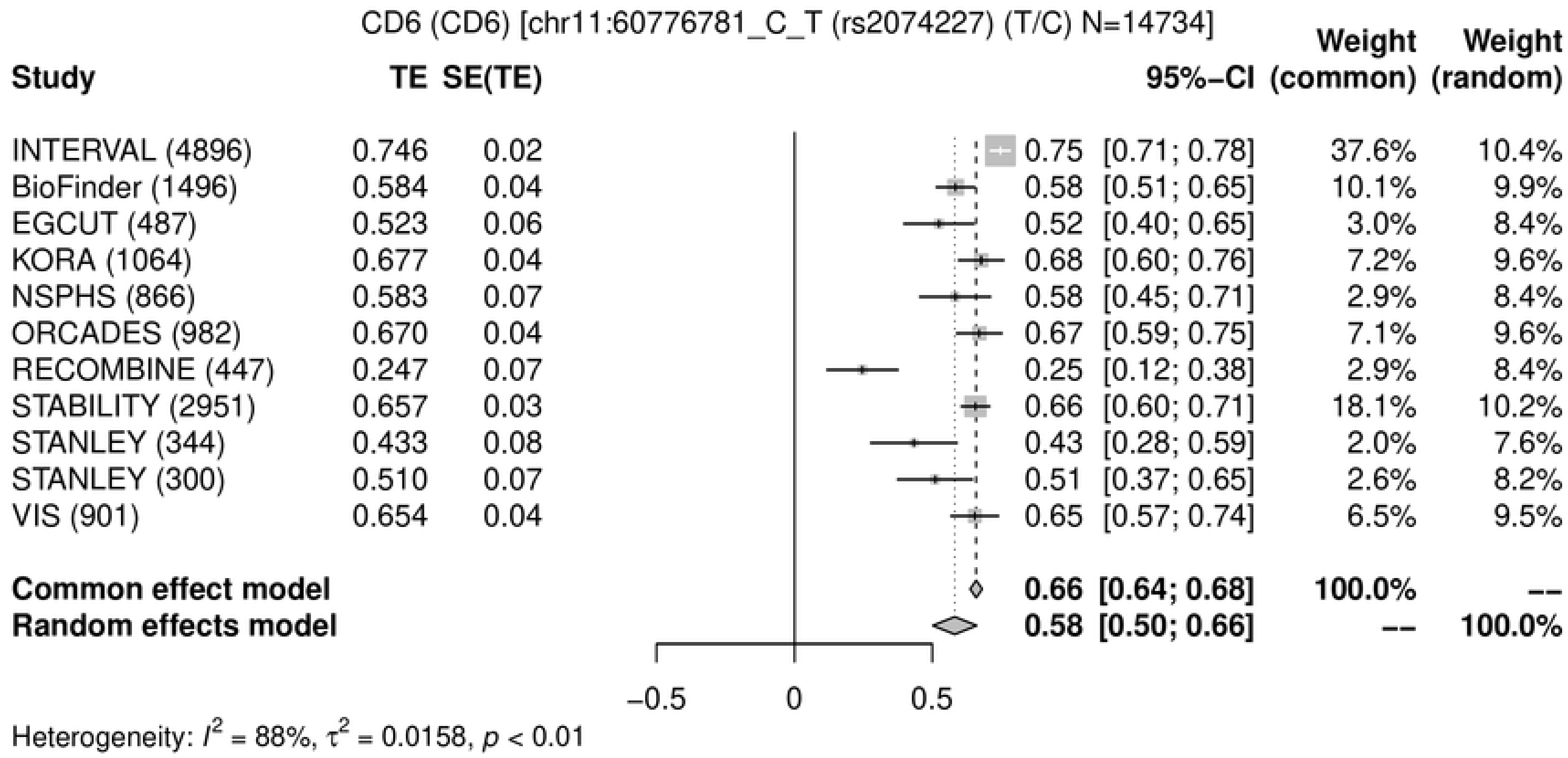
CD6 (CD6) [chr11:60776781_C_T (rs2074227) (T/C) N=14734]
Study
TE
SE(TE)
95%−CI
Weight (common)
Weight (random)
INTERVAL (4896) 0.746 0.02 0.75 [0.71; 0.78] 37.6% 10.4%
BioFinder (1496) 0.584 0.04 0.58 [0.51; 0.65] 10.1% 9.9%
EGCUT (487) 0.523 0.06 0.52 [0.40; 0.65] 3.0% 8.4%
KORA (1064) 0.677 0.04 0.68 [0.60; 0.76] 7.2% 9.6%
NSPHS (866) 0.583 0.07 0.58 [0.45; 0.71] 2.9% 8.4%
ORCADES (982) 0.670 0.04 0.67 [0.59; 0.75] 7.1% 9.6%
RECOMBINE (447) 0.247 0.07 0.25 [0.12; 0.38] 2.9% 8.4%
STABILITY (2951) 0.657 0.03 0.66 [0.60; 0.71] 18.1% 10.2%
STANLEY (344) 0.433 0.08 0.43 [0.28; 0.59] 2.0% 7.6%
STANLEY (300) 0.510 0.07 0.51 [0.37; 0.65] 2.6% 8.2%
VIS (901) 0.654 0.04 0.65 [0.57; 0.74] 6.5% 9.5%
Common effect model 0.66 [0.64; 0.68] 100.0% --
Random effects model 0.58 [0.50; 0.66] -- 100.0%
−0.5
0
0.5
Heterogeneity: $I^2$ = 88%, $\tau^2$ = 0.0158, $p$ < 0.01

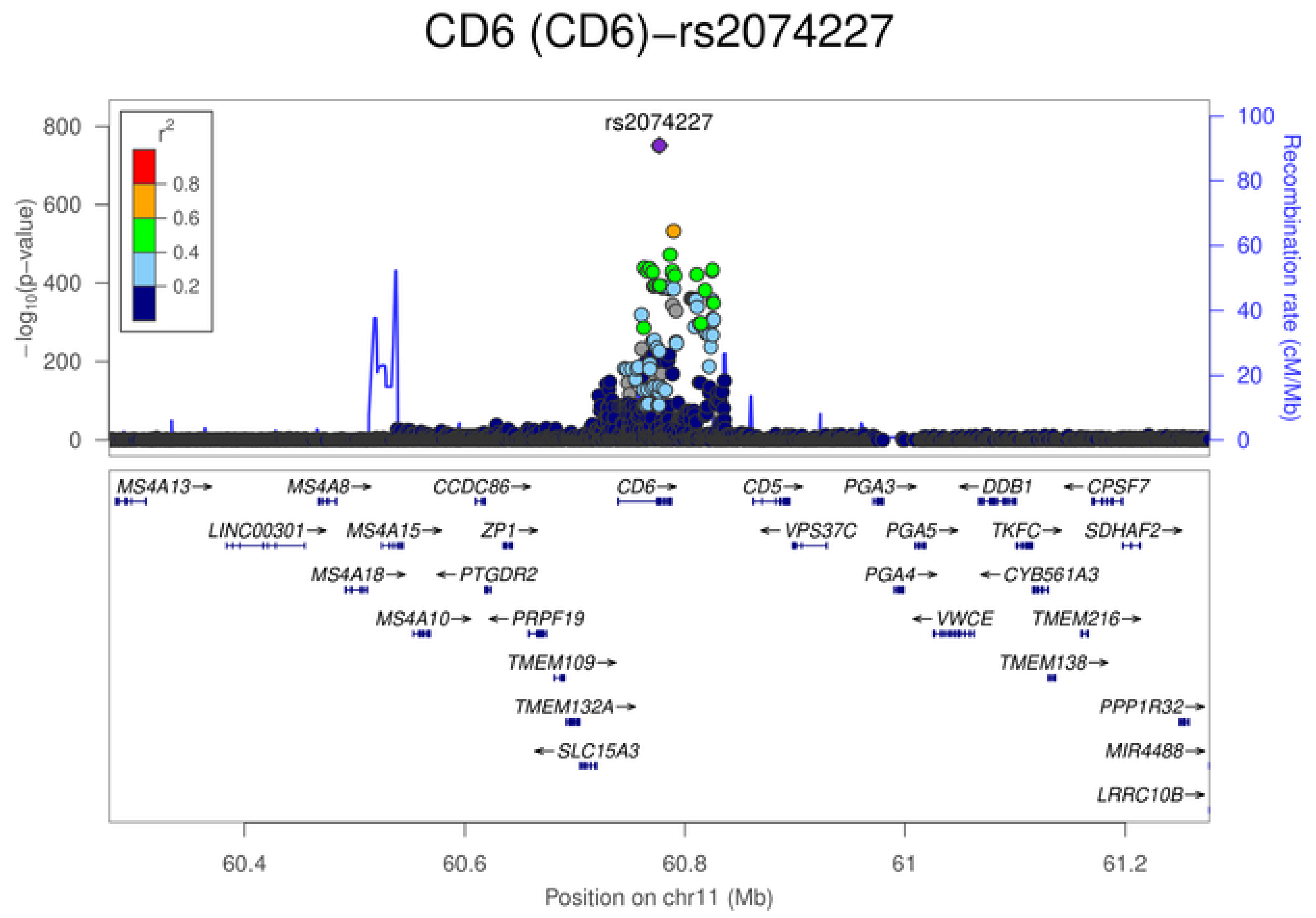
CD6 (CD6)−rs2074227
rs2074227
$r^2$
0.8
0.6
0.4
0.2
$-\log_{10}$(p−value)
Recombination rate (cM/Mb)
MS4A13→
MS4A8→
CCDC86→
CD6→
CD5→
PGA3→
←DDB1
←CPSF7
LINC00301→
MS4A15→
ZP1→
←VPS37C
PGA5→
TKFC→
SDHAF2→
MS4A18→
←PTGDR2
PGA4→
←CYB561A3
MS4A10→
←PRPF19
←VWCE
TMEM216→
TMEM109→
TMEM138→
TMEM132A→
PPP1R32→
←SLC15A3
MIR4488→
LRRC10B→
60.4
60.6
60.8
61
61.2
Position on chr11 (Mb)

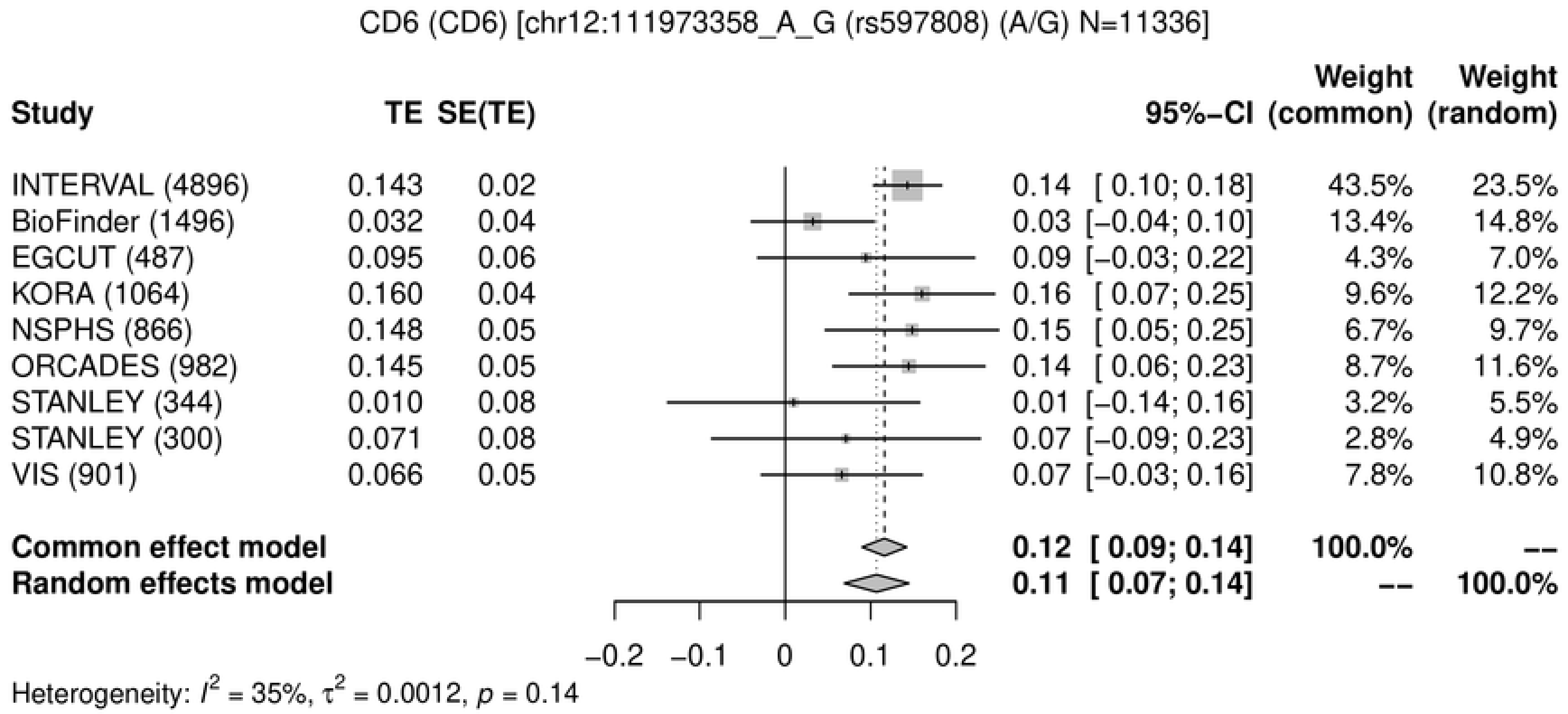
CD6 (CD6) [chr12:111973358_A_G (rs597808) (A/G) N=11336]
Study
TE
SE(TE)
95%−CI
Weight (common)
Weight (random)
INTERVAL (4896) 0.143 0.02 0.14 [ 0.10; 0.18] 43.5% 23.5%
BioFinder (1496) 0.032 0.04 0.03 [−0.04; 0.10] 13.4% 14.8%
EGCUT (487) 0.095 0.06 0.09 [−0.03; 0.22] 4.3% 7.0%
KORA (1064) 0.160 0.04 0.16 [ 0.07; 0.25] 9.6% 12.2%
NSPHS (866) 0.148 0.05 0.15 [ 0.05; 0.25] 6.7% 9.7%
ORCADES (982) 0.145 0.05 0.14 [ 0.06; 0.23] 8.7% 11.6%
STANLEY (344) 0.010 0.08 0.01 [−0.14; 0.16] 3.2% 5.5%
STANLEY (300) 0.071 0.08 0.07 [−0.09; 0.23] 2.8% 4.9%
VIS (901) 0.066 0.05 0.07 [−0.03; 0.16] 7.8% 10.8%
Common effect model 0.12 [ 0.09; 0.14] 100.0% --
Random effects model 0.11 [ 0.07; 0.14] -- 100.0%
−0.2 −0.1 0 0.1 0.2
Heterogeneity: $I^2$ = 35%, $\tau^2$ = 0.0012, $p$ = 0.14

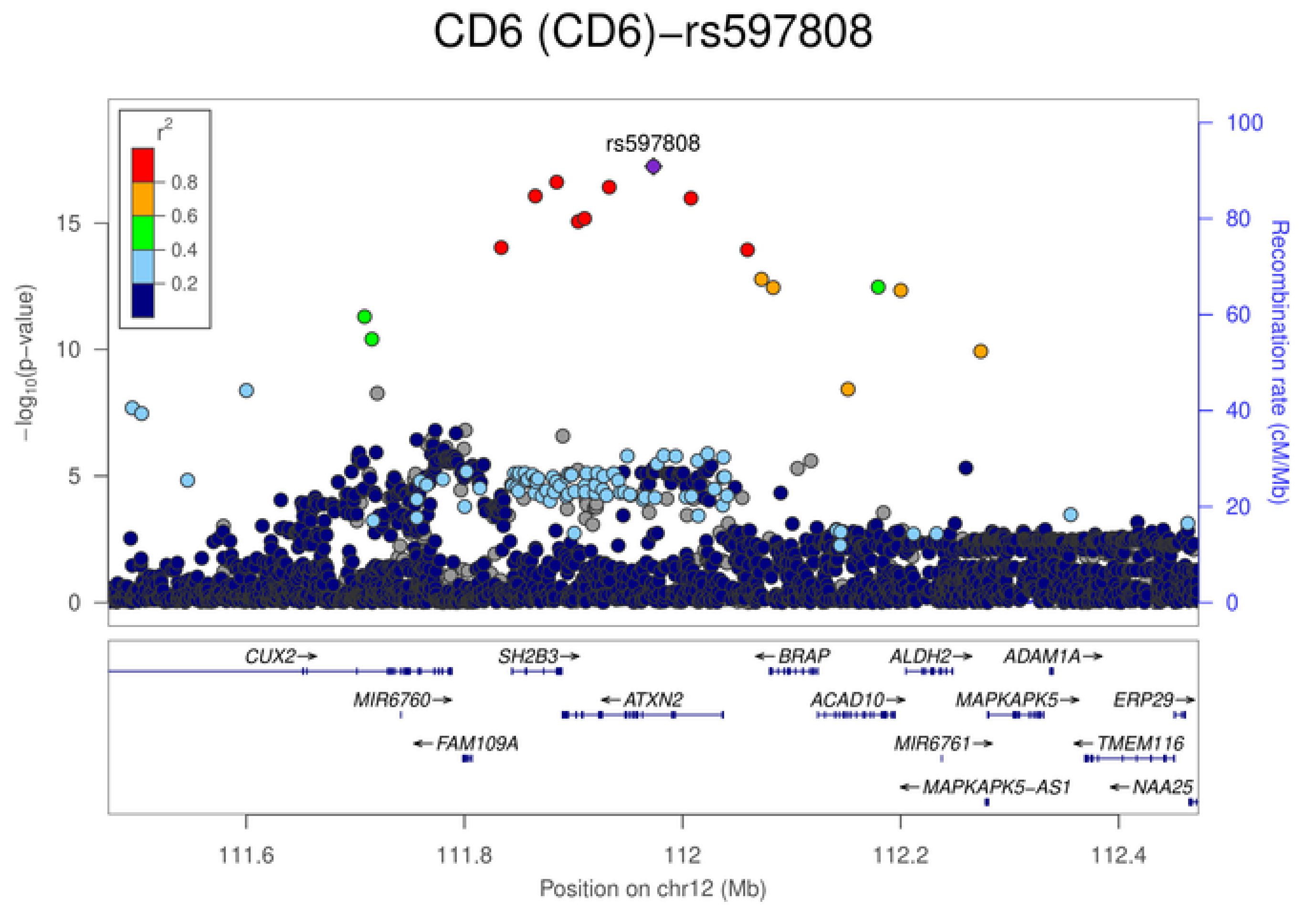
CD6 (CD6)−rs597808
rs597808
$r^2$
0.8
0.6
0.4
0.2
$-\log_{10}$(p−value)
Recombination rate (cM/Mb)
0
5
10
15
20
40
60
80
100
CUX2→
SH2B3→
←BRAP
ALDH2→
ADAM1A→
MIR6760→
←ATXN2
ACAD10→
MAPKAPK5→
ERP29→
←FAM109A
MIR6761→
←TMEM116
←MAPKAPK5−AS1
←NAA25
111.6
111.8
112
112.2
112.4
Position on chr12 (Mb)

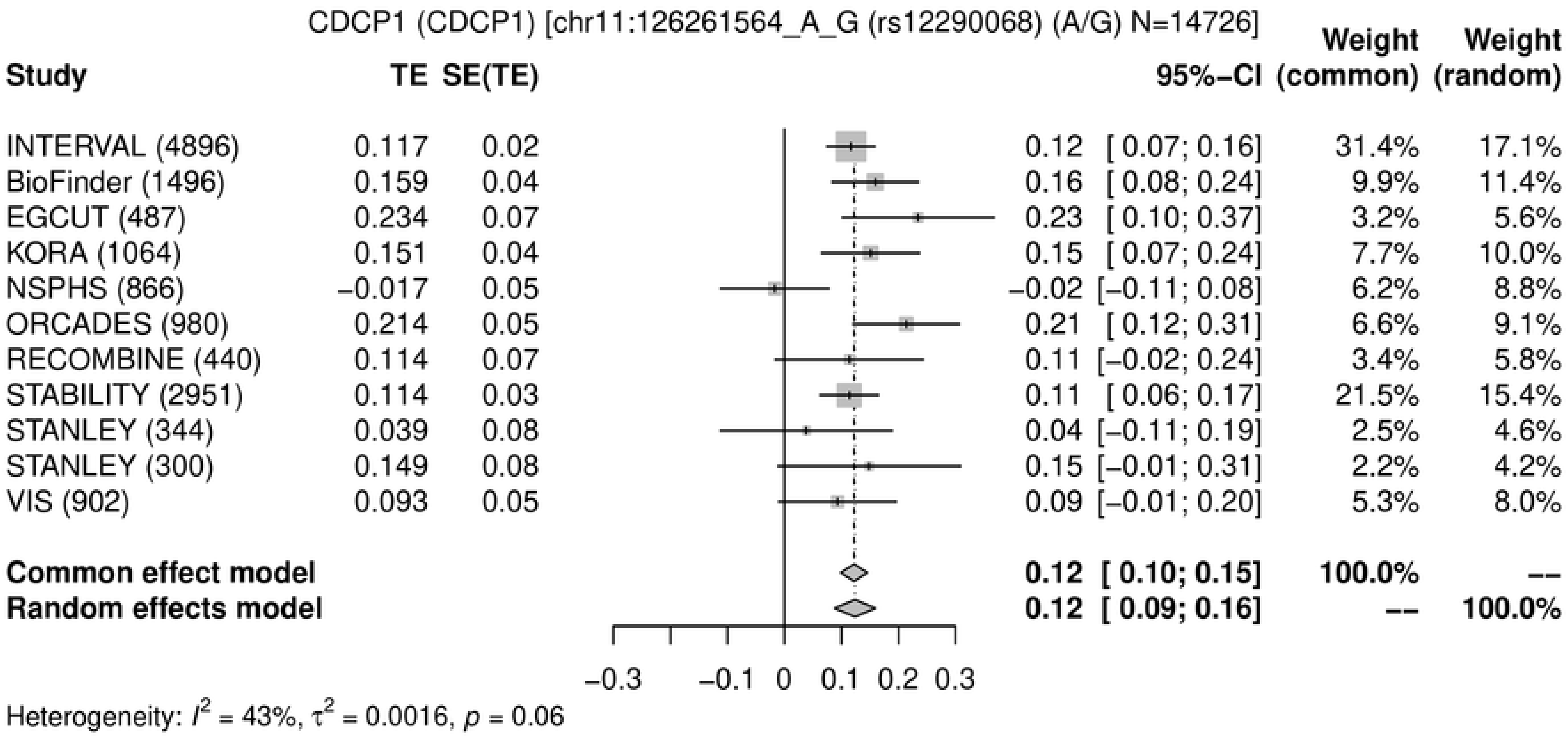
CDCP1 (CDCP1) [chr11:126261564_A_G (rs12290068) (A/G) N=14726]
Study | TE | SE(TE) | 95%-CI | Weight (common) | Weight (random)
INTERVAL (4896) | 0.117 | 0.02 | 0.12 [ 0.07; 0.16] | 31.4% | 17.1%
BioFinder (1496) | 0.159 | 0.04 | 0.16 [ 0.08; 0.24] | 9.9% | 11.4%
EGCUT (487) | 0.234 | 0.07 | 0.23 [ 0.10; 0.37] | 3.2% | 5.6%
KORA (1064) | 0.151 | 0.04 | 0.15 [ 0.07; 0.24] | 7.7% | 10.0%
NSPHS (866) | −0.017 | 0.05 | −0.02 [−0.11; 0.08] | 6.2% | 8.8%
ORCADES (980) | 0.214 | 0.05 | 0.21 [ 0.12; 0.31] | 6.6% | 9.1%
RECOMBINE (440) | 0.114 | 0.07 | 0.11 [−0.02; 0.24] | 3.4% | 5.8%
STABILITY (2951) | 0.114 | 0.03 | 0.11 [ 0.06; 0.17] | 21.5% | 15.4%
STANLEY (344) | 0.039 | 0.08 | 0.04 [−0.11; 0.19] | 2.5% | 4.6%
STANLEY (300) | 0.149 | 0.08 | 0.15 [−0.01; 0.31] | 2.2% | 4.2%
VIS (902) | 0.093 | 0.05 | 0.09 [−0.01; 0.20] | 5.3% | 8.0%
Common effect model | 0.12 [ 0.10; 0.15] | 100.0% | --
Random effects model | 0.12 [ 0.09; 0.16] | -- | 100.0%
−0.3 −0.1 0 0.1 0.2 0.3
Heterogeneity: $I^2$ = 43%, $\tau^2$ = 0.0016, $p$ = 0.06

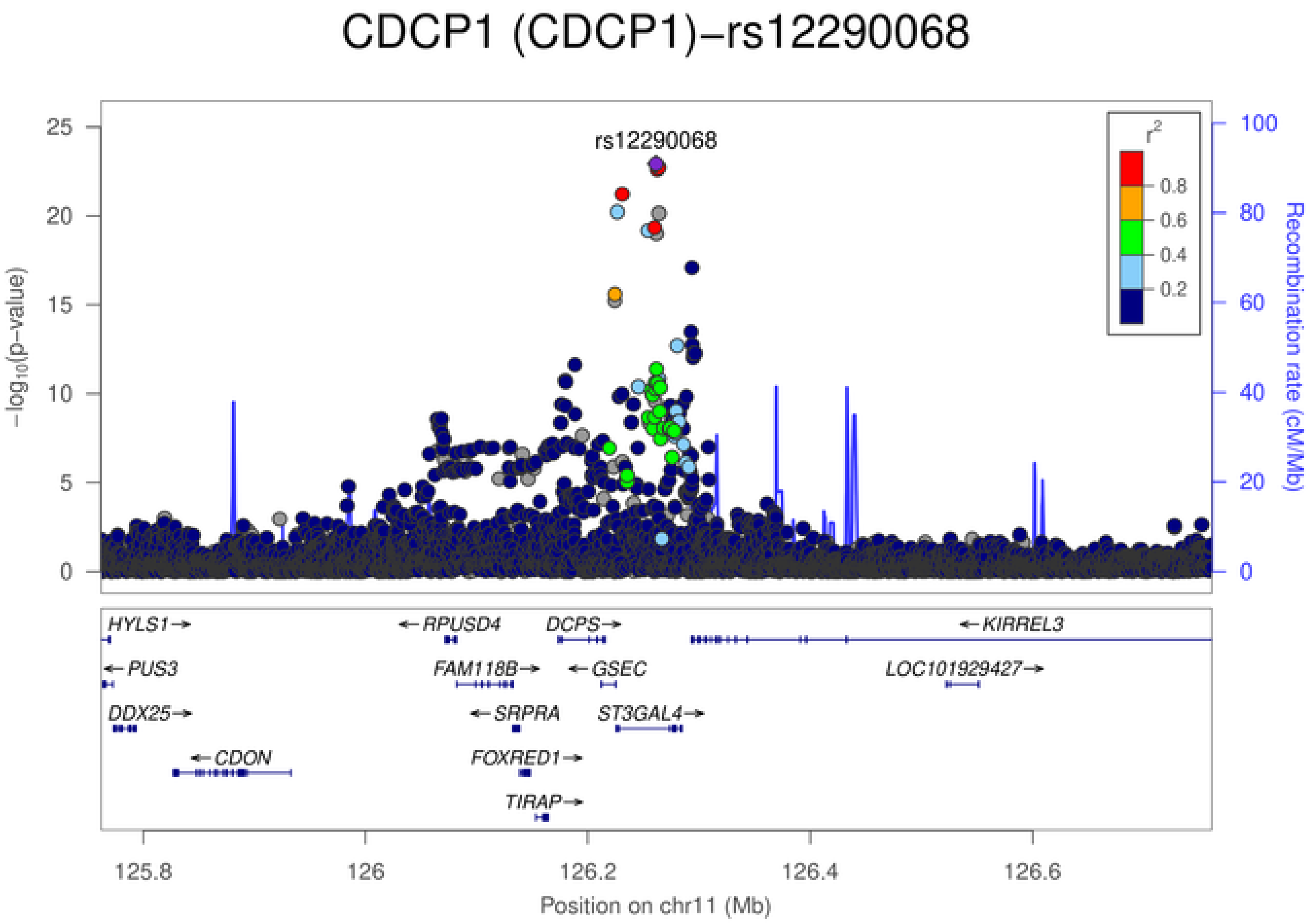
CDCP1 (CDCP1)−rs12290068
rs12290068
$-\log_{10}$(p−value)
Recombination rate (cM/Mb)
$r^2$
0.8
0.6
0.4
0.2
HYLS1→
←PUS3
DDX25→
←CDON
←RPUSD4
FAM118B→
←SRPRA
FOXRED1→
TIRAP→
DCPS→
←GSEC
ST3GAL4→
←KIRREL3
LOC101929427→
125.8 126 126.2 126.4 126.6
Position on chr11 (Mb)

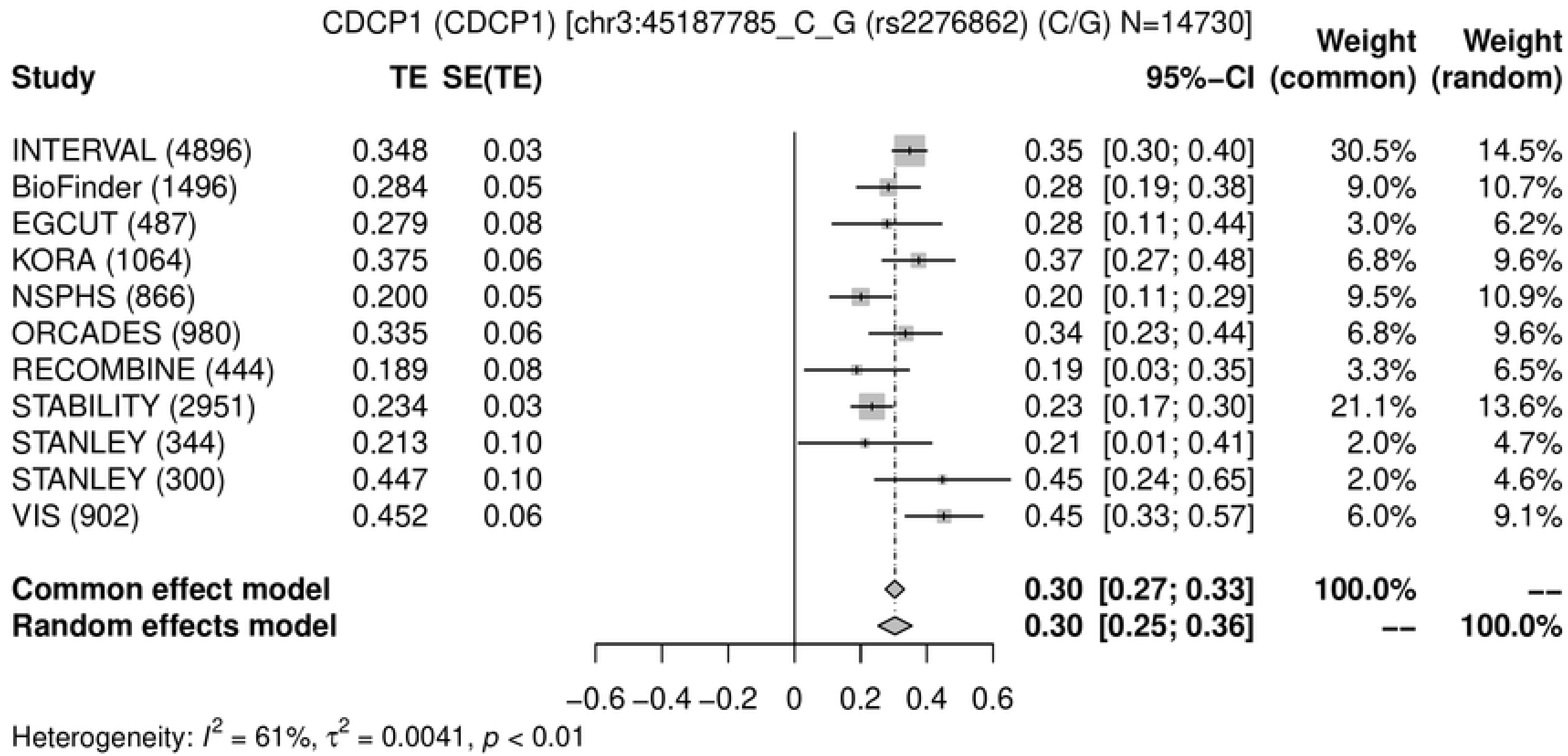
CDCP1 (CDCP1) [chr3:45187785_C_G (rs2276862) (C/G) N=14730]
Study
TE
SE(TE)
95%−CI
Weight (common)
Weight (random)
INTERVAL (4896) 0.348 0.03 0.35 [0.30; 0.40] 30.5% 14.5%
BioFinder (1496) 0.284 0.05 0.28 [0.19; 0.38] 9.0% 10.7%
EGCUT (487) 0.279 0.08 0.28 [0.11; 0.44] 3.0% 6.2%
KORA (1064) 0.375 0.06 0.37 [0.27; 0.48] 6.8% 9.6%
NSPHS (866) 0.200 0.05 0.20 [0.11; 0.29] 9.5% 10.9%
ORCADES (980) 0.335 0.06 0.34 [0.23; 0.44] 6.8% 9.6%
RECOMBINE (444) 0.189 0.08 0.19 [0.03; 0.35] 3.3% 6.5%
STABILITY (2951) 0.234 0.03 0.23 [0.17; 0.30] 21.1% 13.6%
STANLEY (344) 0.213 0.10 0.21 [0.01; 0.41] 2.0% 4.7%
STANLEY (300) 0.447 0.10 0.45 [0.24; 0.65] 2.0% 4.6%
VIS (902) 0.452 0.06 0.45 [0.33; 0.57] 6.0% 9.1%
Common effect model 0.30 [0.27; 0.33] 100.0% --
Random effects model 0.30 [0.25; 0.36] -- 100.0%
−0.6 −0.4 −0.2 0 0.2 0.4 0.6
Heterogeneity: $I^2$ = 61%, $\tau^2$ = 0.0041, $p$ < 0.01

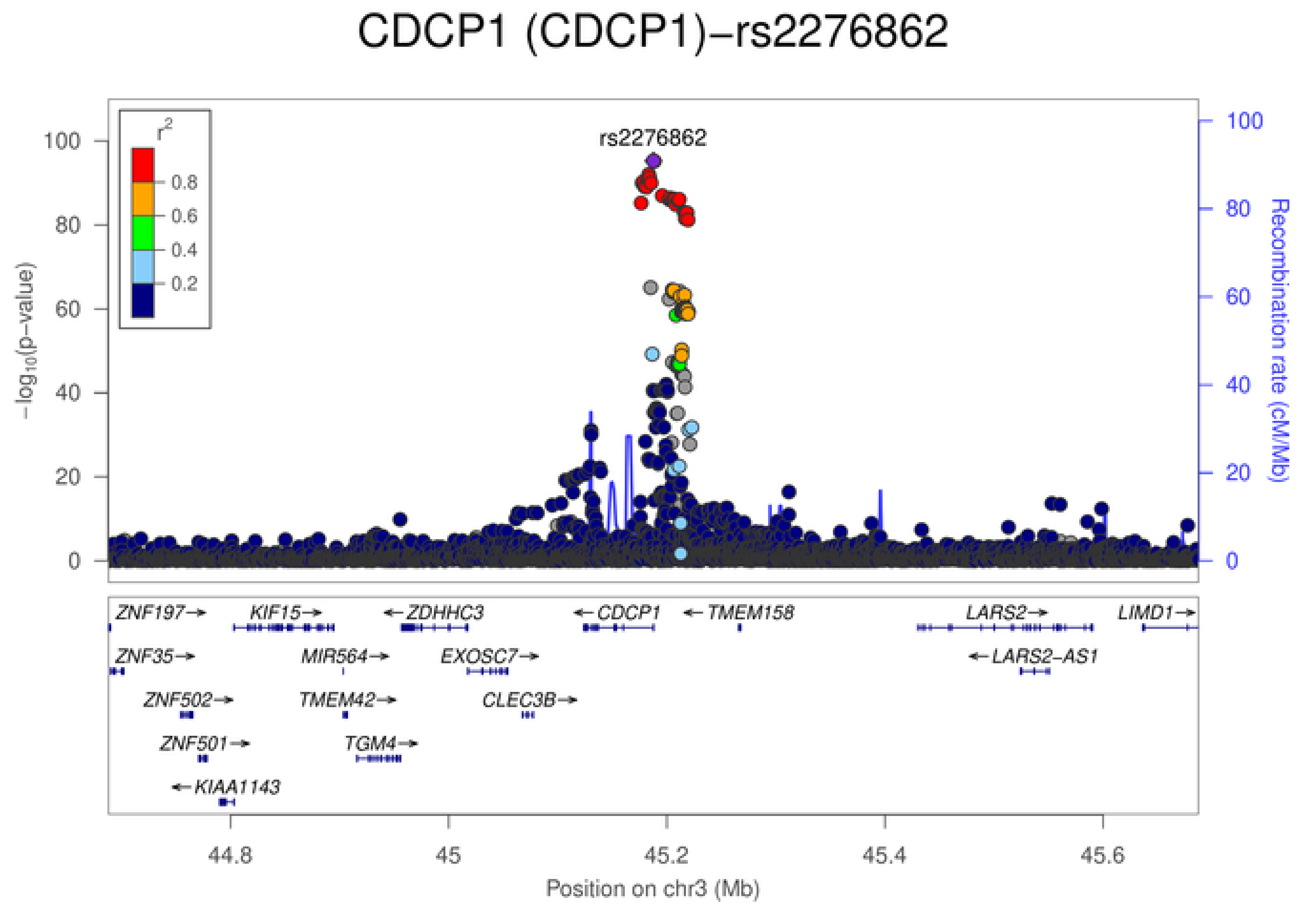
CDCP1 (CDCP1)−rs2276862
rs2276862
$r^2$
0.8
0.6
0.4
0.2
$-\log_{10}$(p−value)
Recombination rate (cM/Mb)
100
80
60
40
20
0
ZNF197→
KIF15→
←ZDHHC3
←CDCP1
←TMEM158
LARS2→
LIMD1→
ZNF35→
MIR564→
EXOSC7→
←LARS2-AS1
ZNF502→
TMEM42→
CLEC3B→
ZNF501→
TGM4→
←KIAA1143
44.8
45
45.2
45.4
45.6
Position on chr3 (Mb)

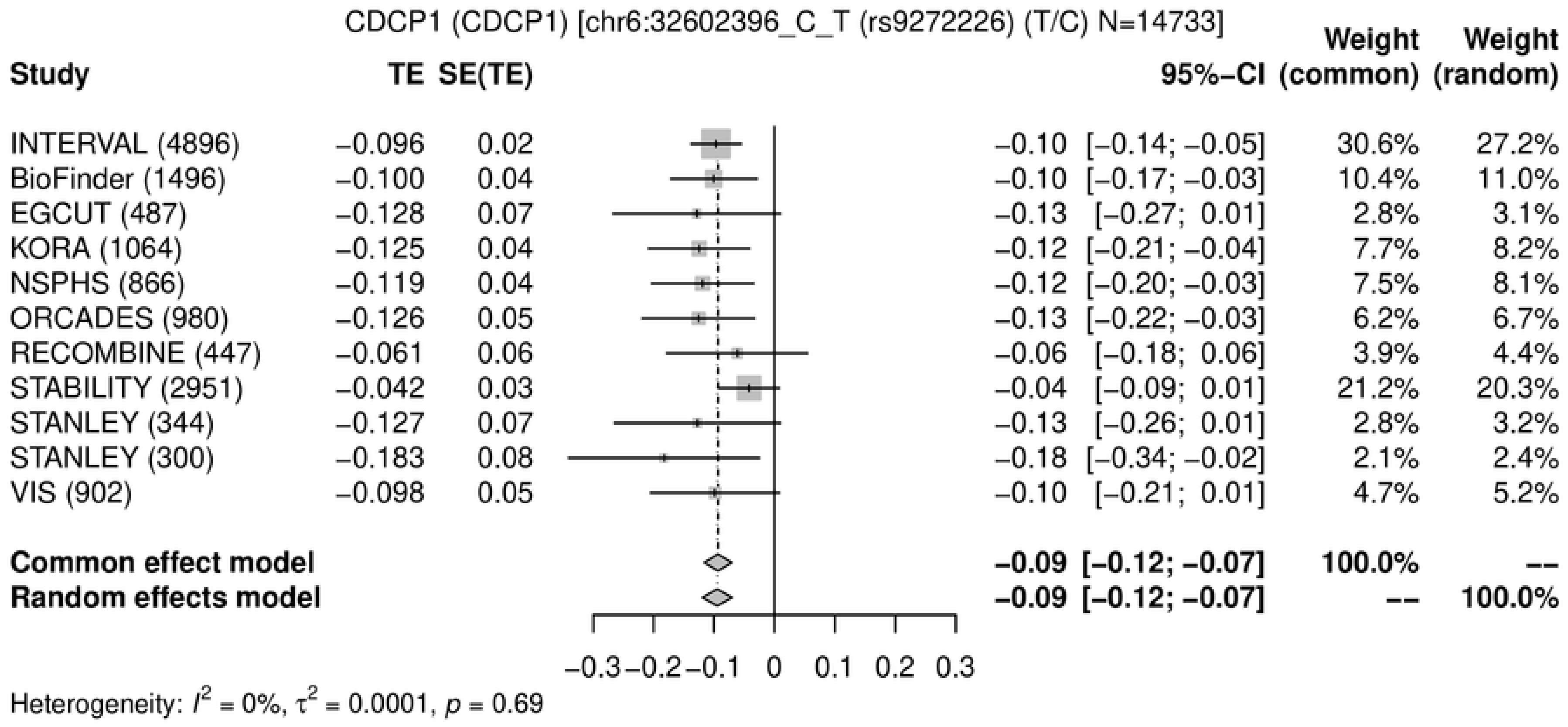
CDCP1 (CDCP1) [chr6:32602396_C_T (rs9272226) (T/C) N=14733]
Study
TE SE(TE)
95%−CI
Weight (common)
Weight (random)
INTERVAL (4896) −0.096 0.02 −0.10 [−0.14; −0.05] 30.6% 27.2%
BioFinder (1496) −0.100 0.04 −0.10 [−0.17; −0.03] 10.4% 11.0%
EGCUT (487) −0.128 0.07 −0.13 [−0.27; 0.01] 2.8% 3.1%
KORA (1064) −0.125 0.04 −0.12 [−0.21; −0.04] 7.7% 8.2%
NSPHS (866) −0.119 0.04 −0.12 [−0.20; −0.03] 7.5% 8.1%
ORCADES (980) −0.126 0.05 −0.13 [−0.22; −0.03] 6.2% 6.7%
RECOMBINE (447) −0.061 0.06 −0.06 [−0.18; 0.06] 3.9% 4.4%
STABILITY (2951) −0.042 0.03 −0.04 [−0.09; 0.01] 21.2% 20.3%
STANLEY (344) −0.127 0.07 −0.13 [−0.26; 0.01] 2.8% 3.2%
STANLEY (300) −0.183 0.08 −0.18 [−0.34; −0.02] 2.1% 2.4%
VIS (902) −0.098 0.05 −0.10 [−0.21; 0.01] 4.7% 5.2%
Common effect model −0.09 [−0.12; −0.07] 100.0% --
Random effects model −0.09 [−0.12; −0.07] -- 100.0%
−0.3 −0.2 −0.1 0 0.1 0.2 0.3
Heterogeneity: $I^2$ = 0%, $\tau^2$ = 0.0001, $p$ = 0.69

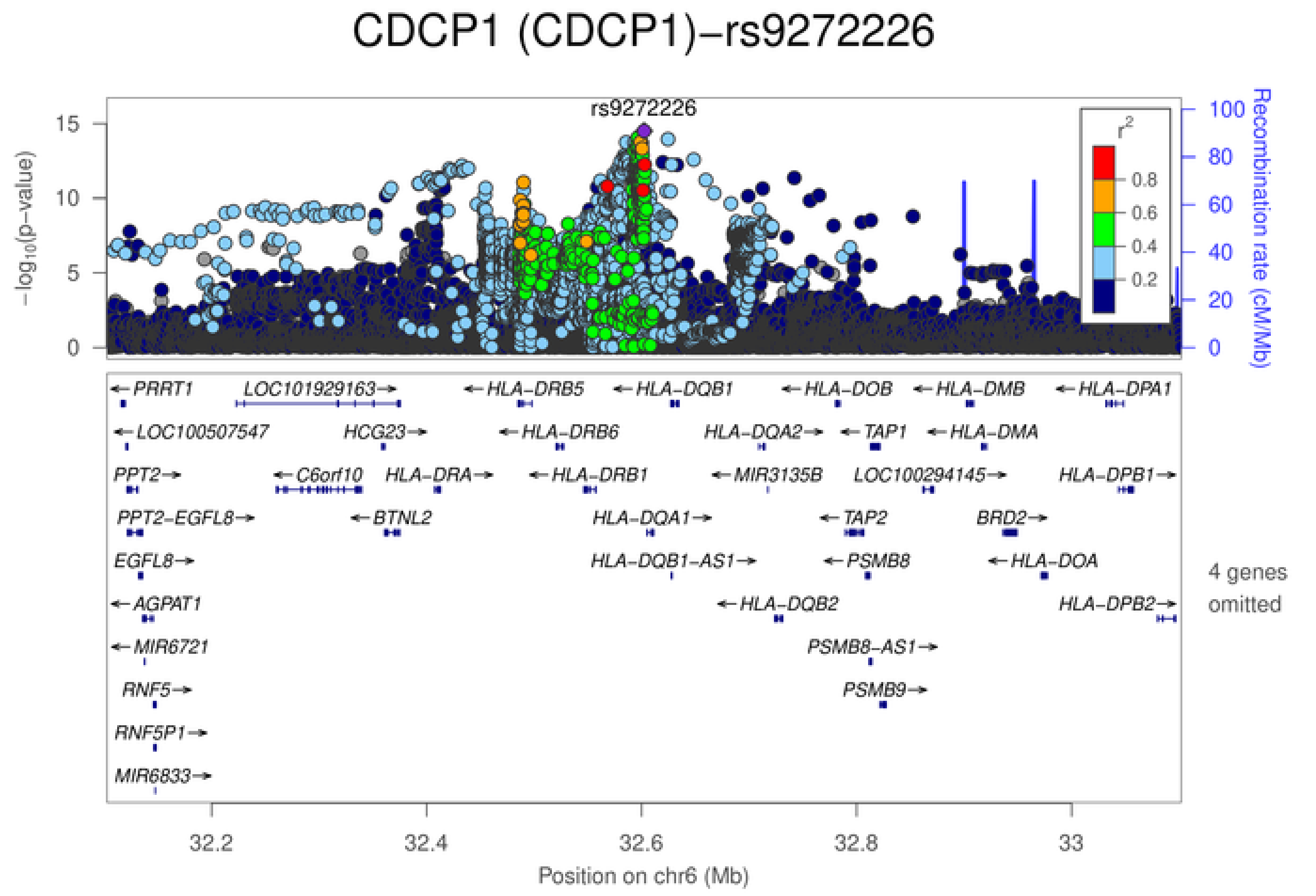
CDCP1 (CDCP1)−rs9272226
rs9272226
$-\log_{10}$(p−value)
Recombination rate (cM/Mb)
$r^2$
0.8
0.6
0.4
0.2
PRRT1
LOC101929163
HLA−DRB5
HLA−DQB1
HLA−DOB
HLA−DMB
HLA−DPA1
LOC100507547
HCG23
HLA−DRB6
HLA−DQA2
TAP1
HLA−DMA
PPT2
C6orf10
HLA−DRA
HLA−DRB1
MIR3135B
LOC100294145
HLA−DPB1
PPT2−EGFL8
BTNL2
HLA−DQA1
TAP2
BRD2
EGFL8
HLA−DQB1−AS1
PSMB8
HLA−DOA
AGPAT1
HLA−DQB2
HLA−DPB2
4 genes omitted
MIR6721
PSMB8−AS1
RNF5
PSMB9
RNF5P1
MIR6833
32.2
32.4
32.6
32.8
33
Position on chr6 (Mb)

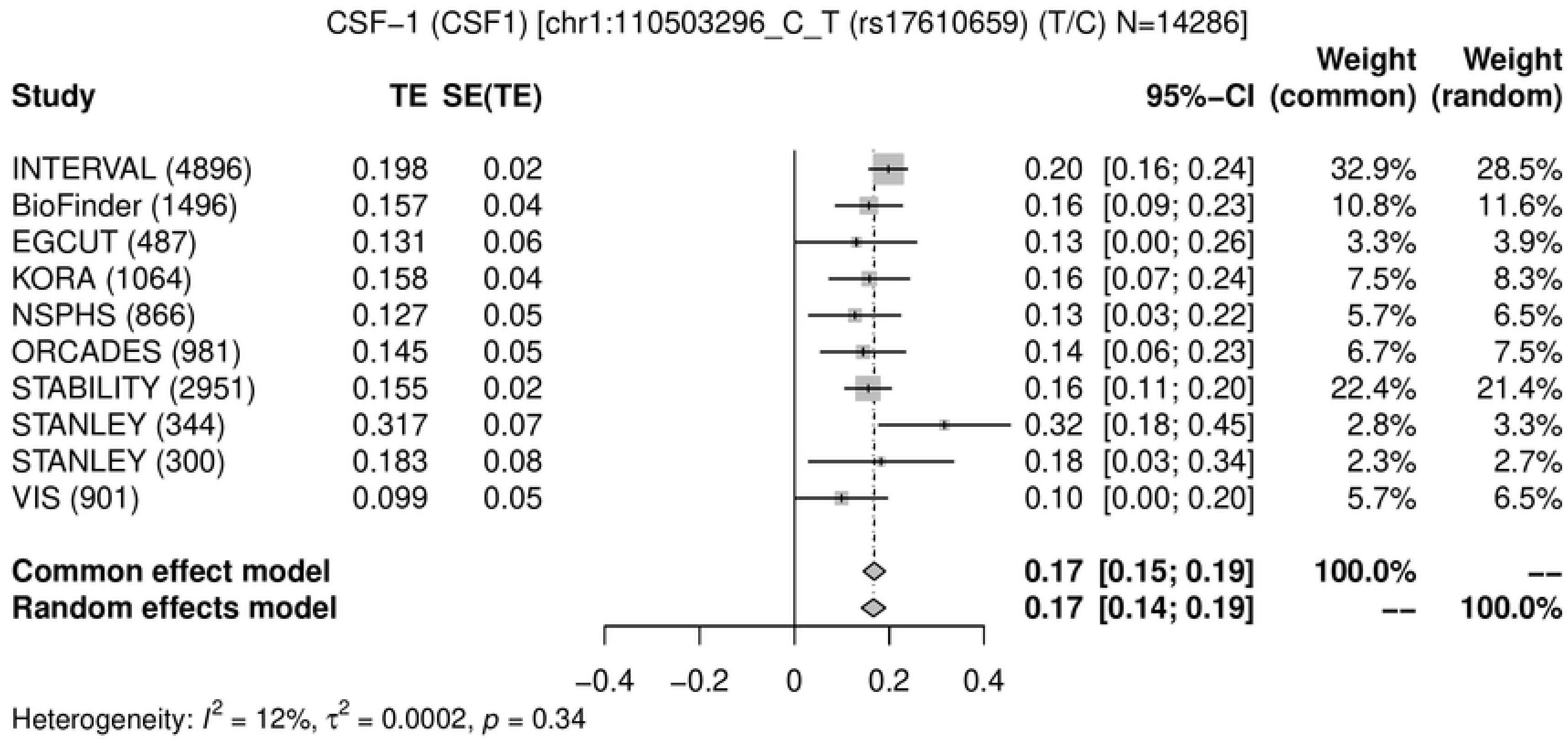
CSF−1 (CSF1) [chr1:110503296_C_T (rs17610659) (T/C) N=14286]
Study
TE
SE(TE)
95%−CI
Weight (common)
Weight (random)
INTERVAL (4896) 0.198 0.02 0.20 [0.16; 0.24] 32.9% 28.5%
BioFinder (1496) 0.157 0.04 0.16 [0.09; 0.23] 10.8% 11.6%
EGCUT (487) 0.131 0.06 0.13 [0.00; 0.26] 3.3% 3.9%
KORA (1064) 0.158 0.04 0.16 [0.07; 0.24] 7.5% 8.3%
NSPHS (866) 0.127 0.05 0.13 [0.03; 0.22] 5.7% 6.5%
ORCADES (981) 0.145 0.05 0.14 [0.06; 0.23] 6.7% 7.5%
STABILITY (2951) 0.155 0.02 0.16 [0.11; 0.20] 22.4% 21.4%
STANLEY (344) 0.317 0.07 0.32 [0.18; 0.45] 2.8% 3.3%
STANLEY (300) 0.183 0.08 0.18 [0.03; 0.34] 2.3% 2.7%
VIS (901) 0.099 0.05 0.10 [0.00; 0.20] 5.7% 6.5%
Common effect model 0.17 [0.15; 0.19] 100.0% --
Random effects model 0.17 [0.14; 0.19] -- 100.0%
−0.4 −0.2 0 0.2 0.4
Heterogeneity: $I^2$ = 12%, $\tau^2$ = 0.0002, $p$ = 0.34

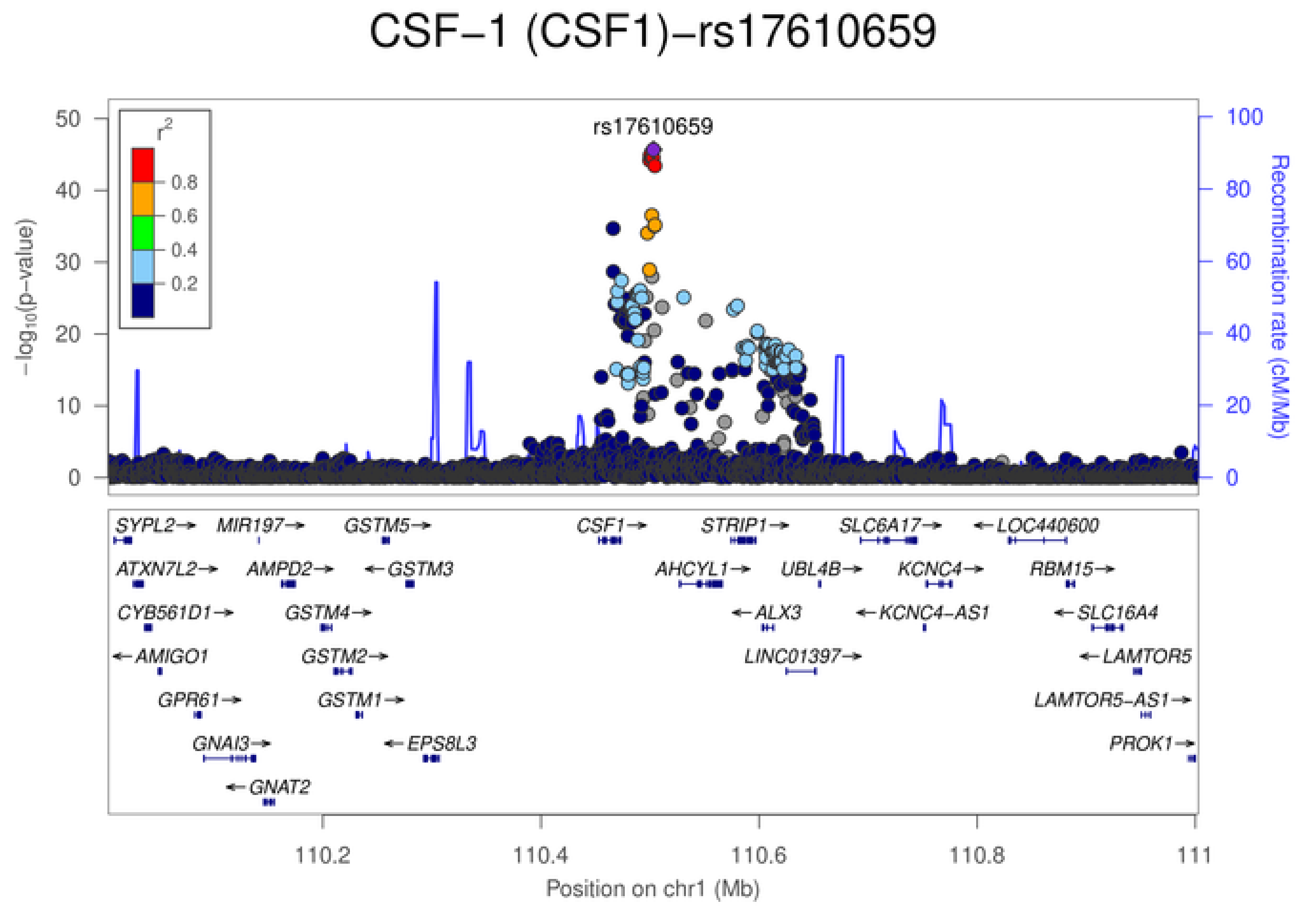
CSF−1 (CSF1)−rs17610659
rs17610659
$r^2$
0.8
0.6
0.4
0.2
$-\log_{10}$(p−value)
Recombination rate (cM/Mb)
50
40
30
20
10
0
100
80
60
40
20
0
SYPL2→
MIR197→
GSTM5→
CSF1→
STRIP1→
SLC6A17→
←LOC440600
ATXN7L2→
AMPD2→
←GSTM3
AHCYL1→
UBL4B→
KCNC4→
RBM15→
CYB561D1→
GSTM4→
←ALX3
←KCNC4−AS1
←SLC16A4
←AMIGO1
GSTM2→
LINC01397→
←LAMTOR5
GPR61→
GSTM1→
LAMTOR5−AS1→
GNAI3→
←EPS8L3
PROK1→
←GNAT2
110.2
110.4
110.6
110.8
111
Position on chr1 (Mb)

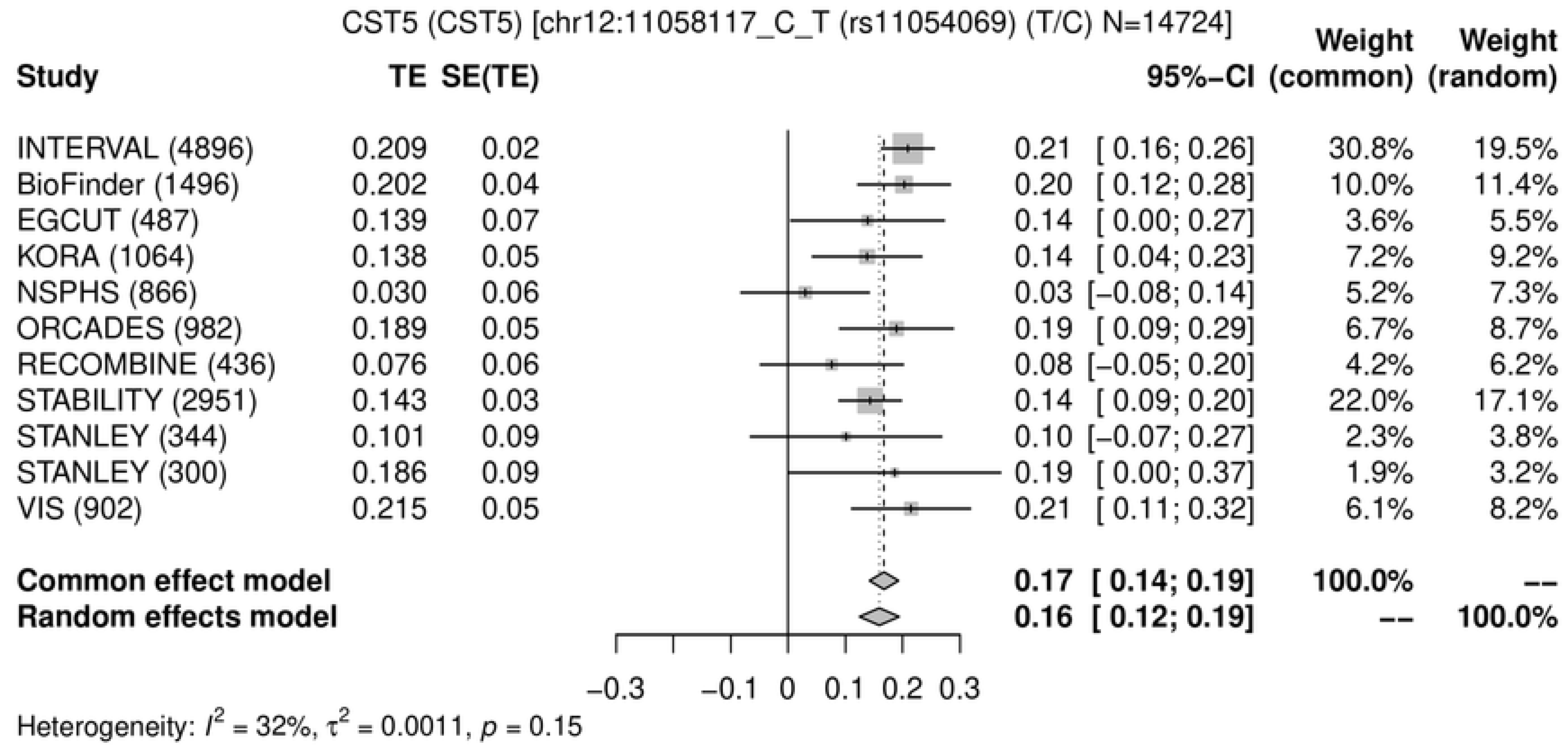
CST5 (CST5) [chr12:11058117_C_T (rs11054069) (T/C) N=14724]
Study
TE
SE(TE)
95%−CI
Weight (common)
Weight (random)
INTERVAL (4896) 0.209 0.02 0.21 [ 0.16; 0.26] 30.8% 19.5%
BioFinder (1496) 0.202 0.04 0.20 [ 0.12; 0.28] 10.0% 11.4%
EGCUT (487) 0.139 0.07 0.14 [ 0.00; 0.27] 3.6% 5.5%
KORA (1064) 0.138 0.05 0.14 [ 0.04; 0.23] 7.2% 9.2%
NSPHS (866) 0.030 0.06 0.03 [−0.08; 0.14] 5.2% 7.3%
ORCADES (982) 0.189 0.05 0.19 [ 0.09; 0.29] 6.7% 8.7%
RECOMBINE (436) 0.076 0.06 0.08 [−0.05; 0.20] 4.2% 6.2%
STABILITY (2951) 0.143 0.03 0.14 [ 0.09; 0.20] 22.0% 17.1%
STANLEY (344) 0.101 0.09 0.10 [−0.07; 0.27] 2.3% 3.8%
STANLEY (300) 0.186 0.09 0.19 [ 0.00; 0.37] 1.9% 3.2%
VIS (902) 0.215 0.05 0.21 [ 0.11; 0.32] 6.1% 8.2%
Common effect model 0.17 [ 0.14; 0.19] 100.0% --
Random effects model 0.16 [ 0.12; 0.19] -- 100.0%
−0.3 −0.1 0 0.1 0.2 0.3
Heterogeneity: $I^2$ = 32%, $\tau^2$ = 0.0011, $p$ = 0.15

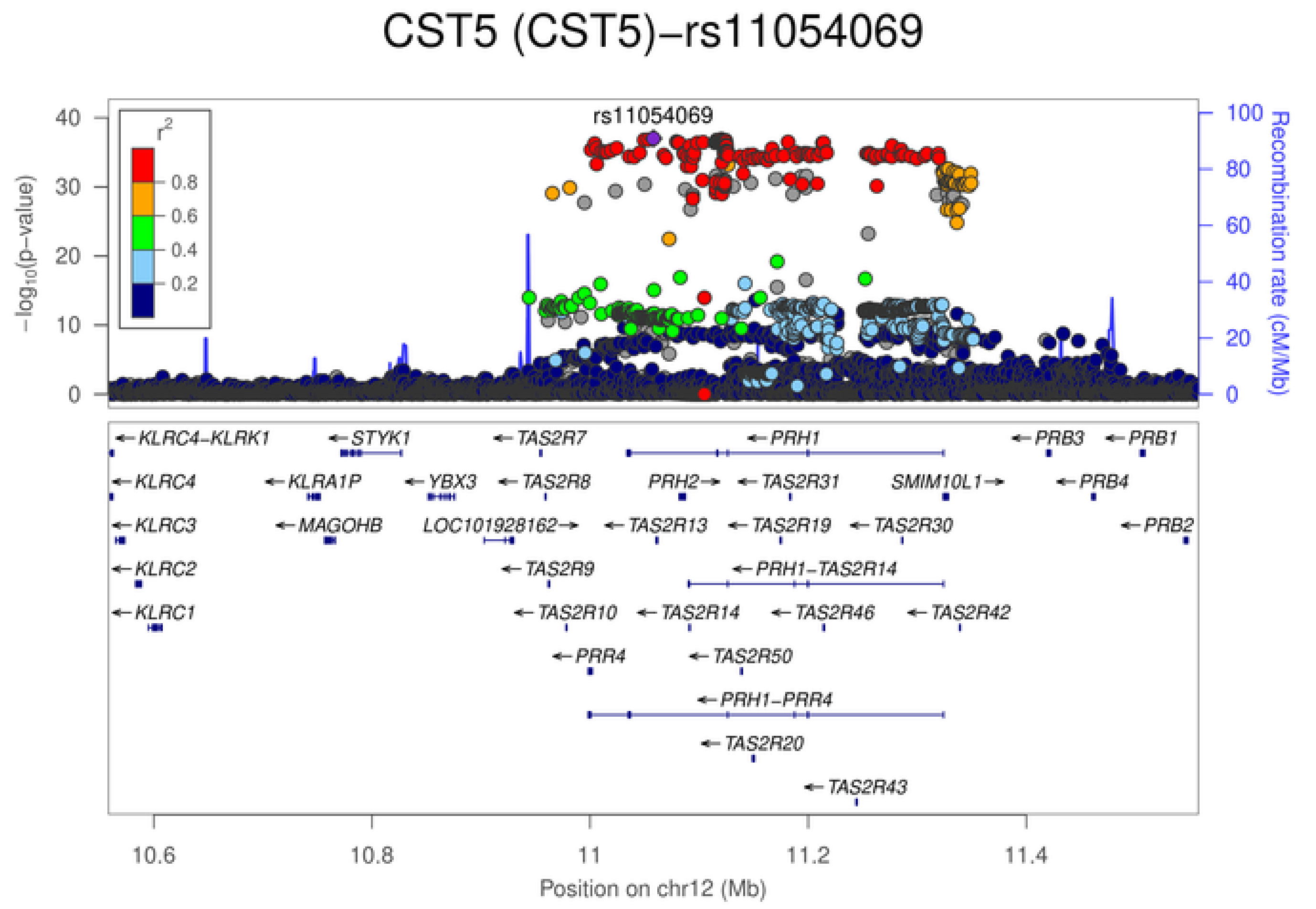
CST5 (CST5)−rs11054069
rs11054069
$r^2$
0.8
0.6
0.4
0.2
$-\log_{10}$(p−value)
Recombination rate (cM/Mb)
KLRC4−KLRK1
STYK1
TAS2R7
PRH1
PRB3
PRB1
KLRC4
KLRA1P
YBX3
TAS2R8
PRH2
TAS2R31
SMIM10L1
PRB4
KLRC3
MAGOHB
LOC101928162
TAS2R13
TAS2R19
TAS2R30
PRB2
KLRC2
TAS2R9
PRH1−TAS2R14
KLRC1
TAS2R10
TAS2R14
TAS2R46
TAS2R42
PRR4
TAS2R50
PRH1−PRR4
TAS2R20
TAS2R43
10.6
10.8
11
11.2
11.4
Position on chr12 (Mb)

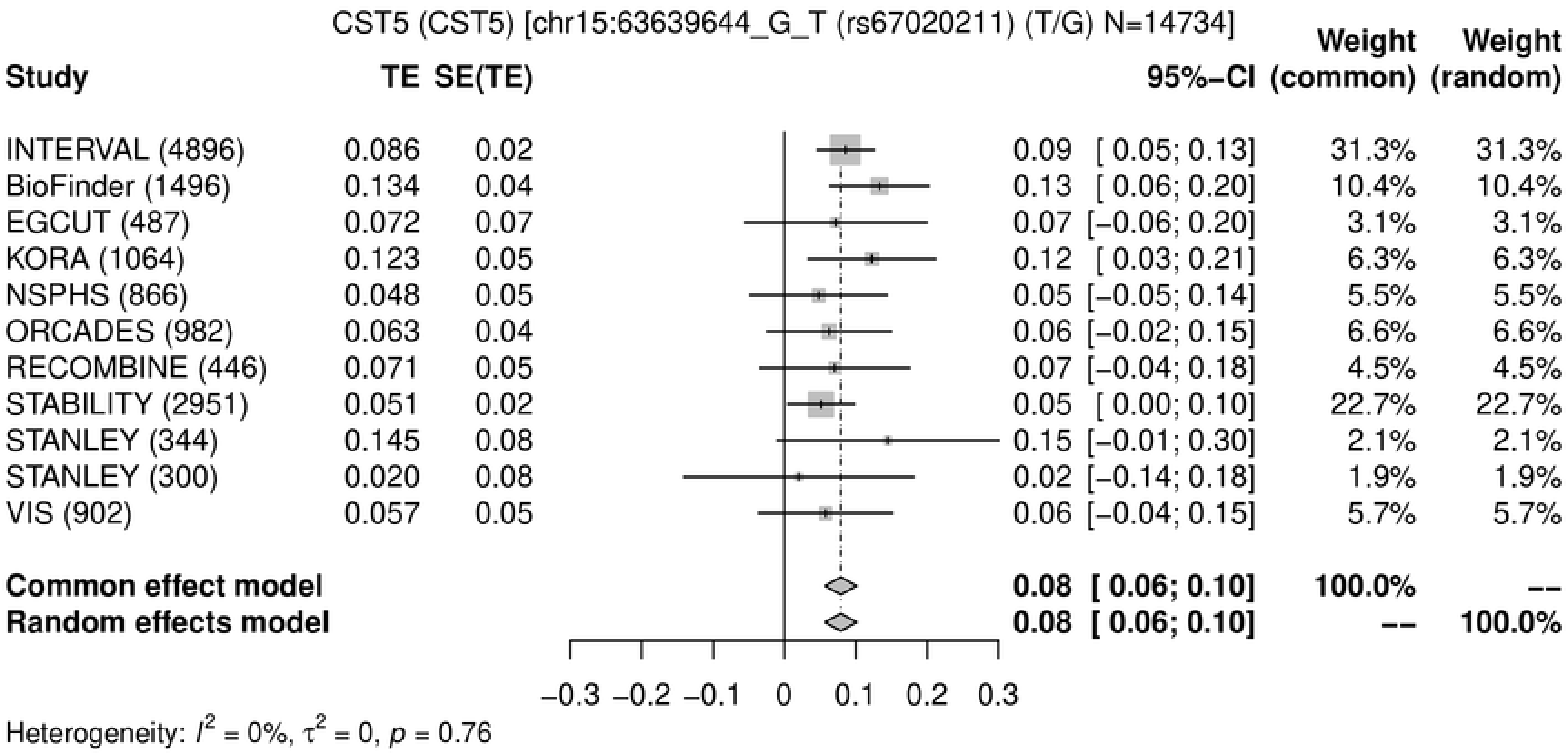
CST5 (CST5) [chr15:63639644_G_T (rs67020211) (T/G) N=14734]
Study
TE SE(TE)
95%−CI
Weight (common)
Weight (random)
INTERVAL (4896) 0.086 0.02 0.09 [ 0.05; 0.13] 31.3% 31.3%
BioFinder (1496) 0.134 0.04 0.13 [ 0.06; 0.20] 10.4% 10.4%
EGCUT (487) 0.072 0.07 0.07 [−0.06; 0.20] 3.1% 3.1%
KORA (1064) 0.123 0.05 0.12 [ 0.03; 0.21] 6.3% 6.3%
NSPHS (866) 0.048 0.05 0.05 [−0.05; 0.14] 5.5% 5.5%
ORCADES (982) 0.063 0.04 0.06 [−0.02; 0.15] 6.6% 6.6%
RECOMBINE (446) 0.071 0.05 0.07 [−0.04; 0.18] 4.5% 4.5%
STABILITY (2951) 0.051 0.02 0.05 [ 0.00; 0.10] 22.7% 22.7%
STANLEY (344) 0.145 0.08 0.15 [−0.01; 0.30] 2.1% 2.1%
STANLEY (300) 0.020 0.08 0.02 [−0.14; 0.18] 1.9% 1.9%
VIS (902) 0.057 0.05 0.06 [−0.04; 0.15] 5.7% 5.7%
Common effect model 0.08 [ 0.06; 0.10] 100.0% --
Random effects model 0.08 [ 0.06; 0.10] -- 100.0%
−0.3 −0.2 −0.1 0 0.1 0.2 0.3
Heterogeneity: $I^2$ = 0%, $\tau^2$ = 0, $p$ = 0.76

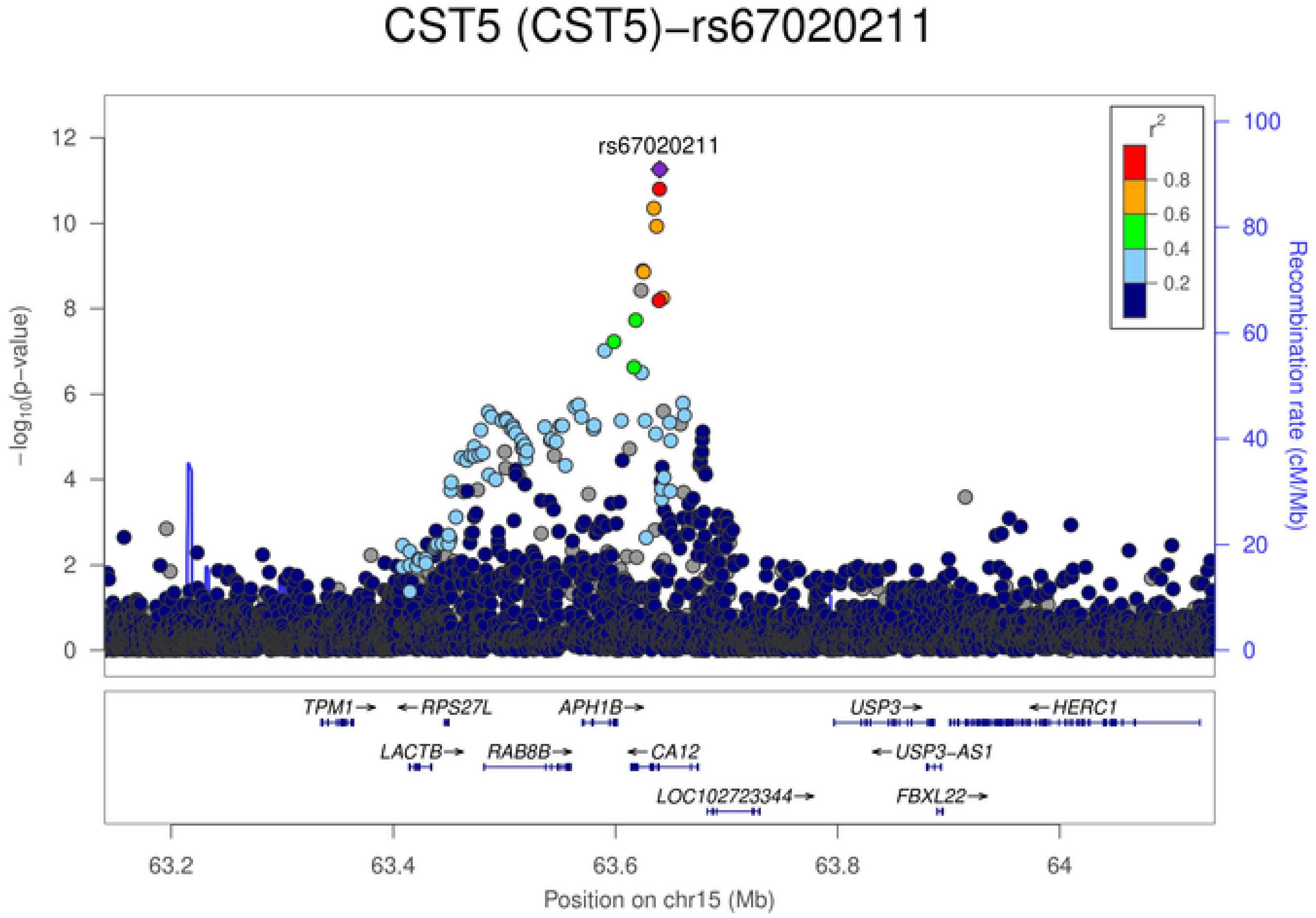
CST5 (CST5)−rs67020211
rs67020211
$-\log_{10}$(p−value)
Recombination rate (cM/Mb)
$r^2$
0.8
0.6
0.4
0.2
TPM1→
←RPS27L
APH1B→
USP3→
←HERC1
LACTB→
RAB8B→
←CA12
←USP3−AS1
LOC102723344→
FBXL22→
63.2
63.4
63.6
63.8
64
Position on chr15 (Mb)

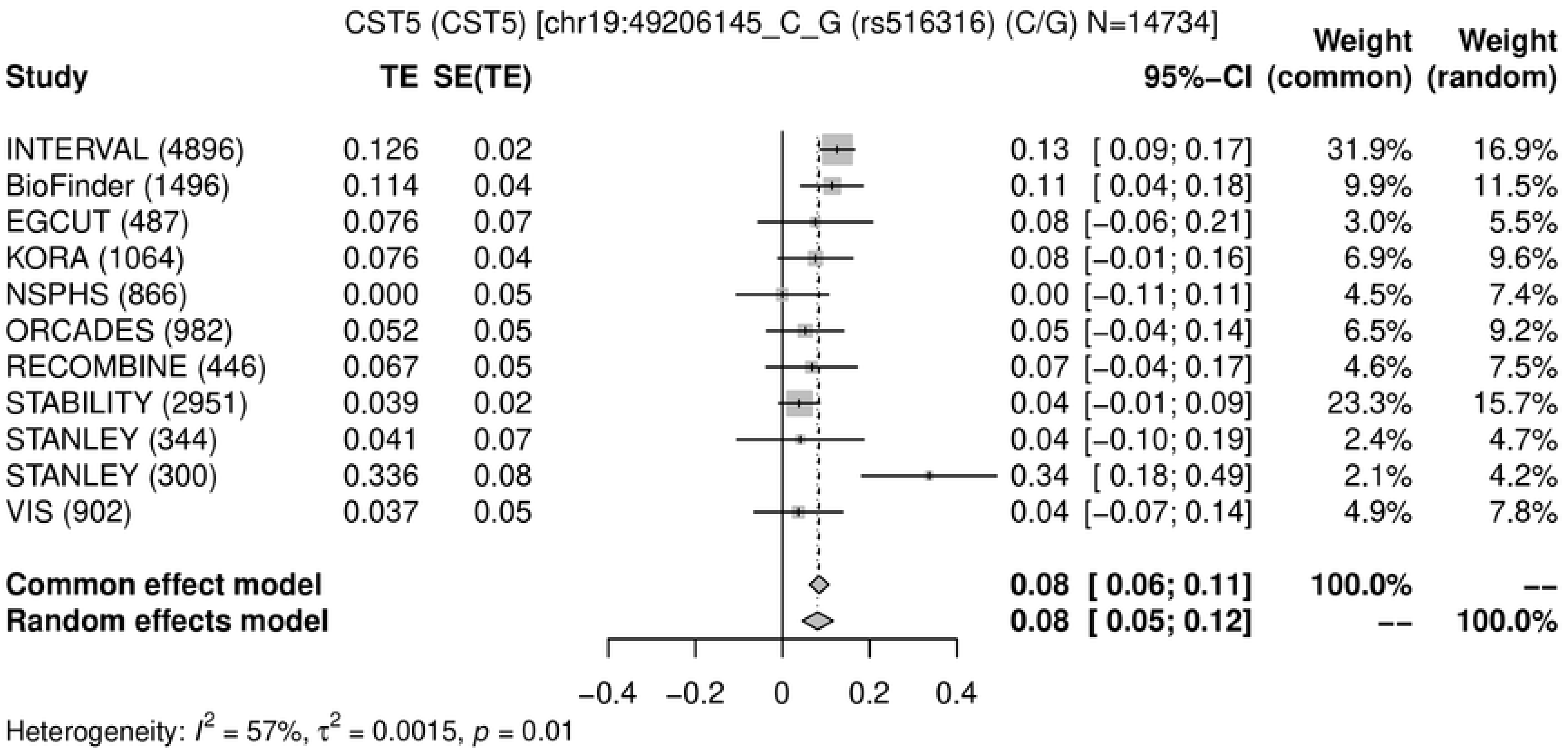

CST5 (CST5) [chr19:49206145_C_G (rs516316) (C/G) N=14734]
Study TE SE(TE) 95%−CI Weight (common) Weight (random)
INTERVAL (4896) 0.126 0.02 0.13 [ 0.09; 0.17] 31.9% 16.9%
BioFinder (1496) 0.114 0.04 0.11 [ 0.04; 0.18] 9.9% 11.5%
EGCUT (487) 0.076 0.07 0.08 [−0.06; 0.21] 3.0% 5.5%
KORA (1064) 0.076 0.04 0.08 [−0.01; 0.16] 6.9% 9.6%
NSPHS (866) 0.000 0.05 0.00 [−0.11; 0.11] 4.5% 7.4%
ORCADES (982) 0.052 0.05 0.05 [−0.04; 0.14] 6.5% 9.2%
RECOMBINE (446) 0.067 0.05 0.07 [−0.04; 0.17] 4.6% 7.5%
STABILITY (2951) 0.039 0.02 0.04 [−0.01; 0.09] 23.3% 15.7%
STANLEY (344) 0.041 0.07 0.04 [−0.10; 0.19] 2.4% 4.7%
STANLEY (300) 0.336 0.08 0.34 [ 0.18; 0.49] 2.1% 4.2%
VIS (902) 0.037 0.05 0.04 [−0.07; 0.14] 4.9% 7.8%
Common effect model 0.08 [ 0.06; 0.11] 100.0% --
Random effects model 0.08 [ 0.05; 0.12] -- 100.0%
−0.4 −0.2 0 0.2 0.4
Heterogeneity: $I^2$ = 57%, $\tau^2$ = 0.0015, $p$ = 0.01


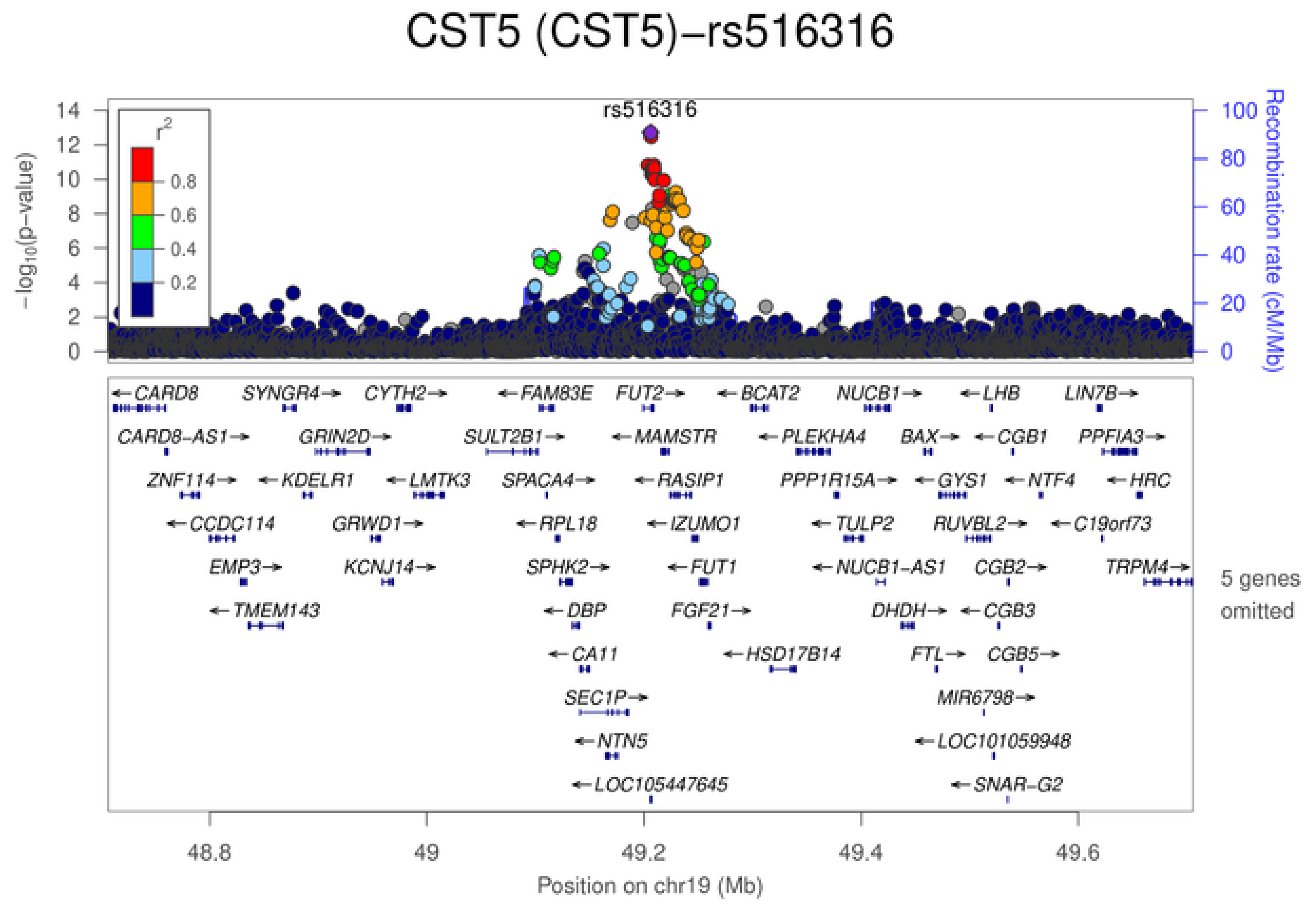

CST5 (CST5)−rs516316
rs516316
$-\log_{10}(p\text{-value})$
Recombination rate (cM/Mb)
$r^2$
0.8
0.6
0.4
0.2
5 genes omitted
48.8
49
49.2
49.4
49.6
Position on chr19 (Mb)

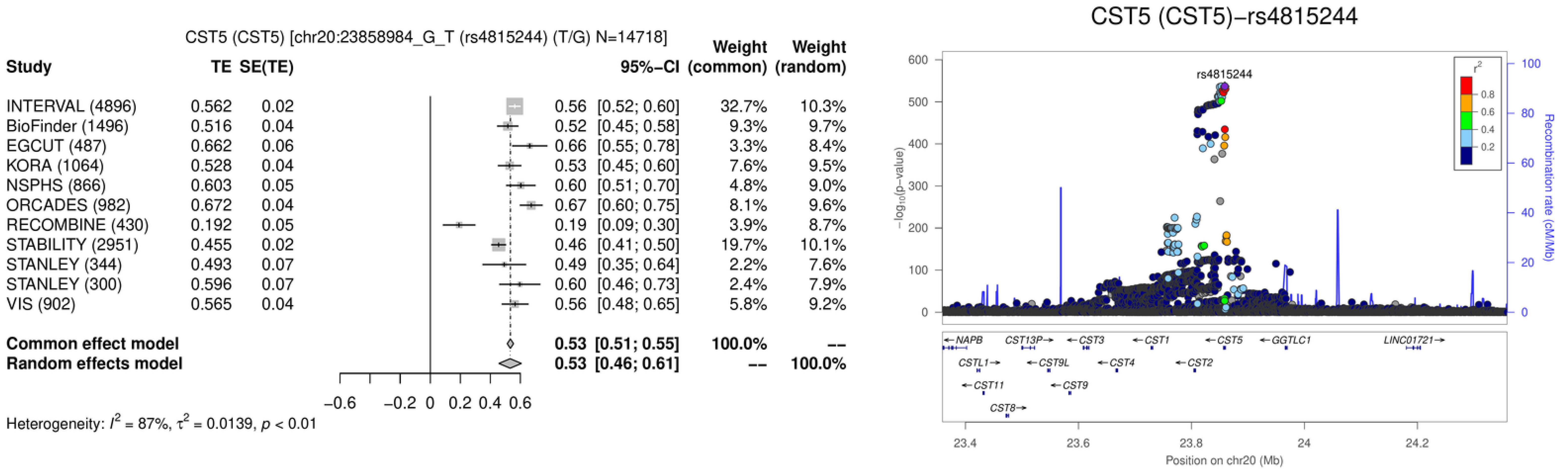
CST5 (CST5) [chr20:23858984_G_T (rs4815244) (T/G) N=14718]
Study
TE
SE(TE)
95%−CI
Weight (common)
Weight (random)
INTERVAL (4896) 0.562 0.02 0.56 [0.52; 0.60] 32.7% 10.3%
BioFinder (1496) 0.516 0.04 0.52 [0.45; 0.58] 9.3% 9.7%
EGCUT (487) 0.662 0.06 0.66 [0.55; 0.78] 3.3% 8.4%
KORA (1064) 0.528 0.04 0.53 [0.45; 0.60] 7.6% 9.5%
NSPHS (866) 0.603 0.05 0.60 [0.51; 0.70] 4.8% 9.0%
ORCADES (982) 0.672 0.04 0.67 [0.60; 0.75] 8.1% 9.6%
RECOMBINE (430) 0.192 0.05 0.19 [0.09; 0.30] 3.9% 8.7%
STABILITY (2951) 0.455 0.02 0.46 [0.41; 0.50] 19.7% 10.1%
STANLEY (344) 0.493 0.07 0.49 [0.35; 0.64] 2.2% 7.6%
STANLEY (300) 0.596 0.07 0.60 [0.46; 0.73] 2.4% 7.9%
VIS (902) 0.565 0.04 0.56 [0.48; 0.65] 5.8% 9.2%
Common effect model 0.53 [0.51; 0.55] 100.0% --
Random effects model 0.53 [0.46; 0.61] -- 100.0%
−0.6 −0.2 0 0.2 0.4 0.6
Heterogeneity: $I^2$ = 87%, $\tau^2$ = 0.0139, $p$ < 0.01
CST5 (CST5)−rs4815244
rs4815244
$-\log_{10}$(p−value)
Recombination rate (cM/Mb)
$r^2$
0.8
0.6
0.4
0.2
←NAPB
CST13P→
←CST3
←CST1
←CST5
←GGTLC1
LINC01721→
CSTL1→
←CST9L
←CST4
←CST2
←CST11
←CST9
CST8→
23.4 23.6 23.8 24 24.2
Position on chr20 (Mb)

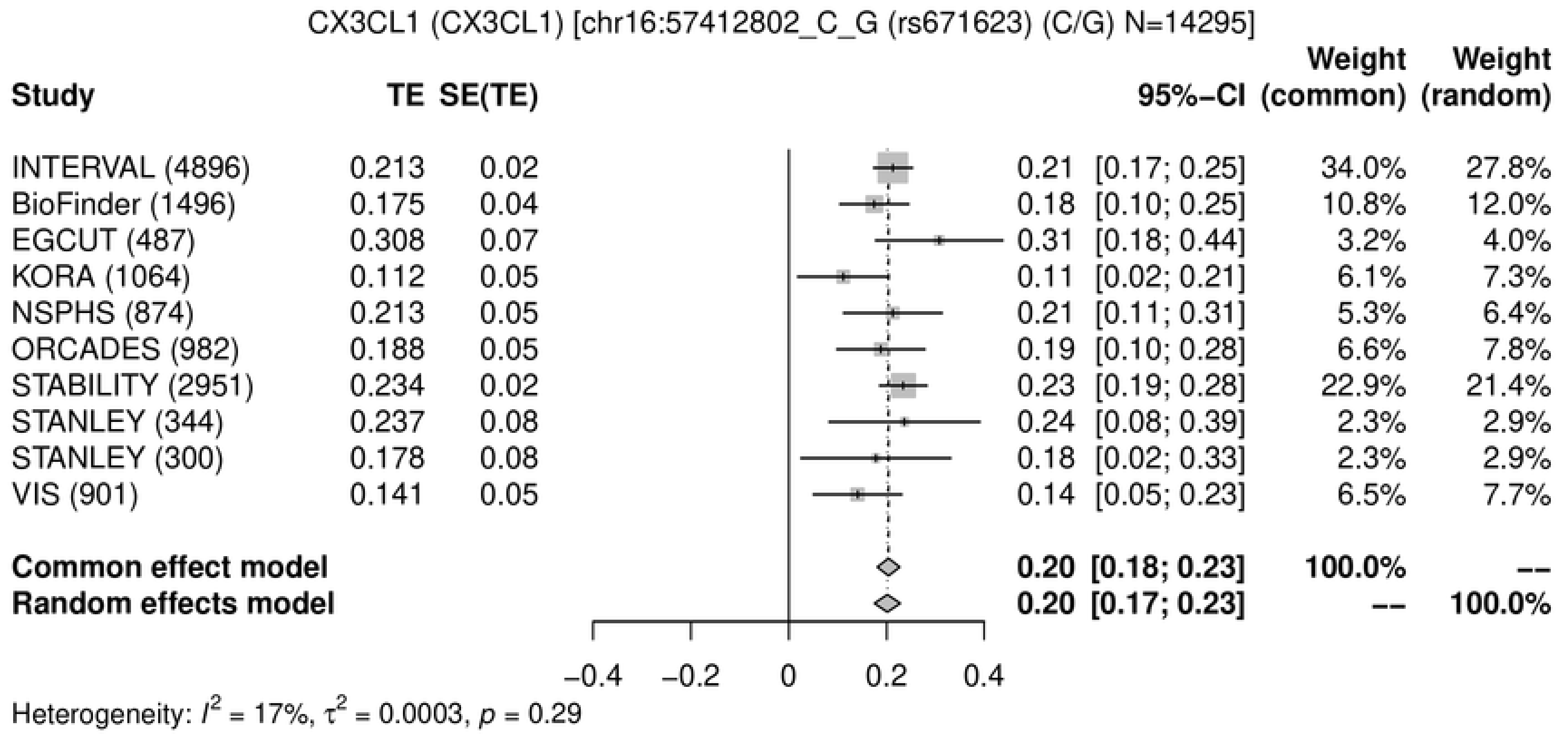
CX3CL1 (CX3CL1) [chr16:57412802_C_G (rs671623) (C/G) N=14295]
Study
TE
SE(TE)
95%−CI
Weight (common)
Weight (random)
INTERVAL (4896) 0.213 0.02 0.21 [0.17; 0.25] 34.0% 27.8%
BioFinder (1496) 0.175 0.04 0.18 [0.10; 0.25] 10.8% 12.0%
EGCUT (487) 0.308 0.07 0.31 [0.18; 0.44] 3.2% 4.0%
KORA (1064) 0.112 0.05 0.11 [0.02; 0.21] 6.1% 7.3%
NSPHS (874) 0.213 0.05 0.21 [0.11; 0.31] 5.3% 6.4%
ORCADES (982) 0.188 0.05 0.19 [0.10; 0.28] 6.6% 7.8%
STABILITY (2951) 0.234 0.02 0.23 [0.19; 0.28] 22.9% 21.4%
STANLEY (344) 0.237 0.08 0.24 [0.08; 0.39] 2.3% 2.9%
STANLEY (300) 0.178 0.08 0.18 [0.02; 0.33] 2.3% 2.9%
VIS (901) 0.141 0.05 0.14 [0.05; 0.23] 6.5% 7.7%
Common effect model 0.20 [0.18; 0.23] 100.0% --
Random effects model 0.20 [0.17; 0.23] -- 100.0%
−0.4 −0.2 0 0.2 0.4
Heterogeneity: $I^2$ = 17%, $\tau^2$ = 0.0003, $p$ = 0.29

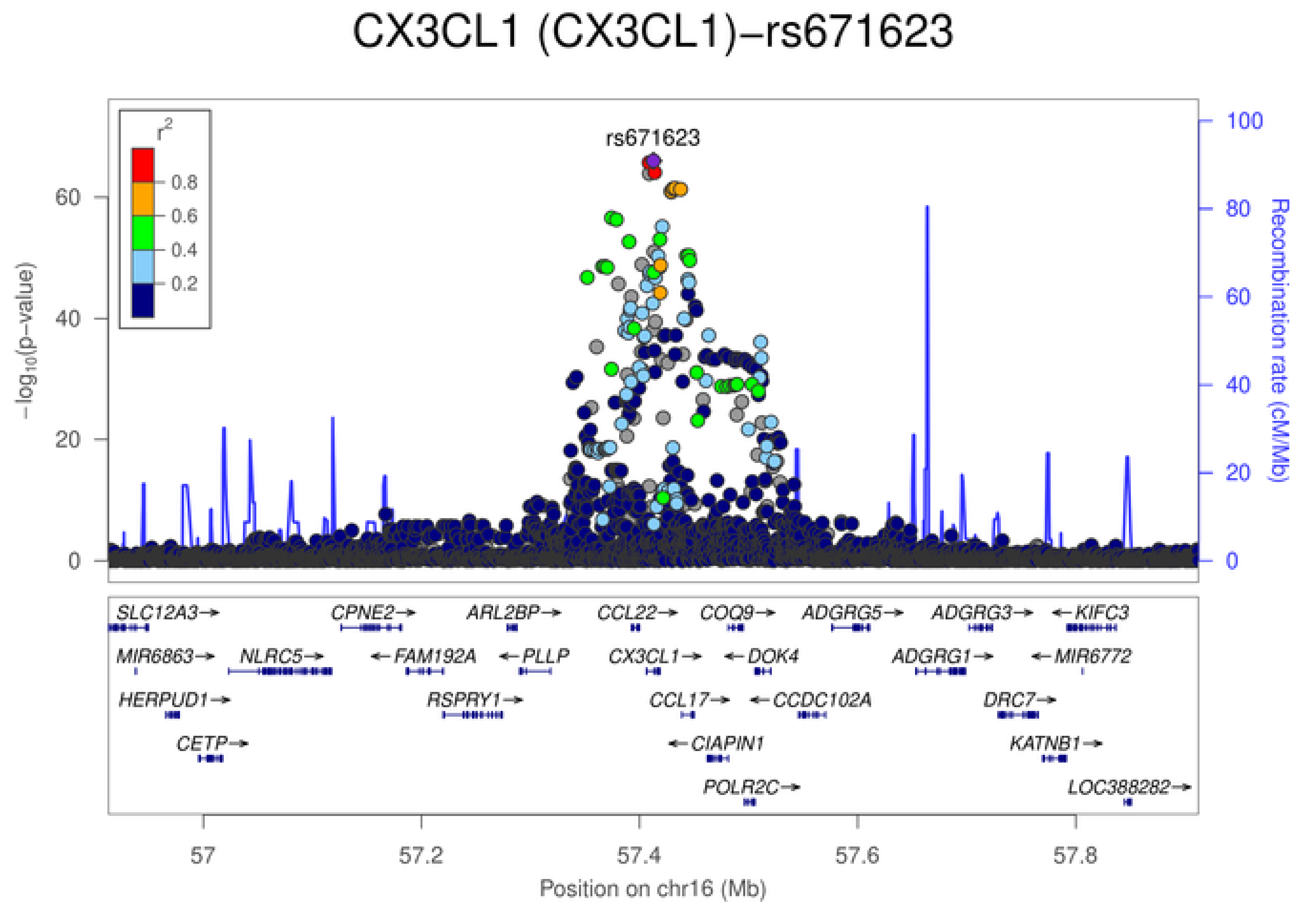
CX3CL1 (CX3CL1)−rs671623
rs671623
$r^2$
0.8
0.6
0.4
0.2
$-\log_{10}$(p−value)
Recombination rate (cM/Mb)
SLC12A3→
CPNE2→
ARL2BP→
CCL22→
COQ9→
ADGRG5→
ADGRG3→
←KIFC3
MIR6863→
NLRC5→
←FAM192A
←PLLP
CX3CL1→
←DOK4
ADGRG1→
←MIR6772
HERPUD1→
RSPRY1→
CCL17→
←CCDC102A
DRC7→
CETP→
←CIAPIN1
KATNB1→
POLR2C→
LOC388282→
57 57.2 57.4 57.6 57.8
Position on chr16 (Mb)

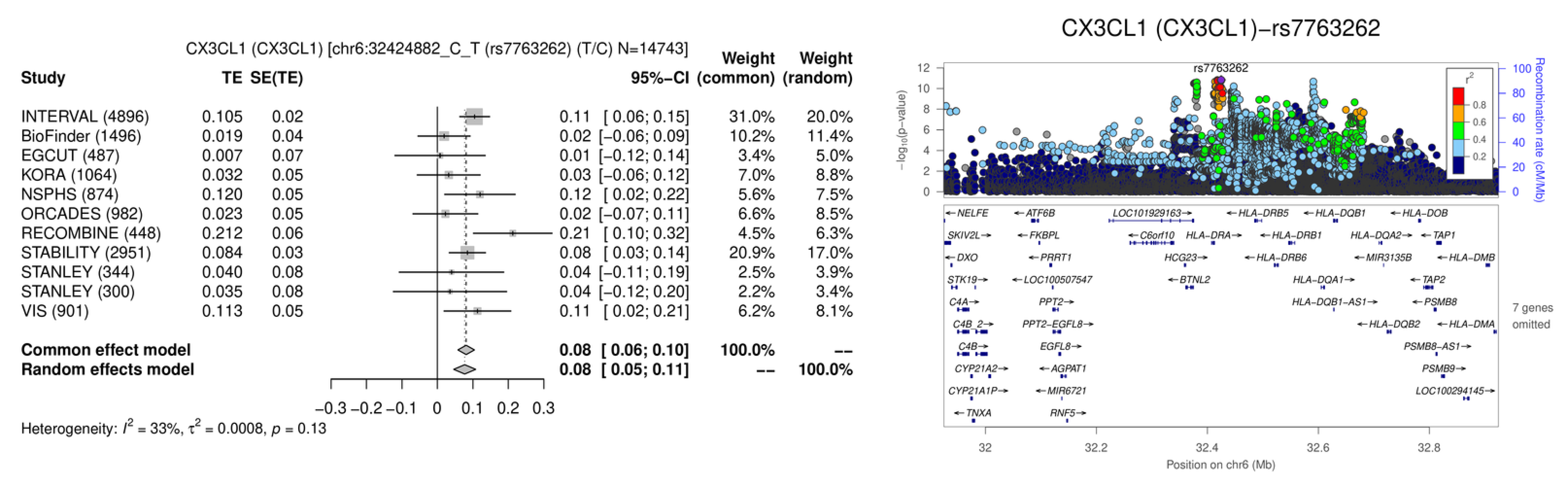
CX3CL1 (CX3CL1) [chr6:32424882_C_T (rs7763262) (T/C) N=14743]
Study
TE
SE(TE)
95%−CI
Weight (common)
Weight (random)
INTERVAL (4896) 0.105 0.02 0.11 [ 0.06; 0.15] 31.0% 20.0%
BioFinder (1496) 0.019 0.04 0.02 [−0.06; 0.09] 10.2% 11.4%
EGCUT (487) 0.007 0.07 0.01 [−0.12; 0.14] 3.4% 5.0%
KORA (1064) 0.032 0.05 0.03 [−0.06; 0.12] 7.0% 8.8%
NSPHS (874) 0.120 0.05 0.12 [ 0.02; 0.22] 5.6% 7.5%
ORCADES (982) 0.023 0.05 0.02 [−0.07; 0.11] 6.6% 8.5%
RECOMBINE (448) 0.212 0.06 0.21 [ 0.10; 0.32] 4.5% 6.3%
STABILITY (2951) 0.084 0.03 0.08 [ 0.03; 0.14] 20.9% 17.0%
STANLEY (344) 0.040 0.08 0.04 [−0.11; 0.19] 2.5% 3.9%
STANLEY (300) 0.035 0.08 0.04 [−0.12; 0.20] 2.2% 3.4%
VIS (901) 0.113 0.05 0.11 [ 0.02; 0.21] 6.2% 8.1%
Common effect model 0.08 [ 0.06; 0.10] 100.0% --
Random effects model 0.08 [ 0.05; 0.11] -- 100.0%
−0.3 −0.2 −0.1 0 0.1 0.2 0.3
Heterogeneity: $I^2$ = 33%, $\tau^2$ = 0.0008, $p$ = 0.13
CX3CL1 (CX3CL1)−rs7763262
rs7763262
$-\log_{10}$(p−value)
Recombination rate (cM/Mb)
$r^2$
0.8
0.6
0.4
0.2
NELFE ATF6B LOC101929163 HLA-DRB5 HLA-DQB1 HLA-DOB
SKIV2L FKBPL C6orf10 HLA-DRA HLA-DRB1 HLA-DQA2 TAP1
DXO PRRT1 HCG23 HLA-DRB6 MIR3135B HLA-DMB
STK19 LOC100507547 BTNL2 HLA-DQA1 TAP2
C4A PPT2 HLA-DQB1-AS1 PSMB8
C4B_2 PPT2-EGFL8 HLA-DQB2 HLA-DMA
C4B EGFL8 PSMB8-AS1
CYP21A2 AGPAT1 PSMB9
CYP21A1P MIR6721 LOC100294145
TNXA RNF5
7 genes omitted
32 32.2 32.4 32.6 32.8
Position on chr6 (Mb)

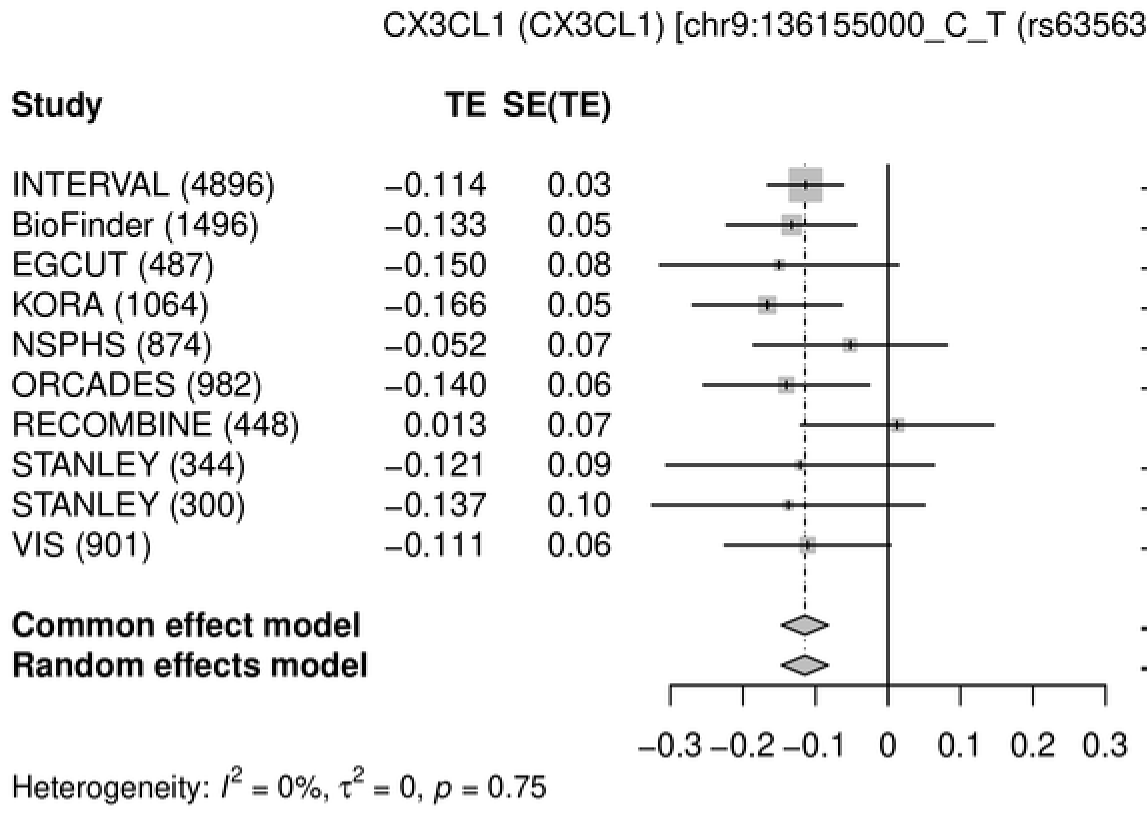
CX3CL1 (CX3CL1) [chr9:136155000_C_T (rs635634) (T/C) N=11792]
Study
TE
SE(TE)
95%-CI
Weight (common)
Weight (random)
INTERVAL (4896) −0.114 0.03 −0.11 [−0.17; −0.06] 38.8% 38.8%
BioFinder (1496) −0.133 0.05 −0.13 [−0.22; −0.04] 13.2% 13.2%
EGCUT (487) −0.150 0.08 −0.15 [−0.31; 0.01] 3.9% 3.9%
KORA (1064) −0.166 0.05 −0.17 [−0.27; −0.06] 10.0% 10.0%
NSPHS (874) −0.052 0.07 −0.05 [−0.19; 0.08] 5.9% 5.9%
ORCADES (982) −0.140 0.06 −0.14 [−0.25; −0.03] 8.1% 8.1%
RECOMBINE (448) 0.013 0.07 0.01 [−0.12; 0.15] 6.0% 6.0%
STANLEY (344) −0.121 0.09 −0.12 [−0.31; 0.06] 3.1% 3.1%
STANLEY (300) −0.137 0.10 −0.14 [−0.33; 0.05] 3.0% 3.0%
VIS (901) −0.111 0.06 −0.11 [−0.23; 0.00] 8.1% 8.1%
Common effect model −0.11 [−0.15; −0.08] 100.0% --
Random effects model −0.11 [−0.15; −0.08] -- 100.0%
−0.3 −0.2 −0.1 0 0.1 0.2 0.3
Heterogeneity: $I^2$ = 0%, $\tau^2$ = 0, $p$ = 0.75

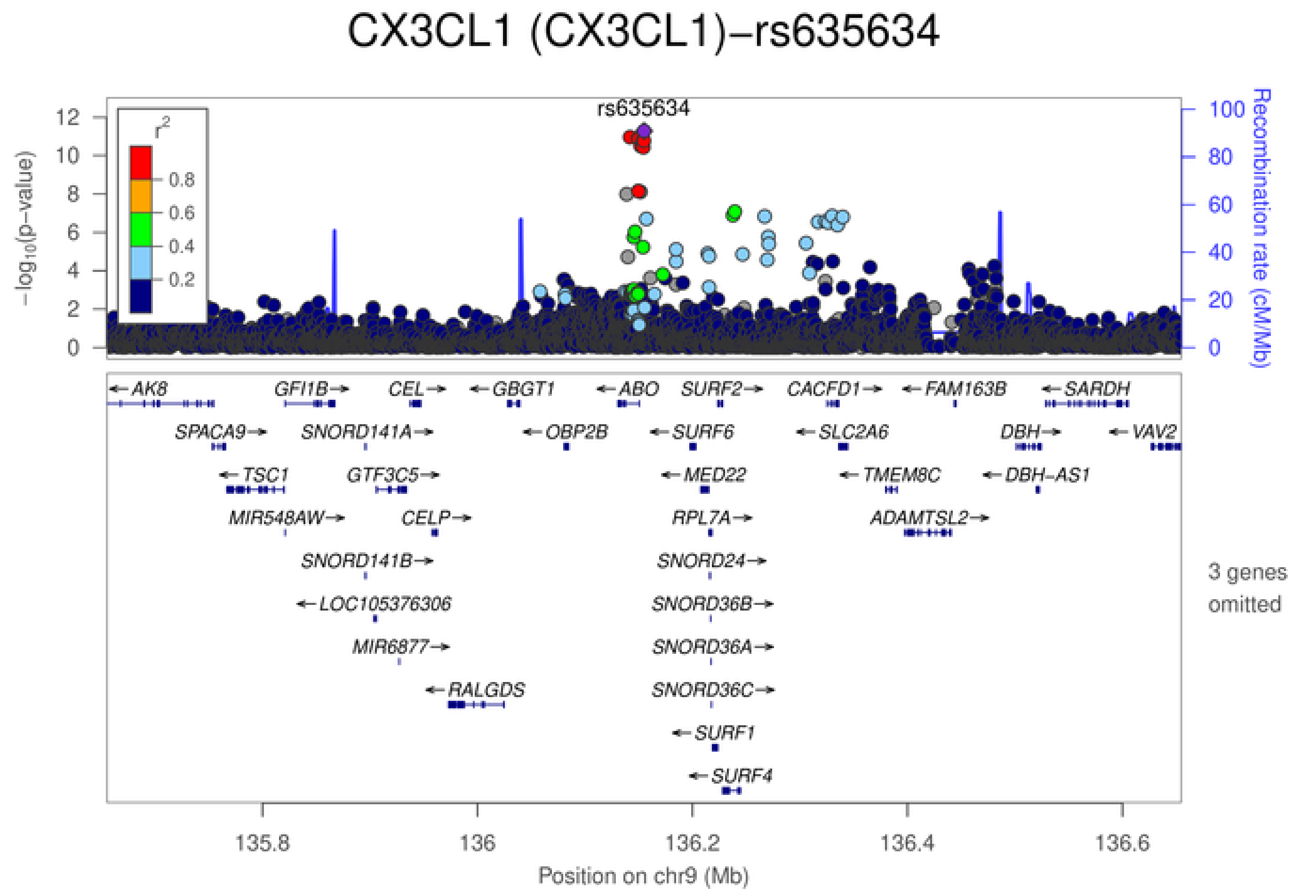
CX3CL1 (CX3CL1)−rs635634
rs635634
$-\log_{10}$(p-value)
Recombination rate (cM/Mb)
$r^2$
0.8
0.6
0.4
0.2
AK8
GFI1B
CEL
GBGT1
ABO
SURF2
CACFD1
FAM163B
SARDH
SPACA9
SNORD141A
OBP2B
SURF6
SLC2A6
DBH
VAV2
TSC1
GTF3C5
MED22
TMEM8C
DBH−AS1
MIR548AW
CELP
RPL7A
ADAMTSL2
SNORD141B
SNORD24
LOC105376306
SNORD36B
MIR6877
SNORD36A
RALGDS
SNORD36C
SURF1
SURF4
3 genes omitted
135.8
136
136.2
136.4
136.6
Position on chr9 (Mb)

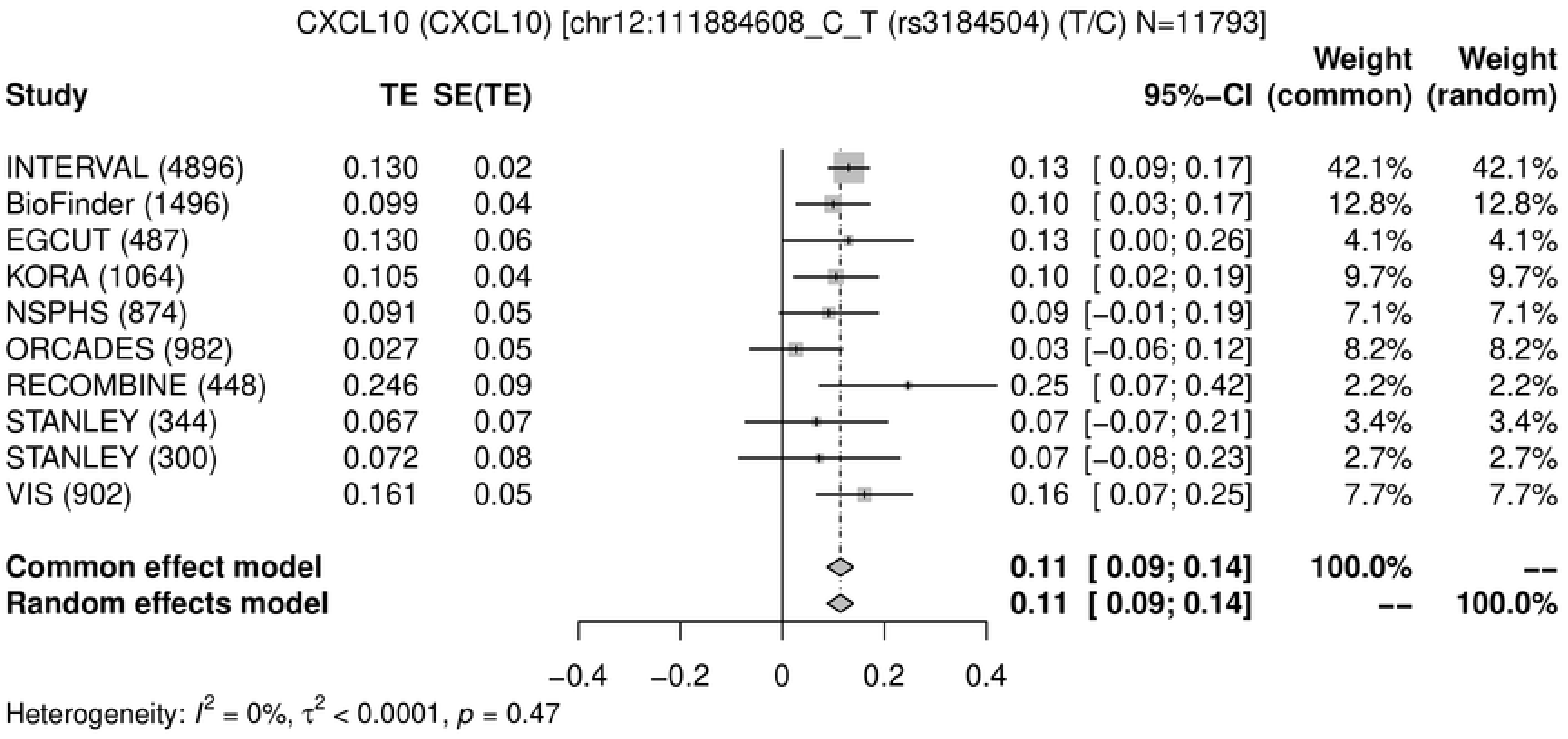
CXCL10 (CXCL10) [chr12:111884608_C_T (rs3184504) (T/C) N=11793]
Study
TE
SE(TE)
95%−CI
Weight (common)
Weight (random)
INTERVAL (4896) 0.130 0.02 0.13 [ 0.09; 0.17] 42.1% 42.1%
BioFinder (1496) 0.099 0.04 0.10 [ 0.03; 0.17] 12.8% 12.8%
EGCUT (487) 0.130 0.06 0.13 [ 0.00; 0.26] 4.1% 4.1%
KORA (1064) 0.105 0.04 0.10 [ 0.02; 0.19] 9.7% 9.7%
NSPHS (874) 0.091 0.05 0.09 [−0.01; 0.19] 7.1% 7.1%
ORCADES (982) 0.027 0.05 0.03 [−0.06; 0.12] 8.2% 8.2%
RECOMBINE (448) 0.246 0.09 0.25 [ 0.07; 0.42] 2.2% 2.2%
STANLEY (344) 0.067 0.07 0.07 [−0.07; 0.21] 3.4% 3.4%
STANLEY (300) 0.072 0.08 0.07 [−0.08; 0.23] 2.7% 2.7%
VIS (902) 0.161 0.05 0.16 [ 0.07; 0.25] 7.7% 7.7%
Common effect model 0.11 [ 0.09; 0.14] 100.0% --
Random effects model 0.11 [ 0.09; 0.14] -- 100.0%
−0.4
−0.2
0
0.2
0.4
Heterogeneity: $I^2$ = 0%, $\tau^2$ < 0.0001, $p$ = 0.47

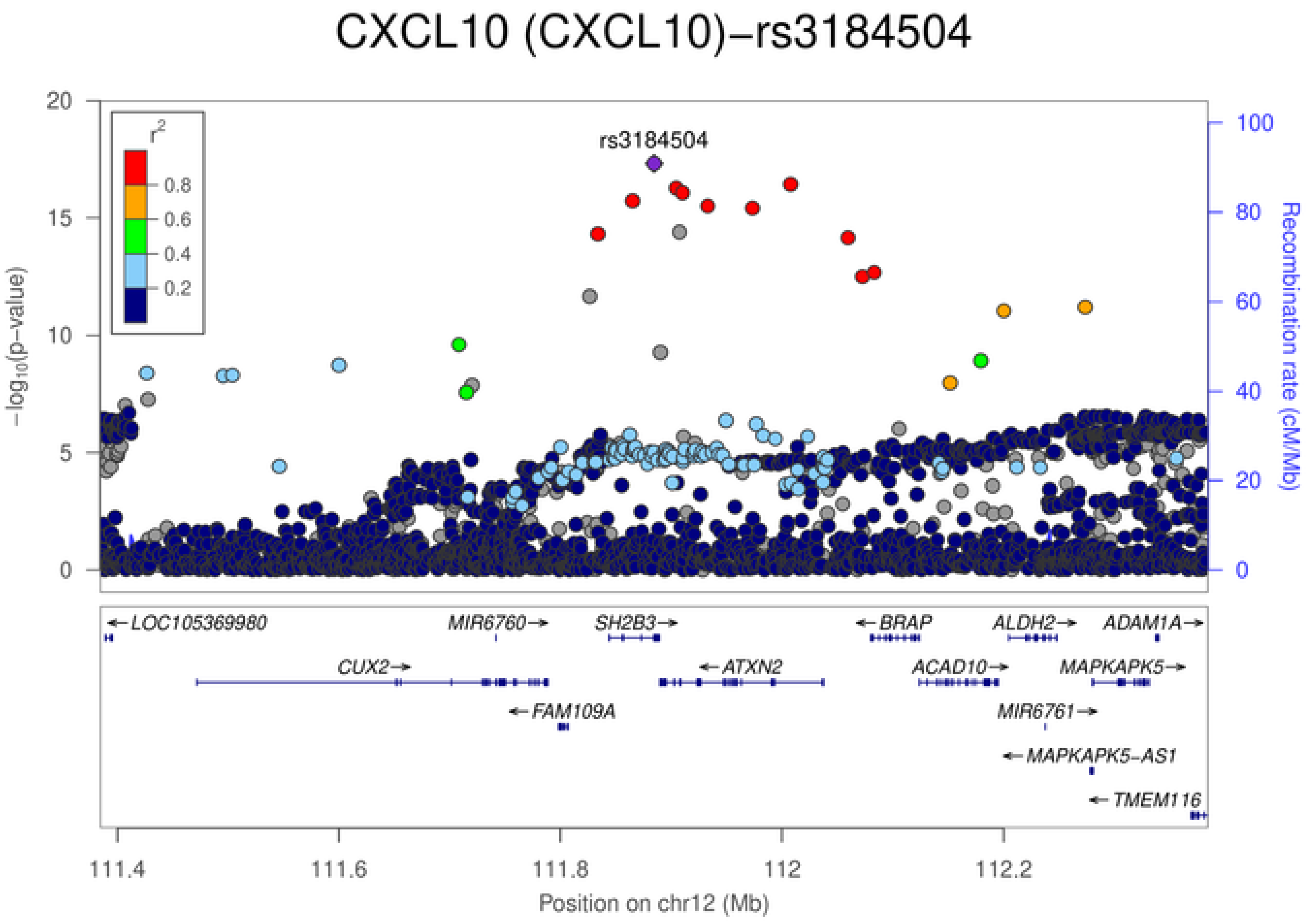
CXCL10 (CXCL10)−rs3184504
rs3184504
$r^2$
0.8
0.6
0.4
0.2
−log10(p−value)
Recombination rate (cM/Mb)
LOC105369980
MIR6760
SH2B3
BRAP
ALDH2
ADAM1A
CUX2
ATXN2
ACAD10
MAPKAPK5
FAM109A
MIR6761
MAPKAPK5-AS1
TMEM116
111.4
111.6
111.8
112
112.2
Position on chr12 (Mb)

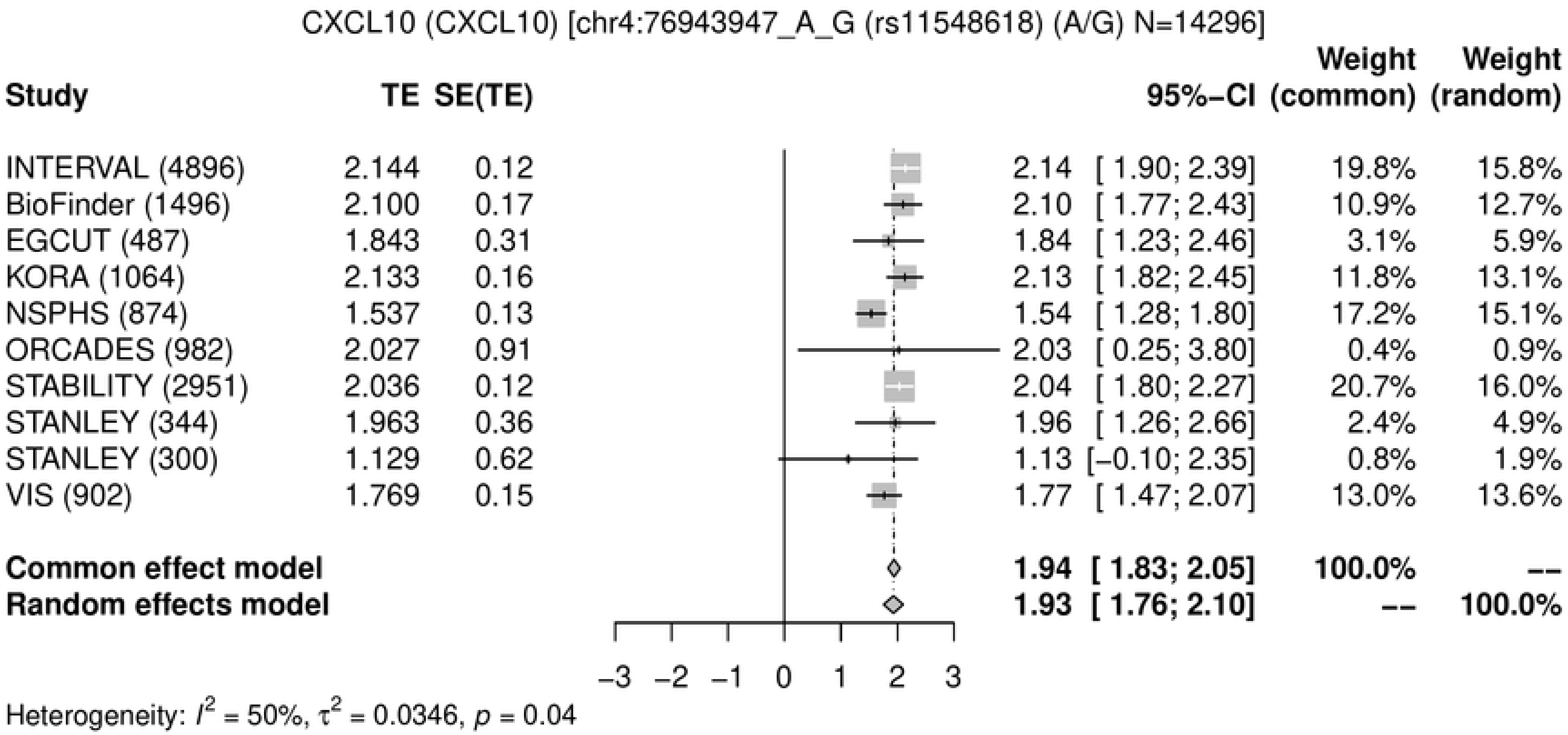
CXCL10 (CXCL10) [chr4:76943947_A_G (rs11548618) (A/G) N=14296]
Study
TE
SE(TE)
95%-CI
Weight (common)
Weight (random)
INTERVAL (4896) 2.144 0.12 2.14 [ 1.90; 2.39] 19.8% 15.8%
BioFinder (1496) 2.100 0.17 2.10 [ 1.77; 2.43] 10.9% 12.7%
EGCUT (487) 1.843 0.31 1.84 [ 1.23; 2.46] 3.1% 5.9%
KORA (1064) 2.133 0.16 2.13 [ 1.82; 2.45] 11.8% 13.1%
NSPHS (874) 1.537 0.13 1.54 [ 1.28; 1.80] 17.2% 15.1%
ORCADES (982) 2.027 0.91 2.03 [ 0.25; 3.80] 0.4% 0.9%
STABILITY (2951) 2.036 0.12 2.04 [ 1.80; 2.27] 20.7% 16.0%
STANLEY (344) 1.963 0.36 1.96 [ 1.26; 2.66] 2.4% 4.9%
STANLEY (300) 1.129 0.62 1.13 [−0.10; 2.35] 0.8% 1.9%
VIS (902) 1.769 0.15 1.77 [ 1.47; 2.07] 13.0% 13.6%
Common effect model 1.94 [ 1.83; 2.05] 100.0% --
Random effects model 1.93 [ 1.76; 2.10] -- 100.0%
−3 −2 −1 0 1 2 3
Heterogeneity: $I^2$ = 50%, $\tau^2$ = 0.0346, $p$ = 0.04
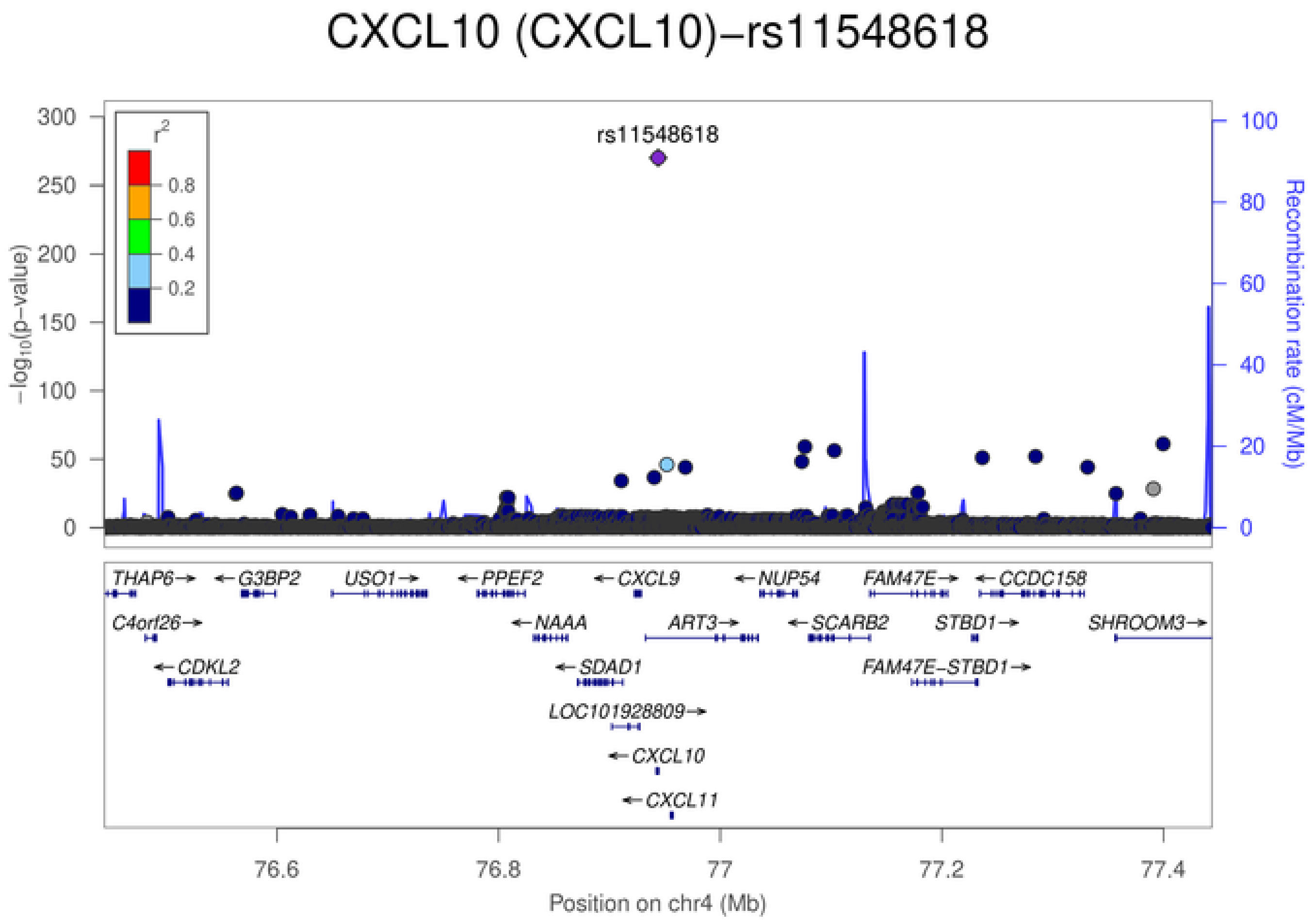
CXCL10 (CXCL10)−rs11548618
rs11548618
$r^2$
0.8
0.6
0.4
0.2
−log10(p−value)
Recombination rate (cM/Mb)
THAP6 G3BP2 USO1 PPEF2 CXCL9 NUP54 FAM47E CCDC158
C4orf26 NAAA ART3 SCARB2 STBD1 SHROOM3
CDKL2 SDAD1 FAM47E−STBD1
LOC101928809
CXCL10
CXCL11
76.6 76.8 77 77.2 77.4
Position on chr4 (Mb)

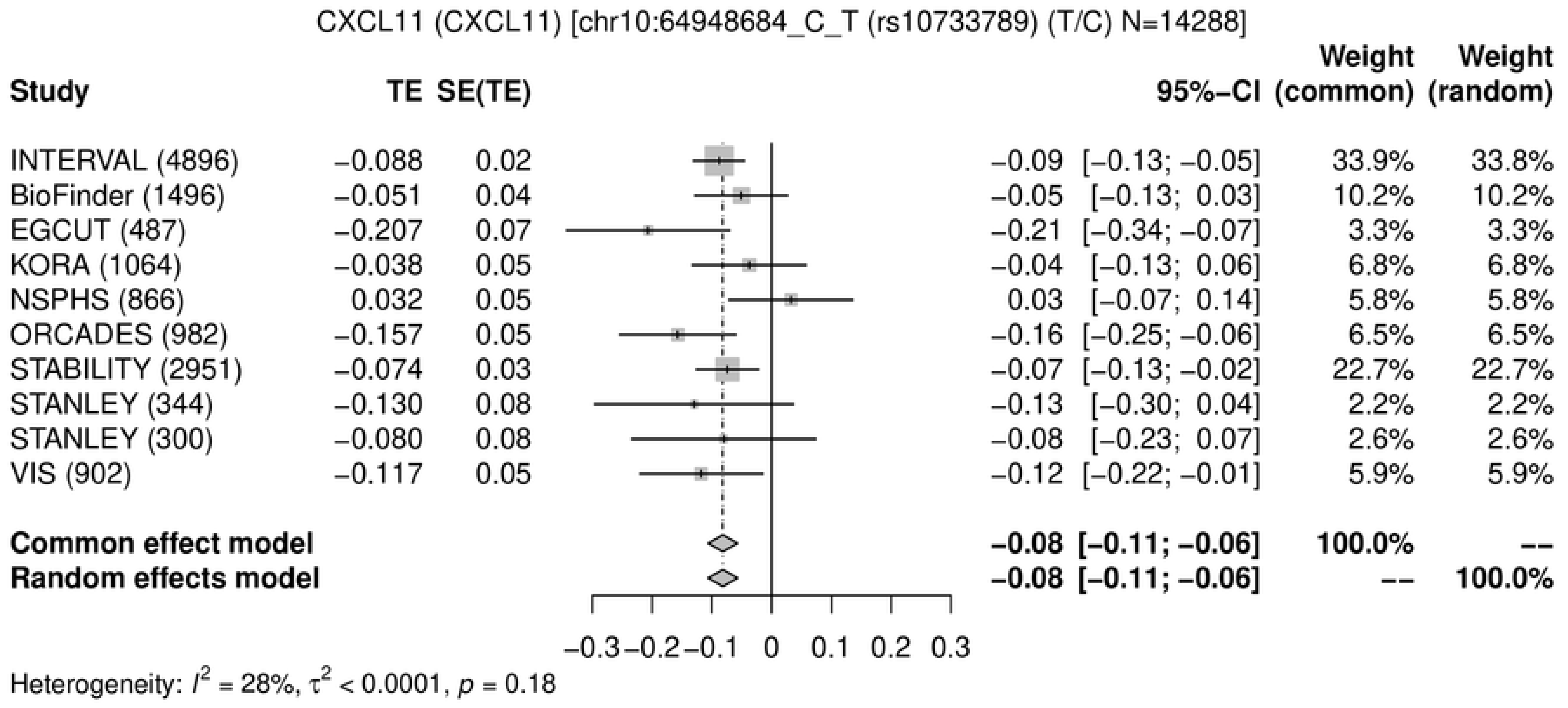
CXCL11 (CXCL11) [chr10:64948684_C_T (rs10733789) (T/C) N=14288]
Study
TE
SE(TE)
95%−CI
Weight (common)
Weight (random)
INTERVAL (4896) −0.088 0.02 −0.09 [−0.13; −0.05] 33.9% 33.8%
BioFinder (1496) −0.051 0.04 −0.05 [−0.13; 0.03] 10.2% 10.2%
EGCUT (487) −0.207 0.07 −0.21 [−0.34; −0.07] 3.3% 3.3%
KORA (1064) −0.038 0.05 −0.04 [−0.13; 0.06] 6.8% 6.8%
NSPHS (866) 0.032 0.05 0.03 [−0.07; 0.14] 5.8% 5.8%
ORCADES (982) −0.157 0.05 −0.16 [−0.25; −0.06] 6.5% 6.5%
STABILITY (2951) −0.074 0.03 −0.07 [−0.13; −0.02] 22.7% 22.7%
STANLEY (344) −0.130 0.08 −0.13 [−0.30; 0.04] 2.2% 2.2%
STANLEY (300) −0.080 0.08 −0.08 [−0.23; 0.07] 2.6% 2.6%
VIS (902) −0.117 0.05 −0.12 [−0.22; −0.01] 5.9% 5.9%
Common effect model −0.08 [−0.11; −0.06] 100.0% --
Random effects model −0.08 [−0.11; −0.06] -- 100.0%
−0.3 −0.2 −0.1 0 0.1 0.2 0.3
Heterogeneity: $I^2$ = 28%, $\tau^2$ < 0.0001, $p$ = 0.18

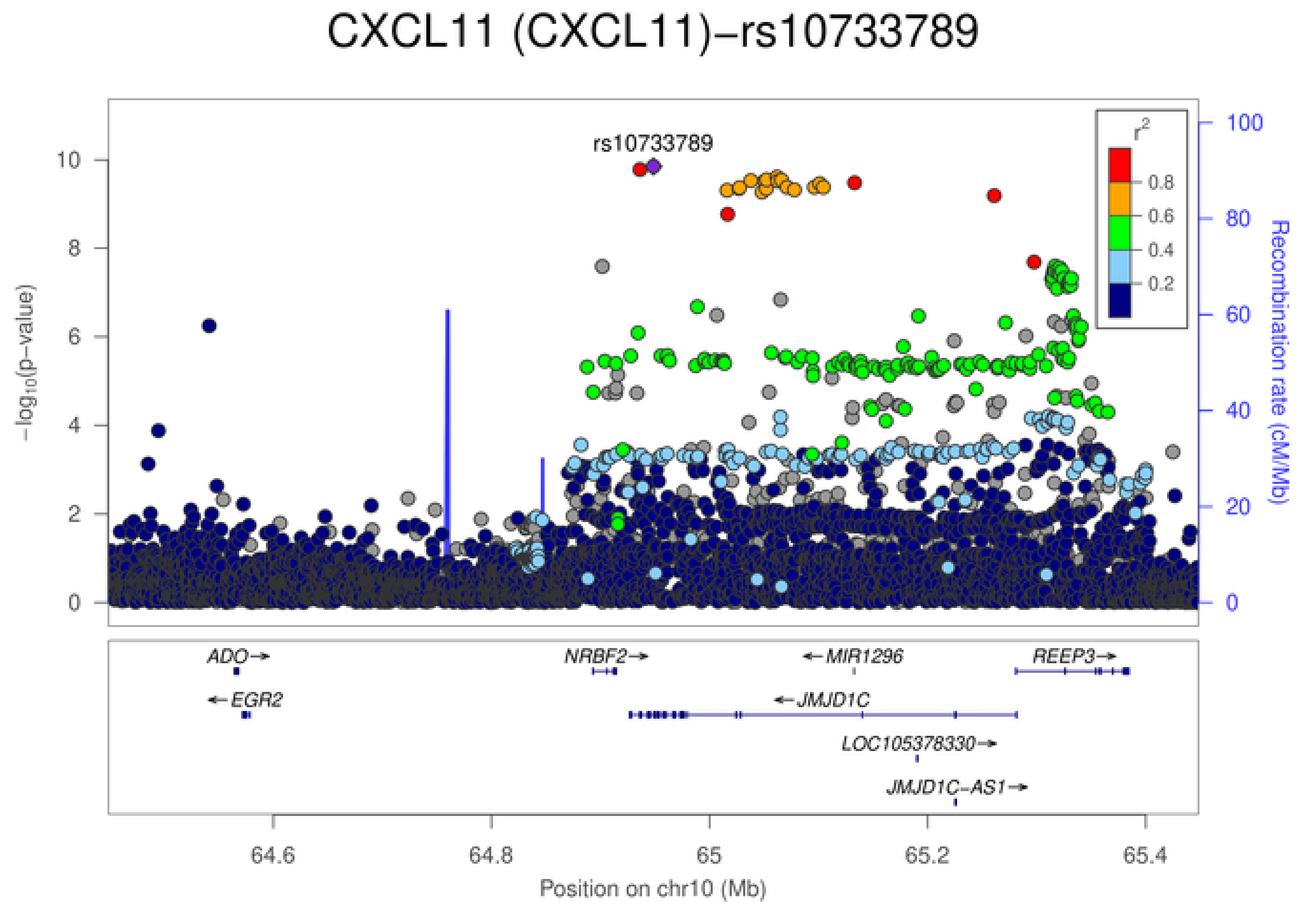
CXCL11 (CXCL11)−rs10733789
rs10733789
−log10(p−value)
Recombination rate (cM/Mb)
Position on chr10 (Mb)

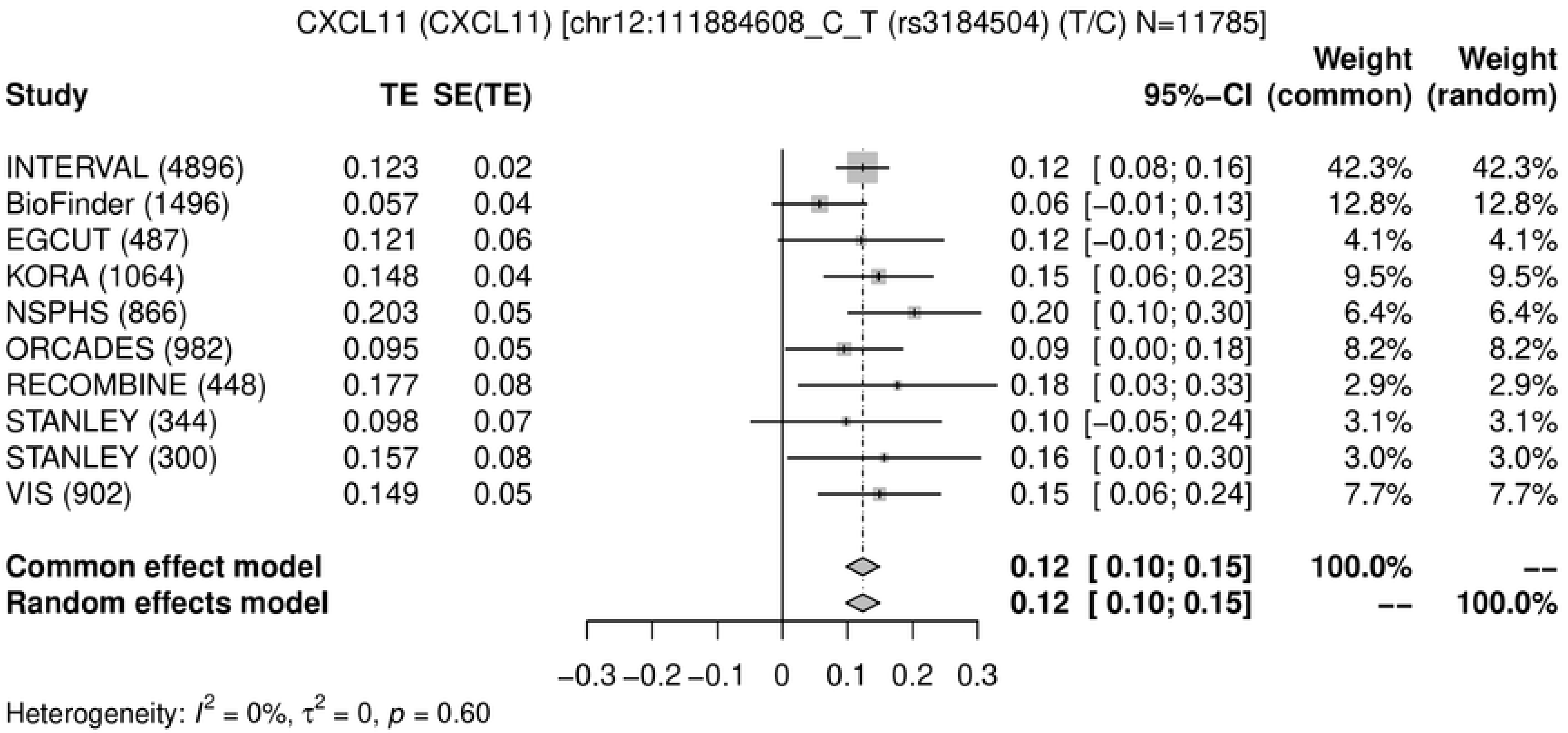
CXCL11 (CXCL11) [chr12:111884608_C_T (rs3184504) (T/C) N=11785]
Study
TE
SE(TE)
95%−CI
Weight (common)
Weight (random)
INTERVAL (4896) 0.123 0.02 0.12 [ 0.08; 0.16] 42.3% 42.3%
BioFinder (1496) 0.057 0.04 0.06 [−0.01; 0.13] 12.8% 12.8%
EGCUT (487) 0.121 0.06 0.12 [−0.01; 0.25] 4.1% 4.1%
KORA (1064) 0.148 0.04 0.15 [ 0.06; 0.23] 9.5% 9.5%
NSPHS (866) 0.203 0.05 0.20 [ 0.10; 0.30] 6.4% 6.4%
ORCADES (982) 0.095 0.05 0.09 [ 0.00; 0.18] 8.2% 8.2%
RECOMBINE (448) 0.177 0.08 0.18 [ 0.03; 0.33] 2.9% 2.9%
STANLEY (344) 0.098 0.07 0.10 [−0.05; 0.24] 3.1% 3.1%
STANLEY (300) 0.157 0.08 0.16 [ 0.01; 0.30] 3.0% 3.0%
VIS (902) 0.149 0.05 0.15 [ 0.06; 0.24] 7.7% 7.7%
Common effect model 0.12 [ 0.10; 0.15] 100.0% --
Random effects model 0.12 [ 0.10; 0.15] -- 100.0%
−0.3 −0.2 −0.1 0 0.1 0.2 0.3
Heterogeneity: $I^2$ = 0%, $\tau^2$ = 0, $p$ = 0.60

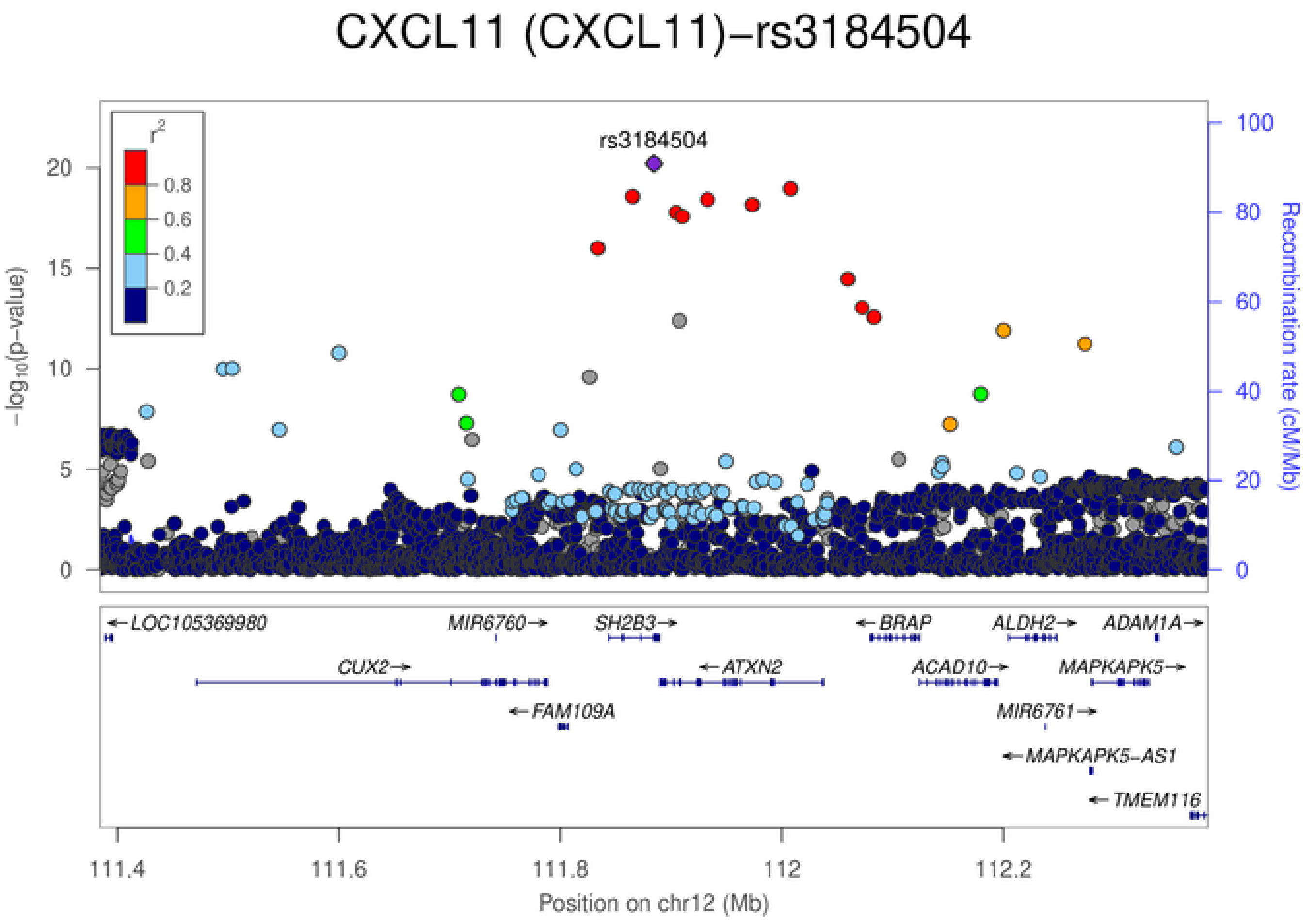
CXCL11 (CXCL11)−rs3184504
rs3184504
$r^2$
0.8
0.6
0.4
0.2
−log10(p−value)
Recombination rate (cM/Mb)
LOC105369980
MIR6760
SH2B3
BRAP
ALDH2
ADAM1A
CUX2
ATXN2
ACAD10
MAPKAPK5
FAM109A
MIR6761
MAPKAPK5−AS1
TMEM116
111.4 111.6 111.8 112 112.2
Position on chr12 (Mb)

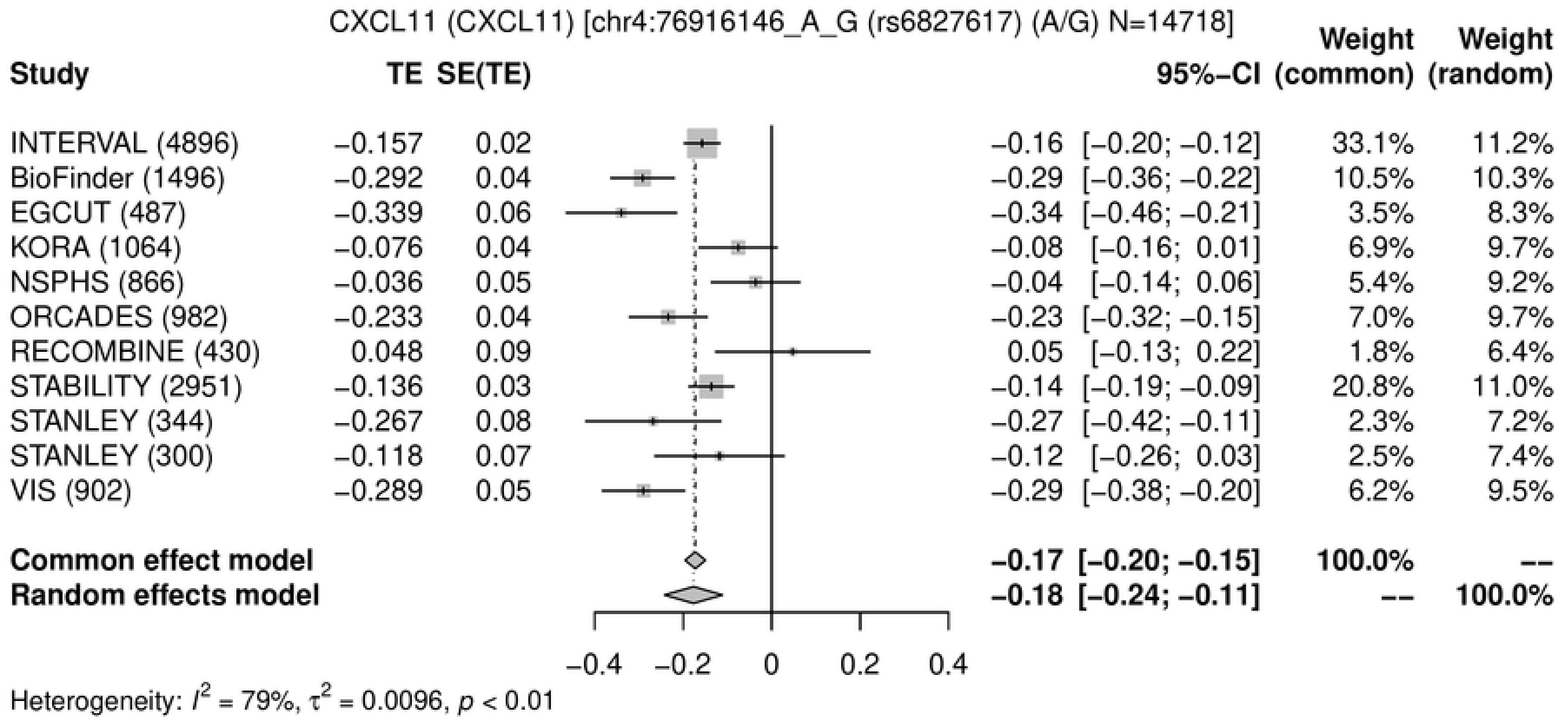
CXCL11 (CXCL11) [chr4:76916146_A_G (rs6827617) (A/G) N=14718]
Study
TE
SE(TE)
95%−CI
Weight (common)
Weight (random)
INTERVAL (4896) −0.157 0.02 −0.16 [−0.20; −0.12] 33.1% 11.2%
BioFinder (1496) −0.292 0.04 −0.29 [−0.36; −0.22] 10.5% 10.3%
EGCUT (487) −0.339 0.06 −0.34 [−0.46; −0.21] 3.5% 8.3%
KORA (1064) −0.076 0.04 −0.08 [−0.16; 0.01] 6.9% 9.7%
NSPHS (866) −0.036 0.05 −0.04 [−0.14; 0.06] 5.4% 9.2%
ORCADES (982) −0.233 0.04 −0.23 [−0.32; −0.15] 7.0% 9.7%
RECOMBINE (430) 0.048 0.09 0.05 [−0.13; 0.22] 1.8% 6.4%
STABILITY (2951) −0.136 0.03 −0.14 [−0.19; −0.09] 20.8% 11.0%
STANLEY (344) −0.267 0.08 −0.27 [−0.42; −0.11] 2.3% 7.2%
STANLEY (300) −0.118 0.07 −0.12 [−0.26; 0.03] 2.5% 7.4%
VIS (902) −0.289 0.05 −0.29 [−0.38; −0.20] 6.2% 9.5%
Common effect model −0.17 [−0.20; −0.15] 100.0% --
Random effects model −0.18 [−0.24; −0.11] -- 100.0%
−0.4
−0.2
0
0.2
0.4
Heterogeneity: $I^2$ = 79%, $\tau^2$ = 0.0096, $p$ < 0.01

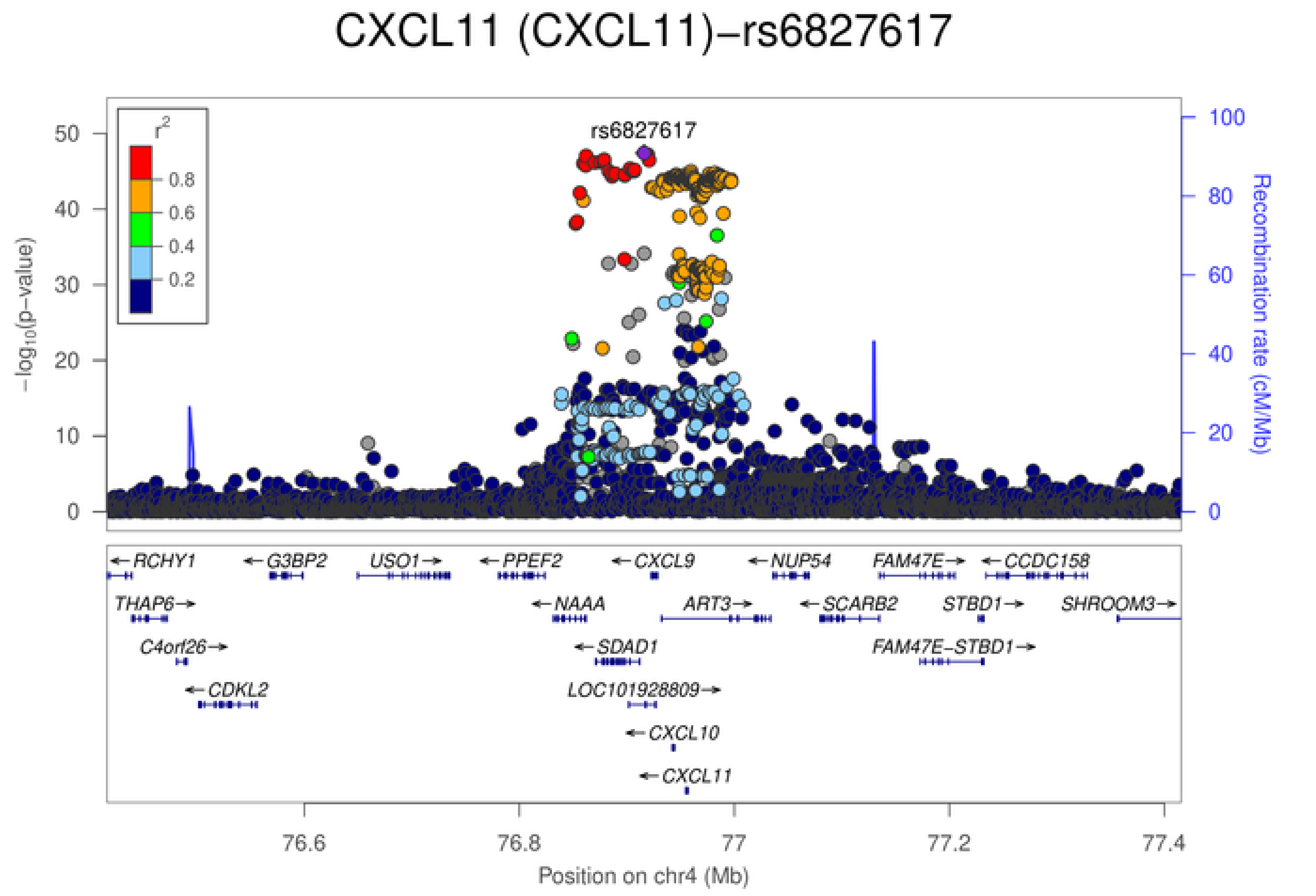
CXCL11 (CXCL11)−rs6827617
rs6827617
$r^2$
0.8
0.6
0.4
0.2
50
40
30
20
10
0
$-\log_{10}$(p−value)
100
80
60
40
20
0
Recombination rate (cM/Mb)
←RCHY1
←G3BP2
USO1→
←PPEF2
←CXCL9
←NUP54
FAM47E→
←CCDC158
THAP6→
←NAAA
ART3→
←SCARB2
STBD1→
SHROOM3→
C4orf26→
←SDAD1
FAM47E−STBD1→
←CDKL2
LOC101928809→
←CXCL10
←CXCL11
76.6
76.8
77
77.2
77.4
Position on chr4 (Mb)

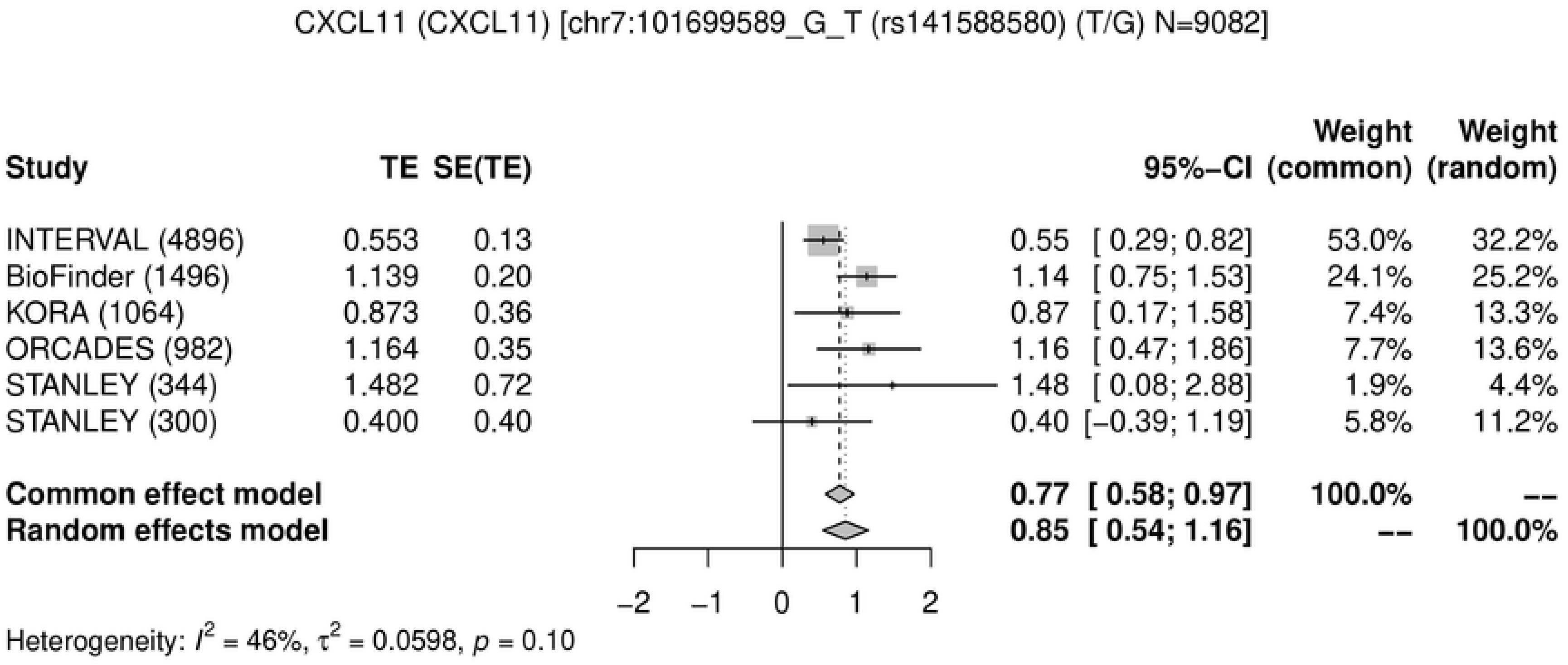
CXCL11 (CXCL11) [chr7:101699589_G_T (rs141588580) (T/G) N=9082]
Study
TE
SE(TE)
95%−CI
Weight (common)
Weight (random)
INTERVAL (4896) 0.553 0.13 0.55 [ 0.29; 0.82] 53.0% 32.2%
BioFinder (1496) 1.139 0.20 1.14 [ 0.75; 1.53] 24.1% 25.2%
KORA (1064) 0.873 0.36 0.87 [ 0.17; 1.58] 7.4% 13.3%
ORCADES (982) 1.164 0.35 1.16 [ 0.47; 1.86] 7.7% 13.6%
STANLEY (344) 1.482 0.72 1.48 [ 0.08; 2.88] 1.9% 4.4%
STANLEY (300) 0.400 0.40 0.40 [−0.39; 1.19] 5.8% 11.2%
Common effect model 0.77 [ 0.58; 0.97] 100.0% --
Random effects model 0.85 [ 0.54; 1.16] -- 100.0%
−2
−1
0
1
2
Heterogeneity: $I^2$ = 46%, $\tau^2$ = 0.0598, $p$ = 0.10
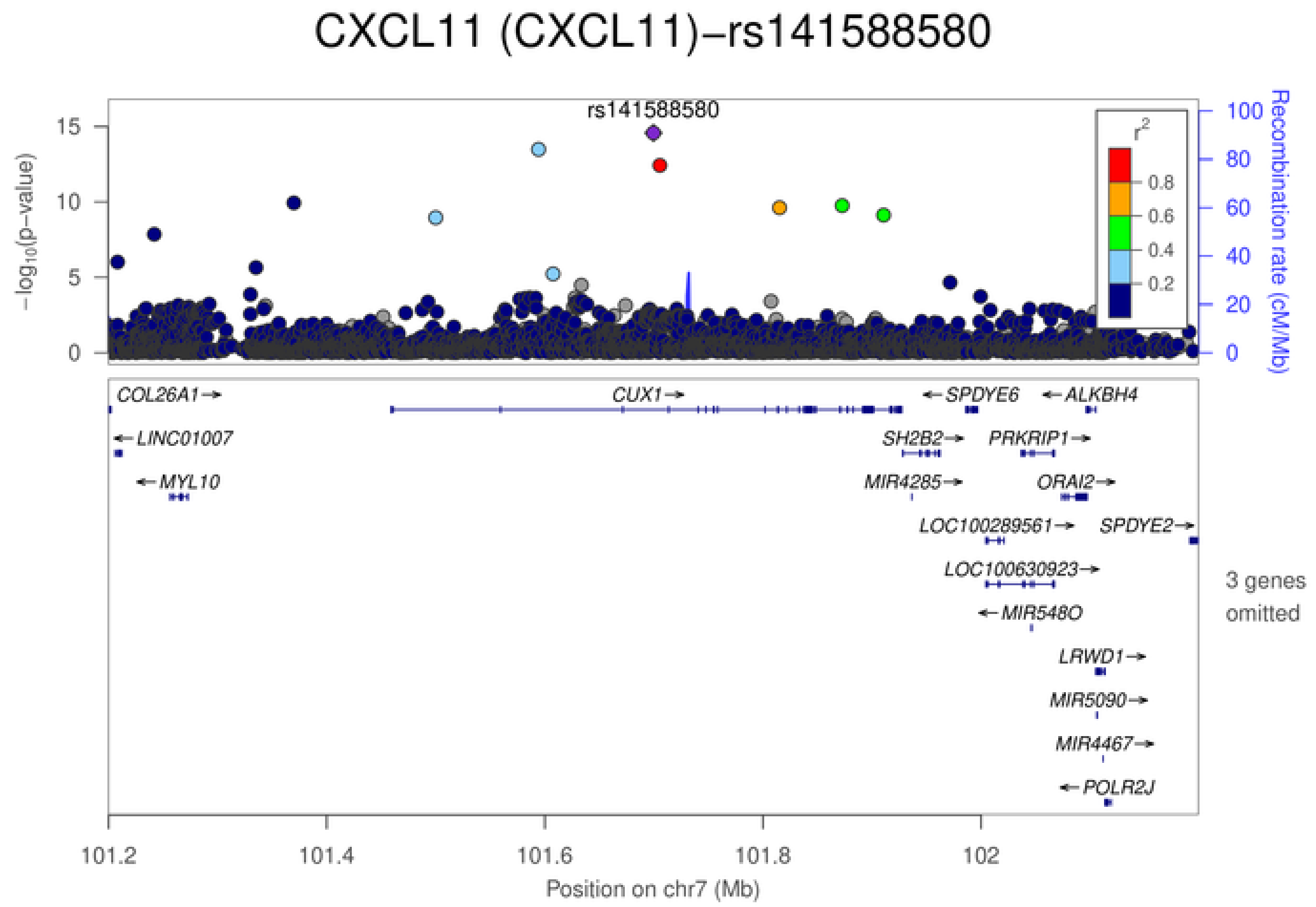
CXCL11 (CXCL11)−rs141588580
rs141588580
$-\log_{10}$(p−value)
Recombination rate (cM/Mb)
$r^2$
0.8
0.6
0.4
0.2
COL26A1
CUX1
SPDYE6
ALKBH4
LINC01007
SH2B2
PRKRIP1
MYL10
MIR4285
ORAI2
LOC100289561
SPDYE2
LOC100630923
MIR548O
LRWD1
MIR5090
MIR4467
POLR2J
3 genes omitted
101.2
101.4
101.6
101.8
102
Position on chr7 (Mb)

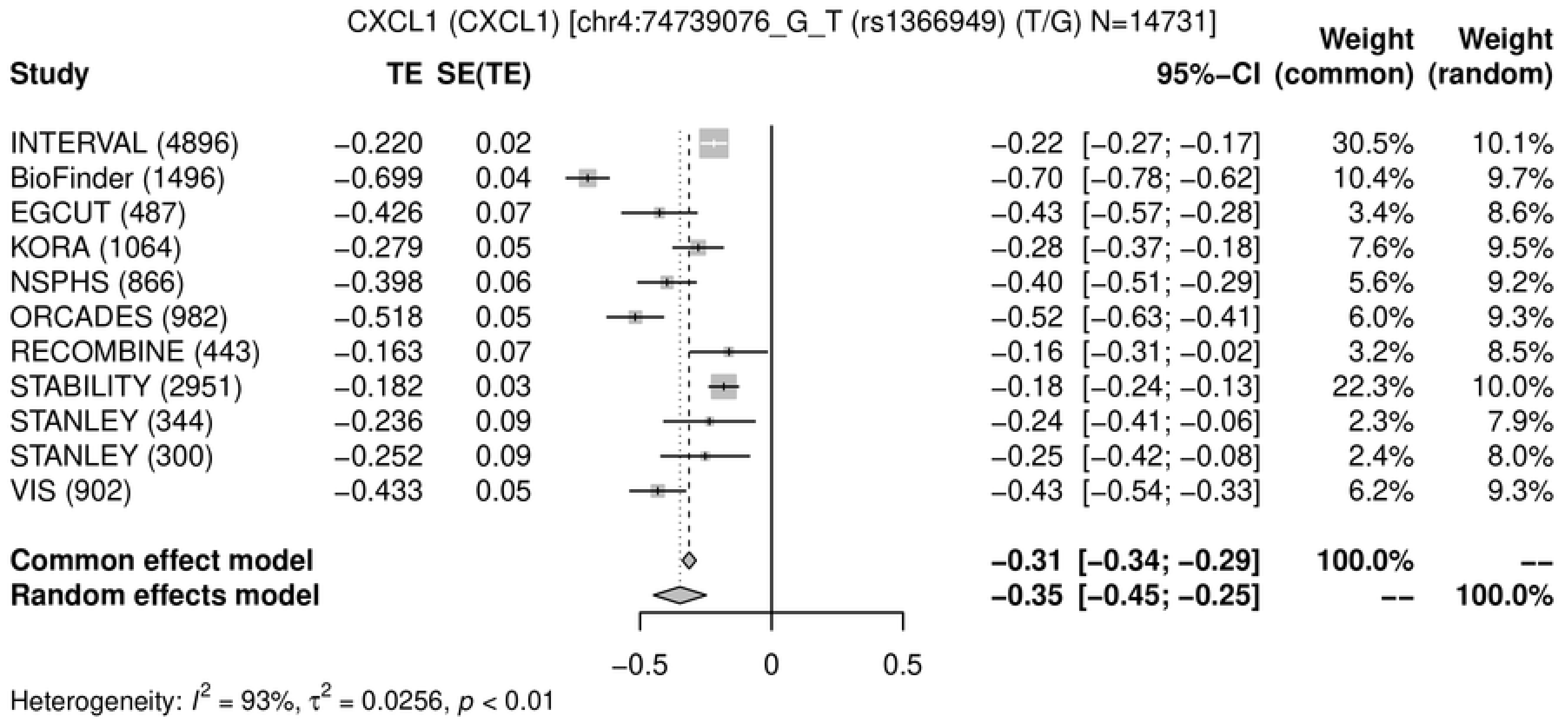
CXCL1 (CXCL1) [chr4:74739076_G_T (rs1366949) (T/G) N=14731]
Study
TE
SE(TE)
95%−CI
Weight (common)
Weight (random)
INTERVAL (4896) −0.220 0.02 −0.22 [−0.27; −0.17] 30.5% 10.1%
BioFinder (1496) −0.699 0.04 −0.70 [−0.78; −0.62] 10.4% 9.7%
EGCUT (487) −0.426 0.07 −0.43 [−0.57; −0.28] 3.4% 8.6%
KORA (1064) −0.279 0.05 −0.28 [−0.37; −0.18] 7.6% 9.5%
NSPHS (866) −0.398 0.06 −0.40 [−0.51; −0.29] 5.6% 9.2%
ORCADES (982) −0.518 0.05 −0.52 [−0.63; −0.41] 6.0% 9.3%
RECOMBINE (443) −0.163 0.07 −0.16 [−0.31; −0.02] 3.2% 8.5%
STABILITY (2951) −0.182 0.03 −0.18 [−0.24; −0.13] 22.3% 10.0%
STANLEY (344) −0.236 0.09 −0.24 [−0.41; −0.06] 2.3% 7.9%
STANLEY (300) −0.252 0.09 −0.25 [−0.42; −0.08] 2.4% 8.0%
VIS (902) −0.433 0.05 −0.43 [−0.54; −0.33] 6.2% 9.3%
Common effect model −0.31 [−0.34; −0.29] 100.0% --
Random effects model −0.35 [−0.45; −0.25] -- 100.0%
−0.5
0
0.5
Heterogeneity: I² = 93%, τ² = 0.0256, p < 0.01

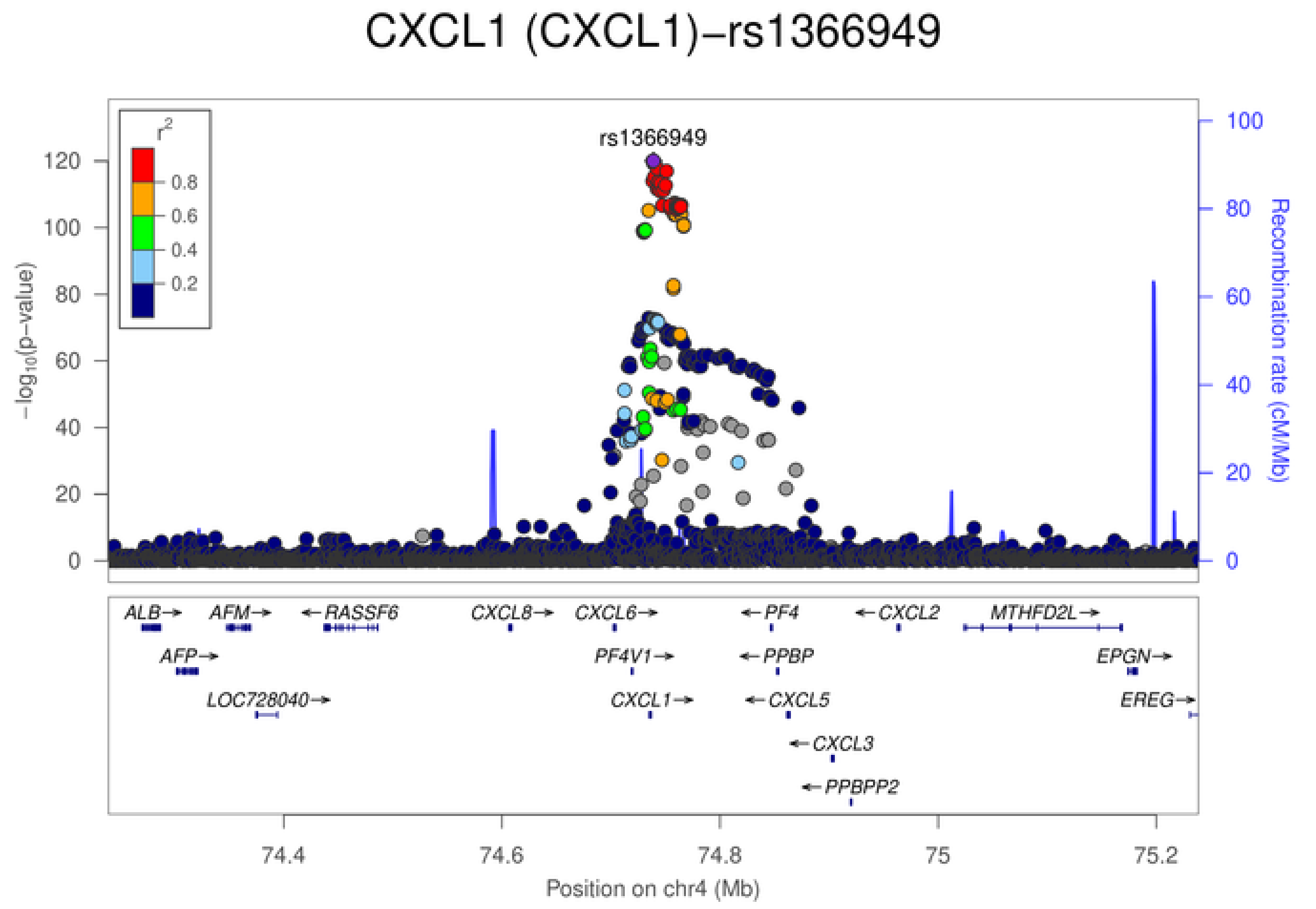
CXCL1 (CXCL1)−rs1366949
rs1366949
r²
0.8
0.6
0.4
0.2
−log10(p−value)
Recombination rate (cM/Mb)
0
20
40
60
80
100
120
ALB→
AFM→
←RASSF6
CXCL8→
CXCL6→
←PF4
←CXCL2
MTHFD2L→
AFP→
PF4V1→
←PPBP
EPGN→
LOC728040→
CXCL1→
←CXCL5
EREG→
←CXCL3
←PPBPP2
74.4
74.6
74.8
75
75.2
Position on chr4 (Mb)

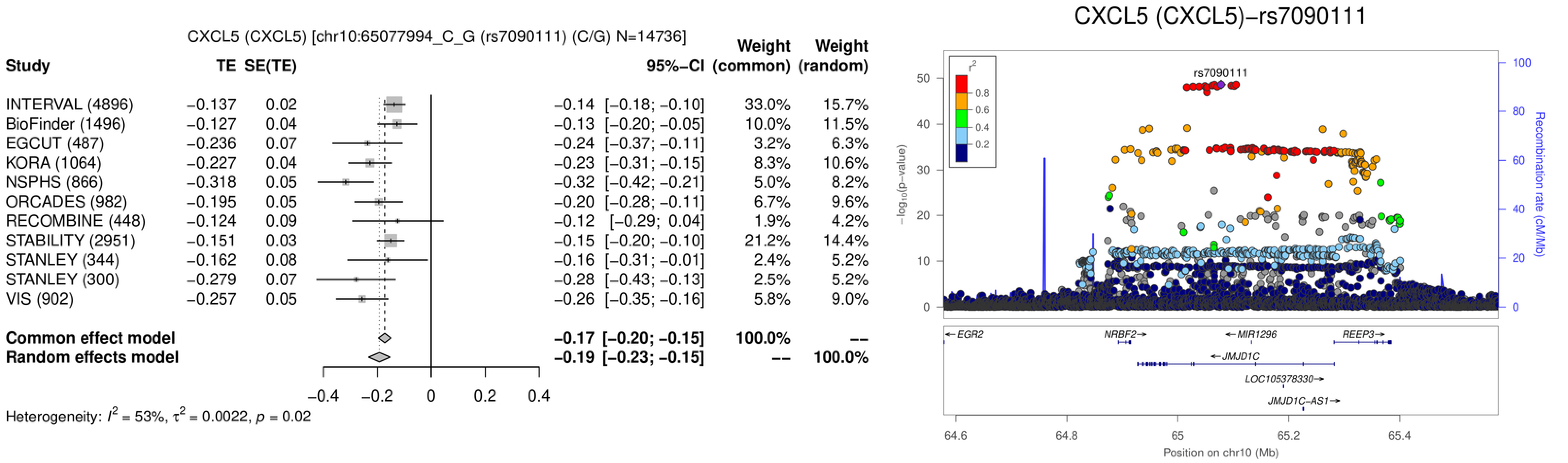
CXCL5 (CXCL5) [chr10:65077994_C_G (rs7090111) (C/G) N=14736]
Study TE SE(TE) 95%−CI Weight (common) Weight (random)
INTERVAL (4896) −0.137 0.02 −0.14 [−0.18; −0.10] 33.0% 15.7%
BioFinder (1496) −0.127 0.04 −0.13 [−0.20; −0.05] 10.0% 11.5%
EGCUT (487) −0.236 0.07 −0.24 [−0.37; −0.11] 3.2% 6.3%
KORA (1064) −0.227 0.04 −0.23 [−0.31; −0.15] 8.3% 10.6%
NSPHS (866) −0.318 0.05 −0.32 [−0.42; −0.21] 5.0% 8.2%
ORCADES (982) −0.195 0.05 −0.20 [−0.28; −0.11] 6.7% 9.6%
RECOMBINE (448) −0.124 0.09 −0.12 [−0.29; 0.04] 1.9% 4.2%
STABILITY (2951) −0.151 0.03 −0.15 [−0.20; −0.10] 21.2% 14.4%
STANLEY (344) −0.162 0.08 −0.16 [−0.31; −0.01] 2.4% 5.2%
STANLEY (300) −0.279 0.07 −0.28 [−0.43; −0.13] 2.5% 5.2%
VIS (902) −0.257 0.05 −0.26 [−0.35; −0.16] 5.8% 9.0%
Common effect model −0.17 [−0.20; −0.15] 100.0% --
Random effects model −0.19 [−0.23; −0.15] -- 100.0%
−0.4 −0.2 0 0.2 0.4
Heterogeneity: $I^2$ = 53%, $\tau^2$ = 0.0022, $p$ = 0.02
CXCL5 (CXCL5)−rs7090111
rs7090111
$r^2$
0.8
0.6
0.4
0.2
$-\log_{10}$(p-value)
0
10
20
30
40
50
Recombination rate (cM/Mb)
0
20
40
60
80
100
←EGR2
NRBF2→
←MIR1296
REEP3→
←JMJD1C
LOC105378330→
JMJD1C−AS1→
64.6
64.8
65
65.2
65.4
Position on chr10 (Mb)

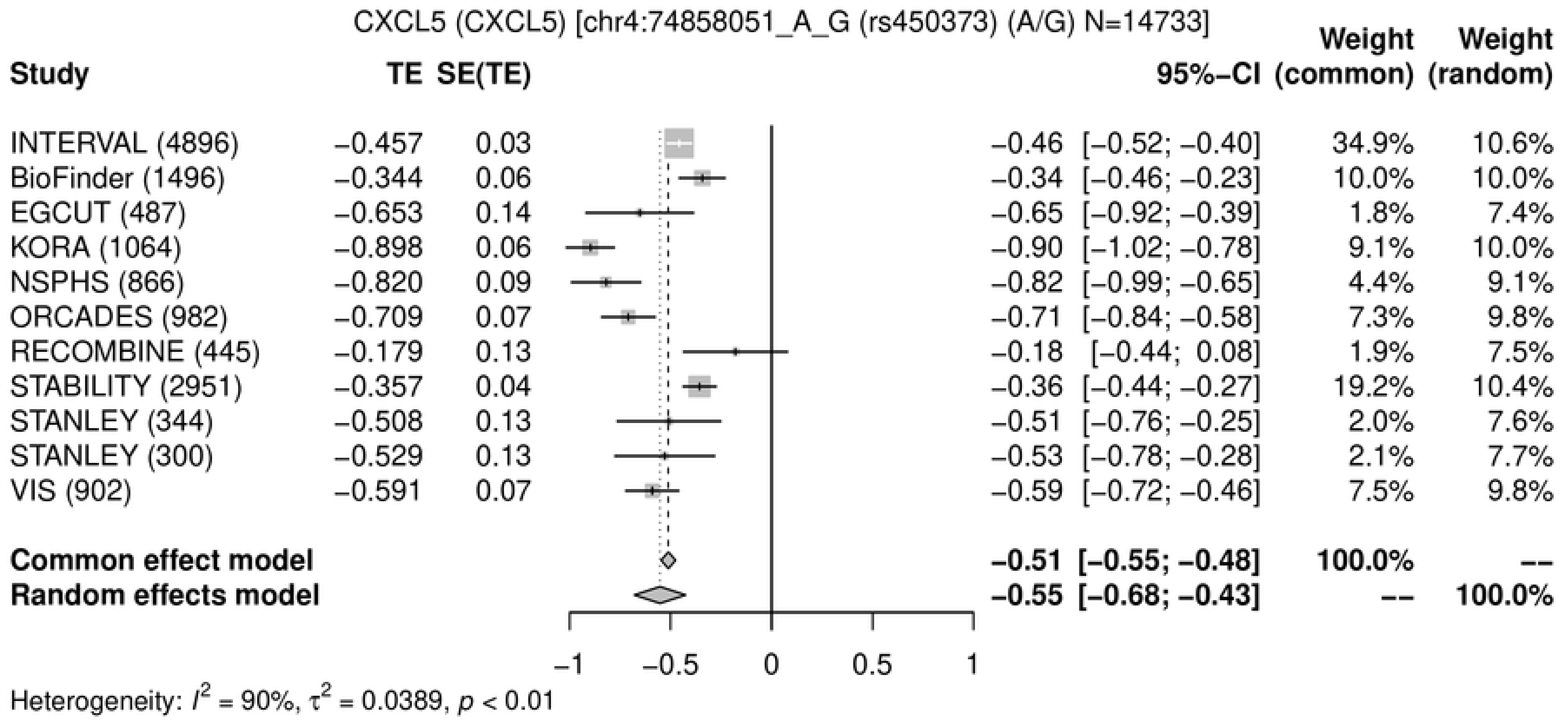
CXCL5 (CXCL5) [chr4:74858051_A_G (rs450373) (A/G) N=14733]
Study
TE
SE(TE)
95%-CI
Weight (common)
Weight (random)
INTERVAL (4896) −0.457 0.03 −0.46 [−0.52; −0.40] 34.9% 10.6%
BioFinder (1496) −0.344 0.06 −0.34 [−0.46; −0.23] 10.0% 10.0%
EGCUT (487) −0.653 0.14 −0.65 [−0.92; −0.39] 1.8% 7.4%
KORA (1064) −0.898 0.06 −0.90 [−1.02; −0.78] 9.1% 10.0%
NSPHS (866) −0.820 0.09 −0.82 [−0.99; −0.65] 4.4% 9.1%
ORCADES (982) −0.709 0.07 −0.71 [−0.84; −0.58] 7.3% 9.8%
RECOMBINE (445) −0.179 0.13 −0.18 [−0.44; 0.08] 1.9% 7.5%
STABILITY (2951) −0.357 0.04 −0.36 [−0.44; −0.27] 19.2% 10.4%
STANLEY (344) −0.508 0.13 −0.51 [−0.76; −0.25] 2.0% 7.6%
STANLEY (300) −0.529 0.13 −0.53 [−0.78; −0.28] 2.1% 7.7%
VIS (902) −0.591 0.07 −0.59 [−0.72; −0.46] 7.5% 9.8%
Common effect model −0.51 [−0.55; −0.48] 100.0% --
Random effects model −0.55 [−0.68; −0.43] -- 100.0%
−1
−0.5
0
0.5
1
Heterogeneity: $I^2$ = 90%, $\tau^2$ = 0.0389, $p$ < 0.01

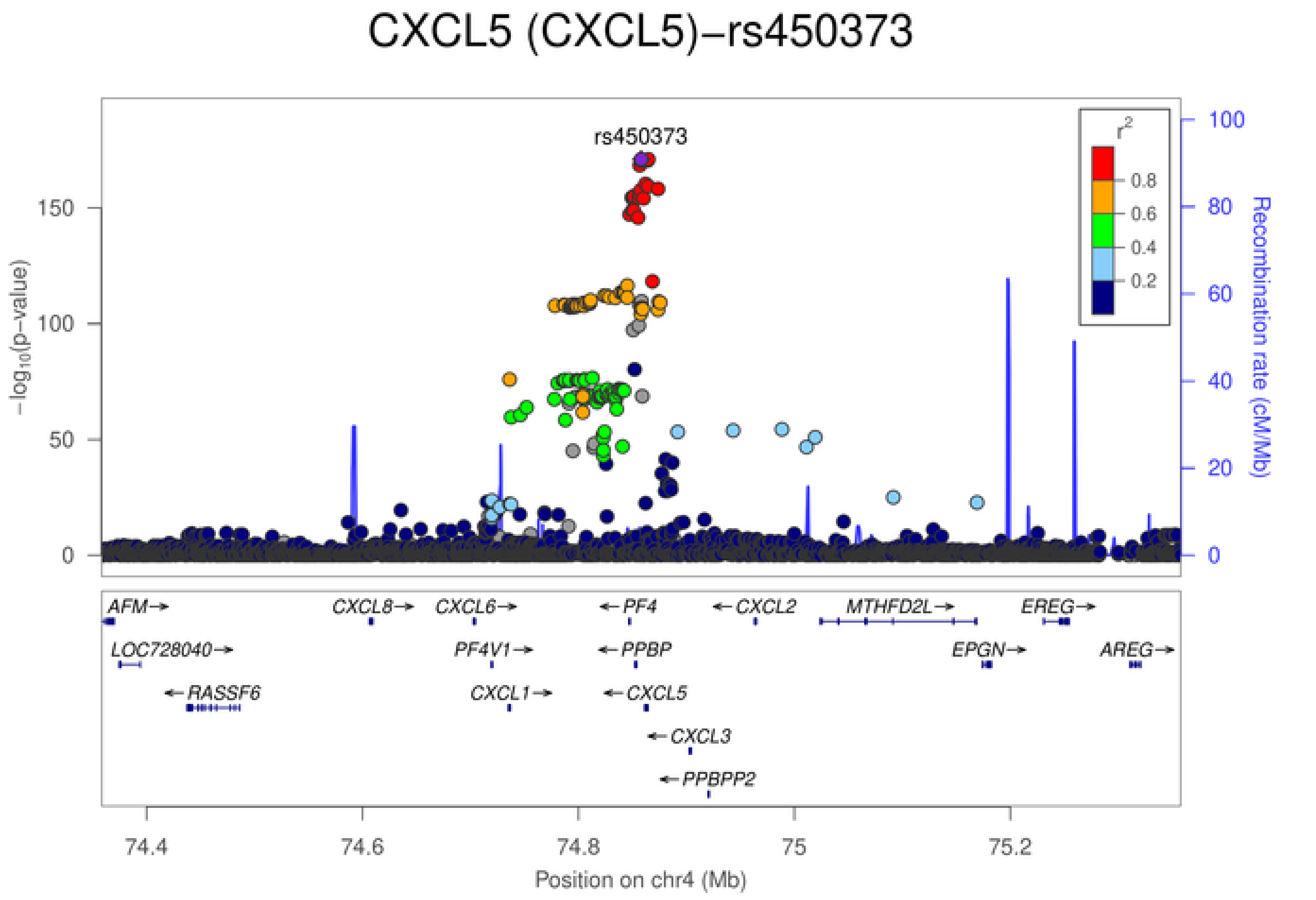
CXCL5 (CXCL5)−rs450373
rs450373
$-\log_{10}$(p-value)
Recombination rate (cM/Mb)
$r^2$
0.8
0.6
0.4
0.2
0
50
100
150
20
40
60
80
100
AFM→
CXCL8→
CXCL6→
←PF4
←CXCL2
MTHFD2L→
EREG→
LOC728040→
PF4V1→
←PPBP
EPGN→
AREG→
←RASSF6
CXCL1→
←CXCL5
←CXCL3
←PPBPP2
74.4
74.6
74.8
75
75.2
Position on chr4 (Mb)

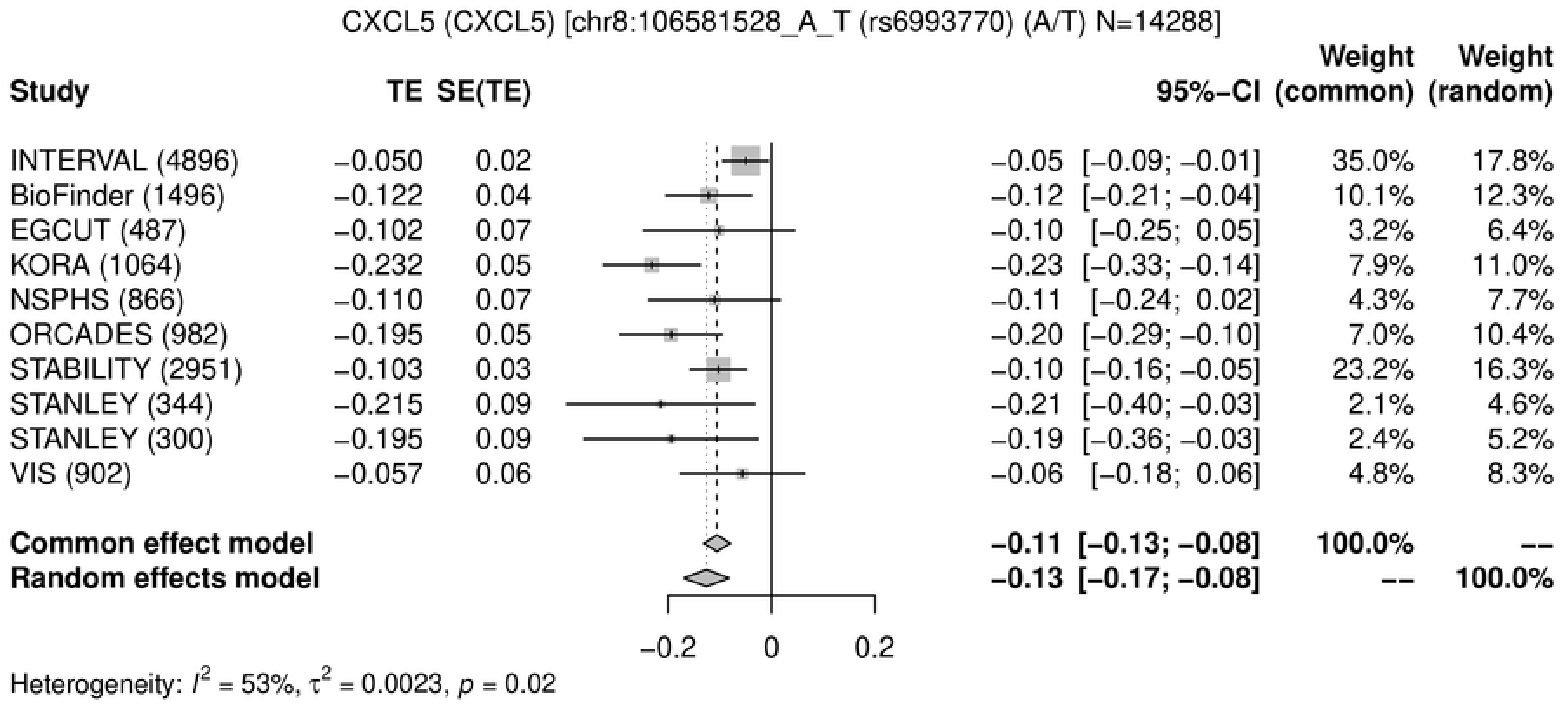
CXCL5 (CXCL5) [chr8:106581528_A_T (rs6993770) (A/T) N=14288]
Study
TE
SE(TE)
95%-CI
Weight (common)
Weight (random)
INTERVAL (4896) −0.050 0.02 −0.05 [−0.09; −0.01] 35.0% 17.8%
BioFinder (1496) −0.122 0.04 −0.12 [−0.21; −0.04] 10.1% 12.3%
EGCUT (487) −0.102 0.07 −0.10 [−0.25; 0.05] 3.2% 6.4%
KORA (1064) −0.232 0.05 −0.23 [−0.33; −0.14] 7.9% 11.0%
NSPHS (866) −0.110 0.07 −0.11 [−0.24; 0.02] 4.3% 7.7%
ORCADES (982) −0.195 0.05 −0.20 [−0.29; −0.10] 7.0% 10.4%
STABILITY (2951) −0.103 0.03 −0.10 [−0.16; −0.05] 23.2% 16.3%
STANLEY (344) −0.215 0.09 −0.21 [−0.40; −0.03] 2.1% 4.6%
STANLEY (300) −0.195 0.09 −0.19 [−0.36; −0.03] 2.4% 5.2%
VIS (902) −0.057 0.06 −0.06 [−0.18; 0.06] 4.8% 8.3%
Common effect model −0.11 [−0.13; −0.08] 100.0% --
Random effects model −0.13 [−0.17; −0.08] -- 100.0%
−0.2
0
0.2
Heterogeneity: $I^2$ = 53%, $\tau^2$ = 0.0023, $p$ = 0.02
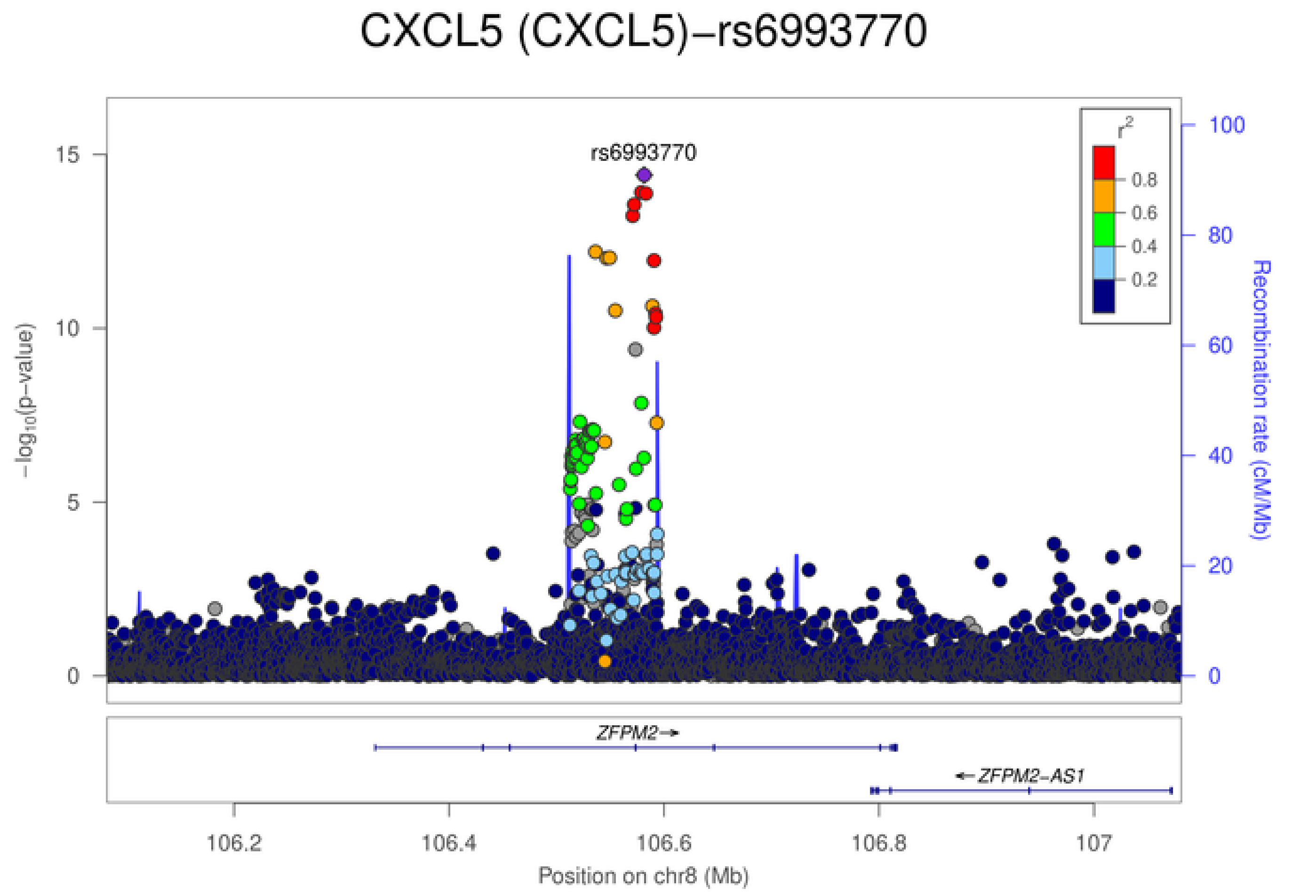
CXCL5 (CXCL5)−rs6993770
rs6993770
$-\log_{10}$(p−value)
Recombination rate (cM/Mb)
$r^2$
0.8
0.6
0.4
0.2
ZFPM2→
←ZFPM2−AS1
106.2
106.4
106.6
106.8
107
Position on chr8 (Mb)

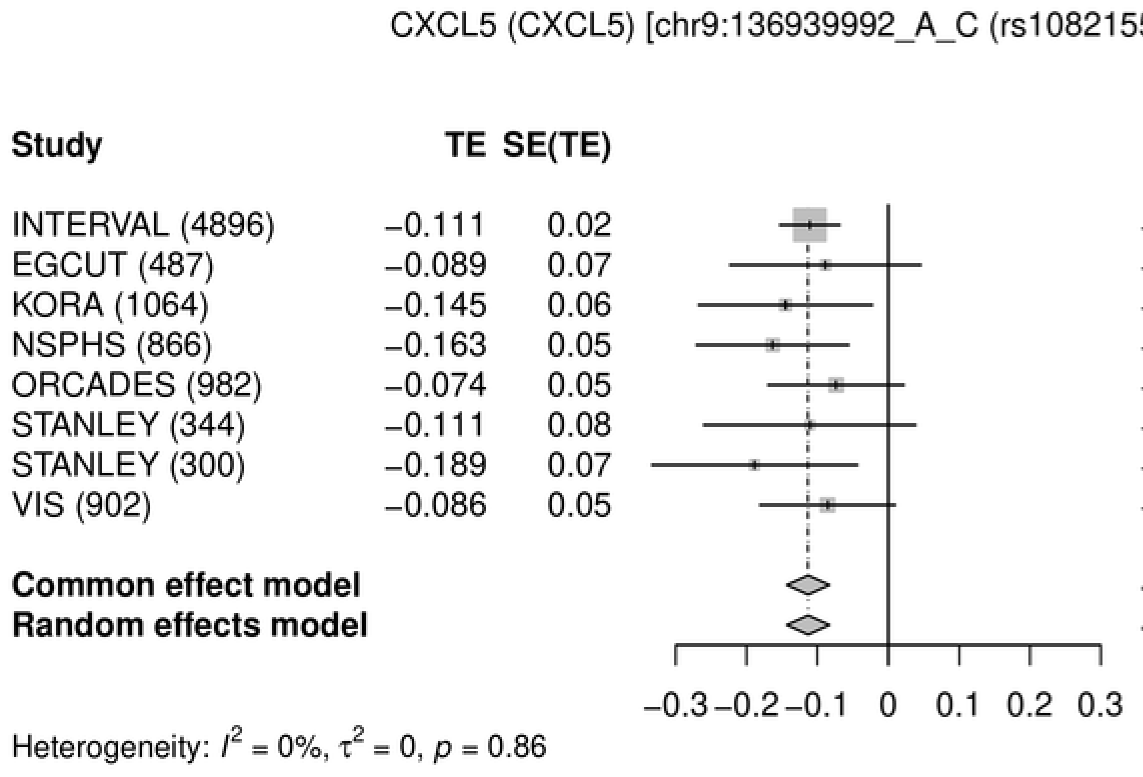
CXCL5 (CXCL5) [chr9:136939992_A_C (rs10821552) (A/C) N=9841]
Study
TE
SE(TE)
95%-CI
Weight (common)
Weight (random)
INTERVAL (4896) −0.111 0.02 −0.11 [−0.15; −0.07] 51.9% 51.9%
EGCUT (487) −0.089 0.07 −0.09 [−0.22; 0.05] 5.1% 5.1%
KORA (1064) −0.145 0.06 −0.15 [−0.27; −0.02] 6.2% 6.2%
NSPHS (866) −0.163 0.05 −0.16 [−0.27; −0.06] 8.0% 8.0%
ORCADES (982) −0.074 0.05 −0.07 [−0.17; 0.02] 10.1% 10.1%
STANLEY (344) −0.111 0.08 −0.11 [−0.26; 0.04] 4.1% 4.1%
STANLEY (300) −0.189 0.07 −0.19 [−0.33; −0.04] 4.4% 4.4%
VIS (902) −0.086 0.05 −0.09 [−0.18; 0.01] 10.2% 10.2%
Common effect model −0.11 [−0.14; −0.08] 100.0% --
Random effects model −0.11 [−0.14; −0.08] -- 100.0%
−0.3 −0.2 −0.1 0 0.1 0.2 0.3
Heterogeneity: $I^2$ = 0%, $\tau^2$ = 0, $p$ = 0.86

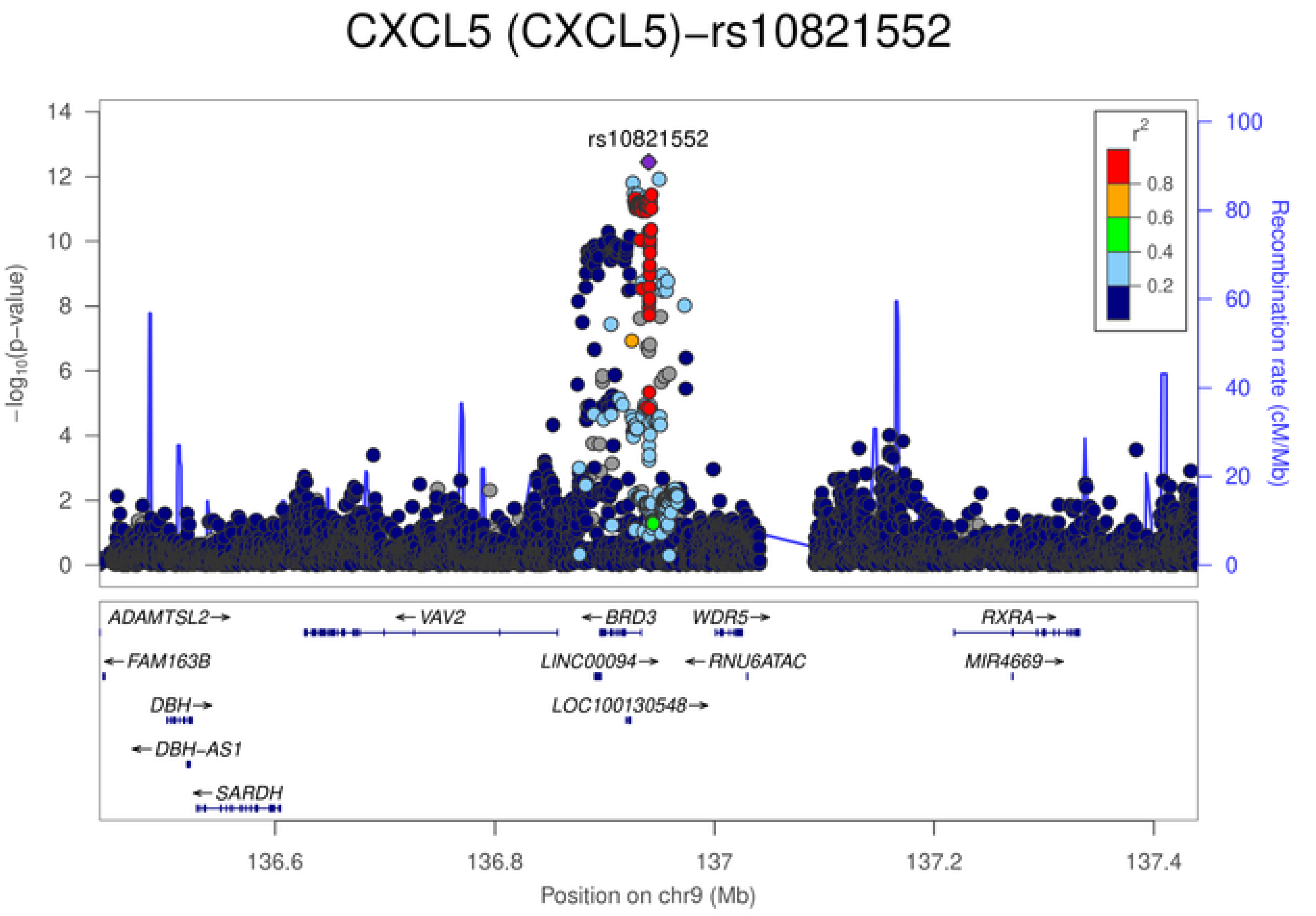
CXCL5 (CXCL5)−rs10821552
rs10821552
$-\log_{10}$(p-value)
Recombination rate (cM/Mb)
$r^2$
0.8
0.6
0.4
0.2
ADAMTSL2→
←FAM163B
DBH→
←DBH-AS1
←SARDH
←VAV2
←BRD3
LINC00094→
LOC100130548→
WDR5→
←RNU6ATAC
RXRA→
MIR4669→
136.6
136.8
137
137.2
137.4
Position on chr9 (Mb)

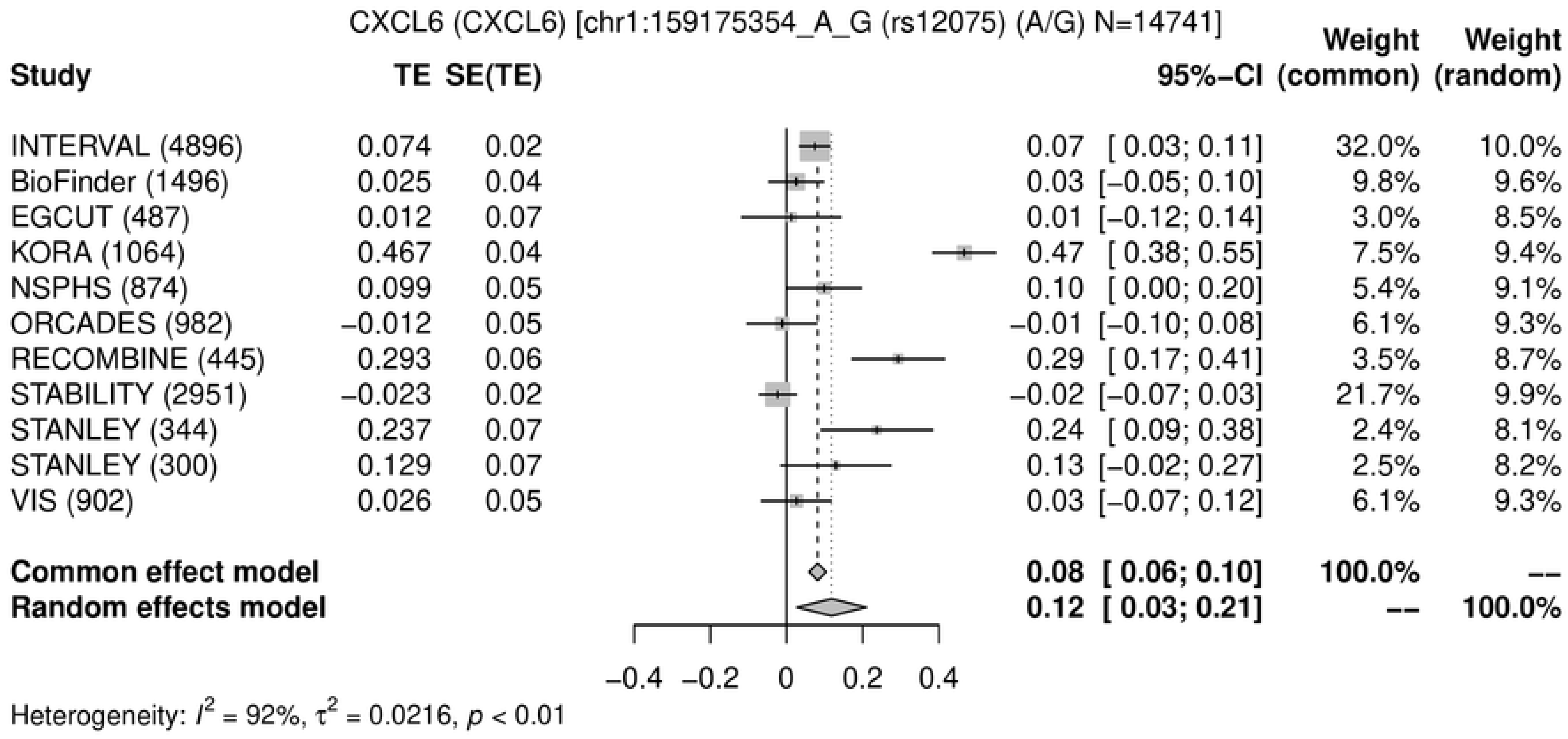
CXCL6 (CXCL6) [chr1:159175354_A_G (rs12075) (A/G) N=14741]
Study | TE | SE(TE) | 95%-CI | Weight (common) | Weight (random)
INTERVAL (4896) | 0.074 | 0.02 | 0.07 [ 0.03; 0.11] | 32.0% | 10.0%
BioFinder (1496) | 0.025 | 0.04 | 0.03 [−0.05; 0.10] | 9.8% | 9.6%
EGCUT (487) | 0.012 | 0.07 | 0.01 [−0.12; 0.14] | 3.0% | 8.5%
KORA (1064) | 0.467 | 0.04 | 0.47 [ 0.38; 0.55] | 7.5% | 9.4%
NSPHS (874) | 0.099 | 0.05 | 0.10 [ 0.00; 0.20] | 5.4% | 9.1%
ORCADES (982) | −0.012 | 0.05 | −0.01 [−0.10; 0.08] | 6.1% | 9.3%
RECOMBINE (445) | 0.293 | 0.06 | 0.29 [ 0.17; 0.41] | 3.5% | 8.7%
STABILITY (2951) | −0.023 | 0.02 | −0.02 [−0.07; 0.03] | 21.7% | 9.9%
STANLEY (344) | 0.237 | 0.07 | 0.24 [ 0.09; 0.38] | 2.4% | 8.1%
STANLEY (300) | 0.129 | 0.07 | 0.13 [−0.02; 0.27] | 2.5% | 8.2%
VIS (902) | 0.026 | 0.05 | 0.03 [−0.07; 0.12] | 6.1% | 9.3%
Common effect model | 0.08 [ 0.06; 0.10] | 100.0% | --
Random effects model | 0.12 [ 0.03; 0.21] | -- | 100.0%
−0.4 −0.2 0 0.2 0.4
Heterogeneity: $I^2$ = 92%, $\tau^2$ = 0.0216, $p$ < 0.01

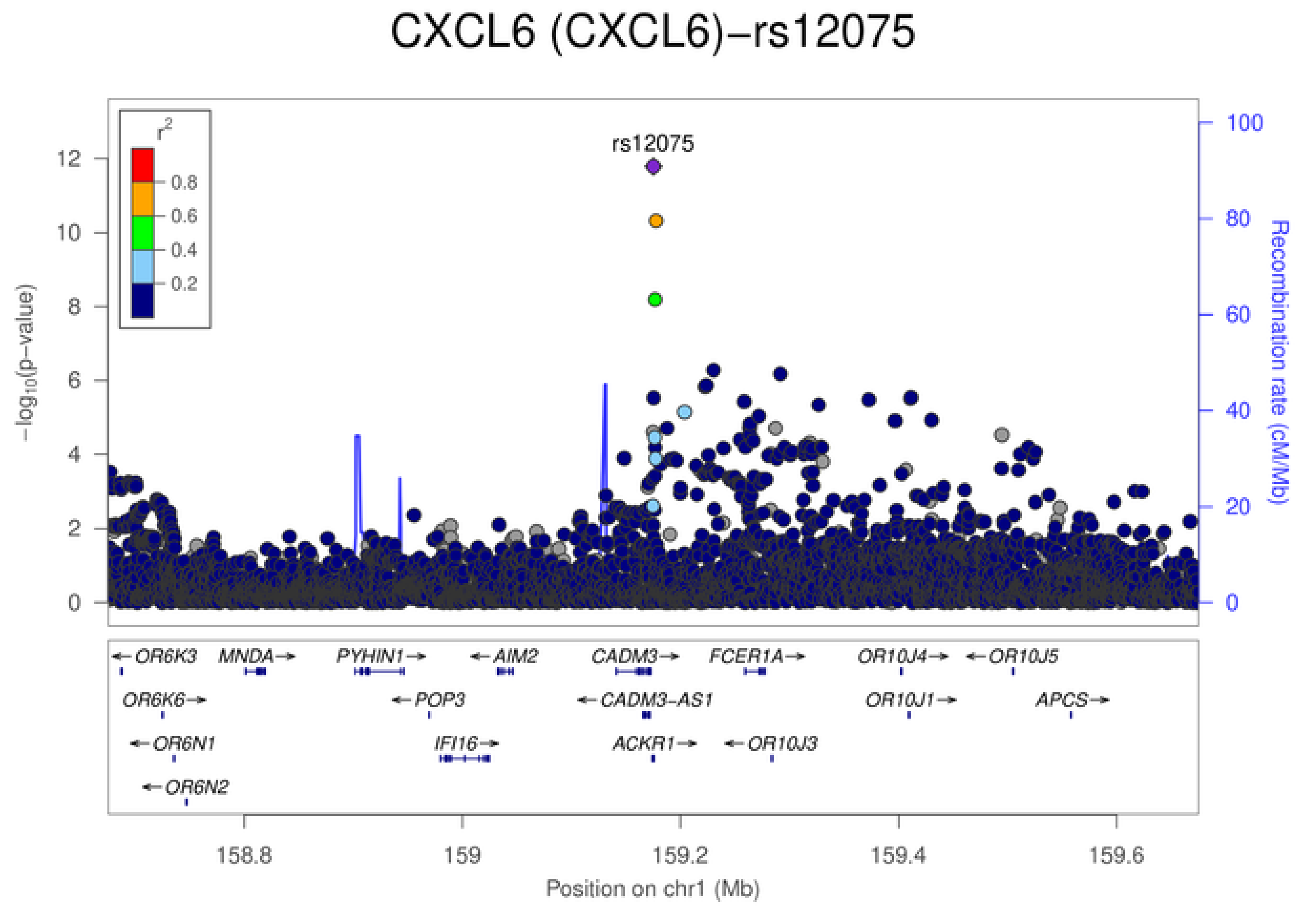
CXCL6 (CXCL6)−rs12075
rs12075
$r^2$
0.8
0.6
0.4
0.2
$-\log_{10}$(p−value)
Recombination rate (cM/Mb)
OR6K3
MNDA
PYHIN1
AIM2
CADM3
FCER1A
OR10J4
OR10J5
OR6K6
POP3
CADM3-AS1
OR10J1
APCS
OR6N1
IFI16
ACKR1
OR10J3
OR6N2
158.8
159
159.2
159.4
159.6
Position on chr1 (Mb)

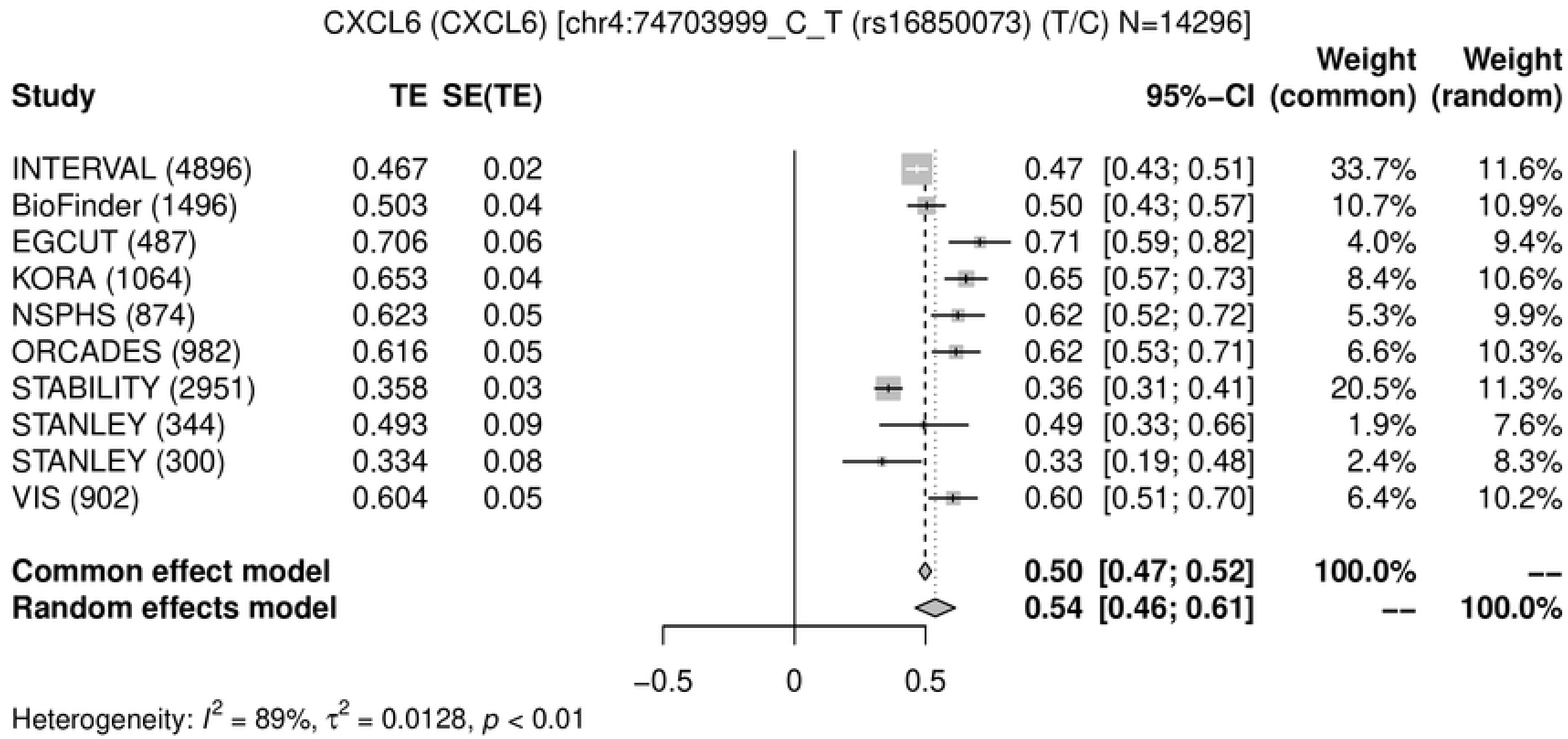

CXCL6 (CXCL6) [chr4:74703999_C_T (rs16850073) (T/C) N=14296]
Study TE SE(TE) 95%−CI Weight (common) Weight (random)
INTERVAL (4896) 0.467 0.02 0.47 [0.43; 0.51] 33.7% 11.6%
BioFinder (1496) 0.503 0.04 0.50 [0.43; 0.57] 10.7% 10.9%
EGCUT (487) 0.706 0.06 0.71 [0.59; 0.82] 4.0% 9.4%
KORA (1064) 0.653 0.04 0.65 [0.57; 0.73] 8.4% 10.6%
NSPHS (874) 0.623 0.05 0.62 [0.52; 0.72] 5.3% 9.9%
ORCADES (982) 0.616 0.05 0.62 [0.53; 0.71] 6.6% 10.3%
STABILITY (2951) 0.358 0.03 0.36 [0.31; 0.41] 20.5% 11.3%
STANLEY (344) 0.493 0.09 0.49 [0.33; 0.66] 1.9% 7.6%
STANLEY (300) 0.334 0.08 0.33 [0.19; 0.48] 2.4% 8.3%
VIS (902) 0.604 0.05 0.60 [0.51; 0.70] 6.4% 10.2%
Common effect model 0.50 [0.47; 0.52] 100.0% --
Random effects model 0.54 [0.46; 0.61] -- 100.0%
−0.5 0 0.5
Heterogeneity: $I^2$ = 89%, $\tau^2$ = 0.0128, $p$ < 0.01


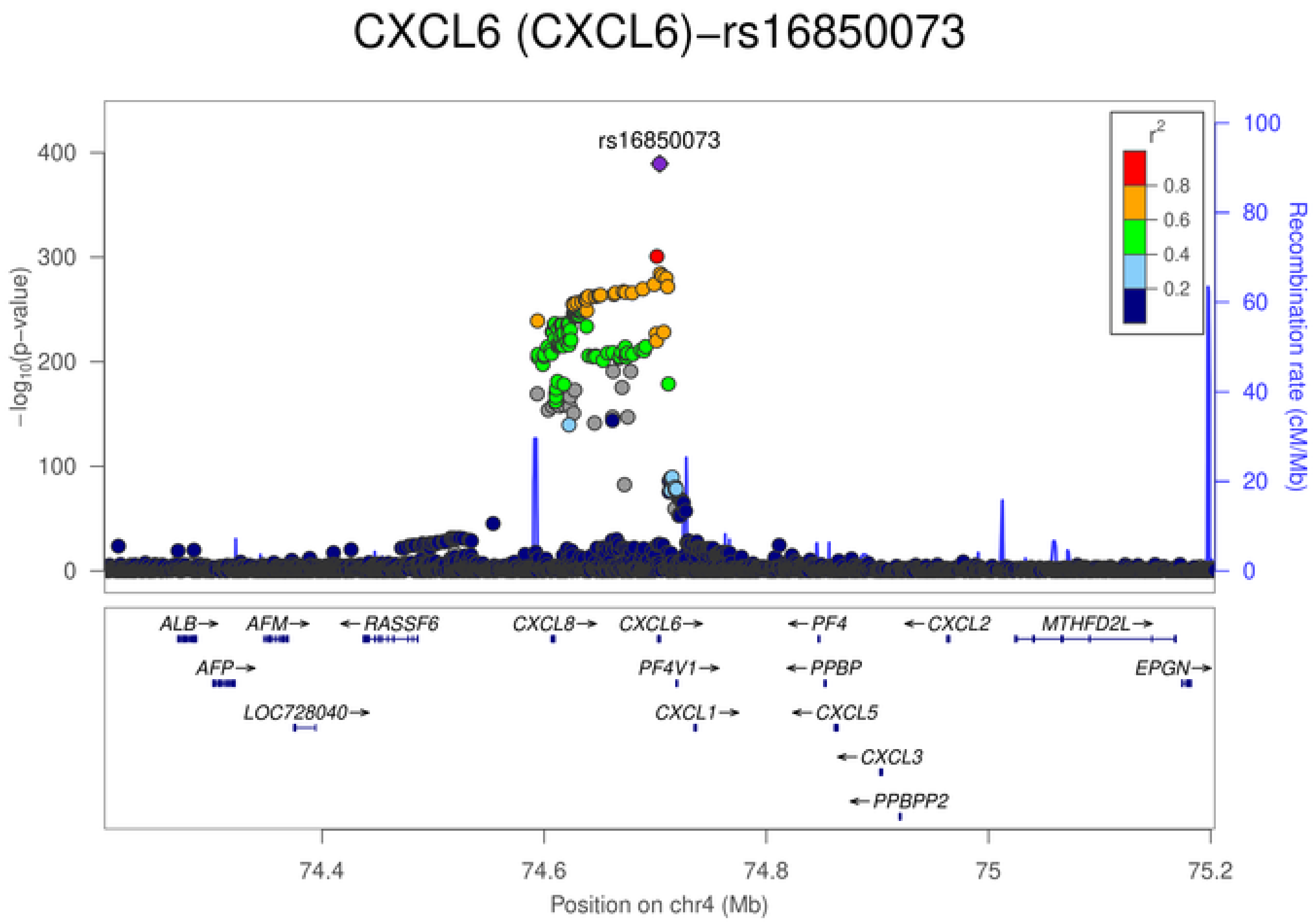

CXCL6 (CXCL6)−rs16850073
rs16850073
$-\log_{10}$(p−value)
Recombination rate (cM/Mb)
$r^2$
0.8
0.6
0.4
0.2
ALB→
AFM→
←RASSF6
CXCL8→
CXCL6→
←PF4
←CXCL2
MTHFD2L→
AFP→
PF4V1→
←PPBP
EPGN→
LOC728040→
CXCL1→
←CXCL5
←CXCL3
←PPBPP2
74.4
74.6
74.8
75
75.2
Position on chr4 (Mb)

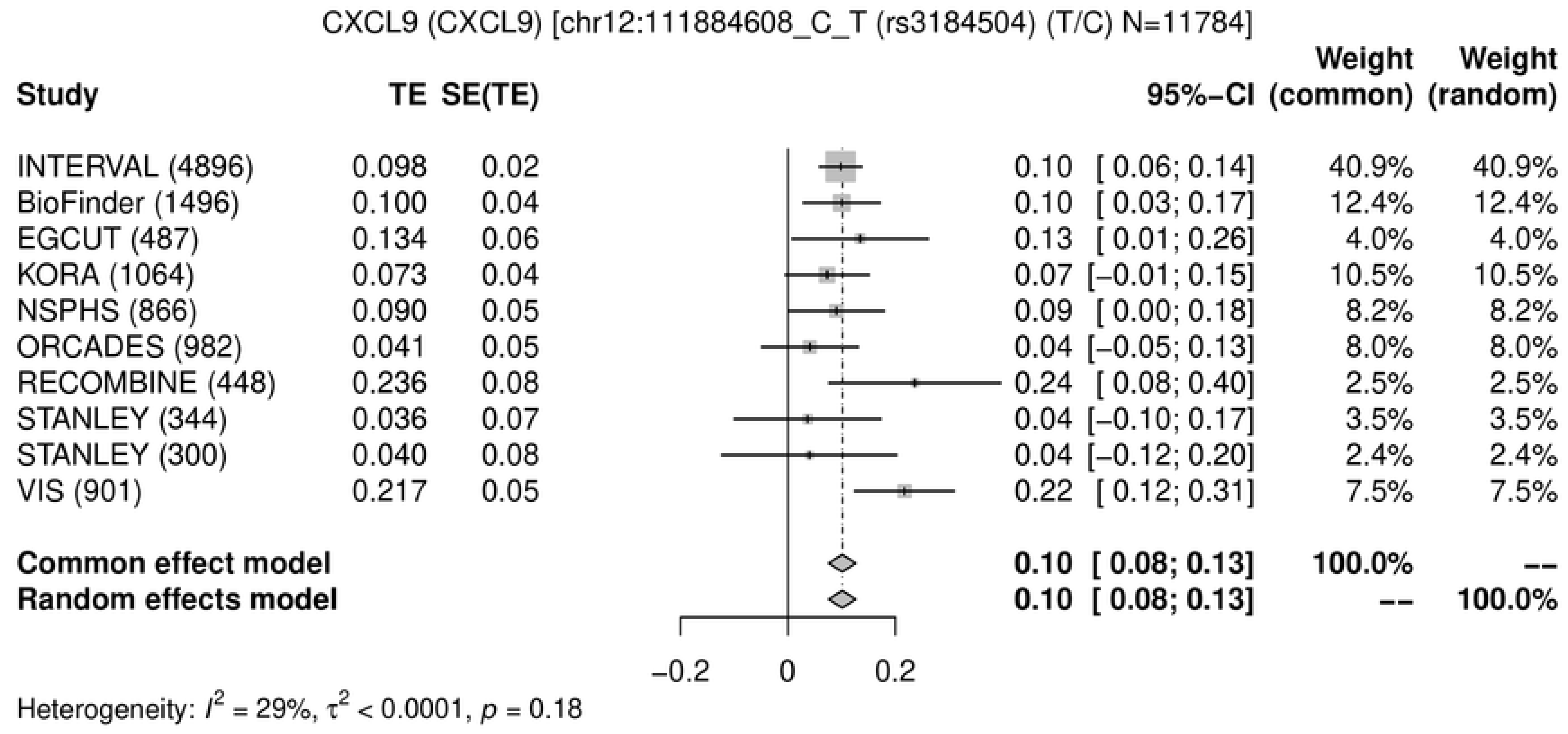
CXCL9 (CXCL9) [chr12:111884608_C_T (rs3184504) (T/C) N=11784]
Study TE SE(TE) 95%-CI Weight (common) Weight (random)
INTERVAL (4896) 0.098 0.02 0.10 [ 0.06; 0.14] 40.9% 40.9%
BioFinder (1496) 0.100 0.04 0.10 [ 0.03; 0.17] 12.4% 12.4%
EGCUT (487) 0.134 0.06 0.13 [ 0.01; 0.26] 4.0% 4.0%
KORA (1064) 0.073 0.04 0.07 [−0.01; 0.15] 10.5% 10.5%
NSPHS (866) 0.090 0.05 0.09 [ 0.00; 0.18] 8.2% 8.2%
ORCADES (982) 0.041 0.05 0.04 [−0.05; 0.13] 8.0% 8.0%
RECOMBINE (448) 0.236 0.08 0.24 [ 0.08; 0.40] 2.5% 2.5%
STANLEY (344) 0.036 0.07 0.04 [−0.10; 0.17] 3.5% 3.5%
STANLEY (300) 0.040 0.08 0.04 [−0.12; 0.20] 2.4% 2.4%
VIS (901) 0.217 0.05 0.22 [ 0.12; 0.31] 7.5% 7.5%
Common effect model 0.10 [ 0.08; 0.13] 100.0% --
Random effects model 0.10 [ 0.08; 0.13] -- 100.0%
−0.2
0
0.2
Heterogeneity: $I^2$ = 29%, $\tau^2$ < 0.0001, $p$ = 0.18

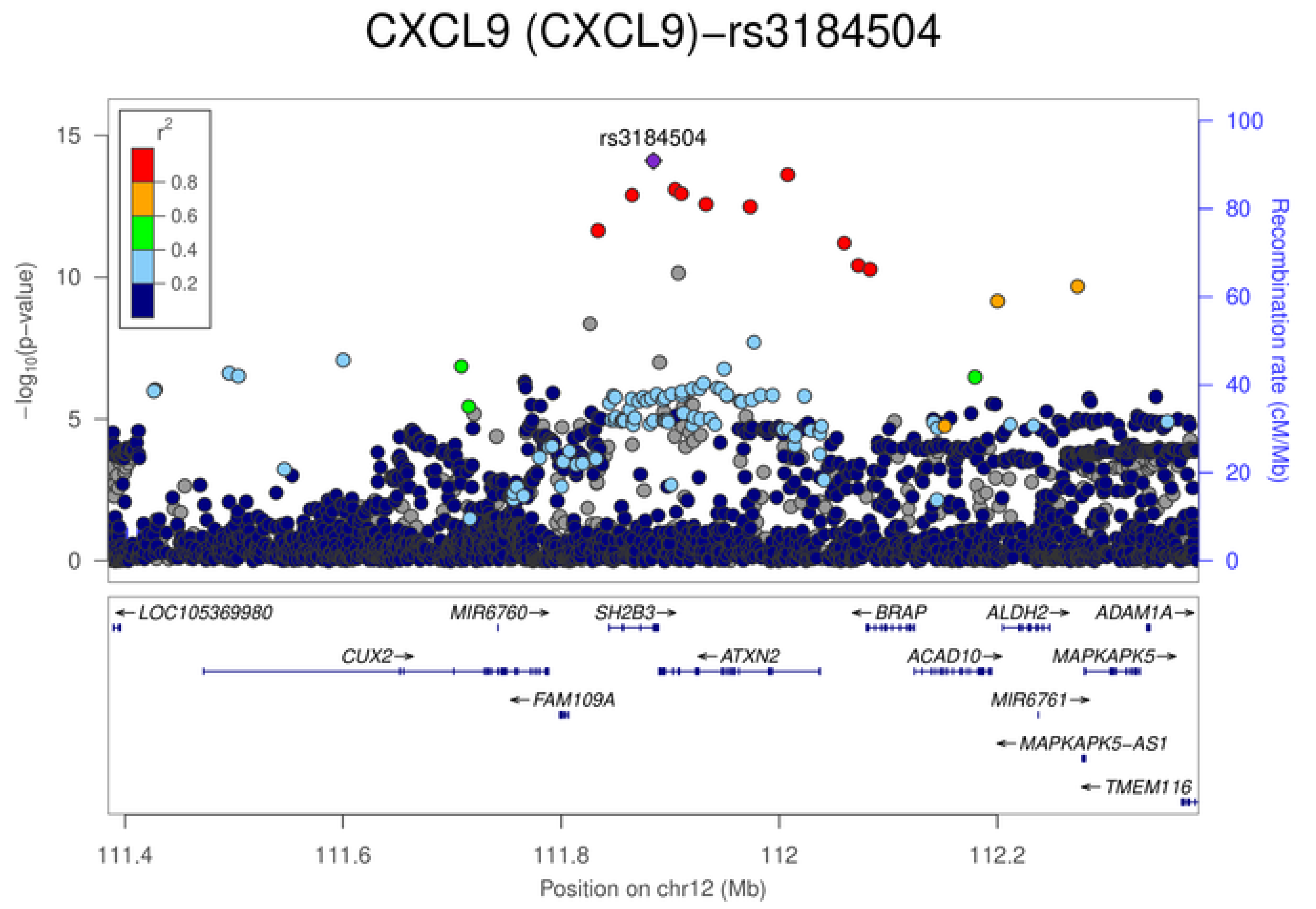
CXCL9 (CXCL9)−rs3184504
rs3184504
$r^2$
0.8
0.6
0.4
0.2
−log10(p−value)
Recombination rate (cM/Mb)
0
5
10
15
0
20
40
60
80
100
LOC105369980
MIR6760
SH2B3
BRAP
ALDH2
ADAM1A
CUX2
ATXN2
ACAD10
MAPKAPK5
FAM109A
MIR6761
MAPKAPK5-AS1
TMEM116
111.4
111.6
111.8
112
112.2
Position on chr12 (Mb)

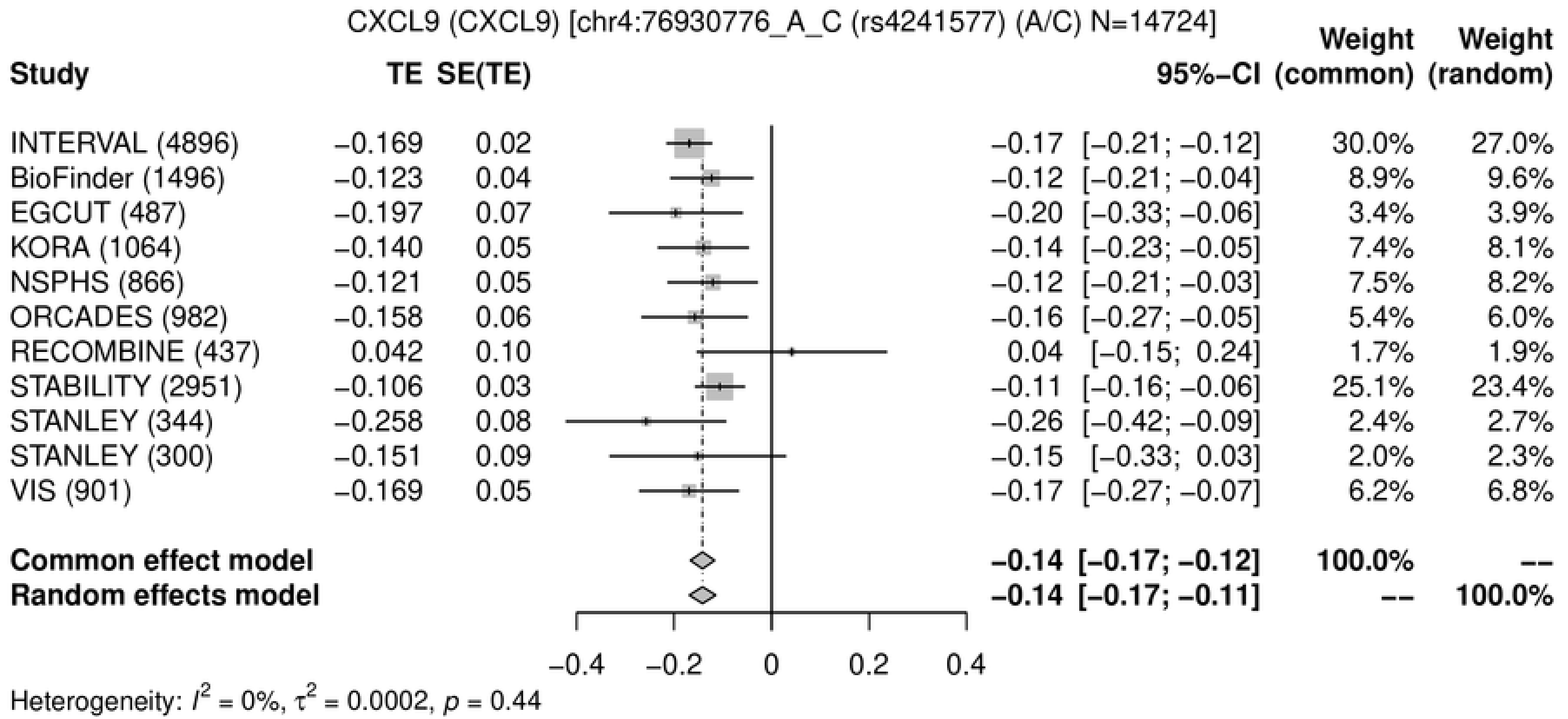
CXCL9 (CXCL9) [chr4:76930776_A_C (rs4241577) (A/C) N=14724]
Study TE SE(TE) 95%−CI Weight (common) Weight (random)
INTERVAL (4896) −0.169 0.02 −0.17 [−0.21; −0.12] 30.0% 27.0%
BioFinder (1496) −0.123 0.04 −0.12 [−0.21; −0.04] 8.9% 9.6%
EGCUT (487) −0.197 0.07 −0.20 [−0.33; −0.06] 3.4% 3.9%
KORA (1064) −0.140 0.05 −0.14 [−0.23; −0.05] 7.4% 8.1%
NSPHS (866) −0.121 0.05 −0.12 [−0.21; −0.03] 7.5% 8.2%
ORCADES (982) −0.158 0.06 −0.16 [−0.27; −0.05] 5.4% 6.0%
RECOMBINE (437) 0.042 0.10 0.04 [−0.15; 0.24] 1.7% 1.9%
STABILITY (2951) −0.106 0.03 −0.11 [−0.16; −0.06] 25.1% 23.4%
STANLEY (344) −0.258 0.08 −0.26 [−0.42; −0.09] 2.4% 2.7%
STANLEY (300) −0.151 0.09 −0.15 [−0.33; 0.03] 2.0% 2.3%
VIS (901) −0.169 0.05 −0.17 [−0.27; −0.07] 6.2% 6.8%
Common effect model −0.14 [−0.17; −0.12] 100.0% --
Random effects model −0.14 [−0.17; −0.11] -- 100.0%
−0.4 −0.2 0 0.2 0.4
Heterogeneity: $I^2$ = 0%, $\tau^2$ = 0.0002, $p$ = 0.44

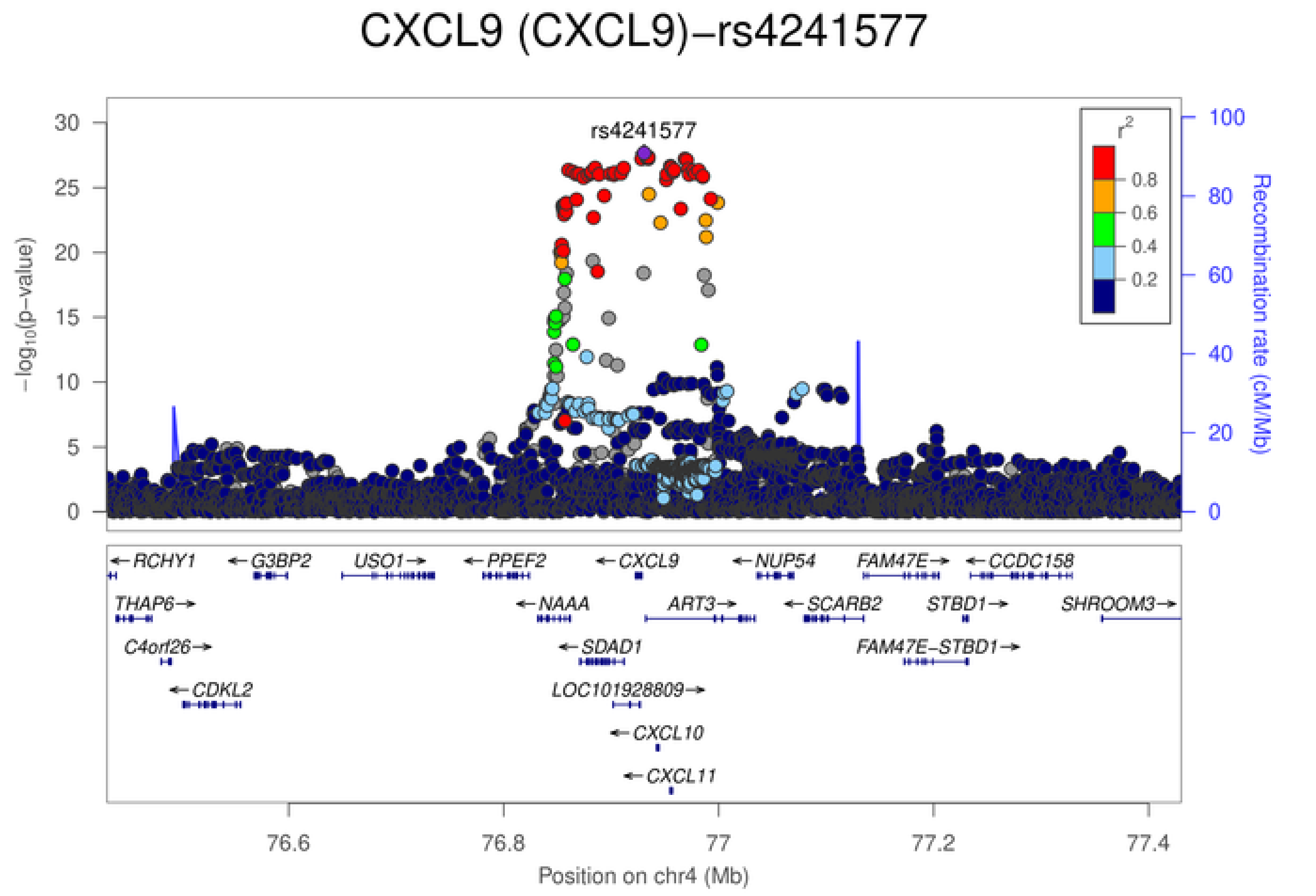
CXCL9 (CXCL9)−rs4241577
rs4241577
$-\log_{10}$(p-value)
Recombination rate (cM/Mb)
$r^2$
0.8
0.6
0.4
0.2
0 5 10 15 20 25 30
0 20 40 60 80 100
RCHY1
G3BP2
USO1
PPEF2
CXCL9
NUP54
FAM47E
CCDC158
THAP6
NAAA
ART3
SCARB2
STBD1
SHROOM3
C4orf26
SDAD1
FAM47E−STBD1
CDKL2
LOC101928809
CXCL10
CXCL11
76.6 76.8 77 77.2 77.4
Position on chr4 (Mb)

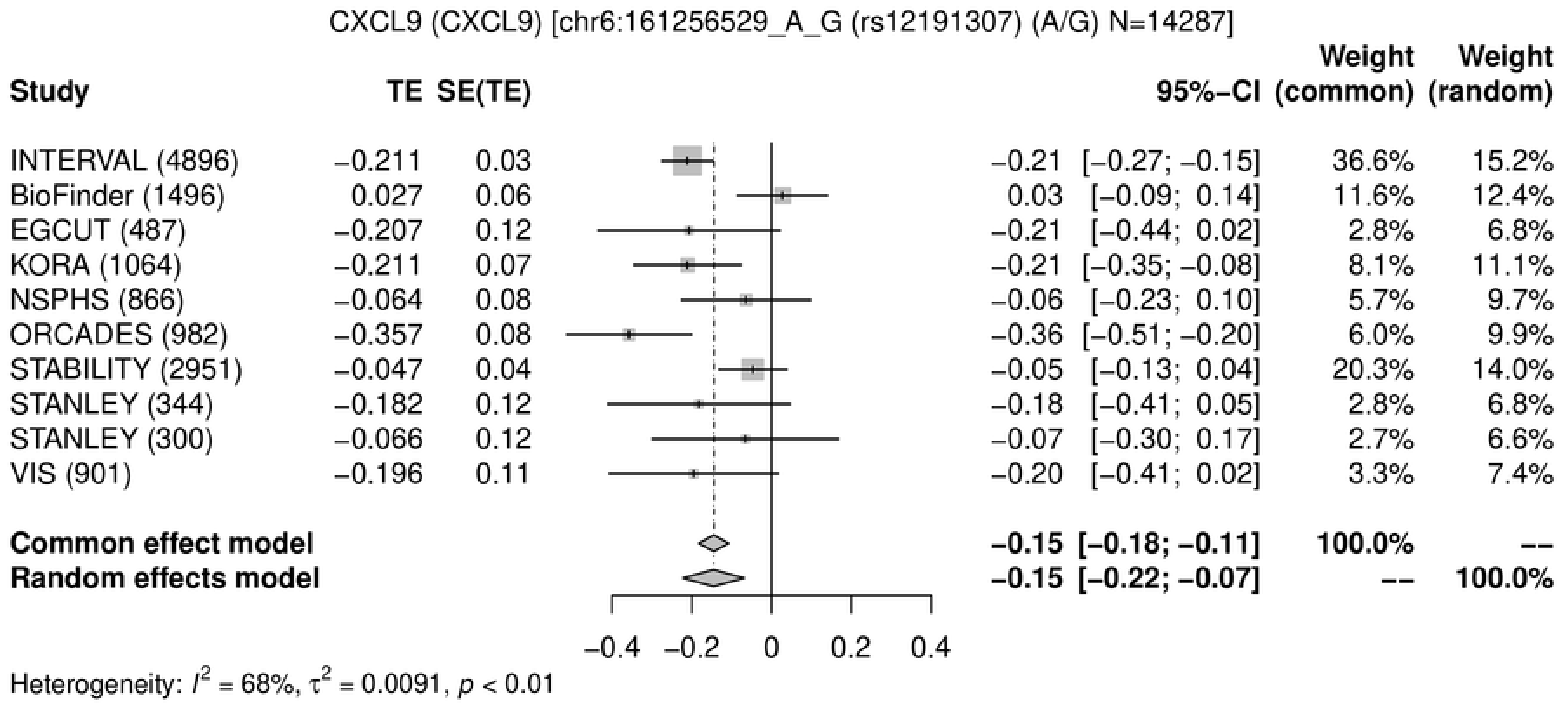
CXCL9 (CXCL9) [chr6:161256529_A_G (rs12191307) (A/G) N=14287]
Study
TE
SE(TE)
95%-CI
Weight (common)
Weight (random)
INTERVAL (4896) −0.211 0.03 −0.21 [−0.27; −0.15] 36.6% 15.2%
BioFinder (1496) 0.027 0.06 0.03 [−0.09; 0.14] 11.6% 12.4%
EGCUT (487) −0.207 0.12 −0.21 [−0.44; 0.02] 2.8% 6.8%
KORA (1064) −0.211 0.07 −0.21 [−0.35; −0.08] 8.1% 11.1%
NSPHS (866) −0.064 0.08 −0.06 [−0.23; 0.10] 5.7% 9.7%
ORCADES (982) −0.357 0.08 −0.36 [−0.51; −0.20] 6.0% 9.9%
STABILITY (2951) −0.047 0.04 −0.05 [−0.13; 0.04] 20.3% 14.0%
STANLEY (344) −0.182 0.12 −0.18 [−0.41; 0.05] 2.8% 6.8%
STANLEY (300) −0.066 0.12 −0.07 [−0.30; 0.17] 2.7% 6.6%
VIS (901) −0.196 0.11 −0.20 [−0.41; 0.02] 3.3% 7.4%
Common effect model −0.15 [−0.18; −0.11] 100.0% --
Random effects model −0.15 [−0.22; −0.07] -- 100.0%
−0.4 −0.2 0 0.2 0.4
Heterogeneity: $I^2$ = 68%, $\tau^2$ = 0.0091, $p$ < 0.01

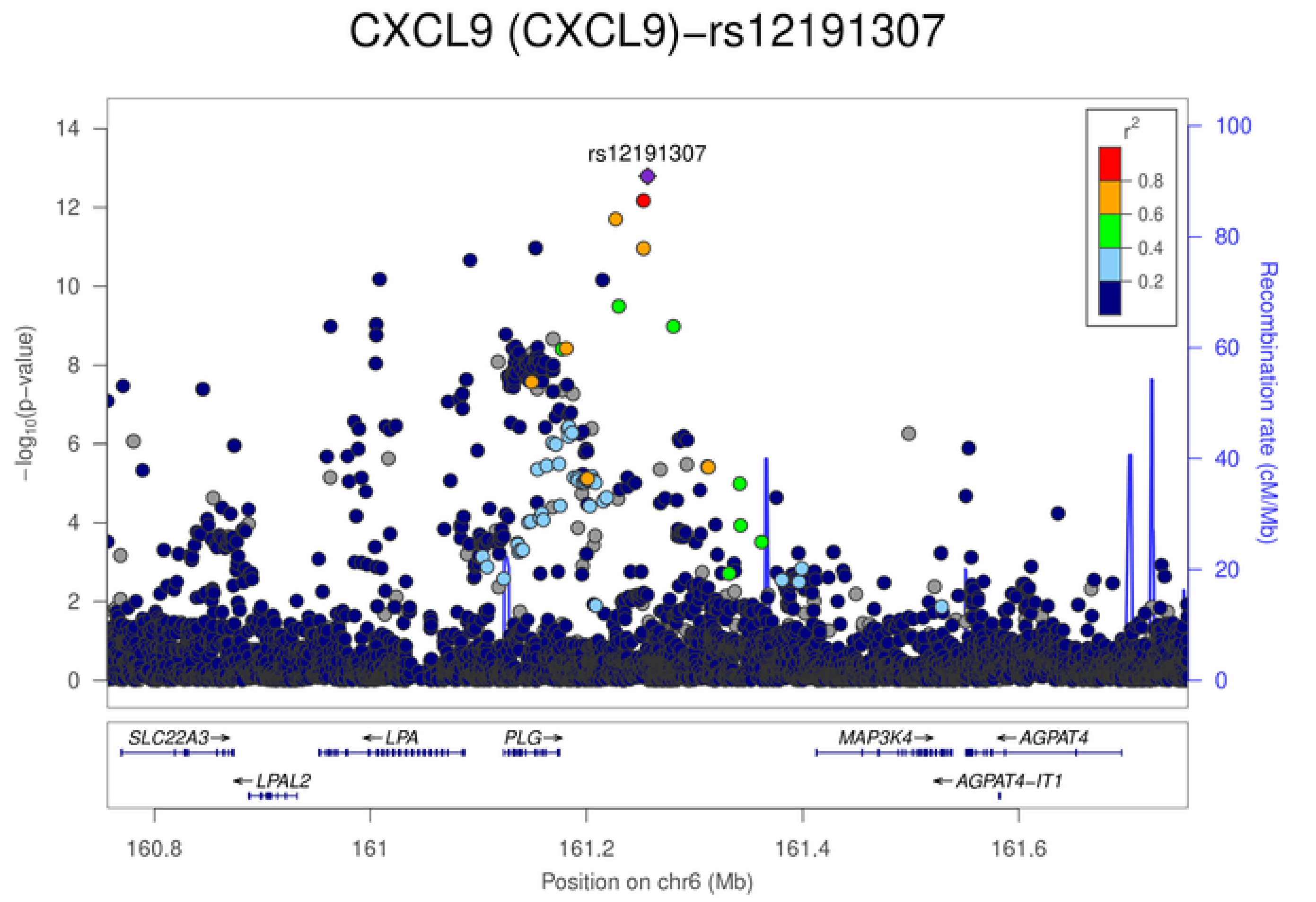
CXCL9 (CXCL9)−rs12191307
rs12191307
$-\log_{10}$(p-value)
Recombination rate (cM/Mb)
$r^2$
0.8
0.6
0.4
0.2
SLC22A3
LPA
PLG
MAP3K4
AGPAT4
LPAL2
AGPAT4-IT1
160.8
161
161.2
161.4
161.6
Position on chr6 (Mb)

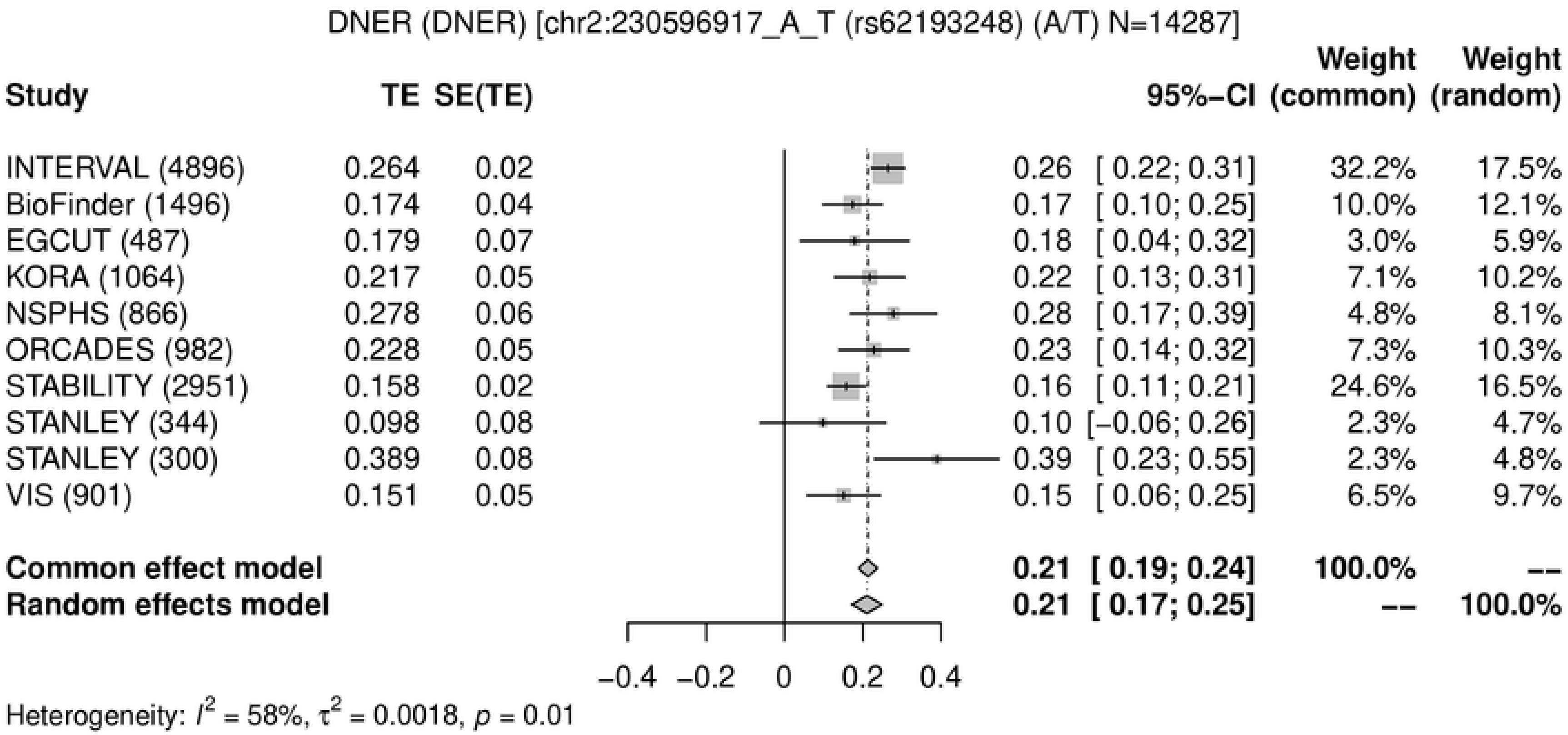
DNER (DNER) [chr2:230596917_A_T (rs62193248) (A/T) N=14287]
Study
TE
SE(TE)
95%−CI
Weight (common)
Weight (random)
INTERVAL (4896) 0.264 0.02 0.26 [ 0.22; 0.31] 32.2% 17.5%
BioFinder (1496) 0.174 0.04 0.17 [ 0.10; 0.25] 10.0% 12.1%
EGCUT (487) 0.179 0.07 0.18 [ 0.04; 0.32] 3.0% 5.9%
KORA (1064) 0.217 0.05 0.22 [ 0.13; 0.31] 7.1% 10.2%
NSPHS (866) 0.278 0.06 0.28 [ 0.17; 0.39] 4.8% 8.1%
ORCADES (982) 0.228 0.05 0.23 [ 0.14; 0.32] 7.3% 10.3%
STABILITY (2951) 0.158 0.02 0.16 [ 0.11; 0.21] 24.6% 16.5%
STANLEY (344) 0.098 0.08 0.10 [−0.06; 0.26] 2.3% 4.7%
STANLEY (300) 0.389 0.08 0.39 [ 0.23; 0.55] 2.3% 4.8%
VIS (901) 0.151 0.05 0.15 [ 0.06; 0.25] 6.5% 9.7%
Common effect model 0.21 [ 0.19; 0.24] 100.0% --
Random effects model 0.21 [ 0.17; 0.25] -- 100.0%
−0.4 −0.2 0 0.2 0.4
Heterogeneity: $I^2$ = 58%, $\tau^2$ = 0.0018, $p$ = 0.01

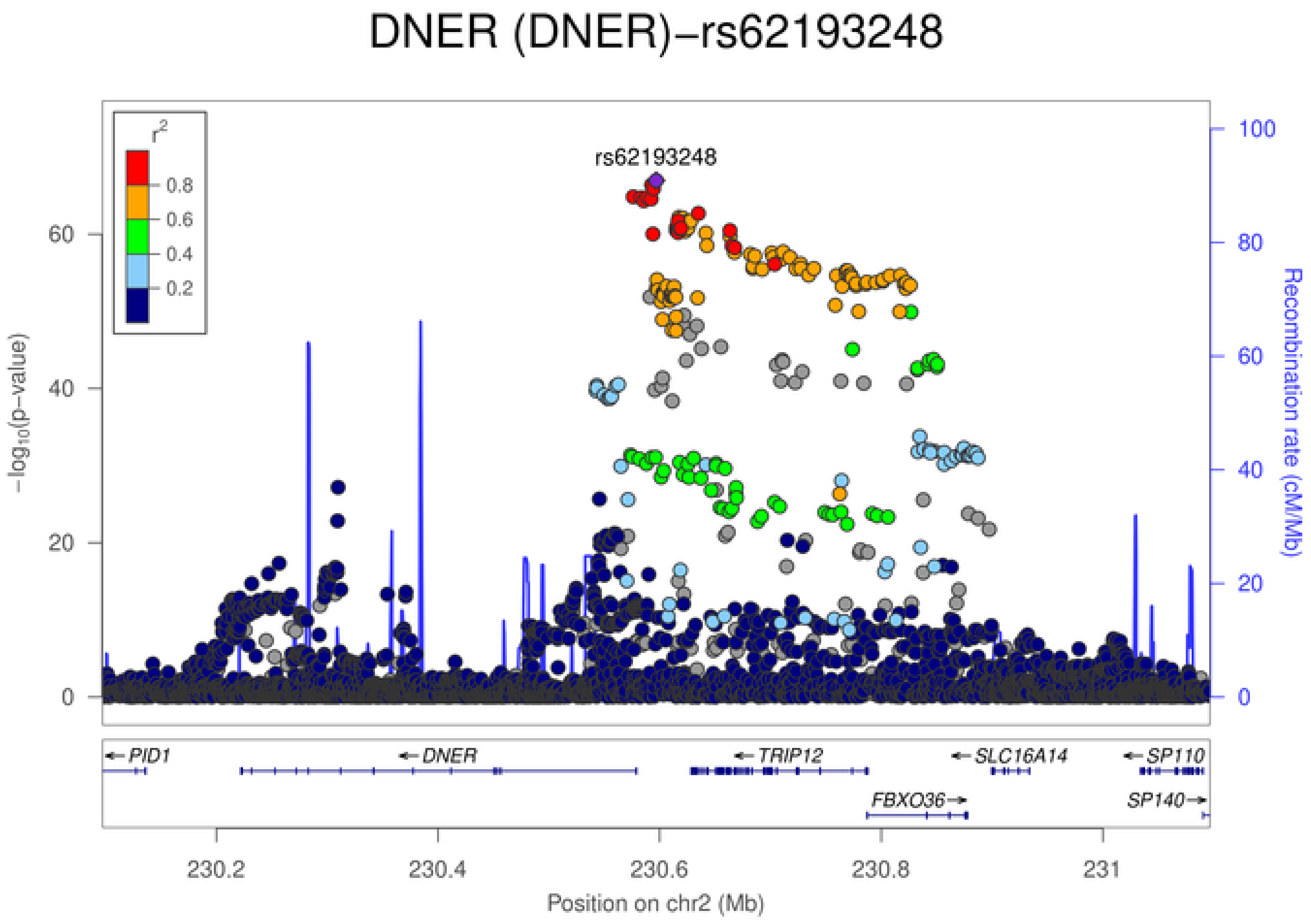
DNER (DNER)−rs62193248
rs62193248
$r^2$
0.8
0.6
0.4
0.2
$-\log_{10}$(p−value)
Recombination rate (cM/Mb)
0
20
40
60
80
100
PID1
DNER
TRIP12
SLC16A14
SP110
FBXO36
SP140
230.2
230.4
230.6
230.8
231
Position on chr2 (Mb)

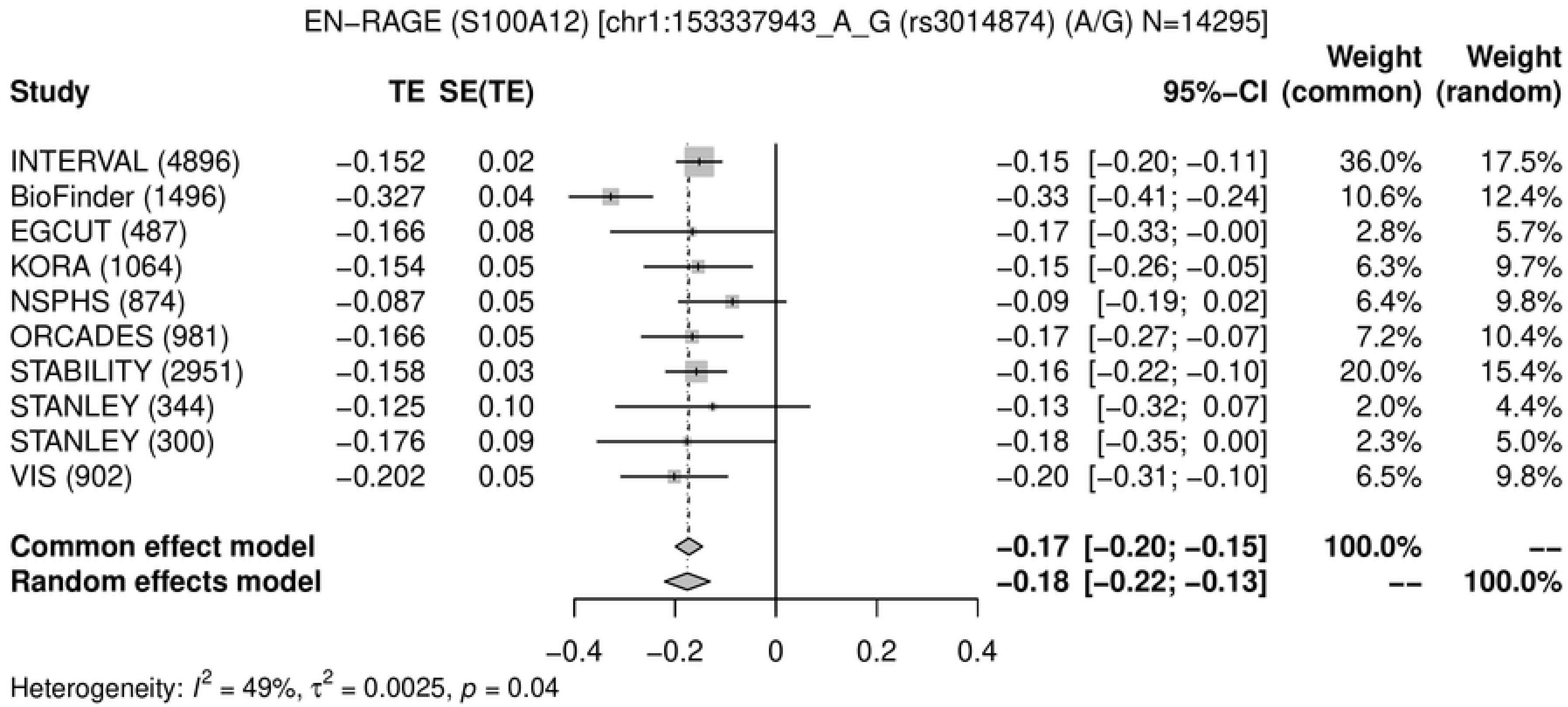
EN−RAGE (S100A12) [chr1:153337943_A_G (rs3014874) (A/G) N=14295]
Study
TE
SE(TE)
95%−CI
Weight (common)
Weight (random)
INTERVAL (4896) −0.152 0.02 −0.15 [−0.20; −0.11] 36.0% 17.5%
BioFinder (1496) −0.327 0.04 −0.33 [−0.41; −0.24] 10.6% 12.4%
EGCUT (487) −0.166 0.08 −0.17 [−0.33; −0.00] 2.8% 5.7%
KORA (1064) −0.154 0.05 −0.15 [−0.26; −0.05] 6.3% 9.7%
NSPHS (874) −0.087 0.05 −0.09 [−0.19; 0.02] 6.4% 9.8%
ORCADES (981) −0.166 0.05 −0.17 [−0.27; −0.07] 7.2% 10.4%
STABILITY (2951) −0.158 0.03 −0.16 [−0.22; −0.10] 20.0% 15.4%
STANLEY (344) −0.125 0.10 −0.13 [−0.32; 0.07] 2.0% 4.4%
STANLEY (300) −0.176 0.09 −0.18 [−0.35; 0.00] 2.3% 5.0%
VIS (902) −0.202 0.05 −0.20 [−0.31; −0.10] 6.5% 9.8%
Common effect model −0.17 [−0.20; −0.15] 100.0% --
Random effects model −0.18 [−0.22; −0.13] -- 100.0%
−0.4 −0.2 0 0.2 0.4
Heterogeneity: $I^2$ = 49%, $\tau^2$ = 0.0025, $p$ = 0.04

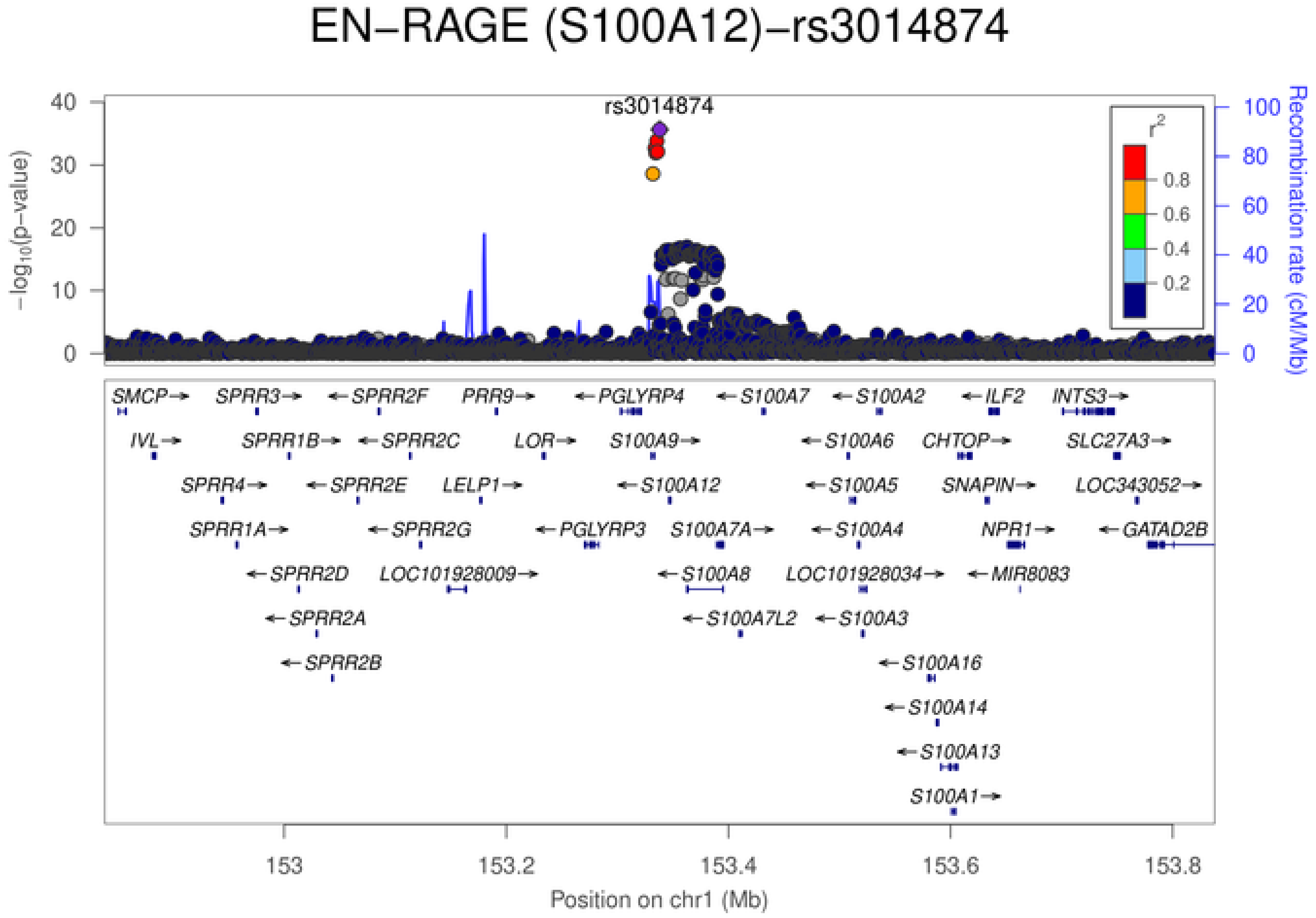
EN−RAGE (S100A12)−rs3014874
rs3014874
$-\log_{10}$(p−value)
Recombination rate (cM/Mb)
$r^2$
0.8
0.6
0.4
0.2
SMCP SPRR3 SPRR2F PRR9 PGLYRP4 S100A7 S100A2 ILF2 INTS3
IVL SPRR1B SPRR2C LOR S100A9 S100A6 CHTOP SLC27A3
SPRR4 SPRR2E LELP1 S100A12 S100A5 SNAPIN LOC343052
SPRR1A SPRR2G PGLYRP3 S100A7A S100A4 NPR1 GATAD2B
SPRR2D LOC101928009 S100A8 LOC101928034 MIR8083
SPRR2A S100A7L2 S100A3
SPRR2B S100A16
S100A14
S100A13
S100A1
153 153.2 153.4 153.6 153.8
Position on chr1 (Mb)

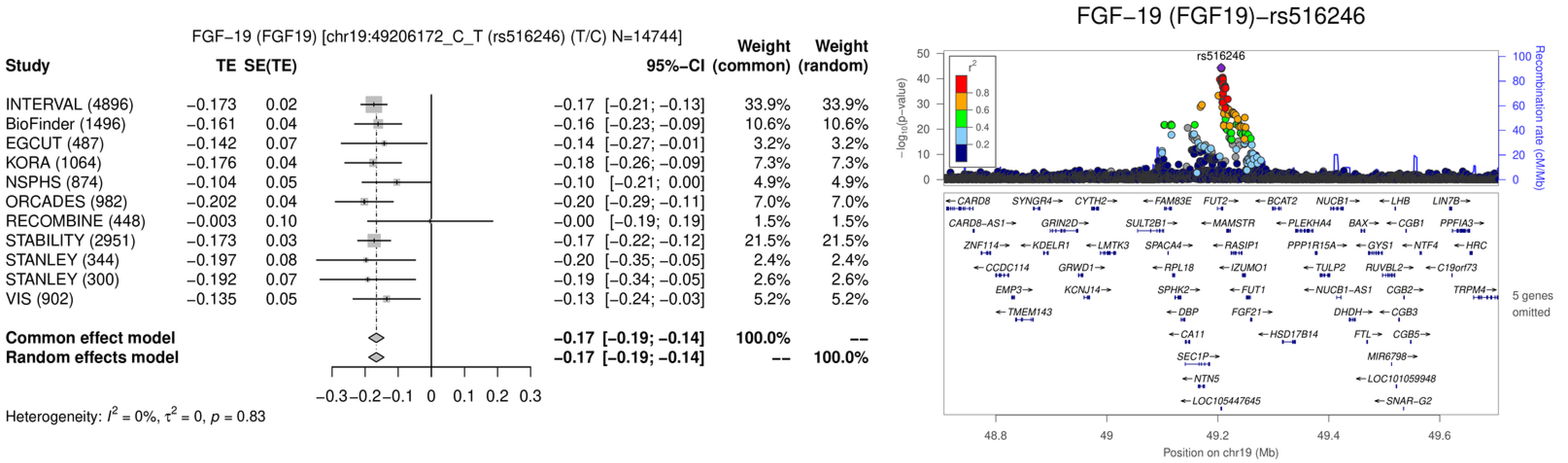
FGF−19 (FGF19) [chr19:49206172_C_T (rs516246) (T/C) N=14744]
Study
TE
SE(TE)
95%−CI
Weight (common)
Weight (random)
INTERVAL (4896) −0.173 0.02 −0.17 [−0.21; −0.13] 33.9% 33.9%
BioFinder (1496) −0.161 0.04 −0.16 [−0.23; −0.09] 10.6% 10.6%
EGCUT (487) −0.142 0.07 −0.14 [−0.27; −0.01] 3.2% 3.2%
KORA (1064) −0.176 0.04 −0.18 [−0.26; −0.09] 7.3% 7.3%
NSPHS (874) −0.104 0.05 −0.10 [−0.21; 0.00] 4.9% 4.9%
ORCADES (982) −0.202 0.04 −0.20 [−0.29; −0.11] 7.0% 7.0%
RECOMBINE (448) −0.003 0.10 −0.00 [−0.19; 0.19] 1.5% 1.5%
STABILITY (2951) −0.173 0.03 −0.17 [−0.22; −0.12] 21.5% 21.5%
STANLEY (344) −0.197 0.08 −0.20 [−0.35; −0.05] 2.4% 2.4%
STANLEY (300) −0.192 0.07 −0.19 [−0.34; −0.05] 2.6% 2.6%
VIS (902) −0.135 0.05 −0.13 [−0.24; −0.03] 5.2% 5.2%
Common effect model −0.17 [−0.19; −0.14] 100.0% --
Random effects model −0.17 [−0.19; −0.14] -- 100.0%
−0.3 −0.2 −0.1 0 0.1 0.2 0.3
Heterogeneity: $I^2$ = 0%, $\tau^2$ = 0, $p$ = 0.83
FGF−19 (FGF19)−rs516246
rs516246
$-\log_{10}$(p−value)
Recombination rate (cM/Mb)
$r^2$
0.8
0.6
0.4
0.2
CARD8, SYNGR4, CYTH2, FAM83E, FUT2, BCAT2, NUCB1, LHB, LIN7B
CARD8−AS1, GRIN2D, SULT2B1, MAMSTR, PLEKHA4, BAX, CGB1, PPFIA3
ZNF114, KDELR1, LMTK3, SPACA4, RASIP1, PPP1R15A, GYS1, NTF4, HRC
CCDC114, GRWD1, RPL18, IZUMO1, TULP2, RUVBL2, C19orf73
EMP3, KCNJ14, SPHK2, FUT1, NUCB1−AS1, CGB2, TRPM4
TMEM143, DBP, FGF21, DHDH, CGB3
CA11, HSD17B14, FTL, CGB5
SEC1P, MIR6798
NTN5, LOC101059948
LOC105447645, SNAR−G2
5 genes omitted
48.8
49
49.2
49.4
49.6
Position on chr19 (Mb)

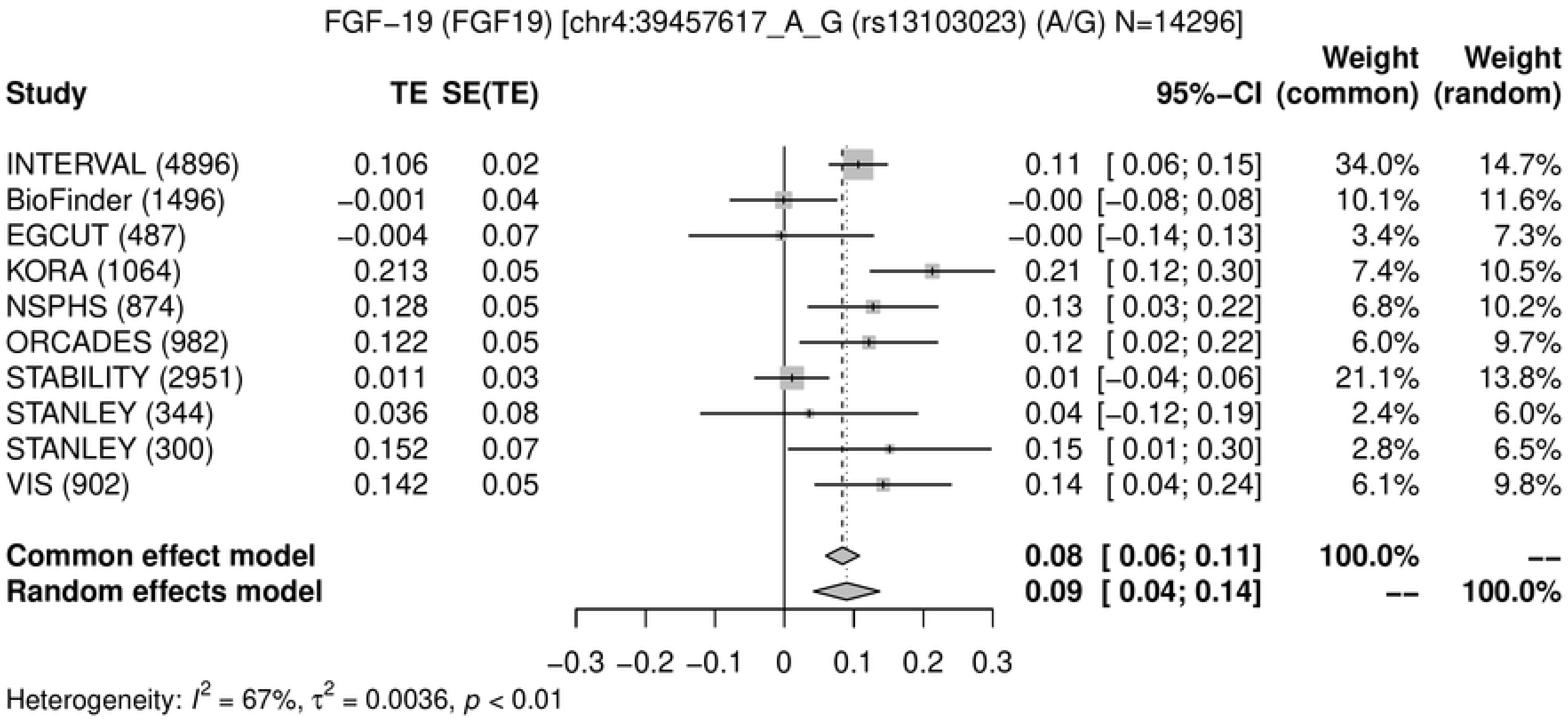
FGF−19 (FGF19) [chr4:39457617_A_G (rs13103023) (A/G) N=14296]
Study TE SE(TE) 95%−CI Weight (common) Weight (random)
INTERVAL (4896) 0.106 0.02 0.11 [ 0.06; 0.15] 34.0% 14.7%
BioFinder (1496) −0.001 0.04 −0.00 [−0.08; 0.08] 10.1% 11.6%
EGCUT (487) −0.004 0.07 −0.00 [−0.14; 0.13] 3.4% 7.3%
KORA (1064) 0.213 0.05 0.21 [ 0.12; 0.30] 7.4% 10.5%
NSPHS (874) 0.128 0.05 0.13 [ 0.03; 0.22] 6.8% 10.2%
ORCADES (982) 0.122 0.05 0.12 [ 0.02; 0.22] 6.0% 9.7%
STABILITY (2951) 0.011 0.03 0.01 [−0.04; 0.06] 21.1% 13.8%
STANLEY (344) 0.036 0.08 0.04 [−0.12; 0.19] 2.4% 6.0%
STANLEY (300) 0.152 0.07 0.15 [ 0.01; 0.30] 2.8% 6.5%
VIS (902) 0.142 0.05 0.14 [ 0.04; 0.24] 6.1% 9.8%
Common effect model 0.08 [ 0.06; 0.11] 100.0% --
Random effects model 0.09 [ 0.04; 0.14] -- 100.0%
−0.3 −0.2 −0.1 0 0.1 0.2 0.3
Heterogeneity: $I^2$ = 67%, $\tau^2$ = 0.0036, $p$ < 0.01

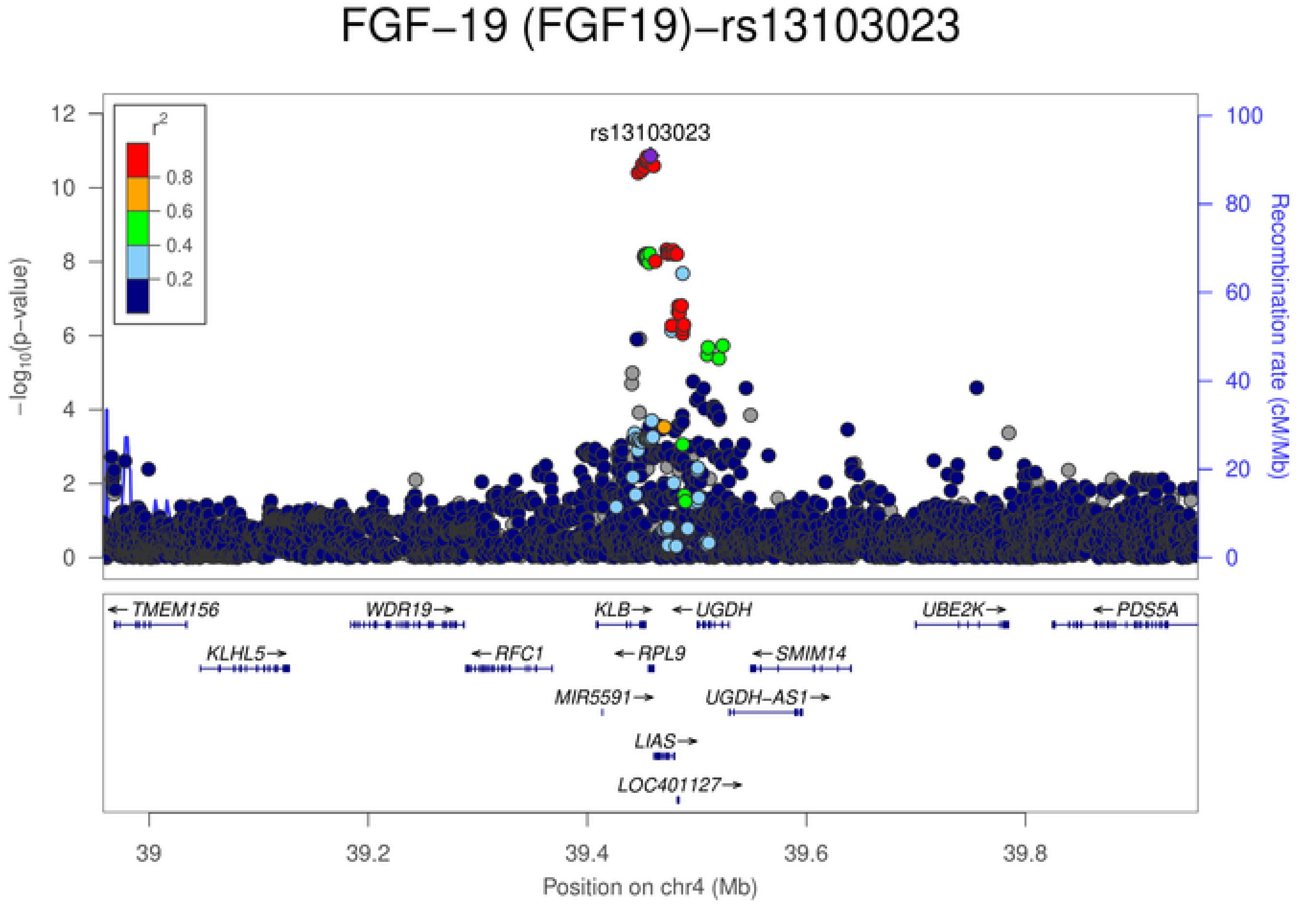
FGF−19 (FGF19)−rs13103023
rs13103023
$r^2$
0.8
0.6
0.4
0.2
$-\log_{10}$(p−value)
Recombination rate (cM/Mb)
TMEM156
WDR19
KLB
UGDH
UBE2K
PDS5A
KLHL5
RFC1
RPL9
SMIM14
MIR5591
UGDH−AS1
LIAS
LOC401127
39 39.2 39.4 39.6 39.8
Position on chr4 (Mb)

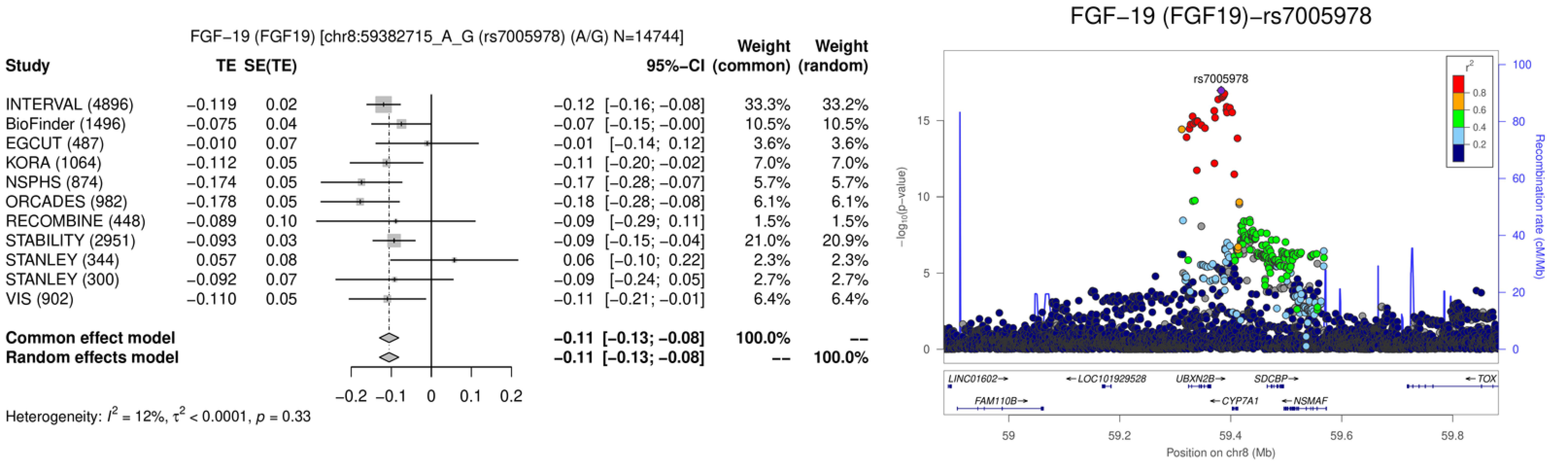
FGF−19 (FGF19) [chr8:59382715_A_G (rs7005978) (A/G) N=14744]
Study
TE
SE(TE)
95%−CI
Weight (common)
Weight (random)
INTERVAL (4896) −0.119 0.02 −0.12 [−0.16; −0.08] 33.3% 33.2%
BioFinder (1496) −0.075 0.04 −0.07 [−0.15; −0.00] 10.5% 10.5%
EGCUT (487) −0.010 0.07 −0.01 [−0.14; 0.12] 3.6% 3.6%
KORA (1064) −0.112 0.05 −0.11 [−0.20; −0.02] 7.0% 7.0%
NSPHS (874) −0.174 0.05 −0.17 [−0.28; −0.07] 5.7% 5.7%
ORCADES (982) −0.178 0.05 −0.18 [−0.28; −0.08] 6.1% 6.1%
RECOMBINE (448) −0.089 0.10 −0.09 [−0.29; 0.11] 1.5% 1.5%
STABILITY (2951) −0.093 0.03 −0.09 [−0.15; −0.04] 21.0% 20.9%
STANLEY (344) 0.057 0.08 0.06 [−0.10; 0.22] 2.3% 2.3%
STANLEY (300) −0.092 0.07 −0.09 [−0.24; 0.05] 2.7% 2.7%
VIS (902) −0.110 0.05 −0.11 [−0.21; −0.01] 6.4% 6.4%
Common effect model −0.11 [−0.13; −0.08] 100.0% --
Random effects model −0.11 [−0.13; −0.08] -- 100.0%
−0.2 −0.1 0 0.1 0.2
Heterogeneity: $I^2$ = 12%, $\tau^2$ < 0.0001, $p$ = 0.33
FGF−19 (FGF19)−rs7005978
rs7005978
$r^2$
0.8
0.6
0.4
0.2
0
5
10
15
$-\log_{10}$(p−value)
0
20
40
60
80
100
Recombination rate (cM/Mb)
LINC01602
LOC101929528
UBXN2B
SDCBP
TOX
FAM110B
CYP7A1
NSMAF
59
59.2
59.4
59.6
59.8
Position on chr8 (Mb)

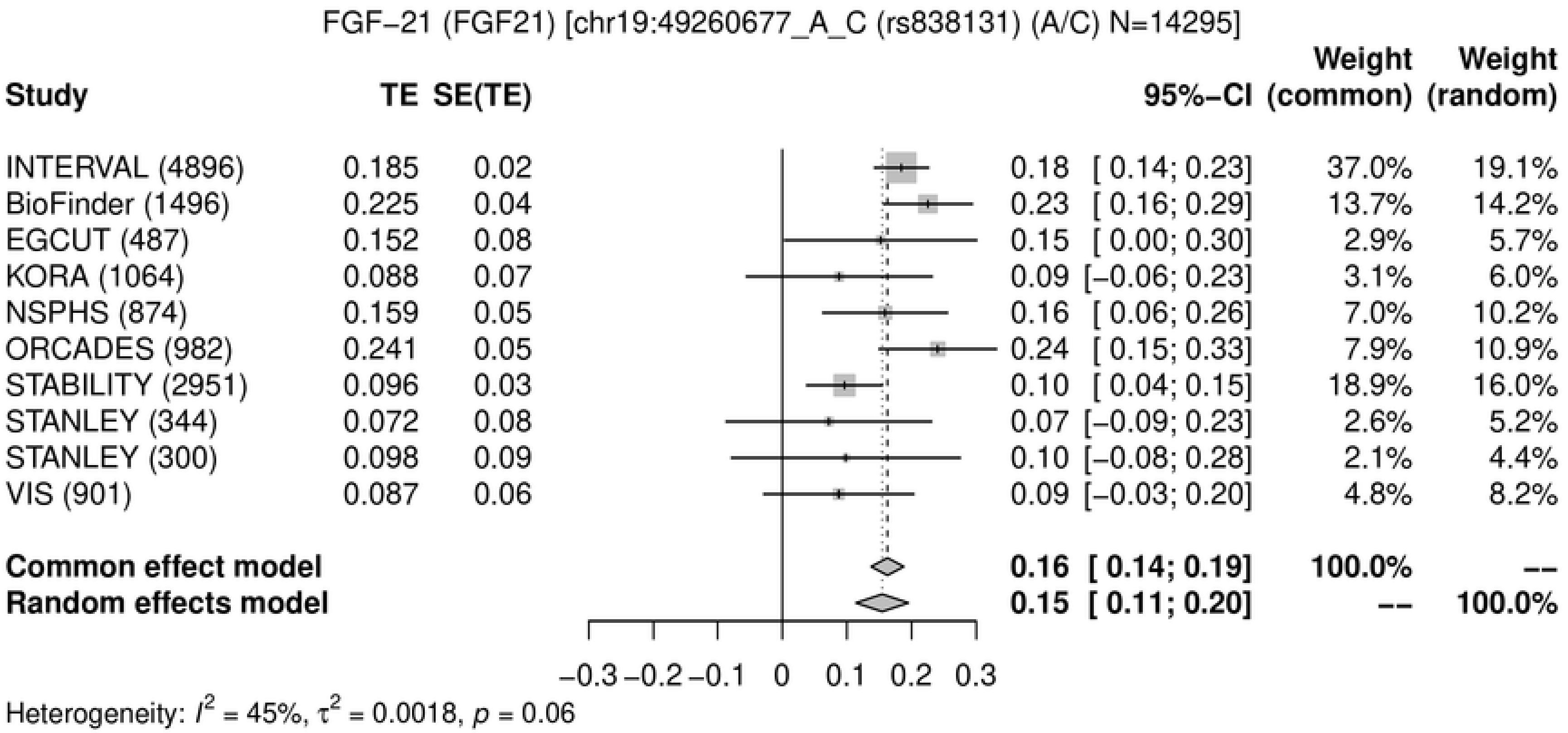
FGF−21 (FGF21) [chr19:49260677_A_C (rs838131) (A/C) N=14295]
Study
TE
SE(TE)
95%−CI
Weight (common)
Weight (random)
INTERVAL (4896) 0.185 0.02 0.18 [ 0.14; 0.23] 37.0% 19.1%
BioFinder (1496) 0.225 0.04 0.23 [ 0.16; 0.29] 13.7% 14.2%
EGCUT (487) 0.152 0.08 0.15 [ 0.00; 0.30] 2.9% 5.7%
KORA (1064) 0.088 0.07 0.09 [−0.06; 0.23] 3.1% 6.0%
NSPHS (874) 0.159 0.05 0.16 [ 0.06; 0.26] 7.0% 10.2%
ORCADES (982) 0.241 0.05 0.24 [ 0.15; 0.33] 7.9% 10.9%
STABILITY (2951) 0.096 0.03 0.10 [ 0.04; 0.15] 18.9% 16.0%
STANLEY (344) 0.072 0.08 0.07 [−0.09; 0.23] 2.6% 5.2%
STANLEY (300) 0.098 0.09 0.10 [−0.08; 0.28] 2.1% 4.4%
VIS (901) 0.087 0.06 0.09 [−0.03; 0.20] 4.8% 8.2%
Common effect model 0.16 [ 0.14; 0.19] 100.0% --
Random effects model 0.15 [ 0.11; 0.20] -- 100.0%
−0.3 −0.2 −0.1 0 0.1 0.2 0.3
Heterogeneity: $I^2$ = 45%, $\tau^2$ = 0.0018, $p$ = 0.06

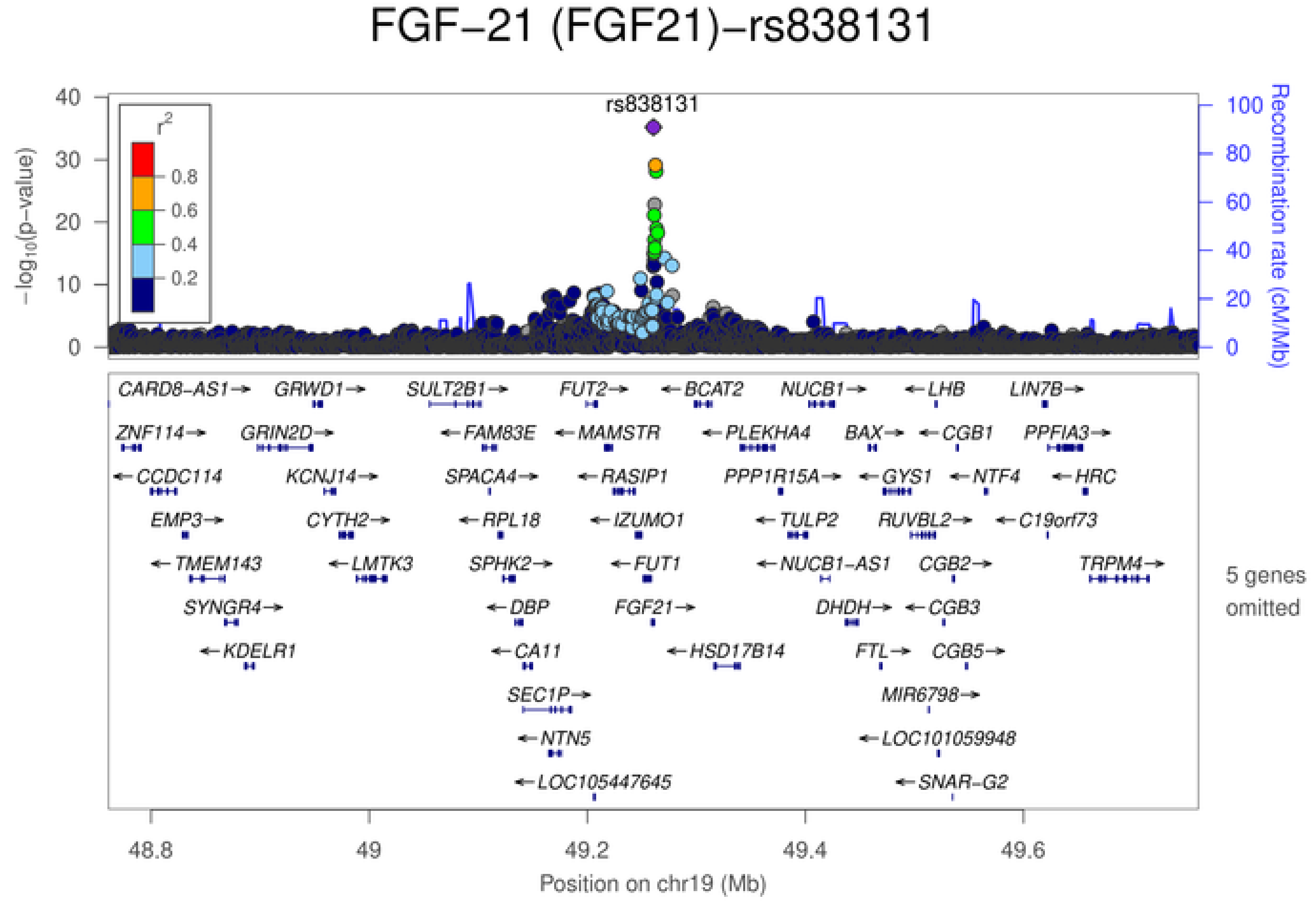
FGF−21 (FGF21)−rs838131
rs838131
$r^2$
0.8
0.6
0.4
0.2
$-\log_{10}(p\text{-value})$
Recombination rate (cM/Mb)
CARD8−AS1 GRWD1 SULT2B1 FUT2 BCAT2 NUCB1 LHB LIN7B
ZNF114 GRIN2D FAM83E MAMSTR PLEKHA4 BAX CGB1 PPFIA3
CCDC114 KCNJ14 SPACA4 RASIP1 PPP1R15A GYS1 NTF4 HRC
EMP3 CYTH2 RPL18 IZUMO1 TULP2 RUVBL2 C19orf73
TMEM143 LMTK3 SPHK2 FUT1 NUCB1−AS1 CGB2 TRPM4
SYNGR4 DBP FGF21 DHDH CGB3
KDELR1 CA11 HSD17B14 FTL CGB5
SEC1P MIR6798
NTN5 LOC101059948
LOC105447645 SNAR−G2
5 genes omitted
48.8 49 49.2 49.4 49.6
Position on chr19 (Mb)

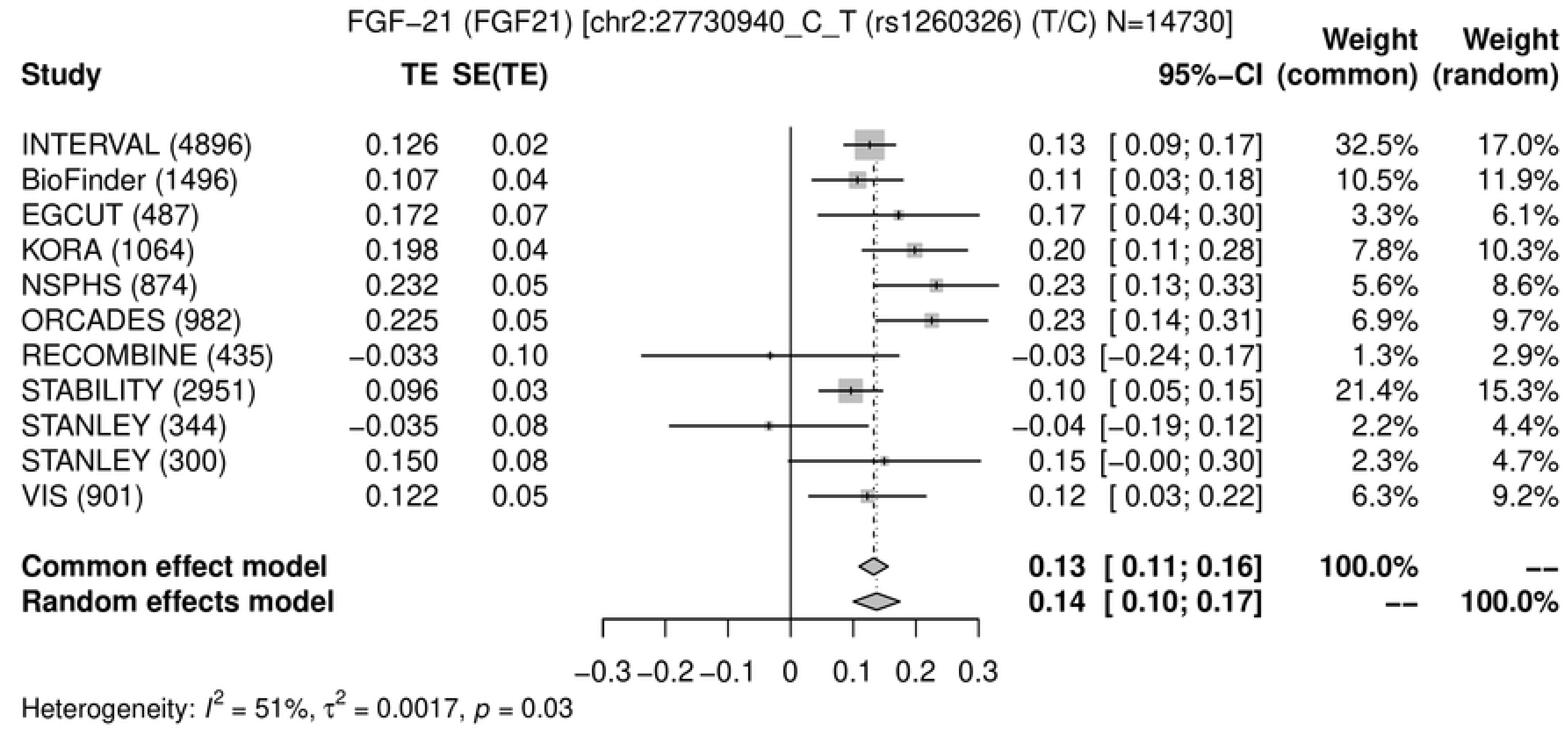
FGF−21 (FGF21) [chr2:27730940_C_T (rs1260326) (T/C) N=14730]
Study
TE
SE(TE)
95%−CI
Weight (common)
Weight (random)
INTERVAL (4896) 0.126 0.02 0.13 [ 0.09; 0.17] 32.5% 17.0%
BioFinder (1496) 0.107 0.04 0.11 [ 0.03; 0.18] 10.5% 11.9%
EGCUT (487) 0.172 0.07 0.17 [ 0.04; 0.30] 3.3% 6.1%
KORA (1064) 0.198 0.04 0.20 [ 0.11; 0.28] 7.8% 10.3%
NSPHS (874) 0.232 0.05 0.23 [ 0.13; 0.33] 5.6% 8.6%
ORCADES (982) 0.225 0.05 0.23 [ 0.14; 0.31] 6.9% 9.7%
RECOMBINE (435) −0.033 0.10 −0.03 [−0.24; 0.17] 1.3% 2.9%
STABILITY (2951) 0.096 0.03 0.10 [ 0.05; 0.15] 21.4% 15.3%
STANLEY (344) −0.035 0.08 −0.04 [−0.19; 0.12] 2.2% 4.4%
STANLEY (300) 0.150 0.08 0.15 [−0.00; 0.30] 2.3% 4.7%
VIS (901) 0.122 0.05 0.12 [ 0.03; 0.22] 6.3% 9.2%
Common effect model 0.13 [ 0.11; 0.16] 100.0% --
Random effects model 0.14 [ 0.10; 0.17] -- 100.0%
−0.3 −0.2 −0.1 0 0.1 0.2 0.3
Heterogeneity: $I^2$ = 51%, $\tau^2$ = 0.0017, $p$ = 0.03

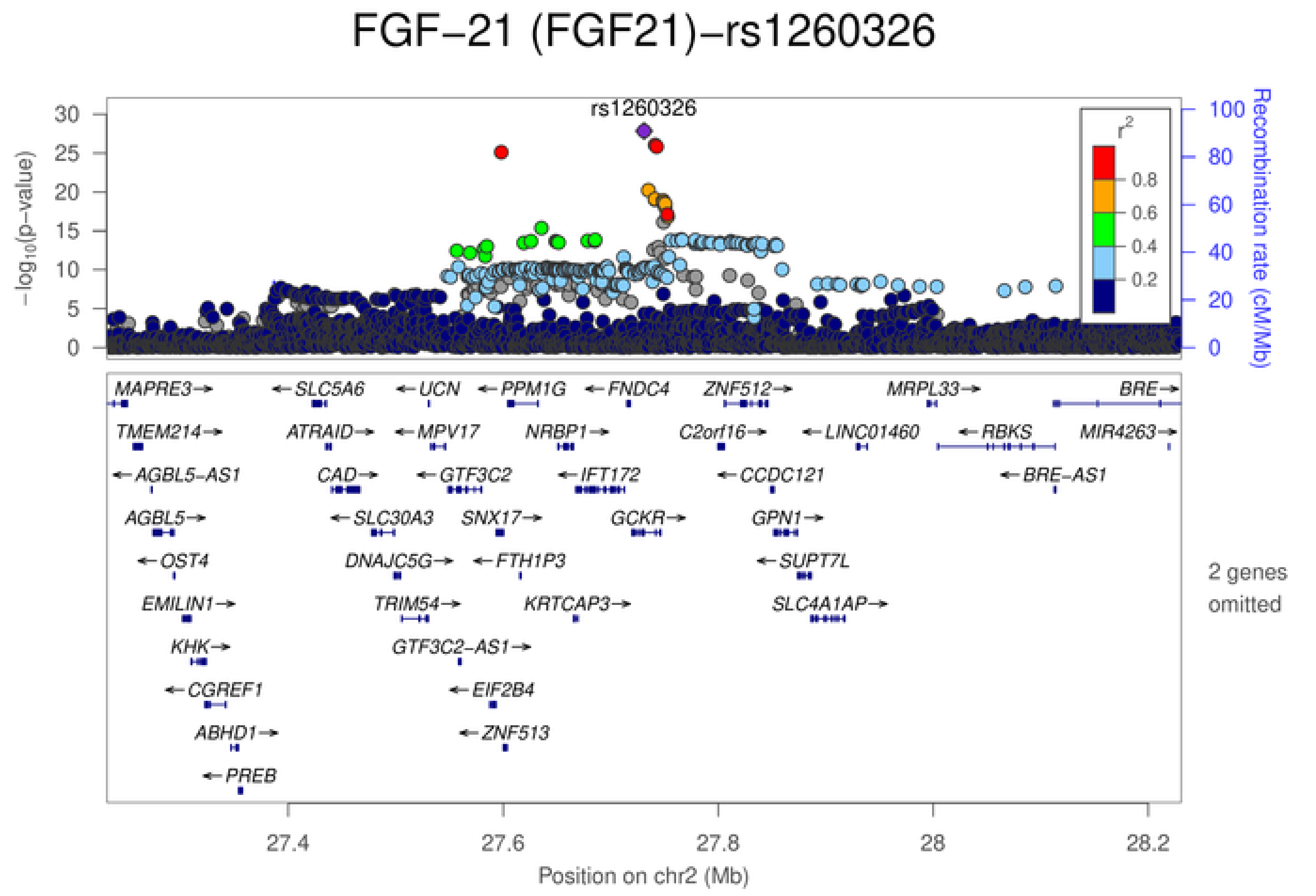
FGF−21 (FGF21)−rs1260326
rs1260326
$-\log_{10}$(p−value)
Recombination rate (cM/Mb)
$r^2$
0.8
0.6
0.4
0.2
MAPRE3→
←SLC5A6
←UCN
←PPM1G
←FNDC4
ZNF512→
MRPL33→
BRE→
TMEM214→
ATRAID→
←MPV17
NRBP1→
C2orf16→
←LINC01460
←RBKS
MIR4263→
←AGBL5−AS1
CAD→
←GTF3C2
←IFT172
←CCDC121
←BRE−AS1
AGBL5→
←SLC30A3
SNX17→
GCKR→
GPN1→
←OST4
DNAJC5G→
←FTH1P3
←SUPT7L
EMILIN1→
TRIM54→
KRTCAP3→
SLC4A1AP→
KHK→
GTF3C2−AS1→
←CGREF1
←EIF2B4
ABHD1→
←ZNF513
←PREB
2 genes omitted
27.4
27.6
27.8
28
28.2
Position on chr2 (Mb)

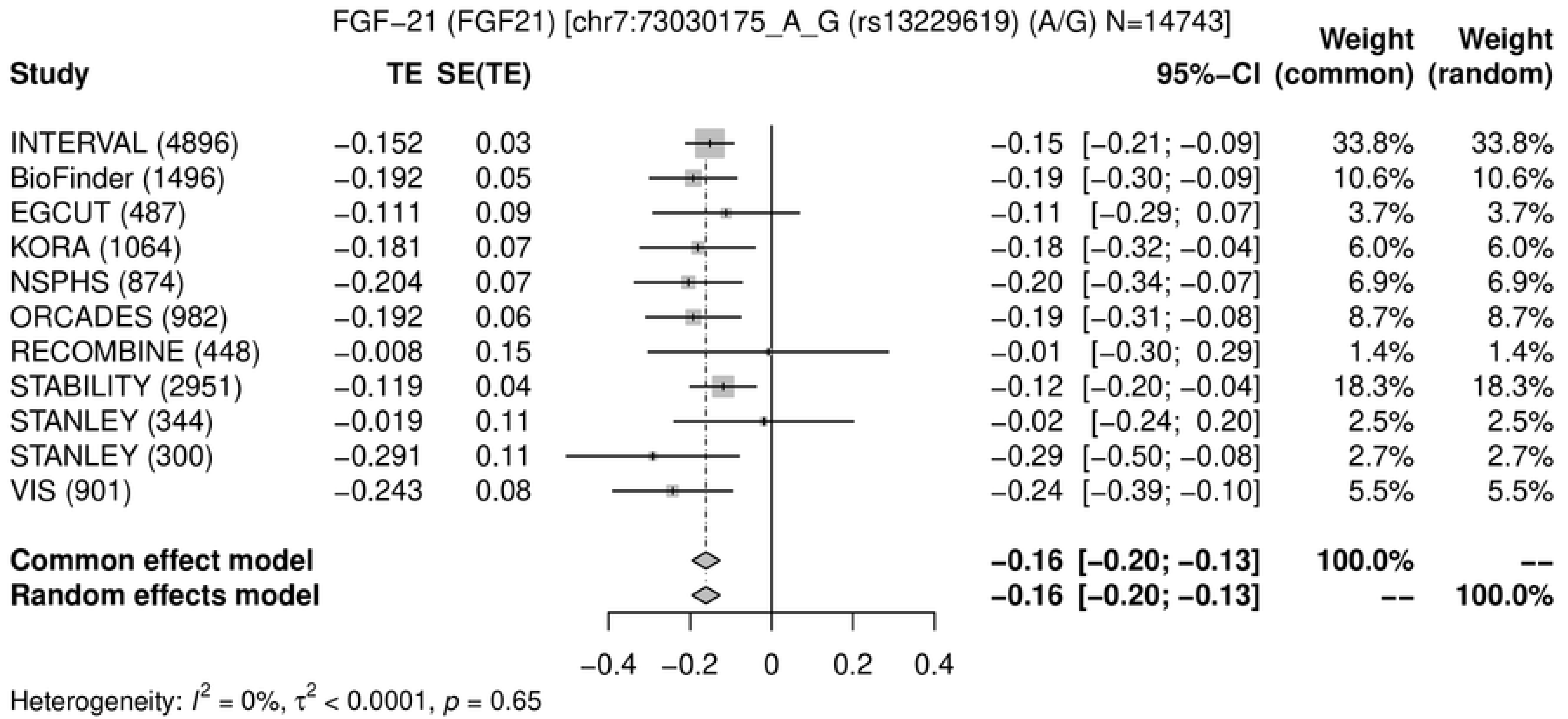
FGF-21 (FGF21) [chr7:73030175_A_G (rs13229619) (A/G) N=14743]
Study
TE SE(TE)
95%-CI
Weight (common)
Weight (random)
INTERVAL (4896) −0.152 0.03 −0.15 [−0.21; −0.09] 33.8% 33.8%
BioFinder (1496) −0.192 0.05 −0.19 [−0.30; −0.09] 10.6% 10.6%
EGCUT (487) −0.111 0.09 −0.11 [−0.29; 0.07] 3.7% 3.7%
KORA (1064) −0.181 0.07 −0.18 [−0.32; −0.04] 6.0% 6.0%
NSPHS (874) −0.204 0.07 −0.20 [−0.34; −0.07] 6.9% 6.9%
ORCADES (982) −0.192 0.06 −0.19 [−0.31; −0.08] 8.7% 8.7%
RECOMBINE (448) −0.008 0.15 −0.01 [−0.30; 0.29] 1.4% 1.4%
STABILITY (2951) −0.119 0.04 −0.12 [−0.20; −0.04] 18.3% 18.3%
STANLEY (344) −0.019 0.11 −0.02 [−0.24; 0.20] 2.5% 2.5%
STANLEY (300) −0.291 0.11 −0.29 [−0.50; −0.08] 2.7% 2.7%
VIS (901) −0.243 0.08 −0.24 [−0.39; −0.10] 5.5% 5.5%
Common effect model −0.16 [−0.20; −0.13] 100.0% --
Random effects model −0.16 [−0.20; −0.13] -- 100.0%
−0.4 −0.2 0 0.2 0.4
Heterogeneity: $I^2 = 0\%$, $\tau^2 < 0.0001$, $p = 0.65$

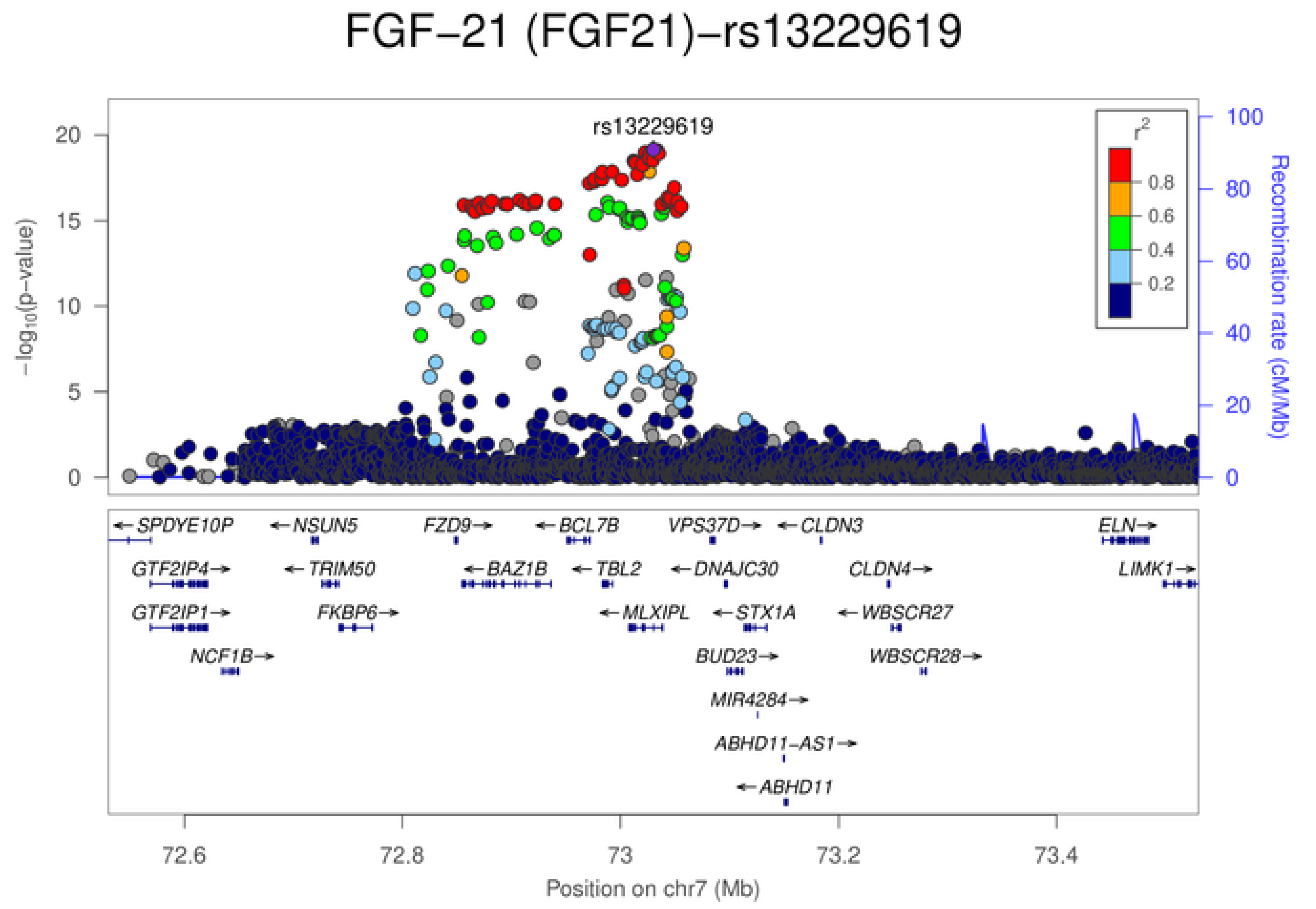
FGF−21 (FGF21)−rs13229619
rs13229619
$-\log_{10}$(p-value)
Recombination rate (cM/Mb)
$r^2$
0.8
0.6
0.4
0.2
SPDYE10P
NSUN5
FZD9
BCL7B
VPS37D
CLDN3
ELN
GTF2IP4
TRIM50
BAZ1B
TBL2
DNAJC30
CLDN4
LIMK1
GTF2IP1
FKBP6
MLXIPL
STX1A
WBSCR27
NCF1B
BUD23
WBSCR28
MIR4284
ABHD11-AS1
ABHD11
72.6
72.8
73
73.2
73.4
Position on chr7 (Mb)

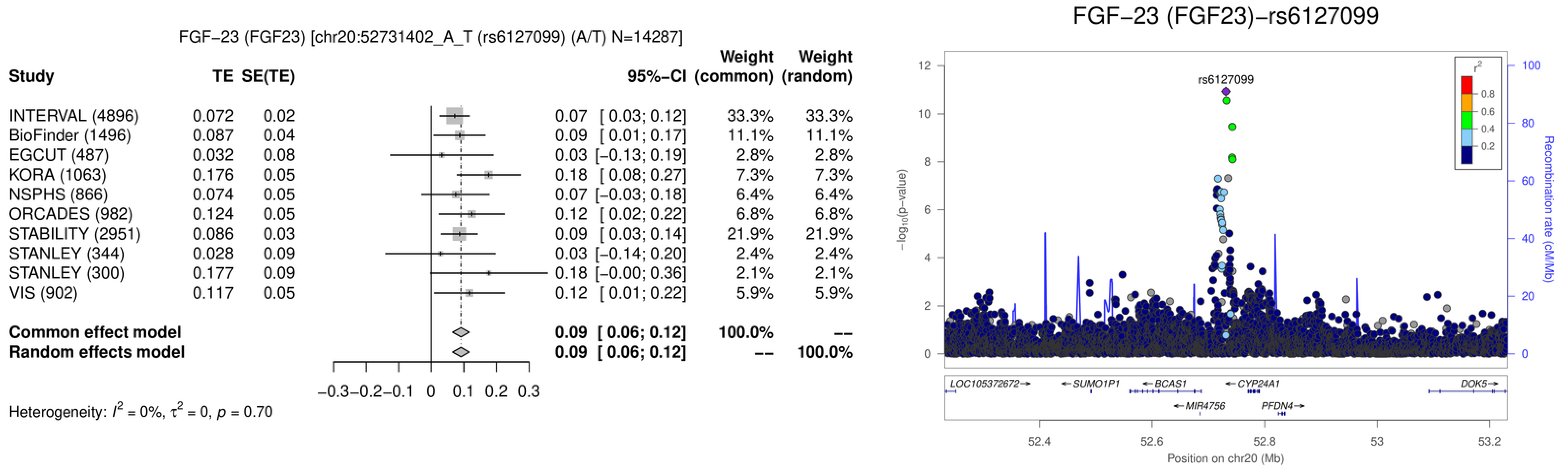
FGF−23 (FGF23) [chr20:52731402_A_T (rs6127099) (A/T) N=14287]
Study
TE
SE(TE)
95%−CI
Weight (common)
Weight (random)
INTERVAL (4896) 0.072 0.02 0.07 [ 0.03; 0.12] 33.3% 33.3%
BioFinder (1496) 0.087 0.04 0.09 [ 0.01; 0.17] 11.1% 11.1%
EGCUT (487) 0.032 0.08 0.03 [−0.13; 0.19] 2.8% 2.8%
KORA (1063) 0.176 0.05 0.18 [ 0.08; 0.27] 7.3% 7.3%
NSPHS (866) 0.074 0.05 0.07 [−0.03; 0.18] 6.4% 6.4%
ORCADES (982) 0.124 0.05 0.12 [ 0.02; 0.22] 6.8% 6.8%
STABILITY (2951) 0.086 0.03 0.09 [ 0.03; 0.14] 21.9% 21.9%
STANLEY (344) 0.028 0.09 0.03 [−0.14; 0.20] 2.4% 2.4%
STANLEY (300) 0.177 0.09 0.18 [−0.00; 0.36] 2.1% 2.1%
VIS (902) 0.117 0.05 0.12 [ 0.01; 0.22] 5.9% 5.9%
Common effect model 0.09 [ 0.06; 0.12] 100.0% --
Random effects model 0.09 [ 0.06; 0.12] -- 100.0%
−0.3 −0.2 −0.1 0 0.1 0.2 0.3
Heterogeneity: $I^2$ = 0%, $\tau^2$ = 0, $p$ = 0.70
FGF−23 (FGF23)−rs6127099
rs6127099
$-\log_{10}(p\text{-value})$
Recombination rate (cM/Mb)
$r^2$
0.8
0.6
0.4
0.2
LOC105372672
SUMO1P1
BCAS1
CYP24A1
DOK5
MIR4756
PFDN4
52.4
52.6
52.8
53
53.2
Position on chr20 (Mb)

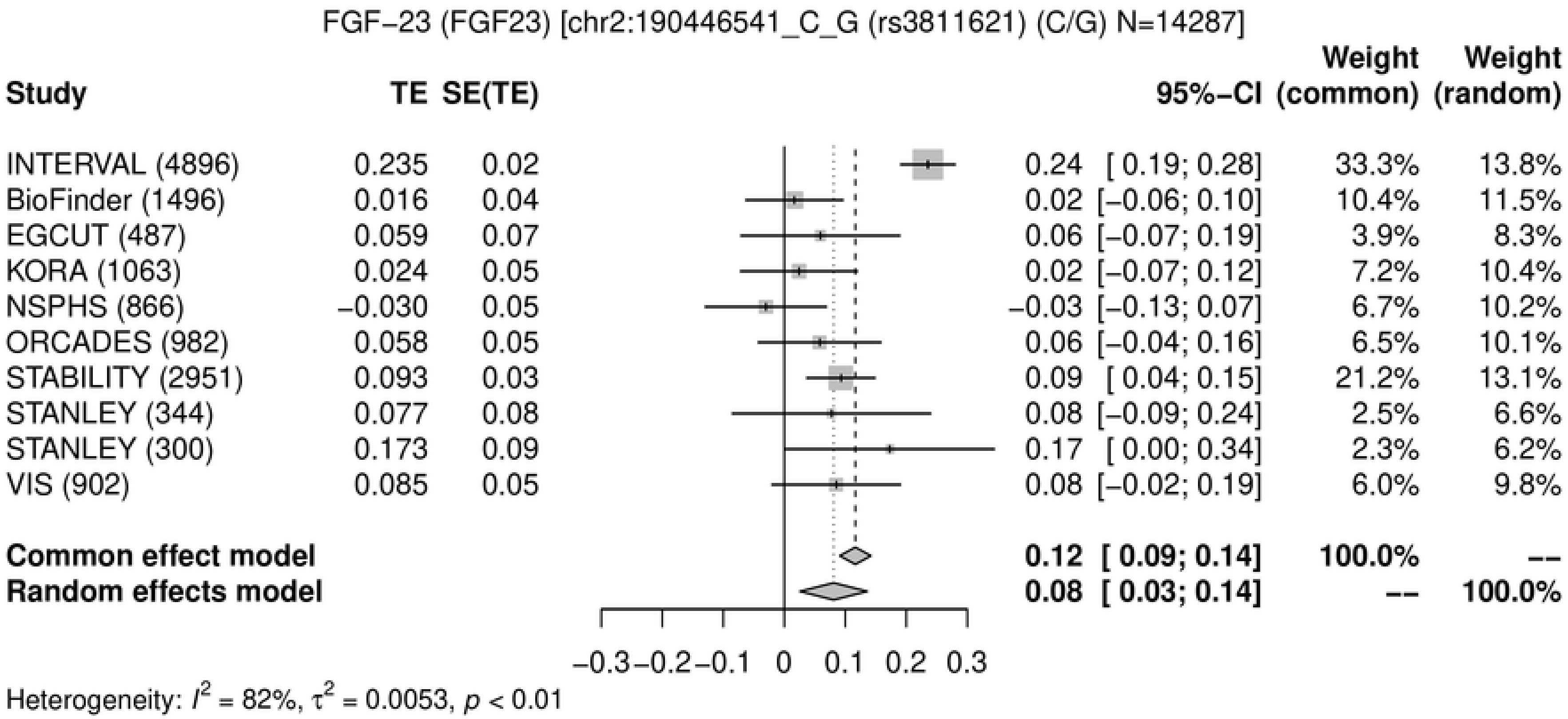
FGF−23 (FGF23) [chr2:190446541_C_G (rs3811621) (C/G) N=14287]
Study
TE
SE(TE)
95%−CI
Weight (common)
Weight (random)
INTERVAL (4896) 0.235 0.02 0.24 [ 0.19; 0.28] 33.3% 13.8%
BioFinder (1496) 0.016 0.04 0.02 [−0.06; 0.10] 10.4% 11.5%
EGCUT (487) 0.059 0.07 0.06 [−0.07; 0.19] 3.9% 8.3%
KORA (1063) 0.024 0.05 0.02 [−0.07; 0.12] 7.2% 10.4%
NSPHS (866) −0.030 0.05 −0.03 [−0.13; 0.07] 6.7% 10.2%
ORCADES (982) 0.058 0.05 0.06 [−0.04; 0.16] 6.5% 10.1%
STABILITY (2951) 0.093 0.03 0.09 [ 0.04; 0.15] 21.2% 13.1%
STANLEY (344) 0.077 0.08 0.08 [−0.09; 0.24] 2.5% 6.6%
STANLEY (300) 0.173 0.09 0.17 [ 0.00; 0.34] 2.3% 6.2%
VIS (902) 0.085 0.05 0.08 [−0.02; 0.19] 6.0% 9.8%
Common effect model 0.12 [ 0.09; 0.14] 100.0% --
Random effects model 0.08 [ 0.03; 0.14] -- 100.0%
−0.3 −0.2 −0.1 0 0.1 0.2 0.3
Heterogeneity: $I^2$ = 82%, $\tau^2$ = 0.0053, $p$ < 0.01

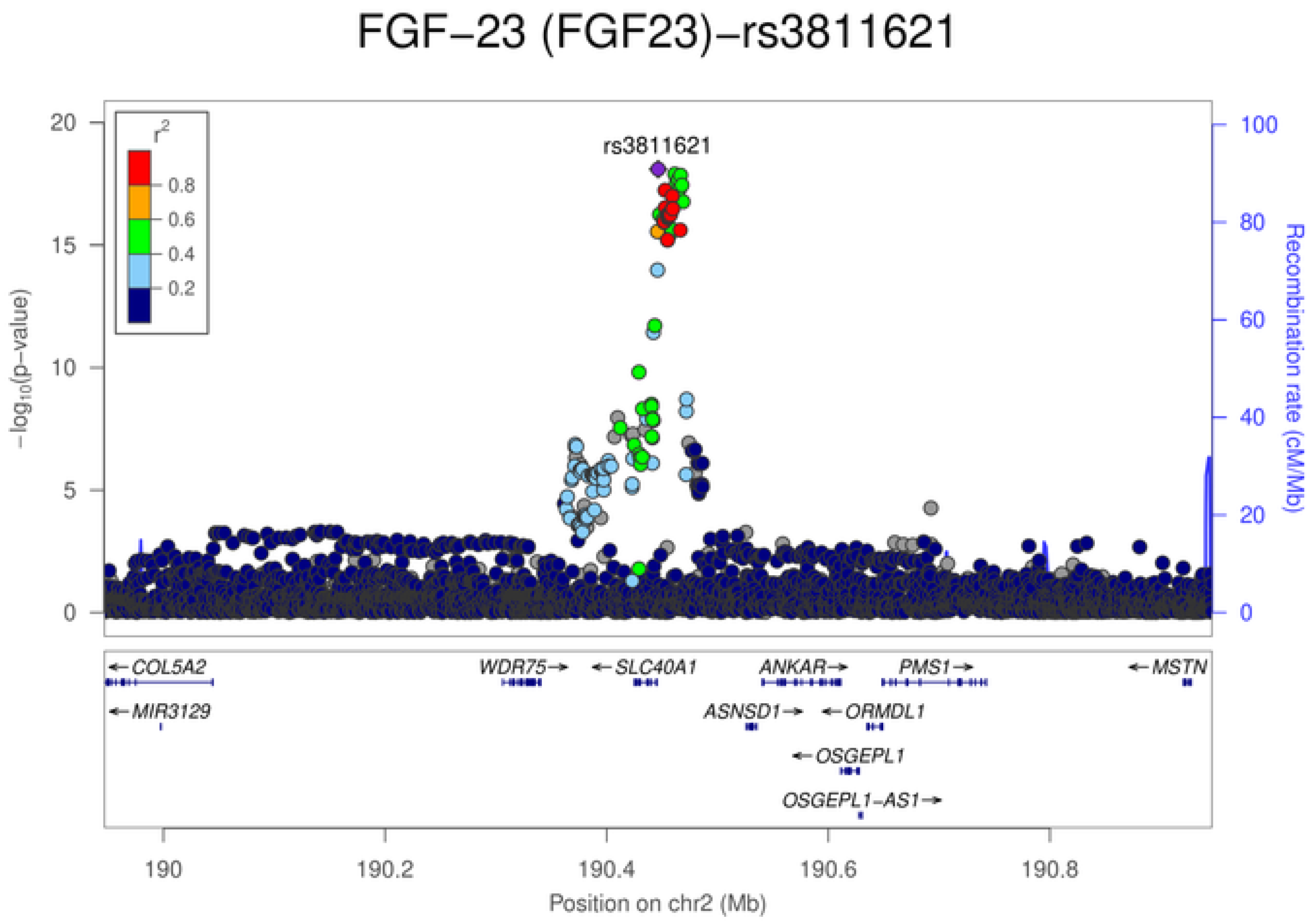
FGF−23 (FGF23)−rs3811621
rs3811621
$r^2$
0.8
0.6
0.4
0.2
20
15
10
5
0
$-\log_{10}$(p−value)
100
80
60
40
20
0
Recombination rate (cM/Mb)
COL5A2
WDR75
SLC40A1
ANKAR
PMS1
MSTN
MIR3129
ASNSD1
ORMDL1
OSGEPL1
OSGEPL1−AS1
190
190.2
190.4
190.6
190.8
Position on chr2 (Mb)

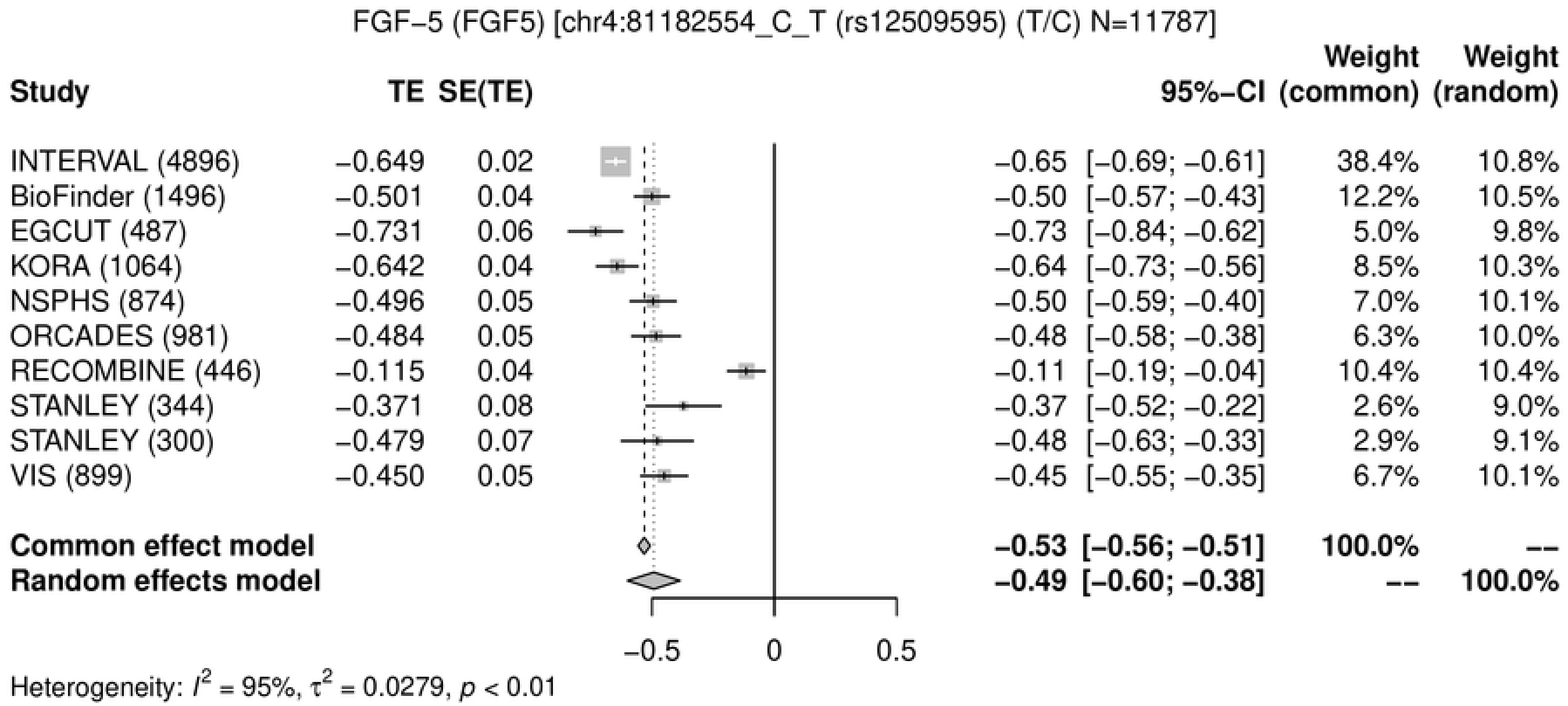
FGF−5 (FGF5) [chr4:81182554_C_T (rs12509595) (T/C) N=11787]
Study
TE
SE(TE)
95%−CI
Weight (common)
Weight (random)
INTERVAL (4896) −0.649 0.02 −0.65 [−0.69; −0.61] 38.4% 10.8%
BioFinder (1496) −0.501 0.04 −0.50 [−0.57; −0.43] 12.2% 10.5%
EGCUT (487) −0.731 0.06 −0.73 [−0.84; −0.62] 5.0% 9.8%
KORA (1064) −0.642 0.04 −0.64 [−0.73; −0.56] 8.5% 10.3%
NSPHS (874) −0.496 0.05 −0.50 [−0.59; −0.40] 7.0% 10.1%
ORCADES (981) −0.484 0.05 −0.48 [−0.58; −0.38] 6.3% 10.0%
RECOMBINE (446) −0.115 0.04 −0.11 [−0.19; −0.04] 10.4% 10.4%
STANLEY (344) −0.371 0.08 −0.37 [−0.52; −0.22] 2.6% 9.0%
STANLEY (300) −0.479 0.07 −0.48 [−0.63; −0.33] 2.9% 9.1%
VIS (899) −0.450 0.05 −0.45 [−0.55; −0.35] 6.7% 10.1%
Common effect model −0.53 [−0.56; −0.51] 100.0% --
Random effects model −0.49 [−0.60; −0.38] -- 100.0%
−0.5
0
0.5
Heterogeneity: $I^2$ = 95%, $\tau^2$ = 0.0279, $p$ < 0.01

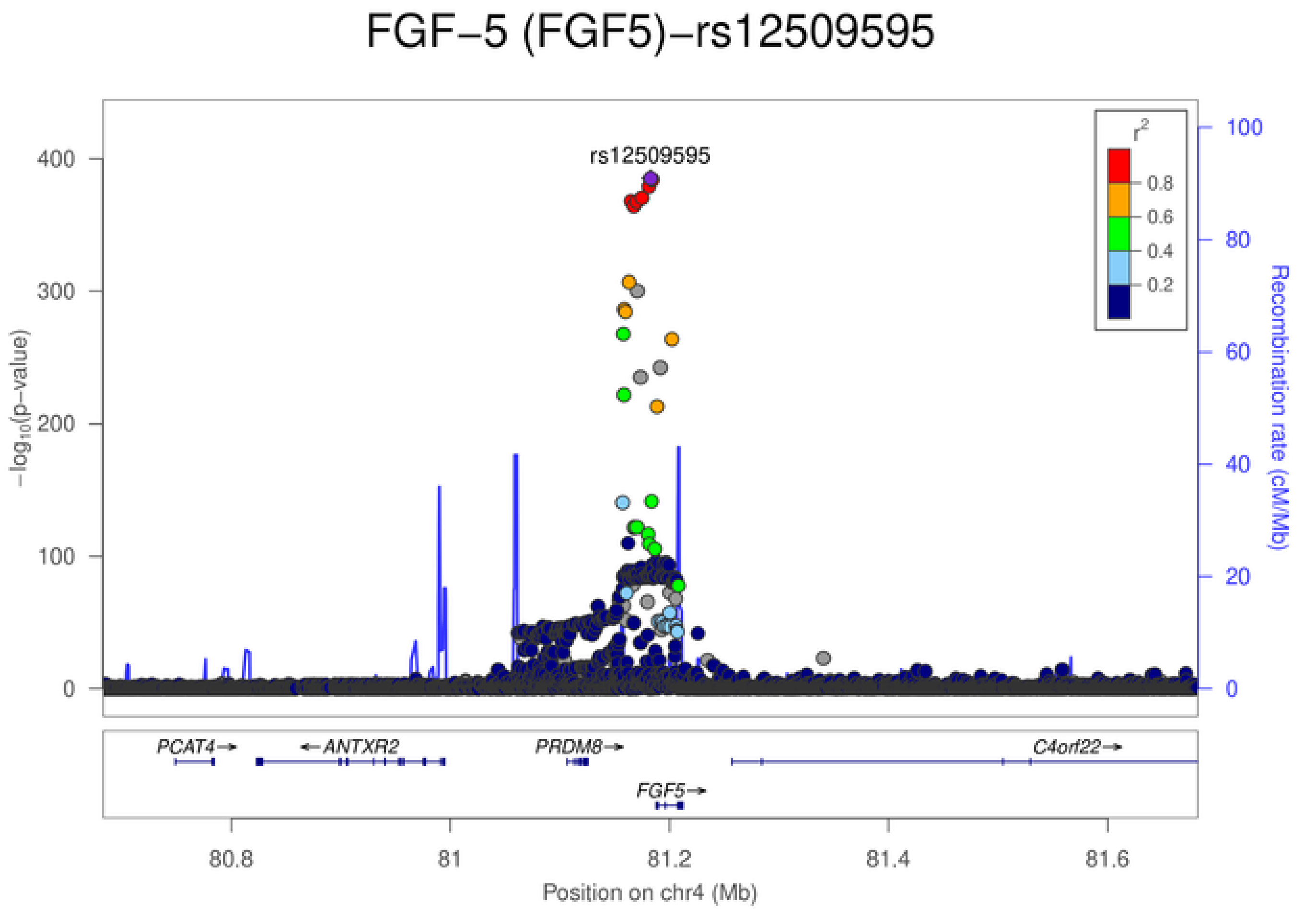
FGF−5 (FGF5)−rs12509595
rs12509595
$-\log_{10}$(p−value)
0
100
200
300
400
Recombination rate (cM/Mb)
0
20
40
60
80
100
$r^2$
0.8
0.6
0.4
0.2
PCAT4→
←ANTXR2
PRDM8→
C4orf22→
FGF5→
80.8
81
81.2
81.4
81.6
Position on chr4 (Mb)

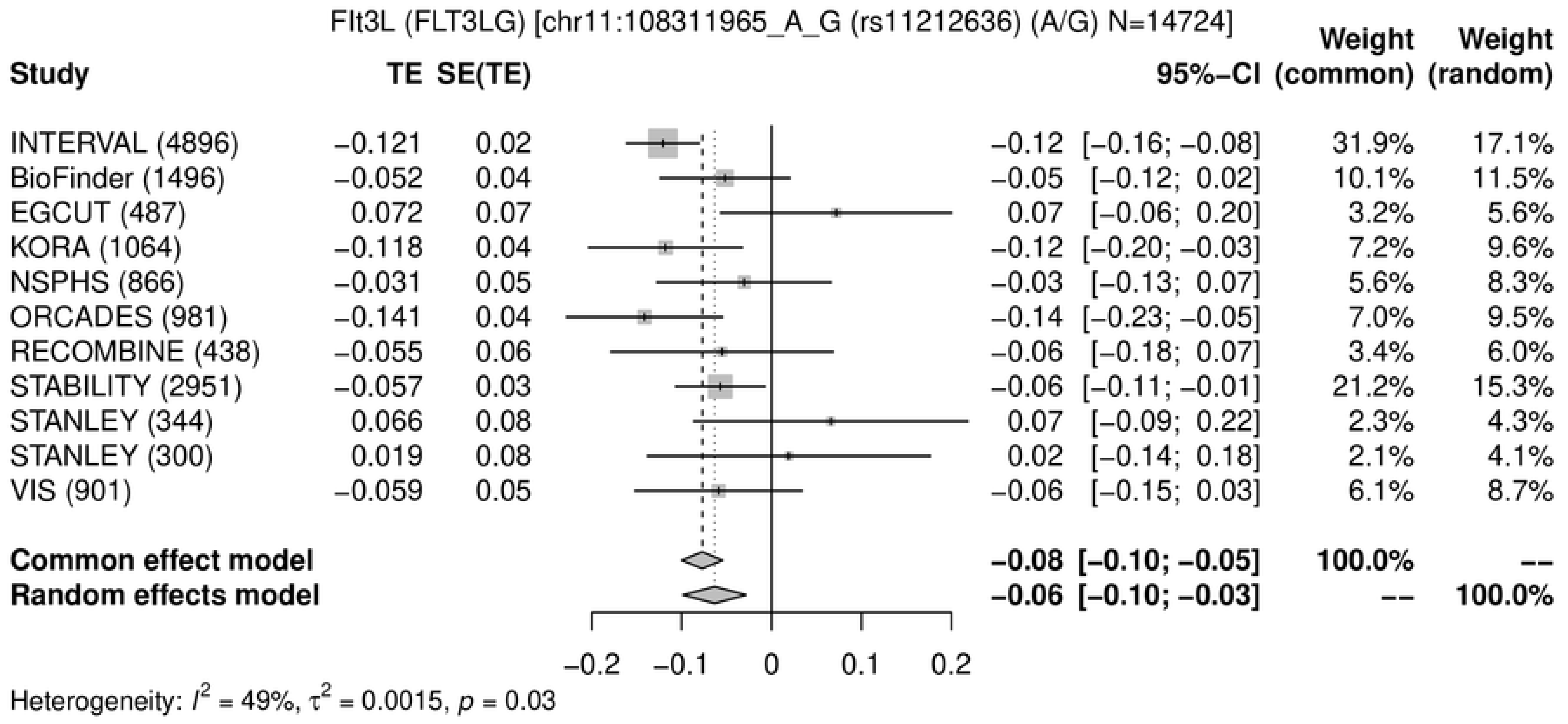
Flt3L (FLT3LG) [chr11:108311965_A_G (rs11212636) (A/G) N=14724]
Study
TE
SE(TE)
95%−CI
Weight (common)
Weight (random)
INTERVAL (4896) −0.121 0.02 −0.12 [−0.16; −0.08] 31.9% 17.1%
BioFinder (1496) −0.052 0.04 −0.05 [−0.12; 0.02] 10.1% 11.5%
EGCUT (487) 0.072 0.07 0.07 [−0.06; 0.20] 3.2% 5.6%
KORA (1064) −0.118 0.04 −0.12 [−0.20; −0.03] 7.2% 9.6%
NSPHS (866) −0.031 0.05 −0.03 [−0.13; 0.07] 5.6% 8.3%
ORCADES (981) −0.141 0.04 −0.14 [−0.23; −0.05] 7.0% 9.5%
RECOMBINE (438) −0.055 0.06 −0.06 [−0.18; 0.07] 3.4% 6.0%
STABILITY (2951) −0.057 0.03 −0.06 [−0.11; −0.01] 21.2% 15.3%
STANLEY (344) 0.066 0.08 0.07 [−0.09; 0.22] 2.3% 4.3%
STANLEY (300) 0.019 0.08 0.02 [−0.14; 0.18] 2.1% 4.1%
VIS (901) −0.059 0.05 −0.06 [−0.15; 0.03] 6.1% 8.7%
Common effect model −0.08 [−0.10; −0.05] 100.0% --
Random effects model −0.06 [−0.10; −0.03] -- 100.0%
−0.2 −0.1 0 0.1 0.2
Heterogeneity: $I^2$ = 49%, $\tau^2$ = 0.0015, $p$ = 0.03

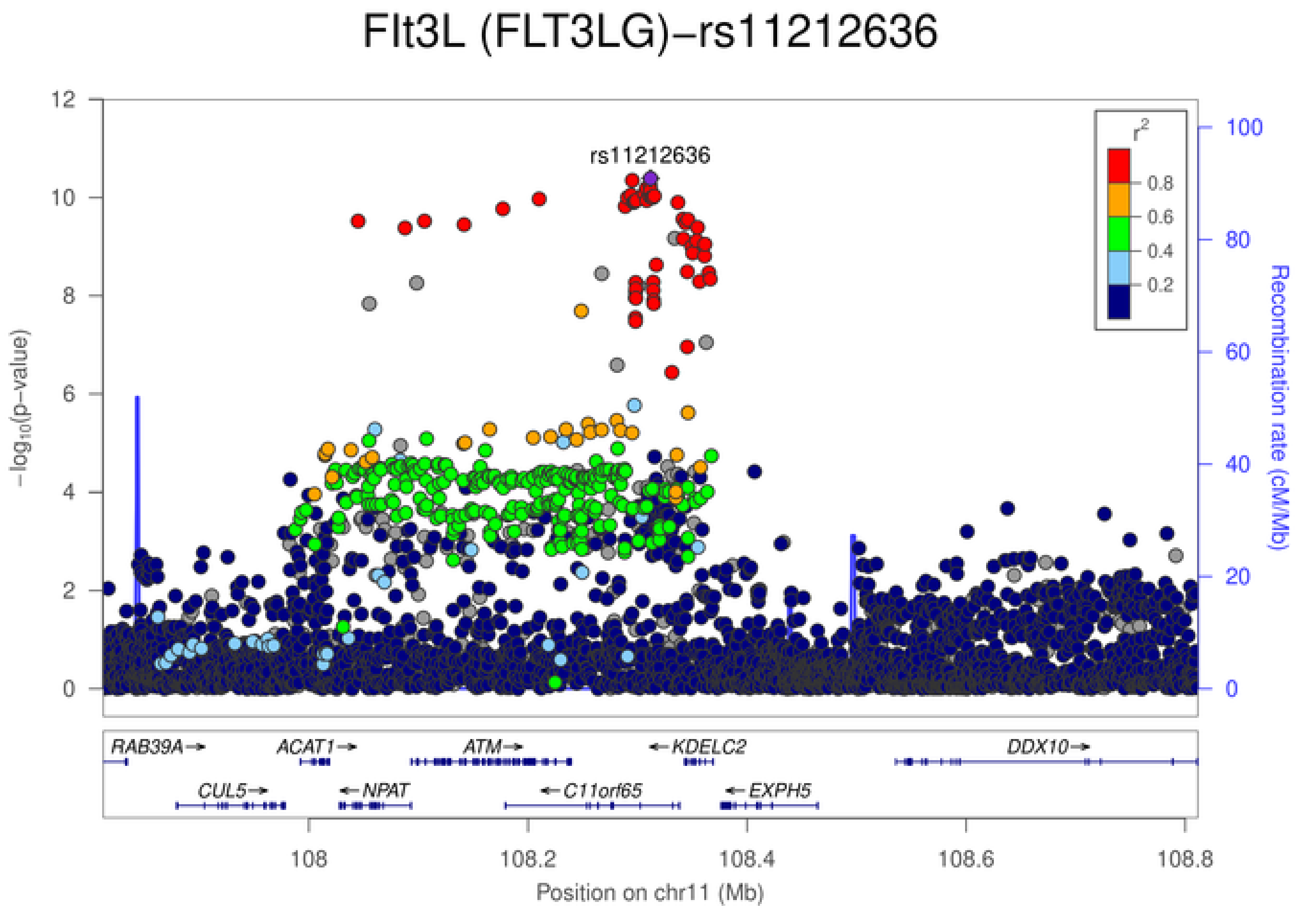
Flt3L (FLT3LG)−rs11212636
rs11212636
$-\log_{10}$(p−value)
Recombination rate (cM/Mb)
$r^2$
0.8
0.6
0.4
0.2
RAB39A→
ACAT1→
ATM→
←KDELC2
DDX10→
CUL5→
←NPAT
←C11orf65
←EXPH5
108
108.2
108.4
108.6
108.8
Position on chr11 (Mb)

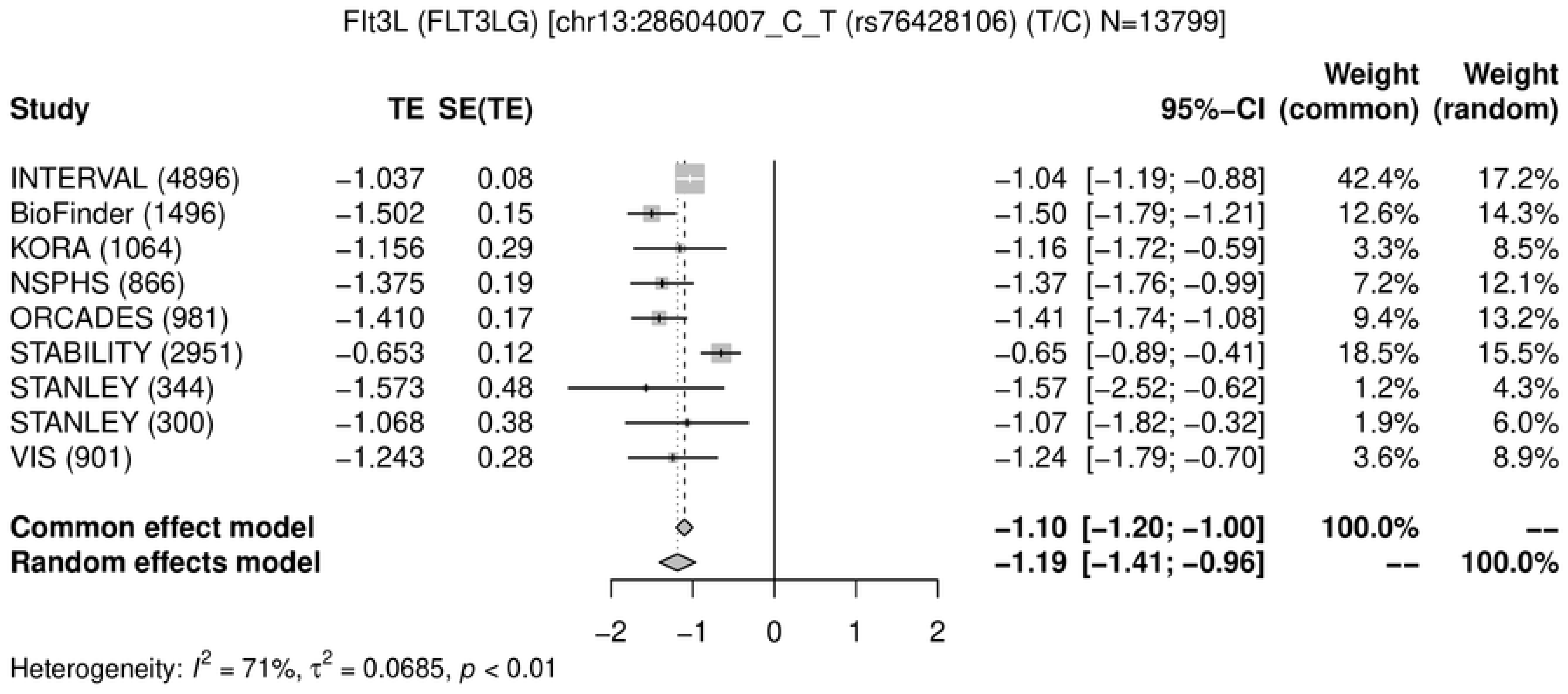

Flt3L (FLT3LG) [chr13:28604007_C_T (rs76428106) (T/C) N=13799]
Study TE SE(TE) 95%-CI Weight (common) Weight (random)
INTERVAL (4896) −1.037 0.08 −1.04 [−1.19; −0.88] 42.4% 17.2%
BioFinder (1496) −1.502 0.15 −1.50 [−1.79; −1.21] 12.6% 14.3%
KORA (1064) −1.156 0.29 −1.16 [−1.72; −0.59] 3.3% 8.5%
NSPHS (866) −1.375 0.19 −1.37 [−1.76; −0.99] 7.2% 12.1%
ORCADES (981) −1.410 0.17 −1.41 [−1.74; −1.08] 9.4% 13.2%
STABILITY (2951) −0.653 0.12 −0.65 [−0.89; −0.41] 18.5% 15.5%
STANLEY (344) −1.573 0.48 −1.57 [−2.52; −0.62] 1.2% 4.3%
STANLEY (300) −1.068 0.38 −1.07 [−1.82; −0.32] 1.9% 6.0%
VIS (901) −1.243 0.28 −1.24 [−1.79; −0.70] 3.6% 8.9%
Common effect model −1.10 [−1.20; −1.00] 100.0% --
Random effects model −1.19 [−1.41; −0.96] -- 100.0%
−2 −1 0 1 2
Heterogeneity: $I^2$ = 71%, $\tau^2$ = 0.0685, $p$ < 0.01


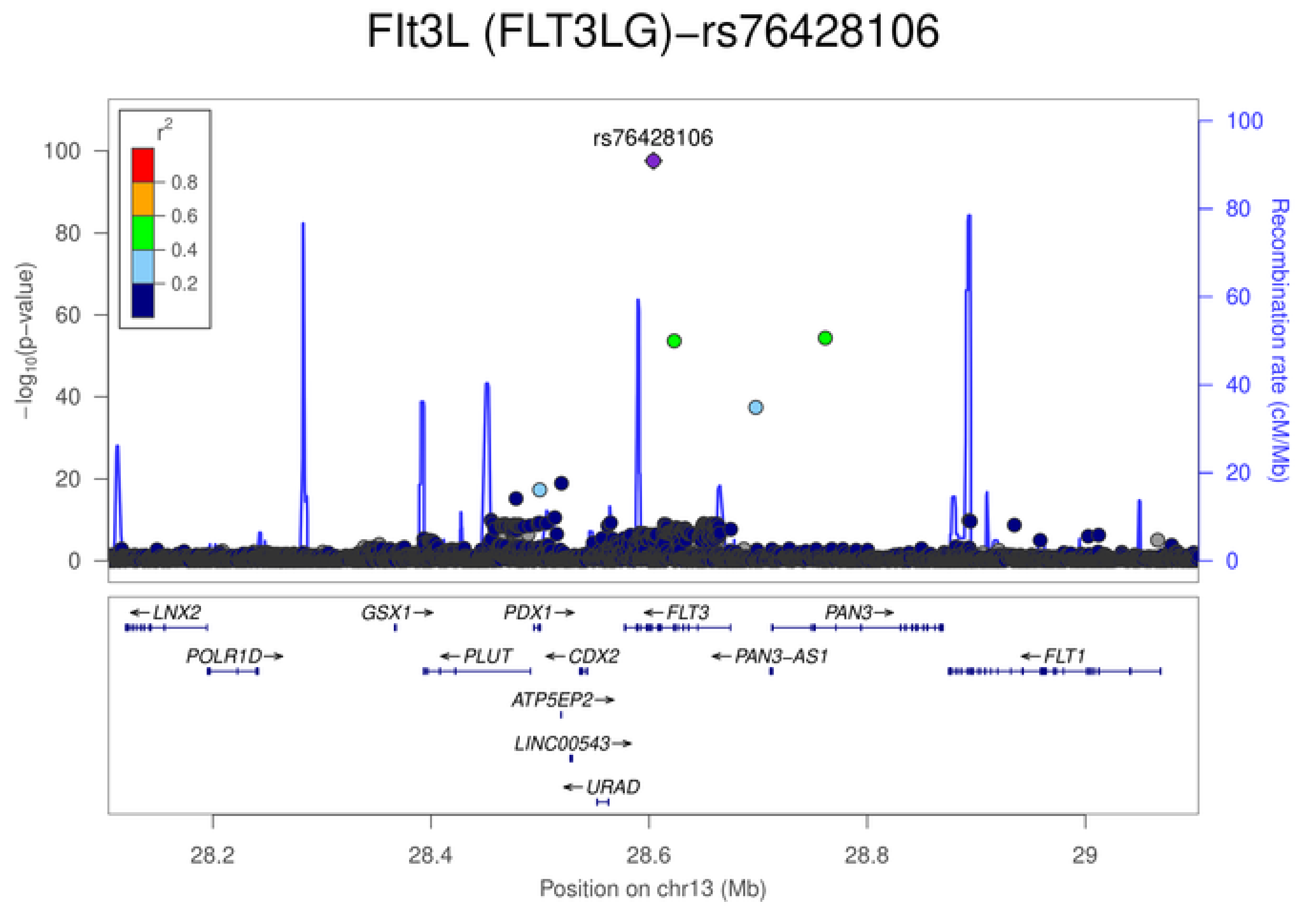

Flt3L (FLT3LG)−rs76428106
rs76428106
$r^2$
0.8
0.6
0.4
0.2
$-\log_{10}$(p−value)
Recombination rate (cM/Mb)
0 20 40 60 80 100
LNX2
GSX1
PDX1
FLT3
PAN3
POLR1D
PLUT
CDX2
PAN3-AS1
FLT1
ATP5EP2
LINC00543
URAD
28.2 28.4 28.6 28.8 29
Position on chr13 (Mb)

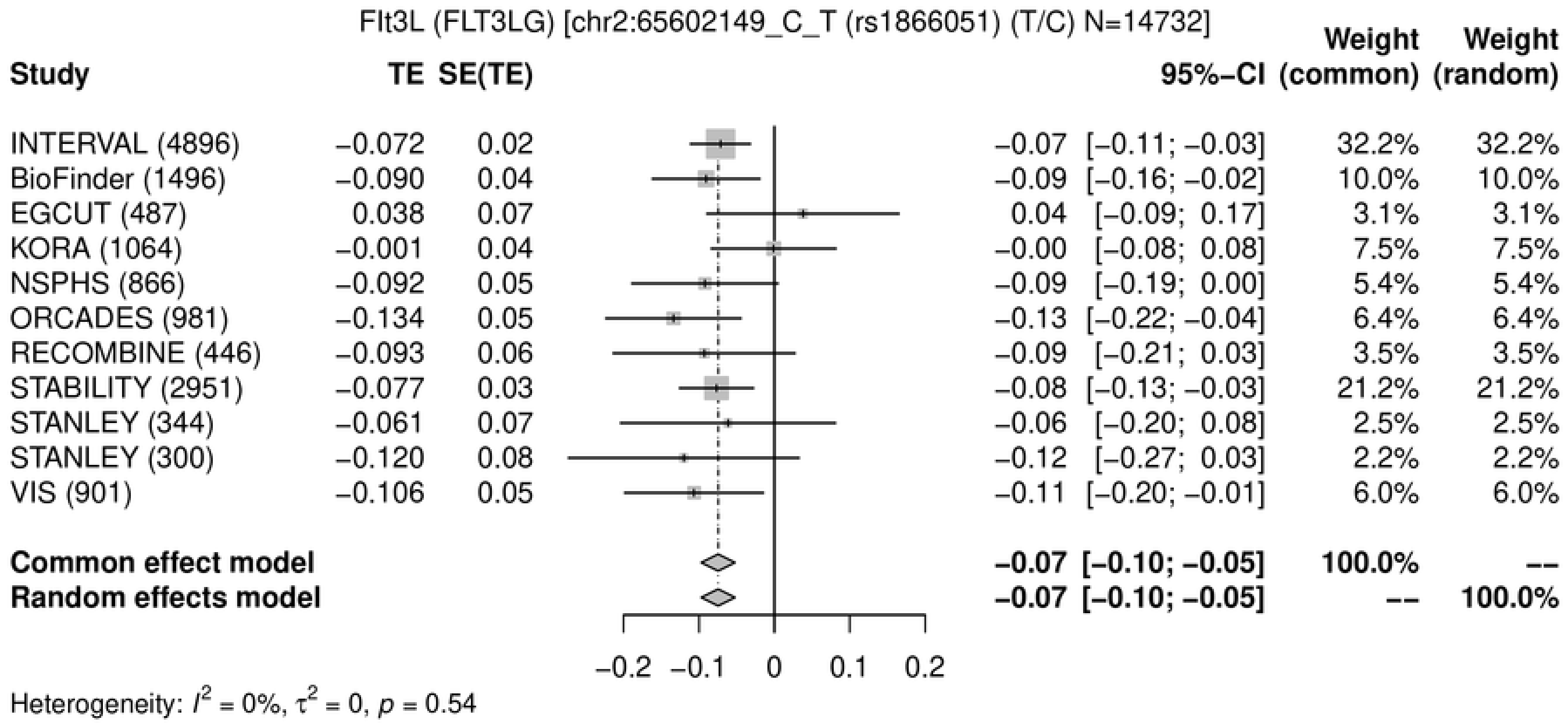
Flt3L (FLT3LG) [chr2:65602149_C_T (rs1866051) (T/C) N=14732]
Study
TE
SE(TE)
95%−CI
Weight (common)
Weight (random)
INTERVAL (4896) −0.072 0.02 −0.07 [−0.11; −0.03] 32.2% 32.2%
BioFinder (1496) −0.090 0.04 −0.09 [−0.16; −0.02] 10.0% 10.0%
EGCUT (487) 0.038 0.07 0.04 [−0.09; 0.17] 3.1% 3.1%
KORA (1064) −0.001 0.04 −0.00 [−0.08; 0.08] 7.5% 7.5%
NSPHS (866) −0.092 0.05 −0.09 [−0.19; 0.00] 5.4% 5.4%
ORCADES (981) −0.134 0.05 −0.13 [−0.22; −0.04] 6.4% 6.4%
RECOMBINE (446) −0.093 0.06 −0.09 [−0.21; 0.03] 3.5% 3.5%
STABILITY (2951) −0.077 0.03 −0.08 [−0.13; −0.03] 21.2% 21.2%
STANLEY (344) −0.061 0.07 −0.06 [−0.20; 0.08] 2.5% 2.5%
STANLEY (300) −0.120 0.08 −0.12 [−0.27; 0.03] 2.2% 2.2%
VIS (901) −0.106 0.05 −0.11 [−0.20; −0.01] 6.0% 6.0%
Common effect model −0.07 [−0.10; −0.05] 100.0% --
Random effects model −0.07 [−0.10; −0.05] -- 100.0%
−0.2 −0.1 0 0.1 0.2
Heterogeneity: $I^2 = 0\%$, $\tau^2 = 0$, $p = 0.54$

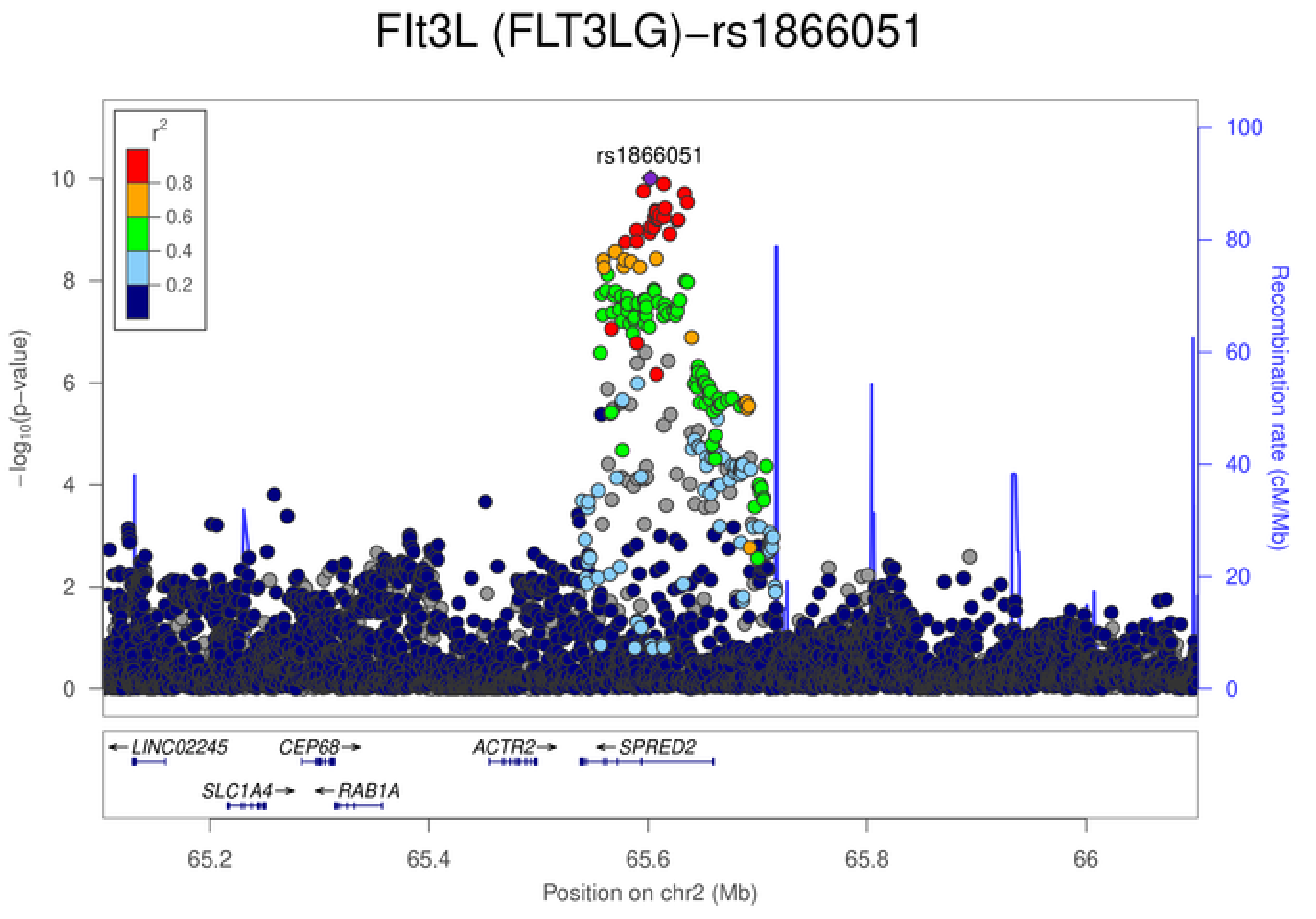
Flt3L (FLT3LG)−rs1866051
rs1866051
$r^2$
0.8
0.6
0.4
0.2
$-\log_{10}$(p−value)
Recombination rate (cM/Mb)
LINC02245
CEP68
ACTR2
SPRED2
SLC1A4
RAB1A
65.2
65.4
65.6
65.8
66
Position on chr2 (Mb)

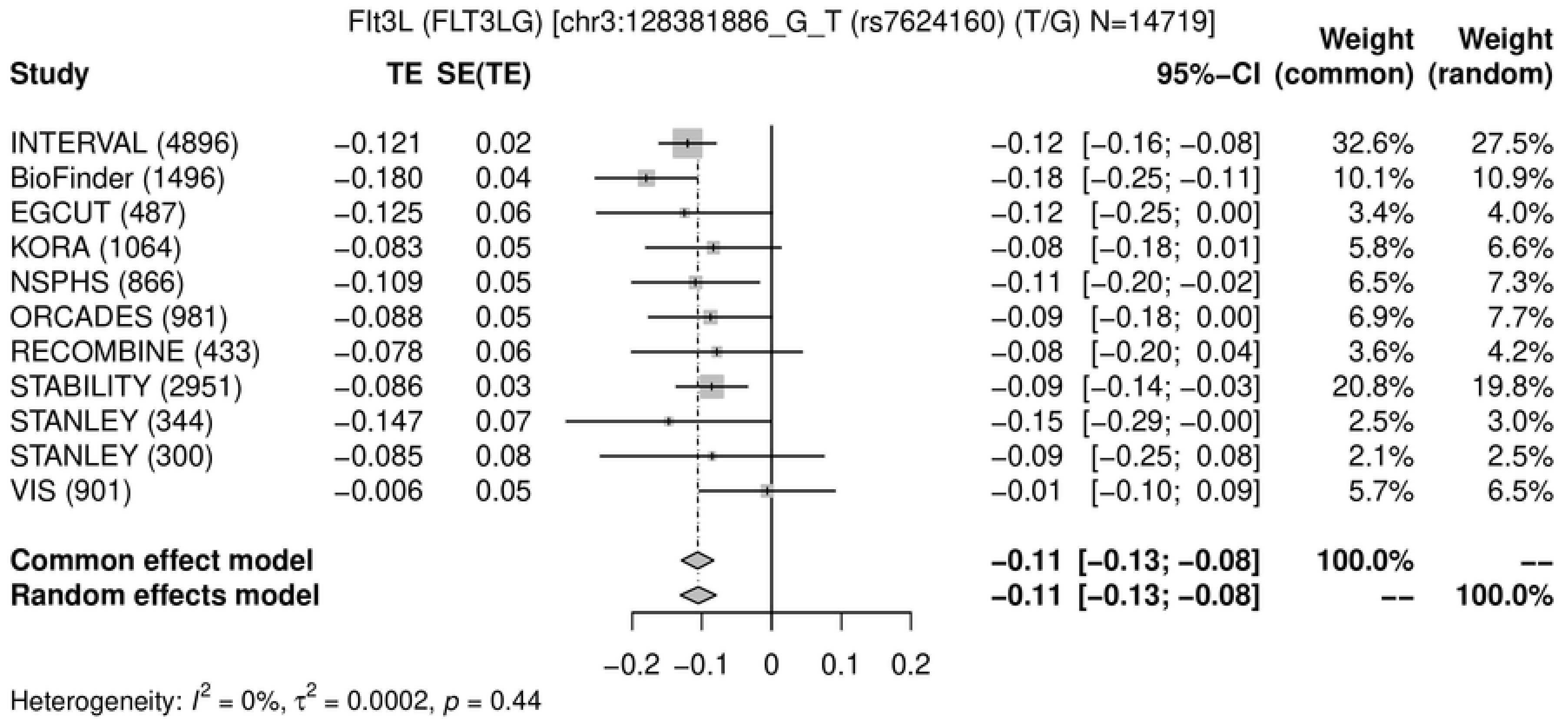
Flt3L (FLT3LG) [chr3:128381886_G_T (rs7624160) (T/G) N=14719]
Study
TE
SE(TE)
95%-CI
Weight (common)
Weight (random)
INTERVAL (4896) −0.121 0.02 −0.12 [−0.16; −0.08] 32.6% 27.5%
BioFinder (1496) −0.180 0.04 −0.18 [−0.25; −0.11] 10.1% 10.9%
EGCUT (487) −0.125 0.06 −0.12 [−0.25; 0.00] 3.4% 4.0%
KORA (1064) −0.083 0.05 −0.08 [−0.18; 0.01] 5.8% 6.6%
NSPHS (866) −0.109 0.05 −0.11 [−0.20; −0.02] 6.5% 7.3%
ORCADES (981) −0.088 0.05 −0.09 [−0.18; 0.00] 6.9% 7.7%
RECOMBINE (433) −0.078 0.06 −0.08 [−0.20; 0.04] 3.6% 4.2%
STABILITY (2951) −0.086 0.03 −0.09 [−0.14; −0.03] 20.8% 19.8%
STANLEY (344) −0.147 0.07 −0.15 [−0.29; −0.00] 2.5% 3.0%
STANLEY (300) −0.085 0.08 −0.09 [−0.25; 0.08] 2.1% 2.5%
VIS (901) −0.006 0.05 −0.01 [−0.10; 0.09] 5.7% 6.5%
Common effect model −0.11 [−0.13; −0.08] 100.0% --
Random effects model −0.11 [−0.13; −0.08] -- 100.0%
−0.2 −0.1 0 0.1 0.2
Heterogeneity: $I^2$ = 0%, $\tau^2$ = 0.0002, $p$ = 0.44
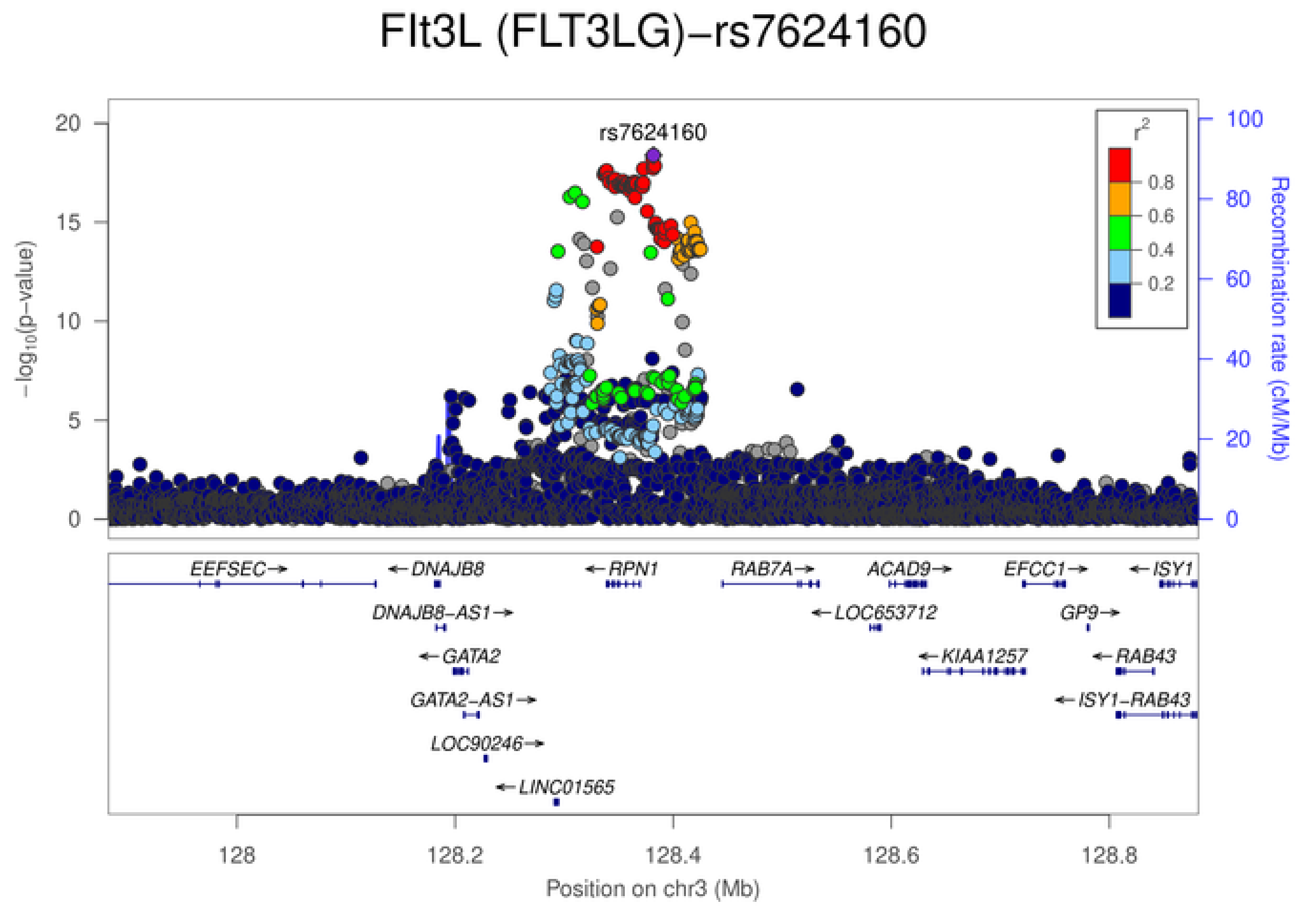
Flt3L (FLT3LG)−rs7624160
rs7624160
$-\log_{10}$(p-value)
Recombination rate (cM/Mb)
$r^2$
0.8
0.6
0.4
0.2
EEFSEC→
←DNAJB8
←RPN1
RAB7A→
ACAD9→
EFCC1→
←ISY1
DNAJB8−AS1→
←LOC653712
GP9→
←GATA2
←KIAA1257
←RAB43
GATA2−AS1→
←ISY1−RAB43
LOC90246→
←LINC01565
128 128.2 128.4 128.6 128.8
Position on chr3 (Mb)

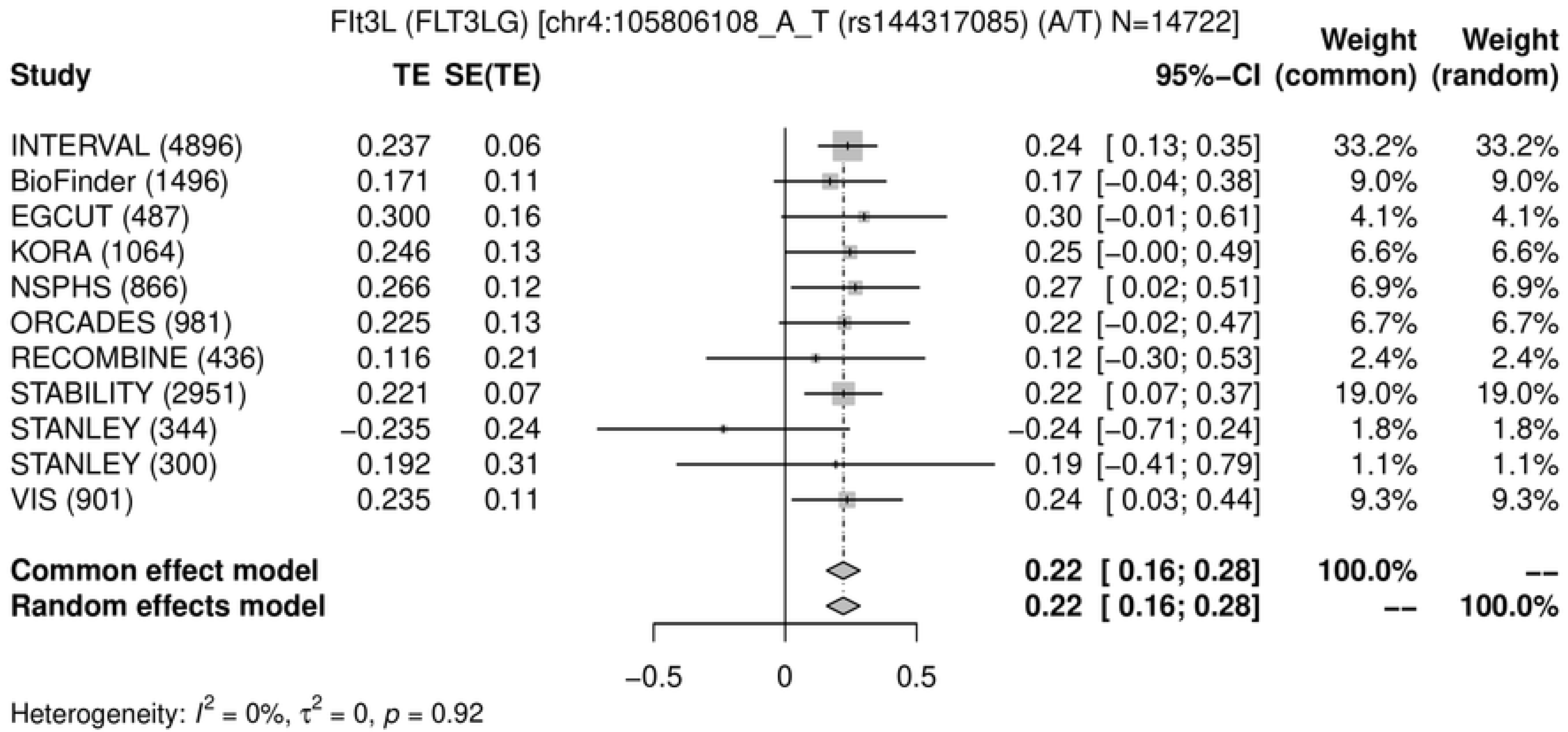
Flt3L (FLT3LG) [chr4:105806108_A_T (rs144317085) (A/T) N=14722]
Study
TE
SE(TE)
95%-CI
Weight (common)
Weight (random)
INTERVAL (4896) 0.237 0.06 0.24 [ 0.13; 0.35] 33.2% 33.2%
BioFinder (1496) 0.171 0.11 0.17 [−0.04; 0.38] 9.0% 9.0%
EGCUT (487) 0.300 0.16 0.30 [−0.01; 0.61] 4.1% 4.1%
KORA (1064) 0.246 0.13 0.25 [−0.00; 0.49] 6.6% 6.6%
NSPHS (866) 0.266 0.12 0.27 [ 0.02; 0.51] 6.9% 6.9%
ORCADES (981) 0.225 0.13 0.22 [−0.02; 0.47] 6.7% 6.7%
RECOMBINE (436) 0.116 0.21 0.12 [−0.30; 0.53] 2.4% 2.4%
STABILITY (2951) 0.221 0.07 0.22 [ 0.07; 0.37] 19.0% 19.0%
STANLEY (344) −0.235 0.24 −0.24 [−0.71; 0.24] 1.8% 1.8%
STANLEY (300) 0.192 0.31 0.19 [−0.41; 0.79] 1.1% 1.1%
VIS (901) 0.235 0.11 0.24 [ 0.03; 0.44] 9.3% 9.3%
Common effect model 0.22 [ 0.16; 0.28] 100.0% --
Random effects model 0.22 [ 0.16; 0.28] -- 100.0%
−0.5
0
0.5
Heterogeneity: $I^2$ = 0%, $\tau^2$ = 0, $p$ = 0.92

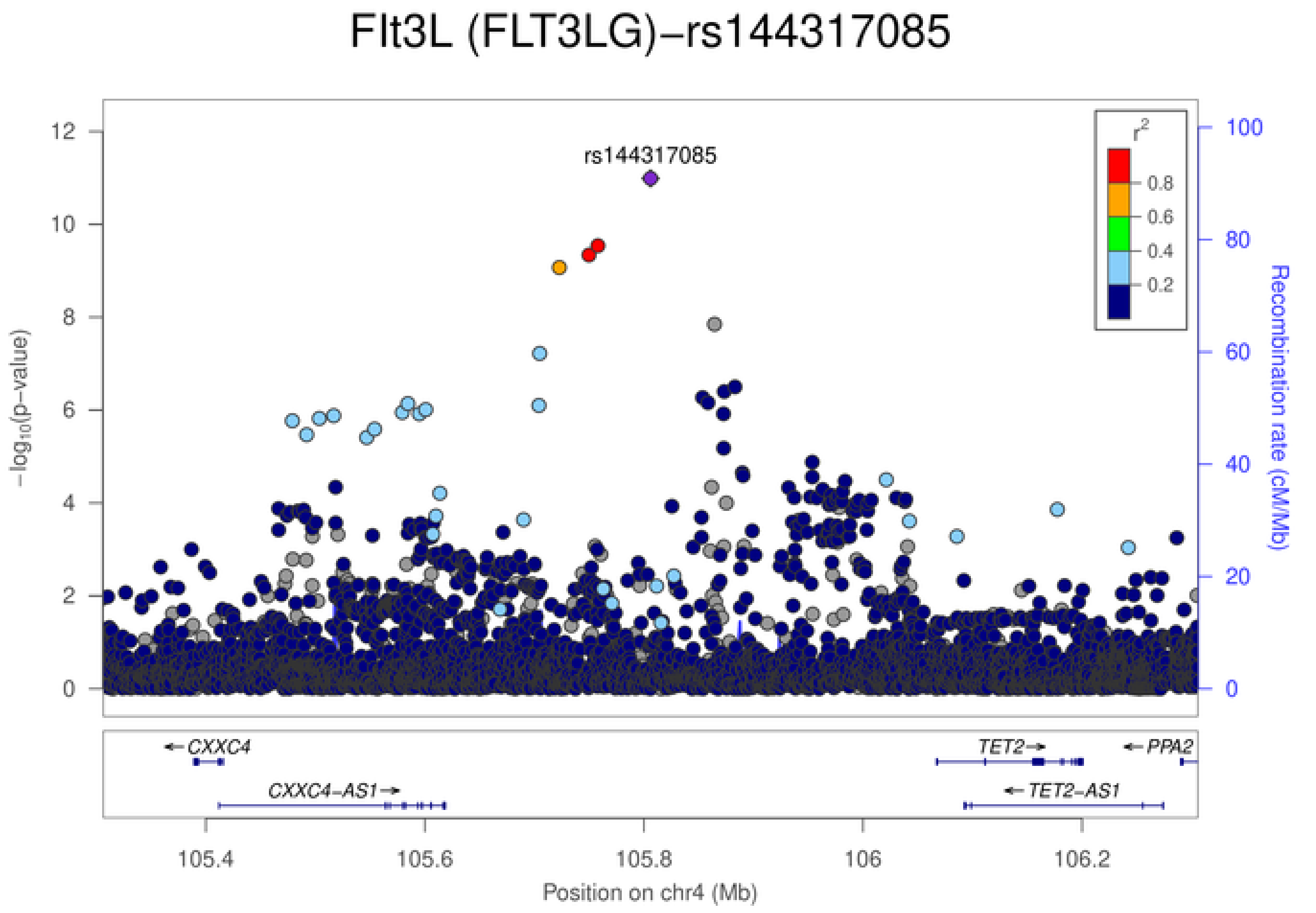
Flt3L (FLT3LG)−rs144317085
rs144317085
$-\log_{10}$(p−value)
Recombination rate (cM/Mb)
$r^2$
0.8
0.6
0.4
0.2
CXXC4
CXXC4−AS1
TET2
TET2−AS1
PPA2
105.4
105.6
105.8
106
106.2
Position on chr4 (Mb)

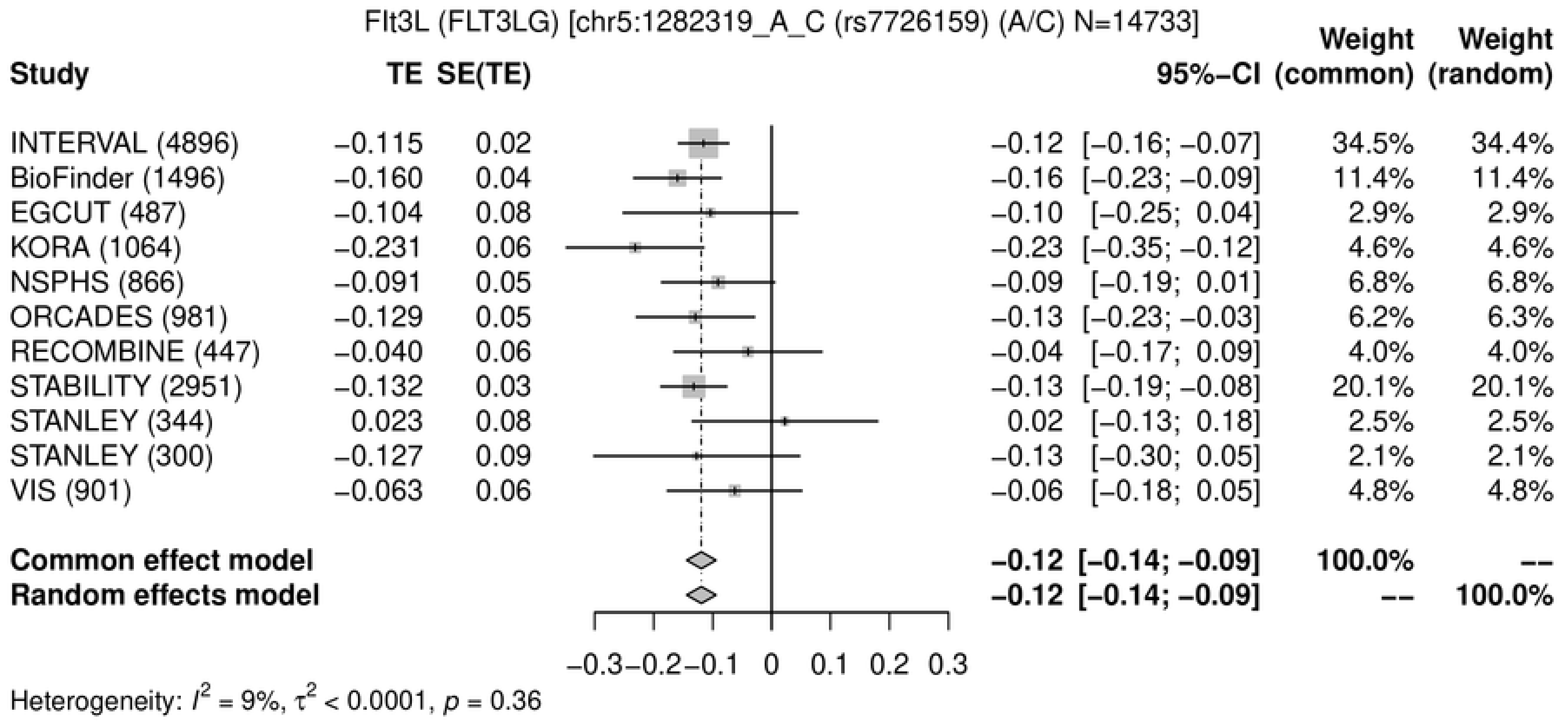
Flt3L (FLT3LG) [chr5:1282319_A_C (rs7726159) (A/C) N=14733]
Study
TE
SE(TE)
95%−CI
Weight (common)
Weight (random)
INTERVAL (4896) −0.115 0.02 −0.12 [−0.16; −0.07] 34.5% 34.4%
BioFinder (1496) −0.160 0.04 −0.16 [−0.23; −0.09] 11.4% 11.4%
EGCUT (487) −0.104 0.08 −0.10 [−0.25; 0.04] 2.9% 2.9%
KORA (1064) −0.231 0.06 −0.23 [−0.35; −0.12] 4.6% 4.6%
NSPHS (866) −0.091 0.05 −0.09 [−0.19; 0.01] 6.8% 6.8%
ORCADES (981) −0.129 0.05 −0.13 [−0.23; −0.03] 6.2% 6.3%
RECOMBINE (447) −0.040 0.06 −0.04 [−0.17; 0.09] 4.0% 4.0%
STABILITY (2951) −0.132 0.03 −0.13 [−0.19; −0.08] 20.1% 20.1%
STANLEY (344) 0.023 0.08 0.02 [−0.13; 0.18] 2.5% 2.5%
STANLEY (300) −0.127 0.09 −0.13 [−0.30; 0.05] 2.1% 2.1%
VIS (901) −0.063 0.06 −0.06 [−0.18; 0.05] 4.8% 4.8%
Common effect model −0.12 [−0.14; −0.09] 100.0% --
Random effects model −0.12 [−0.14; −0.09] -- 100.0%
−0.3 −0.2 −0.1 0 0.1 0.2 0.3
Heterogeneity: $I^2$ = 9%, $\tau^2$ < 0.0001, $p$ = 0.36

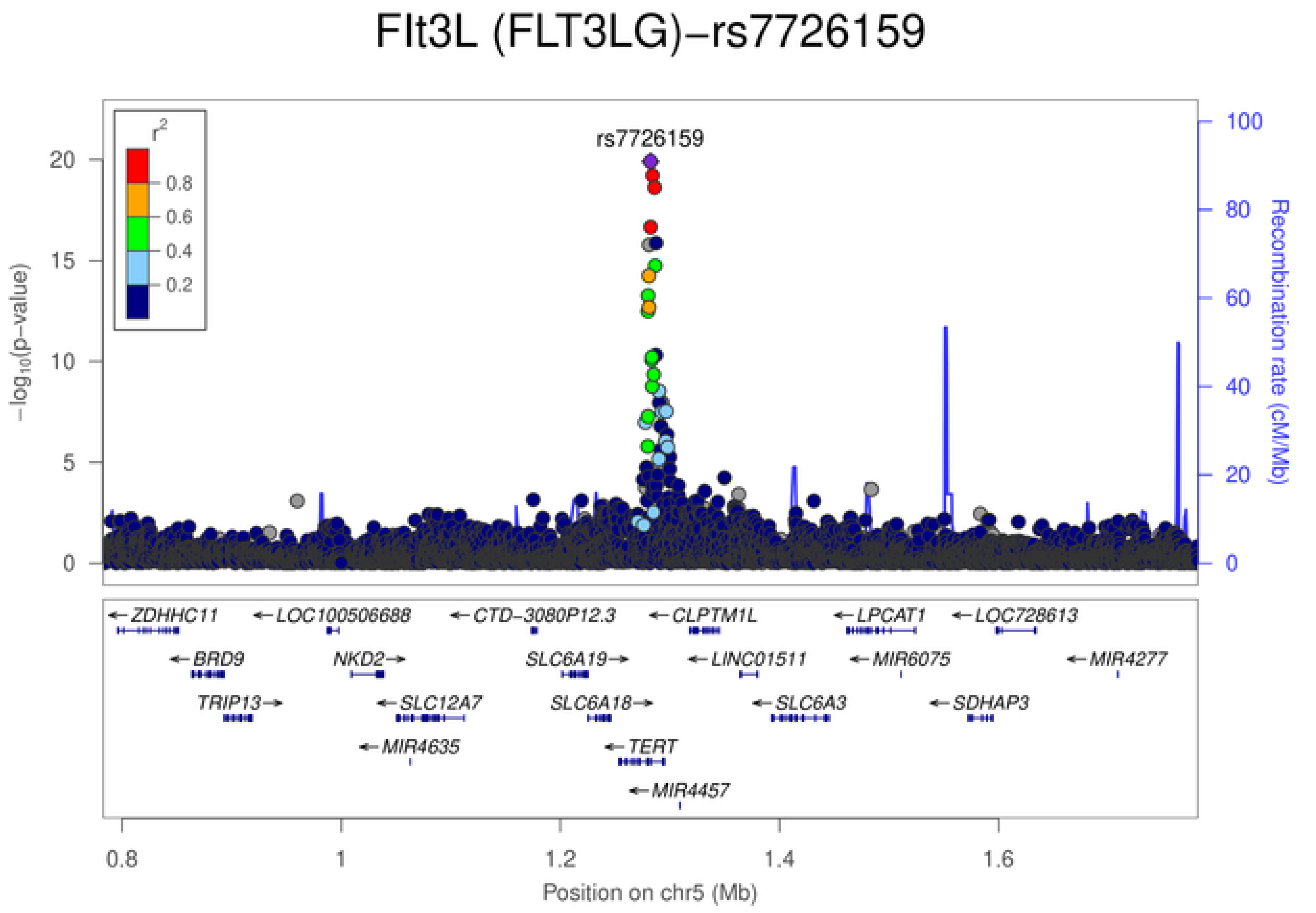
Flt3L (FLT3LG)−rs7726159
rs7726159
$r^2$
0.8
0.6
0.4
0.2
$-\log_{10}$(p-value)
Recombination rate (cM/Mb)
ZDHHC11
LOC100506688
CTD−3080P12.3
CLPTM1L
LPCAT1
LOC728613
BRD9
NKD2
SLC6A19
LINC01511
MIR6075
MIR4277
TRIP13
SLC12A7
SLC6A18
SLC6A3
SDHAP3
MIR4635
TERT
MIR4457
0.8
1
1.2
1.4
1.6
Position on chr5 (Mb)

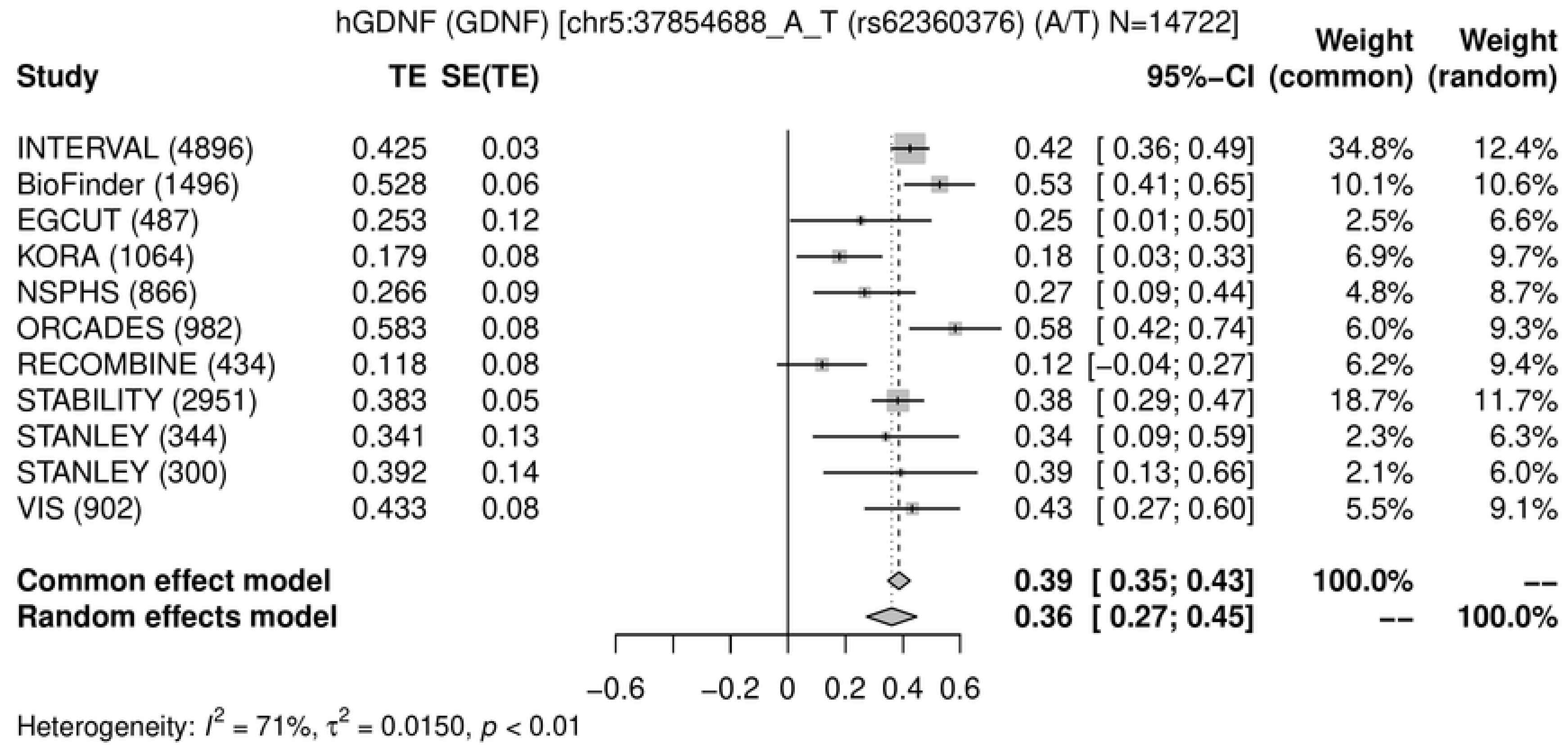
hGDNF (GDNF) [chr5:37854688_A_T (rs62360376) (A/T) N=14722]
Study TE SE(TE) 95%-CI Weight (common) Weight (random)
INTERVAL (4896) 0.425 0.03 0.42 [ 0.36; 0.49] 34.8% 12.4%
BioFinder (1496) 0.528 0.06 0.53 [ 0.41; 0.65] 10.1% 10.6%
EGCUT (487) 0.253 0.12 0.25 [ 0.01; 0.50] 2.5% 6.6%
KORA (1064) 0.179 0.08 0.18 [ 0.03; 0.33] 6.9% 9.7%
NSPHS (866) 0.266 0.09 0.27 [ 0.09; 0.44] 4.8% 8.7%
ORCADES (982) 0.583 0.08 0.58 [ 0.42; 0.74] 6.0% 9.3%
RECOMBINE (434) 0.118 0.08 0.12 [−0.04; 0.27] 6.2% 9.4%
STABILITY (2951) 0.383 0.05 0.38 [ 0.29; 0.47] 18.7% 11.7%
STANLEY (344) 0.341 0.13 0.34 [ 0.09; 0.59] 2.3% 6.3%
STANLEY (300) 0.392 0.14 0.39 [ 0.13; 0.66] 2.1% 6.0%
VIS (902) 0.433 0.08 0.43 [ 0.27; 0.60] 5.5% 9.1%
Common effect model 0.39 [ 0.35; 0.43] 100.0% --
Random effects model 0.36 [ 0.27; 0.45] -- 100.0%
−0.6 −0.2 0 0.2 0.4 0.6
Heterogeneity: $I^2$ = 71%, $\tau^2$ = 0.0150, $p$ < 0.01

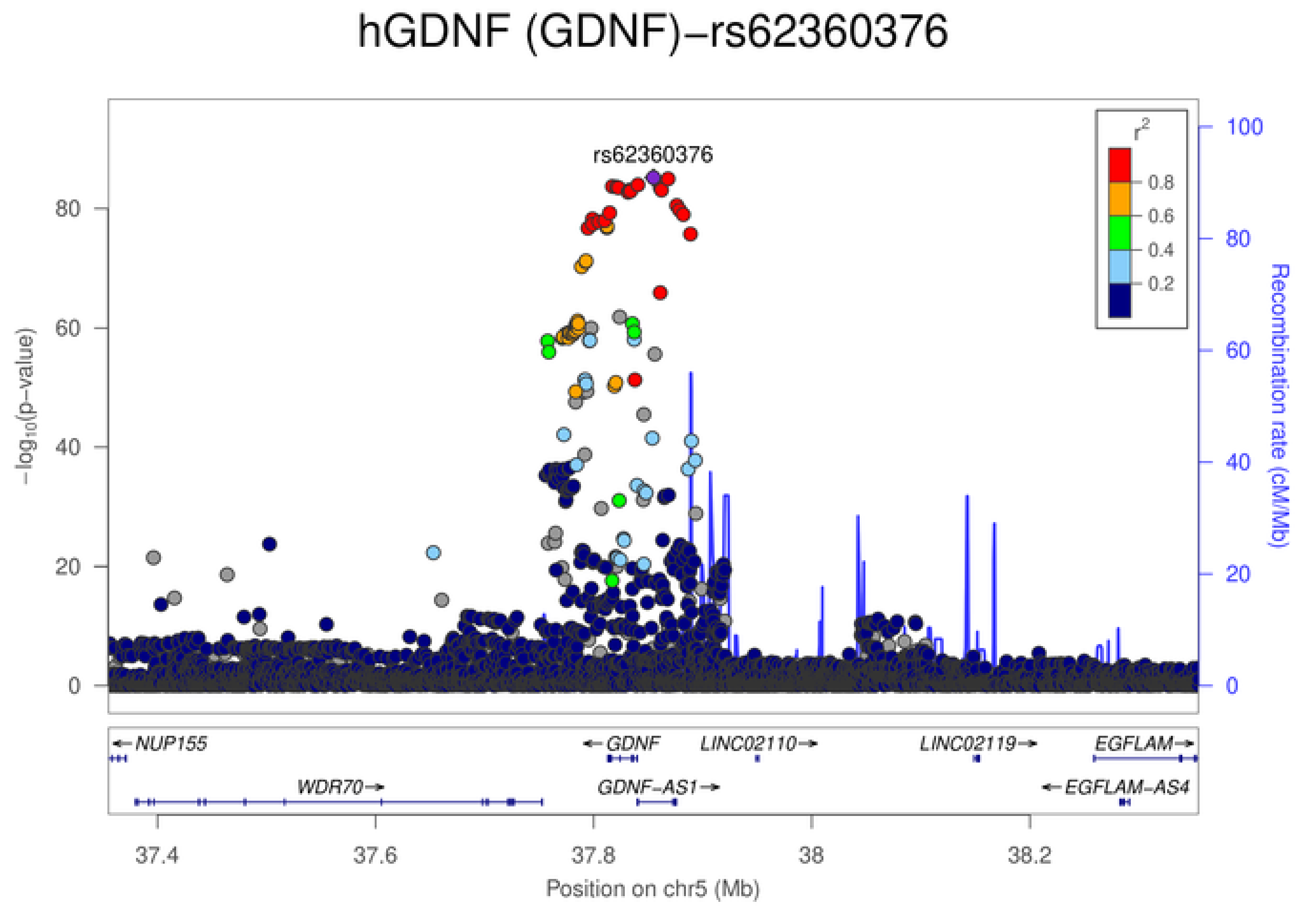
hGDNF (GDNF)−rs62360376
rs62360376
−log10(p−value)
Recombination rate (cM/Mb)
r²
0.8
0.6
0.4
0.2
0
20
40
60
80
100
NUP155
GDNF
LINC02110
LINC02119
EGFLAM
WDR70
GDNF-AS1
EGFLAM-AS4
37.4
37.6
37.8
38
38.2
Position on chr5 (Mb)

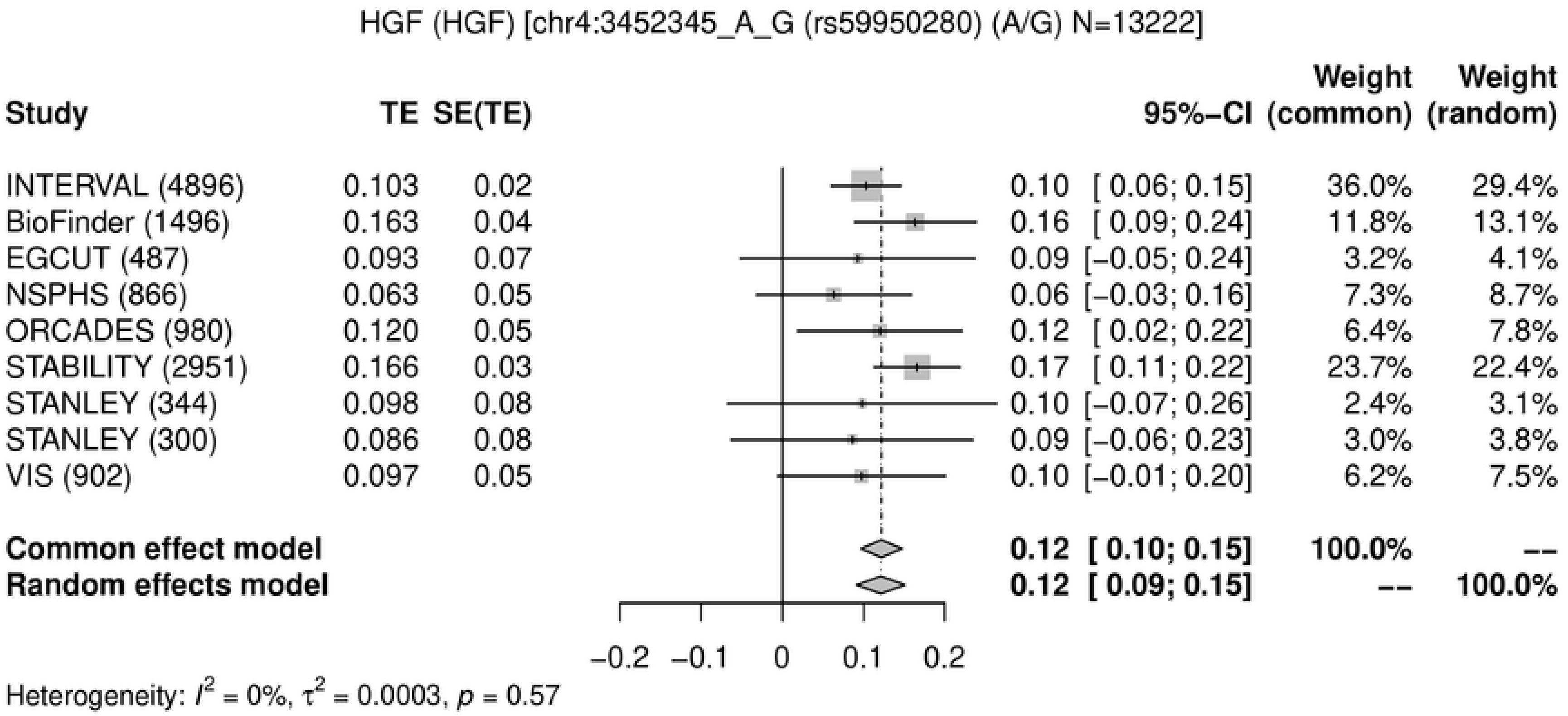
HGF (HGF) [chr4:3452345_A_G (rs59950280) (A/G) N=13222]
Study
TE
SE(TE)
95%-CI
Weight (common)
Weight (random)
INTERVAL (4896) 0.103 0.02 0.10 [ 0.06; 0.15] 36.0% 29.4%
BioFinder (1496) 0.163 0.04 0.16 [ 0.09; 0.24] 11.8% 13.1%
EGCUT (487) 0.093 0.07 0.09 [−0.05; 0.24] 3.2% 4.1%
NSPHS (866) 0.063 0.05 0.06 [−0.03; 0.16] 7.3% 8.7%
ORCADES (980) 0.120 0.05 0.12 [ 0.02; 0.22] 6.4% 7.8%
STABILITY (2951) 0.166 0.03 0.17 [ 0.11; 0.22] 23.7% 22.4%
STANLEY (344) 0.098 0.08 0.10 [−0.07; 0.26] 2.4% 3.1%
STANLEY (300) 0.086 0.08 0.09 [−0.06; 0.23] 3.0% 3.8%
VIS (902) 0.097 0.05 0.10 [−0.01; 0.20] 6.2% 7.5%
Common effect model 0.12 [ 0.10; 0.15] 100.0% --
Random effects model 0.12 [ 0.09; 0.15] -- 100.0%
−0.2 −0.1 0 0.1 0.2
Heterogeneity: I^2 = 0%, τ^2 = 0.0003, p = 0.57

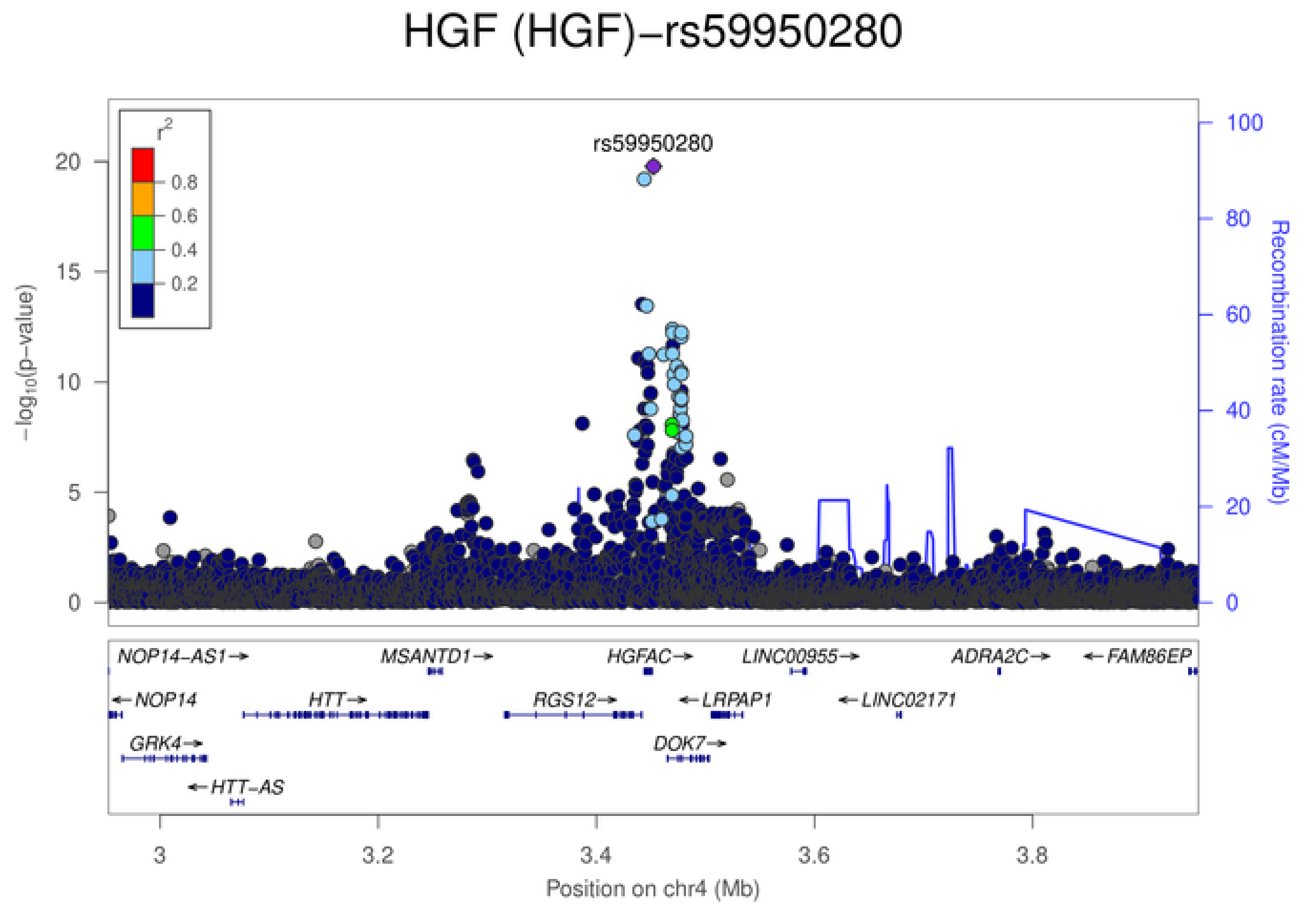
HGF (HGF)−rs59950280
rs59950280
r^2
0.8
0.6
0.4
0.2
−log10(p−value)
Recombination rate (cM/Mb)
NOP14−AS1→
MSANTD1→
HGFAC→
LINC00955→
ADRA2C→
←FAM86EP
←NOP14
HTT→
RGS12→
←LRPAP1
←LINC02171
GRK4→
DOK7→
←HTT−AS
3
3.2
3.4
3.6
3.8
Position on chr4 (Mb)

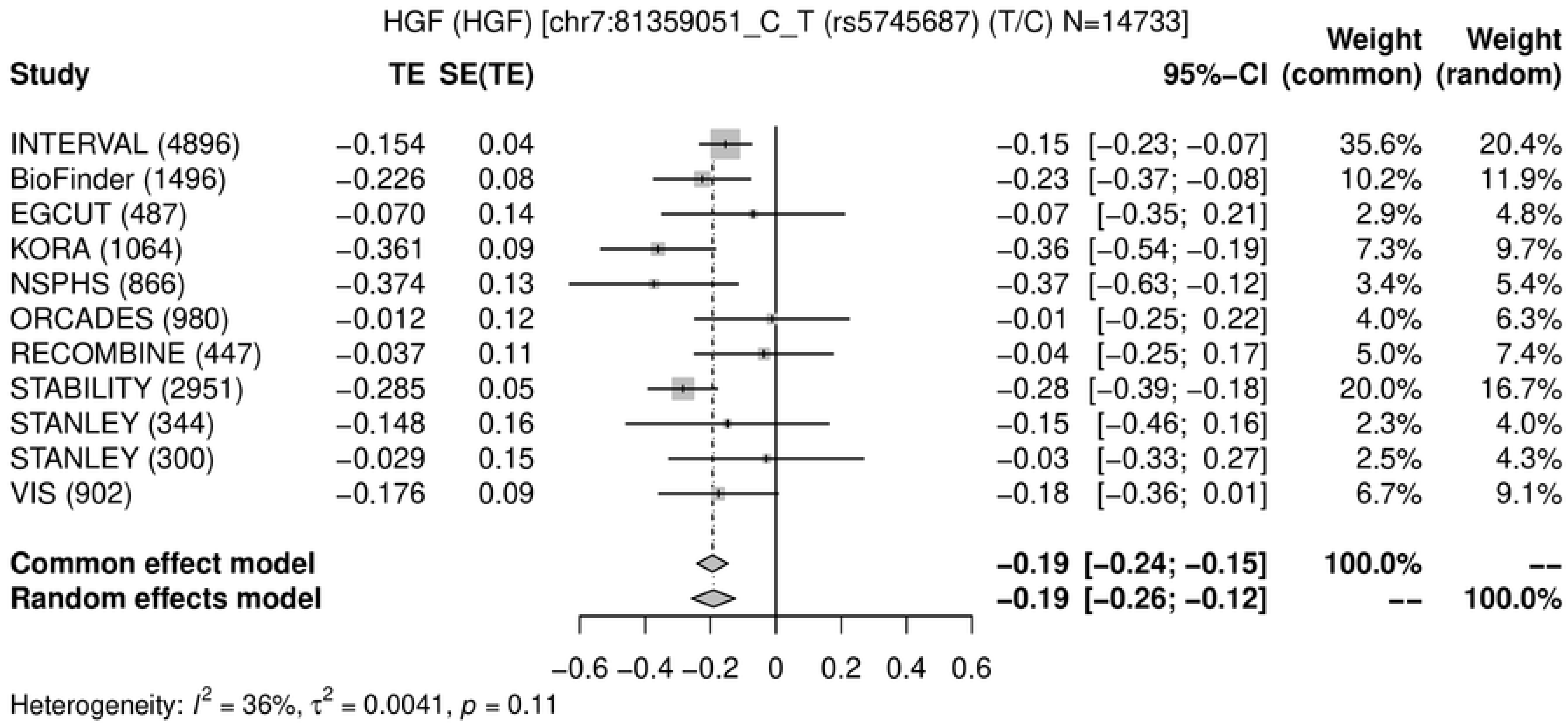
HGF (HGF) [chr7:81359051_C_T (rs5745687) (T/C) N=14733]
Study
TE SE(TE)
95%-CI
Weight (common)
Weight (random)
INTERVAL (4896) −0.154 0.04 −0.15 [−0.23; −0.07] 35.6% 20.4%
BioFinder (1496) −0.226 0.08 −0.23 [−0.37; −0.08] 10.2% 11.9%
EGCUT (487) −0.070 0.14 −0.07 [−0.35; 0.21] 2.9% 4.8%
KORA (1064) −0.361 0.09 −0.36 [−0.54; −0.19] 7.3% 9.7%
NSPHS (866) −0.374 0.13 −0.37 [−0.63; −0.12] 3.4% 5.4%
ORCADES (980) −0.012 0.12 −0.01 [−0.25; 0.22] 4.0% 6.3%
RECOMBINE (447) −0.037 0.11 −0.04 [−0.25; 0.17] 5.0% 7.4%
STABILITY (2951) −0.285 0.05 −0.28 [−0.39; −0.18] 20.0% 16.7%
STANLEY (344) −0.148 0.16 −0.15 [−0.46; 0.16] 2.3% 4.0%
STANLEY (300) −0.029 0.15 −0.03 [−0.33; 0.27] 2.5% 4.3%
VIS (902) −0.176 0.09 −0.18 [−0.36; 0.01] 6.7% 9.1%
Common effect model −0.19 [−0.24; −0.15] 100.0% --
Random effects model −0.19 [−0.26; −0.12] -- 100.0%
−0.6 −0.4 −0.2 0 0.2 0.4 0.6
Heterogeneity: $I^2$ = 36%, $\tau^2$ = 0.0041, $p$ = 0.11

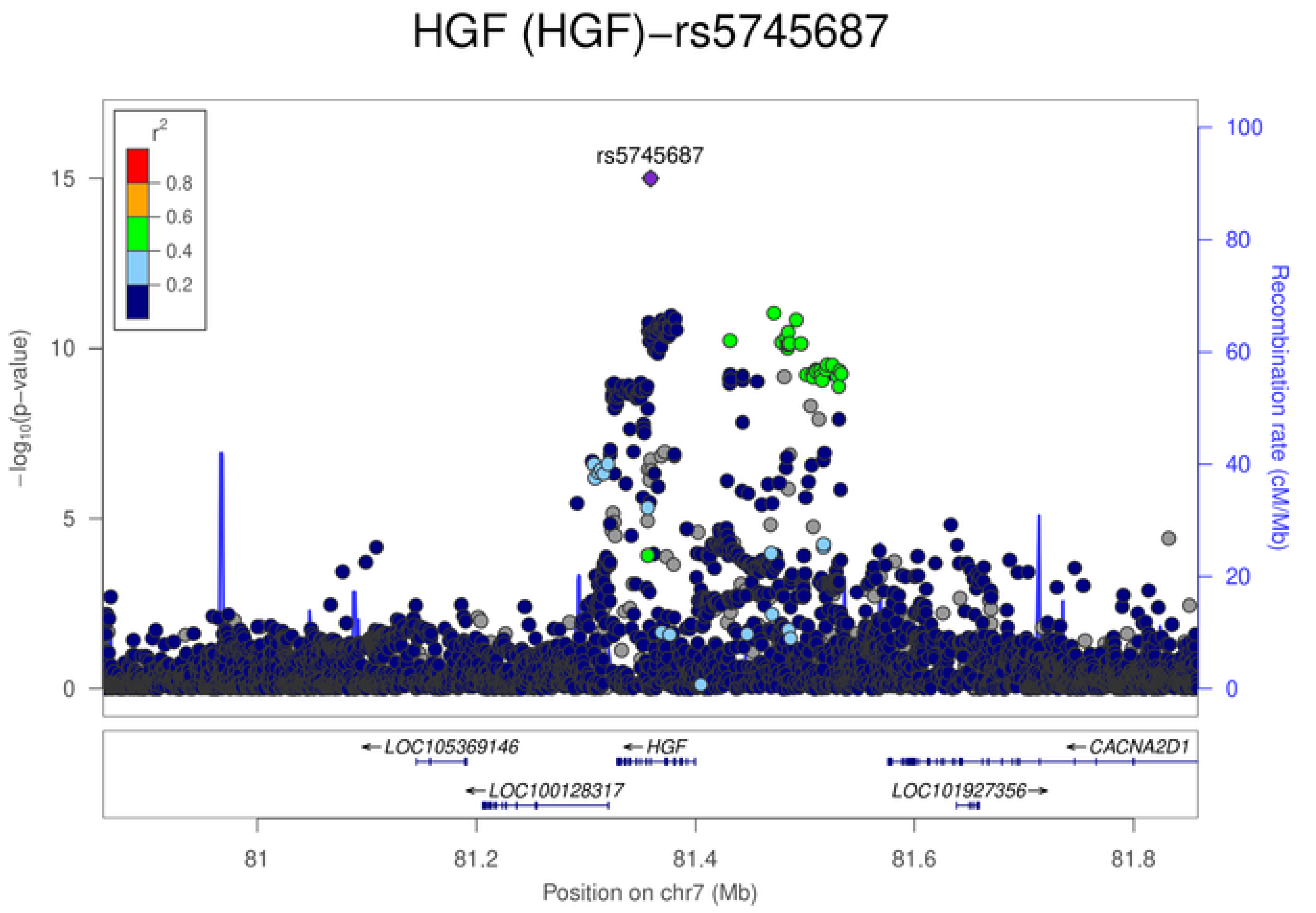
HGF (HGF)−rs5745687
$r^2$
0.8
0.6
0.4
0.2
rs5745687
$-\log_{10}$(p−value)
Recombination rate (cM/Mb)
LOC105369146
HGF
CACNA2D1
LOC100128317
LOC101927356
81 81.2 81.4 81.6 81.8
Position on chr7 (Mb)

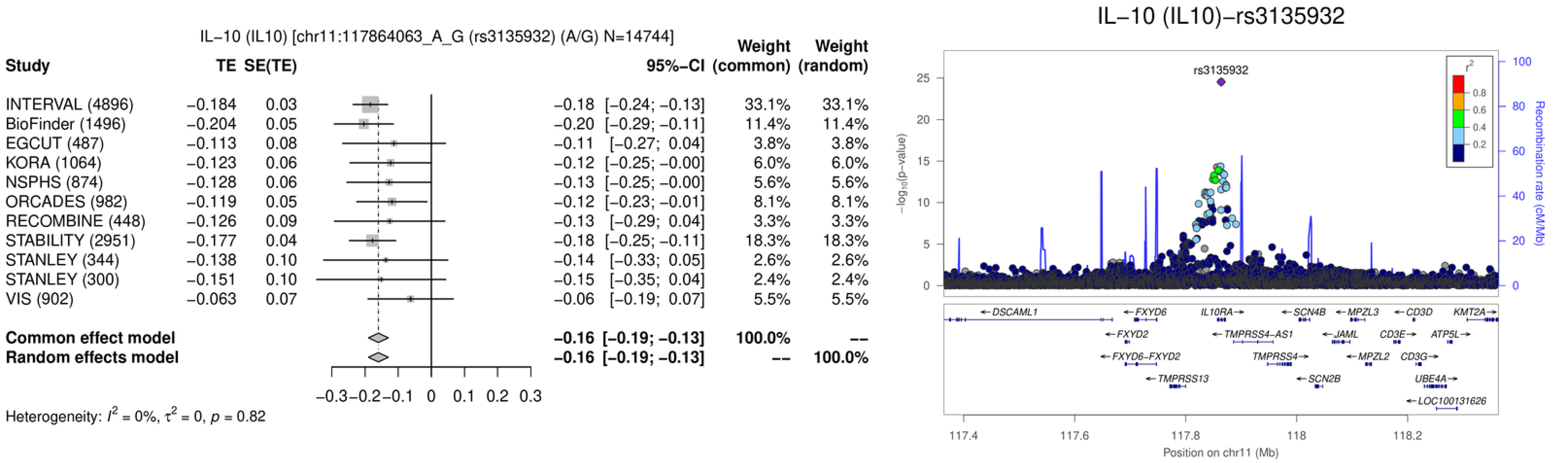
IL−10 (IL10) [chr11:117864063_A_G (rs3135932) (A/G) N=14744]
Study
TE
SE(TE)
95%−CI
Weight (common)
Weight (random)
INTERVAL (4896) −0.184 0.03 −0.18 [−0.24; −0.13] 33.1% 33.1%
BioFinder (1496) −0.204 0.05 −0.20 [−0.29; −0.11] 11.4% 11.4%
EGCUT (487) −0.113 0.08 −0.11 [−0.27; 0.04] 3.8% 3.8%
KORA (1064) −0.123 0.06 −0.12 [−0.25; −0.00] 6.0% 6.0%
NSPHS (874) −0.128 0.06 −0.13 [−0.25; −0.00] 5.6% 5.6%
ORCADES (982) −0.119 0.05 −0.12 [−0.23; −0.01] 8.1% 8.1%
RECOMBINE (448) −0.126 0.09 −0.13 [−0.29; 0.04] 3.3% 3.3%
STABILITY (2951) −0.177 0.04 −0.18 [−0.25; −0.11] 18.3% 18.3%
STANLEY (344) −0.138 0.10 −0.14 [−0.33; 0.05] 2.6% 2.6%
STANLEY (300) −0.151 0.10 −0.15 [−0.35; 0.04] 2.4% 2.4%
VIS (902) −0.063 0.07 −0.06 [−0.19; 0.07] 5.5% 5.5%
Common effect model −0.16 [−0.19; −0.13] 100.0% --
Random effects model −0.16 [−0.19; −0.13] -- 100.0%
−0.3 −0.2 −0.1 0 0.1 0.2 0.3
Heterogeneity: $I^2$ = 0%, $\tau^2$ = 0, $p$ = 0.82
IL−10 (IL10)−rs3135932
rs3135932
$-\log_{10}$(p−value)
Recombination rate (cM/Mb)
$r^2$
0.8
0.6
0.4
0.2
DSCAML1
FXYD6
IL10RA
SCN4B
MPZL3
CD3D
KMT2A
FXYD2
TMPRSS4-AS1
JAML
CD3E
ATP5L
FXYD6-FXYD2
TMPRSS4
MPZL2
CD3G
TMPRSS13
SCN2B
UBE4A
LOC100131626
117.4 117.6 117.8 118 118.2
Position on chr11 (Mb)

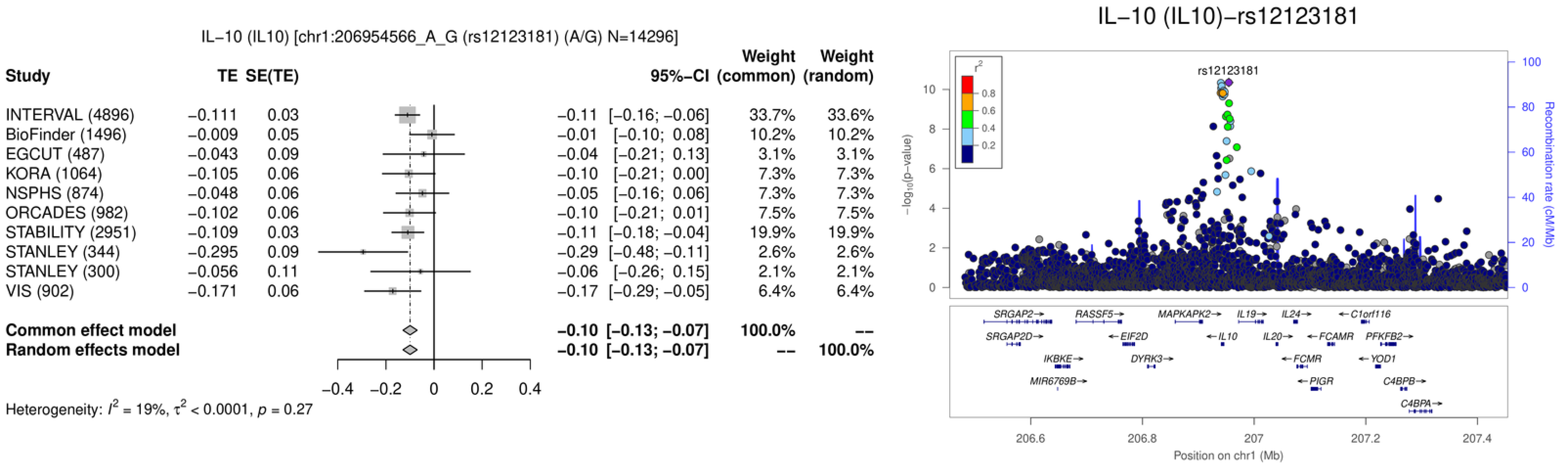
IL−10 (IL10) [chr1:206954566_A_G (rs12123181) (A/G) N=14296]
Study TE SE(TE) 95%−CI Weight (common) Weight (random)
INTERVAL (4896) −0.111 0.03 −0.11 [−0.16; −0.06] 33.7% 33.6%
BioFinder (1496) −0.009 0.05 −0.01 [−0.10; 0.08] 10.2% 10.2%
EGCUT (487) −0.043 0.09 −0.04 [−0.21; 0.13] 3.1% 3.1%
KORA (1064) −0.105 0.06 −0.10 [−0.21; 0.00] 7.3% 7.3%
NSPHS (874) −0.048 0.06 −0.05 [−0.16; 0.06] 7.3% 7.3%
ORCADES (982) −0.102 0.06 −0.10 [−0.21; 0.01] 7.5% 7.5%
STABILITY (2951) −0.109 0.03 −0.11 [−0.18; −0.04] 19.9% 19.9%
STANLEY (344) −0.295 0.09 −0.29 [−0.48; −0.11] 2.6% 2.6%
STANLEY (300) −0.056 0.11 −0.06 [−0.26; 0.15] 2.1% 2.1%
VIS (902) −0.171 0.06 −0.17 [−0.29; −0.05] 6.4% 6.4%
Common effect model −0.10 [−0.13; −0.07] 100.0% --
Random effects model −0.10 [−0.13; −0.07] -- 100.0%
−0.4 −0.2 0 0.2 0.4
Heterogeneity: $I^2$ = 19%, $\tau^2$ < 0.0001, $p$ = 0.27
IL−10 (IL10)−rs12123181
rs12123181
$r^2$
0.8 0.6 0.4 0.2
$-\log_{10}$(p−value)
0 2 4 6 8 10
Recombination rate (cM/Mb)
0 20 40 60 80 100
SRGAP2→ RASSF5→ MAPKAPK2→ IL19→ IL24→ ←C1orf116
SRGAP2D→ ←EIF2D ←IL10 IL20→ ←FCAMR PFKFB2→
IKBKE→ DYRK3→ ←FCMR ←YOD1
MIR6769B→ ←PIGR C4BPB→
C4BPA→
206.6 206.8 207 207.2 207.4
Position on chr1 (Mb)

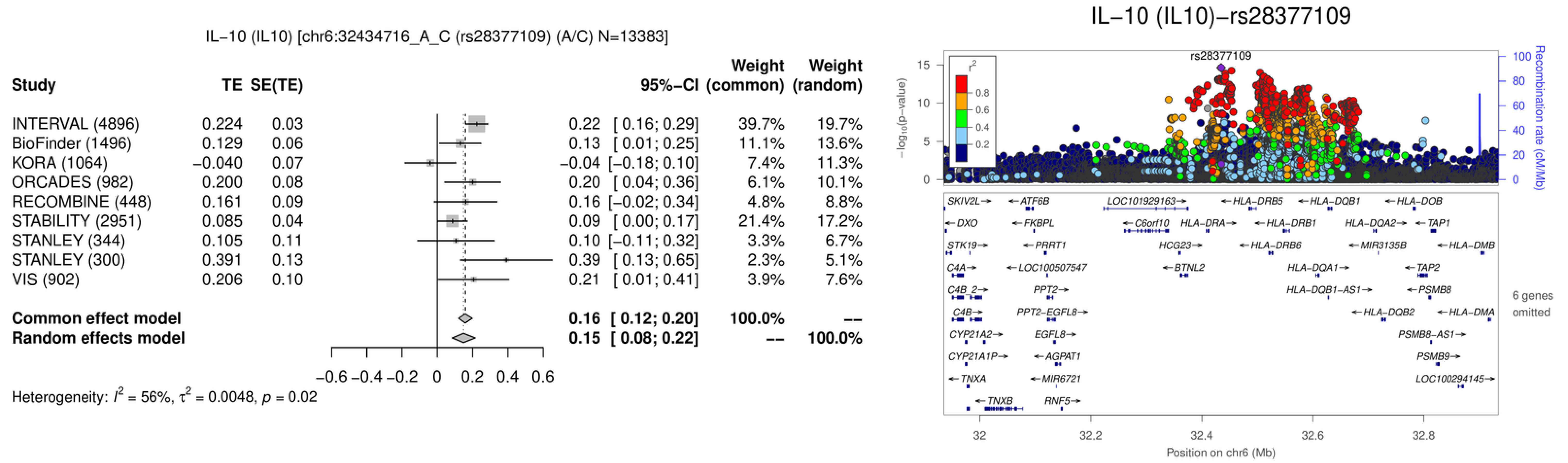
IL−10 (IL10) [chr6:32434716_A_C (rs28377109) (A/C) N=13383]
Study
TE
SE(TE)
95%−CI
Weight (common)
Weight (random)
INTERVAL (4896) 0.224 0.03 0.22 [ 0.16; 0.29] 39.7% 19.7%
BioFinder (1496) 0.129 0.06 0.13 [ 0.01; 0.25] 11.1% 13.6%
KORA (1064) −0.040 0.07 −0.04 [−0.18; 0.10] 7.4% 11.3%
ORCADES (982) 0.200 0.08 0.20 [ 0.04; 0.36] 6.1% 10.1%
RECOMBINE (448) 0.161 0.09 0.16 [−0.02; 0.34] 4.8% 8.8%
STABILITY (2951) 0.085 0.04 0.09 [ 0.00; 0.17] 21.4% 17.2%
STANLEY (344) 0.105 0.11 0.10 [−0.11; 0.32] 3.3% 6.7%
STANLEY (300) 0.391 0.13 0.39 [ 0.13; 0.65] 2.3% 5.1%
VIS (902) 0.206 0.10 0.21 [ 0.01; 0.41] 3.9% 7.6%
Common effect model 0.16 [ 0.12; 0.20] 100.0% --
Random effects model 0.15 [ 0.08; 0.22] -- 100.0%
−0.6 −0.4 −0.2 0 0.2 0.4 0.6
Heterogeneity: $I^2$ = 56%, $\tau^2$ = 0.0048, $p$ = 0.02
IL−10 (IL10)−rs28377109
rs28377109
$-\log_{10}$(p−value)
Recombination rate (cM/Mb)
$r^2$
0.8
0.6
0.4
0.2
SKIV2L, ATF6B, LOC101929163, HLA−DRB5, HLA−DQB1, HLA−DOB
DXO, FKBPL, C6orf10, HLA−DRA, HLA−DRB1, HLA−DQA2, TAP1
STK19, PRRT1, HCG23, HLA−DRB6, MIR3135B, HLA−DMB
C4A, LOC100507547, BTNL2, HLA−DQA1, TAP2
C4B_2, PPT2, HLA−DQB1−AS1, PSMB8
C4B, PPT2−EGFL8, HLA−DQB2, HLA−DMA
CYP21A2, EGFL8, PSMB8−AS1
CYP21A1P, AGPAT1, PSMB9
TNXA, MIR6721, LOC100294145
TNXB, RNF5
6 genes omitted
32 32.2 32.4 32.6 32.8
Position on chr6 (Mb)

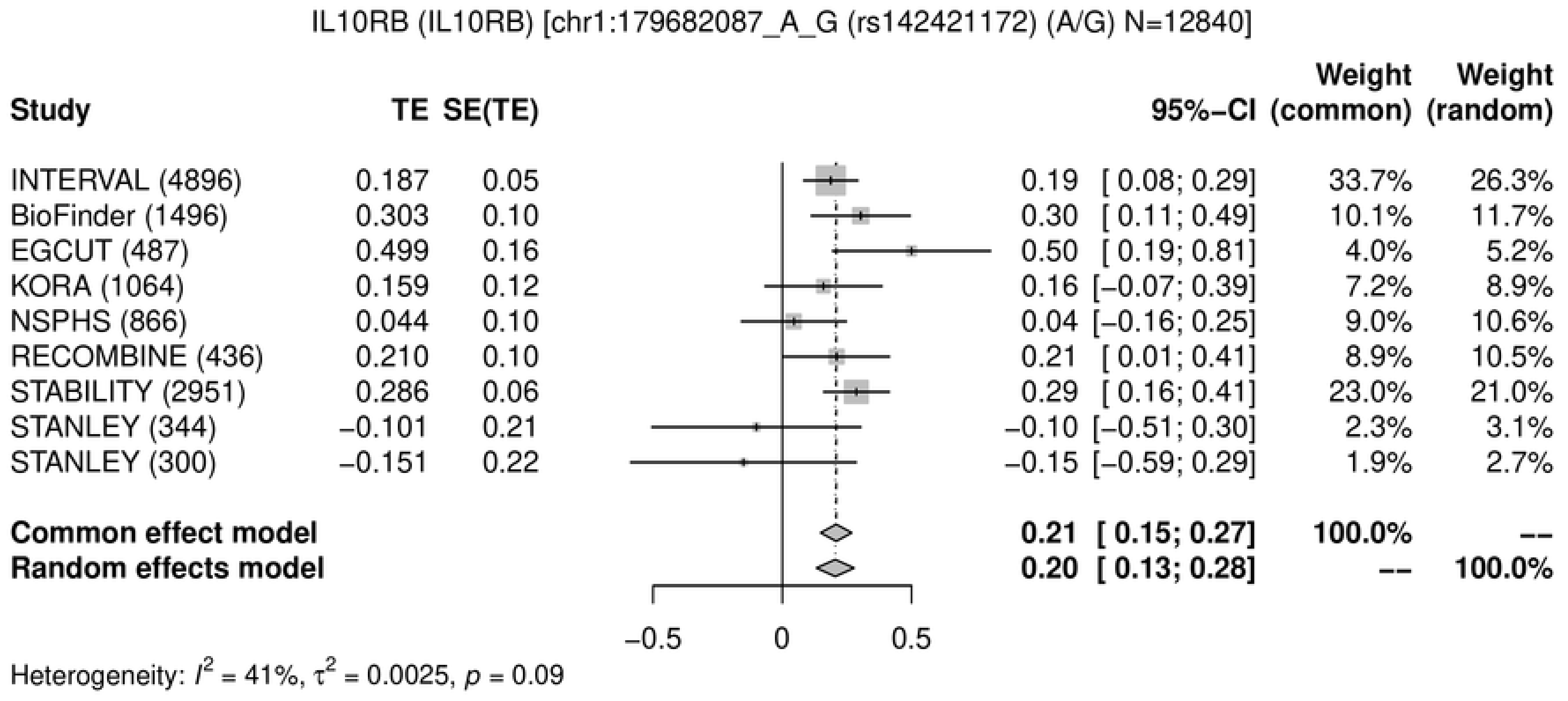
IL10RB (IL10RB) [chr1:179682087_A_G (rs142421172) (A/G) N=12840]
Study TE SE(TE) 95%-CI Weight (common) Weight (random)
INTERVAL (4896) 0.187 0.05 0.19 [ 0.08; 0.29] 33.7% 26.3%
BioFinder (1496) 0.303 0.10 0.30 [ 0.11; 0.49] 10.1% 11.7%
EGCUT (487) 0.499 0.16 0.50 [ 0.19; 0.81] 4.0% 5.2%
KORA (1064) 0.159 0.12 0.16 [−0.07; 0.39] 7.2% 8.9%
NSPHS (866) 0.044 0.10 0.04 [−0.16; 0.25] 9.0% 10.6%
RECOMBINE (436) 0.210 0.10 0.21 [ 0.01; 0.41] 8.9% 10.5%
STABILITY (2951) 0.286 0.06 0.29 [ 0.16; 0.41] 23.0% 21.0%
STANLEY (344) −0.101 0.21 −0.10 [−0.51; 0.30] 2.3% 3.1%
STANLEY (300) −0.151 0.22 −0.15 [−0.59; 0.29] 1.9% 2.7%
Common effect model 0.21 [ 0.15; 0.27] 100.0% --
Random effects model 0.20 [ 0.13; 0.28] -- 100.0%
−0.5
0
0.5
Heterogeneity: $I^2$ = 41%, $\tau^2$ = 0.0025, $p$ = 0.09

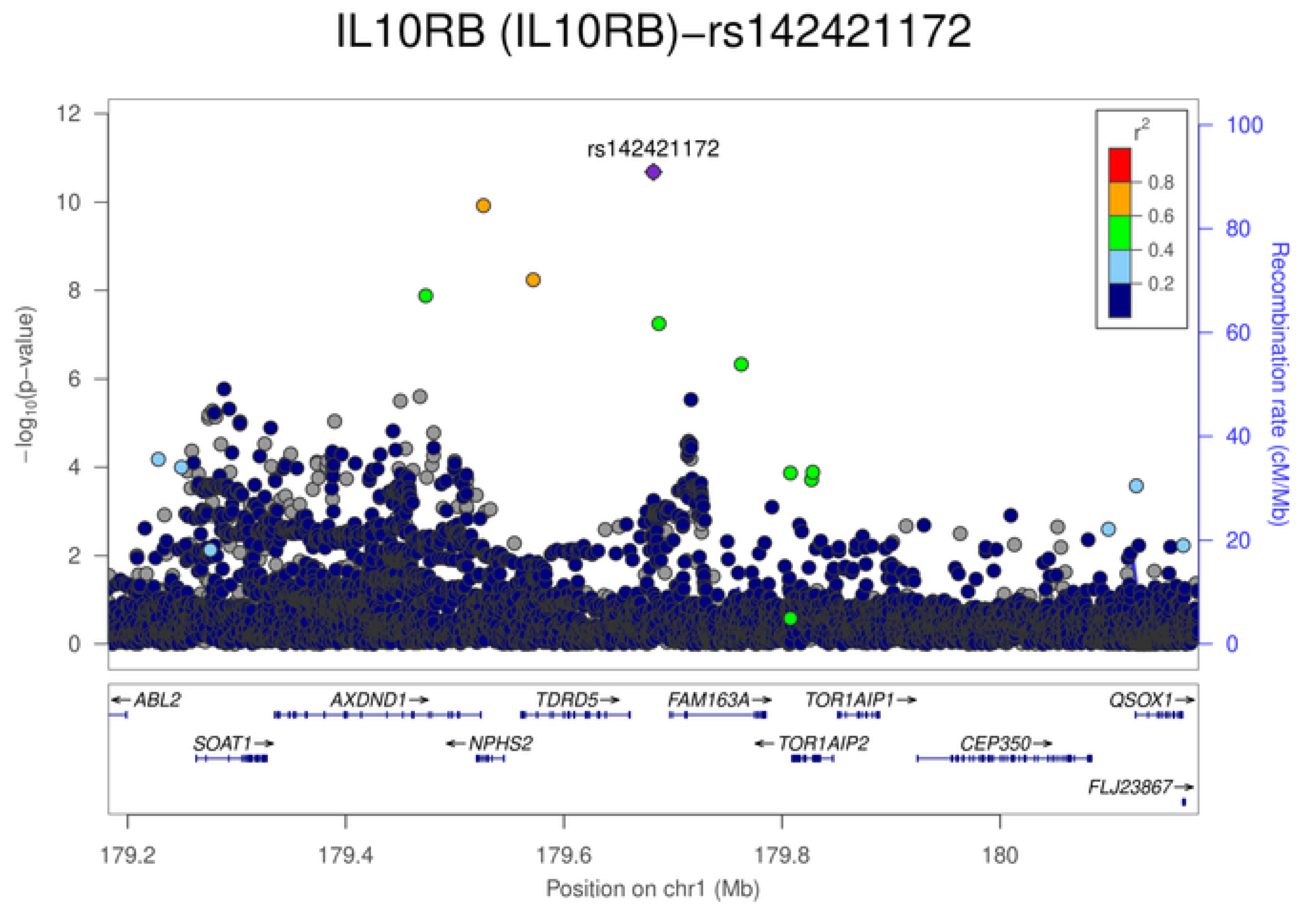
IL10RB (IL10RB)−rs142421172
rs142421172
$-\log_{10}$(p−value)
Recombination rate (cM/Mb)
$r^2$
0.8
0.6
0.4
0.2
ABL2
AXDND1
TDRD5
FAM163A
TOR1AIP1
QSOX1
SOAT1
NPHS2
TOR1AIP2
CEP350
FLJ23867
179.2
179.4
179.6
179.8
180
Position on chr1 (Mb)

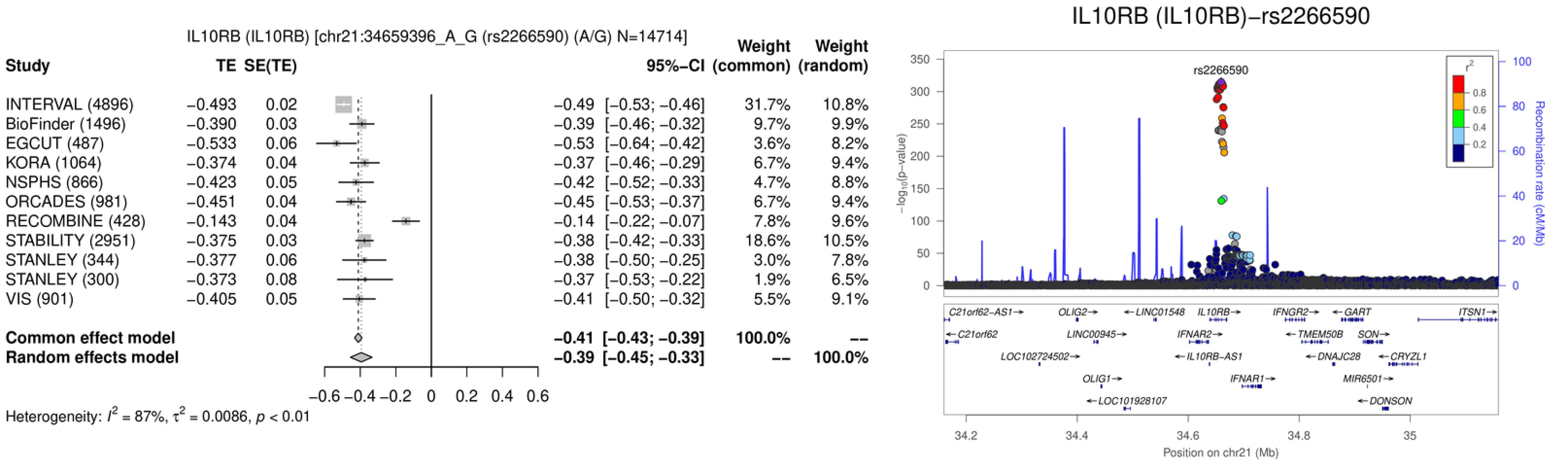
IL10RB (IL10RB) [chr21:34659396_A_G (rs2266590) (A/G) N=14714]
Study
TE
SE(TE)
95%−CI
Weight (common)
Weight (random)
INTERVAL (4896) −0.493 0.02 −0.49 [−0.53; −0.46] 31.7% 10.8%
BioFinder (1496) −0.390 0.03 −0.39 [−0.46; −0.32] 9.7% 9.9%
EGCUT (487) −0.533 0.06 −0.53 [−0.64; −0.42] 3.6% 8.2%
KORA (1064) −0.374 0.04 −0.37 [−0.46; −0.29] 6.7% 9.4%
NSPHS (866) −0.423 0.05 −0.42 [−0.52; −0.33] 4.7% 8.8%
ORCADES (981) −0.451 0.04 −0.45 [−0.53; −0.37] 6.7% 9.4%
RECOMBINE (428) −0.143 0.04 −0.14 [−0.22; −0.07] 7.8% 9.6%
STABILITY (2951) −0.375 0.03 −0.38 [−0.42; −0.33] 18.6% 10.5%
STANLEY (344) −0.377 0.06 −0.38 [−0.50; −0.25] 3.0% 7.8%
STANLEY (300) −0.373 0.08 −0.37 [−0.53; −0.22] 1.9% 6.5%
VIS (901) −0.405 0.05 −0.41 [−0.50; −0.32] 5.5% 9.1%
Common effect model −0.41 [−0.43; −0.39] 100.0% --
Random effects model −0.39 [−0.45; −0.33] -- 100.0%
−0.6 −0.4 −0.2 0 0.2 0.4 0.6
Heterogeneity: $I^2$ = 87%, $\tau^2$ = 0.0086, $p$ < 0.01
IL10RB (IL10RB)−rs2266590
rs2266590
$r^2$
0.8
0.6
0.4
0.2
350
300
250
200
150
100
50
0
$-\log_{10}$(p−value)
100
80
60
40
20
0
Recombination rate (cM/Mb)
C21orf62-AS1→
OLIG2→
←LINC01548
IL10RB→
IFNGR2→
←GART
ITSN1→
←C21orf62
LINC00945→
IFNAR2→
←TMEM50B
SON→
LOC102724502→
←IL10RB-AS1
←DNAJC28
←CRYZL1
OLIG1→
IFNAR1→
MIR6501→
←LOC101928107
←DONSON
34.2
34.4
34.6
34.8
35
Position on chr21 (Mb)

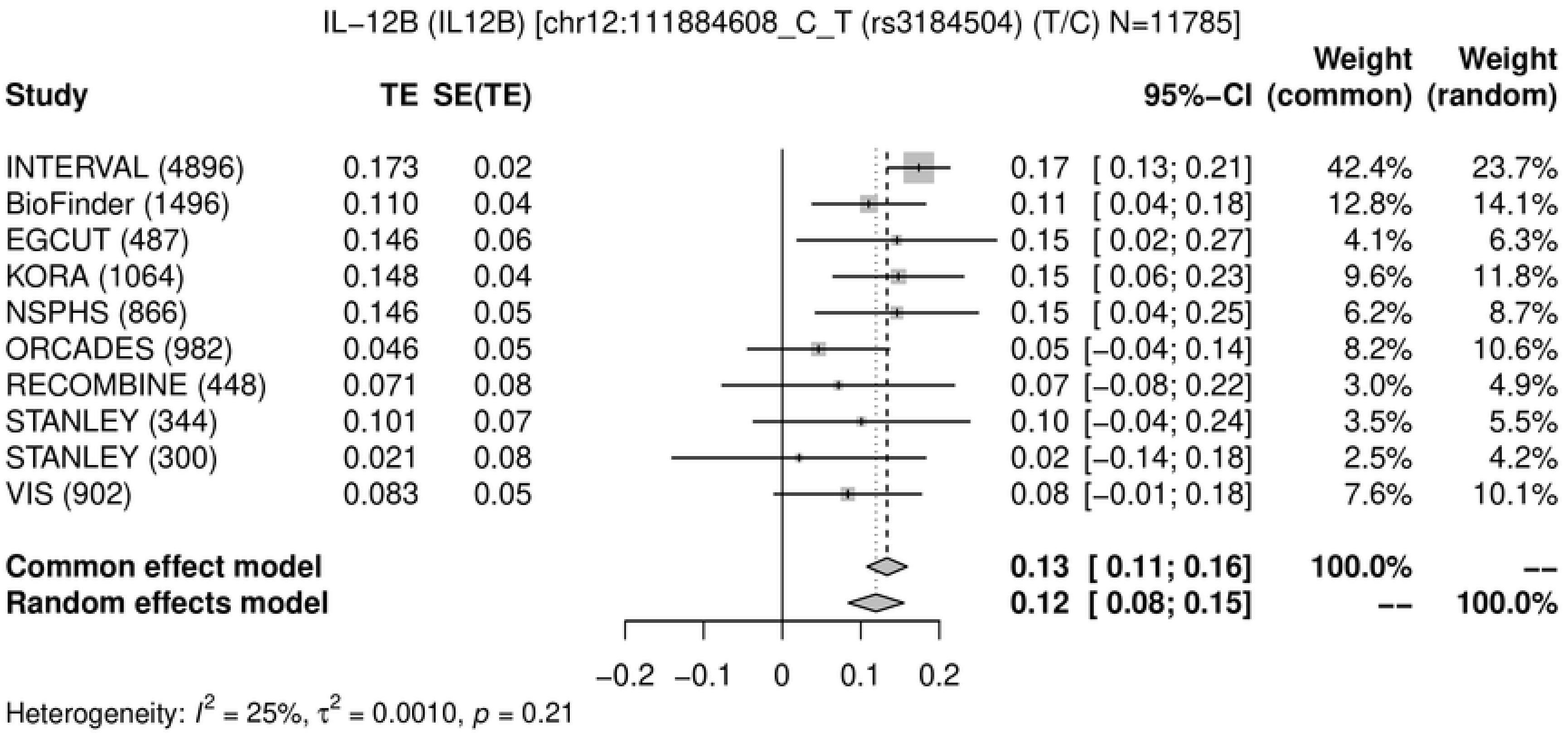
IL−12B (IL12B) [chr12:111884608_C_T (rs3184504) (T/C) N=11785]
Study
TE
SE(TE)
95%−CI
Weight (common)
Weight (random)
INTERVAL (4896) 0.173 0.02 0.17 [ 0.13; 0.21] 42.4% 23.7%
BioFinder (1496) 0.110 0.04 0.11 [ 0.04; 0.18] 12.8% 14.1%
EGCUT (487) 0.146 0.06 0.15 [ 0.02; 0.27] 4.1% 6.3%
KORA (1064) 0.148 0.04 0.15 [ 0.06; 0.23] 9.6% 11.8%
NSPHS (866) 0.146 0.05 0.15 [ 0.04; 0.25] 6.2% 8.7%
ORCADES (982) 0.046 0.05 0.05 [−0.04; 0.14] 8.2% 10.6%
RECOMBINE (448) 0.071 0.08 0.07 [−0.08; 0.22] 3.0% 4.9%
STANLEY (344) 0.101 0.07 0.10 [−0.04; 0.24] 3.5% 5.5%
STANLEY (300) 0.021 0.08 0.02 [−0.14; 0.18] 2.5% 4.2%
VIS (902) 0.083 0.05 0.08 [−0.01; 0.18] 7.6% 10.1%
Common effect model 0.13 [ 0.11; 0.16] 100.0% --
Random effects model 0.12 [ 0.08; 0.15] -- 100.0%
−0.2 −0.1 0 0.1 0.2
Heterogeneity: $I^2$ = 25%, $\tau^2$ = 0.0010, $p$ = 0.21

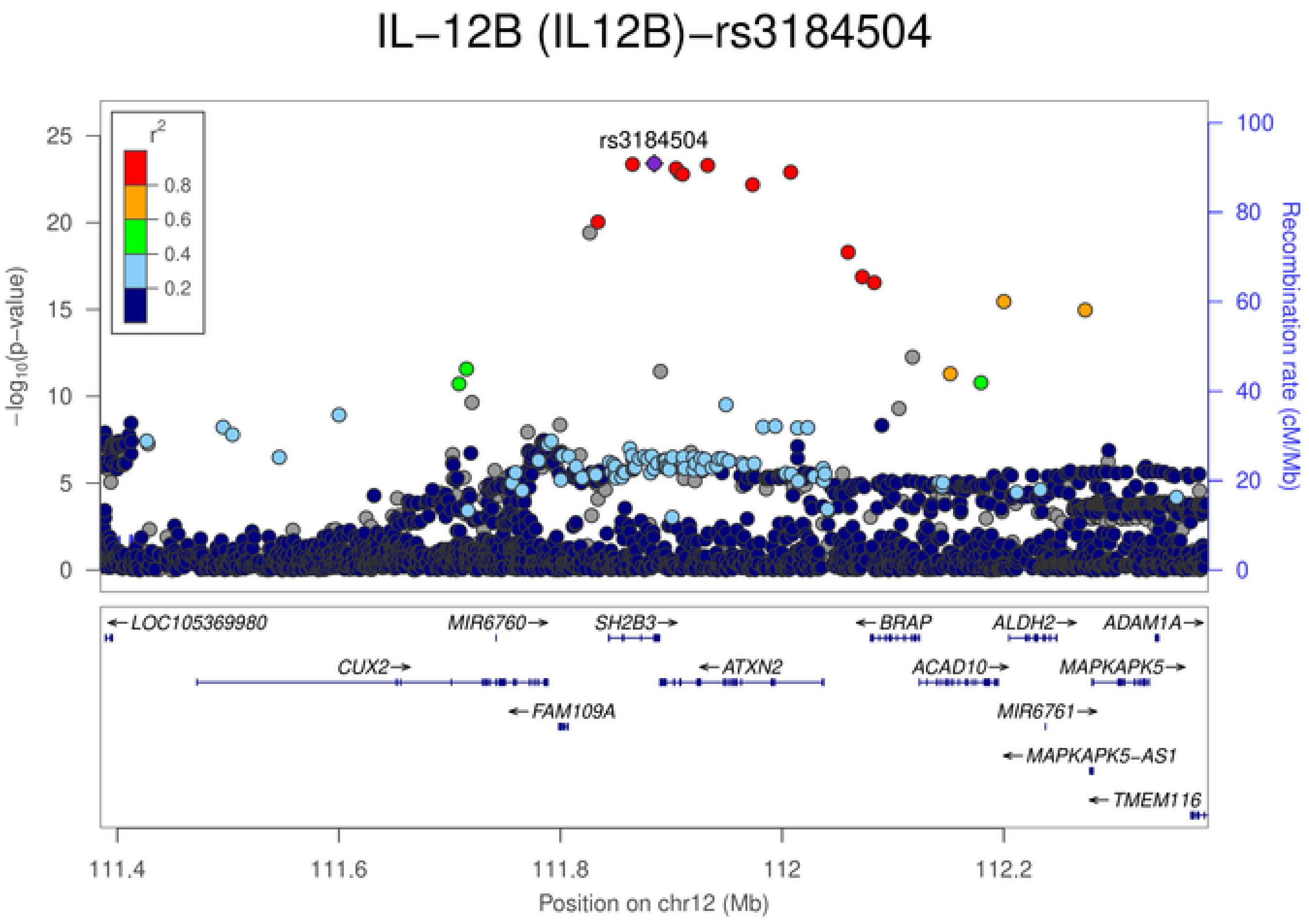
IL−12B (IL12B)−rs3184504
rs3184504
$r^2$
0.8
0.6
0.4
0.2
$-\log_{10}$(p−value)
Recombination rate (cM/Mb)
0
5
10
15
20
25
0
20
40
60
80
100
LOC105369980
MIR6760
SH2B3
BRAP
ALDH2
ADAM1A
CUX2
ATXN2
ACAD10
MAPKAPK5
FAM109A
MIR6761
MAPKAPK5−AS1
TMEM116
111.4
111.6
111.8
112
112.2
Position on chr12 (Mb)

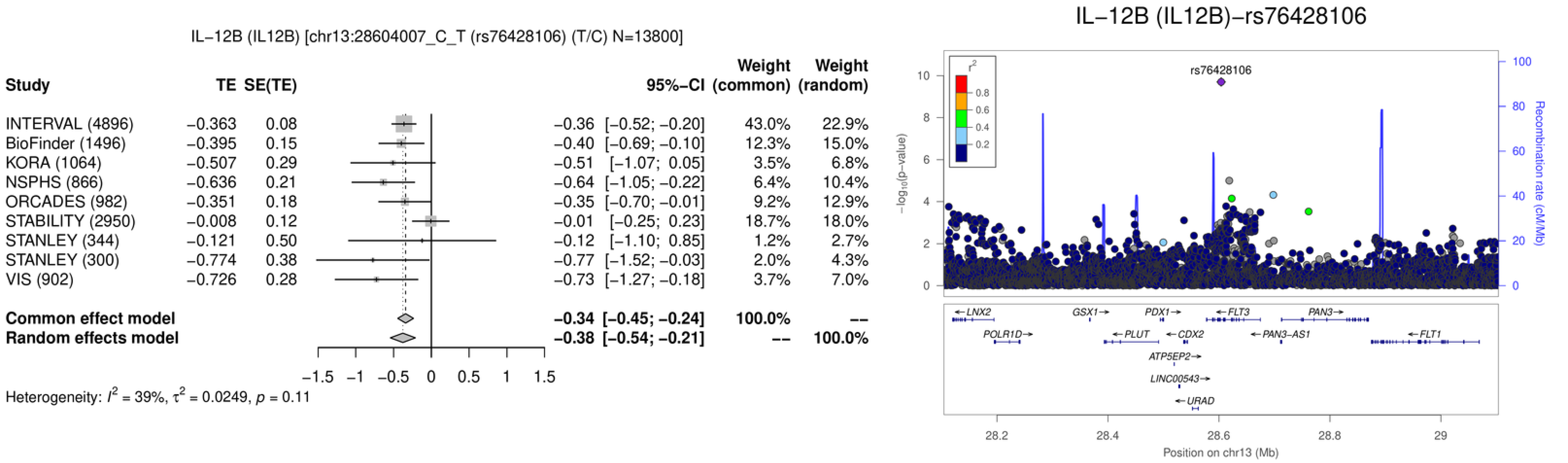
IL−12B (IL12B) [chr13:28604007_C_T (rs76428106) (T/C) N=13800]
Study
TE
SE(TE)
95%−CI
Weight (common)
Weight (random)
INTERVAL (4896) −0.363 0.08 −0.36 [−0.52; −0.20] 43.0% 22.9%
BioFinder (1496) −0.395 0.15 −0.40 [−0.69; −0.10] 12.3% 15.0%
KORA (1064) −0.507 0.29 −0.51 [−1.07; 0.05] 3.5% 6.8%
NSPHS (866) −0.636 0.21 −0.64 [−1.05; −0.22] 6.4% 10.4%
ORCADES (982) −0.351 0.18 −0.35 [−0.70; −0.01] 9.2% 12.9%
STABILITY (2950) −0.008 0.12 −0.01 [−0.25; 0.23] 18.7% 18.0%
STANLEY (344) −0.121 0.50 −0.12 [−1.10; 0.85] 1.2% 2.7%
STANLEY (300) −0.774 0.38 −0.77 [−1.52; −0.03] 2.0% 4.3%
VIS (902) −0.726 0.28 −0.73 [−1.27; −0.18] 3.7% 7.0%
Common effect model −0.34 [−0.45; −0.24] 100.0% --
Random effects model −0.38 [−0.54; −0.21] -- 100.0%
−1.5 −1 −0.5 0 0.5 1 1.5
Heterogeneity: $I^2$ = 39%, $\tau^2$ = 0.0249, $p$ = 0.11
IL−12B (IL12B)−rs76428106
rs76428106
$r^2$
0.8
0.6
0.4
0.2
$-\log_{10}$(p−value)
Recombination rate (cM/Mb)
0 2 4 6 8 10
0 20 40 60 80 100
LNX2
GSX1
PDX1
FLT3
PAN3
POLR1D
PLUT
CDX2
PAN3-AS1
FLT1
ATP5EP2
LINC00543
URAD
28.2 28.4 28.6 28.8 29
Position on chr13 (Mb)

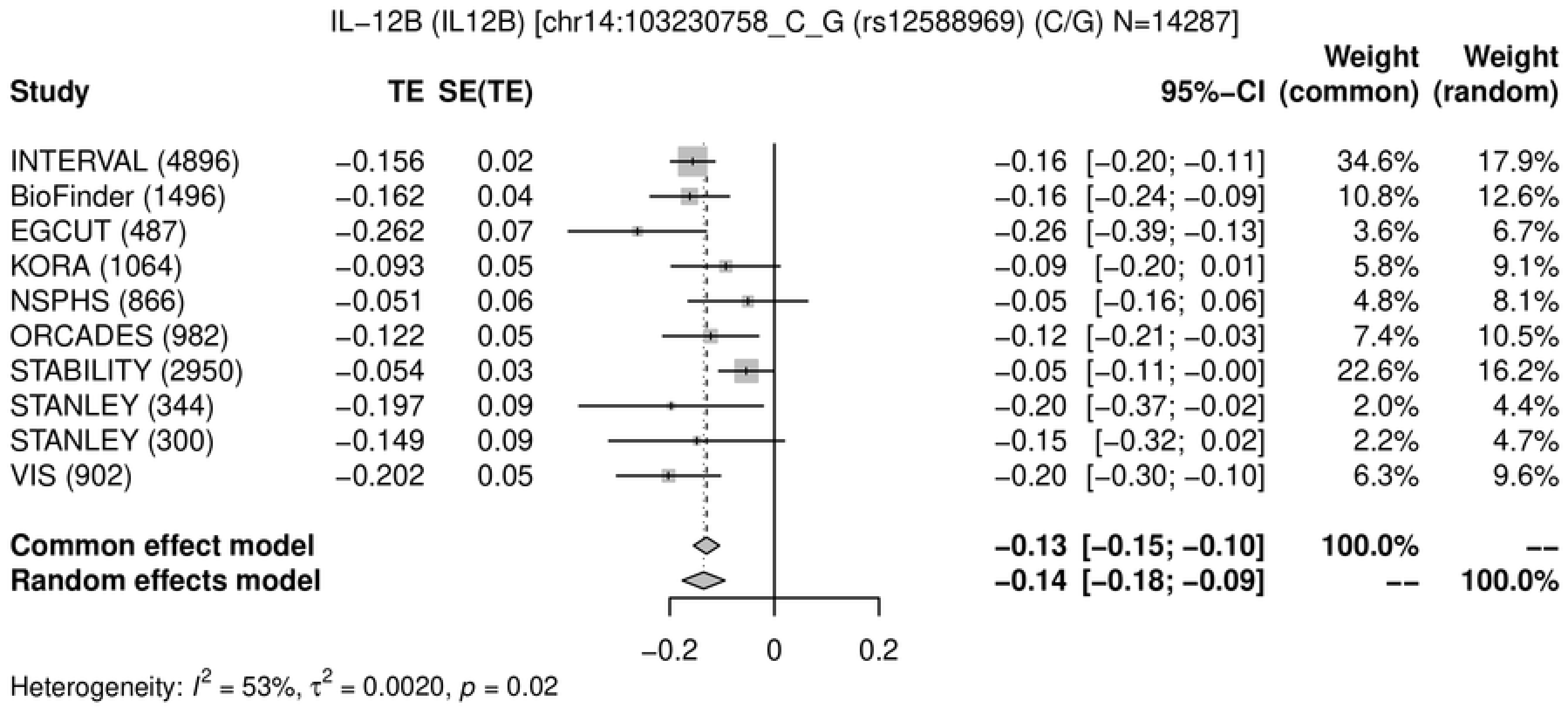

IL−12B (IL12B) [chr14:103230758_C_G (rs12588969) (C/G) N=14287]
Study
TE
SE(TE)
95%−CI
Weight (common)
Weight (random)
INTERVAL (4896) −0.156 0.02 −0.16 [−0.20; −0.11] 34.6% 17.9%
BioFinder (1496) −0.162 0.04 −0.16 [−0.24; −0.09] 10.8% 12.6%
EGCUT (487) −0.262 0.07 −0.26 [−0.39; −0.13] 3.6% 6.7%
KORA (1064) −0.093 0.05 −0.09 [−0.20; 0.01] 5.8% 9.1%
NSPHS (866) −0.051 0.06 −0.05 [−0.16; 0.06] 4.8% 8.1%
ORCADES (982) −0.122 0.05 −0.12 [−0.21; −0.03] 7.4% 10.5%
STABILITY (2950) −0.054 0.03 −0.05 [−0.11; −0.00] 22.6% 16.2%
STANLEY (344) −0.197 0.09 −0.20 [−0.37; −0.02] 2.0% 4.4%
STANLEY (300) −0.149 0.09 −0.15 [−0.32; 0.02] 2.2% 4.7%
VIS (902) −0.202 0.05 −0.20 [−0.30; −0.10] 6.3% 9.6%
Common effect model −0.13 [−0.15; −0.10] 100.0% --
Random effects model −0.14 [−0.18; −0.09] -- 100.0%
−0.2
0
0.2
Heterogeneity: $I^2$ = 53%, $\tau^2$ = 0.0020, $p$ = 0.02


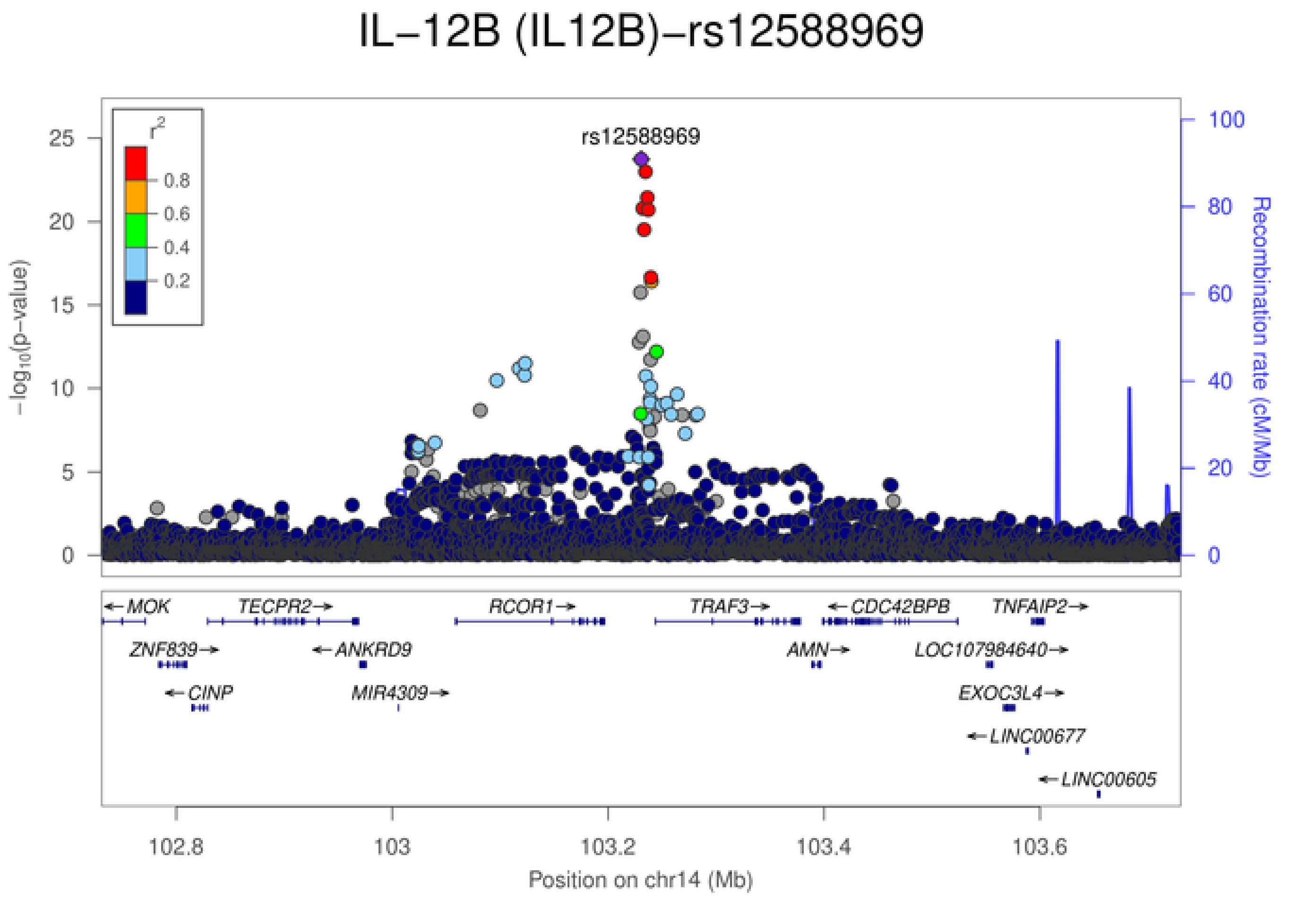

IL−12B (IL12B)−rs12588969
rs12588969
$r^2$
0.8
0.6
0.4
0.2
$-\log_{10}$(p−value)
Recombination rate (cM/Mb)
MOK
TECPR2
RCOR1
TRAF3
CDC42BPB
TNFAIP2
ZNF839
ANKRD9
AMN
LOC107984640
CINP
MIR4309
EXOC3L4
LINC00677
LINC00605
102.8
103
103.2
103.4
103.6
Position on chr14 (Mb)

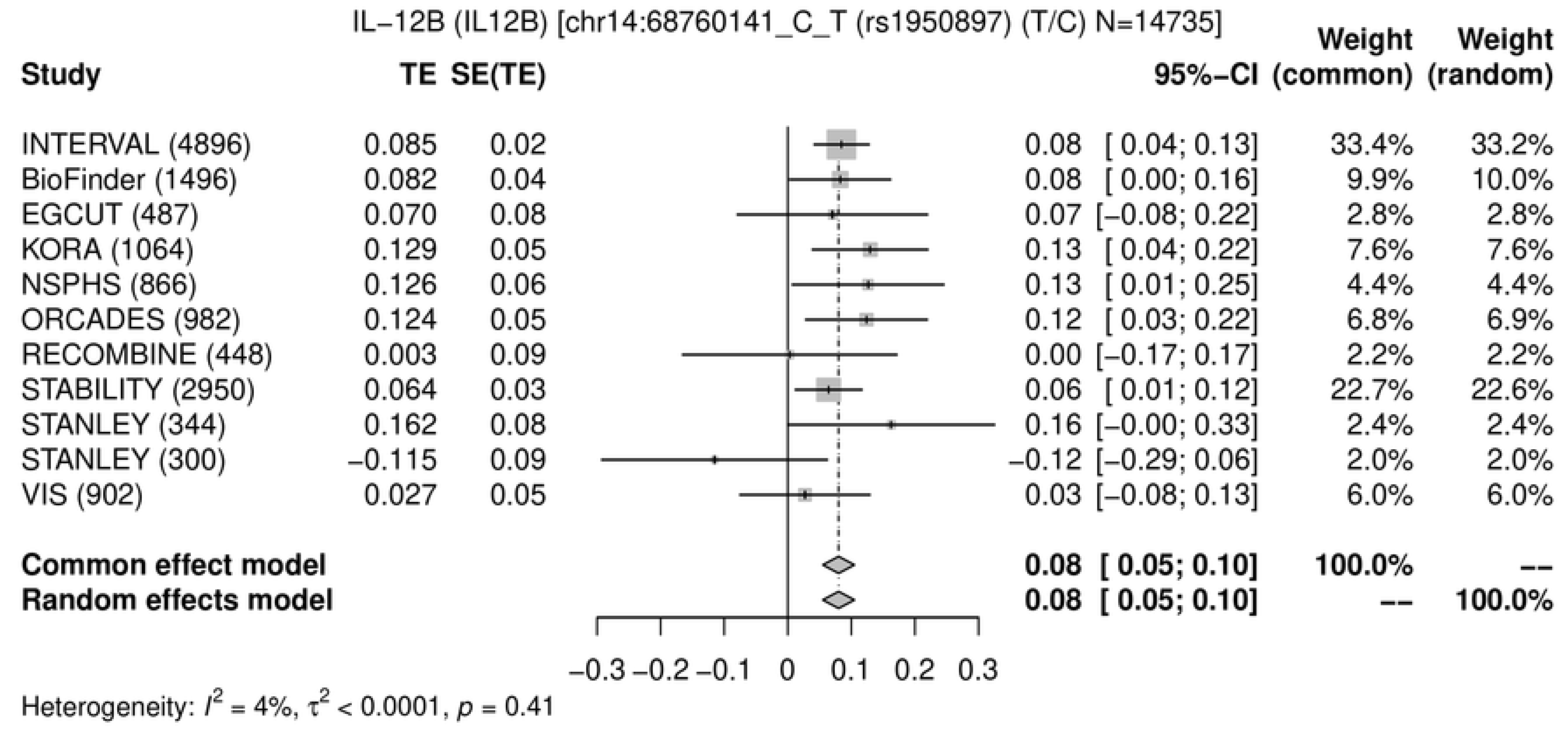
IL−12B (IL12B) [chr14:68760141_C_T (rs1950897) (T/C) N=14735]
Study
TE
SE(TE)
95%−CI
Weight (common)
Weight (random)
INTERVAL (4896) 0.085 0.02 0.08 [ 0.04; 0.13] 33.4% 33.2%
BioFinder (1496) 0.082 0.04 0.08 [ 0.00; 0.16] 9.9% 10.0%
EGCUT (487) 0.070 0.08 0.07 [−0.08; 0.22] 2.8% 2.8%
KORA (1064) 0.129 0.05 0.13 [ 0.04; 0.22] 7.6% 7.6%
NSPHS (866) 0.126 0.06 0.13 [ 0.01; 0.25] 4.4% 4.4%
ORCADES (982) 0.124 0.05 0.12 [ 0.03; 0.22] 6.8% 6.9%
RECOMBINE (448) 0.003 0.09 0.00 [−0.17; 0.17] 2.2% 2.2%
STABILITY (2950) 0.064 0.03 0.06 [ 0.01; 0.12] 22.7% 22.6%
STANLEY (344) 0.162 0.08 0.16 [−0.00; 0.33] 2.4% 2.4%
STANLEY (300) −0.115 0.09 −0.12 [−0.29; 0.06] 2.0% 2.0%
VIS (902) 0.027 0.05 0.03 [−0.08; 0.13] 6.0% 6.0%
Common effect model 0.08 [ 0.05; 0.10] 100.0% --
Random effects model 0.08 [ 0.05; 0.10] -- 100.0%
−0.3 −0.2 −0.1 0 0.1 0.2 0.3
Heterogeneity: $I^2$ = 4%, $\tau^2$ < 0.0001, $p$ = 0.41

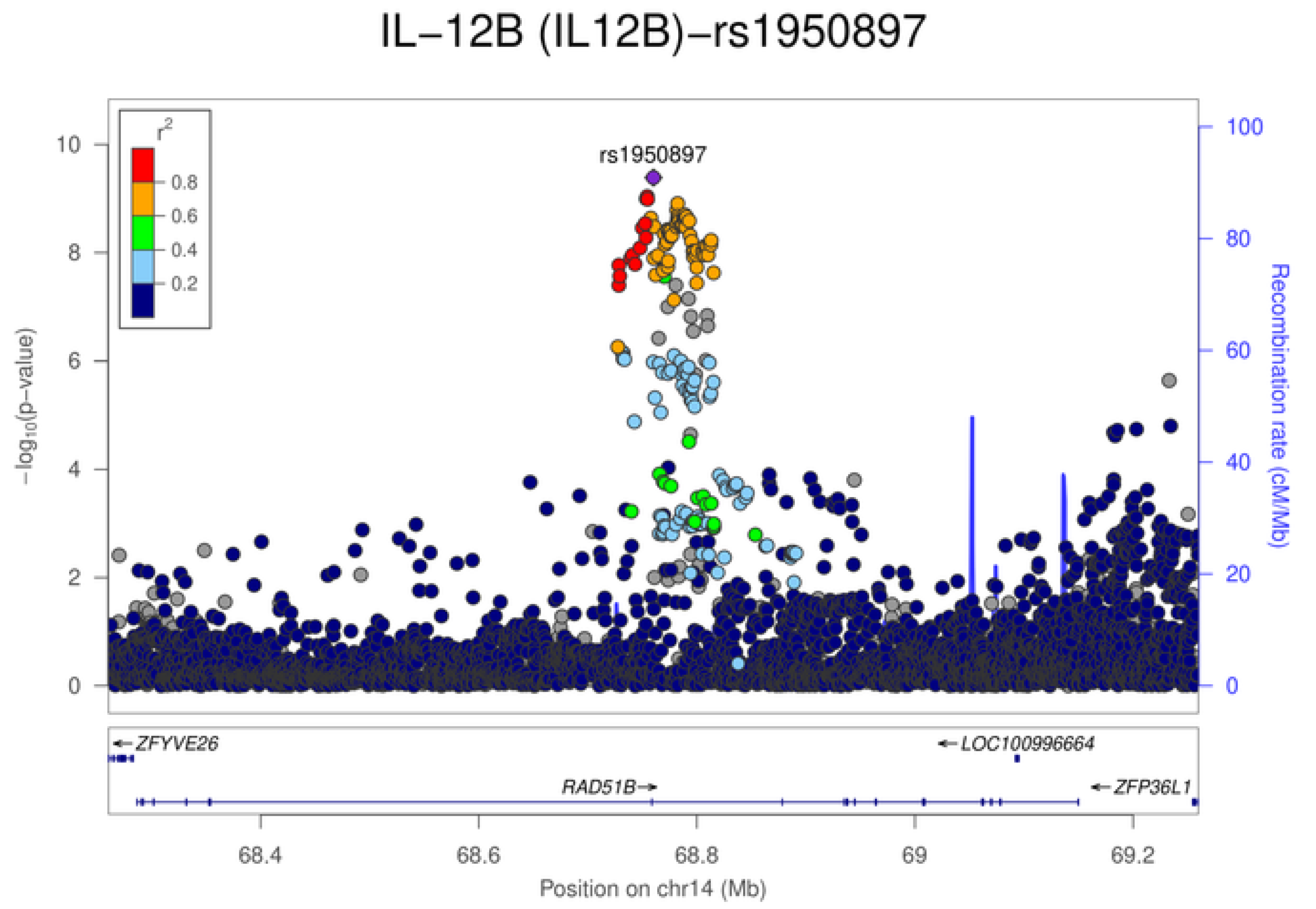
IL−12B (IL12B)−rs1950897
rs1950897
$r^2$
0.8
0.6
0.4
0.2
$-\log_{10}$(p−value)
Recombination rate (cM/Mb)
0
2
4
6
8
10
0
20
40
60
80
100
ZFYVE26
LOC100996664
RAD51B
ZFP36L1
68.4
68.6
68.8
69
69.2
Position on chr14 (Mb)

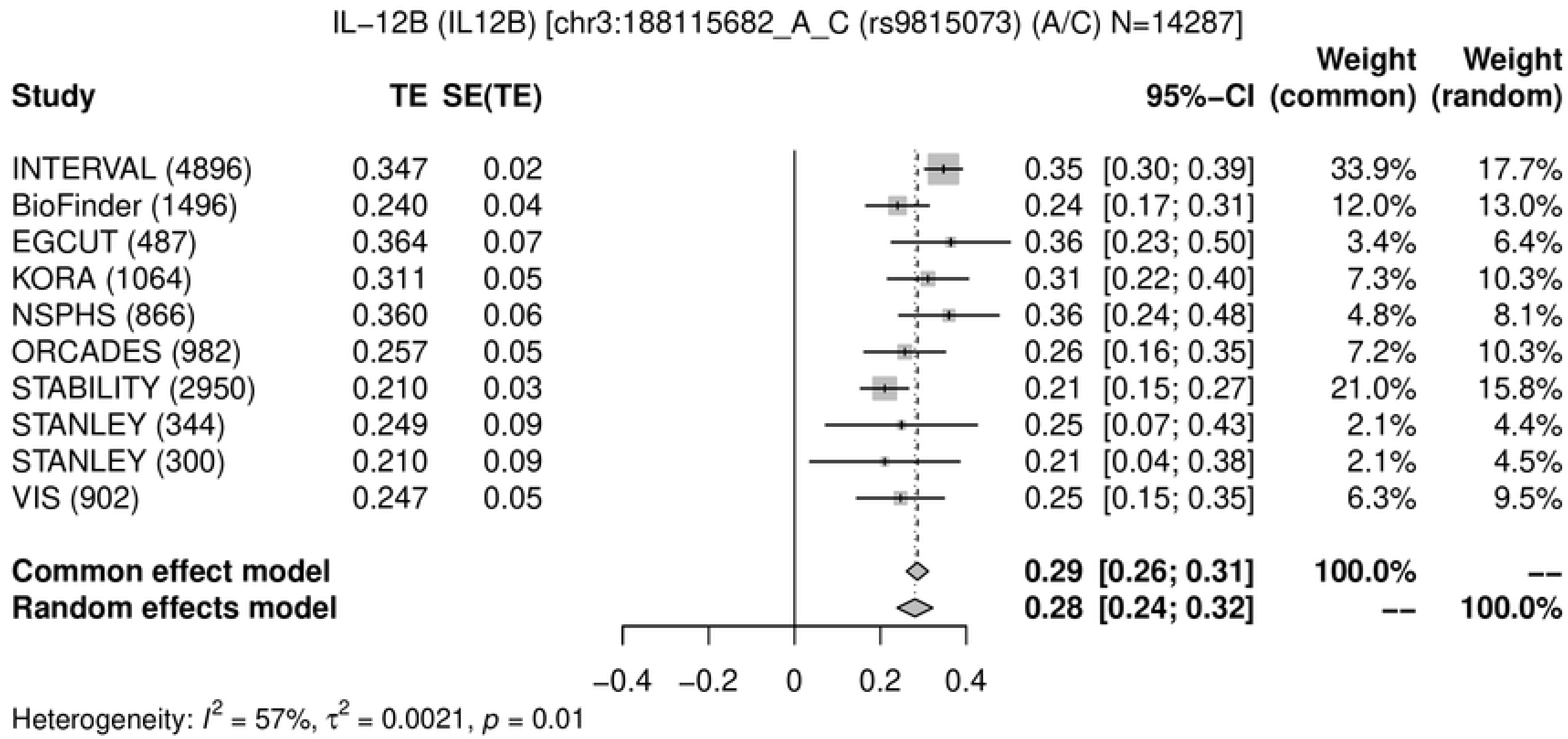
IL−12B (IL12B) [chr3:188115682_A_C (rs9815073) (A/C) N=14287]
Study
TE
SE(TE)
95%−CI
Weight (common)
Weight (random)
INTERVAL (4896) 0.347 0.02 0.35 [0.30; 0.39] 33.9% 17.7%
BioFinder (1496) 0.240 0.04 0.24 [0.17; 0.31] 12.0% 13.0%
EGCUT (487) 0.364 0.07 0.36 [0.23; 0.50] 3.4% 6.4%
KORA (1064) 0.311 0.05 0.31 [0.22; 0.40] 7.3% 10.3%
NSPHS (866) 0.360 0.06 0.36 [0.24; 0.48] 4.8% 8.1%
ORCADES (982) 0.257 0.05 0.26 [0.16; 0.35] 7.2% 10.3%
STABILITY (2950) 0.210 0.03 0.21 [0.15; 0.27] 21.0% 15.8%
STANLEY (344) 0.249 0.09 0.25 [0.07; 0.43] 2.1% 4.4%
STANLEY (300) 0.210 0.09 0.21 [0.04; 0.38] 2.1% 4.5%
VIS (902) 0.247 0.05 0.25 [0.15; 0.35] 6.3% 9.5%
Common effect model 0.29 [0.26; 0.31] 100.0% --
Random effects model 0.28 [0.24; 0.32] -- 100.0%
−0.4 −0.2 0 0.2 0.4
Heterogeneity: I2 = 57%, τ2 = 0.0021, p = 0.01

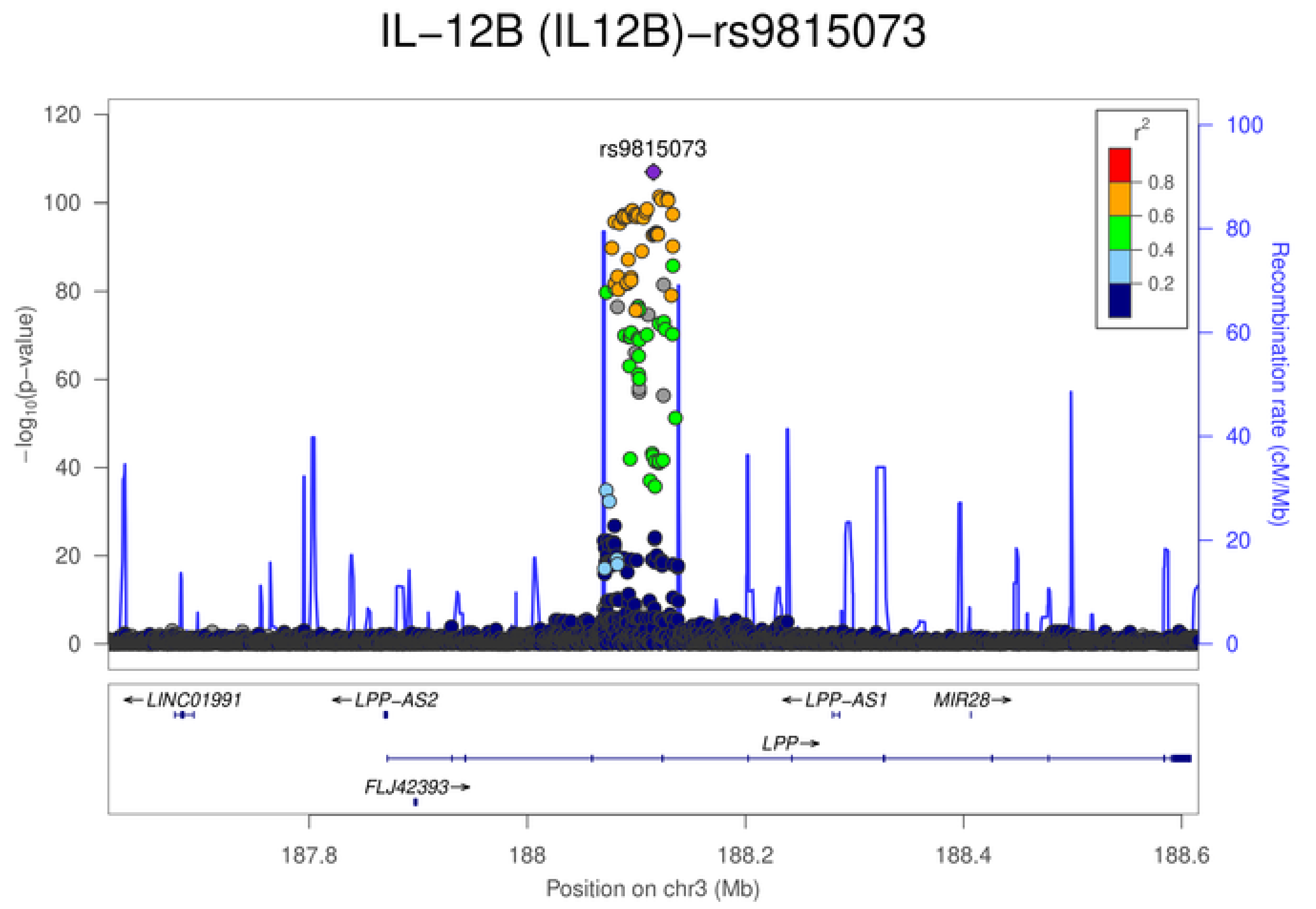
IL−12B (IL12B)−rs9815073
rs9815073
−log10(p−value)
Recombination rate (cM/Mb)
r2
0.8
0.6
0.4
0.2
0
20
40
60
80
100
120
←LINC01991
←LPP-AS2
←LPP-AS1
MIR28→
LPP→
FLJ42393→
187.8
188
188.2
188.4
188.6
Position on chr3 (Mb)

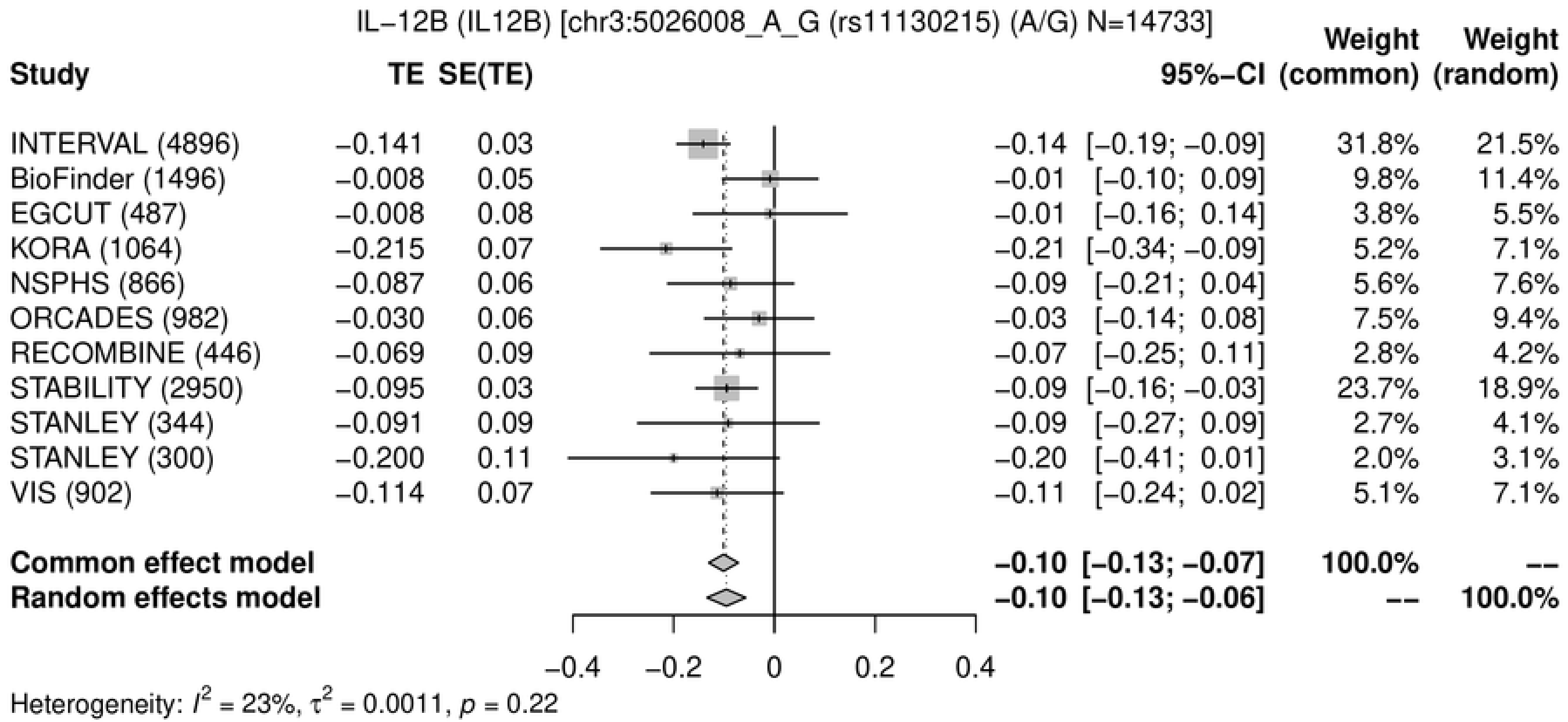
IL−12B (IL12B) [chr3:5026008_A_G (rs11130215) (A/G) N=14733]
Study
TE
SE(TE)
95%−CI
Weight (common)
Weight (random)
INTERVAL (4896) −0.141 0.03 −0.14 [−0.19; −0.09] 31.8% 21.5%
BioFinder (1496) −0.008 0.05 −0.01 [−0.10; 0.09] 9.8% 11.4%
EGCUT (487) −0.008 0.08 −0.01 [−0.16; 0.14] 3.8% 5.5%
KORA (1064) −0.215 0.07 −0.21 [−0.34; −0.09] 5.2% 7.1%
NSPHS (866) −0.087 0.06 −0.09 [−0.21; 0.04] 5.6% 7.6%
ORCADES (982) −0.030 0.06 −0.03 [−0.14; 0.08] 7.5% 9.4%
RECOMBINE (446) −0.069 0.09 −0.07 [−0.25; 0.11] 2.8% 4.2%
STABILITY (2950) −0.095 0.03 −0.09 [−0.16; −0.03] 23.7% 18.9%
STANLEY (344) −0.091 0.09 −0.09 [−0.27; 0.09] 2.7% 4.1%
STANLEY (300) −0.200 0.11 −0.20 [−0.41; 0.01] 2.0% 3.1%
VIS (902) −0.114 0.07 −0.11 [−0.24; 0.02] 5.1% 7.1%
Common effect model −0.10 [−0.13; −0.07] 100.0% --
Random effects model −0.10 [−0.13; −0.06] -- 100.0%
−0.4 −0.2 0 0.2 0.4
Heterogeneity: $I^2$ = 23%, $\tau^2$ = 0.0011, $p$ = 0.22

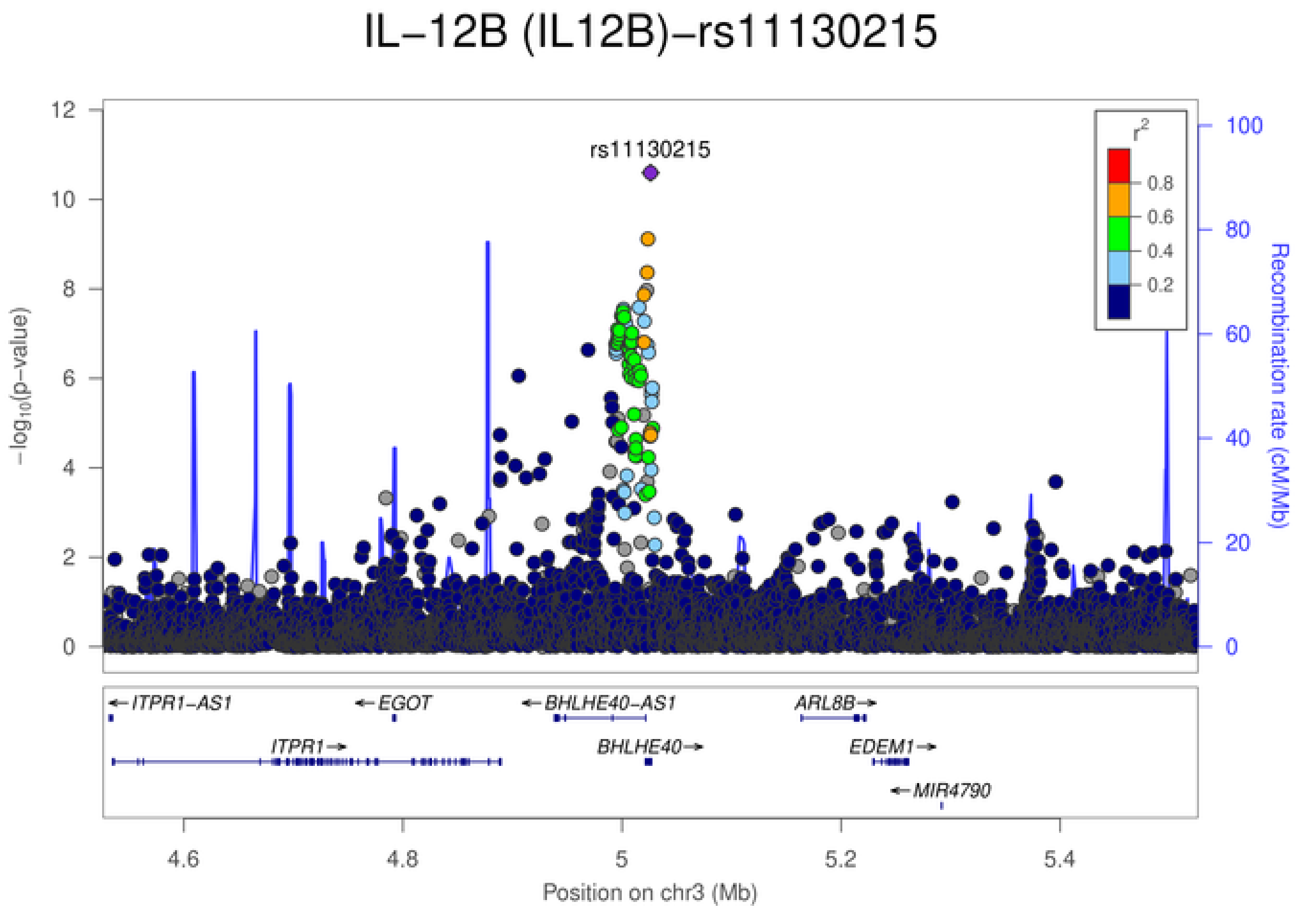
IL−12B (IL12B)−rs11130215
rs11130215
$-\log_{10}$(p-value)
Recombination rate (cM/Mb)
$r^2$
0.8
0.6
0.4
0.2
←ITPR1-AS1
←EGOT
←BHLHE40-AS1
ARL8B→
ITPR1→
BHLHE40→
EDEM1→
←MIR4790
4.6 4.8 5 5.2 5.4
Position on chr3 (Mb)

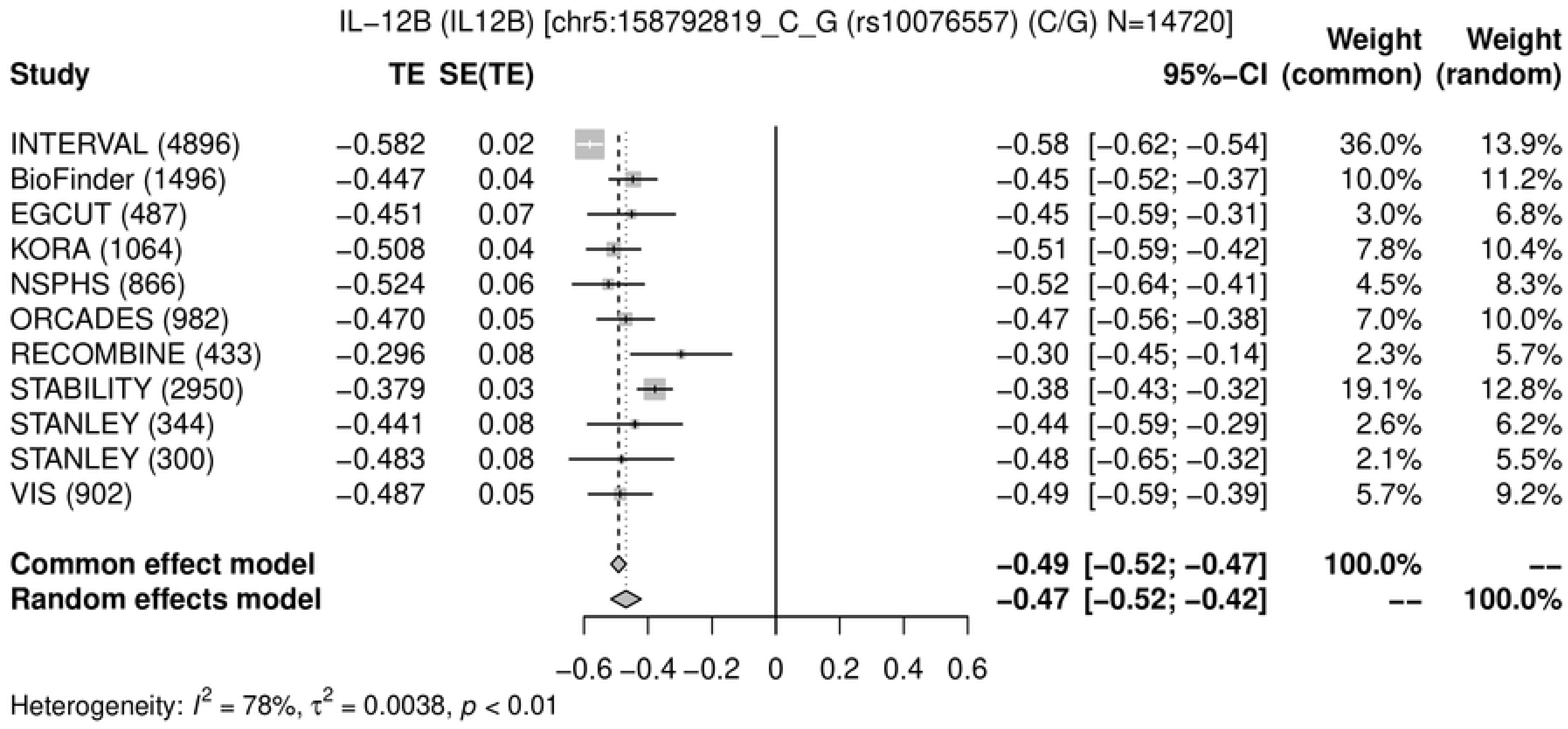
IL−12B (IL12B) [chr5:158792819_C_G (rs10076557) (C/G) N=14720]
Study
TE
SE(TE)
95%−CI
Weight (common)
Weight (random)
INTERVAL (4896) −0.582 0.02 −0.58 [−0.62; −0.54] 36.0% 13.9%
BioFinder (1496) −0.447 0.04 −0.45 [−0.52; −0.37] 10.0% 11.2%
EGCUT (487) −0.451 0.07 −0.45 [−0.59; −0.31] 3.0% 6.8%
KORA (1064) −0.508 0.04 −0.51 [−0.59; −0.42] 7.8% 10.4%
NSPHS (866) −0.524 0.06 −0.52 [−0.64; −0.41] 4.5% 8.3%
ORCADES (982) −0.470 0.05 −0.47 [−0.56; −0.38] 7.0% 10.0%
RECOMBINE (433) −0.296 0.08 −0.30 [−0.45; −0.14] 2.3% 5.7%
STABILITY (2950) −0.379 0.03 −0.38 [−0.43; −0.32] 19.1% 12.8%
STANLEY (344) −0.441 0.08 −0.44 [−0.59; −0.29] 2.6% 6.2%
STANLEY (300) −0.483 0.08 −0.48 [−0.65; −0.32] 2.1% 5.5%
VIS (902) −0.487 0.05 −0.49 [−0.59; −0.39] 5.7% 9.2%
Common effect model −0.49 [−0.52; −0.47] 100.0% --
Random effects model −0.47 [−0.52; −0.42] -- 100.0%
−0.6 −0.4 −0.2 0 0.2 0.4 0.6
Heterogeneity: $I^2$ = 78%, $\tau^2$ = 0.0038, $p$ < 0.01

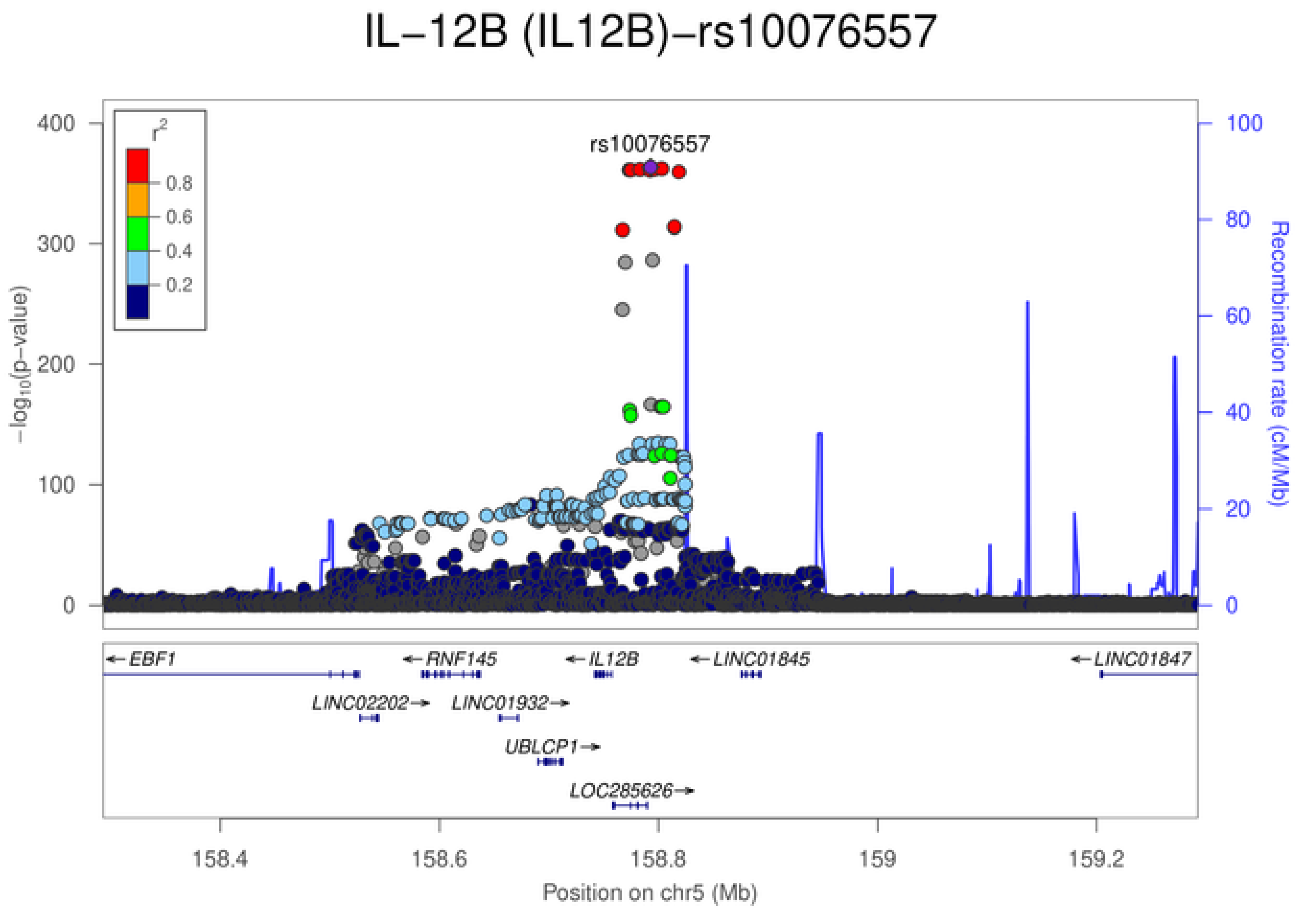
IL−12B (IL12B)−rs10076557
rs10076557
$r^2$
0.8
0.6
0.4
0.2
$-\log_{10}$(p−value)
Recombination rate (cM/Mb)
EBF1
RNF145
IL12B
LINC01845
LINC01847
LINC02202
LINC01932
UBLCP1
LOC285626
158.4
158.6
158.8
159
159.2
Position on chr5 (Mb)

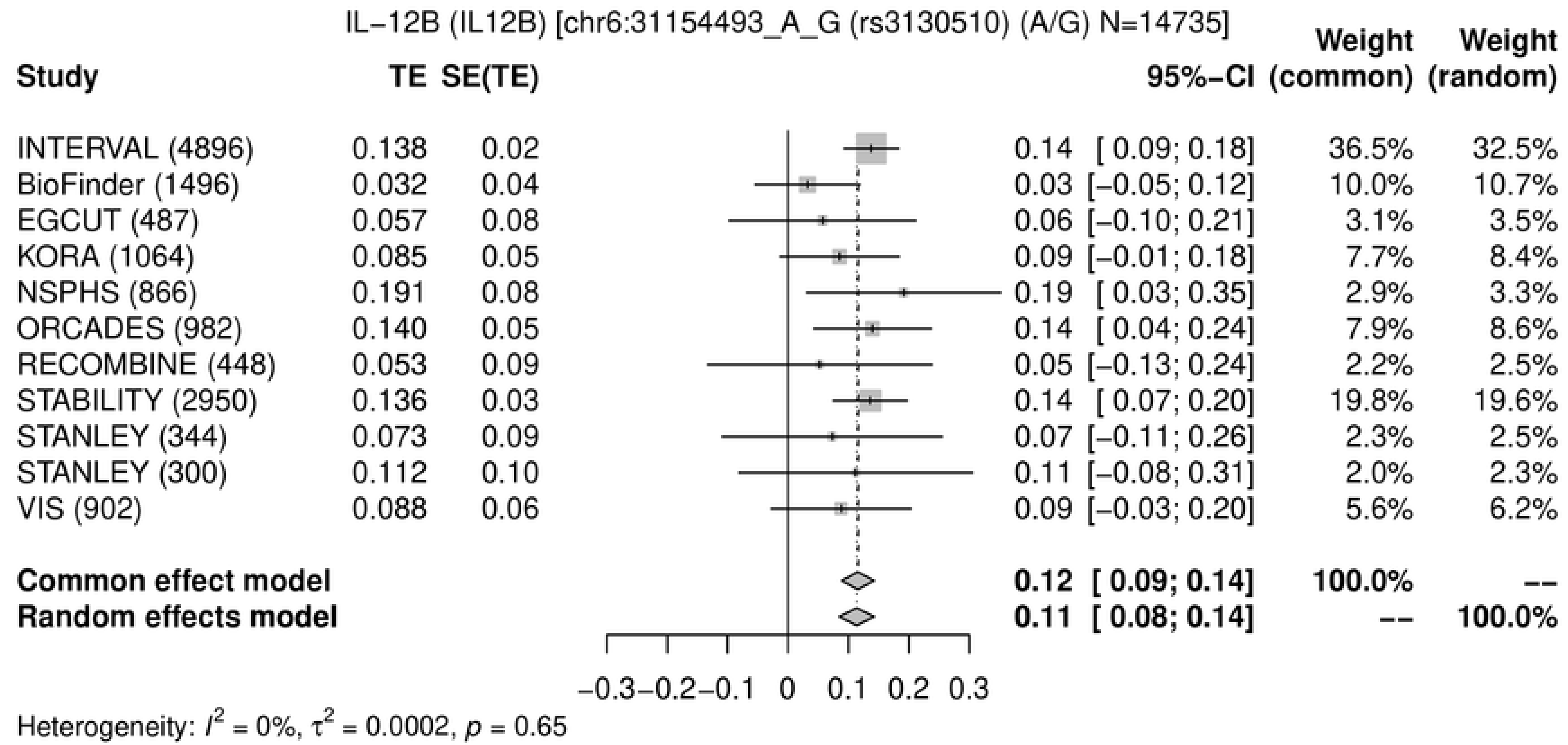
IL−12B (IL12B) [chr6:31154493_A_G (rs3130510) (A/G) N=14735]
Study | TE | SE(TE) | 95%−CI | Weight (common) | Weight (random)
INTERVAL (4896) | 0.138 | 0.02 | 0.14 [ 0.09; 0.18] | 36.5% | 32.5%
BioFinder (1496) | 0.032 | 0.04 | 0.03 [−0.05; 0.12] | 10.0% | 10.7%
EGCUT (487) | 0.057 | 0.08 | 0.06 [−0.10; 0.21] | 3.1% | 3.5%
KORA (1064) | 0.085 | 0.05 | 0.09 [−0.01; 0.18] | 7.7% | 8.4%
NSPHS (866) | 0.191 | 0.08 | 0.19 [ 0.03; 0.35] | 2.9% | 3.3%
ORCADES (982) | 0.140 | 0.05 | 0.14 [ 0.04; 0.24] | 7.9% | 8.6%
RECOMBINE (448) | 0.053 | 0.09 | 0.05 [−0.13; 0.24] | 2.2% | 2.5%
STABILITY (2950) | 0.136 | 0.03 | 0.14 [ 0.07; 0.20] | 19.8% | 19.6%
STANLEY (344) | 0.073 | 0.09 | 0.07 [−0.11; 0.26] | 2.3% | 2.5%
STANLEY (300) | 0.112 | 0.10 | 0.11 [−0.08; 0.31] | 2.0% | 2.3%
VIS (902) | 0.088 | 0.06 | 0.09 [−0.03; 0.20] | 5.6% | 6.2%
Common effect model | 0.12 [ 0.09; 0.14] | 100.0% | --
Random effects model | 0.11 [ 0.08; 0.14] | -- | 100.0%
−0.3 −0.2 −0.1 0 0.1 0.2 0.3
Heterogeneity: $I^2$ = 0%, $\tau^2$ = 0.0002, $p$ = 0.65

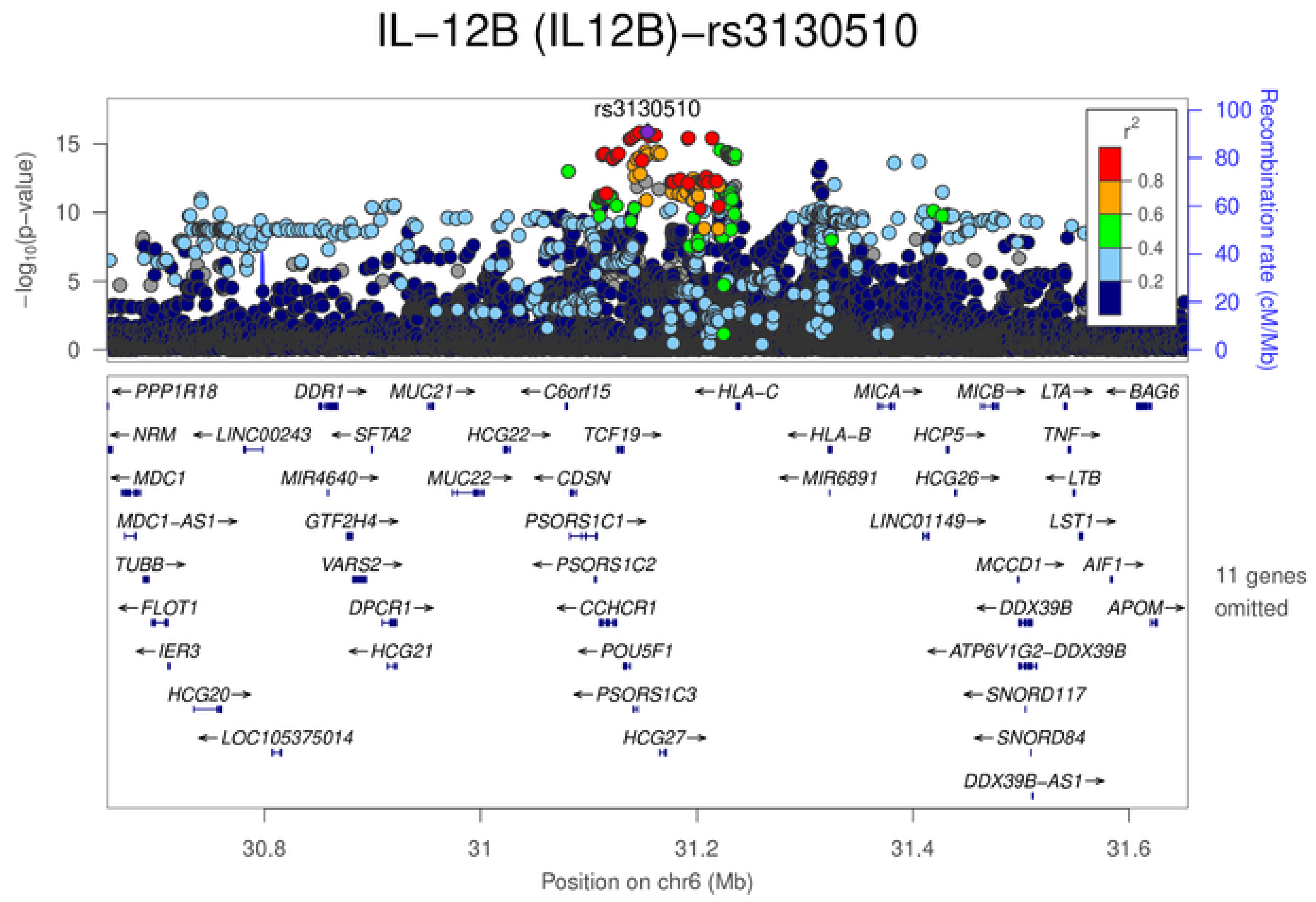
IL−12B (IL12B)−rs3130510
rs3130510
$-\log_{10}$(p−value)
Recombination rate (cM/Mb)
$r^2$
0.8
0.6
0.4
0.2
PPP1R18
DDR1
MUC21
C6orf15
HLA−C
MICA
MICB
LTA
BAG6
NRM
LINC00243
SFTA2
HCG22
TCF19
HLA−B
HCP5
TNF
MDC1
MIR4640
MUC22
CDSN
MIR6891
HCG26
LTB
MDC1−AS1
GTF2H4
PSORS1C1
LINC01149
LST1
TUBB
VARS2
PSORS1C2
MCCD1
AIF1
FLOT1
DPCR1
CCHCR1
DDX39B
APOM
IER3
HCG21
POU5F1
ATP6V1G2−DDX39B
HCG20
PSORS1C3
SNORD117
LOC105375014
HCG27
SNORD84
DDX39B−AS1
11 genes omitted
30.8
31
31.2
31.4
31.6
Position on chr6 (Mb)

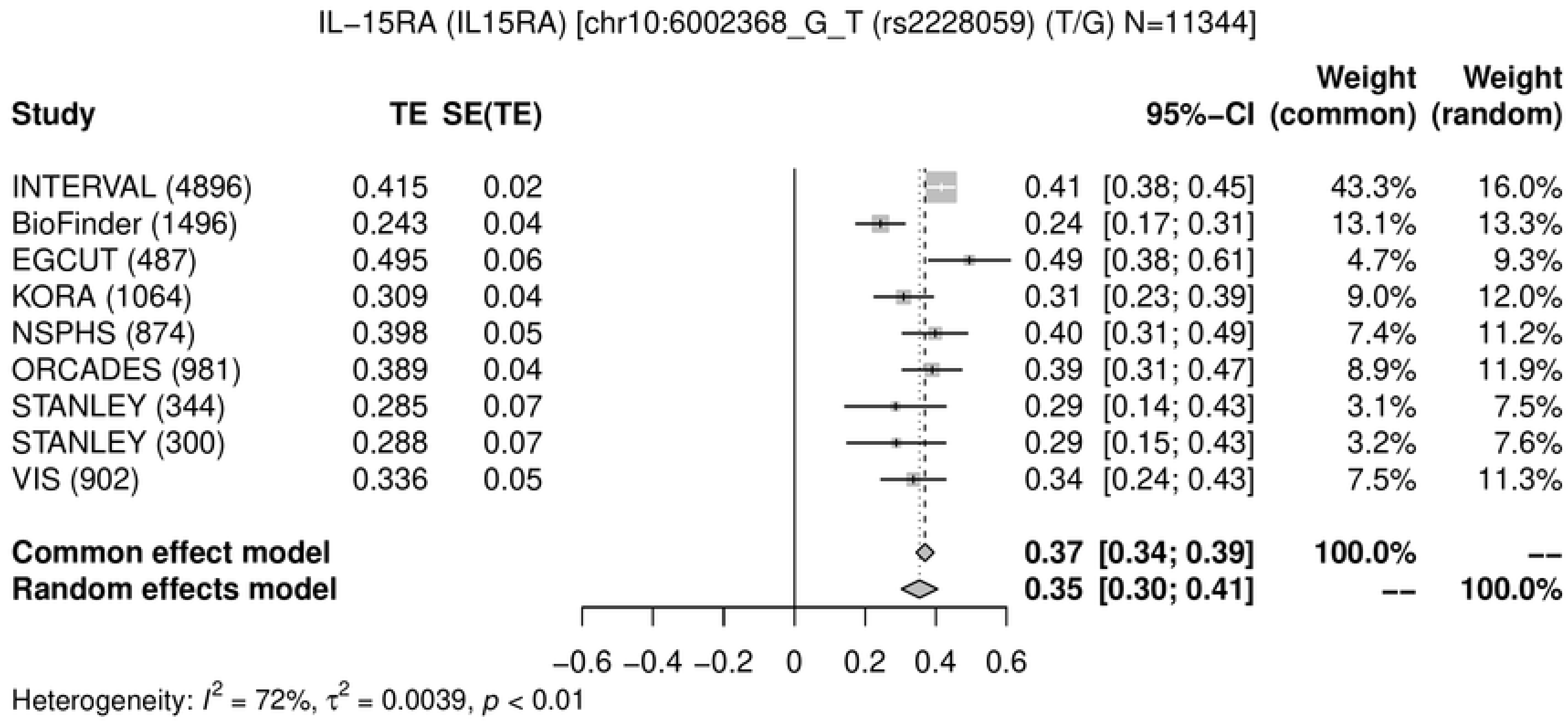
IL−15RA (IL15RA) [chr10:6002368_G_T (rs2228059) (T/G) N=11344]
Study
TE
SE(TE)
95%−CI
Weight (common)
Weight (random)
INTERVAL (4896) 0.415 0.02 0.41 [0.38; 0.45] 43.3% 16.0%
BioFinder (1496) 0.243 0.04 0.24 [0.17; 0.31] 13.1% 13.3%
EGCUT (487) 0.495 0.06 0.49 [0.38; 0.61] 4.7% 9.3%
KORA (1064) 0.309 0.04 0.31 [0.23; 0.39] 9.0% 12.0%
NSPHS (874) 0.398 0.05 0.40 [0.31; 0.49] 7.4% 11.2%
ORCADES (981) 0.389 0.04 0.39 [0.31; 0.47] 8.9% 11.9%
STANLEY (344) 0.285 0.07 0.29 [0.14; 0.43] 3.1% 7.5%
STANLEY (300) 0.288 0.07 0.29 [0.15; 0.43] 3.2% 7.6%
VIS (902) 0.336 0.05 0.34 [0.24; 0.43] 7.5% 11.3%
Common effect model 0.37 [0.34; 0.39] 100.0% --
Random effects model 0.35 [0.30; 0.41] -- 100.0%
−0.6 −0.4 −0.2 0 0.2 0.4 0.6
Heterogeneity: $I^2$ = 72%, $\tau^2$ = 0.0039, $p$ < 0.01

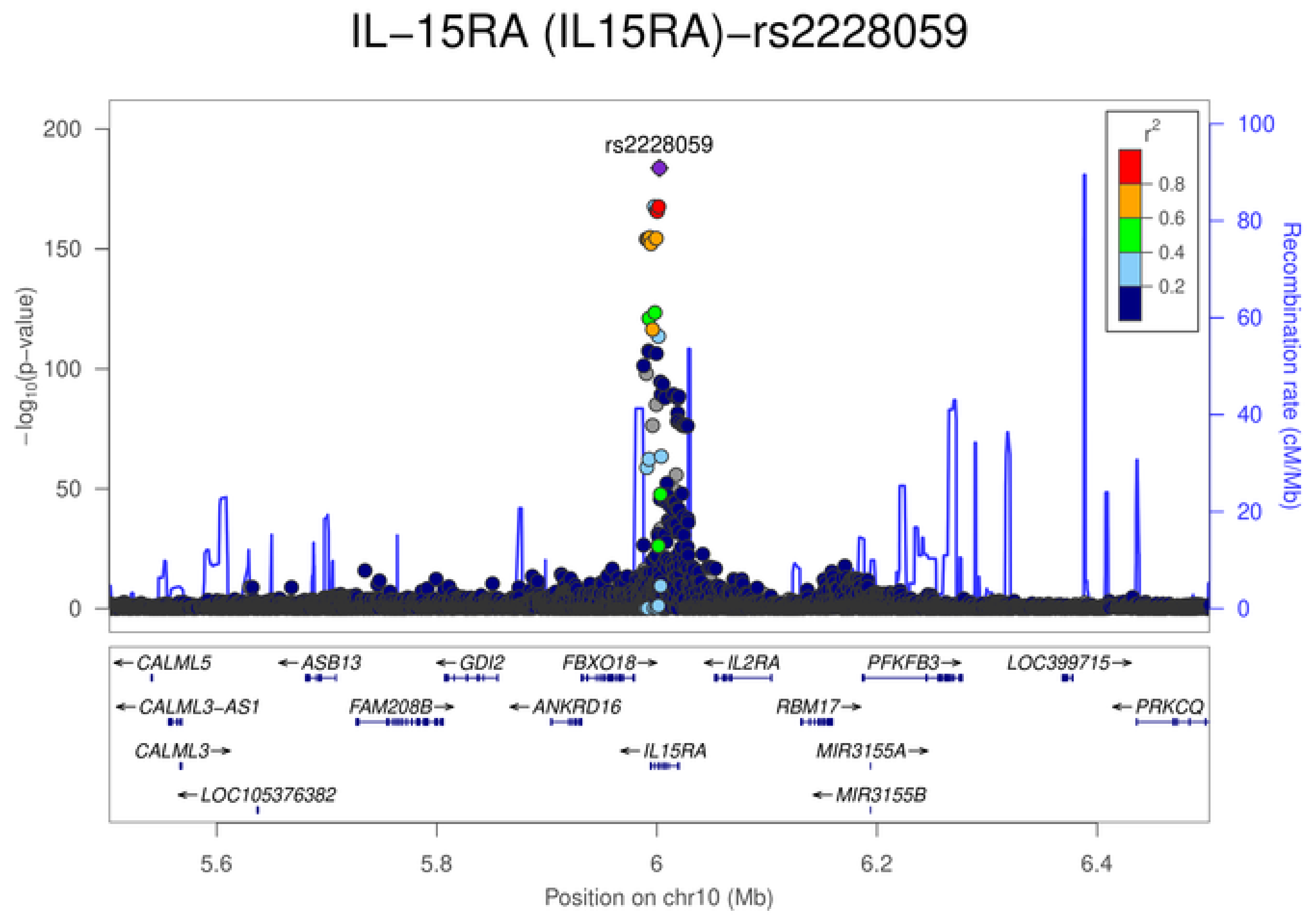
IL−15RA (IL15RA)−rs2228059
rs2228059
$-\log_{10}$(p−value)
Recombination rate (cM/Mb)
$r^2$
0.8
0.6
0.4
0.2
CALML5
ASB13
GDI2
FBXO18
IL2RA
PFKFB3
LOC399715
CALML3-AS1
FAM208B
ANKRD16
RBM17
PRKCQ
CALML3
IL15RA
MIR3155A
LOC105376382
MIR3155B
5.6
5.8
6
6.2
6.4
Position on chr10 (Mb)

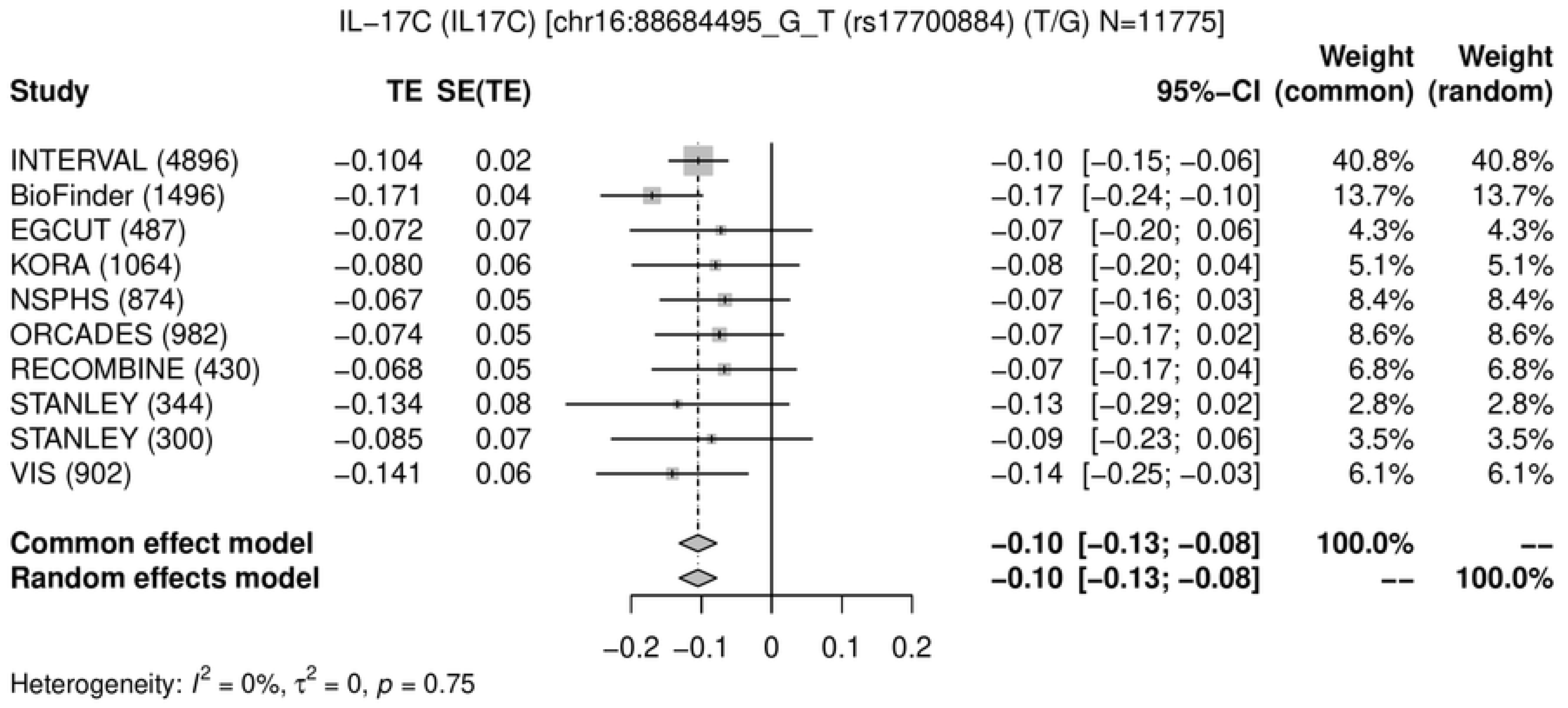
IL−17C (IL17C) [chr16:88684495_G_T (rs17700884) (T/G) N=11775]
Study
TE
SE(TE)
95%−CI
Weight (common)
Weight (random)
INTERVAL (4896) −0.104 0.02 −0.10 [−0.15; −0.06] 40.8% 40.8%
BioFinder (1496) −0.171 0.04 −0.17 [−0.24; −0.10] 13.7% 13.7%
EGCUT (487) −0.072 0.07 −0.07 [−0.20; 0.06] 4.3% 4.3%
KORA (1064) −0.080 0.06 −0.08 [−0.20; 0.04] 5.1% 5.1%
NSPHS (874) −0.067 0.05 −0.07 [−0.16; 0.03] 8.4% 8.4%
ORCADES (982) −0.074 0.05 −0.07 [−0.17; 0.02] 8.6% 8.6%
RECOMBINE (430) −0.068 0.05 −0.07 [−0.17; 0.04] 6.8% 6.8%
STANLEY (344) −0.134 0.08 −0.13 [−0.29; 0.02] 2.8% 2.8%
STANLEY (300) −0.085 0.07 −0.09 [−0.23; 0.06] 3.5% 3.5%
VIS (902) −0.141 0.06 −0.14 [−0.25; −0.03] 6.1% 6.1%
Common effect model −0.10 [−0.13; −0.08] 100.0% --
Random effects model −0.10 [−0.13; −0.08] -- 100.0%
−0.2 −0.1 0 0.1 0.2
Heterogeneity: $I^2 = 0\%$, $\tau^2 = 0$, $p = 0.75$

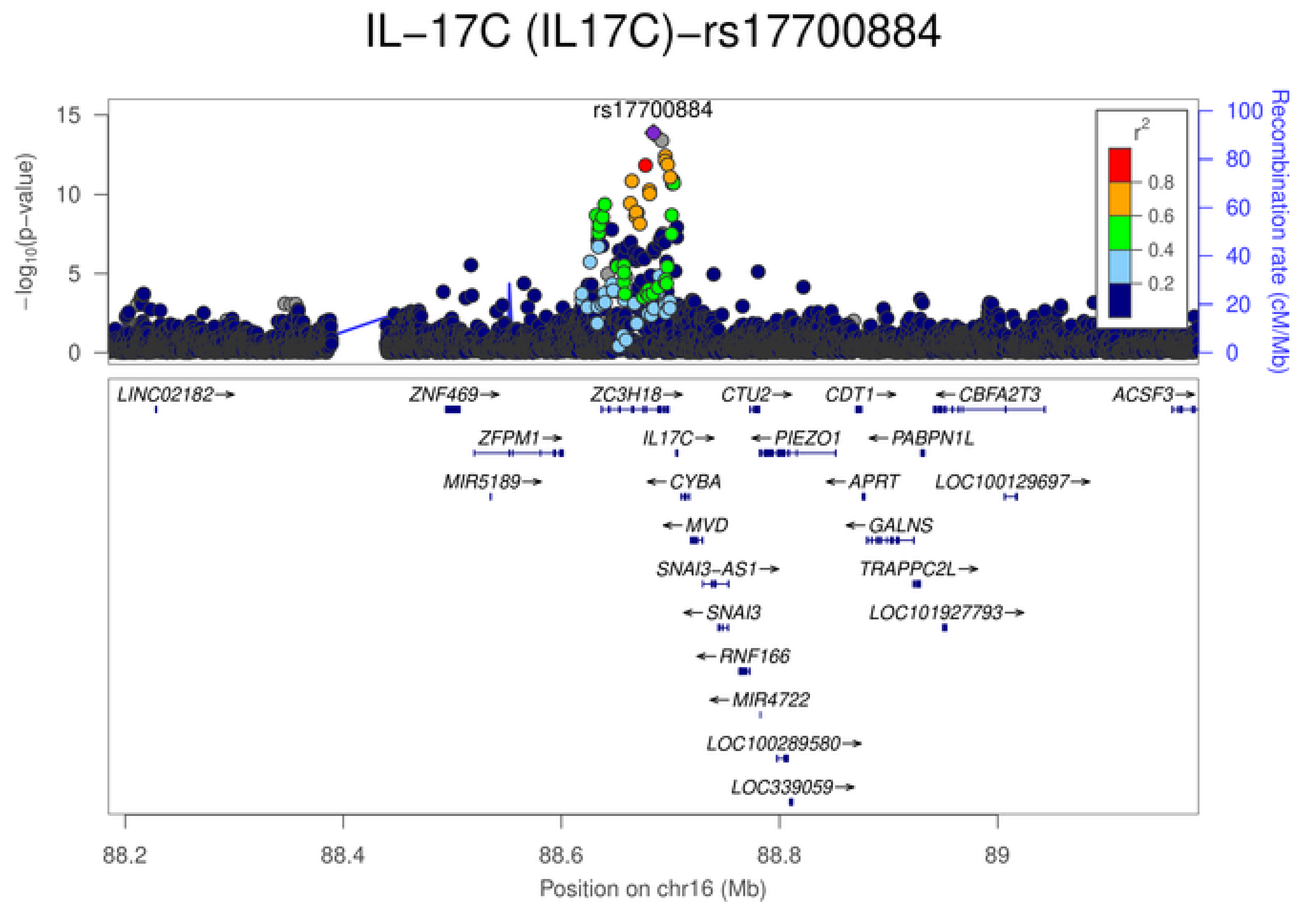
IL−17C (IL17C)−rs17700884
rs17700884
$-\log_{10}(p\text{-value})$
Recombination rate (cM/Mb)
$r^2$
0.8
0.6
0.4
0.2
LINC02182
ZNF469
ZC3H18
CTU2
CDT1
CBFA2T3
ACSF3
ZFPM1
IL17C
PIEZO1
PABPN1L
MIR5189
CYBA
APRT
LOC100129697
MVD
GALNS
SNAI3−AS1
TRAPPC2L
SNAI3
LOC101927793
RNF166
MIR4722
LOC100289580
LOC339059
88.2 88.4 88.6 88.8 89
Position on chr16 (Mb)

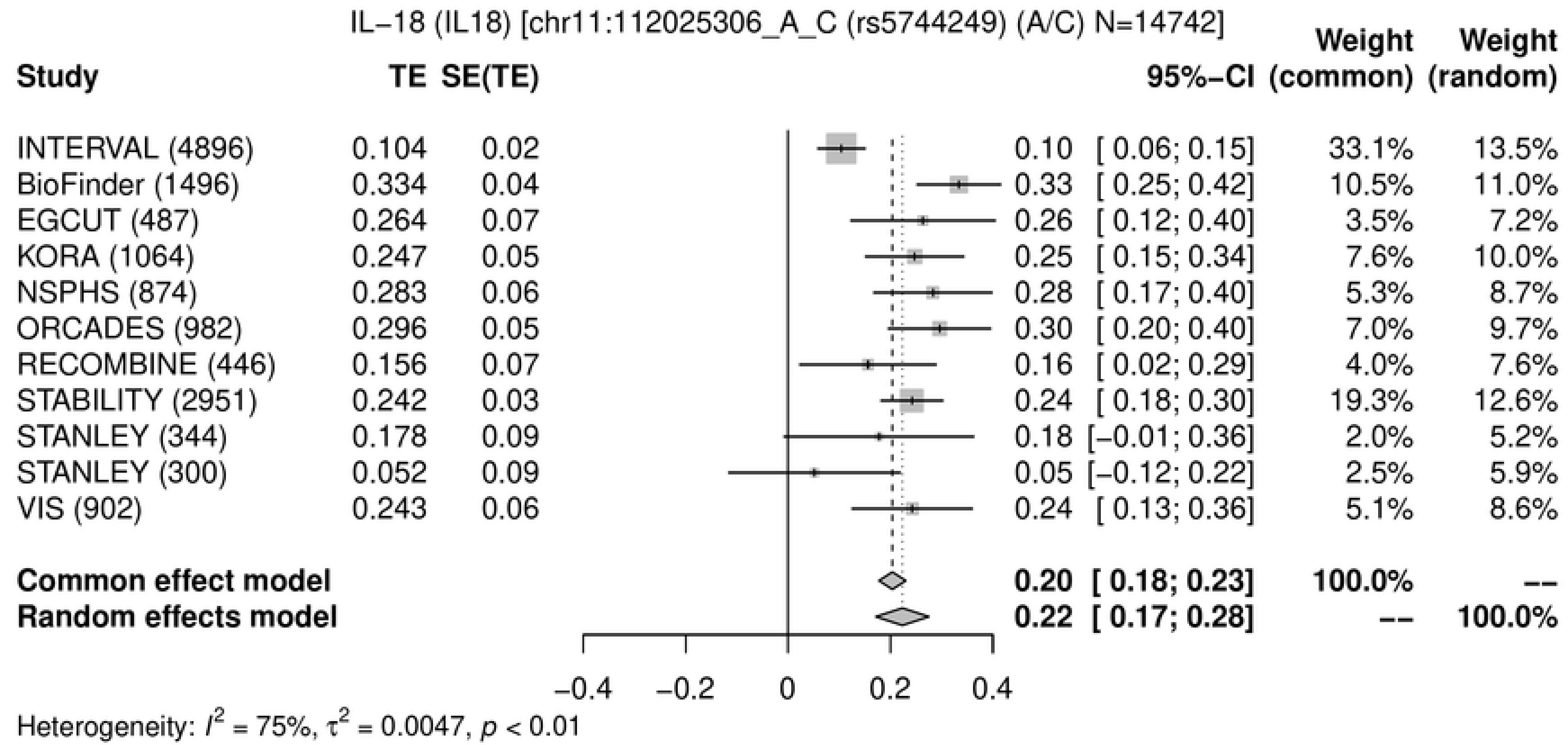
IL–18 (IL18) [chr11:112025306_A_C (rs5744249) (A/C) N=14742]
Study TE SE(TE) 95%–CI Weight (common) Weight (random)
INTERVAL (4896) 0.104 0.02 0.10 [ 0.06; 0.15] 33.1% 13.5%
BioFinder (1496) 0.334 0.04 0.33 [ 0.25; 0.42] 10.5% 11.0%
EGCUT (487) 0.264 0.07 0.26 [ 0.12; 0.40] 3.5% 7.2%
KORA (1064) 0.247 0.05 0.25 [ 0.15; 0.34] 7.6% 10.0%
NSPHS (874) 0.283 0.06 0.28 [ 0.17; 0.40] 5.3% 8.7%
ORCADES (982) 0.296 0.05 0.30 [ 0.20; 0.40] 7.0% 9.7%
RECOMBINE (446) 0.156 0.07 0.16 [ 0.02; 0.29] 4.0% 7.6%
STABILITY (2951) 0.242 0.03 0.24 [ 0.18; 0.30] 19.3% 12.6%
STANLEY (344) 0.178 0.09 0.18 [−0.01; 0.36] 2.0% 5.2%
STANLEY (300) 0.052 0.09 0.05 [−0.12; 0.22] 2.5% 5.9%
VIS (902) 0.243 0.06 0.24 [ 0.13; 0.36] 5.1% 8.6%
Common effect model 0.20 [ 0.18; 0.23] 100.0% --
Random effects model 0.22 [ 0.17; 0.28] -- 100.0%
−0.4 −0.2 0 0.2 0.4
Heterogeneity: $I^2$ = 75%, $\tau^2$ = 0.0047, $p$ < 0.01
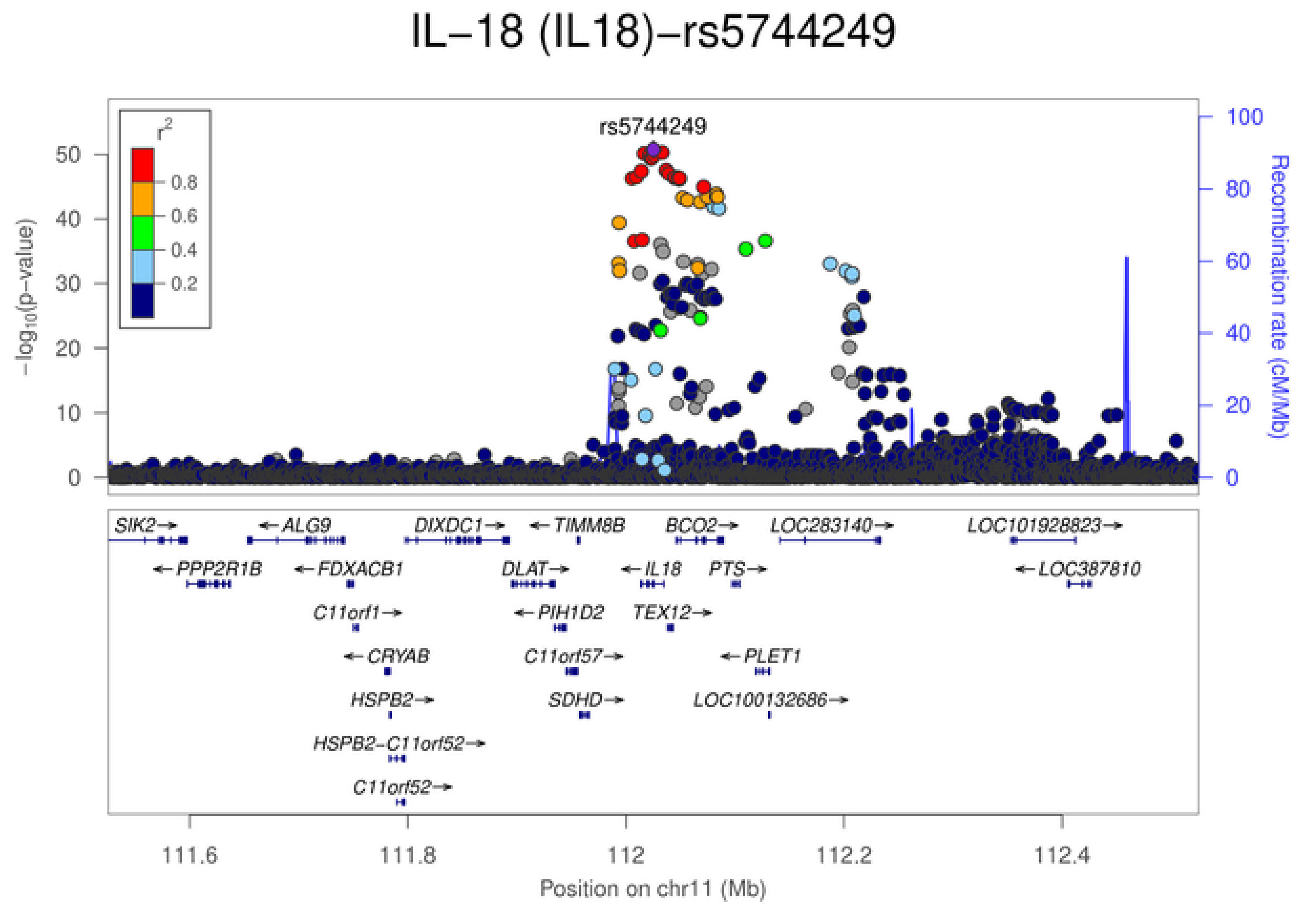
IL–18 (IL18)–rs5744249
rs5744249
$r^2$
0.8
0.6
0.4
0.2
$-\log_{10}$(p–value)
Recombination rate (cM/Mb)
SIK2
ALG9
DIXDC1
TIMM8B
BCO2
LOC283140
LOC101928823
PPP2R1B
FDXACB1
DLAT
IL18
PTS
LOC387810
C11orf1
PIH1D2
TEX12
CRYAB
C11orf57
PLET1
HSPB2
SDHD
LOC100132686
HSPB2–C11orf52
C11orf52
111.6 111.8 112 112.2 112.4
Position on chr11 (Mb)

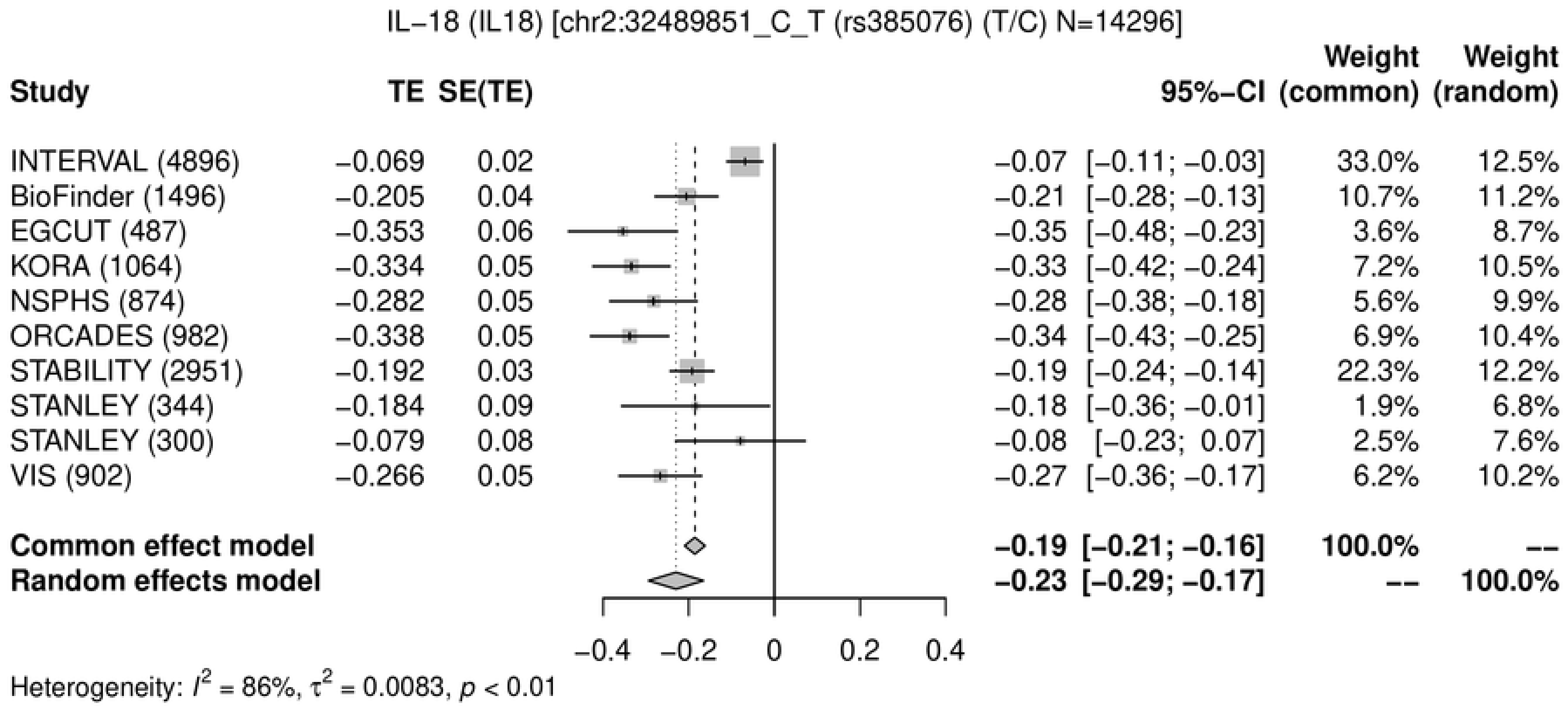
IL−18 (IL18) [chr2:32489851_C_T (rs385076) (T/C) N=14296]
Study TE SE(TE) 95%−CI Weight (common) Weight (random)
INTERVAL (4896) −0.069 0.02 −0.07 [−0.11; −0.03] 33.0% 12.5%
BioFinder (1496) −0.205 0.04 −0.21 [−0.28; −0.13] 10.7% 11.2%
EGCUT (487) −0.353 0.06 −0.35 [−0.48; −0.23] 3.6% 8.7%
KORA (1064) −0.334 0.05 −0.33 [−0.42; −0.24] 7.2% 10.5%
NSPHS (874) −0.282 0.05 −0.28 [−0.38; −0.18] 5.6% 9.9%
ORCADES (982) −0.338 0.05 −0.34 [−0.43; −0.25] 6.9% 10.4%
STABILITY (2951) −0.192 0.03 −0.19 [−0.24; −0.14] 22.3% 12.2%
STANLEY (344) −0.184 0.09 −0.18 [−0.36; −0.01] 1.9% 6.8%
STANLEY (300) −0.079 0.08 −0.08 [−0.23; 0.07] 2.5% 7.6%
VIS (902) −0.266 0.05 −0.27 [−0.36; −0.17] 6.2% 10.2%
Common effect model −0.19 [−0.21; −0.16] 100.0% --
Random effects model −0.23 [−0.29; −0.17] -- 100.0%
−0.4 −0.2 0 0.2 0.4
Heterogeneity: $I^2$ = 86%, $\tau^2$ = 0.0083, $p$ < 0.01

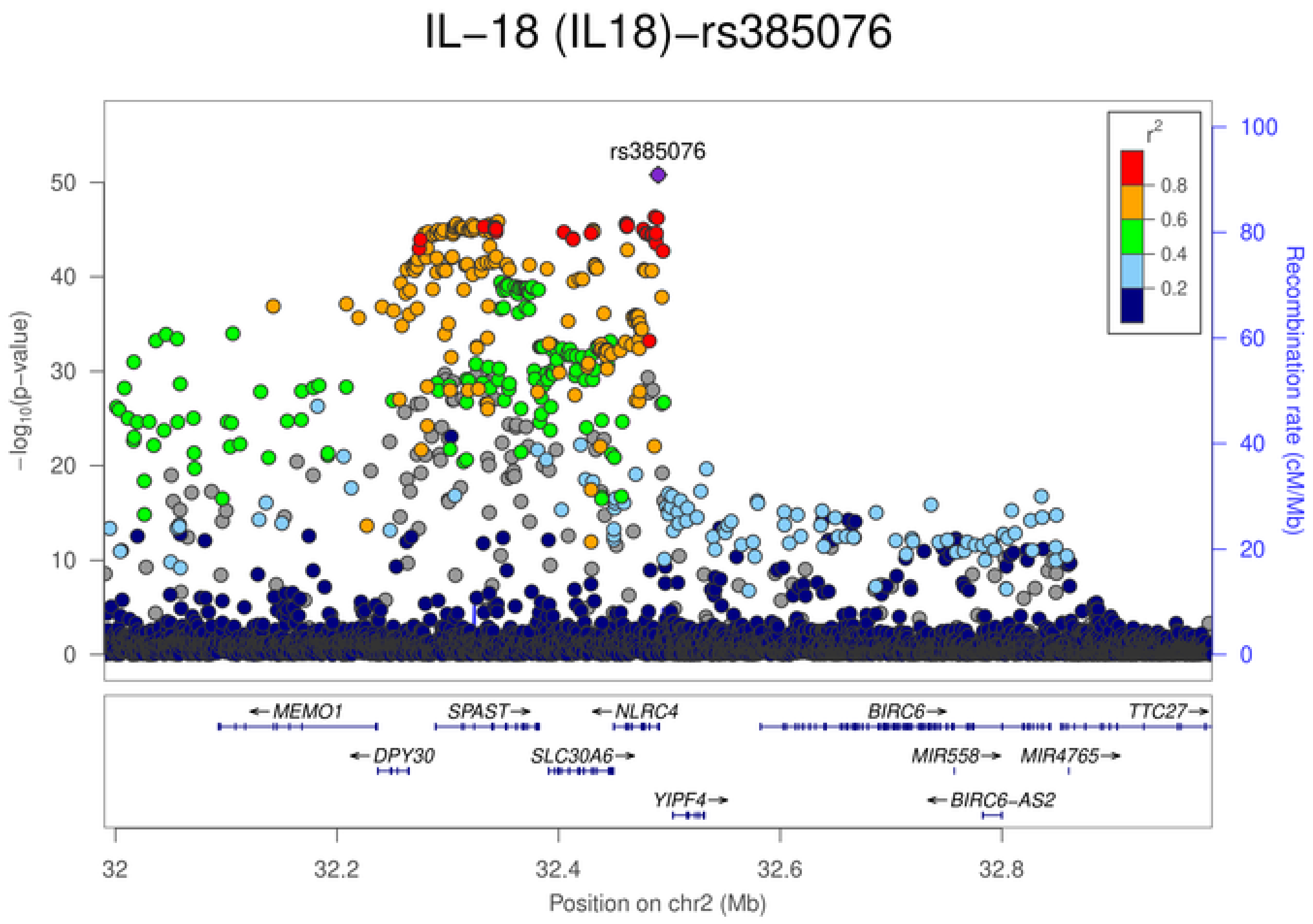
IL−18 (IL18)−rs385076
rs385076
$r^2$
0.8
0.6
0.4
0.2
−log10(p−value)
Recombination rate (cM/Mb)
0 10 20 30 40 50
0 20 40 60 80 100
MEMO1
SPAST
NLRC4
BIRC6
TTC27
DPY30
SLC30A6
MIR558
MIR4765
YIPF4
BIRC6−AS2
32 32.2 32.4 32.6 32.8
Position on chr2 (Mb)

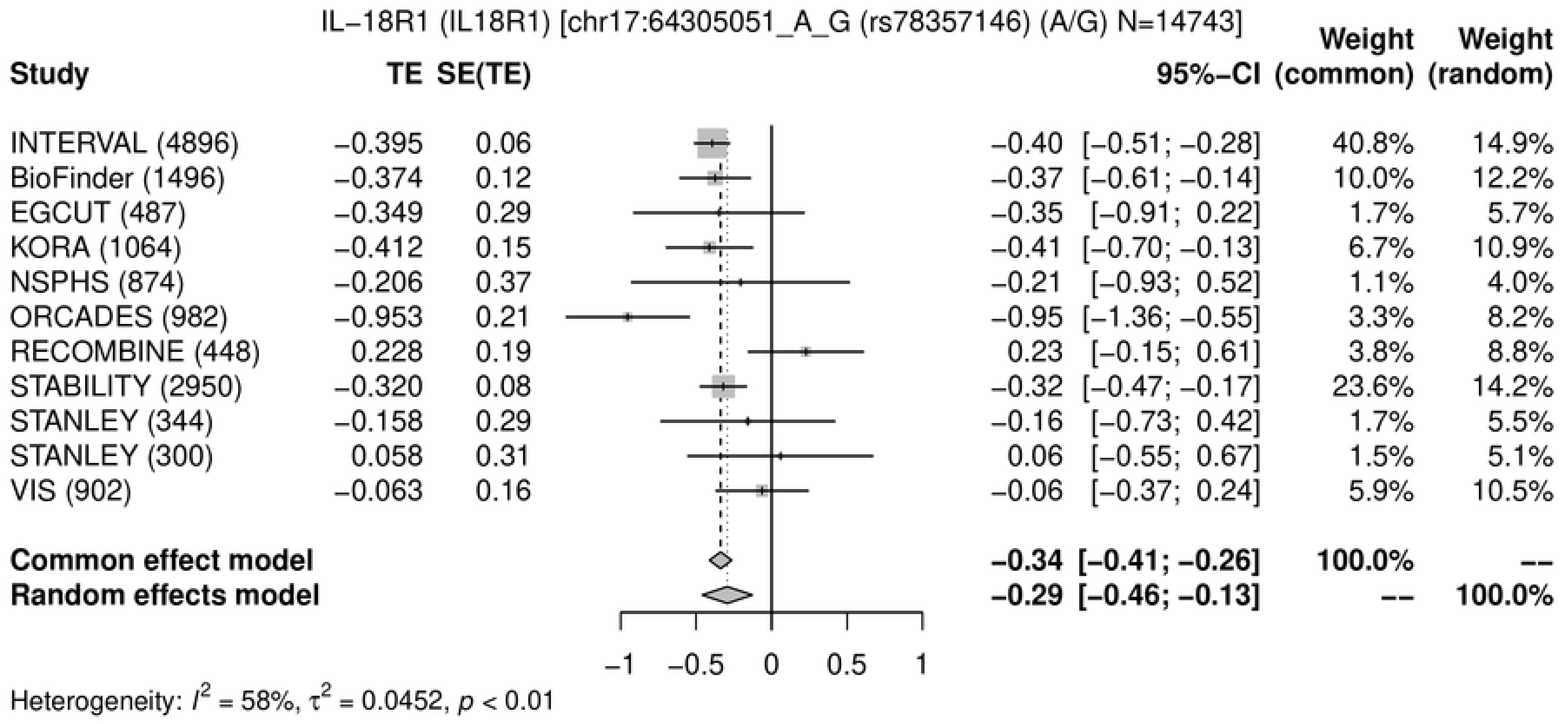
IL−18R1 (IL18R1) [chr17:64305051_A_G (rs78357146) (A/G) N=14743]
Study
TE
SE(TE)
95%−CI
Weight (common)
Weight (random)
INTERVAL (4896) −0.395 0.06 −0.40 [−0.51; −0.28] 40.8% 14.9%
BioFinder (1496) −0.374 0.12 −0.37 [−0.61; −0.14] 10.0% 12.2%
EGCUT (487) −0.349 0.29 −0.35 [−0.91; 0.22] 1.7% 5.7%
KORA (1064) −0.412 0.15 −0.41 [−0.70; −0.13] 6.7% 10.9%
NSPHS (874) −0.206 0.37 −0.21 [−0.93; 0.52] 1.1% 4.0%
ORCADES (982) −0.953 0.21 −0.95 [−1.36; −0.55] 3.3% 8.2%
RECOMBINE (448) 0.228 0.19 0.23 [−0.15; 0.61] 3.8% 8.8%
STABILITY (2950) −0.320 0.08 −0.32 [−0.47; −0.17] 23.6% 14.2%
STANLEY (344) −0.158 0.29 −0.16 [−0.73; 0.42] 1.7% 5.5%
STANLEY (300) 0.058 0.31 0.06 [−0.55; 0.67] 1.5% 5.1%
VIS (902) −0.063 0.16 −0.06 [−0.37; 0.24] 5.9% 10.5%
Common effect model −0.34 [−0.41; −0.26] 100.0% --
Random effects model −0.29 [−0.46; −0.13] -- 100.0%
−1 −0.5 0 0.5 1
Heterogeneity: $I^2$ = 58%, $\tau^2$ = 0.0452, $p$ < 0.01

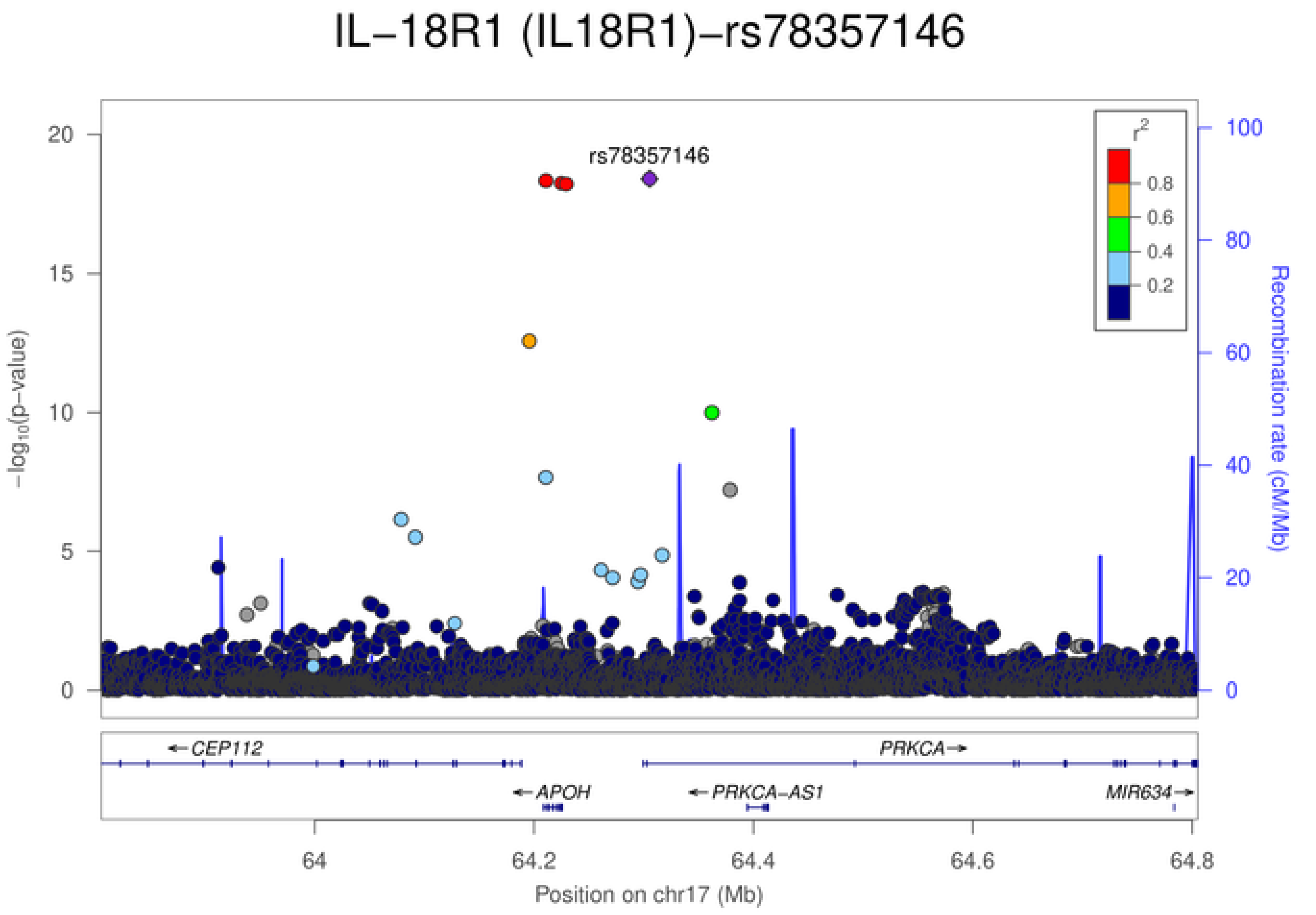
IL−18R1 (IL18R1)−rs78357146
rs78357146
$-\log_{10}$(p−value)
0 5 10 15 20
Recombination rate (cM/Mb)
0 20 40 60 80 100
$r^2$
0.8 0.6 0.4 0.2
CEP112
PRKCA
APOH
PRKCA−AS1
MIR634
64 64.2 64.4 64.6 64.8
Position on chr17 (Mb)

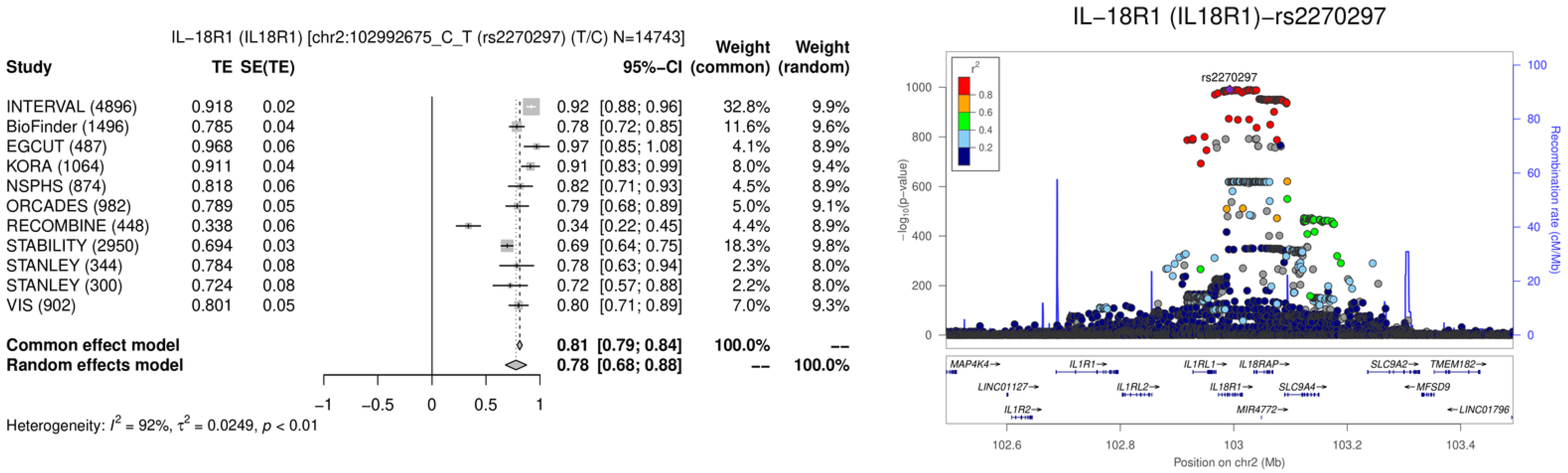
IL−18R1 (IL18R1) [chr2:102992675_C_T (rs2270297) (T/C) N=14743]
Study
TE
SE(TE)
95%−CI
Weight (common)
Weight (random)
INTERVAL (4896) 0.918 0.02 0.92 [0.88; 0.96] 32.8% 9.9%
BioFinder (1496) 0.785 0.04 0.78 [0.72; 0.85] 11.6% 9.6%
EGCUT (487) 0.968 0.06 0.97 [0.85; 1.08] 4.1% 8.9%
KORA (1064) 0.911 0.04 0.91 [0.83; 0.99] 8.0% 9.4%
NSPHS (874) 0.818 0.06 0.82 [0.71; 0.93] 4.5% 8.9%
ORCADES (982) 0.789 0.05 0.79 [0.68; 0.89] 5.0% 9.1%
RECOMBINE (448) 0.338 0.06 0.34 [0.22; 0.45] 4.4% 8.9%
STABILITY (2950) 0.694 0.03 0.69 [0.64; 0.75] 18.3% 9.8%
STANLEY (344) 0.784 0.08 0.78 [0.63; 0.94] 2.3% 8.0%
STANLEY (300) 0.724 0.08 0.72 [0.57; 0.88] 2.2% 8.0%
VIS (902) 0.801 0.05 0.80 [0.71; 0.89] 7.0% 9.3%
Common effect model 0.81 [0.79; 0.84] 100.0% --
Random effects model 0.78 [0.68; 0.88] -- 100.0%
−1 −0.5 0 0.5 1
Heterogeneity: $I^2 = 92\%$, $\tau^2 = 0.0249$, $p < 0.01$
IL−18R1 (IL18R1)−rs2270297
rs2270297
$r^2$
0.8
0.6
0.4
0.2
$-\log_{10}$(p−value)
Recombination rate (cM/Mb)
MAP4K4
IL1R1
IL1RL1
IL18RAP
SLC9A2
TMEM182
LINC01127
IL1RL2
IL18R1
SLC9A4
MFSD9
IL1R2
MIR4772
LINC01796
102.6 102.8 103 103.2 103.4
Position on chr2 (Mb)

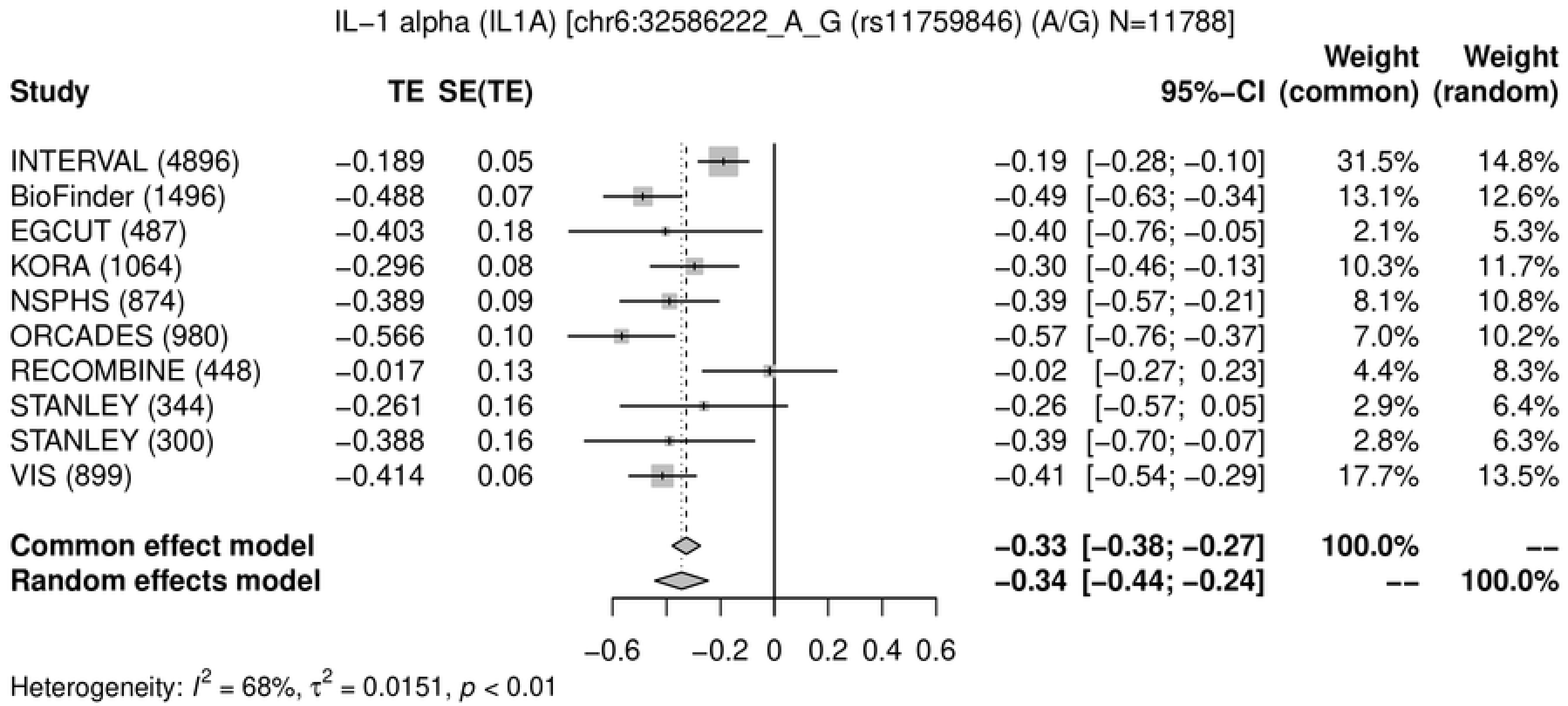
IL−1 alpha (IL1A) [chr6:32586222_A_G (rs11759846) (A/G) N=11788]
Study
TE
SE(TE)
95%−CI
Weight (common)
Weight (random)
INTERVAL (4896) −0.189 0.05 −0.19 [−0.28; −0.10] 31.5% 14.8%
BioFinder (1496) −0.488 0.07 −0.49 [−0.63; −0.34] 13.1% 12.6%
EGCUT (487) −0.403 0.18 −0.40 [−0.76; −0.05] 2.1% 5.3%
KORA (1064) −0.296 0.08 −0.30 [−0.46; −0.13] 10.3% 11.7%
NSPHS (874) −0.389 0.09 −0.39 [−0.57; −0.21] 8.1% 10.8%
ORCADES (980) −0.566 0.10 −0.57 [−0.76; −0.37] 7.0% 10.2%
RECOMBINE (448) −0.017 0.13 −0.02 [−0.27; 0.23] 4.4% 8.3%
STANLEY (344) −0.261 0.16 −0.26 [−0.57; 0.05] 2.9% 6.4%
STANLEY (300) −0.388 0.16 −0.39 [−0.70; −0.07] 2.8% 6.3%
VIS (899) −0.414 0.06 −0.41 [−0.54; −0.29] 17.7% 13.5%
Common effect model −0.33 [−0.38; −0.27] 100.0% --
Random effects model −0.34 [−0.44; −0.24] -- 100.0%
−0.6 −0.2 0 0.2 0.4 0.6
Heterogeneity: $I^2$ = 68%, $\tau^2$ = 0.0151, $p$ < 0.01

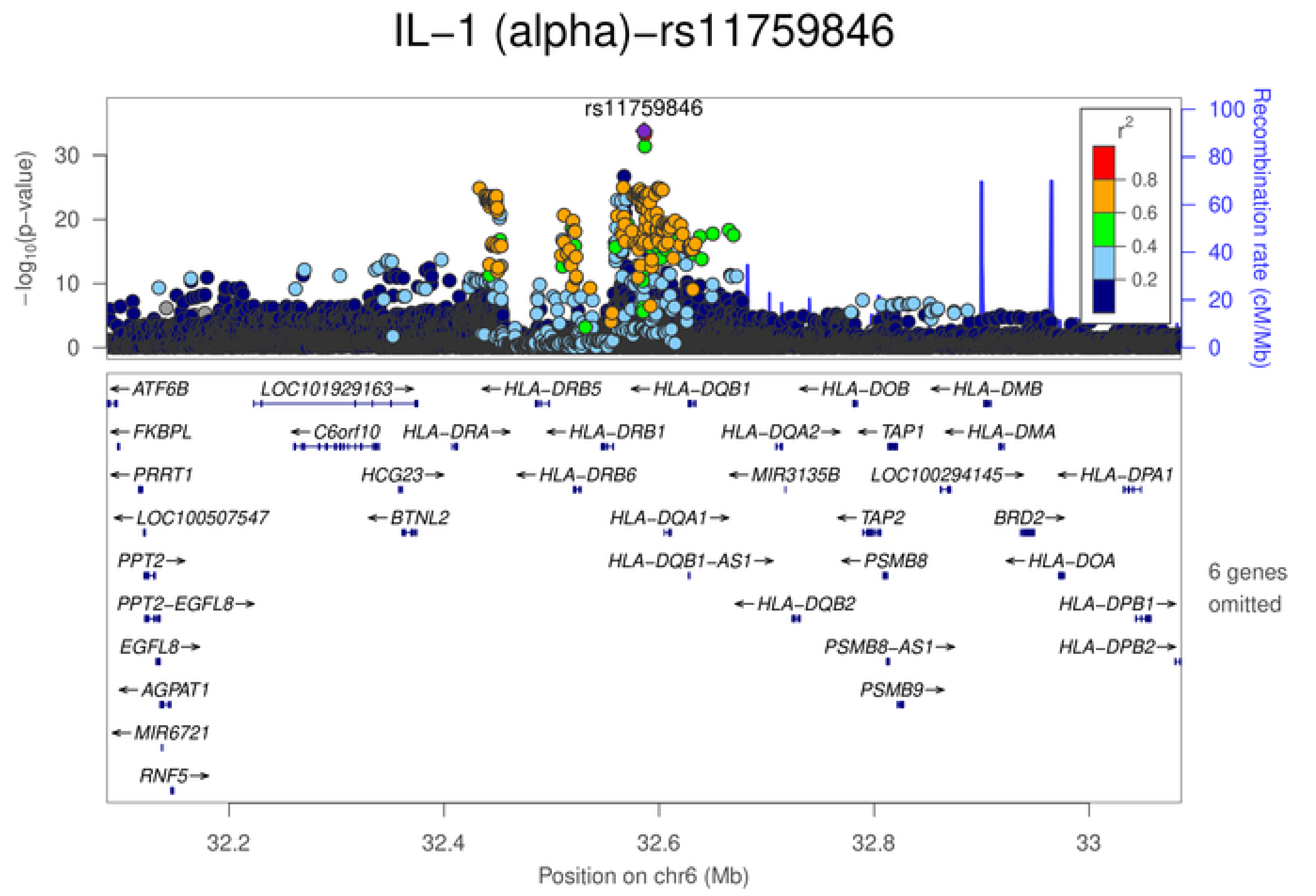
IL−1 (alpha)−rs11759846
rs11759846
$-\log_{10}$(p−value)
Recombination rate (cM/Mb)
$r^2$
0.8
0.6
0.4
0.2
ATF6B
LOC101929163
HLA−DRB5
HLA−DQB1
HLA−DOB
HLA−DMB
FKBPL
C6orf10
HLA−DRA
HLA−DRB1
HLA−DQA2
TAP1
HLA−DMA
PRRT1
HCG23
HLA−DRB6
MIR3135B
LOC100294145
HLA−DPA1
LOC100507547
BTNL2
HLA−DQA1
TAP2
BRD2
PPT2
HLA−DQB1−AS1
PSMB8
HLA−DOA
PPT2−EGFL8
HLA−DQB2
HLA−DPB1
EGFL8
PSMB8−AS1
HLA−DPB2
AGPAT1
PSMB9
MIR6721
RNF5
6 genes omitted
32.2
32.4
32.6
32.8
33
Position on chr6 (Mb)

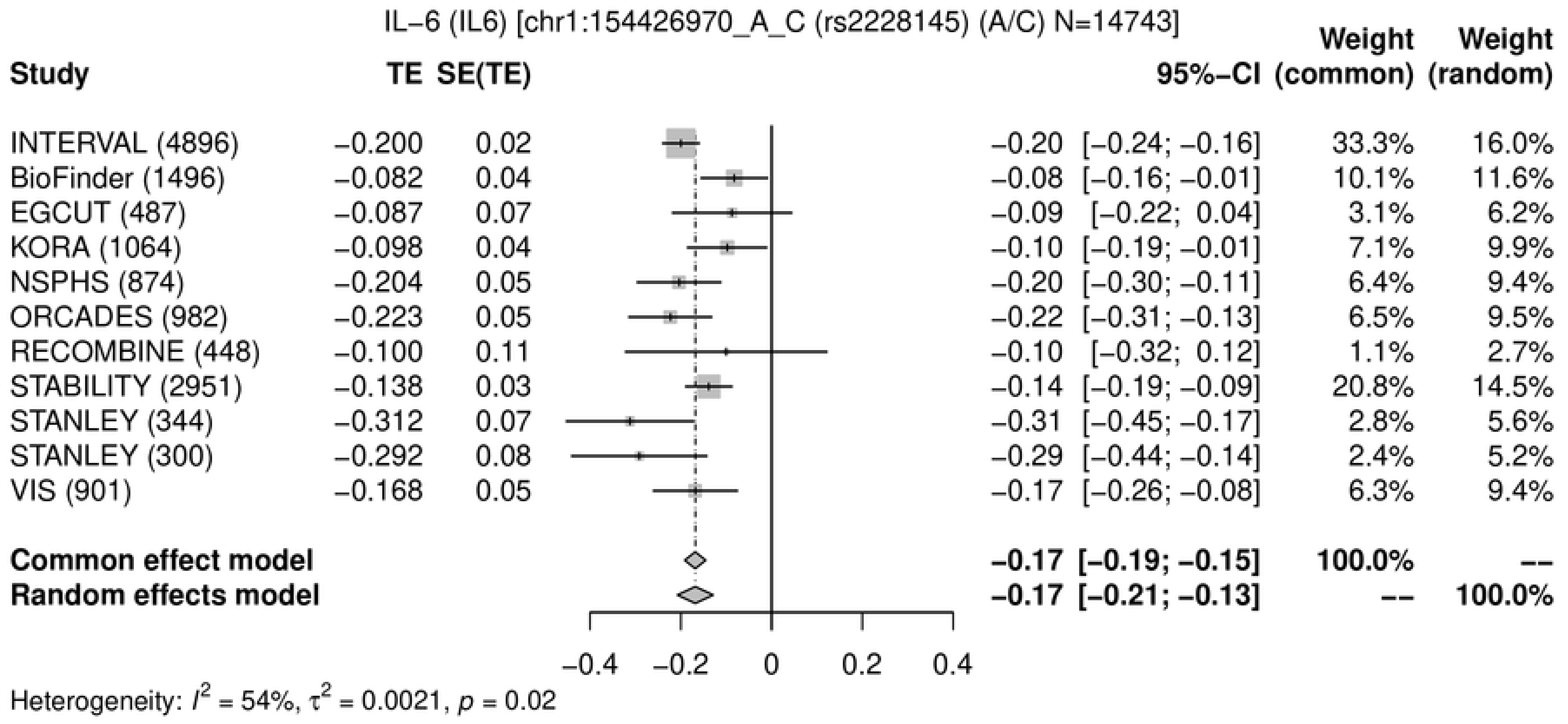
IL−6 (IL6) [chr1:154426970_A_C (rs2228145) (A/C) N=14743]
Study
TE
SE(TE)
95%−CI
Weight (common)
Weight (random)
INTERVAL (4896) −0.200 0.02 −0.20 [−0.24; −0.16] 33.3% 16.0%
BioFinder (1496) −0.082 0.04 −0.08 [−0.16; −0.01] 10.1% 11.6%
EGCUT (487) −0.087 0.07 −0.09 [−0.22; 0.04] 3.1% 6.2%
KORA (1064) −0.098 0.04 −0.10 [−0.19; −0.01] 7.1% 9.9%
NSPHS (874) −0.204 0.05 −0.20 [−0.30; −0.11] 6.4% 9.4%
ORCADES (982) −0.223 0.05 −0.22 [−0.31; −0.13] 6.5% 9.5%
RECOMBINE (448) −0.100 0.11 −0.10 [−0.32; 0.12] 1.1% 2.7%
STABILITY (2951) −0.138 0.03 −0.14 [−0.19; −0.09] 20.8% 14.5%
STANLEY (344) −0.312 0.07 −0.31 [−0.45; −0.17] 2.8% 5.6%
STANLEY (300) −0.292 0.08 −0.29 [−0.44; −0.14] 2.4% 5.2%
VIS (901) −0.168 0.05 −0.17 [−0.26; −0.08] 6.3% 9.4%
Common effect model −0.17 [−0.19; −0.15] 100.0% --
Random effects model −0.17 [−0.21; −0.13] -- 100.0%
−0.4 −0.2 0 0.2 0.4
Heterogeneity: $I^2$ = 54%, $\tau^2$ = 0.0021, $p$ = 0.02

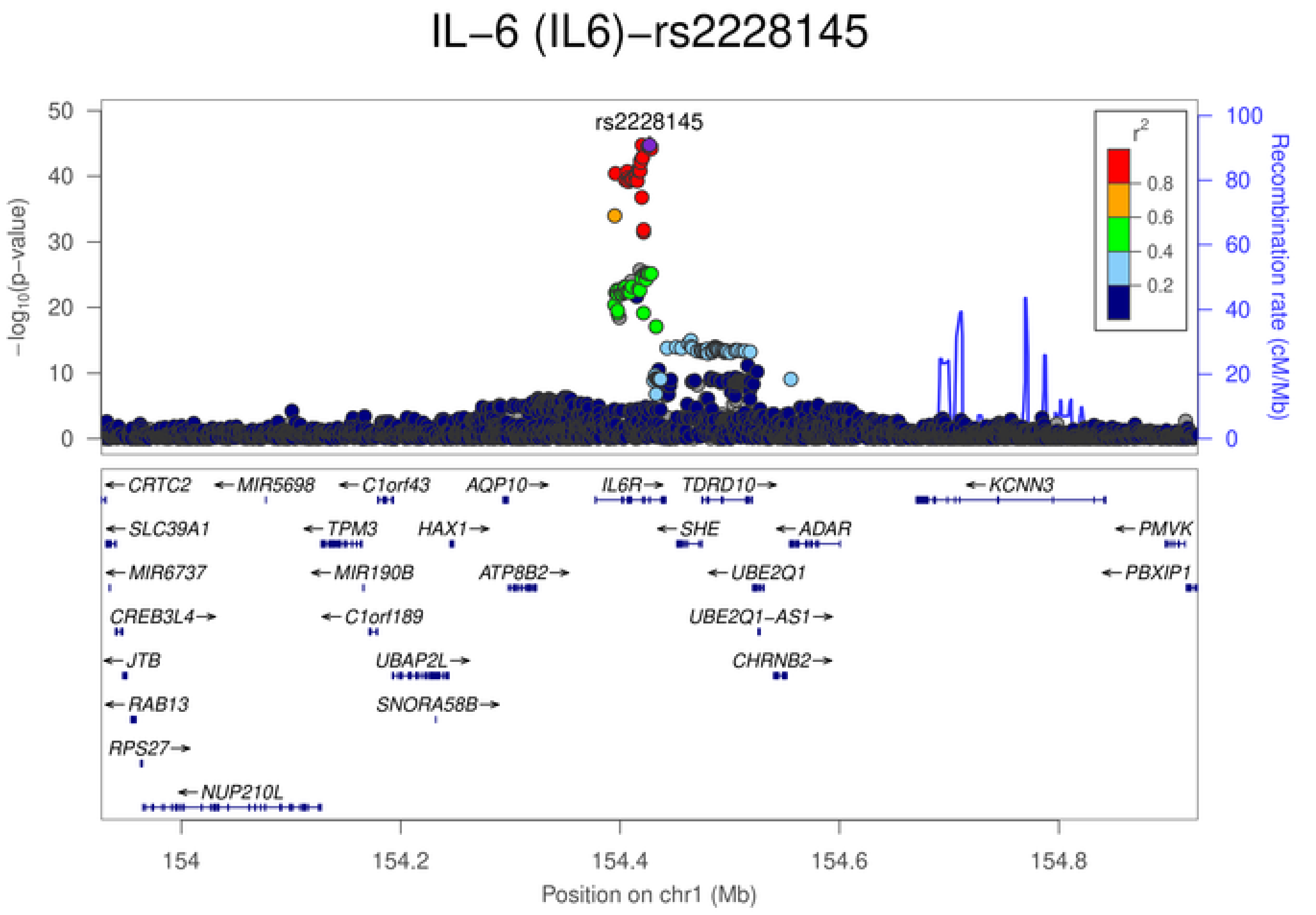
IL−6 (IL6)−rs2228145
rs2228145
$-\log_{10}$(p−value)
Recombination rate (cM/Mb)
$r^2$
0.8
0.6
0.4
0.2
CRTC2
MIR5698
C1orf43
AQP10
IL6R
TDRD10
KCNN3
SLC39A1
TPM3
HAX1
SHE
ADAR
PMVK
MIR6737
MIR190B
ATP8B2
UBE2Q1
PBXIP1
CREB3L4
C1orf189
UBE2Q1−AS1
JTB
UBAP2L
CHRNB2
RAB13
SNORA58B
RPS27
NUP210L
154 154.2 154.4 154.6 154.8
Position on chr1 (Mb)

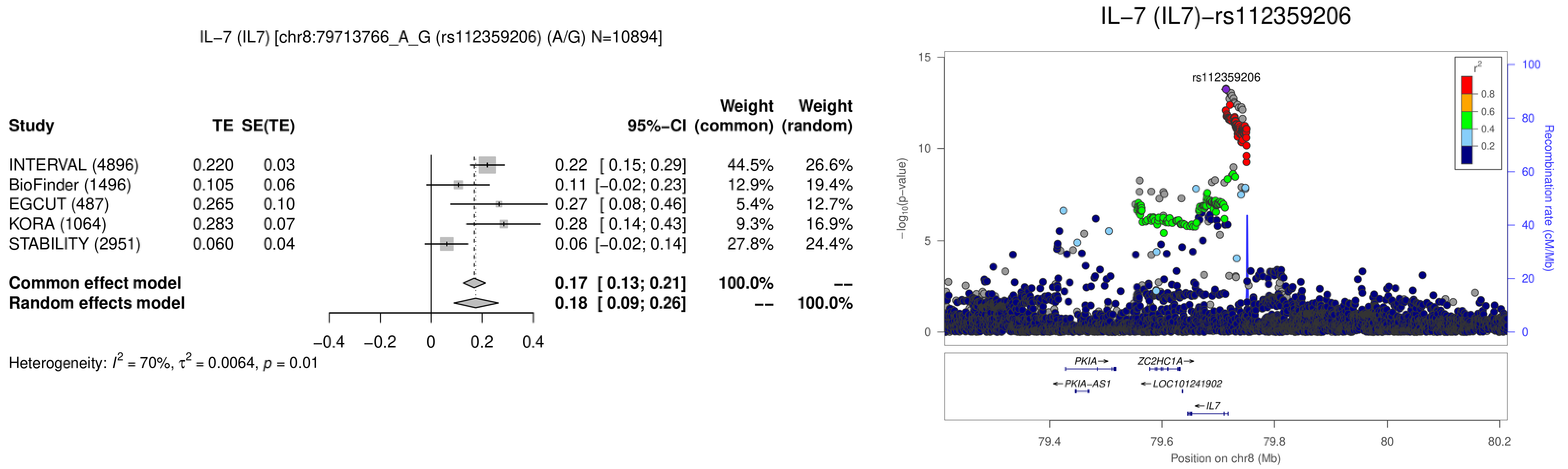
IL−7 (IL7) [chr8:79713766_A_G (rs112359206) (A/G) N=10894]
Study
TE
SE(TE)
95%−CI
Weight (common)
Weight (random)
INTERVAL (4896) 0.220 0.03 0.22 [ 0.15; 0.29] 44.5% 26.6%
BioFinder (1496) 0.105 0.06 0.11 [−0.02; 0.23] 12.9% 19.4%
EGCUT (487) 0.265 0.10 0.27 [ 0.08; 0.46] 5.4% 12.7%
KORA (1064) 0.283 0.07 0.28 [ 0.14; 0.43] 9.3% 16.9%
STABILITY (2951) 0.060 0.04 0.06 [−0.02; 0.14] 27.8% 24.4%
Common effect model 0.17 [ 0.13; 0.21] 100.0% --
Random effects model 0.18 [ 0.09; 0.26] -- 100.0%
−0.4 −0.2 0 0.2 0.4
Heterogeneity: $I^2$ = 70%, $\tau^2$ = 0.0064, $p$ = 0.01
IL−7 (IL7)−rs112359206
rs112359206
$-\log_{10}$(p−value)
Recombination rate (cM/Mb)
$r^2$
0.8
0.6
0.4
0.2
PKIA→
ZC2HC1A→
←PKIA−AS1
←LOC101241902
←IL7
79.4
79.6
79.8
80
80.2
Position on chr8 (Mb)

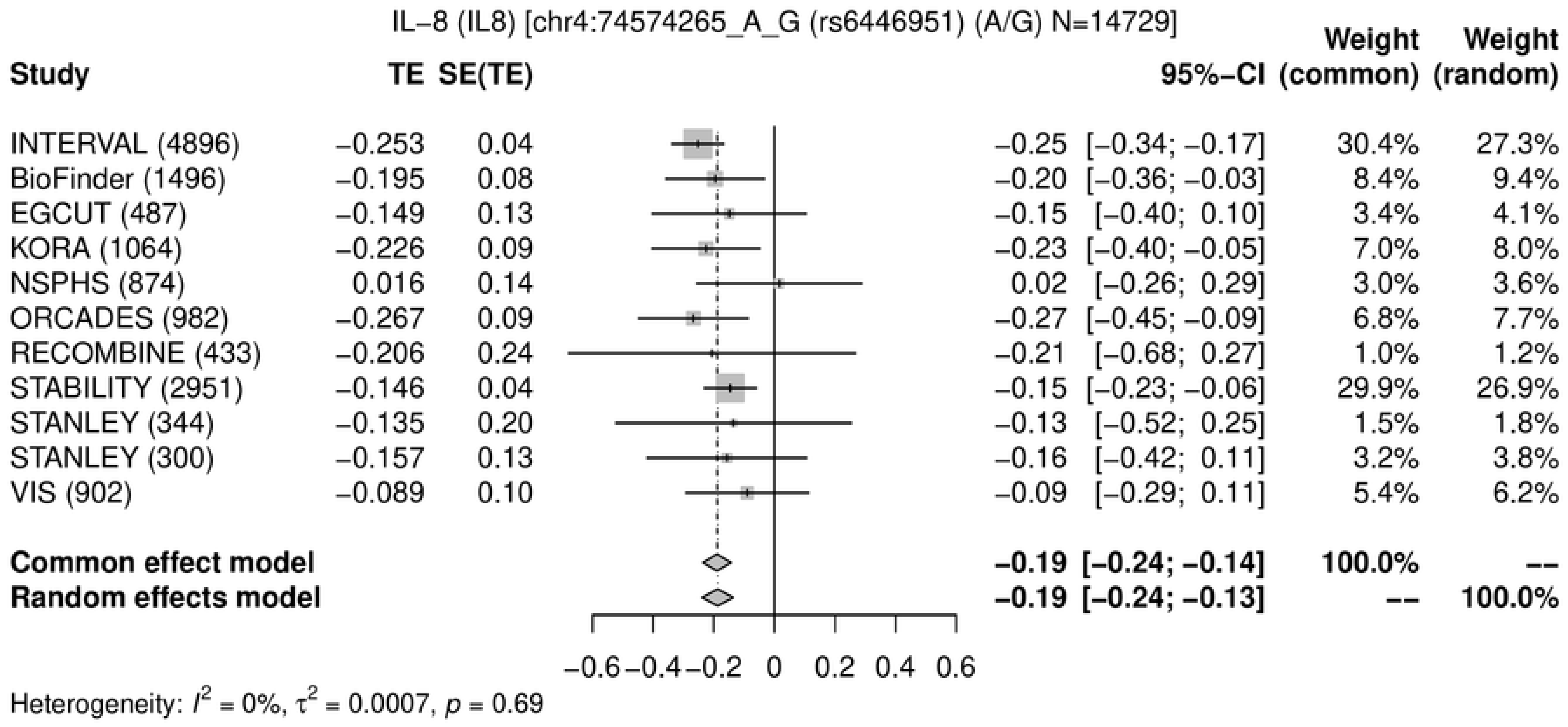
IL−8 (IL8) [chr4:74574265_A_G (rs6446951) (A/G) N=14729]
Study
TE
SE(TE)
95%−CI
Weight (common)
Weight (random)
INTERVAL (4896) −0.253 0.04 −0.25 [−0.34; −0.17] 30.4% 27.3%
BioFinder (1496) −0.195 0.08 −0.20 [−0.36; −0.03] 8.4% 9.4%
EGCUT (487) −0.149 0.13 −0.15 [−0.40; 0.10] 3.4% 4.1%
KORA (1064) −0.226 0.09 −0.23 [−0.40; −0.05] 7.0% 8.0%
NSPHS (874) 0.016 0.14 0.02 [−0.26; 0.29] 3.0% 3.6%
ORCADES (982) −0.267 0.09 −0.27 [−0.45; −0.09] 6.8% 7.7%
RECOMBINE (433) −0.206 0.24 −0.21 [−0.68; 0.27] 1.0% 1.2%
STABILITY (2951) −0.146 0.04 −0.15 [−0.23; −0.06] 29.9% 26.9%
STANLEY (344) −0.135 0.20 −0.13 [−0.52; 0.25] 1.5% 1.8%
STANLEY (300) −0.157 0.13 −0.16 [−0.42; 0.11] 3.2% 3.8%
VIS (902) −0.089 0.10 −0.09 [−0.29; 0.11] 5.4% 6.2%
Common effect model −0.19 [−0.24; −0.14] 100.0% --
Random effects model −0.19 [−0.24; −0.13] -- 100.0%
−0.6 −0.4 −0.2 0 0.2 0.4 0.6
Heterogeneity: I² = 0%, τ² = 0.0007, p = 0.69

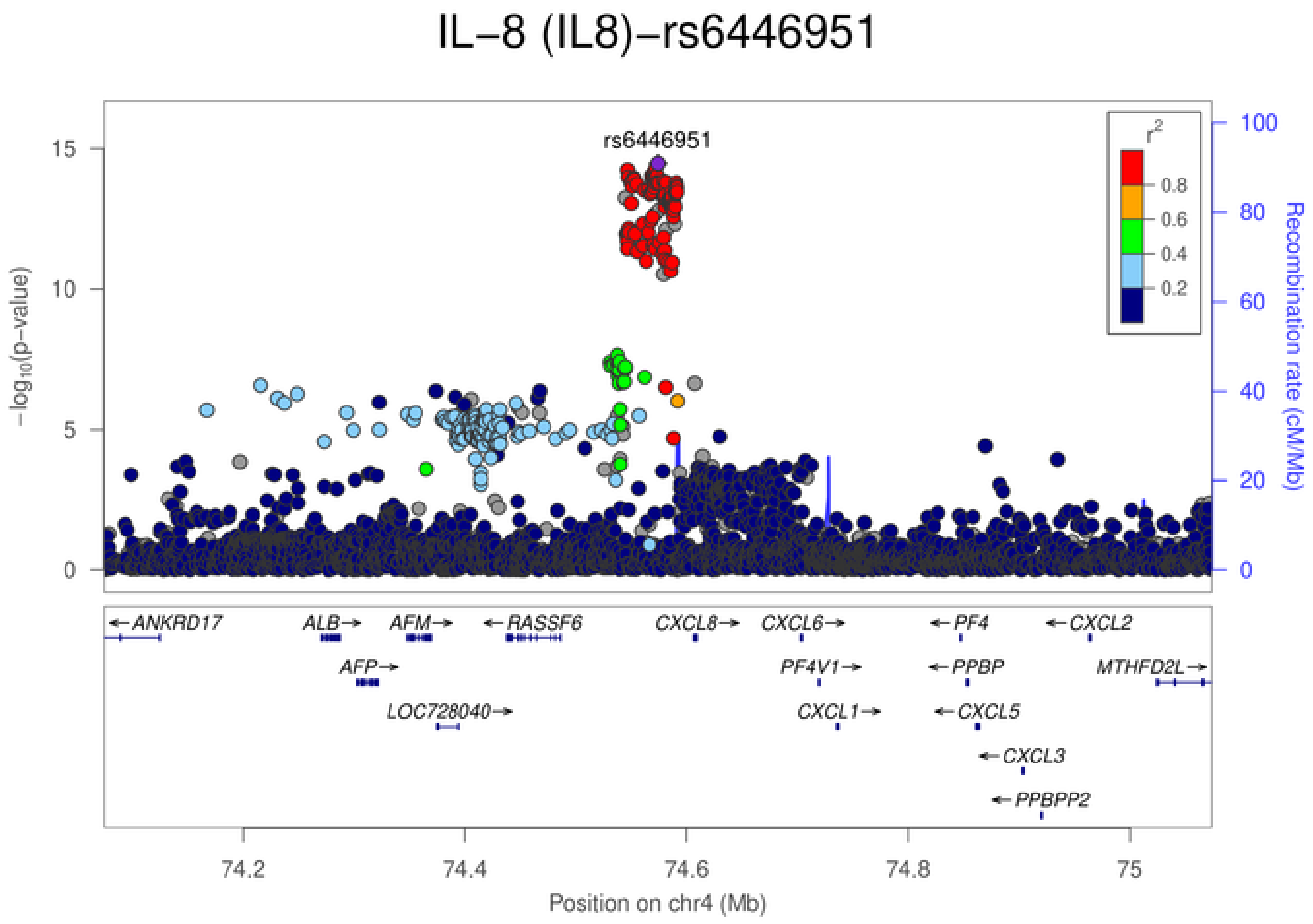
IL−8 (IL8)−rs6446951
rs6446951
r²
0.8
0.6
0.4
0.2
−log10(p−value)
Recombination rate (cM/Mb)
ANKRD17
ALB
AFM
RASSF6
CXCL8
CXCL6
PF4
CXCL2
AFP
PF4V1
PPBP
MTHFD2L
LOC728040
CXCL1
CXCL5
CXCL3
PPBPP2
74.2
74.4
74.6
74.8
75
Position on chr4 (Mb)

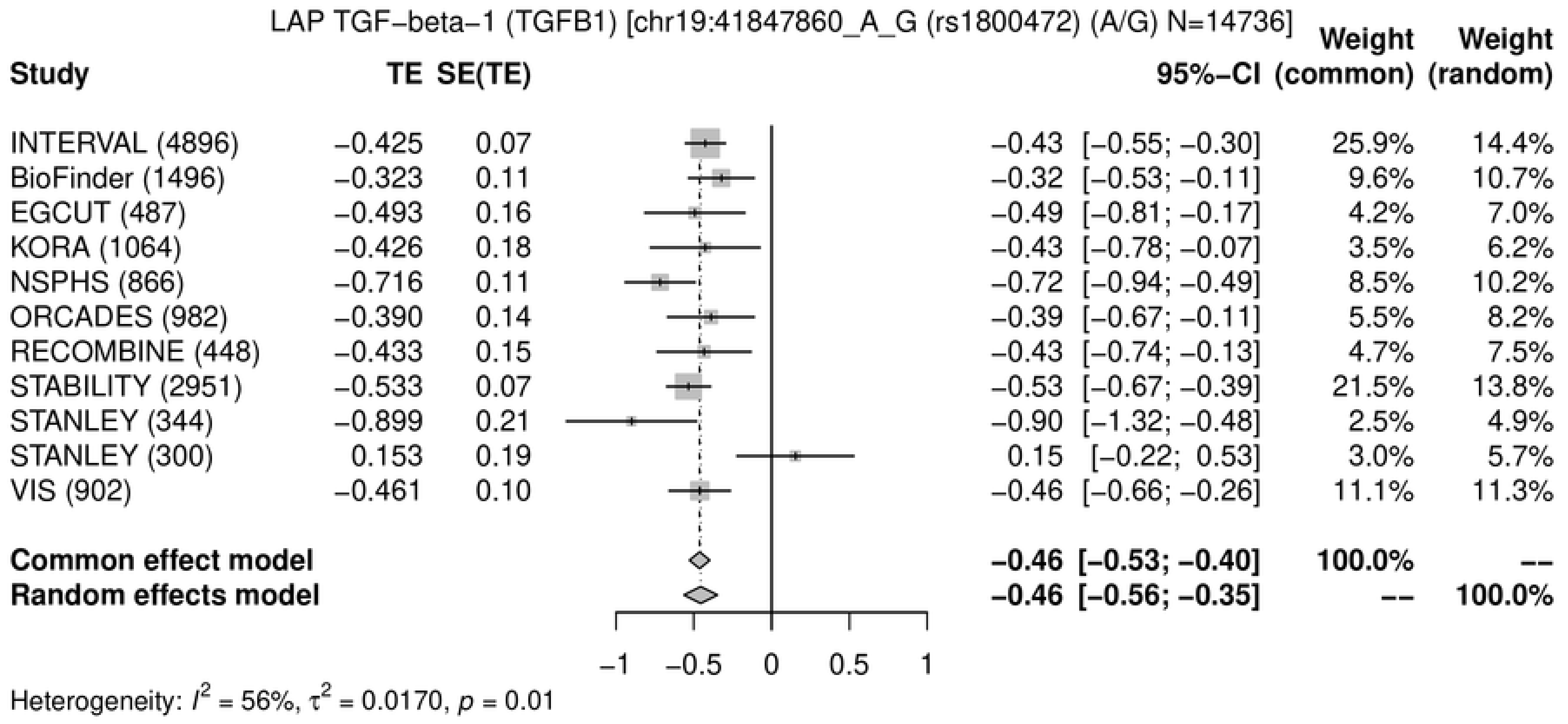
LAP TGF−beta−1 (TGFB1) [chr19:41847860_A_G (rs1800472) (A/G) N=14736]
Study
TE
SE(TE)
95%−CI
Weight (common)
Weight (random)
INTERVAL (4896) −0.425 0.07 −0.43 [−0.55; −0.30] 25.9% 14.4%
BioFinder (1496) −0.323 0.11 −0.32 [−0.53; −0.11] 9.6% 10.7%
EGCUT (487) −0.493 0.16 −0.49 [−0.81; −0.17] 4.2% 7.0%
KORA (1064) −0.426 0.18 −0.43 [−0.78; −0.07] 3.5% 6.2%
NSPHS (866) −0.716 0.11 −0.72 [−0.94; −0.49] 8.5% 10.2%
ORCADES (982) −0.390 0.14 −0.39 [−0.67; −0.11] 5.5% 8.2%
RECOMBINE (448) −0.433 0.15 −0.43 [−0.74; −0.13] 4.7% 7.5%
STABILITY (2951) −0.533 0.07 −0.53 [−0.67; −0.39] 21.5% 13.8%
STANLEY (344) −0.899 0.21 −0.90 [−1.32; −0.48] 2.5% 4.9%
STANLEY (300) 0.153 0.19 0.15 [−0.22; 0.53] 3.0% 5.7%
VIS (902) −0.461 0.10 −0.46 [−0.66; −0.26] 11.1% 11.3%
Common effect model −0.46 [−0.53; −0.40] 100.0% --
Random effects model −0.46 [−0.56; −0.35] -- 100.0%
−1 −0.5 0 0.5 1
Heterogeneity: $I^2$ = 56%, $\tau^2$ = 0.0170, $p$ = 0.01

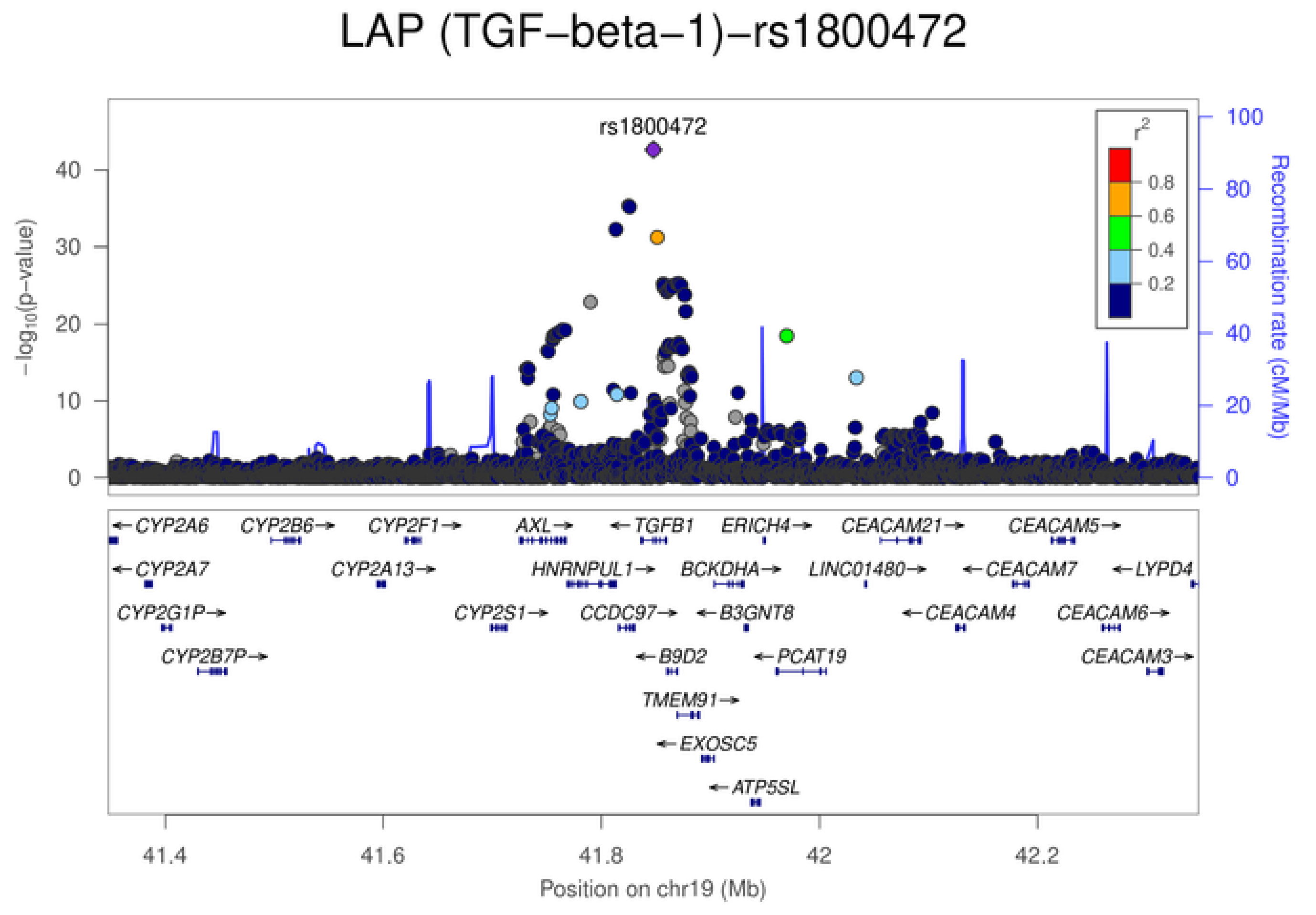
LAP (TGF−beta−1)−rs1800472
rs1800472
$-\log_{10}$(p−value)
0
10
20
30
40
$r^2$
0.8
0.6
0.4
0.2
Recombination rate (cM/Mb)
0
20
40
60
80
100
CYP2A6
CYP2B6
CYP2F1
AXL
TGFB1
ERICH4
CEACAM21
CEACAM5
CYP2A7
CYP2A13
HNRNPUL1
BCKDHA
LINC01480
CEACAM7
LYPD4
CYP2G1P
CYP2S1
CCDC97
B3GNT8
CEACAM4
CEACAM6
CYP2B7P
B9D2
PCAT19
CEACAM3
TMEM91
EXOSC5
ATP5SL
41.4
41.6
41.8
42
42.2
Position on chr19 (Mb)

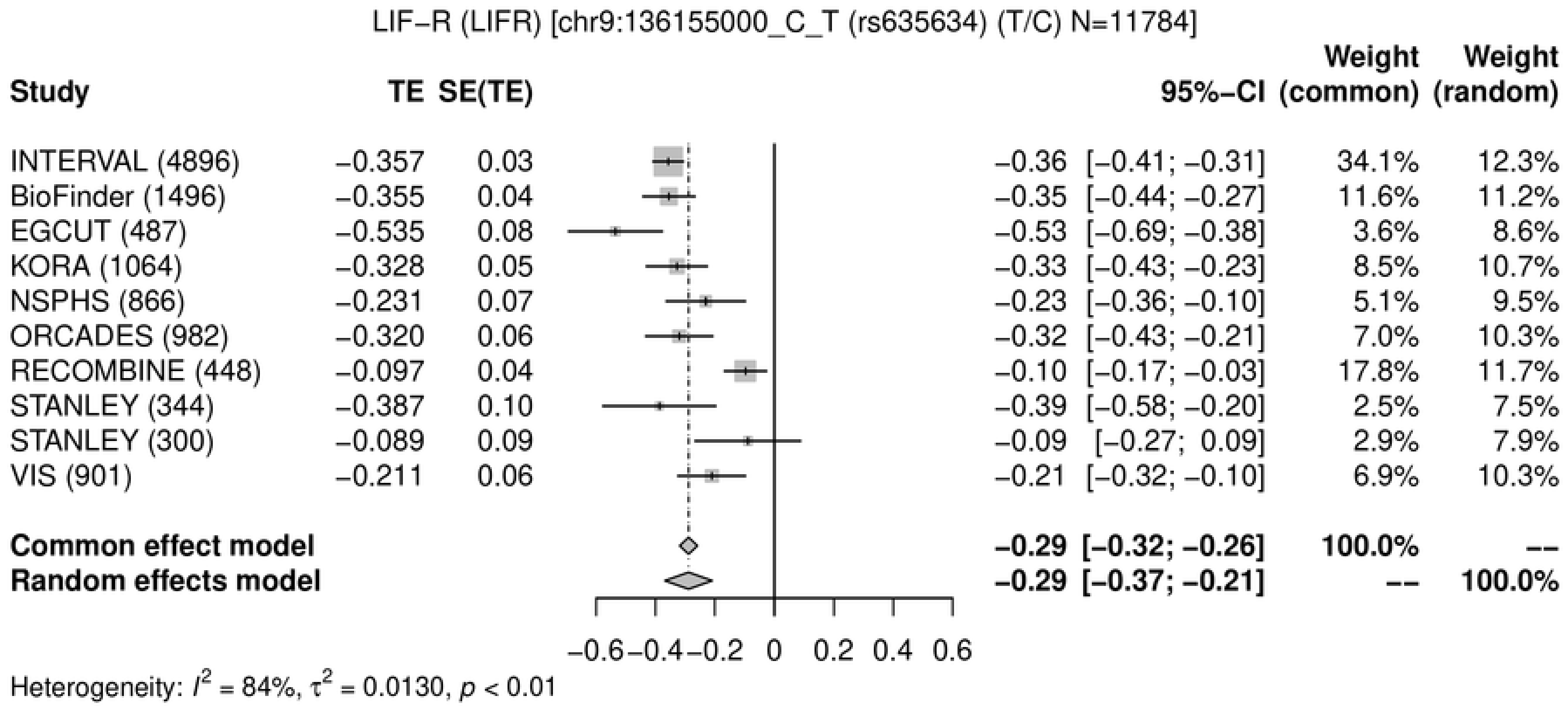
LIF−R (LIFR) [chr9:136155000_C_T (rs635634) (T/C) N=11784]
Study TE SE(TE) 95%−CI Weight (common) Weight (random)
INTERVAL (4896) −0.357 0.03 −0.36 [−0.41; −0.31] 34.1% 12.3%
BioFinder (1496) −0.355 0.04 −0.35 [−0.44; −0.27] 11.6% 11.2%
EGCUT (487) −0.535 0.08 −0.53 [−0.69; −0.38] 3.6% 8.6%
KORA (1064) −0.328 0.05 −0.33 [−0.43; −0.23] 8.5% 10.7%
NSPHS (866) −0.231 0.07 −0.23 [−0.36; −0.10] 5.1% 9.5%
ORCADES (982) −0.320 0.06 −0.32 [−0.43; −0.21] 7.0% 10.3%
RECOMBINE (448) −0.097 0.04 −0.10 [−0.17; −0.03] 17.8% 11.7%
STANLEY (344) −0.387 0.10 −0.39 [−0.58; −0.20] 2.5% 7.5%
STANLEY (300) −0.089 0.09 −0.09 [−0.27; 0.09] 2.9% 7.9%
VIS (901) −0.211 0.06 −0.21 [−0.32; −0.10] 6.9% 10.3%
Common effect model −0.29 [−0.32; −0.26] 100.0% --
Random effects model −0.29 [−0.37; −0.21] -- 100.0%
−0.6 −0.4 −0.2 0 0.2 0.4 0.6
Heterogeneity: $I^2$ = 84%, $\tau^2$ = 0.0130, $p$ < 0.01

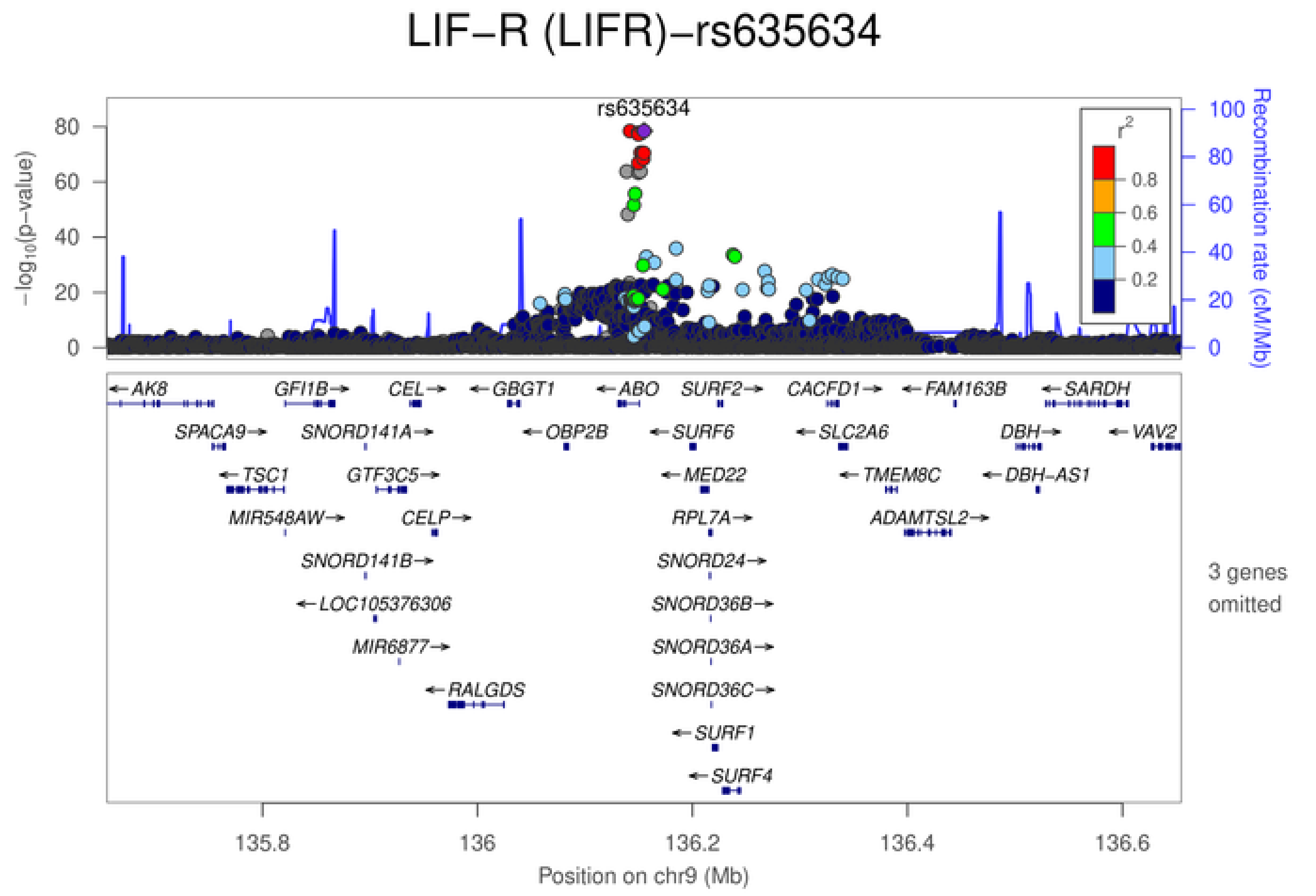
LIF−R (LIFR)−rs635634
rs635634
$-\log_{10}$(p−value)
Recombination rate (cM/Mb)
$r^2$
0.8
0.6
0.4
0.2
AK8 GFI1B CEL GBGT1 ABO SURF2 CACFD1 FAM163B SARDH
SPACA9 SNORD141A OBP2B SURF6 SLC2A6 DBH VAV2
TSC1 GTF3C5 MED22 TMEM8C DBH−AS1
MIR548AW CELP RPL7A ADAMTSL2
SNORD141B SNORD24
LOC105376306 SNORD36B
MIR6877 SNORD36A
RALGDS SNORD36C
SURF1
SURF4
3 genes omitted
135.8 136 136.2 136.4 136.6
Position on chr9 (Mb)

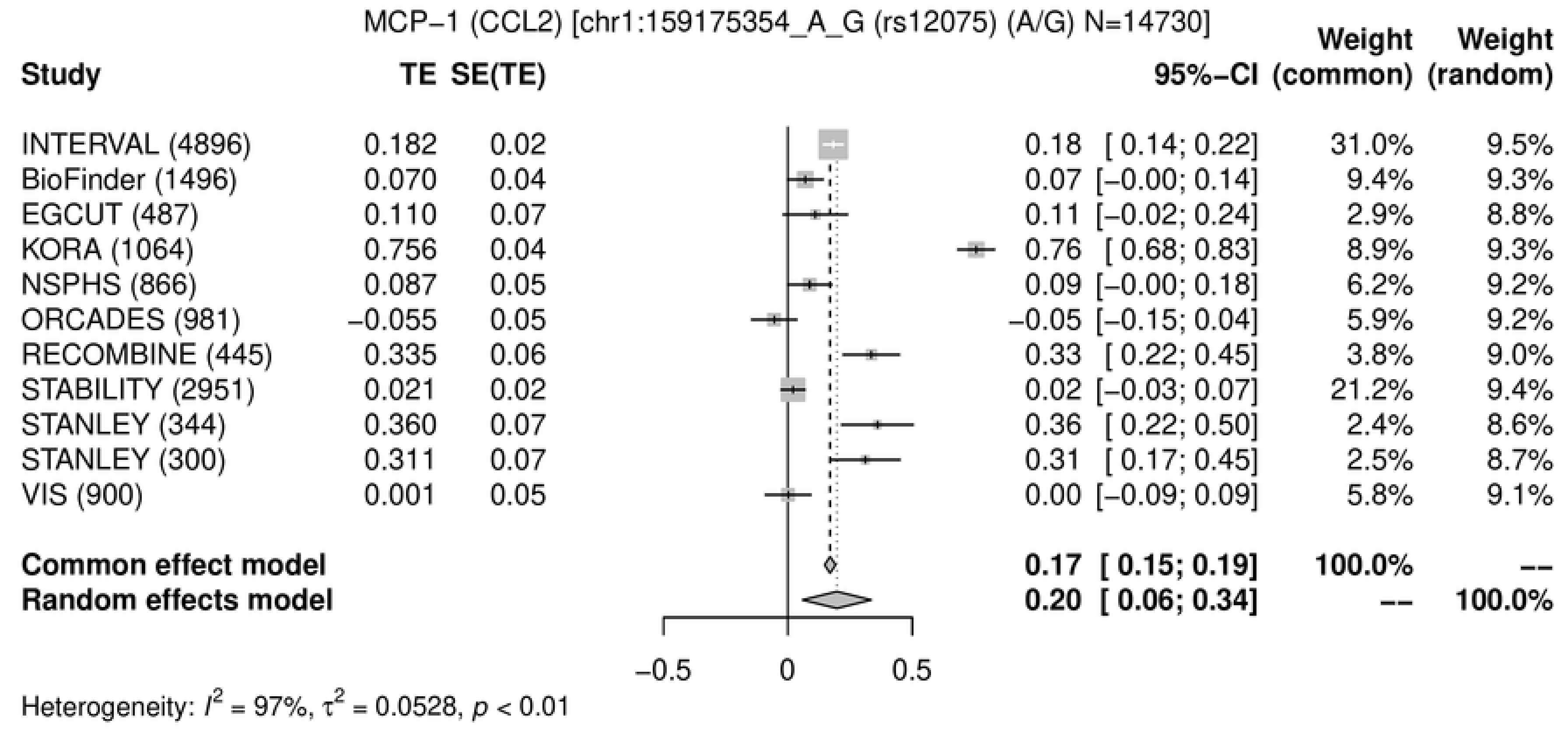
MCP−1 (CCL2) [chr1:159175354_A_G (rs12075) (A/G) N=14730]
Study
TE
SE(TE)
95%−CI
Weight (common)
Weight (random)
INTERVAL (4896) 0.182 0.02 0.18 [ 0.14; 0.22] 31.0% 9.5%
BioFinder (1496) 0.070 0.04 0.07 [−0.00; 0.14] 9.4% 9.3%
EGCUT (487) 0.110 0.07 0.11 [−0.02; 0.24] 2.9% 8.8%
KORA (1064) 0.756 0.04 0.76 [ 0.68; 0.83] 8.9% 9.3%
NSPHS (866) 0.087 0.05 0.09 [−0.00; 0.18] 6.2% 9.2%
ORCADES (981) −0.055 0.05 −0.05 [−0.15; 0.04] 5.9% 9.2%
RECOMBINE (445) 0.335 0.06 0.33 [ 0.22; 0.45] 3.8% 9.0%
STABILITY (2951) 0.021 0.02 0.02 [−0.03; 0.07] 21.2% 9.4%
STANLEY (344) 0.360 0.07 0.36 [ 0.22; 0.50] 2.4% 8.6%
STANLEY (300) 0.311 0.07 0.31 [ 0.17; 0.45] 2.5% 8.7%
VIS (900) 0.001 0.05 0.00 [−0.09; 0.09] 5.8% 9.1%
Common effect model 0.17 [ 0.15; 0.19] 100.0% --
Random effects model 0.20 [ 0.06; 0.34] -- 100.0%
−0.5
0
0.5
Heterogeneity: $I^2$ = 97%, $\tau^2$ = 0.0528, $p$ < 0.01

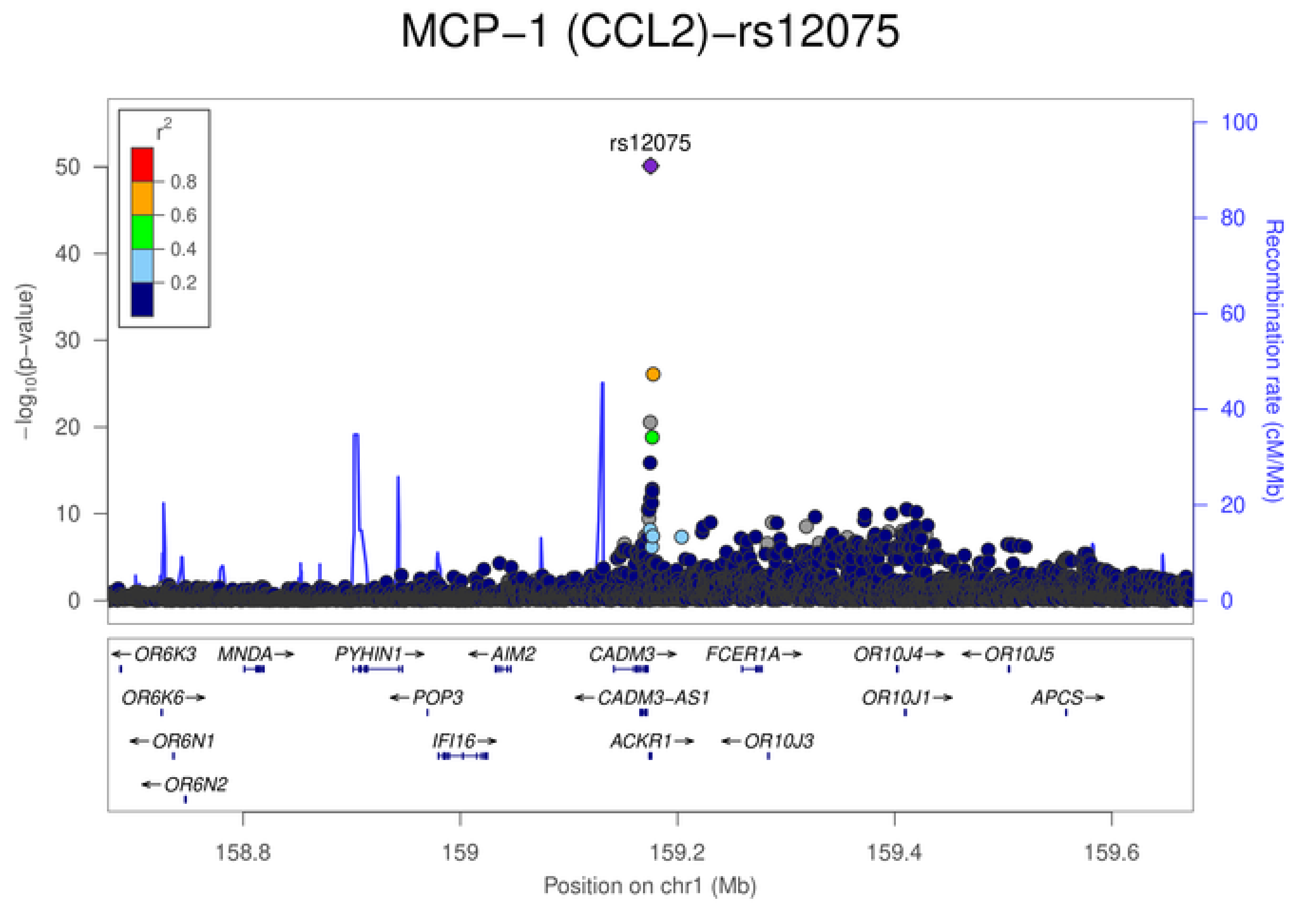
MCP−1 (CCL2)−rs12075
rs12075
$r^2$
0.8
0.6
0.4
0.2
$-\log_{10}$(p−value)
Recombination rate (cM/Mb)
0
10
20
30
40
50
0
20
40
60
80
100
OR6K3
MNDA
PYHIN1
AIM2
CADM3
FCER1A
OR10J4
OR10J5
OR6K6
POP3
CADM3-AS1
OR10J1
APCS
OR6N1
IFI16
ACKR1
OR10J3
OR6N2
158.8
159
159.2
159.4
159.6
Position on chr1 (Mb)

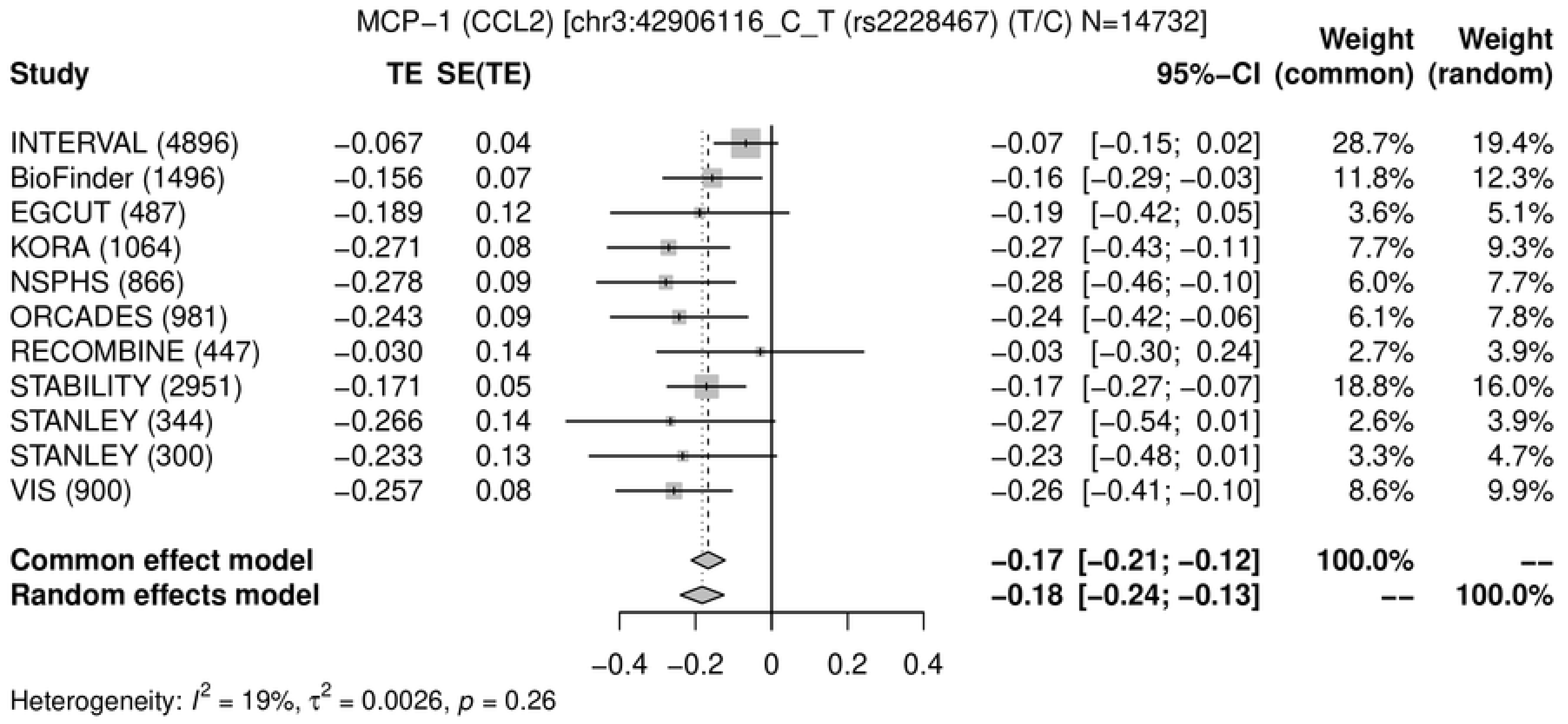
MCP−1 (CCL2) [chr3:42906116_C_T (rs2228467) (T/C) N=14732]
Study TE SE(TE) 95%−CI Weight (common) Weight (random)
INTERVAL (4896) −0.067 0.04 −0.07 [−0.15; 0.02] 28.7% 19.4%
BioFinder (1496) −0.156 0.07 −0.16 [−0.29; −0.03] 11.8% 12.3%
EGCUT (487) −0.189 0.12 −0.19 [−0.42; 0.05] 3.6% 5.1%
KORA (1064) −0.271 0.08 −0.27 [−0.43; −0.11] 7.7% 9.3%
NSPHS (866) −0.278 0.09 −0.28 [−0.46; −0.10] 6.0% 7.7%
ORCADES (981) −0.243 0.09 −0.24 [−0.42; −0.06] 6.1% 7.8%
RECOMBINE (447) −0.030 0.14 −0.03 [−0.30; 0.24] 2.7% 3.9%
STABILITY (2951) −0.171 0.05 −0.17 [−0.27; −0.07] 18.8% 16.0%
STANLEY (344) −0.266 0.14 −0.27 [−0.54; 0.01] 2.6% 3.9%
STANLEY (300) −0.233 0.13 −0.23 [−0.48; 0.01] 3.3% 4.7%
VIS (900) −0.257 0.08 −0.26 [−0.41; −0.10] 8.6% 9.9%
Common effect model −0.17 [−0.21; −0.12] 100.0% --
Random effects model −0.18 [−0.24; −0.13] -- 100.0%
−0.4 −0.2 0 0.2 0.4
Heterogeneity: $I^2$ = 19%, $\tau^2$ = 0.0026, $p$ = 0.26

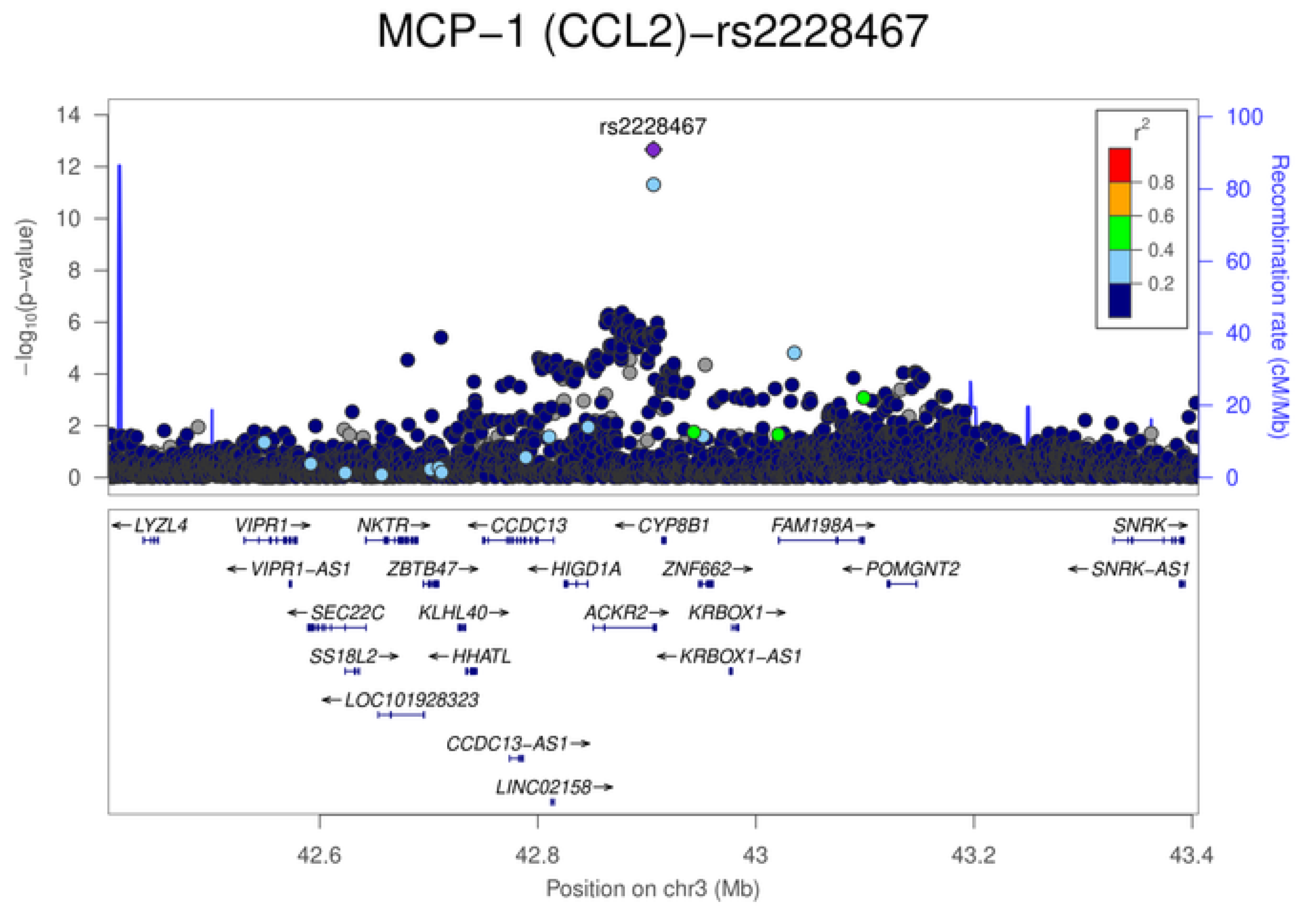
MCP−1 (CCL2)−rs2228467
rs2228467
$-\log_{10}$(p−value)
Recombination rate (cM/Mb)
$r^2$
0.8
0.6
0.4
0.2
LYZL4
VIPR1
NKTR
CCDC13
CYP8B1
FAM198A
SNRK
VIPR1−AS1
ZBTB47
HIGD1A
ZNF662
POMGNT2
SNRK−AS1
SEC22C
KLHL40
ACKR2
KRBOX1
SS18L2
HHATL
KRBOX1−AS1
LOC101928323
CCDC13−AS1
LINC02158
42.6 42.8 43 43.2 43.4
Position on chr3 (Mb)

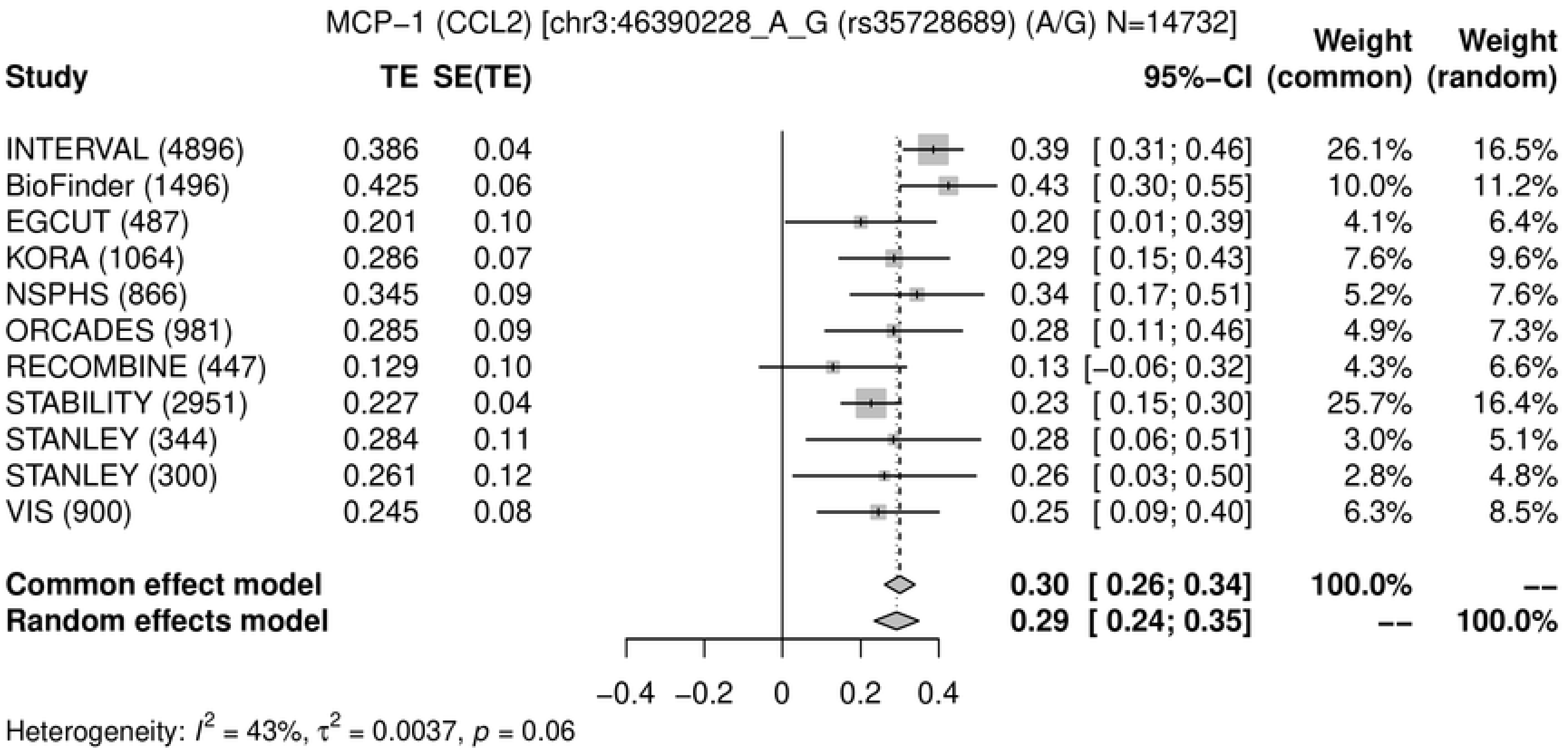
MCP−1 (CCL2) [chr3:46390228_A_G (rs35728689) (A/G) N=14732]
Study
TE
SE(TE)
95%−CI
Weight (common)
Weight (random)
INTERVAL (4896) 0.386 0.04 0.39 [ 0.31; 0.46] 26.1% 16.5%
BioFinder (1496) 0.425 0.06 0.43 [ 0.30; 0.55] 10.0% 11.2%
EGCUT (487) 0.201 0.10 0.20 [ 0.01; 0.39] 4.1% 6.4%
KORA (1064) 0.286 0.07 0.29 [ 0.15; 0.43] 7.6% 9.6%
NSPHS (866) 0.345 0.09 0.34 [ 0.17; 0.51] 5.2% 7.6%
ORCADES (981) 0.285 0.09 0.28 [ 0.11; 0.46] 4.9% 7.3%
RECOMBINE (447) 0.129 0.10 0.13 [−0.06; 0.32] 4.3% 6.6%
STABILITY (2951) 0.227 0.04 0.23 [ 0.15; 0.30] 25.7% 16.4%
STANLEY (344) 0.284 0.11 0.28 [ 0.06; 0.51] 3.0% 5.1%
STANLEY (300) 0.261 0.12 0.26 [ 0.03; 0.50] 2.8% 4.8%
VIS (900) 0.245 0.08 0.25 [ 0.09; 0.40] 6.3% 8.5%
Common effect model 0.30 [ 0.26; 0.34] 100.0% --
Random effects model 0.29 [ 0.24; 0.35] -- 100.0%
−0.4 −0.2 0 0.2 0.4
Heterogeneity: $I^2$ = 43%, $\tau^2$ = 0.0037, $p$ = 0.06

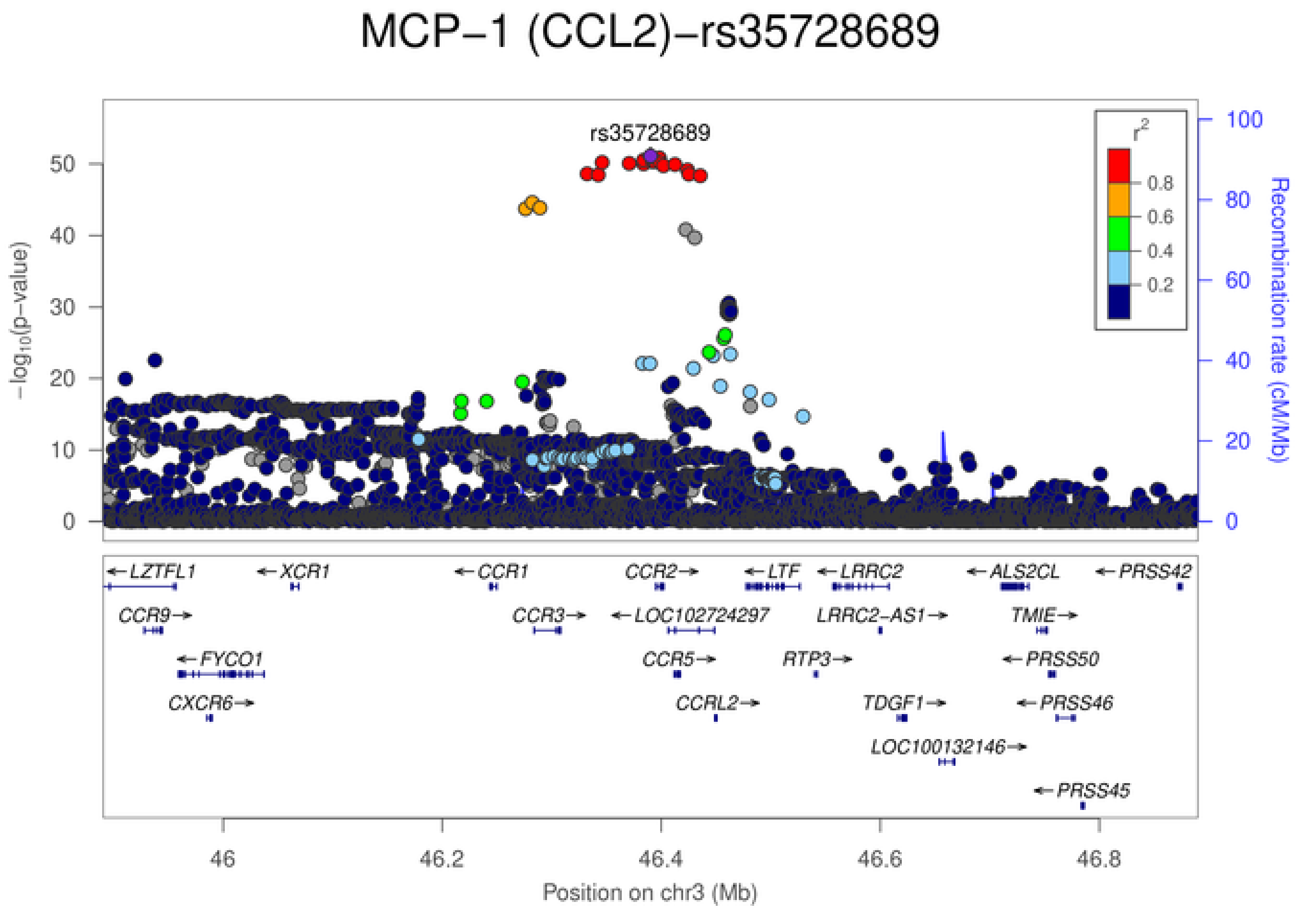
MCP−1 (CCL2)−rs35728689
rs35728689
$-\log_{10}$(p-value)
Recombination rate (cM/Mb)
$r^2$
0.8
0.6
0.4
0.2
0 10 20 30 40 50
0 20 40 60 80 100
←LZTFL1
←XCR1
←CCR1
CCR2→
←LTF
←LRRC2
←ALS2CL
←PRSS42
CCR9→
CCR3→
←LOC102724297
LRRC2−AS1→
TMIE→
←FYCO1
CCR5→
RTP3→
←PRSS50
CXCR6→
CCRL2→
TDGF1→
←PRSS46
LOC100132146→
←PRSS45
46 46.2 46.4 46.6 46.8
Position on chr3 (Mb)

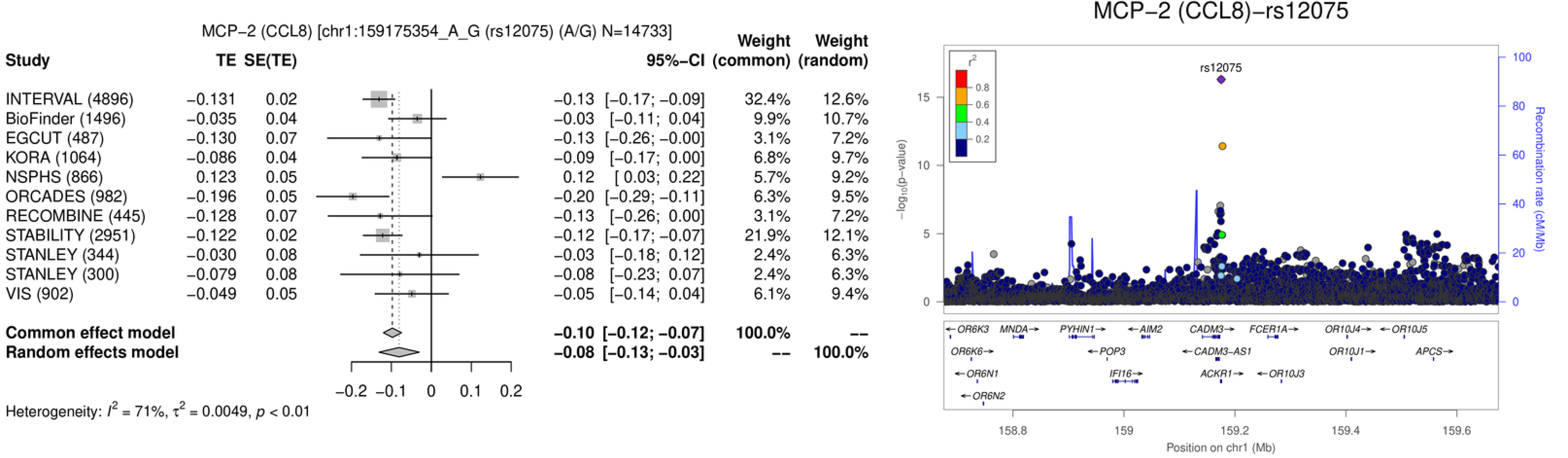
MCP−2 (CCL8) [chr1:159175354_A_G (rs12075) (A/G) N=14733]
Study TE SE(TE) 95%−CI Weight (common) Weight (random)
INTERVAL (4896) −0.131 0.02 −0.13 [−0.17; −0.09] 32.4% 12.6%
BioFinder (1496) −0.035 0.04 −0.03 [−0.11; 0.04] 9.9% 10.7%
EGCUT (487) −0.130 0.07 −0.13 [−0.26; −0.00] 3.1% 7.2%
KORA (1064) −0.086 0.04 −0.09 [−0.17; 0.00] 6.8% 9.7%
NSPHS (866) 0.123 0.05 0.12 [ 0.03; 0.22] 5.7% 9.2%
ORCADES (982) −0.196 0.05 −0.20 [−0.29; −0.11] 6.3% 9.5%
RECOMBINE (445) −0.128 0.07 −0.13 [−0.26; 0.00] 3.1% 7.2%
STABILITY (2951) −0.122 0.02 −0.12 [−0.17; −0.07] 21.9% 12.1%
STANLEY (344) −0.030 0.08 −0.03 [−0.18; 0.12] 2.4% 6.3%
STANLEY (300) −0.079 0.08 −0.08 [−0.23; 0.07] 2.4% 6.3%
VIS (902) −0.049 0.05 −0.05 [−0.14; 0.04] 6.1% 9.4%
Common effect model −0.10 [−0.12; −0.07] 100.0% --
Random effects model −0.08 [−0.13; −0.03] -- 100.0%
−0.2 −0.1 0 0.1 0.2
Heterogeneity: I² = 71%, τ² = 0.0049, p < 0.01
MCP−2 (CCL8)−rs12075
rs12075
r²
0.8
0.6
0.4
0.2
−log10(p−value)
Recombination rate (cM/Mb)
OR6K3
MNDA
PYHIN1
AIM2
CADM3
FCER1A
OR10J4
OR10J5
OR6K6
POP3
CADM3−AS1
OR10J1
APCS
OR6N1
IFI16
ACKR1
OR10J3
OR6N2
158.8
159
159.2
159.4
159.6
Position on chr1 (Mb)

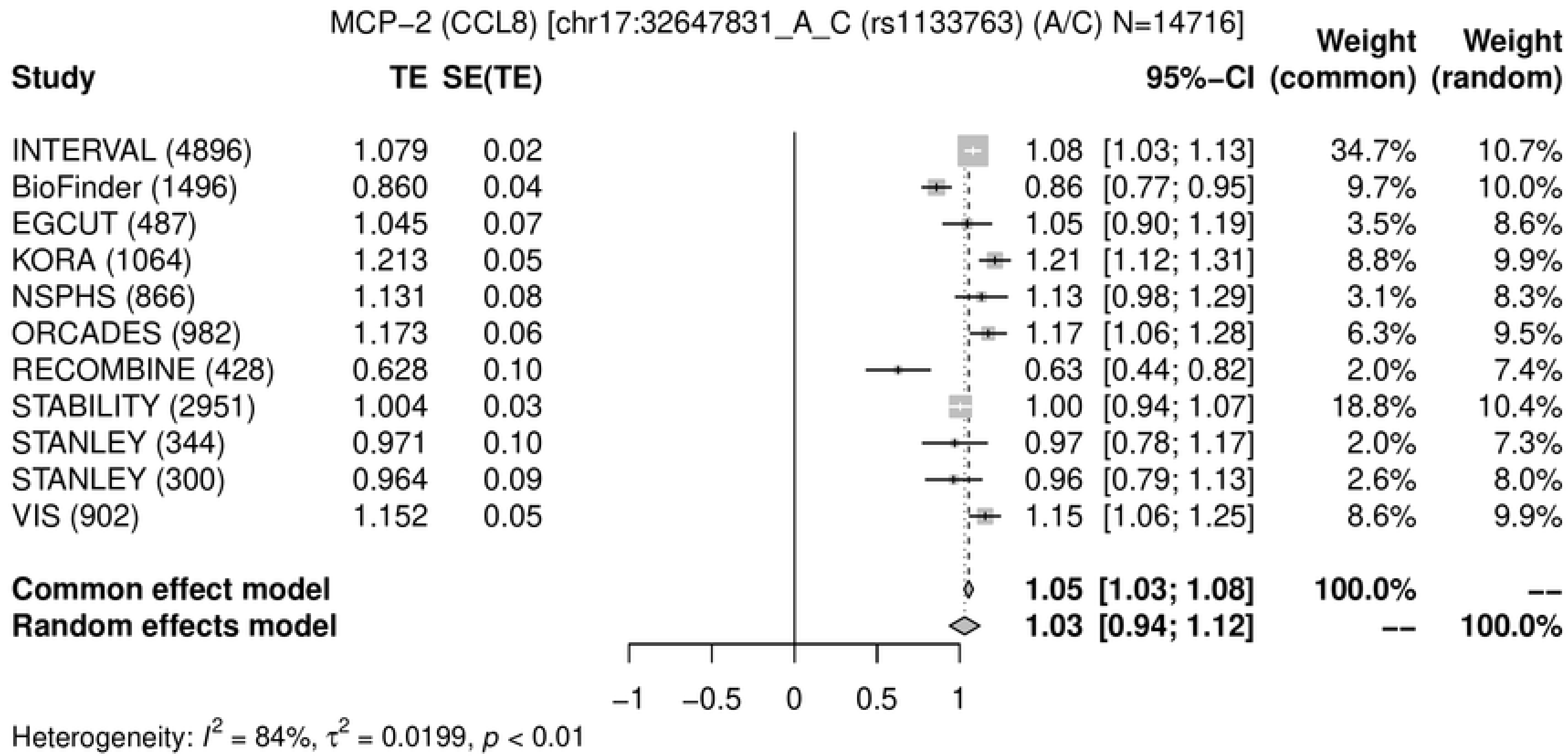
MCP−2 (CCL8) [chr17:32647831_A_C (rs1133763) (A/C) N=14716]
Study
TE
SE(TE)
95%−CI
Weight (common)
Weight (random)
INTERVAL (4896) 1.079 0.02 1.08 [1.03; 1.13] 34.7% 10.7%
BioFinder (1496) 0.860 0.04 0.86 [0.77; 0.95] 9.7% 10.0%
EGCUT (487) 1.045 0.07 1.05 [0.90; 1.19] 3.5% 8.6%
KORA (1064) 1.213 0.05 1.21 [1.12; 1.31] 8.8% 9.9%
NSPHS (866) 1.131 0.08 1.13 [0.98; 1.29] 3.1% 8.3%
ORCADES (982) 1.173 0.06 1.17 [1.06; 1.28] 6.3% 9.5%
RECOMBINE (428) 0.628 0.10 0.63 [0.44; 0.82] 2.0% 7.4%
STABILITY (2951) 1.004 0.03 1.00 [0.94; 1.07] 18.8% 10.4%
STANLEY (344) 0.971 0.10 0.97 [0.78; 1.17] 2.0% 7.3%
STANLEY (300) 0.964 0.09 0.96 [0.79; 1.13] 2.6% 8.0%
VIS (902) 1.152 0.05 1.15 [1.06; 1.25] 8.6% 9.9%
Common effect model 1.05 [1.03; 1.08] 100.0% --
Random effects model 1.03 [0.94; 1.12] -- 100.0%
−1
−0.5
0
0.5
1
Heterogeneity: $I^2$ = 84%, $\tau^2$ = 0.0199, $p$ < 0.01
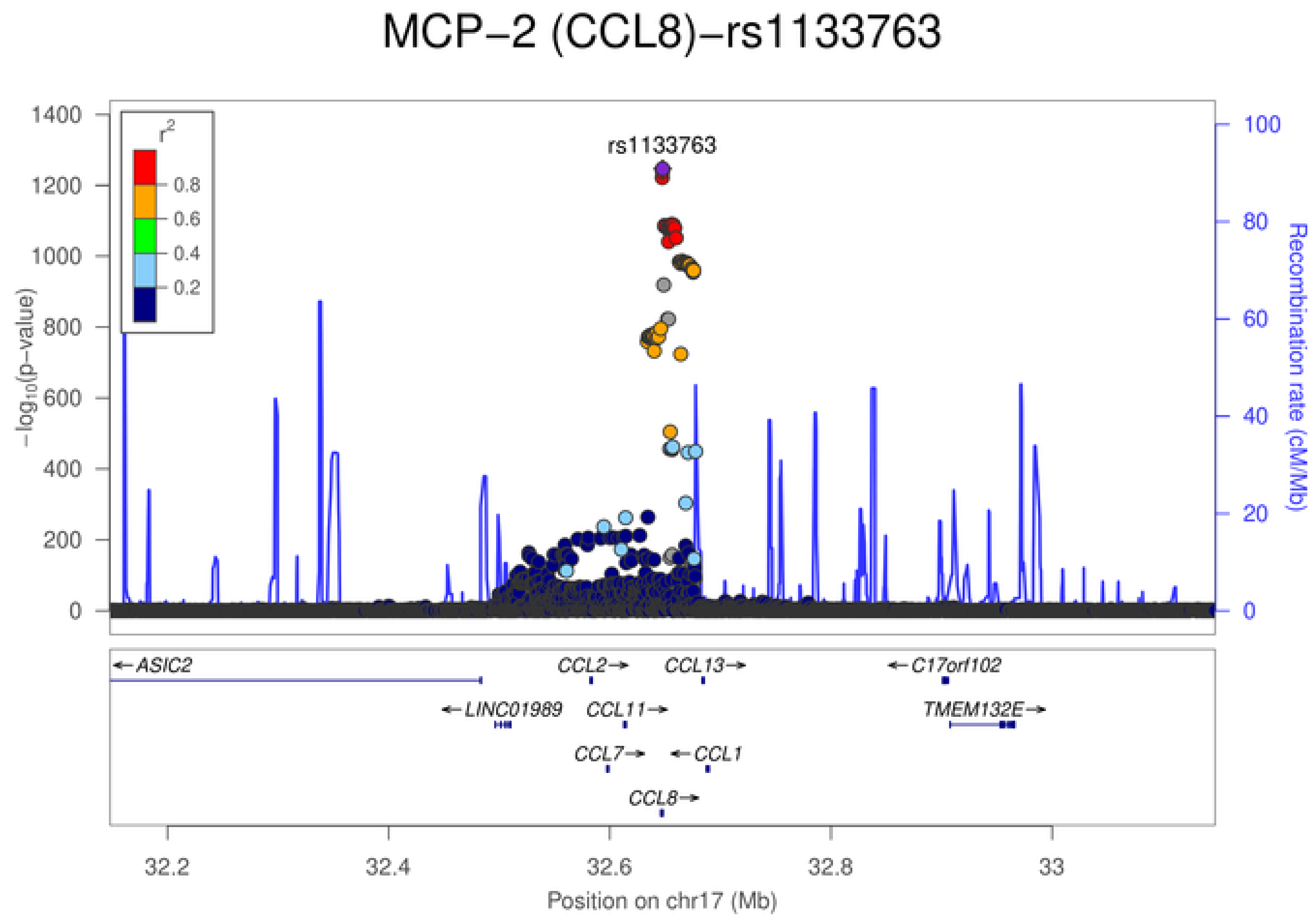
MCP−2 (CCL8)−rs1133763
rs1133763
$r^2$
0.8
0.6
0.4
0.2
1400
1200
1000
800
600
400
200
0
$-\log_{10}$(p−value)
100
80
60
40
20
0
Recombination rate (cM/Mb)
←ASIC2
CCL2→
CCL13→
←C17orf102
←LINC01989
CCL11→
TMEM132E→
CCL7→
←CCL1
CCL8→
32.2
32.4
32.6
32.8
33
Position on chr17 (Mb)

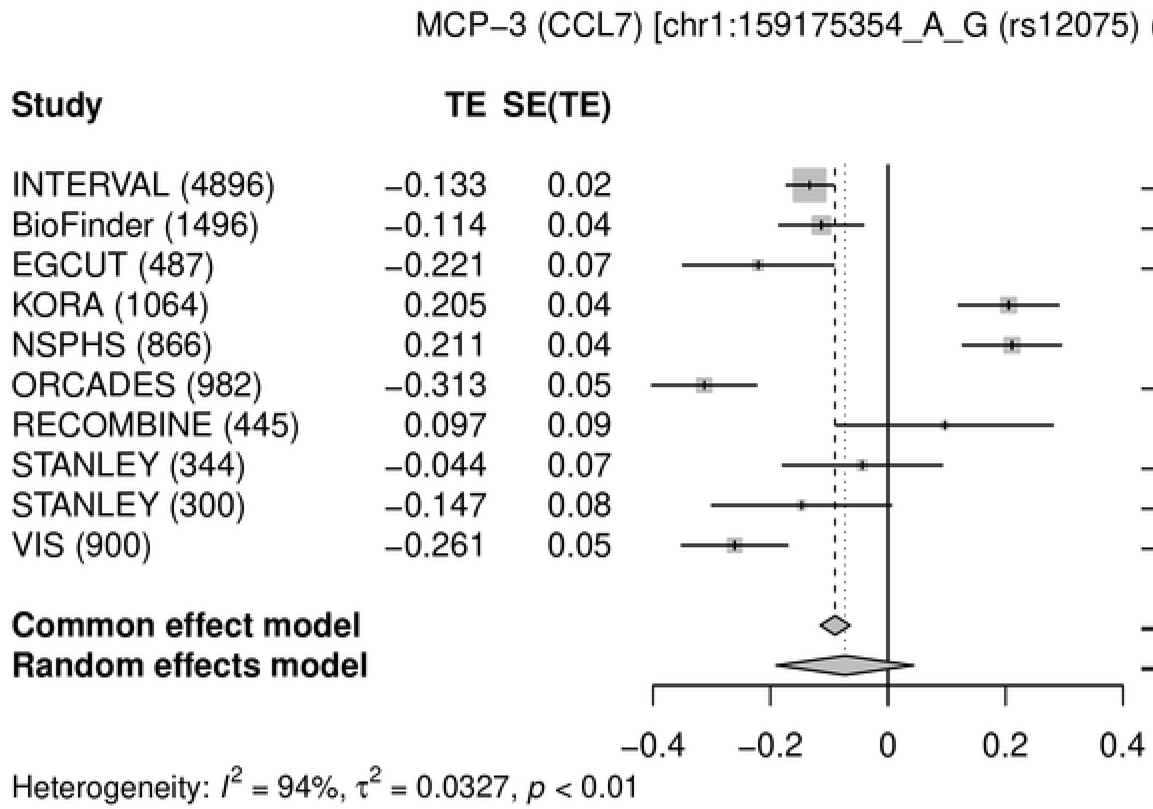
MCP−3 (CCL7) [chr1:159175354_A_G (rs12075) (A/G) N=11780]
Study
TE
SE(TE)
95%−CI
Weight (common)
Weight (random)
INTERVAL (4896) −0.133 0.02 −0.13 [−0.17; −0.09] 40.9% 10.8%
BioFinder (1496) −0.114 0.04 −0.11 [−0.19; −0.04] 12.6% 10.5%
EGCUT (487) −0.221 0.07 −0.22 [−0.35; −0.09] 3.9% 9.7%
KORA (1064) 0.205 0.04 0.21 [ 0.12; 0.29] 8.9% 10.4%
NSPHS (866) 0.211 0.04 0.21 [ 0.13; 0.30] 9.2% 10.4%
ORCADES (982) −0.313 0.05 −0.31 [−0.40; −0.22] 8.2% 10.3%
RECOMBINE (445) 0.097 0.09 0.10 [−0.09; 0.28] 1.9% 8.6%
STANLEY (344) −0.044 0.07 −0.04 [−0.18; 0.09] 3.5% 9.6%
STANLEY (300) −0.147 0.08 −0.15 [−0.30; 0.01] 2.8% 9.3%
VIS (900) −0.261 0.05 −0.26 [−0.35; −0.17] 8.0% 10.3%
Common effect model −0.09 [−0.12; −0.06] 100.0% --
Random effects model −0.07 [−0.19; 0.04] -- 100.0%
−0.4 −0.2 0 0.2 0.4
Heterogeneity: $I^2$ = 94%, $\tau^2$ = 0.0327, $p$ < 0.01

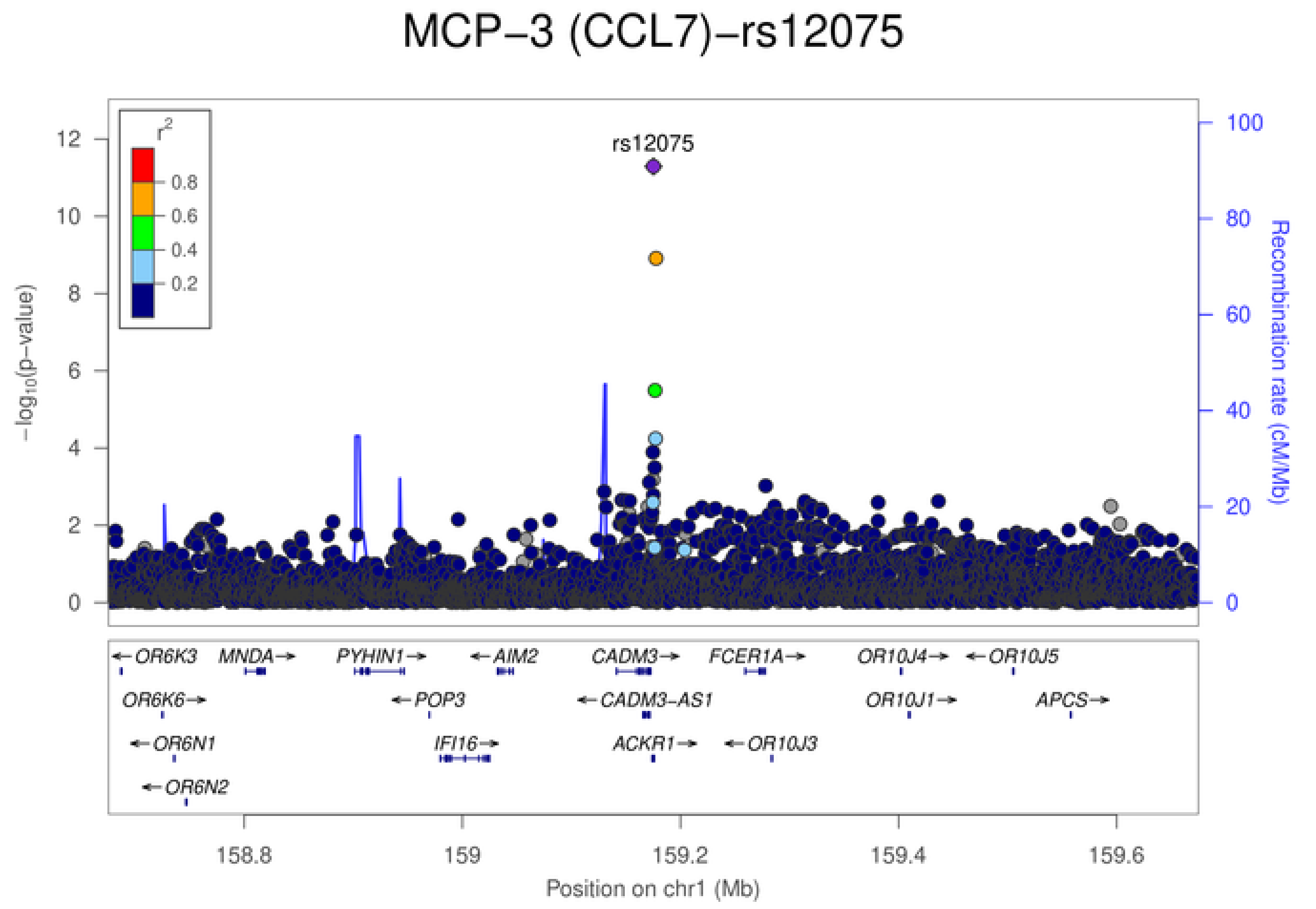
MCP−3 (CCL7)−rs12075
rs12075
$r^2$
0.8
0.6
0.4
0.2
−log10(p−value)
Recombination rate (cM/Mb)
0 2 4 6 8 10 12
0 20 40 60 80 100
OR6K3
MNDA
PYHIN1
AIM2
CADM3
FCER1A
OR10J4
OR10J5
OR6K6
POP3
CADM3−AS1
OR10J1
APCS
OR6N1
IFI16
ACKR1
OR10J3
OR6N2
158.8 159 159.2 159.4 159.6
Position on chr1 (Mb)

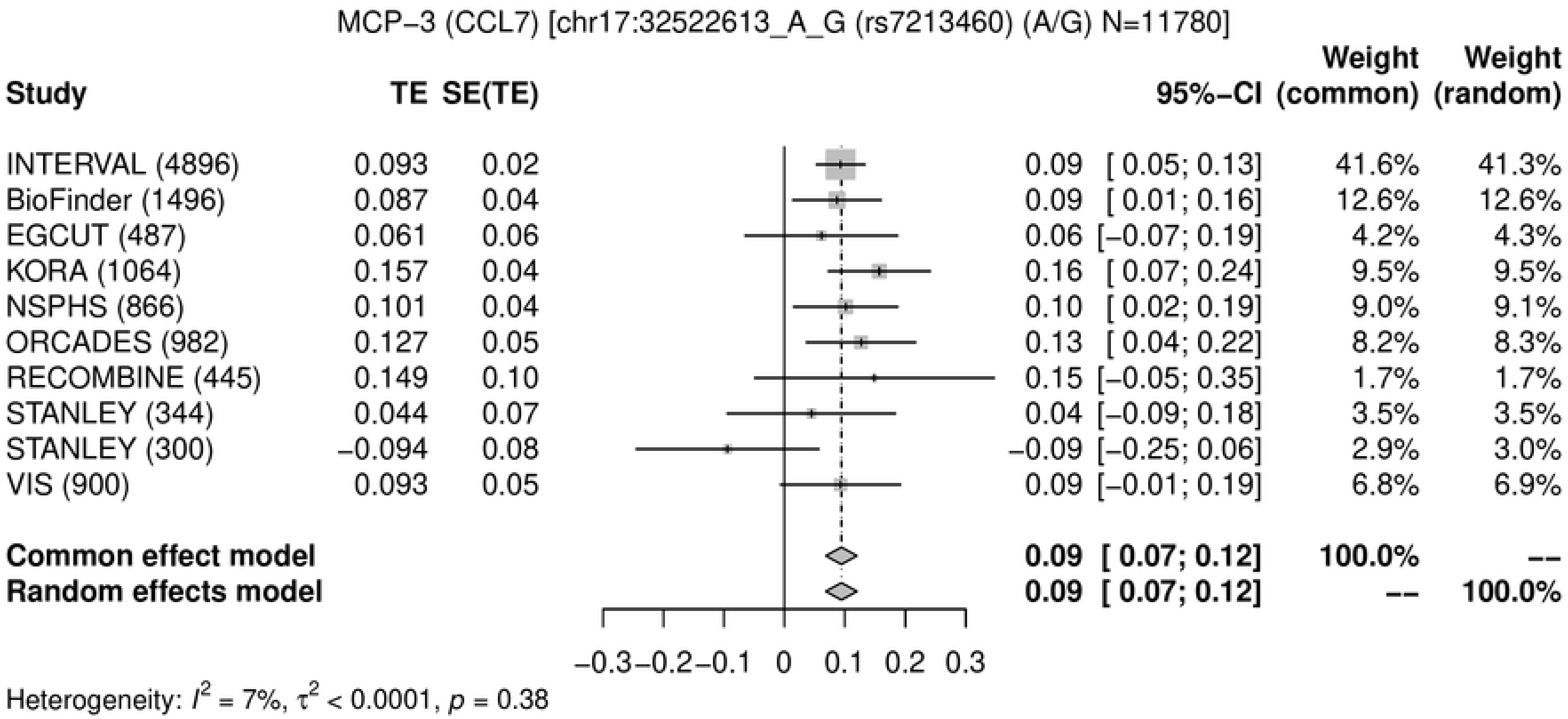
MCP−3 (CCL7) [chr17:32522613_A_G (rs7213460) (A/G) N=11780]
Study
TE
SE(TE)
95%−CI
Weight (common)
Weight (random)
INTERVAL (4896) 0.093 0.02 0.09 [ 0.05; 0.13] 41.6% 41.3%
BioFinder (1496) 0.087 0.04 0.09 [ 0.01; 0.16] 12.6% 12.6%
EGCUT (487) 0.061 0.06 0.06 [−0.07; 0.19] 4.2% 4.3%
KORA (1064) 0.157 0.04 0.16 [ 0.07; 0.24] 9.5% 9.5%
NSPHS (866) 0.101 0.04 0.10 [ 0.02; 0.19] 9.0% 9.1%
ORCADES (982) 0.127 0.05 0.13 [ 0.04; 0.22] 8.2% 8.3%
RECOMBINE (445) 0.149 0.10 0.15 [−0.05; 0.35] 1.7% 1.7%
STANLEY (344) 0.044 0.07 0.04 [−0.09; 0.18] 3.5% 3.5%
STANLEY (300) −0.094 0.08 −0.09 [−0.25; 0.06] 2.9% 3.0%
VIS (900) 0.093 0.05 0.09 [−0.01; 0.19] 6.8% 6.9%
Common effect model 0.09 [ 0.07; 0.12] 100.0% --
Random effects model 0.09 [ 0.07; 0.12] -- 100.0%
−0.3 −0.2 −0.1 0 0.1 0.2 0.3
Heterogeneity: $I^2$ = 7%, $\tau^2$ < 0.0001, $p$ = 0.38
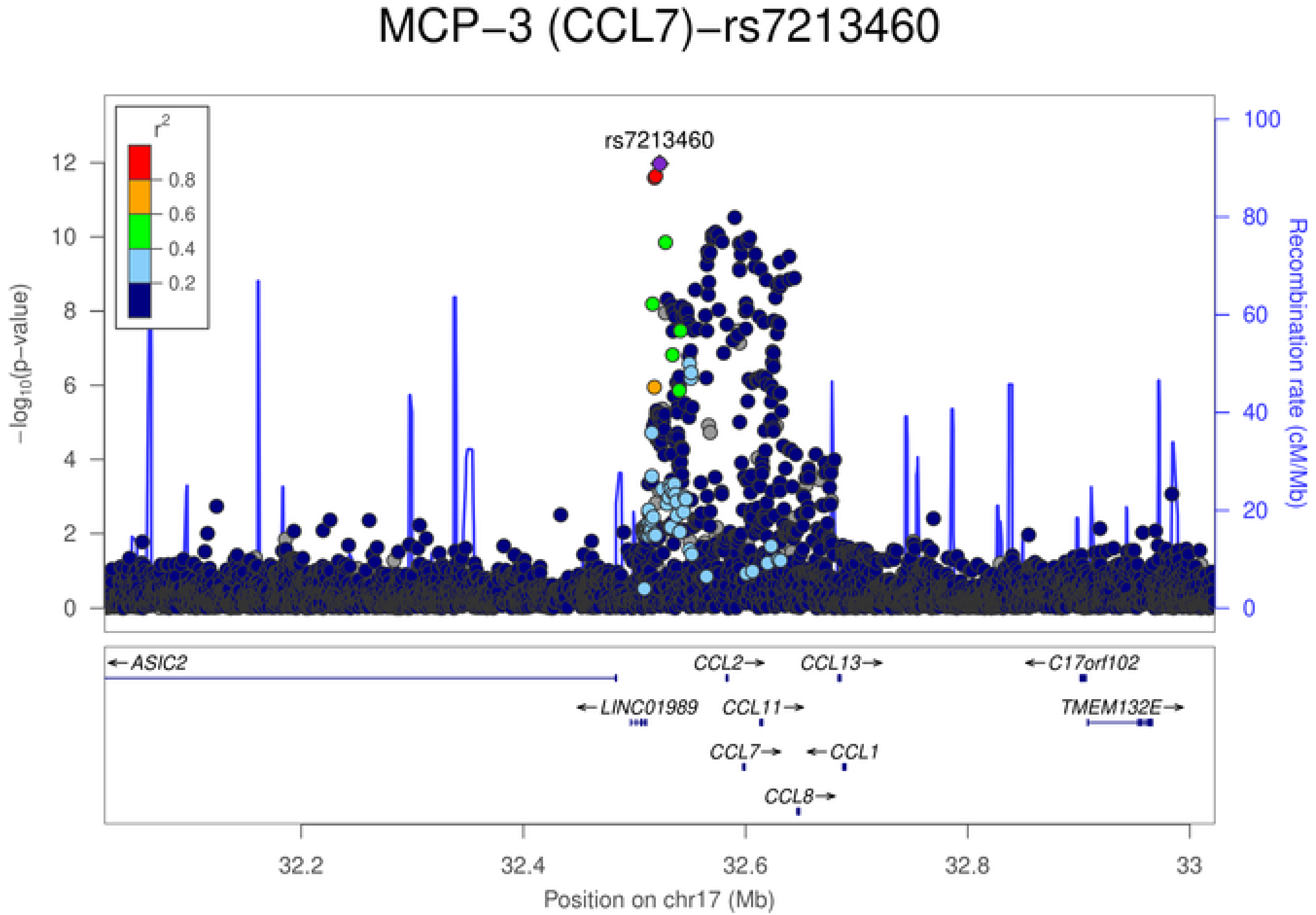
MCP−3 (CCL7)−rs7213460
rs7213460
$r^2$
0.8
0.6
0.4
0.2
$-\log_{10}$(p−value)
Recombination rate (cM/Mb)
←ASIC2
CCL2→
CCL13→
←C17orf102
←LINC01989
CCL11→
TMEM132E→
CCL7→
←CCL1
CCL8→
32.2
32.4
32.6
32.8
33
Position on chr17 (Mb)

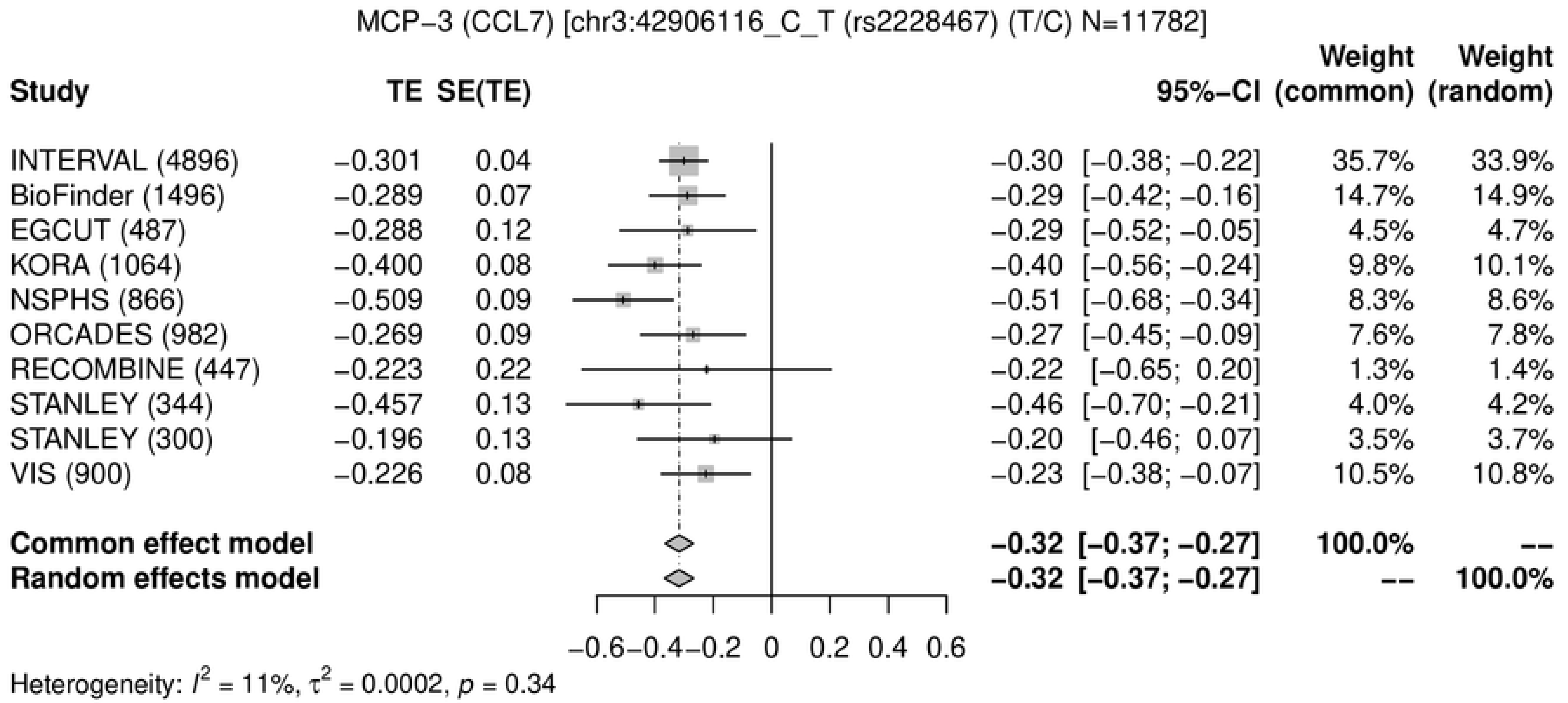
MCP−3 (CCL7) [chr3:42906116_C_T (rs2228467) (T/C) N=11782]
Study
TE
SE(TE)
95%−CI
Weight (common)
Weight (random)
INTERVAL (4896) −0.301 0.04 −0.30 [−0.38; −0.22] 35.7% 33.9%
BioFinder (1496) −0.289 0.07 −0.29 [−0.42; −0.16] 14.7% 14.9%
EGCUT (487) −0.288 0.12 −0.29 [−0.52; −0.05] 4.5% 4.7%
KORA (1064) −0.400 0.08 −0.40 [−0.56; −0.24] 9.8% 10.1%
NSPHS (866) −0.509 0.09 −0.51 [−0.68; −0.34] 8.3% 8.6%
ORCADES (982) −0.269 0.09 −0.27 [−0.45; −0.09] 7.6% 7.8%
RECOMBINE (447) −0.223 0.22 −0.22 [−0.65; 0.20] 1.3% 1.4%
STANLEY (344) −0.457 0.13 −0.46 [−0.70; −0.21] 4.0% 4.2%
STANLEY (300) −0.196 0.13 −0.20 [−0.46; 0.07] 3.5% 3.7%
VIS (900) −0.226 0.08 −0.23 [−0.38; −0.07] 10.5% 10.8%
Common effect model −0.32 [−0.37; −0.27] 100.0% --
Random effects model −0.32 [−0.37; −0.27] -- 100.0%
−0.6 −0.4 −0.2 0 0.2 0.4 0.6
Heterogeneity: $I^2$ = 11%, $\tau^2$ = 0.0002, $p$ = 0.34

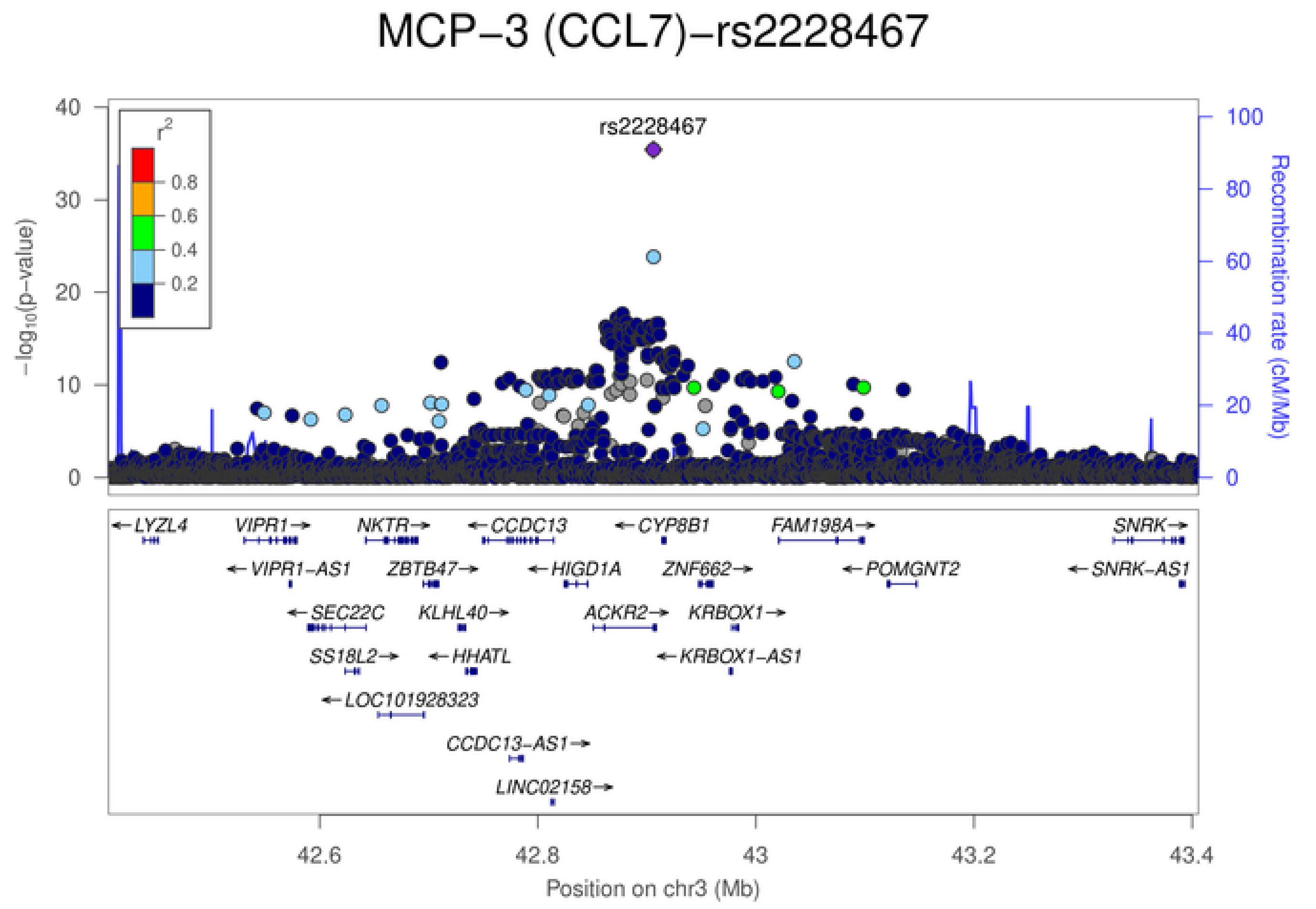
MCP−3 (CCL7)−rs2228467
rs2228467
$r^2$
0.8
0.6
0.4
0.2
$-\log_{10}$(p−value)
Recombination rate (cM/Mb)
LYZL4
VIPR1
NKTR
CCDC13
CYP8B1
FAM198A
SNRK
VIPR1−AS1
ZBTB47
HIGD1A
ZNF662
POMGNT2
SNRK−AS1
SEC22C
KLHL40
ACKR2
KRBOX1
SS18L2
HHATL
KRBOX1−AS1
LOC101928323
CCDC13−AS1
LINC02158
42.6
42.8
43
43.2
43.4
Position on chr3 (Mb)

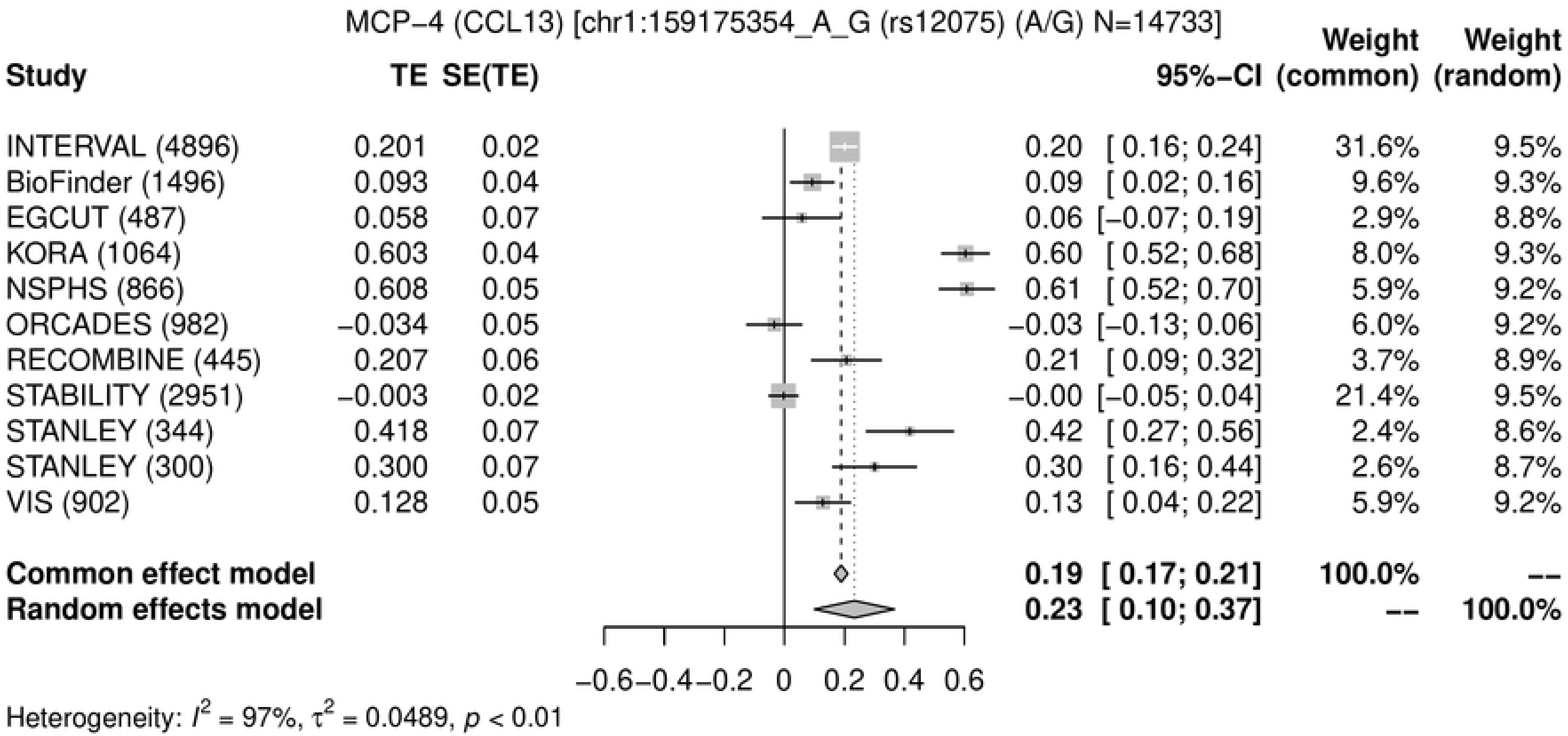
MCP−4 (CCL13) [chr1:159175354_A_G (rs12075) (A/G) N=14733]
Study
TE
SE(TE)
95%−CI
Weight (common)
Weight (random)
INTERVAL (4896) 0.201 0.02 0.20 [ 0.16; 0.24] 31.6% 9.5%
BioFinder (1496) 0.093 0.04 0.09 [ 0.02; 0.16] 9.6% 9.3%
EGCUT (487) 0.058 0.07 0.06 [−0.07; 0.19] 2.9% 8.8%
KORA (1064) 0.603 0.04 0.60 [ 0.52; 0.68] 8.0% 9.3%
NSPHS (866) 0.608 0.05 0.61 [ 0.52; 0.70] 5.9% 9.2%
ORCADES (982) −0.034 0.05 −0.03 [−0.13; 0.06] 6.0% 9.2%
RECOMBINE (445) 0.207 0.06 0.21 [ 0.09; 0.32] 3.7% 8.9%
STABILITY (2951) −0.003 0.02 −0.00 [−0.05; 0.04] 21.4% 9.5%
STANLEY (344) 0.418 0.07 0.42 [ 0.27; 0.56] 2.4% 8.6%
STANLEY (300) 0.300 0.07 0.30 [ 0.16; 0.44] 2.6% 8.7%
VIS (902) 0.128 0.05 0.13 [ 0.04; 0.22] 5.9% 9.2%
Common effect model 0.19 [ 0.17; 0.21] 100.0% --
Random effects model 0.23 [ 0.10; 0.37] -- 100.0%
−0.6 −0.4 −0.2 0 0.2 0.4 0.6
Heterogeneity: $I^2$ = 97%, $\tau^2$ = 0.0489, $p$ < 0.01

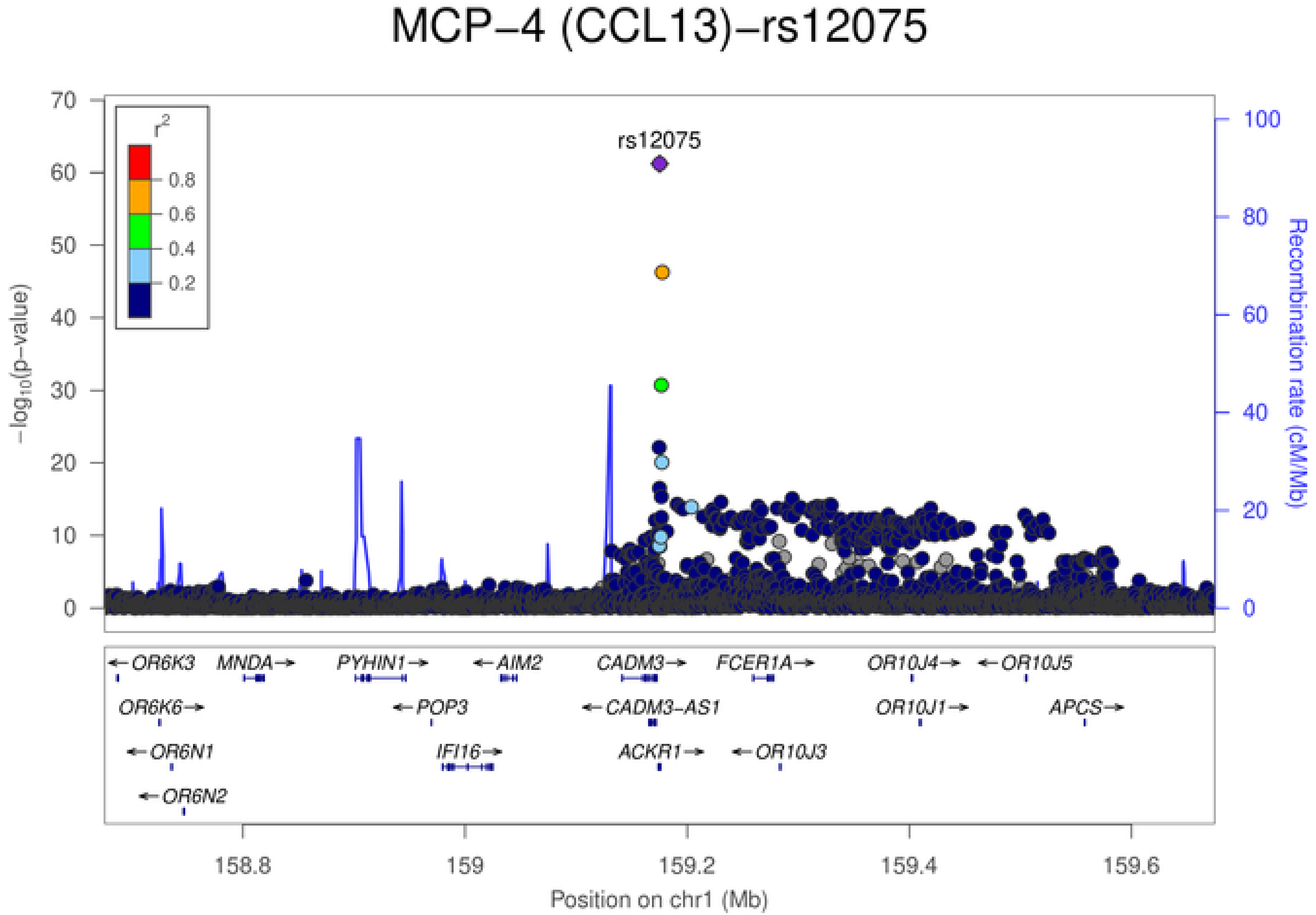
MCP−4 (CCL13)−rs12075
rs12075
$r^2$
0.8
0.6
0.4
0.2
$-\log_{10}$(p−value)
0
10
20
30
40
50
60
70
Recombination rate (cM/Mb)
0
20
40
60
80
100
←OR6K3
MNDA→
PYHIN1→
←AIM2
CADM3→
FCER1A→
OR10J4→
←OR10J5
OR6K6→
←POP3
←CADM3-AS1
OR10J1→
APCS→
←OR6N1
IFI16→
ACKR1→
←OR10J3
←OR6N2
158.8
159
159.2
159.4
159.6
Position on chr1 (Mb)

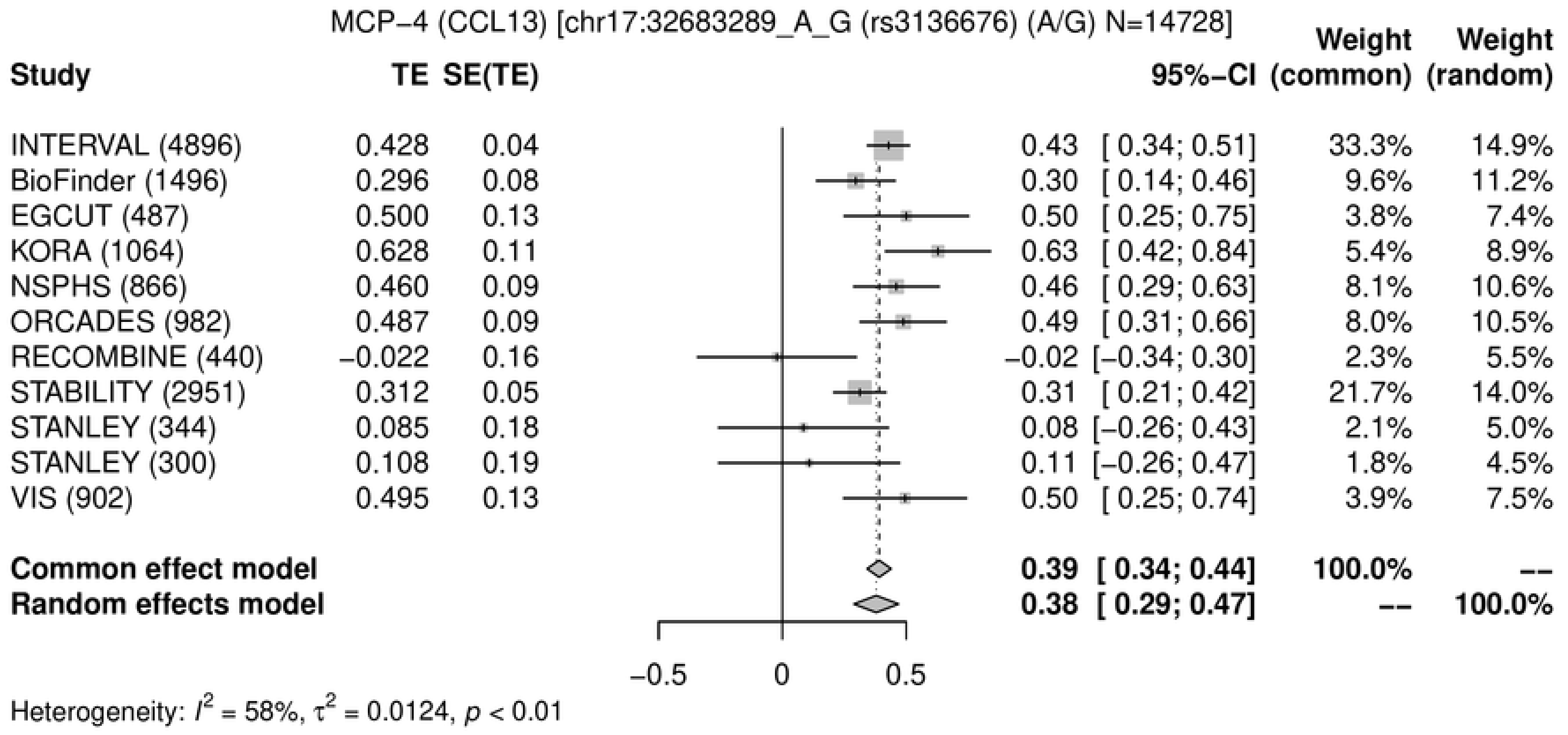
MCP−4 (CCL13) [chr17:32683289_A_G (rs3136676) (A/G) N=14728]
Study TE SE(TE) 95%−CI Weight (common) Weight (random)
INTERVAL (4896) 0.428 0.04 0.43 [ 0.34; 0.51] 33.3% 14.9%
BioFinder (1496) 0.296 0.08 0.30 [ 0.14; 0.46] 9.6% 11.2%
EGCUT (487) 0.500 0.13 0.50 [ 0.25; 0.75] 3.8% 7.4%
KORA (1064) 0.628 0.11 0.63 [ 0.42; 0.84] 5.4% 8.9%
NSPHS (866) 0.460 0.09 0.46 [ 0.29; 0.63] 8.1% 10.6%
ORCADES (982) 0.487 0.09 0.49 [ 0.31; 0.66] 8.0% 10.5%
RECOMBINE (440) −0.022 0.16 −0.02 [−0.34; 0.30] 2.3% 5.5%
STABILITY (2951) 0.312 0.05 0.31 [ 0.21; 0.42] 21.7% 14.0%
STANLEY (344) 0.085 0.18 0.08 [−0.26; 0.43] 2.1% 5.0%
STANLEY (300) 0.108 0.19 0.11 [−0.26; 0.47] 1.8% 4.5%
VIS (902) 0.495 0.13 0.50 [ 0.25; 0.74] 3.9% 7.5%
Common effect model 0.39 [ 0.34; 0.44] 100.0% --
Random effects model 0.38 [ 0.29; 0.47] -- 100.0%
−0.5 0 0.5
Heterogeneity: I2 = 58%, τ2 = 0.0124, p < 0.01

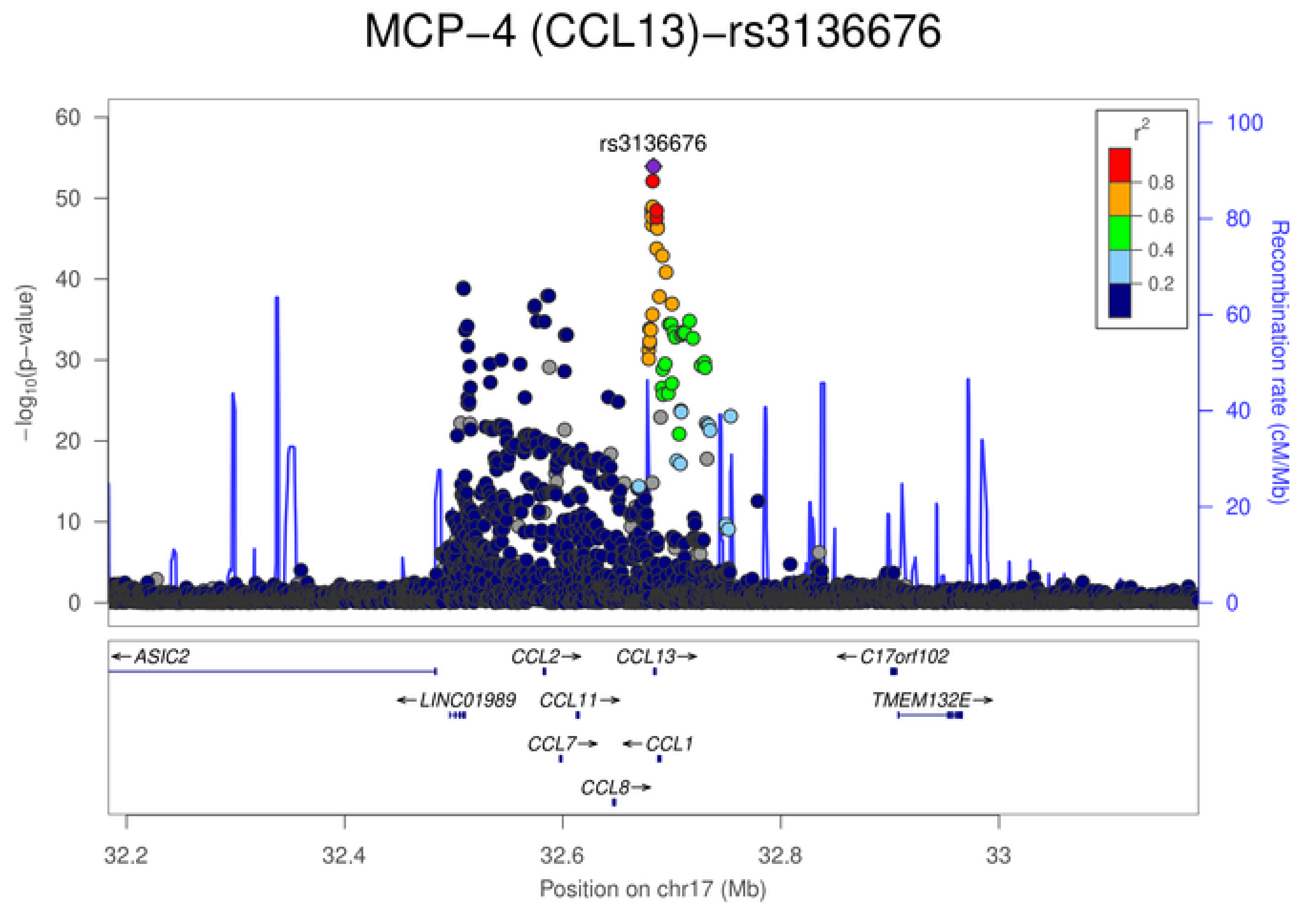
MCP−4 (CCL13)−rs3136676
rs3136676
−log10(p−value)
Recombination rate (cM/Mb)
r2
0.8
0.6
0.4
0.2
ASIC2
CCL2
CCL13
C17orf102
LINC01989
CCL11
TMEM132E
CCL7
CCL1
CCL8
32.2
32.4
32.6
32.8
33
Position on chr17 (Mb)

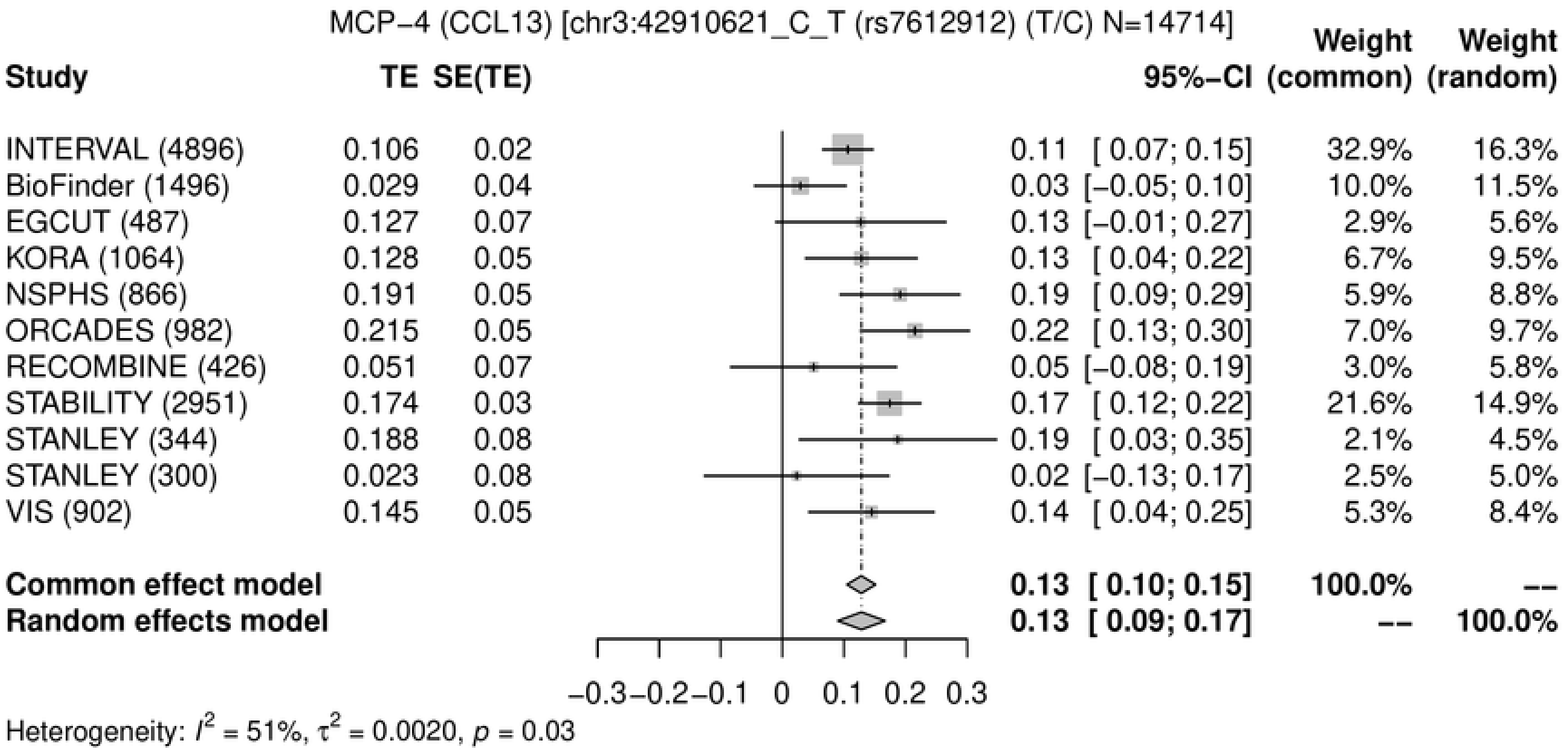
MCP−4 (CCL13) [chr3:42910621_C_T (rs7612912) (T/C) N=14714]
Study
TE
SE(TE)
95%−CI
Weight (common)
Weight (random)
INTERVAL (4896) 0.106 0.02 0.11 [ 0.07; 0.15] 32.9% 16.3%
BioFinder (1496) 0.029 0.04 0.03 [−0.05; 0.10] 10.0% 11.5%
EGCUT (487) 0.127 0.07 0.13 [−0.01; 0.27] 2.9% 5.6%
KORA (1064) 0.128 0.05 0.13 [ 0.04; 0.22] 6.7% 9.5%
NSPHS (866) 0.191 0.05 0.19 [ 0.09; 0.29] 5.9% 8.8%
ORCADES (982) 0.215 0.05 0.22 [ 0.13; 0.30] 7.0% 9.7%
RECOMBINE (426) 0.051 0.07 0.05 [−0.08; 0.19] 3.0% 5.8%
STABILITY (2951) 0.174 0.03 0.17 [ 0.12; 0.22] 21.6% 14.9%
STANLEY (344) 0.188 0.08 0.19 [ 0.03; 0.35] 2.1% 4.5%
STANLEY (300) 0.023 0.08 0.02 [−0.13; 0.17] 2.5% 5.0%
VIS (902) 0.145 0.05 0.14 [ 0.04; 0.25] 5.3% 8.4%
Common effect model 0.13 [ 0.10; 0.15] 100.0% --
Random effects model 0.13 [ 0.09; 0.17] -- 100.0%
−0.3 −0.2 −0.1 0 0.1 0.2 0.3
Heterogeneity: $I^2$ = 51%, $\tau^2$ = 0.0020, $p$ = 0.03

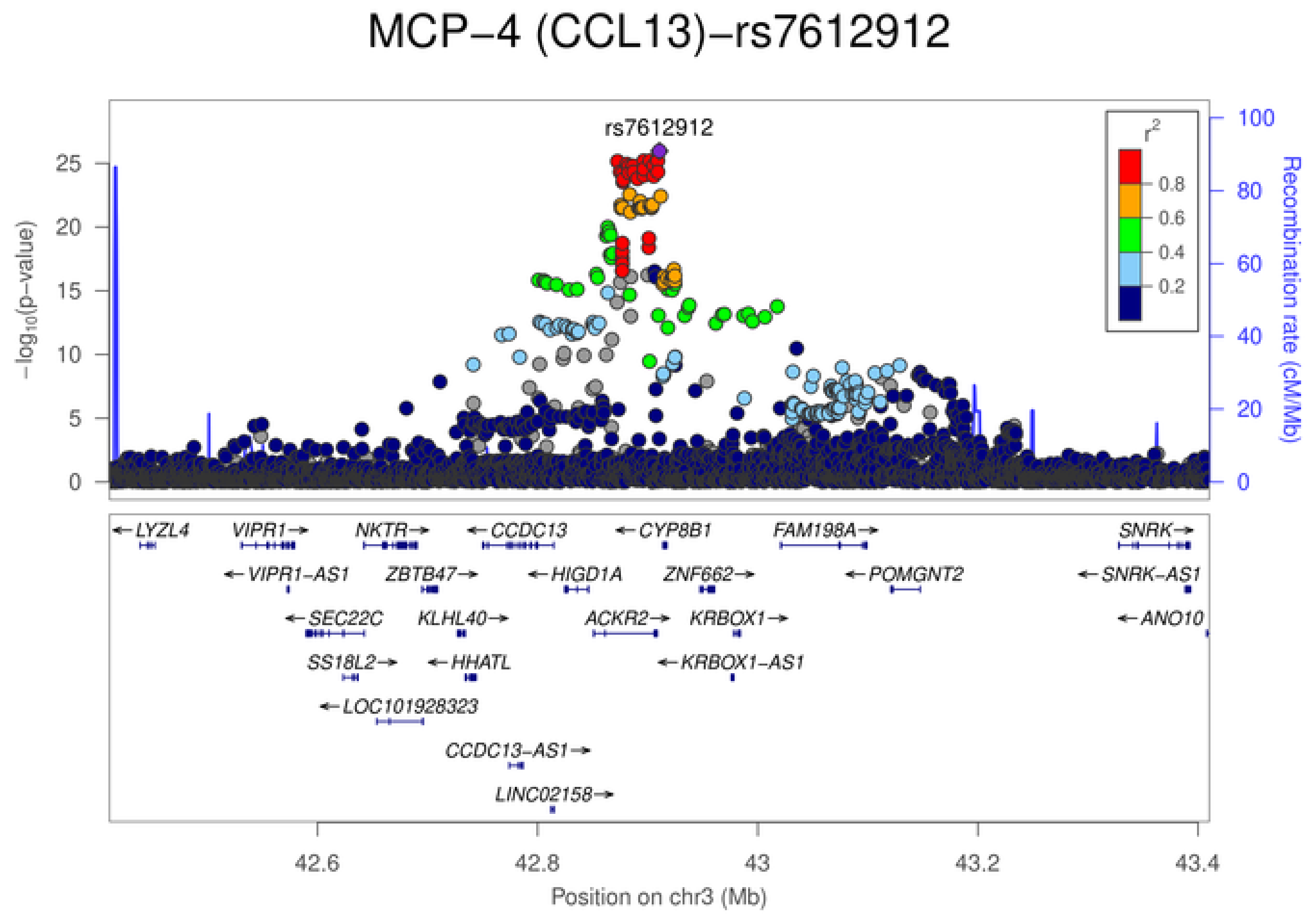
MCP−4 (CCL13)−rs7612912
rs7612912
$-\log_{10}$(p−value)
Recombination rate (cM/Mb)
$r^2$
0.8
0.6
0.4
0.2
LYZL4
VIPR1
NKTR
CCDC13
CYP8B1
FAM198A
SNRK
VIPR1−AS1
ZBTB47
HIGD1A
ZNF662
POMGNT2
SNRK−AS1
SEC22C
KLHL40
ACKR2
KRBOX1
ANO10
SS18L2
HHATL
KRBOX1−AS1
LOC101928323
CCDC13−AS1
LINC02158
42.6
42.8
43
43.2
43.4
Position on chr3 (Mb)

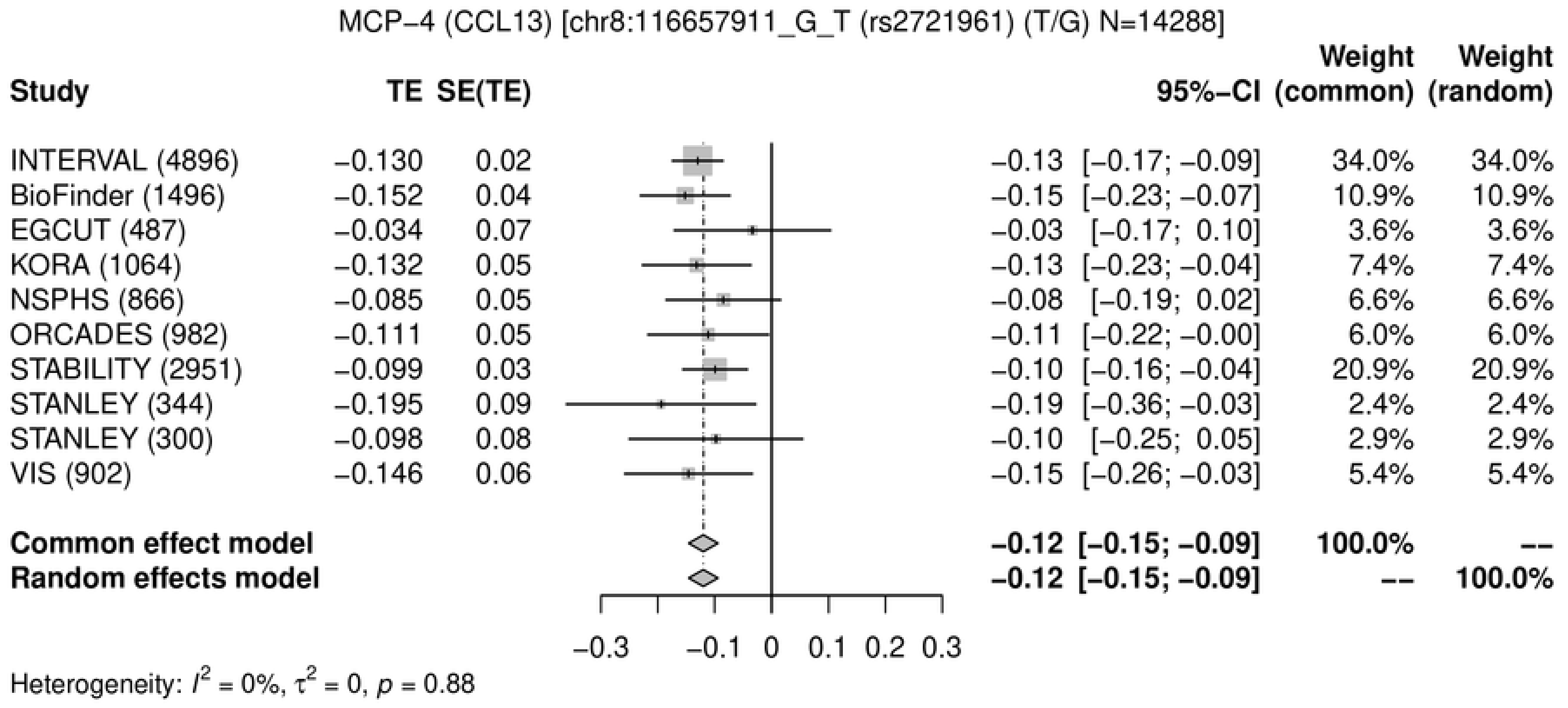
MCP−4 (CCL13) [chr8:116657911_G_T (rs2721961) (T/G) N=14288]
Study
TE
SE(TE)
95%−CI
Weight (common)
Weight (random)
INTERVAL (4896) −0.130 0.02 −0.13 [−0.17; −0.09] 34.0% 34.0%
BioFinder (1496) −0.152 0.04 −0.15 [−0.23; −0.07] 10.9% 10.9%
EGCUT (487) −0.034 0.07 −0.03 [−0.17; 0.10] 3.6% 3.6%
KORA (1064) −0.132 0.05 −0.13 [−0.23; −0.04] 7.4% 7.4%
NSPHS (866) −0.085 0.05 −0.08 [−0.19; 0.02] 6.6% 6.6%
ORCADES (982) −0.111 0.05 −0.11 [−0.22; −0.00] 6.0% 6.0%
STABILITY (2951) −0.099 0.03 −0.10 [−0.16; −0.04] 20.9% 20.9%
STANLEY (344) −0.195 0.09 −0.19 [−0.36; −0.03] 2.4% 2.4%
STANLEY (300) −0.098 0.08 −0.10 [−0.25; 0.05] 2.9% 2.9%
VIS (902) −0.146 0.06 −0.15 [−0.26; −0.03] 5.4% 5.4%
Common effect model −0.12 [−0.15; −0.09] 100.0% --
Random effects model −0.12 [−0.15; −0.09] -- 100.0%
−0.3 −0.1 0 0.1 0.2 0.3
Heterogeneity: $I^2$ = 0%, $\tau^2$ = 0, $p$ = 0.88

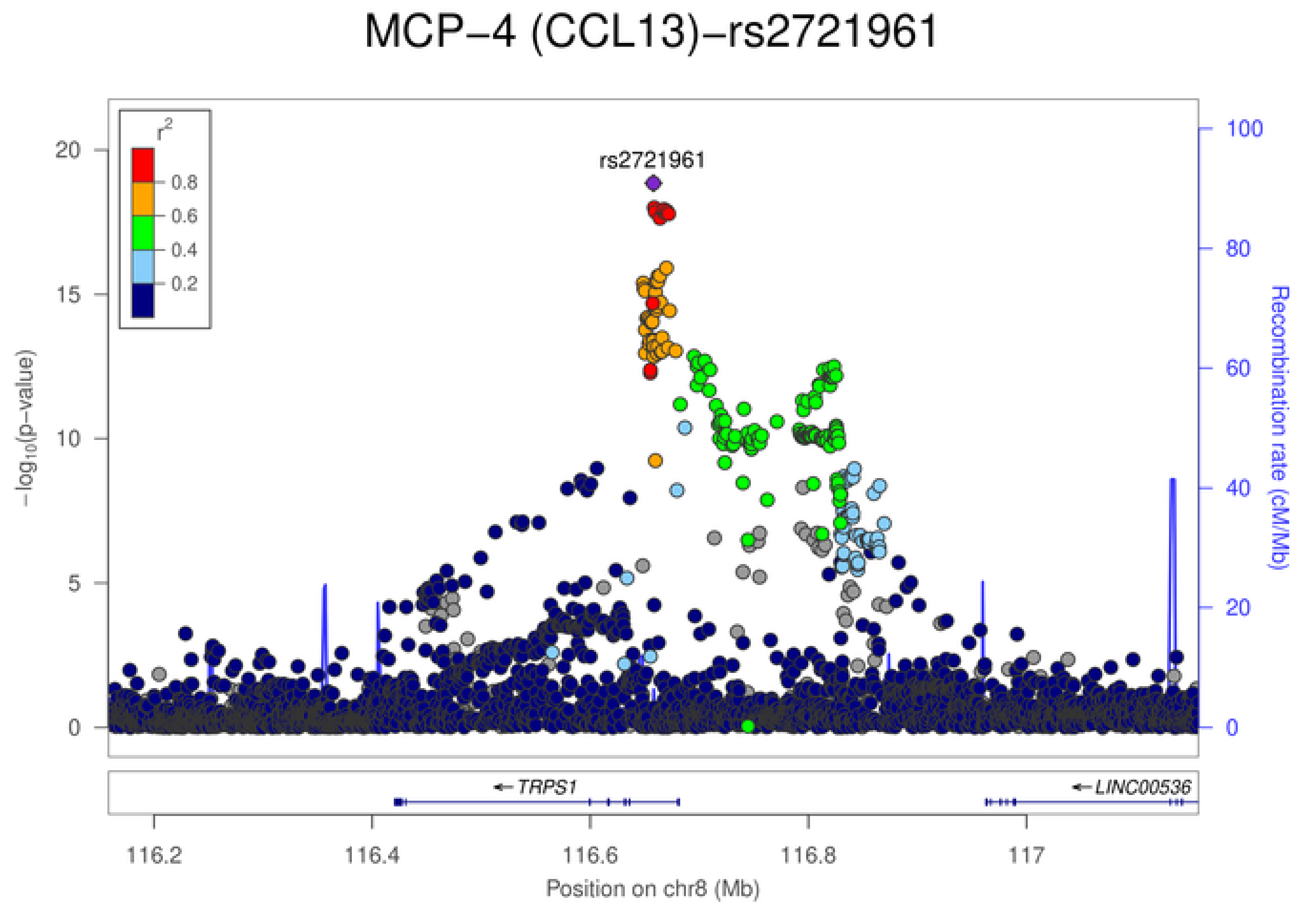
MCP−4 (CCL13)−rs2721961
rs2721961
$r^2$
0.8
0.6
0.4
0.2
20
15
10
5
0
$-\log_{10}$(p−value)
100
80
60
40
20
0
Recombination rate (cM/Mb)
TRPS1
LINC00536
116.2
116.4
116.6
116.8
117
Position on chr8 (Mb)

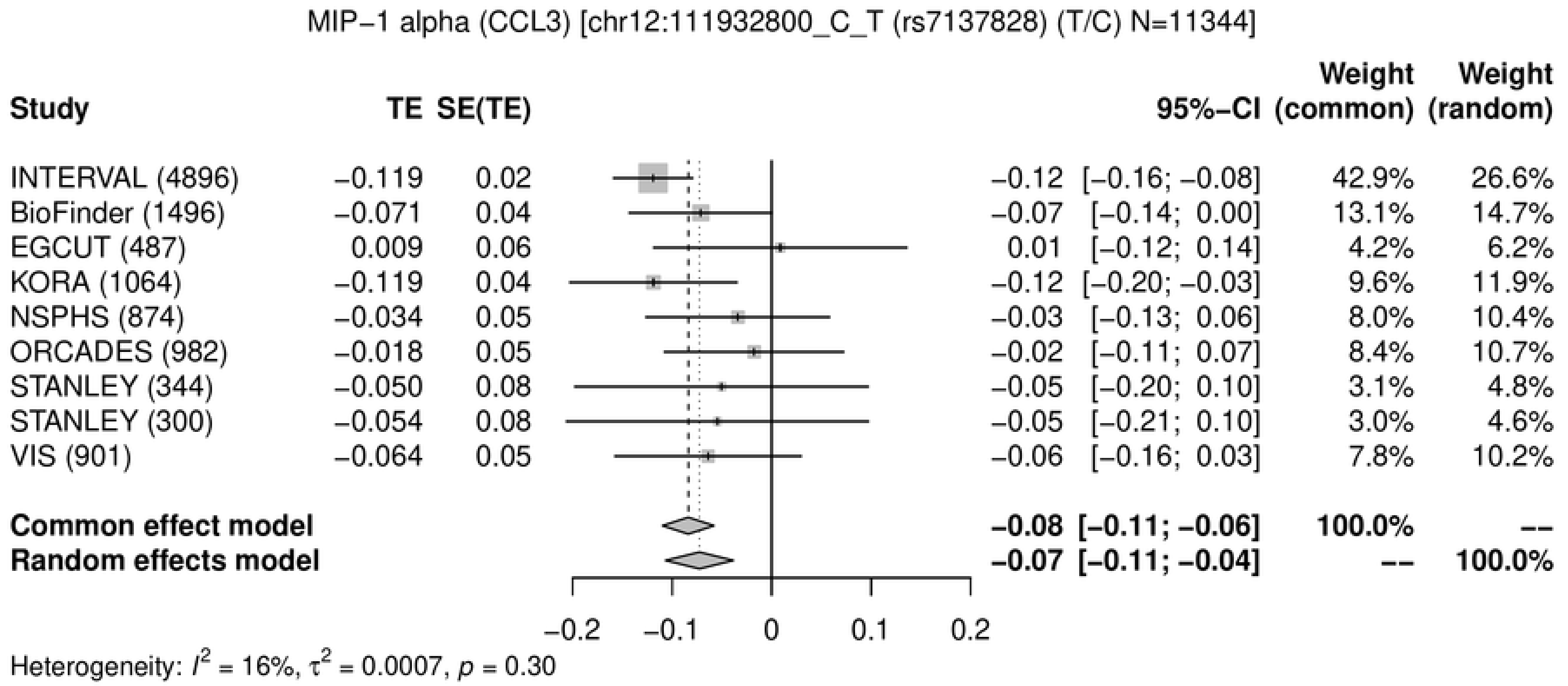
MIP−1 alpha (CCL3) [chr12:111932800_C_T (rs7137828) (T/C) N=11344]
Study
TE
SE(TE)
95%−CI
Weight (common)
Weight (random)
INTERVAL (4896) −0.119 0.02 −0.12 [−0.16; −0.08] 42.9% 26.6%
BioFinder (1496) −0.071 0.04 −0.07 [−0.14; 0.00] 13.1% 14.7%
EGCUT (487) 0.009 0.06 0.01 [−0.12; 0.14] 4.2% 6.2%
KORA (1064) −0.119 0.04 −0.12 [−0.20; −0.03] 9.6% 11.9%
NSPHS (874) −0.034 0.05 −0.03 [−0.13; 0.06] 8.0% 10.4%
ORCADES (982) −0.018 0.05 −0.02 [−0.11; 0.07] 8.4% 10.7%
STANLEY (344) −0.050 0.08 −0.05 [−0.20; 0.10] 3.1% 4.8%
STANLEY (300) −0.054 0.08 −0.05 [−0.21; 0.10] 3.0% 4.6%
VIS (901) −0.064 0.05 −0.06 [−0.16; 0.03] 7.8% 10.2%
Common effect model −0.08 [−0.11; −0.06] 100.0% --
Random effects model −0.07 [−0.11; −0.04] -- 100.0%
−0.2 −0.1 0 0.1 0.2
Heterogeneity: $I^2$ = 16%, $\tau^2$ = 0.0007, $p$ = 0.30

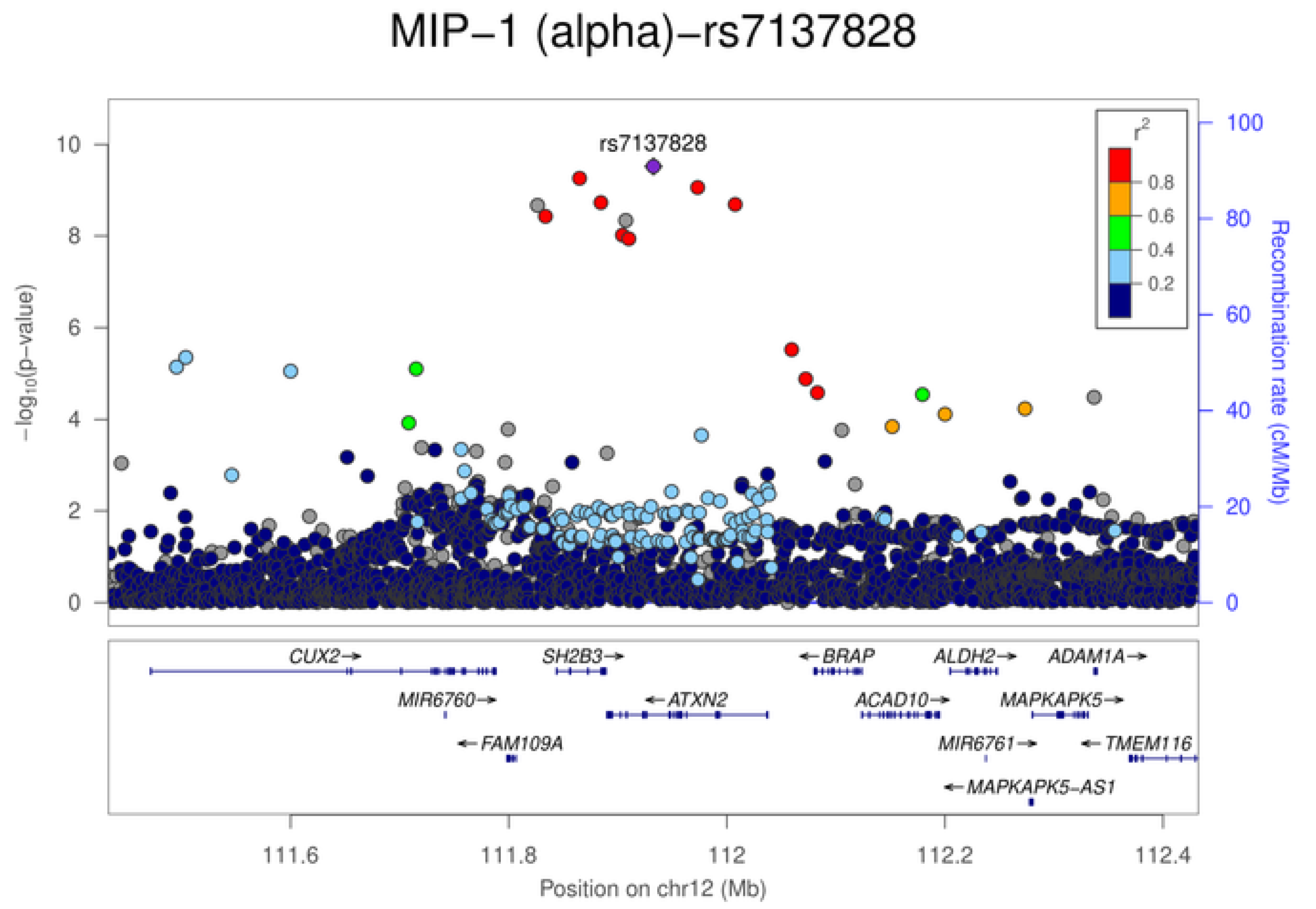
MIP−1 (alpha)−rs7137828
rs7137828
$-\log_{10}$(p−value)
Recombination rate (cM/Mb)
$r^2$
0.8
0.6
0.4
0.2
CUX2
SH2B3
BRAP
ALDH2
ADAM1A
MIR6760
ATXN2
ACAD10
MAPKAPK5
FAM109A
MIR6761
TMEM116
MAPKAPK5−AS1
111.6
111.8
112
112.2
112.4
Position on chr12 (Mb)

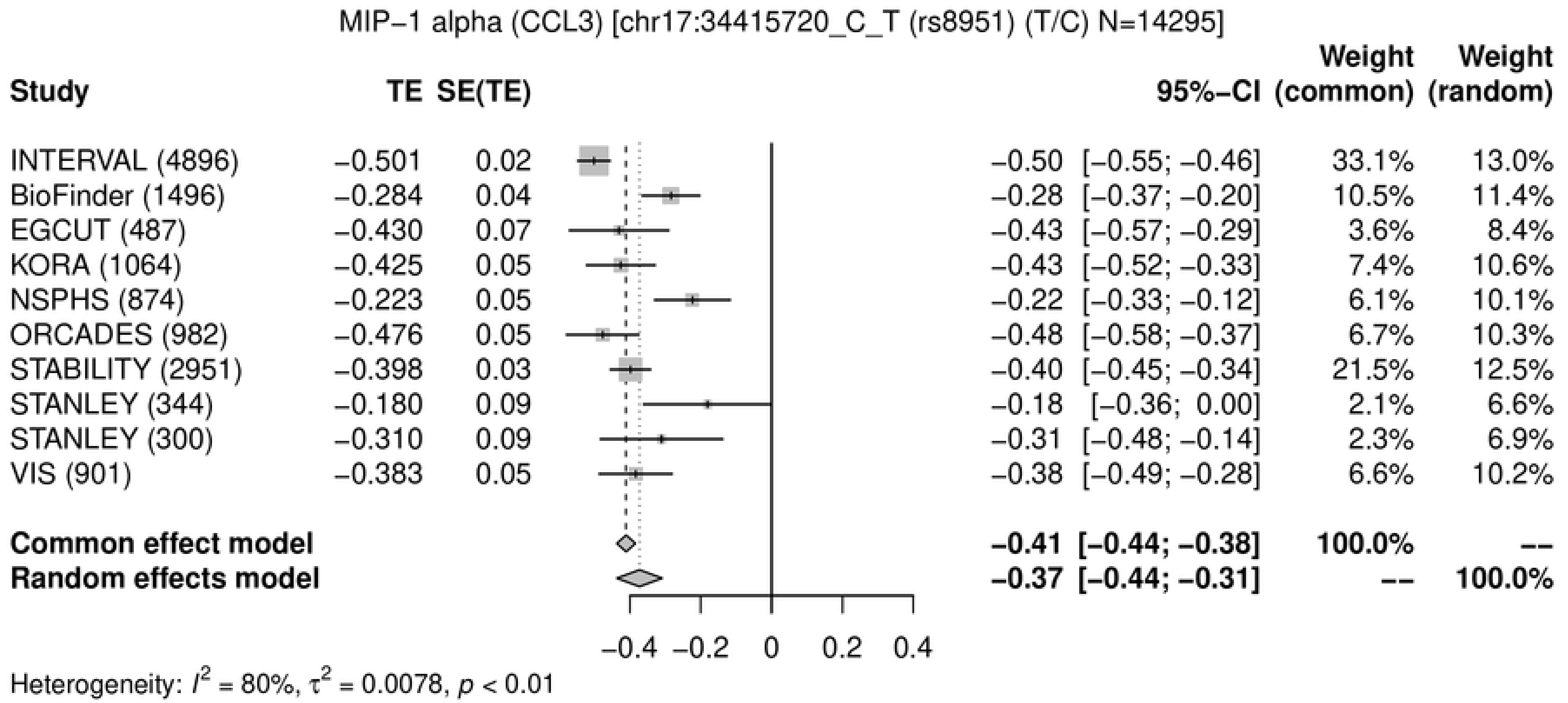
MIP−1 alpha (CCL3) [chr17:34415720_C_T (rs8951) (T/C) N=14295]
Study TE SE(TE) 95%−CI Weight (common) Weight (random)
INTERVAL (4896) −0.501 0.02 −0.50 [−0.55; −0.46] 33.1% 13.0%
BioFinder (1496) −0.284 0.04 −0.28 [−0.37; −0.20] 10.5% 11.4%
EGCUT (487) −0.430 0.07 −0.43 [−0.57; −0.29] 3.6% 8.4%
KORA (1064) −0.425 0.05 −0.43 [−0.52; −0.33] 7.4% 10.6%
NSPHS (874) −0.223 0.05 −0.22 [−0.33; −0.12] 6.1% 10.1%
ORCADES (982) −0.476 0.05 −0.48 [−0.58; −0.37] 6.7% 10.3%
STABILITY (2951) −0.398 0.03 −0.40 [−0.45; −0.34] 21.5% 12.5%
STANLEY (344) −0.180 0.09 −0.18 [−0.36; 0.00] 2.1% 6.6%
STANLEY (300) −0.310 0.09 −0.31 [−0.48; −0.14] 2.3% 6.9%
VIS (901) −0.383 0.05 −0.38 [−0.49; −0.28] 6.6% 10.2%
Common effect model −0.41 [−0.44; −0.38] 100.0% --
Random effects model −0.37 [−0.44; −0.31] -- 100.0%
−0.4 −0.2 0 0.2 0.4
Heterogeneity: $I^2$ = 80%, $\tau^2$ = 0.0078, $p$ < 0.01

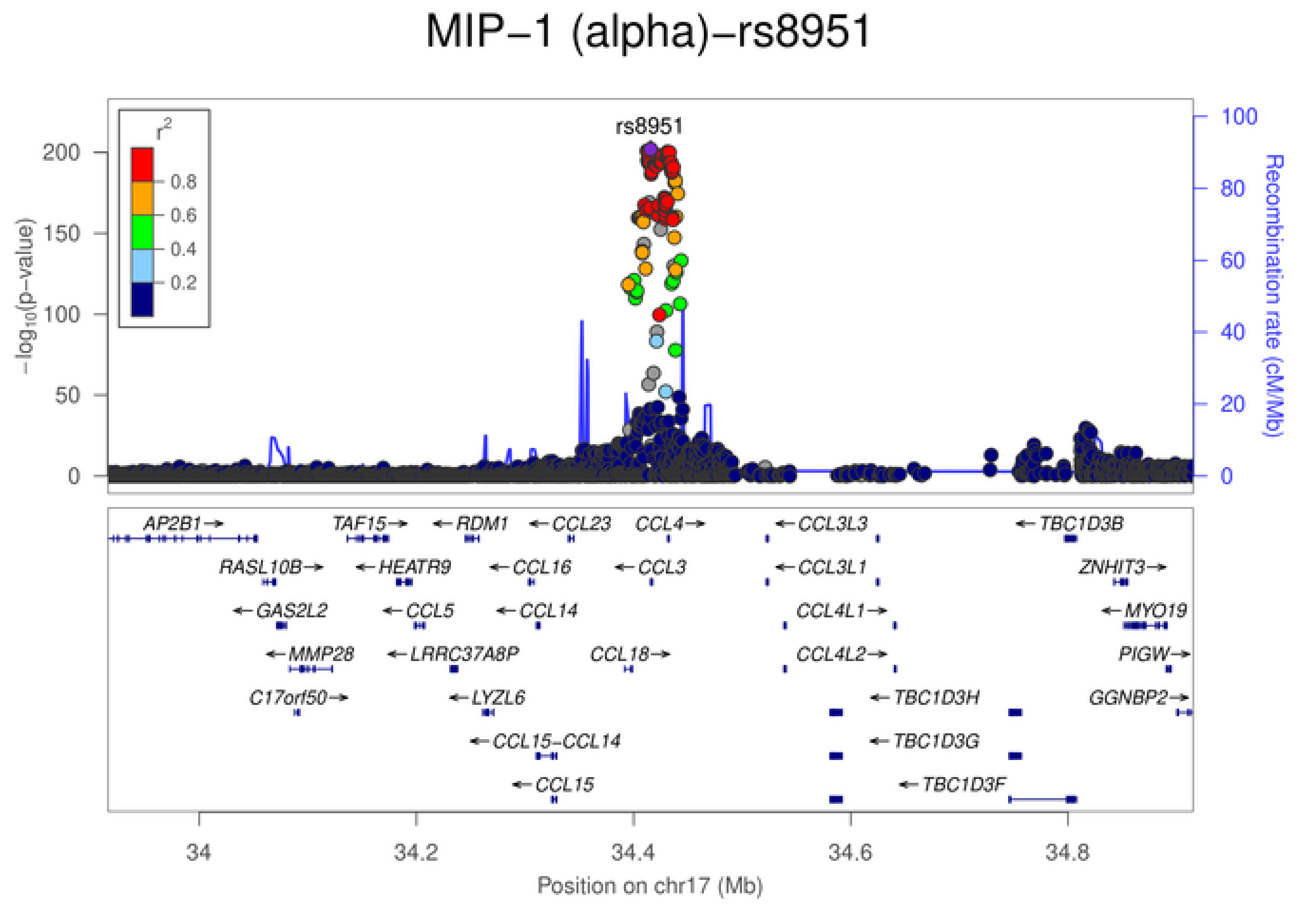
MIP−1 (alpha)−rs8951
rs8951
$r^2$
0.8
0.6
0.4
0.2
$-\log_{10}$(p−value)
Recombination rate (cM/Mb)
AP2B1 TAF15 RDM1 CCL23 CCL4 CCL3L3 TBC1D3B
RASL10B HEATR9 CCL16 CCL3 CCL3L1 ZNHIT3
GAS2L2 CCL5 CCL14 CCL4L1 MYO19
MMP28 LRRC37A8P CCL18 CCL4L2 PIGW
C17orf50 LYZL6 TBC1D3H GGNBP2
CCL15−CCL14 TBC1D3G
CCL15 TBC1D3F
34 34.2 34.4 34.6 34.8
Position on chr17 (Mb)

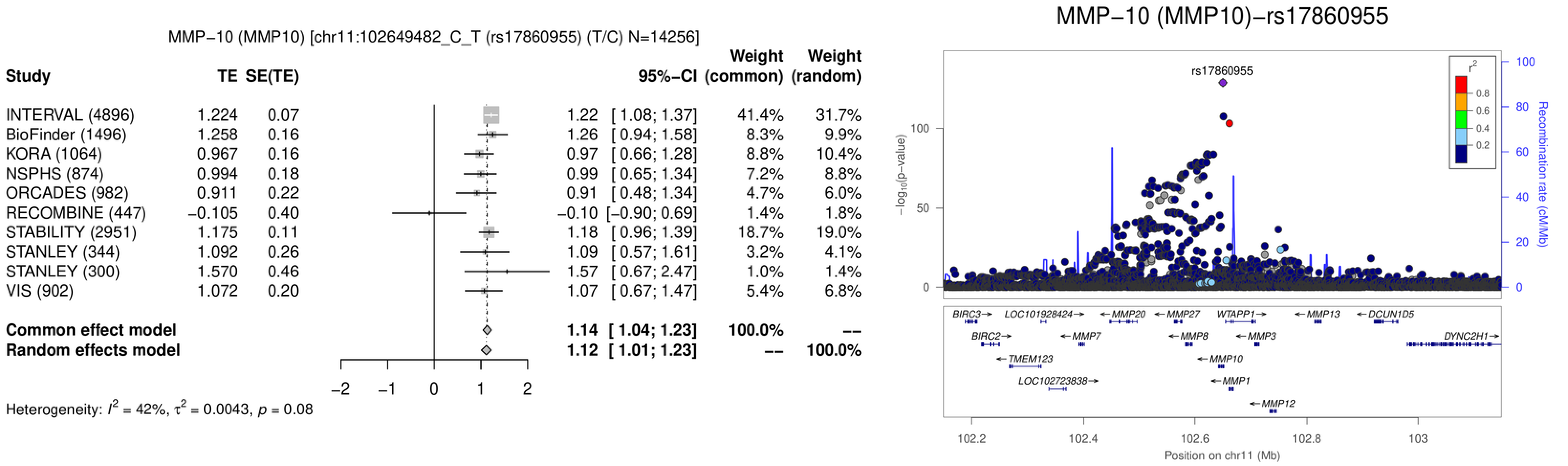
MMP−10 (MMP10) [chr11:102649482_C_T (rs17860955) (T/C) N=14256]
Study
TE
SE(TE)
95%−CI
Weight (common)
Weight (random)
INTERVAL (4896) 1.224 0.07 1.22 [ 1.08; 1.37] 41.4% 31.7%
BioFinder (1496) 1.258 0.16 1.26 [ 0.94; 1.58] 8.3% 9.9%
KORA (1064) 0.967 0.16 0.97 [ 0.66; 1.28] 8.8% 10.4%
NSPHS (874) 0.994 0.18 0.99 [ 0.65; 1.34] 7.2% 8.8%
ORCADES (982) 0.911 0.22 0.91 [ 0.48; 1.34] 4.7% 6.0%
RECOMBINE (447) −0.105 0.40 −0.10 [−0.90; 0.69] 1.4% 1.8%
STABILITY (2951) 1.175 0.11 1.18 [ 0.96; 1.39] 18.7% 19.0%
STANLEY (344) 1.092 0.26 1.09 [ 0.57; 1.61] 3.2% 4.1%
STANLEY (300) 1.570 0.46 1.57 [ 0.67; 2.47] 1.0% 1.4%
VIS (902) 1.072 0.20 1.07 [ 0.67; 1.47] 5.4% 6.8%
Common effect model 1.14 [ 1.04; 1.23] 100.0% --
Random effects model 1.12 [ 1.01; 1.23] -- 100.0%
−2 −1 0 1 2
Heterogeneity: $I^2$ = 42%, $\tau^2$ = 0.0043, $p$ = 0.08
MMP−10 (MMP10)−rs17860955
rs17860955
$-\log_{10}$(p−value)
0
50
100
Recombination rate (cM/Mb)
0
20
40
60
80
100
$r^2$
0.8
0.6
0.4
0.2
BIRC3→
LOC101928424→
←MMP20
←MMP27
WTAPP1→
←MMP13
←DCUN1D5
BIRC2→
←MMP7
←MMP8
←MMP3
DYNC2H1→
←TMEM123
←MMP10
LOC102723838→
←MMP1
←MMP12
102.2
102.4
102.6
102.8
103
Position on chr11 (Mb)

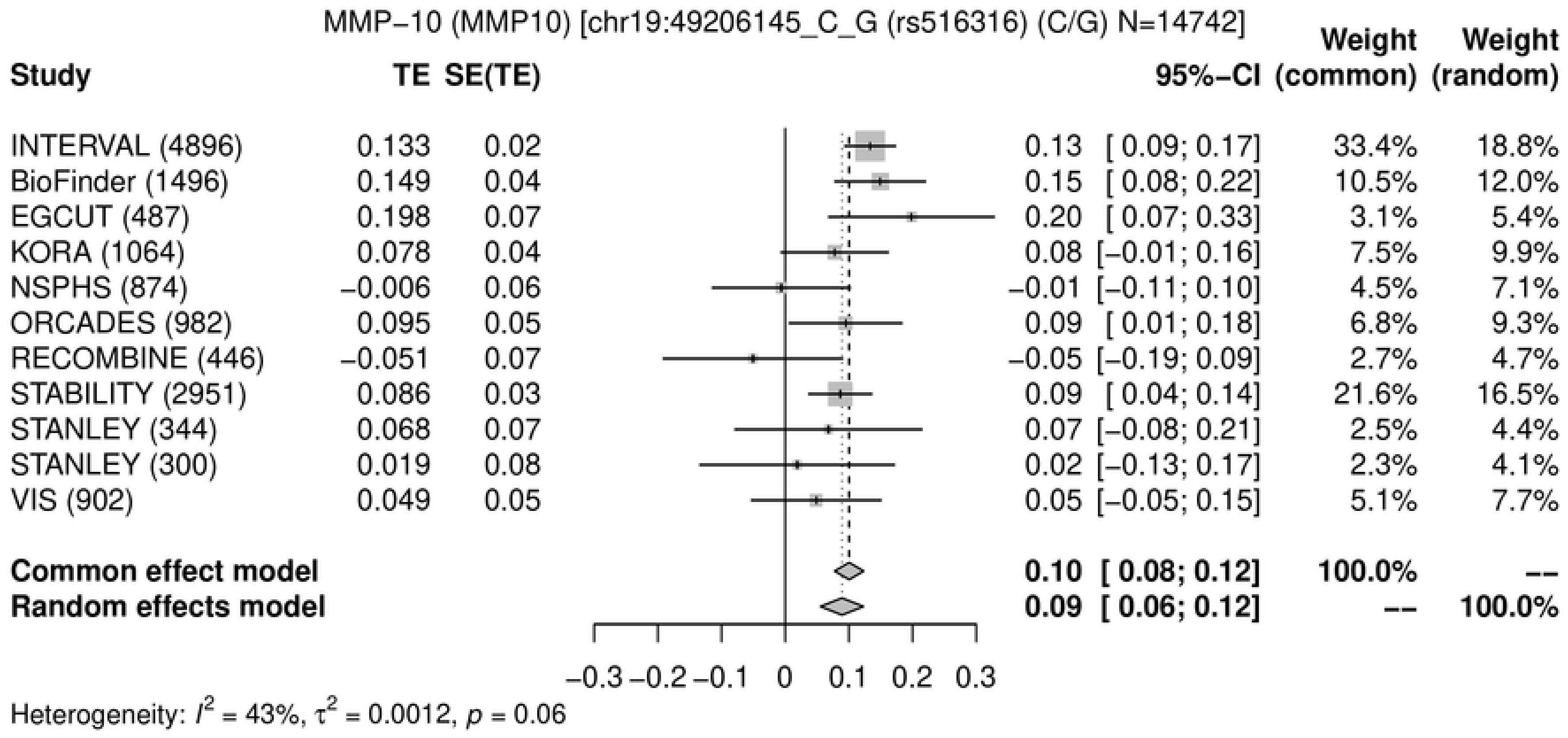
MMP−10 (MMP10) [chr19:49206145_C_G (rs516316) (C/G) N=14742]
Study
TE
SE(TE)
95%−CI
Weight (common)
Weight (random)
INTERVAL (4896) 0.133 0.02 0.13 [ 0.09; 0.17] 33.4% 18.8%
BioFinder (1496) 0.149 0.04 0.15 [ 0.08; 0.22] 10.5% 12.0%
EGCUT (487) 0.198 0.07 0.20 [ 0.07; 0.33] 3.1% 5.4%
KORA (1064) 0.078 0.04 0.08 [−0.01; 0.16] 7.5% 9.9%
NSPHS (874) −0.006 0.06 −0.01 [−0.11; 0.10] 4.5% 7.1%
ORCADES (982) 0.095 0.05 0.09 [ 0.01; 0.18] 6.8% 9.3%
RECOMBINE (446) −0.051 0.07 −0.05 [−0.19; 0.09] 2.7% 4.7%
STABILITY (2951) 0.086 0.03 0.09 [ 0.04; 0.14] 21.6% 16.5%
STANLEY (344) 0.068 0.07 0.07 [−0.08; 0.21] 2.5% 4.4%
STANLEY (300) 0.019 0.08 0.02 [−0.13; 0.17] 2.3% 4.1%
VIS (902) 0.049 0.05 0.05 [−0.05; 0.15] 5.1% 7.7%
Common effect model 0.10 [ 0.08; 0.12] 100.0% --
Random effects model 0.09 [ 0.06; 0.12] -- 100.0%
−0.3 −0.2 −0.1 0 0.1 0.2 0.3
Heterogeneity: $I^2$ = 43%, $\tau^2$ = 0.0012, $p$ = 0.06

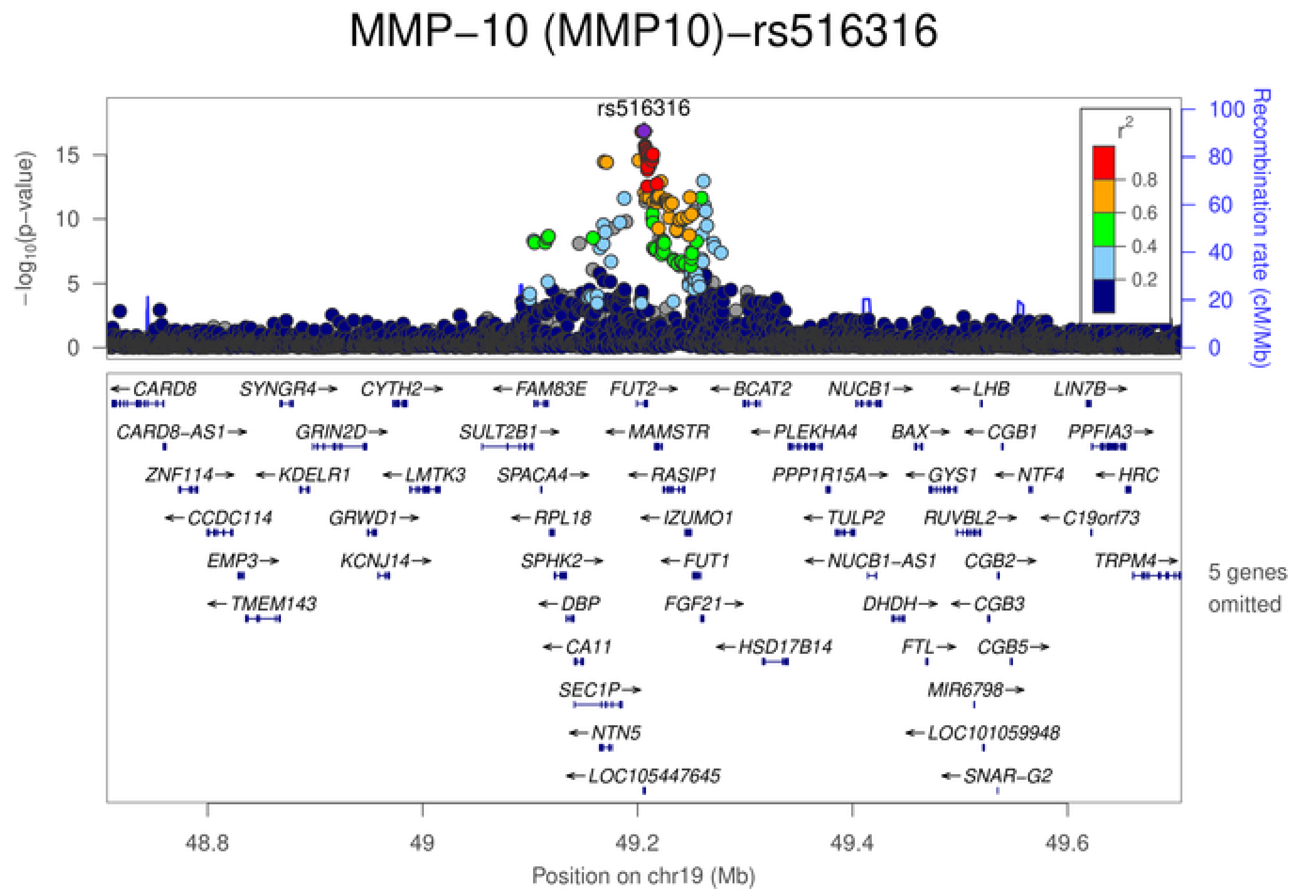
MMP−10 (MMP10)−rs516316
rs516316
$-\log_{10}$(p−value)
Recombination rate (cM/Mb)
$r^2$
0.8
0.6
0.4
0.2
CARD8 SYNGR4 CYTH2 FAM83E FUT2 BCAT2 NUCB1 LHB LIN7B
CARD8-AS1 GRIN2D SULT2B1 MAMSTR PLEKHA4 BAX CGB1 PPFIA3
ZNF114 KDELR1 LMTK3 SPACA4 RASIP1 PPP1R15A GYS1 NTF4 HRC
CCDC114 GRWD1 RPL18 IZUMO1 TULP2 RUVBL2 C19orf73
EMP3 KCNJ14 SPHK2 FUT1 NUCB1-AS1 CGB2 TRPM4
TMEM143 DBP FGF21 DHDH CGB3
CA11 HSD17B14 FTL CGB5
SEC1P MIR6798
NTN5 LOC101059948
LOC105447645 SNAR-G2
5 genes omitted
48.8 49 49.2 49.4 49.6
Position on chr19 (Mb)

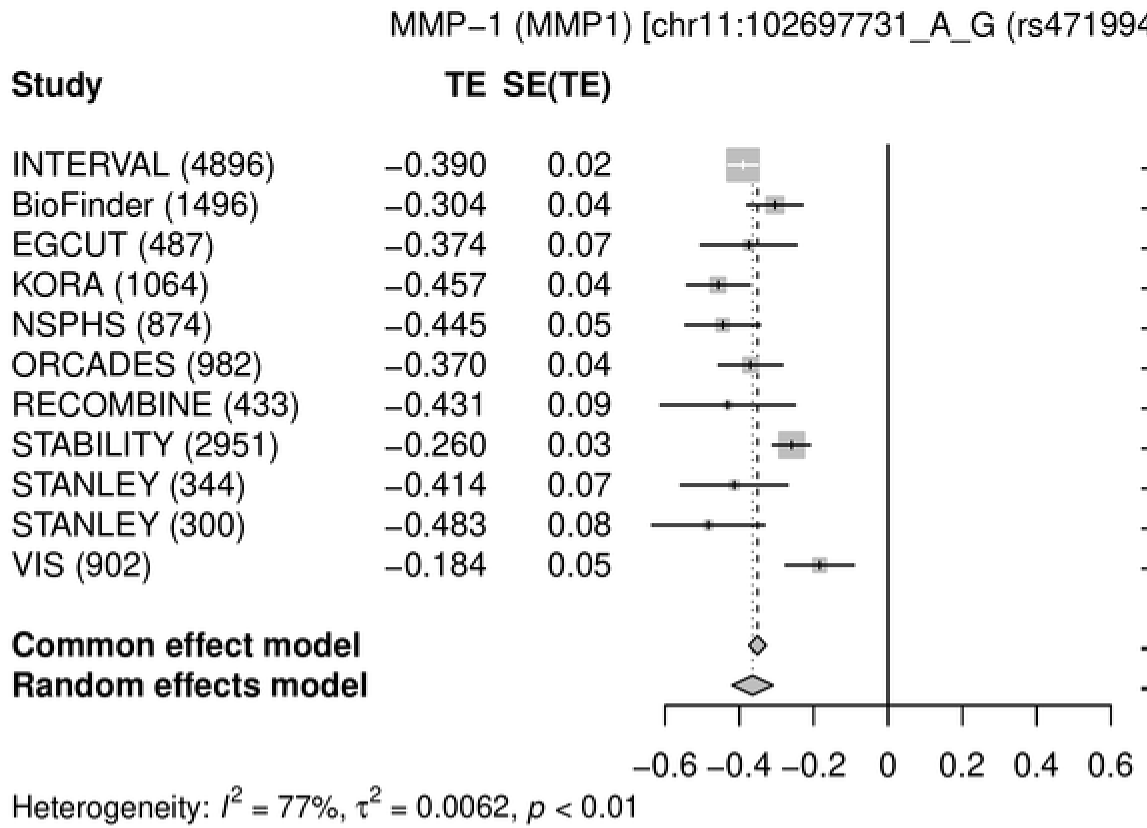
MMP−1 (MMP1) [chr11:102697731_A_G (rs471994) (A/G) N=14729]
Study
TE
SE(TE)
95%−CI
Weight (common)
Weight (random)
INTERVAL (4896) −0.390 0.02 −0.39 [−0.43; −0.35] 33.4% 12.2%
BioFinder (1496) −0.304 0.04 −0.30 [−0.38; −0.23] 9.5% 10.5%
EGCUT (487) −0.374 0.07 −0.37 [−0.50; −0.24] 3.2% 7.6%
KORA (1064) −0.457 0.04 −0.46 [−0.54; −0.37] 7.5% 10.0%
NSPHS (874) −0.445 0.05 −0.44 [−0.55; −0.34] 5.3% 9.1%
ORCADES (982) −0.370 0.04 −0.37 [−0.46; −0.28] 7.2% 9.9%
RECOMBINE (433) −0.431 0.09 −0.43 [−0.61; −0.25] 1.6% 5.5%
STABILITY (2951) −0.260 0.03 −0.26 [−0.31; −0.21] 20.9% 11.8%
STANLEY (344) −0.414 0.07 −0.41 [−0.56; −0.27] 2.6% 7.0%
STANLEY (300) −0.483 0.08 −0.48 [−0.64; −0.33] 2.3% 6.6%
VIS (902) −0.184 0.05 −0.18 [−0.28; −0.09] 6.3% 9.6%
Common effect model −0.35 [−0.37; −0.33] 100.0% --
Random effects model −0.36 [−0.42; −0.31] -- 100.0%
−0.6 −0.4 −0.2 0 0.2 0.4 0.6
Heterogeneity: $I^2$ = 77%, $\tau^2$ = 0.0062, $p$ < 0.01

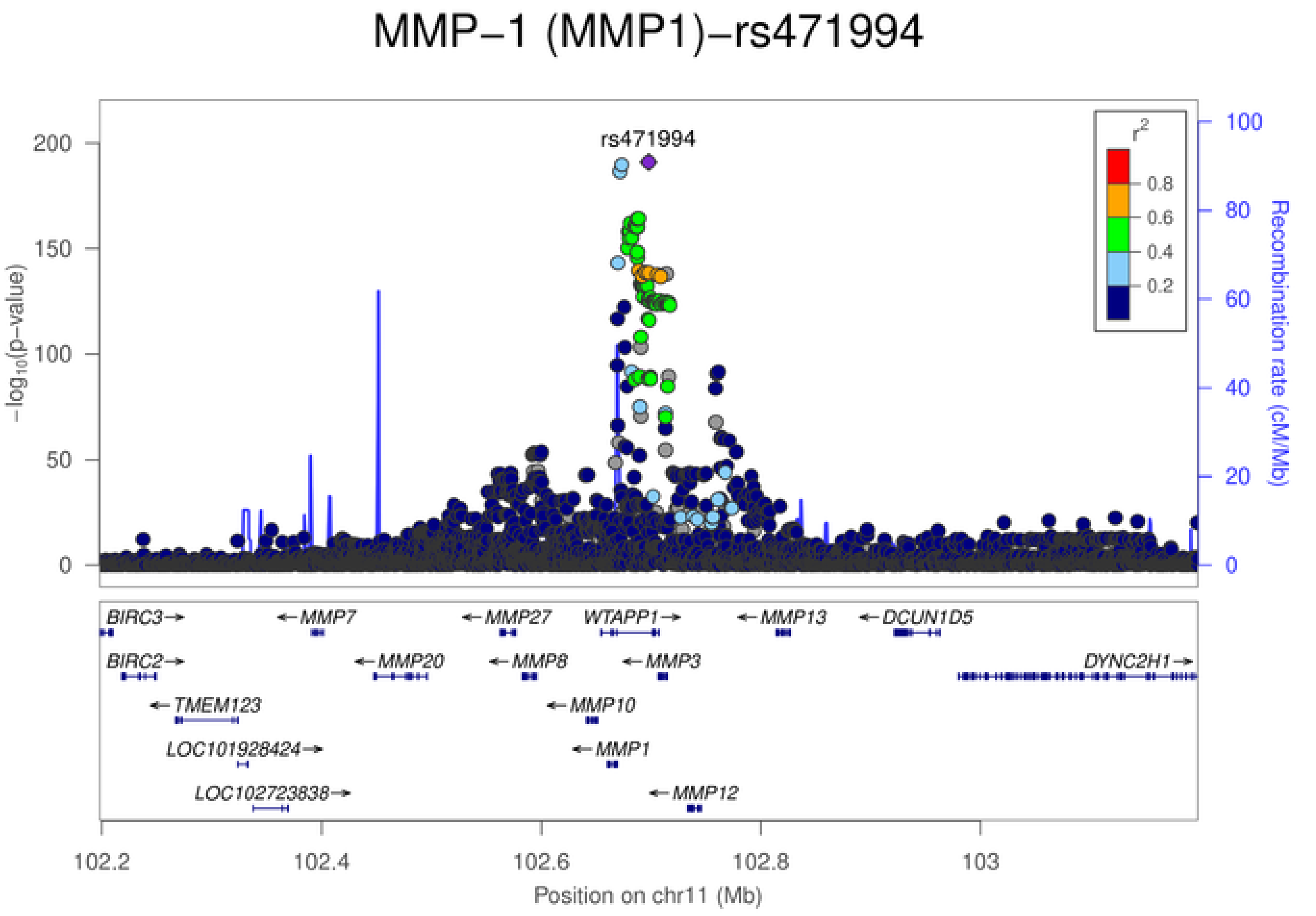
MMP−1 (MMP1)−rs471994
rs471994
$-\log_{10}$(p−value)
Recombination rate (cM/Mb)
$r^2$
0.8
0.6
0.4
0.2
200
150
100
50
0
100
80
60
40
20
0
BIRC3→
←MMP7
←MMP27
WTAPP1→
←MMP13
←DCUN1D5
BIRC2→
←MMP20
←MMP8
←MMP3
DYNC2H1→
←TMEM123
←MMP10
LOC101928424→
←MMP1
LOC102723838→
←MMP12
102.2
102.4
102.6
102.8
103
Position on chr11 (Mb)

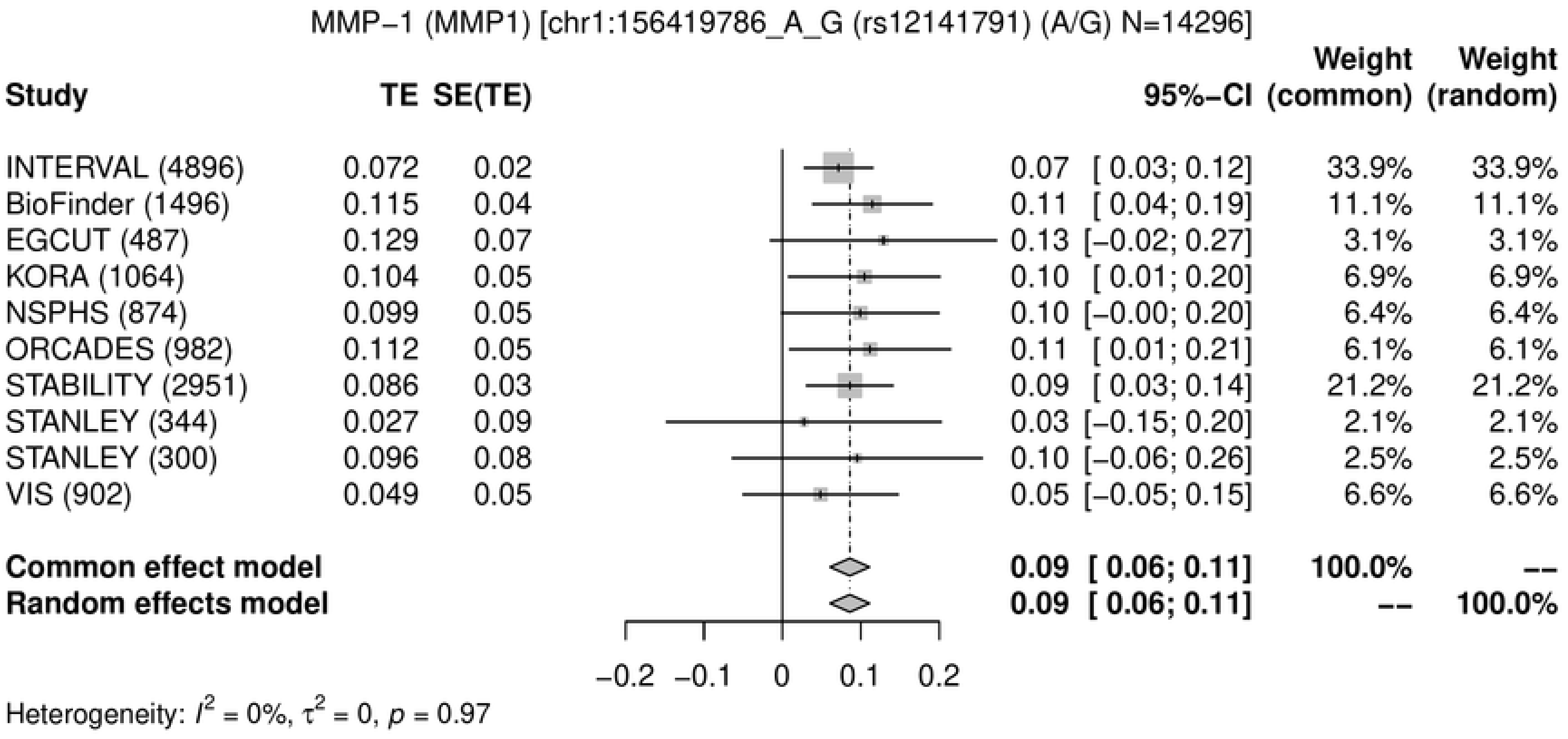
MMP−1 (MMP1) [chr1:156419786_A_G (rs12141791) (A/G) N=14296]
Study | TE | SE(TE) | 95%−CI | Weight (common) | Weight (random)
INTERVAL (4896) | 0.072 | 0.02 | 0.07 [ 0.03; 0.12] | 33.9% | 33.9%
BioFinder (1496) | 0.115 | 0.04 | 0.11 [ 0.04; 0.19] | 11.1% | 11.1%
EGCUT (487) | 0.129 | 0.07 | 0.13 [−0.02; 0.27] | 3.1% | 3.1%
KORA (1064) | 0.104 | 0.05 | 0.10 [ 0.01; 0.20] | 6.9% | 6.9%
NSPHS (874) | 0.099 | 0.05 | 0.10 [−0.00; 0.20] | 6.4% | 6.4%
ORCADES (982) | 0.112 | 0.05 | 0.11 [ 0.01; 0.21] | 6.1% | 6.1%
STABILITY (2951) | 0.086 | 0.03 | 0.09 [ 0.03; 0.14] | 21.2% | 21.2%
STANLEY (344) | 0.027 | 0.09 | 0.03 [−0.15; 0.20] | 2.1% | 2.1%
STANLEY (300) | 0.096 | 0.08 | 0.10 [−0.06; 0.26] | 2.5% | 2.5%
VIS (902) | 0.049 | 0.05 | 0.05 [−0.05; 0.15] | 6.6% | 6.6%
Common effect model | 0.09 [ 0.06; 0.11] | 100.0% | --
Random effects model | 0.09 [ 0.06; 0.11] | -- | 100.0%
−0.2 −0.1 0 0.1 0.2
Heterogeneity: $I^2$ = 0%, $\tau^2$ = 0, $p$ = 0.97

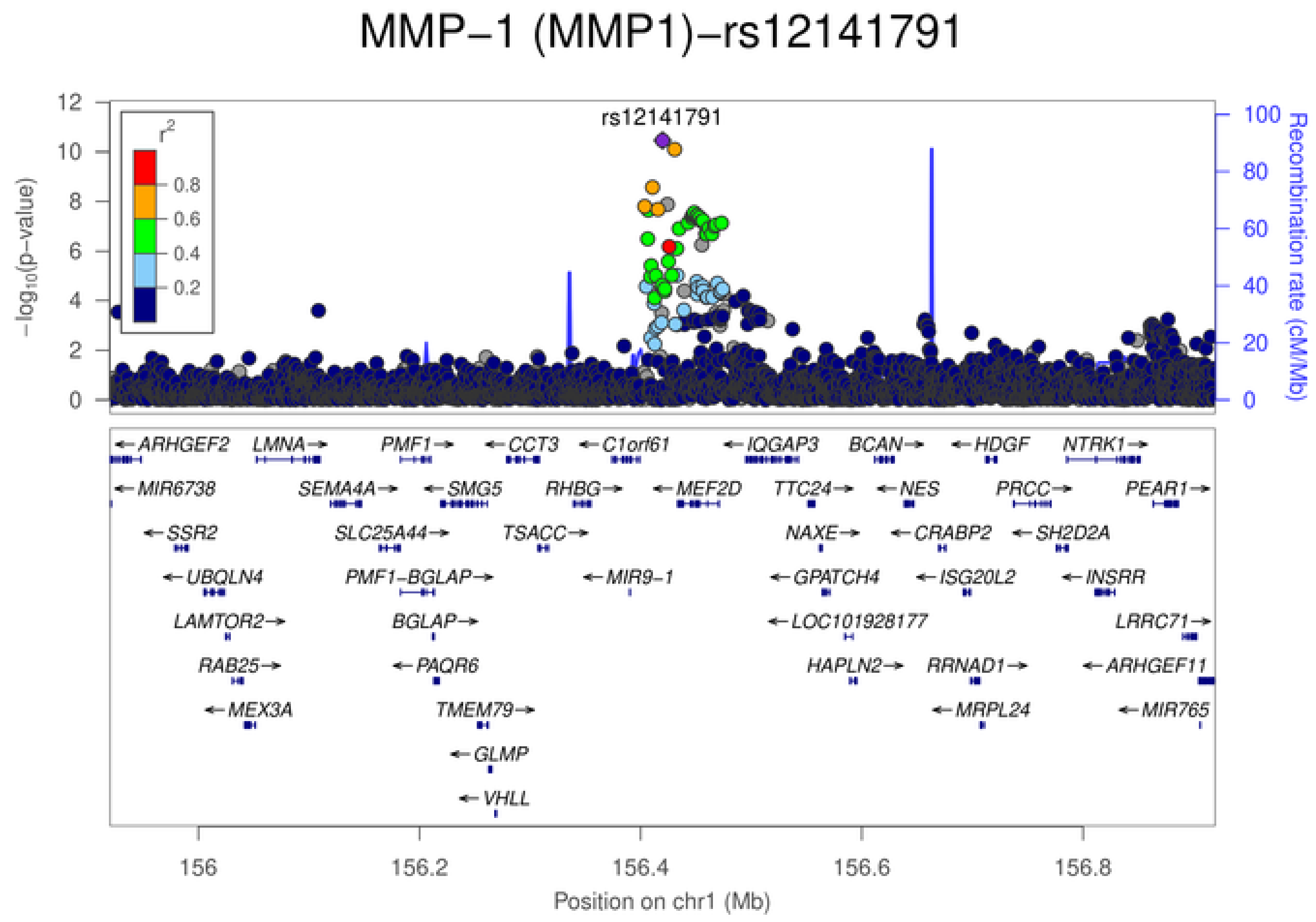
MMP−1 (MMP1)−rs12141791
rs12141791
$-\log_{10}$(p−value)
Recombination rate (cM/Mb)
$r^2$
0.8
0.6
0.4
0.2
ARHGEF2 LMNA PMF1 CCT3 C1orf61 IQGAP3 BCAN HDGF NTRK1
MIR6738 SEMA4A SMG5 RHBG MEF2D TTC24 NES PRCC PEAR1
SSR2 SLC25A44 TSACC NAXE CRABP2 SH2D2A
UBQLN4 PMF1−BGLAP MIR9−1 GPATCH4 ISG20L2 INSRR
LAMTOR2 BGLAP LOC101928177 LRRC71
RAB25 PAQR6 HAPLN2 RRNAD1 ARHGEF11
MEX3A TMEM79 MRPL24 MIR765
GLMP
VHLL
156 156.2 156.4 156.6 156.8
Position on chr1 (Mb)

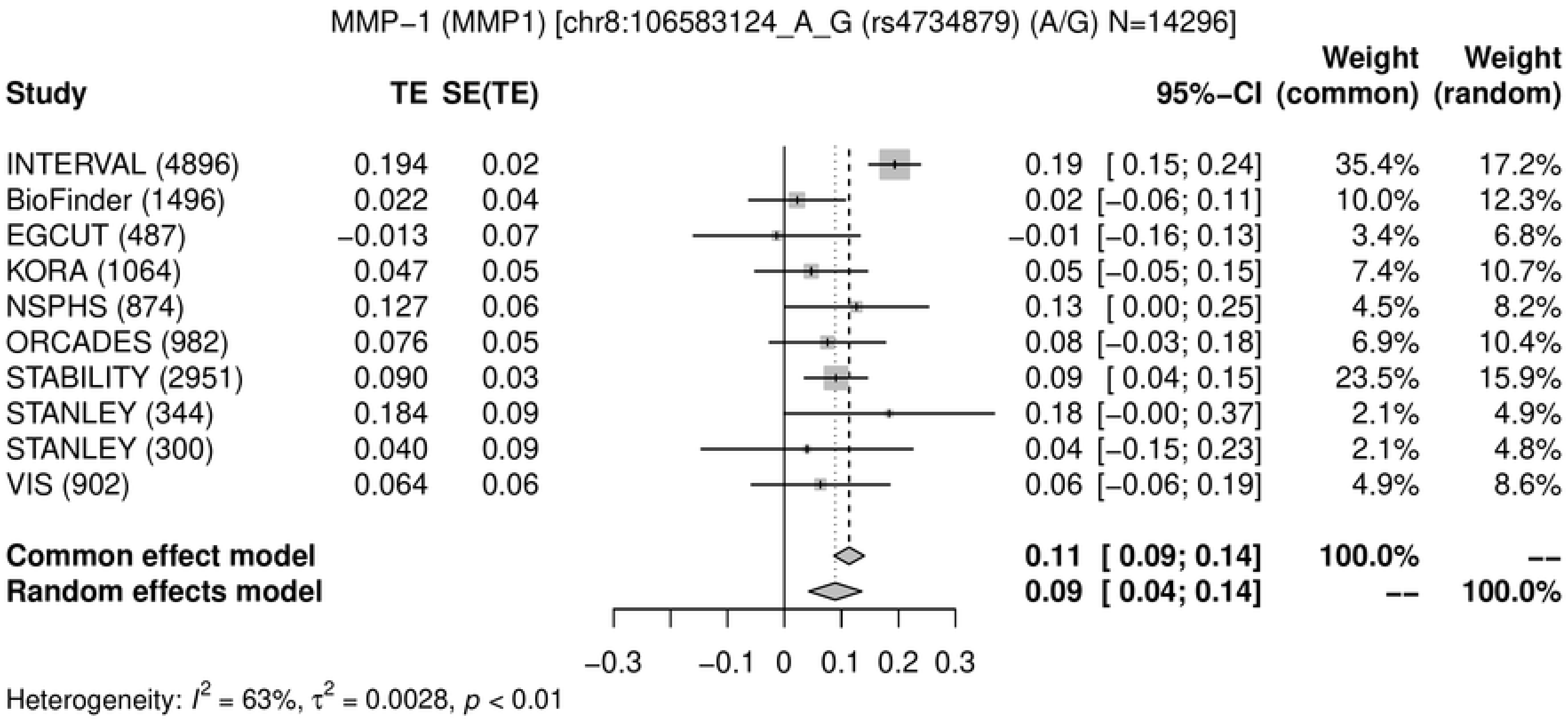
MMP−1 (MMP1) [chr8:106583124_A_G (rs4734879) (A/G) N=14296]
Study TE SE(TE) 95%−CI Weight (common) Weight (random)
INTERVAL (4896) 0.194 0.02 0.19 [ 0.15; 0.24] 35.4% 17.2%
BioFinder (1496) 0.022 0.04 0.02 [−0.06; 0.11] 10.0% 12.3%
EGCUT (487) −0.013 0.07 −0.01 [−0.16; 0.13] 3.4% 6.8%
KORA (1064) 0.047 0.05 0.05 [−0.05; 0.15] 7.4% 10.7%
NSPHS (874) 0.127 0.06 0.13 [ 0.00; 0.25] 4.5% 8.2%
ORCADES (982) 0.076 0.05 0.08 [−0.03; 0.18] 6.9% 10.4%
STABILITY (2951) 0.090 0.03 0.09 [ 0.04; 0.15] 23.5% 15.9%
STANLEY (344) 0.184 0.09 0.18 [−0.00; 0.37] 2.1% 4.9%
STANLEY (300) 0.040 0.09 0.04 [−0.15; 0.23] 2.1% 4.8%
VIS (902) 0.064 0.06 0.06 [−0.06; 0.19] 4.9% 8.6%
Common effect model 0.11 [ 0.09; 0.14] 100.0% --
Random effects model 0.09 [ 0.04; 0.14] -- 100.0%
−0.3 −0.1 0 0.1 0.2 0.3
Heterogeneity: $I^2$ = 63%, $\tau^2$ = 0.0028, $p$ < 0.01

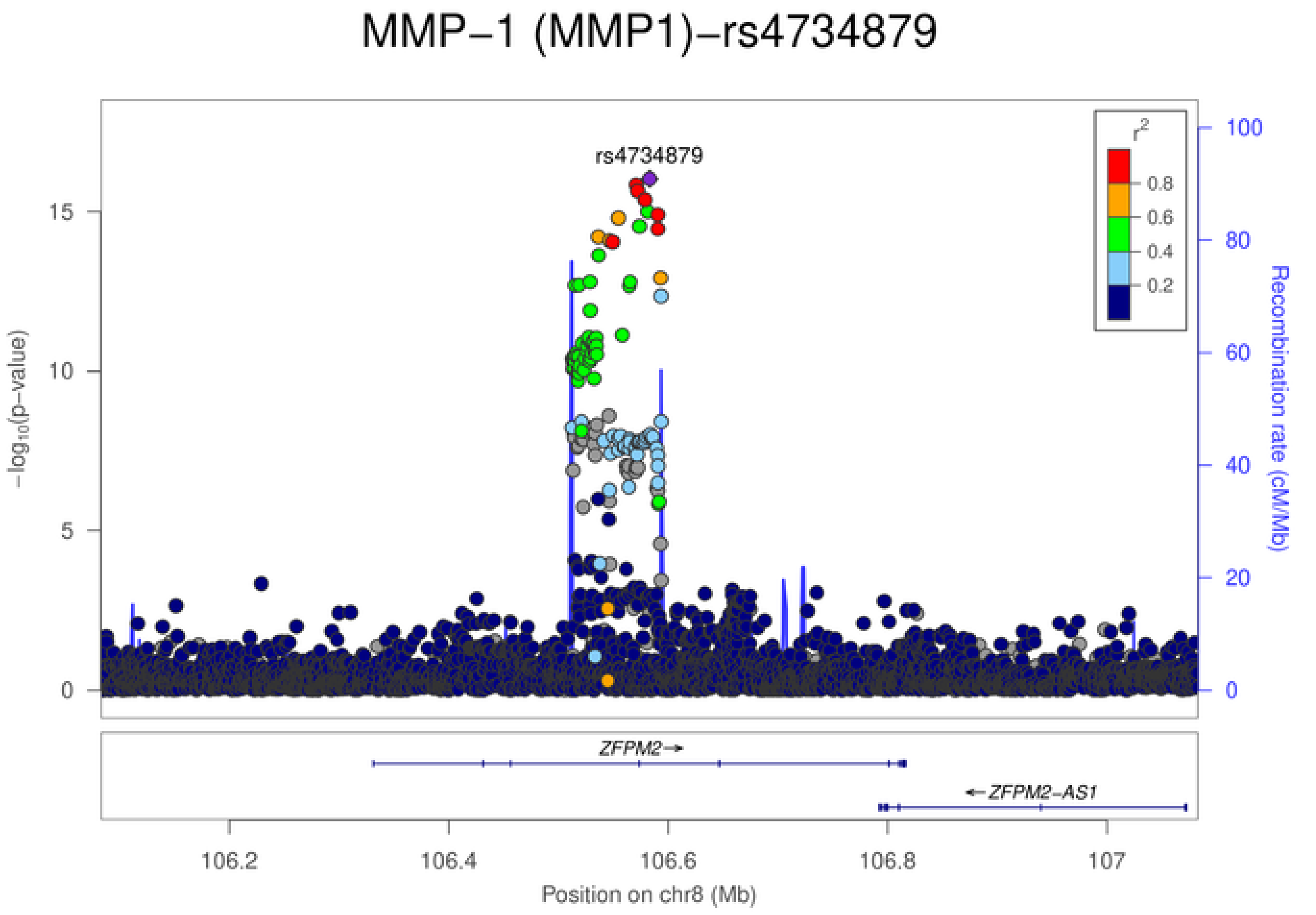
MMP−1 (MMP1)−rs4734879
rs4734879
$r^2$
0.8
0.6
0.4
0.2
$-\log_{10}$(p−value)
Recombination rate (cM/Mb)
ZFPM2→
←ZFPM2-AS1
106.2
106.4
106.6
106.8
107
Position on chr8 (Mb)

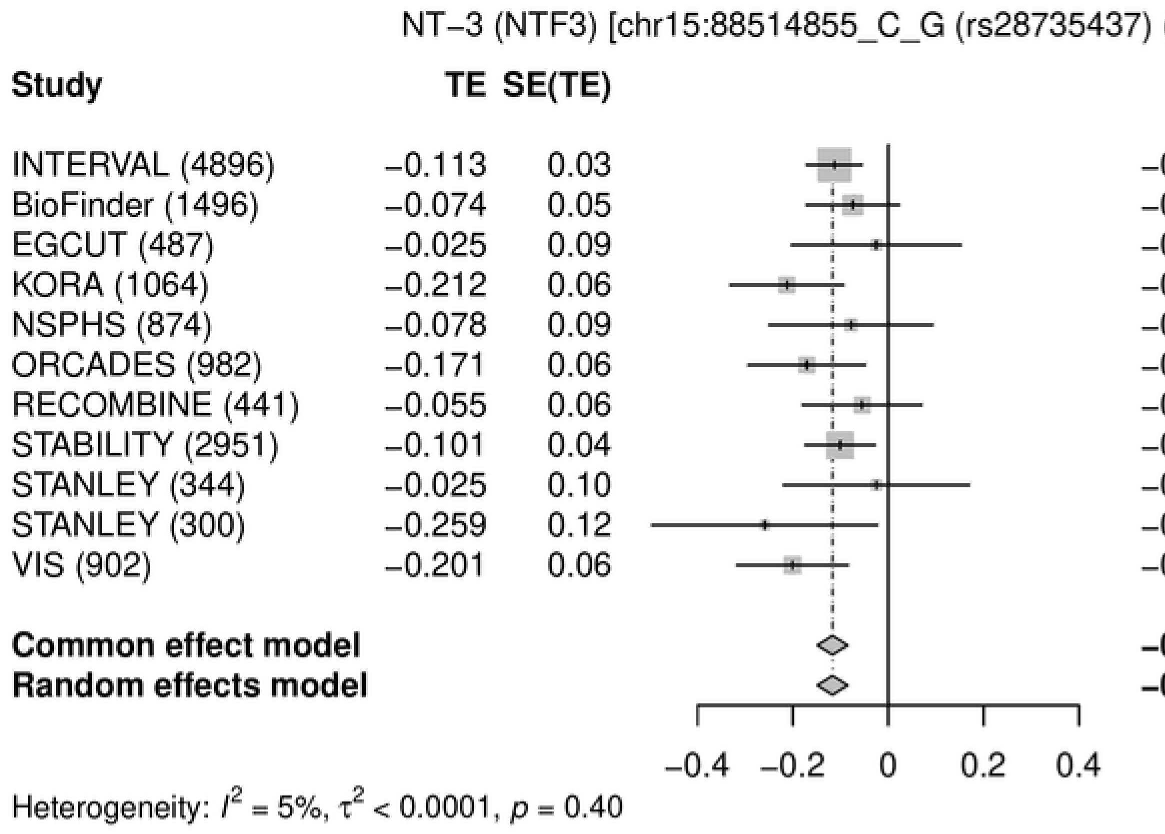
NT−3 (NTF3) [chr15:88514855_C_G (rs28735437) (C/G) N=14737]
Study
TE
SE(TE)
95%−CI
Weight (common)
Weight (random)
INTERVAL (4896) −0.113 0.03 −0.11 [−0.17; −0.05] 30.3% 30.3%
BioFinder (1496) −0.074 0.05 −0.07 [−0.17; 0.02] 10.9% 10.9%
EGCUT (487) −0.025 0.09 −0.03 [−0.20; 0.15] 3.3% 3.3%
KORA (1064) −0.212 0.06 −0.21 [−0.33; −0.09] 7.3% 7.3%
NSPHS (874) −0.078 0.09 −0.08 [−0.25; 0.09] 3.5% 3.5%
ORCADES (982) −0.171 0.06 −0.17 [−0.29; −0.05] 6.9% 6.9%
RECOMBINE (441) −0.055 0.06 −0.05 [−0.18; 0.07] 6.6% 6.6%
STABILITY (2951) −0.101 0.04 −0.10 [−0.17; −0.03] 19.1% 19.1%
STANLEY (344) −0.025 0.10 −0.02 [−0.22; 0.17] 2.7% 2.7%
STANLEY (300) −0.259 0.12 −0.26 [−0.50; −0.02] 1.9% 1.9%
VIS (902) −0.201 0.06 −0.20 [−0.32; −0.08] 7.6% 7.6%
Common effect model −0.12 [−0.15; −0.08] 100.0% --
Random effects model −0.12 [−0.15; −0.08] -- 100.0%
−0.4 −0.2 0 0.2 0.4
Heterogeneity: $I^2$ = 5%, $\tau^2$ < 0.0001, $p$ = 0.40

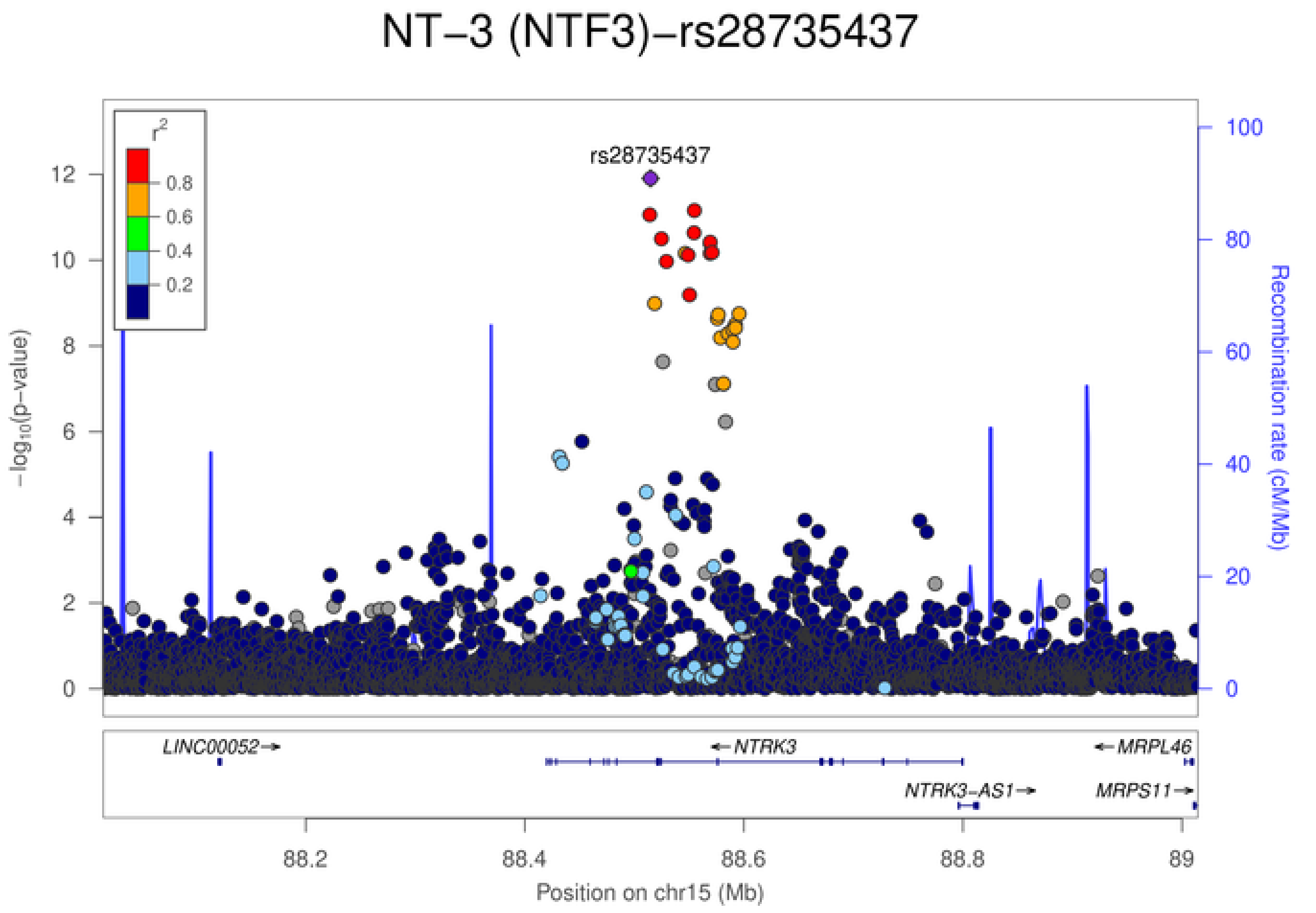
NT−3 (NTF3)−rs28735437
rs28735437
$r^2$
0.8
0.6
0.4
0.2
$-\log_{10}$(p−value)
0
2
4
6
8
10
12
Recombination rate (cM/Mb)
0
20
40
60
80
100
LINC00052→
←NTRK3
←MRPL46
NTRK3−AS1→
MRPS11→
88.2
88.4
88.6
88.8
89
Position on chr15 (Mb)

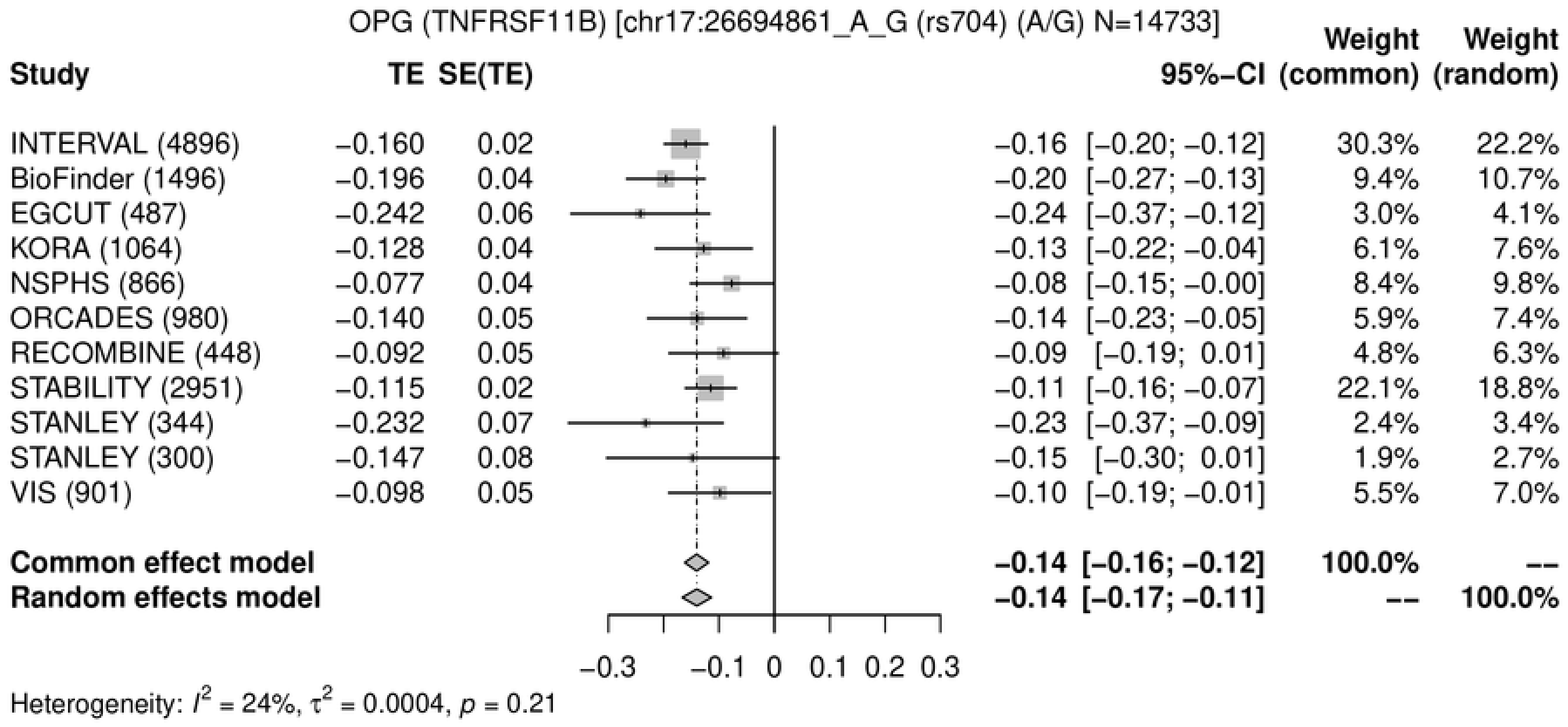
OPG (TNFRSF11B) [chr17:26694861_A_G (rs704) (A/G) N=14733]
Study
TE
SE(TE)
95%−CI
Weight (common)
Weight (random)
INTERVAL (4896) −0.160 0.02 −0.16 [−0.20; −0.12] 30.3% 22.2%
BioFinder (1496) −0.196 0.04 −0.20 [−0.27; −0.13] 9.4% 10.7%
EGCUT (487) −0.242 0.06 −0.24 [−0.37; −0.12] 3.0% 4.1%
KORA (1064) −0.128 0.04 −0.13 [−0.22; −0.04] 6.1% 7.6%
NSPHS (866) −0.077 0.04 −0.08 [−0.15; −0.00] 8.4% 9.8%
ORCADES (980) −0.140 0.05 −0.14 [−0.23; −0.05] 5.9% 7.4%
RECOMBINE (448) −0.092 0.05 −0.09 [−0.19; 0.01] 4.8% 6.3%
STABILITY (2951) −0.115 0.02 −0.11 [−0.16; −0.07] 22.1% 18.8%
STANLEY (344) −0.232 0.07 −0.23 [−0.37; −0.09] 2.4% 3.4%
STANLEY (300) −0.147 0.08 −0.15 [−0.30; 0.01] 1.9% 2.7%
VIS (901) −0.098 0.05 −0.10 [−0.19; −0.01] 5.5% 7.0%
Common effect model −0.14 [−0.16; −0.12] 100.0% --
Random effects model −0.14 [−0.17; −0.11] -- 100.0%
−0.3 −0.1 0 0.1 0.2 0.3
Heterogeneity: $I^2$ = 24%, $\tau^2$ = 0.0004, $p$ = 0.21

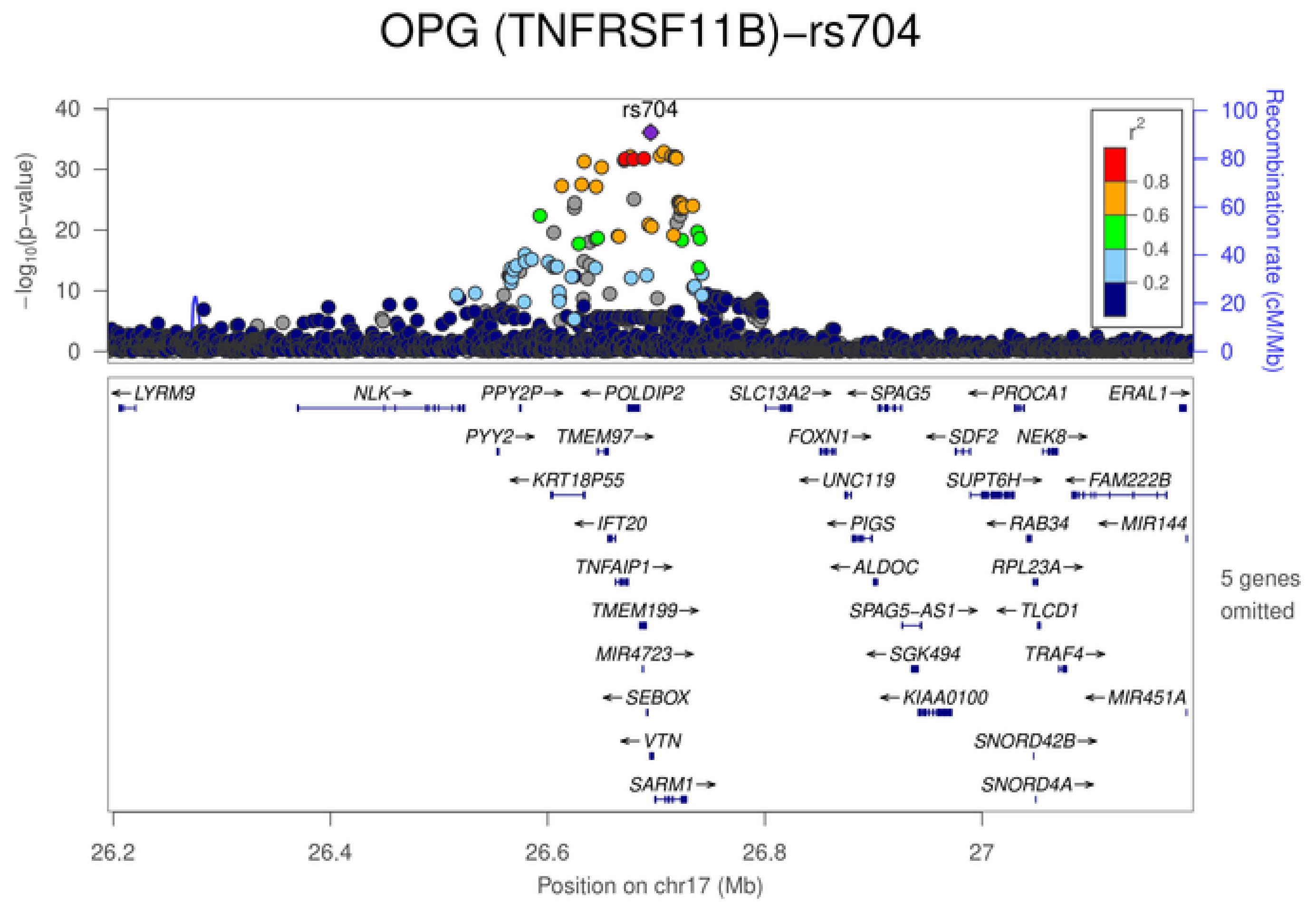
OPG (TNFRSF11B)−rs704
rs704
$-\log_{10}$(p−value)
Recombination rate (cM/Mb)
$r^2$
0.8
0.6
0.4
0.2
LYRM9
NLK
PPY2P
POLDIP2
SLC13A2
SPAG5
PROCA1
ERAL1
PYY2
TMEM97
FOXN1
SDF2
NEK8
KRT18P55
UNC119
SUPT6H
FAM222B
IFT20
PIGS
RAB34
MIR144
TNFAIP1
ALDOC
RPL23A
TMEM199
SPAG5−AS1
TLCD1
MIR4723
SGK494
TRAF4
SEBOX
KIAA0100
MIR451A
VTN
SNORD42B
SARM1
SNORD4A
5 genes omitted
26.2
26.4
26.6
26.8
27
Position on chr17 (Mb)

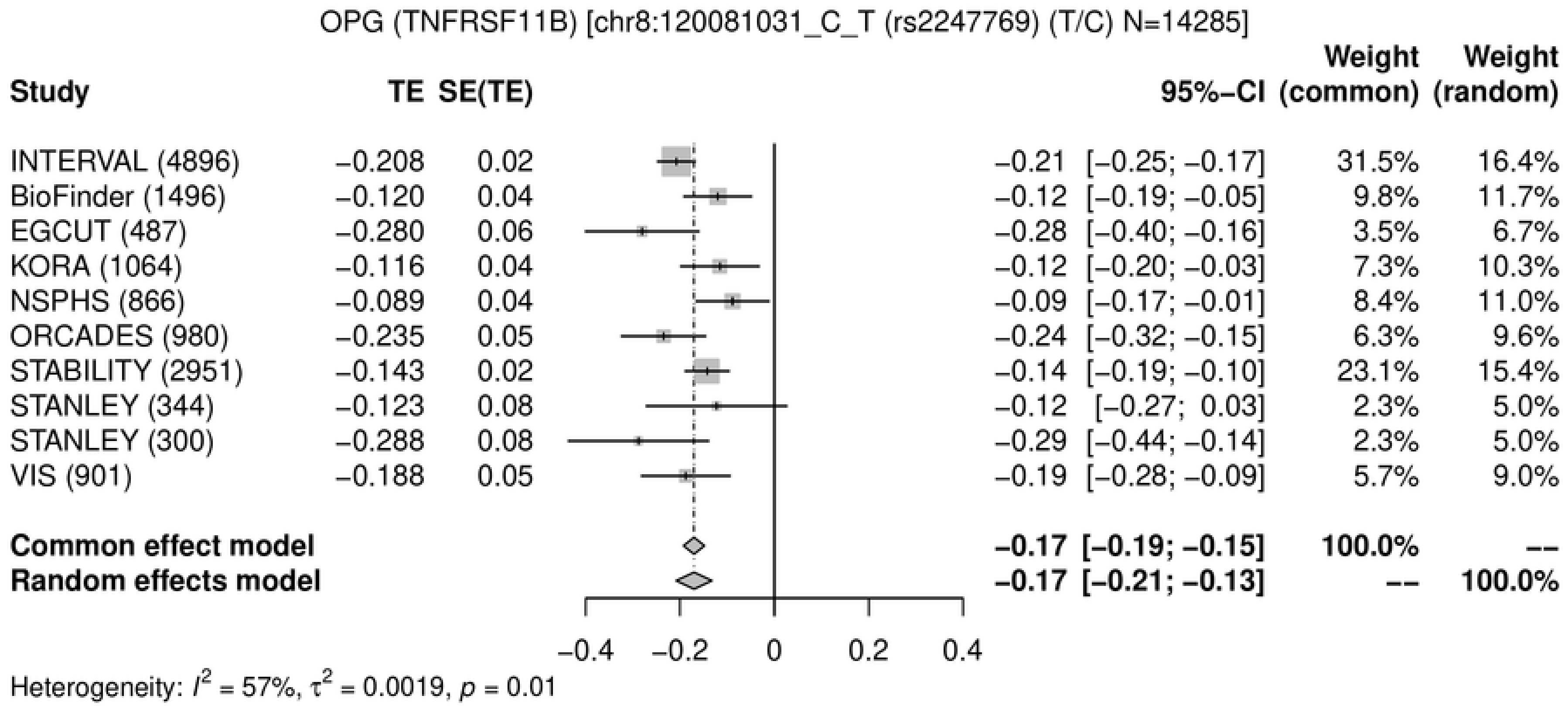
OPG (TNFRSF11B) [chr8:120081031_C_T (rs2247769) (T/C) N=14285]
Study
TE
SE(TE)
95%−CI
Weight (common)
Weight (random)
INTERVAL (4896) −0.208 0.02 −0.21 [−0.25; −0.17] 31.5% 16.4%
BioFinder (1496) −0.120 0.04 −0.12 [−0.19; −0.05] 9.8% 11.7%
EGCUT (487) −0.280 0.06 −0.28 [−0.40; −0.16] 3.5% 6.7%
KORA (1064) −0.116 0.04 −0.12 [−0.20; −0.03] 7.3% 10.3%
NSPHS (866) −0.089 0.04 −0.09 [−0.17; −0.01] 8.4% 11.0%
ORCADES (980) −0.235 0.05 −0.24 [−0.32; −0.15] 6.3% 9.6%
STABILITY (2951) −0.143 0.02 −0.14 [−0.19; −0.10] 23.1% 15.4%
STANLEY (344) −0.123 0.08 −0.12 [−0.27; 0.03] 2.3% 5.0%
STANLEY (300) −0.288 0.08 −0.29 [−0.44; −0.14] 2.3% 5.0%
VIS (901) −0.188 0.05 −0.19 [−0.28; −0.09] 5.7% 9.0%
Common effect model −0.17 [−0.19; −0.15] 100.0% --
Random effects model −0.17 [−0.21; −0.13] -- 100.0%
−0.4 −0.2 0 0.2 0.4
Heterogeneity: $I^2$ = 57%, $\tau^2$ = 0.0019, $p$ = 0.01

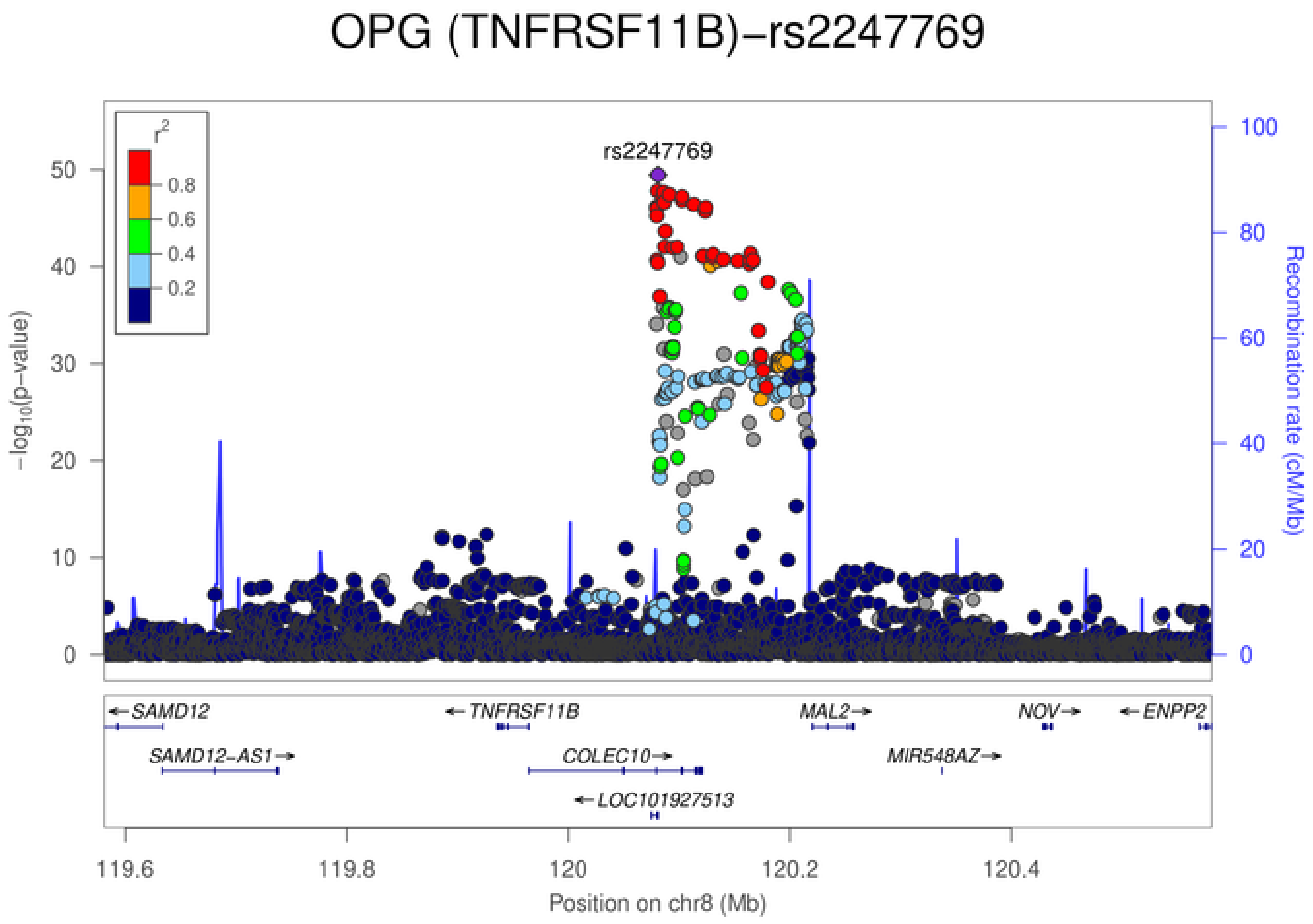
OPG (TNFRSF11B)−rs2247769
rs2247769
$r^2$
0.8
0.6
0.4
0.2
−log10(p−value)
Recombination rate (cM/Mb)
SAMD12
TNFRSF11B
MAL2
NOV
ENPP2
SAMD12−AS1
COLEC10
MIR548AZ
LOC101927513
119.6 119.8 120 120.2 120.4
Position on chr8 (Mb)

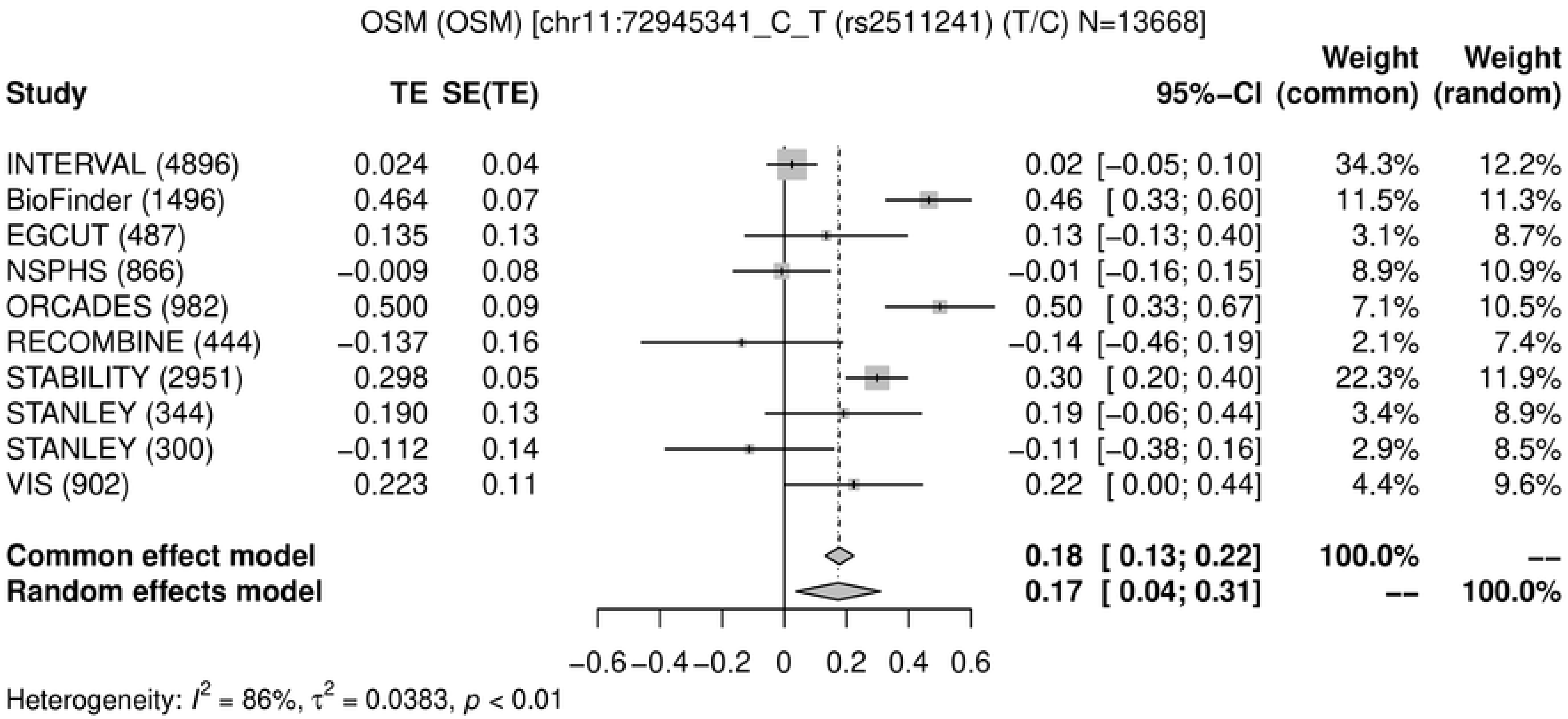
OSM (OSM) [chr11:72945341_C_T (rs2511241) (T/C) N=13668]
Study
TE
SE(TE)
95%−CI
Weight (common)
Weight (random)
INTERVAL (4896) 0.024 0.04 0.02 [−0.05; 0.10] 34.3% 12.2%
BioFinder (1496) 0.464 0.07 0.46 [ 0.33; 0.60] 11.5% 11.3%
EGCUT (487) 0.135 0.13 0.13 [−0.13; 0.40] 3.1% 8.7%
NSPHS (866) −0.009 0.08 −0.01 [−0.16; 0.15] 8.9% 10.9%
ORCADES (982) 0.500 0.09 0.50 [ 0.33; 0.67] 7.1% 10.5%
RECOMBINE (444) −0.137 0.16 −0.14 [−0.46; 0.19] 2.1% 7.4%
STABILITY (2951) 0.298 0.05 0.30 [ 0.20; 0.40] 22.3% 11.9%
STANLEY (344) 0.190 0.13 0.19 [−0.06; 0.44] 3.4% 8.9%
STANLEY (300) −0.112 0.14 −0.11 [−0.38; 0.16] 2.9% 8.5%
VIS (902) 0.223 0.11 0.22 [ 0.00; 0.44] 4.4% 9.6%
Common effect model 0.18 [ 0.13; 0.22] 100.0% --
Random effects model 0.17 [ 0.04; 0.31] -- 100.0%
−0.6 −0.4 −0.2 0 0.2 0.4 0.6
Heterogeneity: $I^2$ = 86%, $\tau^2$ = 0.0383, $p$ < 0.01

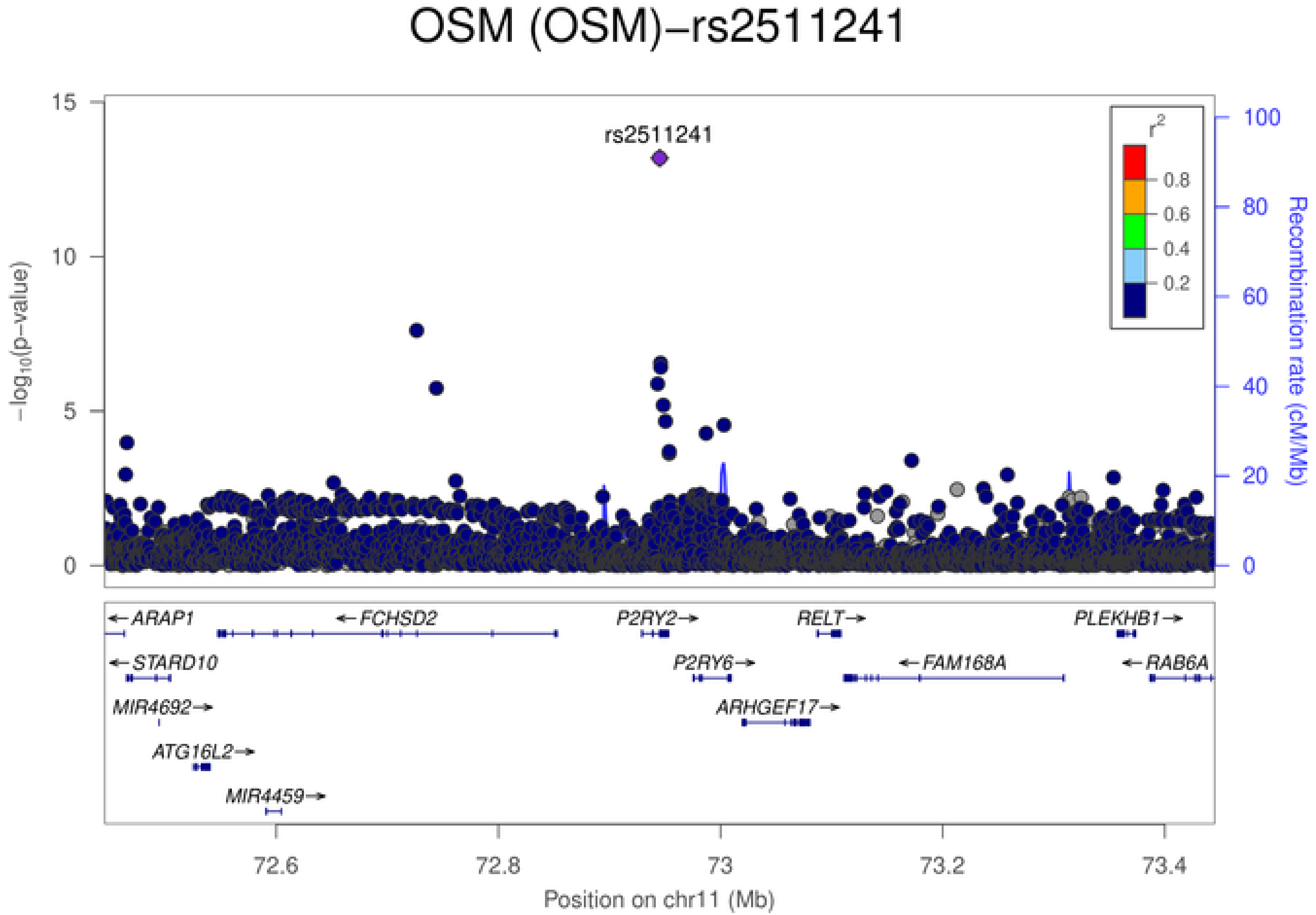
OSM (OSM)−rs2511241
rs2511241
$-\log_{10}$(p-value)
Recombination rate (cM/Mb)
$r^2$
0.8
0.6
0.4
0.2
ARAP1
FCHSD2
P2RY2
RELT
PLEKHB1
STARD10
P2RY6
FAM168A
RAB6A
MIR4692
ARHGEF17
ATG16L2
MIR4459
72.6
72.8
73
73.2
73.4
Position on chr11 (Mb)

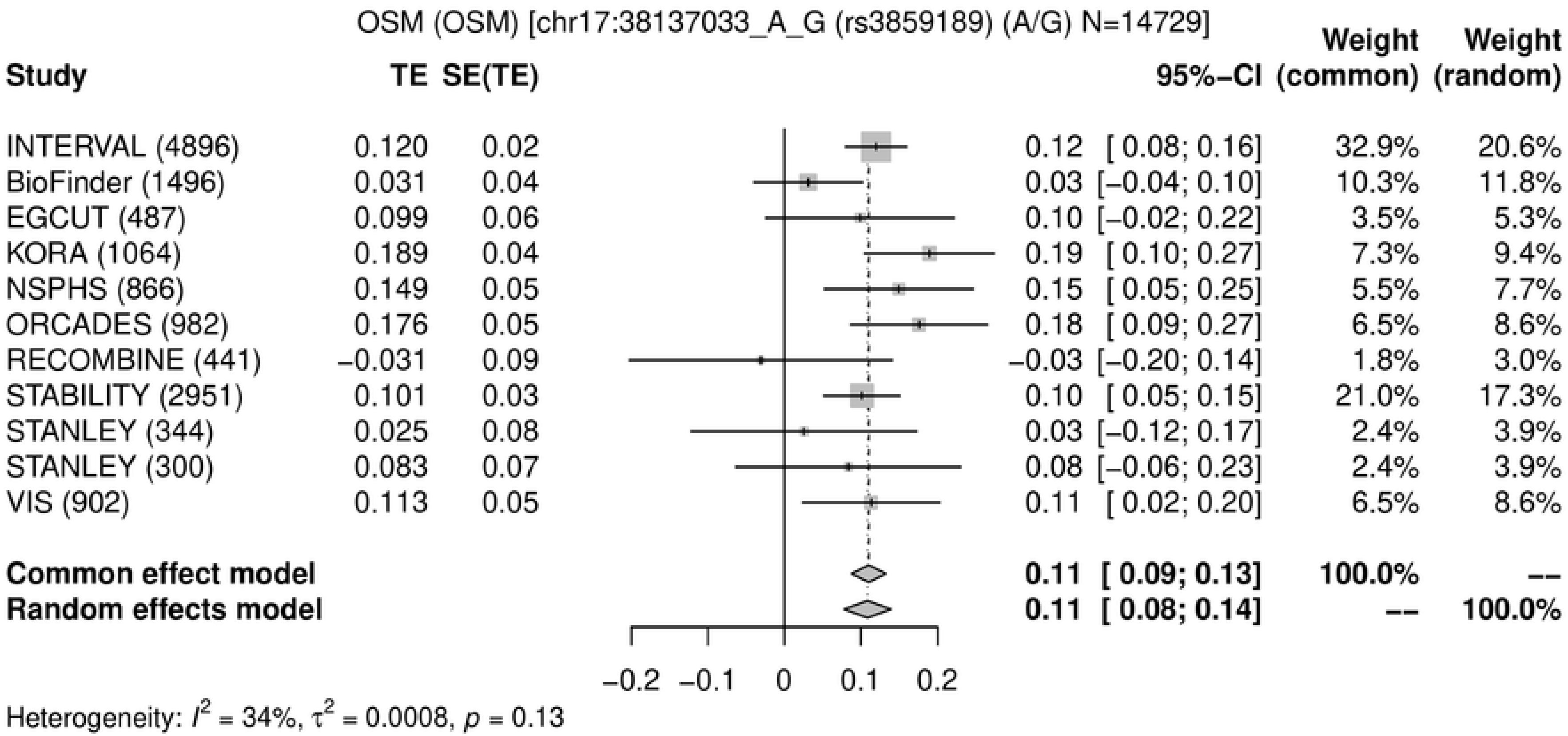

OSM (OSM) [chr17:38137033_A_G (rs3859189) (A/G) N=14729]
Study
TE
SE(TE)
95%-CI
Weight (common)
Weight (random)
INTERVAL (4896) 0.120 0.02 0.12 [ 0.08; 0.16] 32.9% 20.6%
BioFinder (1496) 0.031 0.04 0.03 [−0.04; 0.10] 10.3% 11.8%
EGCUT (487) 0.099 0.06 0.10 [−0.02; 0.22] 3.5% 5.3%
KORA (1064) 0.189 0.04 0.19 [ 0.10; 0.27] 7.3% 9.4%
NSPHS (866) 0.149 0.05 0.15 [ 0.05; 0.25] 5.5% 7.7%
ORCADES (982) 0.176 0.05 0.18 [ 0.09; 0.27] 6.5% 8.6%
RECOMBINE (441) −0.031 0.09 −0.03 [−0.20; 0.14] 1.8% 3.0%
STABILITY (2951) 0.101 0.03 0.10 [ 0.05; 0.15] 21.0% 17.3%
STANLEY (344) 0.025 0.08 0.03 [−0.12; 0.17] 2.4% 3.9%
STANLEY (300) 0.083 0.07 0.08 [−0.06; 0.23] 2.4% 3.9%
VIS (902) 0.113 0.05 0.11 [ 0.02; 0.20] 6.5% 8.6%
Common effect model 0.11 [ 0.09; 0.13] 100.0% --
Random effects model 0.11 [ 0.08; 0.14] -- 100.0%
−0.2 −0.1 0 0.1 0.2
Heterogeneity: $I^2$ = 34%, $\tau^2$ = 0.0008, $p$ = 0.13


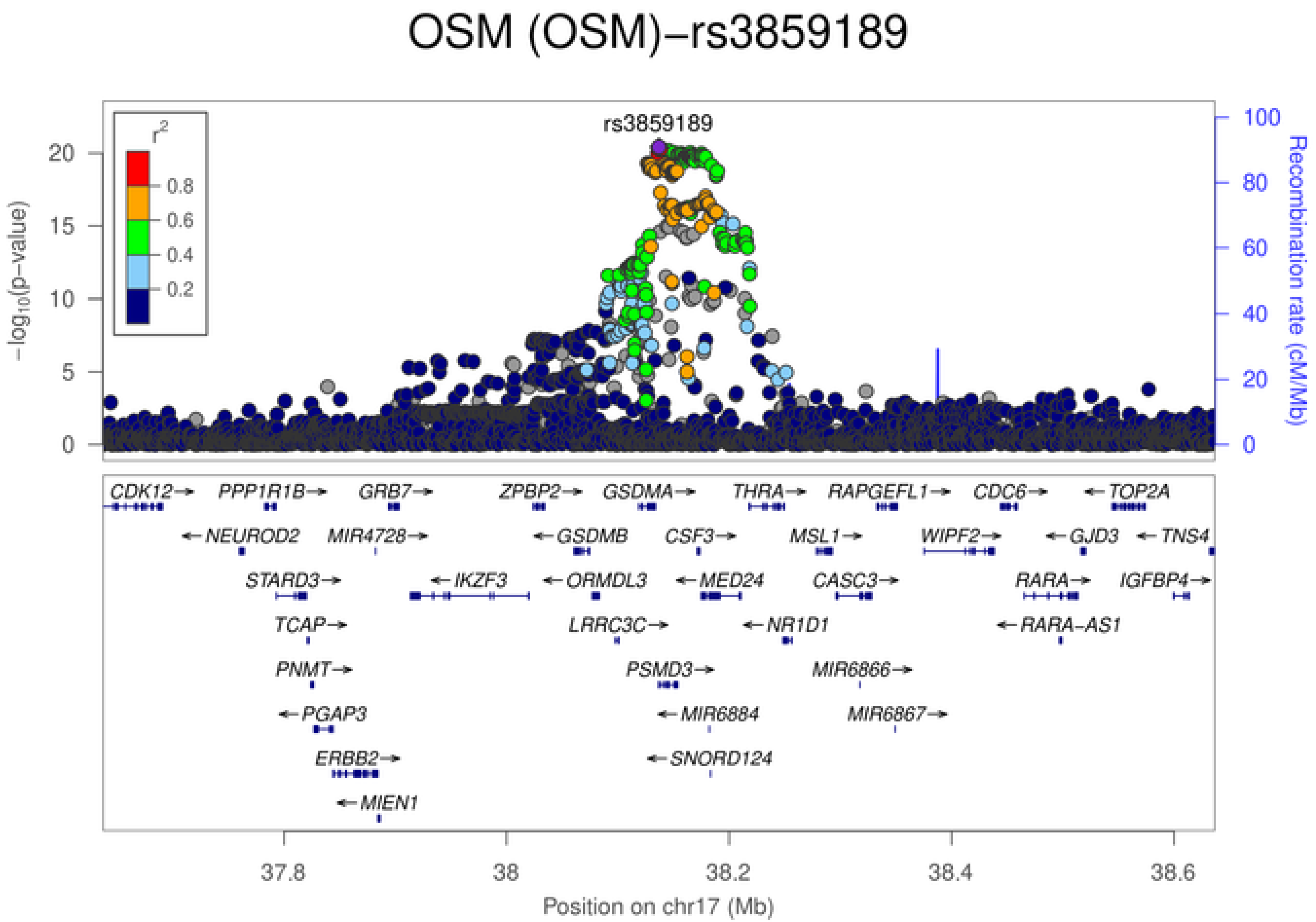

OSM (OSM)−rs3859189
rs3859189
$r^2$
0.8
0.6
0.4
0.2
$-\log_{10}$(p−value)
Recombination rate (cM/Mb)
CDK12 PPP1R1B GRB7 ZPBP2 GSDMA THRA RAPGEFL1 CDC6 TOP2A
NEUROD2 MIR4728 GSDMB CSF3 MSL1 WIPF2 GJD3 TNS4
STARD3 IKZF3 ORMDL3 MED24 CASC3 RARA IGFBP4
TCAP LRRC3C NR1D1 RARA−AS1
PNMT PSMD3 MIR6866
PGAP3 MIR6884 MIR6867
ERBB2 SNORD124
MIEN1
37.8 38 38.2 38.4 38.6
Position on chr17 (Mb)

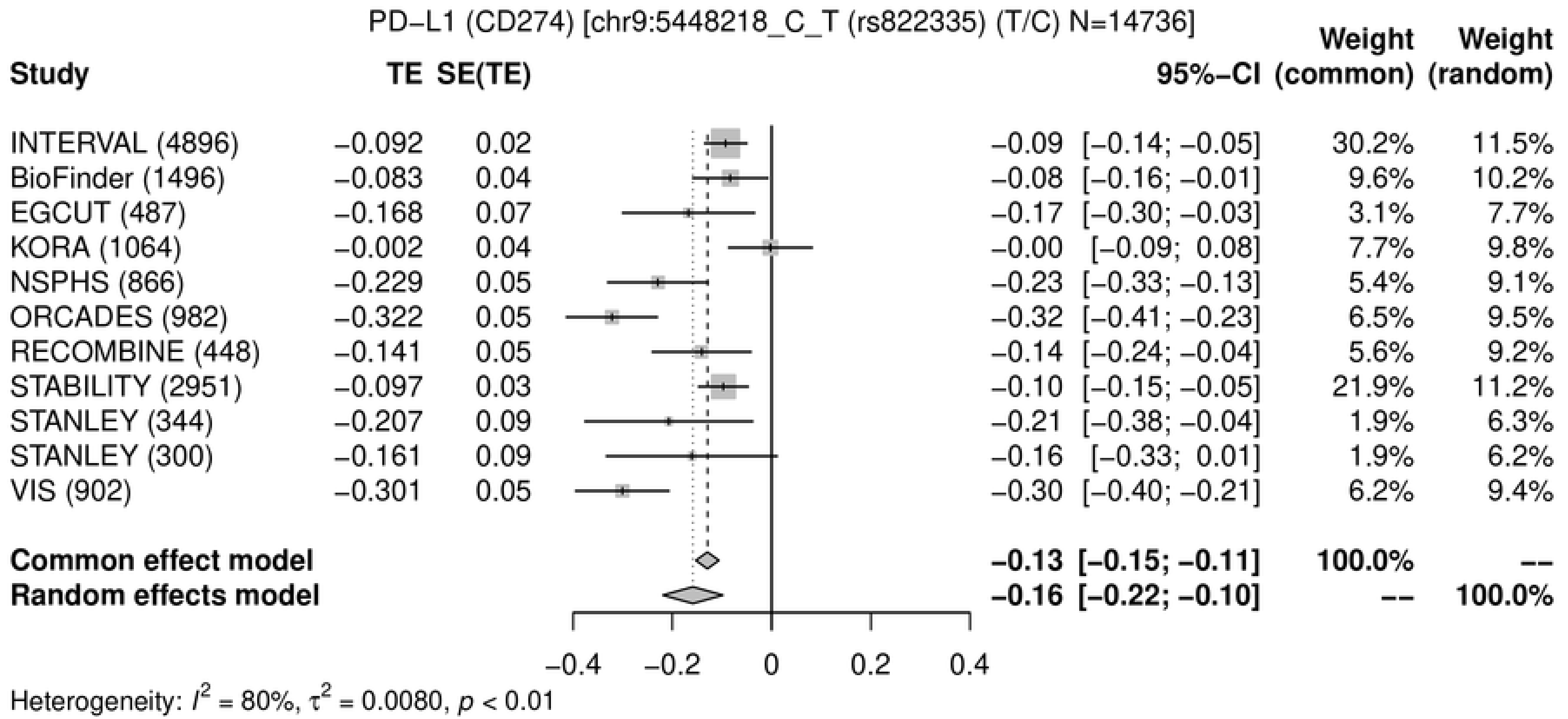
PD−L1 (CD274) [chr9:5448218_C_T (rs822335) (T/C) N=14736]
Study TE SE(TE) 95%−CI Weight (common) Weight (random)
INTERVAL (4896) −0.092 0.02 −0.09 [−0.14; −0.05] 30.2% 11.5%
BioFinder (1496) −0.083 0.04 −0.08 [−0.16; −0.01] 9.6% 10.2%
EGCUT (487) −0.168 0.07 −0.17 [−0.30; −0.03] 3.1% 7.7%
KORA (1064) −0.002 0.04 −0.00 [−0.09; 0.08] 7.7% 9.8%
NSPHS (866) −0.229 0.05 −0.23 [−0.33; −0.13] 5.4% 9.1%
ORCADES (982) −0.322 0.05 −0.32 [−0.41; −0.23] 6.5% 9.5%
RECOMBINE (448) −0.141 0.05 −0.14 [−0.24; −0.04] 5.6% 9.2%
STABILITY (2951) −0.097 0.03 −0.10 [−0.15; −0.05] 21.9% 11.2%
STANLEY (344) −0.207 0.09 −0.21 [−0.38; −0.04] 1.9% 6.3%
STANLEY (300) −0.161 0.09 −0.16 [−0.33; 0.01] 1.9% 6.2%
VIS (902) −0.301 0.05 −0.30 [−0.40; −0.21] 6.2% 9.4%
Common effect model −0.13 [−0.15; −0.11] 100.0% --
Random effects model −0.16 [−0.22; −0.10] -- 100.0%
−0.4 −0.2 0 0.2 0.4
Heterogeneity: $I^2$ = 80%, $\tau^2$ = 0.0080, $p$ < 0.01

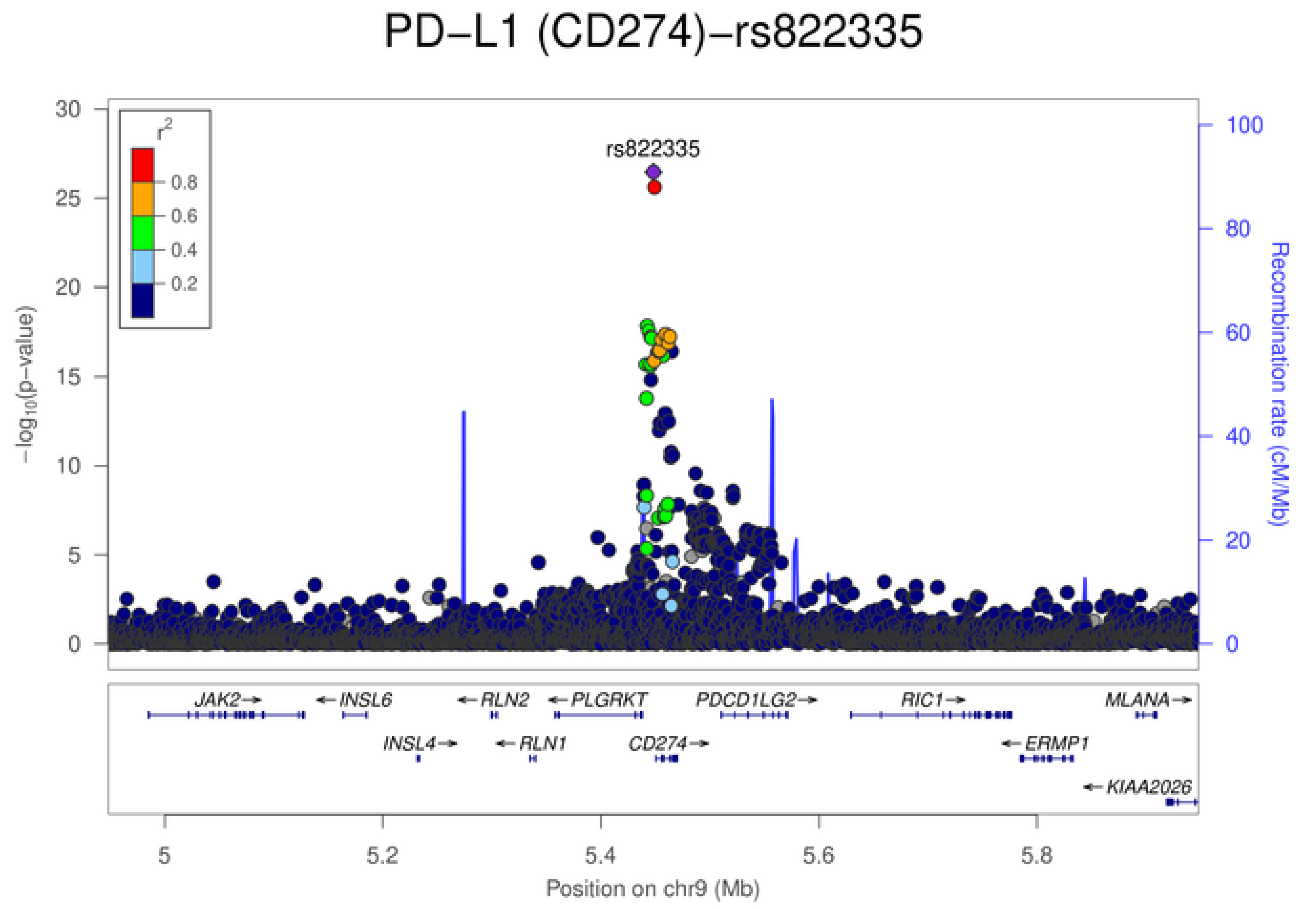
PD−L1 (CD274)−rs822335
rs822335
$r^2$
0.8
0.6
0.4
0.2
$-\log_{10}$(p−value)
0 5 10 15 20 25 30
Recombination rate (cM/Mb)
0 20 40 60 80 100
JAK2→
←INSL6
←RLN2
←PLGRKT
PDCD1LG2→
RIC1→
MLANA→
INSL4→
←RLN1
CD274→
←ERMP1
←KIAA2026
5 5.2 5.4 5.6 5.8
Position on chr9 (Mb)

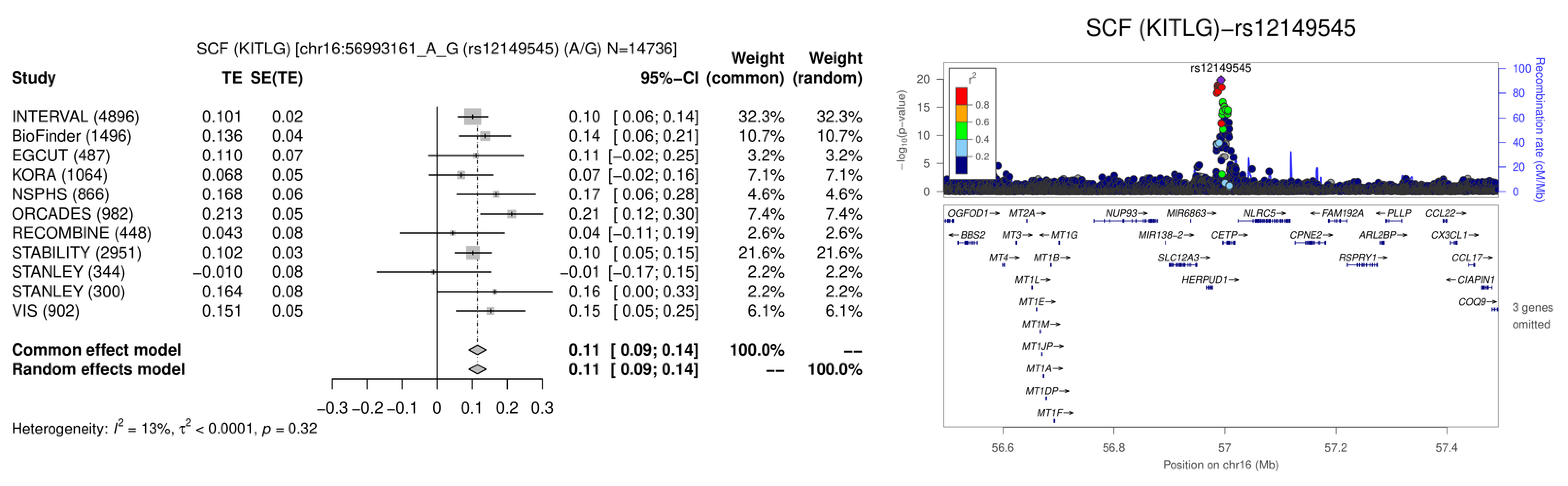
SCF (KITLG) [chr16:56993161_A_G (rs12149545) (A/G) N=14736]
Study
TE
SE(TE)
95%-CI
Weight (common)
Weight (random)
INTERVAL (4896) 0.101 0.02 0.10 [ 0.06; 0.14] 32.3% 32.3%
BioFinder (1496) 0.136 0.04 0.14 [ 0.06; 0.21] 10.7% 10.7%
EGCUT (487) 0.110 0.07 0.11 [−0.02; 0.25] 3.2% 3.2%
KORA (1064) 0.068 0.05 0.07 [−0.02; 0.16] 7.1% 7.1%
NSPHS (866) 0.168 0.06 0.17 [ 0.06; 0.28] 4.6% 4.6%
ORCADES (982) 0.213 0.05 0.21 [ 0.12; 0.30] 7.4% 7.4%
RECOMBINE (448) 0.043 0.08 0.04 [−0.11; 0.19] 2.6% 2.6%
STABILITY (2951) 0.102 0.03 0.10 [ 0.05; 0.15] 21.6% 21.6%
STANLEY (344) −0.010 0.08 −0.01 [−0.17; 0.15] 2.2% 2.2%
STANLEY (300) 0.164 0.08 0.16 [ 0.00; 0.33] 2.2% 2.2%
VIS (902) 0.151 0.05 0.15 [ 0.05; 0.25] 6.1% 6.1%
Common effect model 0.11 [ 0.09; 0.14] 100.0% --
Random effects model 0.11 [ 0.09; 0.14] -- 100.0%
−0.3 −0.2 −0.1 0 0.1 0.2 0.3
Heterogeneity: $I^2$ = 13%, $\tau^2$ < 0.0001, $p$ = 0.32
SCF (KITLG)−rs12149545
rs12149545
$-\log_{10}$(p−value)
Recombination rate (cM/Mb)
$r^2$
0.8
0.6
0.4
0.2
OGFOD1→ MT2A→ NUP93→ MIR6863→ NLRC5→ ←FAM192A ←PLLP CCL22→
←BBS2 MT3→ ←MT1G MIR138−2→ CETP→ CPNE2→ ARL2BP→ CX3CL1→
MT4→ MT1B→ SLC12A3→ RSPRY1→ CCL17→
MT1L→ HERPUD1→ ←CIAPIN1
MT1E→ COQ9→
MT1M→
MT1JP→
MT1A→
MT1DP→
MT1F→
3 genes omitted
56.6 56.8 57 57.2 57.4
Position on chr16 (Mb)

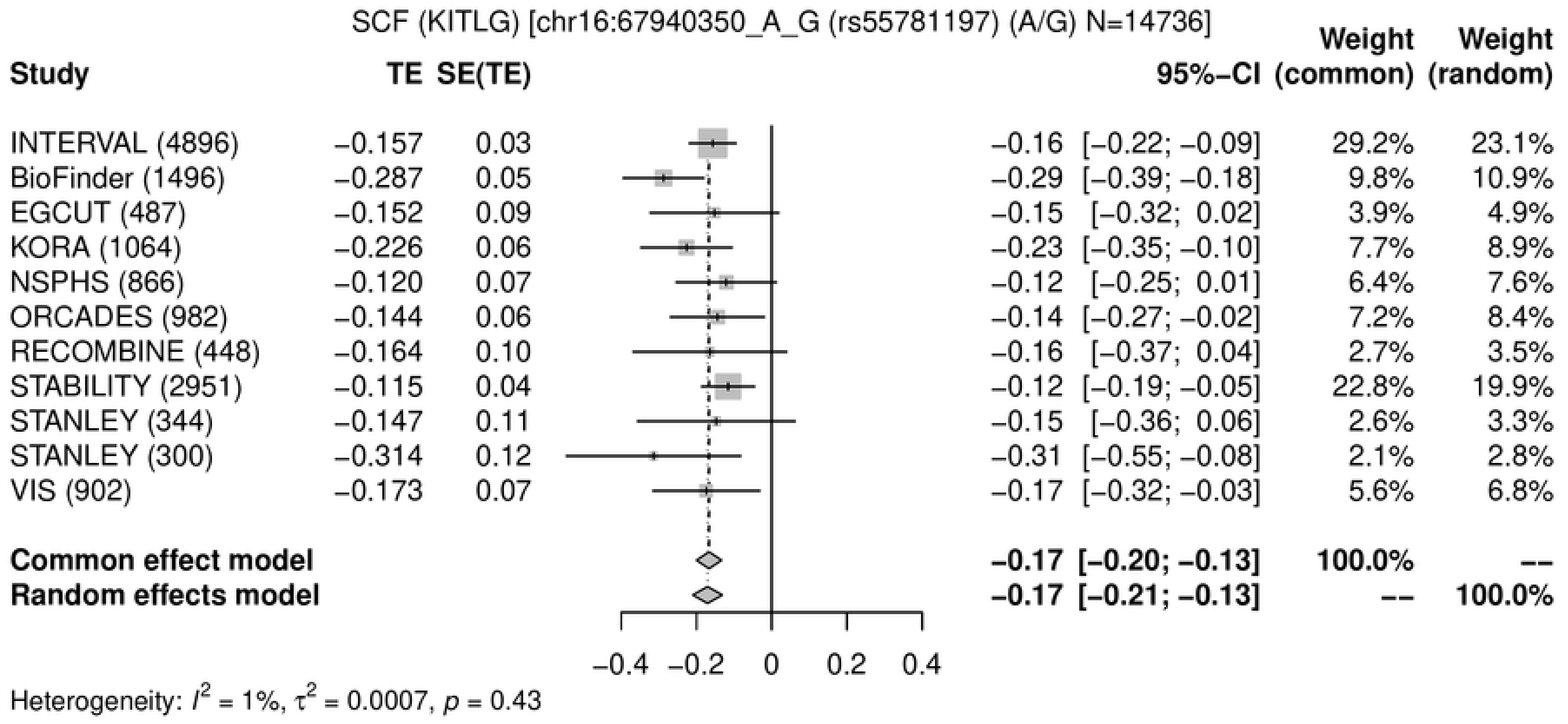
SCF (KITLG) [chr16:67940350_A_G (rs55781197) (A/G) N=14736]
Study
TE
SE(TE)
95%−CI
Weight (common)
Weight (random)
INTERVAL (4896) −0.157 0.03 −0.16 [−0.22; −0.09] 29.2% 23.1%
BioFinder (1496) −0.287 0.05 −0.29 [−0.39; −0.18] 9.8% 10.9%
EGCUT (487) −0.152 0.09 −0.15 [−0.32; 0.02] 3.9% 4.9%
KORA (1064) −0.226 0.06 −0.23 [−0.35; −0.10] 7.7% 8.9%
NSPHS (866) −0.120 0.07 −0.12 [−0.25; 0.01] 6.4% 7.6%
ORCADES (982) −0.144 0.06 −0.14 [−0.27; −0.02] 7.2% 8.4%
RECOMBINE (448) −0.164 0.10 −0.16 [−0.37; 0.04] 2.7% 3.5%
STABILITY (2951) −0.115 0.04 −0.12 [−0.19; −0.05] 22.8% 19.9%
STANLEY (344) −0.147 0.11 −0.15 [−0.36; 0.06] 2.6% 3.3%
STANLEY (300) −0.314 0.12 −0.31 [−0.55; −0.08] 2.1% 2.8%
VIS (902) −0.173 0.07 −0.17 [−0.32; −0.03] 5.6% 6.8%
Common effect model −0.17 [−0.20; −0.13] 100.0% --
Random effects model −0.17 [−0.21; −0.13] -- 100.0%
−0.4 −0.2 0 0.2 0.4
Heterogeneity: I² = 1%, τ² = 0.0007, p = 0.43

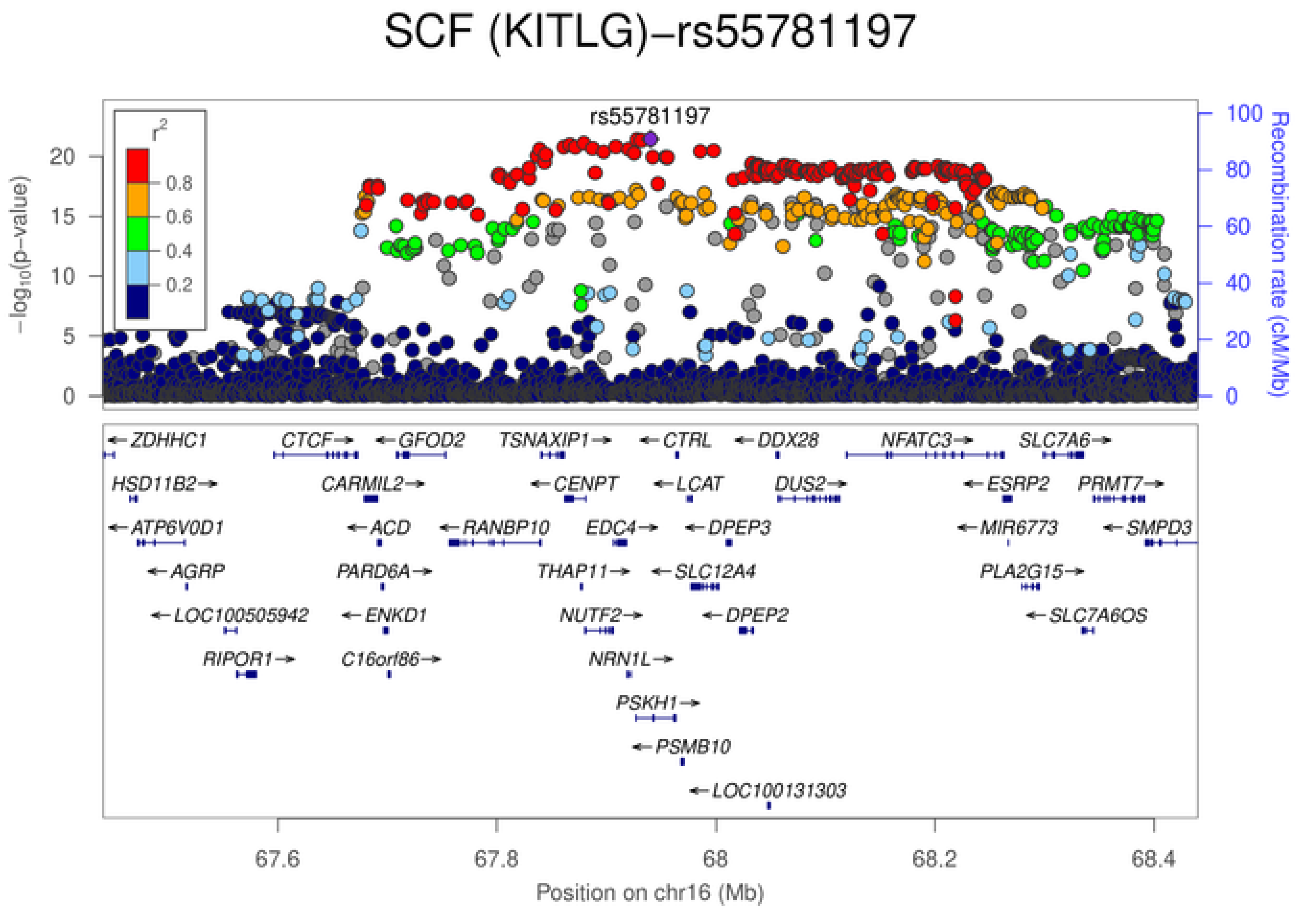
SCF (KITLG)−rs55781197
rs55781197
r²
0.8
0.6
0.4
0.2
−log10(p−value)
Recombination rate (cM/Mb)
ZDHHC1
CTCF
GFOD2
TSNAXIP1
CTRL
DDX28
NFATC3
SLC7A6
HSD11B2
CARMIL2
CENPT
LCAT
DUS2
ESRP2
PRMT7
ATP6V0D1
ACD
RANBP10
EDC4
DPEP3
MIR6773
SMPD3
AGRP
PARD6A
THAP11
SLC12A4
PLA2G15
LOC100505942
ENKD1
NUTF2
DPEP2
SLC7A6OS
RIPOR1
C16orf86
NRN1L
PSKH1
PSMB10
LOC100131303
67.6
67.8
68
68.2
68.4
Position on chr16 (Mb)

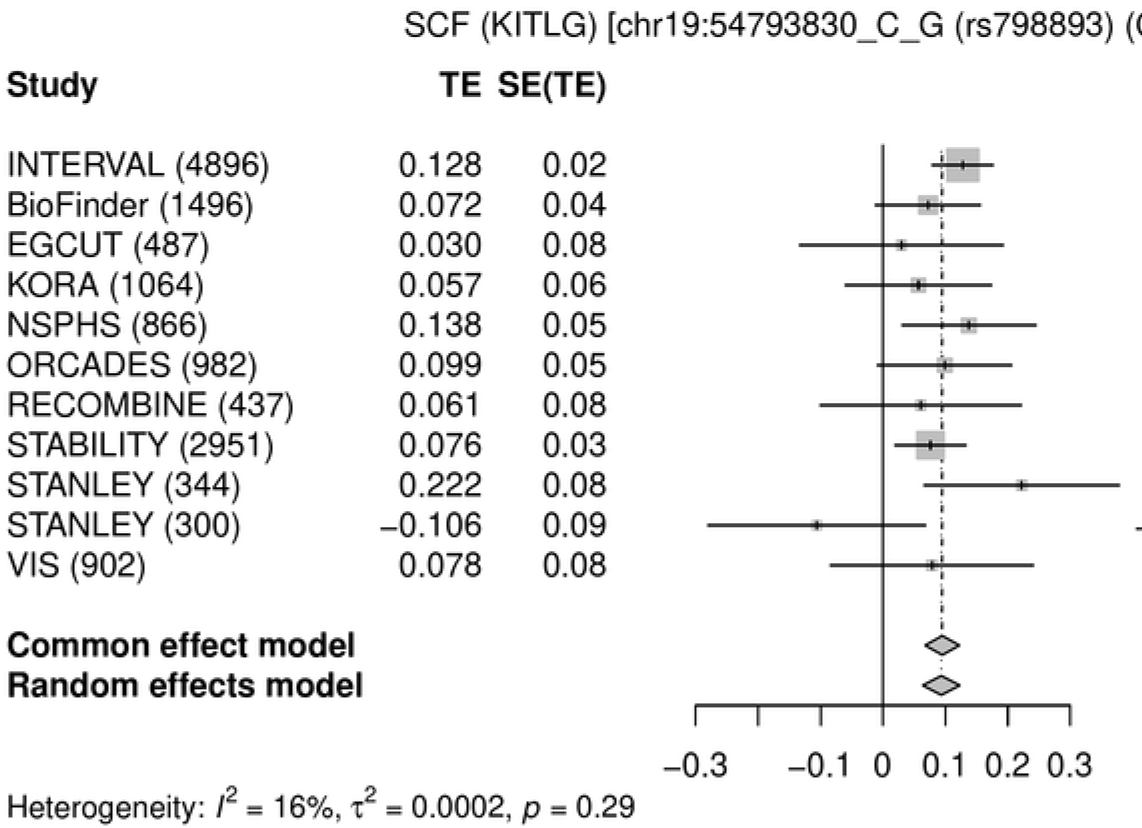
SCF (KITLG) [chr19:54793830_C_G (rs798893) (C/G) N=14725]
Study
TE
SE(TE)
95%-CI
Weight (common)
Weight (random)
INTERVAL (4896) 0.128 0.02 0.13 [ 0.08; 0.18] 32.2% 28.8%
BioFinder (1496) 0.072 0.04 0.07 [−0.01; 0.16] 10.8% 11.5%
EGCUT (487) 0.030 0.08 0.03 [−0.13; 0.19] 2.9% 3.3%
KORA (1064) 0.057 0.06 0.06 [−0.06; 0.17] 5.6% 6.2%
NSPHS (866) 0.138 0.05 0.14 [ 0.03; 0.25] 6.7% 7.3%
ORCADES (982) 0.099 0.05 0.10 [−0.01; 0.21] 6.6% 7.3%
RECOMBINE (437) 0.061 0.08 0.06 [−0.10; 0.22] 3.0% 3.3%
STABILITY (2951) 0.076 0.03 0.08 [ 0.02; 0.13] 23.8% 22.6%
STANLEY (344) 0.222 0.08 0.22 [ 0.07; 0.38] 3.1% 3.6%
STANLEY (300) −0.106 0.09 −0.11 [−0.28; 0.07] 2.5% 2.9%
VIS (902) 0.078 0.08 0.08 [−0.08; 0.24] 2.9% 3.3%
Common effect model 0.10 [ 0.07; 0.12] 100.0% --
Random effects model 0.09 [ 0.06; 0.12] -- 100.0%
−0.3 −0.1 0 0.1 0.2 0.3
Heterogeneity: $I^2$ = 16%, $\tau^2$ = 0.0002, $p$ = 0.29

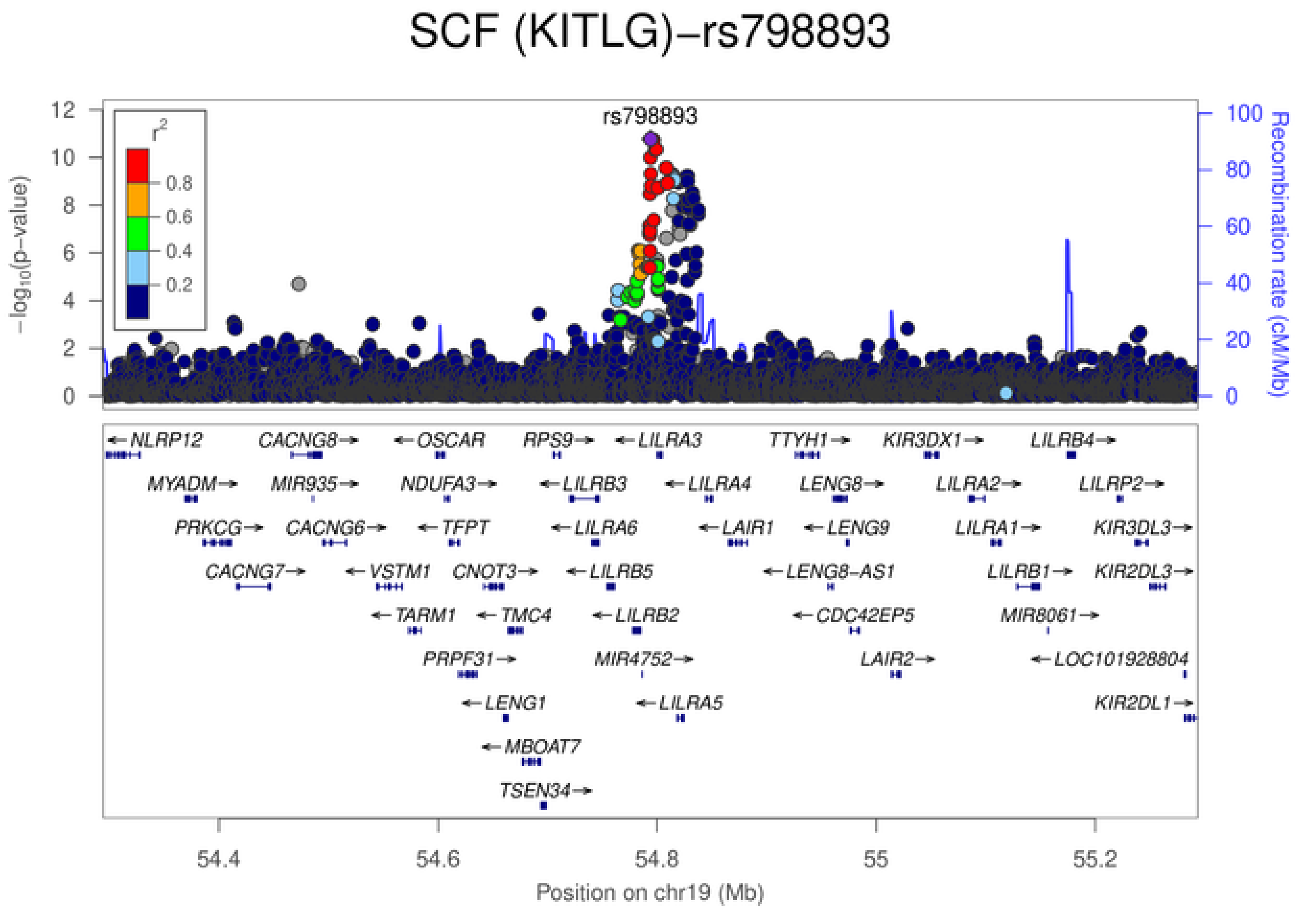
SCF (KITLG)−rs798893
rs798893
$-\log_{10}$(p-value)
Recombination rate (cM/Mb)
$r^2$
0.8
0.6
0.4
0.2
NLRP12
CACNG8
OSCAR
RPS9
LILRA3
TTYH1
KIR3DX1
LILRB4
MYADM
MIR935
NDUFA3
LILRB3
LILRA4
LENG8
LILRA2
LILRP2
PRKCG
CACNG6
TFPT
LILRA6
LAIR1
LENG9
LILRA1
KIR3DL3
CACNG7
VSTM1
CNOT3
LILRB5
LENG8-AS1
LILRB1
KIR2DL3
TARM1
TMC4
LILRB2
CDC42EP5
MIR8061
PRPF31
MIR4752
LAIR2
LOC101928804
LENG1
LILRA5
KIR2DL1
MBOAT7
TSEN34
54.4
54.6
54.8
55
55.2
Position on chr19 (Mb)

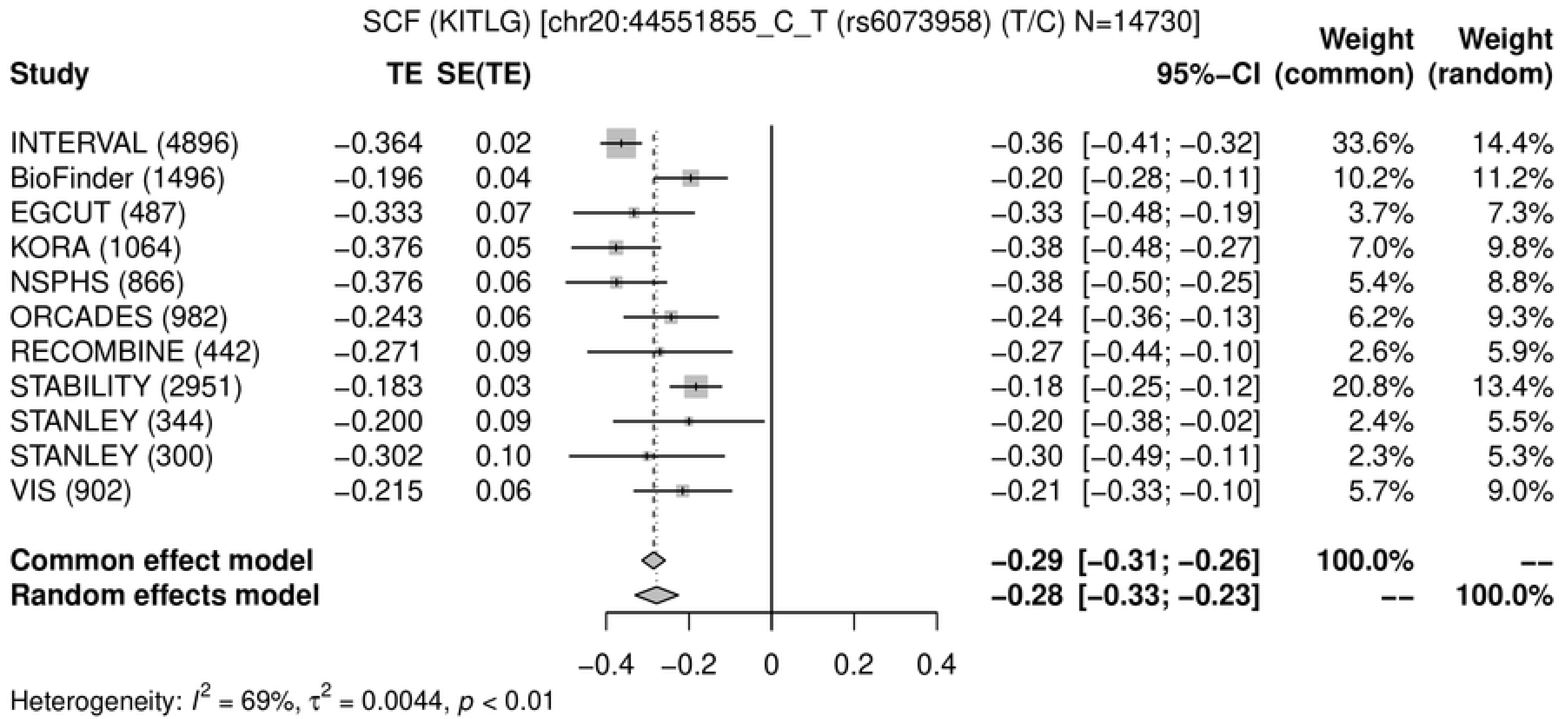
SCF (KITLG) [chr20:44551855_C_T (rs6073958) (T/C) N=14730]
Study TE SE(TE) 95%−CI Weight (common) Weight (random)
INTERVAL (4896) −0.364 0.02 −0.36 [−0.41; −0.32] 33.6% 14.4%
BioFinder (1496) −0.196 0.04 −0.20 [−0.28; −0.11] 10.2% 11.2%
EGCUT (487) −0.333 0.07 −0.33 [−0.48; −0.19] 3.7% 7.3%
KORA (1064) −0.376 0.05 −0.38 [−0.48; −0.27] 7.0% 9.8%
NSPHS (866) −0.376 0.06 −0.38 [−0.50; −0.25] 5.4% 8.8%
ORCADES (982) −0.243 0.06 −0.24 [−0.36; −0.13] 6.2% 9.3%
RECOMBINE (442) −0.271 0.09 −0.27 [−0.44; −0.10] 2.6% 5.9%
STABILITY (2951) −0.183 0.03 −0.18 [−0.25; −0.12] 20.8% 13.4%
STANLEY (344) −0.200 0.09 −0.20 [−0.38; −0.02] 2.4% 5.5%
STANLEY (300) −0.302 0.10 −0.30 [−0.49; −0.11] 2.3% 5.3%
VIS (902) −0.215 0.06 −0.21 [−0.33; −0.10] 5.7% 9.0%
Common effect model −0.29 [−0.31; −0.26] 100.0% --
Random effects model −0.28 [−0.33; −0.23] -- 100.0%
−0.4 −0.2 0 0.2 0.4
Heterogeneity: $I^2$ = 69%, $\tau^2$ = 0.0044, $p$ < 0.01

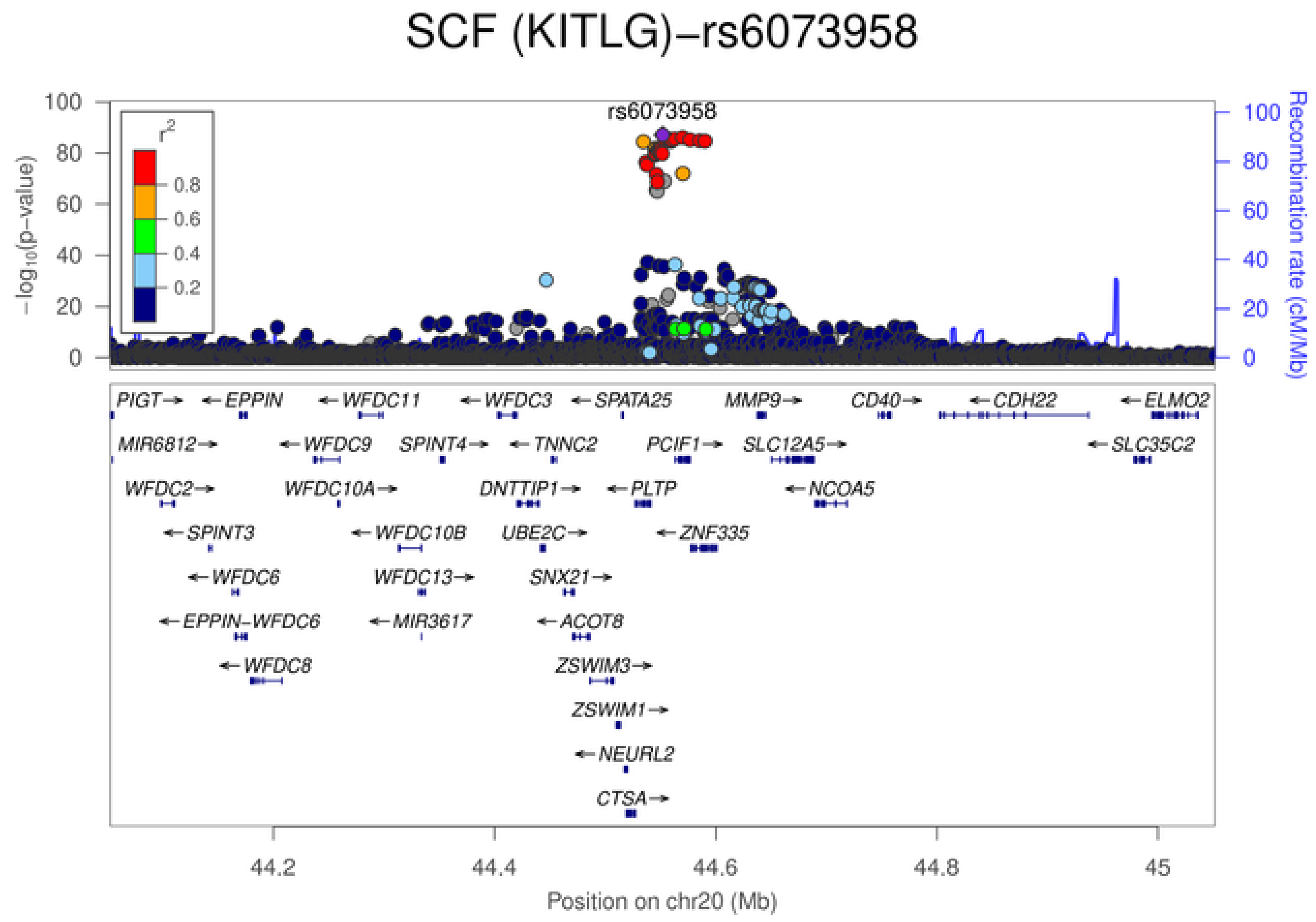
SCF (KITLG)−rs6073958
rs6073958
$-\log_{10}$(p−value)
Recombination rate (cM/Mb)
$r^2$
0.8
0.6
0.4
0.2
0
20
40
60
80
100
PIGT→ ←EPPIN ←WFDC11 ←WFDC3 ←SPATA25 MMP9→ CD40→ ←CDH22 ←ELMO2
MIR6812→ ←WFDC9 SPINT4→ ←TNNC2 PCIF1→ SLC12A5→ ←SLC35C2
WFDC2→ WFDC10A→ DNTTIP1→ ←PLTP ←NCOA5
←SPINT3 ←WFDC10B UBE2C→ ←ZNF335
←WFDC6 WFDC13→ SNX21→
←EPPIN−WFDC6 ←MIR3617 ←ACOT8
←WFDC8 ZSWIM3→
ZSWIM1→
←NEURL2
CTSA→
44.2
44.4
44.6
44.8
45
Position on chr20 (Mb)

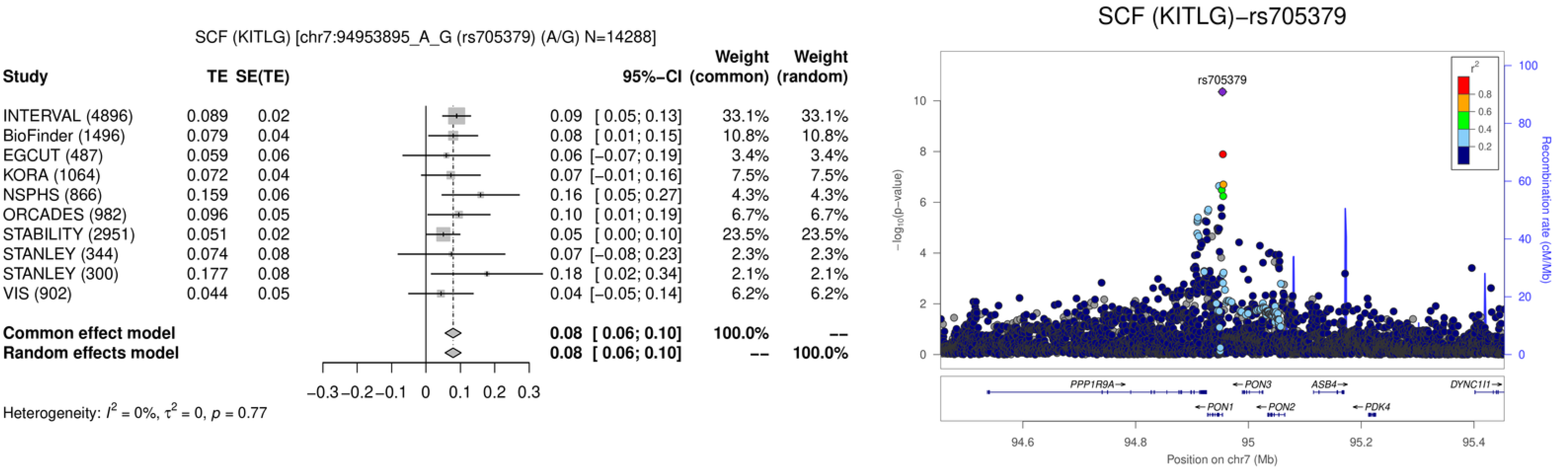

SCF (KITLG) [chr7:94953895_A_G (rs705379) (A/G) N=14288]
Study
TE
SE(TE)
95%−CI
Weight (common)
Weight (random)
INTERVAL (4896) 0.089 0.02 0.09 [ 0.05; 0.13] 33.1% 33.1%
BioFinder (1496) 0.079 0.04 0.08 [ 0.01; 0.15] 10.8% 10.8%
EGCUT (487) 0.059 0.06 0.06 [−0.07; 0.19] 3.4% 3.4%
KORA (1064) 0.072 0.04 0.07 [−0.01; 0.16] 7.5% 7.5%
NSPHS (866) 0.159 0.06 0.16 [ 0.05; 0.27] 4.3% 4.3%
ORCADES (982) 0.096 0.05 0.10 [ 0.01; 0.19] 6.7% 6.7%
STABILITY (2951) 0.051 0.02 0.05 [ 0.00; 0.10] 23.5% 23.5%
STANLEY (344) 0.074 0.08 0.07 [−0.08; 0.23] 2.3% 2.3%
STANLEY (300) 0.177 0.08 0.18 [ 0.02; 0.34] 2.1% 2.1%
VIS (902) 0.044 0.05 0.04 [−0.05; 0.14] 6.2% 6.2%
Common effect model 0.08 [ 0.06; 0.10] 100.0% --
Random effects model 0.08 [ 0.06; 0.10] -- 100.0%
−0.3 −0.2 −0.1 0 0.1 0.2 0.3
Heterogeneity: $I^2$ = 0%, $\tau^2$ = 0, $p$ = 0.77
SCF (KITLG)−rs705379
rs705379
$-\log_{10}$(p−value)
Recombination rate (cM/Mb)
$r^2$
0.8
0.6
0.4
0.2
PPP1R9A→
←PON3
ASB4→
DYNC1I1→
←PON1
←PON2
←PDK4
94.6
94.8
95
95.2
95.4
Position on chr7 (Mb)

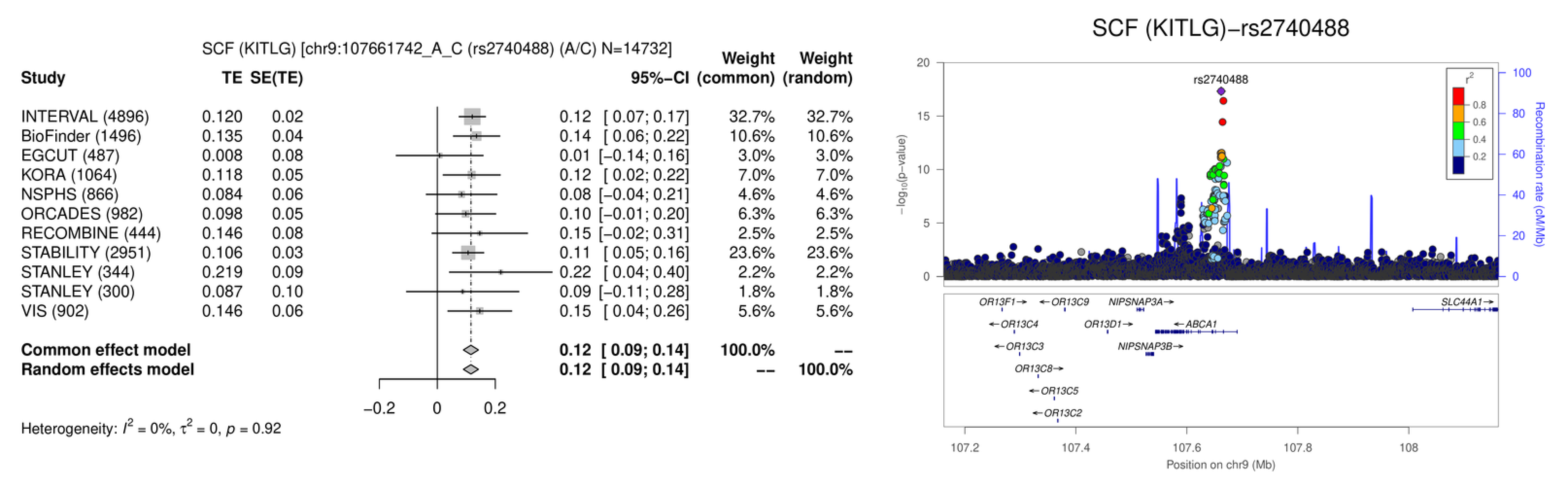
SCF (KITLG) [chr9:107661742_A_C (rs2740488) (A/C) N=14732]
Study
TE
SE(TE)
95%−CI
Weight (common)
Weight (random)
INTERVAL (4896) 0.120 0.02 0.12 [ 0.07; 0.17] 32.7% 32.7%
BioFinder (1496) 0.135 0.04 0.14 [ 0.06; 0.22] 10.6% 10.6%
EGCUT (487) 0.008 0.08 0.01 [−0.14; 0.16] 3.0% 3.0%
KORA (1064) 0.118 0.05 0.12 [ 0.02; 0.22] 7.0% 7.0%
NSPHS (866) 0.084 0.06 0.08 [−0.04; 0.21] 4.6% 4.6%
ORCADES (982) 0.098 0.05 0.10 [−0.01; 0.20] 6.3% 6.3%
RECOMBINE (444) 0.146 0.08 0.15 [−0.02; 0.31] 2.5% 2.5%
STABILITY (2951) 0.106 0.03 0.11 [ 0.05; 0.16] 23.6% 23.6%
STANLEY (344) 0.219 0.09 0.22 [ 0.04; 0.40] 2.2% 2.2%
STANLEY (300) 0.087 0.10 0.09 [−0.11; 0.28] 1.8% 1.8%
VIS (902) 0.146 0.06 0.15 [ 0.04; 0.26] 5.6% 5.6%
Common effect model 0.12 [ 0.09; 0.14] 100.0% --
Random effects model 0.12 [ 0.09; 0.14] -- 100.0%
−0.2
0
0.2
Heterogeneity: $I^2$ = 0%, $\tau^2$ = 0, $p$ = 0.92
SCF (KITLG)−rs2740488
rs2740488
$-\log_{10}$(p-value)
Recombination rate (cM/Mb)
$r^2$
0.8
0.6
0.4
0.2
OR13F1→
←OR13C9
NIPSNAP3A→
SLC44A1→
←OR13C4
OR13D1→
←ABCA1
←OR13C3
NIPSNAP3B→
OR13C8→
←OR13C5
←OR13C2
107.2
107.4
107.6
107.8
108
Position on chr9 (Mb)

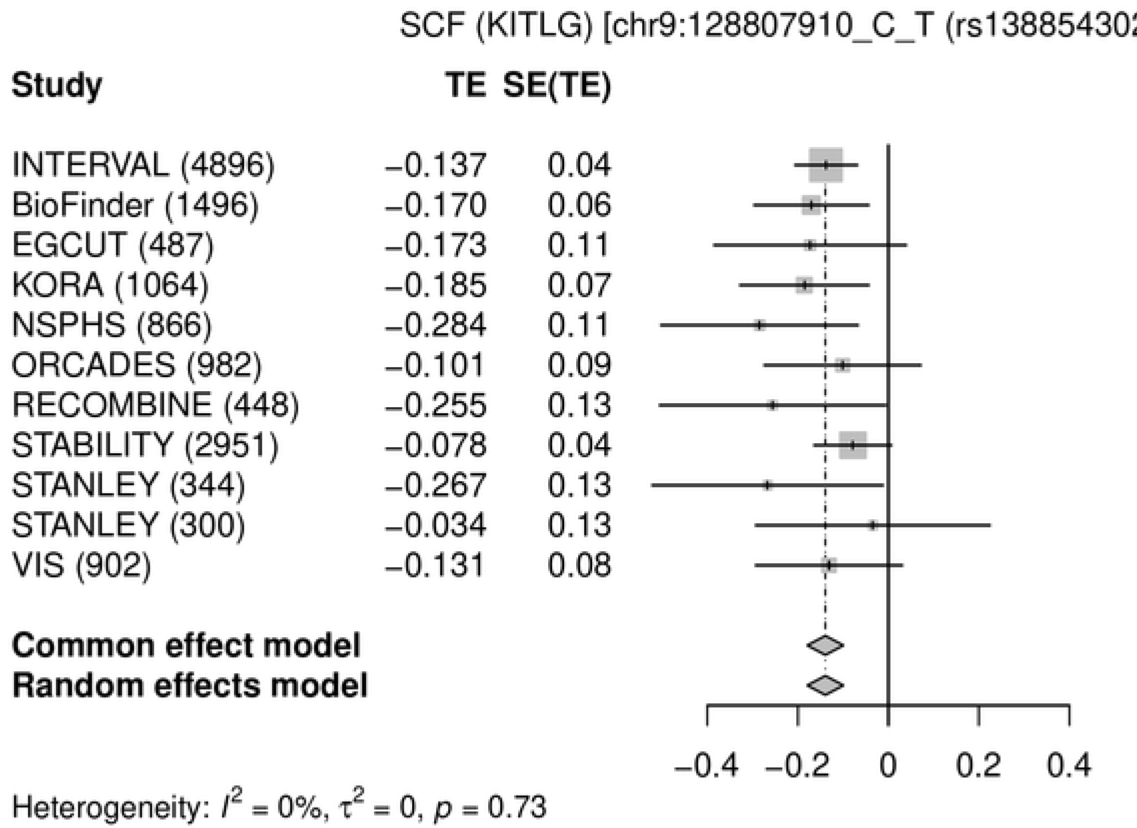

SCF (KITLG) [chr9:128807910_C_T (rs138854302) (T/C) N=14736]
Study
TE
SE(TE)
95%−CI
Weight (common)
Weight (random)
INTERVAL (4896) −0.137 0.04 −0.14 [−0.21; −0.07] 33.9% 33.9%
BioFinder (1496) −0.170 0.06 −0.17 [−0.30; −0.04] 10.1% 10.1%
EGCUT (487) −0.173 0.11 −0.17 [−0.38; 0.04] 3.6% 3.6%
KORA (1064) −0.185 0.07 −0.19 [−0.33; −0.04] 8.0% 8.0%
NSPHS (866) −0.284 0.11 −0.28 [−0.50; −0.07] 3.4% 3.4%
ORCADES (982) −0.101 0.09 −0.10 [−0.27; 0.07] 5.4% 5.4%
RECOMBINE (448) −0.255 0.13 −0.26 [−0.51; −0.01] 2.6% 2.6%
STABILITY (2951) −0.078 0.04 −0.08 [−0.16; 0.01] 22.0% 22.0%
STANLEY (344) −0.267 0.13 −0.27 [−0.52; −0.01] 2.5% 2.5%
STANLEY (300) −0.034 0.13 −0.03 [−0.29; 0.22] 2.4% 2.4%
VIS (902) −0.131 0.08 −0.13 [−0.29; 0.03] 6.2% 6.2%
Common effect model −0.14 [−0.18; −0.10] 100.0% --
Random effects model −0.14 [−0.18; −0.10] -- 100.0%
−0.4 −0.2 0 0.2 0.4
Heterogeneity: $I^2$ = 0%, $\tau^2$ = 0, $p$ = 0.73


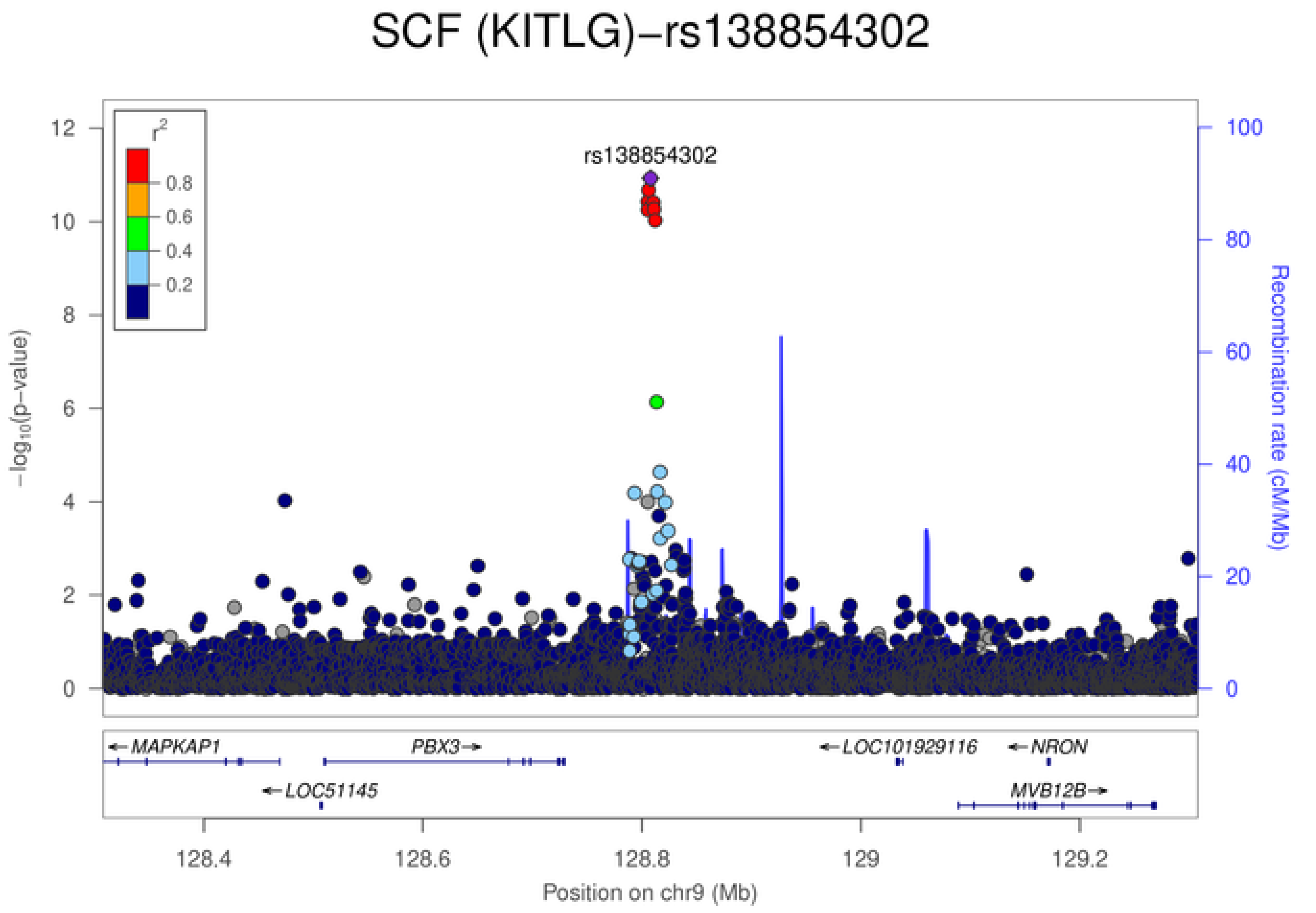

SCF (KITLG)−rs138854302
rs138854302
$r^2$
0.8
0.6
0.4
0.2
$-\log_{10}$(p−value)
Recombination rate (cM/Mb)
MAPKAP1
PBX3
LOC51145
LOC101929116
NRON
MVB12B
128.4
128.6
128.8
129
129.2
Position on chr9 (Mb)

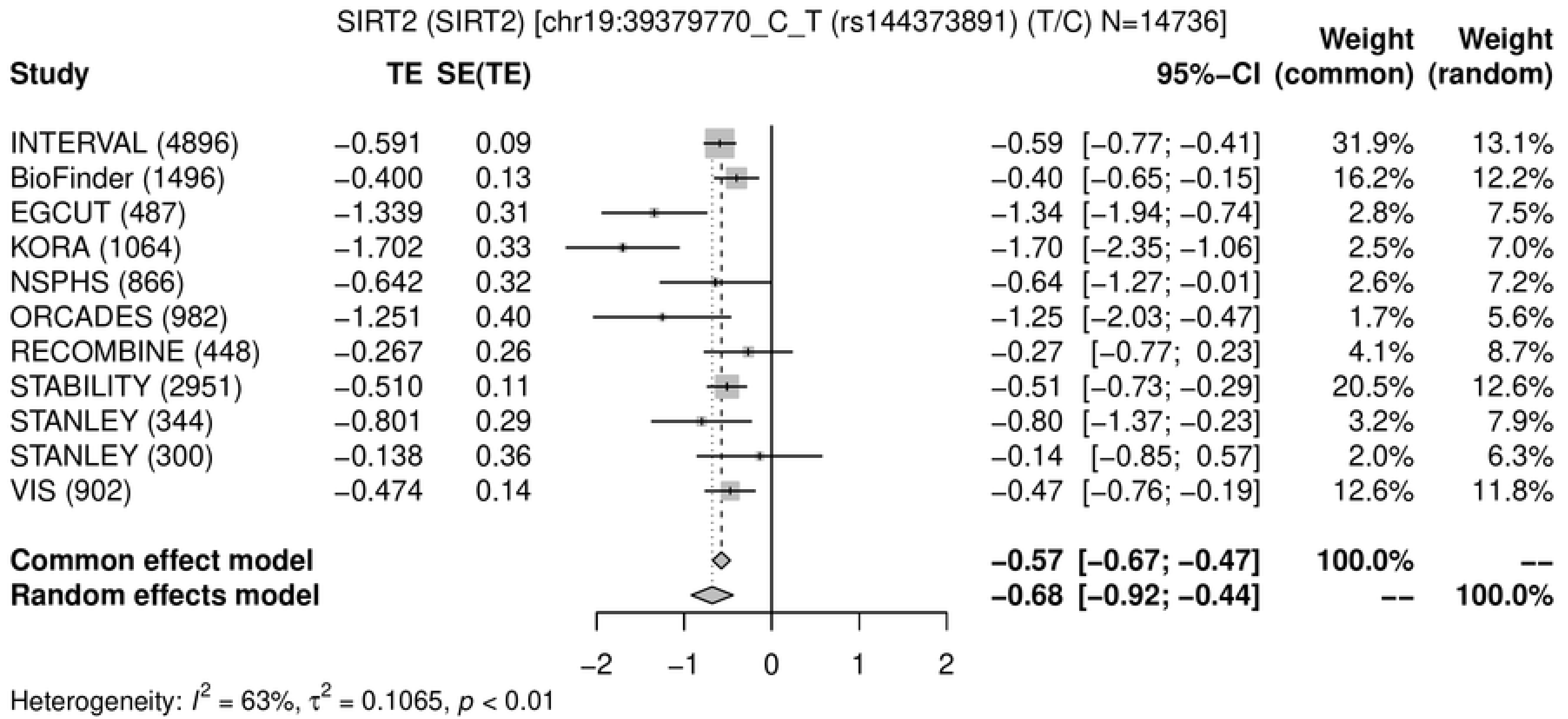
SIRT2 (SIRT2) [chr19:39379770_C_T (rs144373891) (T/C) N=14736]
Study | TE | SE(TE) | 95%-CI | Weight (common) | Weight (random)
INTERVAL (4896) | −0.591 | 0.09 | −0.59 [−0.77; −0.41] | 31.9% | 13.1%
BioFinder (1496) | −0.400 | 0.13 | −0.40 [−0.65; −0.15] | 16.2% | 12.2%
EGCUT (487) | −1.339 | 0.31 | −1.34 [−1.94; −0.74] | 2.8% | 7.5%
KORA (1064) | −1.702 | 0.33 | −1.70 [−2.35; −1.06] | 2.5% | 7.0%
NSPHS (866) | −0.642 | 0.32 | −0.64 [−1.27; −0.01] | 2.6% | 7.2%
ORCADES (982) | −1.251 | 0.40 | −1.25 [−2.03; −0.47] | 1.7% | 5.6%
RECOMBINE (448) | −0.267 | 0.26 | −0.27 [−0.77; 0.23] | 4.1% | 8.7%
STABILITY (2951) | −0.510 | 0.11 | −0.51 [−0.73; −0.29] | 20.5% | 12.6%
STANLEY (344) | −0.801 | 0.29 | −0.80 [−1.37; −0.23] | 3.2% | 7.9%
STANLEY (300) | −0.138 | 0.36 | −0.14 [−0.85; 0.57] | 2.0% | 6.3%
VIS (902) | −0.474 | 0.14 | −0.47 [−0.76; −0.19] | 12.6% | 11.8%
Common effect model | −0.57 [−0.67; −0.47] | 100.0% | --
Random effects model | −0.68 [−0.92; −0.44] | -- | 100.0%
−2 −1 0 1 2
Heterogeneity: $I^2$ = 63%, $\tau^2$ = 0.1065, $p$ < 0.01

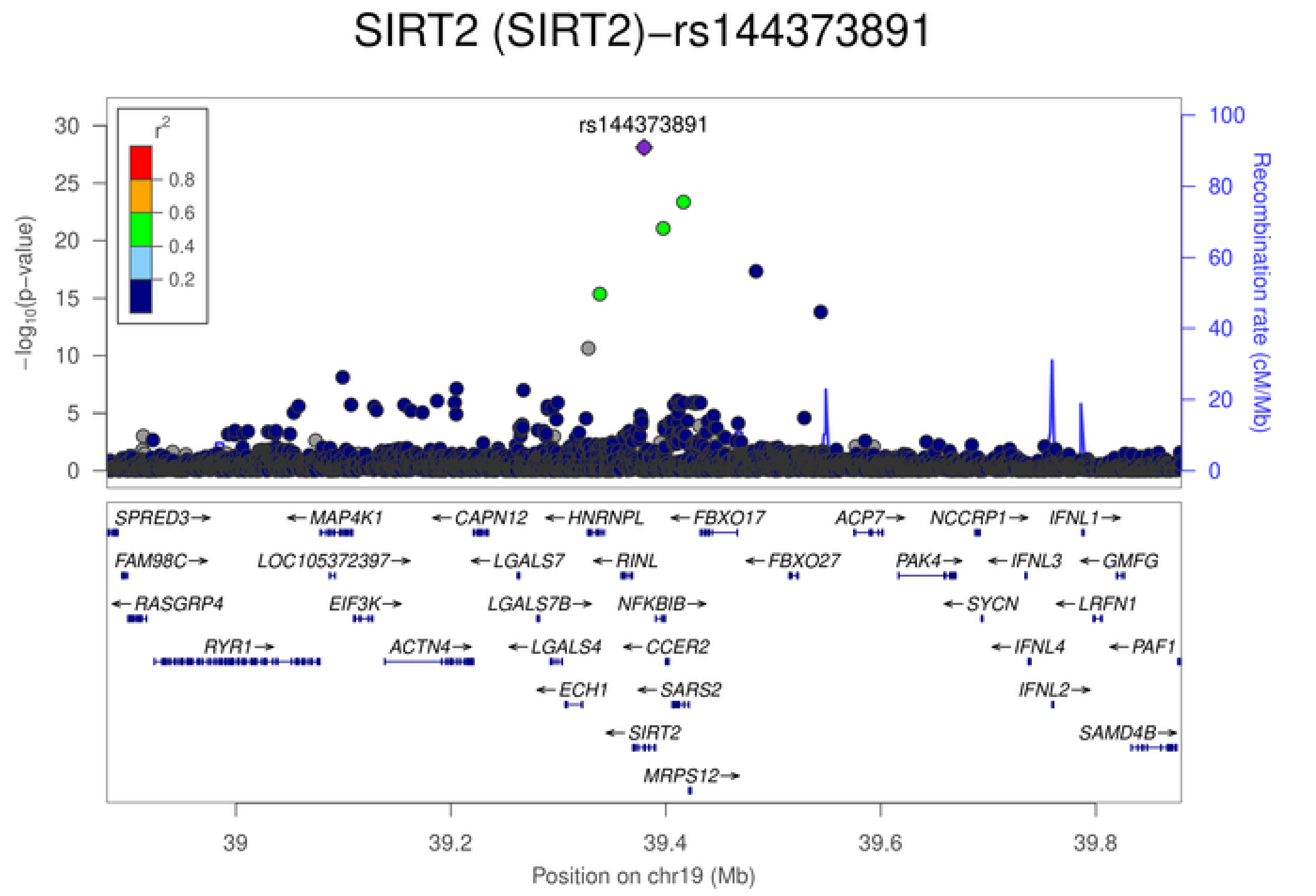
SIRT2 (SIRT2)−rs144373891
rs144373891
$r^2$
0.8
0.6
0.4
0.2
$-\log_{10}$(p-value)
Recombination rate (cM/Mb)
SPRED3 MAP4K1 CAPN12 HNRNPL FBXO17 ACP7 NCCRP1 IFNL1
FAM98C LOC105372397 LGALS7 RINL FBXO27 PAK4 IFNL3 GMFG
RASGRP4 EIF3K LGALS7B NFKBIB SYCN LRFN1
RYR1 ACTN4 LGALS4 CCER2 IFNL4 PAF1
ECH1 SARS2 IFNL2
SIRT2 SAMD4B
MRPS12
39 39.2 39.4 39.6 39.8
Position on chr19 (Mb)

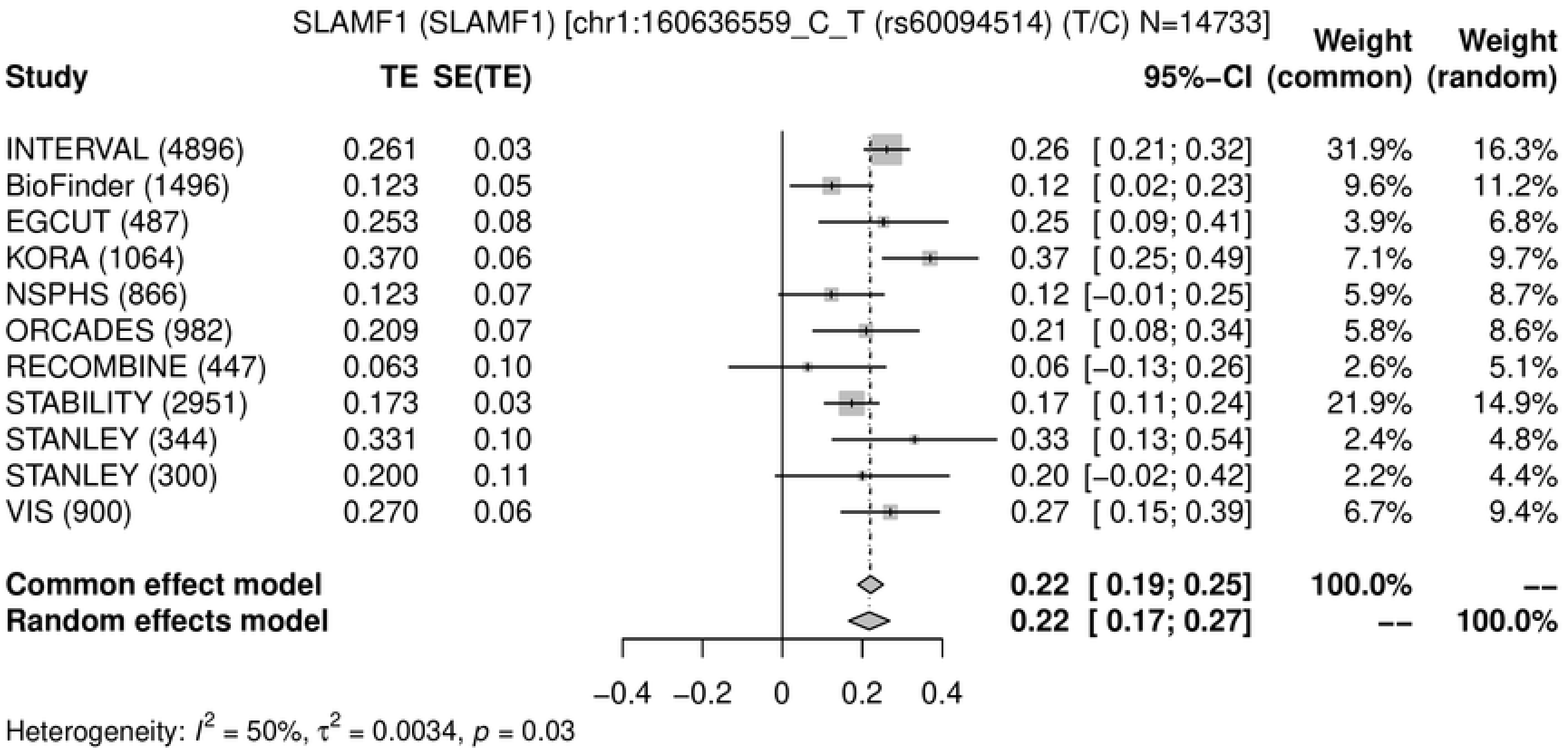
SLAMF1 (SLAMF1) [chr1:160636559_C_T (rs60094514) (T/C) N=14733]
Study
TE
SE(TE)
95%−CI
Weight (common)
Weight (random)
INTERVAL (4896) 0.261 0.03 0.26 [ 0.21; 0.32] 31.9% 16.3%
BioFinder (1496) 0.123 0.05 0.12 [ 0.02; 0.23] 9.6% 11.2%
EGCUT (487) 0.253 0.08 0.25 [ 0.09; 0.41] 3.9% 6.8%
KORA (1064) 0.370 0.06 0.37 [ 0.25; 0.49] 7.1% 9.7%
NSPHS (866) 0.123 0.07 0.12 [−0.01; 0.25] 5.9% 8.7%
ORCADES (982) 0.209 0.07 0.21 [ 0.08; 0.34] 5.8% 8.6%
RECOMBINE (447) 0.063 0.10 0.06 [−0.13; 0.26] 2.6% 5.1%
STABILITY (2951) 0.173 0.03 0.17 [ 0.11; 0.24] 21.9% 14.9%
STANLEY (344) 0.331 0.10 0.33 [ 0.13; 0.54] 2.4% 4.8%
STANLEY (300) 0.200 0.11 0.20 [−0.02; 0.42] 2.2% 4.4%
VIS (900) 0.270 0.06 0.27 [ 0.15; 0.39] 6.7% 9.4%
Common effect model 0.22 [ 0.19; 0.25] 100.0% --
Random effects model 0.22 [ 0.17; 0.27] -- 100.0%
−0.4 −0.2 0 0.2 0.4
Heterogeneity: $I^2$ = 50%, $\tau^2$ = 0.0034, $p$ = 0.03

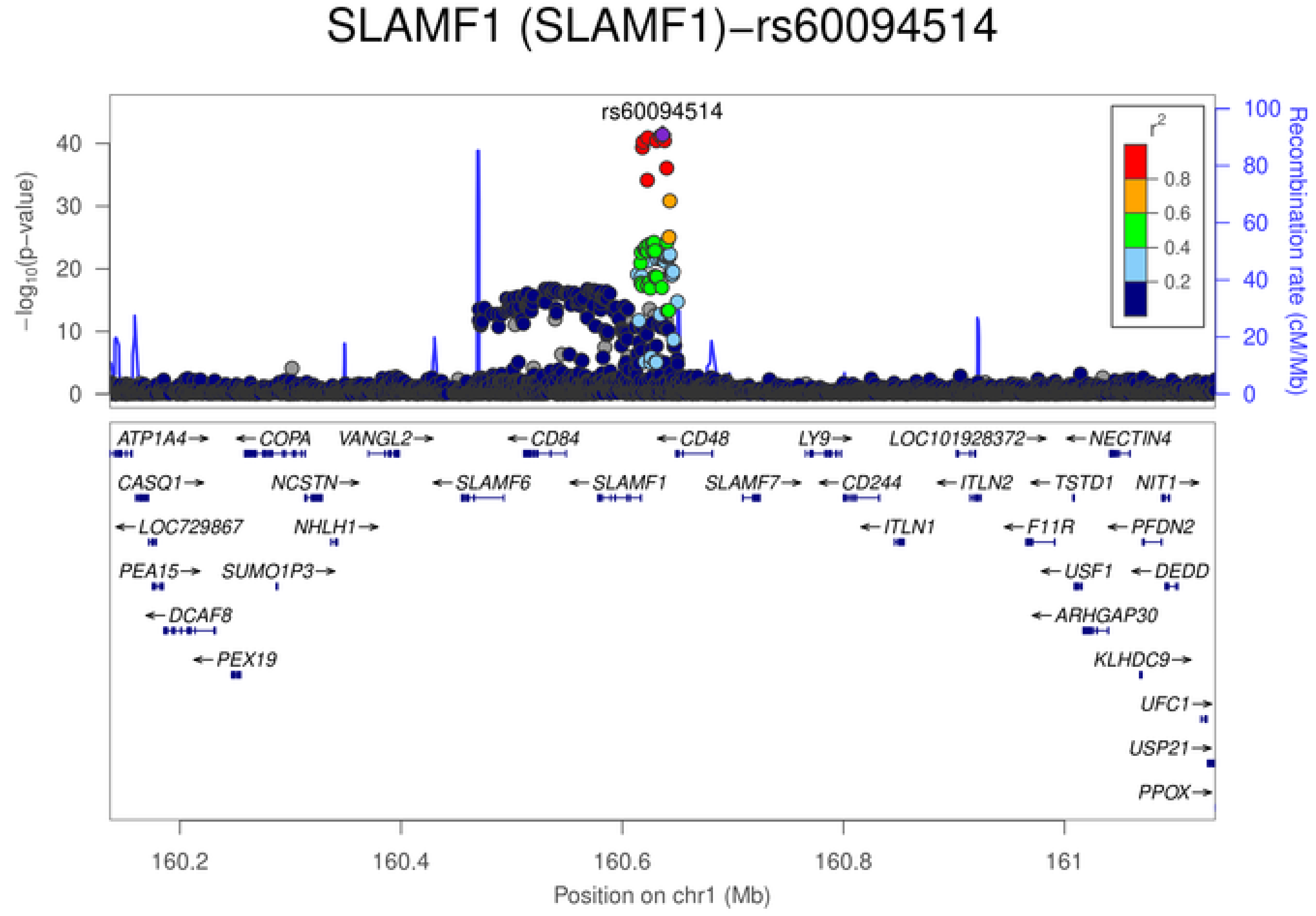
SLAMF1 (SLAMF1)−rs60094514
rs60094514
$-\log_{10}$(p−value)
0
10
20
30
40
Recombination rate (cM/Mb)
0
20
40
60
80
100
$r^2$
0.8
0.6
0.4
0.2
ATP1A4
COPA
VANGL2
CD84
CD48
LY9
LOC101928372
NECTIN4
CASQ1
NCSTN
SLAMF6
SLAMF1
SLAMF7
CD244
ITLN2
TSTD1
NIT1
LOC729867
NHLH1
ITLN1
F11R
PFDN2
PEA15
SUMO1P3
USF1
DEDD
DCAF8
ARHGAP30
PEX19
KLHDC9
UFC1
USP21
PPOX
160.2
160.4
160.6
160.8
161
Position on chr1 (Mb)

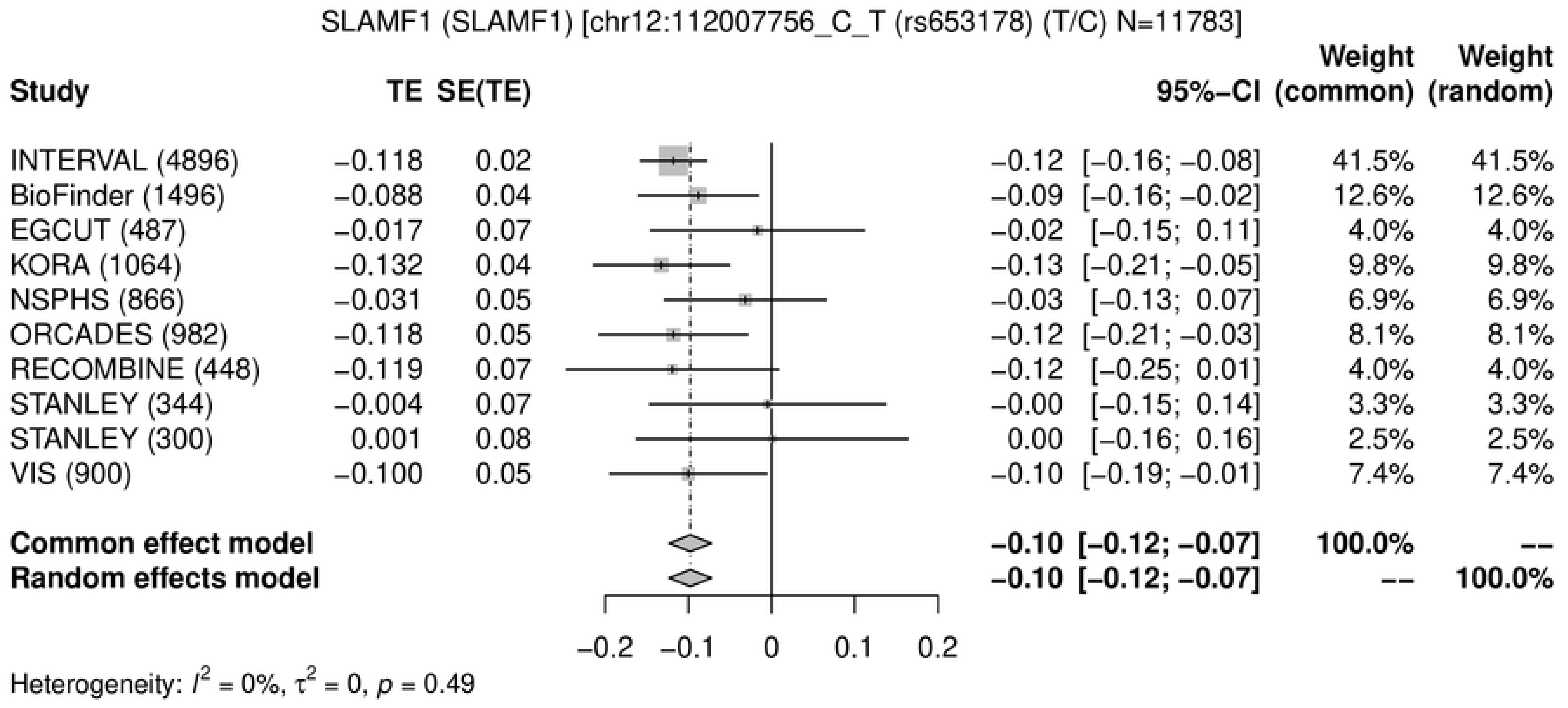
SLAMF1 (SLAMF1) [chr12:112007756_C_T (rs653178) (T/C) N=11783]
Study
TE
SE(TE)
95%-CI
Weight (common)
Weight (random)
INTERVAL (4896) −0.118 0.02 −0.12 [−0.16; −0.08] 41.5% 41.5%
BioFinder (1496) −0.088 0.04 −0.09 [−0.16; −0.02] 12.6% 12.6%
EGCUT (487) −0.017 0.07 −0.02 [−0.15; 0.11] 4.0% 4.0%
KORA (1064) −0.132 0.04 −0.13 [−0.21; −0.05] 9.8% 9.8%
NSPHS (866) −0.031 0.05 −0.03 [−0.13; 0.07] 6.9% 6.9%
ORCADES (982) −0.118 0.05 −0.12 [−0.21; −0.03] 8.1% 8.1%
RECOMBINE (448) −0.119 0.07 −0.12 [−0.25; 0.01] 4.0% 4.0%
STANLEY (344) −0.004 0.07 −0.00 [−0.15; 0.14] 3.3% 3.3%
STANLEY (300) 0.001 0.08 0.00 [−0.16; 0.16] 2.5% 2.5%
VIS (900) −0.100 0.05 −0.10 [−0.19; −0.01] 7.4% 7.4%
Common effect model −0.10 [−0.12; −0.07] 100.0% --
Random effects model −0.10 [−0.12; −0.07] -- 100.0%
−0.2 −0.1 0 0.1 0.2
Heterogeneity: $I^2$ = 0%, $\tau^2$ = 0, $p$ = 0.49

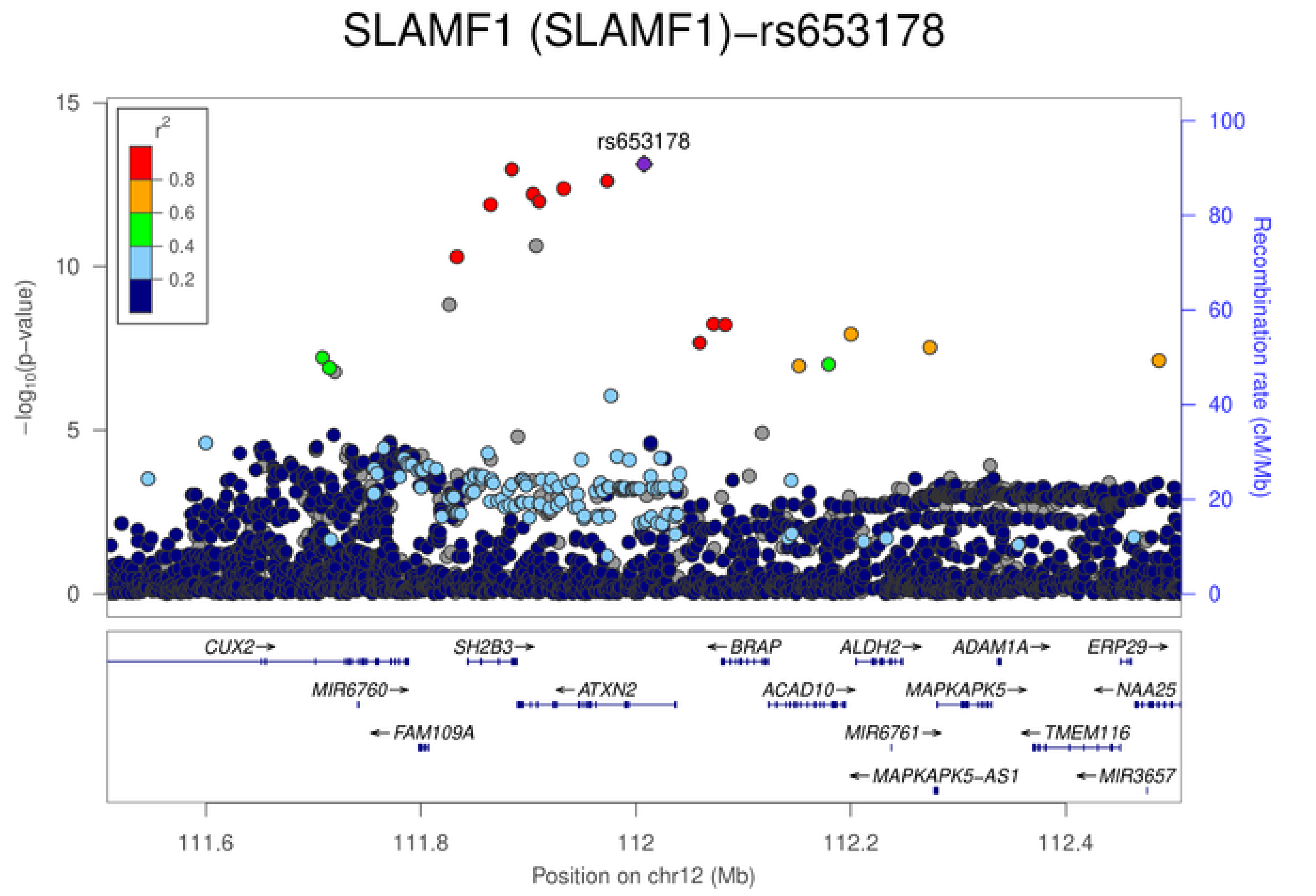
SLAMF1 (SLAMF1)−rs653178
rs653178
$r^2$
0.8
0.6
0.4
0.2
$-\log_{10}$(p-value)
Recombination rate (cM/Mb)
CUX2→
SH2B3→
←BRAP
ALDH2→
ADAM1A→
ERP29→
MIR6760→
←ATXN2
ACAD10→
MAPKAPK5→
←NAA25
←FAM109A
MIR6761→
←TMEM116
←MAPKAPK5-AS1
←MIR3657
111.6
111.8
112
112.2
112.4
Position on chr12 (Mb)

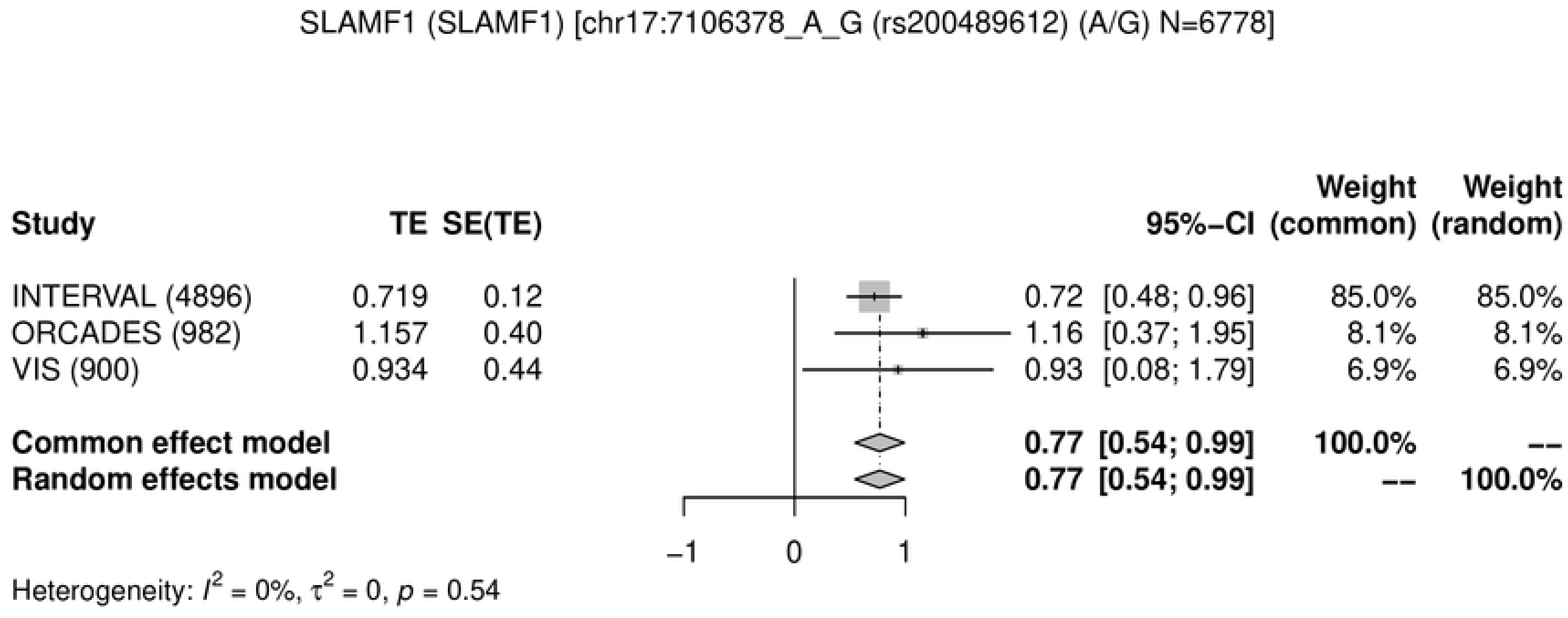
SLAMF1 (SLAMF1) [chr17:7106378_A_G (rs200489612) (A/G) N=6778]
Study
TE
SE(TE)
95%−CI
Weight (common)
Weight (random)
INTERVAL (4896) 0.719 0.12 0.72 [0.48; 0.96] 85.0% 85.0%
ORCADES (982) 1.157 0.40 1.16 [0.37; 1.95] 8.1% 8.1%
VIS (900) 0.934 0.44 0.93 [0.08; 1.79] 6.9% 6.9%
Common effect model 0.77 [0.54; 0.99] 100.0% --
Random effects model 0.77 [0.54; 0.99] -- 100.0%
−1
0
1
Heterogeneity: $I^2$ = 0%, $\tau^2$ = 0, $p$ = 0.54

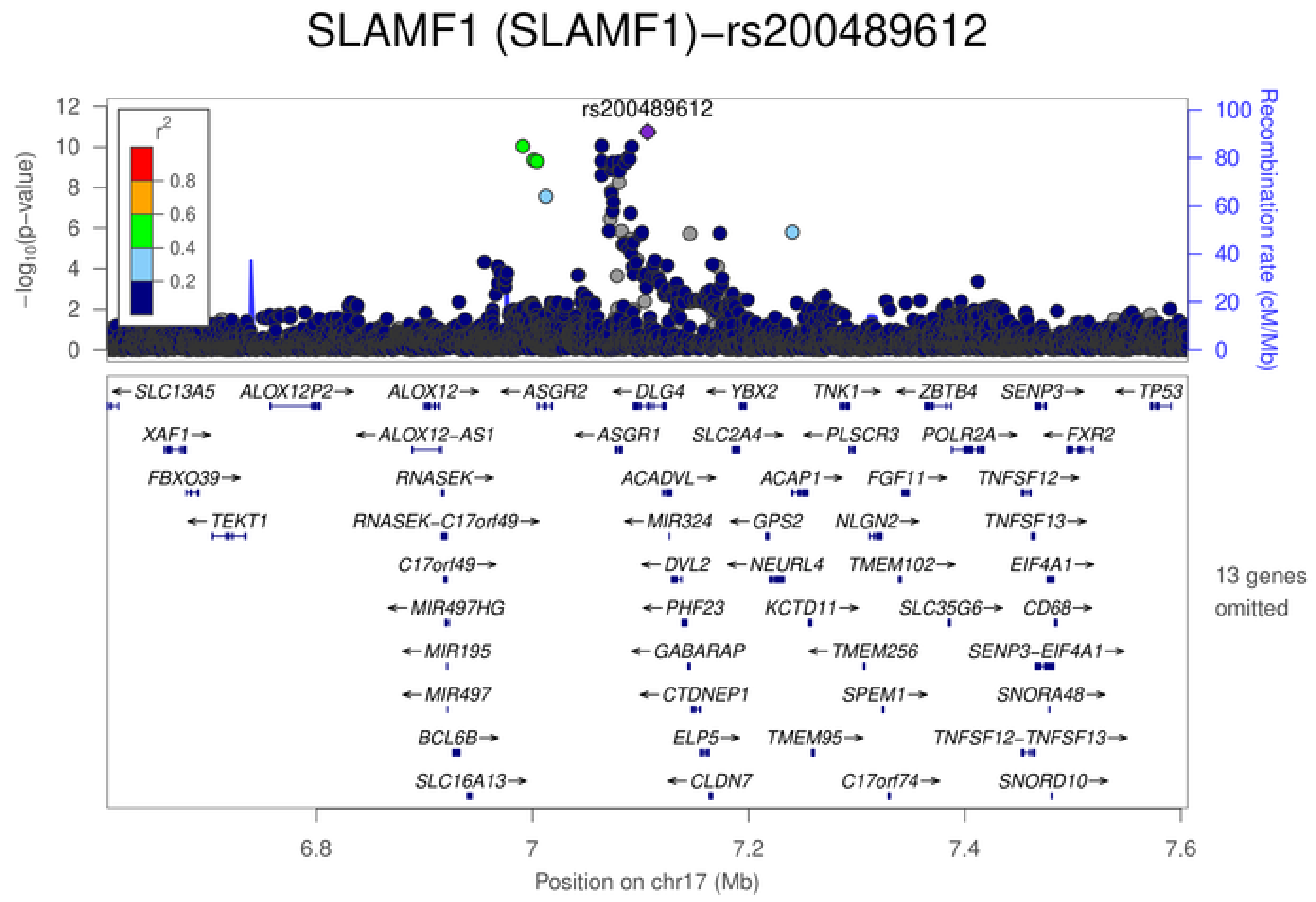
SLAMF1 (SLAMF1)−rs200489612
rs200489612
$-\log_{10}$(p−value)
Recombination rate (cM/Mb)
$r^2$
0.8
0.6
0.4
0.2
SLC13A5
ALOX12P2
ALOX12
ASGR2
DLG4
YBX2
TNK1
ZBTB4
SENP3
TP53
XAF1
ALOX12−AS1
ASGR1
SLC2A4
PLSCR3
POLR2A
FXR2
FBXO39
RNASEK
ACADVL
ACAP1
FGF11
TNFSF12
TEKT1
RNASEK−C17orf49
MIR324
GPS2
NLGN2
TNFSF13
C17orf49
DVL2
NEURL4
TMEM102
EIF4A1
MIR497HG
PHF23
KCTD11
SLC35G6
CD68
MIR195
GABARAP
TMEM256
SENP3−EIF4A1
MIR497
CTDNEP1
SPEM1
SNORA48
BCL6B
ELP5
TMEM95
TNFSF12−TNFSF13
SLC16A13
CLDN7
C17orf74
SNORD10
13 genes omitted
6.8
7
7.2
7.4
7.6
Position on chr17 (Mb)

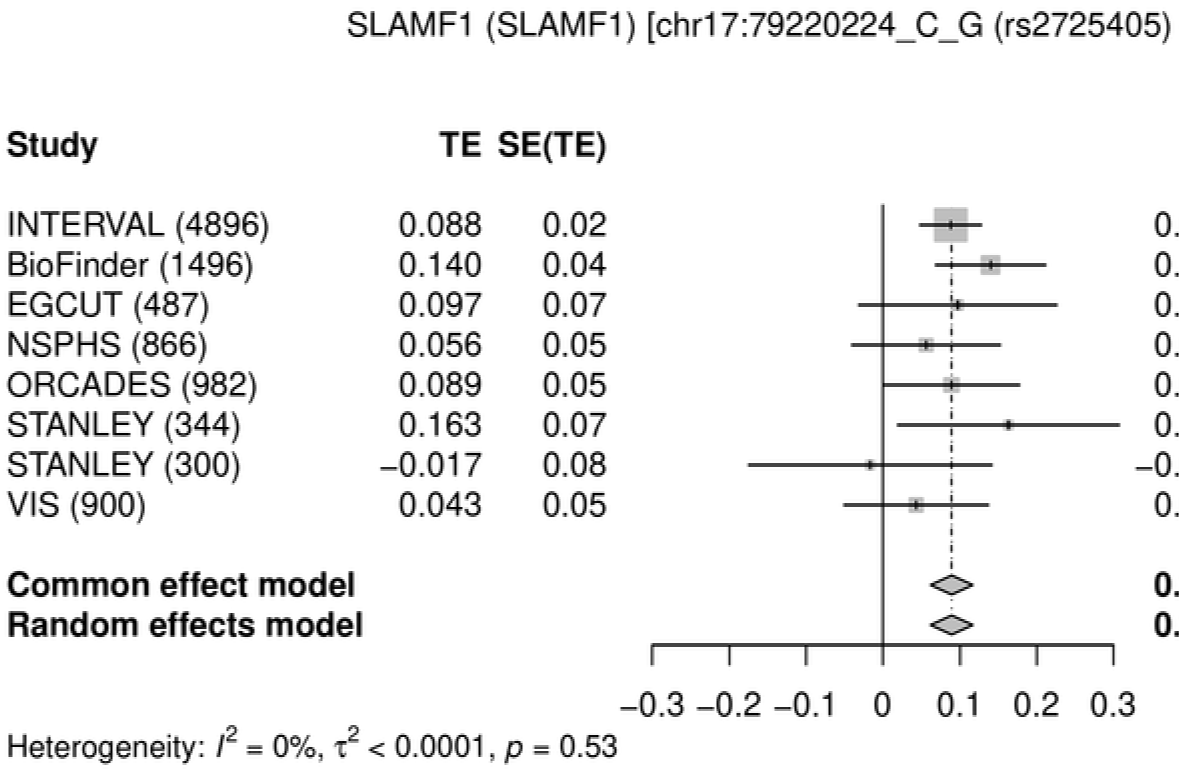
SLAMF1 (SLAMF1) [chr17:79220224_C_G (rs2725405) (C/G) N=10271]
Study
TE
SE(TE)
95%-CI
Weight (common)
Weight (random)
INTERVAL (4896) 0.088 0.02 0.09 [ 0.05; 0.13] 47.5% 47.3%
BioFinder (1496) 0.140 0.04 0.14 [ 0.07; 0.21] 14.8% 14.9%
EGCUT (487) 0.097 0.07 0.10 [−0.03; 0.23] 4.6% 4.6%
NSPHS (866) 0.056 0.05 0.06 [−0.04; 0.15] 8.1% 8.2%
ORCADES (982) 0.089 0.05 0.09 [ 0.00; 0.18] 9.7% 9.7%
STANLEY (344) 0.163 0.07 0.16 [ 0.02; 0.31] 3.7% 3.7%
STANLEY (300) −0.017 0.08 −0.02 [−0.17; 0.14] 3.0% 3.0%
VIS (900) 0.043 0.05 0.04 [−0.05; 0.14] 8.6% 8.6%
Common effect model 0.09 [ 0.06; 0.12] 100.0% --
Random effects model 0.09 [ 0.06; 0.12] -- 100.0%
−0.3 −0.2 −0.1 0 0.1 0.2 0.3
Heterogeneity: $I^2$ = 0%, $\tau^2$ < 0.0001, $p$ = 0.53

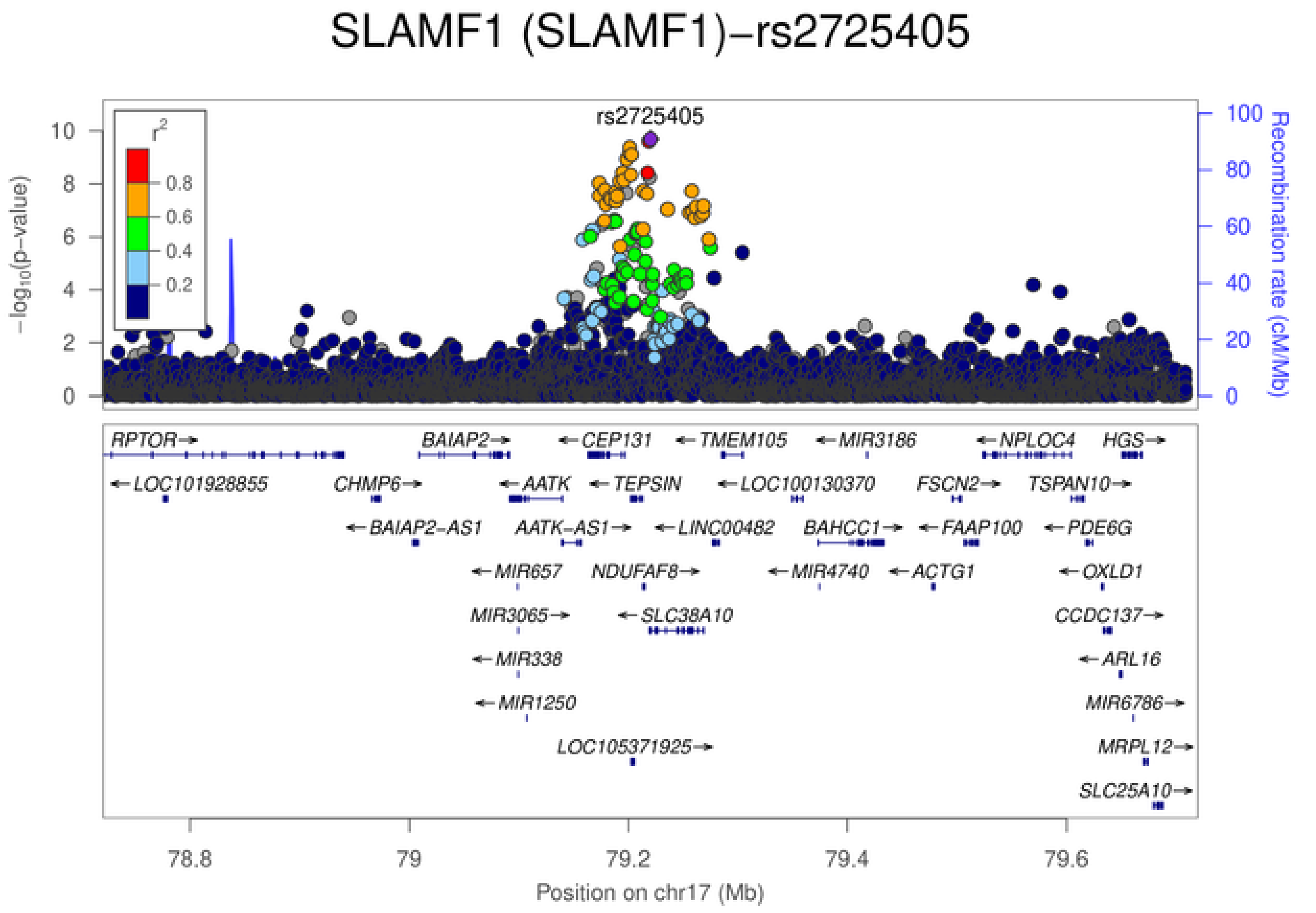
SLAMF1 (SLAMF1)−rs2725405
rs2725405
$r^2$
0.8
0.6
0.4
0.2
$-\log_{10}$(p−value)
Recombination rate (cM/Mb)
RPTOR→
BAIAP2→
←CEP131
←TMEM105
←MIR3186
←NPLOC4
HGS→
←LOC101928855
CHMP6→
←AATK
←TEPSIN
←LOC100130370
FSCN2→
TSPAN10→
←BAIAP2-AS1
AATK-AS1→
←LINC00482
BAHCC1→
←FAAP100
←PDE6G
←MIR657
NDUFAF8→
←MIR4740
←ACTG1
←OXLD1
MIR3065→
←SLC38A10
CCDC137→
←MIR338
←ARL16
←MIR1250
MIR6786→
LOC105371925→
MRPL12→
SLC25A10→
78.8
79
79.2
79.4
79.6
Position on chr17 (Mb)

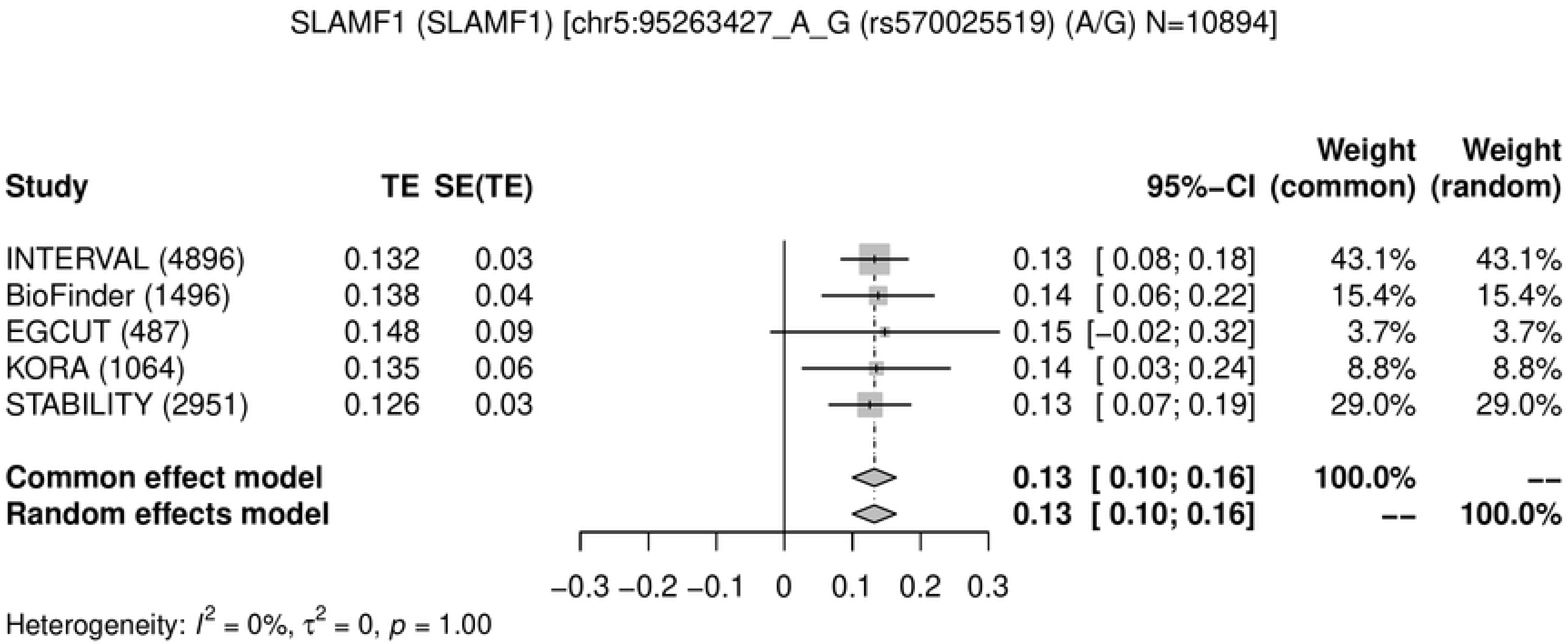
SLAMF1 (SLAMF1) [chr5:95263427_A_G (rs570025519) (A/G) N=10894]
Study
TE
SE(TE)
95%-CI
Weight (common)
Weight (random)
INTERVAL (4896) 0.132 0.03 0.13 [ 0.08; 0.18] 43.1% 43.1%
BioFinder (1496) 0.138 0.04 0.14 [ 0.06; 0.22] 15.4% 15.4%
EGCUT (487) 0.148 0.09 0.15 [−0.02; 0.32] 3.7% 3.7%
KORA (1064) 0.135 0.06 0.14 [ 0.03; 0.24] 8.8% 8.8%
STABILITY (2951) 0.126 0.03 0.13 [ 0.07; 0.19] 29.0% 29.0%
Common effect model 0.13 [ 0.10; 0.16] 100.0% --
Random effects model 0.13 [ 0.10; 0.16] -- 100.0%
−0.3 −0.2 −0.1 0 0.1 0.2 0.3
Heterogeneity: $I^2$ = 0%, $\tau^2$ = 0, $p$ = 1.00

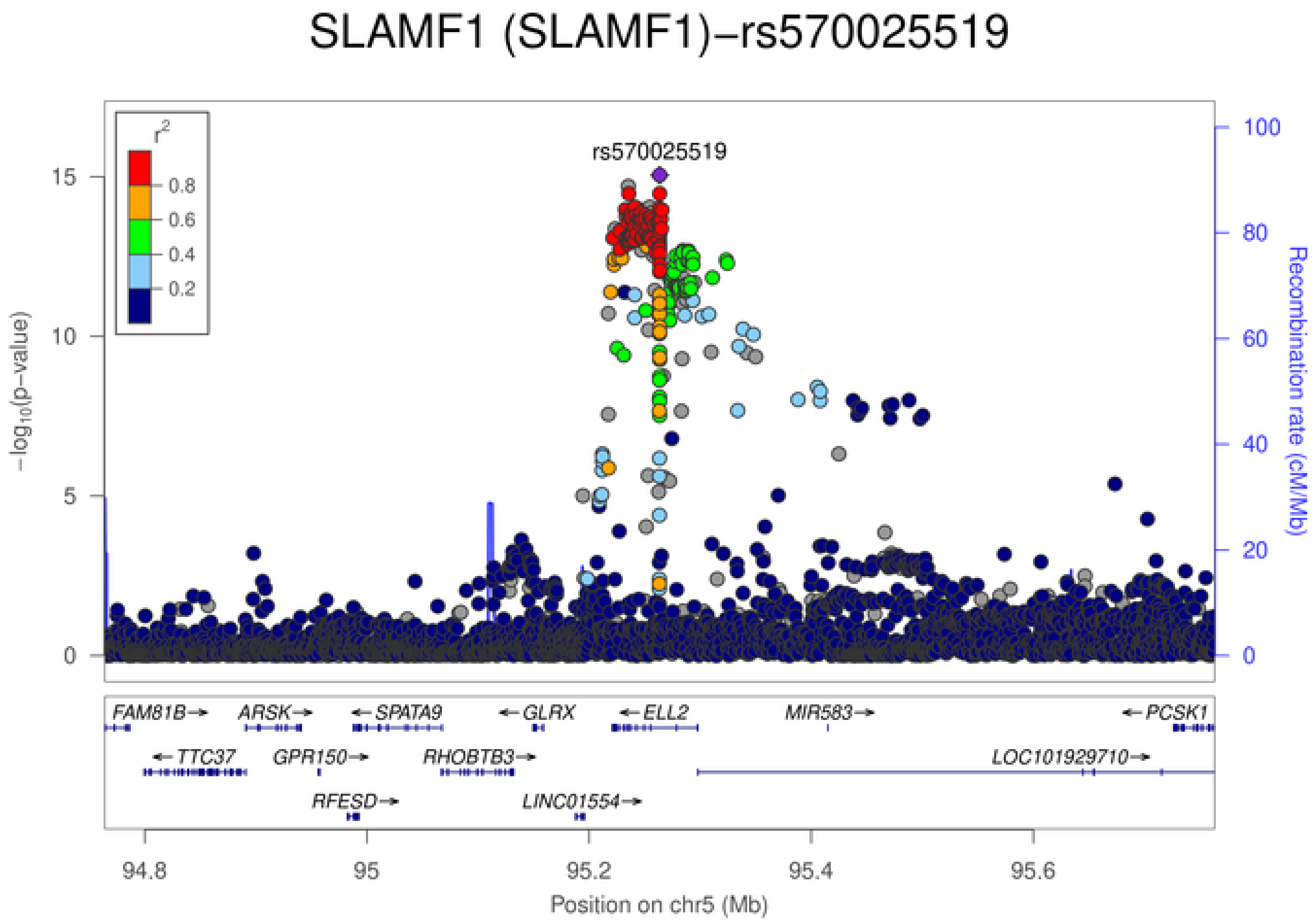
SLAMF1 (SLAMF1)−rs570025519
rs570025519
$r^2$
0.8
0.6
0.4
0.2
$-\log_{10}$(p−value)
0
5
10
15
Recombination rate (cM/Mb)
0
20
40
60
80
100
FAM81B→
ARSK→
←SPATA9
←GLRX
←ELL2
MIR583→
←PCSK1
←TTC37
GPR150→
RHOBTB3→
LOC101929710→
RFESD→
LINC01554→
94.8
95
95.2
95.4
95.6
Position on chr5 (Mb)

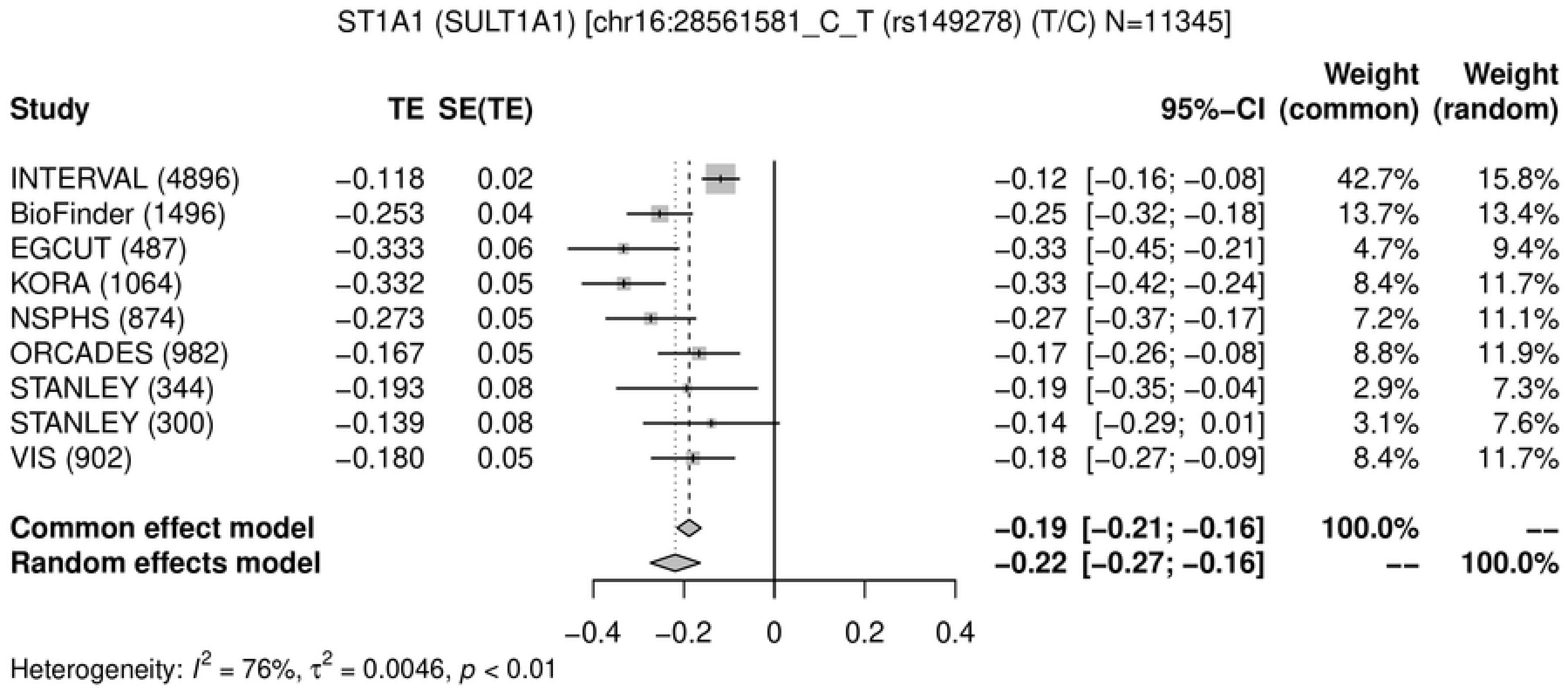
ST1A1 (SULT1A1) [chr16:28561581_C_T (rs149278) (T/C) N=11345]
Study
TE
SE(TE)
95%−CI
Weight (common)
Weight (random)
INTERVAL (4896) −0.118 0.02 −0.12 [−0.16; −0.08] 42.7% 15.8%
BioFinder (1496) −0.253 0.04 −0.25 [−0.32; −0.18] 13.7% 13.4%
EGCUT (487) −0.333 0.06 −0.33 [−0.45; −0.21] 4.7% 9.4%
KORA (1064) −0.332 0.05 −0.33 [−0.42; −0.24] 8.4% 11.7%
NSPHS (874) −0.273 0.05 −0.27 [−0.37; −0.17] 7.2% 11.1%
ORCADES (982) −0.167 0.05 −0.17 [−0.26; −0.08] 8.8% 11.9%
STANLEY (344) −0.193 0.08 −0.19 [−0.35; −0.04] 2.9% 7.3%
STANLEY (300) −0.139 0.08 −0.14 [−0.29; 0.01] 3.1% 7.6%
VIS (902) −0.180 0.05 −0.18 [−0.27; −0.09] 8.4% 11.7%
Common effect model −0.19 [−0.21; −0.16] 100.0% --
Random effects model −0.22 [−0.27; −0.16] -- 100.0%
−0.4 −0.2 0 0.2 0.4
Heterogeneity: $I^2$ = 76%, $\tau^2$ = 0.0046, $p$ < 0.01

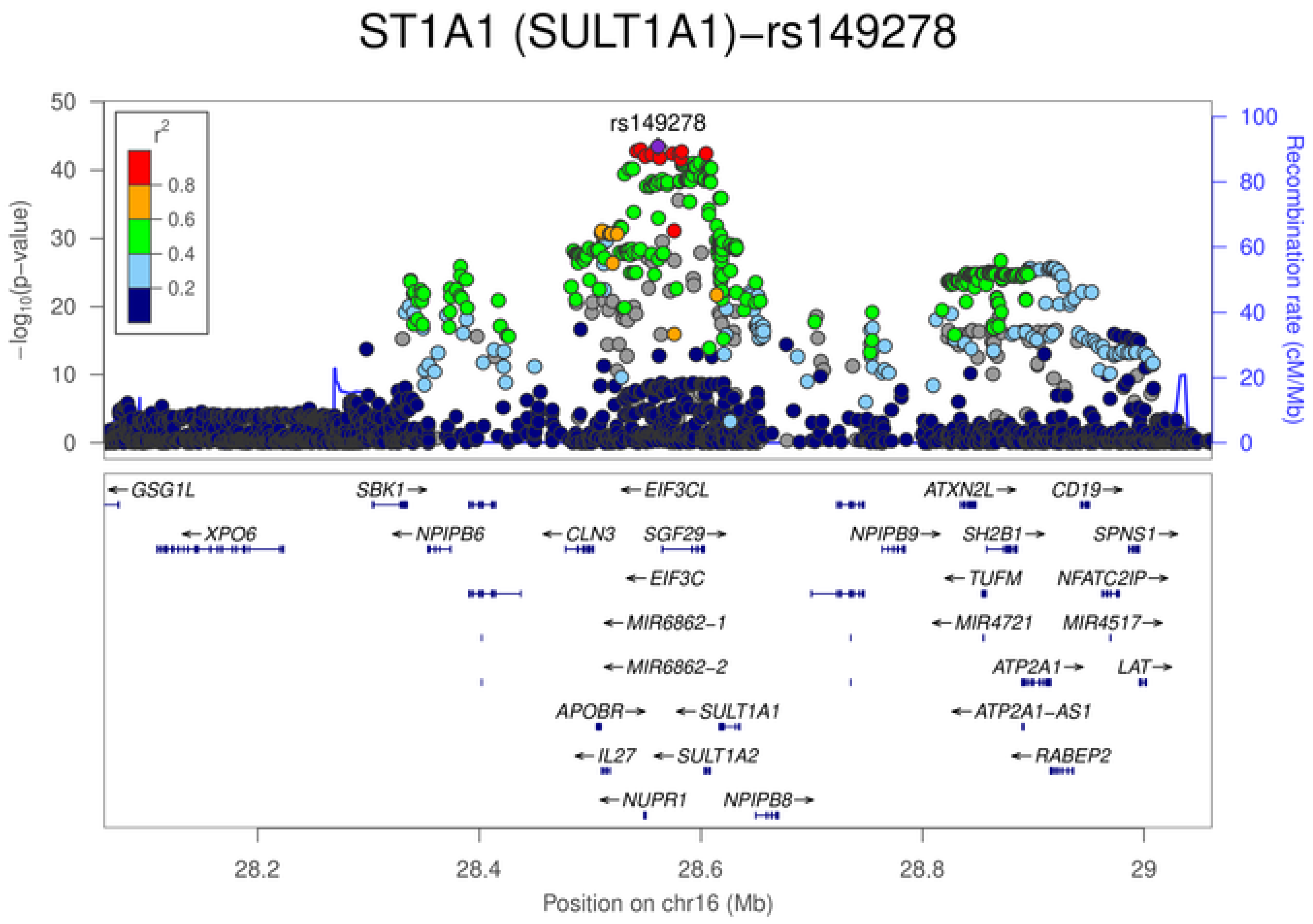
ST1A1 (SULT1A1)−rs149278
rs149278
$r^2$
0.8
0.6
0.4
0.2
$-\log_{10}$(p−value)
Recombination rate (cM/Mb)
GSG1L
SBK1
EIF3CL
ATXN2L
CD19
XPO6
NPIPB6
CLN3
SGF29
NPIPB9
SH2B1
SPNS1
EIF3C
TUFM
NFATC2IP
MIR6862-1
MIR4721
MIR4517
MIR6862-2
ATP2A1
LAT
APOBR
SULT1A1
ATP2A1-AS1
IL27
SULT1A2
RABEP2
NUPR1
NPIPB8
28.2
28.4
28.6
28.8
29
Position on chr16 (Mb)

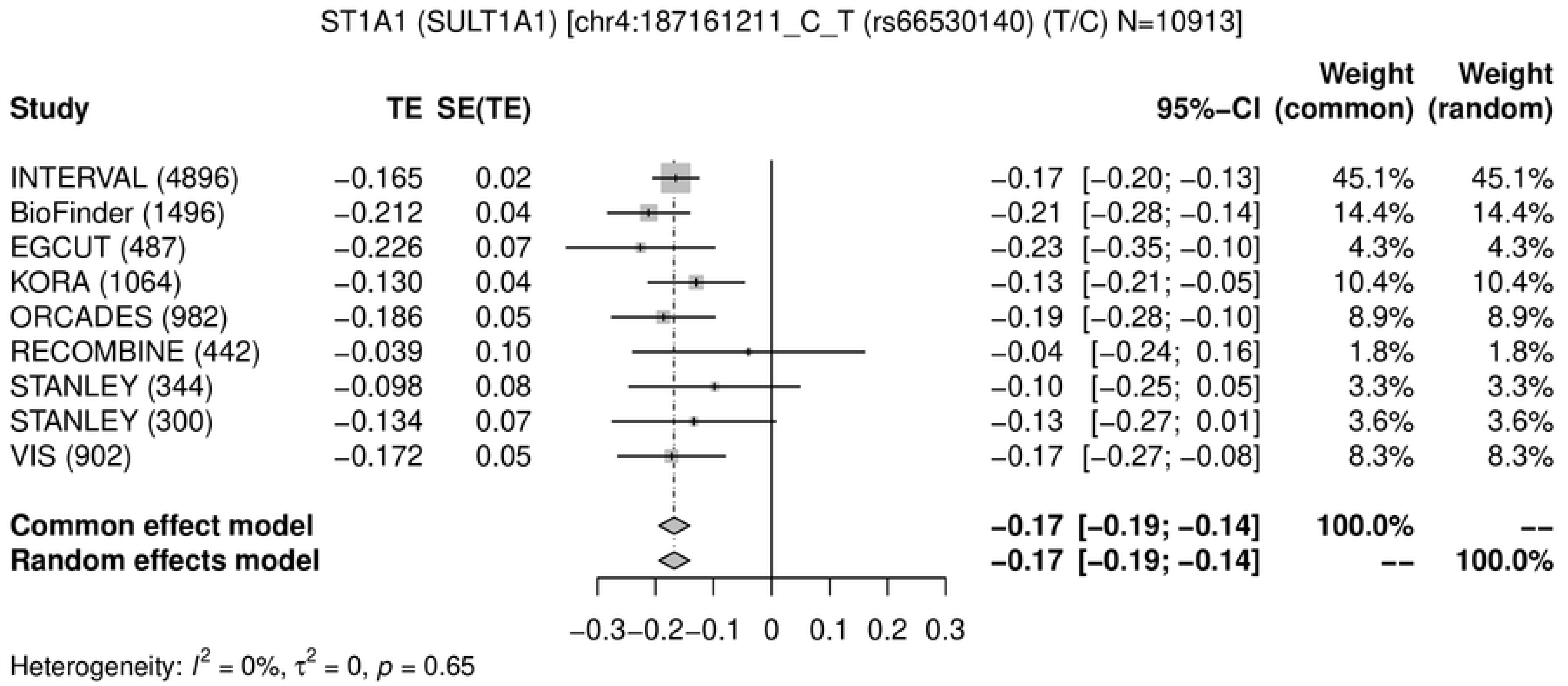
ST1A1 (SULT1A1) [chr4:187161211_C_T (rs66530140) (T/C) N=10913]
Study
TE
SE(TE)
95%−CI
Weight (common)
Weight (random)
INTERVAL (4896) −0.165 0.02 −0.17 [−0.20; −0.13] 45.1% 45.1%
BioFinder (1496) −0.212 0.04 −0.21 [−0.28; −0.14] 14.4% 14.4%
EGCUT (487) −0.226 0.07 −0.23 [−0.35; −0.10] 4.3% 4.3%
KORA (1064) −0.130 0.04 −0.13 [−0.21; −0.05] 10.4% 10.4%
ORCADES (982) −0.186 0.05 −0.19 [−0.28; −0.10] 8.9% 8.9%
RECOMBINE (442) −0.039 0.10 −0.04 [−0.24; 0.16] 1.8% 1.8%
STANLEY (344) −0.098 0.08 −0.10 [−0.25; 0.05] 3.3% 3.3%
STANLEY (300) −0.134 0.07 −0.13 [−0.27; 0.01] 3.6% 3.6%
VIS (902) −0.172 0.05 −0.17 [−0.27; −0.08] 8.3% 8.3%
Common effect model −0.17 [−0.19; −0.14] 100.0% --
Random effects model −0.17 [−0.19; −0.14] -- 100.0%
−0.3 −0.2 −0.1 0 0.1 0.2 0.3
Heterogeneity: $I^2$ = 0%, $\tau^2$ = 0, $p$ = 0.65

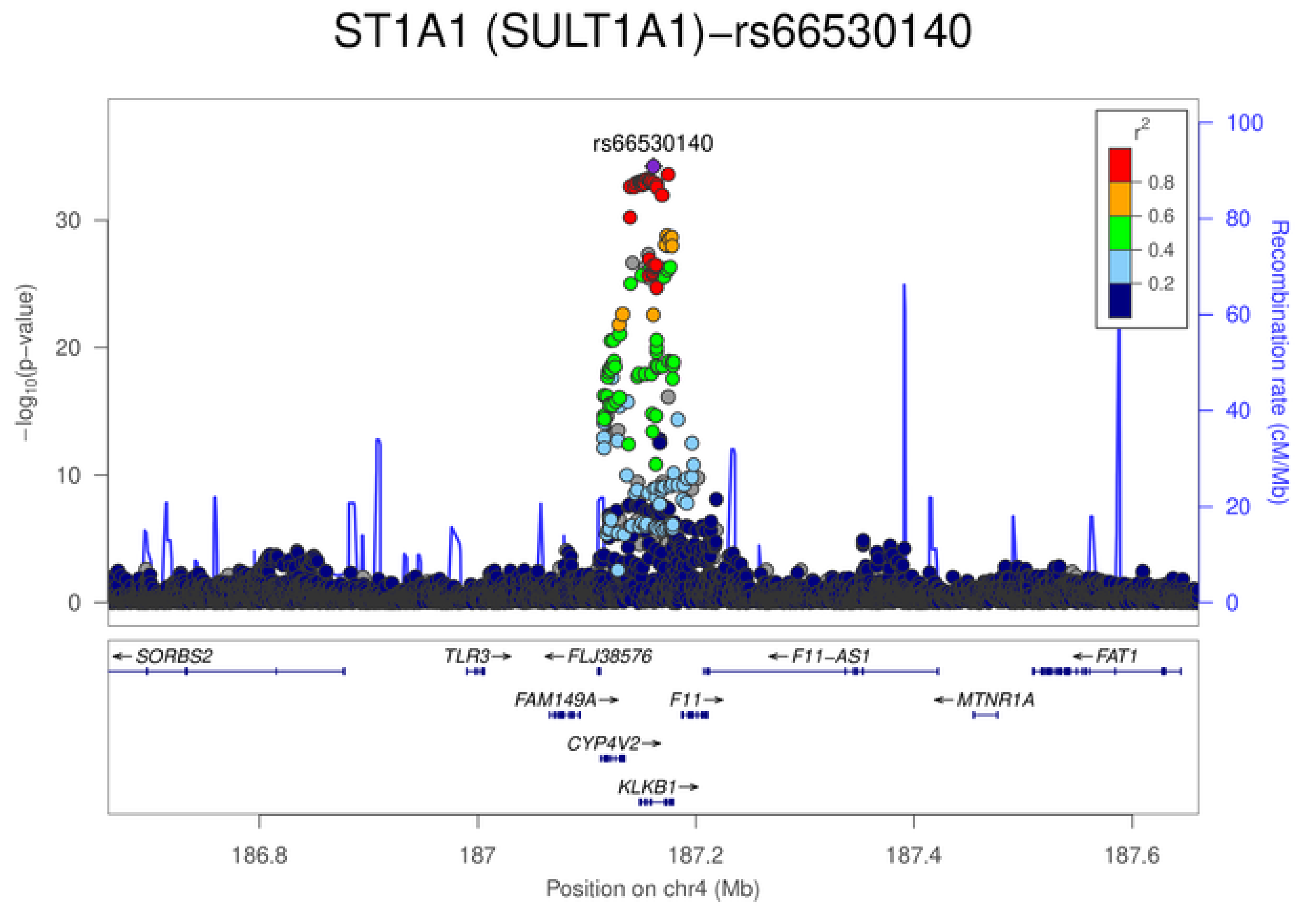
ST1A1 (SULT1A1)−rs66530140
rs66530140
$-\log_{10}$(p−value)
Recombination rate (cM/Mb)
$r^2$
0.8
0.6
0.4
0.2
SORBS2
TLR3
FLJ38576
F11−AS1
FAT1
FAM149A
F11
MTNR1A
CYP4V2
KLKB1
186.8
187
187.2
187.4
187.6
Position on chr4 (Mb)

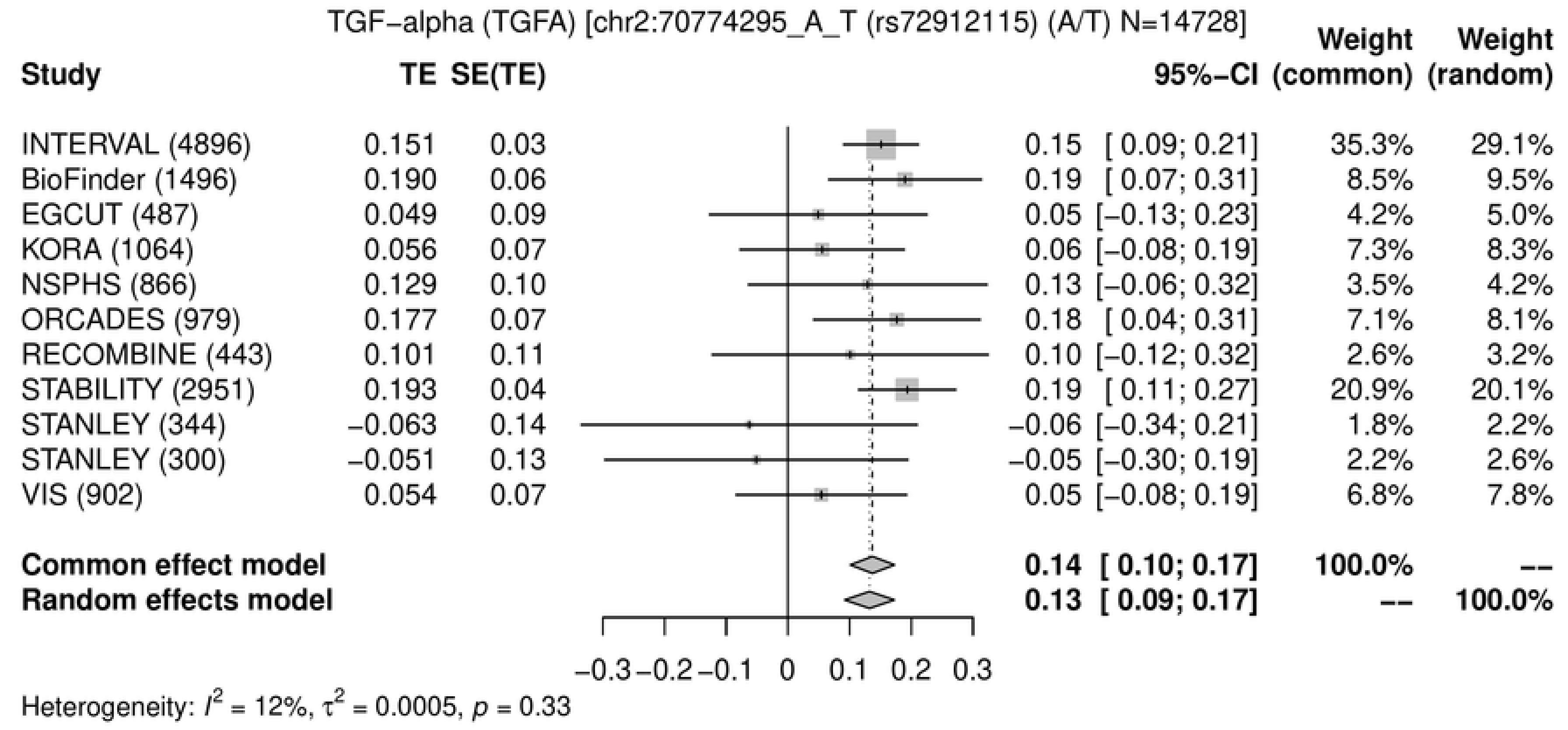
TGF−alpha (TGFA) [chr2:70774295_A_T (rs72912115) (A/T) N=14728]
Study
TE
SE(TE)
95%−CI
Weight (common)
Weight (random)
INTERVAL (4896) 0.151 0.03 0.15 [ 0.09; 0.21] 35.3% 29.1%
BioFinder (1496) 0.190 0.06 0.19 [ 0.07; 0.31] 8.5% 9.5%
EGCUT (487) 0.049 0.09 0.05 [−0.13; 0.23] 4.2% 5.0%
KORA (1064) 0.056 0.07 0.06 [−0.08; 0.19] 7.3% 8.3%
NSPHS (866) 0.129 0.10 0.13 [−0.06; 0.32] 3.5% 4.2%
ORCADES (979) 0.177 0.07 0.18 [ 0.04; 0.31] 7.1% 8.1%
RECOMBINE (443) 0.101 0.11 0.10 [−0.12; 0.32] 2.6% 3.2%
STABILITY (2951) 0.193 0.04 0.19 [ 0.11; 0.27] 20.9% 20.1%
STANLEY (344) −0.063 0.14 −0.06 [−0.34; 0.21] 1.8% 2.2%
STANLEY (300) −0.051 0.13 −0.05 [−0.30; 0.19] 2.2% 2.6%
VIS (902) 0.054 0.07 0.05 [−0.08; 0.19] 6.8% 7.8%
Common effect model 0.14 [ 0.10; 0.17] 100.0% --
Random effects model 0.13 [ 0.09; 0.17] -- 100.0%
−0.3 −0.2 −0.1 0 0.1 0.2 0.3
Heterogeneity: $I^2$ = 12%, $\tau^2$ = 0.0005, $p$ = 0.33

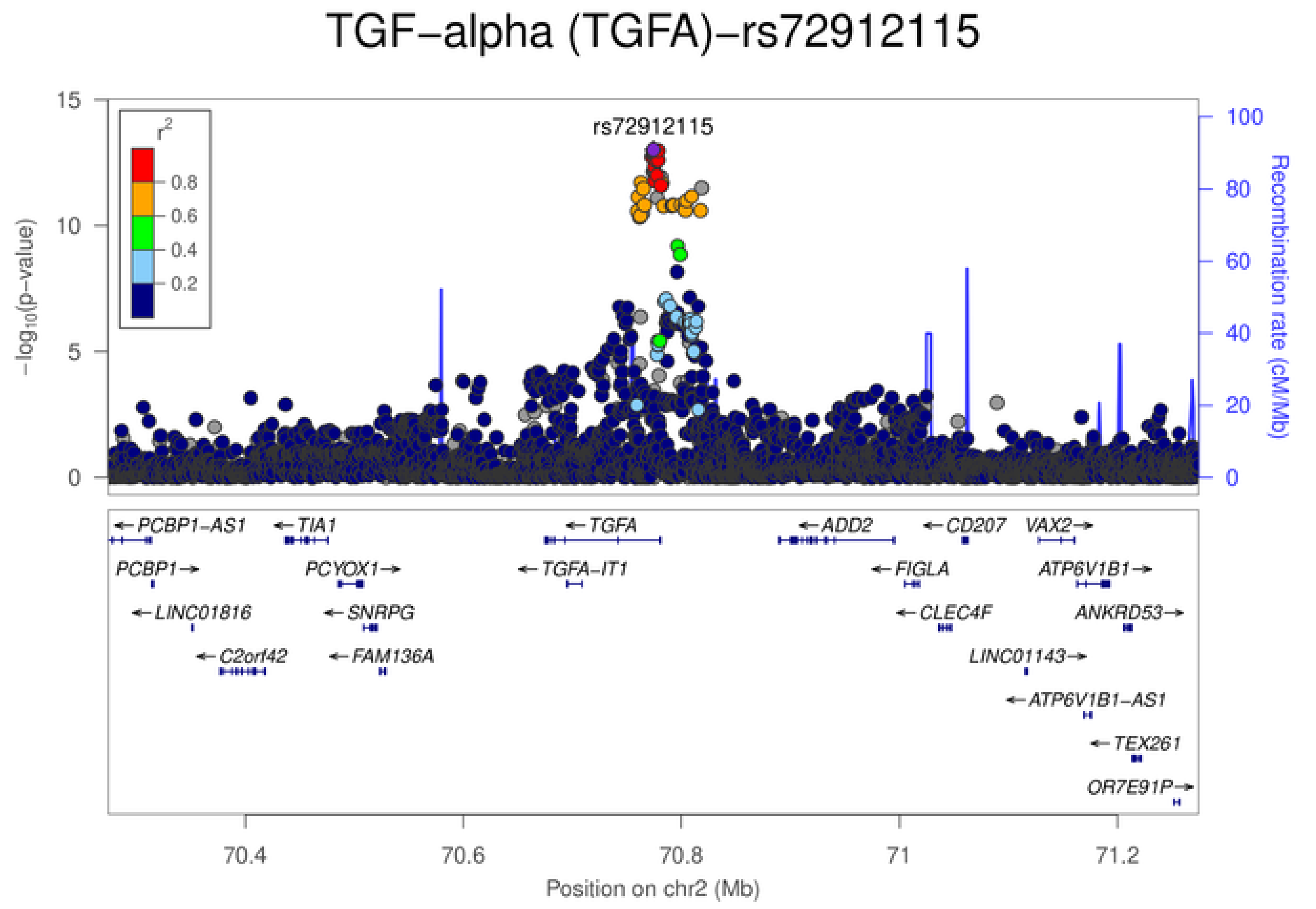
TGF−alpha (TGFA)−rs72912115
rs72912115
$r^2$
0.8
0.6
0.4
0.2
$-\log_{10}$(p−value)
Recombination rate (cM/Mb)
PCBP1−AS1
TIA1
TGFA
ADD2
CD207
VAX2
PCBP1
PCYOX1
TGFA−IT1
FIGLA
ATP6V1B1
LINC01816
SNRPG
CLEC4F
ANKRD53
C2orf42
FAM136A
LINC01143
ATP6V1B1−AS1
TEX261
OR7E91P
70.4
70.6
70.8
71
71.2
Position on chr2 (Mb)

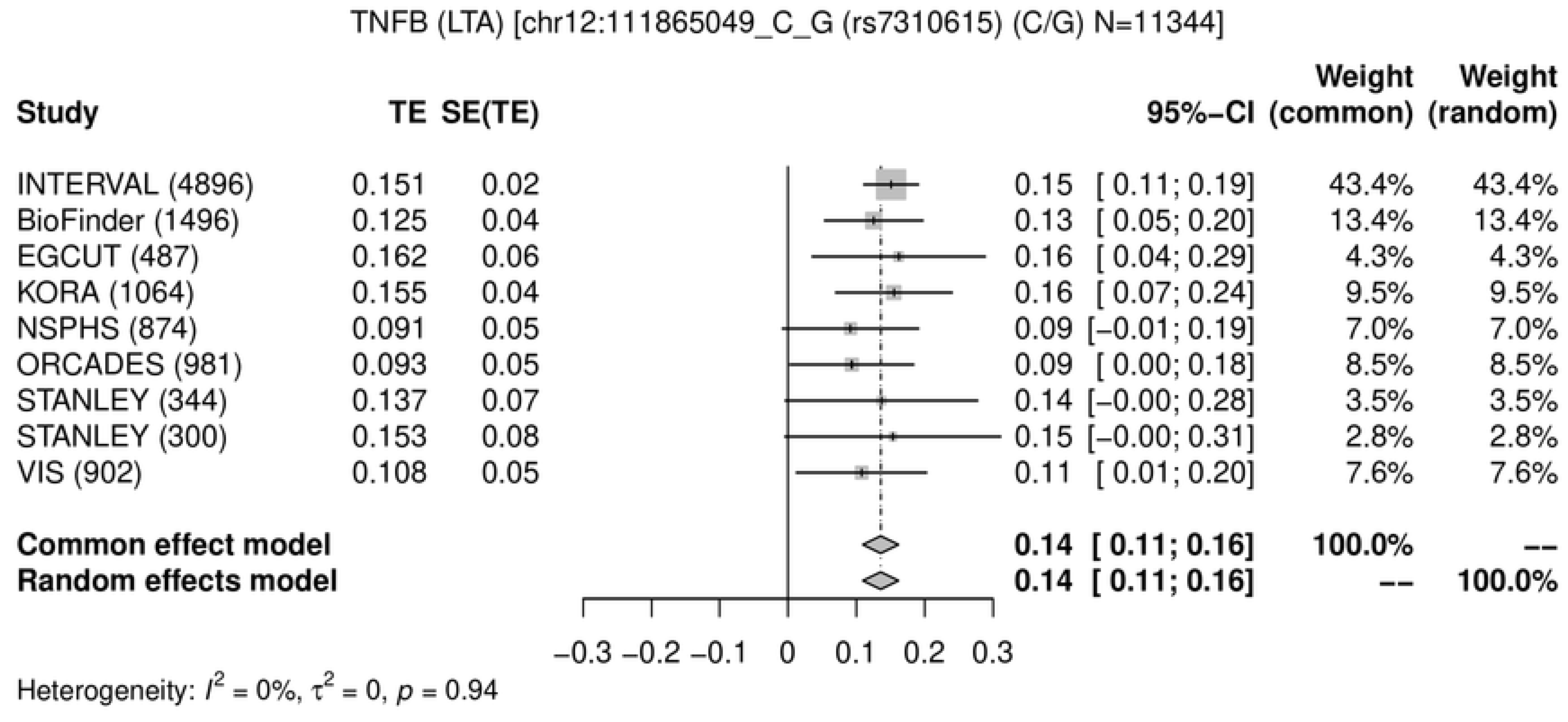
TNFB (LTA) [chr12:111865049_C_G (rs7310615) (C/G) N=11344]
Study TE SE(TE) 95%-CI Weight (common) Weight (random)
INTERVAL (4896) 0.151 0.02 0.15 [ 0.11; 0.19] 43.4% 43.4%
BioFinder (1496) 0.125 0.04 0.13 [ 0.05; 0.20] 13.4% 13.4%
EGCUT (487) 0.162 0.06 0.16 [ 0.04; 0.29] 4.3% 4.3%
KORA (1064) 0.155 0.04 0.16 [ 0.07; 0.24] 9.5% 9.5%
NSPHS (874) 0.091 0.05 0.09 [−0.01; 0.19] 7.0% 7.0%
ORCADES (981) 0.093 0.05 0.09 [ 0.00; 0.18] 8.5% 8.5%
STANLEY (344) 0.137 0.07 0.14 [−0.00; 0.28] 3.5% 3.5%
STANLEY (300) 0.153 0.08 0.15 [−0.00; 0.31] 2.8% 2.8%
VIS (902) 0.108 0.05 0.11 [ 0.01; 0.20] 7.6% 7.6%
Common effect model 0.14 [ 0.11; 0.16] 100.0% --
Random effects model 0.14 [ 0.11; 0.16] -- 100.0%
−0.3 −0.2 −0.1 0 0.1 0.2 0.3
Heterogeneity: $I^2 = 0\%$, $\tau^2 = 0$, $p = 0.94$

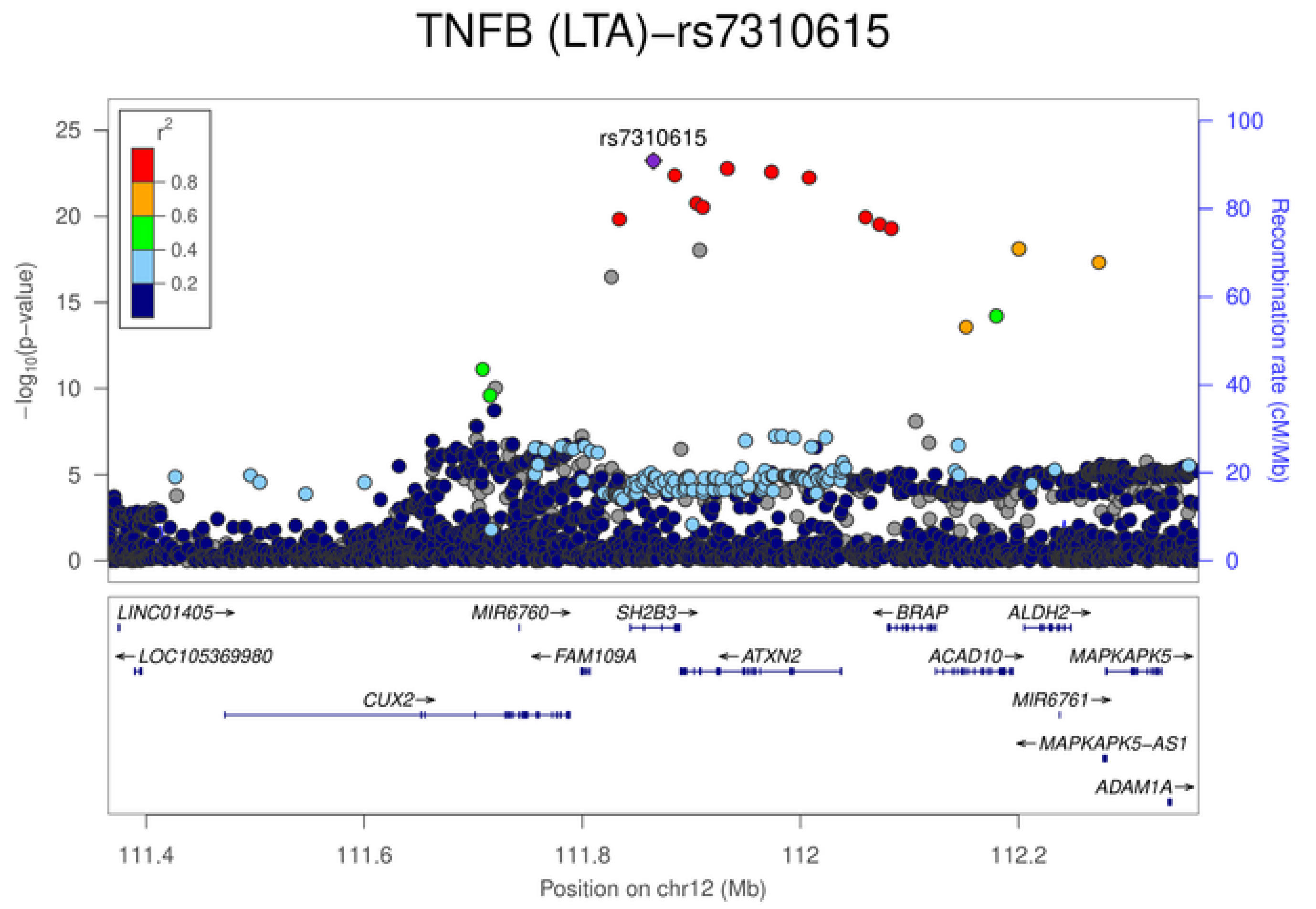
TNFB (LTA)−rs7310615
rs7310615
$r^2$
0.8
0.6
0.4
0.2
$-\log_{10}$(p-value)
Recombination rate (cM/Mb)
LINC01405→
←LOC105369980
CUX2→
MIR6760→
←FAM109A
SH2B3→
←ATXN2
←BRAP
ACAD10→
ALDH2→
MAPKAPK5→
MIR6761→
←MAPKAPK5-AS1
ADAM1A→
111.4 111.6 111.8 112 112.2
Position on chr12 (Mb)

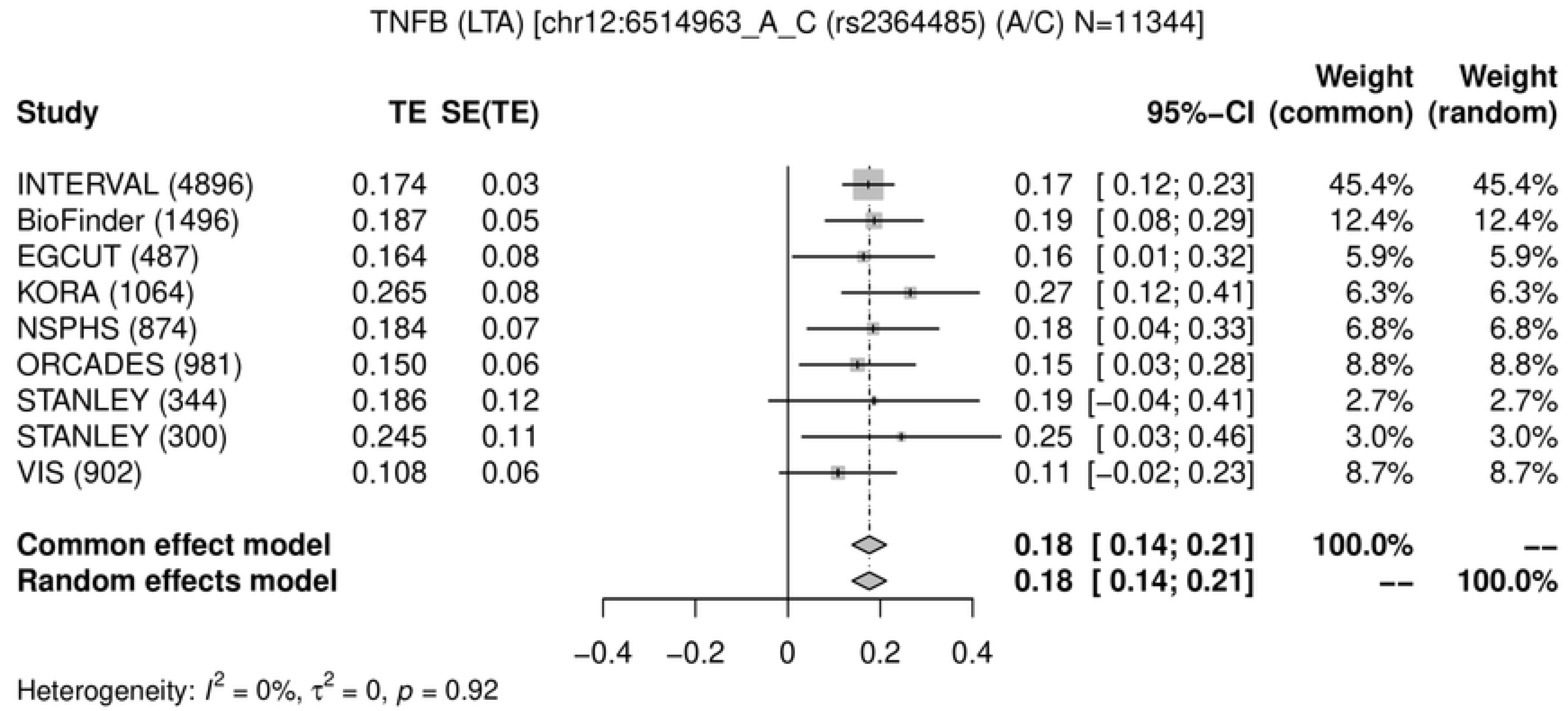
TNFB (LTA) [chr12:6514963_A_C (rs2364485) (A/C) N=11344]
Study
TE
SE(TE)
95%-CI
Weight (common)
Weight (random)
INTERVAL (4896) 0.174 0.03 0.17 [ 0.12; 0.23] 45.4% 45.4%
BioFinder (1496) 0.187 0.05 0.19 [ 0.08; 0.29] 12.4% 12.4%
EGCUT (487) 0.164 0.08 0.16 [ 0.01; 0.32] 5.9% 5.9%
KORA (1064) 0.265 0.08 0.27 [ 0.12; 0.41] 6.3% 6.3%
NSPHS (874) 0.184 0.07 0.18 [ 0.04; 0.33] 6.8% 6.8%
ORCADES (981) 0.150 0.06 0.15 [ 0.03; 0.28] 8.8% 8.8%
STANLEY (344) 0.186 0.12 0.19 [−0.04; 0.41] 2.7% 2.7%
STANLEY (300) 0.245 0.11 0.25 [ 0.03; 0.46] 3.0% 3.0%
VIS (902) 0.108 0.06 0.11 [−0.02; 0.23] 8.7% 8.7%
Common effect model 0.18 [ 0.14; 0.21] 100.0% --
Random effects model 0.18 [ 0.14; 0.21] -- 100.0%
−0.4 −0.2 0 0.2 0.4
Heterogeneity: $I^2$ = 0%, $\tau^2$ = 0, $p$ = 0.92

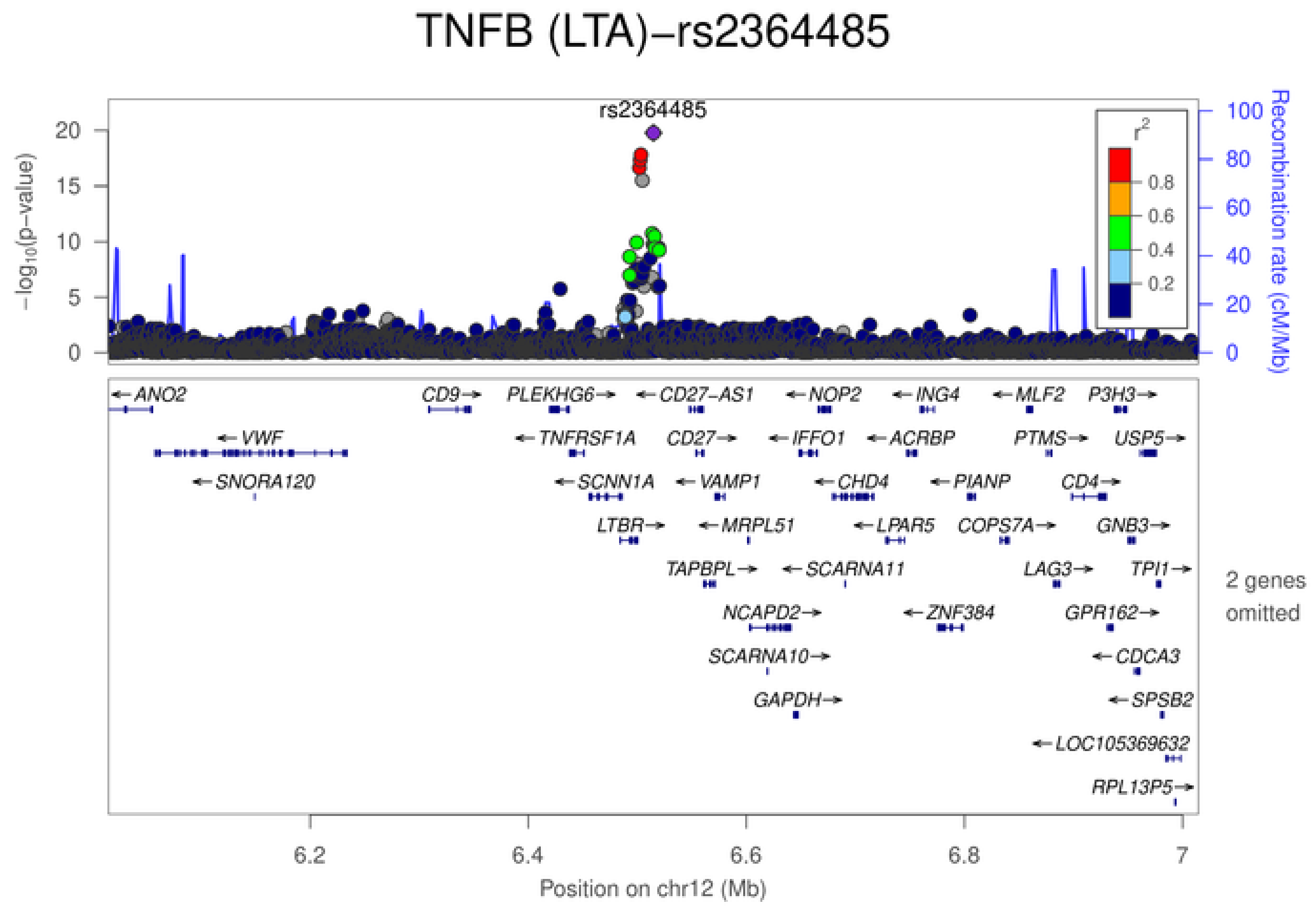
TNFB (LTA)−rs2364485
rs2364485
$-\log_{10}$(p−value)
Recombination rate (cM/Mb)
$r^2$
0.8
0.6
0.4
0.2
ANO2
VWF
SNORA120
CD9
PLEKHG6
TNFRSF1A
SCNN1A
LTBR
CD27−AS1
CD27
VAMP1
MRPL51
TAPBPL
NCAPD2
SCARNA10
GAPDH
NOP2
IFFO1
CHD4
SCARNA11
LPAR5
ZNF384
ING4
ACRBP
PIANP
COPS7A
MLF2
PTMS
CD4
LAG3
GPR162
P3H3
USP5
GNB3
TPI1
CDCA3
SPSB2
LOC105369632
RPL13P5
2 genes omitted
6.2
6.4
6.6
6.8
7
Position on chr12 (Mb)

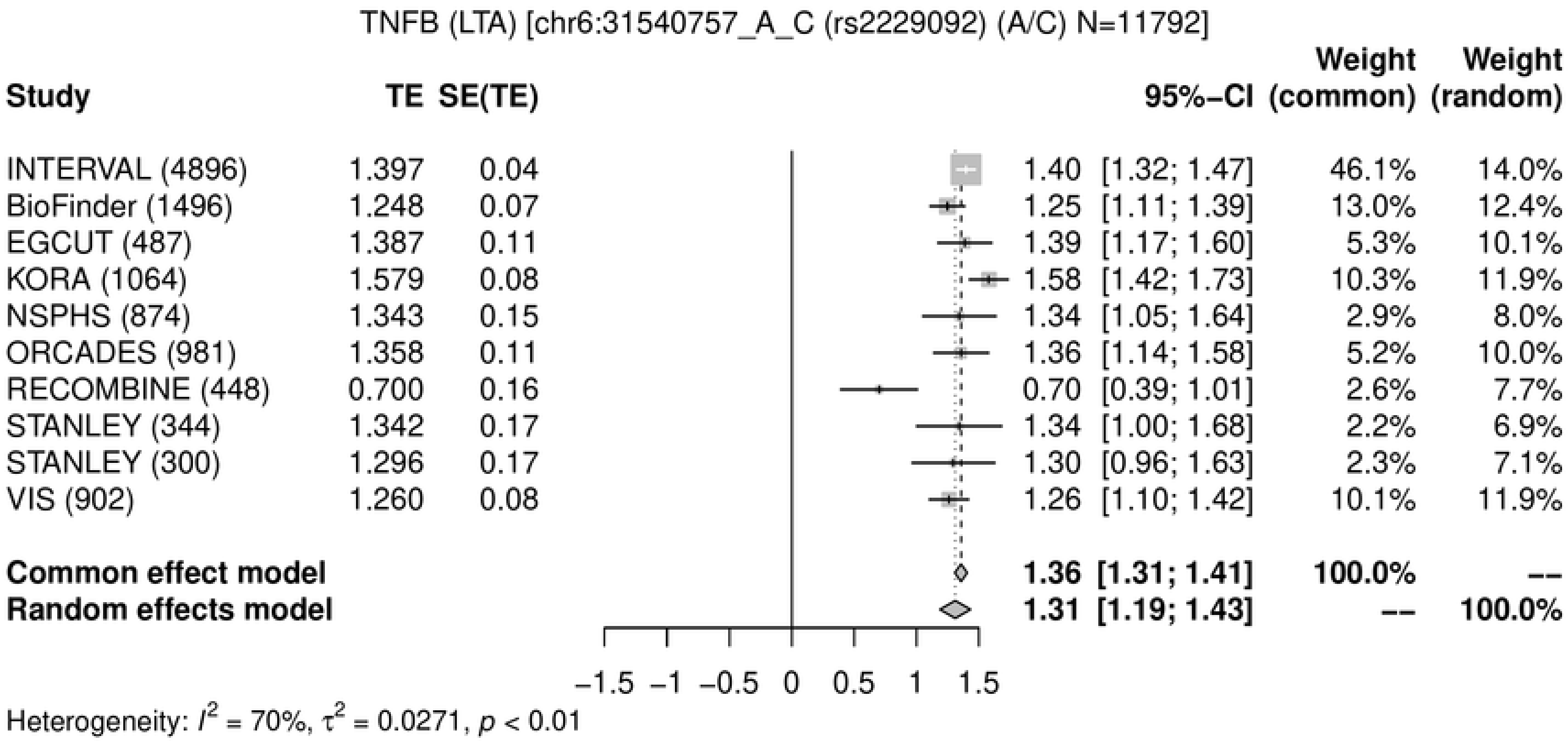
TNFB (LTA) [chr6:31540757_A_C (rs2229092) (A/C) N=11792]
Study | TE | SE(TE) | 95%-CI | Weight (common) | Weight (random)
INTERVAL (4896) 1.397 0.04 1.40 [1.32; 1.47] 46.1% 14.0%
BioFinder (1496) 1.248 0.07 1.25 [1.11; 1.39] 13.0% 12.4%
EGCUT (487) 1.387 0.11 1.39 [1.17; 1.60] 5.3% 10.1%
KORA (1064) 1.579 0.08 1.58 [1.42; 1.73] 10.3% 11.9%
NSPHS (874) 1.343 0.15 1.34 [1.05; 1.64] 2.9% 8.0%
ORCADES (981) 1.358 0.11 1.36 [1.14; 1.58] 5.2% 10.0%
RECOMBINE (448) 0.700 0.16 0.70 [0.39; 1.01] 2.6% 7.7%
STANLEY (344) 1.342 0.17 1.34 [1.00; 1.68] 2.2% 6.9%
STANLEY (300) 1.296 0.17 1.30 [0.96; 1.63] 2.3% 7.1%
VIS (902) 1.260 0.08 1.26 [1.10; 1.42] 10.1% 11.9%
Common effect model 1.36 [1.31; 1.41] 100.0% --
Random effects model 1.31 [1.19; 1.43] -- 100.0%
−1.5 −1 −0.5 0 0.5 1 1.5
Heterogeneity: $I^2 = 70\%$, $\tau^2 = 0.0271$, $p < 0.01$

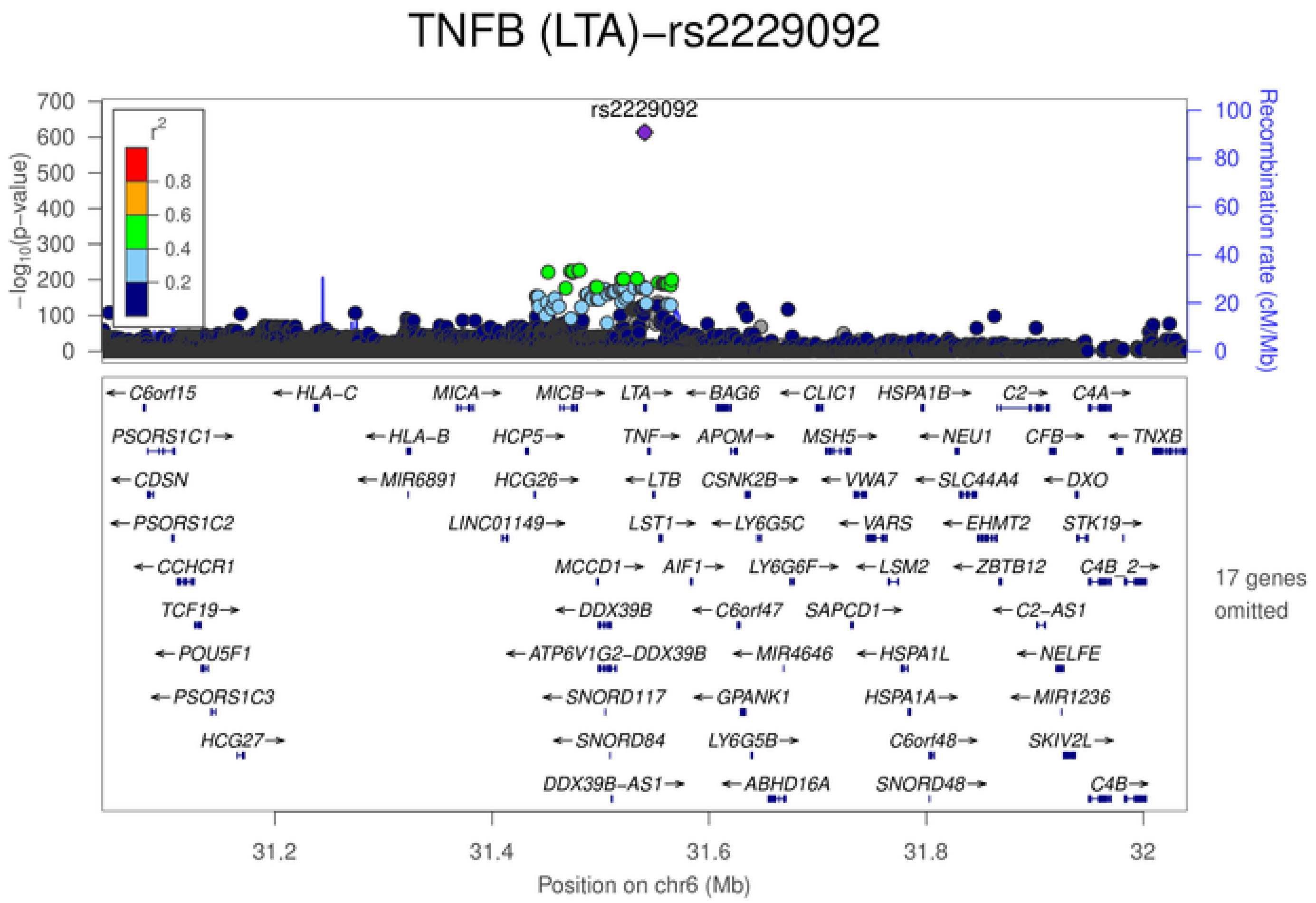
TNFB (LTA)−rs2229092
rs2229092
$-\log_{10}(p\text{-value})$
Recombination rate (cM/Mb)
$r^2$
0.8
0.6
0.4
0.2
17 genes omitted
31.2
31.4
31.6
31.8
32
Position on chr6 (Mb)

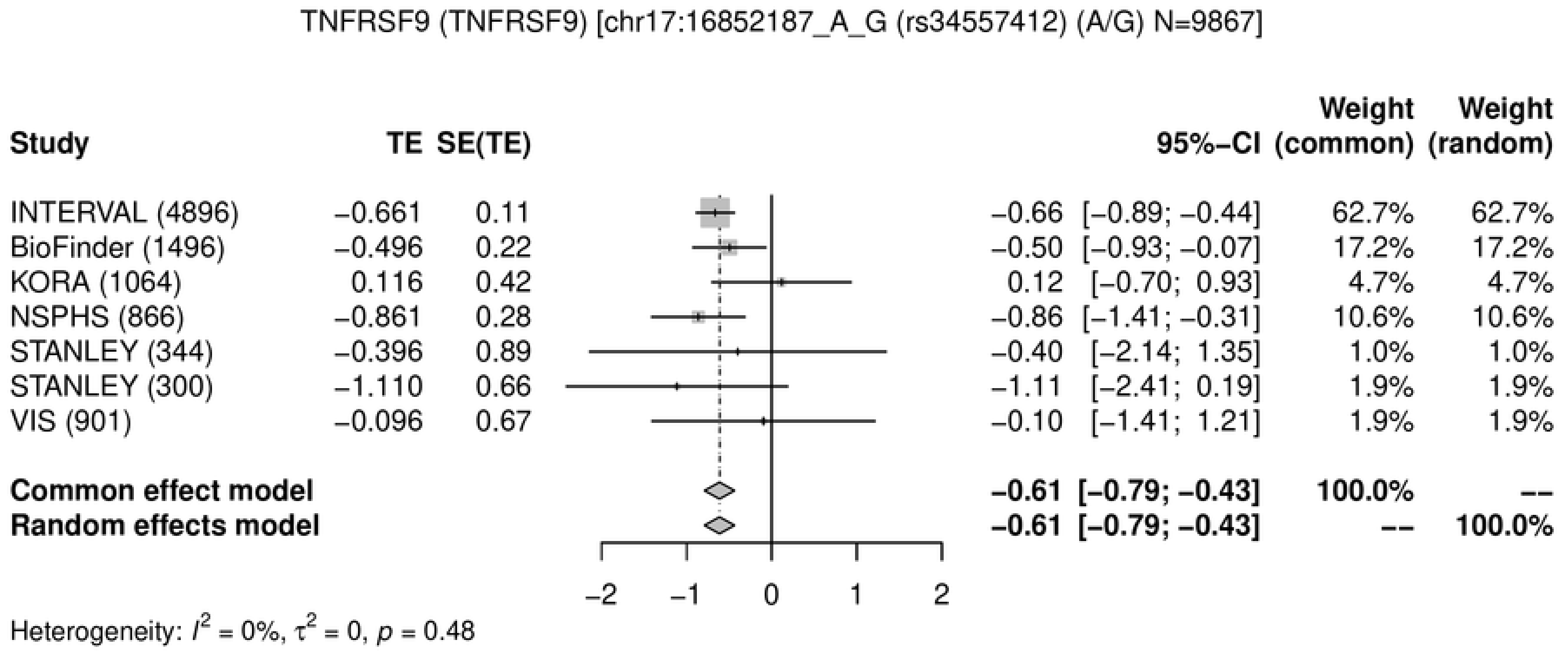
TNFRSF9 (TNFRSF9) [chr17:16852187_A_G (rs34557412) (A/G) N=9867]
Study
TE
SE(TE)
95%−CI
Weight (common)
Weight (random)
INTERVAL (4896) −0.661 0.11 −0.66 [−0.89; −0.44] 62.7% 62.7%
BioFinder (1496) −0.496 0.22 −0.50 [−0.93; −0.07] 17.2% 17.2%
KORA (1064) 0.116 0.42 0.12 [−0.70; 0.93] 4.7% 4.7%
NSPHS (866) −0.861 0.28 −0.86 [−1.41; −0.31] 10.6% 10.6%
STANLEY (344) −0.396 0.89 −0.40 [−2.14; 1.35] 1.0% 1.0%
STANLEY (300) −1.110 0.66 −1.11 [−2.41; 0.19] 1.9% 1.9%
VIS (901) −0.096 0.67 −0.10 [−1.41; 1.21] 1.9% 1.9%
Common effect model −0.61 [−0.79; −0.43] 100.0% --
Random effects model −0.61 [−0.79; −0.43] -- 100.0%
−2
−1
0
1
2
Heterogeneity: $I^2$ = 0%, $\tau^2$ = 0, $p$ = 0.48

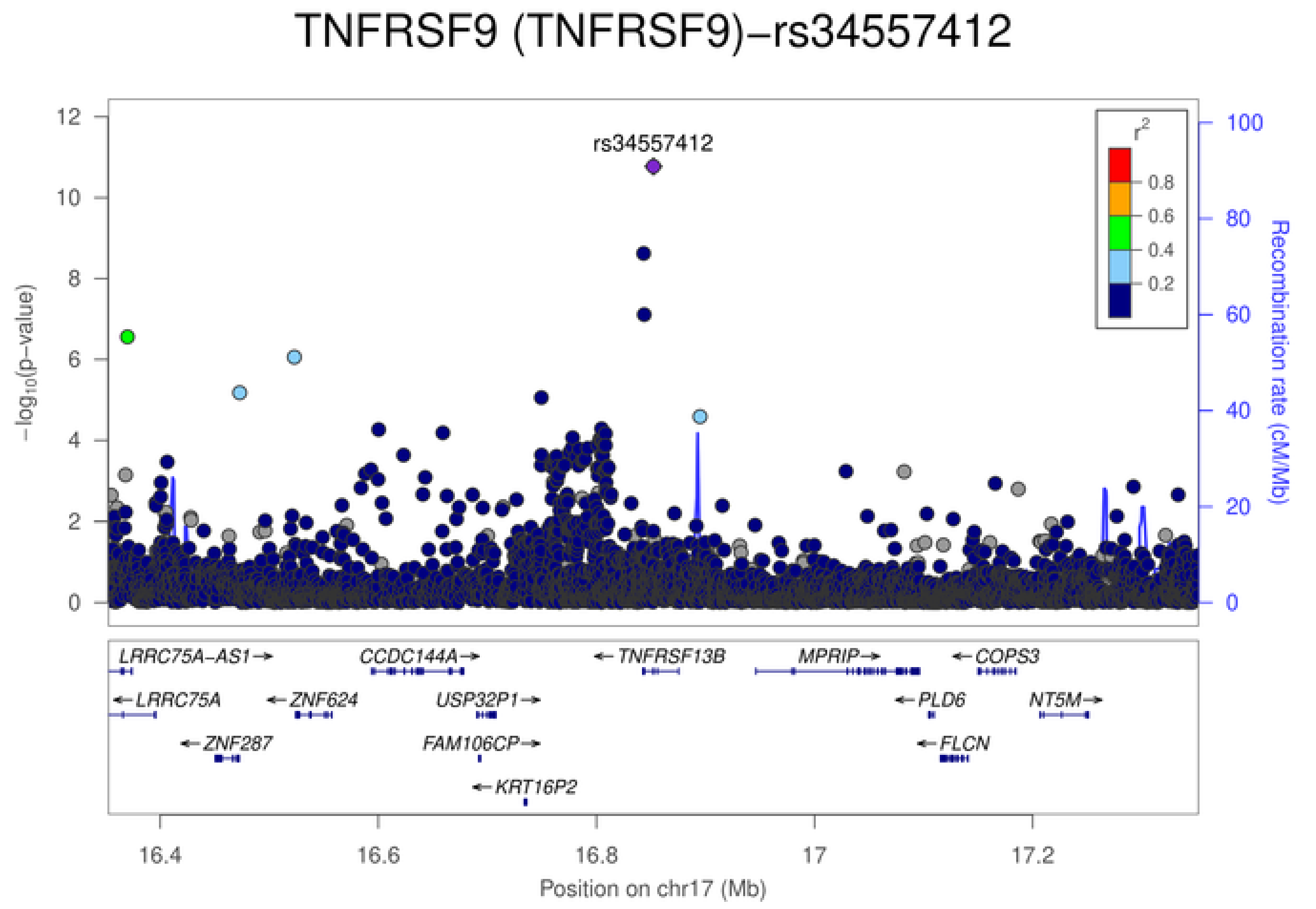
TNFRSF9 (TNFRSF9)−rs34557412
rs34557412
$-\log_{10}$(p−value)
Recombination rate (cM/Mb)
$r^2$
0.8
0.6
0.4
0.2
LRRC75A-AS1→
CCDC144A→
←TNFRSF13B
MPRIP→
←COPS3
←LRRC75A
←ZNF624
USP32P1→
←PLD6
NT5M→
←ZNF287
FAM106CP→
←FLCN
←KRT16P2
16.4
16.6
16.8
17
17.2
Position on chr17 (Mb)

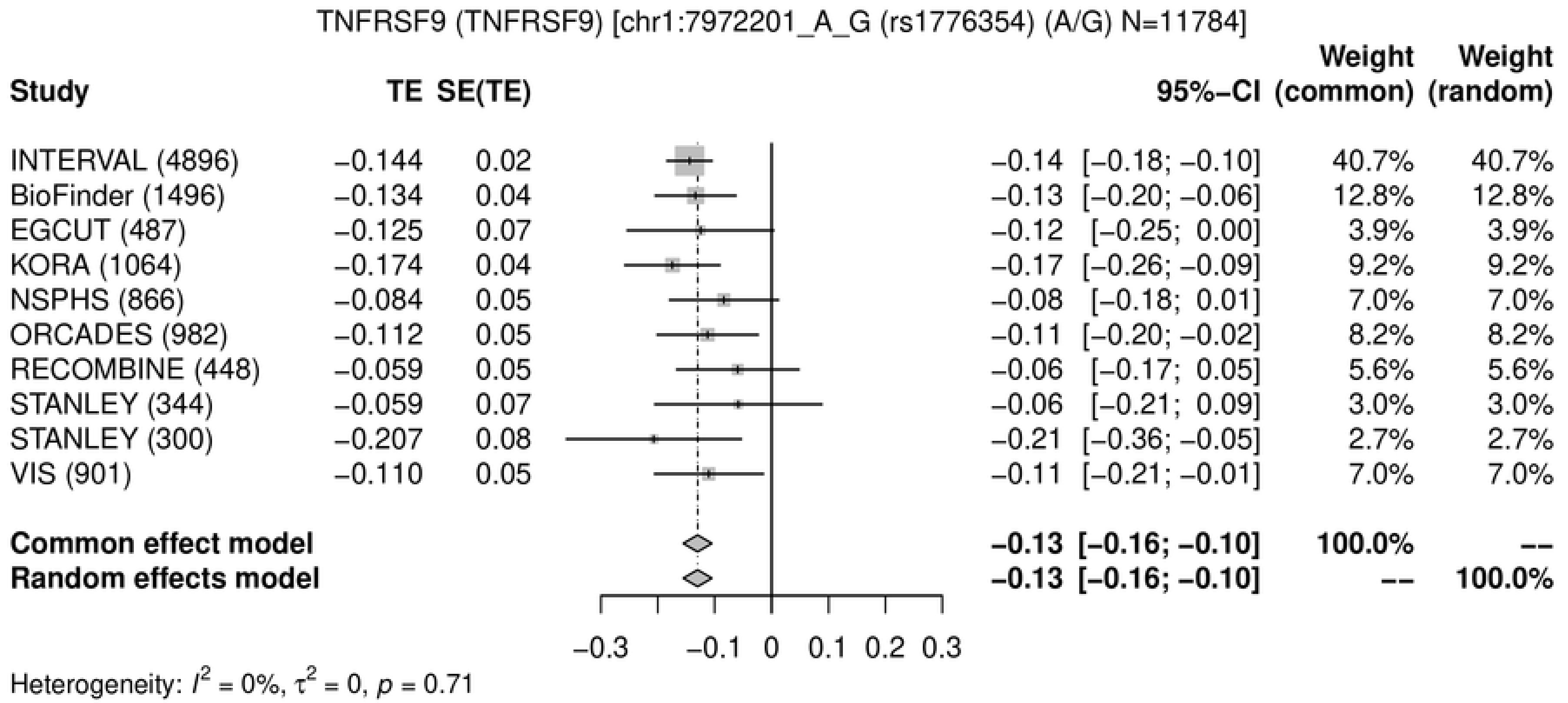
TNFRSF9 (TNFRSF9) [chr1:7972201_A_G (rs1776354) (A/G) N=11784]
Study
TE
SE(TE)
95%−CI
Weight (common)
Weight (random)
INTERVAL (4896) −0.144 0.02 −0.14 [−0.18; −0.10] 40.7% 40.7%
BioFinder (1496) −0.134 0.04 −0.13 [−0.20; −0.06] 12.8% 12.8%
EGCUT (487) −0.125 0.07 −0.12 [−0.25; 0.00] 3.9% 3.9%
KORA (1064) −0.174 0.04 −0.17 [−0.26; −0.09] 9.2% 9.2%
NSPHS (866) −0.084 0.05 −0.08 [−0.18; 0.01] 7.0% 7.0%
ORCADES (982) −0.112 0.05 −0.11 [−0.20; −0.02] 8.2% 8.2%
RECOMBINE (448) −0.059 0.05 −0.06 [−0.17; 0.05] 5.6% 5.6%
STANLEY (344) −0.059 0.07 −0.06 [−0.21; 0.09] 3.0% 3.0%
STANLEY (300) −0.207 0.08 −0.21 [−0.36; −0.05] 2.7% 2.7%
VIS (901) −0.110 0.05 −0.11 [−0.21; −0.01] 7.0% 7.0%
Common effect model −0.13 [−0.16; −0.10] 100.0% --
Random effects model −0.13 [−0.16; −0.10] -- 100.0%
−0.3 −0.1 0 0.1 0.2 0.3
Heterogeneity: $I^2$ = 0%, $\tau^2$ = 0, $p$ = 0.71

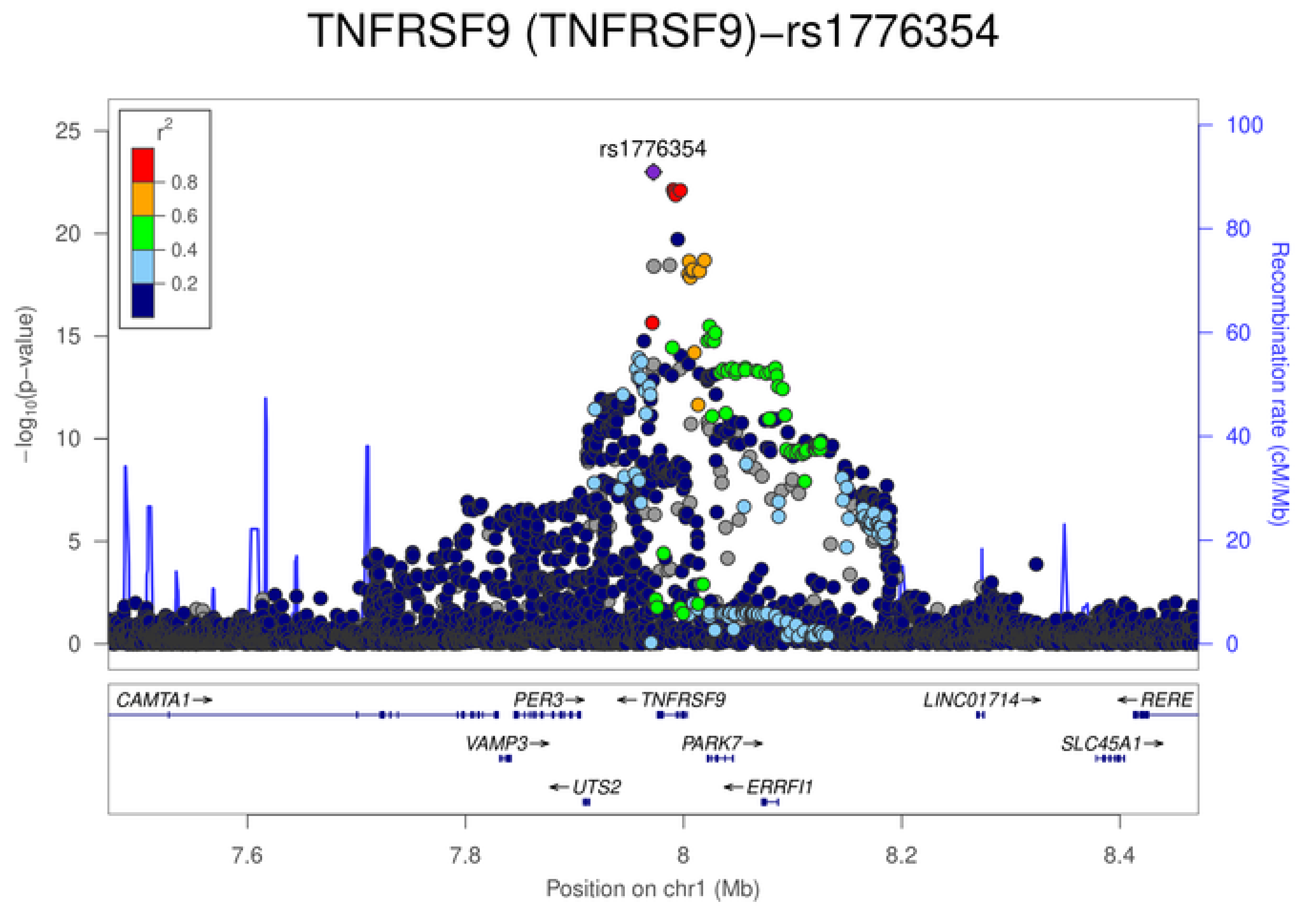
TNFRSF9 (TNFRSF9)−rs1776354
rs1776354
$r^2$
0.8
0.6
0.4
0.2
$-\log_{10}$(p−value)
Recombination rate (cM/Mb)
0 5 10 15 20 25
0 20 40 60 80 100
CAMTA1→
PER3→
← TNFRSF9
LINC01714→
← RERE
VAMP3→
PARK7→
SLC45A1→
← UTS2
← ERRFI1
7.6 7.8 8 8.2 8.4
Position on chr1 (Mb)

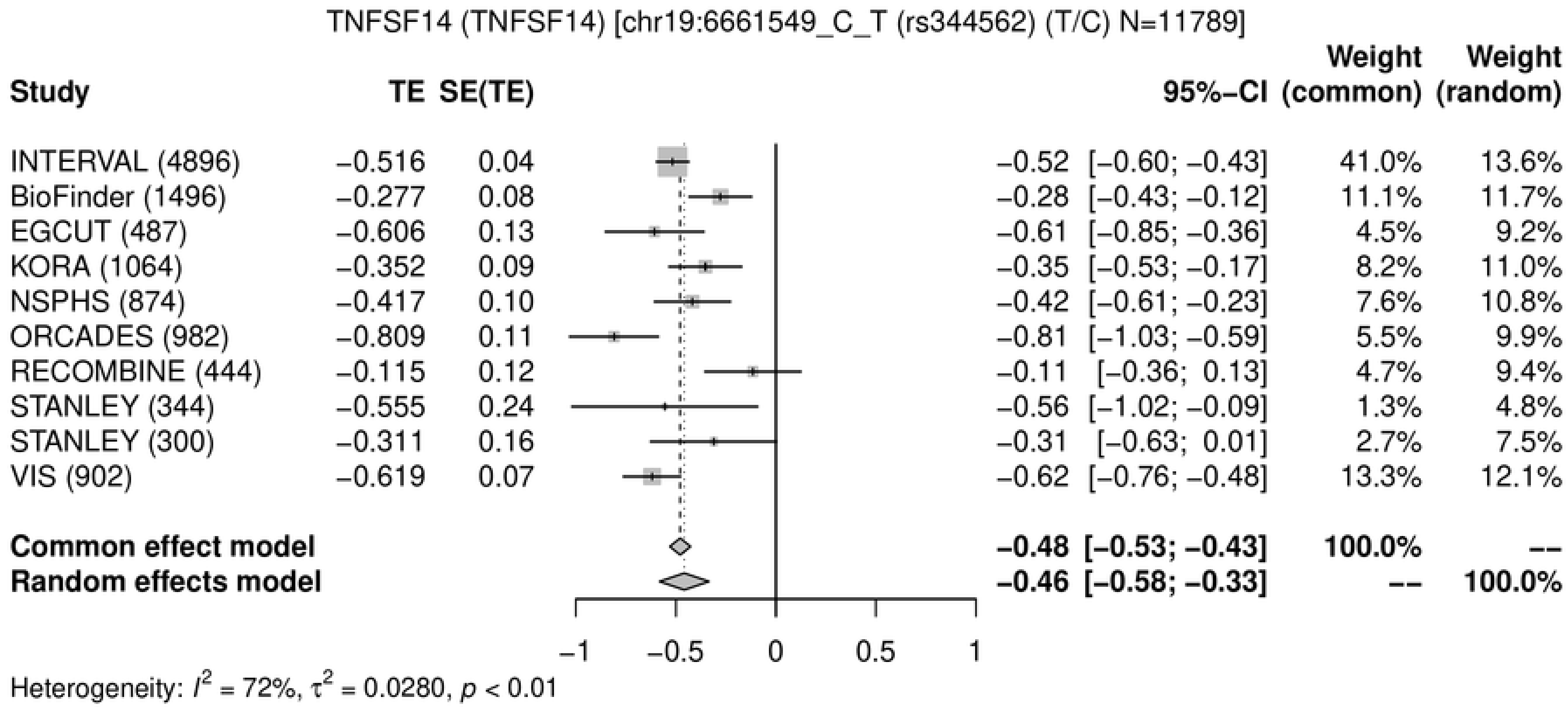
TNFSF14 (TNFSF14) [chr19:6661549_C_T (rs344562) (T/C) N=11789]
Study TE SE(TE) 95%-CI Weight (common) Weight (random)
INTERVAL (4896) −0.516 0.04 −0.52 [−0.60; −0.43] 41.0% 13.6%
BioFinder (1496) −0.277 0.08 −0.28 [−0.43; −0.12] 11.1% 11.7%
EGCUT (487) −0.606 0.13 −0.61 [−0.85; −0.36] 4.5% 9.2%
KORA (1064) −0.352 0.09 −0.35 [−0.53; −0.17] 8.2% 11.0%
NSPHS (874) −0.417 0.10 −0.42 [−0.61; −0.23] 7.6% 10.8%
ORCADES (982) −0.809 0.11 −0.81 [−1.03; −0.59] 5.5% 9.9%
RECOMBINE (444) −0.115 0.12 −0.11 [−0.36; 0.13] 4.7% 9.4%
STANLEY (344) −0.555 0.24 −0.56 [−1.02; −0.09] 1.3% 4.8%
STANLEY (300) −0.311 0.16 −0.31 [−0.63; 0.01] 2.7% 7.5%
VIS (902) −0.619 0.07 −0.62 [−0.76; −0.48] 13.3% 12.1%
Common effect model −0.48 [−0.53; −0.43] 100.0% --
Random effects model −0.46 [−0.58; −0.33] -- 100.0%
−1 −0.5 0 0.5 1
Heterogeneity: $I^2$ = 72%, $\tau^2$ = 0.0280, $p$ < 0.01

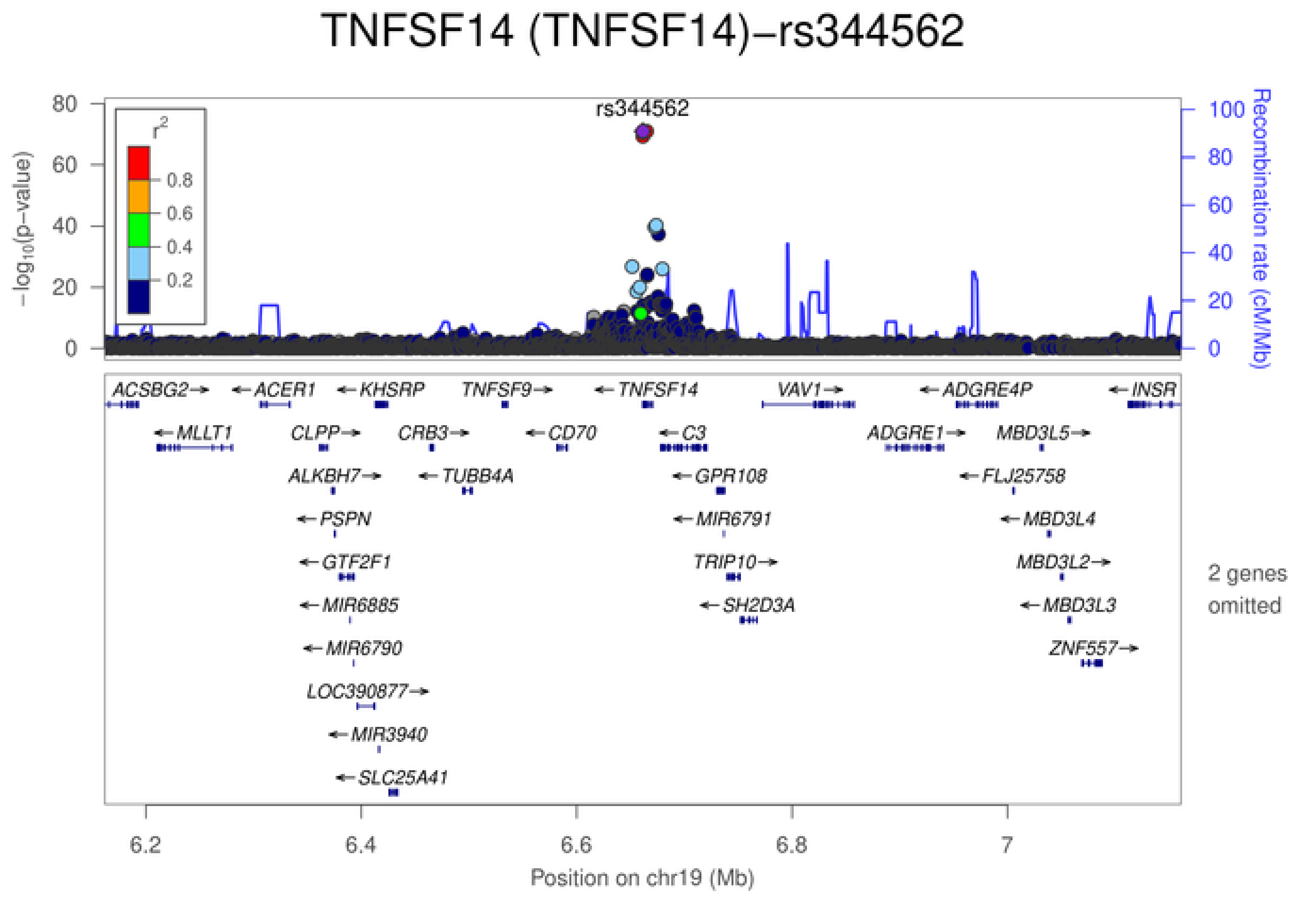
TNFSF14 (TNFSF14)−rs344562
rs344562
$r^2$
0.8
0.6
0.4
0.2
$-\log_{10}$(p-value)
Recombination rate (cM/Mb)
ACSBG2 ACER1 KHSRP TNFSF9 TNFSF14 VAV1 ADGRE4P INSR
MLLT1 CLPP CRB3 CD70 C3 ADGRE1 MBD3L5
ALKBH7 TUBB4A GPR108 FLJ25758
PSPN MIR6791 MBD3L4
GTF2F1 TRIP10 MBD3L2
MIR6885 SH2D3A MBD3L3
MIR6790 ZNF557
LOC390877
MIR3940
SLC25A41
2 genes omitted
6.2 6.4 6.6 6.8 7
Position on chr19 (Mb)

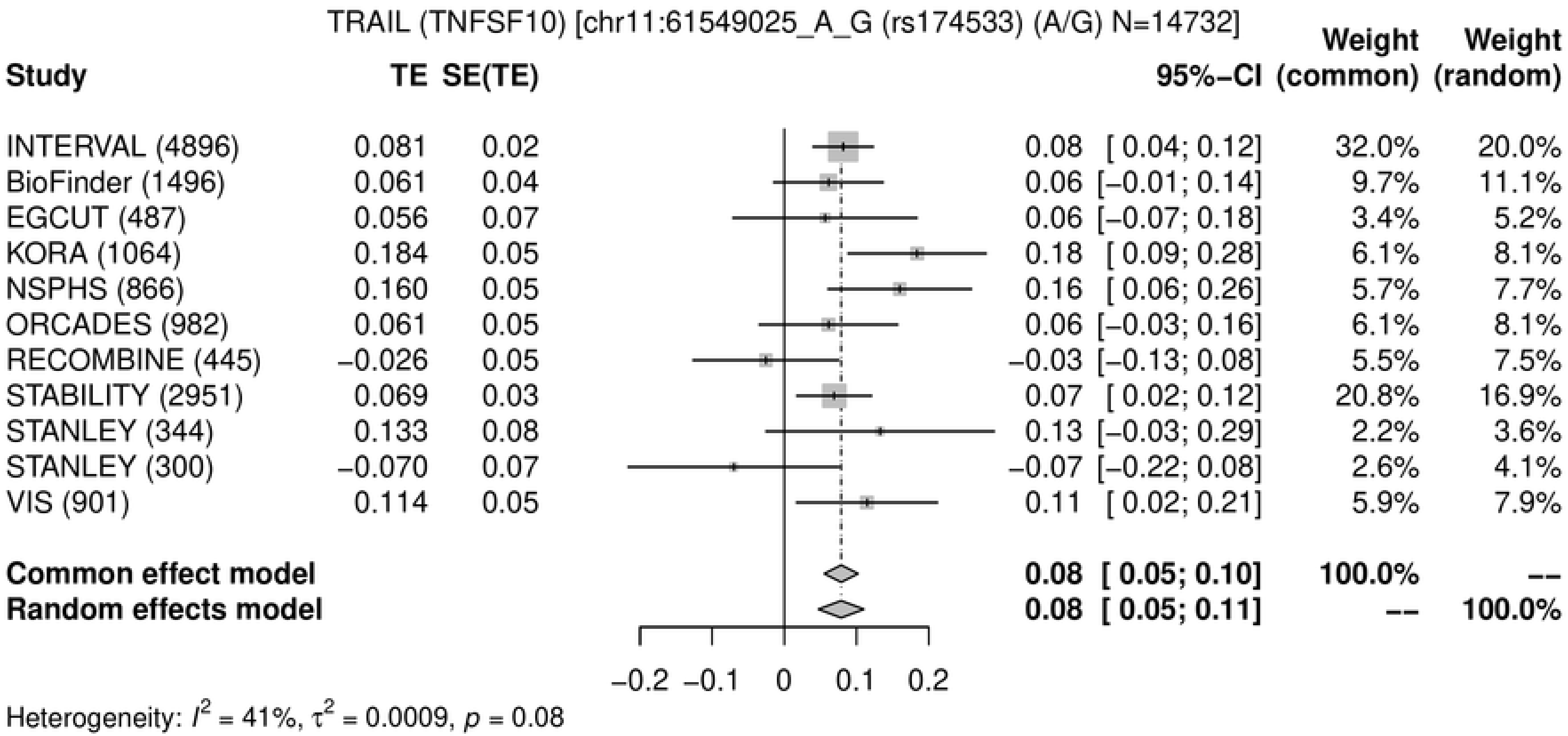
TRAIL (TNFSF10) [chr11:61549025_A_G (rs174533) (A/G) N=14732]
Study TE SE(TE) 95%-CI Weight (common) Weight (random)
INTERVAL (4896) 0.081 0.02 0.08 [ 0.04; 0.12] 32.0% 20.0%
BioFinder (1496) 0.061 0.04 0.06 [−0.01; 0.14] 9.7% 11.1%
EGCUT (487) 0.056 0.07 0.06 [−0.07; 0.18] 3.4% 5.2%
KORA (1064) 0.184 0.05 0.18 [ 0.09; 0.28] 6.1% 8.1%
NSPHS (866) 0.160 0.05 0.16 [ 0.06; 0.26] 5.7% 7.7%
ORCADES (982) 0.061 0.05 0.06 [−0.03; 0.16] 6.1% 8.1%
RECOMBINE (445) −0.026 0.05 −0.03 [−0.13; 0.08] 5.5% 7.5%
STABILITY (2951) 0.069 0.03 0.07 [ 0.02; 0.12] 20.8% 16.9%
STANLEY (344) 0.133 0.08 0.13 [−0.03; 0.29] 2.2% 3.6%
STANLEY (300) −0.070 0.07 −0.07 [−0.22; 0.08] 2.6% 4.1%
VIS (901) 0.114 0.05 0.11 [ 0.02; 0.21] 5.9% 7.9%
Common effect model 0.08 [ 0.05; 0.10] 100.0% --
Random effects model 0.08 [ 0.05; 0.11] -- 100.0%
−0.2 −0.1 0 0.1 0.2
Heterogeneity: $I^2$ = 41%, $\tau^2$ = 0.0009, $p$ = 0.08

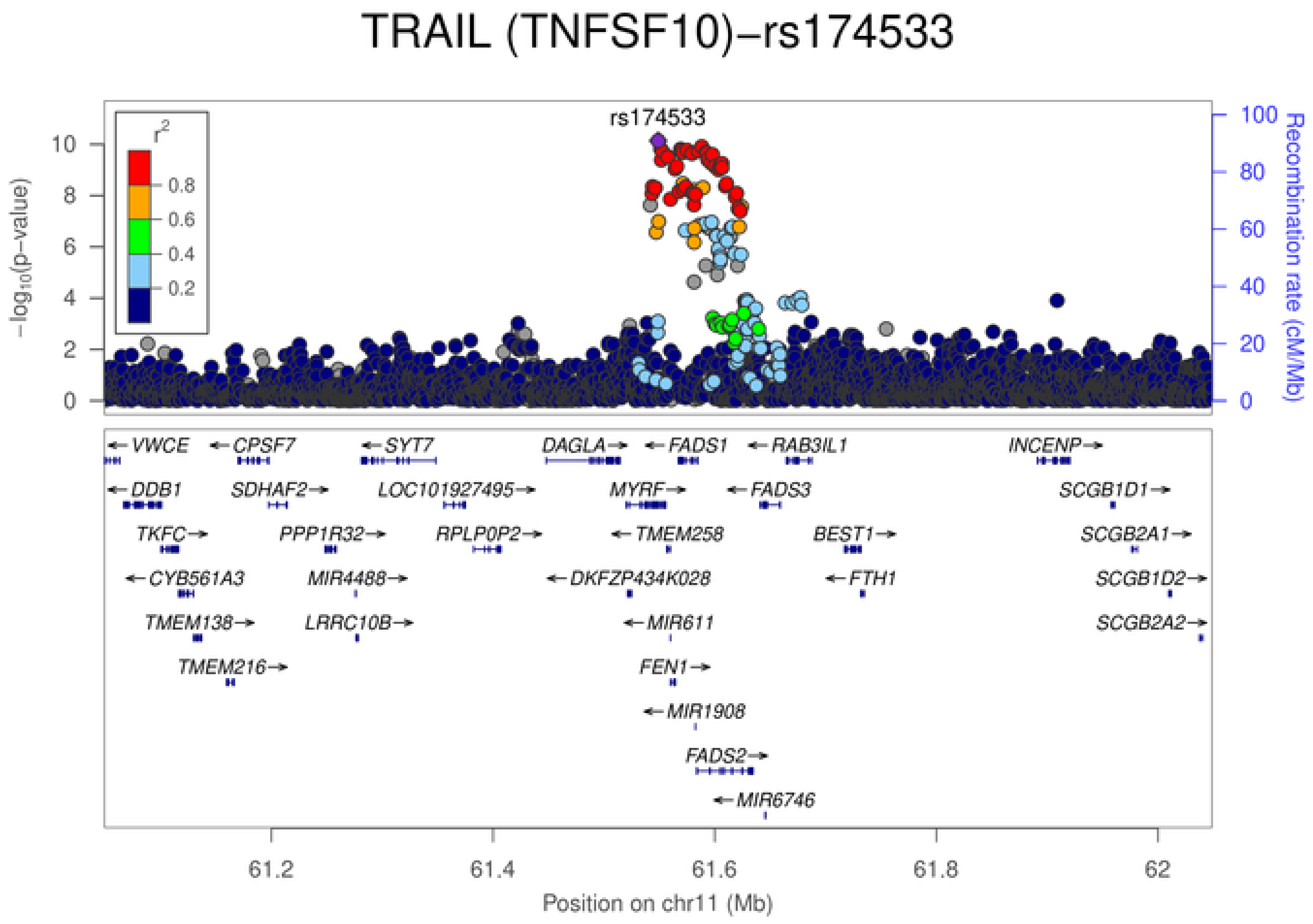
TRAIL (TNFSF10)−rs174533
rs174533
$r^2$
0.8
0.6
0.4
0.2
$-\log_{10}$(p-value)
Recombination rate (cM/Mb)
VWCE
CPSF7
SYT7
DAGLA
FADS1
RAB3IL1
INCENP
DDB1
SDHAF2
LOC101927495
MYRF
FADS3
SCGB1D1
TKFC
PPP1R32
RPLP0P2
TMEM258
BEST1
SCGB2A1
CYB561A3
MIR4488
DKFZP434K028
FTH1
SCGB1D2
TMEM138
LRRC10B
MIR611
SCGB2A2
TMEM216
FEN1
MIR1908
FADS2
MIR6746
61.2
61.4
61.6
61.8
62
Position on chr11 (Mb)

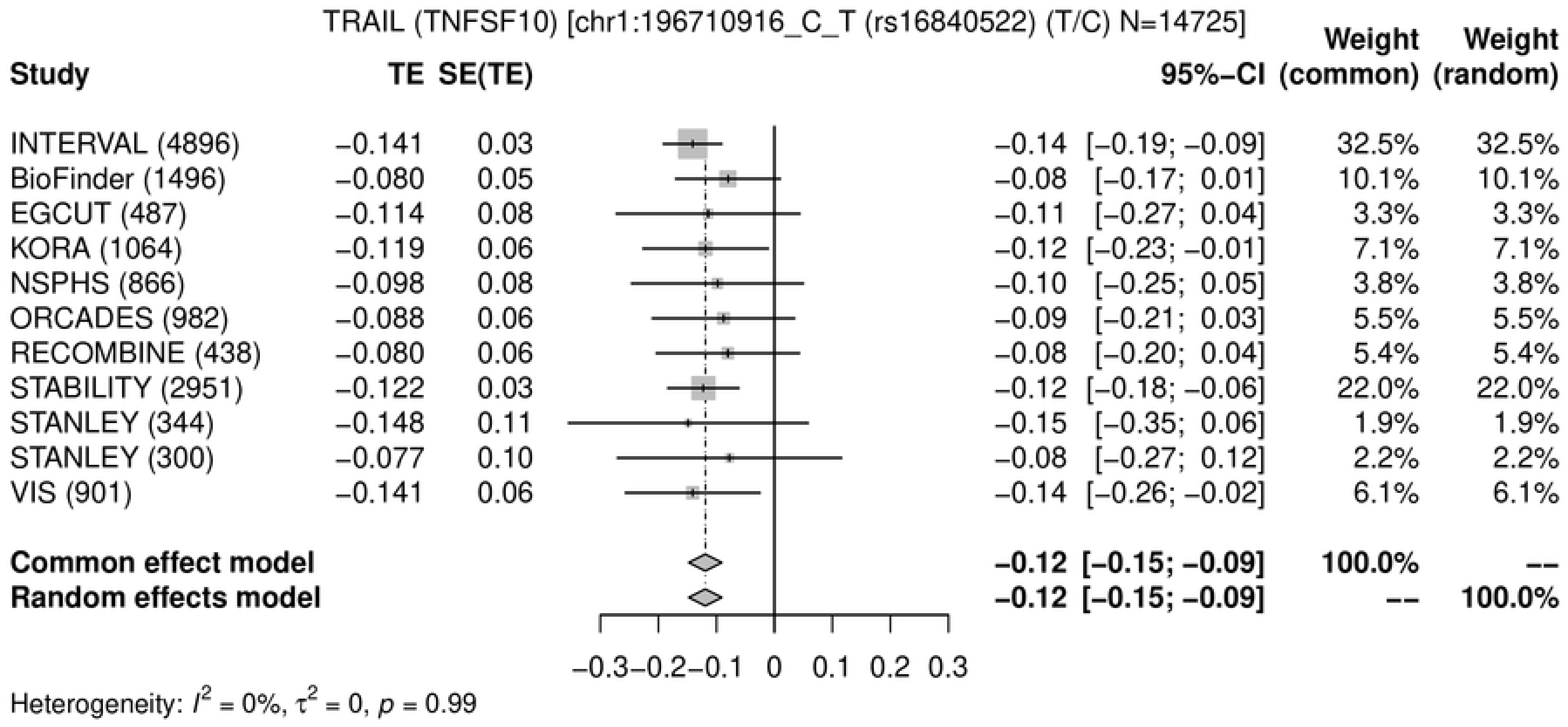
TRAIL (TNFSF10) [chr1:196710916_C_T (rs16840522) (T/C) N=14725]
Study
TE
SE(TE)
95%-CI
Weight (common)
Weight (random)
INTERVAL (4896) −0.141 0.03 −0.14 [−0.19; −0.09] 32.5% 32.5%
BioFinder (1496) −0.080 0.05 −0.08 [−0.17; 0.01] 10.1% 10.1%
EGCUT (487) −0.114 0.08 −0.11 [−0.27; 0.04] 3.3% 3.3%
KORA (1064) −0.119 0.06 −0.12 [−0.23; −0.01] 7.1% 7.1%
NSPHS (866) −0.098 0.08 −0.10 [−0.25; 0.05] 3.8% 3.8%
ORCADES (982) −0.088 0.06 −0.09 [−0.21; 0.03] 5.5% 5.5%
RECOMBINE (438) −0.080 0.06 −0.08 [−0.20; 0.04] 5.4% 5.4%
STABILITY (2951) −0.122 0.03 −0.12 [−0.18; −0.06] 22.0% 22.0%
STANLEY (344) −0.148 0.11 −0.15 [−0.35; 0.06] 1.9% 1.9%
STANLEY (300) −0.077 0.10 −0.08 [−0.27; 0.12] 2.2% 2.2%
VIS (901) −0.141 0.06 −0.14 [−0.26; −0.02] 6.1% 6.1%
Common effect model −0.12 [−0.15; −0.09] 100.0% --
Random effects model −0.12 [−0.15; −0.09] -- 100.0%
−0.3 −0.2 −0.1 0 0.1 0.2 0.3
Heterogeneity: $I^2$ = 0%, $\tau^2$ = 0, $p$ = 0.99

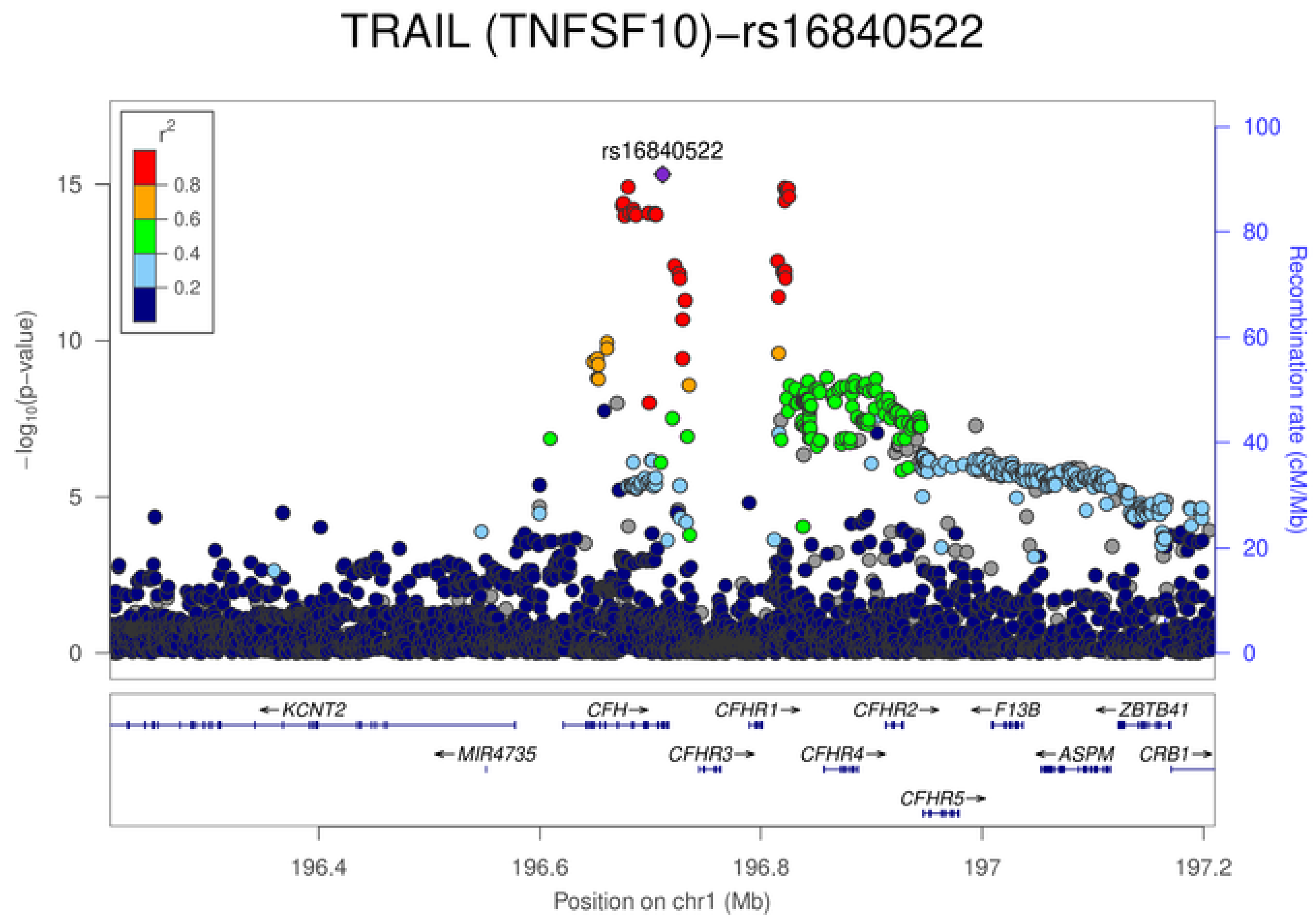
TRAIL (TNFSF10)−rs16840522
rs16840522
$r^2$
0.8
0.6
0.4
0.2
$-\log_{10}$(p−value)
Recombination rate (cM/Mb)
KCNT2
CFH
CFHR1
CFHR2
F13B
ZBTB41
MIR4735
CFHR3
CFHR4
ASPM
CRB1
CFHR5
196.4
196.6
196.8
197
197.2
Position on chr1 (Mb)

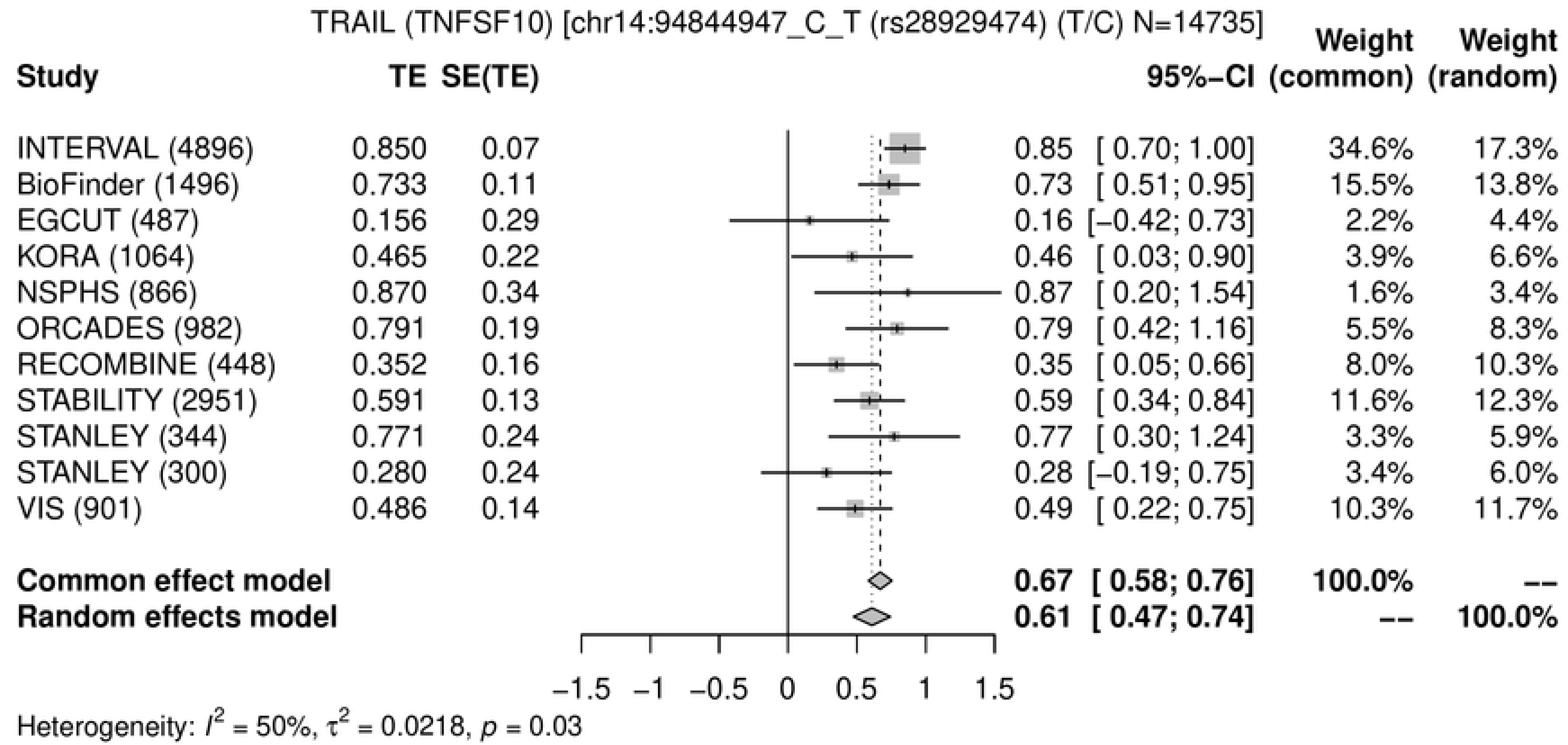
TRAIL (TNFSF10) [chr14:94844947_C_T (rs28929474) (T/C) N=14735]
Study TE SE(TE) 95%−CI Weight (common) Weight (random)
INTERVAL (4896) 0.850 0.07 0.85 [ 0.70; 1.00] 34.6% 17.3%
BioFinder (1496) 0.733 0.11 0.73 [ 0.51; 0.95] 15.5% 13.8%
EGCUT (487) 0.156 0.29 0.16 [−0.42; 0.73] 2.2% 4.4%
KORA (1064) 0.465 0.22 0.46 [ 0.03; 0.90] 3.9% 6.6%
NSPHS (866) 0.870 0.34 0.87 [ 0.20; 1.54] 1.6% 3.4%
ORCADES (982) 0.791 0.19 0.79 [ 0.42; 1.16] 5.5% 8.3%
RECOMBINE (448) 0.352 0.16 0.35 [ 0.05; 0.66] 8.0% 10.3%
STABILITY (2951) 0.591 0.13 0.59 [ 0.34; 0.84] 11.6% 12.3%
STANLEY (344) 0.771 0.24 0.77 [ 0.30; 1.24] 3.3% 5.9%
STANLEY (300) 0.280 0.24 0.28 [−0.19; 0.75] 3.4% 6.0%
VIS (901) 0.486 0.14 0.49 [ 0.22; 0.75] 10.3% 11.7%
Common effect model 0.67 [ 0.58; 0.76] 100.0% --
Random effects model 0.61 [ 0.47; 0.74] -- 100.0%
−1.5 −1 −0.5 0 0.5 1 1.5
Heterogeneity: $I^2$ = 50%, $\tau^2$ = 0.0218, $p$ = 0.03

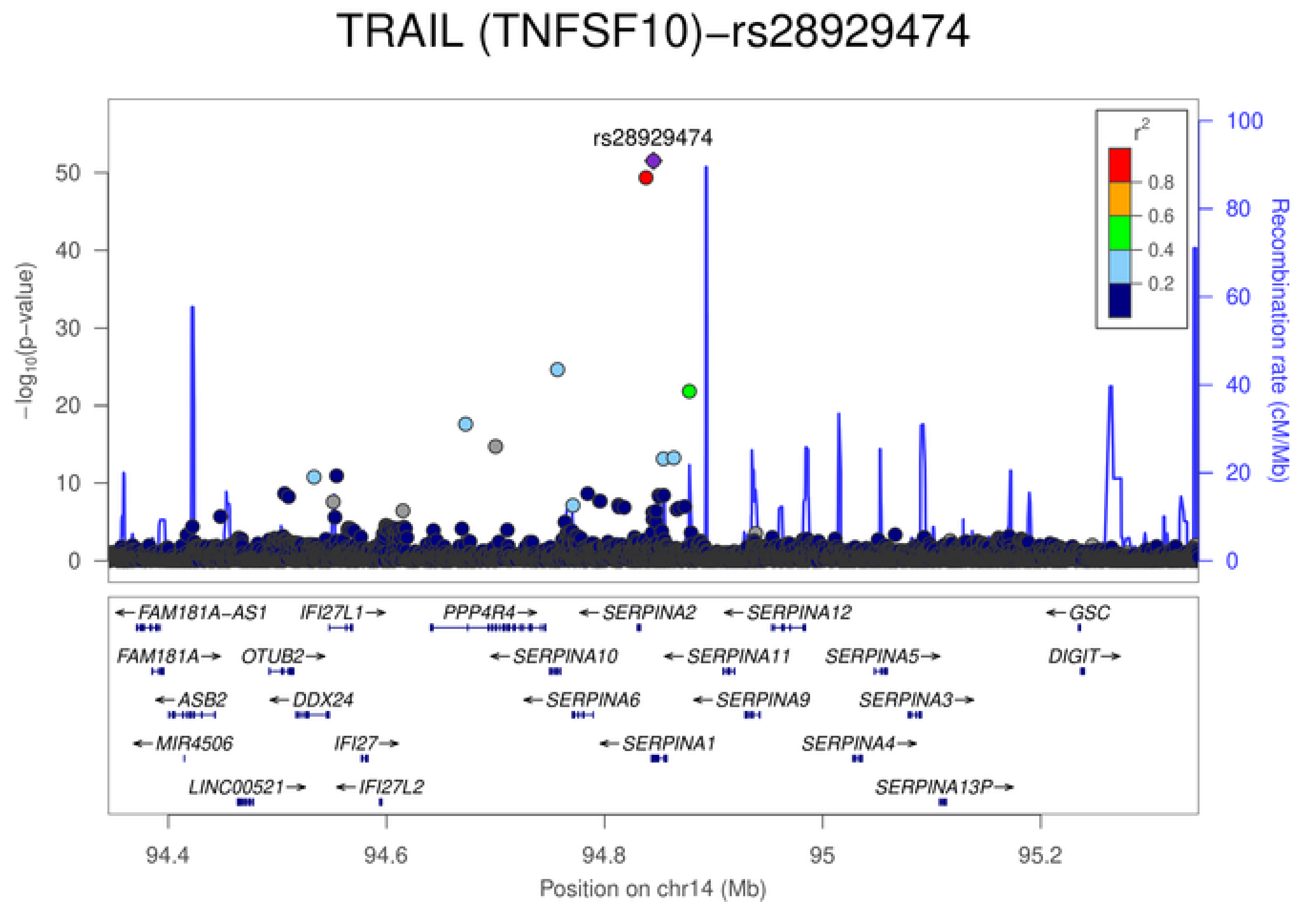
TRAIL (TNFSF10)−rs28929474
rs28929474
$-\log_{10}$(p−value)
Recombination rate (cM/Mb)
$r^2$
0.8
0.6
0.4
0.2
FAM181A−AS1
IFI27L1
PPP4R4
SERPINA2
SERPINA12
GSC
FAM181A
OTUB2
SERPINA10
SERPINA11
SERPINA5
DIGIT
ASB2
DDX24
SERPINA6
SERPINA9
SERPINA3
MIR4506
IFI27
SERPINA1
SERPINA4
LINC00521
IFI27L2
SERPINA13P
94.4 94.6 94.8 95 95.2
Position on chr14 (Mb)

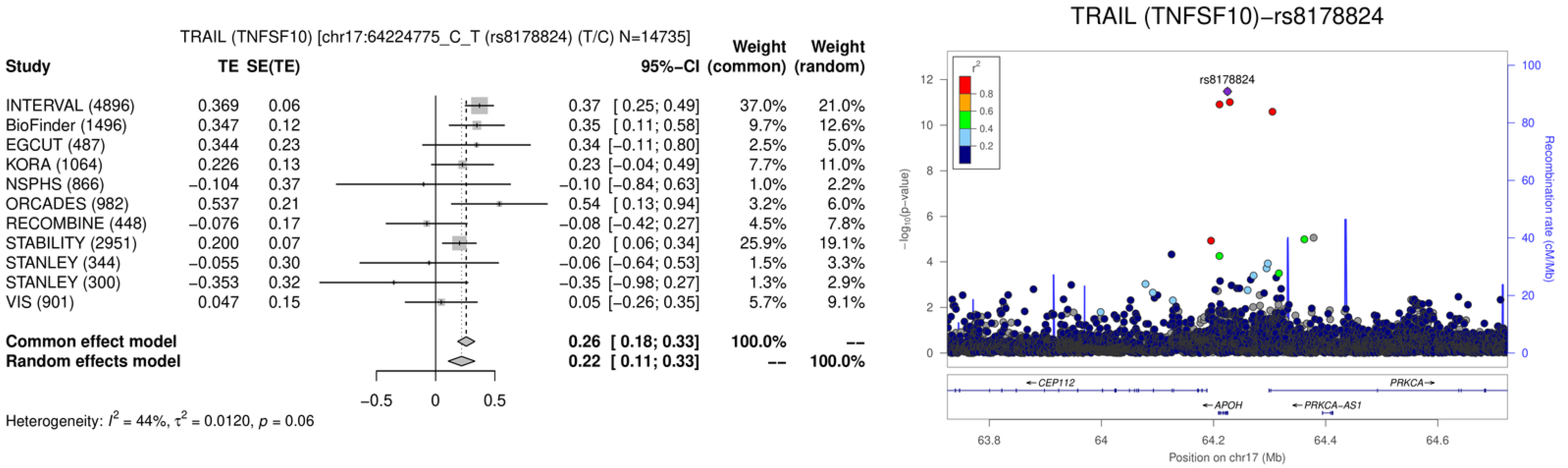
TRAIL (TNFSF10) [chr17:64224775_C_T (rs8178824) (T/C) N=14735]
Study
TE
SE(TE)
95%–CI
Weight (common)
Weight (random)
INTERVAL (4896) 0.369 0.06 0.37 [ 0.25; 0.49] 37.0% 21.0%
BioFinder (1496) 0.347 0.12 0.35 [ 0.11; 0.58] 9.7% 12.6%
EGCUT (487) 0.344 0.23 0.34 [–0.11; 0.80] 2.5% 5.0%
KORA (1064) 0.226 0.13 0.23 [–0.04; 0.49] 7.7% 11.0%
NSPHS (866) –0.104 0.37 –0.10 [–0.84; 0.63] 1.0% 2.2%
ORCADES (982) 0.537 0.21 0.54 [ 0.13; 0.94] 3.2% 6.0%
RECOMBINE (448) –0.076 0.17 –0.08 [–0.42; 0.27] 4.5% 7.8%
STABILITY (2951) 0.200 0.07 0.20 [ 0.06; 0.34] 25.9% 19.1%
STANLEY (344) –0.055 0.30 –0.06 [–0.64; 0.53] 1.5% 3.3%
STANLEY (300) –0.353 0.32 –0.35 [–0.98; 0.27] 1.3% 2.9%
VIS (901) 0.047 0.15 0.05 [–0.26; 0.35] 5.7% 9.1%
Common effect model 0.26 [ 0.18; 0.33] 100.0% --
Random effects model 0.22 [ 0.11; 0.33] -- 100.0%
–0.5
0
0.5
Heterogeneity: I² = 44%, τ² = 0.0120, p = 0.06
TRAIL (TNFSF10)–rs8178824
rs8178824
r²
0.8
0.6
0.4
0.2
–log10(p–value)
Recombination rate (cM/Mb)
CEP112
PRKCA
APOH
PRKCA-AS1
63.8
64
64.2
64.4
64.6
Position on chr17 (Mb)

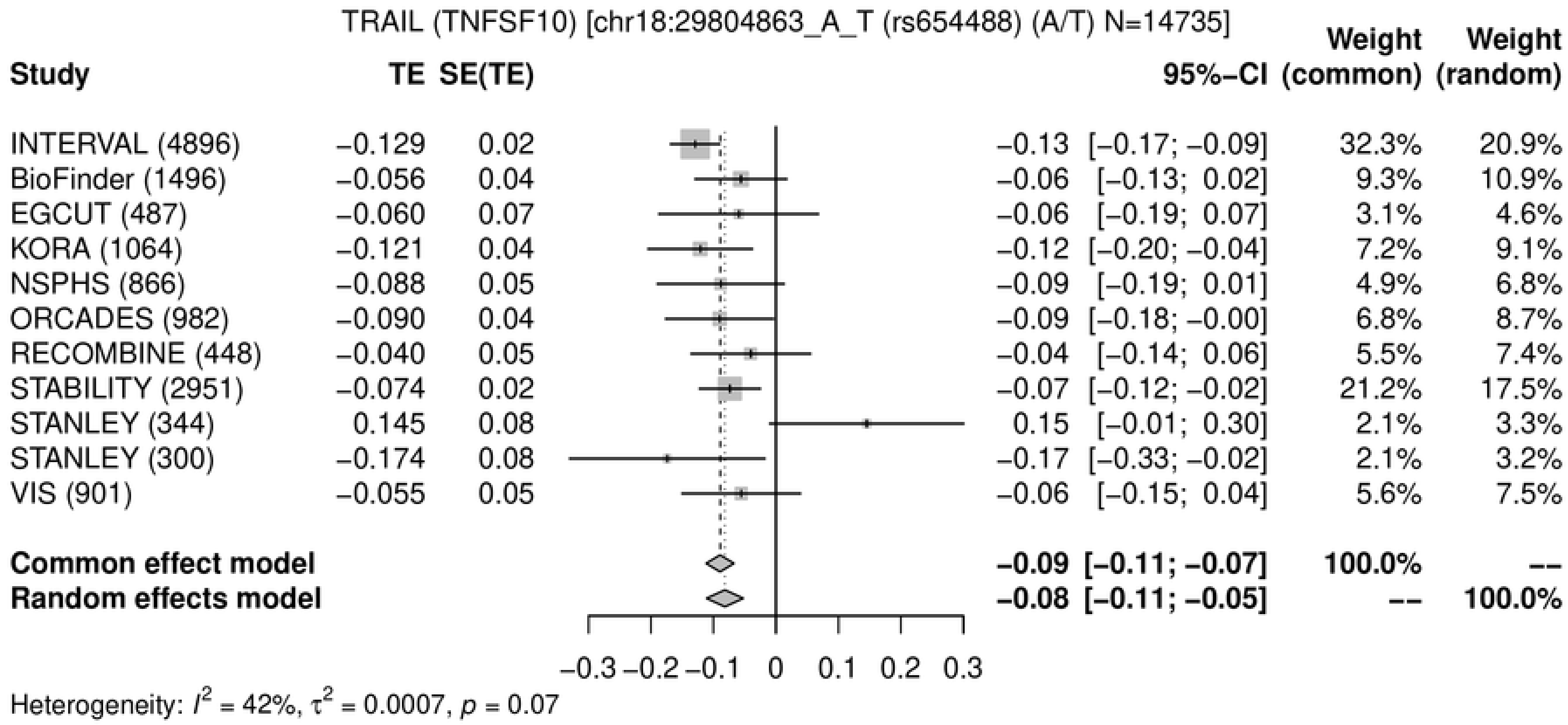

TRAIL (TNFSF10) [chr18:29804863_A_T (rs654488) (A/T) N=14735]
Study
TE
SE(TE)
95%-CI
Weight (common)
Weight (random)
INTERVAL (4896) −0.129 0.02 −0.13 [−0.17; −0.09] 32.3% 20.9%
BioFinder (1496) −0.056 0.04 −0.06 [−0.13; 0.02] 9.3% 10.9%
EGCUT (487) −0.060 0.07 −0.06 [−0.19; 0.07] 3.1% 4.6%
KORA (1064) −0.121 0.04 −0.12 [−0.20; −0.04] 7.2% 9.1%
NSPHS (866) −0.088 0.05 −0.09 [−0.19; 0.01] 4.9% 6.8%
ORCADES (982) −0.090 0.04 −0.09 [−0.18; −0.00] 6.8% 8.7%
RECOMBINE (448) −0.040 0.05 −0.04 [−0.14; 0.06] 5.5% 7.4%
STABILITY (2951) −0.074 0.02 −0.07 [−0.12; −0.02] 21.2% 17.5%
STANLEY (344) 0.145 0.08 0.15 [−0.01; 0.30] 2.1% 3.3%
STANLEY (300) −0.174 0.08 −0.17 [−0.33; −0.02] 2.1% 3.2%
VIS (901) −0.055 0.05 −0.06 [−0.15; 0.04] 5.6% 7.5%
Common effect model −0.09 [−0.11; −0.07] 100.0% --
Random effects model −0.08 [−0.11; −0.05] -- 100.0%
−0.3 −0.2 −0.1 0 0.1 0.2 0.3
Heterogeneity: $I^2$ = 42%, $\tau^2$ = 0.0007, $p$ = 0.07


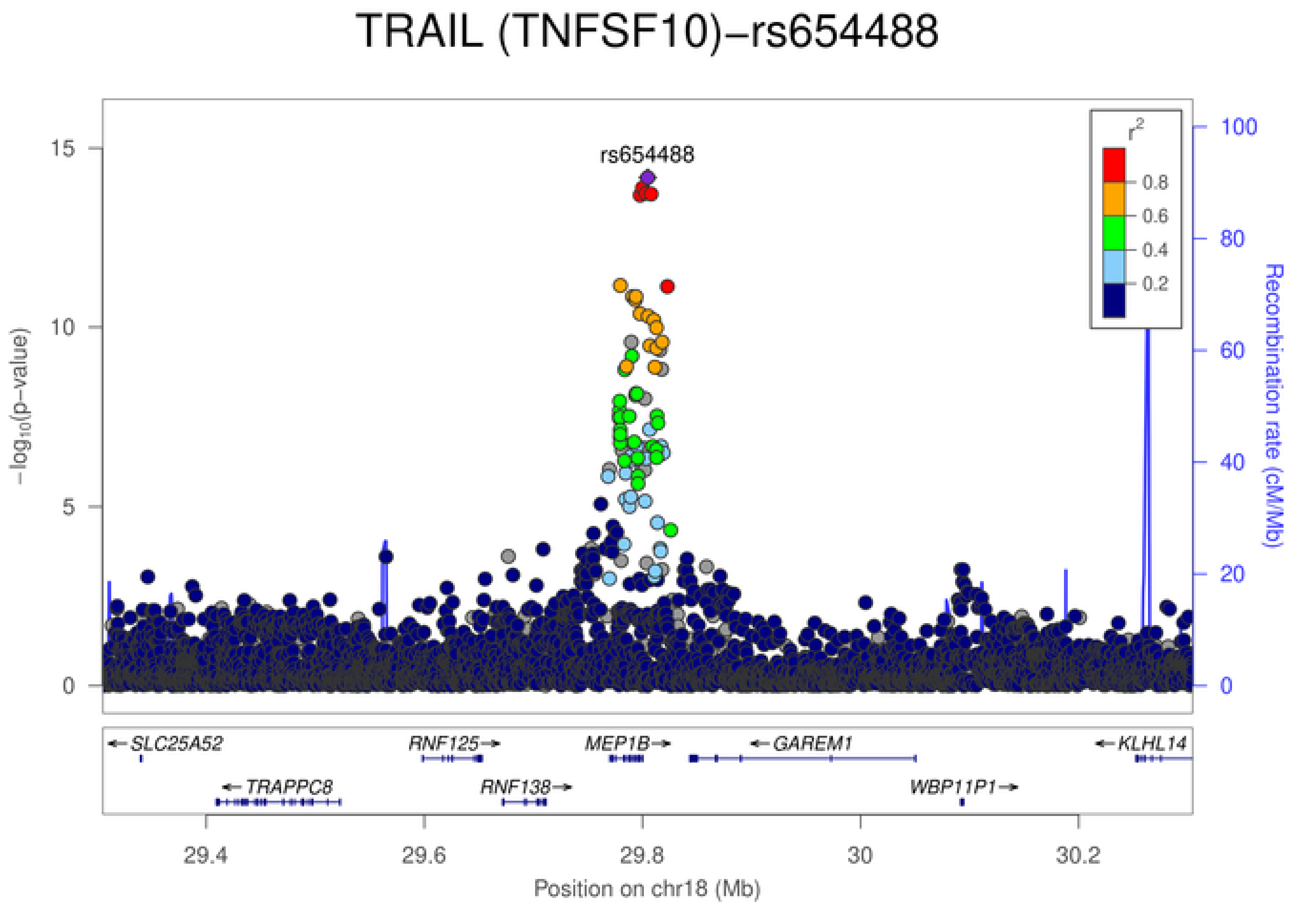

TRAIL (TNFSF10)−rs654488
rs654488
$-\log_{10}$(p−value)
Recombination rate (cM/Mb)
$r^2$
0.8
0.6
0.4
0.2
SLC25A52
RNF125
MEP1B
GAREM1
KLHL14
TRAPPC8
RNF138
WBP11P1
29.4 29.6 29.8 30 30.2
Position on chr18 (Mb)

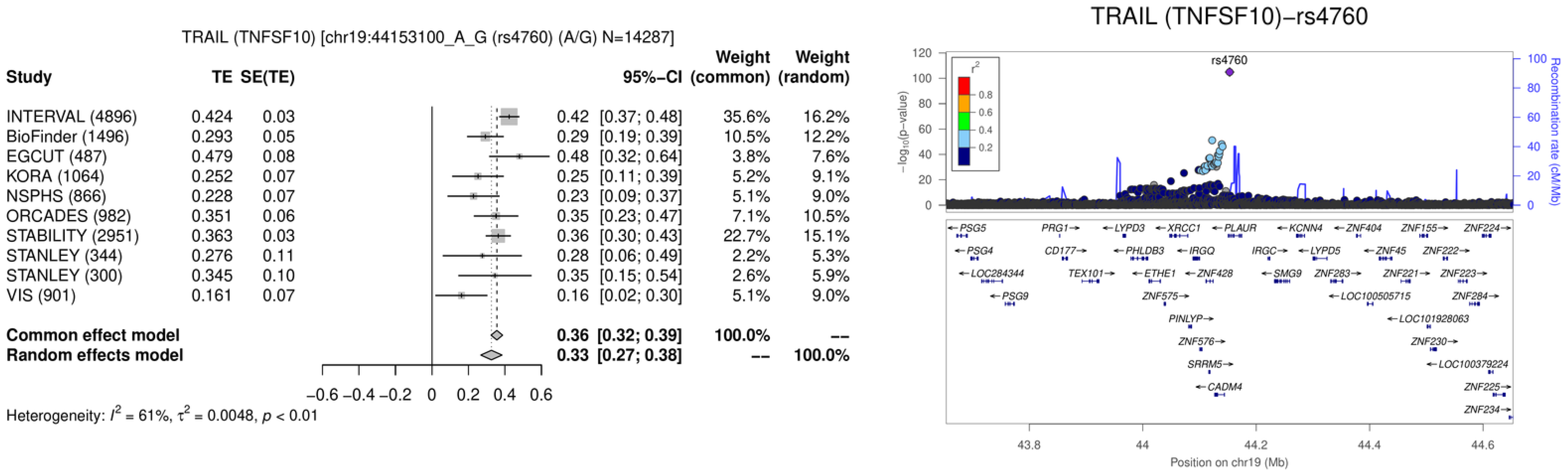

TRAIL (TNFSF10) [chr19:44153100_A_G (rs4760) (A/G) N=14287]
Study TE SE(TE) 95%-CI Weight (common) Weight (random)
INTERVAL (4896) 0.424 0.03 0.42 [0.37; 0.48] 35.6% 16.2%
BioFinder (1496) 0.293 0.05 0.29 [0.19; 0.39] 10.5% 12.2%
EGCUT (487) 0.479 0.08 0.48 [0.32; 0.64] 3.8% 7.6%
KORA (1064) 0.252 0.07 0.25 [0.11; 0.39] 5.2% 9.1%
NSPHS (866) 0.228 0.07 0.23 [0.09; 0.37] 5.1% 9.0%
ORCADES (982) 0.351 0.06 0.35 [0.23; 0.47] 7.1% 10.5%
STABILITY (2951) 0.363 0.03 0.36 [0.30; 0.43] 22.7% 15.1%
STANLEY (344) 0.276 0.11 0.28 [0.06; 0.49] 2.2% 5.3%
STANLEY (300) 0.345 0.10 0.35 [0.15; 0.54] 2.6% 5.9%
VIS (901) 0.161 0.07 0.16 [0.02; 0.30] 5.1% 9.0%
Common effect model 0.36 [0.32; 0.39] 100.0% --
Random effects model 0.33 [0.27; 0.38] -- 100.0%
−0.6 −0.4 −0.2 0 0.2 0.4 0.6
Heterogeneity: $I^2$ = 61%, $\tau^2$ = 0.0048, $p$ < 0.01
TRAIL (TNFSF10)−rs4760
rs4760
$r^2$
0.8
0.6
0.4
0.2
$-\log_{10}$(p-value)
Recombination rate (cM/Mb)
PSG5 PRG1 LYPD3 XRCC1 PLAUR KCNN4 ZNF404 ZNF155 ZNF224
PSG4 CD177 PHLDB3 IRGQ IRGC LYPD5 ZNF45 ZNF222
LOC284344 TEX101 ETHE1 ZNF428 SMG9 ZNF283 ZNF221 ZNF223
PSG9 ZNF575 LOC100505715 ZNF284
PINLYP LOC101928063
ZNF576 ZNF230
SRRM5 LOC100379224
CADM4 ZNF225
ZNF234
43.8 44 44.2 44.4 44.6
Position on chr19 (Mb)

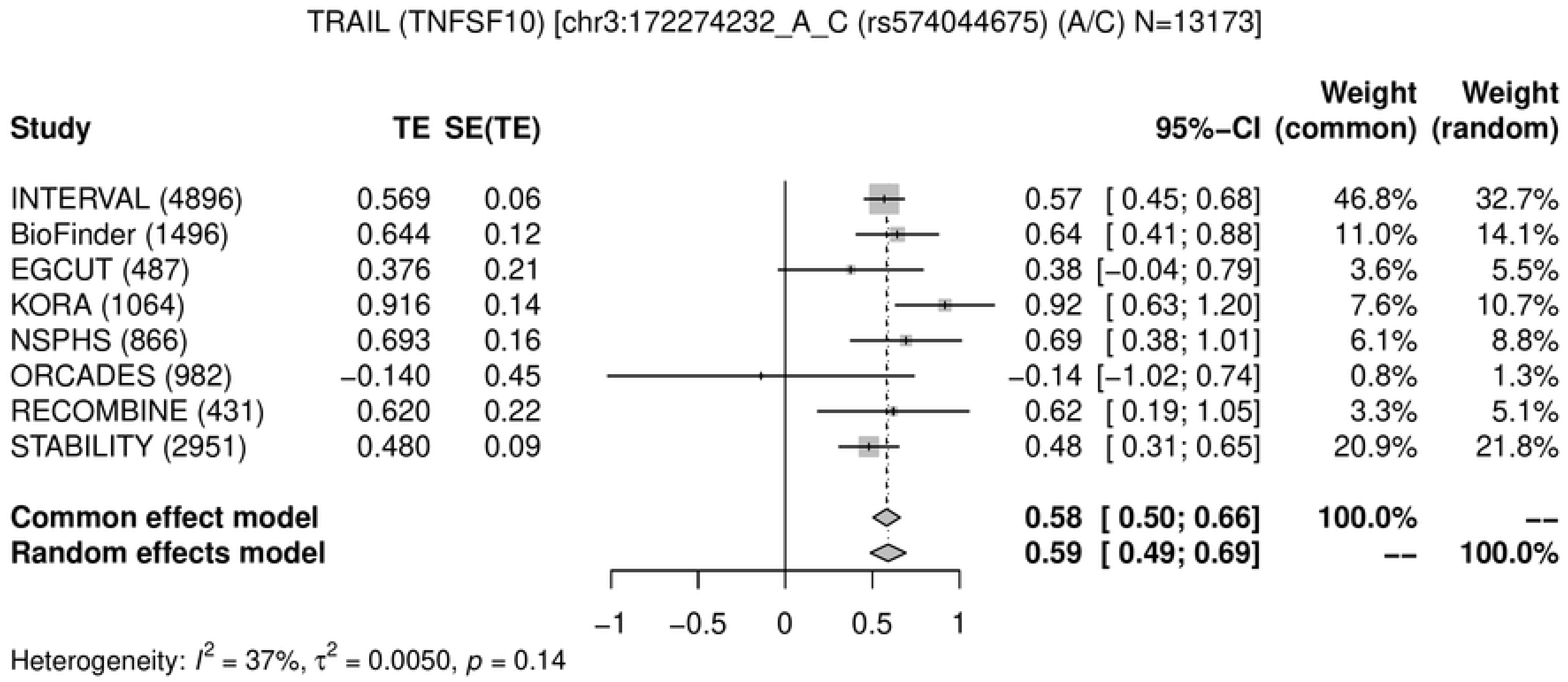
TRAIL (TNFSF10) [chr3:172274232_A_C (rs574044675) (A/C) N=13173]
Study TE SE(TE) 95%-CI Weight (common) Weight (random)
INTERVAL (4896) 0.569 0.06 0.57 [ 0.45; 0.68] 46.8% 32.7%
BioFinder (1496) 0.644 0.12 0.64 [ 0.41; 0.88] 11.0% 14.1%
EGCUT (487) 0.376 0.21 0.38 [−0.04; 0.79] 3.6% 5.5%
KORA (1064) 0.916 0.14 0.92 [ 0.63; 1.20] 7.6% 10.7%
NSPHS (866) 0.693 0.16 0.69 [ 0.38; 1.01] 6.1% 8.8%
ORCADES (982) −0.140 0.45 −0.14 [−1.02; 0.74] 0.8% 1.3%
RECOMBINE (431) 0.620 0.22 0.62 [ 0.19; 1.05] 3.3% 5.1%
STABILITY (2951) 0.480 0.09 0.48 [ 0.31; 0.65] 20.9% 21.8%
Common effect model 0.58 [ 0.50; 0.66] 100.0% --
Random effects model 0.59 [ 0.49; 0.69] -- 100.0%
−1 −0.5 0 0.5 1
Heterogeneity: $I^2$ = 37%, $\tau^2$ = 0.0050, $p$ = 0.14

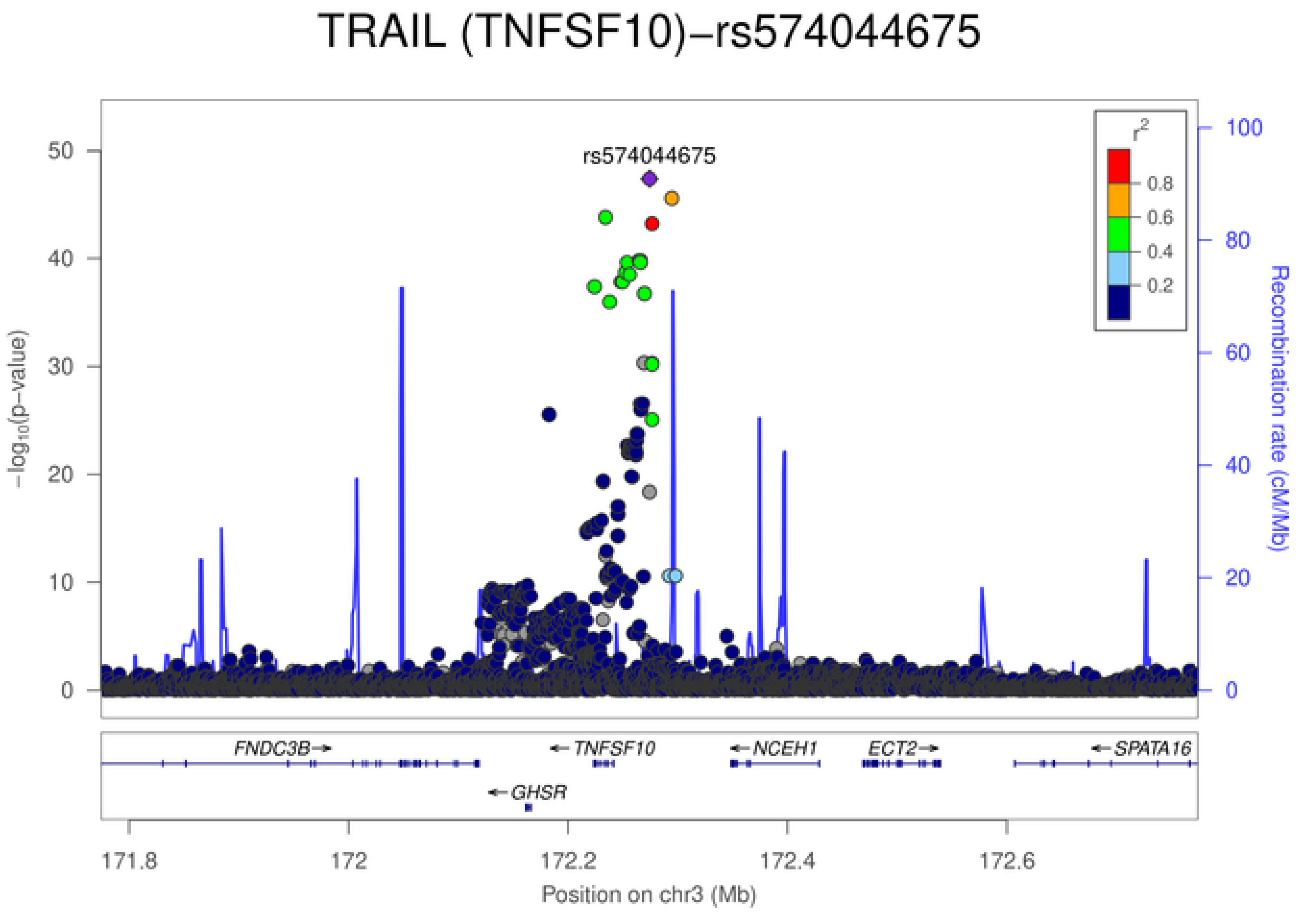
TRAIL (TNFSF10)−rs574044675
rs574044675
$-\log_{10}$(p−value)
Recombination rate (cM/Mb)
$r^2$
0.8
0.6
0.4
0.2
FNDC3B
TNFSF10
NCEH1
ECT2
SPATA16
GHSR
171.8
172
172.2
172.4
172.6
Position on chr3 (Mb)

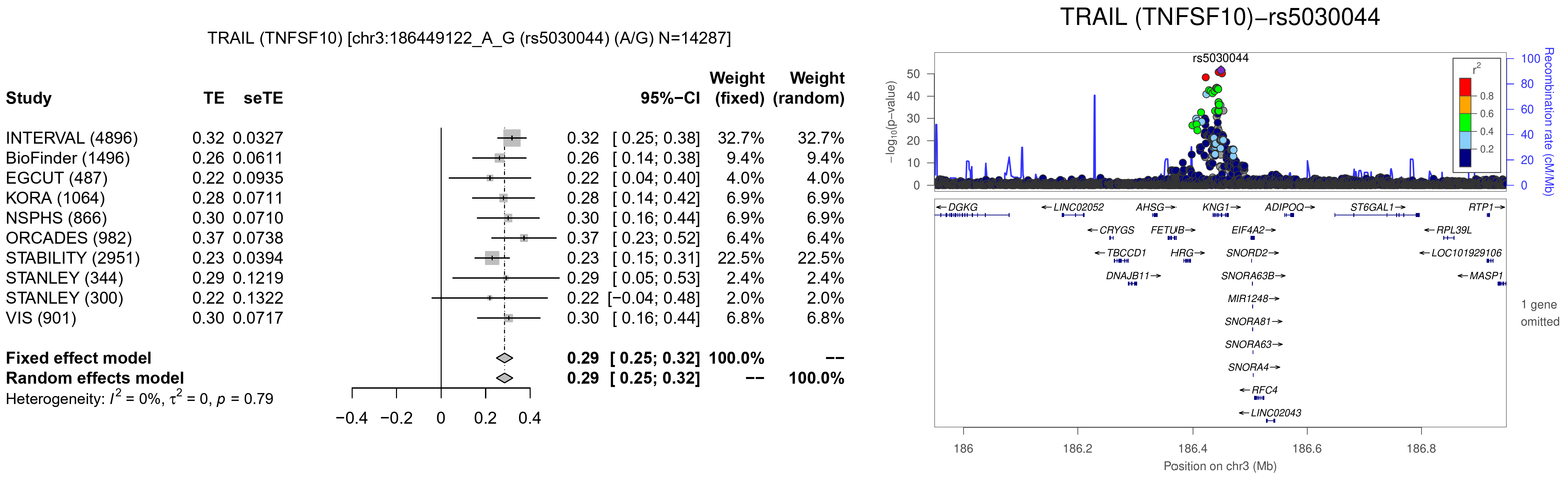

TRAIL (TNFSF10) [chr3:186449122_A_G (rs5030044) (A/G) N=14287]
Study
TE
seTE
95%−CI
Weight (fixed)
Weight (random)
INTERVAL (4896) 0.32 0.0327 0.32 [ 0.25; 0.38] 32.7% 32.7%
BioFinder (1496) 0.26 0.0611 0.26 [ 0.14; 0.38] 9.4% 9.4%
EGCUT (487) 0.22 0.0935 0.22 [ 0.04; 0.40] 4.0% 4.0%
KORA (1064) 0.28 0.0711 0.28 [ 0.14; 0.42] 6.9% 6.9%
NSPHS (866) 0.30 0.0710 0.30 [ 0.16; 0.44] 6.9% 6.9%
ORCADES (982) 0.37 0.0738 0.37 [ 0.23; 0.52] 6.4% 6.4%
STABILITY (2951) 0.23 0.0394 0.23 [ 0.15; 0.31] 22.5% 22.5%
STANLEY (344) 0.29 0.1219 0.29 [ 0.05; 0.53] 2.4% 2.4%
STANLEY (300) 0.22 0.1322 0.22 [−0.04; 0.48] 2.0% 2.0%
VIS (901) 0.30 0.0717 0.30 [ 0.16; 0.44] 6.8% 6.8%
Fixed effect model 0.29 [ 0.25; 0.32] 100.0% --
Random effects model 0.29 [ 0.25; 0.32] -- 100.0%
Heterogeneity: $I^2 = 0\%$, $\tau^2 = 0$, $p = 0.79$
−0.4 −0.2 0 0.2 0.4
TRAIL (TNFSF10)−rs5030044
rs5030044
$-\log_{10}$(p−value)
Recombination rate (cM/Mb)
$r^2$
0.8
0.6
0.4
0.2
DGKG
LINC02052
AHSG
KNG1
ADIPOQ
ST6GAL1
RTP1
CRYGS
FETUB
EIF4A2
RPL39L
TBCCD1
HRG
SNORD2
LOC101929106
DNAJB11
SNORA63B
MASP1
MIR1248
SNORA81
SNORA63
SNORA4
RFC4
LINC02043
1 gene omitted
186
186.2
186.4
186.6
186.8
Position on chr3 (Mb)

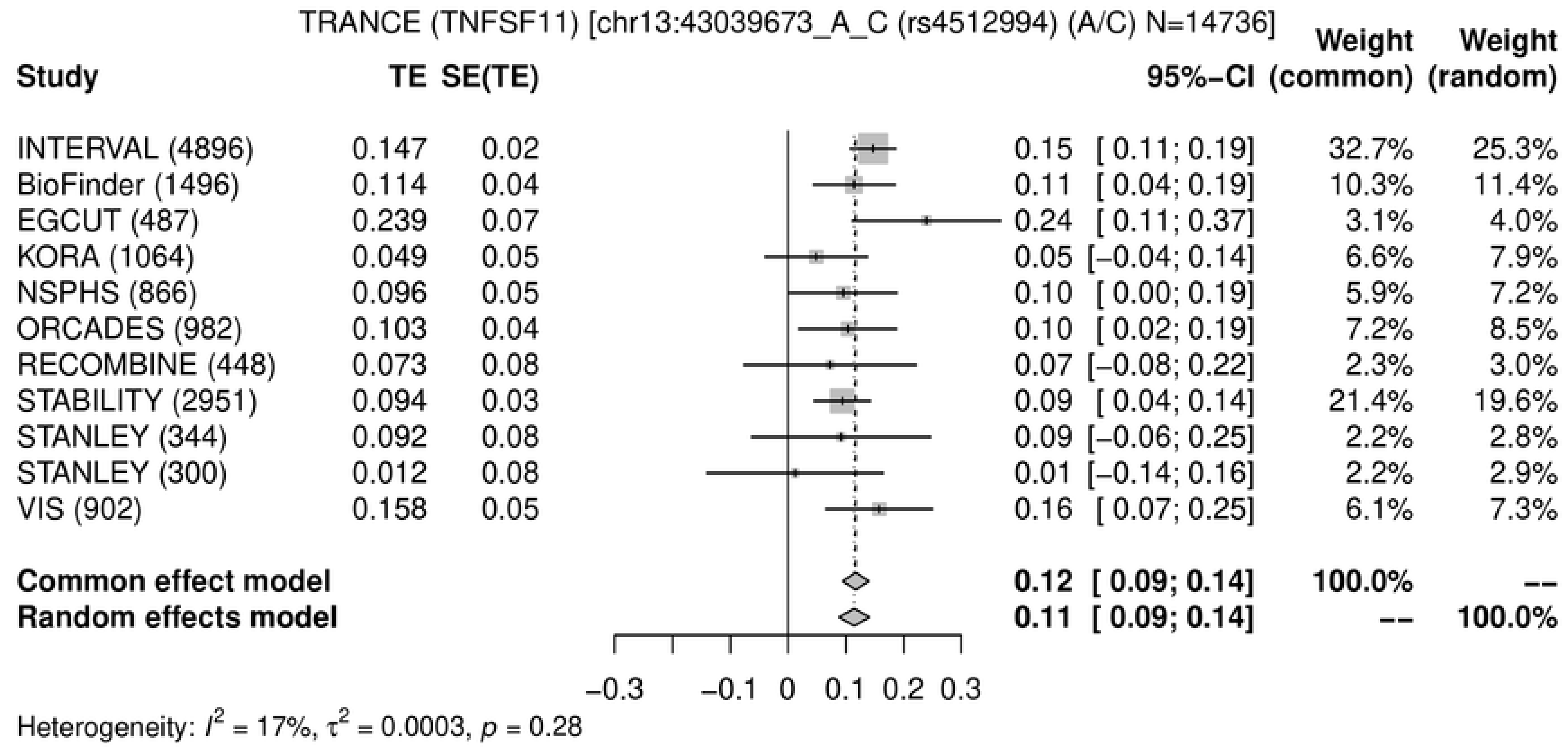
TRANCE (TNFSF11) [chr13:43039673_A_C (rs4512994) (A/C) N=14736]
Study
TE
SE(TE)
95%−CI
Weight (common)
Weight (random)
INTERVAL (4896) 0.147 0.02 0.15 [ 0.11; 0.19] 32.7% 25.3%
BioFinder (1496) 0.114 0.04 0.11 [ 0.04; 0.19] 10.3% 11.4%
EGCUT (487) 0.239 0.07 0.24 [ 0.11; 0.37] 3.1% 4.0%
KORA (1064) 0.049 0.05 0.05 [−0.04; 0.14] 6.6% 7.9%
NSPHS (866) 0.096 0.05 0.10 [ 0.00; 0.19] 5.9% 7.2%
ORCADES (982) 0.103 0.04 0.10 [ 0.02; 0.19] 7.2% 8.5%
RECOMBINE (448) 0.073 0.08 0.07 [−0.08; 0.22] 2.3% 3.0%
STABILITY (2951) 0.094 0.03 0.09 [ 0.04; 0.14] 21.4% 19.6%
STANLEY (344) 0.092 0.08 0.09 [−0.06; 0.25] 2.2% 2.8%
STANLEY (300) 0.012 0.08 0.01 [−0.14; 0.16] 2.2% 2.9%
VIS (902) 0.158 0.05 0.16 [ 0.07; 0.25] 6.1% 7.3%
Common effect model 0.12 [ 0.09; 0.14] 100.0% --
Random effects model 0.11 [ 0.09; 0.14] -- 100.0%
−0.3 −0.1 0 0.1 0.2 0.3
Heterogeneity: $I^2$ = 17%, $\tau^2$ = 0.0003, $p$ = 0.28

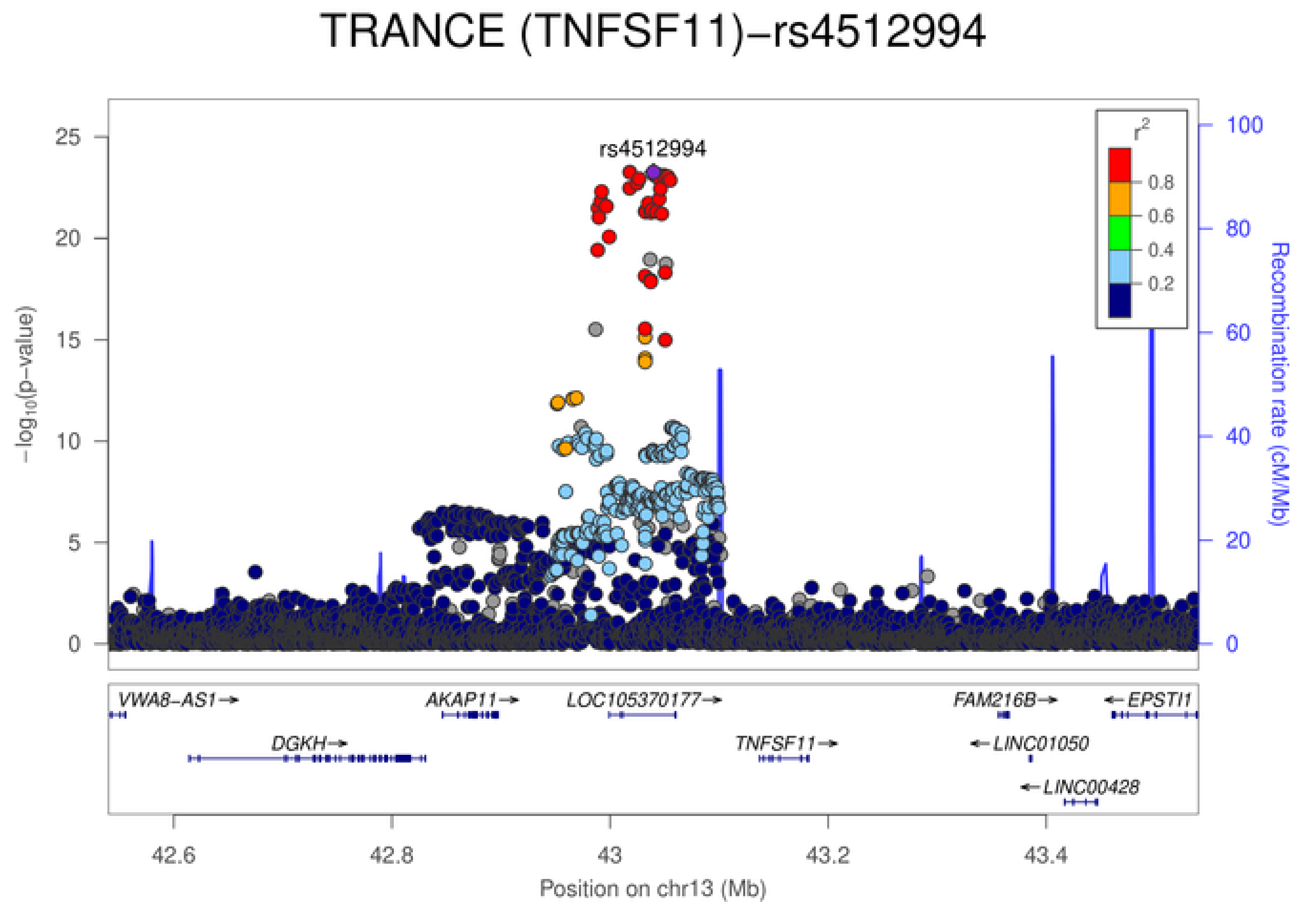
TRANCE (TNFSF11)−rs4512994
rs4512994
$-\log_{10}$(p−value)
Recombination rate (cM/Mb)
$r^2$
0.8
0.6
0.4
0.2
0
5
10
15
20
25
0
20
40
60
80
100
VWA8−AS1→
AKAP11→
LOC105370177→
FAM216B→
←EPSTI1
DGKH→
TNFSF11→
←LINC01050
←LINC00428
42.6
42.8
43
43.2
43.4
Position on chr13 (Mb)

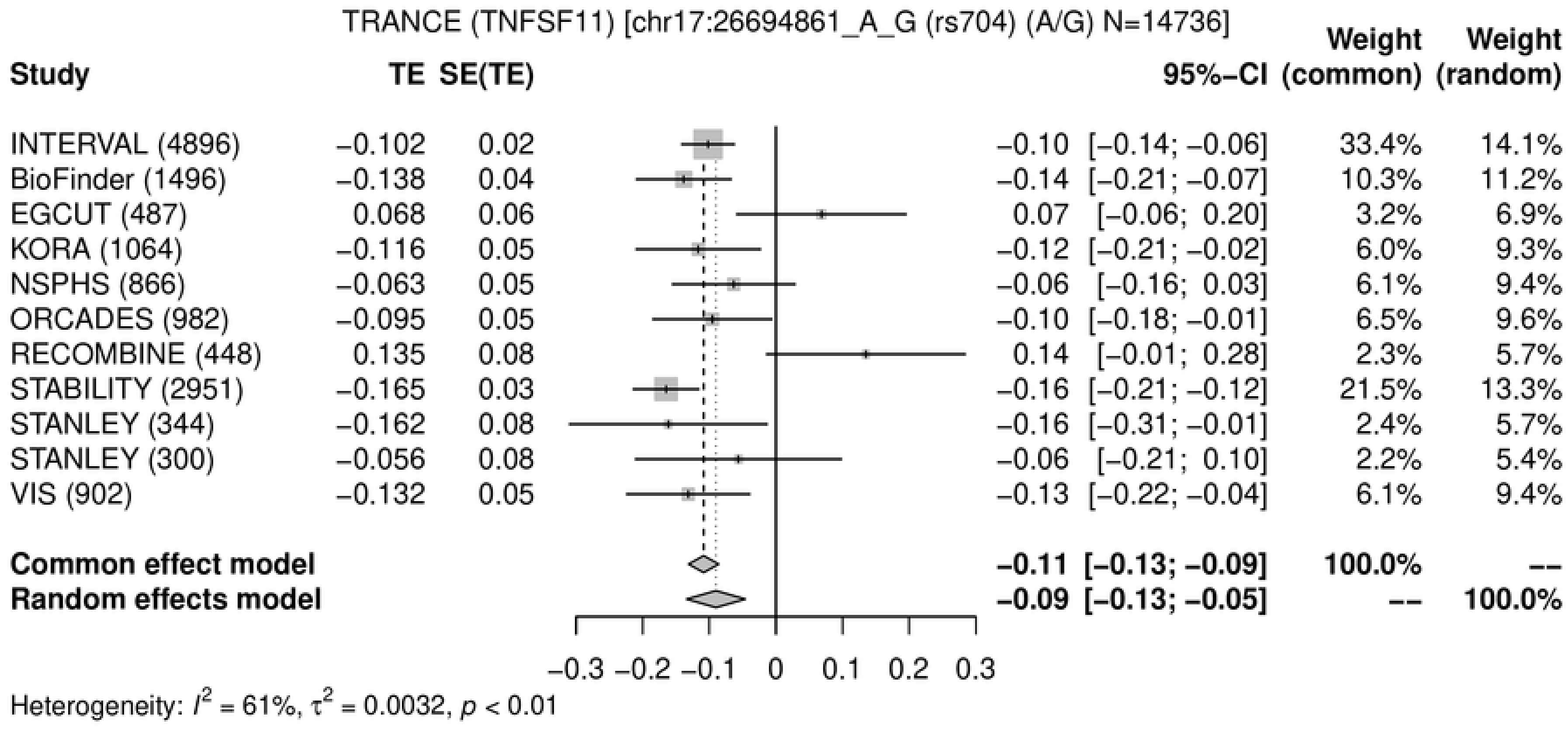
TRANCE (TNFSF11) [chr17:26694861_A_G (rs704) (A/G) N=14736]
Study
TE
SE(TE)
95%-CI
Weight (common)
Weight (random)
INTERVAL (4896) −0.102 0.02 −0.10 [−0.14; −0.06] 33.4% 14.1%
BioFinder (1496) −0.138 0.04 −0.14 [−0.21; −0.07] 10.3% 11.2%
EGCUT (487) 0.068 0.06 0.07 [−0.06; 0.20] 3.2% 6.9%
KORA (1064) −0.116 0.05 −0.12 [−0.21; −0.02] 6.0% 9.3%
NSPHS (866) −0.063 0.05 −0.06 [−0.16; 0.03] 6.1% 9.4%
ORCADES (982) −0.095 0.05 −0.10 [−0.18; −0.01] 6.5% 9.6%
RECOMBINE (448) 0.135 0.08 0.14 [−0.01; 0.28] 2.3% 5.7%
STABILITY (2951) −0.165 0.03 −0.16 [−0.21; −0.12] 21.5% 13.3%
STANLEY (344) −0.162 0.08 −0.16 [−0.31; −0.01] 2.4% 5.7%
STANLEY (300) −0.056 0.08 −0.06 [−0.21; 0.10] 2.2% 5.4%
VIS (902) −0.132 0.05 −0.13 [−0.22; −0.04] 6.1% 9.4%
Common effect model −0.11 [−0.13; −0.09] 100.0% --
Random effects model −0.09 [−0.13; −0.05] -- 100.0%
−0.3 −0.2 −0.1 0 0.1 0.2 0.3
Heterogeneity: $I^2$ = 61%, $\tau^2$ = 0.0032, $p$ < 0.01

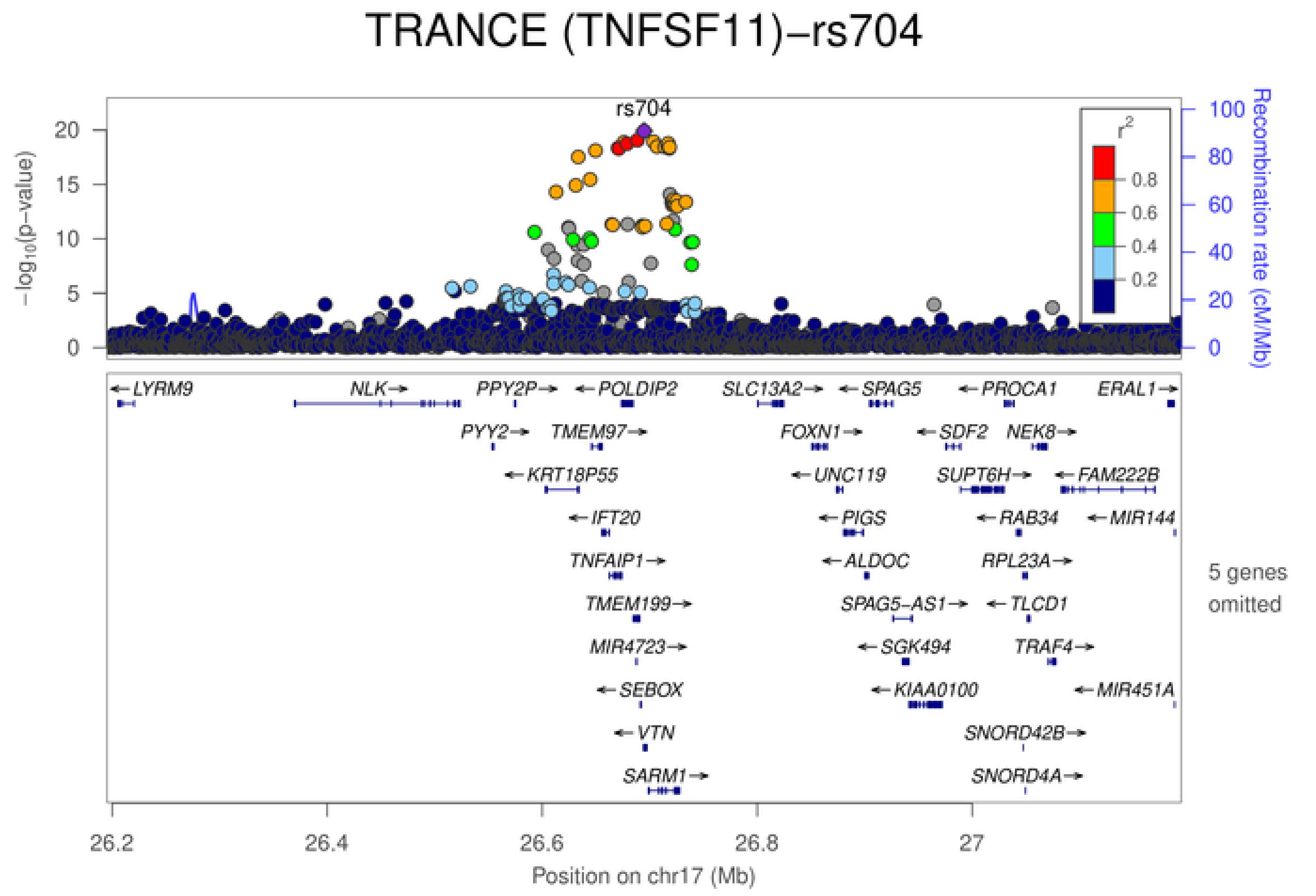
TRANCE (TNFSF11)−rs704
rs704
$-\log_{10}$(p−value)
Recombination rate (cM/Mb)
$r^2$
0.8
0.6
0.4
0.2
LYRM9
NLK
PPY2P
POLDIP2
SLC13A2
SPAG5
PROCA1
ERAL1
PYY2
TMEM97
FOXN1
SDF2
NEK8
KRT18P55
UNC119
SUPT6H
FAM222B
IFT20
PIGS
RAB34
MIR144
TNFAIP1
ALDOC
RPL23A
TMEM199
SPAG5−AS1
TLCD1
MIR4723
SGK494
TRAF4
SEBOX
KIAA0100
MIR451A
VTN
SNORD42B
SARM1
SNORD4A
5 genes omitted
26.2 26.4 26.6 26.8 27
Position on chr17 (Mb)

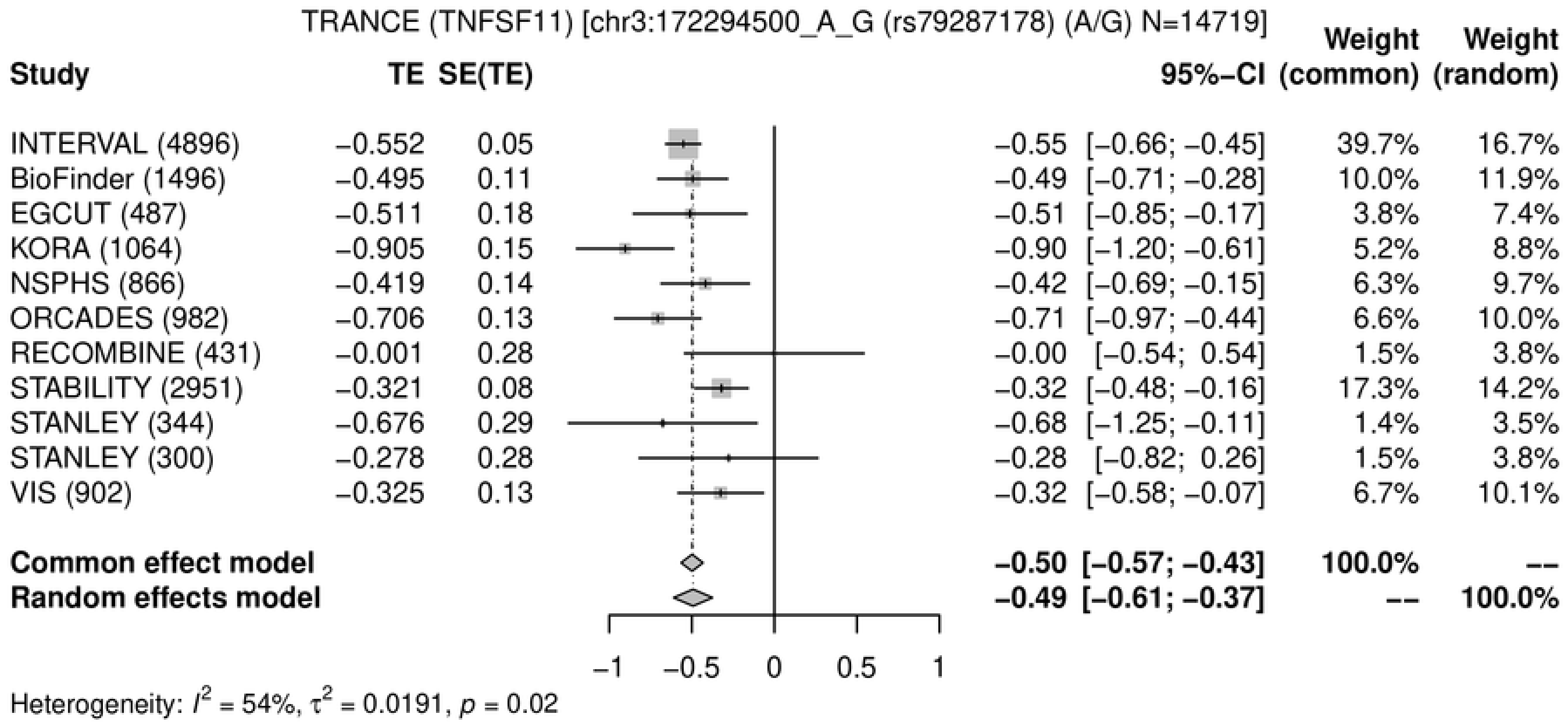
TRANCE (TNFSF11) [chr3:172294500_A_G (rs79287178) (A/G) N=14719]
Study
TE
SE(TE)
95%−CI
Weight (common)
Weight (random)
INTERVAL (4896) −0.552 0.05 −0.55 [−0.66; −0.45] 39.7% 16.7%
BioFinder (1496) −0.495 0.11 −0.49 [−0.71; −0.28] 10.0% 11.9%
EGCUT (487) −0.511 0.18 −0.51 [−0.85; −0.17] 3.8% 7.4%
KORA (1064) −0.905 0.15 −0.90 [−1.20; −0.61] 5.2% 8.8%
NSPHS (866) −0.419 0.14 −0.42 [−0.69; −0.15] 6.3% 9.7%
ORCADES (982) −0.706 0.13 −0.71 [−0.97; −0.44] 6.6% 10.0%
RECOMBINE (431) −0.001 0.28 −0.00 [−0.54; 0.54] 1.5% 3.8%
STABILITY (2951) −0.321 0.08 −0.32 [−0.48; −0.16] 17.3% 14.2%
STANLEY (344) −0.676 0.29 −0.68 [−1.25; −0.11] 1.4% 3.5%
STANLEY (300) −0.278 0.28 −0.28 [−0.82; 0.26] 1.5% 3.8%
VIS (902) −0.325 0.13 −0.32 [−0.58; −0.07] 6.7% 10.1%
Common effect model −0.50 [−0.57; −0.43] 100.0% --
Random effects model −0.49 [−0.61; −0.37] -- 100.0%
−1 −0.5 0 0.5 1
Heterogeneity: I² = 54%, τ² = 0.0191, p = 0.02

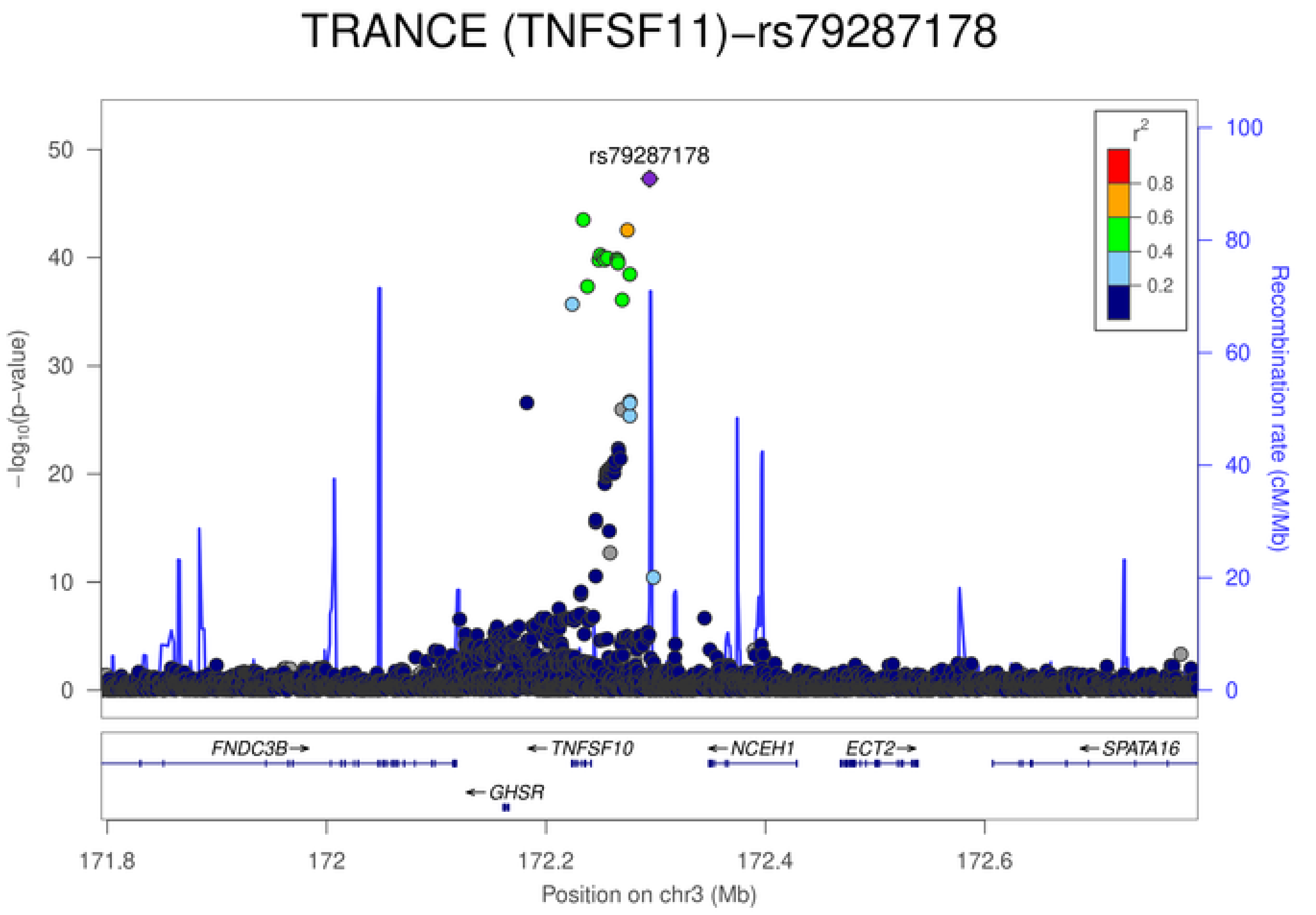
TRANCE (TNFSF11)−rs79287178
rs79287178
−log10(p−value)
Recombination rate (cM/Mb)
r²
0.8
0.6
0.4
0.2
FNDC3B
TNFSF10
NCEH1
ECT2
SPATA16
GHSR
171.8
172
172.2
172.4
172.6
Position on chr3 (Mb)

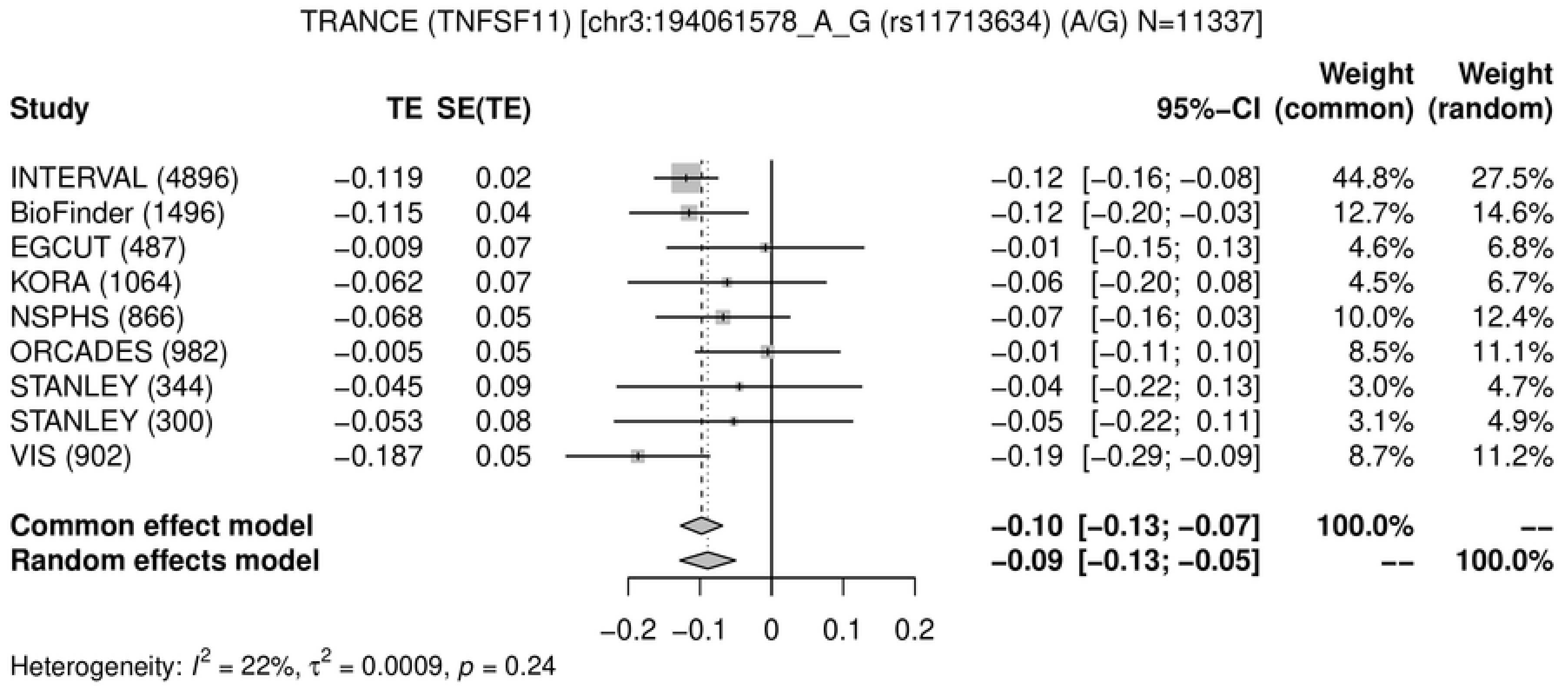
TRANCE (TNFSF11) [chr3:194061578_A_G (rs11713634) (A/G) N=11337]
Study
TE
SE(TE)
95%-CI
Weight (common)
Weight (random)
INTERVAL (4896) −0.119 0.02 −0.12 [−0.16; −0.08] 44.8% 27.5%
BioFinder (1496) −0.115 0.04 −0.12 [−0.20; −0.03] 12.7% 14.6%
EGCUT (487) −0.009 0.07 −0.01 [−0.15; 0.13] 4.6% 6.8%
KORA (1064) −0.062 0.07 −0.06 [−0.20; 0.08] 4.5% 6.7%
NSPHS (866) −0.068 0.05 −0.07 [−0.16; 0.03] 10.0% 12.4%
ORCADES (982) −0.005 0.05 −0.01 [−0.11; 0.10] 8.5% 11.1%
STANLEY (344) −0.045 0.09 −0.04 [−0.22; 0.13] 3.0% 4.7%
STANLEY (300) −0.053 0.08 −0.05 [−0.22; 0.11] 3.1% 4.9%
VIS (902) −0.187 0.05 −0.19 [−0.29; −0.09] 8.7% 11.2%
Common effect model −0.10 [−0.13; −0.07] 100.0% --
Random effects model −0.09 [−0.13; −0.05] -- 100.0%
−0.2 −0.1 0 0.1 0.2
Heterogeneity: $I^2$ = 22%, $\tau^2$ = 0.0009, $p$ = 0.24

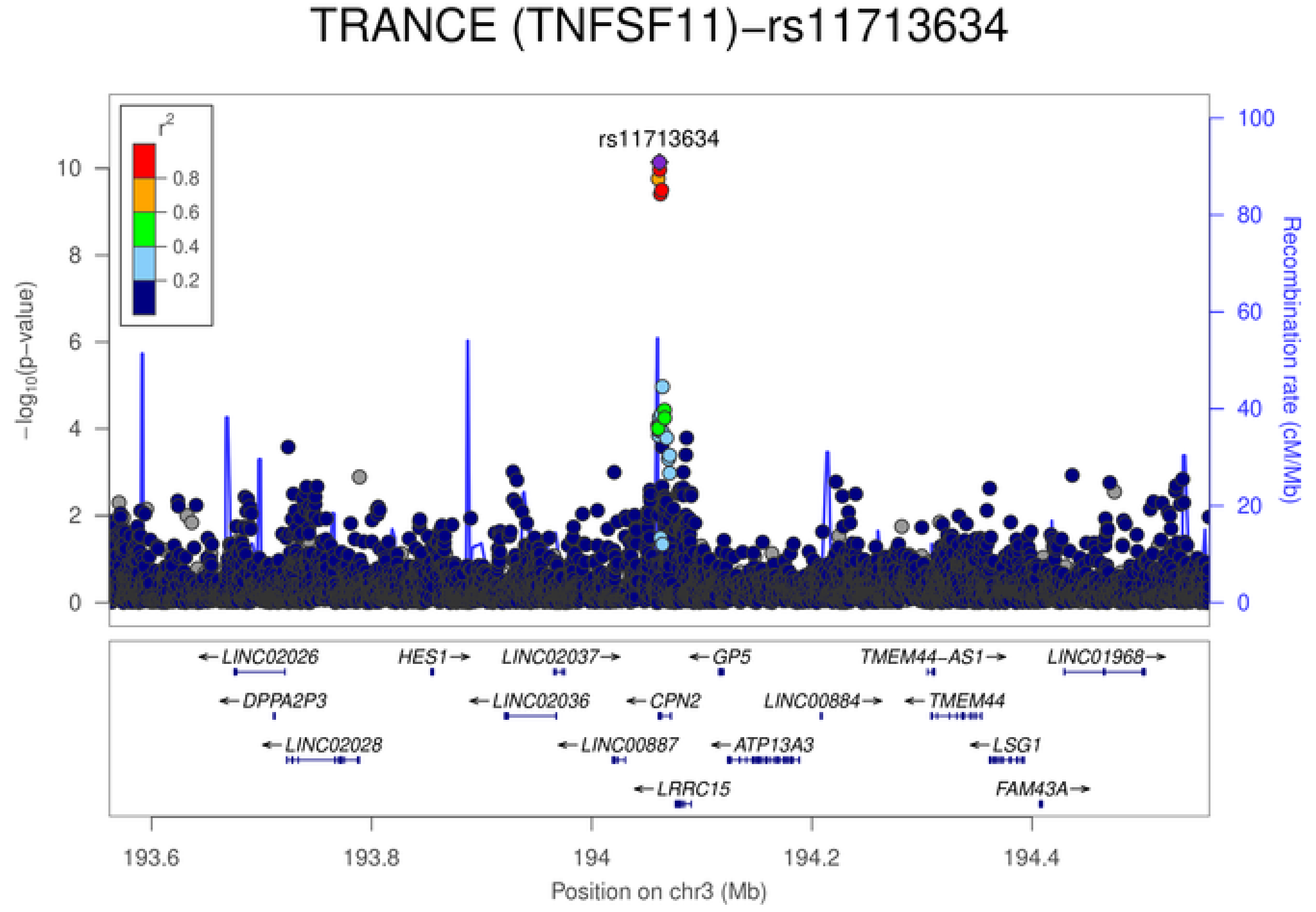
TRANCE (TNFSF11)−rs11713634
rs11713634
$r^2$
0.8
0.6
0.4
0.2
$-\log_{10}$(p−value)
Recombination rate (cM/Mb)
LINC02026
HES1
LINC02037
GP5
TMEM44-AS1
LINC01968
DPPA2P3
LINC02036
CPN2
LINC00884
TMEM44
LINC02028
LINC00887
ATP13A3
LSG1
LRRC15
FAM43A
193.6
193.8
194
194.2
194.4
Position on chr3 (Mb)

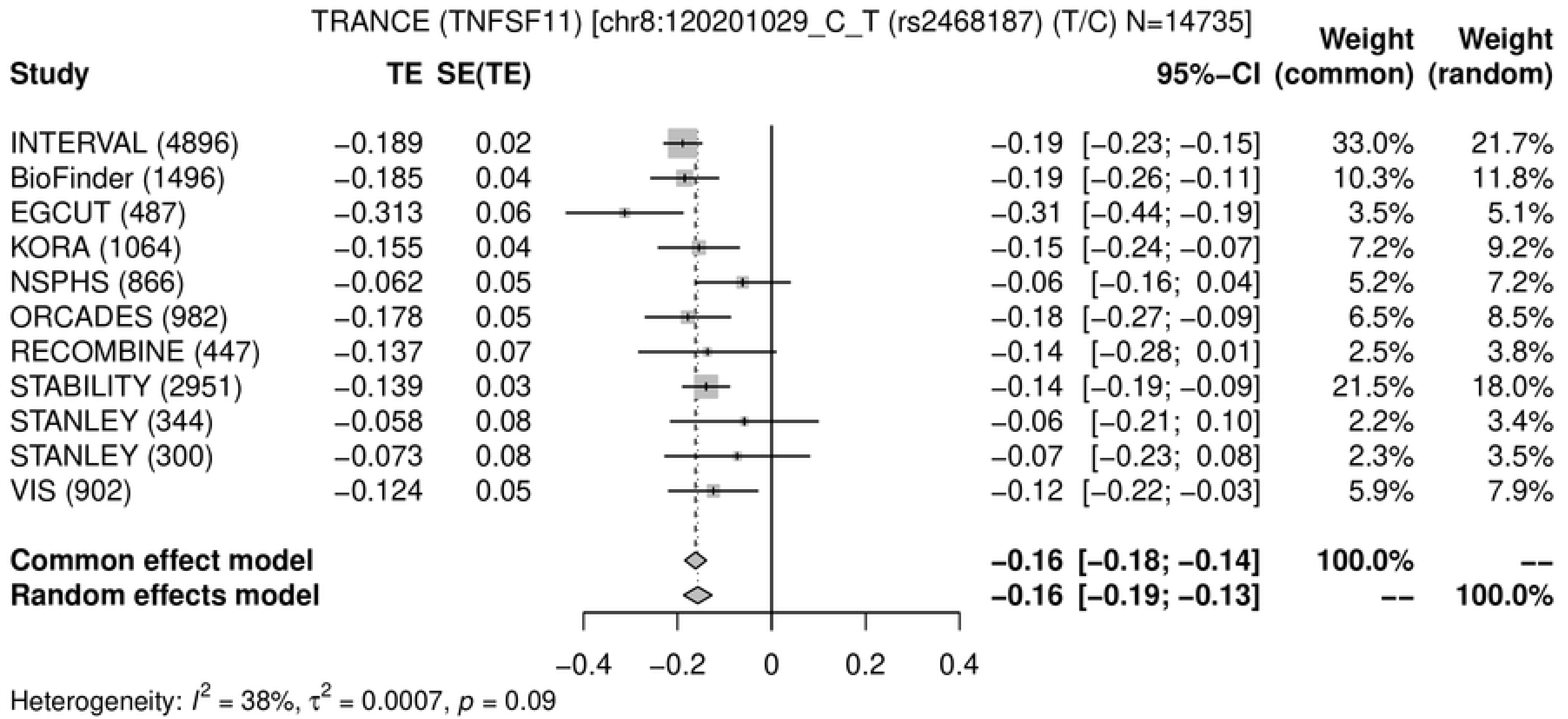
TRANCE (TNFSF11) [chr8:120201029_C_T (rs2468187) (T/C) N=14735]
Study
TE
SE(TE)
95%−CI
Weight (common)
Weight (random)
INTERVAL (4896) −0.189 0.02 −0.19 [−0.23; −0.15] 33.0% 21.7%
BioFinder (1496) −0.185 0.04 −0.19 [−0.26; −0.11] 10.3% 11.8%
EGCUT (487) −0.313 0.06 −0.31 [−0.44; −0.19] 3.5% 5.1%
KORA (1064) −0.155 0.04 −0.15 [−0.24; −0.07] 7.2% 9.2%
NSPHS (866) −0.062 0.05 −0.06 [−0.16; 0.04] 5.2% 7.2%
ORCADES (982) −0.178 0.05 −0.18 [−0.27; −0.09] 6.5% 8.5%
RECOMBINE (447) −0.137 0.07 −0.14 [−0.28; 0.01] 2.5% 3.8%
STABILITY (2951) −0.139 0.03 −0.14 [−0.19; −0.09] 21.5% 18.0%
STANLEY (344) −0.058 0.08 −0.06 [−0.21; 0.10] 2.2% 3.4%
STANLEY (300) −0.073 0.08 −0.07 [−0.23; 0.08] 2.3% 3.5%
VIS (902) −0.124 0.05 −0.12 [−0.22; −0.03] 5.9% 7.9%
Common effect model −0.16 [−0.18; −0.14] 100.0% --
Random effects model −0.16 [−0.19; −0.13] -- 100.0%
−0.4
−0.2
0
0.2
0.4
Heterogeneity: $I^2$ = 38%, $\tau^2$ = 0.0007, $p$ = 0.09

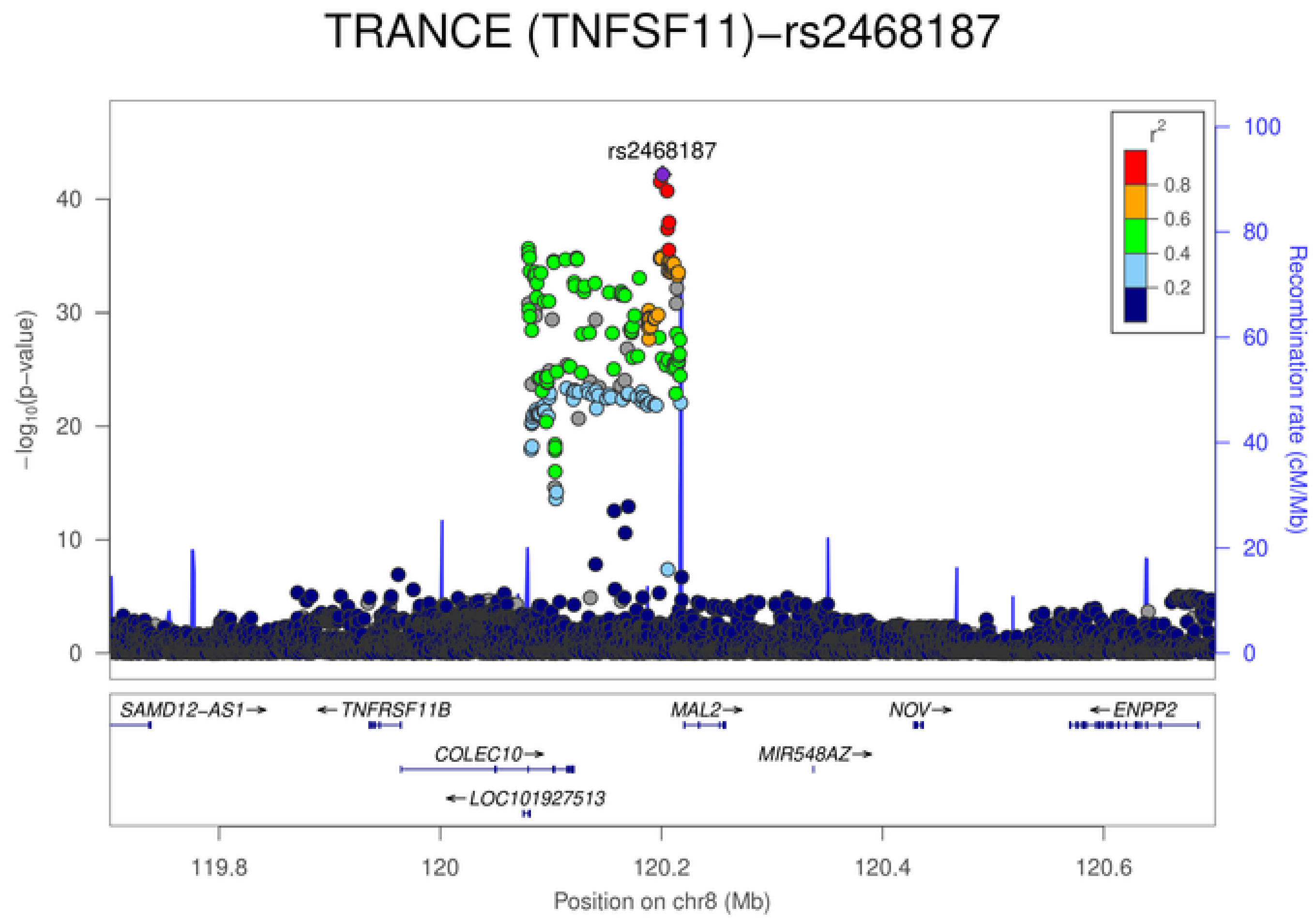
TRANCE (TNFSF11)−rs2468187
rs2468187
$-\log_{10}$(p−value)
Recombination rate (cM/Mb)
$r^2$
0.8
0.6
0.4
0.2
0
10
20
30
40
60
80
100
SAMD12−AS1→
←TNFRSF11B
COLEC10→
←LOC101927513
MAL2→
MIR548AZ→
NOV→
←ENPP2
119.8
120
120.2
120.4
120.6
Position on chr8 (Mb)

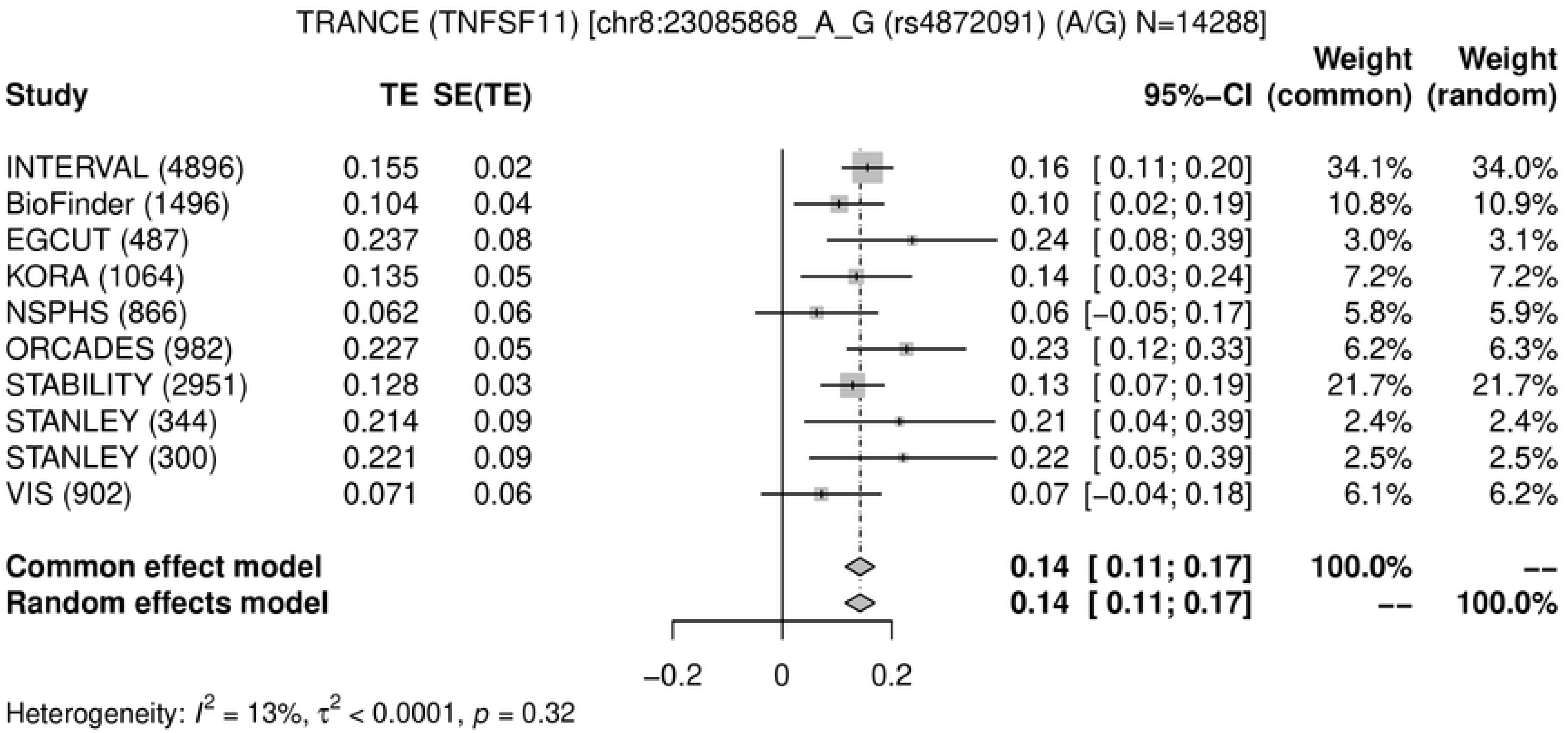
TRANCE (TNFSF11) [chr8:23085868_A_G (rs4872091) (A/G) N=14288]
Study
TE
SE(TE)
95%−CI
Weight (common)
Weight (random)
INTERVAL (4896) 0.155 0.02 0.16 [ 0.11; 0.20] 34.1% 34.0%
BioFinder (1496) 0.104 0.04 0.10 [ 0.02; 0.19] 10.8% 10.9%
EGCUT (487) 0.237 0.08 0.24 [ 0.08; 0.39] 3.0% 3.1%
KORA (1064) 0.135 0.05 0.14 [ 0.03; 0.24] 7.2% 7.2%
NSPHS (866) 0.062 0.06 0.06 [−0.05; 0.17] 5.8% 5.9%
ORCADES (982) 0.227 0.05 0.23 [ 0.12; 0.33] 6.2% 6.3%
STABILITY (2951) 0.128 0.03 0.13 [ 0.07; 0.19] 21.7% 21.7%
STANLEY (344) 0.214 0.09 0.21 [ 0.04; 0.39] 2.4% 2.4%
STANLEY (300) 0.221 0.09 0.22 [ 0.05; 0.39] 2.5% 2.5%
VIS (902) 0.071 0.06 0.07 [−0.04; 0.18] 6.1% 6.2%
Common effect model 0.14 [ 0.11; 0.17] 100.0% --
Random effects model 0.14 [ 0.11; 0.17] -- 100.0%
−0.2
0
0.2
Heterogeneity: $I^2$ = 13%, $\tau^2$ < 0.0001, $p$ = 0.32

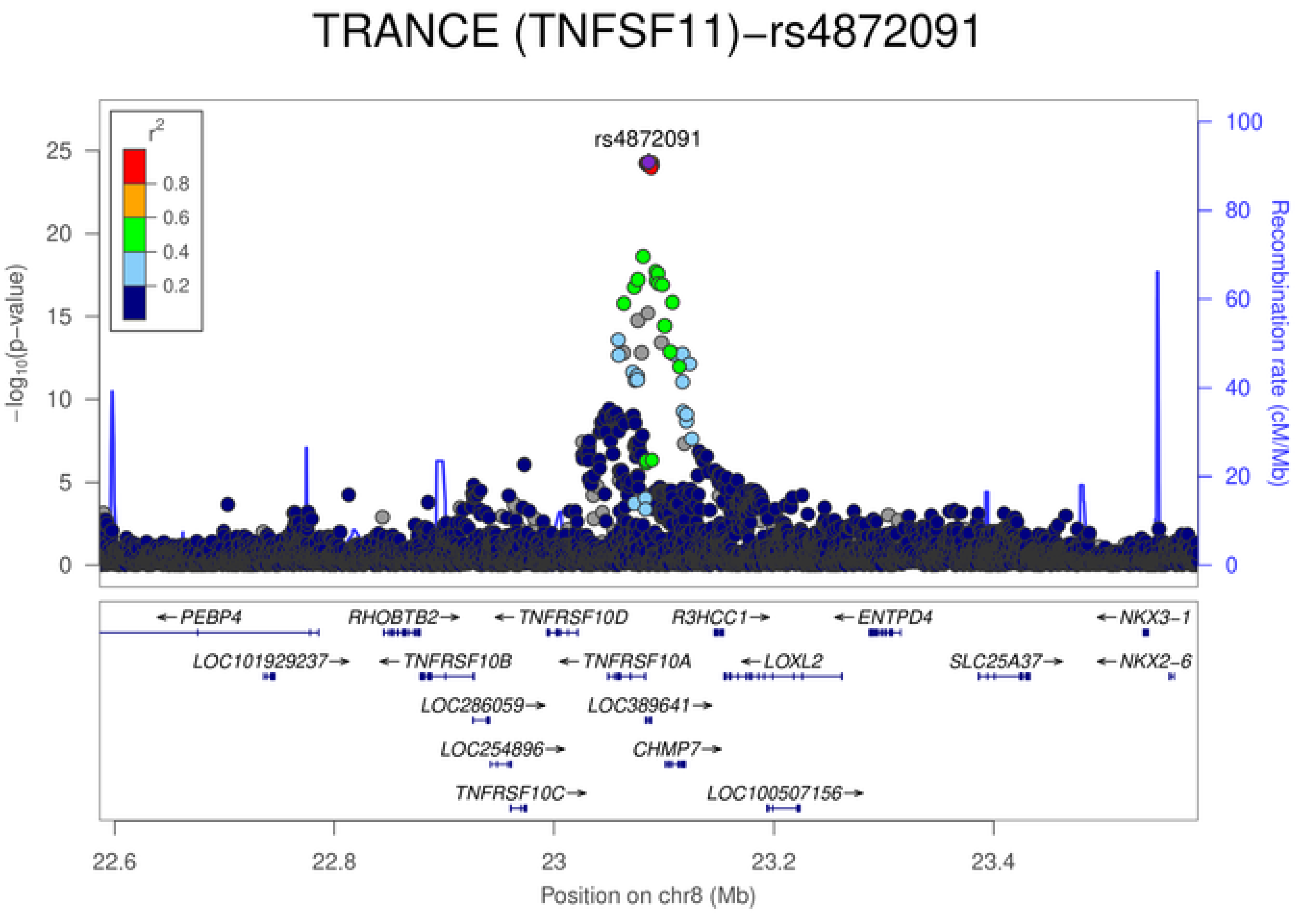
TRANCE (TNFSF11)−rs4872091
rs4872091
$r^2$
0.8
0.6
0.4
0.2
$-\log_{10}$(p−value)
Recombination rate (cM/Mb)
0
5
10
15
20
25
0
20
40
60
80
100
←PEBP4
RHOBTB2→
←TNFRSF10D
R3HCC1→
←ENTPD4
←NKX3−1
LOC101929237→
←TNFRSF10B
←TNFRSF10A
←LOXL2
SLC25A37→
←NKX2−6
LOC286059→
LOC389641→
LOC254896→
CHMP7→
TNFRSF10C→
LOC100507156→
22.6
22.8
23
23.2
23.4
Position on chr8 (Mb)

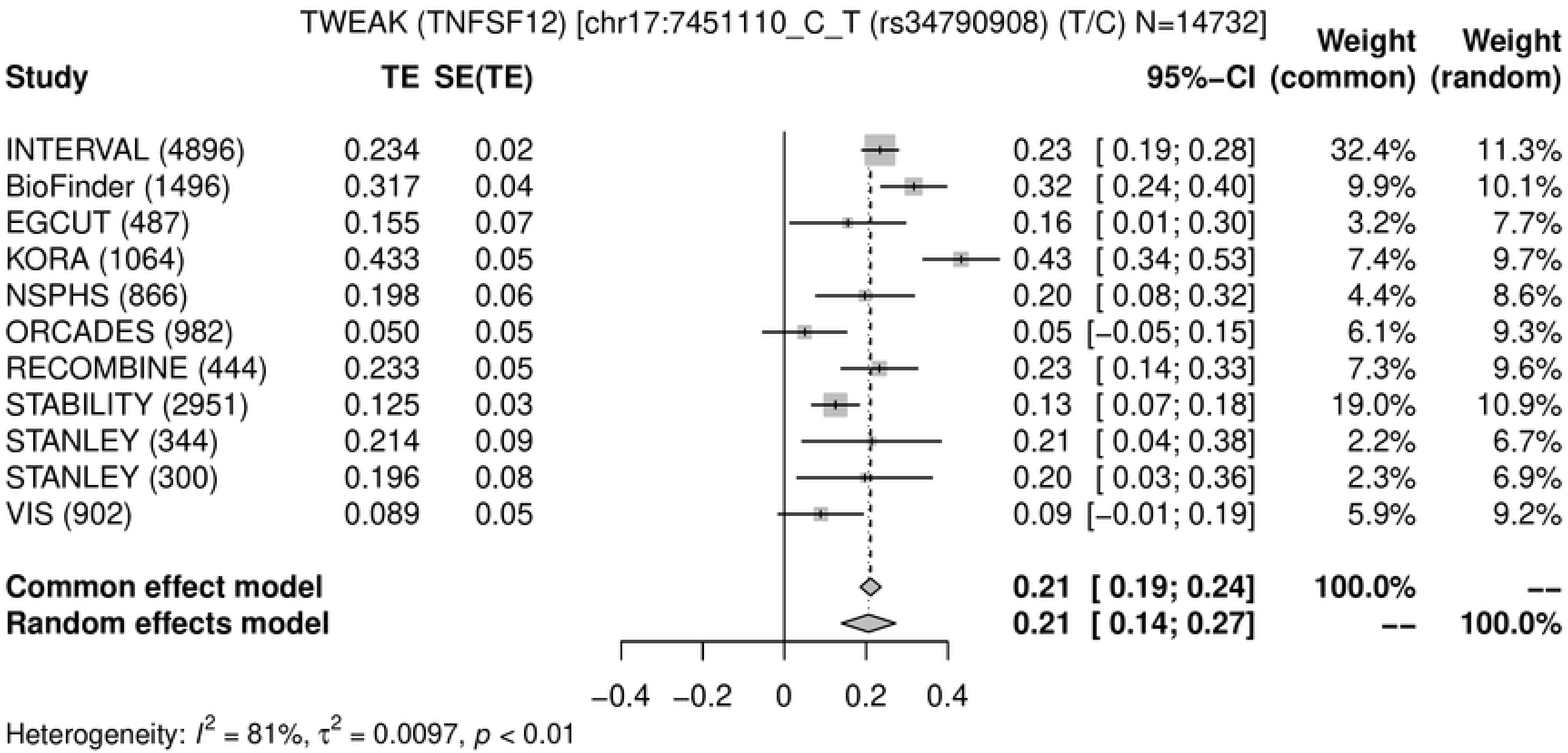
TWEAK (TNFSF12) [chr17:7451110_C_T (rs34790908) (T/C) N=14732]
Study
TE
SE(TE)
95%−CI
Weight (common)
Weight (random)
INTERVAL (4896) 0.234 0.02 0.23 [ 0.19; 0.28] 32.4% 11.3%
BioFinder (1496) 0.317 0.04 0.32 [ 0.24; 0.40] 9.9% 10.1%
EGCUT (487) 0.155 0.07 0.16 [ 0.01; 0.30] 3.2% 7.7%
KORA (1064) 0.433 0.05 0.43 [ 0.34; 0.53] 7.4% 9.7%
NSPHS (866) 0.198 0.06 0.20 [ 0.08; 0.32] 4.4% 8.6%
ORCADES (982) 0.050 0.05 0.05 [−0.05; 0.15] 6.1% 9.3%
RECOMBINE (444) 0.233 0.05 0.23 [ 0.14; 0.33] 7.3% 9.6%
STABILITY (2951) 0.125 0.03 0.13 [ 0.07; 0.18] 19.0% 10.9%
STANLEY (344) 0.214 0.09 0.21 [ 0.04; 0.38] 2.2% 6.7%
STANLEY (300) 0.196 0.08 0.20 [ 0.03; 0.36] 2.3% 6.9%
VIS (902) 0.089 0.05 0.09 [−0.01; 0.19] 5.9% 9.2%
Common effect model 0.21 [ 0.19; 0.24] 100.0% --
Random effects model 0.21 [ 0.14; 0.27] -- 100.0%
−0.4 −0.2 0 0.2 0.4
Heterogeneity: $I^2$ = 81%, $\tau^2$ = 0.0097, $p$ < 0.01

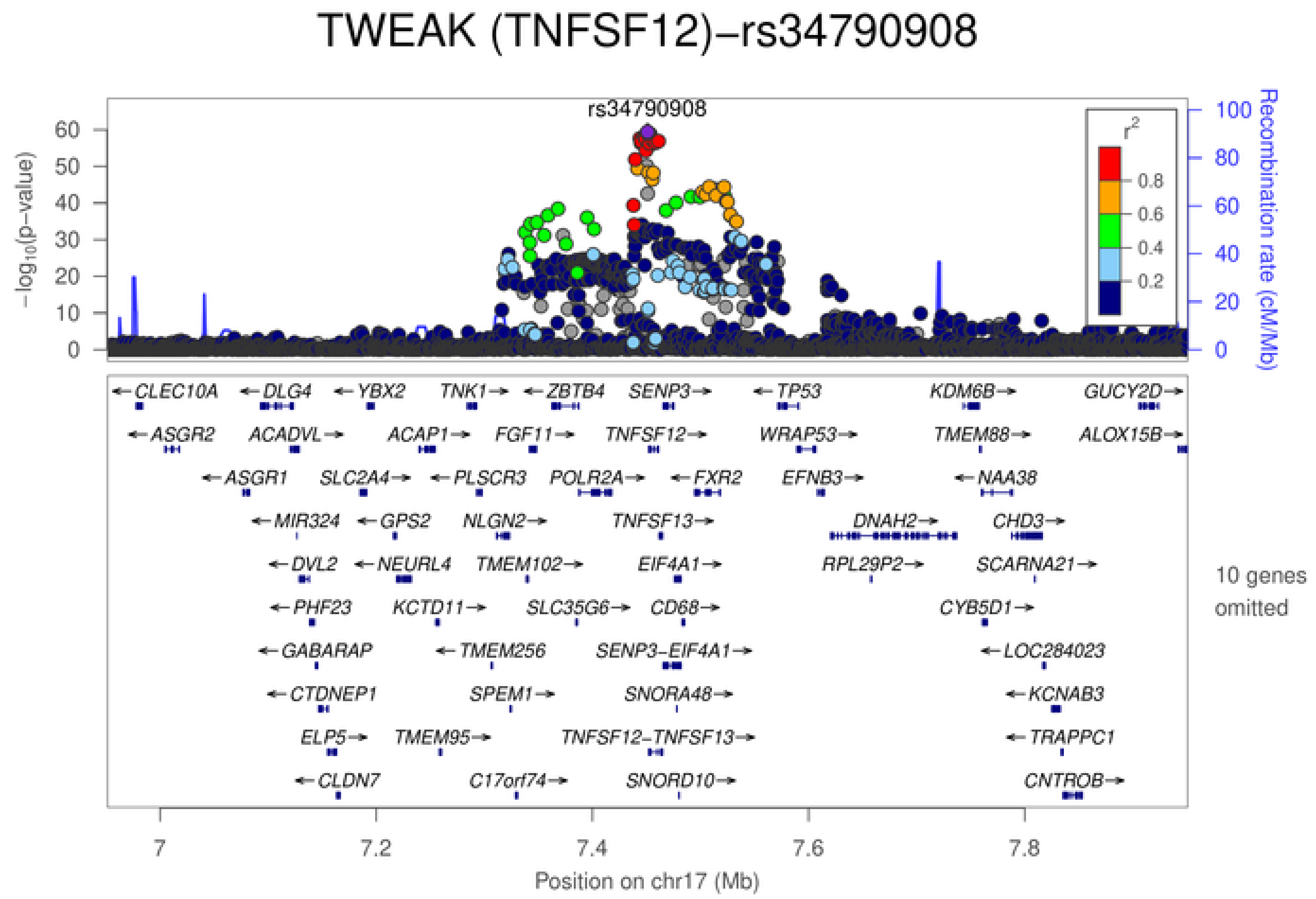
TWEAK (TNFSF12)−rs34790908
rs34790908
$-\log_{10}$(p−value)
Recombination rate (cM/Mb)
$r^2$
0.8
0.6
0.4
0.2
CLEC10A DLG4 YBX2 TNK1 ZBTB4 SENP3 TP53 KDM6B GUCY2D
ASGR2 ACADVL ACAP1 FGF11 TNFSF12 WRAP53 TMEM88 ALOX15B
ASGR1 SLC2A4 PLSCR3 POLR2A FXR2 EFNB3 NAA38
MIR324 GPS2 NLGN2 TNFSF13 DNAH2 CHD3
DVL2 NEURL4 TMEM102 EIF4A1 RPL29P2 SCARNA21
PHF23 KCTD11 SLC35G6 CD68 CYB5D1
GABARAP TMEM256 SENP3−EIF4A1 LOC284023
CTDNEP1 SPEM1 SNORA48 KCNAB3
ELP5 TMEM95 TNFSF12−TNFSF13 TRAPPC1
CLDN7 C17orf74 SNORD10 CNTROB
10 genes omitted
7 7.2 7.4 7.6 7.8
Position on chr17 (Mb)

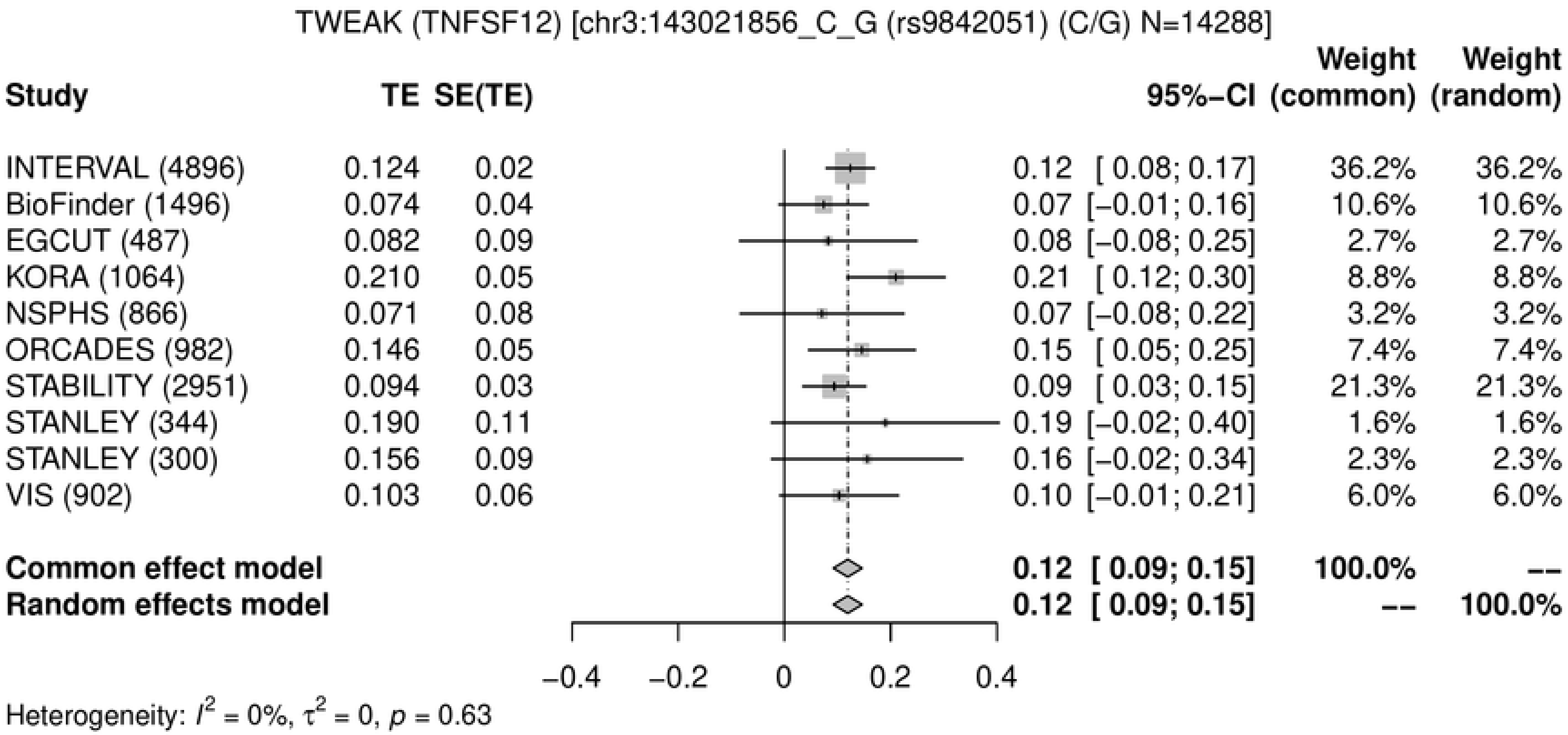
TWEAK (TNFSF12) [chr3:143021856_C_G (rs9842051) (C/G) N=14288]
Study
TE
SE(TE)
95%-CI
Weight (common)
Weight (random)
INTERVAL (4896) 0.124 0.02 0.12 [ 0.08; 0.17] 36.2% 36.2%
BioFinder (1496) 0.074 0.04 0.07 [−0.01; 0.16] 10.6% 10.6%
EGCUT (487) 0.082 0.09 0.08 [−0.08; 0.25] 2.7% 2.7%
KORA (1064) 0.210 0.05 0.21 [ 0.12; 0.30] 8.8% 8.8%
NSPHS (866) 0.071 0.08 0.07 [−0.08; 0.22] 3.2% 3.2%
ORCADES (982) 0.146 0.05 0.15 [ 0.05; 0.25] 7.4% 7.4%
STABILITY (2951) 0.094 0.03 0.09 [ 0.03; 0.15] 21.3% 21.3%
STANLEY (344) 0.190 0.11 0.19 [−0.02; 0.40] 1.6% 1.6%
STANLEY (300) 0.156 0.09 0.16 [−0.02; 0.34] 2.3% 2.3%
VIS (902) 0.103 0.06 0.10 [−0.01; 0.21] 6.0% 6.0%
Common effect model 0.12 [ 0.09; 0.15] 100.0% --
Random effects model 0.12 [ 0.09; 0.15] -- 100.0%
−0.4
−0.2
0
0.2
0.4
Heterogeneity: $I^2$ = 0%, $\tau^2$ = 0, $p$ = 0.63

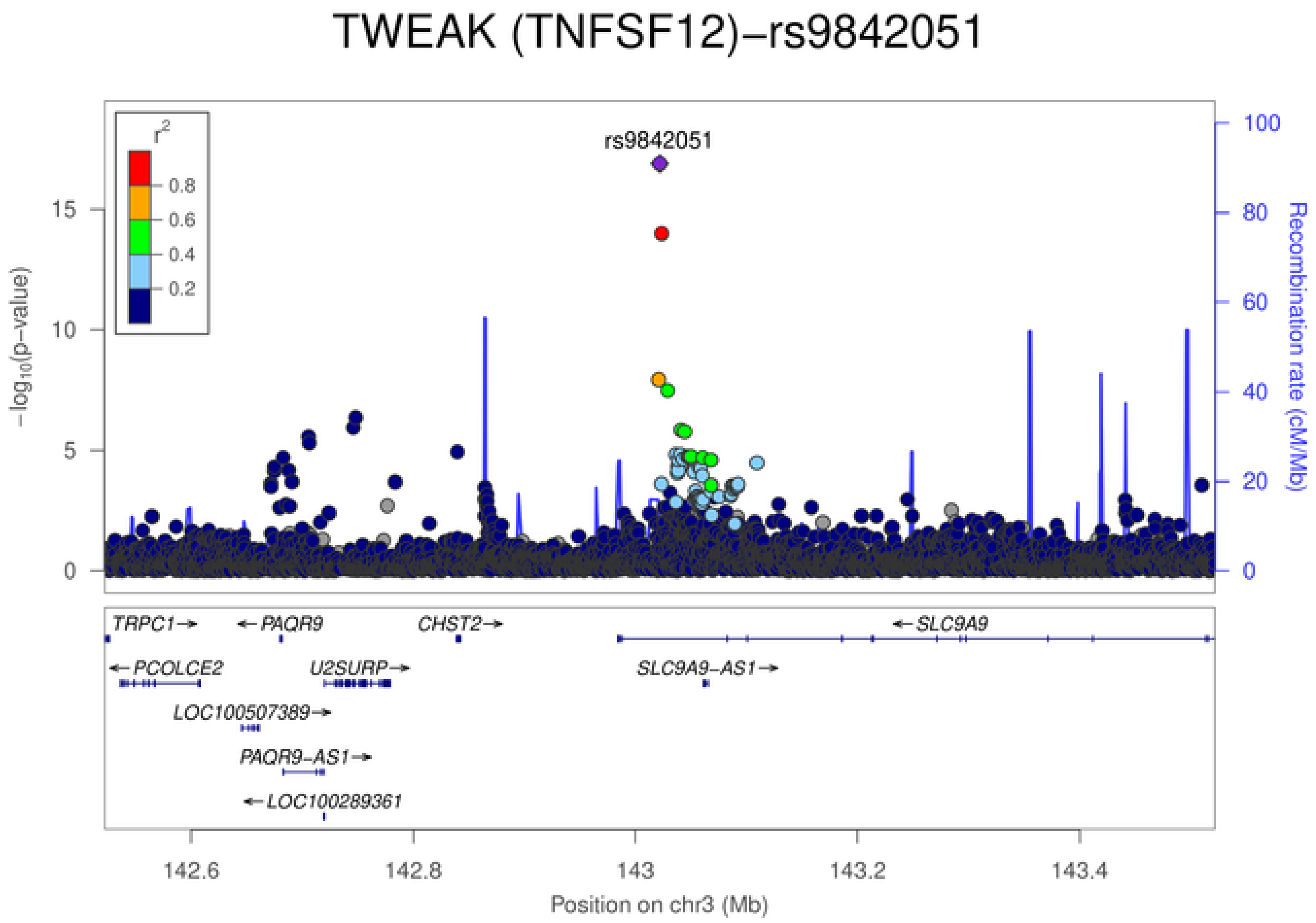
TWEAK (TNFSF12)−rs9842051
rs9842051
$r^2$
0.8
0.6
0.4
0.2
15
10
5
0
$-\log_{10}$(p-value)
100
80
60
40
20
0
Recombination rate (cM/Mb)
TRPC1→
←PAQR9
CHST2→
←SLC9A9
←PCOLCE2
U2SURP→
SLC9A9−AS1→
LOC100507389→
PAQR9−AS1→
←LOC100289361
142.6
142.8
143
143.2
143.4
Position on chr3 (Mb)

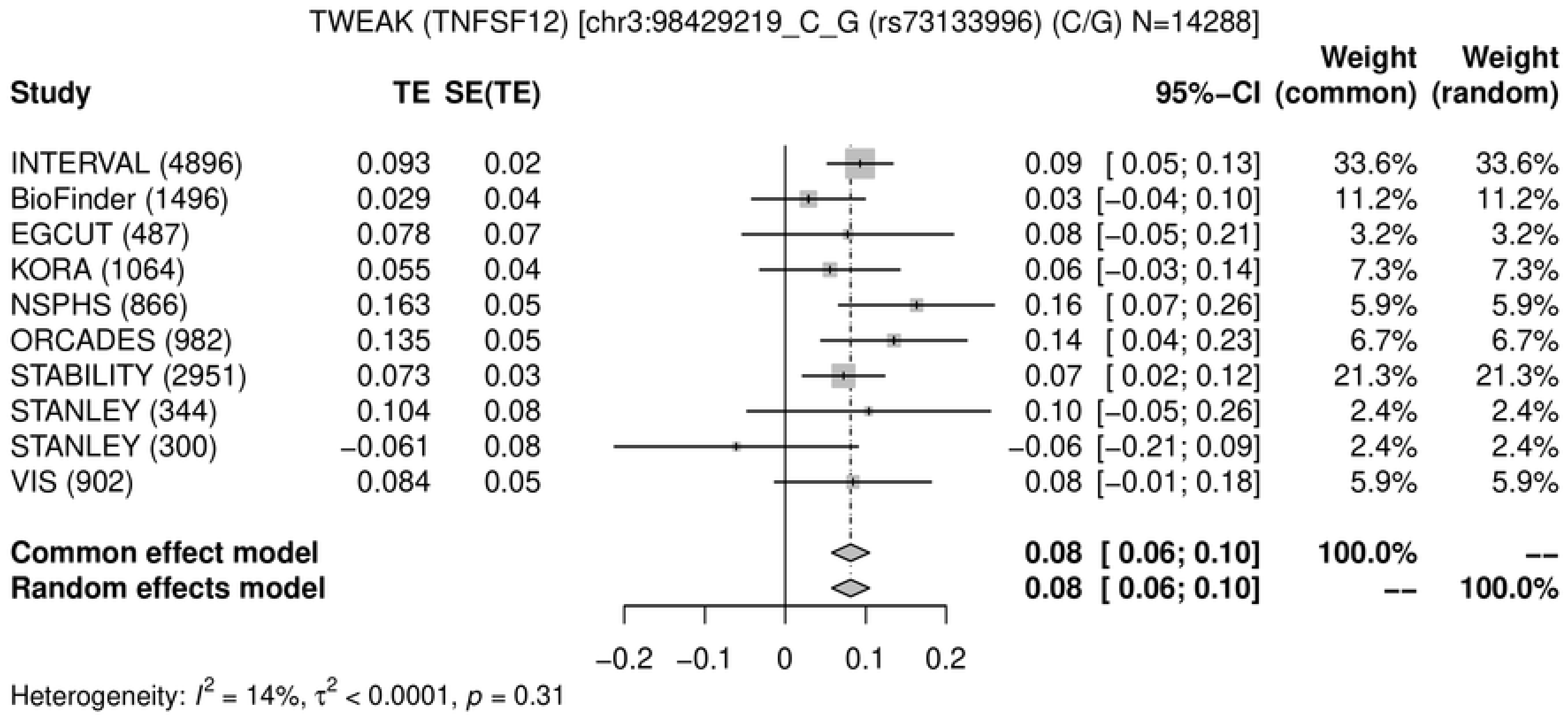
TWEAK (TNFSF12) [chr3:98429219_C_G (rs73133996) (C/G) N=14288]
Study TE SE(TE) 95%-CI Weight (common) Weight (random)
INTERVAL (4896) 0.093 0.02 0.09 [ 0.05; 0.13] 33.6% 33.6%
BioFinder (1496) 0.029 0.04 0.03 [−0.04; 0.10] 11.2% 11.2%
EGCUT (487) 0.078 0.07 0.08 [−0.05; 0.21] 3.2% 3.2%
KORA (1064) 0.055 0.04 0.06 [−0.03; 0.14] 7.3% 7.3%
NSPHS (866) 0.163 0.05 0.16 [ 0.07; 0.26] 5.9% 5.9%
ORCADES (982) 0.135 0.05 0.14 [ 0.04; 0.23] 6.7% 6.7%
STABILITY (2951) 0.073 0.03 0.07 [ 0.02; 0.12] 21.3% 21.3%
STANLEY (344) 0.104 0.08 0.10 [−0.05; 0.26] 2.4% 2.4%
STANLEY (300) −0.061 0.08 −0.06 [−0.21; 0.09] 2.4% 2.4%
VIS (902) 0.084 0.05 0.08 [−0.01; 0.18] 5.9% 5.9%
Common effect model 0.08 [ 0.06; 0.10] 100.0% --
Random effects model 0.08 [ 0.06; 0.10] -- 100.0%
−0.2 −0.1 0 0.1 0.2
Heterogeneity: $I^2$ = 14%, $\tau^2$ < 0.0001, $p$ = 0.31

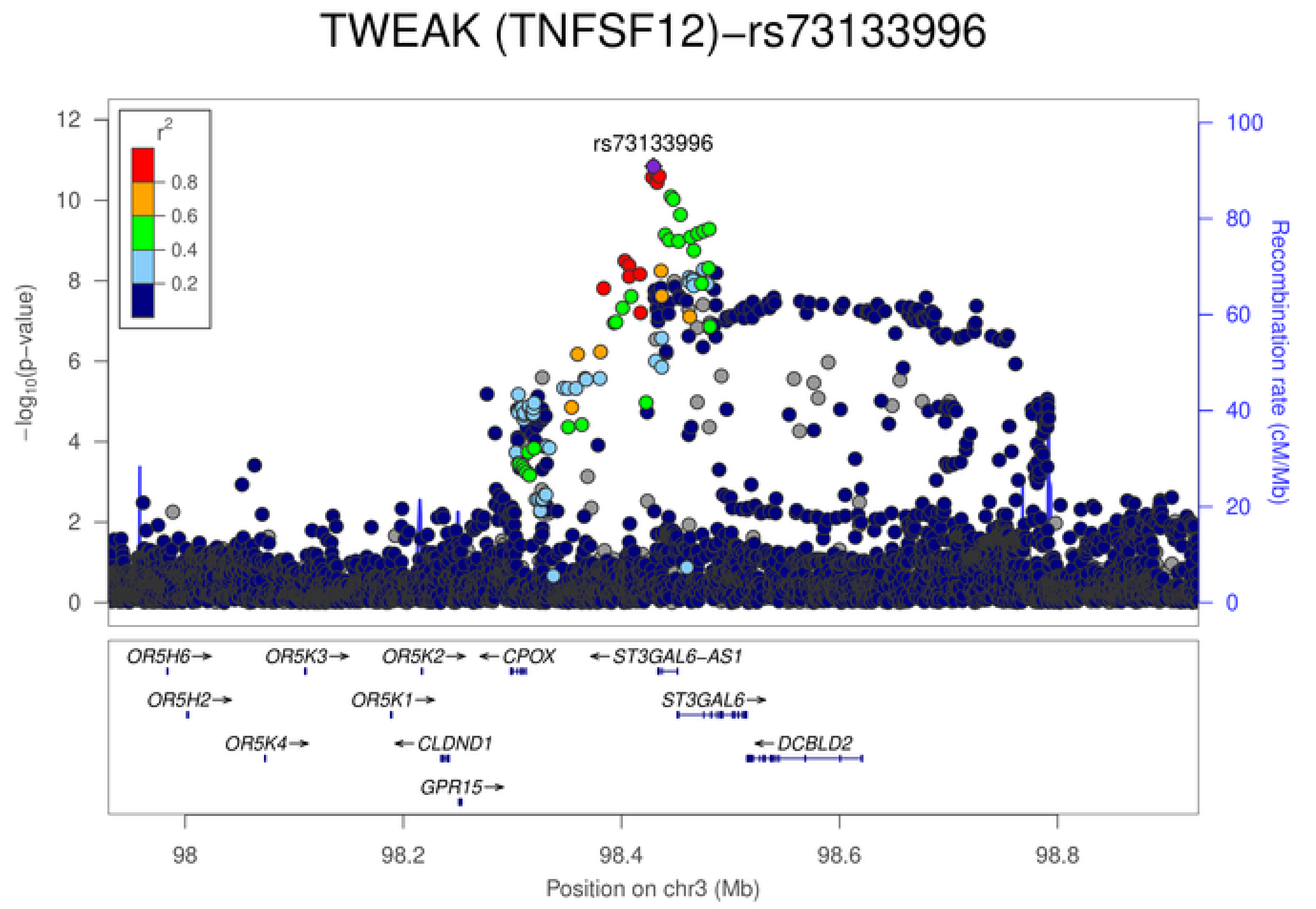
TWEAK (TNFSF12)−rs73133996
rs73133996
$r^2$
0.8
0.6
0.4
0.2
−log10(p−value)
Recombination rate (cM/Mb)
OR5H6→
OR5K3→
OR5K2→
←CPOX
←ST3GAL6−AS1
OR5H2→
OR5K1→
ST3GAL6→
OR5K4→
←CLDND1
←DCBLD2
GPR15→
98 98.2 98.4 98.6 98.8
Position on chr3 (Mb)

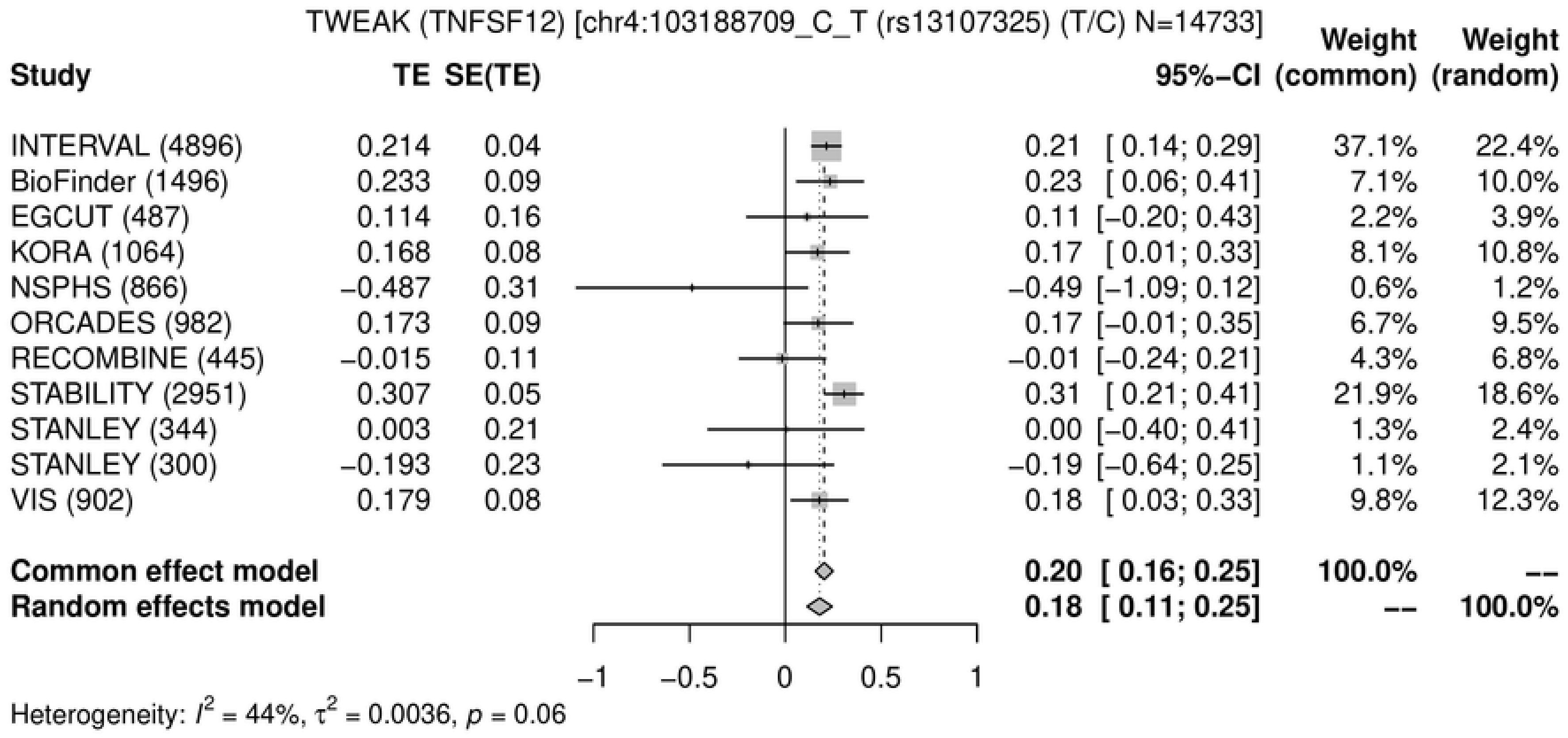
TWEAK (TNFSF12) [chr4:103188709_C_T (rs13107325) (T/C) N=14733]
Study
TE
SE(TE)
95%-CI
Weight (common)
Weight (random)
INTERVAL (4896) 0.214 0.04 0.21 [ 0.14; 0.29] 37.1% 22.4%
BioFinder (1496) 0.233 0.09 0.23 [ 0.06; 0.41] 7.1% 10.0%
EGCUT (487) 0.114 0.16 0.11 [−0.20; 0.43] 2.2% 3.9%
KORA (1064) 0.168 0.08 0.17 [ 0.01; 0.33] 8.1% 10.8%
NSPHS (866) −0.487 0.31 −0.49 [−1.09; 0.12] 0.6% 1.2%
ORCADES (982) 0.173 0.09 0.17 [−0.01; 0.35] 6.7% 9.5%
RECOMBINE (445) −0.015 0.11 −0.01 [−0.24; 0.21] 4.3% 6.8%
STABILITY (2951) 0.307 0.05 0.31 [ 0.21; 0.41] 21.9% 18.6%
STANLEY (344) 0.003 0.21 0.00 [−0.40; 0.41] 1.3% 2.4%
STANLEY (300) −0.193 0.23 −0.19 [−0.64; 0.25] 1.1% 2.1%
VIS (902) 0.179 0.08 0.18 [ 0.03; 0.33] 9.8% 12.3%
Common effect model 0.20 [ 0.16; 0.25] 100.0% --
Random effects model 0.18 [ 0.11; 0.25] -- 100.0%
−1 −0.5 0 0.5 1
Heterogeneity: $I^2$ = 44%, $\tau^2$ = 0.0036, $p$ = 0.06

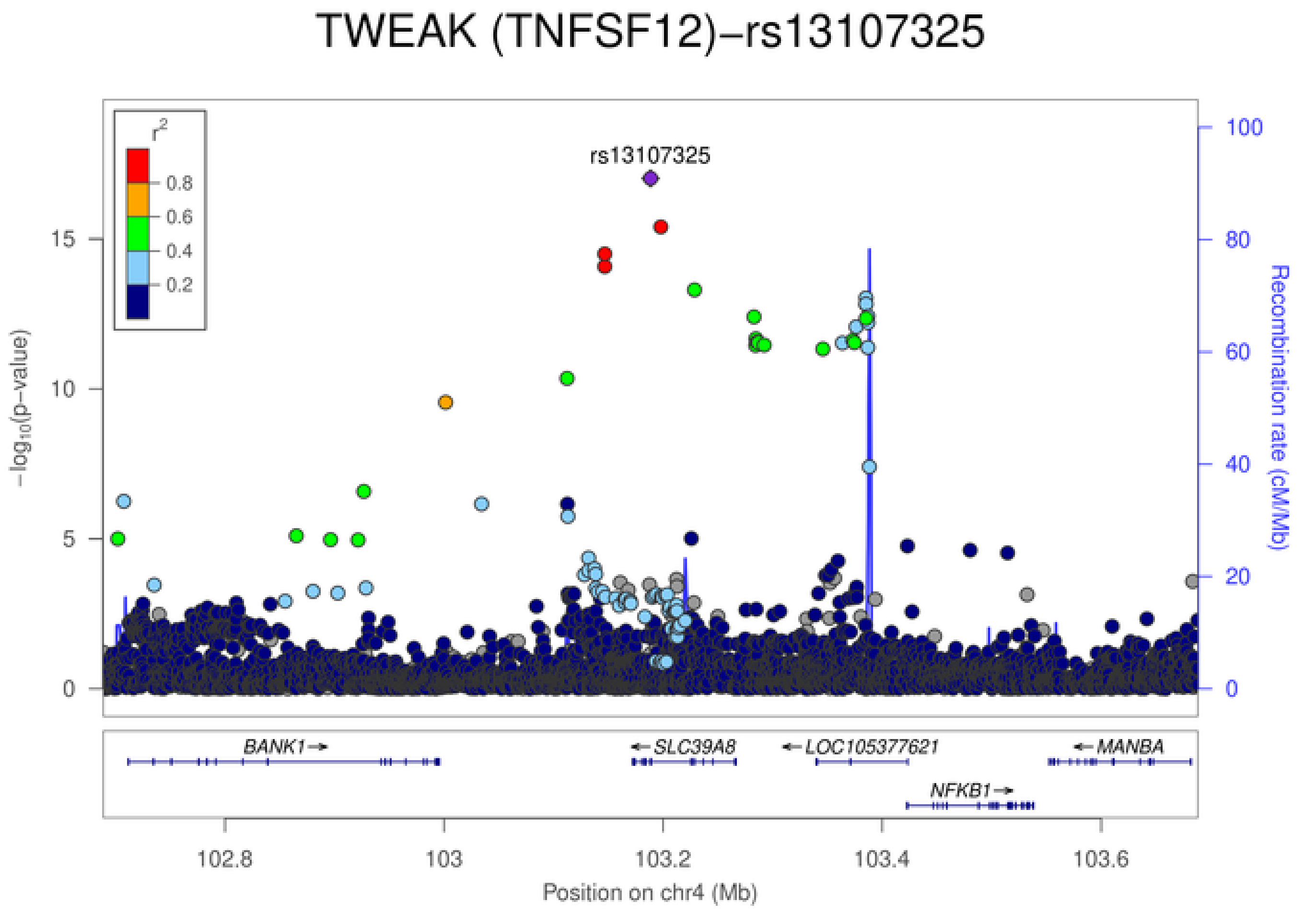
TWEAK (TNFSF12)−rs13107325
rs13107325
$r^2$
0.8
0.6
0.4
0.2
$-\log_{10}$(p−value)
Recombination rate (cM/Mb)
BANK1
SLC39A8
LOC105377621
MANBA
NFKB1
102.8
103
103.2
103.4
103.6
Position on chr4 (Mb)

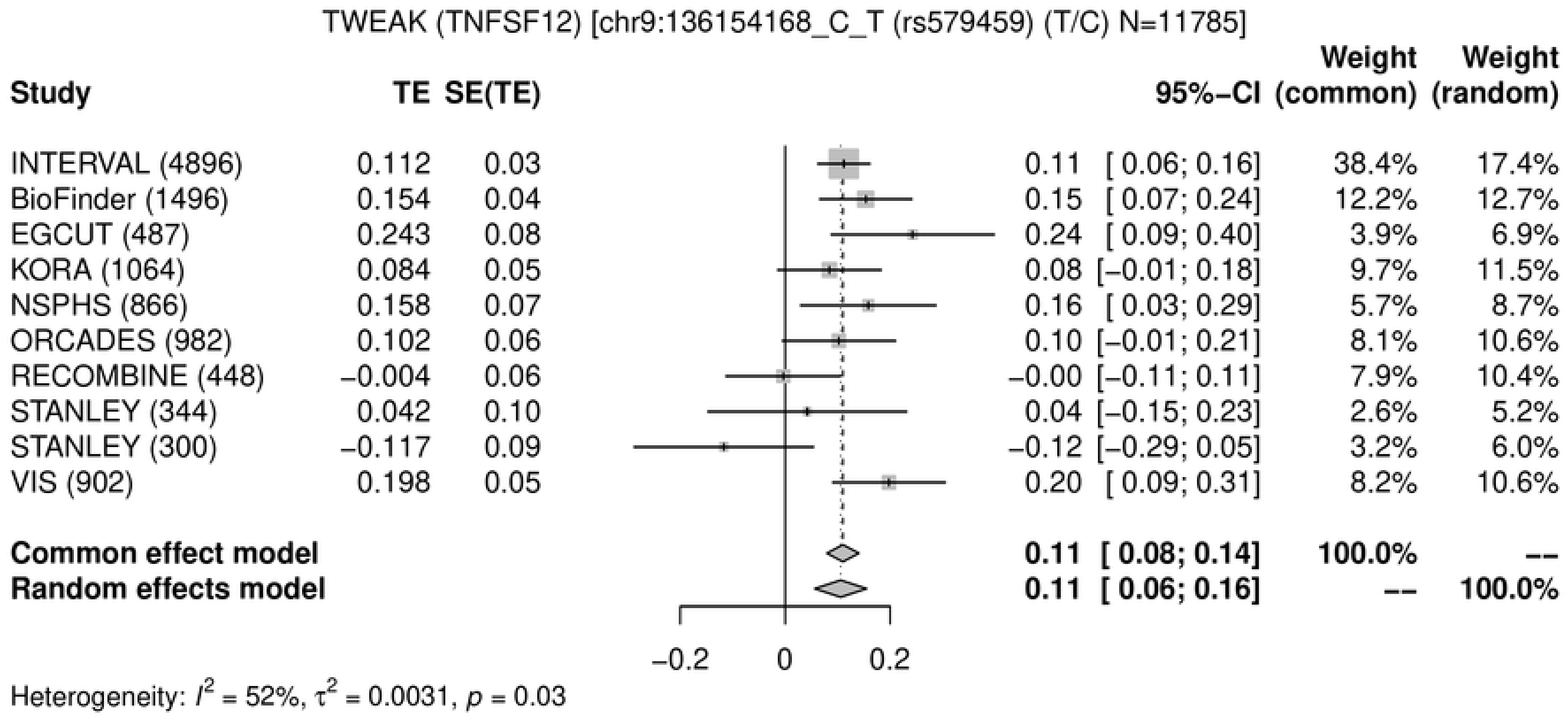
TWEAK (TNFSF12) [chr9:136154168_C_T (rs579459) (T/C) N=11785]
Study | TE | SE(TE) | 95%-CI | Weight (common) | Weight (random)
INTERVAL (4896) 0.112 0.03 0.11 [ 0.06; 0.16] 38.4% 17.4%
BioFinder (1496) 0.154 0.04 0.15 [ 0.07; 0.24] 12.2% 12.7%
EGCUT (487) 0.243 0.08 0.24 [ 0.09; 0.40] 3.9% 6.9%
KORA (1064) 0.084 0.05 0.08 [−0.01; 0.18] 9.7% 11.5%
NSPHS (866) 0.158 0.07 0.16 [ 0.03; 0.29] 5.7% 8.7%
ORCADES (982) 0.102 0.06 0.10 [−0.01; 0.21] 8.1% 10.6%
RECOMBINE (448) −0.004 0.06 −0.00 [−0.11; 0.11] 7.9% 10.4%
STANLEY (344) 0.042 0.10 0.04 [−0.15; 0.23] 2.6% 5.2%
STANLEY (300) −0.117 0.09 −0.12 [−0.29; 0.05] 3.2% 6.0%
VIS (902) 0.198 0.05 0.20 [ 0.09; 0.31] 8.2% 10.6%
Common effect model 0.11 [ 0.08; 0.14] 100.0% --
Random effects model 0.11 [ 0.06; 0.16] -- 100.0%
−0.2 0 0.2
Heterogeneity: $I^2$ = 52%, $\tau^2$ = 0.0031, $p$ = 0.03

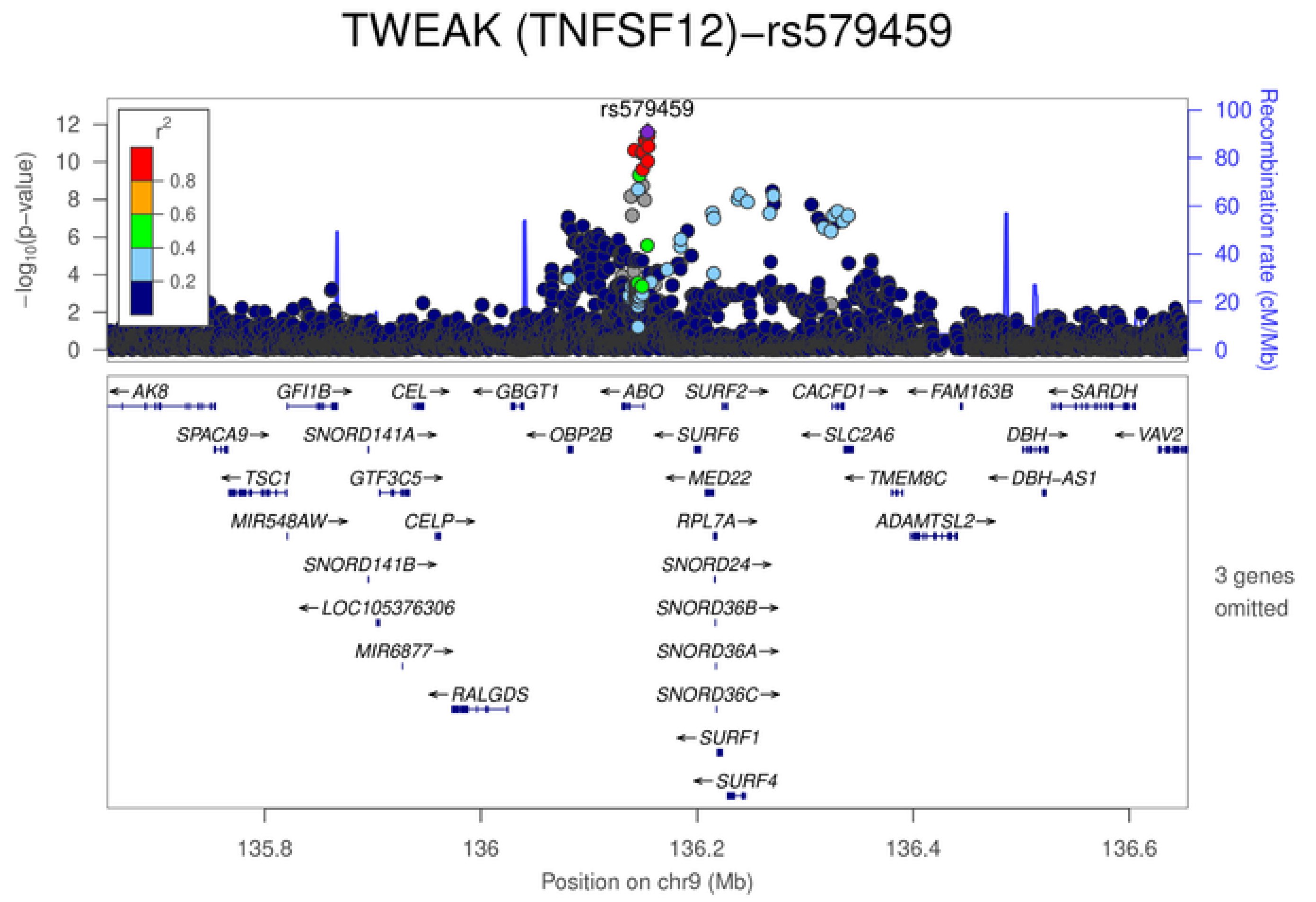
TWEAK (TNFSF12)−rs579459
rs579459
$-\log_{10}$(p−value)
Recombination rate (cM/Mb)
$r^2$
0.8
0.6
0.4
0.2
AK8
GFI1B
CEL
GBGT1
ABO
SURF2
CACFD1
FAM163B
SARDH
SPACA9
SNORD141A
OBP2B
SURF6
SLC2A6
DBH
VAV2
TSC1
GTF3C5
MED22
TMEM8C
DBH−AS1
MIR548AW
CELP
RPL7A
ADAMTSL2
SNORD141B
SNORD24
LOC105376306
SNORD36B
MIR6877
SNORD36A
RALGDS
SNORD36C
SURF1
SURF4
3 genes omitted
135.8
136
136.2
136.4
136.6
Position on chr9 (Mb)

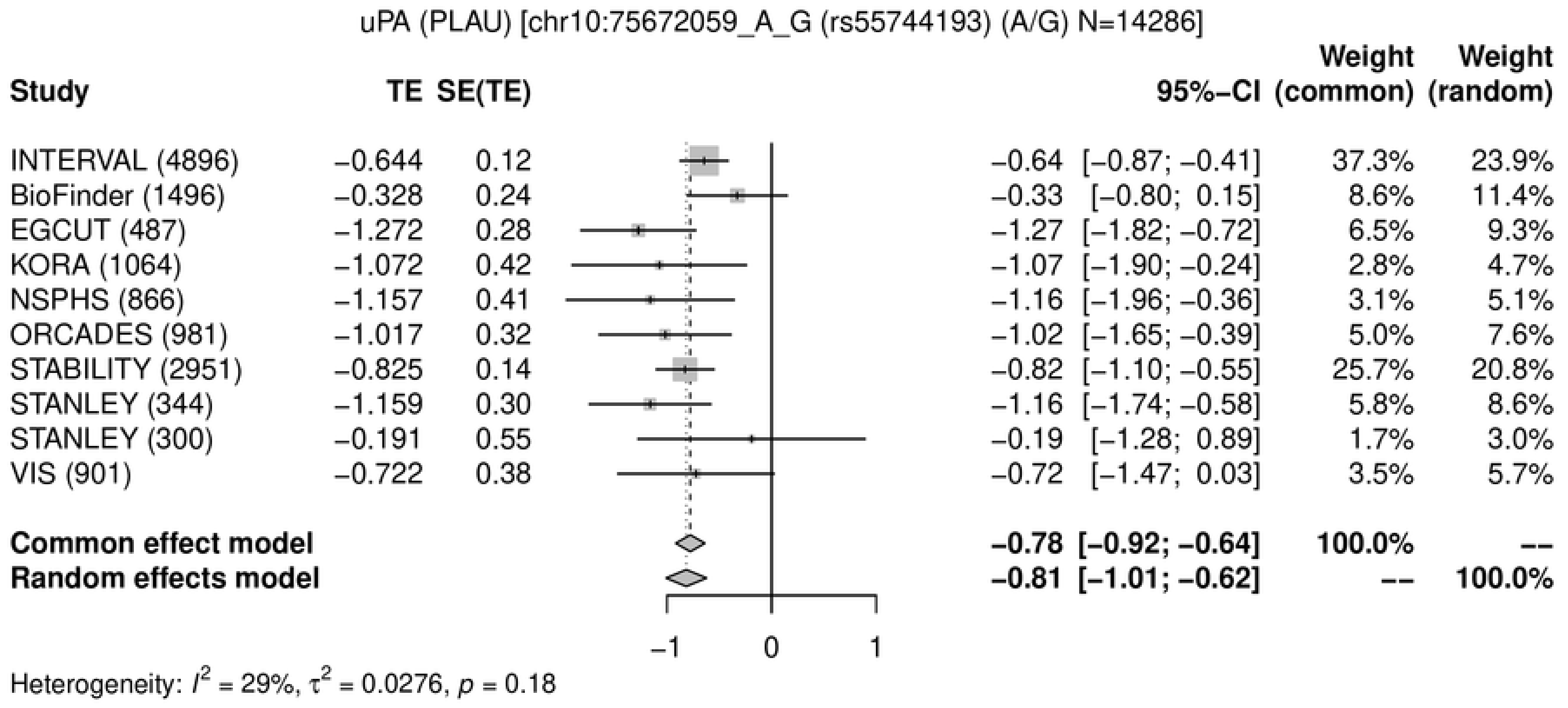
uPA (PLAU) [chr10:75672059_A_G (rs55744193) (A/G) N=14286]
Study TE SE(TE) 95%−CI Weight (common) Weight (random)
INTERVAL (4896) −0.644 0.12 −0.64 [−0.87; −0.41] 37.3% 23.9%
BioFinder (1496) −0.328 0.24 −0.33 [−0.80; 0.15] 8.6% 11.4%
EGCUT (487) −1.272 0.28 −1.27 [−1.82; −0.72] 6.5% 9.3%
KORA (1064) −1.072 0.42 −1.07 [−1.90; −0.24] 2.8% 4.7%
NSPHS (866) −1.157 0.41 −1.16 [−1.96; −0.36] 3.1% 5.1%
ORCADES (981) −1.017 0.32 −1.02 [−1.65; −0.39] 5.0% 7.6%
STABILITY (2951) −0.825 0.14 −0.82 [−1.10; −0.55] 25.7% 20.8%
STANLEY (344) −1.159 0.30 −1.16 [−1.74; −0.58] 5.8% 8.6%
STANLEY (300) −0.191 0.55 −0.19 [−1.28; 0.89] 1.7% 3.0%
VIS (901) −0.722 0.38 −0.72 [−1.47; 0.03] 3.5% 5.7%
Common effect model −0.78 [−0.92; −0.64] 100.0% --
Random effects model −0.81 [−1.01; −0.62] -- 100.0%
−1 0 1
Heterogeneity: $I^2$ = 29%, $\tau^2$ = 0.0276, $p$ = 0.18

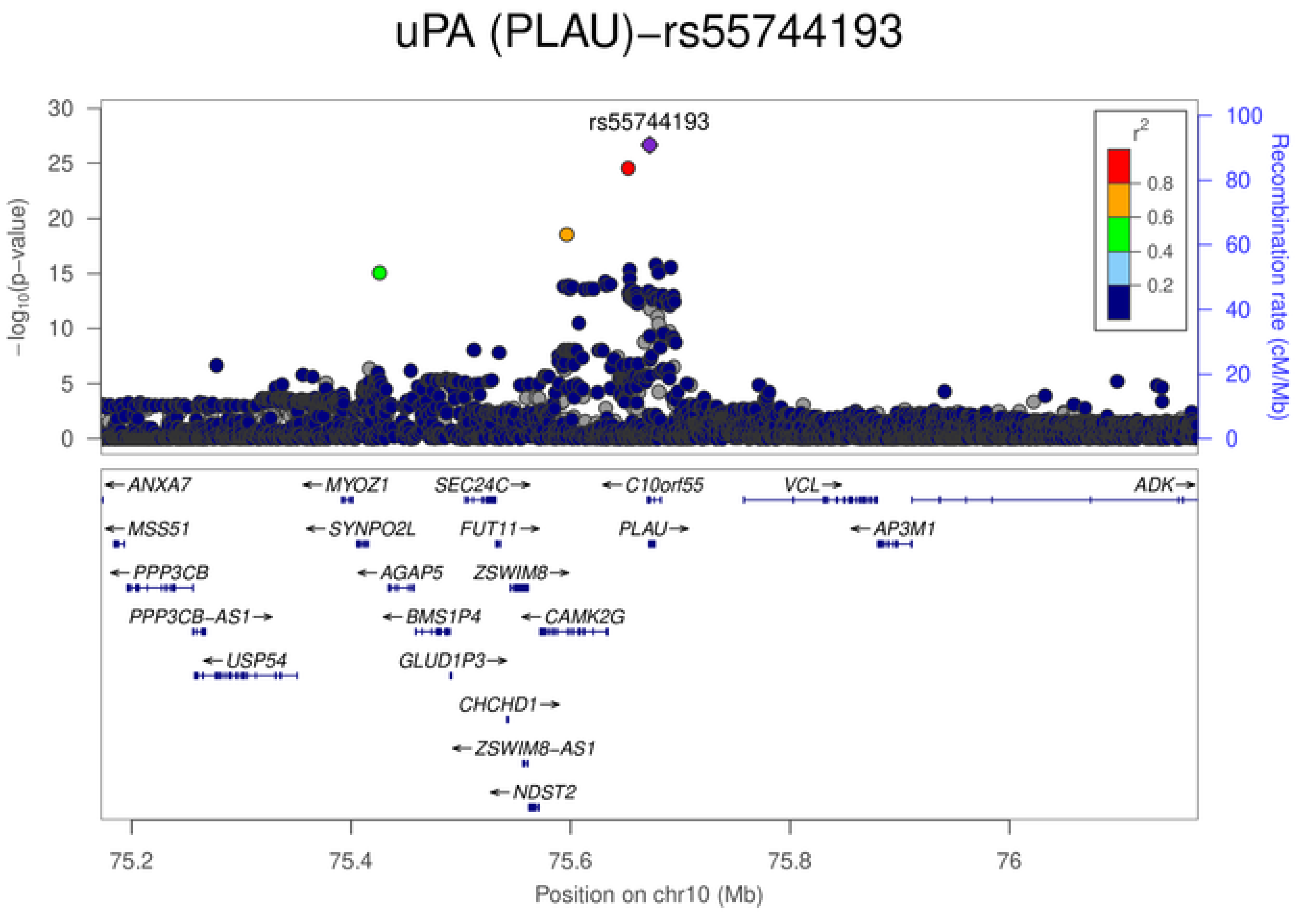
uPA (PLAU)−rs55744193
rs55744193
−log10(p−value)
Recombination rate (cM/Mb)
r2
0.8
0.6
0.4
0.2
ANXA7
MYOZ1
SEC24C
C10orf55
VCL
ADK
MSS51
SYNPO2L
FUT11
PLAU
AP3M1
PPP3CB
AGAP5
ZSWIM8
PPP3CB−AS1
BMS1P4
CAMK2G
USP54
GLUD1P3
CHCHD1
ZSWIM8−AS1
NDST2
75.2
75.4
75.6
75.8
76
Position on chr10 (Mb)

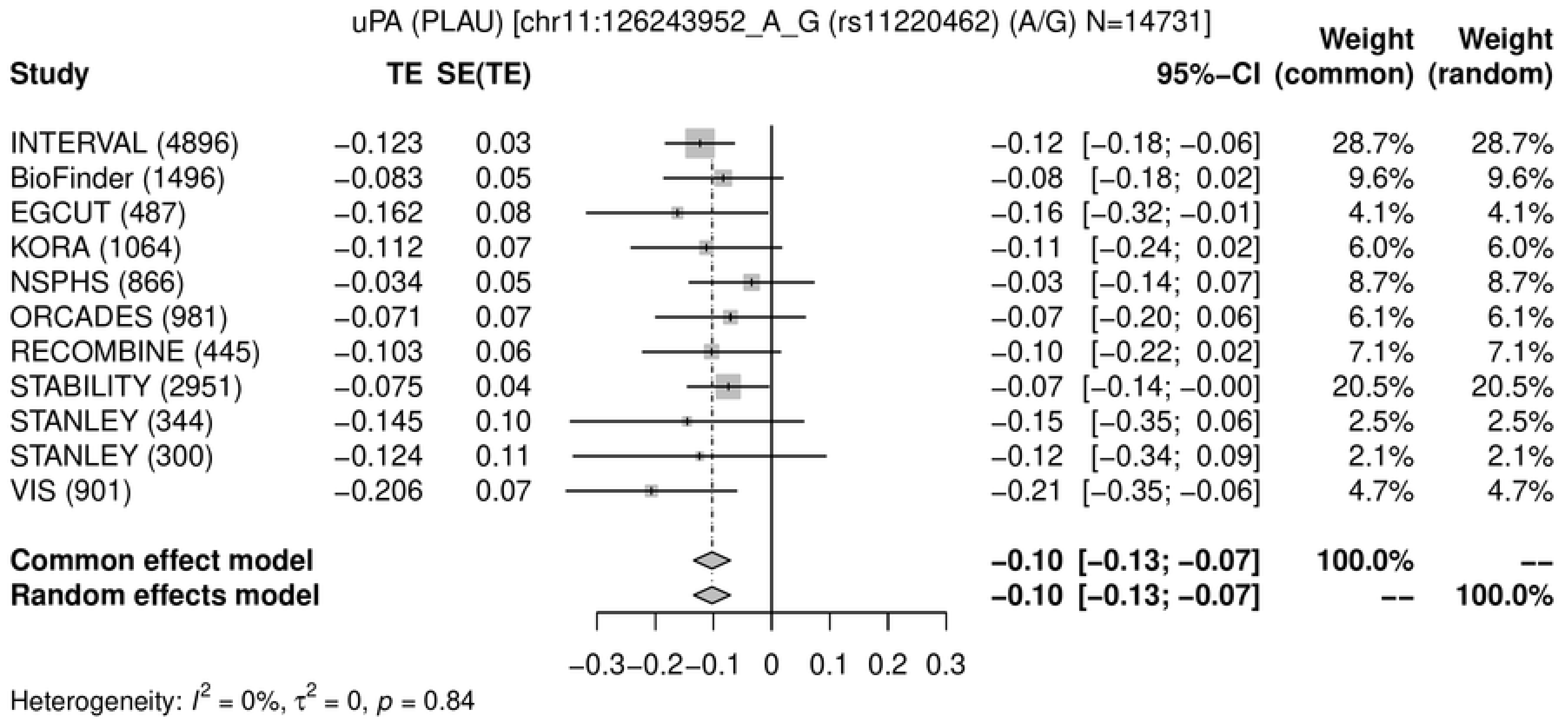
uPA (PLAU) [chr11:126243952_A_G (rs11220462) (A/G) N=14731]
Study
TE
SE(TE)
95%-CI
Weight (common)
Weight (random)
INTERVAL (4896) −0.123 0.03 −0.12 [−0.18; −0.06] 28.7% 28.7%
BioFinder (1496) −0.083 0.05 −0.08 [−0.18; 0.02] 9.6% 9.6%
EGCUT (487) −0.162 0.08 −0.16 [−0.32; −0.01] 4.1% 4.1%
KORA (1064) −0.112 0.07 −0.11 [−0.24; 0.02] 6.0% 6.0%
NSPHS (866) −0.034 0.05 −0.03 [−0.14; 0.07] 8.7% 8.7%
ORCADES (981) −0.071 0.07 −0.07 [−0.20; 0.06] 6.1% 6.1%
RECOMBINE (445) −0.103 0.06 −0.10 [−0.22; 0.02] 7.1% 7.1%
STABILITY (2951) −0.075 0.04 −0.07 [−0.14; −0.00] 20.5% 20.5%
STANLEY (344) −0.145 0.10 −0.15 [−0.35; 0.06] 2.5% 2.5%
STANLEY (300) −0.124 0.11 −0.12 [−0.34; 0.09] 2.1% 2.1%
VIS (901) −0.206 0.07 −0.21 [−0.35; −0.06] 4.7% 4.7%
Common effect model −0.10 [−0.13; −0.07] 100.0% --
Random effects model −0.10 [−0.13; −0.07] -- 100.0%
−0.3 −0.2 −0.1 0 0.1 0.2 0.3
Heterogeneity: $I^2$ = 0%, $\tau^2$ = 0, $p$ = 0.84

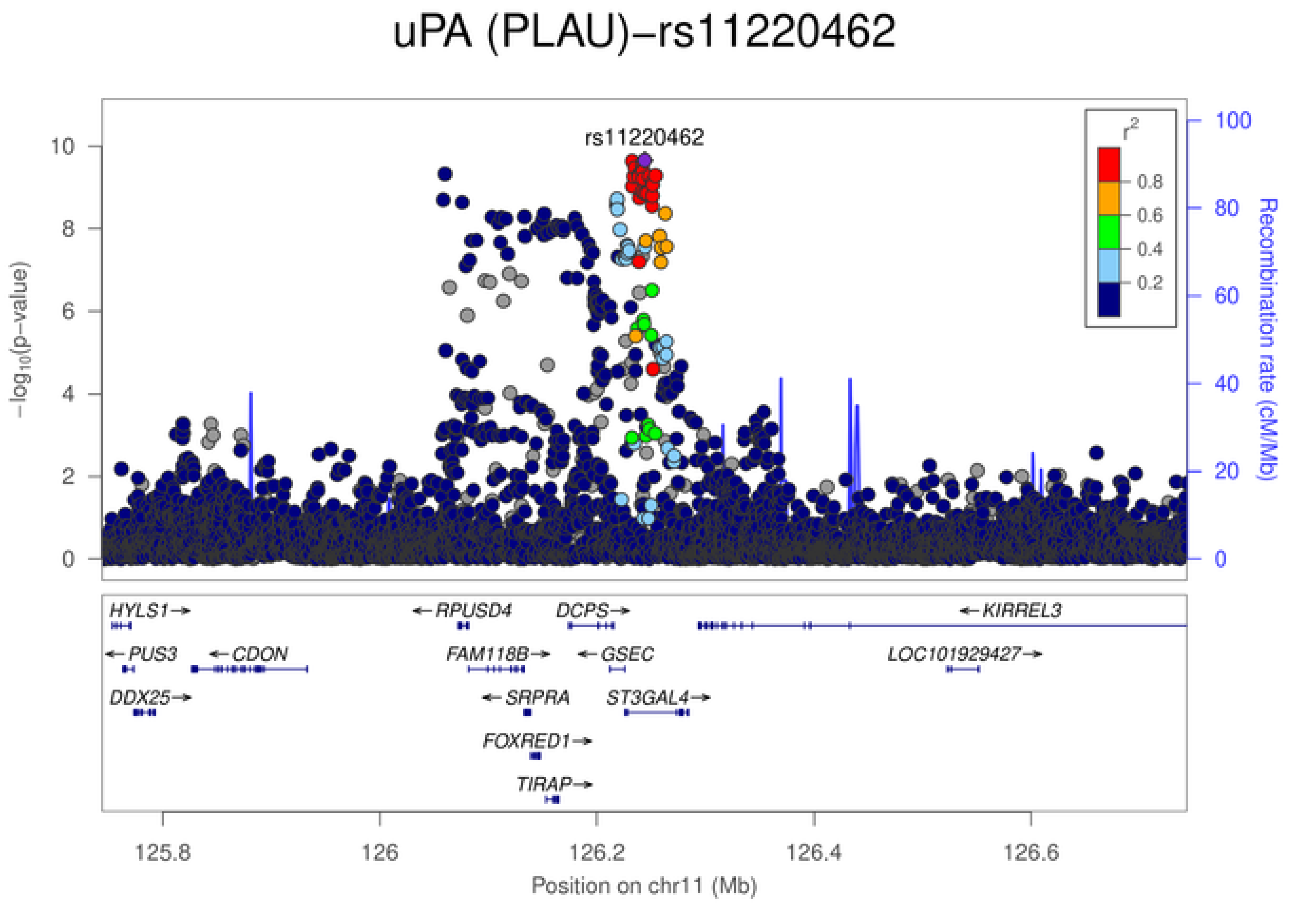
uPA (PLAU)−rs11220462
rs11220462
$-\log_{10}$(p−value)
Recombination rate (cM/Mb)
$r^2$
0.8
0.6
0.4
0.2
HYLS1
PUS3
CDON
DDX25
RPUSD4
FAM118B
SRPRA
FOXRED1
TIRAP
DCPS
GSEC
ST3GAL4
KIRREL3
LOC101929427
125.8
126
126.2
126.4
126.6
Position on chr11 (Mb)

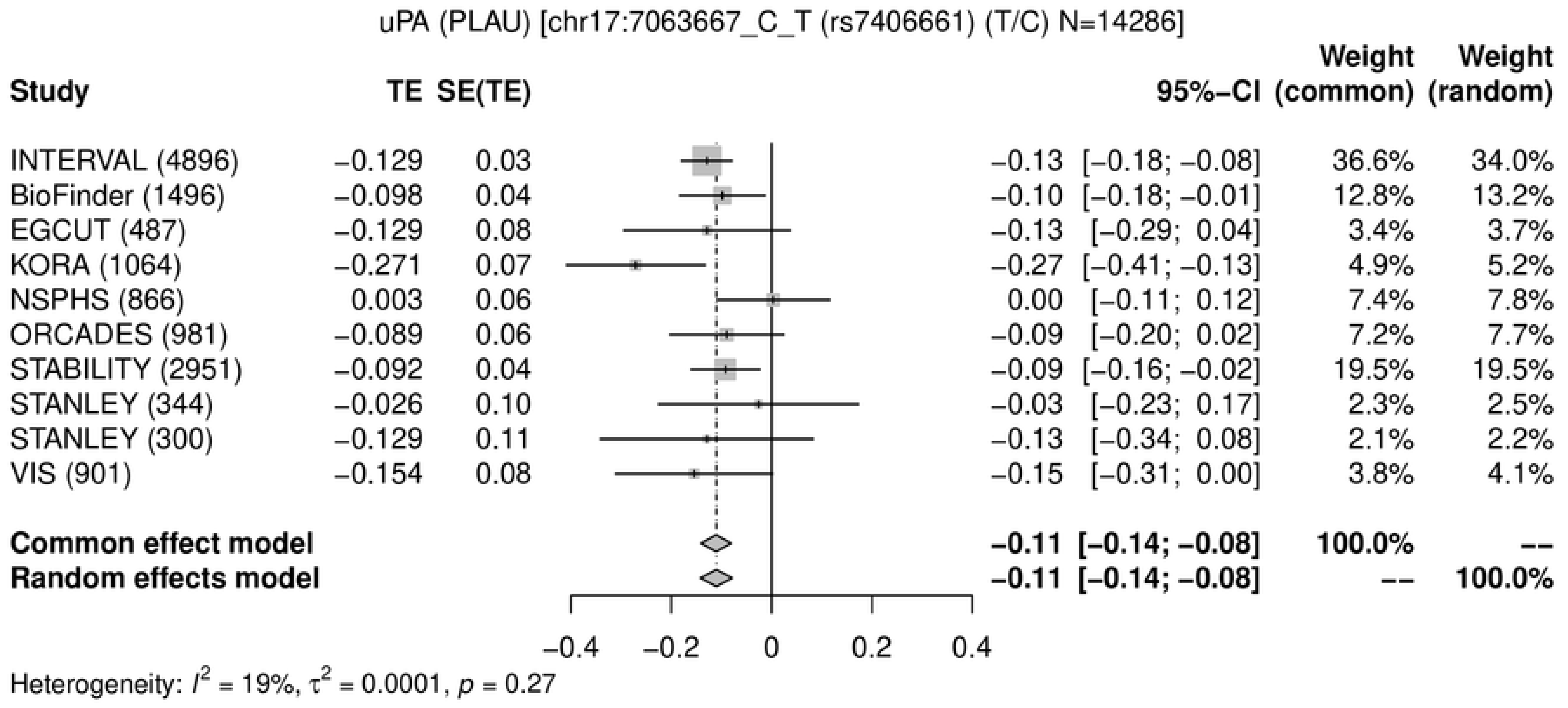
uPA (PLAU) [chr17:7063667_C_T (rs7406661) (T/C) N=14286]
Study
TE
SE(TE)
95%−CI
Weight (common)
Weight (random)
INTERVAL (4896) −0.129 0.03 −0.13 [−0.18; −0.08] 36.6% 34.0%
BioFinder (1496) −0.098 0.04 −0.10 [−0.18; −0.01] 12.8% 13.2%
EGCUT (487) −0.129 0.08 −0.13 [−0.29; 0.04] 3.4% 3.7%
KORA (1064) −0.271 0.07 −0.27 [−0.41; −0.13] 4.9% 5.2%
NSPHS (866) 0.003 0.06 0.00 [−0.11; 0.12] 7.4% 7.8%
ORCADES (981) −0.089 0.06 −0.09 [−0.20; 0.02] 7.2% 7.7%
STABILITY (2951) −0.092 0.04 −0.09 [−0.16; −0.02] 19.5% 19.5%
STANLEY (344) −0.026 0.10 −0.03 [−0.23; 0.17] 2.3% 2.5%
STANLEY (300) −0.129 0.11 −0.13 [−0.34; 0.08] 2.1% 2.2%
VIS (901) −0.154 0.08 −0.15 [−0.31; 0.00] 3.8% 4.1%
Common effect model −0.11 [−0.14; −0.08] 100.0% --
Random effects model −0.11 [−0.14; −0.08] -- 100.0%
−0.4
−0.2
0
0.2
0.4
Heterogeneity: $I^2$ = 19%, $\tau^2$ = 0.0001, $p$ = 0.27

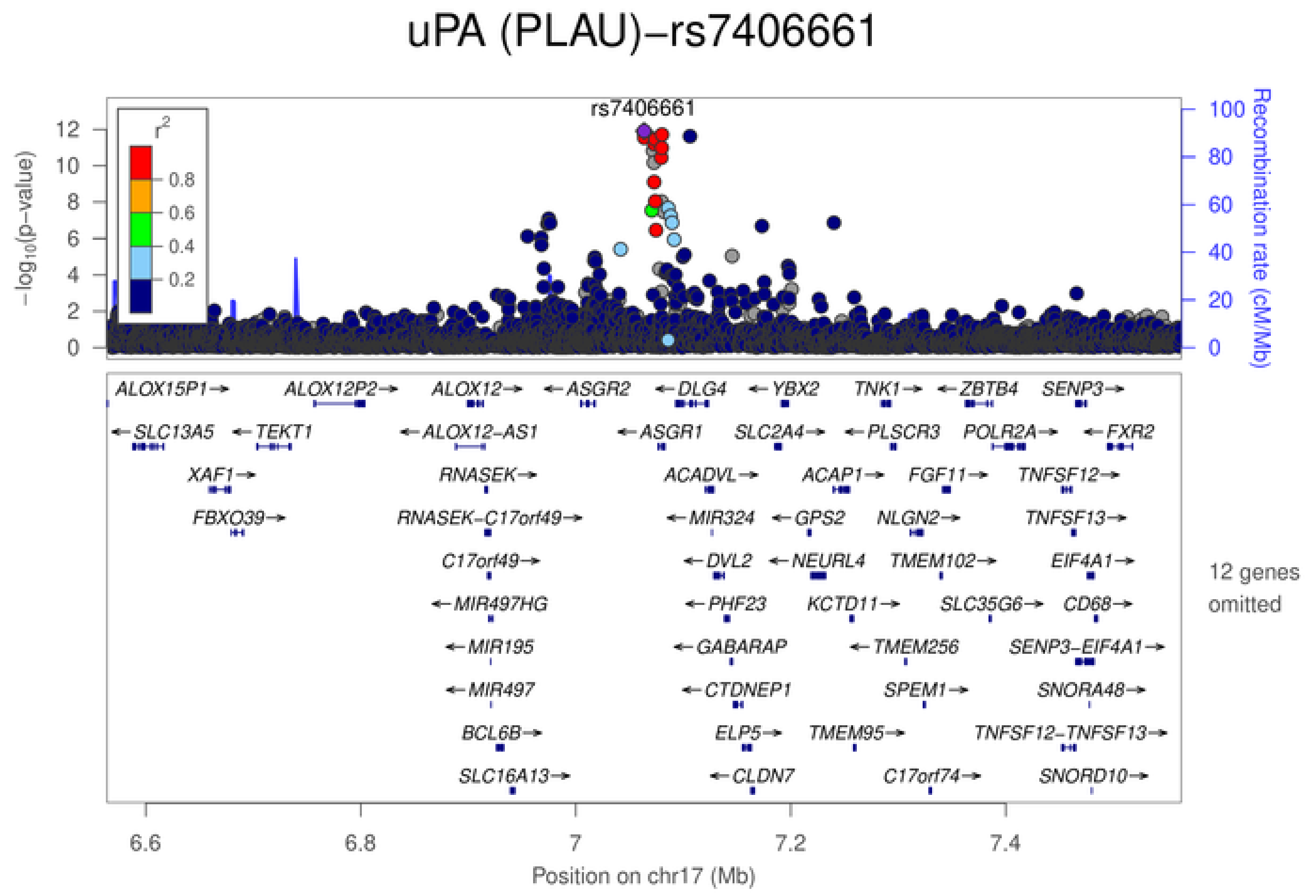
uPA (PLAU)−rs7406661
rs7406661
$-\log_{10}$(p−value)
Recombination rate (cM/Mb)
$r^2$
0.8
0.6
0.4
0.2
ALOX15P1→ ALOX12P2→ ALOX12→ ←ASGR2 ←DLG4 ←YBX2 TNK1→ ←ZBTB4 SENP3→
←SLC13A5 ←TEKT1 ←ALOX12-AS1 ←ASGR1 SLC2A4→ ←PLSCR3 POLR2A→ ←FXR2
XAF1→ RNASEK→ ACADVL→ ACAP1→ FGF11→ TNFSF12→
FBXO39→ RNASEK-C17orf49→ ←MIR324 ←GPS2 NLGN2→ TNFSF13→
C17orf49→ ←DVL2 ←NEURL4 TMEM102→ EIF4A1→
←MIR497HG ←PHF23 KCTD11→ SLC35G6→ CD68→
←MIR195 ←GABARAP ←TMEM256 SENP3-EIF4A1→
←MIR497 ←CTDNEP1 SPEM1→ SNORA48→
BCL6B→ ELP5→ TMEM95→ TNFSF12-TNFSF13→
SLC16A13→ ←CLDN7 C17orf74→ SNORD10→
12 genes omitted
6.6
6.8
7
7.2
7.4
Position on chr17 (Mb)

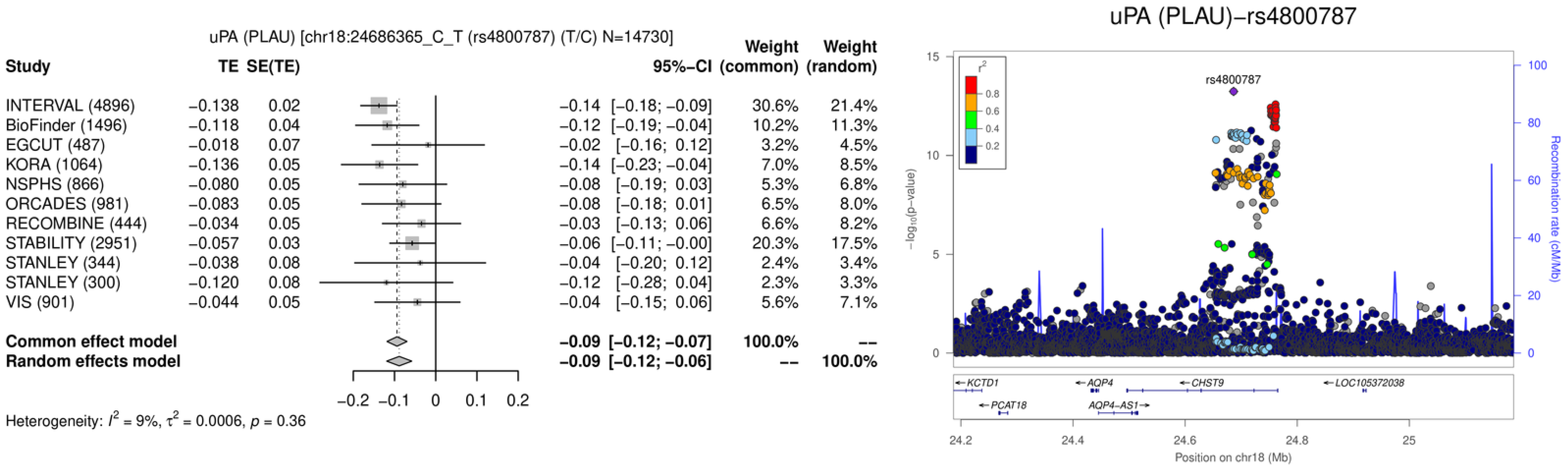
uPA (PLAU) [chr18:24686365_C_T (rs4800787) (T/C) N=14730]
Study
TE
SE(TE)
95%-CI
Weight (common)
Weight (random)
INTERVAL (4896) −0.138 0.02 −0.14 [−0.18; −0.09] 30.6% 21.4%
BioFinder (1496) −0.118 0.04 −0.12 [−0.19; −0.04] 10.2% 11.3%
EGCUT (487) −0.018 0.07 −0.02 [−0.16; 0.12] 3.2% 4.5%
KORA (1064) −0.136 0.05 −0.14 [−0.23; −0.04] 7.0% 8.5%
NSPHS (866) −0.080 0.05 −0.08 [−0.19; 0.03] 5.3% 6.8%
ORCADES (981) −0.083 0.05 −0.08 [−0.18; 0.01] 6.5% 8.0%
RECOMBINE (444) −0.034 0.05 −0.03 [−0.13; 0.06] 6.6% 8.2%
STABILITY (2951) −0.057 0.03 −0.06 [−0.11; −0.00] 20.3% 17.5%
STANLEY (344) −0.038 0.08 −0.04 [−0.20; 0.12] 2.4% 3.4%
STANLEY (300) −0.120 0.08 −0.12 [−0.28; 0.04] 2.3% 3.3%
VIS (901) −0.044 0.05 −0.04 [−0.15; 0.06] 5.6% 7.1%
Common effect model −0.09 [−0.12; −0.07] 100.0% --
Random effects model −0.09 [−0.12; −0.06] -- 100.0%
−0.2 −0.1 0 0.1 0.2
Heterogeneity: $I^2$ = 9%, $\tau^2$ = 0.0006, $p$ = 0.36
uPA (PLAU)−rs4800787
$r^2$
0.8
0.6
0.4
0.2
rs4800787
$-\log_{10}$(p−value)
Recombination rate (cM/Mb)
KCTD1
PCAT18
AQP4
AQP4−AS1
CHST9
LOC105372038
24.2 24.4 24.6 24.8 25
Position on chr18 (Mb)

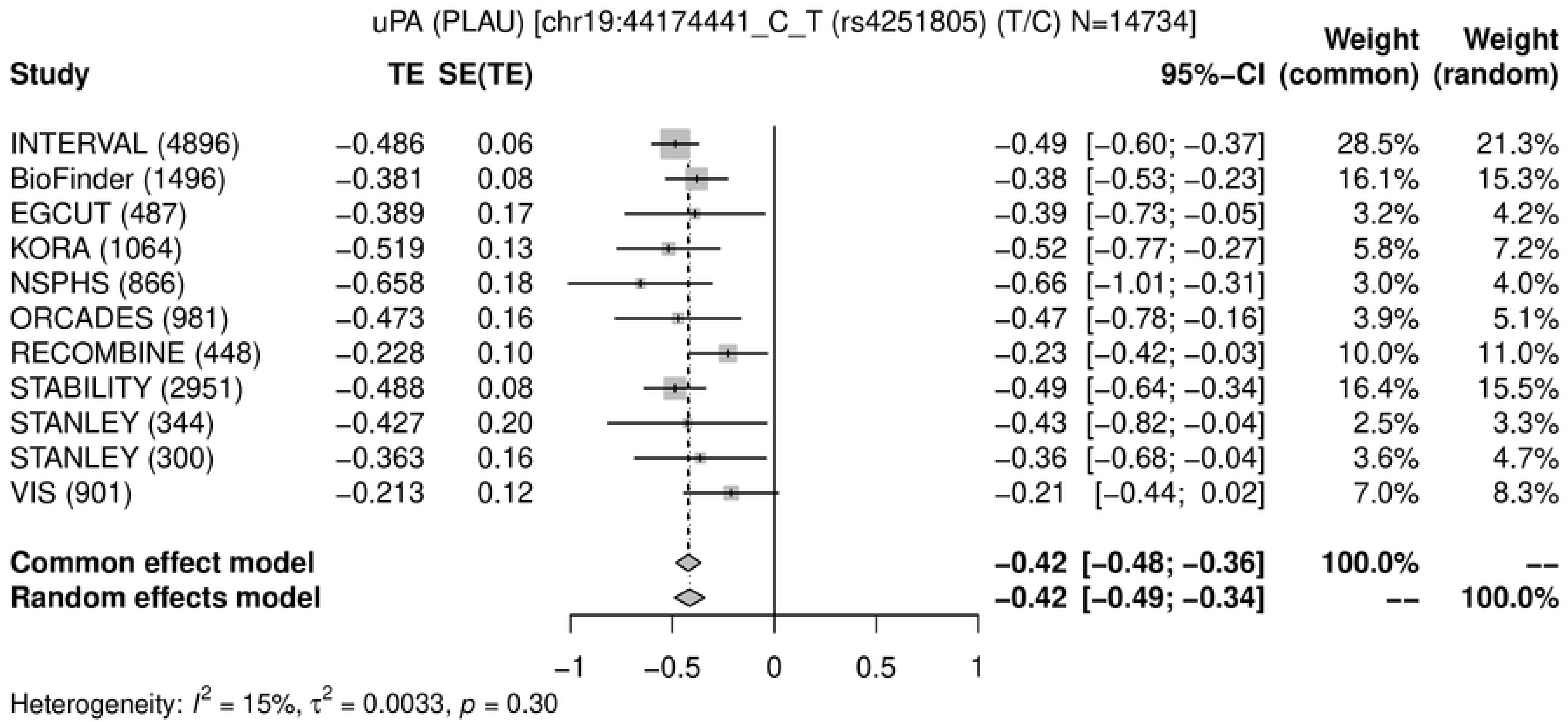
uPA (PLAU) [chr19:44174441_C_T (rs4251805) (T/C) N=14734]
Study
TE
SE(TE)
95%−CI
Weight (common)
Weight (random)
INTERVAL (4896) −0.486 0.06 −0.49 [−0.60; −0.37] 28.5% 21.3%
BioFinder (1496) −0.381 0.08 −0.38 [−0.53; −0.23] 16.1% 15.3%
EGCUT (487) −0.389 0.17 −0.39 [−0.73; −0.05] 3.2% 4.2%
KORA (1064) −0.519 0.13 −0.52 [−0.77; −0.27] 5.8% 7.2%
NSPHS (866) −0.658 0.18 −0.66 [−1.01; −0.31] 3.0% 4.0%
ORCADES (981) −0.473 0.16 −0.47 [−0.78; −0.16] 3.9% 5.1%
RECOMBINE (448) −0.228 0.10 −0.23 [−0.42; −0.03] 10.0% 11.0%
STABILITY (2951) −0.488 0.08 −0.49 [−0.64; −0.34] 16.4% 15.5%
STANLEY (344) −0.427 0.20 −0.43 [−0.82; −0.04] 2.5% 3.3%
STANLEY (300) −0.363 0.16 −0.36 [−0.68; −0.04] 3.6% 4.7%
VIS (901) −0.213 0.12 −0.21 [−0.44; 0.02] 7.0% 8.3%
Common effect model −0.42 [−0.48; −0.36] 100.0% --
Random effects model −0.42 [−0.49; −0.34] -- 100.0%
−1 −0.5 0 0.5 1
Heterogeneity: $I^2$ = 15%, $\tau^2$ = 0.0033, $p$ = 0.30

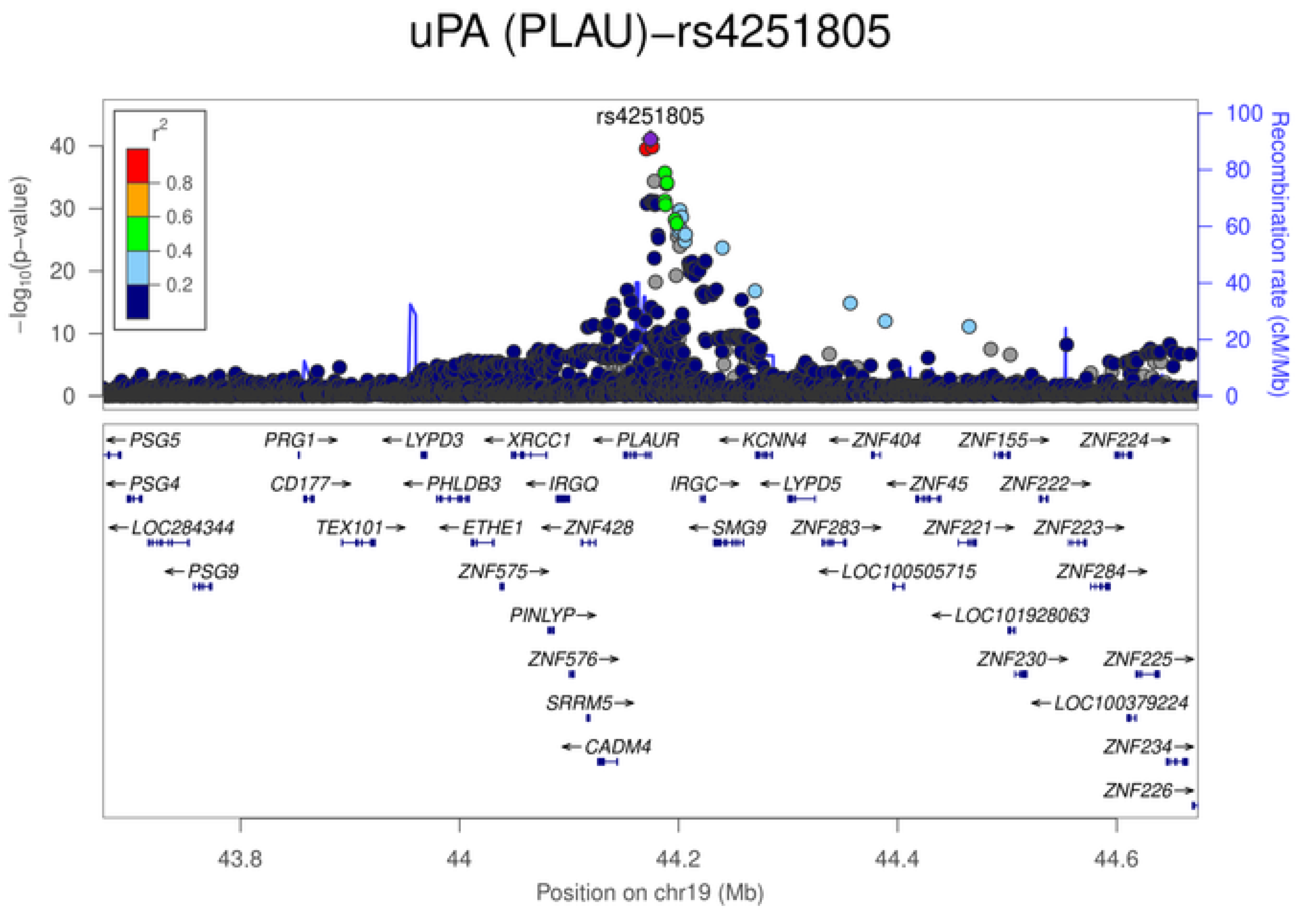
uPA (PLAU)−rs4251805
rs4251805
$r^2$
0.8
0.6
0.4
0.2
$-\log_{10}$(p-value)
Recombination rate (cM/Mb)
0 10 20 30 40
0 20 40 60 80 100
PSG5
PRG1
LYPD3
XRCC1
PLAUR
KCNN4
ZNF404
ZNF155
ZNF224
PSG4
CD177
PHLDB3
IRGQ
IRGC
LYPD5
ZNF45
ZNF222
LOC284344
TEX101
ETHE1
ZNF428
SMG9
ZNF283
ZNF221
ZNF223
PSG9
ZNF575
LOC100505715
ZNF284
PINLYP
LOC101928063
ZNF576
ZNF230
ZNF225
SRRM5
LOC100379224
CADM4
ZNF234
ZNF226
43.8 44 44.2 44.4 44.6
Position on chr19 (Mb)

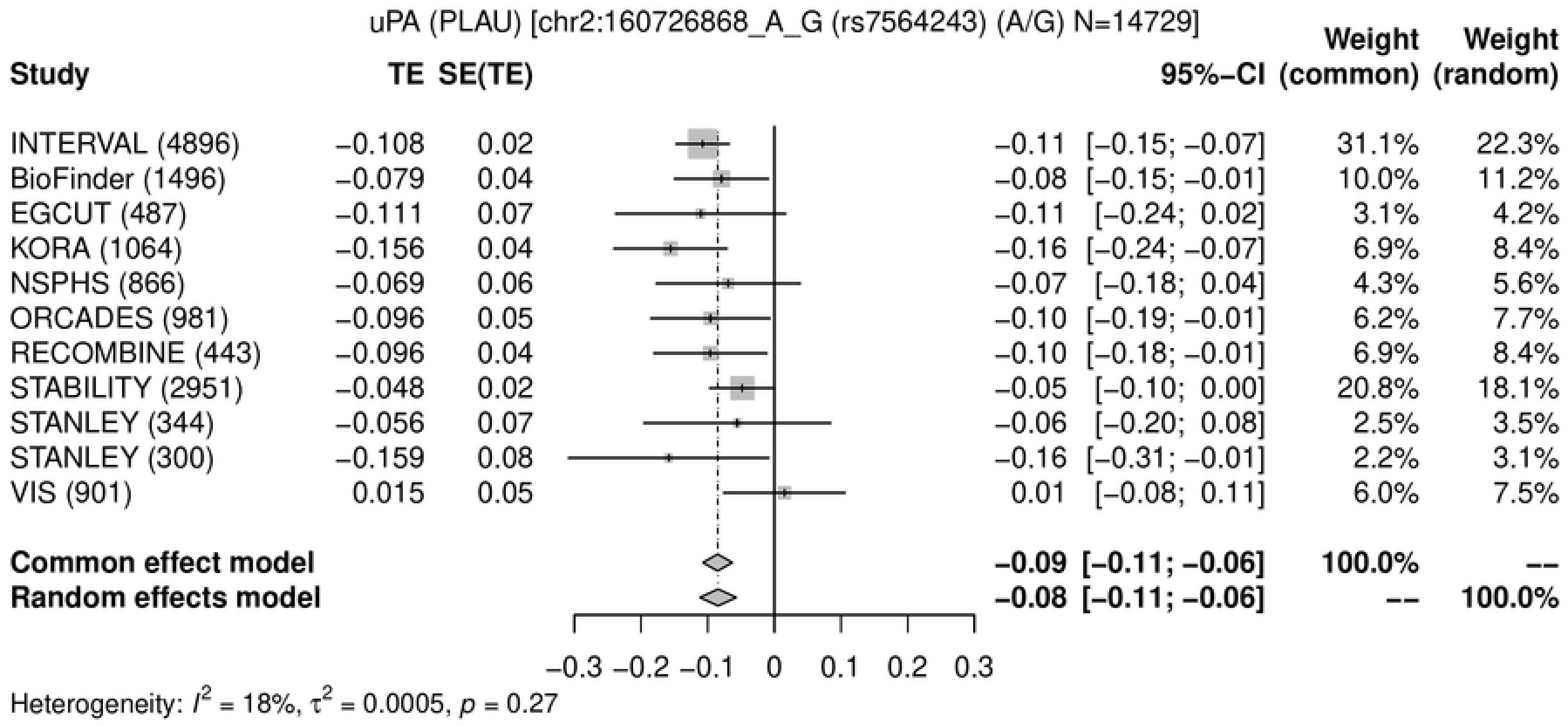
uPA (PLAU) [chr2:160726868_A_G (rs7564243) (A/G) N=14729]
Study
TE
SE(TE)
95%−CI
Weight (common)
Weight (random)
INTERVAL (4896) −0.108 0.02 −0.11 [−0.15; −0.07] 31.1% 22.3%
BioFinder (1496) −0.079 0.04 −0.08 [−0.15; −0.01] 10.0% 11.2%
EGCUT (487) −0.111 0.07 −0.11 [−0.24; 0.02] 3.1% 4.2%
KORA (1064) −0.156 0.04 −0.16 [−0.24; −0.07] 6.9% 8.4%
NSPHS (866) −0.069 0.06 −0.07 [−0.18; 0.04] 4.3% 5.6%
ORCADES (981) −0.096 0.05 −0.10 [−0.19; −0.01] 6.2% 7.7%
RECOMBINE (443) −0.096 0.04 −0.10 [−0.18; −0.01] 6.9% 8.4%
STABILITY (2951) −0.048 0.02 −0.05 [−0.10; 0.00] 20.8% 18.1%
STANLEY (344) −0.056 0.07 −0.06 [−0.20; 0.08] 2.5% 3.5%
STANLEY (300) −0.159 0.08 −0.16 [−0.31; −0.01] 2.2% 3.1%
VIS (901) 0.015 0.05 0.01 [−0.08; 0.11] 6.0% 7.5%
Common effect model −0.09 [−0.11; −0.06] 100.0% --
Random effects model −0.08 [−0.11; −0.06] -- 100.0%
−0.3 −0.2 −0.1 0 0.1 0.2 0.3
Heterogeneity: $I^2$ = 18%, $\tau^2$ = 0.0005, $p$ = 0.27

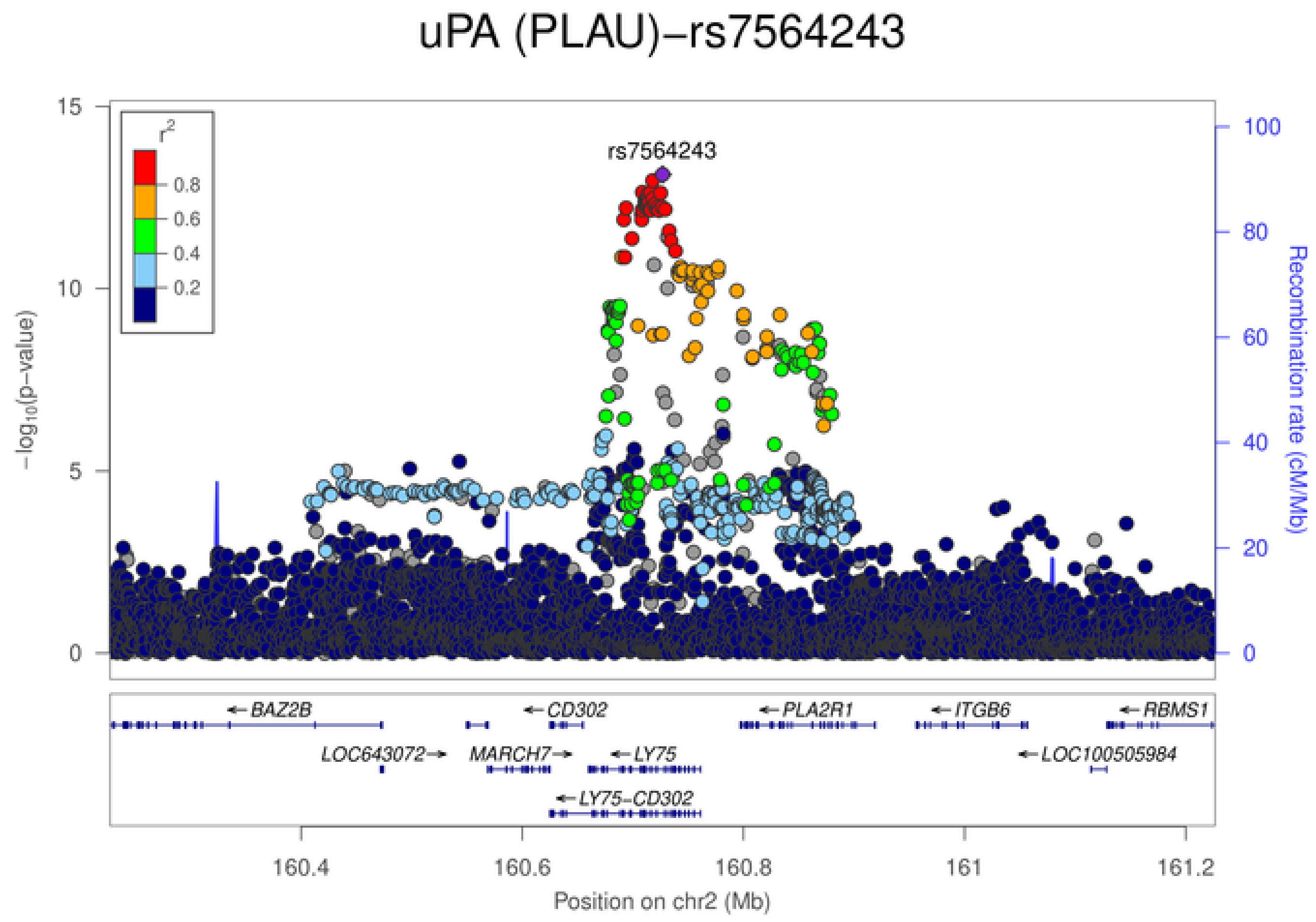
uPA (PLAU)−rs7564243
rs7564243
$r^2$
0.8
0.6
0.4
0.2
15
10
5
0
$-\log_{10}$(p-value)
100
80
60
40
20
0
Recombination rate (cM/Mb)
←BAZ2B
←CD302
←PLA2R1
←ITGB6
←RBMS1
LOC643072→
MARCH7→
←LY75
←LOC100505984
←LY75−CD302
160.4
160.6
160.8
161
161.2
Position on chr2 (Mb)

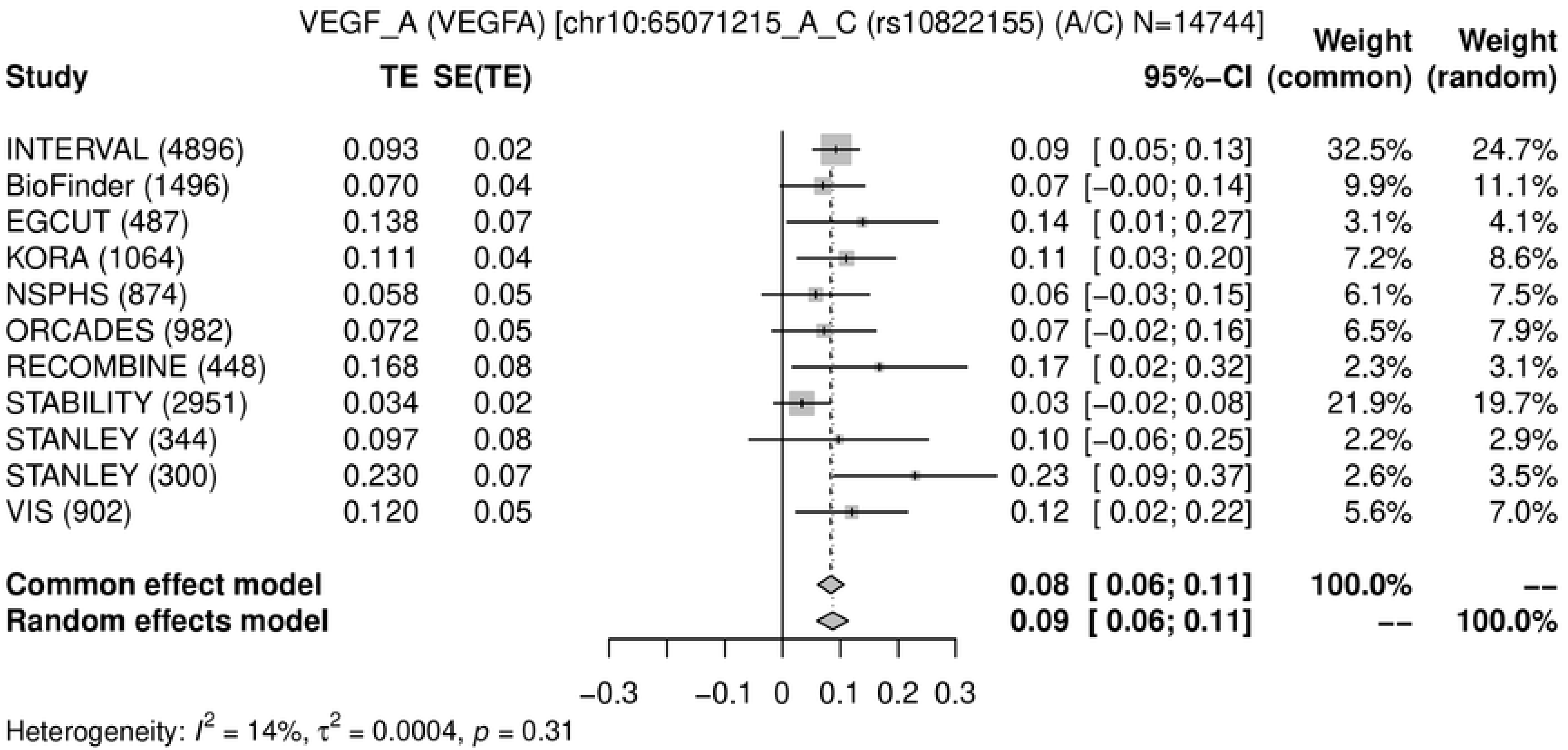
VEGF_A (VEGFA) [chr10:65071215_A_C (rs10822155) (A/C) N=14744]
Study
TE
SE(TE)
95%−CI
Weight (common)
Weight (random)
INTERVAL (4896) 0.093 0.02 0.09 [ 0.05; 0.13] 32.5% 24.7%
BioFinder (1496) 0.070 0.04 0.07 [−0.00; 0.14] 9.9% 11.1%
EGCUT (487) 0.138 0.07 0.14 [ 0.01; 0.27] 3.1% 4.1%
KORA (1064) 0.111 0.04 0.11 [ 0.03; 0.20] 7.2% 8.6%
NSPHS (874) 0.058 0.05 0.06 [−0.03; 0.15] 6.1% 7.5%
ORCADES (982) 0.072 0.05 0.07 [−0.02; 0.16] 6.5% 7.9%
RECOMBINE (448) 0.168 0.08 0.17 [ 0.02; 0.32] 2.3% 3.1%
STABILITY (2951) 0.034 0.02 0.03 [−0.02; 0.08] 21.9% 19.7%
STANLEY (344) 0.097 0.08 0.10 [−0.06; 0.25] 2.2% 2.9%
STANLEY (300) 0.230 0.07 0.23 [ 0.09; 0.37] 2.6% 3.5%
VIS (902) 0.120 0.05 0.12 [ 0.02; 0.22] 5.6% 7.0%
Common effect model 0.08 [ 0.06; 0.11] 100.0% --
Random effects model 0.09 [ 0.06; 0.11] -- 100.0%
−0.3 −0.1 0 0.1 0.2 0.3
Heterogeneity: $I^2$ = 14%, $\tau^2$ = 0.0004, $p$ = 0.31

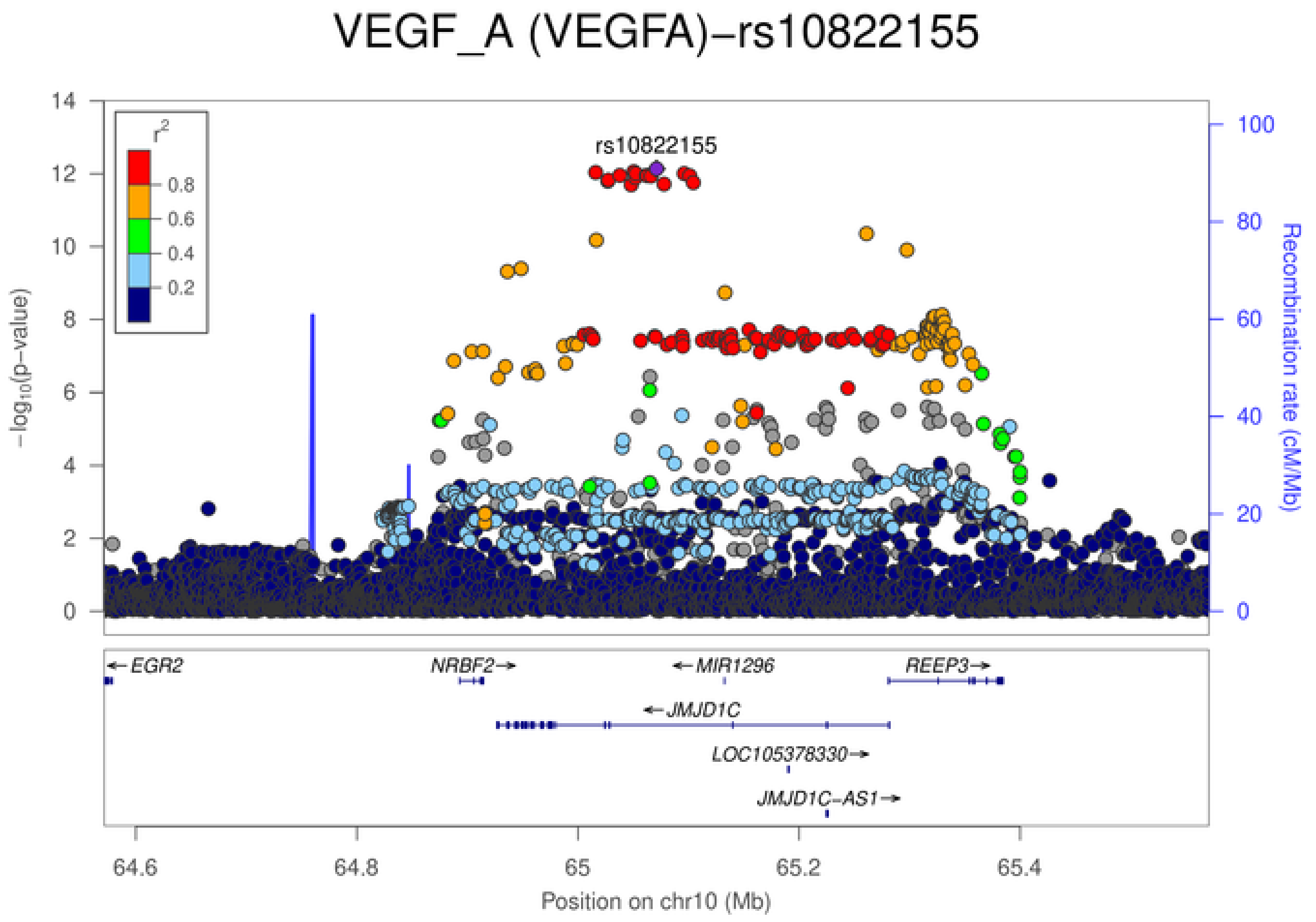
VEGF_A (VEGFA)−rs10822155
rs10822155
$r^2$
0.8
0.6
0.4
0.2
$-\log_{10}$(p−value)
Recombination rate (cM/Mb)
EGR2
NRBF2
MIR1296
REEP3
JMJD1C
LOC105378330
JMJD1C−AS1
64.6
64.8
65
65.2
65.4
Position on chr10 (Mb)

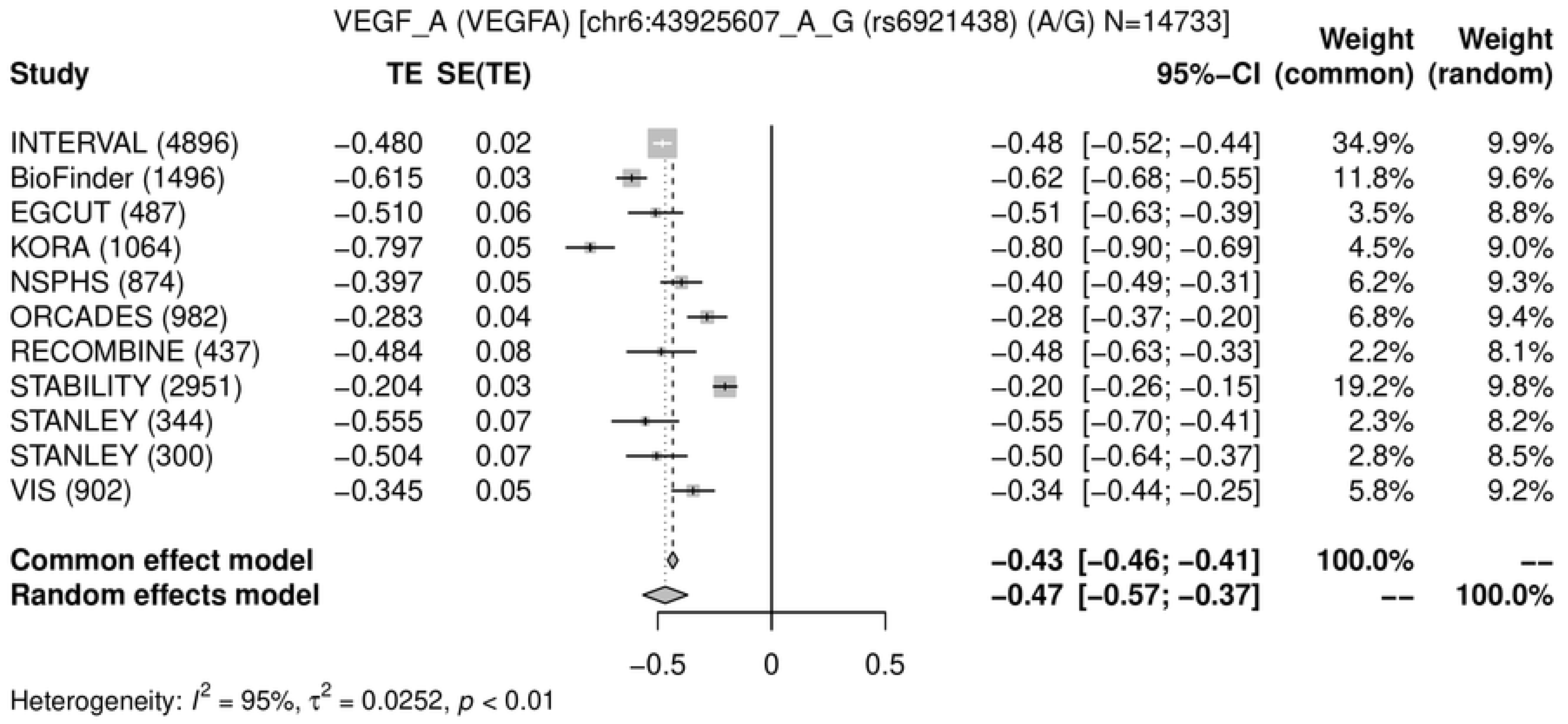
VEGF_A (VEGFA) [chr6:43925607_A_G (rs6921438) (A/G) N=14733]
Study
TE
SE(TE)
95%−CI
Weight (common)
Weight (random)
INTERVAL (4896) −0.480 0.02 −0.48 [−0.52; −0.44] 34.9% 9.9%
BioFinder (1496) −0.615 0.03 −0.62 [−0.68; −0.55] 11.8% 9.6%
EGCUT (487) −0.510 0.06 −0.51 [−0.63; −0.39] 3.5% 8.8%
KORA (1064) −0.797 0.05 −0.80 [−0.90; −0.69] 4.5% 9.0%
NSPHS (874) −0.397 0.05 −0.40 [−0.49; −0.31] 6.2% 9.3%
ORCADES (982) −0.283 0.04 −0.28 [−0.37; −0.20] 6.8% 9.4%
RECOMBINE (437) −0.484 0.08 −0.48 [−0.63; −0.33] 2.2% 8.1%
STABILITY (2951) −0.204 0.03 −0.20 [−0.26; −0.15] 19.2% 9.8%
STANLEY (344) −0.555 0.07 −0.55 [−0.70; −0.41] 2.3% 8.2%
STANLEY (300) −0.504 0.07 −0.50 [−0.64; −0.37] 2.8% 8.5%
VIS (902) −0.345 0.05 −0.34 [−0.44; −0.25] 5.8% 9.2%
Common effect model −0.43 [−0.46; −0.41] 100.0% --
Random effects model −0.47 [−0.57; −0.37] -- 100.0%
−0.5
0
0.5
Heterogeneity: $I^2$ = 95%, $\tau^2$ = 0.0252, $p$ < 0.01

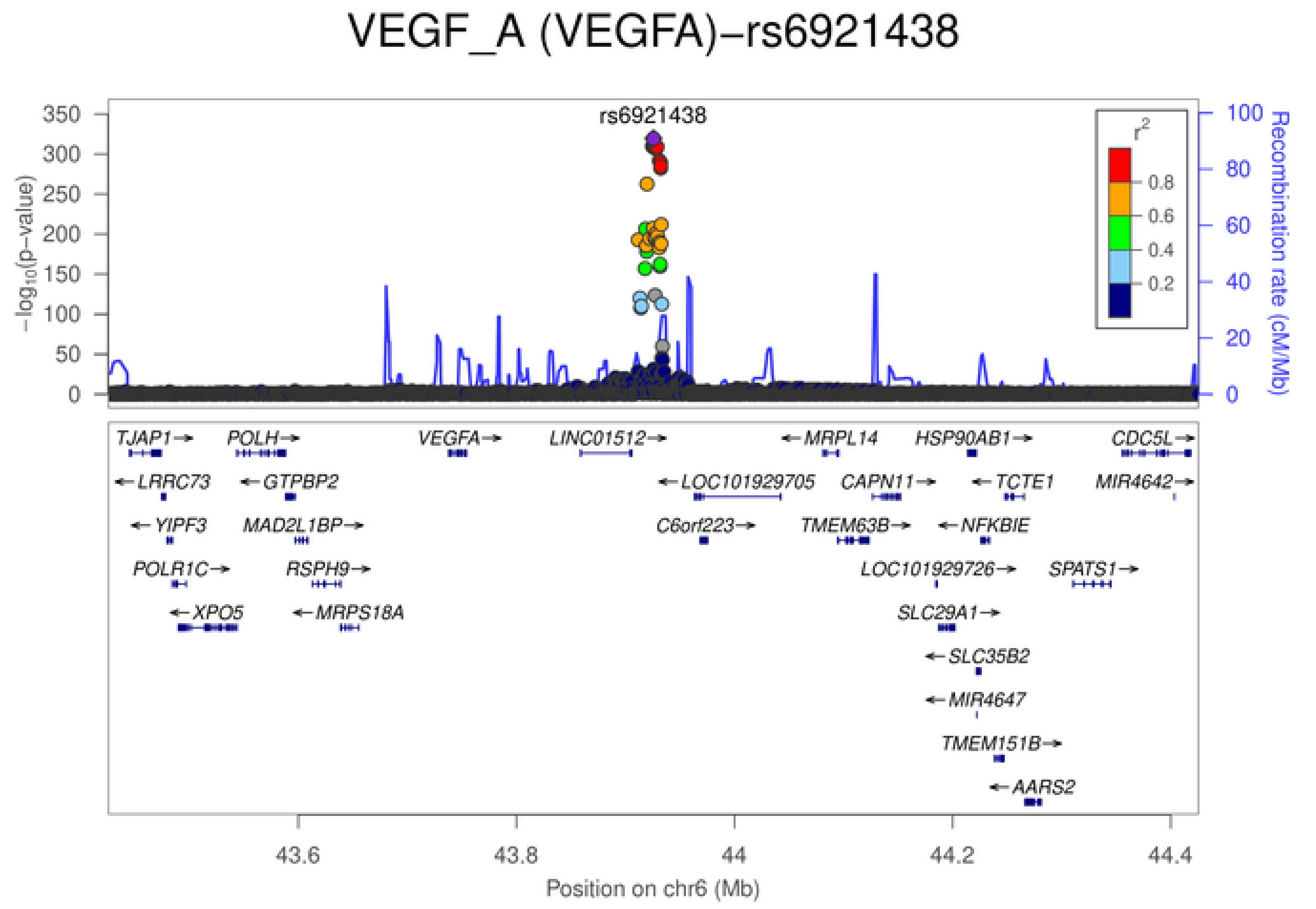
VEGF_A (VEGFA)−rs6921438
rs6921438
$-\log_{10}$(p−value)
Recombination rate (cM/Mb)
$r^2$
0.8
0.6
0.4
0.2
TJAP1
POLH
VEGFA
LINC01512
MRPL14
HSP90AB1
CDC5L
LRRC73
GTPBP2
LOC101929705
CAPN11
TCTE1
MIR4642
YIPF3
MAD2L1BP
C6orf223
TMEM63B
NFKBIE
POLR1C
RSPH9
LOC101929726
SPATS1
XPO5
MRPS18A
SLC29A1
SLC35B2
MIR4647
TMEM151B
AARS2
43.6
43.8
44
44.2
44.4
Position on chr6 (Mb)

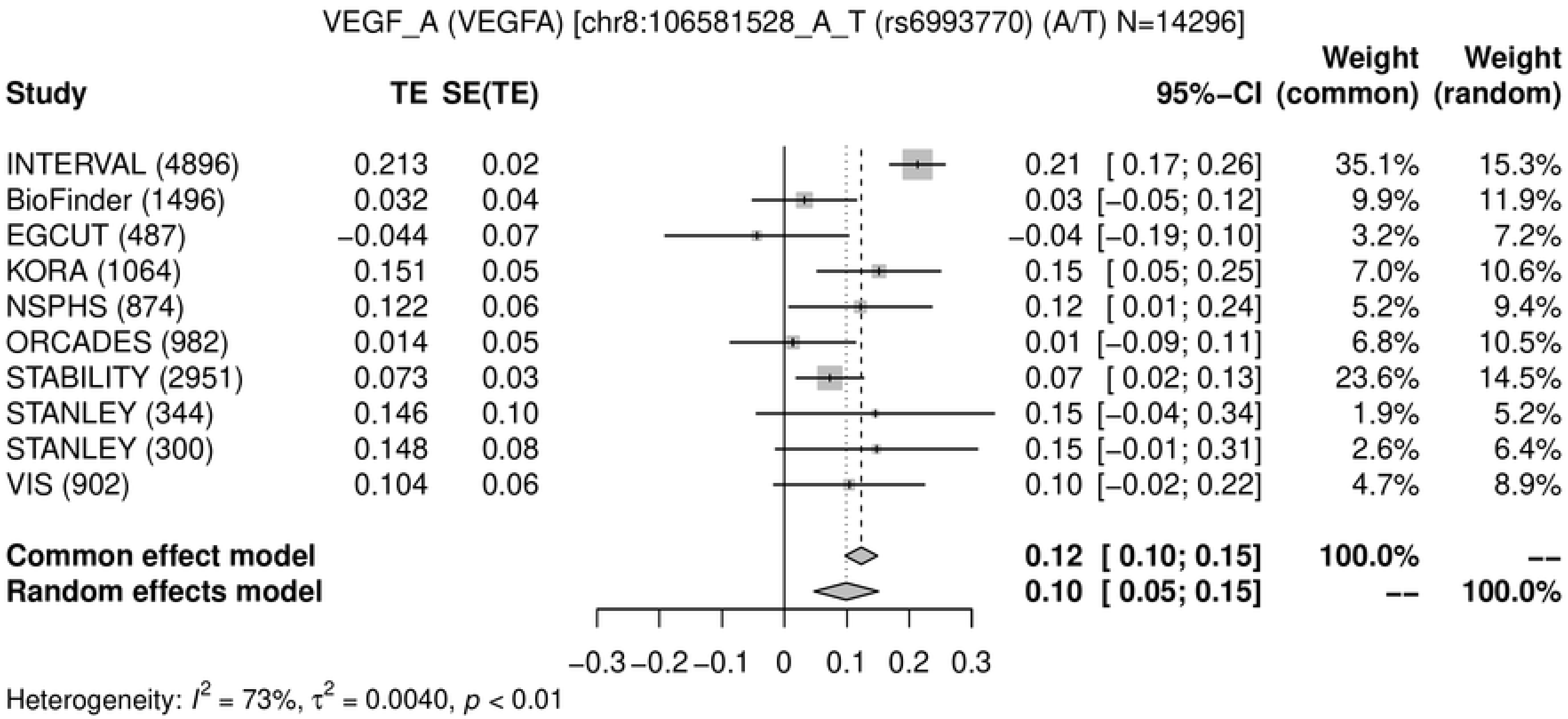
VEGF_A (VEGFA) [chr8:106581528_A_T (rs6993770) (A/T) N=14296]
Study | TE | SE(TE) | 95%-CI | Weight (common) | Weight (random)
INTERVAL (4896) | 0.213 | 0.02 | 0.21 [ 0.17; 0.26] | 35.1% | 15.3%
BioFinder (1496) | 0.032 | 0.04 | 0.03 [−0.05; 0.12] | 9.9% | 11.9%
EGCUT (487) | −0.044 | 0.07 | −0.04 [−0.19; 0.10] | 3.2% | 7.2%
KORA (1064) | 0.151 | 0.05 | 0.15 [ 0.05; 0.25] | 7.0% | 10.6%
NSPHS (874) | 0.122 | 0.06 | 0.12 [ 0.01; 0.24] | 5.2% | 9.4%
ORCADES (982) | 0.014 | 0.05 | 0.01 [−0.09; 0.11] | 6.8% | 10.5%
STABILITY (2951) | 0.073 | 0.03 | 0.07 [ 0.02; 0.13] | 23.6% | 14.5%
STANLEY (344) | 0.146 | 0.10 | 0.15 [−0.04; 0.34] | 1.9% | 5.2%
STANLEY (300) | 0.148 | 0.08 | 0.15 [−0.01; 0.31] | 2.6% | 6.4%
VIS (902) | 0.104 | 0.06 | 0.10 [−0.02; 0.22] | 4.7% | 8.9%
Common effect model | 0.12 [ 0.10; 0.15] | 100.0% | --
Random effects model | 0.10 [ 0.05; 0.15] | -- | 100.0%
−0.3 −0.2 −0.1 0 0.1 0.2 0.3
Heterogeneity: $I^2$ = 73%, $\tau^2$ = 0.0040, $p$ < 0.01

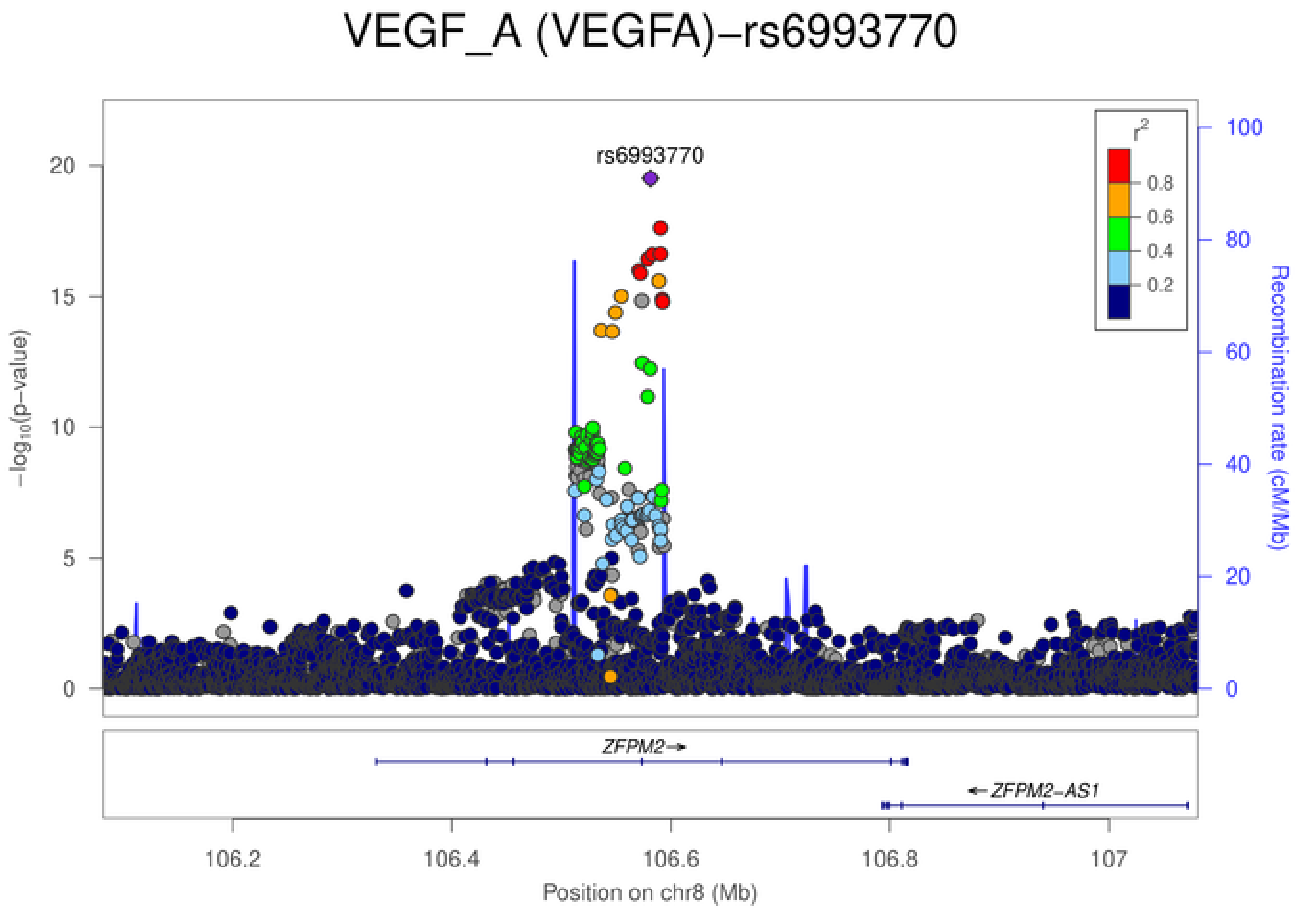
VEGF_A (VEGFA)−rs6993770
rs6993770
$-\log_{10}$(p−value)
Recombination rate (cM/Mb)
$r^2$
0.8
0.6
0.4
0.2
ZFPM2→
←ZFPM2−AS1
106.2
106.4
106.6
106.8
107
Position on chr8 (Mb)

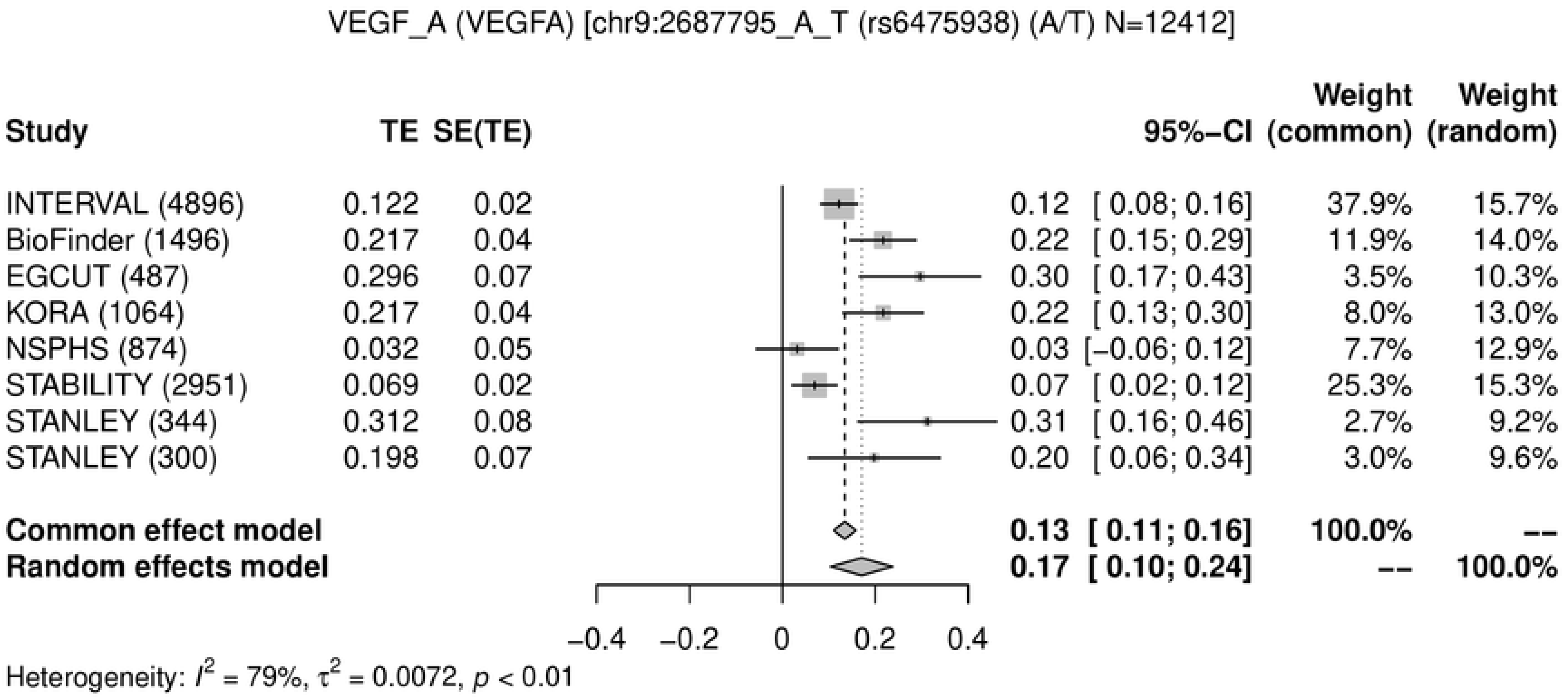
VEGF_A (VEGFA) [chr9:2687795_A_T (rs6475938) (A/T) N=12412]
Study
TE
SE(TE)
95%−CI
Weight (common)
Weight (random)
INTERVAL (4896) 0.122 0.02 0.12 [ 0.08; 0.16] 37.9% 15.7%
BioFinder (1496) 0.217 0.04 0.22 [ 0.15; 0.29] 11.9% 14.0%
EGCUT (487) 0.296 0.07 0.30 [ 0.17; 0.43] 3.5% 10.3%
KORA (1064) 0.217 0.04 0.22 [ 0.13; 0.30] 8.0% 13.0%
NSPHS (874) 0.032 0.05 0.03 [−0.06; 0.12] 7.7% 12.9%
STABILITY (2951) 0.069 0.02 0.07 [ 0.02; 0.12] 25.3% 15.3%
STANLEY (344) 0.312 0.08 0.31 [ 0.16; 0.46] 2.7% 9.2%
STANLEY (300) 0.198 0.07 0.20 [ 0.06; 0.34] 3.0% 9.6%
Common effect model 0.13 [ 0.11; 0.16] 100.0% --
Random effects model 0.17 [ 0.10; 0.24] -- 100.0%
−0.4 −0.2 0 0.2 0.4
Heterogeneity: I² = 79%, τ² = 0.0072, p < 0.01

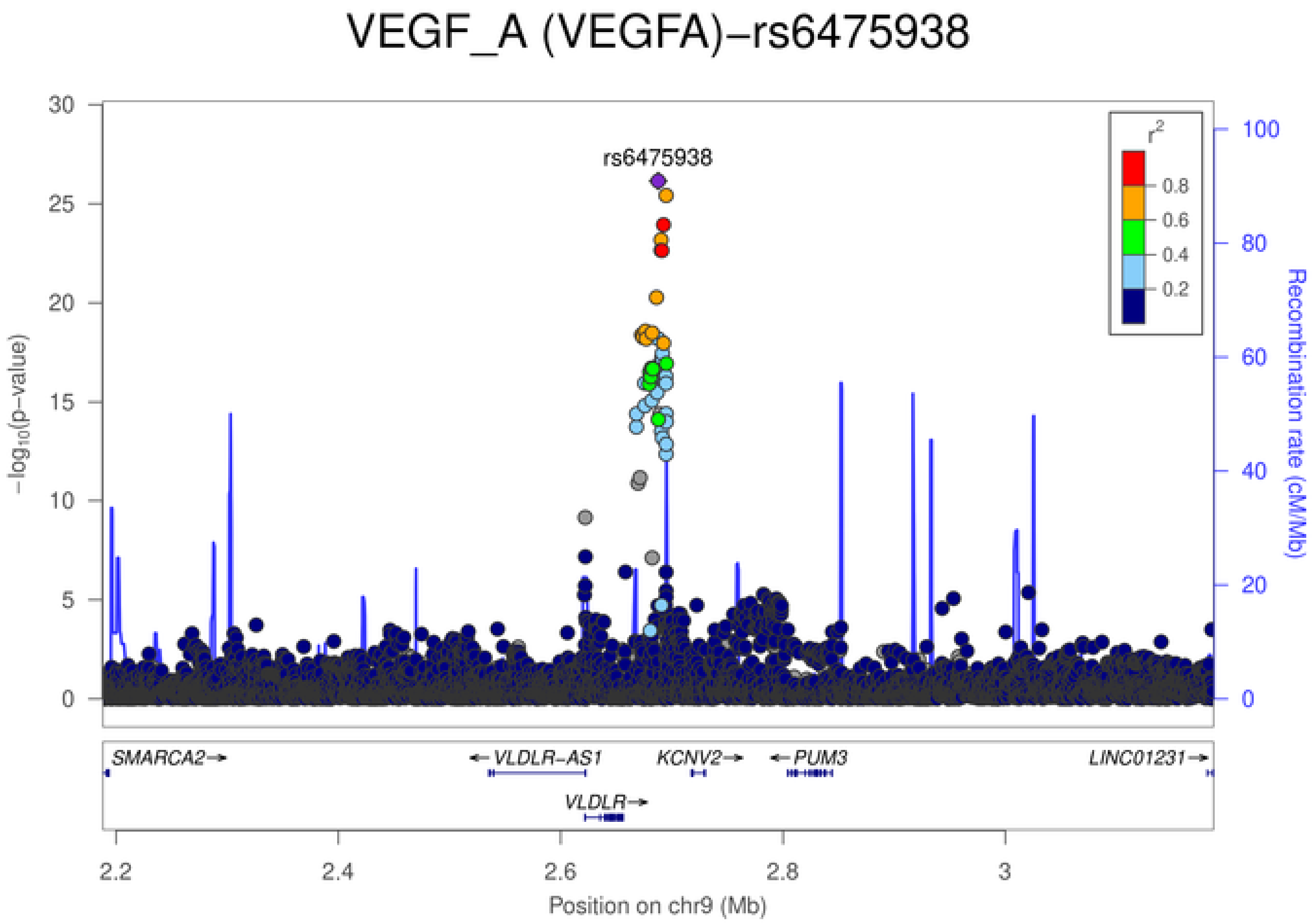
VEGF_A (VEGFA)−rs6475938
rs6475938
r²
0.8
0.6
0.4
0.2
−log10(p−value)
Recombination rate (cM/Mb)
SMARCA2→
←VLDLR-AS1
KCNV2→
←PUM3
LINC01231→
VLDLR→
2.2 2.4 2.6 2.8 3
Position on chr9 (Mb)
